# Declines, and Pronounced Regional Disparities, in Meperidine Use in the United States

**DOI:** 10.1101/2020.11.30.20241182

**Authors:** John M. Boyle, Kenneth L. McCall, Stephanie D. Nichols, Brian J. Piper

## Abstract

**Purpose:** There have been increasing concerns about adverse effects and drug interactions with meperidine including removal from the World Health Organization’s list of essential medications. The goal of this study was to characterize pharmacoepidemiological patterns in meperidine use in the United States.

**Methods:** Meperidine distribution data was obtained from the Drug Enforcement Administration’s Automation of Reports and Consolidated Orders System (ARCOS). Medicare Part D Prescriber Public Use Files (PUF) were utilized to capture overall trends in national meperidine prescriptions.

**Results:** National meperidine distribution decreased from 2001 to 2019 by 94.6%. In 2019 Arkansas, Alabama, Oklahoma, and Mississippi saw significantly greater distribution per person when compared to the average state (9.27, SD = 6.82). Meperidine per ten persons showed an eighteen-fold difference between the highest (Arkansas = 36.8 mg) and lowest (Minnesota = 2.1 mg) states. Five of the six lowest states were in the northeast. Meperidine distribution per state was significantly correlated with the prevalence of adult obesity (*r*(47) = +0.47, *p* < 0.001).

Family medicine and internal medicine physicians accounted for 28.9% and 20.5% of Medicare Part D total daily supply (TDS) of meperidine in 2017. However, interventional pain management (5.66) and pain management (3.48) physicians accounted for the longest while family medicine (0.69) and internal medicine (0.40) accounted for the shortest TDS per provider.

**Conclusion:** Use of meperidine has been declining over the last two-decades. Meperidine distribution varied on a geographical level with south/south-central, and more obese, states showing appreciably greater distribution per person. Primary care doctors continue to account for the majority of meperidine daily supply, but specialists like interventional pain management were the most likely to prescribe meperidine to Medicare patients. Increasing knowledge of meperidine’s undesirable adverse effects (e.g. seizures) and serious drug-drug interactions likely are responsible for these pronounced reductions.

## Introduction

Meperidine was first synthesized in 1938 by Otto Eisleb as a novel anticholinergic [1]. Chemically, meperidine is a phenylpiperidine with an ethyl ester moiety attached at the four carbon. Meperidine’s analgesic properties were later discovered and in 1942 it was approved by the US FDA [2]. Meperidine has an oral bioavailability of 30 – 60% with about one-third the analgesic potency of morphine. It was the analgesic of choice in the United States in the later-half of the 20^th^ century [3]. Meperidine was thought to carry lower adverse effect and misuse profiles compared to traditional opioids. However, it was later discovered that claims of decreased risk of addiction were untrue [4] as meperidine has appreciable misuse potential. The US Drug Enforcement Agency (DEA) categorizes meperidine as a Schedule II substance with a high potential for abuse, with use potentially leading to severe psychological or physical dependence [5]. Moreover, meperidine’s misuse potential is higher than that of many other opioids due to its rapid onset of action [6]. Attempts at home synthesis of a meperidine analogue resulted in MPTP which became an import research tool to study Parkinson’s in experimental animals [7-8].

Like all phenylpiperidine opioids, meperidine is a weak serotonin reuptake inhibitor. However, the most serious adverse effects associated with meperidine are due to its metabolite normeperidine, which possess agonist activity at the serotonin 5-HT_2A_ receptor. Thus, build-up of normeperidine or concurrent administration of meperidine with drugs possessing serotonergic activity (SSRIs, MAOIs, etc) may precipitate serotonin syndrome [9-13]. Serotonin syndrome is marked by a triad of symptoms (altered mental status, neuromuscular abnormalities and autonomic hyperactivity). In mild cases, the symptoms are generally limited to autonomic disturbances such as tachycardia, hypertension, mydriasis, etc. If activity at serotonin receptors continues to increase, serotonin syndrome symptomatology may proceed to seizures, coma, rhabdomyolysis, metabolic acidosis, or even death [13].

Meperidine is metabolized by N-demethylation via cytochrome P450 2B6, 3A4, and 2C19. While meperidine has a short half-life of 2.5 – 4 hours, its neurotoxic metabolite normeperidine has a half-life is 4 – 21 hours [14]. Due to its relatively short duration of action, multiple doses of meperidine are needed for chronic pain control, leading to a build-up of normeperidine. This build up is exaggerated in patients with renal disease and in the elderly as renal function declines with age [15]. The 2012-2019 updates of the American Geriatric Society’s Beers Criteria strongly recommended avoiding meperidine use in older adults [16]. Meperidine was also removed from the World Health Organization’s List of Essential Medicines in 2003 (Supplemental Table 1).

With a prominent adverse effect profile, especially in vulnerable populations, and little added benefit compared to other analgesics, the question must be asked why is meperidine still being used in the US? Its status as an analgesic of choice for so long could result in older physicians being more likely to continue to use this agent. Additionally, traditional medical teaching has taught that meperidine causes less spasm at the Sphincter of Oddi compared to other opioids. Following this logic, meperidine should be employed in the setting of acute pancreatitis. Although surgeons have been following this practice for decades, there is actually no evidence to support the claim that meperidine does not cause sphincter spasm. Furthermore, no study has been done to evaluate patient outcomes in acute pancreatitis patients given traditional opiates like morphine versus meperidine [17].

As no national pharmacoepidemiological studies conducted in the United States have focused on meperidine, our goals were to: 1) characterize changes in meperidine distribution and use between 2001 and 2019, 2) uncover geographical disparities in meperidine use as reported to the DEA, and 3) determine meperidine prescriber characteristics from Medicare Part D Public Use Files (PUF).

## Methods

### Procedures

Mass of meperidine distributed was obtained from the DEA’s Automation of Reports and Consolidated Orders System (ARCOS) retail drug summary reports for the years 2001-2019 in fifty states. Prior research has shown a high correspondence between ARCOS and state Prescription Drug Monitoring Programs [18-19]. Data was obtained by state (Report 2) and retail type (Report 7) [20]. The yearly final aggregate production quotas for meperidine and intermediates was also obtained from 2001-2021. Medicare Part D Prescriber PUF were obtained for 2013 – 2017 [21]. The year 2017 was chosen because it is the last year data was available when data analysis was completed (7/2020). The IRB of the University of New England deemed these data sources to be exempt.

### Data Analysis

ARCOS retail distribution was reported for the total and by three categories: hospitals, pharmacies and practitioners. Teaching institutions, a category used for non-human use, was not displayed separately due to minute and sporadic volumes and is available elsewhere [22]. The mass of meperidine distributed each year as reported by ARCOS was corrected with US Census Bureau population estimates. State were ranked and values outside of 1.96 standard deviations from the average were considered statistically significant. Adult obesity per state in 2019 was obtained from the Behavioral Risk Factor Surveillance System and correlated with per capita meperidine distribution [23]. Heat maps were created with Excel to visualize distribution disparities.

Medicare Part D PUF data was used to plot two variables over time, total daily supply (TDS) and total drug cost (TDC). In addition, for each year, TDS was examined by prescriber specialty. This allowed us to analyze which specialties accounted for the largest amount of the TDS. Finally, TDS for each specialty was divided by the number of Medicare providers in that specialty to calculate TDS per provider. Specialties with less than 200 Medicare prescribers were excluded from the analysis. Linear regressions over time and figures were completed with GraphPad Prism.

## Results

The total distribution of meperidine as reported to the DEA from 2001 to 2019 decreased by 94.6%. A linear regression of national distribution over time was significant (R^2^(18) = 0.978, *p* < .0001, Figure 1A). All states experienced appreciable decreases in meperidine distributed per capita from ‘01 to ‘19. Iowa (−87.5%), Vermont (−90.2%), and Arkansas (−92.2%) had the smallest reductions while Alaska (−97.7%), Connecticut (−98.0%), and Rhode Island (−98.1%) had the largest (Supplemental Figure 1). Further analysis examined distribution by business activity. In 2001, hospitals and pharmacies accounted for almost half, 49.0% and 49.3% respectively, while practitioners were responsible for 1.7% of distribution. The 2019 distribution was similar for pharmacies (48.2%) but lower for hospitals (41.3%) and higher for practitioners (10.4%, Supplemental Figure 2). The decline from ‘01 to ‘19 for practitioners (−67.2%) was less than that for pharmacies (−94.7%) or hospitals (−95.5%). Hospital reductions began in ‘03 while those of pharmacies did not become pronounced until ‘09 (Figure 1B).

**Figure 1.**
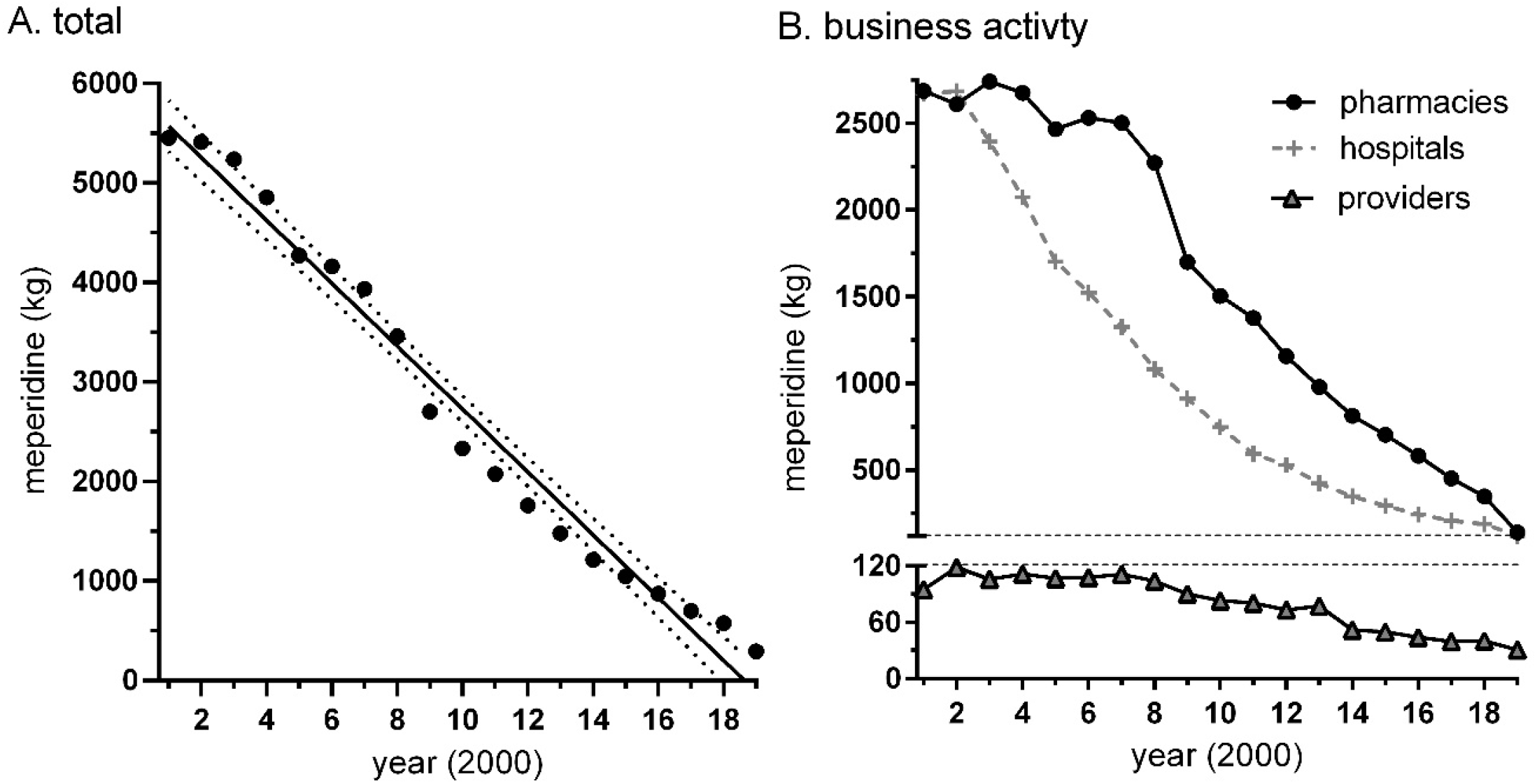
Meperidine total distribution in the United States by weight over time (*r*(17) = −0.989, *p* <.0001) with 95 percent confidence interval (A) and by business activity (B) from 2001 to 2019 as reported by the Drug Enforcement Administration’s Automated Reports and Consolidated Orders System.

**Figure 2.**
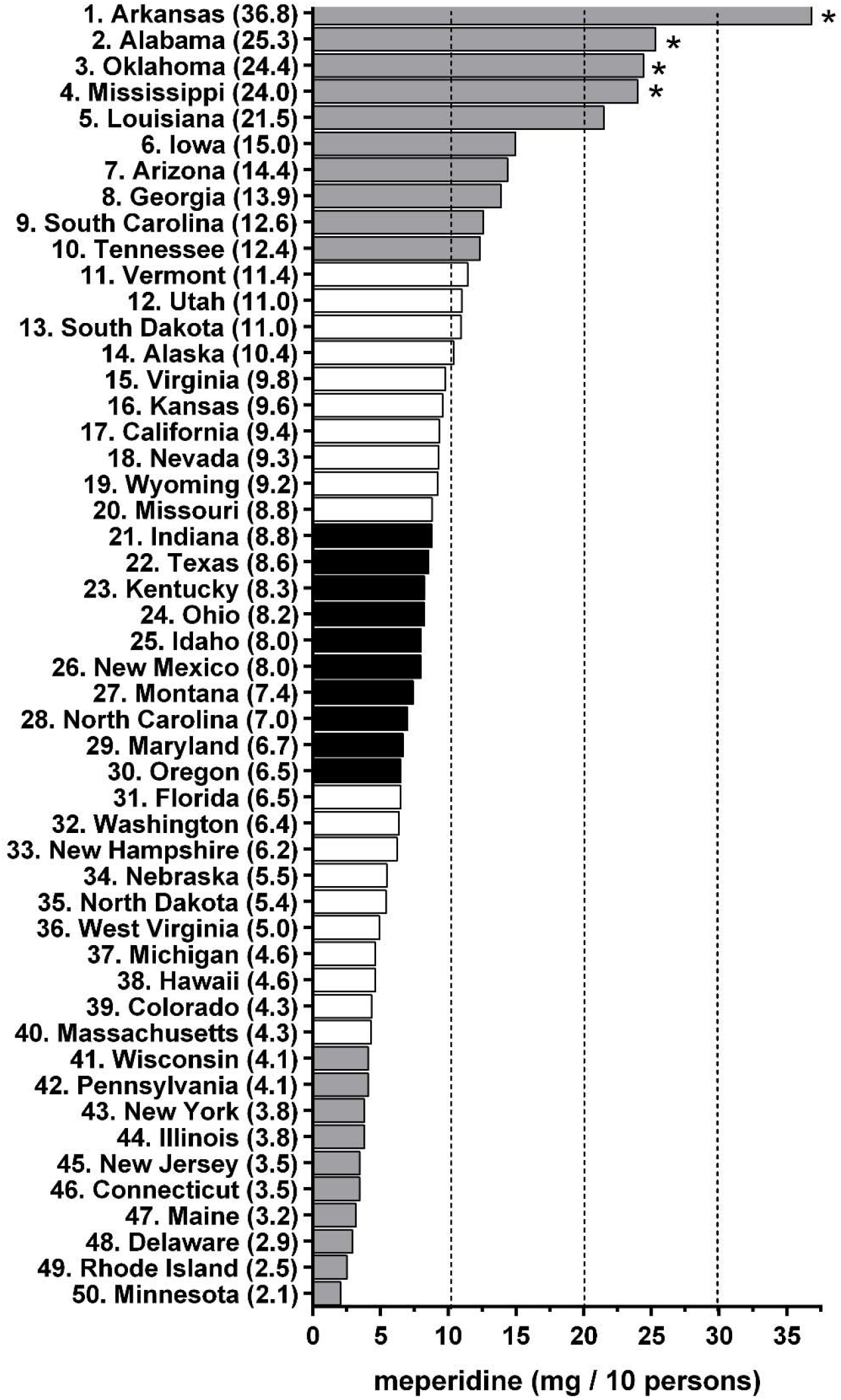
Meperidine distribution per ten persons by state in 2019 as reported by the Drug Enforcement Administration’s Automated Reports and Consolidated Orders System. * *p* < .05 versus the average (9.28 ± 6.82).

Pronounced regional variation was observed in the 2019 meperidine distribution when corrected for population. Meperidine distribution was highest in Arkansas (36.8 mg/10 persons) which was 17.9 fold larger than the lowest (Minnesota = 2.1) state. Furthermore, regional analysis showed that states around Arkansas (Alabama, Oklahoma, and Mississippi) represented the 2nd to 4th largest distribution per 10 persons (Supplemental Figure 3). Each of these four states were significantly elevated relative to the average of the states (Figure 2). This four-state region accounted for 13.8% of the meperidine distributed in 2019, even though only 4.5% of the US population resided in these states. Five of the six lowest states were in the North-East.

**Figure 3.**
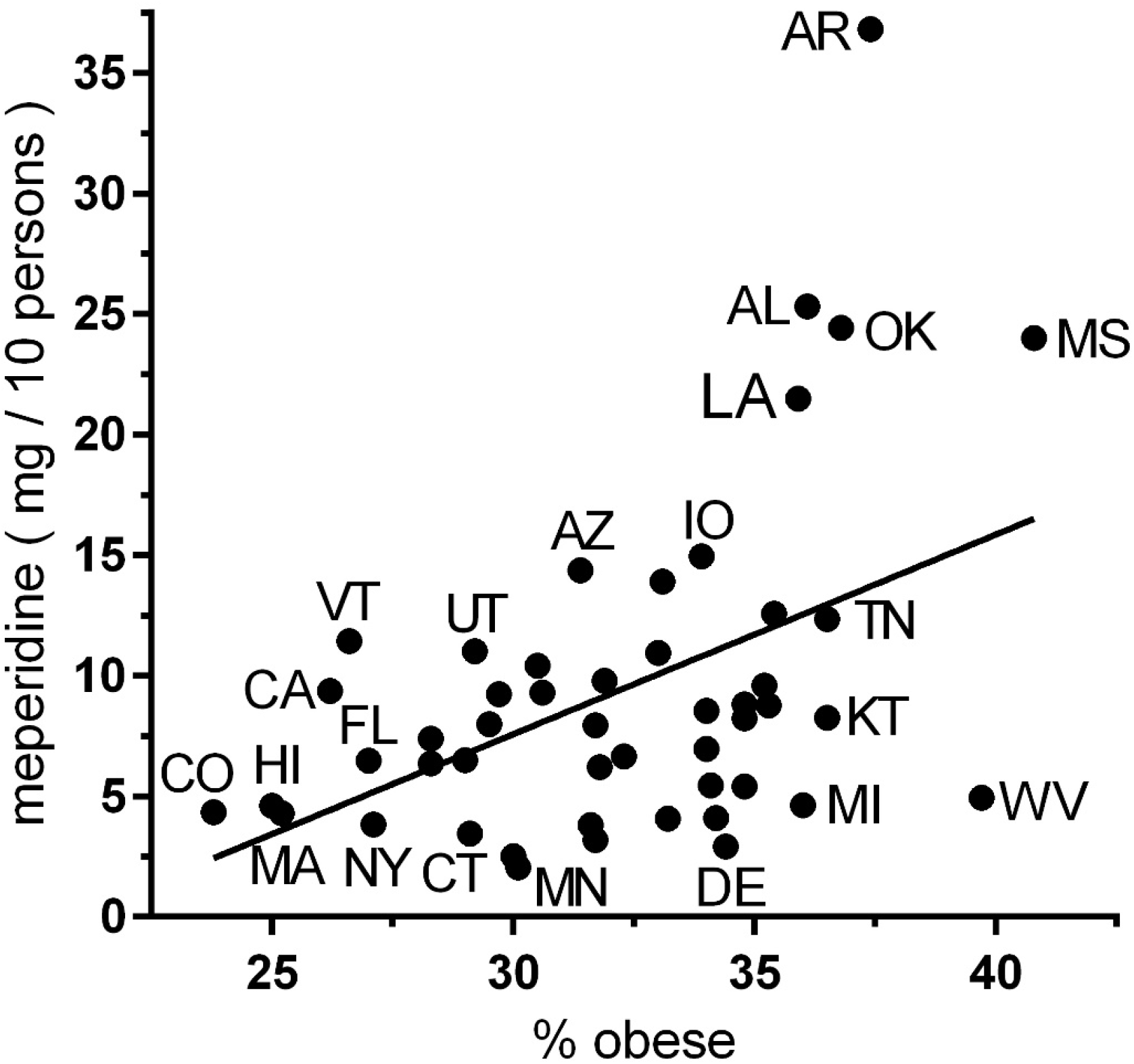
Scatterplot depicting the association of percent obesity according to the Behavioral Risk Factor Surveillance System [23] by meperidine distribution per ten persons per state in 2019 as reported by the Drug Enforcement Administration’s Automated Reports and Consolidated Orders System (*r*(47) = +0.467, *p* < .001).

An exploratory analysis was completed testing if there was an association between meperidine distribution and obesity per state in 2019. States with more obesity also had higher meperidine distribution (R^2^(47) = 0.218, *p* < .001, Figure 3). DEA’s aggregate production quota for meperidine decreased 91.6% from 2001 (10,168 kg) until 2021 (856.7 kg, Supplemental Figure 4).

Analysis of Medicare revealed that between 2013 and 2017, TDS of meperidine decreased by 30.3% while TDC increased by 34.9% ($948,702.10 in 2017). In 2017, Medicare family practice (28.9%) and internal medicine (20.5%) physicians accounted for the largest portion of TDS. However, interventional pain management (5.66) and pain management (3.48) physicians accounted for the longest TDS per provider compared to family medicine (0.69) and internal medicine (0.40). Further information may be found in Supplemental Figures 5-9.

## Discussion

Overall, the US saw a nearly 95% decrease in distribution between 2001 - 2019. Meperidine has played an important role in the history of US medicine and pain management. Meperidine was also involved in the 1976 Barry Kidston case, and a handful of others, with the development of early onset Parkinson’s symptomology and corresponding neuropathology in a young chemistry graduate student attempting to produce synthetic heroin [24]. The fatal 1984 Libby Zion serotonin syndrome case involving meperidine, phenelzine (and possibly cocaine) changed US residency training [25]. The American Pain Society began advocating for restricted meperidine use in the late 1990s [26]. Although hospitals, pharmacies, and practitioners have all decreased their use substantially, individual practitioners have done so at a reduced rate compared to hospitals and pharmacies. There may be a need to provide continuing education classes for these providers to address this problem. As the DEA’s final aggregate production quotas for meperidine (856.7 kg) for 2021 are a 55.2% reduction relative to 2018 [27, 28], this may provide an impetus to encourage health care systems to consider alternative agents.

While it was not surprising to see the large overall decrease in meperidine use as reported to the DEA [18], it was surprising to find pronounced geographical variance. The identification of the Arkansas, Alabama, Oklahoma, and Mississippi region is crucial to understanding meperidine’s pharmacoepidemiological trends and presents a target for further mitigation of meperidine use. The reason for the exaggerated use in this region is unclear. However, we do know meperidine is “indicated” in the setting of acute pancreatitis [29]. Although it is difficult to know rates of acute pancreatitis geographically in the US, it is possible to study risk factors on a geographic level. Risk factors associated with acute pancreatitis include alcohol use, tobacco use, and obesity [30, 31]. Mississippi is the most obese state in the US with an adult rate of 40.8% with Arkansas, Alabama, and Oklahoma having obesity rates over 36% [23]. Due to the prevalence of obesity in the region, it is logical to infer that there may also be concurrent increased prevalence in acute pancreatitis. In fact, we found that meperidine distribution per person significantly corresponded with adult obesity prevalence. While this can not fully explain the inflated use in the four-state region, it is likely a piece of the puzzle. The eighteen-fold difference in meperidine use between states is comparable to the twenty-fold difference identified for buprenorphine [32] and larger than the five-fold difference in the per capita morphine mg equivalent for ten opioids [18].

Medicare Part D data indicated that total day supply decreased substantially from 2013 to 2017. This is exactly what we would expect after looking at the nationwide trends per ARCOS. However, Medicare Part D data also revealed that the total drug cost increased 34.9% over the same period to over nine-hundred thousand in 2017. The reason for the divergent trend in drug cost compared to total day supply is unclear. It is possible that the increasing drug cost of meperidine is due to policies which discourage its use. Analysis of Medicare Part D revealed that family medicine and internal medicine specialties contributed the greatest proportion of total day supply. This is most likely because many physicians in the US practice in primary care. Moreover, the two largest specialties in the realm of primary care are family medicine and internal medicine. In 2010, 21.5% of primary care doctors practiced internal medicine while 38.2% practiced family medicine [21]. Even though the large amount of total day supply coming from these fields is likely accounted for by their large physician populations, it may still present a target for continuing education to restrain meperidine use. When accounting for physician population, we found that pain specialists like interventional pain management and pain management accounted for the largest amount of total day supply per provider. However, it is important to note that they contribute a modest amount of TDS to the sum. Furthermore, these specialists undergo extensive training that may better inform them of risks versus reward when prescribing meperidine relative to primary care physicians.

There are some strengths and limitations to this study. ARCOS is a comprehensive and publically available data source frequently used in research [18, 22, 32]. While meperidine is used for acute pain in non-humans, use by veterinarians is very modest [22, 33]. Although mereridine is always considered inappropriate for older adults (Supplemental Table 1), the version of the Medicare dataset we employed does not allow us to determine how many of the million TDS meperidine recipients were older (≥ age 65) versus disabled. However, older adults account for over eighty percent of Medicare beneficiaries [34]. One limitation from this study was the lack of full prescribing patterns to compare all meperidine distributions across specialties with the Medicare Part D PUF. Additional studies using other health insurances or electronic medical records can help gain further insights into who continues to use meperidine and for which conditions [35-37].

## Conclusion

While the use of meperidine continues to decrease, our analysis has revealed possible areas for further mitigation efforts. The evidence is clear that meperidine puts patients at increased risk with little to no added benefit. Combating adult obesity may also play a role in reducing meperidine use. Eliminating Medicaid reimbursement for older-adult patients and targeted education for health care systems in the four-state (Arkansas, Alabama, Oklahoma, and Mississippi) region identified may lead to improved patient safety.

## Supporting information

Example DEA ARCOS from 2016

## Data Availability

Original data is available at the DEA's ARCOS and Medicare.

https://www.deadiversion.usdoj.gov/arcos/retail_drug_summary/report_yr_2019.pdf

https://www.cms.gov/Research-Statistics-Data-and-Systems/Statistics-Trends-and-Reports/Medicare-Provider-Charge-Data/Part-D-Prescriber

## Acknowledgements

JMB was supported by the Geisinger Commonwealth School of Medicine Summer Research Immersion Program. BJP was supported by the Health Resources and Services Administration (D34HP31025). The National Institute of Environmental Health Sciences (T32 ES007060-31A1) provided software used for figure construction. Technical assistance was provided by Lavinia R. Harrison, Trinidy R. Anthony, Josiah K. Charles, and Iris Johnston.

---

**Supplemental Figure 1.**
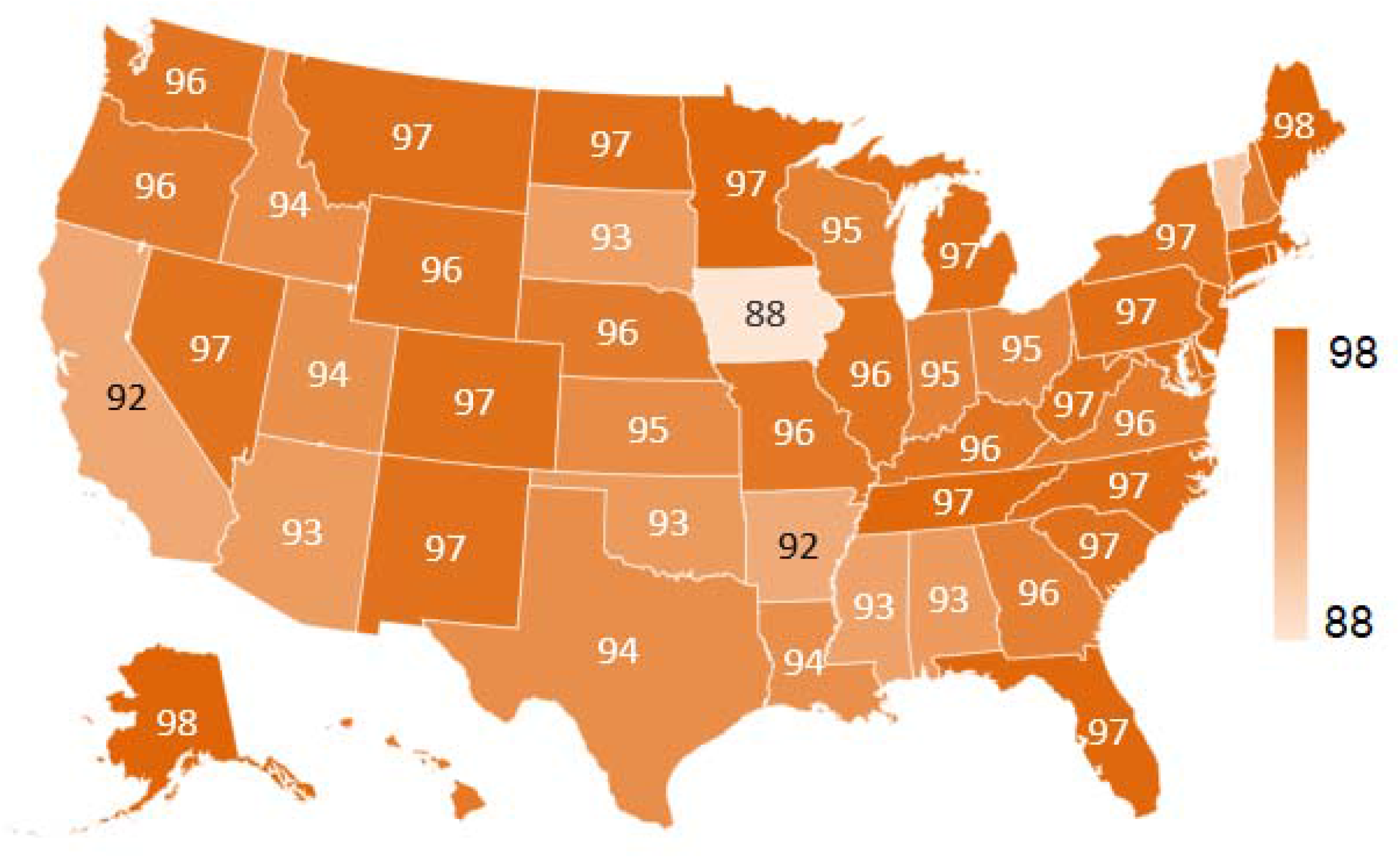
Percent reduction in the distribution of meperidine, corrected for population according to the American Community Survey from 2002 to 2019 as reported to the US Drug Enforcement Administration’s Automated Reports and Consolidated Orders System. Connecticut: 98, Delaware: 97, Hawaii: 96, Maryland: 97, Massachusetts: 97, New Hampshire: 96, New Jersey: 97, Rhode Island: 98, Vermont: 90.

**Supplemental Figure 2.**
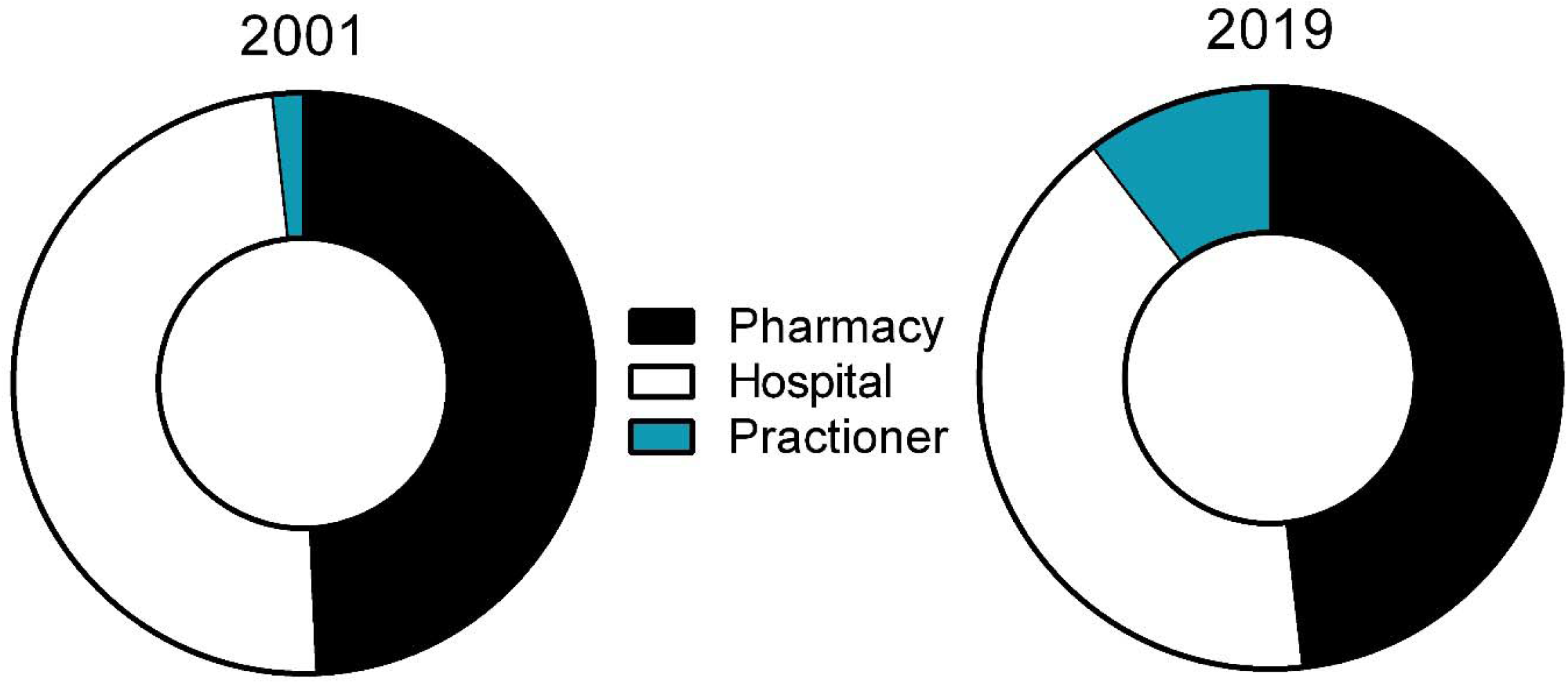
Distribution of meperidine by business activity as reported to the Drug Enforcement Administration’s Automated Reports and Consolidated Orders System showed a six-fold increase to practioners from 2001 (1.74%) to 2019 (10.44%).

**Supplemental Figure 3.**
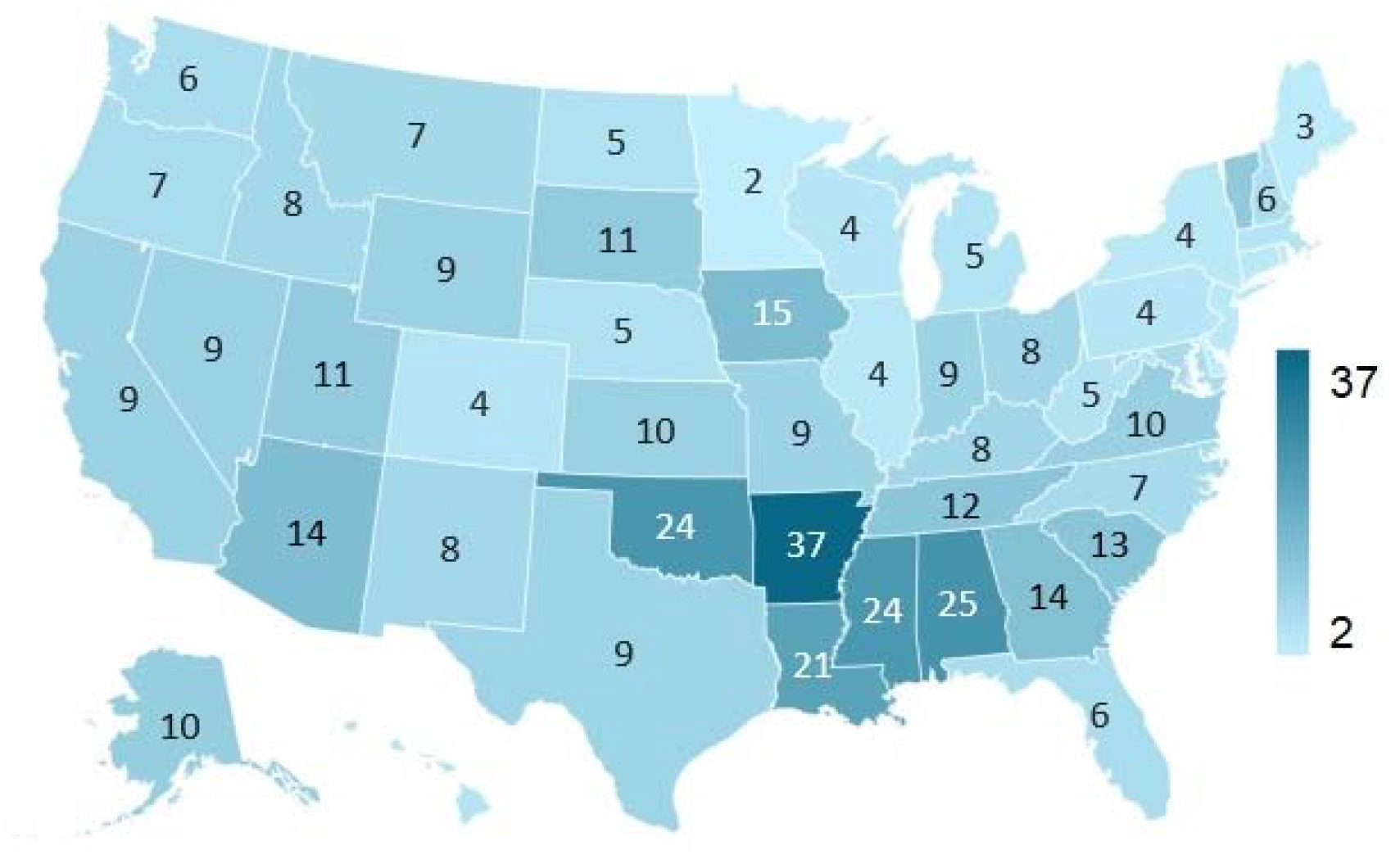
Distribution of meperidine (mg) in 2019 as reported to the US Drug Enforcement Administration’s Automated Reports and Consolidated Orders System (ARCOS). per 10 persons according to the American Community Survey. Connecticut: 3, Delaware: 3, Hawaii: 5, Maryland: 7, Rhode Island: 3, Vermont: 11.

**Supplemental Figure 4.**
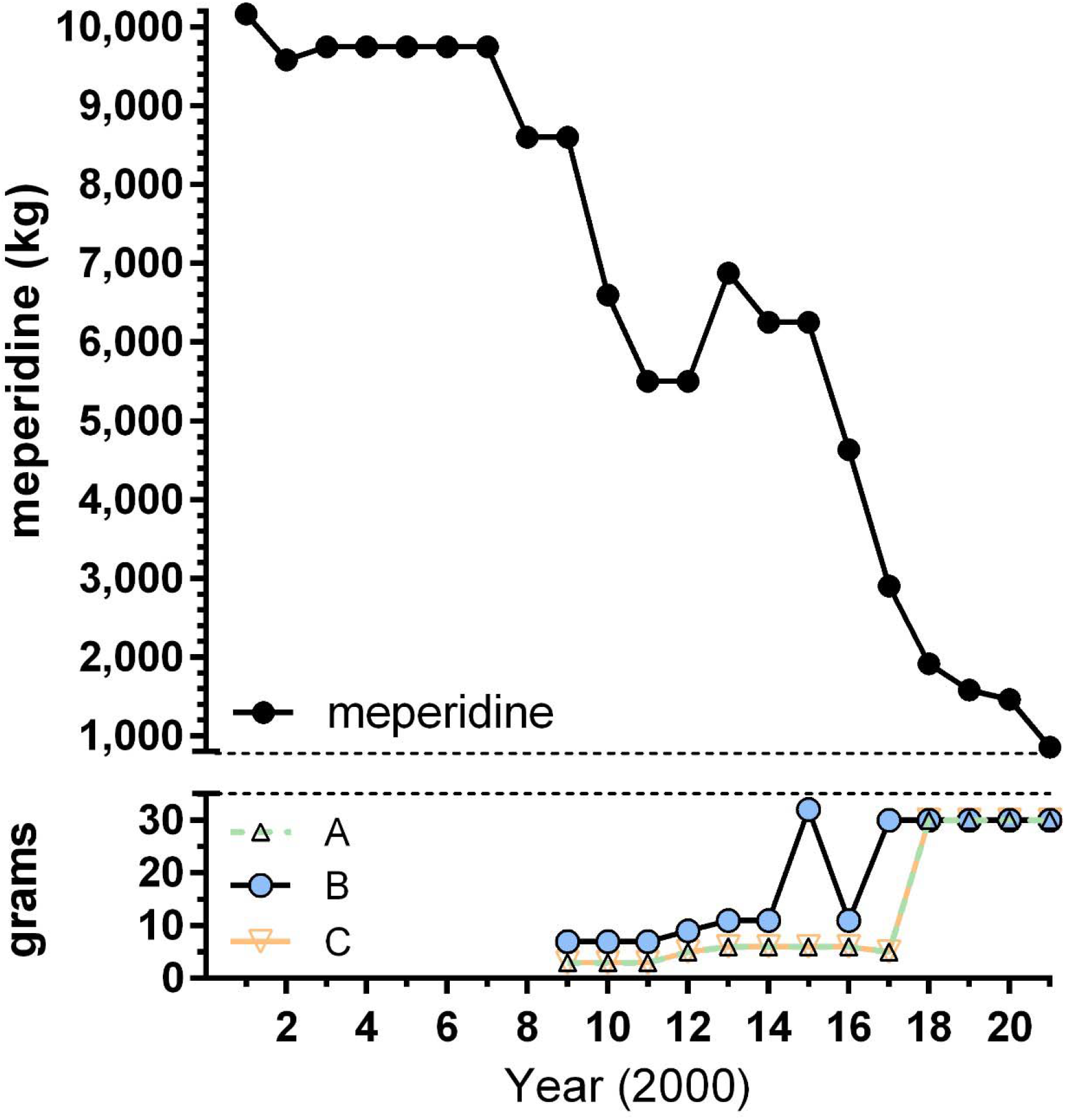
The US Drug Enforcement Administration’s aggregate production quotas for meperidine (kg) and meperidine intermediates A, B, and C (grams). The 2001 to 2017 values were the final quotas, 2019 and 2020 were proposed and 2021 was established.

**Supplemental Figure 5.**
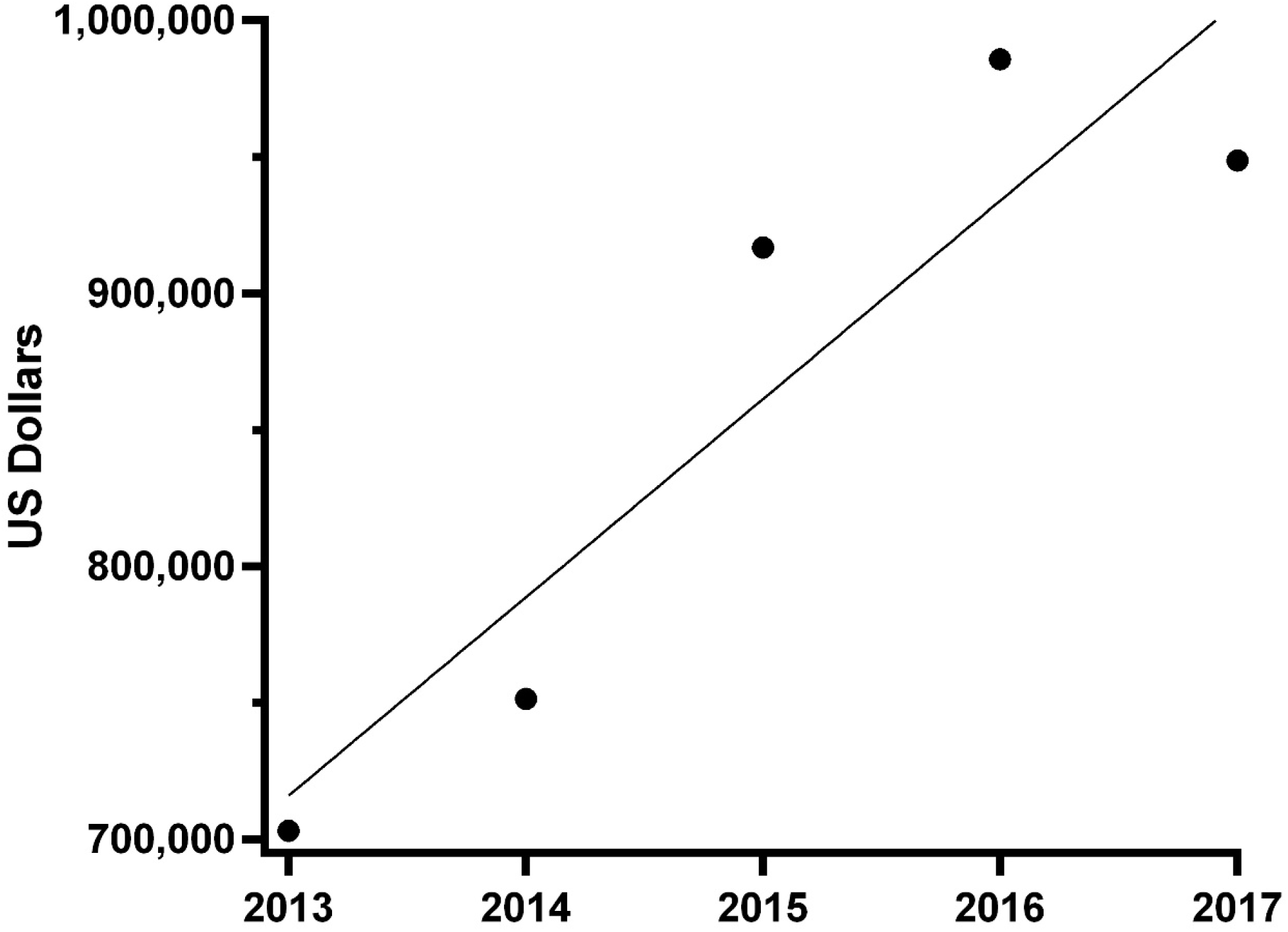
Total drug cost per Medicare Part D Public Use File from 2013 to 2017. Total drug cost is defined as “The aggregate drug cost paid for all associated claims. This amount includes ingredient cost, dispensing fee, sales tax, and any applicable vaccine administration fees and is based on the amounts paid by the Part D plan, Medicare beneficiary, government subsidies, and any other third-party payers”. The year by cost correlation was significant (R^2^(3) = 0.831, *p* < .05).

**Supplemental Figure 6.**
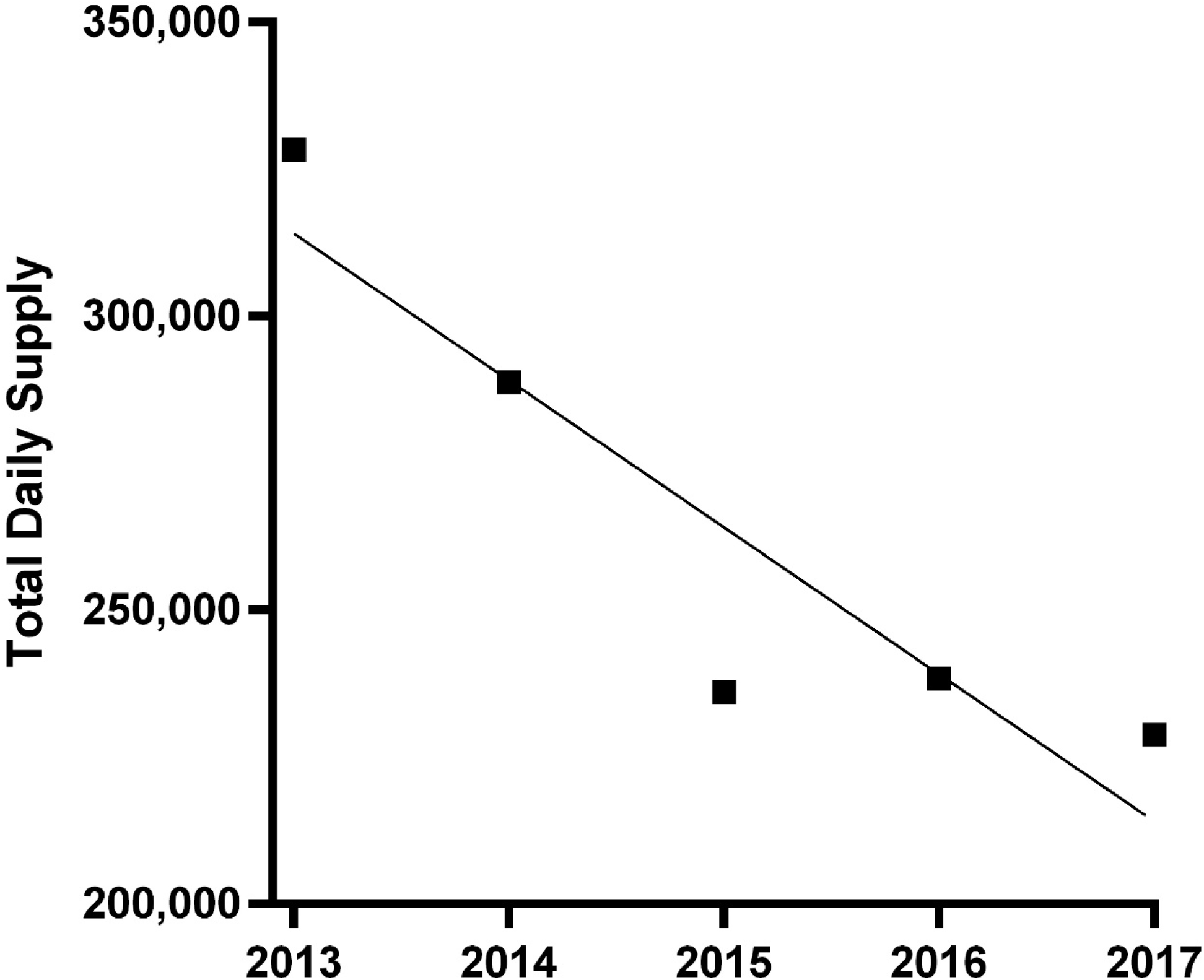
Total daily supply per Medicare Part D Public Use File from 2013 to 2017. Total daily supply is defined as “The aggregate number of day’s supply for which this drug was dispensed”. The year by total daily supply correlation was significant (R^2^(3) = 0.838, *p* < .05).

**Supplemental Figure 7.**
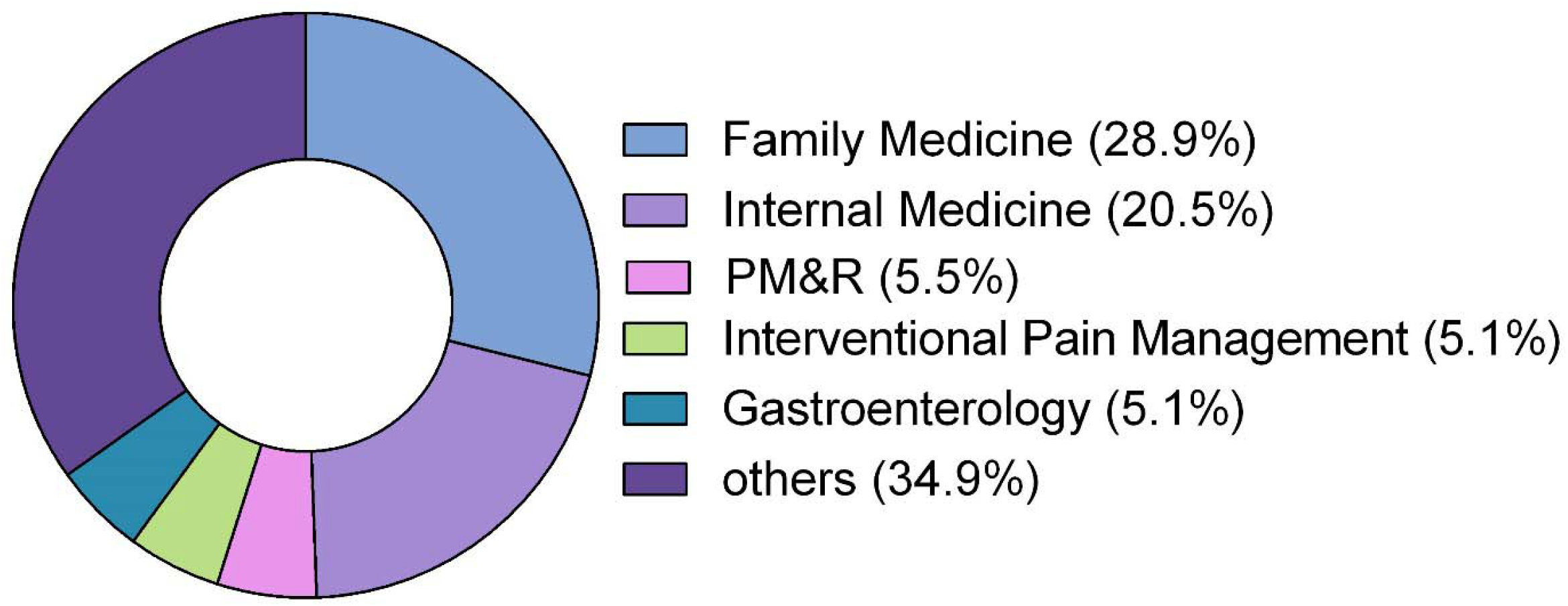
Breakdown of Medicare Part D total daily supply of meperidine in 2017 by specialty. Specialties contributing less than 5% of the whole were grouped as “other”. Physical Medicine and Rehabilitation: PM&R.

**Supplemental Figure 8.**
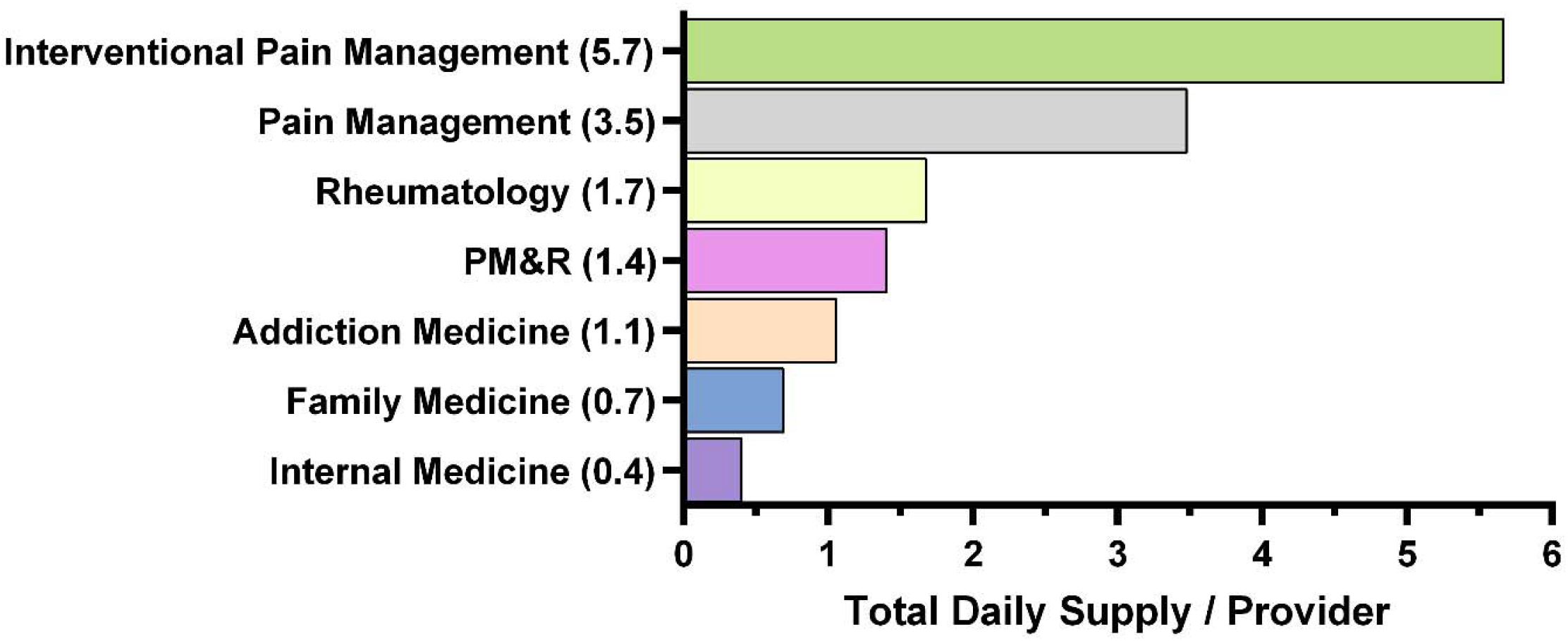
Total daily supply of meperidine prescribed per provider by specialty in Medicare Part D in 2017. The top five specialties for TDS/provider are included on the figure as well as family medicine and internal medicine. Physical Medicine and Rehabilitation: PM&R.

**Supplemental Table 1.**
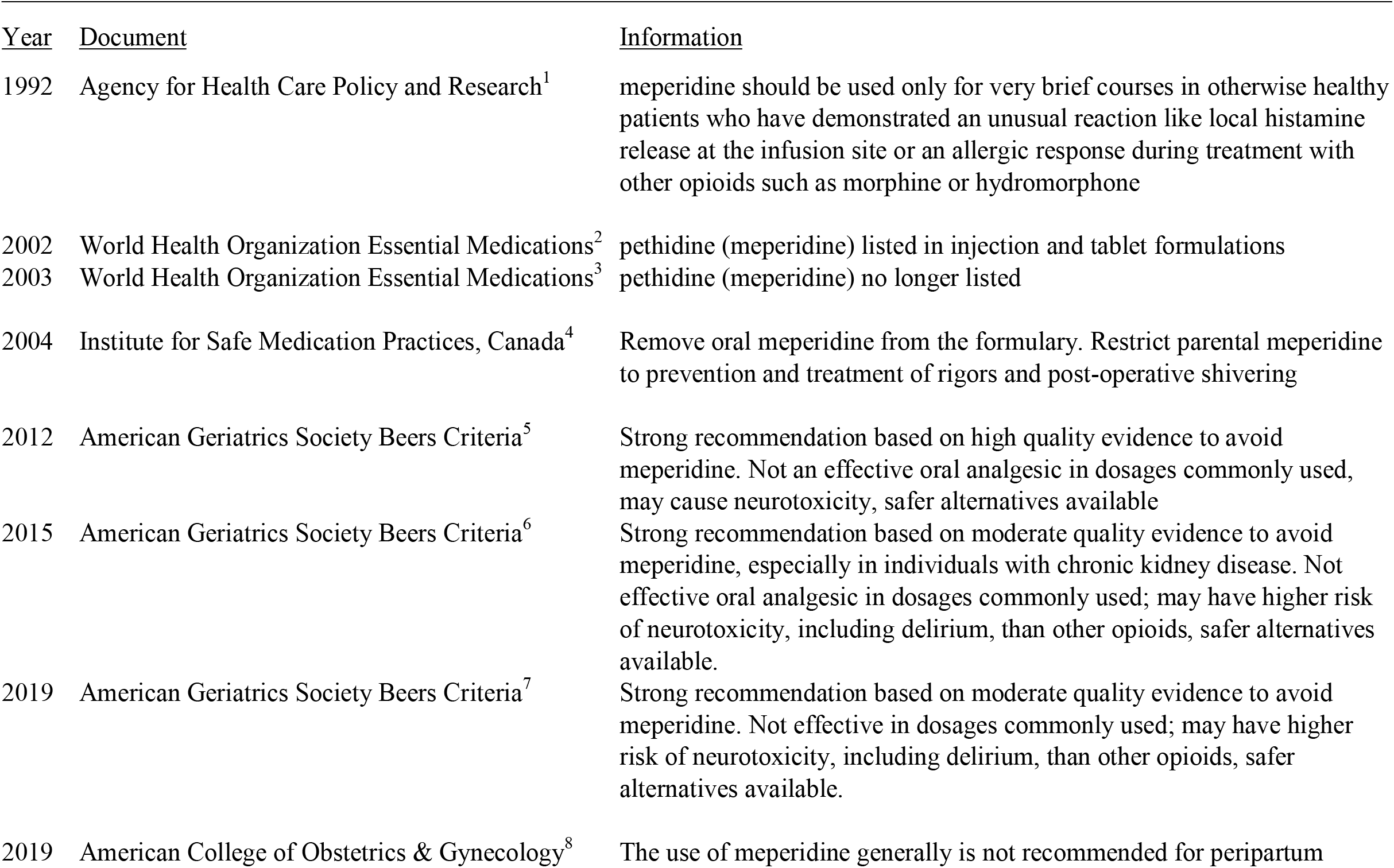
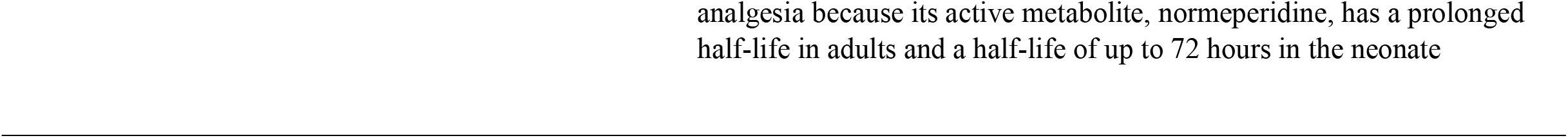
Guidelines and other influential documents and meperidine relevant information.

| <u>Year</u> | <u>Document</u> | <u>Information</u> |
| --- | --- | --- |
| 1992 | Agency for Health Care Policy and Research<sup>1</sup> | meperidine should be used only for very brief courses in otherwise healthy patients who have demonstrated an unusual reaction like local histamine release at the infusion site or an allergic response during treatment with other opioids such as morphine or hydromorphone |
| 2002 | World Health Organization Essential Medications<sup>2</sup> | pethidine (meperidine) listed in injection and tablet formulations |
| 2003 | World Health Organization Essential Medications<sup>3</sup> | pethidine (meperidine) no longer listed |
| 2004 | Institute for Safe Medication Practices, Canada<sup>4</sup> | Remove oral meperidine from the formulary. Restrict parental meperidine to prevention and treatment of rigors and post-operative shivering |
| 2012 | American Geriatrics Society Beers Criteria<sup>5</sup> | Strong recommendation based on high quality evidence to avoid meperidine. Not an effective oral analgesic in dosages commonly used, may cause neurotoxicity, safer alternatives available |
| 2015 | American Geriatrics Society Beers Criteria<sup>6</sup> | Strong recommendation based on moderate quality evidence to avoid meperidine, especially in individuals with chronic kidney disease. Not effective oral analgesic in dosages commonly used; may have higher risk of neurotoxicity, including delirium, than other opioids, safer alternatives available. |
| 2019 | American Geriatrics Society Beers Criteria<sup>7</sup> | Strong recommendation based on moderate quality evidence to avoid meperidine. Not effective in dosages commonly used; may have higher risk of neurotoxicity, including delirium, than other opioids, safer alternatives available. |
| 2019 | American College of Obstetrics & Gynecology<sup>8</sup> | The use of meperidine generally is not recommended for peripartum |

## Notes

**Disclosures:** BJP is part of an osteoarthritis research team supported by Pfizer and Eli Lilly. The other authors have no relevant disclosures.

### Competing Interest Statement

BJP is part of an osteoarthritis research team supported by Pfizer and Eli Lilly. The other authors have no relevant disclosures.

### Clinical Trial

This was not a clinical trial.

### Funding Statement

The Geisinger Commonwealth School of Medicine Summer Research Immersion Program supported the summer stipend of the first author.

### Author Declarations

#20180410-009

