## Supplementary material for "Declines, and Pronounced Regional Disparities, in Meperidine Use in the United States": Example DEA ARCOS from 2016

### RETAIL DRUG DISTRIBUTION BY ZIP CODE WITHIN STATE BY GRAMS WT

DRUG CODE:1100DRUG NAME:AMPHETAMINE

STATE: ALABAMA

| ZIP CODE | QUARTER 1 | QUARTER 2 | QUARTER 3 | QUARTER 4 | TOTAL GRAMS |
|---|---|---|---|---|---|
| 350 | 16,034.16 | 17,001.13 | 17,145.20 | 17,316.18 | 67,496.67 |
| 351 | 8,320.60 | 8,680.25 | 8,732.47 | 8,811.46 | 34,544.78 |
| 352 | 18,259.26 | 18,260.23 | 18,954.25 | 18,976.99 | 74,450.73 |
| 354 | 8,950.98 | 8,671.19 | 8,812.31 | 8,678.92 | 35,113.40 |
| 355 | 3,658.87 | 3,569.32 | 3,521.05 | 3,454.39 | 14,203.63 |
| 356 | 9,395.70 | 9,066.40 | 9,285.71 | 9,142.92 | 36,890.73 |
| 357 | 3,533.20 | 3,732.59 | 3,908.27 | 3,930.91 | 15,104.97 |
| 358 | 4,576.76 | 4,872.61 | 5,201.27 | 5,170.70 | 19,821.34 |
| 359 | 4,415.85 | 4,464.63 | 4,680 | 4,715.34 | 18,275.82 |
| 360 | 6,553.46 | 6,354.41 | 6,657.92 | 6,740.83 | 26,306.62 |
| 361 | 5,712.76 | 5,722.80 | 5,741.89 | 5,759.22 | 22,936.67 |
| 362 | 2,256.28 | 2,259.66 | 2,329.36 | 2,404.22 | 9,249.52 |
| 363 | 4,539.24 | 4,874.87 | 5,098.30 | 4,909.51 | 19,421.92 |
| 364 | 1,267.86 | 1,265.92 | 1,313.17 | 1,353.52 | 5,200.47 |
| 365 | 10,632.38 | 11,347.94 | 11,483.36 | 11,407.55 | 44,871.23 |
| 366 | 8,189.59 | 8,775.67 | 8,456.50 | 7,691.25 | 33,113.01 |
| 367 | 1,462.66 | 1,406.82 | 1,438.83 | 1,502.95 | 5,811.26 |
| 368 | 5,892.66 | 5,945.01 | 6,082.76 | 6,165.69 | 24,086.12 |
| 369 | 124 | 134.59 | 126.98 | 110.87 | 496.44 |
| TOTAL | 123,776.27 | 126,406.04 | 128,969.60 | 128,243.42 | 507,395.33 |

STATE: ALASKA

| ZIP CODE | QUARTER 1 | QUARTER 2 | QUARTER 3 | QUARTER 4 | TOTAL GRAMS |
|---|---|---|---|---|---|
| 995 | 4,567.30 | 4,858.50 | 4,763.02 | 4,747.28 | 18,936.10 |
| 996 | 1,208.11 | 1,394.64 | 1,384.60 | 1,313.36 | 5,300.71 |
| 997 | 1,183.30 | 1,255.83 | 1,241.63 | 1,077.46 | 4,758.22 |
| 998 | 378.46 | 438.45 | 431.46 | 403 | 1,651.37 |
| 999 | 161.8 | 178.87 | 151.49 | 188.96 | 681.12 |
| TOTAL | 7,498.97 | 8,126.29 | 7,972.20 | 7,730.06 | 31,327.52 |

STATE: AMERICAN SAMOA

| ZIP CODE | QUARTER 1 | QUARTER 2 | QUARTER 3 | QUARTER 4 | TOTAL GRAMS |
|---|---|---|---|---|---|
| 967 | 2.53 | 0 | 1.5 | 2.5 | 6.53 |
| TOTAL | 2.53 | 0 | 1.5 | 2.5 | 6.53 |

STATE: ARIZONA

| ZIP CODE | QUARTER 1 | QUARTER 2 | QUARTER 3 | QUARTER 4 | TOTAL GRAMS |
|---|---|---|---|---|---|
| 850 | 15,036.73 | 15,779.70 | 15,873.93 | 16,581.34 | 63,271.70 |
| 851 | 2,490.73 | 2,549.33 | 2,640.82 | 2,771.85 | 10,452.73 |
| 852 | 29,506.29 | 30,174.16 | 30,808.61 | 31,588.50 | 122,077.56 |
| 853 | 9,942.10 | 10,273.60 | 10,235.48 | 10,663.71 | 41,114.89 |
| 855 | 408.5 | 423.48 | 421.39 | 453.09 | 1,706.46 |
| 856 | 1,637.61 | 1,616.48 | 1,533.64 | 1,606.41 | 6,394.14 |
| 857 | 10,153.92 | 10,733.13 | 10,410.67 | 10,690.67 | 41,988.39 |
| 859 | 407.04 | 423.81 | 445.52 | 543.42 | 1,819.79 |
| 860 | 1,245.61 | 1,395.80 | 1,325.35 | 1,301.01 | 5,267.77 |
| 863 | 1,348.15 | 1,534.32 | 1,509.86 | 1,592.85 | 5,985.18 |
| 864 | 948.69 | 897.88 | 881.53 | 936.74 | 3,664.84 |
| 865 | 39.02 | 23.64 | 27.7 | 60.12 | 150.48 |
| TOTAL | 73,164.39 | 75,825.33 | 76,114.50 | 78,789.71 | 303,893.93 |

STATE: ARKANSAS

| ZIP CODE | QUARTER 1 | QUARTER 2 | QUARTER 3 | QUARTER 4 | TOTAL GRAMS |
|---|---|---|---|---|---|
| 716 | 2,662.62 | 2,550.14 | 2,527.95 | 2,788.78 | 10,529.49 |
| 717 | 887.53 | 882.96 | 860.07 | 985.29 | 3,615.85 |
| 718 | 855.97 | 856.88 | 830.82 | 940.19 | 3,483.86 |
| 719 | 3,461.36 | 3,520.20 | 3,369.38 | 3,216.15 | 13,567.09 |

| | | | | | |
|---|---|---|---|---|---|
| 720 | 7,007.26 | 7,130.49 | 6,988.92 | 7,473.19 | 28,599.86 |
| 721 | 5,751.17 | 5,977.15 | 5,931.96 | 5,860.12 | 23,520.40 |
| 722 | 6,730.51 | 7,009.61 | 6,858.61 | 7,020.89 | 27,619.62 |
| 723 | 1,406.42 | 1,352 | 1,470.09 | 1,523.05 | 5,751.56 |
| 724 | 2,653.31 | 2,568.85 | 2,648.86 | 2,766.51 | 10,637.53 |
| 725 | 699.25 | 737.37 | 715.26 | 763.01 | 2,914.89 |

DRUG CODE:1100DRUG NAME:AMPHETAMINE
STATE:ARKANSAS

| ZIP CODE | QUARTER 1 | QUARTER 2 | QUARTER 3 | QUARTER 4 | TOTAL GRAMS |
|---|---|---|---|---|---|
| 726 | 1,517.48 | 1,387.01 | 1,516.42 | 1,469.13 | 5,890.04 |
| 727 | 10,989.94 | 11,312.68 | 11,280.74 | 11,840.18 | 45,423.54 |
| 728 | 913.79 | 914.64 | 892.39 | 881.91 | 3,602.73 |
| 729 | 1,870.74 | 1,795.45 | 1,851.44 | 2,014.24 | 7,531.87 |
| TOTAL | 47,407.35 | 47,995.43 | 47,742.91 | 49,542.64 | 192,688.33 |

STATE: CALIFORNIA

| ZIP CODE | QUARTER 1 | QUARTER 2 | QUARTER 3 | QUARTER 4 | TOTAL GRAMS |
|---|---|---|---|---|---|
| 900 | 18,948.06 | 20,284.14 | 20,373.06 | 20,809.46 | 80,414.72 |
| 902 | 11,891.81 | 12,482.57 | 12,436.65 | 12,397.27 | 49,208.30 |
| 903 | 248.67 | 292.17 | 240.32 | 263.44 | 1,044.60 |
| 904 | 3,953.61 | 3,857.72 | 3,983 | 3,929.95 | 15,724.28 |
| 905 | 1,639.81 | 1,665.85 | 1,692.10 | 1,761.52 | 6,759.28 |
| 906 | 2,303.50 | 2,219.88 | 2,326.37 | 2,336.52 | 9,186.27 |
| 907 | 3,101.47 | 3,110.92 | 3,089.88 | 3,207.04 | 12,509.31 |
| 908 | 2,737.60 | 2,844.97 | 2,854.36 | 3,006.11 | 11,443.04 |
| 910 | 1,621.59 | 1,650.85 | 1,770.51 | 1,555.35 | 6,598.30 |
| 911 | 2,407.55 | 2,372.76 | 2,413.27 | 2,500.55 | 9,694.13 |
| 912 | 1,407.54 | 1,581.90 | 1,614.28 | 1,526.86 | 6,130.58 |
| 913 | 13,419.97 | 13,866.82 | 13,889.90 | 14,103.60 | 55,280.29 |
| 914 | 3,807.25 | 3,828.78 | 3,883.18 | 4,029.47 | 15,548.68 |
| 915 | 1,669.34 | 1,676.17 | 1,784.32 | 1,763.78 | 6,893.61 |
| 916 | 1,800.57 | 1,944.04 | 1,935.59 | 1,868.02 | 7,548.22 |
| 917 | 6,714.99 | 6,746.25 | 6,503.42 | 6,803.23 | 26,767.89 |
| 918 | 320.26 | 332.46 | 327.76 | 378.82 | 1,359.30 |
| 919 | 2,484.16 | 2,623.22 | 2,731.98 | 2,702.14 | 10,541.50 |
| 920 | 25,344.73 | 27,504 | 27,373.07 | 27,435.55 | 107,657.35 |
| 921 | 16,517.51 | 16,807.51 | 16,998.64 | 17,312.69 | 67,636.35 |
| 922 | 3,410.97 | 3,349.17 | 3,126.28 | 3,513.88 | 13,400.30 |
| 923 | 3,910 | 3,954.28 | 3,925.07 | 4,137.27 | 15,926.62 |
| 924 | 422.05 | 332.9 | 390.4 | 351.49 | 1,496.84 |
| 925 | 5,976.84 | 6,126.97 | 6,023.60 | 6,501.81 | 24,629.22 |
| 926 | 20,589.10 | 20,913.30 | 21,027.40 | 21,870.05 | 84,399.85 |
| 927 | 4,666.09 | 4,663.23 | 4,788.58 | 4,752.80 | 18,870.70 |
| 928 | 7,693.73 | 8,084.42 | 7,689.78 | 8,129.82 | 31,597.75 |
| 930 | 5,577.46 | 5,560.54 | 5,592.94 | 5,664.72 | 22,395.66 |
| 931 | 3,366.81 | 3,502.31 | 3,187.52 | 3,433.75 | 13,490.39 |
| 932 | 1,798.62 | 1,704.72 | 1,719.95 | 1,801.99 | 7,025.28 |
| 933 | 1,909.99 | 1,874.97 | 1,877.38 | 1,947.15 | 7,609.49 |
| 934 | 3,218.37 | 3,492.90 | 3,225.09 | 3,444.79 | 13,381.15 |
| 935 | 2,236.49 | 2,258.97 | 2,126.22 | 2,375.12 | 8,996.80 |
| 936 | 2,098.58 | 2,168.05 | 2,106.85 | 2,151.84 | 8,525.32 |
| 937 | 3,013.44 | 3,049.93 | 3,146.61 | 3,307.46 | 12,517.44 |
| 939 | 1,656.30 | 1,670.99 | 1,777.19 | 1,683.08 | 6,787.56 |
| 940 | 5,684.85 | 5,706.41 | 5,753.45 | 5,755.91 | 22,900.62 |
| 941 | 10,974.54 | 11,487.88 | 11,961.76 | 11,402.10 | 45,826.28 |
| 943 | 979.57 | 1,064.40 | 1,025.78 | 1,030.76 | 4,100.51 |
| 944 | 1,079.48 | 1,156.93 | 1,241.05 | 1,204.41 | 4,681.87 |
| 945 | 15,672.12 | 15,360.85 | 15,065.79 | 15,671.73 | 61,770.49 |
| 946 | 3,417.94 | 3,328.53 | 3,341.63 | 3,412.16 | 13,500.26 |

| | | | | | |
|---|---|---|---|---|---|
| 947 | 1,273.72 | 1,354.07 | 1,266.11 | 1,461.86 | 5,355.76 |
| 948 | 710.95 | 520.46 | 686.94 | 627.13 | 2,545.48 |
| 949 | 4,462.56 | 4,665.61 | 4,742.15 | 4,681.79 | 18,552.11 |
| 950 | 7,058.84 | 7,074.52 | 6,959.92 | 7,142.94 | 28,236.22 |
| 951 | 3,781.53 | 3,903.09 | 4,107.49 | 4,054.56 | 15,846.67 |
| 952 | 2,075.06 | 1,926.68 | 1,833.04 | 1,976.10 | 7,810.88 |
| 953 | 4,986.71 | 4,997.65 | 4,737.16 | 5,130.77 | 19,852.29 |
| 954 | 3,651.88 | 3,484 | 3,756.72 | 3,652.72 | 14,545.32 |
| 955 | 828.84 | 840.3 | 784.55 | 822.54 | 3,276.23 |
| 956 | 10,621.07 | 10,411.36 | 10,134.61 | 10,681.54 | 41,848.58 |
| 957 | 2,332.13 | 2,148.86 | 2,225.90 | 2,452.39 | 9,159.28 |
| 958 | 4,874.61 | 4,768.73 | 4,796.21 | 5,082.24 | 19,521.79 |
| 959 | 4,370.11 | 4,436.35 | 4,320.96 | 4,409.48 | 17,536.90 |
| 960 | 1,526.79 | 1,529.22 | 1,578.75 | 1,618.08 | 6,252.84 |
| 961 | 491.65 | 505.7 | 499.29 | 559.98 | 2,056.62 |
| TOTAL | 284,739.78 | 291,072.23 | 290,771.78 | 297,589.58 | 1,164,173.37 |

DRUG CODE:1100DRUG NAME:AMPHETAMINE
STATE: COLORADO

| ZIP CODE | QUARTER 1 | QUARTER 2 | QUARTER 3 | QUARTER 4 | TOTAL GRAMS |
|---|---|---|---|---|---|
| 800 | 10,818.40 | 11,329.79 | 11,598.84 | 11,622.96 | 45,369.99 |
| 801 | 12,329.39 | 13,205.83 | 13,737.90 | 13,610.14 | 52,883.26 |
| 802 | 17,253.61 | 18,467.64 | 18,866.91 | 18,786.97 | 73,375.13 |
| 803 | 3,251.11 | 3,349.94 | 3,274.98 | 3,429.89 | 13,305.92 |
| 804 | 2,756.03 | 2,892.30 | 2,938.76 | 2,895.59 | 11,482.68 |
| 805 | 6,721.54 | 7,110.02 | 7,191.01 | 7,498.32 | 28,520.89 |
| 806 | 2,180.17 | 2,226.84 | 2,283.24 | 2,266.98 | 8,957.23 |
| 807 | 300.8 | 341.58 | 299.4 | 359.03 | 1,300.81 |
| 808 | 1,115.61 | 1,164.67 | 1,155.14 | 1,118.75 | 4,554.17 |
| 809 | 6,869.41 | 7,195.41 | 7,522.74 | 7,167.36 | 28,754.92 |
| 810 | 1,219.78 | 1,269.16 | 1,316.40 | 1,344.16 | 5,149.50 |
| 811 | 209.64 | 216.36 | 225.17 | 229.04 | 880.21 |
| 812 | 532.66 | 597.62 | 595.11 | 564.6 | 2,289.99 |
| 813 | 652.91 | 724.97 | 712.9 | 676.96 | 2,767.74 |
| 814 | 478.83 | 446.76 | 470.73 | 497.82 | 1,894.14 |
| 815 | 1,518.68 | 1,495.23 | 1,626.91 | 1,553.22 | 6,194.04 |
| 816 | 2,062.95 | 2,145.55 | 2,273.82 | 2,307.62 | 8,789.94 |
| TOTAL | 70,271.52 | 74,179.67 | 76,089.96 | 75,929.41 | 296,470.56 |

STATE: CONNECTICUT

| ZIP CODE | QUARTER 1 | QUARTER 2 | QUARTER 3 | QUARTER 4 | TOTAL GRAMS |
|---|---|---|---|---|---|
| 60 | 8,395.31 | 8,519.29 | 8,733.70 | 8,822.96 | 34,471.26 |
| 61 | 3,153.49 | 3,236.56 | 3,069.74 | 3,317.38 | 12,777.17 |
| 62 | 2,078.94 | 2,044.42 | 2,050.11 | 2,130.97 | 8,304.44 |
| 63 | 4,420.23 | 4,649.47 | 4,996.96 | 4,997.01 | 19,063.67 |
| 64 | 11,509 | 12,064.83 | 12,288.03 | 12,107.41 | 47,969.27 |
| 65 | 3,860.19 | 3,797.40 | 3,800.97 | 3,773.30 | 15,231.86 |
| 66 | 1,286.48 | 1,338.99 | 1,313.01 | 1,430.41 | 5,368.89 |
| 67 | 3,036.95 | 3,141.63 | 3,123.63 | 3,265.93 | 12,568.14 |
| 68 | 9,281.19 | 9,483.09 | 9,786.66 | 9,828.30 | 38,379.24 |
| 69 | 1,487.16 | 1,620.45 | 1,729.30 | 1,631.10 | 6,468.01 |
| TOTAL | 48,508.94 | 49,896.13 | 50,892.11 | 51,304.77 | 200,601.95 |

STATE: DELAWARE

| ZIP CODE | QUARTER 1 | QUARTER 2 | QUARTER 3 | QUARTER 4 | TOTAL GRAMS |
|---|---|---|---|---|---|
| 197 | 8,964.29 | 9,293.91 | 9,080.34 | 9,263.22 | 36,601.76 |
| 198 | 6,609.55 | 6,832.45 | 6,768.44 | 6,788.26 | 26,998.70 |
| 199 | 5,291.74 | 5,550.91 | 5,742.81 | 5,779.65 | 22,365.11 |
| TOTAL | 20,865.58 | 21,677.27 | 21,591.59 | 21,831.13 | 85,965.57 |

RETAIL DRUG DISTRIBUTION BY ZIP CODE WITHIN STATE BY GRAMS WT

STATE: DISTRICT OF COLUMBIA

| ZIP CODE | QUARTER 1 | QUARTER 2 | QUARTER 3 | QUARTER 4 | TOTAL GRAMS |
|---|---|---|---|---|---|
| 200 | 10,391.51 | 10,893.11 | 10,814.41 | 10,894.90 | 42,993.93 |
| 203 | 115.51 | 159.31 | 171.02 | 51.93 | 497.77 |
| 204 | 346.13 | 348.04 | 438.96 | 341.67 | 1,474.80 |
| 205 | 0 | 0 | 0.37 | 0 | 0.37 |
| TOTAL | 10,853.15 | 11,400.46 | 11,424.76 | 11,288.50 | 44,966.87 |

STATE: FLORIDA

| ZIP CODE | QUARTER 1 | QUARTER 2 | QUARTER 3 | QUARTER 4 | TOTAL GRAMS |
|---|---|---|---|---|---|
| 320 | 7,894.40 | 8,375.31 | 8,610.48 | 8,751.12 | 33,631.31 |
| 321 | 6,865.23 | 7,275.23 | 7,044.18 | 7,404.32 | 28,588.96 |
| 322 | 19,270.97 | 20,518.17 | 21,691.49 | 20,751.12 | 82,231.75 |
| 323 | 9,032.07 | 9,425.65 | 8,915.38 | 9,567.84 | 36,940.94 |
| 324 | 7,729.92 | 8,018.40 | 8,020.36 | 8,277.27 | 32,045.95 |
| 325 | 16,982.47 | 17,728.92 | 18,523.97 | 18,084.55 | 71,319.91 |
| 326 | 5,180.71 | 5,393.62 | 5,465.37 | 5,785.09 | 21,824.79 |
| 327 | 14,231.73 | 15,175.32 | 15,851.94 | 16,308.10 | 61,567.09 |
| 328 | 11,424.59 | 12,668.88 | 12,556.22 | 13,032.61 | 49,682.30 |
| 329 | 11,506.17 | 11,870.44 | 12,264.63 | 12,576.71 | 48,217.95 |
| 330 | 12,548.51 | 13,804.69 | 13,705.93 | 14,220.67 | 54,279.80 |
| 331 | 14,060.12 | 14,974.21 | 15,057.16 | 15,546.89 | 59,638.38 |
| 333 | 10,106.29 | 11,331.46 | 11,475.04 | 11,376.29 | 44,289.08 |
| 334 | 21,157.43 | 22,522.59 | 22,290.99 | 23,205.25 | 89,176.26 |
| 335 | 8,258.92 | 9,139.63 | 9,442.79 | 9,835.49 | 36,676.83 |
| 336 | 11,894.98 | 13,021.63 | 13,393.66 | 13,564.93 | 51,875.20 |
| 337 | 11,433.23 | 12,507.07 | 12,709.20 | 12,796.20 | 49,445.70 |
| 338 | 6,055.51 | 6,408.38 | 6,256.23 | 6,907.11 | 25,627.23 |
| 339 | 10,541.31 | 11,116.90 | 11,488.77 | 12,164.64 | 45,311.62 |
| 341 | 5,681.51 | 5,897.19 | 5,832.26 | 6,437.07 | 23,848.03 |

DRUG CODE:1100DRUG NAME:AMPHETAMINE
STATE:FLORIDA

| ZIP CODE | QUARTER 1 | QUARTER 2 | QUARTER 3 | QUARTER 4 | TOTAL GRAMS |
|---|---|---|---|---|---|
| 342 | 11,408.55 | 12,320.61 | 12,198.62 | 12,829.65 | 48,757.43 |
| 344 | 3,450.40 | 3,886.76 | 3,810.75 | 3,967.86 | 15,115.77 |
| 346 | 7,420.44 | 7,900.07 | 7,984.69 | 8,181.66 | 31,486.86 |
| 347 | 6,768.27 | 7,229.40 | 7,381.95 | 7,580.73 | 28,960.35 |
| 349 | 6,039.03 | 6,146.65 | 6,111.28 | 6,438.41 | 24,735.37 |
| TOTAL | 256,942.76 | 274,657.18 | 278,083.34 | 285,591.58 | 1,095,274.86 |

STATE: GEORGIA

| ZIP CODE | QUARTER 1 | QUARTER 2 | QUARTER 3 | QUARTER 4 | TOTAL GRAMS |
|---|---|---|---|---|---|
| 300 | 34,841.27 | 36,460.68 | 37,233.62 | 37,645.25 | 146,180.82 |
| 301 | 20,130.87 | 19,709.95 | 21,010.64 | 21,297.69 | 82,149.15 |
| 302 | 12,240.18 | 12,265.69 | 12,497.73 | 12,759.02 | 49,762.62 |
| 303 | 19,930.45 | 21,143.57 | 21,380.17 | 21,725.66 | 84,179.85 |
| 304 | 6,716.84 | 6,596.65 | 6,786.04 | 6,875.45 | 26,974.98 |
| 305 | 10,301.79 | 10,801.51 | 11,019.56 | 11,550.24 | 43,673.10 |
| 306 | 7,322.87 | 7,319.23 | 7,407.97 | 7,755.34 | 29,805.41 |
| 307 | 5,722.53 | 6,034.41 | 6,226.90 | 6,497.57 | 24,481.41 |
| 308 | 3,227.61 | 3,097.18 | 3,295.14 | 3,401.95 | 13,021.88 |
| 309 | 5,153.45 | 5,279.54 | 5,570.49 | 5,468.02 | 21,471.50 |
| 310 | 6,462.72 | 6,413.70 | 6,669.80 | 7,046.01 | 26,592.23 |
| 312 | 2,671.05 | 2,636.79 | 2,695.12 | 2,790.16 | 10,793.12 |
| 313 | 5,267.64 | 5,415.69 | 5,819.75 | 5,685.24 | 22,188.32 |
| 314 | 5,565.26 | 5,822.85 | 5,754.90 | 5,809.28 | 22,952.29 |
| 315 | 6,985.39 | 6,987.42 | 7,469.81 | 7,506.17 | 28,948.79 |
| 316 | 3,178.56 | 3,160.29 | 3,311.45 | 3,365.65 | 13,015.95 |

| | | | | | |
|---|---|---|---|---|---|
| 317 | 6,272.73 | 5,845.33 | 6,233.70 | 6,269.08 | 24,620.84 |
| 318 | 547.23 | 451.91 | 511.9 | 549.68 | 2,060.72 |
| 319 | 6,884.78 | 6,865.19 | 6,912.29 | 6,576.82 | 27,239.08 |
| 398 | 1,522.72 | 1,404.41 | 1,436.26 | 1,492.46 | 5,855.85 |
| TOTAL | 170,945.94 | 173,711.99 | 179,243.24 | 182,066.74 | 705,967.91 |

STATE: GUAM

| ZIP CODE | QUARTER 1 | QUARTER 2 | QUARTER 3 | QUARTER 4 | TOTAL GRAMS |
|---|---|---|---|---|---|
| 969 | 397.32 | 392.44 | 490.58 | 474.36 | 1,754.70 |
| TOTAL | 397.32 | 392.44 | 490.58 | 474.36 | 1,754.70 |

STATE: HAWAII

| ZIP CODE | QUARTER 1 | QUARTER 2 | QUARTER 3 | QUARTER 4 | TOTAL GRAMS |
|---|---|---|---|---|---|
| 967 | 4,351.21 | 4,220.99 | 4,437.32 | 4,446.29 | 17,455.81 |
| 968 | 4,740.93 | 4,395.45 | 5,333.17 | 4,525.04 | 18,994.59 |
| TOTAL | 9,092.14 | 8,616.44 | 9,770.49 | 8,971.33 | 36,450.40 |

STATE: IDAHO

| ZIP CODE | QUARTER 1 | QUARTER 2 | QUARTER 3 | QUARTER 4 | TOTAL GRAMS |
|---|---|---|---|---|---|
| 832 | 1,803.52 | 1,829.38 | 1,787.38 | 1,915.52 | 7,335.80 |
| 833 | 1,752.79 | 1,786.57 | 1,726.77 | 1,703.14 | 6,969.27 |
| 834 | 4,013.68 | 4,084.59 | 4,086.13 | 4,295.59 | 16,479.99 |
| 835 | 308.89 | 344.26 | 356.48 | 367.01 | 1,376.64 |
| 836 | 6,258.75 | 6,823.55 | 6,792.40 | 6,763 | 26,637.70 |
| 837 | 6,421.47 | 7,006.19 | 6,847.68 | 6,947.71 | 27,223.05 |
| 838 | 1,788.41 | 1,874.01 | 1,852.45 | 1,964.42 | 7,479.29 |
| TOTAL | 22,347.51 | 23,748.55 | 23,449.29 | 23,956.39 | 93,501.74 |

STATE: ILLINOIS

| ZIP CODE | QUARTER 1 | QUARTER 2 | QUARTER 3 | QUARTER 4 | TOTAL GRAMS |
|---|---|---|---|---|---|
| 600 | 29,348.04 | 31,171.04 | 31,796.33 | 31,740.62 | 124,056.03 |
| 601 | 19,386.03 | 20,342.86 | 21,180.74 | 21,815.65 | 82,725.28 |
| 602 | 1,538.76 | 1,504.75 | 1,330.43 | 1,455.04 | 5,828.98 |
| 603 | 1,105.34 | 1,116.47 | 1,170.52 | 1,223.97 | 4,616.30 |
| 604 | 16,206.25 | 17,122.54 | 17,410.89 | 17,968.29 | 68,707.97 |
| 605 | 14,947.27 | 16,167.19 | 16,234.11 | 16,761.46 | 64,110.03 |
| 606 | 32,470.37 | 35,483.08 | 35,634.86 | 35,771.43 | 139,359.74 |
| 607 | 1,843.41 | 2,021.19 | 2,121.71 | 2,121.17 | 8,107.48 |
| 608 | 524.74 | 535.88 | 523.89 | 594.16 | 2,178.67 |
| 609 | 1,711.62 | 1,863.50 | 1,858.55 | 1,964.63 | 7,398.30 |
| 610 | 3,311.58 | 3,365.88 | 3,304.05 | 3,436.49 | 13,418 |
| 611 | 3,526.27 | 3,579.36 | 3,477.54 | 3,702.52 | 14,285.69 |
| 612 | 2,940.07 | 2,965.14 | 3,110.85 | 3,187.50 | 12,203.56 |
| 613 | 2,008.29 | 2,203.95 | 2,197.39 | 2,278.35 | 8,687.98 |
| 614 | 1,854.10 | 1,773.82 | 1,876.83 | 1,819.94 | 7,324.69 |
| 615 | 3,097.81 | 3,130.96 | 3,105.37 | 3,281.43 | 12,615.57 |

DRUG CODE:1100DRUG NAME:AMPHETAMINE
STATE:ILLINOIS

| ZIP CODE | QUARTER 1 | QUARTER 2 | QUARTER 3 | QUARTER 4 | TOTAL GRAMS |
|---|---|---|---|---|---|
| 616 | 3,859.13 | 3,977.09 | 4,018.03 | 3,932.70 | 15,786.95 |
| 617 | 3,502.17 | 3,642.83 | 3,481.60 | 3,732.16 | 14,358.76 |
| 618 | 3,561.27 | 3,560.96 | 3,543.85 | 3,826.41 | 14,492.49 |
| 619 | 1,262.67 | 1,238.77 | 1,282.65 | 1,268.18 | 5,052.27 |
| 620 | 3,461.84 | 3,564.86 | 3,437.65 | 3,531.51 | 13,995.86 |
| 622 | 5,097.63 | 5,064.92 | 4,944.90 | 5,114.09 | 20,221.54 |
| 623 | 1,756.76 | 1,874.36 | 1,813.42 | 1,862.31 | 7,306.85 |
| 624 | 1,176.43 | 1,161.07 | 1,186.21 | 1,202.62 | 4,726.33 |
| 625 | 1,732.50 | 1,702.88 | 1,655.42 | 1,767 | 6,857.80 |
| 626 | 1,515.22 | 1,494.16 | 1,576.53 | 1,520.23 | 6,106.14 |

| | | | | | | |
|---|---|---|---|---|---|---|
| | 627 | 2,225.48 | 2,256.93 | 2,302.10 | 2,395.27 | 9,179.78 |
| | 628 | 2,335.58 | 2,410.09 | 2,255.89 | 2,367.33 | 9,368.89 |
| | 629 | 3,735.12 | 3,694.57 | 3,680.29 | 3,852.74 | 14,962.72 |
| TOTAL | | 171,041.75 | 179,991.10 | 181,512.60 | 185,495.20 | 718,040.65 |

STATE: INDIANA

| ZIP CODE | QUARTER 1 | QUARTER 2 | QUARTER 3 | QUARTER 4 | TOTAL GRAMS |
|---|---|---|---|---|---|
| 460 | 13,184.90 | 13,473.94 | 13,613.59 | 13,876.08 | 54,148.51 |
| 461 | 7,461.42 | 7,853.50 | 7,925.73 | 8,342.26 | 31,582.91 |
| 462 | 14,870.34 | 15,457 | 15,272.02 | 15,897.36 | 61,496.72 |
| 463 | 10,432.22 | 10,968.06 | 11,387.32 | 11,638.32 | 44,425.92 |
| 464 | 1,039.25 | 1,046.87 | 1,184.59 | 1,288.80 | 4,559.51 |
| 465 | 16,160.02 | 16,419.02 | 16,589.38 | 17,080.87 | 66,249.29 |
| 466 | 4,126.58 | 4,308.06 | 4,428.37 | 4,485.63 | 17,348.64 |
| 467 | 3,576.64 | 3,659.79 | 3,644.69 | 3,597.09 | 14,478.21 |
| 468 | 5,276.53 | 5,392.21 | 5,452.34 | 5,601.66 | 21,722.74 |
| 469 | 4,899 | 5,072.14 | 5,107.29 | 5,131.22 | 20,209.65 |
| 470 | 1,360.51 | 1,417.43 | 1,559.55 | 1,466.70 | 5,804.19 |
| 471 | 6,792.75 | 6,782.49 | 6,615.33 | 6,595.77 | 26,786.34 |
| 472 | 2,055.02 | 2,133.45 | 2,095.69 | 2,307.52 | 8,591.68 |
| 473 | 4,574.41 | 4,678.23 | 4,523.74 | 4,979.35 | 18,755.73 |
| 474 | 3,811.74 | 3,907.46 | 3,907.80 | 3,907.34 | 15,534.34 |
| 475 | 2,107.32 | 2,225.06 | 2,235.46 | 2,368.37 | 8,936.21 |
| 476 | 2,523.44 | 2,522.03 | 2,507.19 | 2,449.51 | 10,002.17 |
| 477 | 5,347.01 | 5,404.33 | 5,594.47 | 5,497.15 | 21,842.96 |
| 478 | 2,253.39 | 2,170.26 | 2,153.82 | 2,291.72 | 8,869.19 |
| 479 | 6,474.32 | 6,566.38 | 6,451.13 | 6,621.67 | 26,113.50 |
| TOTAL | 118,326.81 | 121,457.71 | 122,249.50 | 125,424.39 | 487,458.41 |

STATE: IOWA

| ZIP CODE | QUARTER 1 | QUARTER 2 | QUARTER 3 | QUARTER 4 | TOTAL GRAMS |
|---|---|---|---|---|---|
| 500 | 3,466.10 | 3,524.66 | 3,732.62 | 3,871.74 | 14,595.12 |
| 501 | 1,590.06 | 1,680.80 | 1,612.61 | 1,663.01 | 6,546.48 |
| 502 | 2,717.24 | 2,763.27 | 2,799.57 | 2,857.58 | 11,137.66 |
| 503 | 4,918.81 | 5,188.97 | 5,437.52 | 5,352.87 | 20,898.17 |
| 504 | 873.49 | 824.95 | 792.73 | 837.76 | 3,328.93 |
| 505 | 1,128.93 | 1,049.89 | 1,088.68 | 1,157.07 | 4,424.57 |
| 506 | 1,518.90 | 1,384.78 | 1,360.68 | 1,438.78 | 5,703.14 |
| 507 | 1,065.25 | 1,095.45 | 1,126.72 | 1,203.74 | 4,491.16 |
| 508 | 171.39 | 169.61 | 175.77 | 170.71 | 687.48 |
| 510 | 946.74 | 928.43 | 901.6 | 864.86 | 3,641.63 |
| 511 | 2,573.39 | 2,637.69 | 2,597.57 | 2,663.63 | 10,472.28 |
| 512 | 454.95 | 393.43 | 367.33 | 416.9 | 1,632.61 |
| 513 | 631.85 | 656.79 | 693.93 | 656.8 | 2,639.37 |
| 514 | 372.62 | 396.47 | 383.79 | 356.25 | 1,509.13 |
| 515 | 1,839.62 | 1,870.17 | 1,868.42 | 1,953.11 | 7,531.32 |
| 516 | 189.88 | 204.16 | 172.05 | 203.67 | 769.76 |
| 520 | 2,852.65 | 2,823.95 | 2,867.44 | 2,984.96 | 11,529 |
| 521 | 475.02 | 452.77 | 399.97 | 449.94 | 1,777.70 |
| 522 | 4,461.80 | 4,656.72 | 4,750.68 | 4,746.93 | 18,616.13 |
| 523 | 2,094.16 | 2,116.51 | 2,063.73 | 2,278.50 | 8,552.90 |
| 524 | 3,714.92 | 3,763.11 | 3,825.37 | 3,827.20 | 15,130.60 |
| 525 | 958.3 | 930.31 | 942.21 | 889.41 | 3,720.23 |
| 526 | 1,489.96 | 1,563.79 | 1,681.94 | 1,588.24 | 6,323.93 |
| 527 | 3,083.07 | 3,130.03 | 3,053.17 | 3,088.37 | 12,354.64 |
| 528 | 2,260.09 | 2,379.28 | 2,385.64 | 2,443.54 | 9,468.55 |
| TOTAL | 45,849.19 | 46,585.99 | 47,081.74 | 47,965.57 | 187,482.49 |

STATE: KANSAS

RETAIL DRUG DISTRIBUTION BY ZIP CODE WITHIN STATE BY GRAMS WT

STATE:KANSAS

| ZIP CODE | QUARTER 1 | QUARTER 2 | QUARTER 3 | QUARTER 4 | TOTAL GRAMS |
|---|---|---|---|---|---|
| 660 | 12,982.99 | 13,092.93 | 13,425.48 | 13,866.67 | 53,368.07 |
| 661 | 1,647.50 | 1,691.05 | 1,635.45 | 1,732.46 | 6,706.46 |
| 662 | 16,182.04 | 16,857.88 | 17,673.70 | 17,513.34 | 68,226.96 |
| 664 | 1,131.15 | 1,077.10 | 1,056.34 | 1,109.76 | 4,374.35 |
| 665 | 1,476.44 | 1,412.06 | 1,299.73 | 1,449.14 | 5,637.37 |
| 666 | 4,527.90 | 4,863.52 | 4,873.02 | 4,813.63 | 19,078.07 |
| 667 | 1,765.32 | 1,707.41 | 1,653.24 | 1,737 | 6,862.97 |
| 668 | 561.42 | 595.87 | 593.79 | 634.99 | 2,386.07 |
| 669 | 123.7 | 108.8 | 113.98 | 82.39 | 428.87 |
| 670 | 2,113.10 | 2,201.79 | 2,220.54 | 2,315.18 | 8,850.61 |
| 671 | 840.52 | 796.24 | 871.17 | 886.82 | 3,394.75 |
| 672 | 6,928.08 | 7,380.37 | 7,315.34 | 7,395.78 | 29,019.57 |
| 673 | 768.83 | 779.62 | 741.67 | 758.78 | 3,048.90 |
| 674 | 1,182.02 | 1,188.77 | 1,206.23 | 1,228.82 | 4,805.84 |
| 675 | 977.34 | 984.72 | 1,015.07 | 959.66 | 3,936.79 |
| 676 | 727.74 | 495.77 | 610.82 | 598.35 | 2,432.68 |
| 677 | 298.31 | 314.24 | 332.24 | 330.77 | 1,275.56 |
| 678 | 608.77 | 635.21 | 609.77 | 669.37 | 2,523.12 |
| 679 | 220.13 | 197.82 | 236.48 | 207.68 | 862.11 |
| TOTAL | 55,063.30 | 56,381.17 | 57,484.06 | 58,290.59 | 227,219.12 |

STATE: KENTUCKY

| ZIP CODE | QUARTER 1 | QUARTER 2 | QUARTER 3 | QUARTER 4 | TOTAL GRAMS |
|---|---|---|---|---|---|
| 400 | 3,362.05 | 3,449.85 | 3,620.77 | 3,543.21 | 13,975.88 |
| 401 | 1,557.50 | 1,600.37 | 1,814.37 | 1,367.28 | 6,339.52 |
| 402 | 15,203.65 | 15,575.39 | 15,356.07 | 16,036.50 | 62,171.61 |
| 403 | 3,561.10 | 3,594.94 | 3,638.56 | 3,892.26 | 14,686.86 |
| 404 | 1,479.40 | 1,385.69 | 1,403.41 | 1,599.08 | 5,867.58 |
| 405 | 5,924.18 | 6,095.12 | 6,215.51 | 6,553.77 | 24,788.58 |
| 406 | 649.1 | 647.89 | 702.4 | 747.2 | 2,746.59 |
| 407 | 476.2 | 514.43 | 504.55 | 543.05 | 2,038.23 |
| 408 | 166.3 | 210.39 | 159.37 | 205.85 | 741.91 |
| 409 | 173.91 | 158.08 | 169.81 | 173.36 | 675.16 |
| 410 | 8,773.62 | 8,914.97 | 8,880.46 | 9,275.85 | 35,844.90 |
| 411 | 1,370.19 | 1,371.38 | 1,383 | 1,409.80 | 5,534.37 |
| 412 | 386.49 | 340.56 | 382.53 | 329.92 | 1,439.50 |
| 413 | 124.72 | 136.08 | 125.4 | 147.61 | 533.81 |
| 414 | 146.35 | 128.26 | 132.21 | 142.35 | 549.17 |
| 415 | 415.66 | 416.89 | 404.35 | 465.15 | 1,702.05 |
| 416 | 283.4 | 292.51 | 297.49 | 300.07 | 1,173.47 |
| 417 | 285.56 | 281.51 | 323.17 | 303.67 | 1,193.91 |
| 418 | 111.43 | 112.3 | 108.72 | 118.29 | 450.74 |
| 420 | 4,036.62 | 4,013.69 | 4,063.17 | 4,155.25 | 16,268.73 |
| 421 | 2,668.89 | 2,726.82 | 2,635.38 | 2,786.70 | 10,817.79 |
| 422 | 2,020.90 | 2,071.69 | 2,128.72 | 2,088.28 | 8,309.59 |
| 423 | 3,245.60 | 3,486.11 | 3,480.65 | 3,543.72 | 13,756.08 |
| 424 | 3,342.11 | 3,327.06 | 3,280.46 | 3,366.40 | 13,316.03 |
| 425 | 400.83 | 405.19 | 396.8 | 375.26 | 1,578.08 |
| 426 | 265.39 | 235.68 | 245.61 | 238.43 | 985.11 |
| 427 | 2,232.01 | 2,273.52 | 2,234 | 2,293.90 | 9,033.43 |
| TOTAL | 62,663.16 | 63,766.37 | 64,086.94 | 66,002.21 | 256,518.68 |

STATE: LOUISIANA

| ZIP CODE | QUARTER 1 | QUARTER 2 | QUARTER 3 | QUARTER 4 | TOTAL GRAMS |
|---|---|---|---|---|---|
| 700 | 17,813.21 | 17,879.28 | 18,576.31 | 18,719.76 | 72,988.56 |
| 701 | 8,728.82 | 9,128.15 | 9,182.17 | 9,516.25 | 36,555.39 |
| 703 | 5,541.89 | 5,447.58 | 5,793.29 | 5,793.48 | 22,576.24 |

| | | | | | |
|---|---|---|---|---|---|
| 704 | 16,480.67 | 16,274.57 | 17,458.80 | 17,743.01 | 67,957.05 |
| 705 | 28,576.28 | 29,161.39 | 29,967.63 | 30,873.99 | 118,579.29 |
| 706 | 9,138.08 | 9,484.43 | 10,299.85 | 10,581.49 | 39,503.85 |
| 707 | 17,337.17 | 17,567.24 | 17,526.92 | 17,884.70 | 70,316.03 |
| 708 | 14,452.65 | 15,013.37 | 15,006.12 | 15,650.17 | 60,122.31 |
| 710 | 1,767.28 | 1,704 | 1,630.62 | 1,886.99 | 6,988.89 |
| 711 | 6,408.71 | 6,361.28 | 6,743.10 | 6,834.83 | 26,347.92 |
| 712 | 6,054.22 | 6,028.44 | 6,262.82 | 6,393.73 | 24,739.21 |
| 713 | 4,899.14 | 4,826.81 | 5,016.39 | 5,247.86 | 19,990.20 |
| 714 | 1,774.17 | 1,709.78 | 1,966.18 | 1,875.66 | 7,325.79 |
| TOTAL | 138,972.29 | 140,586.32 | 145,430.20 | 149,001.92 | 573,990.73 |

DRUG CODE:1100DRUG NAME:AMPHETAMINE
STATE: MAINE

| ZIP CODE | QUARTER 1 | QUARTER 2 | QUARTER 3 | QUARTER 4 | TOTAL GRAMS |
|---|---|---|---|---|---|
| 39 | 829.44 | 876.6 | 814.33 | 810.63 | 3,331 |
| 40 | 6,977.04 | 7,294.93 | 7,348.87 | 7,425.57 | 29,046.41 |
| 41 | 3,716.48 | 3,816.33 | 3,803.91 | 3,842.81 | 15,179.53 |
| 42 | 1,779.05 | 1,778.91 | 1,738 | 1,839.62 | 7,135.58 |
| 43 | 1,192.87 | 1,360.65 | 1,271.50 | 1,287.48 | 5,112.50 |
| 44 | 2,276.71 | 2,209.64 | 2,273.34 | 2,251.19 | 9,010.88 |
| 45 | 624.52 | 638.47 | 672.1 | 611.97 | 2,547.06 |
| 46 | 747.72 | 706.48 | 822.15 | 730.02 | 3,006.37 |
| 47 | 482.21 | 503.45 | 511.14 | 483.53 | 1,980.33 |
| 48 | 480.87 | 557.33 | 572 | 532.4 | 2,142.60 |
| 49 | 1,717.96 | 1,695.11 | 1,714.61 | 1,811.62 | 6,939.30 |
| TOTAL | 20,824.87 | 21,437.90 | 21,541.95 | 21,626.84 | 85,431.56 |

STATE: MARYLAND

| ZIP CODE | QUARTER 1 | QUARTER 2 | QUARTER 3 | QUARTER 4 | TOTAL GRAMS |
|---|---|---|---|---|---|
| 206 | 3,923.99 | 4,034.90 | 3,962.35 | 4,210.56 | 16,131.80 |
| 207 | 5,337.19 | 5,629.02 | 5,748.95 | 5,630.44 | 22,345.60 |
| 208 | 13,835.27 | 14,512.49 | 14,597.59 | 14,271.78 | 57,217.13 |
| 209 | 2,464.67 | 2,570.59 | 2,664.50 | 2,709.95 | 10,409.71 |
| 210 | 16,140.42 | 16,776.37 | 16,461.39 | 16,348.05 | 65,726.23 |
| 211 | 7,360.20 | 7,651.91 | 7,828.58 | 7,994.68 | 30,835.37 |
| 212 | 14,508.18 | 15,152.80 | 15,639.25 | 15,795.14 | 61,095.37 |
| 214 | 2,379.40 | 2,450.46 | 2,327.18 | 2,396.81 | 9,553.85 |
| 215 | 1,545.59 | 1,575.27 | 1,659.99 | 1,663.13 | 6,443.98 |
| 216 | 2,754.16 | 2,838.88 | 2,929.96 | 2,944.40 | 11,467.40 |
| 217 | 9,321.68 | 9,758.86 | 9,893.90 | 9,933.69 | 38,908.13 |
| 218 | 1,787 | 2,003.96 | 2,075.64 | 1,976.73 | 7,843.33 |
| 219 | 2,401.91 | 2,461.60 | 2,399.23 | 2,458.19 | 9,720.93 |
| TOTAL | 83,759.66 | 87,417.11 | 88,188.51 | 88,333.55 | 347,698.83 |

STATE: MASSACHUSETTS

| ZIP CODE | QUARTER 1 | QUARTER 2 | QUARTER 3 | QUARTER 4 | TOTAL GRAMS |
|---|---|---|---|---|---|
| 10 | 8,634.13 | 8,726.07 | 8,424.66 | 8,982.65 | 34,767.51 |
| 11 | 2,149.96 | 2,204.50 | 2,202.85 | 2,074.27 | 8,631.58 |
| 12 | 2,266.06 | 2,363.84 | 2,316.93 | 2,376.91 | 9,323.74 |
| 13 | 2,477.14 | 2,481.11 | 2,368.79 | 2,474.62 | 9,801.66 |
| 14 | 5,074.76 | 5,175.65 | 4,984.39 | 5,061.77 | 20,296.57 |
| 15 | 6,633.28 | 7,017.31 | 6,854.49 | 7,204.51 | 27,709.59 |
| 16 | 5,325.67 | 5,628.13 | 5,850.67 | 5,771.73 | 22,576.20 |
| 17 | 10,773.70 | 11,341.89 | 11,133.54 | 11,132.15 | 44,381.28 |
| 18 | 18,173.66 | 18,697.26 | 19,276.18 | 19,369.03 | 75,516.13 |
| 19 | 16,938.50 | 17,643.96 | 18,273.67 | 17,681.88 | 70,538.01 |
| 20 | 12,616.54 | 13,038.22 | 13,332.02 | 13,272 | 52,258.78 |
| 21 | 31,012.03 | 33,023.25 | 32,924.17 | 32,561.59 | 129,521.04 |

| | | | | | |
|---|---|---|---|---|---|
| 22 | 1,358.83 | 1,357.93 | 1,335.78 | 1,586.54 | 5,639.08 |
| 23 | 12,381.46 | 12,691.34 | 12,806.63 | 12,948.16 | 50,827.59 |
| 24 | 10,859.01 | 11,241.87 | 11,024.33 | 11,444.52 | 44,569.73 |
| 25 | 3,640.07 | 3,818.92 | 4,145.05 | 4,024.58 | 15,628.62 |
| 26 | 4,586.32 | 4,555.81 | 5,028.27 | 4,576.82 | 18,747.22 |
| 27 | 11,056.36 | 11,455.96 | 11,597.88 | 11,767.49 | 45,877.69 |
| TOTAL | 165,957.48 | 172,463.02 | 173,880.30 | 174,311.22 | 686,612.02 |

STATE: MICHIGAN

| ZIP CODE | QUARTER 1 | QUARTER 2 | QUARTER 3 | QUARTER 4 | TOTAL GRAMS |
|---|---|---|---|---|---|
| 480 | 36,844.86 | 37,835.95 | 37,263.67 | 38,452.24 | 150,396.72 |
| 481 | 37,499.61 | 38,614.18 | 37,025.56 | 39,440.72 | 152,580.07 |
| 482 | 6,542.46 | 6,664.84 | 6,608.92 | 6,781.94 | 26,598.16 |
| 483 | 30,372.85 | 31,636.26 | 30,454.32 | 31,737.80 | 124,201.23 |
| 484 | 7,926.25 | 8,042.57 | 7,831.54 | 8,433.15 | 32,233.51 |
| 485 | 2,608.70 | 2,622.14 | 2,565.68 | 2,907.32 | 10,703.84 |
| 486 | 4,603.74 | 4,848.23 | 4,576.18 | 4,926.77 | 18,954.92 |
| 487 | 2,945.34 | 2,935.85 | 2,905.63 | 3,066.41 | 11,853.23 |
| 488 | 14,442.61 | 14,128.88 | 12,160.82 | 14,707.53 | 55,439.84 |
| 489 | 3,863.28 | 4,092.88 | 3,865.63 | 4,243.97 | 16,065.76 |
| 490 | 17,462.77 | 17,590.76 | 16,373.78 | 17,929.69 | 69,357 |
| 491 | 2,141.58 | 2,120 | 2,021.39 | 2,188.75 | 8,471.72 |
| 492 | 3,896.99 | 3,978.48 | 3,683.91 | 4,045.02 | 15,604.40 |
| 493 | 6,927.15 | 6,822.81 | 6,088.95 | 7,351.77 | 27,190.68 |
| 494 | 17,930.86 | 18,611.63 | 17,159 | 18,979.86 | 72,681.35 |

DRUG CODE:1100DRUG NAME:AMPHETAMINE
STATE:MICHIGAN

| ZIP CODE | QUARTER 1 | QUARTER 2 | QUARTER 3 | QUARTER 4 | TOTAL GRAMS |
|---|---|---|---|---|---|
| 495 | 18,470.41 | 18,253.66 | 17,079.43 | 19,185.81 | 72,989.31 |
| 496 | 4,849.03 | 5,317.50 | 5,131.73 | 5,269.07 | 20,567.33 |
| 497 | 2,960.13 | 3,120.16 | 3,192.69 | 3,200.53 | 12,473.51 |
| 498 | 3,171.34 | 3,153.33 | 3,328.68 | 3,240.10 | 12,893.45 |
| 499 | 1,558.04 | 1,549.91 | 1,658.62 | 1,702.45 | 6,469.02 |
| TOTAL | 227,018 | 231,940.02 | 220,976.13 | 237,790.90 | 917,725.05 |

STATE: MINNESOTA

| ZIP CODE | QUARTER 1 | QUARTER 2 | QUARTER 3 | QUARTER 4 | TOTAL GRAMS |
|---|---|---|---|---|---|
| 550 | 11,915.74 | 12,352.58 | 12,094.72 | 12,344.29 | 48,707.33 |
| 551 | 17,353.11 | 17,870.46 | 17,915.99 | 17,637.66 | 70,777.22 |
| 553 | 21,274.23 | 21,549.35 | 22,083.91 | 22,353.95 | 87,261.44 |
| 554 | 24,147.44 | 25,126.95 | 25,934.83 | 26,023.28 | 101,232.50 |
| 556 | 127.28 | 114.96 | 113.28 | 101.37 | 456.89 |
| 557 | 3,100.16 | 3,021.98 | 3,243.17 | 2,745.97 | 12,111.28 |
| 558 | 1,999.66 | 2,009.57 | 1,983.16 | 2,197.50 | 8,189.89 |
| 559 | 5,297.70 | 5,187.68 | 5,075.67 | 5,147.40 | 20,708.45 |
| 560 | 4,988.99 | 5,153.40 | 5,158.37 | 5,148.28 | 20,449.04 |
| 561 | 699.45 | 731.64 | 744.5 | 753.68 | 2,929.27 |
| 562 | 1,621.33 | 1,618.57 | 1,594.65 | 1,643.43 | 6,477.98 |
| 563 | 4,105.45 | 4,212.15 | 4,172.84 | 4,120.11 | 16,610.55 |
| 564 | 2,113.16 | 2,218.22 | 2,272.03 | 2,113.97 | 8,717.38 |
| 565 | 2,602.87 | 2,580.49 | 2,517.87 | 2,567.32 | 10,268.55 |
| 566 | 1,209.33 | 1,176.49 | 1,099.35 | 1,177.61 | 4,662.78 |
| 567 | 660.1 | 663.72 | 619.58 | 645.01 | 2,588.41 |
| TOTAL | 103,216 | 105,588.21 | 106,623.92 | 106,720.83 | 422,148.96 |

STATE: MISSISSIPPI

| ZIP CODE | QUARTER 1 | QUARTER 2 | QUARTER 3 | QUARTER 4 | TOTAL GRAMS |
|---|---|---|---|---|---|
| 386 | 8,367.89 | 8,580.20 | 8,707.48 | 8,923.18 | 34,578.75 |
| 387 | 1,399.99 | 1,573.12 | 1,513.62 | 1,589.27 | 6,076 |

| | | | | | |
|---|---|---|---|---|---|
| 388 | 4,785.82 | 5,040.98 | 5,075.60 | 5,003 | 19,905.40 |
| 389 | 1,392.41 | 1,382.49 | 1,446.99 | 1,500.19 | 5,722.08 |
| 390 | 4,875.98 | 4,622.91 | 4,857.09 | 4,876.99 | 19,232.97 |
| 391 | 5,881.76 | 5,885.22 | 6,187.30 | 5,966.92 | 23,921.20 |
| 392 | 5,010.33 | 4,947.05 | 5,088.90 | 5,348.78 | 20,395.06 |
| 393 | 4,152.73 | 3,974.60 | 4,190.11 | 4,172.49 | 16,489.93 |
| 394 | 8,126.72 | 8,370.92 | 8,593.82 | 8,829.67 | 33,921.13 |
| 395 | 8,500.01 | 8,171.03 | 8,714.60 | 8,612.69 | 33,998.33 |
| 396 | 2,335.80 | 2,310.85 | 2,498.62 | 2,397.98 | 9,543.25 |
| 397 | 3,870.73 | 3,748.06 | 3,889.82 | 3,666.20 | 15,174.81 |
| TOTAL | 58,700.17 | 58,607.43 | 60,763.95 | 60,887.36 | 238,958.91 |

STATE: MISSOURI

| ZIP CODE | QUARTER 1 | QUARTER 2 | QUARTER 3 | QUARTER 4 | TOTAL GRAMS |
|---|---|---|---|---|---|
| 630 | 18,305.31 | 18,640.66 | 18,890.19 | 19,222.42 | 75,058.58 |
| 631 | 37,218.65 | 39,078.03 | 39,496.37 | 36,556.60 | 152,349.65 |
| 633 | 12,681.41 | 12,941.53 | 13,517.06 | 13,373.69 | 52,513.69 |
| 634 | 1,217.35 | 1,192.23 | 1,121.27 | 1,149.40 | 4,680.25 |
| 635 | 1,226.73 | 1,191.24 | 1,151.11 | 1,165.53 | 4,734.61 |
| 636 | 1,998.53 | 2,008.22 | 1,853.22 | 1,977.50 | 7,837.47 |
| 637 | 2,206.73 | 2,249.33 | 2,157.38 | 2,238.40 | 8,851.84 |
| 638 | 1,458.15 | 1,487.90 | 1,515.11 | 1,598.83 | 6,059.99 |
| 639 | 494.73 | 425.59 | 490.71 | 508.75 | 1,919.78 |
| 640 | 12,267.11 | 12,710.15 | 13,231.84 | 13,469.99 | 51,679.09 |
| 641 | 15,707.90 | 16,481.41 | 16,773.67 | 17,186.07 | 66,149.05 |
| 644 | 690.29 | 664.71 | 672.63 | 736.11 | 2,763.74 |
| 645 | 1,109.50 | 1,095.24 | 1,089.75 | 1,117.55 | 4,412.04 |
| 646 | 394.96 | 424.45 | 384.78 | 401.45 | 1,605.64 |
| 647 | 941.31 | 895.44 | 937.09 | 943.32 | 3,717.16 |
| 648 | 1,570.55 | 1,559.34 | 1,585.21 | 1,585.93 | 6,301.03 |
| 650 | 1,022.05 | 1,045.05 | 1,156.29 | 1,023.63 | 4,247.02 |
| 651 | 1,226.88 | 1,145.88 | 1,158.52 | 1,201.25 | 4,732.53 |
| 652 | 5,235.06 | 5,012.97 | 4,986.52 | 5,289.81 | 20,524.36 |
| 653 | 974.82 | 910.9 | 957.06 | 960.3 | 3,803.08 |
| 654 | 839.56 | 811.91 | 871.22 | 844.42 | 3,367.11 |
| 655 | 609.93 | 624.05 | 631.44 | 719.67 | 2,585.09 |
| 656 | 1,266.53 | 1,311.92 | 1,320.98 | 1,336.84 | 5,236.27 |
| 657 | 1,615.50 | 1,715.02 | 1,711.46 | 1,730.92 | 6,772.90 |

DRUG CODE:1100DRUG NAME:AMPHETAMINE
STATE:MISSOURI

| ZIP CODE | QUARTER 1 | QUARTER 2 | QUARTER 3 | QUARTER 4 | TOTAL GRAMS |
|---|---|---|---|---|---|
| 658 | 3,066.82 | 3,188.07 | 3,332.71 | 3,417.34 | 13,004.94 |
| TOTAL | 125,346.36 | 128,811.24 | 130,993.59 | 129,755.72 | 514,906.91 |

STATE: MONTANA

| ZIP CODE | QUARTER 1 | QUARTER 2 | QUARTER 3 | QUARTER 4 | TOTAL GRAMS |
|---|---|---|---|---|---|
| 590 | 483.29 | 562.14 | 502.01 | 533.78 | 2,081.22 |
| 591 | 2,161.69 | 2,210.98 | 2,233.17 | 2,351.67 | 8,957.51 |
| 592 | 329.15 | 348.13 | 323.37 | 344.76 | 1,345.41 |
| 593 | 302.25 | 313.99 | 332.69 | 293.38 | 1,242.31 |
| 594 | 1,111.72 | 1,185.14 | 1,104.48 | 1,062.51 | 4,463.85 |
| 595 | 189.59 | 186.04 | 172.21 | 211.94 | 759.78 |
| 596 | 946.41 | 1,128.13 | 1,019.46 | 1,021.25 | 4,115.25 |
| 597 | 2,701.78 | 2,810.67 | 2,832.04 | 2,915.21 | 11,259.70 |
| 598 | 3,244.46 | 3,518.08 | 3,375.44 | 3,500.90 | 13,638.88 |
| 599 | 928.97 | 1,036.09 | 1,128.60 | 1,059.13 | 4,152.79 |
| TOTAL | 12,399.31 | 13,299.39 | 13,023.47 | 13,294.53 | 52,016.70 |

STATE: NEBRASKA

RETAIL DRUG DISTRIBUTION BY ZIP CODE WITHIN STATE BY GRAMS WT

| ZIP CODE | QUARTER 1 | QUARTER 2 | QUARTER 3 | QUARTER 4 | TOTAL GRAMS |
|---|---|---|---|---|---|
| 680 | 2,082.43 | 2,101.40 | 2,165.06 | 2,228.64 | 8,577.53 |
| 681 | 11,002.91 | 11,439.08 | 11,700.29 | 11,653.27 | 45,795.55 |
| 683 | 548.05 | 500.84 | 487.5 | 473.8 | 2,010.19 |
| 684 | 409.12 | 377.64 | 373.79 | 390.97 | 1,551.52 |
| 685 | 4,979.82 | 5,009.51 | 5,072.83 | 5,258.30 | 20,320.46 |
| 686 | 462.44 | 421.37 | 418.99 | 446.65 | 1,749.45 |
| 687 | 1,022.74 | 997.58 | 980.56 | 1,016.60 | 4,017.48 |
| 688 | 1,398.27 | 1,422.43 | 1,456.85 | 1,437.95 | 5,715.50 |
| 689 | 486.86 | 443.42 | 459.01 | 470.6 | 1,859.89 |
| 690 | 188.42 | 195.3 | 200.97 | 198.37 | 783.06 |
| 691 | 581.5 | 576.79 | 671.98 | 634.06 | 2,464.33 |
| 692 | 16.3 | 16.58 | 35.66 | 49.57 | 118.11 |
| 693 | 467.64 | 493.47 | 513.57 | 469.77 | 1,944.45 |
| TOTAL | 23,646.50 | 23,995.41 | 24,537.06 | 24,728.55 | 96,907.52 |

STATE: NEVADA

| ZIP CODE | QUARTER 1 | QUARTER 2 | QUARTER 3 | QUARTER 4 | TOTAL GRAMS |
|---|---|---|---|---|---|
| 890 | 5,064.91 | 5,249.98 | 5,246.79 | 5,485.92 | 21,047.60 |
| 891 | 12,962.26 | 13,944.85 | 13,819.28 | 14,380.35 | 55,106.74 |
| 893 | 44.17 | 63.21 | 56.46 | 53.45 | 217.29 |
| 894 | 1,437.22 | 1,547.18 | 1,533.43 | 1,560.02 | 6,077.85 |
| 895 | 2,416.24 | 2,652.01 | 2,611.10 | 2,599.22 | 10,278.57 |
| 897 | 410.59 | 444.04 | 431.03 | 471.86 | 1,757.52 |
| 898 | 385.08 | 421.64 | 404.09 | 449.01 | 1,659.82 |
| TOTAL | 22,720.47 | 24,322.91 | 24,102.18 | 24,999.83 | 96,145.39 |

STATE: NEW HAMPSHIRE

| ZIP CODE | QUARTER 1 | QUARTER 2 | QUARTER 3 | QUARTER 4 | TOTAL GRAMS |
|---|---|---|---|---|---|
| 30 | 10,523.56 | 10,889.03 | 11,149.78 | 11,434.20 | 43,996.57 |
| 31 | 5,550.86 | 5,836.43 | 5,798.59 | 5,769.58 | 22,955.46 |
| 32 | 2,511.36 | 2,674.02 | 2,744.36 | 2,576.04 | 10,505.78 |
| 33 | 1,758.79 | 1,814.48 | 1,934.93 | 1,916.65 | 7,424.85 |
| 34 | 1,339.51 | 1,485.60 | 1,385.86 | 1,460.69 | 5,671.66 |
| 35 | 422.62 | 458.47 | 495.08 | 497.35 | 1,873.52 |
| 36 | 86.58 | 88.53 | 87.39 | 88.25 | 350.75 |
| 37 | 1,676.24 | 1,669 | 1,611.63 | 1,681.15 | 6,638.02 |
| 38 | 10,664.59 | 11,100.52 | 11,425.42 | 11,342.30 | 44,532.83 |
| TOTAL | 34,534.11 | 36,016.08 | 36,633.04 | 36,766.21 | 143,949.44 |

STATE: NEW JERSEY

| ZIP CODE | QUARTER 1 | QUARTER 2 | QUARTER 3 | QUARTER 4 | TOTAL GRAMS |
|---|---|---|---|---|---|
| 70 | 14,662.04 | 15,289.67 | 15,228.46 | 15,825.38 | 61,005.55 |
| 71 | 1,211.52 | 1,345.77 | 1,241.88 | 1,358.80 | 5,157.97 |
| 72 | 529.5 | 522.02 | 491.15 | 548.98 | 2,091.65 |
| 73 | 1,213.83 | 1,365.32 | 1,340.45 | 1,429.72 | 5,349.32 |
| 74 | 5,593.40 | 5,638.35 | 5,781.31 | 5,857.67 | 22,870.73 |
| 75 | 573.81 | 614.19 | 524.25 | 593.36 | 2,305.61 |
| 76 | 5,011.03 | 4,976.54 | 4,930.03 | 5,201.92 | 20,119.52 |
| 77 | 7,294.96 | 7,525.31 | 7,705.19 | 7,881.05 | 30,406.51 |
| 78 | 5,038.30 | 5,218.12 | 5,413 | 5,288.29 | 20,957.71 |
| 79 | 4,359.68 | 4,562.34 | 4,619.26 | 4,726.66 | 18,267.94 |
| 80 | 22,189.27 | 23,615.80 | 23,163.65 | 23,304.68 | 92,273.40 |

DRUG CODE:1100DRUG NAME:AMPHETAMINE
STATE:NEW JERSEY

| ZIP CODE | QUARTER 1 | QUARTER 2 | QUARTER 3 | QUARTER 4 | TOTAL GRAMS |
|---|---|---|---|---|---|
| 81 | 1,365.86 | 1,393.22 | 1,442.72 | 1,660.37 | 5,862.17 |

RETAIL DRUG DISTRIBUTION BY ZIP CODE WITHIN STATE BY GRAMS WT

| | | | | | |
|---|---|---|---|---|---|
| 82 | 3,544.87 | 3,973.82 | 4,182.32 | 3,889.33 | 15,590.34 |
| 83 | 2,382.37 | 2,490.93 | 2,456.89 | 2,589.01 | 9,919.20 |
| 84 | 319.74 | 384.04 | 412.25 | 352.81 | 1,468.84 |
| 85 | 3,398.52 | 3,557.80 | 3,354.34 | 3,550.15 | 13,860.81 |
| 86 | 2,496.37 | 2,614.07 | 2,558.81 | 2,686.74 | 10,355.99 |
| 87 | 4,155.64 | 4,539.77 | 4,833.23 | 4,999.76 | 18,528.40 |
| 88 | 8,405.31 | 8,777.02 | 8,723.28 | 9,128.96 | 35,034.57 |
| 89 | 899.33 | 863.99 | 909.21 | 1,019.50 | 3,692.03 |
| TOTAL | 94,645.35 | 99,268.09 | 99,311.68 | 101,893.14 | 395,118.26 |

STATE: NEW MEXICO

| ZIP CODE | QUARTER 1 | QUARTER 2 | QUARTER 3 | QUARTER 4 | TOTAL GRAMS |
|---|---|---|---|---|---|
| 870 | 765.49 | 823.94 | 738.55 | 686.89 | 3,014.87 |
| 871 | 10,996.31 | 11,570.44 | 10,623.29 | 10,847.21 | 44,037.25 |
| 873 | 121.6 | 124.72 | 128.45 | 127.79 | 502.56 |
| 874 | 606.79 | 651.82 | 623.47 | 661.03 | 2,543.11 |
| 875 | 1,851.64 | 1,882.65 | 1,964.41 | 1,999.21 | 7,697.91 |
| 877 | 161.35 | 135.38 | 164.21 | 162.03 | 622.97 |
| 878 | 56.65 | 51.88 | 48.63 | 56.63 | 213.79 |
| 879 | 81.84 | 84.16 | 73.47 | 77.45 | 316.92 |
| 880 | 1,695.25 | 1,753.25 | 1,853.88 | 1,908.98 | 7,211.36 |
| 881 | 290.12 | 307.98 | 274.12 | 329.39 | 1,201.61 |
| 882 | 1,549.19 | 1,409.23 | 1,474.71 | 1,702.56 | 6,135.69 |
| 883 | 833.33 | 805.89 | 880 | 859.05 | 3,378.27 |
| 884 | 57.54 | 64.27 | 58.96 | 58.9 | 239.67 |
| TOTAL | 19,067.10 | 19,665.61 | 18,906.15 | 19,477.12 | 77,115.98 |

STATE: NEW YORK

| ZIP CODE | QUARTER 1 | QUARTER 2 | QUARTER 3 | QUARTER 4 | TOTAL GRAMS |
|---|---|---|---|---|---|
| 100 | 46,379.70 | 48,477.37 | 47,856.50 | 48,514.75 | 191,228.32 |
| 101 | 2,760.52 | 2,758.42 | 2,790.36 | 2,865.51 | 11,174.81 |
| 102 | 386.08 | 425.13 | 419.9 | 434.1 | 1,665.21 |
| 103 | 2,831.59 | 3,050.17 | 3,072.68 | 3,190.20 | 12,144.64 |
| 104 | 3,583.45 | 4,035.69 | 3,589.04 | 3,886.82 | 15,095 |
| 105 | 8,361.89 | 8,808.88 | 8,875.24 | 9,157.10 | 35,203.11 |
| 106 | 1,298.25 | 1,209.22 | 1,233.08 | 1,192.94 | 4,933.49 |
| 107 | 1,652.78 | 1,709.75 | 1,657.97 | 1,719.31 | 6,739.81 |
| 108 | 796.19 | 782.37 | 781.73 | 717.94 | 3,078.23 |
| 109 | 5,866.95 | 6,082.15 | 6,114.78 | 6,111.20 | 24,175.08 |
| 110 | 2,399.20 | 2,291.72 | 2,414.32 | 2,381.95 | 9,487.19 |
| 111 | 2,301.90 | 2,363.86 | 2,356.99 | 2,494.71 | 9,517.46 |
| 112 | 14,045.39 | 15,283.72 | 14,936.95 | 15,456.22 | 59,722.28 |
| 113 | 4,921.01 | 5,234.25 | 5,110.32 | 5,156.52 | 20,422.10 |
| 114 | 1,305.37 | 1,386.97 | 1,294.80 | 1,313.01 | 5,300.15 |
| 115 | 6,942.08 | 6,989.58 | 7,087.48 | 7,127.71 | 28,146.85 |
| 116 | 468.15 | 476.84 | 464.85 | 503.57 | 1,913.41 |
| 117 | 19,000.35 | 19,522.96 | 19,974.34 | 19,577.29 | 78,074.94 |
| 118 | 999.23 | 1,011.05 | 1,009.02 | 1,034.25 | 4,053.55 |
| 119 | 2,600.48 | 2,764.51 | 3,072.79 | 2,727.84 | 11,165.62 |
| 120 | 4,622.07 | 4,943.03 | 4,988.93 | 4,906.09 | 19,460.12 |
| 121 | 3,317.67 | 3,376.37 | 3,434.87 | 3,376.06 | 13,504.97 |
| 122 | 2,753.30 | 2,719.79 | 2,783.16 | 2,718.39 | 10,974.64 |
| 123 | 2,163.69 | 2,262.64 | 2,182.20 | 2,174.42 | 8,782.95 |
| 124 | 1,730.26 | 1,764.94 | 1,769.42 | 1,728.28 | 6,992.90 |
| 125 | 4,820.09 | 4,995.30 | 5,116.51 | 5,099.72 | 20,031.62 |
| 126 | 1,119.73 | 1,143.50 | 1,098.80 | 1,191.64 | 4,553.67 |
| 127 | 957.1 | 1,107.13 | 1,113.40 | 1,099.12 | 4,276.75 |
| 128 | 2,804.19 | 2,985.62 | 3,122.67 | 3,008.26 | 11,920.74 |
| 129 | 1,816.17 | 1,974.59 | 1,982.48 | 2,205.85 | 7,979.09 |
| 130 | 5,560.28 | 5,731.76 | 5,719.54 | 5,739.23 | 22,750.81 |
| 131 | 1,730.45 | 1,758.40 | 1,709.93 | 1,775.92 | 6,974.70 |
| 132 | 2,958.63 | 2,987.16 | 2,896.95 | 3,077.01 | 11,919.75 |

| | | | | | |
|---|---|---|---|---|---|
| 133 | 1,030.98 | 1,031.62 | 1,012.45 | 1,014.64 | 4,089.69 |
| 134 | 1,806.40 | 1,845.23 | 1,790.76 | 1,865.65 | 7,308.04 |
| 135 | 942.11 | 975.44 | 936 | 909.42 | 3,762.97 |
| 136 | 2,715.96 | 2,971.89 | 2,761.50 | 2,628.22 | 11,077.57 |
| 137 | 1,381.30 | 1,231.69 | 1,231.73 | 1,211.80 | 5,056.52 |

DRUG CODE:1100DRUG NAME:AMPHETAMINE
STATE:NEW YORK

| ZIP CODE | QUARTER 1 | QUARTER 2 | QUARTER 3 | QUARTER 4 | TOTAL GRAMS |
|---|---|---|---|---|---|
| 138 | 1,695.95 | 1,612.83 | 1,555.41 | 1,570.80 | 6,434.99 |
| 139 | 1,094.65 | 1,125.65 | 1,087.63 | 1,071.11 | 4,379.04 |
| 140 | 5,990.21 | 6,134.42 | 6,098.89 | 6,074.10 | 24,297.62 |
| 141 | 2,642.03 | 2,762.58 | 2,724 | 2,883.40 | 11,012.01 |
| 142 | 8,129.83 | 8,365.62 | 8,327.70 | 8,539.92 | 33,363.07 |
| 143 | 937.4 | 930.54 | 942.05 | 1,029.69 | 3,839.68 |
| 144 | 5,416.11 | 5,461.71 | 5,431.08 | 5,370.94 | 21,679.84 |
| 145 | 4,929.01 | 5,015.40 | 4,970.47 | 5,028.62 | 19,943.50 |
| 146 | 9,394.72 | 9,846.59 | 9,677.46 | 9,551.69 | 38,470.46 |
| 147 | 1,705.71 | 1,707.09 | 1,744.95 | 1,653.79 | 6,811.54 |
| 148 | 3,461.22 | 3,549.44 | 3,474.53 | 3,542.93 | 14,028.12 |
| 149 | 930.94 | 854.47 | 946.21 | 898.36 | 3,629.98 |
| TOTAL | 219,458.72 | 227,837.05 | 226,744.82 | 228,508.01 | 902,548.60 |

STATE: NORTH CAROLINA

| ZIP CODE | QUARTER 1 | QUARTER 2 | QUARTER 3 | QUARTER 4 | TOTAL GRAMS |
|---|---|---|---|---|---|
| 270 | 3,535.56 | 3,701.15 | 3,673.47 | 3,922.98 | 14,833.16 |
| 271 | 5,236.18 | 5,374.98 | 5,263.33 | 5,461.46 | 21,335.95 |
| 272 | 10,039.08 | 10,429.63 | 10,331.71 | 10,658.37 | 41,458.79 |
| 273 | 5,450.26 | 5,522.80 | 5,545.32 | 5,696.07 | 22,214.45 |
| 274 | 7,139.30 | 7,497.52 | 7,530.56 | 7,581.21 | 29,748.59 |
| 275 | 20,341.03 | 21,680.76 | 21,683.15 | 22,282.95 | 85,987.89 |
| 276 | 12,637.14 | 13,188.90 | 13,244.97 | 13,692.69 | 52,763.70 |
| 277 | 5,326.62 | 5,535.02 | 5,431.87 | 5,617.23 | 21,910.74 |
| 278 | 8,449.43 | 8,618.42 | 8,767.99 | 8,872.24 | 34,708.08 |
| 279 | 2,626.45 | 2,968.34 | 3,056.31 | 2,932.97 | 11,584.07 |
| 280 | 16,003.97 | 16,922.28 | 17,040.82 | 17,479.12 | 67,446.19 |
| 281 | 11,001.79 | 11,180.01 | 11,369.17 | 11,564.41 | 45,115.38 |
| 282 | 17,478.83 | 18,486.76 | 18,758.02 | 19,025.72 | 73,749.33 |
| 283 | 14,013.51 | 13,823.36 | 14,144.70 | 14,268.95 | 56,250.52 |
| 284 | 14,697.37 | 15,967.04 | 16,605.10 | 16,303.54 | 63,573.05 |
| 285 | 10,544.79 | 11,178.48 | 11,769.23 | 10,885.42 | 44,377.92 |
| 286 | 7,205.21 | 7,481.62 | 7,387.87 | 7,764.76 | 29,839.46 |
| 287 | 6,903.46 | 6,947.83 | 6,962.14 | 7,136.81 | 27,950.24 |
| 288 | 4,280.18 | 4,270.01 | 4,271.44 | 4,426.20 | 17,247.83 |
| 289 | 331.28 | 357.47 | 417.11 | 361.74 | 1,467.60 |
| TOTAL | 183,241.44 | 191,132.38 | 193,254.28 | 195,934.84 | 763,562.94 |

STATE: NORTH DAKOTA

| ZIP CODE | QUARTER 1 | QUARTER 2 | QUARTER 3 | QUARTER 4 | TOTAL GRAMS |
|---|---|---|---|---|---|
| 580 | 654.21 | 671.31 | 660.53 | 620.35 | 2,606.40 |
| 581 | 2,158.34 | 2,073.66 | 2,106.10 | 1,959.40 | 8,297.50 |
| 582 | 1,652.66 | 1,649.38 | 1,522.93 | 1,409.98 | 6,234.95 |
| 583 | 588.44 | 576.5 | 591.49 | 446.91 | 2,203.34 |
| 584 | 278.15 | 276.94 | 281.79 | 249.73 | 1,086.61 |
| 585 | 1,566.15 | 1,518 | 1,558.71 | 1,347.59 | 5,990.45 |
| 586 | 367.19 | 345.92 | 354.2 | 322.27 | 1,389.58 |
| 587 | 1,432.49 | 1,421.47 | 1,367.81 | 1,294.29 | 5,516.06 |
| 588 | 474.75 | 483.7 | 519.33 | 445.06 | 1,922.84 |
| TOTAL | 9,172.38 | 9,016.88 | 8,962.89 | 8,095.58 | 35,247.73 |

STATE: OHIO

| ZIP CODE | QUARTER 1 | QUARTER 2 | QUARTER 3 | QUARTER 4 | TOTAL GRAMS |
|---|---|---|---|---|---|
| 430 | 14,841.23 | 14,866.13 | 14,576.43 | 15,481.26 | 59,765.05 |
| 431 | 5,482.79 | 5,357.66 | 5,370.41 | 5,822.28 | 22,033.14 |
| 432 | 35,626.96 | 33,379.59 | 33,747.48 | 34,256.76 | 137,010.79 |
| 433 | 2,278.94 | 2,293.01 | 2,309.83 | 2,326.16 | 9,207.94 |
| 434 | 1,839.36 | 1,819.84 | 1,774.43 | 2,011.45 | 7,445.08 |
| 435 | 4,598.52 | 4,941.97 | 4,729.53 | 5,119.72 | 19,389.74 |
| 436 | 7,753.62 | 7,917.83 | 7,364.50 | 7,986.50 | 31,022.45 |
| 437 | 1,517.86 | 1,538.76 | 1,568.30 | 1,519.29 | 6,144.21 |
| 438 | 231.55 | 264.81 | 234.39 | 211.36 | 942.11 |
| 439 | 1,716.38 | 1,699.09 | 1,800.09 | 1,794.87 | 7,010.43 |
| 440 | 13,424.86 | 12,737.31 | 13,585.47 | 14,109.42 | 53,857.06 |
| 441 | 18,102.68 | 17,838.23 | 18,862.09 | 19,609.34 | 74,412.34 |
| 442 | 8,537.85 | 7,561.56 | 7,968.36 | 8,402.36 | 32,470.13 |
| 443 | 4,211.82 | 3,921.07 | 3,987.81 | 4,200.98 | 16,321.68 |
| 444 | 3,947.77 | 3,711.35 | 3,981.21 | 4,291.03 | 15,931.36 |
| 445 | 3,705.63 | 3,494.79 | 3,876.64 | 4,271.75 | 15,348.81 |

DRUG CODE:1100DRUG NAME:AMPHETAMINE
STATE:OHIO

| ZIP CODE | QUARTER 1 | QUARTER 2 | QUARTER 3 | QUARTER 4 | TOTAL GRAMS |
|---|---|---|---|---|---|
| 446 | 5,895.48 | 5,522.81 | 5,590 | 5,812.86 | 22,821.15 |
| 447 | 4,806.70 | 4,628.15 | 4,862.56 | 5,100.14 | 19,397.55 |
| 448 | 3,070.04 | 3,036.54 | 3,057.50 | 3,189.39 | 12,353.47 |
| 449 | 818 | 867.34 | 818.05 | 871.88 | 3,375.27 |
| 450 | 14,460.78 | 14,802.96 | 14,633.43 | 14,881.42 | 58,778.59 |
| 451 | 5,224.92 | 5,157.12 | 5,124.02 | 5,256.15 | 20,762.21 |
| 452 | 18,891.66 | 19,402.27 | 20,161.96 | 20,706.17 | 79,162.06 |
| 453 | 4,692 | 4,761.05 | 4,658.87 | 4,991.02 | 19,102.94 |
| 454 | 5,536.20 | 5,561.82 | 5,540.83 | 5,927.82 | 22,566.67 |
| 455 | 1,571.87 | 1,559.36 | 1,552.46 | 1,495.49 | 6,179.18 |
| 456 | 3,603.30 | 3,649.70 | 3,530.28 | 3,590.32 | 14,373.60 |
| 457 | 3,188.89 | 2,970.08 | 3,078.61 | 3,106.32 | 12,343.90 |
| 458 | 3,007.47 | 3,029.85 | 3,019.29 | 3,265.79 | 12,322.40 |
| TOTAL | 202,585.13 | 198,292.05 | 201,364.83 | 209,609.30 | 811,851.31 |

STATE: OKLAHOMA

| ZIP CODE | QUARTER 1 | QUARTER 2 | QUARTER 3 | QUARTER 4 | TOTAL GRAMS |
|---|---|---|---|---|---|
| 730 | 9,404.01 | 9,771.46 | 9,997.77 | 10,195.75 | 39,368.99 |
| 731 | 8,145.41 | 8,661.05 | 8,817.39 | 8,715.08 | 34,338.93 |
| 734 | 776.8 | 783.16 | 780.06 | 830.35 | 3,170.37 |
| 735 | 2,580.29 | 2,217.76 | 2,477.83 | 2,292.16 | 9,568.04 |
| 736 | 250.25 | 238.12 | 253.31 | 268.28 | 1,009.96 |
| 737 | 875.14 | 837.5 | 800.66 | 914.81 | 3,428.11 |
| 738 | 167.02 | 142.54 | 151.72 | 155.89 | 617.17 |
| 739 | 119.14 | 81.4 | 91.42 | 86.14 | 378.1 |
| 740 | 11,303.31 | 11,303.77 | 11,601.03 | 11,824.18 | 46,032.29 |
| 741 | 9,102.37 | 9,819.95 | 10,133.54 | 9,677.69 | 38,733.55 |
| 743 | 982.46 | 1,030.56 | 996.02 | 1,069.73 | 4,078.77 |
| 744 | 2,623.46 | 2,556.64 | 2,648.11 | 2,549.54 | 10,377.75 |
| 745 | 416.47 | 487.87 | 459.34 | 475.99 | 1,839.67 |
| 746 | 262.96 | 262.45 | 265.08 | 248.71 | 1,039.20 |
| 747 | 581.81 | 600.55 | 566.98 | 585.14 | 2,334.48 |
| 748 | 1,346.92 | 1,233.20 | 1,226.08 | 1,356.17 | 5,162.37 |
| 749 | 394.28 | 415.08 | 402.49 | 437.49 | 1,649.34 |
| TOTAL | 49,332.10 | 50,443.06 | 51,668.83 | 51,683.10 | 203,127.09 |

STATE: OREGON

| ZIP CODE | QUARTER 1 | QUARTER 2 | QUARTER 3 | QUARTER 4 | TOTAL GRAMS |
|---|---|---|---|---|---|

| | | | | | |
|---|---|---|---|---|---|
| 970 | 11,778.26 | 12,398.89 | 12,455.87 | 12,481.20 | 49,114.22 |
| 971 | 4,545.53 | 4,968.68 | 4,884.17 | 4,726.55 | 19,124.93 |
| 972 | 16,329.64 | 18,058.15 | 17,528.22 | 17,432.82 | 69,348.83 |
| 973 | 8,343.72 | 8,658.33 | 8,440.92 | 8,447.37 | 33,890.34 |
| 974 | 6,404.98 | 6,828.60 | 6,484.86 | 6,413.87 | 26,132.31 |
| 975 | 2,018.31 | 2,165.65 | 2,158.02 | 2,108.31 | 8,450.29 |
| 976 | 427.79 | 486.35 | 401.71 | 471.15 | 1,787 |
| 977 | 3,401.99 | 3,639.93 | 3,726.75 | 3,610.91 | 14,379.58 |
| 978 | 888.1 | 919.57 | 877.08 | 866.61 | 3,551.36 |
| 979 | 338.48 | 374.58 | 307.04 | 339 | 1,359.10 |
| TOTAL | 54,476.80 | 58,498.73 | 57,264.64 | 56,897.79 | 227,137.96 |

STATE: PENNSYLVANIA

| ZIP CODE | QUARTER 1 | QUARTER 2 | QUARTER 3 | QUARTER 4 | TOTAL GRAMS |
|---|---|---|---|---|---|
| 150 | 6,265.47 | 5,703 | 5,733.33 | 6,141.38 | 23,843.18 |
| 151 | 4,816.20 | 4,049 | 4,368.06 | 4,921.45 | 18,154.71 |
| 152 | 10,122.21 | 8,876.49 | 9,435.74 | 10,451.38 | 38,885.82 |
| 153 | 2,502.20 | 2,088.06 | 2,165.52 | 2,411.51 | 9,167.29 |
| 154 | 2,021.73 | 2,031.78 | 1,964.78 | 1,984.39 | 8,002.68 |
| 155 | 853.31 | 797 | 780.39 | 915.82 | 3,346.52 |
| 156 | 4,123.25 | 3,862.03 | 4,052.29 | 4,381.33 | 16,418.90 |
| 157 | 1,542.18 | 1,420.30 | 1,370.13 | 1,372.72 | 5,705.33 |
| 158 | 1,387.96 | 1,369.35 | 1,328.46 | 1,286.74 | 5,372.51 |
| 159 | 1,313.53 | 1,184.64 | 1,290.91 | 1,392.27 | 5,181.35 |
| 160 | 2,849.37 | 2,666.15 | 2,780.36 | 3,111.40 | 11,407.28 |
| 161 | 2,519.04 | 2,322.87 | 2,326.19 | 2,414.10 | 9,582.20 |
| 162 | 1,039.50 | 1,050.42 | 991.73 | 987.4 | 4,069.05 |
| 163 | 2,139.10 | 2,000.23 | 1,931.01 | 2,001.79 | 8,072.13 |
| 164 | 1,115.54 | 910.53 | 955.6 | 1,020.43 | 4,002.10 |
| 165 | 3,972.63 | 3,761.60 | 3,968.14 | 3,930.16 | 15,632.53 |
| 166 | 2,622.19 | 2,523.80 | 2,544.72 | 2,736.48 | 10,427.19 |

DRUG CODE:1100DRUG NAME:AMPHETAMINE
STATE:PENNSYLVANIA

| ZIP CODE | QUARTER 1 | QUARTER 2 | QUARTER 3 | QUARTER 4 | TOTAL GRAMS |
|---|---|---|---|---|---|
| 167 | 570.38 | 584.07 | 588.71 | 593.69 | 2,336.85 |
| 168 | 2,819.57 | 2,769.86 | 2,671.41 | 2,884.32 | 11,145.16 |
| 169 | 484.09 | 480.61 | 452.02 | 464.12 | 1,880.84 |
| 170 | 11,539.12 | 11,875.73 | 11,868.16 | 11,689.86 | 46,972.87 |
| 171 | 2,958.11 | 3,132.99 | 3,014.87 | 2,982.87 | 12,088.84 |
| 172 | 3,205.61 | 3,242.22 | 3,135.28 | 3,216.92 | 12,800.03 |
| 173 | 5,131.01 | 5,324.47 | 5,367.04 | 5,353.34 | 21,175.86 |
| 174 | 3,981.38 | 4,228.05 | 4,025.57 | 4,148.69 | 16,383.69 |
| 175 | 3,949.91 | 3,792.55 | 3,686.13 | 3,720.63 | 15,149.22 |
| 176 | 3,930.95 | 4,153.85 | 4,113.20 | 3,918.83 | 16,116.83 |
| 177 | 2,164.89 | 2,071.67 | 2,042.01 | 2,045.74 | 8,324.31 |
| 178 | 2,575.47 | 2,617.55 | 2,474.86 | 2,694.95 | 10,362.83 |
| 179 | 1,095.33 | 1,067.52 | 975 | 1,040.04 | 4,177.89 |
| 180 | 5,526.95 | 5,566.52 | 5,469.80 | 5,538.84 | 22,102.11 |
| 181 | 2,969.62 | 3,000.42 | 3,070 | 3,012.69 | 12,052.73 |
| 182 | 1,043.27 | 1,071.07 | 1,039.61 | 1,053.22 | 4,207.17 |
| 183 | 1,249.36 | 1,238.05 | 1,291.33 | 1,309.44 | 5,088.18 |
| 184 | 1,987.54 | 2,072.15 | 2,117.70 | 2,130.94 | 8,308.33 |
| 185 | 3,742.07 | 3,926.96 | 3,786 | 3,698.60 | 15,153.63 |
| 186 | 1,756.69 | 1,751.16 | 1,795.24 | 1,692.91 | 6,996 |
| 187 | 18,221.46 | 16,928.39 | 14,903.25 | 15,786.77 | 65,839.87 |
| 188 | 681.97 | 676.83 | 687.09 | 673.12 | 2,719.01 |
| 189 | 7,756.83 | 7,970.32 | 7,929.54 | 7,534.14 | 31,190.83 |
| 190 | 24,178.12 | 25,071.51 | 24,519.78 | 23,316.41 | 97,085.82 |
| 191 | 21,277.86 | 22,215.46 | 21,295.33 | 20,545.48 | 85,334.13 |

| | | | | | |
|---|---|---|---|---|---|
| 193 | 10,203.54 | 10,670.09 | 10,441.30 | 10,119.68 | 41,434.61 |
| 194 | 11,330.50 | 11,691.91 | 11,549.35 | 11,136.01 | 45,707.77 |
| 195 | 2,011.26 | 1,898.62 | 1,922.63 | 1,920.29 | 7,752.80 |
| 196 | 3,072.81 | 3,053.82 | 3,048.49 | 2,886.71 | 12,061.83 |
| TOTAL | 212,621.08 | 210,761.67 | 207,268.06 | 208,570 | 839,220.81 |

STATE: PUERTO RICO

| ZIP CODE | QUARTER 1 | QUARTER 2 | QUARTER 3 | QUARTER 4 | TOTAL GRAMS |
|---|---|---|---|---|---|
| 6 | 1,850.79 | 1,721.44 | 1,990.79 | 1,981.31 | 7,544.33 |
| 7 | 1,837.02 | 1,952.48 | 1,938.39 | 2,010.84 | 7,738.73 |
| 9 | 4,184.87 | 4,210.33 | 4,400.24 | 4,492.65 | 17,288.09 |
| TOTAL | 7,872.68 | 7,884.25 | 8,329.42 | 8,484.80 | 32,571.15 |

STATE: RHODE ISLAND

| ZIP CODE | QUARTER 1 | QUARTER 2 | QUARTER 3 | QUARTER 4 | TOTAL GRAMS |
|---|---|---|---|---|---|
| 28 | 21,636.56 | 22,440.20 | 22,519.39 | 22,617.43 | 89,213.58 |
| 29 | 11,989.03 | 12,517.48 | 12,701.72 | 12,834.54 | 50,042.77 |
| TOTAL | 33,625.59 | 34,957.68 | 35,221.11 | 35,451.97 | 139,256.35 |

STATE: SOUTH CAROLINA

| ZIP CODE | QUARTER 1 | QUARTER 2 | QUARTER 3 | QUARTER 4 | TOTAL GRAMS |
|---|---|---|---|---|---|
| 290 | 10,987.12 | 11,449.21 | 11,820.35 | 11,731.32 | 45,988 |
| 291 | 9,266.59 | 9,382.41 | 9,558.84 | 9,606.33 | 37,814.17 |
| 292 | 9,976.71 | 9,958.24 | 10,102.14 | 10,354.10 | 40,391.19 |
| 293 | 8,718.66 | 8,388.27 | 8,696.60 | 8,585.81 | 34,389.34 |
| 294 | 30,759.56 | 32,319.41 | 32,761.34 | 31,883.41 | 127,723.72 |
| 295 | 16,883.40 | 17,538.84 | 18,249.05 | 17,608.32 | 70,279.61 |
| 296 | 25,872.60 | 27,093.60 | 27,199.94 | 27,580.82 | 107,746.96 |
| 297 | 7,013.13 | 7,186.09 | 7,309.93 | 7,519.42 | 29,028.57 |
| 298 | 4,281.27 | 4,329.43 | 4,447.86 | 4,515.94 | 17,574.50 |
| 299 | 3,313.46 | 3,635.15 | 3,888.77 | 3,638.45 | 14,475.83 |
| TOTAL | 127,072.50 | 131,280.65 | 134,034.82 | 133,023.92 | 525,411.89 |

STATE: SOUTH DAKOTA

| ZIP CODE | QUARTER 1 | QUARTER 2 | QUARTER 3 | QUARTER 4 | TOTAL GRAMS |
|---|---|---|---|---|---|
| 570 | 1,982.36 | 1,966.91 | 2,049.01 | 1,988.55 | 7,986.83 |
| 571 | 10,852.31 | 8,086.03 | 9,412.08 | 8,906.35 | 37,256.77 |
| 572 | 621.83 | 612.05 | 658.85 | 630.64 | 2,523.37 |
| 573 | 602.98 | 636.88 | 664.13 | 668.21 | 2,572.20 |
| 574 | 538.77 | 567.34 | 600.46 | 580.98 | 2,287.55 |
| 575 | 255.73 | 231.16 | 222.22 | 277.37 | 986.48 |
| 576 | 57.94 | 53.44 | 46.13 | 29.94 | 187.45 |
| 577 | 2,196.19 | 2,259.78 | 2,323.10 | 2,280.95 | 9,060.02 |
| TOTAL | 17,108.11 | 14,413.59 | 15,975.98 | 15,362.99 | 62,860.67 |

STATE: TENNESSEE

DRUG CODE:1100DRUG NAME:AMPHETAMINE
STATE:TENNESSEE

| ZIP CODE | QUARTER 1 | QUARTER 2 | QUARTER 3 | QUARTER 4 | TOTAL GRAMS |
|---|---|---|---|---|---|
| 370 | 17,925.48 | 19,043.12 | 19,278.85 | 19,293.09 | 75,540.54 |
| 371 | 11,304.89 | 11,880.63 | 12,374.71 | 12,338.58 | 47,898.81 |
| 372 | 12,371.63 | 13,371.85 | 13,850.29 | 13,584.14 | 53,177.91 |
| 373 | 11,464.19 | 11,727.19 | 11,920.51 | 11,909.77 | 47,021.66 |
| 374 | 8,831.48 | 9,263.66 | 9,108.42 | 9,033.02 | 36,236.58 |
| 376 | 3,767.77 | 3,738.84 | 3,935.97 | 4,001.28 | 15,443.86 |
| 377 | 4,245.96 | 4,254.16 | 4,527.19 | 4,352.18 | 17,379.49 |
| 378 | 6,261.35 | 6,483.54 | 6,520.30 | 6,861.85 | 26,127.04 |
| 379 | 10,404 | 10,551.45 | 10,872.23 | 10,971.60 | 42,799.28 |

RETAIL DRUG DISTRIBUTION BY ZIP CODE WITHIN STATE BY GRAMS WT

| | | | | | | |
|---|---|---|---|---|---|---|
| | 380 | 8,989.04 | 9,181.70 | 9,460.21 | 9,459.80 | 37,090.75 |
| | 381 | 11,377.99 | 11,703.17 | 12,072.81 | 12,213.28 | 47,367.25 |
| | 382 | 3,049.15 | 2,975.54 | 3,274.39 | 3,125.65 | 12,424.73 |
| | 383 | 4,794.70 | 4,488.69 | 4,837.85 | 4,861.39 | 18,982.63 |
| | 384 | 3,101.74 | 3,230.94 | 3,346.39 | 3,464.31 | 13,143.38 |
| | 385 | 2,102.94 | 2,177.76 | 2,141.06 | 2,227 | 8,648.76 |
| TOTAL | | 119,992.31 | 124,072.24 | 127,521.18 | 127,696.94 | 499,282.67 |

STATE: TEXAS

| ZIP CODE | QUARTER 1 | QUARTER 2 | QUARTER 3 | QUARTER 4 | TOTAL GRAMS |
|---|---|---|---|---|---|
| 750 | 46,517.47 | 49,620.16 | 50,316.59 | 50,138.14 | 196,592.36 |
| 751 | 7,640.98 | 8,080.28 | 8,257.49 | 8,523.63 | 32,502.38 |
| 752 | 22,855.70 | 24,286.48 | 25,218 | 25,000.30 | 97,360.48 |
| 753 | 22.89 | 23.24 | 23.85 | 26.69 | 96.67 |
| 754 | 3,627.42 | 3,649.81 | 3,785.60 | 3,807.86 | 14,870.69 |
| 755 | 1,085.95 | 1,039.16 | 1,019.93 | 1,127.37 | 4,272.41 |
| 756 | 3,262.13 | 3,541.37 | 3,454.57 | 3,565.79 | 13,823.86 |
| 757 | 4,299.90 | 4,270.75 | 4,235.31 | 4,685.05 | 17,491.01 |
| 758 | 766.21 | 799.2 | 750.56 | 840.21 | 3,156.18 |
| 759 | 2,525.52 | 2,512.03 | 2,341.09 | 2,515.17 | 9,893.81 |
| 760 | 19,425.86 | 20,984.64 | 21,131.73 | 21,768.66 | 83,310.89 |
| 761 | 12,141.61 | 12,875.95 | 12,801.64 | 13,333.92 | 51,153.12 |
| 762 | 9,538.66 | 10,035.91 | 10,393.85 | 10,721.90 | 40,690.32 |
| 763 | 2,812.62 | 2,920.32 | 2,950.28 | 3,052.26 | 11,735.48 |
| 764 | 1,251.34 | 1,294.12 | 1,340.51 | 1,308.14 | 5,194.11 |
| 765 | 5,626.67 | 6,382.59 | 5,715.83 | 5,629.58 | 23,354.67 |
| 766 | 1,021.80 | 1,026.91 | 1,045.12 | 1,061.28 | 4,155.11 |
| 767 | 3,370.88 | 3,446.08 | 3,361.95 | 3,393.68 | 13,572.59 |
| 768 | 679.87 | 655.92 | 695.44 | 766.6 | 2,797.83 |
| 769 | 1,508.64 | 1,495.53 | 1,552.07 | 1,599.30 | 6,155.54 |
| 770 | 35,287.98 | 36,953.90 | 37,672.76 | 37,842.64 | 147,757.28 |
| 772 | 56.93 | 52.88 | 56.31 | 64.13 | 230.25 |
| 773 | 25,002.45 | 26,916.80 | 27,599.14 | 27,118.54 | 106,636.93 |
| 774 | 17,661.64 | 18,036.73 | 18,973.43 | 19,444.14 | 74,115.94 |
| 775 | 20,515.65 | 21,425.29 | 21,909.04 | 21,855.59 | 85,705.57 |
| 776 | 4,588.88 | 4,932.95 | 4,991.39 | 4,888.12 | 19,401.34 |
| 777 | 2,623.24 | 2,696.16 | 2,636.42 | 2,625.17 | 10,580.99 |
| 778 | 6,326.82 | 6,483.46 | 6,316.43 | 6,886.89 | 26,013.60 |
| 779 | 1,917.49 | 2,012.24 | 1,906.37 | 2,002.24 | 7,838.34 |
| 780 | 4,077.83 | 4,211.16 | 4,252.86 | 4,426.38 | 16,968.23 |
| 781 | 5,282.27 | 5,487.48 | 5,473.85 | 5,656.76 | 21,900.36 |
| 782 | 23,755.52 | 25,077.55 | 24,365.07 | 25,312.51 | 98,510.65 |
| 783 | 1,852.99 | 1,783.31 | 1,899.57 | 1,888.35 | 7,424.22 |
| 784 | 4,711.23 | 4,558.18 | 4,774.72 | 4,938.62 | 18,982.75 |
| 785 | 2,657.60 | 2,697.32 | 2,717.68 | 2,891 | 10,963.60 |
| 786 | 15,259.42 | 15,252.63 | 15,777.88 | 16,114.88 | 62,404.81 |
| 787 | 25,638.87 | 27,213.13 | 27,292.86 | 27,365.93 | 107,510.79 |
| 788 | 600.16 | 634.36 | 554.95 | 619.98 | 2,409.45 |
| 789 | 594.4 | 638.58 | 639.81 | 674.83 | 2,547.62 |
| 790 | 893.89 | 855.97 | 766.97 | 887.96 | 3,404.79 |
| 791 | 1,588.67 | 1,593.16 | 1,624.67 | 1,820.02 | 6,626.52 |
| 792 | 72.72 | 80.88 | 61.95 | 96.84 | 312.39 |
| 793 | 487.08 | 446.82 | 422.03 | 451.55 | 1,807.48 |
| 794 | 4,186.33 | 4,108.02 | 4,327.03 | 4,347.74 | 16,969.12 |
| 795 | 470.03 | 499.88 | 461.16 | 491.49 | 1,922.56 |
| 796 | 2,018.26 | 2,026.32 | 2,026.60 | 2,071.91 | 8,143.09 |
| 797 | 5,317.99 | 5,827.76 | 5,275.28 | 5,625.07 | 22,046.10 |
| 798 | 255.94 | 216.68 | 227.48 | 239.12 | 939.22 |

DRUG CODE:1100DRUG NAME:AMPHETAMINE
STATE:TEXAS

RETAIL DRUG DISTRIBUTION BY ZIP CODE WITHIN STATE BY GRAMS WT

| ZIP CODE | QUARTER 1 | QUARTER 2 | QUARTER 3 | QUARTER 4 | TOTAL GRAMS |
|---|---|---|---|---|---|
| 799 | 2,889.95 | 2,964.53 | 3,137.96 | 2,937 | 11,929.44 |
| TOTAL | 366,574.35 | 384,624.58 | 388,533.08 | 394,450.93 | 1,534,182.94 |
| STATE: | UTAH | | | | |
| ZIP CODE | QUARTER 1 | QUARTER 2 | QUARTER 3 | QUARTER 4 | TOTAL GRAMS |
| 840 | 35,638.38 | 37,455.08 | 39,099.12 | 39,135.63 | 151,328.21 |
| 841 | 14,893.56 | 15,711.39 | 15,814.82 | 15,727.93 | 62,147.70 |
| 843 | 4,012.86 | 4,112.29 | 4,022.23 | 4,230.45 | 16,377.83 |
| 844 | 3,285.41 | 3,473.56 | 3,553.06 | 3,505.36 | 13,817.39 |
| 845 | 712.39 | 744.64 | 656.08 | 691.7 | 2,804.81 |
| 846 | 7,449.74 | 7,706.11 | 7,776.86 | 8,012.68 | 30,945.39 |
| 847 | 5,359.38 | 5,528.28 | 5,772.56 | 6,106.62 | 22,766.84 |
| TOTAL | 71,351.72 | 74,731.35 | 76,694.73 | 77,410.37 | 300,188.17 |
| STATE: | VERMONT | | | | |
| ZIP CODE | QUARTER 1 | QUARTER 2 | QUARTER 3 | QUARTER 4 | TOTAL GRAMS |
| 50 | 517.09 | 592.73 | 556.48 | 623.41 | 2,289.71 |
| 51 | 520.19 | 484.89 | 484.64 | 543.31 | 2,033.03 |
| 52 | 622.72 | 708.67 | 669.47 | 676.49 | 2,677.35 |
| 53 | 1,658.56 | 1,699.52 | 1,570.42 | 1,635.26 | 6,563.76 |
| 54 | 3,247.58 | 3,238.32 | 3,291.29 | 3,278.64 | 13,055.83 |
| 56 | 1,679.87 | 1,712.14 | 1,719.15 | 1,702.65 | 6,813.81 |
| 57 | 888.76 | 867.09 | 785.49 | 806.1 | 3,347.44 |
| 58 | 535.59 | 552.32 | 557.13 | 618.98 | 2,264.02 |
| TOTAL | 9,670.36 | 9,855.68 | 9,634.07 | 9,884.84 | 39,044.95 |
| STATE: | VIRGIN ISLANDS | | | | |
| ZIP CODE | QUARTER 1 | QUARTER 2 | QUARTER 3 | QUARTER 4 | TOTAL GRAMS |
| 8 | 308.21 | 311.32 | 296.41 | 335.32 | 1,251.26 |
| TOTAL | 308.21 | 311.32 | 296.41 | 335.32 | 1,251.26 |
| STATE: | VIRGINIA | | | | |
| ZIP CODE | QUARTER 1 | QUARTER 2 | QUARTER 3 | QUARTER 4 | TOTAL GRAMS |
| 201 | 16,535.54 | 17,326.76 | 17,636.90 | 17,726.22 | 69,225.42 |
| 220 | 10,036.02 | 10,744.30 | 10,468.43 | 10,054.78 | 41,303.53 |
| 221 | 6,709.94 | 6,838.83 | 6,868.24 | 6,744.82 | 27,161.83 |
| 222 | 4,504.33 | 5,008.01 | 4,821.77 | 4,752.48 | 19,086.59 |
| 223 | 3,795.79 | 3,689.12 | 3,879.16 | 3,985.30 | 15,349.37 |
| 224 | 4,130.05 | 4,336.20 | 4,138.55 | 4,439.57 | 17,044.37 |
| 225 | 1,869.98 | 2,000.92 | 1,949.40 | 2,027.13 | 7,847.43 |
| 226 | 2,812.59 | 3,048.40 | 2,961.29 | 3,105.81 | 11,928.09 |
| 227 | 873.59 | 1,027.76 | 1,021.85 | 1,058.33 | 3,981.53 |
| 228 | 3,091 | 3,037.12 | 2,877.13 | 3,150.90 | 12,156.15 |
| 229 | 5,996.09 | 6,274.87 | 6,080.92 | 6,425.73 | 24,777.61 |
| 230 | 4,671.51 | 4,835.58 | 5,078.69 | 5,000.55 | 19,586.33 |
| 231 | 10,412.64 | 10,998.14 | 11,358.31 | 11,211.24 | 43,980.33 |
| 232 | 14,803.99 | 15,467.48 | 15,442.27 | 15,873.05 | 61,586.79 |
| 233 | 9,253.96 | 9,660.69 | 9,532.26 | 9,381.28 | 37,828.19 |
| 234 | 23,465.40 | 24,663.50 | 24,917.75 | 24,555.14 | 97,601.79 |
| 235 | 6,054.46 | 6,276.17 | 6,295.72 | 6,393.56 | 25,019.91 |
| 236 | 8,648.17 | 8,821.35 | 8,988.28 | 8,911.61 | 35,369.41 |
| 237 | 3,967.04 | 3,988.11 | 4,218.14 | 3,953.54 | 16,126.83 |
| 238 | 6,026.07 | 6,299.18 | 6,249.72 | 6,418.29 | 24,993.26 |
| 239 | 1,107.08 | 1,016.80 | 999.78 | 1,091.47 | 4,215.13 |
| 240 | 4,757.71 | 5,117.82 | 5,073.50 | 5,145.36 | 20,094.39 |
| 241 | 2,625.44 | 2,624.44 | 2,642.58 | 2,802.47 | 10,694.93 |
| 242 | 1,327.02 | 1,454.55 | 1,403.24 | 1,444.73 | 5,629.54 |
| 243 | 866.52 | 888.74 | 876.33 | 917.43 | 3,549.02 |
| 244 | 3,232.82 | 3,358.97 | 3,262.10 | 3,301.19 | 13,155.08 |

| | | | | | |
|---|---|---|---|---|---|
| 245 | 4,992.57 | 5,098.79 | 5,120.91 | 5,365.10 | 20,577.37 |
| 246 | 335.28 | 310.11 | 313.42 | 366.98 | 1,325.79 |
| TOTAL | 166,902.60 | 174,212.71 | 174,476.64 | 175,604.06 | 691,196.01 |

STATE: WASHINGTON

| ZIP CODE | QUARTER 1 | QUARTER 2 | QUARTER 3 | QUARTER 4 | TOTAL GRAMS |
|---|---|---|---|---|---|
| 980 | 24,109.31 | 25,418.84 | 25,122.29 | 26,226.85 | 100,877.29 |
| 981 | 17,472.92 | 18,295.20 | 18,166.20 | 16,809.43 | 70,743.75 |
| 982 | 14,902.51 | 15,842.69 | 15,494.37 | 15,705.76 | 61,945.33 |
| 983 | 9,611.51 | 10,175.86 | 9,897.56 | 10,014.45 | 39,699.38 |
| 984 | 6,853.33 | 7,401.89 | 7,086.27 | 7,091.71 | 28,433.20 |
| 985 | 5,162.47 | 5,088.96 | 5,280.96 | 5,058.30 | 20,590.69 |

DRUG CODE:1100DRUG NAME:AMPHETAMINE
STATE:WASHINGTON

| ZIP CODE | QUARTER 1 | QUARTER 2 | QUARTER 3 | QUARTER 4 | TOTAL GRAMS |
|---|---|---|---|---|---|
| 986 | 7,287.06 | 7,898.56 | 7,843.85 | 7,977.84 | 31,007.31 |
| 988 | 1,670.62 | 1,762.53 | 1,729.41 | 1,697.16 | 6,859.72 |
| 989 | 2,465.57 | 2,549.45 | 2,376.14 | 2,402.28 | 9,793.44 |
| 990 | 956.45 | 1,037.01 | 1,038.52 | 900.23 | 3,932.21 |
| 991 | 1,142.51 | 1,016.37 | 1,088.25 | 1,141.94 | 4,389.07 |
| 992 | 4,752.48 | 5,118.54 | 4,949.98 | 5,242.05 | 20,063.05 |
| 993 | 4,966.58 | 5,376.08 | 5,303.38 | 5,318.72 | 20,964.76 |
| 994 | 157.63 | 179.62 | 166.81 | 175.03 | 679.09 |
| TOTAL | 101,510.95 | 107,161.60 | 105,543.99 | 105,761.75 | 419,978.29 |

STATE: WEST VIRGINIA

| ZIP CODE | QUARTER 1 | QUARTER 2 | QUARTER 3 | QUARTER 4 | TOTAL GRAMS |
|---|---|---|---|---|---|
| 247 | 295.63 | 287.43 | 303.09 | 306.53 | 1,192.68 |
| 248 | 196.08 | 206.22 | 195.1 | 215.8 | 813.2 |
| 249 | 411.41 | 395.04 | 430.77 | 463.49 | 1,700.71 |
| 250 | 978.76 | 919.33 | 887.96 | 816.44 | 3,602.49 |
| 251 | 749.53 | 768.31 | 711.46 | 705.13 | 2,934.43 |
| 252 | 503.09 | 517.3 | 534.69 | 491.87 | 2,046.95 |
| 253 | 3,777.62 | 3,680.74 | 3,899.31 | 3,583.95 | 14,941.62 |
| 254 | 2,366.89 | 2,439.02 | 2,429.19 | 2,468.38 | 9,703.48 |
| 255 | 2,290.30 | 2,196.64 | 2,167.28 | 2,294.91 | 8,949.13 |
| 256 | 310.51 | 336.1 | 246.55 | 272.8 | 1,165.96 |
| 257 | 1,709.98 | 1,727.55 | 1,710.26 | 1,649.50 | 6,797.29 |
| 258 | 1,350.12 | 1,295.91 | 1,360.54 | 1,314.96 | 5,321.53 |
| 259 | 431.23 | 370.91 | 390.77 | 339.96 | 1,532.87 |
| 260 | 1,445.67 | 1,437.71 | 1,438.90 | 1,464.50 | 5,786.78 |
| 261 | 2,762 | 2,628.67 | 2,767.37 | 2,514.25 | 10,672.29 |
| 262 | 343.39 | 344.85 | 334.51 | 354.58 | 1,377.33 |
| 263 | 951.6 | 934.24 | 927.79 | 967.24 | 3,780.87 |
| 264 | 239.08 | 248.33 | 254.42 | 247.7 | 989.53 |
| 265 | 2,684.49 | 2,712.37 | 2,665.72 | 2,754.21 | 10,816.79 |
| 266 | 138.98 | 150.69 | 168.79 | 168.65 | 627.11 |
| 267 | 244.59 | 265.79 | 274.37 | 311.37 | 1,096.12 |
| 268 | 229.72 | 202.86 | 205.97 | 244.83 | 883.38 |
| TOTAL | 24,410.67 | 24,066.01 | 24,304.81 | 23,951.05 | 96,732.54 |

STATE: WISCONSIN

| ZIP CODE | QUARTER 1 | QUARTER 2 | QUARTER 3 | QUARTER 4 | TOTAL GRAMS |
|---|---|---|---|---|---|
| 530 | 14,366.62 | 14,600.71 | 14,523.84 | 14,689.06 | 58,180.23 |
| 531 | 15,635.86 | 15,714.82 | 15,648.52 | 15,323.45 | 62,322.65 |
| 532 | 13,488.28 | 13,792.88 | 13,956.18 | 13,913 | 55,150.34 |
| 534 | 5,023.10 | 5,185.14 | 4,924.09 | 4,505.58 | 19,637.91 |
| 535 | 10,622.51 | 10,301.56 | 10,098.10 | 9,860.84 | 40,883.01 |
| 537 | 8,633.99 | 8,652.72 | 8,568.30 | 8,548.40 | 34,403.41 |

| | | | | | |
|---|---|---|---|---|---|
| 538 | 1,323.69 | 1,273.63 | 1,245.09 | 1,238.54 | 5,080.95 |
| 539 | 3,859.26 | 3,690.27 | 3,781.98 | 3,479.46 | 14,810.97 |
| 540 | 2,649.90 | 2,478.43 | 2,629.89 | 2,574.07 | 10,332.29 |
| 541 | 7,402.73 | 7,187.37 | 7,023.54 | 6,930.76 | 28,544.40 |
| 542 | 3,063.14 | 2,974.23 | 2,845.41 | 2,849.85 | 11,732.63 |
| 543 | 8,661.59 | 8,736.89 | 8,516.78 | 8,649.14 | 34,564.40 |
| 544 | 9,358.33 | 9,218.22 | 9,096.97 | 9,010.05 | 36,683.57 |
| 545 | 2,681.64 | 2,618.22 | 2,635.66 | 2,415.39 | 10,350.91 |
| 546 | 4,754.73 | 4,795.06 | 4,480.98 | 4,662.67 | 18,693.44 |
| 547 | 5,006.44 | 4,863.85 | 4,785.86 | 4,860.13 | 19,516.28 |
| 548 | 2,571.55 | 2,706.94 | 2,592.61 | 2,523.47 | 10,394.57 |
| 549 | 16,278.01 | 16,428.14 | 16,366.34 | 16,230.75 | 65,303.24 |
| TOTAL | 135,381.37 | 135,219.08 | 133,720.14 | 132,264.61 | 536,585.20 |

STATE: WYOMING

| ZIP CODE | QUARTER 1 | QUARTER 2 | QUARTER 3 | QUARTER 4 | TOTAL GRAMS |
|---|---|---|---|---|---|
| 820 | 1,301.86 | 1,298.42 | 1,296.02 | 1,375.43 | 5,271.73 |
| 822 | 98.84 | 106.12 | 95.67 | 86.02 | 386.65 |
| 823 | 82.21 | 118.27 | 112.78 | 123.21 | 436.47 |
| 824 | 794.8 | 846.7 | 820.77 | 830.43 | 3,292.70 |
| 825 | 275.8 | 267.8 | 342.42 | 303.99 | 1,190.01 |
| 826 | 1,029.69 | 1,015.86 | 1,030.01 | 951.36 | 4,026.92 |
| 827 | 564.51 | 533.83 | 495.54 | 500.03 | 2,093.91 |
| 828 | 715.03 | 665.96 | 560.78 | 573.17 | 2,514.94 |
| 829 | 877.58 | 897.02 | 983.76 | 982.06 | 3,740.42 |

DRUG CODE:1100DRUG NAME:AMPHETAMINE
STATE:WYOMING

| ZIP CODE | QUARTER 1 | QUARTER 2 | QUARTER 3 | QUARTER 4 | TOTAL GRAMS |
|---|---|---|---|---|---|
| 830 | 403.58 | 478.26 | 502.9 | 457.75 | 1,842.49 |
| 831 | 151.45 | 146.69 | 152.48 | 133.16 | 583.78 |
| TOTAL | 6,295.35 | 6,374.93 | 6,393.13 | 6,316.61 | 25,380.02 |

DRUG CODE:1105BDRUG NAME:DL-METHAMPHETAMINE RACEMIC BASE
STATE: ALABAMA

| ZIP CODE | QUARTER 1 | QUARTER 2 | QUARTER 3 | QUARTER 4 | TOTAL GRAMS |
|---|---|---|---|---|---|
| 350 | 1.21 | 1.61 | 1.21 | 1.61 | 5.64 |
| 351 | 2.41 | 2.81 | 1.61 | 2.41 | 9.24 |
| 352 | 2.41 | 0.8 | 1.21 | 1.21 | 5.63 |
| 356 | 4.02 | 2.41 | 0.8 | 1.61 | 8.84 |
| 361 | 0.4 | 1.21 | 0.8 | 1.21 | 3.62 |
| TOTAL | 10.45 | 8.84 | 5.63 | 8.05 | 32.97 |

STATE: ALASKA

| ZIP CODE | QUARTER 1 | QUARTER 2 | QUARTER 3 | QUARTER 4 | TOTAL GRAMS |
|---|---|---|---|---|---|
| 995 | 0 | 0.8 | 0 | 0 | 0.8 |
| TOTAL | 0 | 0.8 | 0 | 0 | 0.8 |

STATE: ARIZONA

| ZIP CODE | QUARTER 1 | QUARTER 2 | QUARTER 3 | QUARTER 4 | TOTAL GRAMS |
|---|---|---|---|---|---|
| 850 | 0 | 1.21 | 2.01 | 2.41 | 5.63 |
| 852 | 17.28 | 16.07 | 10.45 | 16.48 | 60.28 |
| 853 | 3.21 | 2.81 | 4.42 | 5.22 | 15.66 |
| 856 | 0 | 0 | 0 | 1.21 | 1.21 |
| 857 | 0 | 0 | 0.8 | 0.4 | 1.2 |
| TOTAL | 20.49 | 20.09 | 17.68 | 25.72 | 83.98 |

STATE: ARKANSAS

| ZIP CODE | QUARTER 1 | QUARTER 2 | QUARTER 3 | QUARTER 4 | TOTAL GRAMS |
|---|---|---|---|---|---|

| | | | | | |
|---|---|---|---|---|---|
| 720 | 1.21 | 0.8 | 2.41 | 0.8 | 5.22 |
| 729 | 0 | 0 | 0.4 | 0.4 | 0.8 |
| TOTAL | 1.21 | 0.8 | 2.81 | 1.2 | 6.02 |

STATE: CALIFORNIA

| ZIP CODE | QUARTER 1 | QUARTER 2 | QUARTER 3 | QUARTER 4 | TOTAL GRAMS |
|---|---|---|---|---|---|
| 900 | 12.46 | 19.69 | 16.07 | 20.9 | 69.12 |
| 902 | 6.03 | 8.04 | 5.63 | 6.03 | 25.73 |
| 904 | 2.81 | 0 | 0 | 0 | 2.81 |
| 907 | 0.8 | 0 | 0 | 0 | 0.8 |
| 913 | 14.06 | 13.66 | 14.06 | 15.27 | 57.05 |
| 914 | 2.01 | 0.4 | 1.21 | 2.01 | 5.63 |
| 917 | 16.48 | 22.91 | 19.29 | 16.88 | 75.56 |
| 920 | 3.21 | 8.44 | 5.22 | 6.43 | 23.3 |
| 921 | 4.02 | 0 | 0 | 4.82 | 8.84 |
| 922 | 2.41 | 2.01 | 1.61 | 1.21 | 7.24 |
| 925 | 0.8 | 0.4 | 0.8 | 0.4 | 2.4 |
| 926 | 2.01 | 0.4 | 2.01 | 1.21 | 5.63 |
| 927 | 1.61 | 1.61 | 2.81 | 1.61 | 7.64 |
| 928 | 2.41 | 2.81 | 2.81 | 2.81 | 10.84 |
| 930 | 0.8 | 0.4 | 0 | 0 | 1.2 |
| 931 | 1.21 | 2.41 | 1.61 | 2.41 | 7.64 |
| 935 | 0 | 0 | 0 | 0.8 | 0.8 |
| 940 | 1.21 | 1.61 | 0.4 | 2.41 | 5.63 |
| 941 | 0.8 | 0 | 0 | 0.8 | 1.6 |
| 945 | 4.82 | 2.81 | 0 | 0 | 7.63 |
| 949 | 3.21 | 3.62 | 2.81 | 0.8 | 10.44 |
| 950 | 0.4 | 0.4 | 0 | 0 | 0.8 |
| 953 | 3.62 | 2.41 | 2.01 | 4.02 | 12.06 |
| 958 | 0.8 | 1.21 | 0 | 0 | 2.01 |
| TOTAL | 87.99 | 95.24 | 78.35 | 90.82 | 352.4 |

STATE: COLORADO

| ZIP CODE | QUARTER 1 | QUARTER 2 | QUARTER 3 | QUARTER 4 | TOTAL GRAMS |
|---|---|---|---|---|---|
| 800 | 2.41 | 0 | 0 | 0 | 2.41 |
| 802 | 0 | 0 | 0.8 | 0.4 | 1.2 |
| 803 | 4.02 | 1.61 | 0 | 0.8 | 6.43 |
| 806 | 0 | 0 | 0 | 0.4 | 0.4 |
| TOTAL | 6.43 | 1.61 | 0.8 | 1.6 | 10.44 |

STATE: CONNECTICUT

| ZIP CODE | QUARTER 1 | QUARTER 2 | QUARTER 3 | QUARTER 4 | TOTAL GRAMS |
|---|---|---|---|---|---|
| 60 | 0 | 0.4 | 1.21 | 0 | 1.61 |

DRUG CODE:1105BDRUG NAME:DL-METHAMPHETAMINE RACEMIC BASE
STATE:CONNECTICUT

| ZIP CODE | QUARTER 1 | QUARTER 2 | QUARTER 3 | QUARTER 4 | TOTAL GRAMS |
|---|---|---|---|---|---|
| 64 | 1.61 | 2.41 | 2.01 | 0.8 | 6.83 |
| 65 | 4.82 | 2.81 | 3.21 | 4.02 | 14.86 |
| 68 | 5.22 | 3.21 | 0 | 4.42 | 12.85 |
| TOTAL | 11.65 | 8.83 | 6.43 | 9.24 | 36.15 |

STATE: DISTRICT OF COLUMBIA

| ZIP CODE | QUARTER 1 | QUARTER 2 | QUARTER 3 | QUARTER 4 | TOTAL GRAMS |
|---|---|---|---|---|---|
| 200 | 3.62 | 3.62 | 2.81 | 1.21 | 11.26 |
| TOTAL | 3.62 | 3.62 | 2.81 | 1.21 | 11.26 |

STATE: FLORIDA

RETAIL DRUG DISTRIBUTION BY ZIP CODE WITHIN STATE BY GRAMS WT

| ZIP CODE | QUARTER 1 | QUARTER 2 | QUARTER 3 | QUARTER 4 | TOTAL GRAMS |
|---|---|---|---|---|---|
| 320 | 0 | 0.4 | 0 | 0 | 0.4 |
| 324 | 0.4 | 0 | 0 | 0 | 0.4 |
| 325 | 2.01 | 2.41 | 2.81 | 4.02 | 11.25 |
| 326 | 2.01 | 1.61 | 1.21 | 2.01 | 6.84 |
| 327 | 0.4 | 0 | 0 | 0 | 0.4 |
| 329 | 0.8 | 0 | 0 | 0.4 | 1.2 |
| 330 | 1.21 | 0 | 0 | 0 | 1.21 |
| 331 | 1.21 | 0.4 | 0.4 | 0 | 2.01 |
| 334 | 0.4 | 0 | 0 | 0 | 0.4 |
| 337 | 0.4 | 1.21 | 1.21 | 1.21 | 4.03 |
| 338 | 0.8 | 0 | 0 | 0 | 0.8 |
| 339 | 0.8 | 0 | 0 | 0 | 0.8 |
| 342 | 1.21 | 0.8 | 1.21 | 0.8 | 4.02 |
| 346 | 1.21 | 0.4 | 0 | 0 | 1.61 |
| 349 | 0.4 | 0 | 0 | 0 | 0.4 |
| TOTAL | 13.26 | 7.23 | 6.84 | 8.44 | 35.77 |

STATE: GEORGIA

| ZIP CODE | QUARTER 1 | QUARTER 2 | QUARTER 3 | QUARTER 4 | TOTAL GRAMS |
|---|---|---|---|---|---|
| 300 | 0.8 | 0 | 0 | 0 | 0.8 |
| 301 | 0.8 | 1.21 | 2.01 | 0 | 4.02 |
| 303 | 0.8 | 0 | 0 | 0 | 0.8 |
| 308 | 0.8 | 0 | 0 | 0 | 0.8 |
| 310 | 1.21 | 2.81 | 1.61 | 3.62 | 9.25 |
| 315 | 2.01 | 2.41 | 1.21 | 2.81 | 8.44 |
| 319 | 0 | 0 | 0 | 0.4 | 0.4 |
| TOTAL | 6.42 | 6.43 | 4.83 | 6.83 | 24.51 |

STATE: HAWAII

| ZIP CODE | QUARTER 1 | QUARTER 2 | QUARTER 3 | QUARTER 4 | TOTAL GRAMS |
|---|---|---|---|---|---|
| 967 | 2.41 | 2.01 | 1.61 | 1.61 | 7.64 |
| 968 | 1.21 | 1.21 | 1.21 | 1.21 | 4.84 |
| TOTAL | 3.62 | 3.22 | 2.82 | 2.82 | 12.48 |

STATE: IDAHO

| ZIP CODE | QUARTER 1 | QUARTER 2 | QUARTER 3 | QUARTER 4 | TOTAL GRAMS |
|---|---|---|---|---|---|
| 836 | 0.8 | 0 | 0 | 0 | 0.8 |
| 838 | 3.62 | 5.63 | 4.02 | 4.02 | 17.29 |
| TOTAL | 4.42 | 5.63 | 4.02 | 4.02 | 18.09 |

STATE: ILLINOIS

| ZIP CODE | QUARTER 1 | QUARTER 2 | QUARTER 3 | QUARTER 4 | TOTAL GRAMS |
|---|---|---|---|---|---|
| 601 | 1.61 | 1.21 | 1.61 | 0.8 | 5.23 |
| 605 | 0.8 | 0.4 | 1.21 | 0 | 2.41 |
| 606 | 0.4 | 0.8 | 1.21 | 0.8 | 3.21 |
| 620 | 0.4 | 1.61 | 0.8 | 1.21 | 4.02 |
| TOTAL | 3.21 | 4.02 | 4.83 | 2.81 | 14.87 |

STATE: INDIANA

| ZIP CODE | QUARTER 1 | QUARTER 2 | QUARTER 3 | QUARTER 4 | TOTAL GRAMS |
|---|---|---|---|---|---|
| 460 | 1.61 | 1.21 | 4.42 | 4.42 | 11.66 |
| 465 | 0 | 0.4 | 0.4 | 0 | 0.8 |
| 467 | 1.21 | 1.61 | 1.21 | 0.8 | 4.83 |
| 471 | 0.8 | 0.8 | 0 | 0 | 1.6 |
| 474 | 0.4 | 0 | 0 | 0 | 0.4 |
| 477 | 0 | 0 | 0.4 | 0.4 | 0.8 |
| TOTAL | 4.02 | 4.02 | 6.43 | 5.62 | 20.09 |

STATE: IOWA

| ZIP CODE | QUARTER 1 | QUARTER 2 | QUARTER 3 | QUARTER 4 | TOTAL GRAMS |
|---|---|---|---|---|---|

| | | | | | |
|---|---|---|---|---|---|
| 502 | 5.63 | 7.64 | 6.03 | 7.64 | 26.94 |
| 503 | 2.01 | 1.21 | 0.8 | 1.21 | 5.23 |

DRUG CODE:1105BDRUG NAME:DL-METHAMPHETAMINE RACEMIC BASE
STATE:IOWA

| ZIP CODE | QUARTER 1 | QUARTER 2 | QUARTER 3 | QUARTER 4 | TOTAL GRAMS |
|---|---|---|---|---|---|
| 505 | 0.4 | 0.8 | 0 | 0 | 1.2 |
| TOTAL | 8.04 | 9.65 | 6.83 | 8.85 | 33.37 |

STATE: KANSAS

| ZIP CODE | QUARTER 1 | QUARTER 2 | QUARTER 3 | QUARTER 4 | TOTAL GRAMS |
|---|---|---|---|---|---|
| 666 | 0 | 0 | 0.4 | 0 | 0.4 |
| 672 | 1.61 | 2.01 | 1.21 | 0.8 | 5.63 |
| TOTAL | 1.61 | 2.01 | 1.61 | 0.8 | 6.03 |

STATE: KENTUCKY

| ZIP CODE | QUARTER 1 | QUARTER 2 | QUARTER 3 | QUARTER 4 | TOTAL GRAMS |
|---|---|---|---|---|---|
| 408 | 2.41 | 1.61 | 1.61 | 1.61 | 7.24 |
| 420 | 0 | 0 | 0.8 | 0.8 | 1.6 |
| 424 | 1.61 | 0 | 0 | 0 | 1.61 |
| TOTAL | 4.02 | 1.61 | 2.41 | 2.41 | 10.45 |

STATE: LOUISIANA

| ZIP CODE | QUARTER 1 | QUARTER 2 | QUARTER 3 | QUARTER 4 | TOTAL GRAMS |
|---|---|---|---|---|---|
| 700 | 0.4 | 0 | 0.4 | 0 | 0.8 |
| 703 | 0 | 0 | 0.4 | 0 | 0.4 |
| 704 | 1.61 | 1.61 | 0 | 0 | 3.22 |
| 706 | 0 | 0 | 0 | 0.4 | 0.4 |
| 708 | 0 | 0 | 0 | 0.4 | 0.4 |
| 710 | 0.4 | 0 | 0 | 0 | 0.4 |
| 713 | 1.21 | 2.01 | 0.8 | 1.61 | 5.63 |
| TOTAL | 3.62 | 3.62 | 1.6 | 2.41 | 11.25 |

STATE: MAINE

| ZIP CODE | QUARTER 1 | QUARTER 2 | QUARTER 3 | QUARTER 4 | TOTAL GRAMS |
|---|---|---|---|---|---|
| 40 | 2.41 | 0 | 0 | 0 | 2.41 |
| 41 | 0 | 0 | 0.4 | 0 | 0.4 |
| TOTAL | 2.41 | 0 | 0.4 | 0 | 2.81 |

STATE: MARYLAND

| ZIP CODE | QUARTER 1 | QUARTER 2 | QUARTER 3 | QUARTER 4 | TOTAL GRAMS |
|---|---|---|---|---|---|
| 208 | 2.81 | 2.81 | 1.61 | 4.82 | 12.05 |
| TOTAL | 2.81 | 2.81 | 1.61 | 4.82 | 12.05 |

STATE: MASSACHUSETTS

| ZIP CODE | QUARTER 1 | QUARTER 2 | QUARTER 3 | QUARTER 4 | TOTAL GRAMS |
|---|---|---|---|---|---|
| 10 | 2.41 | 2.01 | 2.01 | 2.41 | 8.84 |
| 18 | 2.41 | 0.4 | 0.8 | 0 | 3.61 |
| 19 | 0.4 | 0.4 | 0 | 0 | 0.8 |
| 21 | 0.8 | 1.21 | 1.61 | 0.4 | 4.02 |
| 25 | 0 | 0 | 0.4 | 0 | 0.4 |
| TOTAL | 6.02 | 4.02 | 4.82 | 2.81 | 17.67 |

STATE: MICHIGAN

| ZIP CODE | QUARTER 1 | QUARTER 2 | QUARTER 3 | QUARTER 4 | TOTAL GRAMS |
|---|---|---|---|---|---|
| 480 | 0 | 0 | 1.61 | 0 | 1.61 |
| 481 | 0 | 0 | 0.8 | 0 | 0.8 |
| 483 | 0 | 0 | 1.61 | 0 | 1.61 |

| | | | | | |
|---|---|---|---|---|---|
| 490 | 0.4 | 0 | 0 | 0 | 0.4 |
| 495 | 0.4 | 0 | 0 | 0 | 0.4 |
| TOTAL | 0.8 | 0 | 4.02 | 0 | 4.82 |

STATE: MINNESOTA

| ZIP CODE | QUARTER 1 | QUARTER 2 | QUARTER 3 | QUARTER 4 | TOTAL GRAMS |
|---|---|---|---|---|---|
| 551 | 9.24 | 10.05 | 9.64 | 8.44 | 37.37 |
| 554 | 24.91 | 6.83 | 21.7 | 23.31 | 76.75 |
| 557 | 1.61 | 1.21 | 4.42 | 1.61 | 8.85 |
| 558 | 0 | 0.4 | 0 | 0 | 0.4 |
| 563 | 1.61 | 1.61 | 1.61 | 0.8 | 5.63 |
| 564 | 2.41 | 3.62 | 3.62 | 3.62 | 13.27 |
| TOTAL | 39.78 | 23.72 | 40.99 | 37.78 | 142.27 |

STATE: MISSISSIPPI

| ZIP CODE | QUARTER 1 | QUARTER 2 | QUARTER 3 | QUARTER 4 | TOTAL GRAMS |
|---|---|---|---|---|---|
| 395 | 0 | 0.4 | 0 | 0 | 0.4 |
| 397 | 0.4 | 1.21 | 0.4 | 0.4 | 2.41 |
| TOTAL | 0.4 | 1.61 | 0.4 | 0.4 | 2.81 |

STATE: MISSOURI

| ZIP CODE | QUARTER 1 | QUARTER 2 | QUARTER 3 | QUARTER 4 | TOTAL GRAMS |
|---|---|---|---|---|---|
| 630 | 0.8 | 0 | 0 | 0 | 0.8 |
| 631 | 21.3 | 28.93 | 8.04 | 12.06 | 70.33 |

DRUG CODE:1105BDRUG NAME:DL-METHAMPHETAMINE RACEMIC BASE
STATE:MISSOURI

| ZIP CODE | QUARTER 1 | QUARTER 2 | QUARTER 3 | QUARTER 4 | TOTAL GRAMS |
|---|---|---|---|---|---|
| 633 | 0 | 0.4 | 3.62 | 0 | 4.02 |
| 634 | 0.8 | 0 | 0 | 0 | 0.8 |
| 636 | 0 | 0 | 0 | 0.8 | 0.8 |
| 638 | 1.21 | 0 | 0 | 0 | 1.21 |
| 639 | 1.21 | 1.61 | 0.8 | 0.4 | 4.02 |
| 641 | 0 | 0 | 0 | 0.8 | 0.8 |
| 645 | 0 | 0 | 0 | 3.21 | 3.21 |
| 651 | 0 | 0 | 0.4 | 0 | 0.4 |
| 652 | 0 | 1.21 | 1.21 | 0.4 | 2.82 |
| 655 | 0 | 0.8 | 1.61 | 2.01 | 4.42 |
| 658 | 3.21 | 0.4 | 0 | 0 | 3.61 |
| TOTAL | 28.53 | 33.35 | 15.68 | 19.68 | 97.24 |

STATE: NEBRASKA

| ZIP CODE | QUARTER 1 | QUARTER 2 | QUARTER 3 | QUARTER 4 | TOTAL GRAMS |
|---|---|---|---|---|---|
| 680 | 0 | 0.8 | 0 | 0 | 0.8 |
| 681 | 4.02 | 2.81 | 3.21 | 2.01 | 12.05 |
| 685 | 0 | 0.4 | 0 | 0 | 0.4 |
| TOTAL | 4.02 | 4.01 | 3.21 | 2.01 | 13.25 |

STATE: NEVADA

| ZIP CODE | QUARTER 1 | QUARTER 2 | QUARTER 3 | QUARTER 4 | TOTAL GRAMS |
|---|---|---|---|---|---|
| 891 | 4.42 | 2.01 | 1.21 | 3.21 | 10.85 |
| 895 | 0.8 | 0.8 | 0.4 | 1.21 | 3.21 |
| TOTAL | 5.22 | 2.81 | 1.61 | 4.42 | 14.06 |

STATE: NEW HAMPSHIRE

| ZIP CODE | QUARTER 1 | QUARTER 2 | QUARTER 3 | QUARTER 4 | TOTAL GRAMS |
|---|---|---|---|---|---|
| 30 | 0.4 | 0 | 0 | 0 | 0.4 |
| 32 | 0 | 0.8 | 0 | 0 | 0.8 |
| 37 | 0 | 0 | 0.8 | 0.4 | 1.2 |

RETAIL DRUG DISTRIBUTION BY ZIP CODE WITHIN STATE BY GRAMS WT

| | | | | | |
|---|---|---|---|---|---|
| TOTAL | 0.4 | 0.8 | 0.8 | 0.4 | 2.4 |

STATE: NEW JERSEY

| ZIP CODE | QUARTER 1 | QUARTER 2 | QUARTER 3 | QUARTER 4 | TOTAL GRAMS |
|---|---|---|---|---|---|
| 70 | 3.62 | 0.8 | 1.21 | 3.21 | 8.84 |
| 74 | 5.63 | 7.23 | 0 | 1.21 | 14.07 |
| 76 | 2.01 | 1.61 | 0 | 0 | 3.62 |
| 77 | 0 | 0.4 | 0 | 0 | 0.4 |
| 79 | 1.61 | 1.21 | 0.8 | 0.8 | 4.42 |
| 81 | 0.8 | 0 | 0 | 0 | 0.8 |
| 85 | 0 | 0 | 0 | 0.4 | 0.4 |
| 87 | 0 | 0 | 0.8 | 0 | 0.8 |
| TOTAL | 13.67 | 11.25 | 2.81 | 5.62 | 33.35 |

STATE: NEW MEXICO

| ZIP CODE | QUARTER 1 | QUARTER 2 | QUARTER 3 | QUARTER 4 | TOTAL GRAMS |
|---|---|---|---|---|---|
| 871 | 14.47 | 9.64 | 10.05 | 11.65 | 45.81 |
| 875 | 0.4 | 0 | 0.8 | 0 | 1.2 |
| 880 | 0.4 | 0 | 0.4 | 1.21 | 2.01 |
| 882 | 0 | 0.4 | 0 | 0 | 0.4 |
| 883 | 0.4 | 0 | 0 | 2.01 | 2.41 |
| TOTAL | 15.67 | 10.04 | 11.25 | 14.87 | 51.83 |

STATE: NEW YORK

| ZIP CODE | QUARTER 1 | QUARTER 2 | QUARTER 3 | QUARTER 4 | TOTAL GRAMS |
|---|---|---|---|---|---|
| 100 | 4.02 | 3.62 | 4.42 | 3.62 | 15.68 |
| 105 | 0.4 | 0.8 | 0.8 | 0.4 | 2.4 |
| 106 | 0.4 | 0.8 | 0.8 | 0.8 | 2.8 |
| 107 | 0 | 0.8 | 0 | 0 | 0.8 |
| 108 | 0.4 | 0 | 0 | 0 | 0.4 |
| 109 | 0 | 0.4 | 0.4 | 0 | 0.8 |
| 112 | 0.4 | 0 | 0 | 0 | 0.4 |
| 113 | 2.41 | 3.21 | 2.81 | 2.81 | 11.24 |
| 115 | 0.8 | 0 | 0 | 0 | 0.8 |
| 117 | 0.4 | 2.41 | 2.41 | 6.83 | 12.05 |
| 118 | 4.02 | 2.81 | 4.02 | 2.41 | 13.26 |
| 130 | 0 | 0.4 | 0 | 0.8 | 1.2 |
| 142 | 0.4 | 0 | 0 | 0 | 0.4 |
| 143 | 0 | 0 | 0 | 0.4 | 0.4 |
| TOTAL | 13.65 | 15.25 | 15.66 | 18.07 | 62.63 |

STATE: NORTH CAROLINA

DRUG CODE:1105BDRUG NAME:DL-METHAMPHETAMINE RACEMIC BASE
STATE:NORTH CAROLINA

| ZIP CODE | QUARTER 1 | QUARTER 2 | QUARTER 3 | QUARTER 4 | TOTAL GRAMS |
|---|---|---|---|---|---|
| 275 | 3.62 | 3.62 | 4.42 | 3.62 | 15.28 |
| 276 | 0.4 | 0 | 1.61 | 2.01 | 4.02 |
| 280 | 2.41 | 2.01 | 0.8 | 1.61 | 6.83 |
| 281 | 1.61 | 2.41 | 2.81 | 0.8 | 7.63 |
| 282 | 2.01 | 2.01 | 0 | 0 | 4.02 |
| 283 | 0 | 0 | 0 | 0.4 | 0.4 |
| 284 | 0.4 | 0 | 0 | 0 | 0.4 |
| 286 | 2.01 | 2.01 | 2.41 | 1.61 | 8.04 |
| 288 | 2.01 | 2.01 | 1.61 | 1.21 | 6.84 |
| TOTAL | 14.47 | 14.07 | 13.66 | 11.26 | 53.46 |

STATE: NORTH DAKOTA

| ZIP CODE | QUARTER 1 | QUARTER 2 | QUARTER 3 | QUARTER 4 | TOTAL GRAMS |
|---|---|---|---|---|---|

| | | | | | | |
|---|---|---|---|---|---|---|
| | 581 | 1.21 | 1.21 | 1.21 | 1.21 | 4.84 |
| TOTAL | | 1.21 | 1.21 | 1.21 | 1.21 | 4.84 |
| STATE: | OHIO | | | | | |
| ZIP CODE | QUARTER 1 | | QUARTER 2 | QUARTER 3 | QUARTER 4 | TOTAL GRAMS |
| | 432 | 6.03 | 7.23 | 1.61 | 4.82 | 19.69 |
| | 450 | 2.01 | 1.61 | 3.62 | 2.01 | 9.25 |
| | 452 | 0.8 | 0.4 | 0.8 | 0.4 | 2.4 |
| | 454 | 0 | 0 | 0.4 | 0 | 0.4 |
| | 457 | 1.61 | 0.8 | 1.61 | 1.61 | 5.63 |
| TOTAL | | 10.45 | 10.04 | 8.04 | 8.84 | 37.37 |
| STATE: | OKLAHOMA | | | | | |
| ZIP CODE | QUARTER 1 | | QUARTER 2 | QUARTER 3 | QUARTER 4 | TOTAL GRAMS |
| | 730 | 0 | 0 | 0.4 | 0 | 0.4 |
| | 731 | 0 | 0.8 | 0 | 0.4 | 1.2 |
| | 740 | 2.81 | 4.42 | 4.42 | 2.81 | 14.46 |
| | 741 | 3.62 | 3.21 | 3.21 | 2.01 | 12.05 |
| | 747 | 0.8 | 0 | 0 | 0 | 0.8 |
| TOTAL | | 7.23 | 8.43 | 8.03 | 5.22 | 28.91 |
| STATE: | OREGON | | | | | |
| ZIP CODE | QUARTER 1 | | QUARTER 2 | QUARTER 3 | QUARTER 4 | TOTAL GRAMS |
| | 972 | 1.21 | 2.41 | 2.01 | 2.01 | 7.64 |
| | 973 | 3.21 | 2.81 | 1.61 | 0 | 7.63 |
| | 974 | 0 | 0.4 | 0 | 0 | 0.4 |
| | 975 | 0 | 0.4 | 0 | 0 | 0.4 |
| | 978 | 14.47 | 12.06 | 12.06 | 12.06 | 50.65 |
| TOTAL | | 18.89 | 18.08 | 15.68 | 14.07 | 66.72 |
| STATE: | PENNSYLVANIA | | | | | |
| ZIP CODE | QUARTER 1 | | QUARTER 2 | QUARTER 3 | QUARTER 4 | TOTAL GRAMS |
| | 151 | 1.61 | 2.41 | 1.61 | 0.8 | 6.43 |
| | 186 | 2.01 | 2.41 | 0 | 0 | 4.42 |
| | 187 | 7.64 | 4.02 | 7.64 | 6.03 | 25.33 |
| | 190 | 4.42 | 4.02 | 5.63 | 1.21 | 15.28 |
| | 194 | 0 | 1.21 | 0 | 2.01 | 3.22 |
| TOTAL | | 15.68 | 14.07 | 14.88 | 10.05 | 54.68 |
| STATE: | RHODE ISLAND | | | | | |
| ZIP CODE | QUARTER 1 | | QUARTER 2 | QUARTER 3 | QUARTER 4 | TOTAL GRAMS |
| | 28 | 1.61 | 0.4 | 0.4 | 1.21 | 3.62 |
| | 29 | 1.21 | 0.4 | 0.4 | 0 | 2.01 |
| TOTAL | | 2.82 | 0.8 | 0.8 | 1.21 | 5.63 |
| STATE: | SOUTH CAROLINA | | | | | |
| ZIP CODE | QUARTER 1 | | QUARTER 2 | QUARTER 3 | QUARTER 4 | TOTAL GRAMS |
| | 292 | 2.81 | 2.81 | 4.02 | 2.81 | 12.45 |
| | 296 | 0.8 | 0 | 0 | 0 | 0.8 |
| TOTAL | | 3.61 | 2.81 | 4.02 | 2.81 | 13.25 |
| STATE: | TENNESSEE | | | | | |
| ZIP CODE | QUARTER 1 | | QUARTER 2 | QUARTER 3 | QUARTER 4 | TOTAL GRAMS |
| | 370 | 1.21 | 0 | 0 | 0 | 1.21 |
| | 380 | 0.8 | 0.8 | 0 | 0 | 1.6 |
| TOTAL | | 2.01 | 0.8 | 0 | 0 | 2.81 |
| STATE: | TEXAS | | | | | |
| ZIP CODE | QUARTER 1 | | QUARTER 2 | QUARTER 3 | QUARTER 4 | TOTAL GRAMS |
| | 750 | 3.21 | 2.81 | 1.61 | 0.8 | 8.43 |

| | | | | | |
|---|---|---|---|---|---|
| 752 | 15.27 | 15.27 | 11.65 | 12.86 | 55.05 |
| 756 | 0 | 1.21 | 0.8 | 1.21 | 3.22 |

DRUG CODE:1105BDRUG NAME:DL-METHAMPHETAMINE RACEMIC BASE
STATE:TEXAS

| ZIP CODE | QUARTER 1 | QUARTER 2 | QUARTER 3 | QUARTER 4 | TOTAL GRAMS |
|---|---|---|---|---|---|
| 757 | 4.02 | 2.81 | 2.01 | 2.41 | 11.25 |
| 760 | 3.21 | 6.03 | 4.82 | 4.02 | 18.08 |
| 761 | 2.01 | 0 | 0.8 | 0 | 2.81 |
| 762 | 2.81 | 2.01 | 0.4 | 0 | 5.22 |
| 764 | 2.01 | 1.21 | 0 | 0 | 3.22 |
| 768 | 0.4 | 0 | 0 | 0 | 0.4 |
| 770 | 5.22 | 2.41 | 2.41 | 2.41 | 12.45 |
| 773 | 2.01 | 1.61 | 2.41 | 2.41 | 8.44 |
| 774 | 0 | 0 | 0.4 | 0.4 | 0.8 |
| 775 | 1.21 | 2.81 | 0.8 | 2.81 | 7.63 |
| 778 | 0 | 0.4 | 0 | 0 | 0.4 |
| 781 | 0 | 0.8 | 0 | 0 | 0.8 |
| 782 | 1.21 | 2.01 | 1.61 | 1.61 | 6.44 |
| 784 | 0 | 0 | 1.21 | 0.4 | 1.61 |
| 787 | 4.82 | 2.81 | 2.81 | 0.8 | 11.24 |
| 788 | 4.02 | 2.81 | 4.02 | 3.21 | 14.06 |
| 799 | 0.8 | 0.8 | 0 | 0 | 1.6 |
| TOTAL | 52.23 | 47.81 | 37.76 | 35.35 | 173.15 |

STATE: UTAH

| ZIP CODE | QUARTER 1 | QUARTER 2 | QUARTER 3 | QUARTER 4 | TOTAL GRAMS |
|---|---|---|---|---|---|
| 840 | 1.21 | 0 | 0 | 0 | 1.21 |
| 841 | 0.8 | 0 | 0 | 0 | 0.8 |
| TOTAL | 2.01 | 0 | 0 | 0 | 2.01 |

STATE: VIRGINIA

| ZIP CODE | QUARTER 1 | QUARTER 2 | QUARTER 3 | QUARTER 4 | TOTAL GRAMS |
|---|---|---|---|---|---|
| 222 | 0 | 0.8 | 1.21 | 0 | 2.01 |
| 234 | 0 | 0.8 | 0 | 0 | 0.8 |
| TOTAL | 0 | 1.6 | 1.21 | 0 | 2.81 |

STATE: WASHINGTON

| ZIP CODE | QUARTER 1 | QUARTER 2 | QUARTER 3 | QUARTER 4 | TOTAL GRAMS |
|---|---|---|---|---|---|
| 980 | 0.4 | 0 | 0.4 | 0 | 0.8 |
| 982 | 0 | 0 | 0.4 | 0 | 0.4 |
| 986 | 0.8 | 1.21 | 0.8 | 0.8 | 3.61 |
| 988 | 2.01 | 1.21 | 1.21 | 0 | 4.43 |
| TOTAL | 3.21 | 2.42 | 2.81 | 0.8 | 9.24 |

STATE: WEST VIRGINIA

| ZIP CODE | QUARTER 1 | QUARTER 2 | QUARTER 3 | QUARTER 4 | TOTAL GRAMS |
|---|---|---|---|---|---|
| 258 | 0 | 0.4 | 0.8 | 0 | 1.2 |
| 260 | 2.01 | 2.41 | 2.01 | 2.01 | 8.44 |
| TOTAL | 2.01 | 2.81 | 2.81 | 2.01 | 9.64 |

STATE: WISCONSIN

| ZIP CODE | QUARTER 1 | QUARTER 2 | QUARTER 3 | QUARTER 4 | TOTAL GRAMS |
|---|---|---|---|---|---|
| 530 | 0 | 1.61 | 2.01 | 2.81 | 6.43 |
| 531 | 0 | 0.8 | 0.4 | 0 | 1.2 |
| 532 | 9.24 | 6.83 | 8.04 | 8.04 | 32.15 |
| 535 | 2.01 | 1.61 | 2.81 | 2.01 | 8.44 |
| 537 | 3.21 | 2.81 | 3.21 | 2.81 | 12.04 |
| 543 | 0.4 | 0 | 0 | 0 | 0.4 |

| | | | | | |
|---|---|---|---|---|---|
| 549 | 3.62 | 4.82 | 3.62 | 3.62 | 15.68 |
| TOTAL | 18.48 | 18.48 | 20.09 | 19.29 | 76.34 |

STATE: WYOMING

| ZIP CODE | QUARTER 1 | QUARTER 2 | QUARTER 3 | QUARTER 4 | TOTAL GRAMS |
|---|---|---|---|---|---|
| 829 | 0.4 | 0 | 0 | 0 | 0.4 |
| TOTAL | 0.4 | 0 | 0 | 0 | 0.4 |

DRUG CODE:1105DDRUG NAME:D-METHAMPHETAMINE

STATE: ALABAMA

| ZIP CODE | QUARTER 1 | QUARTER 2 | QUARTER 3 | QUARTER 4 | TOTAL GRAMS |
|---|---|---|---|---|---|
| 350 | 0 | 0 | 0.4 | 0 | 0.4 |
| 351 | 0.4 | 4.02 | 4.02 | 3.62 | 12.06 |
| 352 | 12.45 | 7.23 | 4.42 | 2.01 | 26.11 |
| 355 | 3.62 | 3.62 | 3.21 | 3.62 | 14.07 |
| 356 | 0 | 0.4 | 0 | 0.4 | 0.8 |
| 358 | 0.8 | 2.81 | 1.61 | 2.81 | 8.03 |
| 359 | 0.8 | 0.4 | 0 | 0 | 1.2 |
| 361 | 0.8 | 0.8 | 2.01 | 0 | 3.61 |
| 362 | 4.42 | 4.82 | 4.02 | 4.42 | 17.68 |

DRUG CODE:1105DDRUG NAME:D-METHAMPHETAMINE
STATE:ALABAMA

| ZIP CODE | QUARTER 1 | QUARTER 2 | QUARTER 3 | QUARTER 4 | TOTAL GRAMS |
|---|---|---|---|---|---|
| 366 | 0 | 0 | 0.4 | 0.8 | 1.2 |
| 368 | 0.4 | 0 | 0 | 1.21 | 1.61 |
| TOTAL | 23.69 | 24.1 | 20.09 | 18.89 | 86.77 |

STATE: ALASKA

| ZIP CODE | QUARTER 1 | QUARTER 2 | QUARTER 3 | QUARTER 4 | TOTAL GRAMS |
|---|---|---|---|---|---|
| 995 | 2.41 | 3.62 | 0.8 | 0.8 | 7.63 |
| 997 | 2.01 | 0.4 | 0.8 | 1.61 | 4.82 |
| TOTAL | 4.42 | 4.02 | 1.6 | 2.41 | 12.45 |

STATE: ARIZONA

| ZIP CODE | QUARTER 1 | QUARTER 2 | QUARTER 3 | QUARTER 4 | TOTAL GRAMS |
|---|---|---|---|---|---|
| 850 | 0.8 | 0 | 0.8 | 0.4 | 2 |
| 852 | 13.26 | 11.25 | 10.44 | 13.66 | 48.61 |
| 853 | 0.4 | 2.41 | 1.21 | 1.61 | 5.63 |
| 855 | 1.61 | 1.21 | 1.61 | 1.21 | 5.64 |
| 856 | 0.4 | 1.21 | 0.4 | 1.61 | 3.62 |
| 860 | 1.21 | 0.4 | 0 | 0 | 1.61 |
| TOTAL | 17.68 | 16.48 | 14.46 | 18.49 | 67.11 |

STATE: ARKANSAS

| ZIP CODE | QUARTER 1 | QUARTER 2 | QUARTER 3 | QUARTER 4 | TOTAL GRAMS |
|---|---|---|---|---|---|
| 720 | 1.21 | 0.8 | 0.8 | 0.8 | 3.61 |
| 721 | 0 | 1.21 | 0.8 | 0.8 | 2.81 |
| 722 | 0.4 | 0 | 0.4 | 1.21 | 2.01 |
| 725 | 0.8 | 0 | 0 | 0 | 0.8 |
| 727 | 0.8 | 0.4 | 0.4 | 1.21 | 2.81 |
| 729 | 0 | 1.61 | 0.4 | 0 | 2.01 |
| TOTAL | 3.21 | 4.02 | 2.8 | 4.02 | 14.05 |

STATE: CALIFORNIA

| ZIP CODE | QUARTER 1 | QUARTER 2 | QUARTER 3 | QUARTER 4 | TOTAL GRAMS |
|---|---|---|---|---|---|
| 900 | 9.24 | 14.06 | 16.87 | 13.66 | 53.83 |
| 902 | 3.62 | 6.43 | 5.22 | 6.83 | 22.1 |

| | | | | | |
|---|---|---|---|---|---|
| 904 | 2.81 | 0.4 | 1.21 | 1.21 | 5.63 |
| 907 | 0 | 0 | 0 | 2.01 | 2.01 |
| 908 | 1.61 | 0.8 | 0.8 | 0.8 | 4.01 |
| 910 | 0 | 4.82 | 0.4 | 2.81 | 8.03 |
| 912 | 0 | 0 | 0 | 2.01 | 2.01 |
| 913 | 24.1 | 20.49 | 25.31 | 24.91 | 94.81 |
| 914 | 6.83 | 3.21 | 12.05 | 6.03 | 28.12 |
| 916 | 0 | 0 | 0 | 0.4 | 0.4 |
| 917 | 4.42 | 0.4 | 4.42 | 2.81 | 12.05 |
| 919 | 2.41 | 3.21 | 2.81 | 3.21 | 11.64 |
| 920 | 43.79 | 32.94 | 43.38 | 48.2 | 168.31 |
| 921 | 5.22 | 4.42 | 7.63 | 11.25 | 28.52 |
| 922 | 7.63 | 8.84 | 4.02 | 5.22 | 25.71 |
| 923 | 2.01 | 0 | 0 | 0 | 2.01 |
| 925 | 7.23 | 3.62 | 5.22 | 9.24 | 25.31 |
| 926 | 20.89 | 32.14 | 26.91 | 15.26 | 95.2 |
| 927 | 8.03 | 5.22 | 12.45 | 5.22 | 30.92 |
| 928 | 4.42 | 7.63 | 6.43 | 8.03 | 26.51 |
| 930 | 1.61 | 5.22 | 4.02 | 4.42 | 15.27 |
| 931 | 4.42 | 5.22 | 3.62 | 1.61 | 14.87 |
| 933 | 0.4 | 0 | 0.8 | 0.4 | 1.6 |
| 934 | 1.61 | 1.21 | 0.8 | 1.61 | 5.23 |
| 935 | 3.62 | 2.41 | 3.62 | 1.21 | 10.86 |
| 936 | 0 | 0 | 0.8 | 2.01 | 2.81 |
| 937 | 5.62 | 1.61 | 3.21 | 3.62 | 14.06 |
| 939 | 0 | 0 | 0 | 1.61 | 1.61 |
| 940 | 10.44 | 10.44 | 13.26 | 9.64 | 43.78 |
| 941 | 8.03 | 16.47 | 16.87 | 12.05 | 53.42 |
| 943 | 0 | 0.08 | 0.8 | 0.4 | 1.28 |
| 945 | 6.43 | 9.24 | 8.44 | 8.84 | 32.95 |
| 946 | 3.62 | 8.03 | 5.22 | 4.82 | 21.69 |
| 947 | 1.21 | 1.61 | 0 | 0 | 2.82 |
| 949 | 0.8 | 2.81 | 5.22 | 3.21 | 12.04 |
| 950 | 8.03 | 10.44 | 16.87 | 10.85 | 46.19 |
| 951 | 5.62 | 4.82 | 5.62 | 12.45 | 28.51 |
| 953 | 1.21 | 2.41 | 1.21 | 0.8 | 5.63 |

DRUG CODE:1105DDRUG NAME:D-METHAMPHETAMINE
STATE:CALIFORNIA

| ZIP CODE | QUARTER 1 | QUARTER 2 | QUARTER 3 | QUARTER 4 | TOTAL GRAMS |
|---|---|---|---|---|---|
| 954 | 1.21 | 0.4 | 0.4 | 0 | 2.01 |
| 955 | 1.21 | 1.21 | 0.8 | 1.21 | 4.43 |
| 956 | 6.83 | 5.62 | 7.63 | 7.23 | 27.31 |
| 957 | 3.21 | 1.21 | 2.81 | 5.22 | 12.45 |
| 958 | 8.44 | 6.03 | 6.83 | 8.03 | 29.33 |
| 959 | 13.26 | 10.04 | 11.65 | 11.25 | 46.2 |
| 960 | 0 | 0 | 2.01 | 0.8 | 2.81 |
| TOTAL | 251.09 | 255.16 | 297.64 | 282.4 | 1,086.29 |

STATE: COLORADO

| ZIP CODE | QUARTER 1 | QUARTER 2 | QUARTER 3 | QUARTER 4 | TOTAL GRAMS |
|---|---|---|---|---|---|
| 800 | 3.21 | 11.65 | 7.63 | 9.24 | 31.73 |
| 801 | 16.07 | 15.26 | 18.48 | 15.26 | 65.07 |
| 802 | 4.42 | 9.64 | 11.65 | 6.83 | 32.54 |
| 803 | 1.21 | 1.21 | 0.4 | 0.4 | 3.22 |
| 804 | 0.8 | 0.8 | 1.21 | 0 | 2.81 |
| 805 | 0.8 | 1.21 | 2.41 | 4.02 | 8.44 |
| 809 | 2.41 | 2.81 | 0.8 | 1.21 | 7.23 |
| 810 | 2.01 | 2.01 | 0.8 | 0.4 | 5.22 |
| 812 | 0.4 | 2.41 | 0.8 | 0.8 | 4.41 |

| | | | | | |
|---|---|---|---|---|---|
| 815 | 0 | 0 | 0 | 0.8 | 0.8 |
| 816 | 0.8 | 2.81 | 2.81 | 2.01 | 8.43 |
| TOTAL | 32.13 | 49.81 | 46.99 | 40.97 | 169.9 |

STATE: CONNECTICUT

| ZIP CODE | QUARTER 1 | QUARTER 2 | QUARTER 3 | QUARTER 4 | TOTAL GRAMS |
|---|---|---|---|---|---|
| 60 | 6.03 | 9.24 | 5.62 | 6.03 | 26.92 |
| 61 | 0.8 | 0.8 | 4.02 | 2.81 | 8.43 |
| 64 | 0 | 1.21 | 0.4 | 1.21 | 2.82 |
| 65 | 0 | 0 | 1.21 | 2.01 | 3.22 |
| 66 | 0 | 0 | 0 | 0.8 | 0.8 |
| 67 | 0.4 | 1.61 | 1.21 | 0.4 | 3.62 |
| 68 | 5.22 | 5.22 | 4.82 | 4.82 | 20.08 |
| TOTAL | 12.45 | 18.08 | 17.28 | 18.08 | 65.89 |

STATE: DELAWARE

| ZIP CODE | QUARTER 1 | QUARTER 2 | QUARTER 3 | QUARTER 4 | TOTAL GRAMS |
|---|---|---|---|---|---|
| 197 | 0.8 | 0.8 | 0.4 | 0.8 | 2.8 |
| 198 | 2.41 | 2.81 | 2.41 | 3.21 | 10.84 |
| 199 | 1.21 | 2.01 | 1.61 | 2.01 | 6.84 |
| TOTAL | 4.42 | 5.62 | 4.42 | 6.02 | 20.48 |

STATE: DISTRICT OF COLUMBIA

| ZIP CODE | QUARTER 1 | QUARTER 2 | QUARTER 3 | QUARTER 4 | TOTAL GRAMS |
|---|---|---|---|---|---|
| 200 | 2.01 | 0 | 0 | 1.61 | 3.62 |
| TOTAL | 2.01 | 0 | 0 | 1.61 | 3.62 |

STATE: FLORIDA

| ZIP CODE | QUARTER 1 | QUARTER 2 | QUARTER 3 | QUARTER 4 | TOTAL GRAMS |
|---|---|---|---|---|---|
| 320 | 2.41 | 3.21 | 2.01 | 1.21 | 8.84 |
| 322 | 3.62 | 3.21 | 3.21 | 4.02 | 14.06 |
| 324 | 0 | 0 | 0.8 | 0.4 | 1.2 |
| 325 | 0 | 0 | 0 | 0.4 | 0.4 |
| 326 | 0.4 | 0.8 | 1.61 | 0.8 | 3.61 |
| 327 | 0.4 | 2.41 | 1.61 | 0.4 | 4.82 |
| 328 | 0.4 | 0 | 0.4 | 0 | 0.8 |
| 329 | 0.8 | 1.21 | 0.8 | 1.61 | 4.42 |
| 330 | 1.21 | 3.62 | 4.82 | 1.21 | 10.86 |
| 331 | 0.8 | 1.61 | 1.61 | 1.61 | 5.63 |
| 333 | 0 | 0 | 0 | 0.4 | 0.4 |
| 334 | 1.61 | 2.41 | 1.61 | 0 | 5.63 |
| 335 | 0.4 | 0 | 0.4 | 0.4 | 1.2 |
| 336 | 0 | 0.8 | 0 | 0 | 0.8 |
| 337 | 0 | 0.4 | 0 | 0 | 0.4 |
| 339 | 1.61 | 2.01 | 1.21 | 2.41 | 7.24 |
| 341 | 0.8 | 0.4 | 0 | 3.21 | 4.41 |
| 342 | 2.41 | 2.41 | 0.8 | 2.81 | 8.43 |
| 347 | 0 | 0.8 | 0 | 0.4 | 1.2 |
| 349 | 0 | 0 | 0.4 | 0 | 0.4 |
| TOTAL | 16.87 | 25.3 | 21.29 | 21.29 | 84.75 |

STATE: GEORGIA

DRUG CODE:1105DDRUG NAME:D-METHAMPHETAMINE
STATE:GEORGIA

| ZIP CODE | QUARTER 1 | QUARTER 2 | QUARTER 3 | QUARTER 4 | TOTAL GRAMS |
|---|---|---|---|---|---|
| 300 | 7.63 | 9.24 | 8.44 | 4.82 | 30.13 |
| 301 | 4.02 | 6.83 | 6.83 | 7.23 | 24.91 |
| 302 | 0.4 | 0 | 0 | 0 | 0.4 |
| 303 | 6.03 | 8.84 | 12.45 | 8.44 | 35.76 |

| | | | | | |
|---|---|---|---|---|---|
| | 304 | 1.21 | 2.41 | 2.41 | 2.01 | 8.04 |
| | 306 | 0.8 | 0.8 | 1.61 | 0.8 | 4.01 |
| | 307 | 0 | 0 | 0 | 0.4 | 0.4 |
| | 308 | 1.21 | 0 | 0 | 0 | 1.21 |
| | 309 | 2.81 | 2.01 | 2.81 | 2.81 | 10.44 |
| | 310 | 0.4 | 0 | 0 | 0 | 0.4 |
| | 313 | 0.4 | 0.4 | 0.8 | 0.4 | 2 |
| | 316 | 0.4 | 0 | 0.4 | 0 | 0.8 |
| | 317 | 0 | 0 | 0 | 0.8 | 0.8 |
| | 319 | 0.4 | 0 | 0.4 | 0 | 0.8 |
| TOTAL | | 25.71 | 30.53 | 36.15 | 27.71 | 120.1 |

STATE: HAWAII

| ZIP CODE | QUARTER 1 | QUARTER 2 | QUARTER 3 | QUARTER 4 | TOTAL GRAMS |
|---|---|---|---|---|---|
| 967 | 0.8 | 0.8 | 2.41 | 2.81 | 6.82 |
| 968 | 1.61 | 2.41 | 1.61 | 2.41 | 8.04 |
| TOTAL | 2.41 | 3.21 | 4.02 | 5.22 | 14.86 |

STATE: IDAHO

| ZIP CODE | QUARTER 1 | QUARTER 2 | QUARTER 3 | QUARTER 4 | TOTAL GRAMS |
|---|---|---|---|---|---|
| 833 | 0.8 | 1.21 | 1.21 | 1.61 | 4.83 |
| 834 | 0 | 0.4 | 2.01 | 0 | 2.41 |
| 836 | 0.8 | 2.81 | 2.41 | 1.61 | 7.63 |
| 837 | 4.42 | 2.01 | 1.21 | 1.21 | 8.85 |
| 838 | 0.4 | 0 | 0.4 | 0 | 0.8 |
| TOTAL | 6.42 | 6.43 | 7.24 | 4.43 | 24.52 |

STATE: ILLINOIS

| ZIP CODE | QUARTER 1 | QUARTER 2 | QUARTER 3 | QUARTER 4 | TOTAL GRAMS |
|---|---|---|---|---|---|
| 600 | 15.26 | 17.67 | 17.27 | 18.08 | 68.28 |
| 601 | 10.04 | 4.02 | 2.41 | 3.21 | 19.68 |
| 603 | 0.8 | 0.4 | 0 | 0 | 1.2 |
| 604 | 1.61 | 2.01 | 1.61 | 1.61 | 6.84 |
| 605 | 2.01 | 16.47 | 15.67 | 12.05 | 46.2 |
| 606 | 8.03 | 12.85 | 12.05 | 11.25 | 44.18 |
| 607 | 0 | 1.21 | 0 | 2.01 | 3.22 |
| 614 | 0 | 0 | 0 | 0.8 | 0.8 |
| 616 | 0.4 | 0 | 1.21 | 0 | 1.61 |
| 620 | 0.8 | 0 | 0 | 0 | 0.8 |
| 623 | 2.01 | 2.01 | 2.01 | 0 | 6.03 |
| 627 | 0.8 | 0.8 | 4.02 | 3.21 | 8.83 |
| TOTAL | 41.76 | 57.44 | 56.25 | 52.22 | 207.67 |

STATE: INDIANA

| ZIP CODE | QUARTER 1 | QUARTER 2 | QUARTER 3 | QUARTER 4 | TOTAL GRAMS |
|---|---|---|---|---|---|
| 460 | 5.22 | 3.62 | 6.03 | 4.82 | 19.69 |
| 462 | 2.01 | 0.8 | 0.8 | 0.4 | 4.01 |
| 463 | 0.8 | 0.8 | 0 | 0 | 1.6 |
| 465 | 0.4 | 0 | 0.4 | 0 | 0.8 |
| 468 | 0 | 0 | 0 | 0.4 | 0.4 |
| 469 | 1.61 | 0.8 | 3.21 | 7.23 | 12.85 |
| 471 | 0 | 4.42 | 0.4 | 0.4 | 5.22 |
| 474 | 7.23 | 6.03 | 6.83 | 4.42 | 24.51 |
| 477 | 0.4 | 0.4 | 0 | 0 | 0.8 |
| 478 | 0 | 0 | 0.4 | 0.8 | 1.2 |
| 479 | 0 | 0 | 0 | 0.8 | 0.8 |
| TOTAL | 17.67 | 16.87 | 18.07 | 19.27 | 71.88 |

STATE: IOWA

| ZIP CODE | QUARTER 1 | QUARTER 2 | QUARTER 3 | QUARTER 4 | TOTAL GRAMS |
|---|---|---|---|---|---|
| 501 | 1.61 | 0 | 0 | 0 | 1.61 |

| | | | | | |
|---|---|---|---|---|---|
| 503 | 2.01 | 2.01 | 3.21 | 0.8 | 8.03 |
| 504 | 2.41 | 3.21 | 3.21 | 4.02 | 12.85 |
| 515 | 0.8 | 2.41 | 4.02 | 2.01 | 9.24 |
| 516 | 0 | 0 | 2.41 | 1.21 | 3.62 |
| 520 | 0 | 0 | 0.4 | 0.4 | 0.8 |
| 522 | 0.8 | 1.61 | 0.8 | 0 | 3.21 |

DRUG CODE:1105DDRUG NAME:D-METHAMPHETAMINE
STATE:IOWA

| ZIP CODE | QUARTER 1 | QUARTER 2 | QUARTER 3 | QUARTER 4 | TOTAL GRAMS |
|---|---|---|---|---|---|
| 525 | 2.41 | 2.81 | 3.62 | 3.62 | 12.46 |
| 528 | 1.61 | 2.41 | 1.61 | 1.61 | 7.24 |
| TOTAL | 11.65 | 14.46 | 19.28 | 13.67 | 59.06 |

STATE: KANSAS

| ZIP CODE | QUARTER 1 | QUARTER 2 | QUARTER 3 | QUARTER 4 | TOTAL GRAMS |
|---|---|---|---|---|---|
| 660 | 3.21 | 3.62 | 6.03 | 4.02 | 16.88 |
| 662 | 0.4 | 2.41 | 0 | 0 | 2.81 |
| 665 | 0.8 | 1.61 | 2.01 | 1.21 | 5.63 |
| 666 | 0.8 | 0.4 | 0.4 | 0 | 1.6 |
| 667 | 0 | 0.8 | 0 | 0 | 0.8 |
| 670 | 0.4 | 0 | 0 | 0 | 0.4 |
| 672 | 6.43 | 4.82 | 6.43 | 4.42 | 22.1 |
| 679 | 0 | 0 | 0.4 | 0.8 | 1.2 |
| TOTAL | 12.04 | 13.66 | 15.27 | 10.45 | 51.42 |

STATE: KENTUCKY

| ZIP CODE | QUARTER 1 | QUARTER 2 | QUARTER 3 | QUARTER 4 | TOTAL GRAMS |
|---|---|---|---|---|---|
| 400 | 0.4 | 0 | 0 | 0 | 0.4 |
| 403 | 1.61 | 0.8 | 1.61 | 1.21 | 5.23 |
| 405 | 1.61 | 1.61 | 1.21 | 0 | 4.43 |
| 410 | 0.8 | 0 | 0 | 1.21 | 2.01 |
| 414 | 0 | 0.8 | 0 | 0 | 0.8 |
| 417 | 0.4 | 1.21 | 0.4 | 0.4 | 2.41 |
| 420 | 0 | 0 | 0.4 | 0.8 | 1.2 |
| 421 | 0.4 | 0.4 | 0 | 0 | 0.8 |
| 424 | 0 | 1.21 | 1.21 | 1.21 | 3.63 |
| TOTAL | 5.22 | 6.03 | 4.83 | 4.83 | 20.91 |

STATE: LOUISIANA

| ZIP CODE | QUARTER 1 | QUARTER 2 | QUARTER 3 | QUARTER 4 | TOTAL GRAMS |
|---|---|---|---|---|---|
| 701 | 0 | 0 | 0.4 | 0 | 0.4 |
| 703 | 2.41 | 2.01 | 2.01 | 1.61 | 8.04 |
| 704 | 0 | 0 | 2.41 | 1.61 | 4.02 |
| 705 | 0 | 0 | 0 | 0.4 | 0.4 |
| 706 | 0 | 1.61 | 0.4 | 0.8 | 2.81 |
| 707 | 0 | 0 | 0 | 0.8 | 0.8 |
| 708 | 4.02 | 23.7 | 4.02 | 5.22 | 36.96 |
| 710 | 2.41 | 2.41 | 2.41 | 2.81 | 10.04 |
| 711 | 0 | 0 | 1.21 | 0 | 1.21 |
| 712 | 0 | 0 | 0.4 | 0 | 0.4 |
| 713 | 0 | 0 | 0.8 | 1.61 | 2.41 |
| TOTAL | 8.84 | 29.73 | 14.06 | 14.86 | 67.49 |

STATE: MAINE

| ZIP CODE | QUARTER 1 | QUARTER 2 | QUARTER 3 | QUARTER 4 | TOTAL GRAMS |
|---|---|---|---|---|---|
| 40 | 0 | 0 | 2.41 | 2.01 | 4.42 |
| 41 | 2.01 | 1.61 | 0 | 0 | 3.62 |

| | | | | | |
|---|---|---|---|---|---|
| 49 | 0.4 | 0 | 0 | 0 | 0.4 |
| TOTAL | 2.41 | 1.61 | 2.41 | 2.01 | 8.44 |

STATE: MARYLAND

| ZIP CODE | QUARTER 1 | QUARTER 2 | QUARTER 3 | QUARTER 4 | TOTAL GRAMS |
|---|---|---|---|---|---|
| 207 | 0 | 0 | 0.4 | 0 | 0.4 |
| 208 | 8.03 | 7.63 | 4.82 | 10.04 | 30.52 |
| 209 | 6.03 | 2.01 | 0.8 | 0 | 8.84 |
| 210 | 12.05 | 12.45 | 9.24 | 14.06 | 47.8 |
| 211 | 8.84 | 8.03 | 8.84 | 8.84 | 34.55 |
| 212 | 13.26 | 12.45 | 16.87 | 12.85 | 55.43 |
| 214 | 0.8 | 0.8 | 0.8 | 0 | 2.4 |
| 217 | 2.41 | 1.21 | 1.21 | 0.8 | 5.63 |
| TOTAL | 51.42 | 44.58 | 42.98 | 46.59 | 185.57 |

STATE: MASSACHUSETTS

| ZIP CODE | QUARTER 1 | QUARTER 2 | QUARTER 3 | QUARTER 4 | TOTAL GRAMS |
|---|---|---|---|---|---|
| 10 | 14.06 | 18.48 | 15.26 | 15.26 | 63.06 |
| 12 | 2.41 | 0.8 | 3.62 | 2.01 | 8.84 |
| 14 | 2.81 | 3.21 | 1.61 | 2.81 | 10.44 |
| 15 | 0.8 | 0 | 0 | 0 | 0.8 |
| 17 | 2.01 | 4.02 | 2.81 | 4.42 | 13.26 |
| 18 | 3.62 | 2.01 | 2.41 | 3.21 | 11.25 |
| 19 | 2.41 | 3.21 | 3.21 | 3.62 | 12.45 |

DRUG CODE:1105DDRUG NAME:D-METHAMPHETAMINE
STATE:MASSACHUSETTS

| ZIP CODE | QUARTER 1 | QUARTER 2 | QUARTER 3 | QUARTER 4 | TOTAL GRAMS |
|---|---|---|---|---|---|
| 21 | 2.01 | 7.23 | 5.22 | 7.63 | 22.09 |
| 23 | 0 | 0 | 0 | 0.4 | 0.4 |
| 24 | 3.21 | 2.01 | 2.81 | 2.41 | 10.44 |
| 27 | 0.8 | 0.8 | 1.21 | 1.21 | 4.02 |
| TOTAL | 34.14 | 41.77 | 38.16 | 42.98 | 157.05 |

STATE: MICHIGAN

| ZIP CODE | QUARTER 1 | QUARTER 2 | QUARTER 3 | QUARTER 4 | TOTAL GRAMS |
|---|---|---|---|---|---|
| 480 | 1.21 | 2.41 | 2.41 | 0.4 | 6.43 |
| 481 | 5.22 | 6.43 | 5.62 | 6.03 | 23.3 |
| 483 | 4.82 | 2.01 | 4.82 | 0 | 11.65 |
| 486 | 0 | 0 | 0 | 0.8 | 0.8 |
| 487 | 7.63 | 7.63 | 4.82 | 4.82 | 24.9 |
| 488 | 0 | 0 | 0.8 | 1.21 | 2.01 |
| 490 | 6.43 | 7.23 | 8.84 | 10.44 | 32.94 |
| 492 | 0 | 0 | 0 | 1.61 | 1.61 |
| 493 | 3.62 | 1.61 | 0 | 2.01 | 7.24 |
| 494 | 0.4 | 0 | 0 | 0 | 0.4 |
| 495 | 0 | 0 | 0.4 | 1.21 | 1.61 |
| 496 | 0 | 0 | 0 | 0.4 | 0.4 |
| TOTAL | 29.33 | 27.32 | 27.71 | 28.93 | 113.29 |

STATE: MINNESOTA

| ZIP CODE | QUARTER 1 | QUARTER 2 | QUARTER 3 | QUARTER 4 | TOTAL GRAMS |
|---|---|---|---|---|---|
| 550 | 0.4 | 0.4 | 0.8 | 0.4 | 2 |
| 551 | 8.44 | 4.42 | 8.84 | 5.22 | 26.92 |
| 553 | 12.85 | 15.67 | 19.68 | 14.06 | 62.26 |
| 554 | 21.69 | 13.66 | 14.46 | 14.06 | 63.87 |
| 557 | 0.8 | 1.61 | 1.21 | 1.21 | 4.83 |
| 559 | 21.69 | 26.91 | 23.3 | 28.12 | 100.02 |
| 560 | 0.8 | 1.61 | 1.21 | 2.41 | 6.03 |

| | | | | | |
|---|---|---|---|---|---|
| 562 | 1.61 | 0.8 | 1.21 | 2.01 | 5.63 |
| 564 | 2.41 | 1.61 | 0 | 2.41 | 6.43 |
| 565 | 1.21 | 2.41 | 1.61 | 2.41 | 7.64 |
| 566 | 1.61 | 0.8 | 0.4 | 0.4 | 3.21 |
| TOTAL | 73.51 | 69.9 | 72.72 | 72.71 | 288.84 |

STATE: MISSISSIPPI

| ZIP CODE | QUARTER 1 | QUARTER 2 | QUARTER 3 | QUARTER 4 | TOTAL GRAMS |
|---|---|---|---|---|---|
| 386 | 0 | 0.4 | 1.21 | 0.8 | 2.41 |
| 391 | 1.21 | 0 | 0 | 0 | 1.21 |
| 392 | 4.42 | 6.03 | 5.62 | 6.83 | 22.9 |
| 394 | 4.42 | 3.21 | 1.21 | 0 | 8.84 |
| 396 | 1.21 | 0 | 1.61 | 1.21 | 4.03 |
| TOTAL | 11.26 | 9.64 | 9.65 | 8.84 | 39.39 |

STATE: MISSOURI

| ZIP CODE | QUARTER 1 | QUARTER 2 | QUARTER 3 | QUARTER 4 | TOTAL GRAMS |
|---|---|---|---|---|---|
| 630 | 0 | 3.62 | 1.61 | 0.8 | 6.03 |
| 631 | 29.73 | 40.97 | 34.95 | 33.74 | 139.39 |
| 634 | 0.4 | 0.8 | 0.8 | 1.61 | 3.61 |
| 636 | 0 | 0 | 0 | 0.4 | 0.4 |
| 641 | 14.86 | 3.21 | 4.42 | 5.22 | 27.71 |
| 645 | 1.21 | 2.81 | 0.8 | 0 | 4.82 |
| 648 | 0 | 0.4 | 0 | 0 | 0.4 |
| 658 | 0.4 | 0.4 | 0.4 | 0.4 | 1.6 |
| TOTAL | 46.6 | 52.21 | 42.98 | 42.17 | 183.96 |

STATE: MONTANA

| ZIP CODE | QUARTER 1 | QUARTER 2 | QUARTER 3 | QUARTER 4 | TOTAL GRAMS |
|---|---|---|---|---|---|
| 591 | 0.8 | 1.61 | 0 | 0 | 2.41 |
| 593 | 0 | 0.4 | 0.4 | 0.4 | 1.2 |
| 596 | 2.01 | 0.8 | 0.8 | 0 | 3.61 |
| 599 | 1.21 | 0.8 | 0.4 | 0 | 2.41 |
| TOTAL | 4.02 | 3.61 | 1.6 | 0.4 | 9.63 |

STATE: NEBRASKA

| ZIP CODE | QUARTER 1 | QUARTER 2 | QUARTER 3 | QUARTER 4 | TOTAL GRAMS |
|---|---|---|---|---|---|
| 680 | 2.41 | 1.61 | 2.01 | 2.01 | 8.04 |
| 681 | 5.22 | 4.42 | 4.82 | 3.21 | 17.67 |
| 685 | 0 | 0.4 | 0 | 0 | 0.4 |
| 686 | 0.8 | 0.8 | 0 | 0 | 1.6 |

DRUG CODE:1105DDRUG NAME:D-METHAMPHETAMINE
STATE:NEBRASKA

| ZIP CODE | QUARTER 1 | QUARTER 2 | QUARTER 3 | QUARTER 4 | TOTAL GRAMS |
|---|---|---|---|---|---|
| 689 | 0.4 | 0 | 0 | 0 | 0.4 |
| TOTAL | 8.83 | 7.23 | 6.83 | 5.22 | 28.11 |

STATE: NEVADA

| ZIP CODE | QUARTER 1 | QUARTER 2 | QUARTER 3 | QUARTER 4 | TOTAL GRAMS |
|---|---|---|---|---|---|
| 890 | 0.8 | 0.8 | 0 | 0 | 1.6 |
| 891 | 14.46 | 18.88 | 18.08 | 18.88 | 70.3 |
| 894 | 0 | 0.4 | 2.01 | 0 | 2.41 |
| 895 | 2.81 | 2.41 | 4.02 | 1.21 | 10.45 |
| 898 | 0 | 0 | 0.4 | 0 | 0.4 |
| TOTAL | 18.07 | 22.49 | 24.51 | 20.09 | 85.16 |

STATE: NEW HAMPSHIRE

RETAIL DRUG DISTRIBUTION BY ZIP CODE WITHIN STATE BY GRAMS WT

| ZIP CODE | QUARTER 1 | QUARTER 2 | QUARTER 3 | QUARTER 4 | TOTAL GRAMS |
|---|---|---|---|---|---|
| 31 | 1.21 | 1.21 | 2.01 | 2.81 | 7.24 |
| 32 | 0 | 0.4 | 0.8 | 0 | 1.2 |
| 37 | 0.8 | 2.41 | 2.01 | 1.61 | 6.83 |
| 38 | 4.02 | 1.21 | 2.41 | 3.29 | 10.93 |
| TOTAL | 6.03 | 5.23 | 7.23 | 7.71 | 26.2 |
| STATE: | NEW JERSEY | | | | |
| ZIP CODE | QUARTER 1 | QUARTER 2 | QUARTER 3 | QUARTER 4 | TOTAL GRAMS |
| 70 | 4.42 | 8.84 | 4.42 | 3.62 | 21.3 |
| 72 | 0 | 0.8 | 0 | 0 | 0.8 |
| 73 | 0.4 | 0 | 0 | 0 | 0.4 |
| 74 | 0.4 | 0 | 0 | 2.01 | 2.41 |
| 76 | 8.03 | 6.83 | 6.43 | 9.64 | 30.93 |
| 77 | 0 | 0 | 1.21 | 2.01 | 3.22 |
| 78 | 5.22 | 4.82 | 4.02 | 4.82 | 18.88 |
| 79 | 0 | 1.21 | 0 | 1.21 | 2.42 |
| 80 | 5.22 | 4.42 | 5.22 | 2.81 | 17.67 |
| 81 | 5.22 | 6.43 | 3.62 | 0.8 | 16.07 |
| 82 | 0.4 | 0.4 | 0.8 | 0 | 1.6 |
| 83 | 0.4 | 0 | 0 | 0 | 0.4 |
| 85 | 0 | 0.8 | 0 | 0 | 0.8 |
| 87 | 0 | 0 | 0 | 0.8 | 0.8 |
| 88 | 3.21 | 3.21 | 2.01 | 3.21 | 11.64 |
| 89 | 0 | 0 | 0 | 1.61 | 1.61 |
| TOTAL | 32.92 | 37.76 | 27.73 | 32.54 | 130.95 |
| STATE: | NEW MEXICO | | | | |
| ZIP CODE | QUARTER 1 | QUARTER 2 | QUARTER 3 | QUARTER 4 | TOTAL GRAMS |
| 870 | 0 | 0.4 | 0 | 0 | 0.4 |
| 871 | 11.25 | 15.67 | 9.24 | 15.26 | 51.42 |
| 874 | 0.8 | 0 | 0 | 0 | 0.8 |
| 875 | 0 | 0.4 | 0 | 0 | 0.4 |
| 880 | 2.41 | 1.21 | 1.61 | 1.21 | 6.44 |
| 882 | 0 | 0 | 0 | 1.21 | 1.21 |
| 883 | 0.4 | 1.21 | 1.21 | 0.4 | 3.22 |
| TOTAL | 14.86 | 18.89 | 12.06 | 18.08 | 63.89 |
| STATE: | NEW YORK | | | | |
| ZIP CODE | QUARTER 1 | QUARTER 2 | QUARTER 3 | QUARTER 4 | TOTAL GRAMS |
| 100 | 13.26 | 12.45 | 13.26 | 16.87 | 55.84 |
| 101 | 2.01 | 2.41 | 0.8 | 0.4 | 5.62 |
| 104 | 0.88 | 0.8 | 0.4 | 0.8 | 2.88 |
| 105 | 3.62 | 4.02 | 2.41 | 2.81 | 12.86 |
| 106 | 11.65 | 16.07 | 9.64 | 12.05 | 49.41 |
| 108 | 0.8 | 0.4 | 0 | 0.8 | 2 |
| 109 | 3.21 | 3.62 | 4.82 | 6.03 | 17.68 |
| 110 | 0 | 0 | 0 | 0.4 | 0.4 |
| 112 | 0 | 0.4 | 0.4 | 0 | 0.8 |
| 114 | 2.41 | 0.8 | 2.41 | 2.41 | 8.03 |
| 115 | 2.41 | 4.42 | 2.41 | 4.02 | 13.26 |
| 117 | 1.21 | 2.01 | 1.21 | 2.81 | 7.24 |
| 119 | 0 | 0.4 | 0.8 | 0.4 | 1.6 |
| 120 | 0 | 0 | 0.4 | 2.41 | 2.81 |
| 121 | 1.21 | 0.8 | 0 | 0 | 2.01 |
| 123 | 1.21 | 3.62 | 1.61 | 3.21 | 9.65 |
| 127 | 0.8 | 1.21 | 0.4 | 1.61 | 4.02 |
| 128 | 2.01 | 0.4 | 1.21 | 1.21 | 4.83 |

RETAIL DRUG DISTRIBUTION BY ZIP CODE WITHIN STATE BY GRAMS WT

STATE:NEW YORK

| ZIP CODE | QUARTER 1 | QUARTER 2 | QUARTER 3 | QUARTER 4 | TOTAL GRAMS |
|---|---|---|---|---|---|
| 130 | 2.01 | 3.62 | 3.62 | 1.61 | 10.86 |
| 131 | 0.4 | 0.4 | 0 | 0.4 | 1.2 |
| 134 | 0.4 | 1.21 | 0 | 0 | 1.61 |
| 139 | 1.61 | 2.41 | 0.4 | 0.8 | 5.22 |
| 140 | 4.02 | 2.81 | 4.02 | 1.21 | 12.06 |
| 141 | 3.62 | 2.41 | 3.62 | 2.01 | 11.66 |
| 142 | 3.21 | 3.21 | 4.82 | 4.82 | 16.06 |
| 144 | 14.46 | 4.82 | 3.21 | 4.02 | 26.51 |
| 146 | 1.21 | 0 | 0 | 0 | 1.21 |
| 148 | 2.01 | 3.21 | 2.01 | 1.61 | 8.84 |
| 149 | 0 | 0 | 0 | 1.21 | 1.21 |
| TOTAL | 79.64 | 77.93 | 63.88 | 75.93 | 297.38 |

STATE: NORTH CAROLINA

| ZIP CODE | QUARTER 1 | QUARTER 2 | QUARTER 3 | QUARTER 4 | TOTAL GRAMS |
|---|---|---|---|---|---|
| 271 | 0 | 0 | 0.8 | 0 | 0.8 |
| 272 | 2.41 | 1.21 | 1.61 | 5.62 | 10.85 |
| 273 | 0 | 0.8 | 0 | 0 | 0.8 |
| 274 | 0 | 0 | 0.4 | 0 | 0.4 |
| 275 | 14.06 | 8.84 | 21.29 | 12.05 | 56.24 |
| 276 | 7.23 | 11.25 | 4.02 | 4.42 | 26.92 |
| 277 | 3.62 | 4.02 | 4.02 | 2.81 | 14.47 |
| 278 | 0 | 0.8 | 1.21 | 0.4 | 2.41 |
| 280 | 5.22 | 3.21 | 5.62 | 3.62 | 17.67 |
| 281 | 6.43 | 6.83 | 6.03 | 8.84 | 28.13 |
| 282 | 5.62 | 2.01 | 2.01 | 2.41 | 12.05 |
| 283 | 2.81 | 3.62 | 2.81 | 3.62 | 12.86 |
| 285 | 0.4 | 1.21 | 1.21 | 0.4 | 3.22 |
| 286 | 0 | 0.4 | 0.8 | 0.8 | 2 |
| 287 | 1.61 | 0.8 | 1.61 | 0.8 | 4.82 |
| TOTAL | 49.41 | 45 | 53.44 | 45.79 | 193.64 |

STATE: NORTH DAKOTA

| ZIP CODE | QUARTER 1 | QUARTER 2 | QUARTER 3 | QUARTER 4 | TOTAL GRAMS |
|---|---|---|---|---|---|
| 581 | 0 | 0 | 0 | 0.4 | 0.4 |
| 584 | 1.61 | 0.8 | 1.21 | 2.81 | 6.43 |
| 587 | 0 | 0 | 0 | 0.4 | 0.4 |
| TOTAL | 1.61 | 0.8 | 1.21 | 3.61 | 7.23 |

STATE: OHIO

| ZIP CODE | QUARTER 1 | QUARTER 2 | QUARTER 3 | QUARTER 4 | TOTAL GRAMS |
|---|---|---|---|---|---|
| 430 | 2.41 | 2.01 | 0.8 | 0 | 5.22 |
| 431 | 0 | 0 | 0.4 | 0.4 | 0.8 |
| 432 | 67.49 | 86.77 | 30.53 | 30.13 | 214.92 |
| 434 | 0 | 0 | 1.21 | 0.8 | 2.01 |
| 437 | 0.8 | 0 | 0 | 0 | 0.8 |
| 440 | 5.62 | 1.61 | 0.8 | 0 | 8.03 |
| 441 | 0.8 | 3.21 | 3.21 | 4.42 | 11.64 |
| 444 | 0 | 0.4 | 0 | 0 | 0.4 |
| 445 | 2.01 | 2.41 | 1.21 | 2.01 | 7.64 |
| 446 | 3.62 | 3.62 | 3.21 | 2.01 | 12.46 |
| 450 | 3.21 | 4.82 | 4.42 | 6.83 | 19.28 |
| 451 | 0.8 | 0 | 2.01 | 0.4 | 3.21 |
| 452 | 1.61 | 0.8 | 1.21 | 0.8 | 4.42 |
| 453 | 2.01 | 1.21 | 0.8 | 1.21 | 5.23 |
| 454 | 6.83 | 4.82 | 5.22 | 3.21 | 20.08 |
| TOTAL | 97.21 | 111.68 | 55.03 | 52.22 | 316.14 |

STATE: OKLAHOMA

RETAIL DRUG DISTRIBUTION BY ZIP CODE WITHIN STATE BY GRAMS WT

| ZIP CODE | QUARTER 1 | QUARTER 2 | QUARTER 3 | QUARTER 4 | TOTAL GRAMS |
|---|---|---|---|---|---|
| 730 | 1.61 | 2.41 | 3.62 | 4.02 | 11.66 |
| 731 | 1.61 | 2.01 | 2.01 | 2.41 | 8.04 |
| 735 | 0 | 1.61 | 2.41 | 2.01 | 6.03 |
| 741 | 0.8 | 0.4 | 0 | 0 | 1.2 |
| 744 | 0.8 | 0 | 0 | 0 | 0.8 |
| 745 | 0 | 0 | 0 | 0.8 | 0.8 |
| 747 | 0.4 | 0 | 0 | 1.21 | 1.61 |
| 748 | 1.21 | 1.61 | 1.21 | 1.61 | 5.64 |
| TOTAL | 6.43 | 8.04 | 9.25 | 12.06 | 35.78 |

STATE: OREGON

DRUG CODE:1105DDRUG NAME:D-METHAMPHETAMINE
STATE:OREGON

| ZIP CODE | QUARTER 1 | QUARTER 2 | QUARTER 3 | QUARTER 4 | TOTAL GRAMS |
|---|---|---|---|---|---|
| 970 | 14.46 | 10.04 | 13.66 | 14.06 | 52.22 |
| 971 | 5.62 | 7.23 | 8.03 | 4.02 | 24.9 |
| 972 | 9.64 | 9.24 | 10.04 | 7.63 | 36.55 |
| 973 | 10.04 | 6.43 | 7.23 | 6.03 | 29.73 |
| 974 | 1.61 | 1.21 | 0.8 | 0.8 | 4.42 |
| 975 | 4.02 | 5.62 | 4.42 | 2.81 | 16.87 |
| 976 | 0.8 | 0 | 0 | 0 | 0.8 |
| 977 | 1.21 | 1.61 | 2.41 | 1.21 | 6.44 |
| 978 | 0.4 | 0.4 | 0.4 | 1.21 | 2.41 |
| TOTAL | 47.8 | 41.78 | 46.99 | 37.77 | 174.34 |

STATE: PENNSYLVANIA

| ZIP CODE | QUARTER 1 | QUARTER 2 | QUARTER 3 | QUARTER 4 | TOTAL GRAMS |
|---|---|---|---|---|---|
| 150 | 4.82 | 6.03 | 6.83 | 4.82 | 22.5 |
| 151 | 2.01 | 4.02 | 5.22 | 2.41 | 13.66 |
| 152 | 14.86 | 15.67 | 14.86 | 15.67 | 61.06 |
| 153 | 2.81 | 2.01 | 1.21 | 0.4 | 6.43 |
| 156 | 0 | 0 | 0 | 0.8 | 0.8 |
| 160 | 4.42 | 4.02 | 1.61 | 2.01 | 12.06 |
| 168 | 2.01 | 3.21 | 2.41 | 5.22 | 12.85 |
| 170 | 0.8 | 0 | 0 | 3.21 | 4.01 |
| 172 | 0.4 | 0 | 0 | 0 | 0.4 |
| 173 | 0.8 | 2.81 | 1.61 | 0.8 | 6.02 |
| 174 | 0.48 | 0 | 0 | 0 | 0.48 |
| 175 | 1.21 | 0.8 | 0.4 | 0 | 2.41 |
| 176 | 0 | 0 | 0 | 1.21 | 1.21 |
| 178 | 0 | 1.21 | 2.81 | 3.62 | 7.64 |
| 180 | 0.8 | 4.02 | 2.01 | 0.4 | 7.23 |
| 181 | 2.01 | 3.21 | 5.22 | 3.21 | 13.65 |
| 183 | 0 | 0 | 0 | 2.01 | 2.01 |
| 184 | 5.62 | 6.43 | 4.42 | 2.41 | 18.88 |
| 185 | 1.61 | 3.21 | 4.82 | 3.62 | 13.26 |
| 186 | 0 | 0 | 1.21 | 2.01 | 3.22 |
| 187 | 37.76 | 53.83 | 39.77 | 38.56 | 169.92 |
| 189 | 3.21 | 4.82 | 3.62 | 3.62 | 15.27 |
| 190 | 7.23 | 10.04 | 8.03 | 6.03 | 31.33 |
| 191 | 11.25 | 10.04 | 10.85 | 12.45 | 44.59 |
| 193 | 1.21 | 1.61 | 0 | 1.21 | 4.03 |
| 194 | 1.61 | 1.61 | 1.21 | 1.61 | 6.04 |
| 195 | 0.8 | 1.61 | 2.01 | 0 | 4.42 |
| 196 | 0 | 0.4 | 0.8 | 0.8 | 2 |
| TOTAL | 107.73 | 140.61 | 120.93 | 118.11 | 487.38 |

STATE: RHODE ISLAND

RETAIL DRUG DISTRIBUTION BY ZIP CODE WITHIN STATE BY GRAMS WT

| ZIP CODE | QUARTER 1 | QUARTER 2 | QUARTER 3 | QUARTER 4 | TOTAL GRAMS |
|---|---|---|---|---|---|
| 28 | 1.21 | 1.21 | 1.61 | 1.61 | 5.64 |
| 29 | 0.8 | 0.8 | 0.8 | 1.21 | 3.61 |
| TOTAL | 2.01 | 2.01 | 2.41 | 2.82 | 9.25 |

STATE: SOUTH CAROLINA

| ZIP CODE | QUARTER 1 | QUARTER 2 | QUARTER 3 | QUARTER 4 | TOTAL GRAMS |
|---|---|---|---|---|---|
| 290 | 2.01 | 2.41 | 2.81 | 1.21 | 8.44 |
| 292 | 0.8 | 1.21 | 2.01 | 5.62 | 9.64 |
| 293 | 1.21 | 0.8 | 0 | 1.61 | 3.62 |
| 294 | 0 | 0.4 | 0 | 0.8 | 1.2 |
| 295 | 0.4 | 0 | 1.61 | 1.61 | 3.62 |
| 296 | 2.81 | 7.63 | 4.42 | 7.23 | 22.09 |
| 299 | 0 | 0 | 0 | 0.8 | 0.8 |
| TOTAL | 7.23 | 12.45 | 10.85 | 18.88 | 49.41 |

STATE: SOUTH DAKOTA

| ZIP CODE | QUARTER 1 | QUARTER 2 | QUARTER 3 | QUARTER 4 | TOTAL GRAMS |
|---|---|---|---|---|---|
| 571 | 9.24 | 5.62 | 9.24 | 5.62 | 29.72 |
| 577 | 4.82 | 3.21 | 4.82 | 6.43 | 19.28 |
| TOTAL | 14.06 | 8.83 | 14.06 | 12.05 | 49 |

STATE: TENNESSEE

| ZIP CODE | QUARTER 1 | QUARTER 2 | QUARTER 3 | QUARTER 4 | TOTAL GRAMS |
|---|---|---|---|---|---|
| 370 | 2.41 | 1.21 | 1.61 | 2.01 | 7.24 |
| 371 | 0.4 | 0 | 0 | 0 | 0.4 |
| 372 | 2.41 | 0 | 0 | 0 | 2.41 |

DRUG CODE:1105DDRUG NAME:D-METHAMPHETAMINE
STATE:TENNESSEE

| ZIP CODE | QUARTER 1 | QUARTER 2 | QUARTER 3 | QUARTER 4 | TOTAL GRAMS |
|---|---|---|---|---|---|
| 373 | 0 | 0.4 | 0 | 0 | 0.4 |
| 376 | 0.8 | 0 | 0 | 0 | 0.8 |
| 377 | 0 | 0.4 | 0 | 0 | 0.4 |
| 378 | 1.61 | 0.8 | 1.61 | 0.8 | 4.82 |
| 379 | 0.4 | 0.8 | 0 | 1.21 | 2.41 |
| 380 | 0 | 0.4 | 0.8 | 1.21 | 2.41 |
| 381 | 6.43 | 6.43 | 6.43 | 4.82 | 24.11 |
| TOTAL | 14.46 | 10.44 | 10.45 | 10.05 | 45.4 |

STATE: TEXAS

| ZIP CODE | QUARTER 1 | QUARTER 2 | QUARTER 3 | QUARTER 4 | TOTAL GRAMS |
|---|---|---|---|---|---|
| 750 | 10.85 | 20.89 | 17.67 | 14.46 | 63.87 |
| 751 | 0 | 0.4 | 0 | 0 | 0.4 |
| 752 | 11.25 | 12.05 | 8.84 | 12.45 | 44.59 |
| 754 | 1.61 | 2.01 | 2.41 | 2.81 | 8.84 |
| 760 | 2.41 | 8.44 | 6.43 | 5.62 | 22.9 |
| 761 | 2.01 | 2.81 | 1.21 | 0.4 | 6.43 |
| 762 | 2.41 | 1.21 | 3.21 | 3.62 | 10.45 |
| 763 | 0 | 0.8 | 1.21 | 1.61 | 3.62 |
| 764 | 0.8 | 0.4 | 1.21 | 1.61 | 4.02 |
| 767 | 0 | 0 | 0 | 0.8 | 0.8 |
| 769 | 0 | 0 | 0.4 | 0 | 0.4 |
| 770 | 1.61 | 4.82 | 6.03 | 6.43 | 18.89 |
| 773 | 0.4 | 0.8 | 0 | 0 | 1.2 |
| 774 | 2.41 | 4.02 | 3.62 | 2.41 | 12.46 |
| 775 | 2.41 | 0.8 | 2.41 | 0 | 5.62 |
| 778 | 2.41 | 0.4 | 0 | 0.4 | 3.21 |
| 780 | 2.01 | 1.61 | 0 | 0.4 | 4.02 |

| | | | | | |
|---|---|---|---|---|---|
| 781 | 0.8 | 0.8 | 2.01 | 0.8 | 4.41 |
| 782 | 6.03 | 5.22 | 4.02 | 1.61 | 16.88 |
| 785 | 0.4 | 3.62 | 5.62 | 2.01 | 11.65 |
| 786 | 3.62 | 3.62 | 2.41 | 3.21 | 12.86 |
| 787 | 8.44 | 4.82 | 6.83 | 9.64 | 29.73 |
| 789 | 1.61 | 0.8 | 1.21 | 1.61 | 5.23 |
| 790 | 1.21 | 0.4 | 1.21 | 1.21 | 4.03 |
| 794 | 1.21 | 0 | 0 | 0 | 1.21 |
| 795 | 0 | 1.21 | 0 | 0 | 1.21 |
| 797 | 1.61 | 1.21 | 2.81 | 2.81 | 8.44 |
| TOTAL | 67.52 | 83.16 | 80.77 | 75.92 | 307.37 |

STATE: UTAH

| ZIP CODE | QUARTER 1 | QUARTER 2 | QUARTER 3 | QUARTER 4 | TOTAL GRAMS |
|---|---|---|---|---|---|
| 840 | 7.23 | 7.23 | 8.44 | 9.64 | 32.54 |
| 841 | 12.05 | 13.26 | 8.03 | 4.42 | 37.76 |
| 843 | 0.4 | 0.8 | 0.8 | 0 | 2 |
| 844 | 2.81 | 1.61 | 0.4 | 0.4 | 5.22 |
| 846 | 6.03 | 4.42 | 5.62 | 4.42 | 20.49 |
| 847 | 8.44 | 9.24 | 6.03 | 8.03 | 31.74 |
| TOTAL | 36.96 | 36.56 | 29.32 | 26.91 | 129.75 |

STATE: VERMONT

| ZIP CODE | QUARTER 1 | QUARTER 2 | QUARTER 3 | QUARTER 4 | TOTAL GRAMS |
|---|---|---|---|---|---|
| 54 | 4.02 | 0 | 0 | 0 | 4.02 |
| 56 | 1.21 | 0.4 | 2.41 | 1.21 | 5.23 |
| 57 | 0 | 0 | 0 | 0.4 | 0.4 |
| TOTAL | 5.23 | 0.4 | 2.41 | 1.61 | 9.65 |

STATE: VIRGINIA

| ZIP CODE | QUARTER 1 | QUARTER 2 | QUARTER 3 | QUARTER 4 | TOTAL GRAMS |
|---|---|---|---|---|---|
| 201 | 7.63 | 6.43 | 6.83 | 9.64 | 30.53 |
| 220 | 2.81 | 4.02 | 4.02 | 3.62 | 14.47 |
| 221 | 2.01 | 4.42 | 2.41 | 0.4 | 9.24 |
| 222 | 0.4 | 0 | 0 | 2.41 | 2.81 |
| 223 | 0 | 1.21 | 1.61 | 2.01 | 4.83 |
| 224 | 0 | 0 | 0.4 | 1.61 | 2.01 |
| 228 | 4.82 | 2.41 | 0 | 0.4 | 7.63 |
| 229 | 0.8 | 0.8 | 0 | 0 | 1.6 |
| 230 | 0 | 0.4 | 0 | 0 | 0.4 |
| 231 | 5.22 | 6.03 | 6.83 | 1.21 | 19.29 |
| 232 | 5.62 | 6.03 | 7.63 | 10.85 | 30.13 |

DRUG CODE:1105DDRUG NAME:D-METHAMPHETAMINE
STATE:VIRGINIA

| ZIP CODE | QUARTER 1 | QUARTER 2 | QUARTER 3 | QUARTER 4 | TOTAL GRAMS |
|---|---|---|---|---|---|
| 233 | 1.61 | 0 | 0.8 | 0 | 2.41 |
| 234 | 5.62 | 8.44 | 9.24 | 7.63 | 30.93 |
| 235 | 2.01 | 2.01 | 3.62 | 2.41 | 10.05 |
| 236 | 0.8 | 1.21 | 0.8 | 1.61 | 4.42 |
| 237 | 0 | 0.4 | 0 | 0 | 0.4 |
| 238 | 6.03 | 6.03 | 3.62 | 3.62 | 19.3 |
| 240 | 1.61 | 2.41 | 4.02 | 4.02 | 12.06 |
| 241 | 1.61 | 2.01 | 1.21 | 2.01 | 6.84 |
| 243 | 0 | 0.4 | 0 | 0 | 0.4 |
| 245 | 2.41 | 2.41 | 1.61 | 1.61 | 8.04 |
| TOTAL | 51.01 | 57.07 | 54.65 | 55.06 | 217.79 |

STATE: WASHINGTON

| ZIP CODE | QUARTER 1 | QUARTER 2 | QUARTER 3 | QUARTER 4 | TOTAL GRAMS |
|---|---|---|---|---|---|

| | | | | | |
|---|---|---|---|---|---|
| 980 | 21.29 | 33.34 | 27.32 | 21.69 | 103.64 |
| 981 | 18.08 | 26.51 | 10.85 | 12.85 | 68.29 |
| 982 | 12.85 | 13.26 | 12.05 | 16.87 | 55.03 |
| 983 | 4.02 | 5.62 | 1.21 | 2.41 | 13.26 |
| 984 | 0.4 | 0 | 1.21 | 2.01 | 3.62 |
| 985 | 4.02 | 3.62 | 2.81 | 2.81 | 13.26 |
| 986 | 14.86 | 7.23 | 22.9 | 22.5 | 67.49 |
| 988 | 0.4 | 0 | 0.8 | 1.21 | 2.41 |
| 989 | 1.21 | 2.81 | 2.01 | 2.01 | 8.04 |
| 992 | 6.83 | 6.83 | 9.24 | 6.03 | 28.93 |
| 993 | 2.41 | 0.8 | 0 | 0 | 3.21 |
| TOTAL | 86.37 | 100.02 | 90.4 | 90.39 | 367.18 |

STATE: WEST VIRGINIA

| ZIP CODE | QUARTER 1 | QUARTER 2 | QUARTER 3 | QUARTER 4 | TOTAL GRAMS |
|---|---|---|---|---|---|
| 254 | 0 | 0 | 0.8 | 0 | 0.8 |
| 258 | 0.4 | 0.4 | 0.4 | 0 | 1.2 |
| TOTAL | 0.4 | 0.4 | 1.2 | 0 | 2 |

STATE: WISCONSIN

| ZIP CODE | QUARTER 1 | QUARTER 2 | QUARTER 3 | QUARTER 4 | TOTAL GRAMS |
|---|---|---|---|---|---|
| 530 | 0.4 | 0.4 | 2.41 | 2.41 | 5.62 |
| 531 | 0.8 | 3.21 | 1.21 | 3.21 | 8.43 |
| 532 | 7.23 | 4.02 | 6.43 | 7.23 | 24.91 |
| 535 | 2.41 | 4.82 | 2.01 | 3.21 | 12.45 |
| 537 | 10.04 | 9.64 | 11.25 | 14.06 | 44.99 |
| 539 | 0.8 | 0.8 | 0.4 | 0.4 | 2.4 |
| 540 | 1.21 | 3.21 | 1.21 | 2.41 | 8.04 |
| 541 | 0.8 | 3.21 | 0.8 | 0 | 4.81 |
| 542 | 0.8 | 1.61 | 2.01 | 2.81 | 7.23 |
| 543 | 1.21 | 2.01 | 5.62 | 10.44 | 19.28 |
| 546 | 3.21 | 2.01 | 2.01 | 1.61 | 8.84 |
| 547 | 0.8 | 0 | 0 | 0 | 0.8 |
| 549 | 6.43 | 14.06 | 8.84 | 6.83 | 36.16 |
| TOTAL | 36.14 | 49 | 44.2 | 54.62 | 183.96 |

STATE: WYOMING

| ZIP CODE | QUARTER 1 | QUARTER 2 | QUARTER 3 | QUARTER 4 | TOTAL GRAMS |
|---|---|---|---|---|---|
| 820 | 4.42 | 4.42 | 3.21 | 5.22 | 17.27 |
| 826 | 0 | 0.8 | 1.61 | 0.8 | 3.21 |
| 829 | 1.61 | 0.8 | 0.8 | 0.8 | 4.01 |
| TOTAL | 6.03 | 6.02 | 5.62 | 6.82 | 24.49 |

DRUG CODE:1205DRUG NAME:LISDEXAMFETAMINE

STATE: ALABAMA

| ZIP CODE | QUARTER 1 | QUARTER 2 | QUARTER 3 | QUARTER 4 | TOTAL GRAMS |
|---|---|---|---|---|---|
| 350 | 6,547.56 | 6,533.69 | 6,475.88 | 6,767.24 | 26,324.37 |
| 351 | 4,358.30 | 4,189.49 | 4,421.89 | 4,428.25 | 17,397.93 |
| 352 | 9,650.80 | 9,049 | 9,449.04 | 9,868.17 | 38,017.01 |
| 354 | 4,707.47 | 4,406.86 | 4,345 | 4,792.45 | 18,251.78 |
| 355 | 1,271.82 | 1,156.78 | 1,282.80 | 1,283.96 | 4,995.36 |
| 356 | 3,827.60 | 3,787.13 | 3,892.93 | 4,008.55 | 15,516.21 |
| 357 | 2,167.30 | 2,169.61 | 2,267.89 | 2,341.31 | 8,946.11 |
| 358 | 2,563.30 | 2,448.25 | 2,404.32 | 2,459.24 | 9,875.11 |
| 359 | 2,531.50 | 2,441.89 | 2,445.94 | 2,570.81 | 9,990.14 |
| 360 | 3,962.30 | 3,842.05 | 3,930.50 | 4,004.50 | 15,739.35 |
| 361 | 2,828.07 | 2,807.83 | 3,018.26 | 2,947.15 | 11,601.31 |

RETAIL DRUG DISTRIBUTION BY ZIP CODE WITHIN STATE BY GRAMS WT

DRUG CODE:1205DRUG NAME:LISDEXAMFETAMINE
STATE:ALABAMA

| ZIP CODE | QUARTER 1 | QUARTER 2 | QUARTER 3 | QUARTER 4 | TOTAL GRAMS |
|---|---|---|---|---|---|
| 362 | 1,905.42 | 1,787.49 | 1,966.12 | 1,967.85 | 7,626.88 |
| 363 | 4,308.58 | 3,873.85 | 4,204.52 | 4,324.77 | 16,711.72 |
| 364 | 1,167.76 | 1,033.64 | 1,124.98 | 1,230.20 | 4,556.58 |
| 365 | 8,030.39 | 7,743.65 | 7,840.77 | 7,766.77 | 31,381.58 |
| 366 | 6,823.31 | 6,653.93 | 6,478.77 | 6,750.47 | 26,706.48 |
| 367 | 771.19 | 616.25 | 679.85 | 750.37 | 2,817.66 |
| 368 | 4,720.19 | 4,242.68 | 4,335.17 | 4,658.33 | 17,956.37 |
| 369 | 176.32 | 161.87 | 149.73 | 142.79 | 630.71 |
| TOTAL | 72,319.18 | 68,945.94 | 70,714.36 | 73,063.18 | 285,042.66 |

STATE: ALASKA

| ZIP CODE | QUARTER 1 | QUARTER 2 | QUARTER 3 | QUARTER 4 | TOTAL GRAMS |
|---|---|---|---|---|---|
| 995 | 1,243.49 | 1,120.94 | 1,157.36 | 1,253.90 | 4,775.69 |
| 996 | 479.82 | 491.96 | 498.32 | 508.15 | 1,978.25 |
| 997 | 375.19 | 391.37 | 341.08 | 376.92 | 1,484.56 |
| 998 | 221.41 | 212.74 | 207.54 | 215.63 | 857.32 |
| 999 | 191.93 | 168.23 | 179.21 | 149.15 | 688.52 |
| TOTAL | 2,511.84 | 2,385.24 | 2,383.51 | 2,503.75 | 9,784.34 |

STATE: ARIZONA

| ZIP CODE | QUARTER 1 | QUARTER 2 | QUARTER 3 | QUARTER 4 | TOTAL GRAMS |
|---|---|---|---|---|---|
| 850 | 6,246.37 | 6,197.23 | 6,053.86 | 6,559.70 | 25,057.16 |
| 851 | 1,060.24 | 1,135.39 | 1,078.73 | 1,198.40 | 4,472.76 |
| 852 | 16,200.67 | 15,871.74 | 16,196.05 | 16,716.34 | 64,984.80 |
| 853 | 3,785.98 | 3,796.96 | 3,785.98 | 4,220.13 | 15,589.05 |
| 855 | 123.71 | 147.99 | 139.32 | 121.98 | 533 |
| 856 | 671.75 | 600.65 | 554.4 | 560.18 | 2,386.98 |
| 857 | 4,550.80 | 4,383.73 | 4,421.31 | 4,613.82 | 17,969.66 |
| 859 | 113.89 | 104.64 | 131.81 | 120.24 | 470.58 |
| 860 | 298.3 | 312.17 | 332.99 | 359 | 1,302.46 |
| 863 | 480.98 | 566.54 | 573.48 | 582.72 | 2,203.72 |
| 864 | 242.8 | 230.66 | 231.82 | 184.99 | 890.27 |
| 865 | 0 | 0 | 0 | 1.73 | 1.73 |
| TOTAL | 33,775.49 | 33,347.70 | 33,499.75 | 35,239.23 | 135,862.17 |

STATE: ARKANSAS

| ZIP CODE | QUARTER 1 | QUARTER 2 | QUARTER 3 | QUARTER 4 | TOTAL GRAMS |
|---|---|---|---|---|---|
| 716 | 1,599.60 | 1,594.98 | 1,581.68 | 1,678.22 | 6,454.48 |
| 717 | 813.39 | 726.09 | 716.27 | 851.54 | 3,107.29 |
| 718 | 796.62 | 678.11 | 668.86 | 823.79 | 2,967.38 |
| 719 | 1,667.82 | 1,560.29 | 1,875.93 | 2,063.82 | 7,167.86 |
| 720 | 4,580.29 | 4,142.09 | 4,316.09 | 4,647.92 | 17,686.39 |
| 721 | 3,392.29 | 3,127.52 | 3,200.36 | 3,295.17 | 13,015.34 |
| 722 | 3,621.80 | 3,468.60 | 3,555.32 | 3,689.43 | 14,335.15 |
| 723 | 1,335.99 | 1,101.86 | 1,194.35 | 1,249.27 | 4,881.47 |
| 724 | 2,904.95 | 2,836.16 | 2,746.55 | 2,779.50 | 11,267.16 |
| 725 | 640.53 | 656.14 | 681.58 | 744.59 | 2,722.84 |
| 726 | 819.75 | 792 | 786.22 | 886.81 | 3,284.78 |
| 727 | 5,319.10 | 5,183.82 | 4,939.29 | 5,573.46 | 21,015.67 |
| 728 | 1,245.81 | 1,123.25 | 1,118.05 | 1,215.17 | 4,702.28 |
| 729 | 2,765.05 | 2,462.71 | 2,402.58 | 2,622.26 | 10,252.60 |
| TOTAL | 31,502.99 | 29,453.62 | 29,783.13 | 32,120.95 | 122,860.69 |

STATE: CALIFORNIA

| ZIP CODE | QUARTER 1 | QUARTER 2 | QUARTER 3 | QUARTER 4 | TOTAL GRAMS |
|---|---|---|---|---|---|
| 900 | 5,490.22 | 5,637.63 | 6,009.93 | 5,929.57 | 23,067.35 |
| 902 | 4,560.05 | 4,738.69 | 4,757.18 | 4,864.13 | 18,920.05 |

| | | | | | |
|---|---|---|---|---|---|
| 903 | 135.28 | 154.93 | 134.7 | 179.79 | 604.7 |
| 904 | 911.66 | 923.8 | 1,009.36 | 984.5 | 3,829.32 |
| 905 | 839.4 | 755 | 796.62 | 873.51 | 3,264.53 |
| 906 | 1,061.97 | 1,086.83 | 958.49 | 1,116.89 | 4,224.18 |
| 907 | 1,349.86 | 1,529.07 | 1,357.96 | 1,464.91 | 5,701.80 |
| 908 | 804.14 | 769.45 | 764.25 | 912.24 | 3,250.08 |
| 910 | 667.13 | 741.7 | 753.84 | 756.15 | 2,918.82 |
| 911 | 1,301.88 | 1,316.91 | 1,355.64 | 1,458.55 | 5,432.98 |
| 912 | 419.7 | 468.26 | 426.64 | 442.82 | 1,757.42 |
| 913 | 5,687.35 | 5,587.34 | 5,559.01 | 5,919.74 | 22,753.44 |
| 914 | 1,057.34 | 1,144.06 | 1,163.72 | 1,178.17 | 4,543.29 |
| 915 | 414.5 | 449.76 | 471.15 | 559.02 | 1,894.43 |
| 916 | 485.03 | 497.74 | 474.62 | 589.08 | 2,046.47 |

DRUG CODE:1205DRUG NAME:LISDEXAMFETAMINE
STATE:CALIFORNIA

| ZIP CODE | QUARTER 1 | QUARTER 2 | QUARTER 3 | QUARTER 4 | TOTAL GRAMS |
|---|---|---|---|---|---|
| 917 | 2,739.04 | 2,832.11 | 2,650.01 | 2,856.97 | 11,078.13 |
| 918 | 106.95 | 119.67 | 128.92 | 106.95 | 462.49 |
| 919 | 894.9 | 989.13 | 941.72 | 1,040 | 3,865.75 |
| 920 | 13,259.30 | 13,439.67 | 14,053.03 | 14,987.24 | 55,739.24 |
| 921 | 4,603.41 | 4,696.48 | 4,610.35 | 4,962.41 | 18,872.65 |
| 922 | 1,266.62 | 1,193.20 | 1,083.94 | 1,175.86 | 4,719.62 |
| 923 | 2,036.07 | 1,974.79 | 1,893.28 | 2,043.01 | 7,947.15 |
| 924 | 219.68 | 204.65 | 190.19 | 187.88 | 802.4 |
| 925 | 1,986.35 | 2,233.78 | 1,976.52 | 2,273.67 | 8,470.32 |
| 926 | 8,211.91 | 8,193.41 | 8,068.54 | 8,637.39 | 33,111.25 |
| 927 | 1,244.07 | 1,256.79 | 1,260.84 | 1,200.14 | 4,961.84 |
| 928 | 2,884.72 | 2,928.65 | 2,832.69 | 3,141.40 | 11,787.46 |
| 930 | 2,462.13 | 2,665.04 | 2,482.94 | 2,617.06 | 10,227.17 |
| 931 | 1,044.05 | 979.3 | 937.68 | 976.41 | 3,937.44 |
| 932 | 1,151.58 | 1,228.46 | 1,114.58 | 1,154.47 | 4,649.09 |
| 933 | 767.72 | 868.31 | 767.14 | 861.37 | 3,264.54 |
| 934 | 1,837.20 | 2,013.52 | 1,896.17 | 2,057.46 | 7,804.35 |
| 935 | 1,110.53 | 1,033.64 | 1,059.66 | 1,070.64 | 4,274.47 |
| 936 | 1,541.21 | 1,409.99 | 1,484.56 | 1,586.31 | 6,022.07 |
| 937 | 1,970.16 | 2,029.71 | 2,064.40 | 2,193.31 | 8,257.58 |
| 939 | 787.95 | 767.72 | 748.64 | 758.47 | 3,062.78 |
| 940 | 1,852.23 | 2,011.79 | 2,047.05 | 2,049.36 | 7,960.43 |
| 941 | 3,160.47 | 3,508.49 | 3,520.05 | 3,583.64 | 13,772.65 |
| 943 | 413.92 | 406.98 | 366.52 | 396.58 | 1,584 |
| 944 | 474.04 | 423.17 | 421.43 | 430.11 | 1,748.75 |
| 945 | 4,435.76 | 4,580.29 | 4,646.77 | 4,884.95 | 18,547.77 |
| 946 | 763.67 | 851.54 | 926.12 | 899.52 | 3,440.85 |
| 947 | 464.21 | 474.62 | 505.26 | 497.74 | 1,941.83 |
| 948 | 42.2 | 72.26 | 55.5 | 52.03 | 221.99 |
| 949 | 1,186.26 | 1,357.96 | 1,246.38 | 1,251.59 | 5,042.19 |
| 950 | 1,956.29 | 2,223.95 | 2,162.09 | 2,199.09 | 8,541.42 |
| 951 | 1,190.89 | 1,167.18 | 1,204.18 | 1,200.14 | 4,762.39 |
| 952 | 870.62 | 857.32 | 874.67 | 937.1 | 3,539.71 |
| 953 | 2,430.91 | 2,337.84 | 2,178.28 | 2,485.25 | 9,432.28 |
| 954 | 578.68 | 669.44 | 703.55 | 645.74 | 2,597.41 |
| 955 | 93.65 | 124.29 | 123.71 | 104.64 | 446.29 |
| 956 | 3,207.30 | 3,122.90 | 3,061.04 | 3,399.81 | 12,791.05 |
| 957 | 924.96 | 1,000.69 | 1,038.27 | 1,026.13 | 3,990.05 |
| 958 | 1,462.59 | 1,453.92 | 1,462.01 | 1,470.11 | 5,848.63 |
| 959 | 1,583.99 | 1,578.21 | 1,463.75 | 1,677.07 | 6,303.02 |
| 960 | 779.86 | 730.72 | 715.69 | 828.42 | 3,054.69 |
| 961 | 161.87 | 131.23 | 140.48 | 156.67 | 590.25 |

RETAIL DRUG DISTRIBUTION BY ZIP CODE WITHIN STATE BY GRAMS WT

| | | | | | |
|---|---|---|---|---|---|
| TOTAL | 105,345.43 | 107,943.98 | 107,101.71 | 113,293.18 | 433,684.30 |

STATE: COLORADO

| ZIP CODE | QUARTER 1 | QUARTER 2 | QUARTER 3 | QUARTER 4 | TOTAL GRAMS |
|---|---|---|---|---|---|
| 800 | 4,304.53 | 4,217.82 | 4,102.20 | 4,261.18 | 16,885.73 |
| 801 | 5,091.90 | 5,257.82 | 5,220.82 | 5,531.84 | 21,102.38 |
| 802 | 5,322.57 | 5,297.71 | 5,642.26 | 5,816.26 | 22,078.80 |
| 803 | 1,582.26 | 1,525.61 | 1,531.97 | 1,617.52 | 6,257.36 |
| 804 | 846.92 | 829 | 833.04 | 897.21 | 3,406.17 |
| 805 | 2,611.28 | 2,697.99 | 2,674.87 | 2,898.59 | 10,882.73 |
| 806 | 1,247.54 | 1,183.95 | 1,140.01 | 1,248.12 | 4,819.62 |
| 807 | 158.98 | 137.01 | 159.56 | 169.96 | 625.51 |
| 808 | 406.98 | 408.72 | 397.73 | 470.57 | 1,684 |
| 809 | 2,655.21 | 2,574.28 | 2,586.42 | 2,836.74 | 10,652.65 |
| 810 | 615.1 | 586.19 | 588.51 | 656.14 | 2,445.94 |
| 811 | 98.86 | 77.47 | 84.98 | 99.43 | 360.74 |
| 812 | 124.87 | 127.76 | 151.46 | 120.82 | 524.91 |
| 813 | 206.96 | 172.27 | 178.05 | 210.43 | 767.71 |
| 814 | 127.76 | 121.4 | 123.14 | 114.46 | 486.76 |
| 815 | 512.77 | 425.48 | 546.3 | 464.79 | 1,949.34 |
| 816 | 663.08 | 717.42 | 770.03 | 708.17 | 2,858.70 |
| TOTAL | 26,577.57 | 26,357.90 | 26,731.35 | 28,122.23 | 107,789.05 |

STATE: CONNECTICUT

| ZIP CODE | QUARTER 1 | QUARTER 2 | QUARTER 3 | QUARTER 4 | TOTAL GRAMS |
|---|---|---|---|---|---|
| 60 | 5,627.80 | 5,491.37 | 5,645.72 | 6,039.99 | 22,804.88 |

DRUG CODE:1205DRUG NAME:LISDEXAMFETAMINE
STATE:CONNECTICUT

| ZIP CODE | QUARTER 1 | QUARTER 2 | QUARTER 3 | QUARTER 4 | TOTAL GRAMS |
|---|---|---|---|---|---|
| 61 | 1,984.04 | 2,103.71 | 2,045.90 | 2,212.97 | 8,346.62 |
| 62 | 1,768.99 | 1,596.13 | 1,582.26 | 1,840.67 | 6,788.05 |
| 63 | 3,371.48 | 3,517.16 | 3,417.15 | 3,447.79 | 13,753.58 |
| 64 | 6,701.91 | 7,168.44 | 6,715.21 | 7,234.34 | 27,819.90 |
| 65 | 1,638.34 | 1,725.63 | 1,734.30 | 1,726.21 | 6,824.48 |
| 66 | 1,369.52 | 1,322.11 | 1,357.96 | 1,496.70 | 5,546.29 |
| 67 | 3,000.34 | 3,082.43 | 2,930.97 | 3,125.21 | 12,138.95 |
| 68 | 8,114.21 | 8,066.81 | 8,105.54 | 8,571.49 | 32,858.05 |
| 69 | 929.58 | 1,087.98 | 1,132.50 | 964.27 | 4,114.33 |
| TOTAL | 34,506.21 | 35,161.77 | 34,667.51 | 36,659.64 | 140,995.13 |

STATE: DELAWARE

| ZIP CODE | QUARTER 1 | QUARTER 2 | QUARTER 3 | QUARTER 4 | TOTAL GRAMS |
|---|---|---|---|---|---|
| 197 | 5,494.84 | 5,258.98 | 5,079.19 | 5,480.97 | 21,313.98 |
| 198 | 3,605.03 | 3,613.13 | 3,442.01 | 3,660.53 | 14,320.70 |
| 199 | 5,407.55 | 5,295.40 | 5,161.28 | 5,723.77 | 21,588 |
| TOTAL | 14,507.42 | 14,167.51 | 13,682.48 | 14,865.27 | 57,222.68 |

STATE: DISTRICT OF COLUMBIA

| ZIP CODE | QUARTER 1 | QUARTER 2 | QUARTER 3 | QUARTER 4 | TOTAL GRAMS |
|---|---|---|---|---|---|
| 200 | 4,568.15 | 4,616.13 | 4,939.86 | 4,995.94 | 19,120.08 |
| 203 | 2.31 | 37.58 | 2.31 | 32.37 | 74.57 |
| 204 | 17.92 | 37 | 28.91 | 35.26 | 119.09 |
| TOTAL | 4,588.38 | 4,690.71 | 4,971.08 | 5,063.57 | 19,313.74 |

STATE: FLORIDA

| ZIP CODE | QUARTER 1 | QUARTER 2 | QUARTER 3 | QUARTER 4 | TOTAL GRAMS |
|---|---|---|---|---|---|
| 320 | 4,689.55 | 4,496.46 | 4,474.49 | 4,730.01 | 18,390.51 |
| 321 | 3,000.34 | 2,881.83 | 2,844.83 | 3,082.43 | 11,809.43 |

| | | | | | |
|---|---|---|---|---|---|
| 322 | 8,857.07 | 8,692.89 | 8,528.71 | 8,783.65 | 34,862.32 |
| 323 | 5,147.98 | 4,783.20 | 4,658.91 | 5,198.85 | 19,788.94 |
| 324 | 3,490.57 | 3,465.13 | 3,247.77 | 3,529.30 | 13,732.77 |
| 325 | 7,708.96 | 7,351.12 | 7,574.27 | 7,853.49 | 30,487.84 |
| 326 | 3,321.18 | 3,303.26 | 3,081.85 | 3,454.15 | 13,160.44 |
| 327 | 9,495.29 | 9,307.99 | 9,298.16 | 9,837.53 | 37,938.97 |
| 328 | 8,113.06 | 8,066.81 | 7,939.05 | 8,579.58 | 32,698.50 |
| 329 | 6,792.68 | 6,622.71 | 6,761.46 | 7,001.37 | 27,178.22 |
| 330 | 6,704.23 | 6,762.04 | 6,233.02 | 6,819.27 | 26,518.56 |
| 331 | 8,155.83 | 7,829.21 | 7,535.53 | 8,354.70 | 31,875.27 |
| 333 | 3,454.15 | 3,640.30 | 3,415.41 | 3,731.64 | 14,241.50 |
| 334 | 9,594.73 | 9,284.86 | 8,879.04 | 9,611.49 | 37,370.12 |
| 335 | 6,683.41 | 6,683.41 | 6,666.65 | 7,143 | 27,176.47 |
| 336 | 6,464.31 | 6,356.21 | 6,585.72 | 6,627.34 | 26,033.58 |
| 337 | 5,049.13 | 4,976.86 | 4,756.03 | 5,032.94 | 19,814.96 |
| 338 | 6,388.01 | 5,900.67 | 5,497.73 | 6,116.30 | 23,902.71 |
| 339 | 4,939.86 | 4,904.60 | 4,634.63 | 5,324.30 | 19,803.39 |
| 341 | 2,216.44 | 2,062.08 | 2,019.30 | 2,323.96 | 8,621.78 |
| 342 | 5,768.86 | 5,593.70 | 5,555.54 | 5,986.80 | 22,904.90 |
| 344 | 2,668.51 | 2,442.47 | 2,444.21 | 2,576.59 | 10,131.78 |
| 346 | 3,969.23 | 4,024.15 | 3,990.05 | 4,187.18 | 16,170.61 |
| 347 | 4,969.35 | 5,113.29 | 4,847.95 | 5,127.75 | 20,058.34 |
| 349 | 2,339.57 | 2,342.46 | 2,199.09 | 2,537.28 | 9,418.40 |
| TOTAL | 139,982.30 | 136,887.71 | 133,669.40 | 143,550.90 | 554,090.31 |

STATE: GEORGIA

| ZIP CODE | QUARTER 1 | QUARTER 2 | QUARTER 3 | QUARTER 4 | TOTAL GRAMS |
|---|---|---|---|---|---|
| 300 | 26,902.46 | 25,757.82 | 26,004.67 | 26,600.69 | 105,265.64 |
| 301 | 12,406.03 | 11,642.93 | 12,104.26 | 12,362.67 | 48,515.89 |
| 302 | 8,012.47 | 7,580.05 | 7,524.55 | 8,048.31 | 31,165.38 |
| 303 | 12,306.59 | 12,433.20 | 12,469.62 | 12,855.79 | 50,065.20 |
| 304 | 3,260.48 | 3,009.01 | 3,270.31 | 3,344.89 | 12,884.69 |
| 305 | 6,464.31 | 6,125.55 | 6,285.68 | 6,712.90 | 25,588.44 |
| 306 | 5,032.94 | 4,942.18 | 4,982.64 | 5,216.20 | 20,173.96 |
| 307 | 2,878.36 | 2,628.62 | 2,778.35 | 2,853.50 | 11,138.83 |
| 308 | 2,415.88 | 2,190.42 | 2,096.77 | 2,355.76 | 9,058.83 |
| 309 | 2,941.37 | 2,680.65 | 2,961.61 | 3,066.82 | 11,650.45 |
| 310 | 5,224.29 | 5,028.31 | 4,974.55 | 5,176.89 | 20,404.04 |
| 312 | 1,993.29 | 1,885.18 | 1,960.92 | 2,121.63 | 7,961.02 |
| 313 | 2,553.47 | 2,404.32 | 2,466.17 | 2,471.96 | 9,895.92 |
| 314 | 3,265.69 | 3,373.21 | 3,416.57 | 3,393.45 | 13,448.92 |

DRUG CODE:1205DRUG NAME:LISDEXAMFETAMINE
STATE:GEORGIA

| ZIP CODE | QUARTER 1 | QUARTER 2 | QUARTER 3 | QUARTER 4 | TOTAL GRAMS |
|---|---|---|---|---|---|
| 315 | 3,315.98 | 3,078.96 | 3,363.39 | 3,188.80 | 12,947.13 |
| 316 | 1,564.34 | 1,509.42 | 1,676.49 | 1,626.77 | 6,377.02 |
| 317 | 3,040.81 | 2,734.41 | 2,869.69 | 3,058.73 | 11,703.64 |
| 318 | 300.61 | 260.15 | 237.02 | 245.11 | 1,042.89 |
| 319 | 2,326.85 | 2,181.17 | 2,246.50 | 2,360.38 | 9,114.90 |
| 398 | 397.73 | 411.61 | 439.36 | 450.34 | 1,699.04 |
| TOTAL | 106,603.95 | 101,857.17 | 104,129.12 | 107,511.59 | 420,101.83 |

STATE: GUAM

| ZIP CODE | QUARTER 1 | QUARTER 2 | QUARTER 3 | QUARTER 4 | TOTAL GRAMS |
|---|---|---|---|---|---|
| 969 | 21.97 | 18.5 | 8.67 | 43.94 | 93.08 |
| TOTAL | 21.97 | 18.5 | 8.67 | 43.94 | 93.08 |

STATE: HAWAII

RETAIL DRUG DISTRIBUTION BY ZIP CODE WITHIN STATE BY GRAMS WT

| ZIP CODE | | QUARTER 1 | QUARTER 2 | QUARTER 3 | QUARTER 4 | TOTAL GRAMS |
|---|---|---|---|---|---|---|
| | 967 | 679.85 | 726.67 | 742.86 | 716.27 | 2,865.65 |
| | 968 | 523.76 | 527.23 | 750.37 | 521.45 | 2,322.81 |
| TOTAL | | 1,203.61 | 1,253.90 | 1,493.23 | 1,237.72 | 5,188.46 |

STATE: IDAHO

| ZIP CODE | | QUARTER 1 | QUARTER 2 | QUARTER 3 | QUARTER 4 | TOTAL GRAMS |
|---|---|---|---|---|---|---|
| | 832 | 486.76 | 496.01 | 419.7 | 522.02 | 1,924.49 |
| | 833 | 772.92 | 723.78 | 683.31 | 782.75 | 2,962.76 |
| | 834 | 1,383.97 | 1,340.61 | 1,323.85 | 1,384.55 | 5,432.98 |
| | 835 | 154.35 | 139.32 | 146.26 | 184.41 | 624.34 |
| | 836 | 2,526.88 | 2,673.71 | 2,575.44 | 2,798.58 | 10,574.61 |
| | 837 | 1,962.07 | 2,135.50 | 1,973.06 | 2,052.83 | 8,123.46 |
| | 838 | 897.21 | 923.8 | 931.32 | 952.71 | 3,705.04 |
| TOTAL | | 8,184.16 | 8,432.73 | 8,052.94 | 8,677.85 | 33,347.68 |

STATE: ILLINOIS

| ZIP CODE | | QUARTER 1 | QUARTER 2 | QUARTER 3 | QUARTER 4 | TOTAL GRAMS |
|---|---|---|---|---|---|---|
| | 600 | 16,705.93 | 16,222.64 | 16,526.14 | 16,783.40 | 66,238.11 |
| | 601 | 9,460.03 | 9,717.28 | 9,664.68 | 9,961.24 | 38,803.23 |
| | 602 | 765.98 | 772.92 | 641.69 | 746.33 | 2,926.92 |
| | 603 | 612.79 | 531.27 | 543.99 | 645.16 | 2,333.21 |
| | 604 | 7,075.94 | 7,226.83 | 7,059.18 | 7,390.43 | 28,752.38 |
| | 605 | 8,894.07 | 8,779.60 | 8,780.76 | 9,321.28 | 35,775.71 |
| | 606 | 10,138.72 | 10,592.53 | 10,339.90 | 10,826.08 | 41,897.23 |
| | 607 | 645.16 | 663.08 | 687.94 | 663.66 | 2,659.84 |
| | 608 | 156.67 | 142.79 | 134.7 | 176.9 | 611.06 |
| | 609 | 1,007.63 | 993.75 | 978.72 | 1,038.27 | 4,018.37 |
| | 610 | 856.74 | 891.43 | 844.03 | 929.58 | 3,521.78 |
| | 611 | 1,073.53 | 1,079.31 | 984.5 | 1,059.66 | 4,197 |
| | 612 | 683.31 | 641.69 | 633.02 | 689.67 | 2,647.69 |
| | 613 | 563.65 | 609.9 | 591.4 | 633.02 | 2,397.97 |
| | 614 | 743.44 | 677.53 | 710.48 | 706.44 | 2,837.89 |
| | 615 | 664.24 | 744.59 | 656.14 | 822.06 | 2,887.03 |
| | 616 | 1,148.68 | 1,177.01 | 1,156.20 | 1,197.25 | 4,679.14 |
| | 617 | 1,363.74 | 1,367.21 | 1,279.34 | 1,452.19 | 5,462.48 |
| | 618 | 1,186.26 | 1,118.05 | 1,219.79 | 1,425.02 | 4,949.12 |
| | 619 | 322.58 | 252.05 | 244.54 | 215.05 | 1,034.22 |
| | 620 | 885.07 | 919.18 | 871.2 | 942.88 | 3,618.33 |
| | 622 | 1,569.54 | 1,507.68 | 1,439.47 | 1,562.60 | 6,079.29 |
| | 623 | 805.29 | 793.73 | 738.23 | 849.23 | 3,186.48 |
| | 624 | 239.91 | 275.18 | 256.1 | 278.07 | 1,049.26 |
| | 625 | 535.9 | 533.01 | 483.29 | 545.73 | 2,097.93 |
| | 626 | 405.83 | 358.42 | 375.77 | 406.98 | 1,547 |
| | 627 | 686.2 | 602.38 | 637.07 | 630.13 | 2,555.78 |
| | 628 | 400.05 | 385.59 | 383.28 | 405.83 | 1,574.75 |
| | 629 | 1,008.21 | 976.41 | 1,031.91 | 1,015.72 | 4,032.25 |
| TOTAL | | 70,605.09 | 70,553.04 | 69,893.46 | 73,319.86 | 284,371.45 |

STATE: INDIANA

| ZIP CODE | | QUARTER 1 | QUARTER 2 | QUARTER 3 | QUARTER 4 | TOTAL GRAMS |
|---|---|---|---|---|---|---|
| | 460 | 9,687.80 | 9,554.84 | 10,072.81 | 10,555.53 | 39,870.98 |
| | 461 | 5,990.85 | 5,826.09 | 6,150.98 | 6,447.55 | 24,415.47 |
| | 462 | 9,502.23 | 9,314.93 | 9,224.74 | 9,754.86 | 37,796.76 |
| | 463 | 7,947.72 | 7,755.21 | 7,834.41 | 8,143.12 | 31,680.46 |
| | 464 | 769.45 | 755.58 | 725.52 | 770.61 | 3,021.16 |
| | 465 | 8,165.08 | 7,755.21 | 7,922.28 | 8,271.45 | 32,114.02 |

RETAIL DRUG DISTRIBUTION BY ZIP CODE WITHIN STATE BY GRAMS WT

| ZIP CODE | QUARTER 1 | QUARTER 2 | QUARTER 3 | QUARTER 4 | TOTAL GRAMS |
|---|---|---|---|---|---|
| 466 | 2,625.15 | 2,652.32 | 2,530.92 | 2,548.84 | 10,357.23 |
| 467 | 2,609.54 | 2,374.26 | 2,511.27 | 2,725.74 | 10,220.81 |
| 468 | 3,279.56 | 3,119.43 | 3,239.09 | 3,550.69 | 13,188.77 |
| 469 | 6,718.10 | 6,434.25 | 6,476.45 | 6,708.27 | 26,337.07 |
| 470 | 663.66 | 663.66 | 635.91 | 652.67 | 2,615.90 |
| 471 | 4,945.07 | 4,769.33 | 4,681.45 | 4,378.53 | 18,774.38 |
| 472 | 1,416.92 | 1,242.92 | 1,288.58 | 1,351.60 | 5,300.02 |
| 473 | 3,528.72 | 3,285.92 | 3,444.90 | 3,610.23 | 13,869.77 |
| 474 | 2,769.10 | 2,828.64 | 2,748.29 | 3,103.24 | 11,449.27 |
| 475 | 1,649.90 | 1,647.59 | 1,611.74 | 1,663.77 | 6,573 |
| 476 | 2,354.60 | 2,233.78 | 2,125.67 | 2,220.48 | 8,934.53 |
| 477 | 4,878.59 | 4,563.52 | 4,742.15 | 4,808.64 | 18,992.90 |
| 478 | 1,397.27 | 1,201.29 | 1,207.07 | 1,260.26 | 5,065.89 |
| 479 | 3,194 | 3,138.50 | 3,041.96 | 3,304.42 | 12,678.88 |
| TOTAL | 84,093.31 | 81,117.27 | 82,216.19 | 85,830.50 | 333,257.27 |

STATE: IOWA

| ZIP CODE | QUARTER 1 | QUARTER 2 | QUARTER 3 | QUARTER 4 | TOTAL GRAMS |
|---|---|---|---|---|---|
| 500 | 2,395.07 | 2,302.57 | 2,422.82 | 2,632.67 | 9,753.13 |
| 501 | 1,431.95 | 1,388.02 | 1,381.66 | 1,566.65 | 5,768.28 |
| 502 | 1,683.43 | 1,668.97 | 1,477.05 | 1,545.26 | 6,374.71 |
| 503 | 2,829.22 | 2,792.80 | 2,770.26 | 3,020.57 | 11,412.85 |
| 504 | 972.36 | 999.53 | 930.74 | 977.57 | 3,880.20 |
| 505 | 1,608.27 | 1,507.11 | 1,437.73 | 1,566.65 | 6,119.76 |
| 506 | 1,338.30 | 1,472.42 | 1,456.23 | 1,559.71 | 5,826.66 |
| 507 | 1,189.73 | 1,245.23 | 1,100.12 | 1,257.95 | 4,793.03 |
| 508 | 158.98 | 179.79 | 139.32 | 173.43 | 651.52 |
| 510 | 1,079.31 | 1,015.14 | 1,029.02 | 1,061.97 | 4,185.44 |
| 511 | 2,314.13 | 2,341.88 | 2,401.43 | 2,626.31 | 9,683.75 |
| 512 | 409.29 | 353.8 | 375.77 | 365.36 | 1,504.22 |
| 513 | 710.48 | 700.08 | 726.67 | 661.92 | 2,799.15 |
| 514 | 361.31 | 380.39 | 354.95 | 365.36 | 1,462.01 |
| 515 | 1,686.90 | 1,676.49 | 1,616.37 | 1,685.16 | 6,664.92 |
| 516 | 218.52 | 196.55 | 182.1 | 211.58 | 808.75 |
| 520 | 1,662.04 | 1,623.30 | 1,659.73 | 1,691.52 | 6,636.59 |
| 521 | 205.8 | 176.32 | 157.24 | 187.88 | 727.24 |
| 522 | 2,596.83 | 2,587.58 | 2,446.52 | 2,654.06 | 10,284.99 |
| 523 | 1,723.32 | 1,702.50 | 1,597.29 | 1,842.98 | 6,866.09 |
| 524 | 2,485.83 | 2,423.40 | 2,422.82 | 2,641.92 | 9,973.97 |
| 525 | 1,086.25 | 1,067.75 | 1,055.03 | 1,148.11 | 4,357.14 |
| 526 | 1,209.96 | 1,155.62 | 1,177.01 | 1,190.31 | 4,732.90 |
| 527 | 1,634.87 | 1,563.76 | 1,561.45 | 1,701.93 | 6,462.01 |
| 528 | 1,508.26 | 1,486.30 | 1,424.44 | 1,576.48 | 5,995.48 |
| TOTAL | 34,500.41 | 34,007.30 | 33,303.77 | 35,913.31 | 137,724.79 |

STATE: KANSAS

| ZIP CODE | QUARTER 1 | QUARTER 2 | QUARTER 3 | QUARTER 4 | TOTAL GRAMS |
|---|---|---|---|---|---|
| 660 | 6,458.53 | 6,694.40 | 6,792.10 | 7,493.33 | 27,438.36 |
| 661 | 855.59 | 982.19 | 982.19 | 946.35 | 3,766.32 |
| 662 | 6,983.45 | 7,062.07 | 7,197.92 | 7,750.01 | 28,993.45 |
| 664 | 437.04 | 417.97 | 412.19 | 509.88 | 1,777.08 |
| 665 | 803.56 | 741.12 | 764.83 | 875.82 | 3,185.33 |
| 666 | 3,401.54 | 3,295.17 | 3,475.54 | 3,575.55 | 13,747.80 |
| 667 | 1,207.07 | 1,177.59 | 1,198.40 | 1,270.66 | 4,853.72 |
| 668 | 509.88 | 501.79 | 450.92 | 530.12 | 1,992.71 |
| 669 | 108.1 | 95.96 | 90.76 | 108.1 | 402.92 |
| 670 | 1,232.51 | 1,314.60 | 1,335.41 | 1,612.90 | 5,495.42 |
| 671 | 564.23 | 549.77 | 550.35 | 665.39 | 2,329.74 |
| 672 | 3,577.86 | 3,581.33 | 3,727.01 | 3,958.83 | 14,845.03 |

| | | | | | |
|---|---|---|---|---|---|
| 673 | 538.79 | 548.04 | 561.34 | 572.9 | 2,221.07 |
| 674 | 894.9 | 860.21 | 907.04 | 900.1 | 3,562.25 |
| 675 | 776.39 | 798.36 | 770.03 | 816.28 | 3,161.06 |
| 676 | 595.44 | 547.46 | 479.82 | 631.86 | 2,254.58 |
| 677 | 146.26 | 168.81 | 134.12 | 120.82 | 570.01 |
| 678 | 375.77 | 357.84 | 386.17 | 375.19 | 1,494.97 |
| 679 | 137.01 | 97.7 | 93.65 | 108.1 | 436.46 |
| TOTAL | 29,603.92 | 29,792.38 | 30,309.79 | 32,822.19 | 122,528.28 |

STATE: KENTUCKY

DRUG CODE:1205DRUG NAME:LISDEXAMFETAMINE
STATE:KENTUCKY

| ZIP CODE | QUARTER 1 | QUARTER 2 | QUARTER 3 | QUARTER 4 | TOTAL GRAMS |
|---|---|---|---|---|---|
| 400 | 3,046.01 | 3,080.12 | 3,077.80 | 3,248.92 | 12,452.85 |
| 401 | 913.98 | 924.38 | 896.63 | 979.88 | 3,714.87 |
| 402 | 10,586.75 | 10,013.85 | 10,032.35 | 10,798.91 | 41,431.86 |
| 403 | 2,270.20 | 2,163.25 | 2,439 | 2,558.67 | 9,431.12 |
| 404 | 929.01 | 907.62 | 897.21 | 1,077.58 | 3,811.42 |
| 405 | 3,296.85 | 3,439.67 | 3,591.71 | 3,803.32 | 14,131.55 |
| 406 | 487.92 | 464.79 | 520.87 | 464.21 | 1,937.79 |
| 407 | 226.62 | 285.58 | 261.88 | 261.3 | 1,035.38 |
| 408 | 102.9 | 140.48 | 96.54 | 138.17 | 478.09 |
| 409 | 105.21 | 100.01 | 101.17 | 138.17 | 444.56 |
| 410 | 4,591.85 | 4,539.24 | 4,517.85 | 4,713.83 | 18,362.77 |
| 411 | 649.78 | 671.17 | 659.61 | 685.05 | 2,665.61 |
| 412 | 315.06 | 226.04 | 289.05 | 239.91 | 1,070.06 |
| 413 | 41.05 | 57.81 | 71.11 | 52.61 | 222.58 |
| 414 | 78.04 | 106.95 | 69.37 | 90.18 | 344.54 |
| 415 | 409.87 | 339.92 | 417.39 | 390.22 | 1,557.40 |
| 416 | 271.71 | 289.05 | 248.58 | 335.88 | 1,145.22 |
| 417 | 149.15 | 134.7 | 167.65 | 131.23 | 582.73 |
| 418 | 53.76 | 47.98 | 41.05 | 39.89 | 182.68 |
| 420 | 4,158.27 | 4,006.23 | 4,224.18 | 4,439.81 | 16,828.49 |
| 421 | 1,646.43 | 1,632.55 | 1,792.11 | 1,736.03 | 6,807.12 |
| 422 | 959.65 | 904.15 | 1,061.39 | 879.87 | 3,805.06 |
| 423 | 2,043.58 | 2,024.51 | 2,105.44 | 2,086.94 | 8,260.47 |
| 424 | 2,863.91 | 2,844.25 | 2,829.80 | 2,763.32 | 11,301.28 |
| 425 | 286.16 | 260.15 | 287.32 | 302.92 | 1,136.55 |
| 426 | 112.73 | 106.37 | 113.89 | 119.67 | 452.66 |
| 427 | 1,577.63 | 1,478.20 | 1,444.09 | 1,468.95 | 5,968.87 |
| TOTAL | 42,174.08 | 41,189.02 | 42,255.04 | 43,945.44 | 169,563.58 |

STATE: LOUISIANA

| ZIP CODE | QUARTER 1 | QUARTER 2 | QUARTER 3 | QUARTER 4 | TOTAL GRAMS |
|---|---|---|---|---|---|
| 700 | 9,228.21 | 8,328.11 | 8,806.20 | 9,131.67 | 35,494.19 |
| 701 | 4,654.86 | 4,579.13 | 4,450.21 | 4,762.97 | 18,447.17 |
| 703 | 5,347.43 | 4,865.29 | 5,287.88 | 5,389.05 | 20,889.65 |
| 704 | 10,228.32 | 9,585.48 | 9,329.38 | 9,699.36 | 38,842.54 |
| 705 | 20,123.08 | 18,646.04 | 18,773.80 | 20,131.75 | 77,674.67 |
| 706 | 10,683.29 | 9,936.38 | 9,627.10 | 9,434.59 | 39,681.36 |
| 707 | 12,547.08 | 11,859.72 | 12,058.59 | 12,492.16 | 48,957.55 |
| 708 | 9,971.07 | 9,780.87 | 9,357.13 | 10,342.21 | 39,451.28 |
| 710 | 1,847.03 | 1,797.89 | 1,836.05 | 1,952.24 | 7,433.21 |
| 711 | 5,890.26 | 5,575.77 | 6,039.99 | 6,344.65 | 23,850.67 |
| 712 | 3,816.04 | 3,557.05 | 3,643.76 | 3,805.05 | 14,821.90 |
| 713 | 3,693.48 | 3,283.61 | 3,487.68 | 3,552.42 | 14,017.19 |
| 714 | 1,729.10 | 1,487.45 | 1,601.92 | 1,659.73 | 6,478.20 |
| TOTAL | 99,759.25 | 93,282.79 | 94,299.69 | 98,697.85 | 386,039.58 |

STATE: MAINE

| ZIP CODE | QUARTER 1 | QUARTER 2 | QUARTER 3 | QUARTER 4 | TOTAL GRAMS |
|---|---|---|---|---|---|
| 39 | 635.91 | 627.24 | 618.57 | 614.52 | 2,496.24 |
| 40 | 5,481.54 | 5,762.50 | 5,464.20 | 5,749.78 | 22,458.02 |
| 41 | 2,044.74 | 2,148.22 | 2,107.75 | 2,129.72 | 8,430.43 |
| 42 | 1,559.14 | 1,666.66 | 1,603.65 | 1,651.63 | 6,481.08 |
| 43 | 653.25 | 746.91 | 753.26 | 730.14 | 2,883.56 |
| 44 | 2,084.05 | 2,114.11 | 2,019.88 | 2,041.27 | 8,259.31 |
| 45 | 414.5 | 407.56 | 395.42 | 456.12 | 1,673.60 |
| 46 | 781.01 | 782.17 | 789.68 | 748.06 | 3,100.92 |
| 47 | 782.75 | 897.21 | 821.48 | 900.1 | 3,401.54 |
| 48 | 310.44 | 326.05 | 311.6 | 272.29 | 1,220.38 |
| 49 | 1,496.12 | 1,499.59 | 1,475.89 | 1,473.58 | 5,945.18 |
| TOTAL | 16,243.45 | 16,978.22 | 16,361.38 | 16,767.21 | 66,350.26 |

STATE: MARYLAND

| ZIP CODE | QUARTER 1 | QUARTER 2 | QUARTER 3 | QUARTER 4 | TOTAL GRAMS |
|---|---|---|---|---|---|
| 206 | 3,067.40 | 3,283.61 | 2,930.97 | 3,234.47 | 12,516.45 |
| 207 | 3,779.62 | 3,783.66 | 3,683.08 | 4,005.65 | 15,252.01 |
| 208 | 7,161.50 | 7,303.14 | 7,377.71 | 7,756.37 | 29,598.72 |
| 209 | 1,280.49 | 1,306.51 | 1,300.15 | 1,387.44 | 5,274.59 |
| 210 | 9,621.90 | 10,232.37 | 9,707.46 | 10,455.52 | 40,017.25 |
| 211 | 4,654.28 | 4,838.70 | 4,709.78 | 4,978.60 | 19,181.36 |

DRUG CODE:1205DRUG NAME:LISDEXAMFETAMINE
STATE:MARYLAND

| ZIP CODE | QUARTER 1 | QUARTER 2 | QUARTER 3 | QUARTER 4 | TOTAL GRAMS |
|---|---|---|---|---|---|
| 212 | 8,905.41 | 8,913.61 | 8,637.18 | 9,234.57 | 35,690.77 |
| 214 | 1,438.31 | 1,460.86 | 1,418.66 | 1,470.11 | 5,787.94 |
| 215 | 1,855.70 | 2,051.68 | 1,973.63 | 2,056.88 | 7,937.89 |
| 216 | 2,129.14 | 2,226.84 | 2,173.08 | 2,360.96 | 8,890.02 |
| 217 | 5,349.74 | 5,197.70 | 5,327.77 | 5,859.04 | 21,734.25 |
| 218 | 1,981.15 | 2,076.54 | 1,960.34 | 2,152.84 | 8,170.87 |
| 219 | 1,234.82 | 1,200.14 | 1,170.07 | 1,264.30 | 4,869.33 |
| TOTAL | 52,459.46 | 53,875.36 | 52,369.88 | 56,216.75 | 214,921.45 |

STATE: MASSACHUSETTS

| ZIP CODE | QUARTER 1 | QUARTER 2 | QUARTER 3 | QUARTER 4 | TOTAL GRAMS |
|---|---|---|---|---|---|
| 10 | 1,631.98 | 1,631.40 | 1,623.88 | 1,812.34 | 6,699.60 |
| 11 | 654.99 | 713.95 | 650.94 | 770.03 | 2,789.91 |
| 12 | 501.21 | 523.76 | 474.04 | 474.62 | 1,973.63 |
| 13 | 312.75 | 347.44 | 261.3 | 297.72 | 1,219.21 |
| 14 | 1,293.79 | 1,352.18 | 1,289.74 | 1,244.65 | 5,180.36 |
| 15 | 1,279.34 | 1,325.01 | 1,334.83 | 1,429.64 | 5,368.82 |
| 16 | 896.63 | 861.37 | 843.45 | 889.7 | 3,491.15 |
| 17 | 2,362.12 | 2,313.56 | 2,350.55 | 2,532.08 | 9,558.31 |
| 18 | 4,125.32 | 4,144.98 | 4,060 | 4,431.71 | 16,762.01 |
| 19 | 3,746.09 | 3,991.20 | 3,818.35 | 4,112.03 | 15,667.67 |
| 20 | 2,959.29 | 3,048.90 | 2,798 | 3,013.06 | 11,819.25 |
| 21 | 4,519.01 | 4,743.31 | 4,695.91 | 5,201.17 | 19,159.40 |
| 22 | 232.4 | 220.83 | 230.08 | 322.58 | 1,005.89 |
| 23 | 2,376.57 | 2,433.80 | 2,408.36 | 2,500.28 | 9,719.01 |
| 24 | 2,370.79 | 2,513 | 2,407.79 | 2,742.51 | 10,034.09 |
| 25 | 647.47 | 705.28 | 680.42 | 740.55 | 2,773.72 |
| 26 | 696.61 | 770.03 | 774.65 | 781.01 | 3,022.30 |
| 27 | 1,903.68 | 2,038.96 | 2,017.57 | 2,221.06 | 8,181.27 |
| TOTAL | 32,510.04 | 33,678.96 | 32,719.86 | 35,516.74 | 134,425.60 |

STATE: MICHIGAN

| ZIP CODE | QUARTER 1 | QUARTER 2 | QUARTER 3 | QUARTER 4 | TOTAL GRAMS |
|---|---|---|---|---|---|

| | | | | | |
|---|---|---|---|---|---|
| 480 | 7,377.71 | 7,446.51 | 7,034.32 | 7,432.63 | 29,291.17 |
| 481 | 7,090.40 | 6,930.84 | 6,644.68 | 7,326.26 | 27,992.18 |
| 482 | 1,755.11 | 1,623.88 | 1,518.67 | 1,690.94 | 6,588.60 |
| 483 | 6,634.28 | 6,608.26 | 6,357.37 | 6,801.92 | 26,401.83 |
| 484 | 2,032.02 | 2,025.66 | 1,929.12 | 2,087.52 | 8,074.32 |
| 485 | 890.27 | 835.35 | 767.72 | 912.24 | 3,405.58 |
| 486 | 2,315.87 | 2,484.67 | 2,290.43 | 2,383.51 | 9,474.48 |
| 487 | 1,063.13 | 1,127.30 | 1,057.92 | 1,137.70 | 4,386.05 |
| 488 | 4,300.49 | 4,213.19 | 4,233.43 | 4,507.45 | 17,254.56 |
| 489 | 983.35 | 1,027.28 | 1,012.25 | 1,118.62 | 4,141.50 |
| 490 | 5,602.95 | 5,517.39 | 5,311.58 | 5,796.61 | 22,228.53 |
| 491 | 941.15 | 936.52 | 835.35 | 909.93 | 3,622.95 |
| 492 | 1,501.33 | 1,507.68 | 1,403.63 | 1,464.91 | 5,877.55 |
| 493 | 1,870.15 | 1,885.18 | 1,837.78 | 1,963.81 | 7,556.92 |
| 494 | 4,715.56 | 4,977.44 | 4,784.93 | 5,137.57 | 19,615.50 |
| 495 | 3,873.85 | 4,023.58 | 3,905.64 | 4,143.24 | 15,946.31 |
| 496 | 2,488.72 | 2,458.66 | 2,430.91 | 2,490.45 | 9,868.74 |
| 497 | 1,236.56 | 1,305.35 | 1,254.48 | 1,347.55 | 5,143.94 |
| 498 | 1,300.15 | 1,175.28 | 1,169.50 | 1,157.36 | 4,802.29 |
| 499 | 345.13 | 402.36 | 363.05 | 413.92 | 1,524.46 |
| TOTAL | 58,318.18 | 58,512.38 | 56,142.76 | 60,224.14 | 233,197.46 |

STATE: MINNESOTA

| ZIP CODE | QUARTER 1 | QUARTER 2 | QUARTER 3 | QUARTER 4 | TOTAL GRAMS |
|---|---|---|---|---|---|
| 550 | 2,543.64 | 2,690.48 | 2,740.19 | 3,038.49 | 11,012.80 |
| 551 | 3,855.93 | 4,133.99 | 3,929.92 | 4,335.17 | 16,255.01 |
| 553 | 5,635.90 | 5,674.05 | 5,726.08 | 6,211.11 | 23,247.14 |
| 554 | 5,663.07 | 5,933.62 | 6,025.54 | 6,703.65 | 24,325.88 |
| 556 | 19.08 | 18.5 | 19.66 | 17.92 | 75.16 |
| 557 | 1,022.08 | 1,029.60 | 1,052.14 | 1,046.94 | 4,150.76 |
| 558 | 406.4 | 382.12 | 359.58 | 448.61 | 1,596.71 |
| 559 | 1,214.01 | 1,238.29 | 1,229.62 | 1,377.61 | 5,059.53 |
| 560 | 2,902.64 | 2,969.70 | 3,002.65 | 3,163.36 | 12,038.35 |
| 561 | 658.46 | 660.77 | 668.28 | 686.78 | 2,674.29 |
| 562 | 804.72 | 893.74 | 883.91 | 856.74 | 3,439.11 |
| 563 | 1,318.07 | 1,322.69 | 1,409.41 | 1,435.42 | 5,485.59 |

DRUG CODE:1205DRUG NAME:LISDEXAMFETAMINE
STATE:MINNESOTA

| ZIP CODE | QUARTER 1 | QUARTER 2 | QUARTER 3 | QUARTER 4 | TOTAL GRAMS |
|---|---|---|---|---|---|
| 564 | 976.41 | 1,083.94 | 1,075.84 | 1,077 | 4,213.19 |
| 565 | 1,912.35 | 1,794.42 | 1,770.14 | 1,797.89 | 7,274.80 |
| 566 | 945.77 | 897.21 | 839.98 | 945.77 | 3,628.73 |
| 567 | 686.2 | 787.37 | 688.52 | 736.5 | 2,898.59 |
| TOTAL | 30,564.73 | 31,510.49 | 31,421.46 | 33,878.96 | 127,375.64 |

STATE: MISSISSIPPI

| ZIP CODE | QUARTER 1 | QUARTER 2 | QUARTER 3 | QUARTER 4 | TOTAL GRAMS |
|---|---|---|---|---|---|
| 386 | 6,425.58 | 6,161.97 | 6,428.47 | 6,993.28 | 26,009.30 |
| 387 | 644 | 522.6 | 533.59 | 625.5 | 2,325.69 |
| 388 | 2,733.83 | 2,532.08 | 2,565.61 | 3,004.39 | 10,835.91 |
| 389 | 823.79 | 749.8 | 779.86 | 947.51 | 3,300.96 |
| 390 | 2,887.03 | 2,651.74 | 2,894.55 | 3,011.32 | 11,444.64 |
| 391 | 3,573.81 | 3,224.06 | 3,308.47 | 3,476.69 | 13,583.03 |
| 392 | 3,196.31 | 2,872.58 | 2,880.09 | 3,166.83 | 12,115.81 |
| 393 | 2,174.23 | 1,934.32 | 2,055.15 | 2,203.72 | 8,367.42 |
| 394 | 6,408.82 | 5,838.81 | 6,349.85 | 6,635.43 | 25,232.91 |
| 395 | 5,662.49 | 5,452.64 | 5,601.21 | 6,236.54 | 22,952.88 |
| 396 | 2,100.82 | 1,859.75 | 1,936.64 | 2,044.74 | 7,941.95 |
| 397 | 2,179.44 | 1,872.47 | 2,021.62 | 1,996.76 | 8,070.29 |

| | | | | | |
|---|---|---|---|---|---|
| TOTAL | 38,810.15 | 35,672.82 | 37,355.11 | 40,342.71 | 152,180.79 |

STATE: MISSOURI

| ZIP CODE | QUARTER 1 | QUARTER 2 | QUARTER 3 | QUARTER 4 | TOTAL GRAMS |
|---|---|---|---|---|---|
| 630 | 7,556.92 | 7,417.02 | 7,602.02 | 8,066.81 | 30,642.77 |
| 631 | 21,484.51 | 20,911.61 | 21,556.77 | 22,683.49 | 86,636.38 |
| 633 | 6,977.09 | 6,863.78 | 6,982.29 | 7,188.10 | 28,011.26 |
| 634 | 498.9 | 504.1 | 497.17 | 583.88 | 2,084.05 |
| 635 | 630.71 | 618.57 | 556.71 | 570.01 | 2,376 |
| 636 | 887.96 | 821.48 | 867.15 | 882.76 | 3,459.35 |
| 637 | 535.9 | 565.96 | 511.62 | 589.66 | 2,203.14 |
| 638 | 1,109.95 | 1,054.45 | 1,035.38 | 1,145.22 | 4,345 |
| 639 | 416.23 | 417.97 | 426.64 | 404.67 | 1,665.51 |
| 640 | 5,448.01 | 5,433.56 | 5,439.92 | 5,815.11 | 22,136.60 |
| 641 | 8,554.72 | 8,913.72 | 8,850.13 | 9,495.29 | 35,813.86 |
| 644 | 396.58 | 450.34 | 400.05 | 461.9 | 1,708.87 |
| 645 | 1,106.48 | 1,082.78 | 1,137.12 | 1,222.10 | 4,548.48 |
| 646 | 181.52 | 182.68 | 192.51 | 191.35 | 748.06 |
| 647 | 752.69 | 741.7 | 653.25 | 779.28 | 2,926.92 |
| 648 | 1,345.82 | 1,189.73 | 1,149.84 | 1,231.93 | 4,917.32 |
| 650 | 538.79 | 456.7 | 499.48 | 498.9 | 1,993.87 |
| 651 | 724.36 | 681.58 | 645.16 | 677.53 | 2,728.63 |
| 652 | 2,394.49 | 2,176.55 | 2,123.36 | 2,286.96 | 8,981.36 |
| 653 | 540.52 | 416.23 | 455.54 | 476.35 | 1,888.64 |
| 654 | 403.51 | 384.44 | 348.59 | 443.98 | 1,580.52 |
| 655 | 631.86 | 610.47 | 552.09 | 612.79 | 2,407.21 |
| 656 | 776.97 | 709.33 | 723.78 | 750.95 | 2,961.03 |
| 657 | 1,158.51 | 1,107.64 | 1,061.39 | 1,168.34 | 4,495.88 |
| 658 | 1,856.28 | 1,708.29 | 1,764.94 | 1,906 | 7,235.51 |
| TOTAL | 66,909.28 | 65,420.68 | 66,032.90 | 70,133.36 | 268,496.22 |

STATE: MONTANA

| ZIP CODE | QUARTER 1 | QUARTER 2 | QUARTER 3 | QUARTER 4 | TOTAL GRAMS |
|---|---|---|---|---|---|
| 590 | 180.37 | 183.26 | 146.84 | 215.05 | 725.52 |
| 591 | 1,034.80 | 1,055.61 | 962.54 | 1,086.83 | 4,139.78 |
| 592 | 126.03 | 123.14 | 113.89 | 119.09 | 482.15 |
| 593 | 171.7 | 154.35 | 197.13 | 182.68 | 705.86 |
| 594 | 712.8 | 665.97 | 610.47 | 737.66 | 2,726.90 |
| 595 | 135.85 | 156.09 | 124.29 | 139.32 | 555.55 |
| 596 | 232.4 | 267.08 | 238.18 | 275.75 | 1,013.41 |
| 597 | 1,020.35 | 985.08 | 974.1 | 1,119.20 | 4,098.73 |
| 598 | 1,251.01 | 1,326.16 | 1,385.71 | 1,392.64 | 5,355.52 |
| 599 | 371.14 | 407.56 | 396 | 451.5 | 1,626.20 |
| TOTAL | 5,236.45 | 5,324.30 | 5,149.15 | 5,719.72 | 21,429.62 |

STATE: NEBRASKA

| ZIP CODE | QUARTER 1 | QUARTER 2 | QUARTER 3 | QUARTER 4 | TOTAL GRAMS |
|---|---|---|---|---|---|
| 680 | 1,387.44 | 1,446.41 | 1,534.28 | 1,522.72 | 5,890.85 |
| 681 | 6,069.47 | 6,075.83 | 6,205.90 | 6,689.20 | 25,040.40 |
| 683 | 378.66 | 408.14 | 411.03 | 431.26 | 1,629.09 |

DRUG CODE:1205DRUG NAME:LISDEXAMFETAMINE
STATE:NEBRASKA

| ZIP CODE | QUARTER 1 | QUARTER 2 | QUARTER 3 | QUARTER 4 | TOTAL GRAMS |
|---|---|---|---|---|---|
| 684 | 352.06 | 341.66 | 354.38 | 400.62 | 1,448.72 |
| 685 | 3,516.58 | 3,557.05 | 3,618.91 | 3,937.44 | 14,629.98 |
| 686 | 442.82 | 375.77 | 390.8 | 422.59 | 1,631.98 |
| 687 | 508.15 | 529.54 | 548.62 | 592.55 | 2,178.86 |
| 688 | 1,219.79 | 1,144.06 | 1,285.12 | 1,290.90 | 4,939.87 |

RETAIL DRUG DISTRIBUTION BY ZIP CODE WITHIN STATE BY GRAMS WT

| | | | | | |
|---|---|---|---|---|---|
| 689 | 532.43 | 486.18 | 482.71 | 522.6 | 2,023.92 |
| 690 | 156.67 | 209.27 | 182.68 | 181.52 | 730.14 |
| 691 | 343.97 | 337.03 | 338.77 | 352.64 | 1,372.41 |
| 692 | 16.76 | 14.45 | 16.19 | 27.17 | 74.57 |
| 693 | 455.54 | 423.17 | 469.42 | 455.54 | 1,803.67 |
| TOTAL | 15,380.34 | 15,348.56 | 15,838.81 | 16,826.75 | 63,394.46 |
| STATE: | NEVADA | | | | |
| ZIP CODE | QUARTER 1 | QUARTER 2 | QUARTER 3 | QUARTER 4 | TOTAL GRAMS |
| 890 | 1,730.83 | 1,711.75 | 1,600.18 | 1,858.59 | 6,901.35 |
| 891 | 3,404.87 | 3,450.08 | 3,288.96 | 3,848.41 | 13,992.32 |
| 893 | 5.78 | 18.5 | 15.03 | 9.25 | 48.56 |
| 894 | 633.6 | 698.34 | 667.71 | 685.63 | 2,685.28 |
| 895 | 921.49 | 950.4 | 967.74 | 1,007.63 | 3,847.26 |
| 897 | 233.55 | 208.69 | 183.84 | 242.22 | 868.3 |
| 898 | 81.51 | 46.25 | 46.25 | 63.01 | 237.02 |
| TOTAL | 7,011.63 | 7,084.01 | 6,769.71 | 7,714.74 | 28,580.09 |
| STATE: | NEW HAMPSHIRE | | | | |
| ZIP CODE | QUARTER 1 | QUARTER 2 | QUARTER 3 | QUARTER 4 | TOTAL GRAMS |
| 30 | 3,920.67 | 3,866.91 | 3,703.31 | 4,006.23 | 15,497.12 |
| 31 | 2,273.67 | 2,129.14 | 2,025.08 | 2,265.57 | 8,693.46 |
| 32 | 1,114.58 | 1,097.81 | 1,145.79 | 1,111.11 | 4,469.29 |
| 33 | 785.06 | 757.89 | 834.78 | 854.43 | 3,232.16 |
| 34 | 343.97 | 353.22 | 301.19 | 389.64 | 1,388.02 |
| 35 | 372.3 | 275.18 | 241.07 | 274.02 | 1,162.57 |
| 36 | 10.41 | 29.48 | 24.86 | 36.42 | 101.17 |
| 37 | 707.59 | 647.47 | 656.72 | 679.27 | 2,691.05 |
| 38 | 4,050.17 | 3,971.55 | 3,910.27 | 4,050.75 | 15,982.74 |
| TOTAL | 13,578.42 | 13,128.65 | 12,843.07 | 13,667.44 | 53,217.58 |
| STATE: | NEW JERSEY | | | | |
| ZIP CODE | QUARTER 1 | QUARTER 2 | QUARTER 3 | QUARTER 4 | TOTAL GRAMS |
| 70 | 8,481.88 | 8,816.03 | 8,226.94 | 9,107.97 | 34,632.82 |
| 71 | 531.85 | 545.15 | 471.15 | 565.96 | 2,114.11 |
| 72 | 365.36 | 375.19 | 271.13 | 325.47 | 1,337.15 |
| 73 | 430.68 | 511.62 | 459.01 | 519.71 | 1,921.02 |
| 74 | 3,531.61 | 3,253.55 | 3,299.22 | 3,445.48 | 13,529.86 |
| 75 | 390.8 | 405.83 | 404.09 | 403.51 | 1,604.23 |
| 76 | 2,998.03 | 3,071.45 | 3,136.77 | 3,314.25 | 12,520.50 |
| 77 | 3,105.55 | 3,202.67 | 3,117.12 | 3,358.18 | 12,783.52 |
| 78 | 2,948.89 | 3,172.61 | 2,878.94 | 2,941.95 | 11,942.39 |
| 79 | 3,225.80 | 3,223.49 | 3,000.34 | 3,370.90 | 12,820.53 |
| 80 | 8,602.13 | 8,840.88 | 8,410.20 | 9,307.41 | 35,160.62 |
| 81 | 589.66 | 572.9 | 541.1 | 550.35 | 2,254.01 |
| 82 | 1,257.37 | 1,314.60 | 1,357.96 | 1,194.35 | 5,124.28 |
| 83 | 955.6 | 1,008.21 | 1,039.42 | 961.38 | 3,964.61 |
| 84 | 86.14 | 112.73 | 121.98 | 109.26 | 430.11 |
| 85 | 2,311.24 | 2,316.45 | 2,103.13 | 2,376.57 | 9,107.39 |
| 86 | 1,510.58 | 1,444.67 | 1,388.02 | 1,508.26 | 5,851.53 |
| 87 | 2,173.08 | 2,183.48 | 2,248.81 | 2,218.17 | 8,823.54 |
| 88 | 4,069.25 | 4,145.56 | 3,856.51 | 4,213.77 | 16,285.09 |
| 89 | 479.82 | 466.53 | 428.37 | 515.67 | 1,890.39 |
| TOTAL | 48,045.32 | 48,983.60 | 46,760.21 | 50,308.57 | 194,097.70 |
| STATE: | NEW MEXICO | | | | |
| ZIP CODE | QUARTER 1 | QUARTER 2 | QUARTER 3 | QUARTER 4 | TOTAL GRAMS |
| 870 | 328.36 | 307.55 | 286.16 | 337.03 | 1,259.10 |
| 871 | 4,529.41 | 4,227.07 | 4,217.82 | 4,321.30 | 17,295.60 |
| 873 | 17.92 | 27.17 | 18.5 | 27.75 | 91.34 |
| 874 | 112.73 | 123.14 | 100.01 | 133.54 | 469.42 |
| 875 | 432.42 | 420.86 | 419.12 | 419.12 | 1,691.52 |

| | | | | | |
|---|---|---|---|---|---|
| 877 | 79.78 | 90.76 | 74 | 83.25 | 327.79 |
| 878 | 52.61 | 40.47 | 41.05 | 35.84 | 169.97 |
| 879 | 24.28 | 43.36 | 19.08 | 16.76 | 103.48 |

DRUG CODE:1205DRUG NAME:LISDEXAMFETAMINE
STATE:NEW MEXICO

| ZIP CODE | QUARTER 1 | QUARTER 2 | QUARTER 3 | QUARTER 4 | TOTAL GRAMS |
|---|---|---|---|---|---|
| 880 | 422.01 | 489.65 | 454.96 | 445.14 | 1,811.76 |
| 881 | 145.68 | 168.81 | 102.9 | 151.46 | 568.85 |
| 882 | 667.71 | 624.93 | 563.65 | 596.6 | 2,452.89 |
| 883 | 202.34 | 201.18 | 209.27 | 235.86 | 848.65 |
| 884 | 39.31 | 45.09 | 26.01 | 34.69 | 145.1 |
| TOTAL | 7,054.56 | 6,810.04 | 6,532.53 | 6,838.34 | 27,235.47 |

STATE: NEW YORK

| ZIP CODE | QUARTER 1 | QUARTER 2 | QUARTER 3 | QUARTER 4 | TOTAL GRAMS |
|---|---|---|---|---|---|
| 100 | 14,413.19 | 15,456.08 | 14,454.23 | 15,806.41 | 60,129.91 |
| 101 | 950.97 | 926.12 | 912.82 | 972.94 | 3,762.85 |
| 102 | 168.23 | 165.91 | 178.63 | 168.81 | 681.58 |
| 103 | 1,097.81 | 1,107.64 | 1,057.92 | 1,161.40 | 4,424.77 |
| 104 | 1,262.57 | 1,213.43 | 1,222.10 | 1,289.74 | 4,987.84 |
| 105 | 4,772.79 | 4,850.84 | 4,786.09 | 5,288.46 | 19,698.18 |
| 106 | 508.73 | 530.12 | 449.76 | 526.65 | 2,015.26 |
| 107 | 575.79 | 681 | 615.68 | 735.34 | 2,607.81 |
| 108 | 454.96 | 445.14 | 424.33 | 437.04 | 1,761.47 |
| 109 | 2,703.20 | 2,942.53 | 2,718.23 | 2,817.08 | 11,181.04 |
| 110 | 1,230.20 | 1,359.11 | 1,134.81 | 1,292.05 | 5,016.17 |
| 111 | 601.22 | 716.27 | 703.55 | 686.78 | 2,707.82 |
| 112 | 5,261.29 | 5,630.12 | 5,026 | 5,515.65 | 21,433.06 |
| 113 | 1,624.46 | 1,653.94 | 1,537.17 | 1,641.23 | 6,456.80 |
| 114 | 609.32 | 675.22 | 501.79 | 626.66 | 2,412.99 |
| 115 | 4,345.58 | 4,379.11 | 4,246.14 | 4,445.01 | 17,415.84 |
| 116 | 319.11 | 307.55 | 286.16 | 300.61 | 1,213.43 |
| 117 | 8,185.90 | 8,358.17 | 8,044.26 | 8,426.39 | 33,014.72 |
| 118 | 511.04 | 541.68 | 459.59 | 561.34 | 2,073.65 |
| 119 | 918.6 | 1,122.67 | 1,239.45 | 1,109.95 | 4,390.67 |
| 120 | 1,908.89 | 2,026.82 | 1,897.32 | 1,841.25 | 7,674.28 |
| 121 | 937.68 | 1,011.68 | 1,086.83 | 975.25 | 4,011.44 |
| 122 | 700.66 | 734.77 | 621.46 | 774.08 | 2,830.97 |
| 123 | 800.09 | 848.07 | 764.83 | 850.96 | 3,263.95 |
| 124 | 400.62 | 492.54 | 472.31 | 520.87 | 1,886.34 |
| 125 | 2,154 | 2,340.73 | 2,132.03 | 2,391.60 | 9,018.36 |
| 126 | 476.93 | 450.92 | 446.29 | 508.15 | 1,882.29 |
| 127 | 440.51 | 523.76 | 445.72 | 426.64 | 1,836.63 |
| 128 | 1,379.84 | 1,322.09 | 1,344.08 | 1,405.33 | 5,451.34 |
| 129 | 486.76 | 482.14 | 479.24 | 507.57 | 1,955.71 |
| 130 | 1,775.92 | 1,697.88 | 1,731.41 | 1,848.76 | 7,053.97 |
| 131 | 652.1 | 689.1 | 622.04 | 722.63 | 2,685.87 |
| 132 | 882.76 | 949.82 | 909.35 | 987.39 | 3,729.32 |
| 133 | 557.87 | 512.2 | 546.3 | 520.29 | 2,136.66 |
| 134 | 618.57 | 659.61 | 616.83 | 702.39 | 2,597.40 |
| 135 | 190.77 | 235.86 | 217.37 | 220.83 | 864.83 |
| 136 | 949.24 | 1,023.82 | 953.29 | 1,122.09 | 4,048.44 |
| 137 | 447.45 | 468.26 | 456.12 | 463.06 | 1,834.89 |
| 138 | 682.16 | 709.33 | 637.07 | 768.29 | 2,796.85 |
| 139 | 528.96 | 603.54 | 513.93 | 530.12 | 2,176.55 |
| 140 | 1,513.47 | 1,536.59 | 1,415.19 | 1,505.37 | 5,970.62 |
| 141 | 556.71 | 519.71 | 566.54 | 597.76 | 2,240.72 |
| 142 | 1,760.31 | 1,731.41 | 1,735.46 | 1,910.62 | 7,137.80 |
| 143 | 146.84 | 119.67 | 110.42 | 156.09 | 533.02 |

| | | | | | |
|---|---|---|---|---|---|
| 144 | 1,786.33 | 1,893.28 | 1,729.68 | 1,996.76 | 7,406.05 |
| 145 | 1,518.09 | 1,611.16 | 1,599.02 | 1,752.80 | 6,481.07 |
| 146 | 2,319.34 | 2,344.77 | 2,430.33 | 2,630.93 | 9,725.37 |
| 147 | 670.6 | 709.91 | 664.82 | 713.38 | 2,758.71 |
| 148 | 1,096.08 | 1,078.16 | 1,016.30 | 1,069.49 | 4,260.03 |
| 149 | 356.69 | 331.83 | 330.67 | 353.22 | 1,372.41 |
| TOTAL | 79,211.20 | 82,722.08 | 78,490.96 | 84,583.51 | 325,007.75 |

STATE: NORTH CAROLINA

| ZIP CODE | QUARTER 1 | QUARTER 2 | QUARTER 3 | QUARTER 4 | TOTAL GRAMS |
|---|---|---|---|---|---|
| 270 | 2,266.15 | 2,409.52 | 2,200.83 | 2,313.56 | 9,190.06 |
| 271 | 3,100.35 | 3,167.41 | 2,916.51 | 3,216.55 | 12,400.82 |
| 272 | 6,020.91 | 5,683.30 | 5,412.17 | 5,976.40 | 23,092.78 |
| 273 | 3,391.13 | 3,371.48 | 3,065.66 | 3,449.52 | 13,277.79 |
| 274 | 3,876.16 | 3,945.53 | 3,505.60 | 3,799.85 | 15,127.14 |

DRUG CODE:1205DRUG NAME:LISDEXAMFETAMINE
STATE:NORTH CAROLINA

| ZIP CODE | QUARTER 1 | QUARTER 2 | QUARTER 3 | QUARTER 4 | TOTAL GRAMS |
|---|---|---|---|---|---|
| 275 | 10,998.93 | 11,611.14 | 10,965.98 | 11,920.42 | 45,496.47 |
| 276 | 5,410.44 | 5,434.72 | 5,245.10 | 5,551.49 | 21,641.75 |
| 277 | 2,204.39 | 2,233.51 | 1,948.20 | 2,138.39 | 8,524.49 |
| 278 | 5,235.27 | 5,045.08 | 4,795.92 | 5,071.67 | 20,147.94 |
| 279 | 1,327.90 | 1,336.57 | 1,257.37 | 1,299.57 | 5,221.41 |
| 280 | 9,375.63 | 9,526.51 | 9,349.61 | 9,576.80 | 37,828.55 |
| 281 | 7,082.88 | 7,214.69 | 6,868.98 | 7,543.63 | 28,710.18 |
| 282 | 7,974.31 | 7,828.05 | 7,783.57 | 8,424.10 | 32,010.03 |
| 283 | 7,946.56 | 7,481.19 | 7,273.08 | 7,804.93 | 30,505.76 |
| 284 | 6,374.71 | 6,414.60 | 6,289.73 | 6,373.55 | 25,452.59 |
| 285 | 4,661.80 | 4,880.90 | 5,282.10 | 4,783.20 | 19,608 |
| 286 | 6,593.23 | 7,021.60 | 6,868.41 | 7,489.86 | 27,973.10 |
| 287 | 3,452.99 | 3,383.04 | 3,316.56 | 3,677.87 | 13,830.46 |
| 288 | 1,363.74 | 1,438.89 | 1,489.19 | 1,507.11 | 5,798.93 |
| 289 | 232.97 | 262.46 | 255.52 | 204.65 | 955.6 |
| TOTAL | 98,890.45 | 99,690.19 | 96,090.09 | 102,123.12 | 396,793.85 |

STATE: NORTH DAKOTA

| ZIP CODE | QUARTER 1 | QUARTER 2 | QUARTER 3 | QUARTER 4 | TOTAL GRAMS |
|---|---|---|---|---|---|
| 580 | 639.96 | 597.18 | 579.26 | 547.46 | 2,363.86 |
| 581 | 1,653.94 | 1,626.20 | 1,689.21 | 1,503.64 | 6,472.99 |
| 582 | 1,186.84 | 1,149.84 | 1,186.26 | 1,105.33 | 4,628.27 |
| 583 | 249.16 | 267.08 | 275.18 | 245.69 | 1,037.11 |
| 584 | 183.26 | 202.34 | 166.49 | 208.69 | 760.78 |
| 585 | 483.87 | 476.93 | 527.23 | 505.84 | 1,993.87 |
| 586 | 118.51 | 107.53 | 82.67 | 101.75 | 410.46 |
| 587 | 1,004.74 | 882.76 | 883.91 | 928.43 | 3,699.84 |
| 588 | 223.15 | 191.93 | 191.93 | 174.01 | 781.02 |
| TOTAL | 5,743.43 | 5,501.79 | 5,582.14 | 5,320.84 | 22,148.20 |

STATE: OHIO

| ZIP CODE | QUARTER 1 | QUARTER 2 | QUARTER 3 | QUARTER 4 | TOTAL GRAMS |
|---|---|---|---|---|---|
| 430 | 9,590.10 | 9,483.73 | 9,306.25 | 9,805.15 | 38,185.23 |
| 431 | 3,732.21 | 3,750.71 | 3,595.78 | 3,716.03 | 14,794.73 |
| 432 | 22,604.29 | 21,479.88 | 22,547.63 | 22,578.27 | 89,210.07 |
| 433 | 922.07 | 881.6 | 920.91 | 1,020.35 | 3,744.93 |
| 434 | 1,016.88 | 1,037.69 | 1,053.88 | 1,126.72 | 4,235.17 |
| 435 | 2,392.76 | 2,380.04 | 2,253.43 | 2,481.21 | 9,507.44 |
| 436 | 3,398.07 | 3,024.04 | 3,018.26 | 3,097.46 | 12,537.83 |
| 437 | 1,096.08 | 1,144.06 | 1,039.42 | 1,103.59 | 4,383.15 |
| 438 | 200.6 | 216.79 | 191.35 | 215.63 | 824.37 |

| | | | | | |
|---|---|---|---|---|---|
| 439 | 1,037.11 | 1,065.44 | 1,024.97 | 1,181.64 | 4,309.16 |
| 440 | 9,969.33 | 9,821.92 | 9,380.25 | 9,696.47 | 38,867.97 |
| 441 | 10,951.87 | 11,362.91 | 10,386.87 | 11,077.55 | 43,779.20 |
| 442 | 4,791.87 | 4,780.89 | 4,578.55 | 5,043.34 | 19,194.65 |
| 443 | 1,805.41 | 1,712.33 | 1,691.52 | 1,705.40 | 6,914.66 |
| 444 | 2,254.59 | 2,236.09 | 2,185.22 | 2,209.50 | 8,885.40 |
| 445 | 1,954.56 | 1,971.90 | 1,804.83 | 2,042.43 | 7,773.72 |
| 446 | 3,228.69 | 3,257.02 | 3,187.64 | 3,377.84 | 13,051.19 |
| 447 | 2,599.14 | 2,717.65 | 2,753.49 | 2,816.50 | 10,886.78 |
| 448 | 1,826.80 | 1,762.05 | 1,685.74 | 1,767.83 | 7,042.42 |
| 449 | 731.3 | 703.55 | 711.64 | 749.22 | 2,895.71 |
| 450 | 6,636.01 | 6,507.67 | 6,539.47 | 6,938.93 | 26,622.08 |
| 451 | 2,389.87 | 2,364.43 | 2,270.20 | 2,496.81 | 9,521.31 |
| 452 | 7,472.88 | 7,412.60 | 7,454.60 | 7,750.59 | 30,090.67 |
| 453 | 3,338.53 | 3,334.48 | 3,158.16 | 3,453.57 | 13,284.74 |
| 454 | 3,669.78 | 3,555.89 | 3,546.07 | 3,786.56 | 14,558.30 |
| 455 | 877.56 | 853.85 | 856.17 | 893.74 | 3,481.32 |
| 456 | 1,726.21 | 1,703.08 | 1,631.98 | 1,838.94 | 6,900.21 |
| 457 | 1,604.23 | 1,467.80 | 1,526.76 | 1,601.34 | 6,200.13 |
| 458 | 1,276.44 | 1,173.54 | 1,195.51 | 1,198.40 | 4,843.89 |
| TOTAL | 115,095.24 | 113,163.63 | 111,496.55 | 116,771.01 | 456,526.43 |

STATE: OKLAHOMA

| ZIP CODE | QUARTER 1 | QUARTER 2 | QUARTER 3 | QUARTER 4 | TOTAL GRAMS |
|---|---|---|---|---|---|
| 730 | 7,779.49 | 7,626.87 | 7,932.11 | 8,409.04 | 31,747.51 |
| 731 | 6,918.70 | 6,993.03 | 6,717.04 | 7,019.78 | 27,648.55 |
| 734 | 752.69 | 657.3 | 721.47 | 815.12 | 2,946.58 |
| 735 | 1,203.60 | 1,200.14 | 1,125.56 | 1,135.39 | 4,664.69 |

DRUG CODE:1205DRUG NAME:LISDEXAMFETAMINE
STATE:OKLAHOMA

| ZIP CODE | QUARTER 1 | QUARTER 2 | QUARTER 3 | QUARTER 4 | TOTAL GRAMS |
|---|---|---|---|---|---|
| 736 | 351.48 | 306.97 | 317.38 | 307.55 | 1,283.38 |
| 737 | 801.25 | 808.18 | 792.58 | 863.1 | 3,265.11 |
| 738 | 89.61 | 98.86 | 75.73 | 93.07 | 357.27 |
| 739 | 68.79 | 53.19 | 67.06 | 83.82 | 272.86 |
| 740 | 6,518.66 | 6,058.49 | 6,351.58 | 7,029.12 | 25,957.85 |
| 741 | 4,283.72 | 4,406.86 | 4,188.91 | 4,478.54 | 17,358.03 |
| 743 | 1,255.63 | 1,067.75 | 1,047.52 | 1,159.09 | 4,529.99 |
| 744 | 1,792.69 | 1,667.24 | 1,689.79 | 1,860.33 | 7,010.05 |
| 745 | 543.41 | 652.67 | 538.21 | 633.02 | 2,367.31 |
| 746 | 465.37 | 413.92 | 430.68 | 445.72 | 1,755.69 |
| 747 | 487.34 | 442.82 | 481.56 | 540.52 | 1,952.24 |
| 748 | 1,255.06 | 1,183.95 | 1,295.52 | 1,482.25 | 5,216.78 |
| 749 | 529.54 | 539.37 | 526.07 | 604.69 | 2,199.67 |
| TOTAL | 35,097.03 | 34,177.61 | 34,298.77 | 36,960.15 | 140,533.56 |

STATE: OREGON

| ZIP CODE | QUARTER 1 | QUARTER 2 | QUARTER 3 | QUARTER 4 | TOTAL GRAMS |
|---|---|---|---|---|---|
| 970 | 3,365.12 | 3,672.67 | 3,569.19 | 3,829.91 | 14,436.89 |
| 971 | 1,027.28 | 1,079.89 | 1,060.24 | 1,158.51 | 4,325.92 |
| 972 | 4,075.61 | 4,449.64 | 4,404.54 | 4,611.50 | 17,541.29 |
| 973 | 1,543.53 | 1,534.86 | 1,611.74 | 1,588.04 | 6,278.17 |
| 974 | 1,389.75 | 1,371.25 | 1,388.60 | 1,419.81 | 5,569.41 |
| 975 | 588.51 | 658.46 | 501.21 | 653.25 | 2,401.43 |
| 976 | 94.23 | 90.18 | 84.4 | 88.45 | 357.26 |
| 977 | 1,401.31 | 1,423.86 | 1,384.55 | 1,319.22 | 5,528.94 |
| 978 | 300.61 | 345.7 | 370.56 | 423.17 | 1,440.04 |
| 979 | 72.26 | 64.17 | 57.81 | 102.32 | 296.56 |

| | | | | | |
|---|---|---|---|---|---|
| TOTAL | 13,858.21 | 14,690.68 | 14,432.84 | 15,194.18 | 58,175.91 |

STATE: PENNSYLVANIA

| ZIP CODE | QUARTER 1 | QUARTER 2 | QUARTER 3 | QUARTER 4 | TOTAL GRAMS |
|---|---|---|---|---|---|
| 150 | 2,114.69 | 2,333.79 | 2,300.84 | 2,317.02 | 9,066.34 |
| 151 | 1,731.99 | 1,670.13 | 1,697.88 | 1,768.41 | 6,868.41 |
| 152 | 3,681.34 | 3,745.51 | 3,790.02 | 4,141.51 | 15,358.38 |
| 153 | 882.76 | 890.85 | 978.15 | 948.66 | 3,700.42 |
| 154 | 672.91 | 676.38 | 702.39 | 727.83 | 2,779.51 |
| 155 | 298.3 | 298.3 | 258.99 | 266.5 | 1,122.09 |
| 156 | 1,485.72 | 1,477.62 | 1,526.18 | 1,562.03 | 6,051.55 |
| 157 | 1,172.39 | 1,108.80 | 1,155.62 | 1,264.30 | 4,701.11 |
| 158 | 1,101.86 | 1,064.28 | 1,042.89 | 1,123.83 | 4,332.86 |
| 159 | 700.66 | 718 | 769.45 | 782.17 | 2,970.28 |
| 160 | 1,106.48 | 1,227.31 | 1,179.90 | 1,235.98 | 4,749.67 |
| 161 | 1,392.06 | 1,516.36 | 1,418.08 | 1,398.42 | 5,724.92 |
| 162 | 553.24 | 483.29 | 548.62 | 599.49 | 2,184.64 |
| 163 | 1,056.19 | 1,118.05 | 1,122.67 | 1,141.17 | 4,438.08 |
| 164 | 639.96 | 619.72 | 591.97 | 678.11 | 2,529.76 |
| 165 | 2,865.64 | 2,711.29 | 2,805.52 | 2,808.41 | 11,190.86 |
| 166 | 1,286.27 | 1,291.48 | 1,222.68 | 1,369.52 | 5,169.95 |
| 167 | 368.25 | 398.31 | 443.4 | 379.81 | 1,589.77 |
| 168 | 1,360.85 | 1,308.82 | 1,342.93 | 1,443.52 | 5,456.12 |
| 169 | 349.17 | 327.2 | 334.14 | 403.51 | 1,414.02 |
| 170 | 4,230.54 | 4,246.72 | 4,177.93 | 4,472.18 | 17,127.37 |
| 171 | 1,142.90 | 1,136.54 | 1,138.28 | 1,206.49 | 4,624.21 |
| 172 | 1,154.47 | 1,033.06 | 1,134.81 | 1,145.79 | 4,468.13 |
| 173 | 1,963.23 | 1,901.37 | 1,895.01 | 2,073.64 | 7,833.25 |
| 174 | 1,133.08 | 1,266.04 | 1,057.92 | 1,271.24 | 4,728.28 |
| 175 | 1,350.44 | 1,199.56 | 1,219.21 | 1,279.34 | 5,048.55 |
| 176 | 1,357.96 | 1,505.95 | 1,395.53 | 1,579.37 | 5,838.81 |
| 177 | 740.55 | 718 | 685.05 | 728.98 | 2,872.58 |
| 178 | 686.78 | 674.64 | 633.02 | 705.28 | 2,699.72 |
| 179 | 258.41 | 291.36 | 276.33 | 388.48 | 1,214.58 |
| 180 | 2,038.96 | 2,203.14 | 2,249.97 | 2,274.25 | 8,766.32 |
| 181 | 721.47 | 899.52 | 823.21 | 898.95 | 3,343.15 |
| 182 | 385.59 | 315.06 | 331.83 | 334.72 | 1,367.20 |
| 183 | 541.1 | 567.12 | 497.17 | 554.4 | 2,159.79 |
| 184 | 561.34 | 606.43 | 622.61 | 627.82 | 2,418.20 |
| 185 | 389.64 | 363.05 | 385.01 | 455.54 | 1,593.24 |
| 186 | 439.36 | 450.34 | 381.55 | 455.54 | 1,726.79 |

DRUG CODE:1205DRUG NAME:LISDEXAMFETAMINE
STATE:PENNSYLVANIA

| ZIP CODE | QUARTER 1 | QUARTER 2 | QUARTER 3 | QUARTER 4 | TOTAL GRAMS |
|---|---|---|---|---|---|
| 187 | 11,287.98 | 10,526.04 | 10,974.97 | 10,750.35 | 43,539.34 |
| 188 | 199.44 | 205.23 | 202.91 | 243.38 | 850.96 |
| 189 | 2,297.95 | 2,350.55 | 2,301.42 | 2,261.53 | 9,211.45 |
| 190 | 7,506.05 | 8,032.70 | 7,532.06 | 8,246.60 | 31,317.41 |
| 191 | 5,196.54 | 5,445.70 | 5,036.99 | 5,809.91 | 21,489.14 |
| 193 | 4,447.32 | 4,723.08 | 4,476.81 | 4,780.31 | 18,427.52 |
| 194 | 3,335.64 | 3,380.15 | 3,242.56 | 3,455.30 | 13,413.65 |
| 195 | 934.21 | 878.71 | 857.9 | 973.52 | 3,644.34 |
| 196 | 1,660.88 | 1,664.93 | 1,582.26 | 1,703.66 | 6,611.73 |
| TOTAL | 80,782.56 | 81,570.48 | 80,344.64 | 85,036.77 | 327,734.45 |

STATE: PUERTO RICO

| ZIP CODE | QUARTER 1 | QUARTER 2 | QUARTER 3 | QUARTER 4 | TOTAL GRAMS |
|---|---|---|---|---|---|
| 6 | 263.61 | 260.72 | 254.94 | 291.94 | 1,071.21 |

| | | | | | | |
|---|---|---|---|---|---|---|
| | 7 | 230.08 | 193.66 | 190.77 | 221.99 | 836.5 |
| | 9 | 1,296.68 | 1,189.73 | 1,182.79 | 1,457.39 | 5,126.59 |
| TOTAL | | 1,790.37 | 1,644.11 | 1,628.50 | 1,971.32 | 7,034.30 |
| STATE: | RHODE ISLAND | | | | | |
| ZIP CODE | QUARTER 1 | | QUARTER 2 | QUARTER 3 | QUARTER 4 | TOTAL GRAMS |
| | 28 | 4,948.54 | 4,832.34 | 5,035.83 | 5,172.84 | 19,989.55 |
| | 29 | 2,199.09 | 2,480.05 | 2,281.18 | 2,441.89 | 9,402.21 |
| TOTAL | | 7,147.63 | 7,312.39 | 7,317.01 | 7,614.73 | 29,391.76 |
| STATE: | SOUTH CAROLINA | | | | | |
| ZIP CODE | QUARTER 1 | | QUARTER 2 | QUARTER 3 | QUARTER 4 | TOTAL GRAMS |
| | 290 | 5,128.90 | 4,963.57 | 5,012.71 | 5,347.43 | 20,452.61 |
| | 291 | 3,418.88 | 3,363.96 | 3,362.23 | 3,762.85 | 13,907.92 |
| | 292 | 4,563.52 | 4,191.80 | 4,210.88 | 4,728.86 | 17,695.06 |
| | 293 | 4,862.40 | 4,870.49 | 4,745.62 | 4,985.53 | 19,464.04 |
| | 294 | 13,924.12 | 13,445.45 | 13,111.31 | 13,554.13 | 54,035.01 |
| | 295 | 7,509.52 | 7,305.45 | 7,312.97 | 7,556.92 | 29,684.86 |
| | 296 | 13,069.11 | 12,974.30 | 13,130.39 | 14,194.09 | 53,367.89 |
| | 297 | 3,263.37 | 3,161.05 | 3,066.82 | 3,369.17 | 12,860.41 |
| | 298 | 2,523.98 | 2,333.21 | 2,337.84 | 2,558.67 | 9,753.70 |
| | 299 | 1,892.70 | 1,775.35 | 1,863.22 | 1,818.12 | 7,349.39 |
| TOTAL | | 60,156.50 | 58,384.63 | 58,153.99 | 61,875.77 | 238,570.89 |
| STATE: | SOUTH DAKOTA | | | | | |
| ZIP CODE | QUARTER 1 | | QUARTER 2 | QUARTER 3 | QUARTER 4 | TOTAL GRAMS |
| | 570 | 1,570.12 | 1,401.89 | 1,489.76 | 1,546.42 | 6,008.19 |
| | 571 | 7,208.91 | 7,129.71 | 7,115.25 | 7,140.69 | 28,594.56 |
| | 572 | 464.21 | 441.67 | 452.65 | 458.43 | 1,816.96 |
| | 573 | 912.24 | 868.31 | 892.01 | 900.1 | 3,572.66 |
| | 574 | 555.55 | 573.48 | 575.79 | 597.76 | 2,302.58 |
| | 575 | 277.49 | 265.93 | 285.58 | 301.19 | 1,130.19 |
| | 576 | 13.87 | 12.72 | 12.14 | 0 | 38.73 |
| | 577 | 925.54 | 985.66 | 930.74 | 983.93 | 3,825.87 |
| TOTAL | | 11,927.93 | 11,679.37 | 11,753.92 | 11,928.52 | 47,289.74 |
| STATE: | TENNESSEE | | | | | |
| ZIP CODE | QUARTER 1 | | QUARTER 2 | QUARTER 3 | QUARTER 4 | TOTAL GRAMS |
| | 370 | 11,750.46 | 11,664.32 | 11,784.57 | 12,296.19 | 47,495.54 |
| | 371 | 7,517.03 | 7,396.79 | 7,722.84 | 8,007.84 | 30,644.50 |
| | 372 | 7,264.98 | 7,316.43 | 7,715.90 | 7,634.39 | 29,931.70 |
| | 373 | 5,840.54 | 5,574.04 | 5,616.82 | 5,797.19 | 22,828.59 |
| | 374 | 3,598.67 | 3,426.98 | 3,517.16 | 3,454.15 | 13,996.96 |
| | 376 | 2,667.35 | 2,486.99 | 2,396.80 | 2,573.70 | 10,124.84 |
| | 377 | 2,754.65 | 2,678.34 | 2,710.13 | 2,845.41 | 10,988.53 |
| | 378 | 4,239.79 | 4,234.58 | 4,183.13 | 4,514.96 | 17,172.46 |
| | 379 | 6,087.97 | 5,816.26 | 6,086.81 | 6,212.26 | 24,203.30 |
| | 380 | 5,800.08 | 5,518.54 | 5,582.13 | 6,017.44 | 22,918.19 |
| | 381 | 7,851.18 | 7,722.84 | 7,596.81 | 8,073.74 | 31,244.57 |
| | 382 | 853.85 | 775.23 | 772.34 | 846.34 | 3,247.76 |
| | 383 | 2,353.45 | 2,141.28 | 2,306.04 | 2,452.30 | 9,253.07 |
| | 384 | 1,842.98 | 1,720.43 | 1,748.75 | 1,822.75 | 7,134.91 |
| | 385 | 1,710.02 | 1,720.43 | 1,632.55 | 1,711.18 | 6,774.18 |
| TOTAL | | 72,133 | 70,193.48 | 71,372.78 | 74,259.84 | 287,959.10 |
| STATE: | TEXAS | | | | | |
| ZIP CODE | QUARTER 1 | | QUARTER 2 | QUARTER 3 | QUARTER 4 | TOTAL GRAMS |
| | 750 | 32,274.74 | 32,203.64 | 31,926.73 | 33,793.99 | 130,199.10 |

RETAIL DRUG DISTRIBUTION BY ZIP CODE WITHIN STATE BY GRAMS WT

STATE:TEXAS

| ZIP CODE | QUARTER 1 | QUARTER 2 | QUARTER 3 | QUARTER 4 | TOTAL GRAMS |
|---|---|---|---|---|---|
| 751 | 8,206.71 | 7,763.88 | 7,871.41 | 8,428.70 | 32,270.70 |
| 752 | 14,472.73 | 14,368.68 | 14,682.58 | 15,456.66 | 58,980.65 |
| 753 | 21.39 | 20.23 | 19.08 | 9.83 | 70.53 |
| 754 | 2,613.59 | 2,368.48 | 2,325.12 | 2,684.70 | 9,991.89 |
| 755 | 925.54 | 883.91 | 842.29 | 898.95 | 3,550.69 |
| 756 | 3,699.84 | 3,580.75 | 3,379.57 | 3,690.59 | 14,350.75 |
| 757 | 3,432.76 | 3,349.51 | 3,073.76 | 3,691.17 | 13,547.20 |
| 758 | 763.09 | 713.95 | 637.64 | 696.61 | 2,811.29 |
| 759 | 2,298.53 | 2,196.78 | 2,158.63 | 2,539.59 | 9,193.53 |
| 760 | 13,620.04 | 13,618.30 | 13,503.26 | 14,470.42 | 55,212.02 |
| 761 | 7,205.44 | 7,232.03 | 6,827.36 | 7,463.27 | 28,728.10 |
| 762 | 5,775.80 | 5,620.29 | 5,719.72 | 6,056.18 | 23,171.99 |
| 763 | 1,979.99 | 1,738.92 | 1,812.34 | 2,014.68 | 7,545.93 |
| 764 | 704.7 | 662.5 | 647.47 | 661.35 | 2,676.02 |
| 765 | 3,157 | 3,093.41 | 2,762.74 | 2,877.78 | 11,890.93 |
| 766 | 689.67 | 698.34 | 657.3 | 671.75 | 2,717.06 |
| 767 | 2,863.33 | 2,708.40 | 2,591.04 | 2,892.23 | 11,055 |
| 768 | 419.12 | 335.3 | 411.03 | 419.7 | 1,585.15 |
| 769 | 1,566.07 | 1,390.91 | 1,633.71 | 1,592.09 | 6,182.78 |
| 770 | 28,024.55 | 27,425.64 | 27,362.05 | 28,304.35 | 111,116.59 |
| 772 | 20.81 | 17.34 | 12.72 | 9.25 | 60.12 |
| 773 | 20,342.76 | 19,355.94 | 19,633.43 | 20,945.14 | 80,277.27 |
| 774 | 17,015.80 | 16,204.14 | 16,009.32 | 17,334.33 | 66,563.59 |
| 775 | 13,452.39 | 13,077.78 | 13,044.25 | 13,765.72 | 53,340.14 |
| 776 | 3,842.05 | 3,352.98 | 3,421.20 | 4,272.16 | 14,888.39 |
| 777 | 1,864.37 | 1,703.08 | 1,751.64 | 1,870.73 | 7,189.82 |
| 778 | 6,294.35 | 5,817.42 | 5,869.45 | 6,464.31 | 24,445.53 |
| 779 | 2,069.60 | 2,111.22 | 1,900.79 | 2,160.36 | 8,241.97 |
| 780 | 5,095.95 | 4,838.12 | 4,823.09 | 5,200.01 | 19,957.17 |
| 781 | 3,790.60 | 3,535.08 | 3,408.48 | 3,798.12 | 14,532.28 |
| 782 | 15,779.24 | 15,280.92 | 15,102.28 | 16,426.13 | 62,588.57 |
| 783 | 2,402.58 | 2,090.41 | 2,159.20 | 2,366.16 | 9,018.35 |
| 784 | 4,062.89 | 3,884.83 | 3,705.04 | 3,869.22 | 15,521.98 |
| 785 | 7,844.24 | 7,326.84 | 6,958.59 | 7,433.21 | 29,562.88 |
| 786 | 8,532.18 | 8,404.42 | 8,196.30 | 9,226.48 | 34,359.38 |
| 787 | 12,815.32 | 12,579.46 | 12,478.29 | 13,726.98 | 51,600.05 |
| 788 | 774.65 | 630.71 | 690.83 | 709.33 | 2,805.52 |
| 789 | 427.79 | 393.11 | 379.81 | 421.43 | 1,622.14 |
| 790 | 663.66 | 705.86 | 576.94 | 594.86 | 2,541.32 |
| 791 | 1,212.28 | 1,219.79 | 1,092.03 | 1,185.68 | 4,709.78 |
| 792 | 94.23 | 81.51 | 72.26 | 60.12 | 308.12 |
| 793 | 389.64 | 366.52 | 331.25 | 376.34 | 1,463.75 |
| 794 | 3,164.52 | 3,082.43 | 3,152.96 | 3,437.38 | 12,837.29 |
| 795 | 434.73 | 402.36 | 437.04 | 427.22 | 1,701.35 |
| 796 | 1,381.08 | 1,293.79 | 1,269.51 | 1,482.25 | 5,426.63 |
| 797 | 2,990.51 | 2,989.93 | 2,778.35 | 2,926.92 | 11,685.71 |
| 798 | 92.5 | 80.93 | 79.2 | 93.65 | 346.28 |
| 799 | 3,147.18 | 2,977.22 | 2,850.03 | 3,106.71 | 12,081.14 |
| TOTAL | 274,712.53 | 265,777.99 | 263,029.11 | 283,004.79 | 1,086,524.42 |

STATE: UTAH

| ZIP CODE | QUARTER 1 | QUARTER 2 | QUARTER 3 | QUARTER 4 | TOTAL GRAMS |
|---|---|---|---|---|---|
| 840 | 11,457.36 | 11,183.34 | 11,656.81 | 13,033.26 | 47,330.77 |
| 841 | 3,693.48 | 3,598.09 | 3,676.72 | 3,891.19 | 14,859.48 |
| 843 | 775.23 | 749.22 | 744.59 | 864.26 | 3,133.30 |
| 844 | 890.85 | 900.1 | 898.95 | 1,026.13 | 3,716.03 |
| 845 | 126.03 | 111.57 | 100.01 | 114.46 | 452.07 |
| 846 | 1,897.32 | 1,851.08 | 1,880.56 | 2,052.83 | 7,681.79 |
| 847 | 717.42 | 700.08 | 812.23 | 800.09 | 3,029.82 |

| | | | | | |
|---|---|---|---|---|---|
| TOTAL | 19,557.69 | 19,093.48 | 19,769.87 | 21,782.22 | 80,203.26 |
| STATE: | VERMONT | | | | |
| ZIP CODE | QUARTER 1 | QUARTER 2 | QUARTER 3 | QUARTER 4 | TOTAL GRAMS |
| 50 | 221.41 | 255.52 | 274.02 | 305.81 | 1,056.76 |
| 51 | 324.31 | 220.26 | 248.58 | 238.18 | 1,031.33 |
| 52 | 597.76 | 591.4 | 579.83 | 577.52 | 2,346.51 |
| 53 | 665.97 | 712.22 | 615.68 | 691.99 | 2,685.86 |
| 54 | 2,423.40 | 2,349.98 | 2,480.05 | 2,445.94 | 9,699.37 |

DRUG CODE:1205DRUG NAME:LISDEXAMFETAMINE
STATE:VERMONT

| ZIP CODE | QUARTER 1 | QUARTER 2 | QUARTER 3 | QUARTER 4 | TOTAL GRAMS |
|---|---|---|---|---|---|
| 56 | 609.9 | 635.91 | 598.91 | 574.63 | 2,419.35 |
| 57 | 874.67 | 889.12 | 835.35 | 919.76 | 3,518.90 |
| 58 | 686.2 | 671.75 | 704.7 | 796.04 | 2,858.69 |
| TOTAL | 6,403.62 | 6,326.16 | 6,337.12 | 6,549.87 | 25,616.77 |
| STATE: | VIRGIN ISLANDS | | | | |
| ZIP CODE | QUARTER 1 | QUARTER 2 | QUARTER 3 | QUARTER 4 | TOTAL GRAMS |
| 8 | 86.72 | 80.93 | 67.06 | 60.7 | 295.41 |
| TOTAL | 86.72 | 80.93 | 67.06 | 60.7 | 295.41 |
| STATE: | VIRGINIA | | | | |
| ZIP CODE | QUARTER 1 | QUARTER 2 | QUARTER 3 | QUARTER 4 | TOTAL GRAMS |
| 201 | 6,341.18 | 6,581.09 | 6,624.45 | 6,989.23 | 26,535.95 |
| 220 | 3,343.73 | 3,525.83 | 3,516.58 | 3,473.80 | 13,859.94 |
| 221 | 2,761.01 | 2,950.04 | 2,832.11 | 2,872.58 | 11,415.74 |
| 222 | 1,733.14 | 1,974.21 | 1,848.19 | 2,112.96 | 7,668.50 |
| 223 | 1,593.82 | 1,698.46 | 1,700.77 | 1,802.52 | 6,795.57 |
| 224 | 2,074.80 | 1,979.99 | 1,978.84 | 2,203.14 | 8,236.77 |
| 225 | 923.8 | 1,054.45 | 967.74 | 980.46 | 3,926.45 |
| 226 | 1,473 | 1,494.97 | 1,326.16 | 1,486.30 | 5,780.43 |
| 227 | 468.26 | 483.29 | 528.38 | 538.79 | 2,018.72 |
| 228 | 1,037.11 | 1,048.10 | 964.27 | 1,092.61 | 4,142.09 |
| 229 | 2,050.98 | 2,062.60 | 2,078.85 | 2,169.61 | 8,362.04 |
| 230 | 1,658.57 | 1,629.09 | 1,633.71 | 1,706.55 | 6,627.92 |
| 231 | 4,780.31 | 4,574.51 | 4,443.28 | 4,657.75 | 18,455.85 |
| 232 | 5,359.57 | 5,391.36 | 5,064.73 | 5,543.40 | 21,359.06 |
| 233 | 3,170.30 | 3,006.12 | 2,901.48 | 3,113.65 | 12,191.55 |
| 234 | 7,346.49 | 7,229.72 | 7,020.45 | 7,384.68 | 28,981.34 |
| 235 | 1,773.03 | 1,822.75 | 1,558.56 | 1,849.92 | 7,004.26 |
| 236 | 2,929.23 | 2,987.62 | 2,967.39 | 3,061.62 | 11,945.86 |
| 237 | 1,152.73 | 1,315.76 | 1,220.95 | 1,239.45 | 4,928.89 |
| 238 | 2,177.70 | 2,230.89 | 2,030.29 | 2,182.91 | 8,621.79 |
| 239 | 523.18 | 556.71 | 549.77 | 607.01 | 2,236.67 |
| 240 | 3,356.45 | 3,100.93 | 3,136.19 | 3,456.46 | 13,050.03 |
| 241 | 2,086.36 | 1,986.35 | 1,884.03 | 2,059.19 | 8,015.93 |
| 242 | 934.21 | 919.18 | 945.19 | 952.71 | 3,751.29 |
| 243 | 894.32 | 905.3 | 895.48 | 894.32 | 3,589.42 |
| 244 | 1,393.80 | 1,357.38 | 1,271.82 | 1,462.01 | 5,485.01 |
| 245 | 3,037.92 | 2,913.62 | 3,075.49 | 3,058.15 | 12,085.18 |
| 246 | 523.18 | 517.98 | 538.21 | 523.18 | 2,102.55 |
| TOTAL | 66,898.18 | 67,298.30 | 65,503.36 | 69,474.96 | 269,174.80 |
| STATE: | WASHINGTON | | | | |
| ZIP CODE | QUARTER 1 | QUARTER 2 | QUARTER 3 | QUARTER 4 | TOTAL GRAMS |
| 980 | 9,427.65 | 9,582.59 | 9,585.48 | 10,036.39 | 38,632.11 |
| 981 | 5,312.16 | 5,548.60 | 5,422.58 | 5,526.64 | 21,809.98 |

| | | | | | |
|---|---|---|---|---|---|
| 982 | 3,091.68 | 3,220.02 | 3,183.02 | 3,314.25 | 12,808.97 |
| 983 | 3,147.18 | 3,333.32 | 3,017.68 | 3,219.44 | 12,717.62 |
| 984 | 1,877.67 | 1,909.46 | 1,889.23 | 1,937.21 | 7,613.57 |
| 985 | 1,710.02 | 1,551.04 | 1,533.70 | 1,714.64 | 6,509.40 |
| 986 | 1,990.40 | 2,119.89 | 2,044.16 | 2,326.85 | 8,481.30 |
| 988 | 185.57 | 213.9 | 220.26 | 184.99 | 804.72 |
| 989 | 392.53 | 411.03 | 390.22 | 434.73 | 1,628.51 |
| 990 | 299.46 | 389.06 | 362.47 | 356.11 | 1,407.10 |
| 991 | 253.21 | 218.52 | 258.41 | 297.72 | 1,027.86 |
| 992 | 1,697.30 | 1,748.17 | 1,805.98 | 2,093.30 | 7,344.75 |
| 993 | 1,581.68 | 1,575.32 | 1,426.17 | 1,701.93 | 6,285.10 |
| 994 | 82.09 | 68.79 | 63.01 | 78.62 | 292.51 |
| TOTAL | 31,048.60 | 31,889.71 | 31,202.37 | 33,222.82 | 127,363.50 |

STATE: WEST VIRGINIA

| ZIP CODE | QUARTER 1 | QUARTER 2 | QUARTER 3 | QUARTER 4 | TOTAL GRAMS |
|---|---|---|---|---|---|
| 247 | 374.61 | 433.58 | 382.7 | 409.29 | 1,600.18 |
| 248 | 274.6 | 274.02 | 235.86 | 306.97 | 1,091.45 |
| 249 | 415.08 | 433.58 | 383.28 | 389.64 | 1,621.58 |
| 250 | 808.76 | 693.72 | 742.86 | 702.39 | 2,947.73 |
| 251 | 587.35 | 621.46 | 575.21 | 621.46 | 2,405.48 |
| 252 | 400.62 | 473.46 | 357.27 | 402.94 | 1,634.29 |
| 253 | 1,997.91 | 1,930.85 | 1,982.30 | 2,000.80 | 7,911.86 |
| 254 | 1,356.22 | 1,327.32 | 1,170.65 | 1,414.03 | 5,268.22 |

DRUG CODE:1205DRUG NAME:LISDEXAMFETAMINE
STATE:WEST VIRGINIA

| ZIP CODE | QUARTER 1 | QUARTER 2 | QUARTER 3 | QUARTER 4 | TOTAL GRAMS |
|---|---|---|---|---|---|
| 255 | 1,793.84 | 1,671.29 | 1,712.33 | 1,761.47 | 6,938.93 |
| 256 | 268.82 | 265.35 | 245.11 | 232.4 | 1,011.68 |
| 257 | 1,630.24 | 1,510.58 | 1,488.61 | 1,573.59 | 6,203.02 |
| 258 | 1,114 | 1,028.44 | 1,059.66 | 1,068.33 | 4,270.43 |
| 259 | 361.31 | 364.2 | 384.44 | 345.7 | 1,455.65 |
| 260 | 955.6 | 989.13 | 987.97 | 1,094.92 | 4,027.62 |
| 261 | 2,524.56 | 2,614.75 | 2,391.60 | 2,406.63 | 9,937.54 |
| 262 | 610.47 | 605.27 | 617.41 | 619.15 | 2,452.30 |
| 263 | 1,182.79 | 1,254.48 | 1,327.32 | 1,404.78 | 5,169.37 |
| 264 | 364.2 | 386.17 | 349.75 | 365.94 | 1,466.06 |
| 265 | 2,061.50 | 1,999.65 | 2,010.05 | 2,123.94 | 8,195.14 |
| 266 | 238.18 | 268.82 | 267.66 | 231.82 | 1,006.48 |
| 267 | 766.56 | 691.41 | 650.94 | 745.17 | 2,854.08 |
| 268 | 431.26 | 418.54 | 395.42 | 412.19 | 1,657.41 |
| TOTAL | 20,518.48 | 20,256.07 | 19,718.40 | 20,633.55 | 81,126.50 |

STATE: WISCONSIN

| ZIP CODE | QUARTER 1 | QUARTER 2 | QUARTER 3 | QUARTER 4 | TOTAL GRAMS |
|---|---|---|---|---|---|
| 530 | 6,063.11 | 6,457.96 | 6,260.82 | 6,775.33 | 25,557.22 |
| 531 | 6,471.83 | 6,723.88 | 6,865.52 | 7,452.87 | 27,514.10 |
| 532 | 7,345.92 | 7,634.39 | 7,451.71 | 8,514.26 | 30,946.28 |
| 534 | 2,323.38 | 2,281.76 | 2,209.50 | 2,180.02 | 8,994.66 |
| 535 | 4,057.11 | 4,246.72 | 4,180.24 | 4,428.82 | 16,912.89 |
| 537 | 2,401.43 | 2,582.37 | 2,595.09 | 2,906.69 | 10,485.58 |
| 538 | 715.69 | 749.22 | 737.66 | 852.7 | 3,055.27 |
| 539 | 1,492.08 | 1,541.79 | 1,585.15 | 1,722.74 | 6,341.76 |
| 540 | 513.93 | 549.2 | 581.57 | 712.8 | 2,357.50 |
| 541 | 3,495.77 | 3,507.91 | 3,504.44 | 3,698.11 | 14,206.23 |
| 542 | 1,516.93 | 1,432.53 | 1,479.94 | 1,583.42 | 6,012.82 |
| 543 | 4,890.73 | 4,869.34 | 4,973.39 | 5,261.29 | 19,994.75 |
| 544 | 4,910.96 | 5,047.39 | 5,079.76 | 5,309.27 | 20,347.38 |
| 545 | 1,137.12 | 1,283.38 | 1,251.01 | 1,183.95 | 4,855.46 |

| | | | | | |
|---|---|---|---|---|---|
| 546 | 1,244.07 | 1,431.95 | 1,388.02 | 1,651.63 | 5,715.67 |
| 547 | 993.18 | 1,051.56 | 1,150.42 | 1,312.29 | 4,507.45 |
| 548 | 1,301.30 | 1,330.21 | 1,331.94 | 1,372.41 | 5,335.86 |
| 549 | 6,303.60 | 6,541.20 | 6,608.26 | 7,032.59 | 26,485.65 |
| TOTAL | 57,178.14 | 59,262.76 | 59,234.44 | 63,951.19 | 239,626.53 |

STATE: WYOMING

| ZIP CODE | QUARTER 1 | QUARTER 2 | QUARTER 3 | QUARTER 4 | TOTAL GRAMS |
|---|---|---|---|---|---|
| 820 | 639.38 | 599.49 | 626.08 | 689.67 | 2,554.62 |
| 822 | 104.64 | 89.03 | 75.15 | 71.68 | 340.5 |
| 823 | 32.95 | 50.29 | 26.01 | 50.29 | 159.54 |
| 824 | 404.09 | 426.06 | 530.7 | 443.98 | 1,804.83 |
| 825 | 65.33 | 83.82 | 52.61 | 82.09 | 283.85 |
| 826 | 287.32 | 348.02 | 309.28 | 353.22 | 1,297.84 |
| 827 | 274.6 | 354.38 | 271.71 | 296.57 | 1,197.26 |
| 828 | 247.43 | 214.48 | 200.02 | 232.97 | 894.9 |
| 829 | 385.59 | 400.05 | 382.12 | 363.62 | 1,531.38 |
| 830 | 130.07 | 124.87 | 113.31 | 118.51 | 486.76 |
| 831 | 29.48 | 28.33 | 39.89 | 25.44 | 123.14 |
| TOTAL | 2,600.88 | 2,718.82 | 2,626.88 | 2,728.04 | 10,674.62 |

DRUG CODE:1724DRUG NAME:METHYLPHENIDATE (DL;D;L;ISOMERS)

STATE: ALABAMA

| ZIP CODE | QUARTER 1 | QUARTER 2 | QUARTER 3 | QUARTER 4 | TOTAL GRAMS |
|---|---|---|---|---|---|
| 350 | 9,015.86 | 8,431.91 | 8,602.62 | 8,780.84 | 34,831.23 |
| 351 | 5,002.63 | 4,682.63 | 4,631.50 | 5,007.62 | 19,324.38 |
| 352 | 9,660.17 | 8,802.94 | 8,558.93 | 9,237.71 | 36,259.75 |
| 354 | 2,668.31 | 2,534.69 | 2,542.67 | 2,797.14 | 10,542.81 |
| 355 | 2,306.90 | 1,995.09 | 2,106.42 | 2,132.27 | 8,540.68 |
| 356 | 4,383.81 | 4,066.46 | 4,174.05 | 4,276.50 | 16,900.82 |
| 357 | 2,557.55 | 2,423.69 | 2,508.08 | 2,531.12 | 10,020.44 |
| 358 | 2,880.46 | 2,887.26 | 2,921.62 | 2,793.28 | 11,482.62 |
| 359 | 4,075.87 | 3,684.48 | 3,808.45 | 4,016.67 | 15,585.47 |
| 360 | 3,973.58 | 3,583.61 | 3,695.20 | 3,893.74 | 15,146.13 |
| 361 | 3,343.71 | 2,716.25 | 3,001.65 | 3,150.93 | 12,212.54 |
| 362 | 2,145.64 | 1,926.18 | 2,099.46 | 2,153.57 | 8,324.85 |

DRUG CODE:1724DRUG NAME:METHYLPHENIDATE (DL;D;L;ISOMERS)
STATE:ALABAMA

| ZIP CODE | QUARTER 1 | QUARTER 2 | QUARTER 3 | QUARTER 4 | TOTAL GRAMS |
|---|---|---|---|---|---|
| 363 | 5,984.77 | 5,547.03 | 6,035.48 | 5,785.99 | 23,353.27 |
| 364 | 1,469.14 | 1,240.40 | 1,326.41 | 1,467.68 | 5,503.63 |
| 365 | 6,272.31 | 6,021.98 | 6,115.30 | 6,553.07 | 24,962.66 |
| 366 | 5,134.34 | 4,766.36 | 4,904.52 | 5,356.72 | 20,161.94 |
| 367 | 736.96 | 602.52 | 679.95 | 659.79 | 2,679.22 |
| 368 | 3,794.21 | 3,345.90 | 3,452.24 | 3,734.90 | 14,327.25 |
| 369 | 167.8 | 152.24 | 159.8 | 220.7 | 700.54 |
| TOTAL | 75,574.02 | 69,411.62 | 71,324.35 | 74,550.24 | 290,860.23 |

STATE: ALASKA

| ZIP CODE | QUARTER 1 | QUARTER 2 | QUARTER 3 | QUARTER 4 | TOTAL GRAMS |
|---|---|---|---|---|---|
| 995 | 4,614.88 | 4,591.27 | 4,053.29 | 4,357.61 | 17,617.05 |
| 996 | 1,573.27 | 1,481.86 | 1,401.77 | 1,576.22 | 6,033.12 |
| 997 | 1,370.07 | 1,354.87 | 1,254.74 | 1,344.43 | 5,324.11 |
| 998 | 525.32 | 612.08 | 442.74 | 522.37 | 2,102.51 |
| 999 | 467.16 | 453.32 | 410.62 | 410.73 | 1,741.83 |
| TOTAL | 8,550.70 | 8,493.40 | 7,563.16 | 8,211.36 | 32,818.62 |

STATE: AMERICAN SAMOA

RETAIL DRUG DISTRIBUTION BY ZIP CODE WITHIN STATE BY GRAMS WT

| ZIP CODE | QUARTER 1 | QUARTER 2 | QUARTER 3 | QUARTER 4 | TOTAL GRAMS |
|---|---|---|---|---|---|
| 967 | 1.3 | 1.73 | 9.08 | 2.59 | 14.7 |
| TOTAL | 1.3 | 1.73 | 9.08 | 2.59 | 14.7 |

STATE: ARIZONA

| ZIP CODE | QUARTER 1 | QUARTER 2 | QUARTER 3 | QUARTER 4 | TOTAL GRAMS |
|---|---|---|---|---|---|
| 850 | 12,612.24 | 12,195.98 | 11,601.56 | 12,348.29 | 48,758.07 |
| 851 | 3,630.65 | 3,411.76 | 3,475.75 | 3,694.71 | 14,212.87 |
| 852 | 25,537.28 | 23,742.70 | 23,696.10 | 25,637.21 | 98,613.29 |
| 853 | 12,882.62 | 11,458.80 | 11,716.08 | 12,167.89 | 48,225.39 |
| 855 | 832.24 | 747.74 | 759.33 | 805.84 | 3,145.15 |
| 856 | 2,926.29 | 2,704.84 | 2,625.77 | 2,944.80 | 11,201.70 |
| 857 | 12,055.08 | 11,624.51 | 11,725.82 | 12,162.61 | 47,568.02 |
| 859 | 764.16 | 682.28 | 759.7 | 792.39 | 2,998.53 |
| 860 | 1,478.62 | 1,441.94 | 1,513.22 | 1,456.96 | 5,890.74 |
| 863 | 1,909.79 | 1,829.70 | 1,892.77 | 2,085.52 | 7,717.78 |
| 864 | 1,205.36 | 1,173.62 | 1,111.12 | 1,141.96 | 4,632.06 |
| 865 | 365.68 | 331.09 | 439.87 | 315.44 | 1,452.08 |
| TOTAL | 76,200.01 | 71,344.96 | 71,317.09 | 75,553.62 | 294,415.68 |

STATE: ARKANSAS

| ZIP CODE | QUARTER 1 | QUARTER 2 | QUARTER 3 | QUARTER 4 | TOTAL GRAMS |
|---|---|---|---|---|---|
| 716 | 1,673.87 | 1,515.66 | 1,491.54 | 1,631.12 | 6,312.19 |
| 717 | 1,172.81 | 974.03 | 975.37 | 1,160.48 | 4,282.69 |
| 718 | 1,170.03 | 1,042.68 | 995.55 | 1,073.25 | 4,281.51 |
| 719 | 2,407.54 | 2,043.42 | 2,085.69 | 2,252.61 | 8,789.26 |
| 720 | 6,871.82 | 5,917.77 | 6,075.35 | 6,548.97 | 25,413.91 |
| 721 | 5,335.48 | 4,890.07 | 4,651.18 | 4,879.65 | 19,756.38 |
| 722 | 4,774.88 | 4,330.30 | 4,275.40 | 4,341.56 | 17,722.14 |
| 723 | 1,701.59 | 1,406.13 | 1,440.45 | 1,451.26 | 5,999.43 |
| 724 | 4,140.18 | 3,663.05 | 3,609.96 | 3,900.50 | 15,313.69 |
| 725 | 1,231.92 | 1,218.38 | 1,069.21 | 1,182.99 | 4,702.50 |
| 726 | 1,502.28 | 1,382.67 | 1,395.22 | 1,483.98 | 5,764.15 |
| 727 | 7,504.28 | 7,099.32 | 6,891.59 | 7,224.22 | 28,719.41 |
| 728 | 1,475.98 | 1,379.66 | 1,389.94 | 1,422.71 | 5,668.29 |
| 729 | 3,457.44 | 2,995.05 | 2,953.27 | 3,126.80 | 12,532.56 |
| TOTAL | 44,420.10 | 39,858.19 | 39,299.72 | 41,680.10 | 165,258.11 |

STATE: CALIFORNIA

| ZIP CODE | QUARTER 1 | QUARTER 2 | QUARTER 3 | QUARTER 4 | TOTAL GRAMS |
|---|---|---|---|---|---|
| 900 | 10,058.47 | 10,221.08 | 9,377.54 | 10,194 | 39,851.09 |
| 902 | 7,727.56 | 7,633.04 | 7,088.45 | 7,334.96 | 29,784.01 |
| 903 | 396.55 | 413.46 | 404.85 | 430.62 | 1,645.48 |
| 904 | 1,260.78 | 1,332.60 | 1,374.44 | 1,369 | 5,336.82 |
| 905 | 1,354.03 | 1,382.04 | 1,170.53 | 1,393.74 | 5,300.34 |
| 906 | 3,949.85 | 3,523.08 | 3,276.03 | 3,417.20 | 14,166.16 |
| 907 | 3,452 | 3,324.15 | 3,312.37 | 3,211.41 | 13,299.93 |
| 908 | 2,224.13 | 2,234.14 | 2,291.41 | 2,268.28 | 9,017.96 |
| 910 | 1,414.62 | 1,446.96 | 1,450.07 | 1,506.98 | 5,818.63 |
| 911 | 2,195.25 | 2,119.13 | 2,029.22 | 2,122.34 | 8,465.94 |
| 912 | 1,033.69 | 1,009.43 | 1,054.75 | 1,013.93 | 4,111.80 |
| 913 | 12,811.71 | 12,592.55 | 11,880.27 | 12,437.39 | 49,721.92 |

DRUG CODE:1724DRUG NAME:METHYLPHENIDATE (DL;D;L;ISOMERS)
STATE:CALIFORNIA

| ZIP CODE | QUARTER 1 | QUARTER 2 | QUARTER 3 | QUARTER 4 | TOTAL GRAMS |
|---|---|---|---|---|---|
| 914 | 2,872.64 | 2,771.49 | 2,634.38 | 2,735.97 | 11,014.48 |
| 915 | 842.05 | 796.5 | 796.67 | 823.02 | 3,258.24 |
| 916 | 864.57 | 858.12 | 748.51 | 811.6 | 3,282.80 |

RETAIL DRUG DISTRIBUTION BY ZIP CODE WITHIN STATE BY GRAMS WT

| | | | | | |
|---|---|---|---|---|---|
| 917 | 9,181.97 | 8,672.75 | 8,154.25 | 8,710.30 | 34,719.27 |
| 918 | 234.68 | 227.83 | 232.69 | 272.16 | 967.36 |
| 919 | 2,694.44 | 2,805.92 | 2,624.76 | 2,874.76 | 10,999.88 |
| 920 | 28,075.90 | 28,088.72 | 27,834.88 | 29,025.82 | 113,025.32 |
| 921 | 11,829.12 | 11,483.16 | 11,161.65 | 11,911.25 | 46,385.18 |
| 922 | 4,551.60 | 4,258.29 | 3,668.68 | 4,422.37 | 16,900.94 |
| 923 | 7,847.70 | 7,523.36 | 6,771.54 | 7,768.42 | 29,911.02 |
| 924 | 1,038.56 | 928.62 | 946.53 | 980.41 | 3,894.12 |
| 925 | 9,090.27 | 8,948.36 | 8,203.76 | 9,065.55 | 35,307.94 |
| 926 | 12,597.72 | 12,595.81 | 11,401.71 | 12,198.70 | 48,793.94 |
| 927 | 3,289.99 | 3,437.23 | 2,862.22 | 3,047.74 | 12,637.18 |
| 928 | 8,012.47 | 8,138.71 | 7,362.75 | 8,401.81 | 31,915.74 |
| 930 | 5,810.48 | 5,735.47 | 5,685.43 | 5,892.94 | 23,124.32 |
| 931 | 1,754.28 | 1,746.60 | 1,616.95 | 1,699.41 | 6,817.24 |
| 932 | 3,455.61 | 3,479.94 | 3,072.03 | 3,314.30 | 13,321.88 |
| 933 | 2,937.92 | 2,962.93 | 2,715.09 | 2,988.20 | 11,604.14 |
| 934 | 3,789.21 | 3,554.40 | 3,433.08 | 3,628.99 | 14,405.68 |
| 935 | 3,459.33 | 3,472.33 | 3,180.76 | 3,364.36 | 13,476.78 |
| 936 | 3,951.61 | 3,759.21 | 3,668.85 | 3,873.58 | 15,253.25 |
| 937 | 5,900.31 | 5,504.28 | 5,318.78 | 5,672.13 | 22,395.50 |
| 939 | 2,273.02 | 2,173.28 | 2,070.35 | 2,122.88 | 8,639.53 |
| 940 | 5,730.05 | 5,495.45 | 5,266.78 | 5,907.70 | 22,399.98 |
| 941 | 6,432.18 | 6,320.74 | 6,025.31 | 6,074.36 | 24,852.59 |
| 943 | 1,242.79 | 1,146.52 | 1,101.34 | 1,251.34 | 4,741.99 |
| 944 | 1,009.13 | 1,004.49 | 1,010.92 | 987.03 | 4,011.57 |
| 945 | 19,912.52 | 19,738.76 | 17,941.88 | 19,433.96 | 77,027.12 |
| 946 | 2,935.27 | 3,229.80 | 3,012.03 | 2,856.58 | 12,033.68 |
| 947 | 868.11 | 798.67 | 729.5 | 876.35 | 3,272.63 |
| 948 | 920.81 | 558.34 | 590.75 | 856.81 | 2,926.71 |
| 949 | 3,624.82 | 3,784.84 | 3,844.84 | 3,869.66 | 15,124.16 |
| 950 | 7,785.12 | 6,955.38 | 6,366.35 | 6,538.57 | 27,645.42 |
| 951 | 4,857.03 | 4,491.21 | 4,025.56 | 4,607.75 | 17,981.55 |
| 952 | 3,282.22 | 3,130.02 | 2,888.63 | 2,986.84 | 12,287.71 |
| 953 | 6,550.70 | 5,994.78 | 5,792.28 | 6,181 | 24,518.76 |
| 954 | 3,558 | 3,417.71 | 3,403.83 | 3,778.18 | 14,157.72 |
| 955 | 895.75 | 1,013.77 | 897.65 | 973.11 | 3,780.28 |
| 956 | 13,988.69 | 14,068.32 | 12,774.73 | 14,844.57 | 55,676.31 |
| 957 | 3,327.39 | 2,854.71 | 2,830.54 | 3,243.17 | 12,255.81 |
| 958 | 6,489.18 | 6,035.90 | 5,702.65 | 6,153.61 | 24,381.34 |
| 959 | 4,179.22 | 4,042.91 | 3,758.05 | 3,872.45 | 15,852.63 |
| 960 | 3,148.93 | 2,891.84 | 2,855.89 | 2,926.76 | 11,823.42 |
| 961 | 445.42 | 447.98 | 411.52 | 502.57 | 1,807.49 |
| TOTAL | 285,417.42 | 278,577.11 | 262,107.26 | 281,028.89 | 1,107,130.68 |

STATE: COLORADO

| ZIP CODE | QUARTER 1 | QUARTER 2 | QUARTER 3 | QUARTER 4 | TOTAL GRAMS |
|---|---|---|---|---|---|
| 800 | 10,330.77 | 9,971.72 | 9,890.54 | 10,236.47 | 40,429.50 |
| 801 | 10,945.49 | 10,590.28 | 10,533.09 | 10,985.63 | 43,054.49 |
| 802 | 10,893.83 | 10,387.73 | 9,899.20 | 10,649.72 | 41,830.48 |
| 803 | 1,795.62 | 1,868.95 | 1,797.14 | 2,013.10 | 7,474.81 |
| 804 | 1,793.36 | 1,640.86 | 1,693.18 | 1,837.73 | 6,965.13 |
| 805 | 6,796.98 | 6,473.08 | 6,296.66 | 6,829.16 | 26,395.88 |
| 806 | 2,320.52 | 2,231.36 | 2,273.97 | 2,379.08 | 9,204.93 |
| 807 | 470.55 | 442.56 | 371.74 | 417.32 | 1,702.17 |
| 808 | 1,662.80 | 1,666.02 | 1,463.98 | 1,658.94 | 6,451.74 |
| 809 | 8,033.72 | 7,829.71 | 8,133.10 | 7,630.64 | 31,627.17 |
| 810 | 2,306.07 | 2,076.43 | 2,069.05 | 2,098.38 | 8,549.93 |
| 811 | 336.34 | 328.74 | 311.66 | 341.32 | 1,318.06 |
| 812 | 771.23 | 702.68 | 651.2 | 649.69 | 2,774.80 |
| 813 | 588.23 | 600.29 | 519.62 | 528.25 | 2,236.39 |
| 814 | 661.3 | 654.15 | 657.29 | 744.96 | 2,717.70 |
| 815 | 2,015.24 | 1,921.41 | 2,048.89 | 1,944.52 | 7,930.06 |

| | | | | | |
|---|---|---|---|---|---|
| 816 | 1,137.64 | 1,051.35 | 1,101.25 | 1,070.65 | 4,360.89 |

DRUG CODE:1724DRUG NAME:METHYLPHENIDATE (DL;D;L;ISOMERS)
STATE:COLORADO

| ZIP CODE | QUARTER 1 | QUARTER 2 | QUARTER 3 | QUARTER 4 | TOTAL GRAMS |
|---|---|---|---|---|---|
| TOTAL | 62,859.69 | 60,437.32 | 59,711.56 | 62,015.56 | 245,024.13 |

STATE: CONNECTICUT

| ZIP CODE | QUARTER 1 | QUARTER 2 | QUARTER 3 | QUARTER 4 | TOTAL GRAMS |
|---|---|---|---|---|---|
| 60 | 12,209.68 | 11,198.73 | 10,925.19 | 11,741.87 | 46,075.47 |
| 61 | 4,453.86 | 4,142.88 | 3,785.54 | 4,202.21 | 16,584.49 |
| 62 | 2,665.45 | 2,552.21 | 2,395.13 | 2,623.69 | 10,236.48 |
| 63 | 4,876.42 | 4,997.04 | 4,924.66 | 4,593.47 | 19,391.59 |
| 64 | 12,862.87 | 12,561 | 12,035.19 | 12,532.12 | 49,991.18 |
| 65 | 3,770.59 | 3,542.90 | 3,184.54 | 3,410.78 | 13,908.81 |
| 66 | 1,704.11 | 1,563.63 | 1,458.46 | 1,610.94 | 6,337.14 |
| 67 | 4,925.93 | 4,871.14 | 4,485.66 | 4,793.45 | 19,076.18 |
| 68 | 9,720.37 | 9,247.15 | 8,269.79 | 9,083.56 | 36,320.87 |
| 69 | 1,094.39 | 1,224.02 | 979.29 | 1,065.08 | 4,362.78 |
| TOTAL | 58,283.67 | 55,900.70 | 52,443.45 | 55,657.17 | 222,284.99 |

STATE: DELAWARE

| ZIP CODE | QUARTER 1 | QUARTER 2 | QUARTER 3 | QUARTER 4 | TOTAL GRAMS |
|---|---|---|---|---|---|
| 197 | 7,466.91 | 7,236.38 | 6,782.85 | 7,426.26 | 28,912.40 |
| 198 | 5,745.04 | 5,609.81 | 5,184.44 | 5,448.73 | 21,988.02 |
| 199 | 6,922.24 | 6,205.47 | 5,907.41 | 6,614.29 | 25,649.41 |
| TOTAL | 20,134.19 | 19,051.66 | 17,874.70 | 19,489.28 | 76,549.83 |

STATE: DISTRICT OF COLUMBIA

| ZIP CODE | QUARTER 1 | QUARTER 2 | QUARTER 3 | QUARTER 4 | TOTAL GRAMS |
|---|---|---|---|---|---|
| 200 | 7,560.65 | 7,443.62 | 7,518.11 | 7,538.80 | 30,061.18 |
| 203 | 311.44 | 151.09 | 304.95 | 124.08 | 891.56 |
| 204 | 517.7 | 407.62 | 605.12 | 364.13 | 1,894.57 |
| TOTAL | 8,389.79 | 8,002.33 | 8,428.18 | 8,027.01 | 32,847.31 |

STATE: FLORIDA

| ZIP CODE | QUARTER 1 | QUARTER 2 | QUARTER 3 | QUARTER 4 | TOTAL GRAMS |
|---|---|---|---|---|---|
| 320 | 8,572.98 | 7,849.54 | 7,799.14 | 8,381.64 | 32,603.30 |
| 321 | 5,830.73 | 5,186.66 | 5,105.83 | 5,273.85 | 21,397.07 |
| 322 | 13,970.64 | 12,557.72 | 13,421.93 | 12,816.75 | 52,767.04 |
| 323 | 5,484.03 | 4,948.24 | 4,786.23 | 5,330.60 | 20,549.10 |
| 324 | 6,753.07 | 6,385.19 | 5,980.45 | 6,494.27 | 25,612.98 |
| 325 | 13,632.20 | 12,601.14 | 12,934.48 | 13,575.80 | 52,743.62 |
| 326 | 5,645.57 | 5,409.82 | 5,156.87 | 5,759.37 | 21,971.63 |
| 327 | 12,494.99 | 11,999.27 | 11,481.64 | 12,517.28 | 48,493.18 |
| 328 | 8,238.08 | 7,709.39 | 7,702.50 | 8,135.46 | 31,785.43 |
| 329 | 10,529.39 | 9,813.78 | 9,626.98 | 9,770.94 | 39,741.09 |
| 330 | 10,967.22 | 10,537.22 | 9,481.74 | 10,407.28 | 41,393.46 |
| 331 | 12,337.02 | 11,051.44 | 9,834.74 | 11,493.55 | 44,716.75 |
| 333 | 4,837.87 | 4,791.67 | 4,501.57 | 4,851.82 | 18,982.93 |
| 334 | 13,963.40 | 13,171 | 12,324.80 | 13,713.93 | 53,173.13 |
| 335 | 9,636.60 | 9,281.49 | 9,375.60 | 10,134.89 | 38,428.58 |
| 336 | 10,007.41 | 9,874.30 | 9,993.33 | 9,903.37 | 39,778.41 |
| 337 | 8,154.32 | 7,294.35 | 6,923.65 | 7,367.54 | 29,739.86 |
| 338 | 9,036.95 | 8,361.53 | 8,142.55 | 8,428.61 | 33,969.64 |
| 339 | 9,057.35 | 8,772.70 | 8,344.96 | 9,120.64 | 35,295.65 |
| 341 | 3,612.93 | 3,250.69 | 2,937.95 | 3,525.61 | 13,327.18 |
| 342 | 9,811.96 | 8,979.30 | 8,767.98 | 9,483.68 | 37,042.92 |
| 344 | 6,051.22 | 5,196.01 | 5,188.36 | 5,776.16 | 22,211.75 |

| | | | | | | |
|---|---|---|---|---|---|---|
| | 346 | 8,032.01 | 7,647.92 | 7,627.18 | 8,320.72 | 31,627.83 |
| | 347 | 6,718.84 | 6,643.99 | 6,370.59 | 6,982.08 | 26,715.50 |
| | 349 | 5,107.35 | 4,624.82 | 4,373.56 | 4,868.91 | 18,974.64 |
| TOTAL | | 218,484.13 | 203,939.18 | 198,184.61 | 212,434.75 | 833,042.67 |

STATE: GEORGIA

| ZIP CODE | QUARTER 1 | QUARTER 2 | QUARTER 3 | QUARTER 4 | TOTAL GRAMS |
|---|---|---|---|---|---|
| 300 | 32,810.15 | 30,221.43 | 30,328.09 | 32,046.01 | 125,405.68 |
| 301 | 20,188.37 | 17,834.29 | 18,689.58 | 19,518.77 | 76,231.01 |
| 302 | 15,787.73 | 14,189.55 | 14,247.96 | 15,091.22 | 59,316.46 |
| 303 | 12,657.20 | 11,611.53 | 12,036.32 | 13,401.58 | 49,706.63 |
| 304 | 4,978.02 | 4,422.36 | 4,550.62 | 4,510.69 | 18,461.69 |
| 305 | 10,593.31 | 9,743.70 | 9,792.02 | 10,241.39 | 40,370.42 |
| 306 | 6,019.45 | 5,601.22 | 5,760.63 | 5,833.64 | 23,214.94 |
| 307 | 6,513.23 | 6,233.47 | 6,442.11 | 6,575.34 | 25,764.15 |
| 308 | 3,341.66 | 2,980.81 | 2,981.31 | 3,354.69 | 12,658.47 |
| 309 | 5,551.36 | 4,809.70 | 5,223.95 | 4,928.30 | 20,513.31 |

DRUG CODE:1724DRUG NAME:METHYLPHENIDATE (DL;D;L;ISOMERS)
STATE:GEORGIA

| ZIP CODE | QUARTER 1 | QUARTER 2 | QUARTER 3 | QUARTER 4 | TOTAL GRAMS |
|---|---|---|---|---|---|
| 310 | 11,998.86 | 10,835.38 | 11,026.89 | 11,600.41 | 45,461.54 |
| 312 | 4,137.99 | 3,850.69 | 3,822.11 | 3,897.95 | 15,708.74 |
| 313 | 5,067.12 | 4,695.78 | 5,651.95 | 4,611.88 | 20,026.73 |
| 314 | 4,389 | 4,163.24 | 4,084.96 | 4,124.43 | 16,761.63 |
| 315 | 9,543.51 | 8,097.60 | 8,778.21 | 8,336.42 | 34,755.74 |
| 316 | 4,189.81 | 3,555.44 | 3,691.42 | 3,998.57 | 15,435.24 |
| 317 | 9,934.71 | 8,091.79 | 8,514.13 | 8,766.99 | 35,307.62 |
| 318 | 762.65 | 667.91 | 562.85 | 581.39 | 2,574.80 |
| 319 | 5,631.24 | 5,106.28 | 5,216.88 | 4,975.39 | 20,929.79 |
| 398 | 1,738.37 | 1,476.81 | 1,490.57 | 1,572.26 | 6,278.01 |
| TOTAL | 175,833.74 | 158,188.98 | 162,892.56 | 167,967.32 | 664,882.60 |

STATE: GUAM

| ZIP CODE | QUARTER 1 | QUARTER 2 | QUARTER 3 | QUARTER 4 | TOTAL GRAMS |
|---|---|---|---|---|---|
| 969 | 437.21 | 397.25 | 438.24 | 572.6 | 1,845.30 |
| TOTAL | 437.21 | 397.25 | 438.24 | 572.6 | 1,845.30 |

STATE: HAWAII

| ZIP CODE | QUARTER 1 | QUARTER 2 | QUARTER 3 | QUARTER 4 | TOTAL GRAMS |
|---|---|---|---|---|---|
| 967 | 6,063.76 | 5,600.28 | 5,474.31 | 5,820.66 | 22,959.01 |
| 968 | 5,639.90 | 5,874.97 | 6,326.36 | 5,362.75 | 23,203.98 |
| TOTAL | 11,703.66 | 11,475.25 | 11,800.67 | 11,183.41 | 46,162.99 |

STATE: IDAHO

| ZIP CODE | QUARTER 1 | QUARTER 2 | QUARTER 3 | QUARTER 4 | TOTAL GRAMS |
|---|---|---|---|---|---|
| 832 | 2,769.94 | 2,691.68 | 2,558.34 | 2,785.12 | 10,805.08 |
| 833 | 2,687.77 | 2,606.02 | 2,482.04 | 2,651.85 | 10,427.68 |
| 834 | 3,466.17 | 3,339.78 | 3,272.86 | 3,304.10 | 13,382.91 |
| 835 | 899.02 | 842.49 | 817.8 | 841.34 | 3,400.65 |
| 836 | 9,194.57 | 9,375.36 | 8,540.87 | 8,992.02 | 36,102.82 |
| 837 | 6,919.01 | 7,035.50 | 6,247.62 | 6,571.91 | 26,774.04 |
| 838 | 2,903.41 | 2,785.83 | 2,642.64 | 2,801.55 | 11,133.43 |
| TOTAL | 28,839.89 | 28,676.66 | 26,562.17 | 27,947.89 | 112,026.61 |

STATE: ILLINOIS

| ZIP CODE | QUARTER 1 | QUARTER 2 | QUARTER 3 | QUARTER 4 | TOTAL GRAMS |
|---|---|---|---|---|---|
| 600 | 28,613.96 | 27,704.77 | 26,608.52 | 27,902.28 | 110,829.53 |

| | | | | | |
|---|---|---|---|---|---|
| 601 | 20,418.91 | 19,472.81 | 18,293.52 | 19,914.69 | 78,099.93 |
| 602 | 1,443.67 | 1,419.33 | 1,311.20 | 1,334.70 | 5,508.90 |
| 603 | 1,417.04 | 1,234.96 | 1,262.46 | 1,324.57 | 5,239.03 |
| 604 | 14,959.09 | 13,692 | 12,973.01 | 14,328.07 | 55,952.17 |
| 605 | 16,800.15 | 15,581.35 | 15,262.50 | 16,578.69 | 64,222.69 |
| 606 | 17,004.66 | 16,544.80 | 15,014.27 | 17,120 | 65,683.73 |
| 607 | 1,252.17 | 1,181.27 | 1,186.16 | 1,354.39 | 4,973.99 |
| 608 | 687.25 | 585.91 | 522.23 | 658.47 | 2,453.86 |
| 609 | 2,246.16 | 2,285.67 | 2,138.77 | 2,384.93 | 9,055.53 |
| 610 | 4,219.39 | 3,923.39 | 3,760.61 | 3,920.26 | 15,823.65 |
| 611 | 4,316.94 | 4,021.34 | 3,942 | 4,183.04 | 16,463.32 |
| 612 | 3,557.36 | 3,249.78 | 3,387.47 | 3,391.27 | 13,585.88 |
| 613 | 2,556.52 | 2,395.83 | 2,304.50 | 2,455.01 | 9,711.86 |
| 614 | 2,527.79 | 2,180.83 | 2,157.60 | 2,302.07 | 9,168.29 |
| 615 | 4,550.04 | 4,327.54 | 4,134.43 | 4,615.96 | 17,627.97 |
| 616 | 5,698.21 | 5,162.92 | 5,075.16 | 5,515.88 | 21,452.17 |
| 617 | 6,524.96 | 6,148.05 | 5,849.26 | 6,459.75 | 24,982.02 |
| 618 | 6,916.42 | 6,406.44 | 6,214.90 | 6,727.61 | 26,265.37 |
| 619 | 2,492.76 | 2,270.10 | 2,208.44 | 2,461.16 | 9,432.46 |
| 620 | 5,909.43 | 5,522.13 | 5,132.84 | 5,706.71 | 22,271.11 |
| 622 | 7,550.43 | 7,014.66 | 6,866.26 | 7,507.96 | 28,939.31 |
| 623 | 2,368.35 | 2,131.79 | 2,135.81 | 2,185.59 | 8,821.54 |
| 624 | 2,279.53 | 2,075 | 2,126.27 | 2,314.06 | 8,794.86 |
| 625 | 3,405.62 | 3,188.20 | 3,006.55 | 3,130.02 | 12,730.39 |
| 626 | 4,274.85 | 3,805.03 | 3,776.79 | 4,202.71 | 16,059.38 |
| 627 | 5,136.70 | 4,687.68 | 4,512.63 | 4,941.31 | 19,278.32 |
| 628 | 3,827.07 | 3,543.36 | 3,369.52 | 3,679.48 | 14,419.43 |
| 629 | 4,875.33 | 4,462.63 | 4,407.29 | 4,439.20 | 18,184.45 |
| TOTAL | 187,830.76 | 176,219.57 | 168,940.97 | 183,039.84 | 716,031.14 |

STATE: INDIANA

| ZIP CODE | QUARTER 1 | QUARTER 2 | QUARTER 3 | QUARTER 4 | TOTAL GRAMS |
|---|---|---|---|---|---|
| 460 | 13,840.59 | 12,966.71 | 12,943.89 | 13,838.72 | 53,589.91 |
| 461 | 11,404.82 | 10,866.22 | 10,569.17 | 11,646.16 | 44,486.37 |

DRUG CODE:1724DRUG NAME:METHYLPHENIDATE (DL;D;L;ISOMERS)
STATE:INDIANA

| ZIP CODE | QUARTER 1 | QUARTER 2 | QUARTER 3 | QUARTER 4 | TOTAL GRAMS |
|---|---|---|---|---|---|
| 462 | 18,573.86 | 17,446.42 | 17,072.20 | 17,376.56 | 70,469.04 |
| 463 | 10,111.84 | 9,795.27 | 9,333.44 | 10,271.98 | 39,512.53 |
| 464 | 997.07 | 940.84 | 873.8 | 924.19 | 3,735.90 |
| 465 | 8,861.48 | 8,180.35 | 8,016.45 | 8,551.14 | 33,609.42 |
| 466 | 2,687.31 | 2,526.38 | 2,434.37 | 2,667.37 | 10,315.43 |
| 467 | 5,087.20 | 4,895.63 | 4,841.15 | 5,205.73 | 20,029.71 |
| 468 | 7,867.07 | 7,119.20 | 7,066 | 7,585.66 | 29,637.93 |
| 469 | 5,812.42 | 5,126.32 | 4,931.59 | 5,460.25 | 21,330.58 |
| 470 | 1,560.89 | 1,437.45 | 1,368.06 | 1,628.91 | 5,995.31 |
| 471 | 8,042.07 | 7,354.27 | 6,715.40 | 6,566.27 | 28,678.01 |
| 472 | 3,869.71 | 3,700.71 | 3,815.86 | 3,897.05 | 15,283.33 |
| 473 | 5,670.50 | 5,292.57 | 5,051.35 | 5,481.28 | 21,495.70 |
| 474 | 3,610.84 | 3,381.68 | 3,471.39 | 3,535.26 | 13,999.17 |
| 475 | 3,248.29 | 3,005.71 | 3,063.33 | 3,186.03 | 12,503.36 |
| 476 | 3,048.66 | 2,668.46 | 2,603.76 | 2,717.22 | 11,038.10 |
| 477 | 5,997.67 | 5,682.26 | 5,884.69 | 5,703.26 | 23,267.88 |
| 478 | 3,488.20 | 3,013.57 | 3,061.26 | 3,150.03 | 12,713.06 |
| 479 | 5,748.93 | 5,421.45 | 5,127.33 | 5,840.36 | 22,138.07 |
| TOTAL | 129,529.42 | 120,821.47 | 118,244.49 | 125,233.43 | 493,828.81 |

STATE: IOWA

| ZIP CODE | QUARTER 1 | QUARTER 2 | QUARTER 3 | QUARTER 4 | TOTAL GRAMS |
|---|---|---|---|---|---|

| | | | | | |
|---|---|---|---|---|---|
| 500 | 6,606.26 | 6,424.22 | 6,471.28 | 6,810.52 | 26,312.28 |
| 501 | 3,876.80 | 3,878.75 | 3,693.67 | 3,906.11 | 15,355.33 |
| 502 | 4,996.31 | 4,675.77 | 4,623.65 | 4,816.03 | 19,111.76 |
| 503 | 8,144.98 | 8,018.81 | 7,807.63 | 8,066.10 | 32,037.52 |
| 504 | 2,097.06 | 2,035.06 | 1,941.48 | 2,181.01 | 8,254.61 |
| 505 | 2,510.33 | 2,358.32 | 2,255.46 | 2,435.07 | 9,559.18 |
| 506 | 3,394.82 | 3,050.74 | 3,076.54 | 3,241.49 | 12,763.59 |
| 507 | 2,197.81 | 2,171.21 | 2,007.27 | 2,190.89 | 8,567.18 |
| 508 | 669.27 | 615.36 | 570.49 | 627.12 | 2,482.24 |
| 510 | 2,229.97 | 2,168.37 | 2,050.77 | 2,041.47 | 8,490.58 |
| 511 | 4,314.92 | 4,309.86 | 4,113.62 | 4,143.89 | 16,882.29 |
| 512 | 1,202.15 | 940.69 | 977.6 | 1,058.25 | 4,178.69 |
| 513 | 1,053.73 | 994.58 | 1,087.45 | 1,058.84 | 4,194.60 |
| 514 | 1,094.21 | 874.8 | 963.19 | 997.73 | 3,929.93 |
| 515 | 3,750.34 | 3,477.15 | 3,367.49 | 3,546.73 | 14,141.71 |
| 516 | 488.58 | 478.43 | 481.26 | 488.53 | 1,936.80 |
| 520 | 4,135.13 | 3,783.85 | 3,700.48 | 3,900.64 | 15,520.10 |
| 521 | 1,136.62 | 1,138.32 | 1,084.69 | 1,175.59 | 4,535.22 |
| 522 | 7,493.72 | 7,141.38 | 6,818.22 | 7,360.05 | 28,813.37 |
| 523 | 4,113.18 | 3,865.41 | 3,723.27 | 3,999.39 | 15,701.25 |
| 524 | 7,097.22 | 6,714.67 | 6,343.34 | 6,710.45 | 26,865.68 |
| 525 | 2,716.89 | 2,378.16 | 2,326.11 | 2,366.20 | 9,787.36 |
| 526 | 3,234.53 | 2,938.76 | 2,960.62 | 3,147.40 | 12,281.31 |
| 527 | 4,917.61 | 4,546.57 | 4,311.49 | 4,860.94 | 18,636.61 |
| 528 | 3,623.11 | 3,443.29 | 3,216.40 | 3,629.90 | 13,912.70 |
| TOTAL | 87,095.55 | 82,422.53 | 79,973.47 | 84,760.34 | 334,251.89 |

STATE: KANSAS

| ZIP CODE | QUARTER 1 | QUARTER 2 | QUARTER 3 | QUARTER 4 | TOTAL GRAMS |
|---|---|---|---|---|---|
| 660 | 9,839.99 | 9,745.41 | 9,746.44 | 10,410.55 | 39,742.39 |
| 661 | 1,564.14 | 1,431.31 | 1,340.80 | 1,422.14 | 5,758.39 |
| 662 | 9,070.11 | 8,509.20 | 8,659.76 | 9,102.68 | 35,341.75 |
| 664 | 1,859.02 | 1,718.31 | 1,371.95 | 1,693.80 | 6,643.08 |
| 665 | 1,775.48 | 1,671.18 | 1,633.32 | 1,809.37 | 6,889.35 |
| 666 | 6,143.38 | 5,875.39 | 6,025.77 | 5,724.44 | 23,768.98 |
| 667 | 1,860.03 | 1,896.44 | 1,754.28 | 1,878.41 | 7,389.16 |
| 668 | 729.65 | 683.41 | 713.18 | 756.53 | 2,882.77 |
| 669 | 320.28 | 296.76 | 313.99 | 294.27 | 1,225.30 |
| 670 | 3,095.27 | 2,850.15 | 2,733.15 | 2,910.07 | 11,588.64 |
| 671 | 1,610.51 | 1,431.09 | 1,495.18 | 1,715.11 | 6,251.89 |
| 672 | 6,931.04 | 6,473.97 | 6,367.52 | 6,919.52 | 26,692.05 |
| 673 | 1,556.40 | 1,438.01 | 1,557.75 | 1,537.44 | 6,089.60 |
| 674 | 2,122.06 | 1,979.44 | 2,001.13 | 2,163.27 | 8,265.90 |
| 675 | 1,904.13 | 1,786.25 | 1,703.74 | 2,009.48 | 7,403.60 |
| 676 | 1,187.35 | 917.7 | 984.81 | 1,070.49 | 4,160.35 |
| 677 | 274.5 | 255.65 | 300.74 | 337.2 | 1,168.09 |

DRUG CODE:1724DRUG NAME:METHYLPHENIDATE (DL;D;L;ISOMERS)
STATE:KANSAS

| ZIP CODE | QUARTER 1 | QUARTER 2 | QUARTER 3 | QUARTER 4 | TOTAL GRAMS |
|---|---|---|---|---|---|
| 678 | 1,005.21 | 921.48 | 844.23 | 939.95 | 3,710.87 |
| 679 | 113.03 | 85.97 | 112.83 | 107.64 | 419.47 |
| TOTAL | 52,961.58 | 49,967.12 | 49,660.57 | 52,802.36 | 205,391.63 |

STATE: KENTUCKY

| ZIP CODE | QUARTER 1 | QUARTER 2 | QUARTER 3 | QUARTER 4 | TOTAL GRAMS |
|---|---|---|---|---|---|
| 400 | 4,667.57 | 4,337.07 | 4,400.54 | 4,360 | 17,765.18 |
| 401 | 2,157.25 | 2,078.14 | 2,721.08 | 1,749.42 | 8,705.89 |
| 402 | 15,214.15 | 13,564.16 | 13,454.94 | 14,451.43 | 56,684.68 |
| 403 | 6,367.92 | 5,834.84 | 6,134.19 | 6,198.26 | 24,535.21 |

| | | | | | |
|---|---|---|---|---|---|
| 404 | 2,376.47 | 2,228.87 | 2,146.26 | 2,350.57 | 9,102.17 |
| 405 | 5,698.39 | 5,386.08 | 5,258.20 | 5,590.80 | 21,933.47 |
| 406 | 1,057.09 | 994.97 | 972.13 | 1,072.60 | 4,096.79 |
| 407 | 1,611.29 | 1,612.44 | 1,605.48 | 1,663.74 | 6,492.95 |
| 408 | 451.35 | 413.84 | 370.31 | 488.98 | 1,724.48 |
| 409 | 702.8 | 635.34 | 598.73 | 638.64 | 2,575.51 |
| 410 | 10,785.36 | 10,027.99 | 9,885.66 | 10,528.11 | 41,227.12 |
| 411 | 2,342.12 | 2,360.02 | 2,176.21 | 2,137.42 | 9,015.77 |
| 412 | 832.26 | 686.12 | 776.58 | 731.59 | 3,026.55 |
| 413 | 432.68 | 350.48 | 400.62 | 369.74 | 1,553.52 |
| 414 | 277.35 | 267.88 | 271.3 | 290.89 | 1,107.42 |
| 415 | 1,151.19 | 1,009.62 | 1,034.75 | 1,065.34 | 4,260.90 |
| 416 | 636.51 | 543.11 | 588.37 | 626.2 | 2,394.19 |
| 417 | 856 | 947.27 | 911.93 | 1,005.87 | 3,721.07 |
| 418 | 274.78 | 250.49 | 314.02 | 293.08 | 1,132.37 |
| 420 | 5,081.27 | 4,795.49 | 4,808.16 | 4,774.04 | 19,458.96 |
| 421 | 3,256.82 | 2,934.18 | 2,826.06 | 2,982.77 | 11,999.83 |
| 422 | 3,103.73 | 2,354.20 | 2,909.73 | 2,490.12 | 10,857.78 |
| 423 | 2,805.66 | 2,767.71 | 2,775.08 | 2,812.61 | 11,161.06 |
| 424 | 3,691.37 | 3,325.92 | 3,525.26 | 3,637.56 | 14,180.11 |
| 425 | 1,084.52 | 980.48 | 836.29 | 1,028.42 | 3,929.71 |
| 426 | 695.42 | 665.87 | 656 | 666.95 | 2,684.24 |
| 427 | 2,915.54 | 2,687.09 | 2,634.41 | 2,684.01 | 10,921.05 |
| TOTAL | 80,526.86 | 74,039.67 | 74,992.29 | 76,689.16 | 306,247.98 |

STATE: LOUISIANA

| ZIP CODE | QUARTER 1 | QUARTER 2 | QUARTER 3 | QUARTER 4 | TOTAL GRAMS |
|---|---|---|---|---|---|
| 700 | 10,603.31 | 9,044.39 | 9,629.29 | 10,057.26 | 39,334.25 |
| 701 | 4,703.59 | 4,213.95 | 4,317.21 | 4,554.95 | 17,789.70 |
| 703 | 5,065.11 | 4,465.28 | 4,677.23 | 4,980.92 | 19,188.54 |
| 704 | 10,751.54 | 9,381.23 | 9,757.95 | 10,270.20 | 40,160.92 |
| 705 | 14,623.38 | 12,095.23 | 12,451.25 | 13,880.38 | 53,050.24 |
| 706 | 9,101.13 | 7,798.16 | 7,945.34 | 8,495.36 | 33,339.99 |
| 707 | 10,937.23 | 9,540.53 | 9,479.71 | 10,347.06 | 40,304.53 |
| 708 | 8,264.19 | 7,283.90 | 6,898.76 | 7,970.23 | 30,417.08 |
| 710 | 2,075.22 | 1,722.90 | 1,705.03 | 1,920.93 | 7,424.08 |
| 711 | 5,258.70 | 4,902.93 | 4,887.04 | 5,157.45 | 20,206.12 |
| 712 | 5,210.58 | 4,367.07 | 4,722.66 | 5,008.32 | 19,308.63 |
| 713 | 3,941.37 | 3,305.10 | 3,451.65 | 3,882.97 | 14,581.09 |
| 714 | 2,843.26 | 2,324.96 | 2,907.91 | 2,687.65 | 10,763.78 |
| TOTAL | 93,378.61 | 80,445.63 | 82,831.03 | 89,213.68 | 345,868.95 |

STATE: MAINE

| ZIP CODE | QUARTER 1 | QUARTER 2 | QUARTER 3 | QUARTER 4 | TOTAL GRAMS |
|---|---|---|---|---|---|
| 39 | 820.78 | 886.81 | 716.42 | 685.71 | 3,109.72 |
| 40 | 8,245.51 | 8,342.50 | 7,725.83 | 7,916.34 | 32,230.18 |
| 41 | 2,951.36 | 3,016.05 | 2,815.90 | 2,924.40 | 11,707.71 |
| 42 | 3,162.92 | 3,200.30 | 3,093.91 | 3,173.57 | 12,630.70 |
| 43 | 2,661.71 | 2,677.71 | 2,708.10 | 2,620.25 | 10,667.77 |
| 44 | 7,363.97 | 7,307.51 | 6,905.04 | 6,617.26 | 28,193.78 |
| 45 | 1,263.54 | 1,219.46 | 1,292.89 | 1,232.58 | 5,008.47 |
| 46 | 2,070.60 | 1,957.54 | 2,129.32 | 2,032.41 | 8,189.87 |
| 47 | 2,031.20 | 2,197.01 | 1,869.12 | 1,991.17 | 8,088.50 |
| 48 | 1,353.27 | 1,207.91 | 1,248.44 | 1,220.17 | 5,029.79 |
| 49 | 5,264 | 5,143.78 | 4,855.87 | 4,907.26 | 20,170.91 |
| TOTAL | 37,188.86 | 37,156.58 | 35,360.84 | 35,321.12 | 145,027.40 |

STATE: MARYLAND

| ZIP CODE | QUARTER 1 | QUARTER 2 | QUARTER 3 | QUARTER 4 | TOTAL GRAMS |
|---|---|---|---|---|---|
| 206 | 4,079.27 | 3,813.52 | 3,688.73 | 4,151.63 | 15,733.15 |

RETAIL DRUG DISTRIBUTION BY ZIP CODE WITHIN STATE BY GRAMS WT

DRUG CODE:1724DRUG NAME:METHYLPHENIDATE (DL;D;L;ISOMERS)
STATE:MARYLAND

| ZIP CODE | QUARTER 1 | QUARTER 2 | QUARTER 3 | QUARTER 4 | TOTAL GRAMS |
|---|---|---|---|---|---|
| 207 | 7,402.10 | 7,275.59 | 7,030.88 | 6,788.02 | 28,496.59 |
| 208 | 16,547.55 | 15,402.75 | 15,372.76 | 15,463.39 | 62,786.45 |
| 209 | 2,767.66 | 2,559.53 | 2,387.57 | 2,710.97 | 10,425.73 |
| 210 | 17,473.74 | 17,380.04 | 16,627.76 | 17,402.19 | 68,883.73 |
| 211 | 9,128.55 | 8,760.12 | 8,246.47 | 9,208.80 | 35,343.94 |
| 212 | 20,091.92 | 19,880.73 | 19,028.74 | 20,283.81 | 79,285.20 |
| 214 | 2,020.54 | 2,007.68 | 1,902.69 | 1,978.85 | 7,909.76 |
| 215 | 3,438.69 | 3,446.26 | 3,530.26 | 3,462.38 | 13,877.59 |
| 216 | 4,094.97 | 4,111.62 | 3,895.04 | 4,139.78 | 16,241.41 |
| 217 | 12,082.51 | 11,758.59 | 11,565.69 | 11,721.69 | 47,128.48 |
| 218 | 3,686.57 | 3,410.59 | 3,319.66 | 3,445.14 | 13,861.96 |
| 219 | 1,822.19 | 1,761.83 | 1,679.20 | 1,768.42 | 7,031.64 |
| TOTAL | 104,636.26 | 101,568.85 | 98,275.45 | 102,525.07 | 407,005.63 |

STATE: MASSACHUSETTS

| ZIP CODE | QUARTER 1 | QUARTER 2 | QUARTER 3 | QUARTER 4 | TOTAL GRAMS |
|---|---|---|---|---|---|
| 10 | 11,674.92 | 10,885.04 | 10,064.59 | 10,924.26 | 43,548.81 |
| 11 | 4,770.82 | 4,838.67 | 4,279.69 | 4,552.38 | 18,441.56 |
| 12 | 4,688.91 | 4,493.74 | 4,387.36 | 4,501.55 | 18,071.56 |
| 13 | 2,194.34 | 2,030.23 | 1,903.02 | 1,995.07 | 8,122.66 |
| 14 | 6,701.07 | 6,534.30 | 5,974.10 | 6,071.18 | 25,280.65 |
| 15 | 8,740.55 | 8,610.71 | 7,791.10 | 8,430.03 | 33,572.39 |
| 16 | 4,657.30 | 4,618.08 | 4,100.07 | 4,469.22 | 17,844.67 |
| 17 | 10,873.44 | 10,731.81 | 9,861.57 | 10,885.63 | 42,352.45 |
| 18 | 16,574.61 | 15,952.54 | 14,669.21 | 16,229.01 | 63,425.37 |
| 19 | 12,582.67 | 12,067.30 | 11,234.26 | 12,240.19 | 48,124.42 |
| 20 | 10,267.77 | 10,082.41 | 9,209.38 | 9,985.24 | 39,544.80 |
| 21 | 21,111.34 | 21,382.75 | 19,280.21 | 21,418.96 | 83,193.26 |
| 22 | 984.88 | 822.49 | 897.89 | 1,025.29 | 3,730.55 |
| 23 | 9,744.05 | 9,381.24 | 8,655.80 | 9,435.17 | 37,216.26 |
| 24 | 12,251.39 | 11,846.32 | 11,011.07 | 12,061.22 | 47,170 |
| 25 | 2,970.51 | 3,072.03 | 3,059.23 | 3,037.56 | 12,139.33 |
| 26 | 3,918.07 | 4,086.61 | 4,129.17 | 3,842.25 | 15,976.10 |
| 27 | 10,060.91 | 9,714.44 | 8,674.78 | 9,821.92 | 38,272.05 |
| TOTAL | 154,767.55 | 151,150.71 | 139,182.50 | 150,926.13 | 596,026.89 |

STATE: MICHIGAN

| ZIP CODE | QUARTER 1 | QUARTER 2 | QUARTER 3 | QUARTER 4 | TOTAL GRAMS |
|---|---|---|---|---|---|
| 480 | 25,796.50 | 25,126.25 | 21,931.56 | 25,437.61 | 98,291.92 |
| 481 | 28,442.94 | 27,381.56 | 23,876.48 | 28,572.49 | 108,273.47 |
| 482 | 5,927.05 | 5,752.26 | 5,121.50 | 5,660.82 | 22,461.63 |
| 483 | 21,013.19 | 19,951.32 | 17,354.84 | 20,145.31 | 78,464.66 |
| 484 | 7,072.97 | 6,625.81 | 5,747.47 | 6,803.74 | 26,249.99 |
| 485 | 2,624.16 | 2,371.59 | 2,042.77 | 2,591.92 | 9,630.44 |
| 486 | 7,577.61 | 7,435.06 | 6,458.12 | 7,703.83 | 29,174.62 |
| 487 | 4,728.51 | 4,671.65 | 3,990.39 | 4,563.56 | 17,954.11 |
| 488 | 13,323.02 | 12,785.44 | 10,106.91 | 13,151.96 | 49,367.33 |
| 489 | 3,193.53 | 3,108.17 | 2,662.16 | 3,264.81 | 12,228.67 |
| 490 | 14,163.67 | 13,634.24 | 12,060.37 | 13,848.52 | 53,706.80 |
| 491 | 1,168.80 | 1,076.80 | 1,052.50 | 1,082.79 | 4,380.89 |
| 492 | 6,274.42 | 5,821.36 | 4,741.93 | 6,042.77 | 22,880.48 |
| 493 | 6,403.26 | 5,872.45 | 4,631 | 6,605.15 | 23,511.86 |
| 494 | 18,166.17 | 17,057.06 | 14,801.99 | 18,025.70 | 68,050.92 |
| 495 | 13,851.50 | 13,222.70 | 11,258.19 | 13,588.02 | 51,920.41 |
| 496 | 7,498.47 | 7,442.70 | 6,864.01 | 7,478.23 | 29,283.41 |
| 497 | 5,795.20 | 5,490.94 | 5,035.37 | 5,438.92 | 21,760.43 |
| 498 | 6,896.55 | 7,222.32 | 7,428.42 | 7,121.01 | 28,668.30 |

| | | | | | |
|---|---|---|---|---|---|
| 499 | 1,715.75 | 1,500.70 | 1,439.25 | 1,593.83 | 6,249.53 |
| TOTAL | 201,633.27 | 193,550.38 | 168,605.23 | 198,720.99 | 762,509.87 |

STATE: MINNESOTA

| ZIP CODE | QUARTER 1 | QUARTER 2 | QUARTER 3 | QUARTER 4 | TOTAL GRAMS |
|---|---|---|---|---|---|
| 550 | 11,198.85 | 10,982.72 | 10,293.61 | 11,665.95 | 44,141.13 |
| 551 | 15,267.09 | 14,998.06 | 14,127.44 | 15,259.59 | 59,652.18 |
| 553 | 20,773.13 | 19,892.42 | 18,774.79 | 20,975.81 | 80,416.15 |
| 554 | 20,246.37 | 19,789.51 | 19,546.88 | 20,811.83 | 80,394.59 |
| 556 | 214.63 | 197.01 | 166.31 | 167.48 | 745.43 |
| 557 | 2,434.63 | 2,158.50 | 2,050.73 | 2,179.77 | 8,823.63 |
| 558 | 2,565.68 | 2,575.44 | 2,502.08 | 2,660.88 | 10,304.08 |

DRUG CODE:1724DRUG NAME:METHYLPHENIDATE (DL;D;L;ISOMERS)
STATE:MINNESOTA

| ZIP CODE | QUARTER 1 | QUARTER 2 | QUARTER 3 | QUARTER 4 | TOTAL GRAMS |
|---|---|---|---|---|---|
| 559 | 8,409.18 | 7,832.44 | 7,585.78 | 8,370.65 | 32,198.05 |
| 560 | 4,740.06 | 4,590.39 | 4,375.85 | 4,729.14 | 18,435.44 |
| 561 | 1,453.46 | 1,330.47 | 1,336.80 | 1,421.32 | 5,542.05 |
| 562 | 2,363.94 | 2,265.66 | 2,116.98 | 2,246.91 | 8,993.49 |
| 563 | 6,228.76 | 6,028.14 | 5,718.48 | 5,930.61 | 23,905.99 |
| 564 | 2,149.76 | 2,025.25 | 2,002.68 | 2,002.70 | 8,180.39 |
| 565 | 3,867.62 | 3,683.25 | 3,485.92 | 3,517.01 | 14,553.80 |
| 566 | 1,061.34 | 1,005.91 | 990.78 | 1,057.94 | 4,115.97 |
| 567 | 1,044.59 | 1,054.77 | 992.29 | 973.84 | 4,065.49 |
| TOTAL | 104,019.09 | 100,409.94 | 96,067.40 | 103,971.43 | 404,467.86 |

STATE: MISSISSIPPI

| ZIP CODE | QUARTER 1 | QUARTER 2 | QUARTER 3 | QUARTER 4 | TOTAL GRAMS |
|---|---|---|---|---|---|
| 386 | 6,441.78 | 5,597.50 | 5,976.53 | 5,994.51 | 24,010.32 |
| 387 | 648.86 | 593.32 | 595.03 | 627.7 | 2,464.91 |
| 388 | 3,445.88 | 2,851.35 | 2,949.29 | 3,162.55 | 12,409.07 |
| 389 | 1,256.01 | 1,164.60 | 1,090.25 | 1,191.66 | 4,702.52 |
| 390 | 3,835.22 | 3,345.45 | 3,756.23 | 3,805.28 | 14,742.18 |
| 391 | 4,433.23 | 3,823.20 | 3,912.49 | 3,970.61 | 16,139.53 |
| 392 | 3,770.51 | 3,290.59 | 3,314.07 | 3,641.57 | 14,016.74 |
| 393 | 2,798.90 | 2,463.68 | 2,688.41 | 2,773.35 | 10,724.34 |
| 394 | 7,477.75 | 6,430.34 | 6,703.80 | 7,025.03 | 27,636.92 |
| 395 | 9,888.24 | 8,607.18 | 8,915.64 | 8,862.17 | 36,273.23 |
| 396 | 2,042.04 | 1,766.03 | 1,808.84 | 1,870.40 | 7,487.31 |
| 397 | 1,638.32 | 1,422.55 | 1,512.44 | 1,521.91 | 6,095.22 |
| TOTAL | 47,676.74 | 41,355.79 | 43,223.02 | 44,446.74 | 176,702.29 |

STATE: MISSOURI

| ZIP CODE | QUARTER 1 | QUARTER 2 | QUARTER 3 | QUARTER 4 | TOTAL GRAMS |
|---|---|---|---|---|---|
| 630 | 17,881.69 | 16,235.61 | 16,118.32 | 17,572.99 | 67,808.61 |
| 631 | 44,012.26 | 44,759.10 | 43,996.11 | 46,547.83 | 179,315.30 |
| 633 | 11,467 | 10,754.77 | 10,469.59 | 11,553.57 | 44,244.93 |
| 634 | 1,384.74 | 1,275.86 | 1,265.92 | 1,407.61 | 5,334.13 |
| 635 | 790.76 | 679.97 | 673.06 | 725.59 | 2,869.38 |
| 636 | 2,346.56 | 2,287.80 | 2,216.74 | 2,259.37 | 9,110.47 |
| 637 | 2,127.96 | 1,962.28 | 1,832.77 | 2,072.57 | 7,995.58 |
| 638 | 2,360.42 | 2,065.94 | 2,053.32 | 2,422.64 | 8,902.32 |
| 639 | 928.6 | 842.33 | 881.55 | 871.99 | 3,524.47 |
| 640 | 9,865.23 | 9,159.73 | 9,051.73 | 9,845.30 | 37,921.99 |
| 641 | 12,363.74 | 11,671.34 | 11,529.55 | 12,424.10 | 47,988.73 |
| 644 | 1,132.89 | 1,072.41 | 899.86 | 1,056.13 | 4,161.29 |
| 645 | 2,093.35 | 1,910.10 | 1,791.35 | 2,018.35 | 7,813.15 |
| 646 | 564.69 | 502.03 | 503.55 | 570.34 | 2,140.61 |
| 647 | 1,008.13 | 940.77 | 954.72 | 948.5 | 3,852.12 |

| | | | | | |
|---|---|---|---|---|---|
| 648 | 3,259.87 | 3,025.79 | 2,958.27 | 3,268.57 | 12,512.50 |
| 650 | 1,367.55 | 1,330.66 | 1,235.42 | 1,415.30 | 5,348.93 |
| 651 | 1,778.35 | 1,587.28 | 1,557.34 | 1,688.01 | 6,610.98 |
| 652 | 3,956.21 | 3,685.61 | 3,712.07 | 4,011.18 | 15,365.07 |
| 653 | 1,230.69 | 1,180.18 | 1,121.34 | 1,265.62 | 4,797.83 |
| 654 | 1,518.43 | 1,473.98 | 1,465.71 | 1,589.69 | 6,047.81 |
| 655 | 1,544.73 | 1,500.33 | 1,381.42 | 1,512.30 | 5,938.78 |
| 656 | 1,719.64 | 1,626.79 | 1,550.47 | 1,699.03 | 6,595.93 |
| 657 | 3,710.01 | 3,576.43 | 3,524.59 | 3,619.57 | 14,430.60 |
| 658 | 4,545.15 | 4,304.77 | 4,056.95 | 4,322.63 | 17,229.50 |
| TOTAL | 134,958.65 | 129,411.86 | 126,801.72 | 136,688.78 | 527,861.01 |

STATE: MONTANA

| ZIP CODE | QUARTER 1 | QUARTER 2 | QUARTER 3 | QUARTER 4 | TOTAL GRAMS |
|---|---|---|---|---|---|
| 590 | 1,045.45 | 920.13 | 885.49 | 1,011.24 | 3,862.31 |
| 591 | 4,571.37 | 4,368.65 | 3,979.84 | 4,373.64 | 17,293.50 |
| 592 | 597.66 | 585.64 | 478.89 | 537.81 | 2,200 |
| 593 | 715.97 | 699.28 | 704.78 | 772.42 | 2,892.45 |
| 594 | 2,757.09 | 2,465.91 | 2,338.86 | 2,595.55 | 10,157.41 |
| 595 | 349.96 | 333.64 | 276.08 | 355.82 | 1,315.50 |
| 596 | 1,274.74 | 1,242.18 | 1,109.26 | 1,206.20 | 4,832.38 |
| 597 | 2,611.83 | 2,367.41 | 2,315.31 | 2,572.64 | 9,867.19 |
| 598 | 3,333.25 | 3,310.31 | 3,141.70 | 3,354.39 | 13,139.65 |
| 599 | 1,290.24 | 1,262.94 | 1,246.16 | 1,332.31 | 5,131.65 |
| TOTAL | 18,547.56 | 17,556.09 | 16,476.37 | 18,112.02 | 70,692.04 |

DRUG CODE:1724DRUG NAME:METHYLPHENIDATE (DL;D;L;ISOMERS)
STATE: NEBRASKA

| ZIP CODE | QUARTER 1 | QUARTER 2 | QUARTER 3 | QUARTER 4 | TOTAL GRAMS |
|---|---|---|---|---|---|
| 680 | 3,349.16 | 3,148.02 | 3,135.25 | 3,326.84 | 12,959.27 |
| 681 | 12,744.42 | 12,378.87 | 12,211.98 | 12,758.41 | 50,093.68 |
| 683 | 992.47 | 997.76 | 913.19 | 968.31 | 3,871.73 |
| 684 | 767.94 | 743.02 | 733.92 | 804.8 | 3,049.68 |
| 685 | 6,048.64 | 5,616.12 | 5,570.92 | 6,001.76 | 23,237.44 |
| 686 | 762.52 | 635.08 | 700.38 | 677.4 | 2,775.38 |
| 687 | 1,659.84 | 1,508 | 1,471.24 | 1,528.84 | 6,167.92 |
| 688 | 2,601.12 | 2,520.47 | 2,383.12 | 2,536.40 | 10,041.11 |
| 689 | 1,315.81 | 1,098.79 | 1,093.04 | 1,228.21 | 4,735.85 |
| 690 | 347.95 | 295.25 | 251.5 | 273.37 | 1,168.07 |
| 691 | 1,318.80 | 1,334.50 | 1,279.45 | 1,306.60 | 5,239.35 |
| 692 | 58.8 | 37.92 | 47.86 | 64.12 | 208.7 |
| 693 | 1,048.54 | 1,040.19 | 993.32 | 986.52 | 4,068.57 |
| TOTAL | 33,016.01 | 31,353.99 | 30,785.17 | 32,461.58 | 127,616.75 |

STATE: NEVADA

| ZIP CODE | QUARTER 1 | QUARTER 2 | QUARTER 3 | QUARTER 4 | TOTAL GRAMS |
|---|---|---|---|---|---|
| 890 | 3,350.01 | 3,046.37 | 2,845.53 | 3,157.70 | 12,399.61 |
| 891 | 7,058.56 | 6,622.52 | 6,063.36 | 6,484.31 | 26,228.75 |
| 893 | 32.34 | 40.04 | 32.08 | 22.05 | 126.51 |
| 894 | 1,708.72 | 1,594.80 | 1,606.66 | 1,716.13 | 6,626.31 |
| 895 | 2,240.69 | 2,187.54 | 2,201.77 | 2,087.02 | 8,717.02 |
| 897 | 489.15 | 574.45 | 501.96 | 522.06 | 2,087.62 |
| 898 | 455.9 | 398.5 | 414.66 | 461.12 | 1,730.18 |
| TOTAL | 15,335.37 | 14,464.22 | 13,666.02 | 14,450.39 | 57,916 |

STATE: NEW HAMPSHIRE

| ZIP CODE | QUARTER 1 | QUARTER 2 | QUARTER 3 | QUARTER 4 | TOTAL GRAMS |
|---|---|---|---|---|---|
| 30 | 8,792.91 | 8,555.47 | 7,929.53 | 8,694.17 | 33,972.08 |

| | | | | | |
|---|---|---|---|---|---|
| 31 | 5,408.36 | 5,229.48 | 4,953.88 | 5,305.40 | 20,897.12 |
| 32 | 3,115.80 | 3,160.11 | 3,141.86 | 3,260.07 | 12,677.84 |
| 33 | 2,304.57 | 2,402.01 | 2,195.32 | 2,247.01 | 9,148.91 |
| 34 | 2,158.28 | 2,214.51 | 1,985.22 | 2,095.61 | 8,453.62 |
| 35 | 1,747.06 | 1,738.29 | 1,761.31 | 1,921.72 | 7,168.38 |
| 36 | 114.53 | 86.04 | 105.67 | 112.32 | 418.56 |
| 37 | 3,524.90 | 3,341.65 | 3,237.82 | 3,321.90 | 13,426.27 |
| 38 | 7,822.78 | 7,911 | 7,488.10 | 7,944.91 | 31,166.79 |
| TOTAL | 34,989.19 | 34,638.56 | 32,798.71 | 34,903.11 | 137,329.57 |

STATE: NEW JERSEY

| ZIP CODE | QUARTER 1 | QUARTER 2 | QUARTER 3 | QUARTER 4 | TOTAL GRAMS |
|---|---|---|---|---|---|
| 70 | 16,394.07 | 16,345.45 | 14,581.55 | 16,678.11 | 63,999.18 |
| 71 | 1,971.45 | 1,861.50 | 1,568.57 | 1,783.60 | 7,185.12 |
| 72 | 1,143.55 | 1,112.48 | 842.63 | 1,006.11 | 4,104.77 |
| 73 | 1,000.69 | 1,193.64 | 928.12 | 1,120.72 | 4,243.17 |
| 74 | 5,421.12 | 5,047.28 | 4,755.84 | 5,255.07 | 20,479.31 |
| 75 | 999.86 | 1,016.94 | 786.04 | 1,019.72 | 3,822.56 |
| 76 | 4,938.75 | 4,728.50 | 4,276.50 | 4,815.19 | 18,758.94 |
| 77 | 6,659.06 | 6,300.63 | 5,802.95 | 6,393.77 | 25,156.41 |
| 78 | 4,968.96 | 5,022.87 | 4,339.53 | 4,932.02 | 19,263.38 |
| 79 | 4,919.30 | 4,912.83 | 4,237.18 | 4,756.37 | 18,825.68 |
| 80 | 18,222.50 | 18,155.14 | 15,952.14 | 17,439.31 | 69,769.09 |
| 81 | 1,512.95 | 1,534.78 | 1,310.91 | 1,562.65 | 5,921.29 |
| 82 | 2,814.32 | 2,911.10 | 2,737.56 | 2,769.87 | 11,232.85 |
| 83 | 2,500.87 | 2,635.18 | 2,137.16 | 2,435.55 | 9,708.76 |
| 84 | 219.81 | 221.8 | 251.05 | 203.97 | 896.63 |
| 85 | 4,126.95 | 4,357.28 | 3,731.60 | 4,132.61 | 16,348.44 |
| 86 | 3,032.56 | 3,077.65 | 2,693.05 | 2,961.47 | 11,764.73 |
| 87 | 5,059.12 | 4,914.02 | 4,744.66 | 4,773.28 | 19,491.08 |
| 88 | 9,054.64 | 8,934.13 | 8,133.88 | 8,854.54 | 34,977.19 |
| 89 | 849.03 | 831.98 | 770.36 | 901.34 | 3,352.71 |
| TOTAL | 95,809.56 | 95,115.18 | 84,581.28 | 93,795.27 | 369,301.29 |

STATE: NEW MEXICO

| ZIP CODE | QUARTER 1 | QUARTER 2 | QUARTER 3 | QUARTER 4 | TOTAL GRAMS |
|---|---|---|---|---|---|
| 870 | 1,551.77 | 1,532.81 | 1,428.88 | 1,528.62 | 6,042.08 |
| 871 | 13,835.17 | 13,271.42 | 12,975.12 | 12,314.27 | 52,395.98 |
| 873 | 660.83 | 506.61 | 541.17 | 607.48 | 2,316.09 |
| 874 | 1,673.44 | 1,578.54 | 1,498.45 | 1,626.92 | 6,377.35 |
| 875 | 2,017.47 | 1,830.07 | 1,774.95 | 1,819.27 | 7,441.76 |

DRUG CODE:1724DRUG NAME:METHYLPHENIDATE (DL;D;L;ISOMERS)
STATE:NEW MEXICO

| ZIP CODE | QUARTER 1 | QUARTER 2 | QUARTER 3 | QUARTER 4 | TOTAL GRAMS |
|---|---|---|---|---|---|
| 877 | 269.99 | 253.05 | 278.43 | 334.22 | 1,135.69 |
| 878 | 162.29 | 63.62 | 66.34 | 139.04 | 431.29 |
| 879 | 109.94 | 101.01 | 72.25 | 99.15 | 382.35 |
| 880 | 1,822.84 | 1,684.50 | 1,658.03 | 1,788.55 | 6,953.92 |
| 881 | 596.89 | 476.78 | 439.65 | 501.69 | 2,015.01 |
| 882 | 1,998.80 | 1,605.76 | 1,599.55 | 1,806.66 | 7,010.77 |
| 883 | 658.89 | 533.26 | 589.88 | 572.25 | 2,354.28 |
| 884 | 119.16 | 135.5 | 88.24 | 109.43 | 452.33 |
| TOTAL | 25,477.48 | 23,572.93 | 23,010.94 | 23,247.55 | 95,308.90 |

STATE: NEW YORK

| ZIP CODE | QUARTER 1 | QUARTER 2 | QUARTER 3 | QUARTER 4 | TOTAL GRAMS |
|---|---|---|---|---|---|
| 100 | 21,946.29 | 22,905.12 | 19,832.81 | 22,055.70 | 86,739.92 |
| 101 | 1,299.79 | 1,419.68 | 1,203.38 | 1,334.29 | 5,257.14 |
| 102 | 125.12 | 139.09 | 105.93 | 127.54 | 497.68 |

| | | | | | |
|---|---|---|---|---|---|
| 103 | 2,771.74 | 2,938.65 | 2,438.30 | 2,728.93 | 10,877.62 |
| 104 | 8,560.15 | 8,821.98 | 7,208.30 | 8,185.16 | 32,775.59 |
| 105 | 10,654.79 | 10,722.25 | 9,480.45 | 10,585.45 | 41,442.94 |
| 106 | 1,124.10 | 1,059.38 | 1,026.63 | 1,208.26 | 4,418.37 |
| 107 | 1,993.32 | 2,011.77 | 1,745.19 | 1,927.37 | 7,677.65 |
| 108 | 1,044.03 | 1,094.55 | 893.25 | 1,046.37 | 4,078.20 |
| 109 | 7,170.16 | 7,159.89 | 6,520.51 | 7,008.94 | 27,859.50 |
| 110 | 3,120.24 | 2,884.23 | 2,596.72 | 2,935.56 | 11,536.75 |
| 111 | 1,197.11 | 1,254.85 | 1,155.17 | 1,219.90 | 4,827.03 |
| 112 | 12,141.90 | 12,856.22 | 10,685.78 | 11,994.43 | 47,678.33 |
| 113 | 5,200.99 | 5,414.26 | 4,168.83 | 5,023.92 | 19,808 |
| 114 | 2,669.41 | 2,674.02 | 2,189.02 | 2,467.29 | 9,999.74 |
| 115 | 8,814.18 | 8,543.93 | 7,836.07 | 8,681.28 | 33,875.46 |
| 116 | 861.04 | 961.48 | 746.6 | 857.94 | 3,427.06 |
| 117 | 21,574.57 | 21,400.69 | 18,972.82 | 20,440.77 | 82,388.85 |
| 118 | 1,427.71 | 1,357.49 | 1,193.16 | 1,412.58 | 5,390.94 |
| 119 | 2,719.23 | 2,968.95 | 2,791.49 | 2,752.96 | 11,232.63 |
| 120 | 7,767.65 | 7,711.35 | 7,169.23 | 7,362.72 | 30,010.95 |
| 121 | 4,985.16 | 4,860.72 | 4,518.09 | 5,004.28 | 19,368.25 |
| 122 | 2,951.92 | 2,975.77 | 2,596.96 | 2,852.40 | 11,377.05 |
| 123 | 3,687.22 | 3,425.46 | 3,202.06 | 3,407.35 | 13,722.09 |
| 124 | 1,843.61 | 1,948.35 | 1,800.21 | 1,793.46 | 7,385.63 |
| 125 | 5,632.23 | 5,631.49 | 5,150.97 | 5,455.08 | 21,869.77 |
| 126 | 1,219.53 | 1,179.76 | 1,063.01 | 1,111.89 | 4,574.19 |
| 127 | 1,033.77 | 1,216.68 | 993.46 | 1,047.68 | 4,291.59 |
| 128 | 4,906.45 | 5,022.90 | 4,490.86 | 4,877.37 | 19,297.58 |
| 129 | 3,987.41 | 4,045.70 | 3,822.93 | 3,990.83 | 15,846.87 |
| 130 | 6,370.89 | 6,503.97 | 5,772.02 | 6,463.23 | 25,110.11 |
| 131 | 2,306.42 | 2,391.95 | 2,118.27 | 2,260.87 | 9,077.51 |
| 132 | 3,673.82 | 3,661.62 | 3,439.72 | 3,480.37 | 14,255.53 |
| 133 | 2,059.94 | 2,100.59 | 1,895.97 | 2,061.29 | 8,117.79 |
| 134 | 2,921.67 | 2,823.88 | 2,587.92 | 2,817.96 | 11,151.43 |
| 135 | 1,080.89 | 1,172.62 | 1,010.75 | 1,028.42 | 4,292.68 |
| 136 | 5,322.42 | 6,029.51 | 5,093.52 | 5,588.39 | 22,033.84 |
| 137 | 1,631.74 | 1,498.28 | 1,452.91 | 1,410.93 | 5,993.86 |
| 138 | 2,772.90 | 2,690.27 | 2,444.51 | 2,691.45 | 10,599.13 |
| 139 | 1,329.49 | 1,277.78 | 1,199.32 | 1,209.20 | 5,015.79 |
| 140 | 5,940.07 | 5,851.58 | 5,360.56 | 5,846.94 | 22,999.15 |
| 141 | 2,900.61 | 2,975.09 | 2,557.28 | 2,848.98 | 11,281.96 |
| 142 | 7,698.05 | 7,507.45 | 6,925.80 | 7,753.79 | 29,885.09 |
| 143 | 751.84 | 658.09 | 599.13 | 650.97 | 2,660.03 |
| 144 | 7,940.91 | 7,858.57 | 7,293.14 | 7,874.94 | 30,967.56 |
| 145 | 8,505.07 | 8,243.69 | 7,773.84 | 8,254.38 | 32,776.98 |
| 146 | 13,317.36 | 12,858.26 | 12,108.89 | 12,884.96 | 51,169.47 |
| 147 | 2,380.93 | 2,247.26 | 2,104.48 | 2,275.09 | 9,007.76 |
| 148 | 5,500.68 | 5,449.74 | 5,044.49 | 5,564.43 | 21,559.34 |
| 149 | 1,325.70 | 1,238.71 | 1,157.11 | 1,257.46 | 4,978.98 |
| TOTAL | 240,162.22 | 241,645.57 | 215,537.82 | 235,121.75 | 932,467.36 |

STATE: NORTH CAROLINA

| ZIP CODE | QUARTER 1 | QUARTER 2 | QUARTER 3 | QUARTER 4 | TOTAL GRAMS |
|---|---|---|---|---|---|
| 270 | 4,955.83 | 4,822.61 | 4,701.01 | 5,221.43 | 19,700.88 |
| 271 | 5,732.25 | 5,501.78 | 5,305.67 | 5,856.47 | 22,396.17 |

DRUG CODE:1724DRUG NAME:METHYLPHENIDATE (DL;D;L;ISOMERS)
STATE:NORTH CAROLINA

| ZIP CODE | QUARTER 1 | QUARTER 2 | QUARTER 3 | QUARTER 4 | TOTAL GRAMS |
|---|---|---|---|---|---|
| 272 | 12,281.14 | 11,578.26 | 10,783.64 | 11,578.70 | 46,221.74 |
| 273 | 7,350.22 | 7,035.99 | 6,395.12 | 7,305.92 | 28,087.25 |
| 274 | 6,012.62 | 5,807.63 | 5,521.28 | 6,092.59 | 23,434.12 |

| | | | | | |
|---|---|---|---|---|---|
| 275 | 25,122.07 | 25,238.75 | 23,199.17 | 25,582.58 | 99,142.57 |
| 276 | 9,306.59 | 9,349.46 | 8,447.71 | 9,538.91 | 36,642.67 |
| 277 | 6,110.27 | 5,815.48 | 5,737.55 | 6,077.38 | 23,740.68 |
| 278 | 9,846.82 | 9,281.44 | 8,593.39 | 9,657.62 | 37,379.27 |
| 279 | 2,508.70 | 2,500.53 | 2,403.25 | 2,528.66 | 9,941.14 |
| 280 | 18,834.85 | 18,290.47 | 16,923.08 | 18,549.74 | 72,598.14 |
| 281 | 14,517.52 | 13,958.71 | 12,795.88 | 14,316.62 | 55,588.73 |
| 282 | 13,390.54 | 12,927.97 | 12,343.95 | 13,953.39 | 52,615.85 |
| 283 | 17,123.79 | 16,374.42 | 15,646.64 | 16,598.74 | 65,743.59 |
| 284 | 9,317.51 | 8,930.68 | 8,459.79 | 9,114.48 | 35,822.46 |
| 285 | 12,506.39 | 12,724.76 | 12,064.44 | 11,891.64 | 49,187.23 |
| 286 | 11,937.81 | 11,541.83 | 10,693.11 | 12,122.66 | 46,295.41 |
| 287 | 8,636.18 | 8,569.78 | 8,167.08 | 8,878.75 | 34,251.79 |
| 288 | 3,945.75 | 3,920.41 | 3,750.71 | 4,022.73 | 15,639.60 |
| 289 | 727.47 | 881.55 | 743.05 | 757.24 | 3,109.31 |
| TOTAL | 200,164.32 | 195,052.51 | 182,675.52 | 199,646.25 | 777,538.60 |

STATE: NORTH DAKOTA

| ZIP CODE | QUARTER 1 | QUARTER 2 | QUARTER 3 | QUARTER 4 | TOTAL GRAMS |
|---|---|---|---|---|---|
| 580 | 1,548.86 | 1,377.06 | 1,457.58 | 1,408.32 | 5,791.82 |
| 581 | 3,845.88 | 3,667.22 | 3,534.73 | 3,236.63 | 14,284.46 |
| 582 | 2,550.53 | 2,541.48 | 2,144.98 | 2,351.52 | 9,588.51 |
| 583 | 1,363.65 | 1,254.03 | 1,123.79 | 1,003.94 | 4,745.41 |
| 584 | 719.39 | 674.23 | 622.63 | 655.77 | 2,672.02 |
| 585 | 3,090.21 | 2,832.74 | 2,645.37 | 2,807.05 | 11,375.37 |
| 586 | 764.88 | 712.8 | 688.79 | 723.1 | 2,889.57 |
| 587 | 1,770.27 | 1,812.47 | 1,818.51 | 1,557.01 | 6,958.26 |
| 588 | 804.74 | 746.38 | 598.1 | 608.11 | 2,757.33 |
| TOTAL | 16,458.41 | 15,618.41 | 14,634.48 | 14,351.45 | 61,062.75 |

STATE: OHIO

| ZIP CODE | QUARTER 1 | QUARTER 2 | QUARTER 3 | QUARTER 4 | TOTAL GRAMS |
|---|---|---|---|---|---|
| 430 | 17,141.92 | 15,374.68 | 15,014.86 | 17,195.74 | 64,727.20 |
| 431 | 8,206.08 | 7,617.95 | 7,099.40 | 8,329.05 | 31,252.48 |
| 432 | 44,903.20 | 41,997.16 | 45,830.32 | 26,582.48 | 159,313.16 |
| 433 | 3,064.61 | 2,841.45 | 2,888.04 | 2,969.21 | 11,763.31 |
| 434 | 2,960.82 | 2,786.49 | 2,433.60 | 2,770.66 | 10,951.57 |
| 435 | 7,015.85 | 6,458.35 | 6,362.55 | 6,707.49 | 26,544.24 |
| 436 | 8,945.56 | 8,237.58 | 7,626.29 | 8,047.30 | 32,856.73 |
| 437 | 2,787.22 | 2,742.34 | 2,574.55 | 2,655.25 | 10,759.36 |
| 438 | 400.94 | 427.13 | 414.32 | 384.4 | 1,626.79 |
| 439 | 2,443.22 | 2,295.39 | 2,302.99 | 2,311.90 | 9,353.50 |
| 440 | 14,087.33 | 12,439.02 | 12,445.31 | 14,138.39 | 53,110.05 |
| 441 | 17,197.06 | 15,010.97 | 14,812.40 | 16,595.38 | 63,615.81 |
| 442 | 8,674.12 | 7,547.06 | 7,549.14 | 8,706.66 | 32,476.98 |
| 443 | 4,402.89 | 3,962.30 | 3,761.91 | 4,184.30 | 16,311.40 |
| 444 | 4,126.07 | 3,484.12 | 3,416.09 | 3,786.07 | 14,812.35 |
| 445 | 3,213.02 | 2,753.45 | 2,714.28 | 2,996.68 | 11,677.43 |
| 446 | 6,484.74 | 5,809.38 | 5,822.87 | 6,565.60 | 24,682.59 |
| 447 | 4,892.58 | 4,441.04 | 4,425.02 | 4,757.95 | 18,516.59 |
| 448 | 5,205.67 | 4,822.80 | 4,659.60 | 4,907.45 | 19,595.52 |
| 449 | 1,763.79 | 1,627.90 | 1,378.11 | 1,667.44 | 6,437.24 |
| 450 | 16,569.49 | 16,072.59 | 15,472.15 | 16,587.81 | 64,702.04 |
| 451 | 6,989.28 | 6,426 | 6,331.24 | 6,830.53 | 26,577.05 |
| 452 | 20,849.10 | 19,202.86 | 19,144.60 | 20,662.93 | 79,859.49 |
| 453 | 7,813.35 | 7,394.61 | 7,396.58 | 7,838.28 | 30,442.82 |
| 454 | 9,563.58 | 9,141.17 | 8,891.14 | 9,670.56 | 37,266.45 |
| 455 | 2,816.31 | 2,627 | 2,496.41 | 2,614.12 | 10,553.84 |
| 456 | 7,137.79 | 6,517.51 | 6,568.08 | 6,784.69 | 27,008.07 |
| 457 | 2,510.66 | 2,341.94 | 2,361.52 | 2,468.64 | 9,682.76 |
| 458 | 5,382.85 | 5,072.33 | 4,804.46 | 5,403.39 | 20,663.03 |

RETAIL DRUG DISTRIBUTION BY ZIP CODE WITHIN STATE BY GRAMS WT

| | | | | | |
|---|---|---|---|---|---|
| TOTAL | 247,549.10 | 227,472.57 | 226,997.83 | 225,120.35 | 927,139.85 |
| STATE: OKLAHOMA | | | | | |
| ZIP CODE | QUARTER 1 | QUARTER 2 | QUARTER 3 | QUARTER 4 | TOTAL GRAMS |
| 730 | 9,783.36 | 9,420.36 | 9,220.84 | 9,913.34 | 38,337.90 |

DRUG CODE:1724DRUG NAME:METHYLPHENIDATE (DL;D;L;ISOMERS)
STATE:OKLAHOMA

| ZIP CODE | QUARTER 1 | QUARTER 2 | QUARTER 3 | QUARTER 4 | TOTAL GRAMS |
|---|---|---|---|---|---|
| 731 | 7,139.07 | 6,734.20 | 6,576.77 | 6,855.90 | 27,305.94 |
| 734 | 772.16 | 718 | 657.42 | 678.78 | 2,826.36 |
| 735 | 3,572.41 | 3,482.63 | 3,323.21 | 3,367.64 | 13,745.89 |
| 736 | 848.77 | 798.59 | 842.96 | 821.4 | 3,311.72 |
| 737 | 1,421.32 | 1,287.77 | 1,186.58 | 1,268.55 | 5,164.22 |
| 738 | 262.6 | 199.9 | 169.71 | 175.58 | 807.79 |
| 739 | 72.36 | 77.73 | 73.71 | 92.26 | 316.06 |
| 740 | 11,084.50 | 9,966.70 | 10,237.68 | 10,747.60 | 42,036.48 |
| 741 | 6,961.77 | 6,660.53 | 6,335.77 | 6,524.86 | 26,482.93 |
| 743 | 2,004.31 | 1,796.75 | 1,720.19 | 1,792.23 | 7,313.48 |
| 744 | 3,235.76 | 2,944.77 | 2,953.79 | 2,916.62 | 12,050.94 |
| 745 | 948.41 | 830.02 | 743.06 | 813.61 | 3,335.10 |
| 746 | 727.59 | 686.56 | 633.39 | 736.81 | 2,784.35 |
| 747 | 768.54 | 663.71 | 679.21 | 719.69 | 2,831.15 |
| 748 | 1,949.94 | 1,690.67 | 1,590.90 | 1,625.44 | 6,856.95 |
| 749 | 858.28 | 789.81 | 776 | 899.88 | 3,323.97 |
| TOTAL | 52,411.15 | 48,748.70 | 47,721.19 | 49,950.19 | 198,831.23 |
| STATE: OREGON | | | | | |
| ZIP CODE | QUARTER 1 | QUARTER 2 | QUARTER 3 | QUARTER 4 | TOTAL GRAMS |
| 970 | 12,971.10 | 13,103.58 | 12,008.17 | 12,772.01 | 50,854.86 |
| 971 | 5,671.71 | 5,587 | 5,318.33 | 5,419.49 | 21,996.53 |
| 972 | 17,073.69 | 17,241.43 | 16,147.17 | 15,554.87 | 66,017.16 |
| 973 | 12,064.32 | 11,728.45 | 11,169.18 | 11,368.83 | 46,330.78 |
| 974 | 7,372.58 | 7,512.74 | 6,794.07 | 7,116.43 | 28,795.82 |
| 975 | 3,074.30 | 3,172.49 | 3,035.21 | 3,040.35 | 12,322.35 |
| 976 | 695.11 | 776.61 | 617.05 | 709.77 | 2,798.54 |
| 977 | 3,404.47 | 3,296.43 | 3,194.31 | 3,353.02 | 13,248.23 |
| 978 | 1,808.60 | 1,690.96 | 1,536.46 | 1,783.37 | 6,819.39 |
| 979 | 590.4 | 570.22 | 547.69 | 544.57 | 2,252.88 |
| TOTAL | 64,726.28 | 64,679.91 | 60,367.64 | 61,662.71 | 251,436.54 |
| STATE: PENNSYLVANIA | | | | | |
| ZIP CODE | QUARTER 1 | QUARTER 2 | QUARTER 3 | QUARTER 4 | TOTAL GRAMS |
| 150 | 8,585.87 | 6,757 | 7,095.78 | 7,679.03 | 30,117.68 |
| 151 | 6,643.66 | 5,367.33 | 5,740.42 | 6,654.57 | 24,405.98 |
| 152 | 12,866.34 | 10,730.30 | 10,999.15 | 12,767.37 | 47,363.16 |
| 153 | 4,034.30 | 3,008.66 | 3,179.86 | 3,692.77 | 13,915.59 |
| 154 | 1,616.98 | 1,563.20 | 1,503.31 | 1,581.04 | 6,264.53 |
| 155 | 1,290.85 | 1,073.77 | 962.87 | 1,053.50 | 4,380.99 |
| 156 | 5,499.58 | 4,724.01 | 4,965.44 | 5,435.82 | 20,624.85 |
| 157 | 2,551.19 | 2,133.84 | 2,187.58 | 2,346.46 | 9,219.07 |
| 158 | 2,429.40 | 2,148.09 | 2,035.83 | 2,231.78 | 8,845.10 |
| 159 | 2,212.85 | 2,018.94 | 2,134.95 | 2,263.36 | 8,630.10 |
| 160 | 4,125.54 | 3,266.20 | 3,506.72 | 4,160.73 | 15,059.19 |
| 161 | 4,299.92 | 3,693.84 | 3,612.50 | 4,179.75 | 15,786.01 |
| 162 | 1,646.57 | 1,598.11 | 1,571.96 | 1,686.93 | 6,503.57 |
| 163 | 2,825.23 | 2,515.13 | 2,392.50 | 2,488.25 | 10,221.11 |
| 164 | 1,129.10 | 961.95 | 1,025.20 | 1,077.95 | 4,194.20 |

| | | | | | |
|---|---|---|---|---|---|
| 165 | 4,910.19 | 4,463.94 | 4,577.54 | 4,821.12 | 18,772.79 |
| 166 | 4,147.72 | 3,634.97 | 3,604.73 | 3,815.66 | 15,203.08 |
| 167 | 707.76 | 764.38 | 725.18 | 743.29 | 2,940.61 |
| 168 | 3,436.86 | 3,210.92 | 3,282.80 | 3,385.33 | 13,315.91 |
| 169 | 751.4 | 723.46 | 736.99 | 792.65 | 3,004.50 |
| 170 | 15,955.30 | 15,292.52 | 15,023.42 | 15,059.93 | 61,331.17 |
| 171 | 3,523.34 | 3,240.28 | 3,157.62 | 3,197 | 13,118.24 |
| 172 | 4,208.58 | 3,991.14 | 3,724.13 | 4,055.63 | 15,979.48 |
| 173 | 6,991.05 | 6,876.15 | 6,456.15 | 6,663 | 26,986.35 |
| 174 | 5,158.19 | 5,331.15 | 4,597.43 | 5,034.09 | 20,120.86 |
| 175 | 5,686.10 | 5,092.57 | 4,703 | 5,131.49 | 20,613.16 |
| 176 | 4,982.96 | 4,932.65 | 4,570.10 | 4,675.58 | 19,161.29 |
| 177 | 3,194.26 | 3,102.29 | 2,820.22 | 3,090.70 | 12,207.47 |
| 178 | 3,765.74 | 3,526.73 | 3,274.93 | 3,625.52 | 14,192.92 |
| 179 | 1,645.17 | 1,504.70 | 1,401.28 | 1,575.85 | 6,127 |
| 180 | 6,114.25 | 6,138.16 | 5,727.72 | 5,941.39 | 23,921.52 |
| 181 | 3,368.37 | 3,394.63 | 3,017.07 | 3,262.53 | 13,042.60 |
| 182 | 1,412.29 | 1,396.95 | 1,286.31 | 1,393.88 | 5,489.43 |
| 183 | 1,702.36 | 1,575.79 | 1,434.05 | 1,587.82 | 6,300.02 |

DRUG CODE:1724DRUG NAME:METHYLPHENIDATE (DL;D;L;ISOMERS)
STATE:PENNSYLVANIA

| ZIP CODE | QUARTER 1 | QUARTER 2 | QUARTER 3 | QUARTER 4 | TOTAL GRAMS |
|---|---|---|---|---|---|
| 184 | 2,381.88 | 2,240.42 | 2,003.03 | 2,335.58 | 8,960.91 |
| 185 | 2,840.26 | 2,716.24 | 2,556.09 | 2,657.87 | 10,770.46 |
| 186 | 2,518.06 | 2,476.64 | 2,260.92 | 2,363.52 | 9,619.14 |
| 187 | 27,368.27 | 26,566.68 | 25,707.33 | 12,701.40 | 92,343.68 |
| 188 | 1,065.18 | 1,018.70 | 868.27 | 1,017.81 | 3,969.96 |
| 189 | 5,979.68 | 5,695.73 | 5,406.82 | 6,147.16 | 23,229.39 |
| 190 | 18,552.83 | 18,728.94 | 17,404.84 | 18,174.39 | 72,861 |
| 191 | 14,420.60 | 14,047.25 | 12,602.23 | 13,672.99 | 54,743.07 |
| 193 | 9,546.32 | 9,068.26 | 8,862.56 | 9,076.97 | 36,554.11 |
| 194 | 8,506.60 | 8,701.82 | 8,206.62 | 8,627.58 | 34,042.62 |
| 195 | 2,674.93 | 2,601.99 | 2,383.01 | 2,590.26 | 10,250.19 |
| 196 | 3,856.04 | 3,795.15 | 3,442.10 | 3,815 | 14,908.29 |
| TOTAL | 243,129.92 | 227,411.57 | 218,738.56 | 220,332.30 | 909,612.35 |

STATE: PUERTO RICO

| ZIP CODE | QUARTER 1 | QUARTER 2 | QUARTER 3 | QUARTER 4 | TOTAL GRAMS |
|---|---|---|---|---|---|
| 6 | 5,820.22 | 5,030.47 | 5,027.18 | 5,280.82 | 21,158.69 |
| 7 | 7,192.68 | 6,438.96 | 6,379.62 | 6,969.49 | 26,980.75 |
| 9 | 10,668.40 | 8,897.46 | 8,719.93 | 10,299.87 | 38,585.66 |
| TOTAL | 23,681.30 | 20,366.89 | 20,126.73 | 22,550.18 | 86,725.10 |

STATE: RHODE ISLAND

| ZIP CODE | QUARTER 1 | QUARTER 2 | QUARTER 3 | QUARTER 4 | TOTAL GRAMS |
|---|---|---|---|---|---|
| 28 | 13,428.13 | 13,053.36 | 12,192.73 | 13,071.60 | 51,745.82 |
| 29 | 6,738.19 | 6,569.56 | 5,853.64 | 6,565.97 | 25,727.36 |
| TOTAL | 20,166.32 | 19,622.92 | 18,046.37 | 19,637.57 | 77,473.18 |

STATE: SOUTH CAROLINA

| ZIP CODE | QUARTER 1 | QUARTER 2 | QUARTER 3 | QUARTER 4 | TOTAL GRAMS |
|---|---|---|---|---|---|
| 290 | 7,284.01 | 6,634.06 | 6,490.09 | 7,067.05 | 27,475.21 |
| 291 | 6,162.57 | 5,715.57 | 5,562.80 | 6,073.99 | 23,514.93 |
| 292 | 6,737.09 | 5,852.92 | 6,003.51 | 6,129.14 | 24,722.66 |
| 293 | 8,313.52 | 7,523.18 | 7,287.47 | 7,735.78 | 30,859.95 |
| 294 | 20,475.62 | 18,392.90 | 17,931.10 | 18,871.46 | 75,671.08 |
| 295 | 11,902.71 | 10,773.79 | 10,913.53 | 11,262.17 | 44,852.20 |
| 296 | 20,759.83 | 19,228.74 | 18,554.52 | 20,151.43 | 78,694.52 |
| 297 | 7,433.10 | 6,807.63 | 6,336.98 | 7,429.05 | 28,006.76 |

| | | | | | |
|---|---|---|---|---|---|
| 298 | 3,473.06 | 2,990.87 | 3,108.92 | 3,193.34 | 12,766.19 |
| 299 | 3,195.91 | 2,947.23 | 2,902.20 | 3,118.63 | 12,163.97 |
| TOTAL | 95,737.42 | 86,866.89 | 85,091.12 | 91,032.04 | 358,727.47 |

STATE: SOUTH DAKOTA

| ZIP CODE | QUARTER 1 | QUARTER 2 | QUARTER 3 | QUARTER 4 | TOTAL GRAMS |
|---|---|---|---|---|---|
| 570 | 2,678.19 | 2,494 | 2,386.30 | 2,627.64 | 10,186.13 |
| 571 | 13,329.28 | 12,429.95 | 12,963.89 | 8,913.47 | 47,636.59 |
| 572 | 1,167.62 | 1,082.04 | 997.3 | 1,113.03 | 4,359.99 |
| 573 | 1,748.01 | 1,603.21 | 1,621.13 | 1,669.46 | 6,641.81 |
| 574 | 1,183.07 | 1,097.59 | 1,141.88 | 1,148.62 | 4,571.16 |
| 575 | 891.5 | 924.77 | 771.61 | 905.52 | 3,493.40 |
| 576 | 193.35 | 165.33 | 173.63 | 119.03 | 651.34 |
| 577 | 3,227.33 | 2,972.46 | 2,784.54 | 2,960.98 | 11,945.31 |
| TOTAL | 24,418.35 | 22,769.35 | 22,840.28 | 19,457.75 | 89,485.73 |

STATE: TENNESSEE

| ZIP CODE | QUARTER 1 | QUARTER 2 | QUARTER 3 | QUARTER 4 | TOTAL GRAMS |
|---|---|---|---|---|---|
| 370 | 13,264.21 | 12,102.89 | 12,220.87 | 12,960.31 | 50,548.28 |
| 371 | 8,592.26 | 8,112.78 | 8,178.80 | 8,597.85 | 33,481.69 |
| 372 | 6,426.43 | 6,035.20 | 5,848.68 | 6,088.42 | 24,398.73 |
| 373 | 8,509.33 | 7,877.85 | 7,913.49 | 8,202.26 | 32,502.93 |
| 374 | 4,030.69 | 3,690 | 3,717.92 | 3,922.73 | 15,361.34 |
| 376 | 5,218.03 | 4,856.49 | 5,122.77 | 5,082.40 | 20,279.69 |
| 377 | 5,064.97 | 4,671.55 | 4,904.58 | 5,014.58 | 19,655.68 |
| 378 | 7,107.82 | 6,683.64 | 6,690.38 | 7,082.69 | 27,564.53 |
| 379 | 7,162.20 | 6,421.62 | 6,714.01 | 6,800 | 27,097.83 |
| 380 | 6,549.60 | 5,889.97 | 5,873.89 | 6,091.77 | 24,405.23 |
| 381 | 8,403.11 | 7,717.92 | 7,401.80 | 7,888.56 | 31,411.39 |
| 382 | 1,819.66 | 1,490.99 | 1,659.52 | 1,578.30 | 6,548.47 |
| 383 | 3,893.23 | 3,291.83 | 3,567.78 | 3,620.08 | 14,372.92 |
| 384 | 2,192.60 | 2,054.10 | 2,079.52 | 2,146 | 8,472.22 |
| 385 | 2,104.34 | 2,053.05 | 2,073.82 | 2,257.96 | 8,489.17 |
| TOTAL | 90,338.48 | 82,949.88 | 83,967.83 | 87,333.91 | 344,590.10 |

DRUG CODE:1724DRUG NAME:METHYLPHENIDATE (DL;D;L;ISOMERS)

STATE: TEXAS

| ZIP CODE | QUARTER 1 | QUARTER 2 | QUARTER 3 | QUARTER 4 | TOTAL GRAMS |
|---|---|---|---|---|---|
| 750 | 35,662.84 | 33,620.32 | 31,732.35 | 35,057.51 | 136,073.02 |
| 751 | 9,848.96 | 9,178.48 | 8,815.10 | 9,620.62 | 37,463.16 |
| 752 | 12,268.66 | 11,027.38 | 10,977.28 | 12,201.42 | 46,474.74 |
| 753 | 15.48 | 8.65 | 22.57 | 16.26 | 62.96 |
| 754 | 5,473.42 | 4,953.60 | 4,906.89 | 5,580.76 | 20,914.67 |
| 755 | 1,749.85 | 1,395.17 | 1,336.17 | 1,375.81 | 5,857 |
| 756 | 4,400.30 | 4,069.40 | 3,892.93 | 4,353.99 | 16,716.62 |
| 757 | 5,907.15 | 5,150.84 | 4,921.02 | 5,323.64 | 21,302.65 |
| 758 | 1,373.62 | 1,225.55 | 1,207.35 | 1,278.43 | 5,084.95 |
| 759 | 4,582.20 | 4,067.85 | 4,033.42 | 4,369.75 | 17,053.22 |
| 760 | 18,674.01 | 17,841.64 | 16,695.51 | 18,204.25 | 71,415.41 |
| 761 | 11,293.97 | 10,291.56 | 9,890.33 | 10,678.17 | 42,154.03 |
| 762 | 7,629.92 | 7,213.89 | 6,754.04 | 7,822.56 | 29,420.41 |
| 763 | 4,255.24 | 3,837.10 | 3,953.92 | 3,856.03 | 15,902.29 |
| 764 | 1,585.45 | 1,398.57 | 1,351.99 | 1,452.40 | 5,788.41 |
| 765 | 10,759.83 | 10,723.32 | 9,855 | 9,601.72 | 40,939.87 |
| 766 | 1,442.91 | 1,247.58 | 1,234.26 | 1,330.64 | 5,255.39 |
| 767 | 4,114.53 | 3,784.17 | 3,737.66 | 4,117.39 | 15,753.75 |
| 768 | 1,515.04 | 1,326.21 | 1,286 | 1,472.83 | 5,600.08 |
| 769 | 3,173.39 | 2,739.37 | 2,567.32 | 2,894.13 | 11,374.21 |

| | | | | | |
|---|---|---|---|---|---|
| 770 | 26,255.28 | 23,657.32 | 22,742.40 | 25,565.21 | 98,220.21 |
| 772 | 6.05 | 0 | 4.32 | 1.73 | 12.1 |
| 773 | 19,025.92 | 17,055.23 | 16,523.49 | 18,244.09 | 70,848.73 |
| 774 | 18,540.49 | 16,156.19 | 15,892.96 | 17,729.07 | 68,318.71 |
| 775 | 17,247.67 | 16,320.79 | 15,200.27 | 17,273.01 | 66,041.74 |
| 776 | 4,362.78 | 3,812.31 | 3,861.30 | 4,070.08 | 16,106.47 |
| 777 | 1,909.08 | 1,712.28 | 1,600.82 | 1,800.35 | 7,022.53 |
| 778 | 4,756.04 | 4,536.84 | 4,354.17 | 4,816.74 | 18,463.79 |
| 779 | 2,387.67 | 2,297.34 | 2,178.37 | 2,415.18 | 9,278.56 |
| 780 | 6,069.85 | 5,289.47 | 5,266.79 | 5,615.12 | 22,241.23 |
| 781 | 6,446.64 | 5,824.97 | 5,521.68 | 6,270.66 | 24,063.95 |
| 782 | 26,676.08 | 25,122.82 | 23,478.96 | 25,935.79 | 101,213.65 |
| 783 | 3,441.82 | 2,989.48 | 2,831.74 | 3,205.38 | 12,468.42 |
| 784 | 6,409.30 | 5,695.94 | 5,438.50 | 5,840.27 | 23,384.01 |
| 785 | 13,093.62 | 11,306.81 | 10,792.94 | 11,759.52 | 46,952.89 |
| 786 | 13,115.14 | 11,838.98 | 10,974.73 | 12,931 | 48,859.85 |
| 787 | 13,336.70 | 12,472.89 | 11,726.55 | 13,472.08 | 51,008.22 |
| 788 | 1,622.31 | 1,464.18 | 1,322.82 | 1,396.03 | 5,805.34 |
| 789 | 699.42 | 629.85 | 616.59 | 691.08 | 2,636.94 |
| 790 | 1,451.02 | 1,315.12 | 1,222.17 | 1,389.71 | 5,378.02 |
| 791 | 2,205.53 | 2,325.58 | 2,010.79 | 2,177.87 | 8,719.77 |
| 792 | 333.97 | 253.4 | 284.58 | 305.8 | 1,177.75 |
| 793 | 1,209.90 | 1,031.71 | 1,074.13 | 1,057.21 | 4,372.95 |
| 794 | 5,315.54 | 4,957.55 | 4,607.06 | 5,298.74 | 20,178.89 |
| 795 | 1,108.71 | 963.48 | 989.19 | 1,050.33 | 4,111.71 |
| 796 | 3,719.07 | 3,422.09 | 3,212.27 | 3,544.15 | 13,897.58 |
| 797 | 4,841.66 | 4,360.66 | 4,026.89 | 4,564.92 | 17,794.13 |
| 798 | 279.94 | 257.4 | 263.58 | 267.08 | 1,068 |
| 799 | 8,524.51 | 8,312.95 | 7,926 | 8,401.70 | 33,165.16 |
| TOTAL | 360,118.48 | 330,484.28 | 315,117.17 | 347,698.21 | 1,353,418.14 |
| STATE: UTAH | | | | | |
| ZIP CODE | QUARTER 1 | QUARTER 2 | QUARTER 3 | QUARTER 4 | TOTAL GRAMS |
| 840 | 24,201.90 | 23,623.70 | 23,420.09 | 24,012.58 | 95,258.27 |
| 841 | 9,713.43 | 9,464.13 | 8,942.90 | 9,150.88 | 37,271.34 |
| 843 | 2,276.57 | 2,172.58 | 2,114.78 | 2,329.27 | 8,893.20 |
| 844 | 2,642.51 | 2,655.73 | 2,619.47 | 2,795.45 | 10,713.16 |
| 845 | 486.7 | 483.3 | 479.1 | 482.04 | 1,931.14 |
| 846 | 4,681.28 | 4,689.05 | 4,607.76 | 4,850.77 | 18,828.86 |
| 847 | 3,353.46 | 3,007.96 | 3,002.58 | 3,229.26 | 12,593.26 |
| TOTAL | 47,355.85 | 46,096.45 | 45,186.68 | 46,850.25 | 185,489.23 |
| STATE: VERMONT | | | | | |
| ZIP CODE | QUARTER 1 | QUARTER 2 | QUARTER 3 | QUARTER 4 | TOTAL GRAMS |
| 50 | 1,523.90 | 1,612.59 | 1,478.98 | 1,384.09 | 5,999.56 |
| 51 | 931.54 | 827.43 | 799.55 | 767.42 | 3,325.94 |
| 52 | 1,210.60 | 1,177.16 | 1,156.55 | 1,229.93 | 4,774.24 |
| 53 | 1,238.26 | 1,296.27 | 1,086.84 | 1,235.94 | 4,857.31 |

DRUG CODE:1724DRUG NAME:METHYLPHENIDATE (DL;D;L;ISOMERS)
STATE:VERMONT

| ZIP CODE | QUARTER 1 | QUARTER 2 | QUARTER 3 | QUARTER 4 | TOTAL GRAMS |
|---|---|---|---|---|---|
| 54 | 6,557.73 | 6,370.94 | 6,207.81 | 6,428.39 | 25,564.87 |
| 56 | 3,612.90 | 3,650.53 | 3,430.83 | 3,356.46 | 14,050.72 |
| 57 | 2,461.98 | 2,340.04 | 2,302.43 | 2,453.43 | 9,557.88 |
| 58 | 1,906.08 | 1,862.48 | 1,831.46 | 1,993.26 | 7,593.28 |
| TOTAL | 19,442.99 | 19,137.44 | 18,294.45 | 18,848.92 | 75,723.80 |

STATE: VIRGIN ISLANDS

RETAIL DRUG DISTRIBUTION BY ZIP CODE WITHIN STATE BY GRAMS WT

| ZIP CODE | QUARTER 1 | QUARTER 2 | QUARTER 3 | QUARTER 4 | TOTAL GRAMS |
|---|---|---|---|---|---|
| 8 | 215.51 | 201.95 | 183.6 | 198.22 | 799.28 |
| TOTAL | 215.51 | 201.95 | 183.6 | 198.22 | 799.28 |

STATE: VIRGINIA

| ZIP CODE | QUARTER 1 | QUARTER 2 | QUARTER 3 | QUARTER 4 | TOTAL GRAMS |
|---|---|---|---|---|---|
| 201 | 12,491.37 | 12,627.08 | 11,619.89 | 12,591.63 | 49,329.97 |
| 220 | 9,069.87 | 9,720.88 | 9,487.66 | 8,520.21 | 36,798.62 |
| 221 | 5,444.29 | 5,567 | 5,436.39 | 5,510.94 | 21,958.62 |
| 222 | 2,879.12 | 2,868.12 | 2,751.84 | 2,792.74 | 11,291.82 |
| 223 | 3,279.72 | 3,291.40 | 2,937.97 | 3,425.41 | 12,934.50 |
| 224 | 4,957.86 | 4,916.36 | 4,372.86 | 4,924.60 | 19,171.68 |
| 225 | 2,360.09 | 2,237.49 | 2,163.96 | 2,337.40 | 9,098.94 |
| 226 | 2,950.85 | 2,841.97 | 2,641.41 | 3,007.87 | 11,442.10 |
| 227 | 953.64 | 905.39 | 956.91 | 992.07 | 3,808.01 |
| 228 | 2,045.41 | 1,951.74 | 1,893.77 | 2,061.78 | 7,952.70 |
| 229 | 6,180.15 | 5,764.50 | 5,598.85 | 6,120.86 | 23,664.36 |
| 230 | 4,005.48 | 3,969.73 | 3,658.74 | 3,975.26 | 15,609.21 |
| 231 | 8,679.01 | 8,373.57 | 7,758.16 | 8,664.12 | 33,474.86 |
| 232 | 10,319.21 | 9,788.92 | 9,314.24 | 10,284.22 | 39,706.59 |
| 233 | 6,176.78 | 6,208.47 | 5,200.47 | 5,767.42 | 23,353.14 |
| 234 | 13,088.20 | 12,717.40 | 10,985.55 | 11,995.80 | 48,786.95 |
| 235 | 3,457.16 | 3,584.03 | 3,080.57 | 3,488.16 | 13,609.92 |
| 236 | 8,556.96 | 7,432.51 | 7,693.76 | 7,227.20 | 30,910.43 |
| 237 | 3,393.13 | 3,435.01 | 2,793.37 | 3,220.19 | 12,841.70 |
| 238 | 5,359.80 | 5,157.62 | 4,522.23 | 5,131.58 | 20,171.23 |
| 239 | 1,312.40 | 1,167.84 | 1,254.55 | 1,297.51 | 5,032.30 |
| 240 | 5,552.37 | 5,237.26 | 5,265.14 | 5,810.74 | 21,865.51 |
| 241 | 3,545.86 | 3,348.38 | 3,314.09 | 3,457.40 | 13,665.73 |
| 242 | 2,668.58 | 2,550.90 | 2,509.43 | 2,589.25 | 10,318.16 |
| 243 | 1,820.52 | 1,804.58 | 1,808.40 | 1,936.91 | 7,370.41 |
| 244 | 2,755.92 | 2,581.38 | 2,474.59 | 2,722.62 | 10,534.51 |
| 245 | 6,031.14 | 5,882.28 | 5,412.06 | 6,237.37 | 23,562.85 |
| 246 | 911.39 | 976.19 | 981.89 | 926.69 | 3,796.16 |
| TOTAL | 140,246.28 | 136,908 | 127,888.75 | 137,017.95 | 542,060.98 |

STATE: WASHINGTON

| ZIP CODE | QUARTER 1 | QUARTER 2 | QUARTER 3 | QUARTER 4 | TOTAL GRAMS |
|---|---|---|---|---|---|
| 980 | 23,212.96 | 22,927.06 | 21,273.39 | 24,643.12 | 92,056.53 |
| 981 | 14,259.42 | 14,683.96 | 13,648.17 | 12,423.77 | 55,015.32 |
| 982 | 15,374.65 | 15,585.60 | 14,555.27 | 15,244.74 | 60,760.26 |
| 983 | 12,308.30 | 11,791.93 | 11,476.96 | 12,087.56 | 47,664.75 |
| 984 | 8,456.14 | 8,175.93 | 7,639.74 | 7,642.89 | 31,914.70 |
| 985 | 7,459.12 | 7,086.01 | 6,699.42 | 7,214.20 | 28,458.75 |
| 986 | 8,854.51 | 9,263.95 | 8,430.36 | 9,109.37 | 35,658.19 |
| 988 | 2,675.63 | 2,687.72 | 2,335.32 | 2,574.47 | 10,273.14 |
| 989 | 3,555.53 | 3,509.22 | 3,112.11 | 3,343.50 | 13,520.36 |
| 990 | 1,750.83 | 1,877.69 | 1,767.14 | 1,708.39 | 7,104.05 |
| 991 | 1,386.33 | 1,212.19 | 1,381.02 | 1,372.61 | 5,352.15 |
| 992 | 8,239.88 | 8,313.25 | 7,661.40 | 7,919.40 | 32,133.93 |
| 993 | 5,473.16 | 5,514.50 | 5,209.59 | 5,151.51 | 21,348.76 |
| 994 | 506.46 | 486.55 | 451.37 | 532.7 | 1,977.08 |
| TOTAL | 113,512.92 | 113,115.56 | 105,641.26 | 110,968.23 | 443,237.97 |

STATE: WEST VIRGINIA

| ZIP CODE | QUARTER 1 | QUARTER 2 | QUARTER 3 | QUARTER 4 | TOTAL GRAMS |
|---|---|---|---|---|---|
| 247 | 1,122.95 | 1,055.89 | 968.83 | 1,060 | 4,207.67 |
| 248 | 415.81 | 419.16 | 411.05 | 446.79 | 1,692.81 |
| 249 | 731.04 | 692.65 | 689.46 | 746.64 | 2,859.79 |
| 250 | 1,213.86 | 1,149.77 | 1,093.26 | 1,111.54 | 4,568.43 |
| 251 | 1,090.68 | 916.55 | 937.62 | 1,023.35 | 3,968.20 |

| | | | | | |
|---|---|---|---|---|---|
| 252 | 789.4 | 794.85 | 754.12 | 730 | 3,068.37 |
| 253 | 3,044.57 | 2,913.25 | 3,053.23 | 2,974.25 | 11,985.30 |

DRUG CODE:1724DRUG NAME:METHYLPHENIDATE (DL;D;L;ISOMERS)
STATE:WEST VIRGINIA

| ZIP CODE | QUARTER 1 | QUARTER 2 | QUARTER 3 | QUARTER 4 | TOTAL GRAMS |
|---|---|---|---|---|---|
| 254 | 3,149.05 | 3,067.03 | 2,741.86 | 3,169.68 | 12,127.62 |
| 255 | 3,118.18 | 2,780.47 | 2,886.97 | 3,092.26 | 11,877.88 |
| 256 | 694.37 | 709.3 | 654.4 | 714.27 | 2,772.34 |
| 257 | 2,789.54 | 2,525.16 | 2,505.61 | 2,625.19 | 10,445.50 |
| 258 | 2,449.97 | 2,348.47 | 2,338.02 | 2,404.61 | 9,541.07 |
| 259 | 855.49 | 921.65 | 916.3 | 910.65 | 3,604.09 |
| 260 | 2,124.41 | 1,973.83 | 1,969.46 | 2,099.23 | 8,166.93 |
| 261 | 2,257.73 | 2,164.21 | 2,155.99 | 2,093.77 | 8,671.70 |
| 262 | 901.89 | 861.64 | 792.53 | 838.29 | 3,394.35 |
| 263 | 1,464.50 | 1,394.93 | 1,310.58 | 1,415.15 | 5,585.16 |
| 264 | 439.4 | 450.44 | 436.02 | 456.86 | 1,782.72 |
| 265 | 3,046.49 | 2,868.65 | 2,745.11 | 2,992.40 | 11,652.65 |
| 266 | 413.88 | 414.87 | 410.72 | 417.03 | 1,656.50 |
| 267 | 931.47 | 816.62 | 751.08 | 836.68 | 3,335.85 |
| 268 | 650.55 | 530.59 | 580.93 | 576.2 | 2,338.27 |
| TOTAL | 33,695.23 | 31,769.98 | 31,103.15 | 32,734.84 | 129,303.20 |

STATE: WISCONSIN

| ZIP CODE | QUARTER 1 | QUARTER 2 | QUARTER 3 | QUARTER 4 | TOTAL GRAMS |
|---|---|---|---|---|---|
| 530 | 12,446.10 | 12,166.47 | 11,522.16 | 12,713.19 | 48,847.92 |
| 531 | 11,537.08 | 10,853.44 | 10,274.62 | 11,045.43 | 43,710.57 |
| 532 | 12,340.55 | 11,825.86 | 10,935.99 | 11,967.37 | 47,069.77 |
| 534 | 2,558.44 | 2,492.83 | 2,353.90 | 2,325.12 | 9,730.29 |
| 535 | 10,778.67 | 10,541.85 | 9,797.46 | 9,910.59 | 41,028.57 |
| 537 | 6,778.21 | 6,674.75 | 6,259.78 | 6,710.15 | 26,422.89 |
| 538 | 1,016.84 | 953.44 | 825.08 | 947.6 | 3,742.96 |
| 539 | 3,725.34 | 3,642.91 | 3,412.29 | 3,404.03 | 14,184.57 |
| 540 | 2,423.52 | 2,300.35 | 2,142.77 | 2,527.61 | 9,394.25 |
| 541 | 3,902.51 | 3,755.93 | 3,490.25 | 4,126.80 | 15,275.49 |
| 542 | 2,369.67 | 2,164.41 | 2,019.19 | 2,170.14 | 8,723.41 |
| 543 | 3,663.50 | 3,575.58 | 3,309.25 | 3,737.88 | 14,286.21 |
| 544 | 8,179.13 | 7,721.20 | 6,967.56 | 7,791.77 | 30,659.66 |
| 545 | 1,627.99 | 1,606.79 | 1,590.55 | 1,515.79 | 6,341.12 |
| 546 | 5,538.25 | 5,457.56 | 5,049.29 | 5,536.76 | 21,581.86 |
| 547 | 5,887.01 | 5,824.38 | 5,398.10 | 5,905.78 | 23,015.27 |
| 548 | 2,541.20 | 2,667.94 | 2,411.04 | 2,795.82 | 10,416 |
| 549 | 10,708.56 | 10,516.39 | 9,841.79 | 10,431.48 | 41,498.22 |
| TOTAL | 108,022.57 | 104,742.08 | 97,601.07 | 105,563.31 | 415,929.03 |

STATE: WYOMING

| ZIP CODE | QUARTER 1 | QUARTER 2 | QUARTER 3 | QUARTER 4 | TOTAL GRAMS |
|---|---|---|---|---|---|
| 820 | 2,483.10 | 2,125.77 | 2,091.65 | 2,349.68 | 9,050.20 |
| 822 | 359.53 | 306.82 | 243.26 | 301.04 | 1,210.65 |
| 823 | 148.73 | 143.96 | 157.85 | 129.99 | 580.53 |
| 824 | 966.73 | 909.09 | 870.53 | 857.02 | 3,603.37 |
| 825 | 514.22 | 468.43 | 496.68 | 450.65 | 1,929.98 |
| 826 | 1,950.18 | 1,678.68 | 1,597.60 | 1,651.68 | 6,878.14 |
| 827 | 719.04 | 729.59 | 654.41 | 699.41 | 2,802.45 |
| 828 | 676.33 | 574.41 | 568.04 | 620.91 | 2,439.69 |
| 829 | 891.02 | 814.06 | 807.67 | 856.07 | 3,368.82 |
| 830 | 284.4 | 282.24 | 292.95 | 301.58 | 1,161.17 |
| 831 | 240.3 | 208.31 | 197.36 | 252.1 | 898.07 |
| TOTAL | 9,233.58 | 8,241.36 | 7,978 | 8,470.13 | 33,923.07 |

RETAIL DRUG DISTRIBUTION BY ZIP CODE WITHIN STATE BY GRAMS WT

DRUG CODE:2012DRUG NAME:GAMMA HYDROXYBUTYRIC ACID(FDA APPROV

STATE: ALABAMA

| ZIP CODE | QUARTER 1 | QUARTER 2 | QUARTER 3 | QUARTER 4 | TOTAL GRAMS |
|---|---|---|---|---|---|
| 352 | 371.52 | 0 | 0 | 0 | 371.52 |
| TOTAL | 371.52 | 0 | 0 | 0 | 371.52 |

STATE: CALIFORNIA

| ZIP CODE | QUARTER 1 | QUARTER 2 | QUARTER 3 | QUARTER 4 | TOTAL GRAMS |
|---|---|---|---|---|---|
| 908 | 0 | 0 | 965.95 | 520.13 | 1,486.08 |
| 928 | 0 | 0 | 371.52 | 1,337.47 | 1,708.99 |
| 940 | 4,829.76 | 3,938.11 | 668.74 | 222.91 | 9,659.52 |
| TOTAL | 4,829.76 | 3,938.11 | 2,006.21 | 2,080.51 | 12,854.59 |

STATE: FLORIDA

| ZIP CODE | QUARTER 1 | QUARTER 2 | QUARTER 3 | QUARTER 4 | TOTAL GRAMS |
|---|---|---|---|---|---|
| 322 | 148.61 | 0 | 0 | 0 | 148.61 |

DRUG CODE:2012DRUG NAME:GAMMA HYDROXYBUTYRIC ACID(FDA APPROV

STATE:FLORIDA

| ZIP CODE | QUARTER 1 | QUARTER 2 | QUARTER 3 | QUARTER 4 | TOTAL GRAMS |
|---|---|---|---|---|---|
| TOTAL | 148.61 | 0 | 0 | 0 | 148.61 |

STATE: GEORGIA

| ZIP CODE | QUARTER 1 | QUARTER 2 | QUARTER 3 | QUARTER 4 | TOTAL GRAMS |
|---|---|---|---|---|---|
| 303 | 297.22 | 1,634.69 | 668.74 | 0 | 2,600.65 |
| TOTAL | 297.22 | 1,634.69 | 668.74 | 0 | 2,600.65 |

STATE: ILLINOIS

| ZIP CODE | QUARTER 1 | QUARTER 2 | QUARTER 3 | QUARTER 4 | TOTAL GRAMS |
|---|---|---|---|---|---|
| 606 | 668.74 | 371.52 | 1,263.17 | 743.04 | 3,046.47 |
| TOTAL | 668.74 | 371.52 | 1,263.17 | 743.04 | 3,046.47 |

STATE: LOUISIANA

| ZIP CODE | QUARTER 1 | QUARTER 2 | QUARTER 3 | QUARTER 4 | TOTAL GRAMS |
|---|---|---|---|---|---|
| 700 | 0 | 0 | 222.91 | 520.13 | 743.04 |
| TOTAL | 0 | 0 | 222.91 | 520.13 | 743.04 |

STATE: MASSACHUSETTS

| ZIP CODE | QUARTER 1 | QUARTER 2 | QUARTER 3 | QUARTER 4 | TOTAL GRAMS |
|---|---|---|---|---|---|
| 21 | 1,337.47 | 1,857.60 | 0 | 0 | 3,195.07 |
| TOTAL | 1,337.47 | 1,857.60 | 0 | 0 | 3,195.07 |

STATE: MICHIGAN

| ZIP CODE | QUARTER 1 | QUARTER 2 | QUARTER 3 | QUARTER 4 | TOTAL GRAMS |
|---|---|---|---|---|---|
| 481 | 0 | 0 | 0 | 297.22 | 297.22 |
| TOTAL | 0 | 0 | 0 | 297.22 | 297.22 |

STATE: MISSOURI

| ZIP CODE | QUARTER 1 | QUARTER 2 | QUARTER 3 | QUARTER 4 | TOTAL GRAMS |
|---|---|---|---|---|---|
| 631 | 12,981,428.93 | 4,943,965.25 | 7,389,458.50 | 0 | 25,314,852.68 |
| TOTAL | 12,981,428.93 | 4,943,965.25 | 7,389,458.50 | 0 | 25,314,852.68 |

STATE: NEBRASKA

| ZIP CODE | QUARTER 1 | QUARTER 2 | QUARTER 3 | QUARTER 4 | TOTAL GRAMS |
|---|---|---|---|---|---|
| 681 | 222.91 | 0 | 0 | 0 | 222.91 |
| TOTAL | 222.91 | 0 | 0 | 0 | 222.91 |

STATE: NEW YORK

RETAIL DRUG DISTRIBUTION BY ZIP CODE WITHIN STATE BY GRAMS WT

| ZIP CODE | QUARTER 1 | QUARTER 2 | QUARTER 3 | QUARTER 4 | TOTAL GRAMS |
|---|---|---|---|---|---|
| 104 | 1,708.99 | 3,715.20 | 3,640.90 | 3,863.81 | 12,928.90 |
| TOTAL | 1,708.99 | 3,715.20 | 3,640.90 | 3,863.81 | 12,928.90 |

STATE: NORTH CAROLINA

| ZIP CODE | QUARTER 1 | QUARTER 2 | QUARTER 3 | QUARTER 4 | TOTAL GRAMS |
|---|---|---|---|---|---|
| 276 | 668.74 | 0 | 0 | 74.3 | 743.04 |
| 277 | 0 | 520.13 | 1,188.86 | 520.13 | 2,229.12 |
| 280 | 891.65 | 594.43 | 0 | 0 | 1,486.08 |
| TOTAL | 1,560.39 | 1,114.56 | 1,188.86 | 594.43 | 4,458.24 |

STATE: OHIO

| ZIP CODE | QUARTER 1 | QUARTER 2 | QUARTER 3 | QUARTER 4 | TOTAL GRAMS |
|---|---|---|---|---|---|
| 441 | 297.22 | 891.65 | 1,114.56 | 817.34 | 3,120.77 |
| 452 | 0 | 520.13 | 1,263.17 | 1,560.38 | 3,343.68 |
| TOTAL | 297.22 | 1,411.78 | 2,377.73 | 2,377.72 | 6,464.45 |

STATE: SOUTH CAROLINA

| ZIP CODE | QUARTER 1 | QUARTER 2 | QUARTER 3 | QUARTER 4 | TOTAL GRAMS |
|---|---|---|---|---|---|
| 292 | 817.34 | 3,104.26 | 1,560.38 | 520.13 | 6,002.11 |
| 296 | 594.43 | 520.13 | 0 | 1,188.86 | 2,303.42 |
| TOTAL | 1,411.77 | 3,624.39 | 1,560.38 | 1,708.99 | 8,305.53 |

STATE: TENNESSEE

| ZIP CODE | QUARTER 1 | QUARTER 2 | QUARTER 3 | QUARTER 4 | TOTAL GRAMS |
|---|---|---|---|---|---|
| 381 | 0 | 297.22 | 743.04 | 1,263.17 | 2,303.43 |
| TOTAL | 0 | 297.22 | 743.04 | 1,263.17 | 2,303.43 |

STATE: TEXAS

| ZIP CODE | QUARTER 1 | QUARTER 2 | QUARTER 3 | QUARTER 4 | TOTAL GRAMS |
|---|---|---|---|---|---|
| 752 | 0 | 668.74 | 222.91 | 1,560.38 | 2,452.03 |
| 770 | 520.13 | 0 | 668.74 | 1,486.08 | 2,674.95 |
| 782 | 0 | 0 | 1,040.26 | 891.65 | 1,931.91 |
| 787 | 0 | 668.74 | 1,486.08 | 965.95 | 3,120.77 |
| TOTAL | 520.13 | 1,337.48 | 3,417.99 | 4,904.06 | 10,179.66 |

STATE: VIRGINIA

| ZIP CODE | QUARTER 1 | QUARTER 2 | QUARTER 3 | QUARTER 4 | TOTAL GRAMS |
|---|---|---|---|---|---|
| 235 | 2,897.86 | 2,154.82 | 1,263.17 | 965.95 | 7,281.80 |
| TOTAL | 2,897.86 | 2,154.82 | 1,263.17 | 965.95 | 7,281.80 |

STATE: WASHINGTON

| ZIP CODE | QUARTER 1 | QUARTER 2 | QUARTER 3 | QUARTER 4 | TOTAL GRAMS |
|---|---|---|---|---|---|

DRUG CODE:2012DRUG NAME:GAMMA HYDROXYBUTYRIC ACID(FDA APPROV
STATE:WASHINGTON

| ZIP CODE | QUARTER 1 | QUARTER 2 | QUARTER 3 | QUARTER 4 | TOTAL GRAMS |
|---|---|---|---|---|---|
| 981 | 1,040.26 | 297.22 | 1,040.26 | 891.65 | 3,269.39 |
| TOTAL | 1,040.26 | 297.22 | 1,040.26 | 891.65 | 3,269.39 |

DRUG CODE:2100DRUG NAME:BARBITURIC ACID DERIVIATIVE OR SALT [PER
STATE: ALABAMA

| ZIP CODE | QUARTER 1 | QUARTER 2 | QUARTER 3 | QUARTER 4 | TOTAL GRAMS |
|---|---|---|---|---|---|
| 351 | 0 | 0 | 0 | 5 | 5 |
| 352 | 0 | 0 | 15 | 5 | 20 |
| 362 | 0 | 0 | 5 | 5 | 10 |
| 363 | 0 | 0 | 10 | 0 | 10 |
| TOTAL | 0 | 0 | 30 | 15 | 45 |

RETAIL DRUG DISTRIBUTION BY ZIP CODE WITHIN STATE BY GRAMS WT

STATE: ALASKA

| ZIP CODE | QUARTER 1 | QUARTER 2 | QUARTER 3 | QUARTER 4 | TOTAL GRAMS |
|---|---|---|---|---|---|
| 997 | 0 | 0 | 5 | 0 | 5 |
| 998 | 0 | 0 | 30 | 25 | 55 |
| TOTAL | 0 | 0 | 35 | 25 | 60 |

STATE: ARIZONA

| ZIP CODE | QUARTER 1 | QUARTER 2 | QUARTER 3 | QUARTER 4 | TOTAL GRAMS |
|---|---|---|---|---|---|
| 850 | 0 | 0 | 5 | 0 | 5 |
| 852 | 0 | 150 | 335 | 0 | 485 |
| 853 | 0 | 15 | 20 | 5 | 40 |
| 857 | 0 | 0 | 20 | 0 | 20 |
| 860 | 0 | 0 | 5 | 0 | 5 |
| 863 | 0 | 10 | 10 | 5 | 25 |
| TOTAL | 0 | 175 | 395 | 10 | 580 |

STATE: ARKANSAS

| ZIP CODE | QUARTER 1 | QUARTER 2 | QUARTER 3 | QUARTER 4 | TOTAL GRAMS |
|---|---|---|---|---|---|
| 716 | 0 | 0 | 5 | 0 | 5 |
| TOTAL | 0 | 0 | 5 | 0 | 5 |

STATE: CALIFORNIA

| ZIP CODE | QUARTER 1 | QUARTER 2 | QUARTER 3 | QUARTER 4 | TOTAL GRAMS |
|---|---|---|---|---|---|
| 900 | 0 | 5 | 0 | 15 | 20 |
| 902 | 0 | 5 | 70 | 5 | 80 |
| 903 | 0 | 3 | 0 | 5 | 8 |
| 905 | 0 | 0 | 5 | 0 | 5 |
| 906 | 0 | 0 | 5 | 0 | 5 |
| 907 | 0 | 0 | 15 | 0 | 15 |
| 908 | 0 | 10 | 15 | 0 | 25 |
| 910 | 0 | 0 | 10 | 0 | 10 |
| 911 | 0 | 5 | 0 | 0 | 5 |
| 912 | 0 | 0 | 5 | 0 | 5 |
| 913 | 0 | 10 | 65 | 35 | 110 |
| 914 | 0 | 10 | 0 | 0 | 10 |
| 915 | 0 | 0 | 0 | 5 | 5 |
| 917 | 0 | 0 | 45 | 5 | 50 |
| 920 | 0 | 15 | 45 | 5 | 65 |
| 921 | 0 | 0 | 0 | 5 | 5 |
| 922 | 0 | 10 | 0 | 0 | 10 |
| 923 | 0 | 5 | 15 | 5 | 25 |
| 925 | 0 | 15 | 15 | 20 | 50 |
| 926 | 0 | 10 | 50 | 45 | 105 |
| 927 | 0 | 0 | 0 | 5 | 5 |
| 928 | 0 | 30 | 15 | 0 | 45 |
| 930 | 0 | 10 | 5 | 5 | 20 |
| 932 | 0 | 0 | 20 | 10 | 30 |
| 933 | 0 | 0 | 5 | 5 | 10 |
| 934 | 0 | 25 | 30 | 20 | 75 |
| 935 | 0 | 5 | 25 | 0 | 30 |
| 940 | 0 | 20 | 25 | 0 | 45 |
| 945 | 0 | 20 | 15 | 10 | 45 |
| 947 | 0 | 0 | 10 | 0 | 10 |
| 948 | 0 | 0 | 0 | 5 | 5 |
| 949 | 0 | 0 | 15 | 0 | 15 |
| 951 | 0 | 0 | 5 | 10 | 15 |
| 952 | 0 | 0 | 5 | 5 | 10 |
| 953 | 0 | 0 | 5 | 0 | 5 |
| 954 | 0 | 5 | 10 | 0 | 15 |

RETAIL DRUG DISTRIBUTION BY ZIP CODE WITHIN STATE BY GRAMS WT

DRUG CODE:2100DRUG NAME:BARBITURIC ACID DERIVIATIVE OR SALT [PER
STATE:CALIFORNIA

| ZIP CODE | QUARTER 1 | QUARTER 2 | QUARTER 3 | QUARTER 4 | TOTAL GRAMS |
|---|---|---|---|---|---|
| 956 | 0 | 20 | 60 | 20 | 100 |
| 959 | 0 | 0 | 5 | 0 | 5 |
| TOTAL | 0 | 238 | 615 | 245 | 1,098 |

STATE: COLORADO

| ZIP CODE | QUARTER 1 | QUARTER 2 | QUARTER 3 | QUARTER 4 | TOTAL GRAMS |
|---|---|---|---|---|---|
| 800 | 0 | 0 | 0 | 10 | 10 |
| 801 | 0 | 0 | 25 | 15 | 40 |
| 805 | 0 | 0 | 20 | 0 | 20 |
| 809 | 0 | 0 | 5 | 0 | 5 |
| 812 | 0 | 0 | 5 | 5 | 10 |
| 815 | 0 | 0 | 5 | 0 | 5 |
| 816 | 0 | 5 | 10 | 15 | 30 |
| TOTAL | 0 | 5 | 70 | 45 | 120 |

STATE: CONNECTICUT

| ZIP CODE | QUARTER 1 | QUARTER 2 | QUARTER 3 | QUARTER 4 | TOTAL GRAMS |
|---|---|---|---|---|---|
| 60 | 0 | 10 | 5 | 5 | 20 |
| 61 | 0 | 0 | 0 | 5 | 5 |
| 64 | 0 | 5 | 20 | 20 | 45 |
| 66 | 0 | 0 | 0 | 5 | 5 |
| 67 | 0 | 0 | 0 | 10 | 10 |
| 68 | 0 | 15 | 5 | 5 | 25 |
| 69 | 0 | 5 | 0 | 0 | 5 |
| TOTAL | 0 | 35 | 30 | 50 | 115 |

STATE: DELAWARE

| ZIP CODE | QUARTER 1 | QUARTER 2 | QUARTER 3 | QUARTER 4 | TOTAL GRAMS |
|---|---|---|---|---|---|
| 199 | 0 | 0 | 0 | 15 | 15 |
| TOTAL | 0 | 0 | 0 | 15 | 15 |

STATE: DISTRICT OF COLUMBIA

| ZIP CODE | QUARTER 1 | QUARTER 2 | QUARTER 3 | QUARTER 4 | TOTAL GRAMS |
|---|---|---|---|---|---|
| 200 | 0 | 5 | 10 | 20 | 35 |
| TOTAL | 0 | 5 | 10 | 20 | 35 |

STATE: FLORIDA

| ZIP CODE | QUARTER 1 | QUARTER 2 | QUARTER 3 | QUARTER 4 | TOTAL GRAMS |
|---|---|---|---|---|---|
| 320 | 0 | 5 | 25 | 15 | 45 |
| 322 | 0 | 0 | 10 | 0 | 10 |
| 325 | 0 | 0 | 0 | 5 | 5 |
| 327 | 0 | 0 | 5 | 0 | 5 |
| 328 | 0 | 0 | 0 | 5 | 5 |
| 329 | 0 | 0 | 0 | 10 | 10 |
| 330 | 0 | 0 | 10 | 15 | 25 |
| 331 | 0 | 0 | 5 | 5 | 10 |
| 333 | 0 | 5 | 5 | 0 | 10 |
| 334 | 0 | 0 | 20 | 20 | 40 |
| 335 | 0 | 0 | 25 | 10 | 35 |
| 336 | 0 | 10 | 0 | 20 | 30 |
| 337 | 0 | 5 | 5 | 5 | 15 |
| 338 | 0 | 0 | 5 | 5 | 10 |
| 339 | 0 | 0 | 10 | 0 | 10 |
| 342 | 0 | 0 | 10 | 5 | 15 |
| 344 | 0 | 0 | 10 | 0 | 10 |
| 346 | 0 | 0 | 10 | 0 | 10 |
| 347 | 0 | 0 | 0 | 15 | 15 |

| | | | | | |
|---|---|---|---|---|---|
| TOTAL | 0 | 25 | 155 | 135 | 315 |

STATE: GEORGIA

| ZIP CODE | QUARTER 1 | QUARTER 2 | QUARTER 3 | QUARTER 4 | TOTAL GRAMS |
|---|---|---|---|---|---|
| 300 | 0 | 15 | 45 | 40 | 100 |
| 301 | 0 | 5 | 0 | 40 | 45 |
| 302 | 0 | 5 | 15 | 20 | 40 |
| 303 | 0 | 0 | 5 | 15 | 20 |
| 305 | 0 | 10 | 25 | 10 | 45 |
| 309 | 0 | 0 | 5 | 0 | 5 |
| 310 | 0 | 5 | 15 | 10 | 30 |
| 314 | 0 | 0 | 10 | 0 | 10 |
| 315 | 0 | 5 | 20 | 0 | 25 |
| 316 | 0 | 0 | 0 | 10 | 10 |
| 317 | 0 | 0 | 0 | 5 | 5 |

DRUG CODE:2100DRUG NAME:BARBITURIC ACID DERIVIATIVE OR SALT [PER
STATE:GEORGIA

| ZIP CODE | QUARTER 1 | QUARTER 2 | QUARTER 3 | QUARTER 4 | TOTAL GRAMS |
|---|---|---|---|---|---|
| TOTAL | 0 | 45 | 140 | 150 | 335 |

STATE: IDAHO

| ZIP CODE | QUARTER 1 | QUARTER 2 | QUARTER 3 | QUARTER 4 | TOTAL GRAMS |
|---|---|---|---|---|---|
| 838 | 0 | 0 | 0 | 10 | 10 |
| TOTAL | 0 | 0 | 0 | 10 | 10 |

STATE: ILLINOIS

| ZIP CODE | QUARTER 1 | QUARTER 2 | QUARTER 3 | QUARTER 4 | TOTAL GRAMS |
|---|---|---|---|---|---|
| 600 | 0 | 10 | 110 | 5 | 125 |
| 601 | 0 | 0 | 30 | 30 | 60 |
| 603 | 0 | 0 | 0 | 5 | 5 |
| 604 | 0 | 0 | 5 | 15 | 20 |
| 605 | 0 | 0 | 15 | 5 | 20 |
| 606 | 0 | 0 | 10 | 5 | 15 |
| 617 | 0 | 0 | 0 | 10 | 10 |
| 618 | 0 | 0 | 15 | 0 | 15 |
| TOTAL | 0 | 10 | 185 | 75 | 270 |

STATE: INDIANA

| ZIP CODE | QUARTER 1 | QUARTER 2 | QUARTER 3 | QUARTER 4 | TOTAL GRAMS |
|---|---|---|---|---|---|
| 461 | 0 | 0 | 5 | 25 | 30 |
| 462 | 0 | 0 | 0 | 10 | 10 |
| 463 | 0 | 0 | 0 | 5 | 5 |
| 465 | 0 | 0 | 15 | 0 | 15 |
| 466 | 0 | 0 | 15 | 0 | 15 |
| 467 | 0 | 0 | 5 | 0 | 5 |
| 468 | 0 | 5 | 25 | 20 | 50 |
| 470 | 0 | 0 | 5 | 15 | 20 |
| 471 | 0 | 0 | 20 | 5 | 25 |
| TOTAL | 0 | 5 | 90 | 80 | 175 |

STATE: KANSAS

| ZIP CODE | QUARTER 1 | QUARTER 2 | QUARTER 3 | QUARTER 4 | TOTAL GRAMS |
|---|---|---|---|---|---|
| 661 | 0 | 0 | 5 | 0 | 5 |
| 662 | 0 | 0 | 10 | 5 | 15 |
| 666 | 0 | 0 | 5 | 5 | 10 |
| 672 | 0 | 0 | 35 | 15 | 50 |
| 674 | 0 | 0 | 5 | 0 | 5 |
| 677 | 0 | 0 | 5 | 0 | 5 |
| 678 | 0 | 0 | 5 | 0 | 5 |

| | | | | | |
|---|---|---|---|---|---|
| TOTAL | 0 | 0 | 70 | 25 | 95 |

STATE: KENTUCKY

| ZIP CODE | QUARTER 1 | QUARTER 2 | QUARTER 3 | QUARTER 4 | TOTAL GRAMS |
|---|---|---|---|---|---|
| 402 | 0 | 5 | 0 | 25 | 30 |
| 403 | 0 | 0 | 0 | 10 | 10 |
| 405 | 0 | 0 | 5 | 0 | 5 |
| 410 | 0 | 0 | 15 | 5 | 20 |
| 418 | 0 | 0 | 5 | 0 | 5 |
| 427 | 0 | 5 | 0 | 10 | 15 |
| TOTAL | 0 | 10 | 25 | 50 | 85 |

STATE: LOUISIANA

| ZIP CODE | QUARTER 1 | QUARTER 2 | QUARTER 3 | QUARTER 4 | TOTAL GRAMS |
|---|---|---|---|---|---|
| 700 | 0 | 0 | 0 | 5 | 5 |
| 701 | 0 | 0 | 5 | 0 | 5 |
| 704 | 0 | 0 | 5 | 15 | 20 |
| 705 | 0 | 0 | 5 | 5 | 10 |
| 708 | 0 | 0 | 5 | 0 | 5 |
| 711 | 0 | 0 | 10 | 0 | 10 |
| 712 | 0 | 0 | 0 | 5 | 5 |
| TOTAL | 0 | 0 | 30 | 30 | 60 |

STATE: MAINE

| ZIP CODE | QUARTER 1 | QUARTER 2 | QUARTER 3 | QUARTER 4 | TOTAL GRAMS |
|---|---|---|---|---|---|
| 40 | 0 | 0 | 0 | 10 | 10 |
| 42 | 0 | 0 | 0 | 5 | 5 |
| 43 | 0 | 0 | 10 | 0 | 10 |
| TOTAL | 0 | 0 | 10 | 15 | 25 |

STATE: MARYLAND

| ZIP CODE | QUARTER 1 | QUARTER 2 | QUARTER 3 | QUARTER 4 | TOTAL GRAMS |
|---|---|---|---|---|---|
| 208 | 0 | 20 | 20 | 15 | 55 |

DRUG CODE:2100DRUG NAME:BARBITURIC ACID DERIVIATIVE OR SALT [PER

STATE:MARYLAND

| ZIP CODE | QUARTER 1 | QUARTER 2 | QUARTER 3 | QUARTER 4 | TOTAL GRAMS |
|---|---|---|---|---|---|
| 210 | 0 | 5 | 5 | 0 | 10 |
| 212 | 0 | 0 | 5 | 10 | 15 |
| 215 | 0 | 0 | 0 | 5 | 5 |
| 217 | 0 | 0 | 0 | 5 | 5 |
| 218 | 0 | 5 | 0 | 10 | 15 |
| TOTAL | 0 | 30 | 30 | 45 | 105 |

STATE: MASSACHUSETTS

| ZIP CODE | QUARTER 1 | QUARTER 2 | QUARTER 3 | QUARTER 4 | TOTAL GRAMS |
|---|---|---|---|---|---|
| 11 | 0 | 0 | 5 | 0 | 5 |
| 15 | 0 | 0 | 15 | 0 | 15 |
| 18 | 0 | 0 | 5 | 0 | 5 |
| 19 | 0 | 0 | 0 | 15 | 15 |
| 20 | 0 | 5 | 0 | 5 | 10 |
| 21 | 0 | 0 | 0 | 5 | 5 |
| 23 | 0 | 0 | 0 | 5 | 5 |
| 24 | 0 | 0 | 20 | 0 | 20 |
| TOTAL | 0 | 5 | 45 | 30 | 80 |

STATE: MICHIGAN

| ZIP CODE | QUARTER 1 | QUARTER 2 | QUARTER 3 | QUARTER 4 | TOTAL GRAMS |
|---|---|---|---|---|---|

| | | | | | |
|---|---|---|---|---|---|
| 480 | 0 | 0 | 20 | 25 | 45 |
| 481 | 0 | 0 | 20 | 0 | 20 |
| 483 | 0 | 0 | 10 | 20 | 30 |
| 484 | 0 | 20 | 10 | 25 | 55 |
| 485 | 0 | 0 | 0 | 5 | 5 |
| 486 | 0 | 5 | 0 | 0 | 5 |
| 487 | 0 | 0 | 10 | 0 | 10 |
| 488 | 0 | 30 | 25 | 5 | 60 |
| 493 | 0 | 0 | 10 | 0 | 10 |
| 494 | 0 | 0 | 10 | 5 | 15 |
| 495 | 0 | 0 | 5 | 0 | 5 |
| 496 | 0 | 5 | 10 | 0 | 15 |
| TOTAL | 0 | 60 | 130 | 85 | 275 |

STATE: MINNESOTA

| ZIP CODE | QUARTER 1 | QUARTER 2 | QUARTER 3 | QUARTER 4 | TOTAL GRAMS |
|---|---|---|---|---|---|
| 551 | 0 | 0 | 20 | 5 | 25 |
| 553 | 0 | 0 | 0 | 25 | 25 |
| TOTAL | 0 | 0 | 20 | 30 | 50 |

STATE: MISSISSIPPI

| ZIP CODE | QUARTER 1 | QUARTER 2 | QUARTER 3 | QUARTER 4 | TOTAL GRAMS |
|---|---|---|---|---|---|
| 386 | 0 | 0 | 5 | 0 | 5 |
| 390 | 0 | 0 | 5 | 5 | 10 |
| 391 | 0 | 0 | 5 | 0 | 5 |
| 394 | 0 | 0 | 5 | 0 | 5 |
| 395 | 0 | 0 | 10 | 0 | 10 |
| TOTAL | 0 | 0 | 30 | 5 | 35 |

STATE: MISSOURI

| ZIP CODE | QUARTER 1 | QUARTER 2 | QUARTER 3 | QUARTER 4 | TOTAL GRAMS |
|---|---|---|---|---|---|
| 631 | 0 | 0 | 0 | 60 | 60 |
| 640 | 0 | 0 | 10 | 5 | 15 |
| 641 | 0 | 0 | 10 | 0 | 10 |
| 645 | 0 | 5 | 0 | 0 | 5 |
| 653 | 0 | 0 | 20 | 0 | 20 |
| TOTAL | 0 | 5 | 40 | 65 | 110 |

STATE: MONTANA

| ZIP CODE | QUARTER 1 | QUARTER 2 | QUARTER 3 | QUARTER 4 | TOTAL GRAMS |
|---|---|---|---|---|---|
| 593 | 0 | 5 | 5 | 15 | 25 |
| TOTAL | 0 | 5 | 5 | 15 | 25 |

STATE: NEVADA

| ZIP CODE | QUARTER 1 | QUARTER 2 | QUARTER 3 | QUARTER 4 | TOTAL GRAMS |
|---|---|---|---|---|---|
| 891 | 0 | 5 | 20 | 5 | 30 |
| 895 | 0 | 10 | 5 | 0 | 15 |
| TOTAL | 0 | 15 | 25 | 5 | 45 |

STATE: NEW HAMPSHIRE

| ZIP CODE | QUARTER 1 | QUARTER 2 | QUARTER 3 | QUARTER 4 | TOTAL GRAMS |
|---|---|---|---|---|---|
| 30 | 0 | 5 | 20 | 5 | 30 |
| 32 | 0 | 0 | 5 | 0 | 5 |

DRUG CODE:2100DRUG NAME:BARBITURIC ACID DERIVIATIVE OR SALT [PER
STATE:NEW HAMPSHIRE

| ZIP CODE | QUARTER 1 | QUARTER 2 | QUARTER 3 | QUARTER 4 | TOTAL GRAMS |
|---|---|---|---|---|---|

| | | | | | |
|---|---|---|---|---|---|
| 33 | 0 | 0 | 5 | 0 | 5 |
| 34 | 0 | 0 | 0 | 5 | 5 |
| TOTAL | 0 | 5 | 30 | 10 | 45 |

STATE: NEW JERSEY

| ZIP CODE | QUARTER 1 | QUARTER 2 | QUARTER 3 | QUARTER 4 | TOTAL GRAMS |
|---|---|---|---|---|---|
| 70 | 0 | 75 | 190 | 230 | 495 |
| 71 | 0 | 0 | 10 | 10 | 20 |
| 72 | 0 | 0 | 0 | 5 | 5 |
| 74 | 0 | 0 | 40 | 25 | 65 |
| 76 | 0 | 25 | 90 | 80 | 195 |
| 77 | 0 | 0 | 5 | 20 | 25 |
| 78 | 0 | 0 | 35 | 0 | 35 |
| 79 | 0 | 0 | 20 | 25 | 45 |
| 80 | 0 | 15 | 40 | 0 | 55 |
| 82 | 0 | 5 | 15 | 5 | 25 |
| 83 | 0 | 0 | 15 | 0 | 15 |
| 85 | 0 | 5 | 15 | 0 | 20 |
| 86 | 0 | 0 | 0 | 5 | 5 |
| 87 | 0 | 0 | 5 | 0 | 5 |
| 88 | 0 | 5 | 35 | 30 | 70 |
| 89 | 0 | 0 | 10 | 0 | 10 |
| TOTAL | 0 | 130 | 525 | 435 | 1,090 |

STATE: NEW MEXICO

| ZIP CODE | QUARTER 1 | QUARTER 2 | QUARTER 3 | QUARTER 4 | TOTAL GRAMS |
|---|---|---|---|---|---|
| 875 | 0 | 0 | 10 | 0 | 10 |
| TOTAL | 0 | 0 | 10 | 0 | 10 |

STATE: NEW YORK

| ZIP CODE | QUARTER 1 | QUARTER 2 | QUARTER 3 | QUARTER 4 | TOTAL GRAMS |
|---|---|---|---|---|---|
| 100 | 0 | 17 | 75 | 75 | 167 |
| 101 | 0 | 0 | 10 | 0 | 10 |
| 103 | 0 | 0 | 10 | 5 | 15 |
| 104 | 0 | 0 | 5 | 20 | 25 |
| 105 | 0 | 0 | 0 | 5 | 5 |
| 107 | 0 | 0 | 20 | 5 | 25 |
| 109 | 0 | 5 | 5 | 5 | 15 |
| 110 | 0 | 0 | 0 | 5 | 5 |
| 112 | 0 | 0 | 55 | 30 | 85 |
| 113 | 0 | 0 | 5 | 5 | 10 |
| 114 | 0 | 5 | 0 | 20 | 25 |
| 115 | 0 | 0 | 5 | 5 | 10 |
| 116 | 0 | 0 | 0 | 5 | 5 |
| 117 | 0 | 0 | 15 | 40 | 55 |
| 118 | 0 | 0 | 15 | 0 | 15 |
| 119 | 0 | 5 | 30 | 25 | 60 |
| 121 | 0 | 0 | 5 | 5 | 10 |
| 125 | 0 | 0 | 0 | 5 | 5 |
| 128 | 0 | 5 | 0 | 0 | 5 |
| 131 | 0 | 5 | 15 | 35 | 55 |
| 134 | 0 | 5 | 5 | 0 | 10 |
| 139 | 0 | 0 | 0 | 10 | 10 |
| 141 | 0 | 5 | 15 | 0 | 20 |
| 144 | 0 | 5 | 30 | 25 | 60 |
| TOTAL | 0 | 57 | 320 | 330 | 707 |

STATE: NORTH CAROLINA

| ZIP CODE | QUARTER 1 | QUARTER 2 | QUARTER 3 | QUARTER 4 | TOTAL GRAMS |
|---|---|---|---|---|---|
| 271 | 0 | 0 | 20 | 15 | 35 |
| 275 | 0 | 5 | 15 | 15 | 35 |

| | | | | | |
|---|---|---|---|---|---|
| 276 | 0 | 0 | 5 | 0 | 5 |
| 277 | 0 | 0 | 5 | 10 | 15 |
| 278 | 0 | 0 | 0 | 10 | 10 |
| 279 | 0 | 0 | 35 | 45 | 80 |
| 280 | 0 | 5 | 5 | 5 | 15 |
| 282 | 0 | 15 | 20 | 10 | 45 |
| 283 | 0 | 0 | 10 | 0 | 10 |
| 284 | 0 | 0 | 5 | 5 | 10 |
| 285 | 0 | 5 | 15 | 20 | 40 |

DRUG CODE:2100DRUG NAME:BARBITURIC ACID DERIVIATIVE OR SALT [PER
STATE:NORTH CAROLINA

| ZIP CODE | QUARTER 1 | QUARTER 2 | QUARTER 3 | QUARTER 4 | TOTAL GRAMS |
|---|---|---|---|---|---|
| TOTAL | 0 | 30 | 135 | 135 | 300 |

STATE: OHIO

| ZIP CODE | QUARTER 1 | QUARTER 2 | QUARTER 3 | QUARTER 4 | TOTAL GRAMS |
|---|---|---|---|---|---|
| 430 | 0 | 0 | 10 | 20 | 30 |
| 431 | 0 | 0 | 0 | 20 | 20 |
| 432 | 0 | 0 | 5 | 65 | 70 |
| 433 | 0 | 0 | 10 | 0 | 10 |
| 436 | 0 | 5 | 0 | 5 | 10 |
| 440 | 0 | 0 | 5 | 5 | 10 |
| 441 | 0 | 0 | 5 | 0 | 5 |
| 444 | 0 | 0 | 45 | 20 | 65 |
| 450 | 0 | 0 | 10 | 5 | 15 |
| 452 | 0 | 0 | 5 | 5 | 10 |
| 453 | 0 | 0 | 5 | 0 | 5 |
| 454 | 0 | 0 | 0 | 5 | 5 |
| 455 | 0 | 10 | 5 | 0 | 15 |
| 457 | 0 | 0 | 5 | 0 | 5 |
| TOTAL | 0 | 15 | 110 | 150 | 275 |

STATE: OKLAHOMA

| ZIP CODE | QUARTER 1 | QUARTER 2 | QUARTER 3 | QUARTER 4 | TOTAL GRAMS |
|---|---|---|---|---|---|
| 731 | 0 | 0 | 5 | 0 | 5 |
| 737 | 0 | 0 | 15 | 0 | 15 |
| 740 | 0 | 0 | 5 | 0 | 5 |
| 741 | 0 | 5 | 10 | 25 | 40 |
| TOTAL | 0 | 5 | 35 | 25 | 65 |

STATE: OREGON

| ZIP CODE | QUARTER 1 | QUARTER 2 | QUARTER 3 | QUARTER 4 | TOTAL GRAMS |
|---|---|---|---|---|---|
| 970 | 0 | 0 | 0 | 10 | 10 |
| 971 | 0 | 0 | 5 | 0 | 5 |
| 972 | 0 | 0 | 5 | 0 | 5 |
| 973 | 0 | 0 | 5 | 0 | 5 |
| TOTAL | 0 | 0 | 15 | 10 | 25 |

STATE: PENNSYLVANIA

| ZIP CODE | QUARTER 1 | QUARTER 2 | QUARTER 3 | QUARTER 4 | TOTAL GRAMS |
|---|---|---|---|---|---|
| 165 | 0 | 0 | 0 | 5 | 5 |
| 169 | 0 | 0 | 0 | 5 | 5 |
| 170 | 0 | 0 | 15 | 10 | 25 |
| 176 | 0 | 0 | 0 | 10 | 10 |
| 177 | 0 | 0 | 15 | 20 | 35 |
| 180 | 0 | 0 | 0 | 15 | 15 |
| 182 | 0 | 0 | 0 | 5 | 5 |
| 184 | 0 | 0 | 10 | 5 | 15 |
| 187 | 0 | 10 | 15 | 20 | 45 |

| | | | | | |
|---|---|---|---|---|---|
| 189 | 0 | 0 | 20 | 20 | 40 |
| 190 | 0 | 0 | 20 | 40 | 60 |
| 191 | 0 | 0 | 20 | 5 | 25 |
| 193 | 0 | 5 | 0 | 5 | 10 |
| 194 | 0 | 0 | 15 | 5 | 20 |
| 195 | 0 | 0 | 0 | 10 | 10 |
| TOTAL | 0 | 15 | 130 | 180 | 325 |

STATE: RHODE ISLAND

| ZIP CODE | QUARTER 1 | QUARTER 2 | QUARTER 3 | QUARTER 4 | TOTAL GRAMS |
|---|---|---|---|---|---|
| 28 | 0 | 10 | 30 | 20 | 60 |
| 29 | 0 | 0 | 15 | 10 | 25 |
| TOTAL | 0 | 10 | 45 | 30 | 85 |

STATE: SOUTH CAROLINA

| ZIP CODE | QUARTER 1 | QUARTER 2 | QUARTER 3 | QUARTER 4 | TOTAL GRAMS |
|---|---|---|---|---|---|
| 293 | 0 | 0 | 5 | 5 | 10 |
| 295 | 0 | 0 | 0 | 5 | 5 |
| 296 | 0 | 0 | 5 | 0 | 5 |
| 298 | 0 | 0 | 0 | 5 | 5 |
| 299 | 0 | 0 | 5 | 0 | 5 |
| TOTAL | 0 | 0 | 15 | 15 | 30 |

STATE: TENNESSEE

| ZIP CODE | QUARTER 1 | QUARTER 2 | QUARTER 3 | QUARTER 4 | TOTAL GRAMS |
|---|---|---|---|---|---|
| 370 | 0 | 0 | 25 | 10 | 35 |

DRUG CODE:2100DRUG NAME:BARBITURIC ACID DERIVIATIVE OR SALT [PER
STATE:TENNESSEE

| ZIP CODE | QUARTER 1 | QUARTER 2 | QUARTER 3 | QUARTER 4 | TOTAL GRAMS |
|---|---|---|---|---|---|
| 371 | 0 | 0 | 10 | 0 | 10 |
| TOTAL | 0 | 0 | 35 | 10 | 45 |

STATE: TEXAS

| ZIP CODE | QUARTER 1 | QUARTER 2 | QUARTER 3 | QUARTER 4 | TOTAL GRAMS |
|---|---|---|---|---|---|
| 750 | 0 | 5 | 20 | 25 | 50 |
| 752 | 0 | 0 | 45 | 5 | 50 |
| 754 | 0 | 0 | 5 | 0 | 5 |
| 759 | 0 | 0 | 10 | 0 | 10 |
| 760 | 0 | 10 | 30 | 5 | 45 |
| 761 | 0 | 0 | 10 | 45 | 55 |
| 762 | 0 | 0 | 5 | 0 | 5 |
| 767 | 0 | 0 | 5 | 0 | 5 |
| 770 | 0 | 0 | 10 | 0 | 10 |
| 773 | 0 | 5 | 15 | 0 | 20 |
| 774 | 0 | 0 | 35 | 20 | 55 |
| 775 | 0 | 0 | 5 | 20 | 25 |
| 776 | 0 | 10 | 20 | 25 | 55 |
| 780 | 0 | 0 | 40 | 20 | 60 |
| 782 | 0 | 0 | 40 | 5 | 45 |
| 785 | 0 | 10 | 40 | 35 | 85 |
| 787 | 0 | 0 | 10 | 0 | 10 |
| 793 | 0 | 0 | 10 | 0 | 10 |
| 794 | 0 | 0 | 10 | 5 | 15 |
| 797 | 0 | 0 | 10 | 20 | 30 |
| TOTAL | 0 | 40 | 375 | 230 | 645 |

STATE: VERMONT

| ZIP CODE | QUARTER 1 | QUARTER 2 | QUARTER 3 | QUARTER 4 | TOTAL GRAMS |
|---|---|---|---|---|---|

| | | | | | |
|---|---|---|---|---|---|
| 54 | 0 | 0 | 5 | 0 | 5 |
| TOTAL | 0 | 0 | 5 | 0 | 5 |

STATE: VIRGINIA

| ZIP CODE | QUARTER 1 | QUARTER 2 | QUARTER 3 | QUARTER 4 | TOTAL GRAMS |
|---|---|---|---|---|---|
| 201 | 0 | 0 | 5 | 5 | 10 |
| 220 | 0 | 15 | 25 | 50 | 90 |
| 221 | 0 | 0 | 5 | 15 | 20 |
| 222 | 0 | 10 | 5 | 0 | 15 |
| 223 | 0 | 0 | 10 | 5 | 15 |
| 225 | 0 | 0 | 5 | 0 | 5 |
| 226 | 0 | 0 | 0 | 10 | 10 |
| 227 | 0 | 0 | 0 | 5 | 5 |
| 231 | 0 | 0 | 5 | 0 | 5 |
| 233 | 0 | 0 | 5 | 0 | 5 |
| 234 | 0 | 0 | 5 | 0 | 5 |
| 235 | 0 | 10 | 20 | 10 | 40 |
| 236 | 0 | 0 | 10 | 5 | 15 |
| 239 | 0 | 0 | 0 | 5 | 5 |
| 240 | 0 | 10 | 10 | 0 | 20 |
| 245 | 0 | 0 | 0 | 25 | 25 |
| TOTAL | 0 | 45 | 110 | 135 | 290 |

STATE: WASHINGTON

| ZIP CODE | QUARTER 1 | QUARTER 2 | QUARTER 3 | QUARTER 4 | TOTAL GRAMS |
|---|---|---|---|---|---|
| 982 | 0 | 0 | 5 | 15 | 20 |
| 993 | 0 | 0 | 5 | 10 | 15 |
| TOTAL | 0 | 0 | 10 | 25 | 35 |

STATE: WEST VIRGINIA

| ZIP CODE | QUARTER 1 | QUARTER 2 | QUARTER 3 | QUARTER 4 | TOTAL GRAMS |
|---|---|---|---|---|---|
| 254 | 0 | 0 | 0 | 40 | 40 |
| TOTAL | 0 | 0 | 0 | 40 | 40 |

STATE: WISCONSIN

| ZIP CODE | QUARTER 1 | QUARTER 2 | QUARTER 3 | QUARTER 4 | TOTAL GRAMS |
|---|---|---|---|---|---|
| 530 | 0 | 0 | 0 | 15 | 15 |
| 531 | 0 | 0 | 5 | 0 | 5 |
| TOTAL | 0 | 0 | 5 | 15 | 20 |

DRUG CODE:2125DRUG NAME:AMOBARBITAL (SCHEDULE 2)
STATE: ALABAMA

| ZIP CODE | QUARTER 1 | QUARTER 2 | QUARTER 3 | QUARTER 4 | TOTAL GRAMS |
|---|---|---|---|---|---|
| 352 | 5.47 | 0 | 0 | 0 | 5.47 |

DRUG CODE:2125DRUG NAME:AMOBARBITAL (SCHEDULE 2)
STATE:ALABAMA

| ZIP CODE | QUARTER 1 | QUARTER 2 | QUARTER 3 | QUARTER 4 | TOTAL GRAMS |
|---|---|---|---|---|---|
| TOTAL | 5.47 | 0 | 0 | 0 | 5.47 |

STATE: ARIZONA

| ZIP CODE | QUARTER 1 | QUARTER 2 | QUARTER 3 | QUARTER 4 | TOTAL GRAMS |
|---|---|---|---|---|---|
| 850 | 10.48 | 11.85 | 10.03 | 1.82 | 34.18 |
| TOTAL | 10.48 | 11.85 | 10.03 | 1.82 | 34.18 |

STATE: ARKANSAS

| ZIP CODE | QUARTER 1 | QUARTER 2 | QUARTER 3 | QUARTER 4 | TOTAL GRAMS |
|---|---|---|---|---|---|
| 722 | 0.46 | 0 | 0 | 0 | 0.46 |

| | | | | | |
|---|---|---|---|---|---|
| TOTAL | 0.46 | 0 | 0 | 0 | 0.46 |
| STATE: | CALIFORNIA | | | | |
| ZIP CODE | QUARTER 1 | QUARTER 2 | QUARTER 3 | QUARTER 4 | TOTAL GRAMS |
| 900 | 22.79 | 7.75 | 10.03 | 1.37 | 41.94 |
| 910 | 0 | 0 | 0 | 0.46 | 0.46 |
| 911 | 0 | 1.82 | 0 | 0 | 1.82 |
| 915 | 0.91 | 1.82 | 0.46 | 0 | 3.19 |
| 920 | 1.37 | 1.37 | 0 | 0 | 2.74 |
| 921 | 0 | 3.19 | 0 | 0 | 3.19 |
| 923 | 1.82 | 2.73 | 0.46 | 0.91 | 5.92 |
| 926 | 1.37 | 1.37 | 0.91 | 0.46 | 4.11 |
| 928 | 6.38 | 1.37 | 0.91 | 0 | 8.66 |
| 941 | 9.11 | 5.92 | 0.46 | 4.1 | 19.59 |
| 958 | 7.75 | 1.37 | 1.82 | 0.46 | 11.4 |
| TOTAL | 51.5 | 28.71 | 15.05 | 7.76 | 103.02 |
| STATE: | COLORADO | | | | |
| ZIP CODE | QUARTER 1 | QUARTER 2 | QUARTER 3 | QUARTER 4 | TOTAL GRAMS |
| 800 | 15.95 | 13.67 | 9.11 | 0 | 38.73 |
| 801 | 5.92 | 5.47 | 0 | 0 | 11.39 |
| 802 | 4.56 | 7.29 | 2.28 | 1.82 | 15.95 |
| TOTAL | 26.43 | 26.43 | 11.39 | 1.82 | 66.07 |
| STATE: | CONNECTICUT | | | | |
| ZIP CODE | QUARTER 1 | QUARTER 2 | QUARTER 3 | QUARTER 4 | TOTAL GRAMS |
| 61 | 0.46 | 0.46 | 0 | 0 | 0.92 |
| TOTAL | 0.46 | 0.46 | 0 | 0 | 0.92 |
| STATE: | DISTRICT OF COLUMBIA | | | | |
| ZIP CODE | QUARTER 1 | QUARTER 2 | QUARTER 3 | QUARTER 4 | TOTAL GRAMS |
| 200 | 0 | 2.28 | 0 | 0 | 2.28 |
| TOTAL | 0 | 2.28 | 0 | 0 | 2.28 |
| STATE: | FLORIDA | | | | |
| ZIP CODE | QUARTER 1 | QUARTER 2 | QUARTER 3 | QUARTER 4 | TOTAL GRAMS |
| 322 | 3.65 | 5.92 | 0 | 0 | 9.57 |
| 325 | 0 | 0 | 0.46 | 1.37 | 1.83 |
| 328 | 0.91 | 4.1 | 0.46 | 7.29 | 12.76 |
| 330 | 1.37 | 2.28 | 0 | 0 | 3.65 |
| 331 | 0 | 0.91 | 0 | 0 | 0.91 |
| TOTAL | 5.93 | 13.21 | 0.92 | 8.66 | 28.72 |
| STATE: | GEORGIA | | | | |
| ZIP CODE | QUARTER 1 | QUARTER 2 | QUARTER 3 | QUARTER 4 | TOTAL GRAMS |
| 301 | 0.46 | 0 | 0 | 0 | 0.46 |
| 303 | 5.47 | 3.65 | 0.91 | 0.91 | 10.94 |
| 309 | 4.56 | 12.3 | 5.92 | 0.46 | 23.24 |
| TOTAL | 10.49 | 15.95 | 6.83 | 1.37 | 34.64 |
| STATE: | IDAHO | | | | |
| ZIP CODE | QUARTER 1 | QUARTER 2 | QUARTER 3 | QUARTER 4 | TOTAL GRAMS |
| 837 | 0.46 | 0 | 0 | 0 | 0.46 |
| TOTAL | 0.46 | 0 | 0 | 0 | 0.46 |
| STATE: | ILLINOIS | | | | |
| ZIP CODE | QUARTER 1 | QUARTER 2 | QUARTER 3 | QUARTER 4 | TOTAL GRAMS |
| 600 | 0 | 0.46 | 0 | 0 | 0.46 |
| 601 | 0.46 | 0 | 0.46 | 0 | 0.92 |
| 602 | 0 | 0 | 0.91 | 0.91 | 1.82 |
| 604 | 0.91 | 0 | 0.46 | 0 | 1.37 |

| | | | | | |
|---|---|---|---|---|---|
| 606 | 6.38 | 10.03 | 5.01 | 0 | 21.42 |
| 617 | 0.91 | 0 | 0 | 0 | 0.91 |
| 618 | 0 | 0 | 0.46 | 0.46 | 0.92 |
| 627 | 2.28 | 0.46 | 0 | 0 | 2.74 |
| TOTAL | 10.94 | 10.95 | 7.3 | 1.37 | 30.56 |

DRUG CODE:2125DRUG NAME:AMOBARBITAL (SCHEDULE 2)
STATE: INDIANA

| ZIP CODE | QUARTER 1 | QUARTER 2 | QUARTER 3 | QUARTER 4 | TOTAL GRAMS |
|---|---|---|---|---|---|
| 462 | 6.38 | 1.37 | 0 | 0 | 7.75 |
| TOTAL | 6.38 | 1.37 | 0 | 0 | 7.75 |

STATE: IOWA

| ZIP CODE | QUARTER 1 | QUARTER 2 | QUARTER 3 | QUARTER 4 | TOTAL GRAMS |
|---|---|---|---|---|---|
| 522 | 0 | 0.46 | 0 | 0 | 0.46 |
| TOTAL | 0 | 0.46 | 0 | 0 | 0.46 |

STATE: KANSAS

| ZIP CODE | QUARTER 1 | QUARTER 2 | QUARTER 3 | QUARTER 4 | TOTAL GRAMS |
|---|---|---|---|---|---|
| 661 | 10.03 | 5.01 | 0.91 | 0 | 15.95 |
| 672 | 0.46 | 1.37 | 0 | 0 | 1.83 |
| TOTAL | 10.49 | 6.38 | 0.91 | 0 | 17.78 |

STATE: KENTUCKY

| ZIP CODE | QUARTER 1 | QUARTER 2 | QUARTER 3 | QUARTER 4 | TOTAL GRAMS |
|---|---|---|---|---|---|
| 402 | 3.65 | 1.82 | 4.1 | 0.46 | 10.03 |
| 405 | 3.19 | 0.91 | 0.91 | 0 | 5.01 |
| TOTAL | 6.84 | 2.73 | 5.01 | 0.46 | 15.04 |

STATE: MAINE

| ZIP CODE | QUARTER 1 | QUARTER 2 | QUARTER 3 | QUARTER 4 | TOTAL GRAMS |
|---|---|---|---|---|---|
| 41 | 1.37 | 1.37 | 0.91 | 0 | 3.65 |
| TOTAL | 1.37 | 1.37 | 0.91 | 0 | 3.65 |

STATE: MARYLAND

| ZIP CODE | QUARTER 1 | QUARTER 2 | QUARTER 3 | QUARTER 4 | TOTAL GRAMS |
|---|---|---|---|---|---|
| 208 | 3.19 | 0 | 0 | 0 | 3.19 |
| 209 | 0 | 0 | 0.91 | 0 | 0.91 |
| 212 | 5.01 | 7.29 | 0.91 | 2.73 | 15.94 |
| TOTAL | 8.2 | 7.29 | 1.82 | 2.73 | 20.04 |

STATE: MASSACHUSETTS

| ZIP CODE | QUARTER 1 | QUARTER 2 | QUARTER 3 | QUARTER 4 | TOTAL GRAMS |
|---|---|---|---|---|---|
| 11 | 0.46 | 0 | 0 | 0 | 0.46 |
| 16 | 3.19 | 0 | 0.91 | 0.91 | 5.01 |
| 18 | 0.46 | 0 | 0 | 0 | 0.46 |
| 21 | 1.82 | 14.13 | 3.19 | 0.46 | 19.6 |
| 22 | 1.82 | 1.37 | 0.91 | 0 | 4.1 |
| TOTAL | 7.75 | 15.5 | 5.01 | 1.37 | 29.63 |

STATE: MICHIGAN

| ZIP CODE | QUARTER 1 | QUARTER 2 | QUARTER 3 | QUARTER 4 | TOTAL GRAMS |
|---|---|---|---|---|---|
| 480 | 0 | 1.82 | 0 | 0 | 1.82 |
| 481 | 0 | 0.46 | 0 | 0 | 0.46 |
| 482 | 5.47 | 2.28 | 3.65 | 0.91 | 12.31 |
| 483 | 1.37 | 0 | 0 | 0 | 1.37 |
| 490 | 0.46 | 0 | 0 | 0 | 0.46 |
| 495 | 1.37 | 1.82 | 0 | 0 | 3.19 |

| | | | | | |
|---|---|---|---|---|---|
| TOTAL | 8.67 | 6.38 | 3.65 | 0.91 | 19.61 |
| STATE: | MINNESOTA | | | | |
| ZIP CODE | QUARTER 1 | QUARTER 2 | QUARTER 3 | QUARTER 4 | TOTAL GRAMS |
| 551 | 0 | 5.92 | 0 | 0.46 | 6.38 |
| 554 | 4.56 | 4.56 | 0.91 | 1.37 | 11.4 |
| 559 | 0.46 | 5.47 | 0 | 0 | 5.93 |
| TOTAL | 5.02 | 15.95 | 0.91 | 1.83 | 23.71 |
| STATE: | MISSISSIPPI | | | | |
| ZIP CODE | QUARTER 1 | QUARTER 2 | QUARTER 3 | QUARTER 4 | TOTAL GRAMS |
| 392 | 3.65 | 0.91 | 1.37 | 0 | 5.93 |
| TOTAL | 3.65 | 0.91 | 1.37 | 0 | 5.93 |
| STATE: | MISSOURI | | | | |
| ZIP CODE | QUARTER 1 | QUARTER 2 | QUARTER 3 | QUARTER 4 | TOTAL GRAMS |
| 631 | 4.56 | 5.47 | 1.82 | 0.46 | 12.31 |
| 641 | 1.82 | 0 | 0 | 0.46 | 2.28 |
| 652 | 0 | 0 | 2.28 | 0 | 2.28 |
| TOTAL | 6.38 | 5.47 | 4.1 | 0.92 | 16.87 |
| STATE: | NEBRASKA | | | | |
| ZIP CODE | QUARTER 1 | QUARTER 2 | QUARTER 3 | QUARTER 4 | TOTAL GRAMS |
| 681 | 0.91 | 5.92 | 4.1 | 1.37 | 12.3 |
| TOTAL | 0.91 | 5.92 | 4.1 | 1.37 | 12.3 |
| STATE: | NEVADA | | | | |
| ZIP CODE | QUARTER 1 | QUARTER 2 | QUARTER 3 | QUARTER 4 | TOTAL GRAMS |
| 891 | 2.28 | 0 | 0.91 | 0 | 3.19 |

DRUG CODE:2125DRUG NAME:AMOBARBITAL (SCHEDULE 2)
STATE:NEVADA

| | | | | | |
|---|---|---|---|---|---|
| ZIP CODE | QUARTER 1 | QUARTER 2 | QUARTER 3 | QUARTER 4 | TOTAL GRAMS |
| TOTAL | 2.28 | 0 | 0.91 | 0 | 3.19 |
| STATE: | NEW HAMPSHIRE | | | | |
| ZIP CODE | QUARTER 1 | QUARTER 2 | QUARTER 3 | QUARTER 4 | TOTAL GRAMS |
| 37 | 4.1 | 5.47 | 0 | 1.37 | 10.94 |
| TOTAL | 4.1 | 5.47 | 0 | 1.37 | 10.94 |
| STATE: | NEW JERSEY | | | | |
| ZIP CODE | QUARTER 1 | QUARTER 2 | QUARTER 3 | QUARTER 4 | TOTAL GRAMS |
| 70 | 0.46 | 0.46 | 3.19 | 0 | 4.11 |
| 71 | 8.2 | 4.56 | 0.91 | 0 | 13.67 |
| 74 | 0.46 | 0.46 | 0 | 0 | 0.92 |
| 76 | 4.1 | 1.37 | 0.91 | 1.82 | 8.2 |
| 79 | 2.28 | 3.19 | 0 | 0.91 | 6.38 |
| 81 | 0.91 | 0.91 | 0 | 0 | 1.82 |
| 86 | 0 | 0.46 | 0 | 0 | 0.46 |
| 89 | 0 | 4.56 | 0 | 0 | 4.56 |
| TOTAL | 16.41 | 15.97 | 5.01 | 2.73 | 40.12 |
| STATE: | NEW MEXICO | | | | |
| ZIP CODE | QUARTER 1 | QUARTER 2 | QUARTER 3 | QUARTER 4 | TOTAL GRAMS |
| 871 | 0 | 2.28 | 0.46 | 0 | 2.74 |
| TOTAL | 0 | 2.28 | 0.46 | 0 | 2.74 |
| STATE: | NEW YORK | | | | |
| ZIP CODE | QUARTER 1 | QUARTER 2 | QUARTER 3 | QUARTER 4 | TOTAL GRAMS |
| 100 | 19.6 | 9.57 | 11.39 | 0.91 | 41.47 |
| 103 | 0.91 | 0.46 | 0 | 0.91 | 2.28 |

| | | | | | |
|---|---|---|---|---|---|
| 104 | 3.65 | 2.28 | 1.37 | 0 | 7.3 |
| 105 | 0 | 2.28 | 0.91 | 0 | 3.19 |
| 106 | 0 | 0.91 | 0.91 | 0 | 1.82 |
| 110 | 2.73 | 5.01 | 4.56 | 0 | 12.3 |
| 112 | 1.82 | 3.65 | 0 | 0 | 5.47 |
| 113 | 0 | 1.37 | 0 | 0 | 1.37 |
| 117 | 9.11 | 14.58 | 2.73 | 1.82 | 28.24 |
| 122 | 4.56 | 2.28 | 2.73 | 0 | 9.57 |
| 123 | 0 | 1.37 | 0 | 0 | 1.37 |
| 126 | 0.46 | 6.84 | 0 | 0 | 7.3 |
| 142 | 8.66 | 5.92 | 2.73 | 5.47 | 22.78 |
| 146 | 5.01 | 0.46 | 2.28 | 0 | 7.75 |
| TOTAL | 56.51 | 56.98 | 29.61 | 9.11 | 152.21 |

STATE: NORTH CAROLINA

| ZIP CODE | QUARTER 1 | QUARTER 2 | QUARTER 3 | QUARTER 4 | TOTAL GRAMS |
|---|---|---|---|---|---|
| 271 | 4.56 | 3.19 | 0 | 1.82 | 9.57 |
| 275 | 2.28 | 4.1 | 0 | 1.37 | 7.75 |
| 277 | 0 | 3.19 | 0 | 0 | 3.19 |
| 280 | 2.73 | 1.37 | 0.91 | 0 | 5.01 |
| 282 | 2.73 | 7.29 | 0 | 4.56 | 14.58 |
| TOTAL | 12.3 | 19.14 | 0.91 | 7.75 | 40.1 |

STATE: OHIO

| ZIP CODE | QUARTER 1 | QUARTER 2 | QUARTER 3 | QUARTER 4 | TOTAL GRAMS |
|---|---|---|---|---|---|
| 432 | 0 | 0.46 | 0 | 0 | 0.46 |
| 436 | 0.91 | 0 | 0 | 0 | 0.91 |
| 443 | 0.46 | 0.91 | 0.46 | 0 | 1.83 |
| 452 | 5.01 | 6.38 | 2.28 | 6.38 | 20.05 |
| 454 | 0 | 1.37 | 0.91 | 0 | 2.28 |
| TOTAL | 6.38 | 9.12 | 3.65 | 6.38 | 25.53 |

STATE: OKLAHOMA

| ZIP CODE | QUARTER 1 | QUARTER 2 | QUARTER 3 | QUARTER 4 | TOTAL GRAMS |
|---|---|---|---|---|---|
| 741 | 0 | 0.46 | 0 | 0 | 0.46 |
| TOTAL | 0 | 0.46 | 0 | 0 | 0.46 |

STATE: OREGON

| ZIP CODE | QUARTER 1 | QUARTER 2 | QUARTER 3 | QUARTER 4 | TOTAL GRAMS |
|---|---|---|---|---|---|
| 972 | 2.73 | 2.73 | 2.73 | 2.28 | 10.47 |
| TOTAL | 2.73 | 2.73 | 2.73 | 2.28 | 10.47 |

STATE: PENNSYLVANIA

| ZIP CODE | QUARTER 1 | QUARTER 2 | QUARTER 3 | QUARTER 4 | TOTAL GRAMS |
|---|---|---|---|---|---|
| 152 | 5.92 | 0 | 0 | 0 | 5.92 |
| 174 | 0.91 | 2.28 | 0.46 | 1.82 | 5.47 |
| 178 | 0 | 5.47 | 1.82 | 1.82 | 9.11 |

DRUG CODE:2125DRUG NAME:AMOBARBITAL (SCHEDULE 2)
STATE:PENNSYLVANIA

| ZIP CODE | QUARTER 1 | QUARTER 2 | QUARTER 3 | QUARTER 4 | TOTAL GRAMS |
|---|---|---|---|---|---|
| 181 | 0 | 0.46 | 0 | 0 | 0.46 |
| 187 | 0 | 2.73 | 0 | 0 | 2.73 |
| 191 | 8.2 | 4.1 | 6.84 | 0 | 19.14 |
| TOTAL | 15.03 | 15.04 | 9.12 | 3.64 | 42.83 |

STATE: PUERTO RICO

| ZIP CODE | QUARTER 1 | QUARTER 2 | QUARTER 3 | QUARTER 4 | TOTAL GRAMS |
|---|---|---|---|---|---|

| | | | | | |
|---|---|---|---|---|---|
| 7 | 0 | 0 | 0.91 | 0 | 0.91 |
| TOTAL | 0 | 0 | 0.91 | 0 | 0.91 |

STATE: SOUTH CAROLINA

| ZIP CODE | QUARTER 1 | QUARTER 2 | QUARTER 3 | QUARTER 4 | TOTAL GRAMS |
|---|---|---|---|---|---|
| 292 | 1.37 | 0.91 | 0 | 0 | 2.28 |
| 294 | 5.01 | 5.47 | 4.1 | 4.1 | 18.68 |
| TOTAL | 6.38 | 6.38 | 4.1 | 4.1 | 20.96 |

STATE: TENNESSEE

| ZIP CODE | QUARTER 1 | QUARTER 2 | QUARTER 3 | QUARTER 4 | TOTAL GRAMS |
|---|---|---|---|---|---|
| 372 | 1.37 | 6.38 | 0 | 1.37 | 9.12 |
| 381 | 1.82 | 0 | 0 | 0 | 1.82 |
| TOTAL | 3.19 | 6.38 | 0 | 1.37 | 10.94 |

STATE: TEXAS

| ZIP CODE | QUARTER 1 | QUARTER 2 | QUARTER 3 | QUARTER 4 | TOTAL GRAMS |
|---|---|---|---|---|---|
| 752 | 6.84 | 2.28 | 2.28 | 2.73 | 14.13 |
| 753 | 0.46 | 0.46 | 0 | 0 | 0.92 |
| 761 | 0 | 0.46 | 0.91 | 0 | 1.37 |
| 765 | 0 | 5.01 | 0 | 2.28 | 7.29 |
| 770 | 6.84 | 4.1 | 3.65 | 2.28 | 16.87 |
| 782 | 4.1 | 6.38 | 1.37 | 2.28 | 14.13 |
| 787 | 2.28 | 9.57 | 4.1 | 0.91 | 16.86 |
| 794 | 0.91 | 0 | 0 | 0 | 0.91 |
| TOTAL | 21.43 | 28.26 | 12.31 | 10.48 | 72.48 |

STATE: UTAH

| ZIP CODE | QUARTER 1 | QUARTER 2 | QUARTER 3 | QUARTER 4 | TOTAL GRAMS |
|---|---|---|---|---|---|
| 841 | 7.29 | 8.66 | 4.1 | 2.28 | 22.33 |
| TOTAL | 7.29 | 8.66 | 4.1 | 2.28 | 22.33 |

STATE: VERMONT

| ZIP CODE | QUARTER 1 | QUARTER 2 | QUARTER 3 | QUARTER 4 | TOTAL GRAMS |
|---|---|---|---|---|---|
| 54 | 1.82 | 0 | 0 | 0 | 1.82 |
| TOTAL | 1.82 | 0 | 0 | 0 | 1.82 |

STATE: VIRGINIA

| ZIP CODE | QUARTER 1 | QUARTER 2 | QUARTER 3 | QUARTER 4 | TOTAL GRAMS |
|---|---|---|---|---|---|
| 229 | 3.65 | 0.91 | 0 | 0 | 4.56 |
| 232 | 2.28 | 1.37 | 4.56 | 0 | 8.21 |
| TOTAL | 5.93 | 2.28 | 4.56 | 0 | 12.77 |

STATE: WASHINGTON

| ZIP CODE | QUARTER 1 | QUARTER 2 | QUARTER 3 | QUARTER 4 | TOTAL GRAMS |
|---|---|---|---|---|---|
| 980 | 1.82 | 0.91 | 0.46 | 0.46 | 3.65 |
| 981 | 14.13 | 20.05 | 0.46 | 7.29 | 41.93 |
| 992 | 0 | 1.82 | 1.37 | 0 | 3.19 |
| TOTAL | 15.95 | 22.78 | 2.29 | 7.75 | 48.77 |

STATE: WEST VIRGINIA

| ZIP CODE | QUARTER 1 | QUARTER 2 | QUARTER 3 | QUARTER 4 | TOTAL GRAMS |
|---|---|---|---|---|---|
| 253 | 0 | 0.46 | 0 | 0 | 0.46 |
| 257 | 0 | 0.46 | 0 | 0 | 0.46 |
| 265 | 0 | 3.65 | 0.91 | 0 | 4.56 |
| TOTAL | 0 | 4.57 | 0.91 | 0 | 5.48 |

STATE: WISCONSIN

| ZIP CODE | QUARTER 1 | QUARTER 2 | QUARTER 3 | QUARTER 4 | TOTAL GRAMS |
|---|---|---|---|---|---|
| 532 | 6.38 | 6.38 | 0 | 1.37 | 14.13 |

| | | | | | |
|---|---|---|---|---|---|
| 537 | 7.75 | 1.82 | 0.46 | 2.73 | 12.76 |
| 549 | 0 | 0 | 0 | 0.91 | 0.91 |
| TOTAL | 14.13 | 8.2 | 0.46 | 5.01 | 27.8 |

DRUG CODE: 2165 DRUG NAME: BUTALBITAL
STATE: ALABAMA

| ZIP CODE | QUARTER 1 | QUARTER 2 | QUARTER 3 | QUARTER 4 | TOTAL GRAMS |
|---|---|---|---|---|---|
| 350 | 855 | 910 | 875 | 930 | 3,570 |
| 351 | 420 | 425 | 395 | 420 | 1,660 |
| 352 | 495 | 580 | 655 | 475 | 2,205 |

DRUG CODE:2165DRUG NAME:BUTALBITAL
STATE:ALABAMA

| ZIP CODE | QUARTER 1 | QUARTER 2 | QUARTER 3 | QUARTER 4 | TOTAL GRAMS |
|---|---|---|---|---|---|
| 354 | 515 | 415 | 420 | 470 | 1,820 |
| 355 | 270 | 240 | 280 | 220 | 1,010 |
| 356 | 445 | 425 | 345 | 535 | 1,750 |
| 357 | 325 | 295 | 265 | 225 | 1,110 |
| 358 | 325 | 355 | 295 | 255 | 1,230 |
| 359 | 395 | 430 | 470 | 430 | 1,725 |
| 360 | 310 | 270 | 205 | 255 | 1,040 |
| 361 | 285 | 250 | 220 | 175 | 930 |
| 362 | 340 | 295 | 300 | 165 | 1,100 |
| 363 | 480 | 345 | 390 | 390 | 1,605 |
| 364 | 120 | 130 | 135 | 130 | 515 |
| 365 | 790 | 720 | 830 | 635 | 2,975 |
| 366 | 480 | 425 | 425 | 315 | 1,645 |
| 367 | 220 | 250 | 255 | 205 | 930 |
| 368 | 500 | 365 | 335 | 395 | 1,595 |
| 369 | 0 | 10 | 5 | 5 | 20 |
| TOTAL | 7,570 | 7,135 | 7,100 | 6,630 | 28,435 |

STATE: ALASKA

| ZIP CODE | QUARTER 1 | QUARTER 2 | QUARTER 3 | QUARTER 4 | TOTAL GRAMS |
|---|---|---|---|---|---|
| 995 | 110 | 125 | 140 | 150 | 525 |
| 996 | 165 | 180 | 125 | 220 | 690 |
| 997 | 55 | 40 | 15 | 30 | 140 |
| 998 | 30 | 30 | 20 | 35 | 115 |
| 999 | 5 | 5 | 0 | 15 | 25 |
| TOTAL | 365 | 380 | 300 | 450 | 1,495 |

STATE: ARIZONA

| ZIP CODE | QUARTER 1 | QUARTER 2 | QUARTER 3 | QUARTER 4 | TOTAL GRAMS |
|---|---|---|---|---|---|
| 850 | 1,900 | 1,680 | 1,425 | 1,640 | 6,645 |
| 851 | 785 | 615 | 675 | 530 | 2,605 |
| 852 | 3,985 | 4,225 | 4,385 | 3,785 | 16,380 |
| 853 | 2,135 | 1,745 | 1,715 | 1,900 | 7,495 |
| 855 | 270 | 320 | 310 | 275 | 1,175 |
| 856 | 410 | 350 | 365 | 475 | 1,600 |
| 857 | 1,525 | 1,425 | 1,515 | 1,470 | 5,935 |
| 859 | 165 | 175 | 210 | 170 | 720 |
| 860 | 145 | 100 | 65 | 85 | 395 |
| 863 | 485 | 385 | 485 | 425 | 1,780 |
| 864 | 705 | 595 | 640 | 660 | 2,600 |
| TOTAL | 12,510 | 11,615 | 11,790 | 11,415 | 47,330 |

STATE: ARKANSAS

| ZIP CODE | QUARTER 1 | QUARTER 2 | QUARTER 3 | QUARTER 4 | TOTAL GRAMS |
|---|---|---|---|---|---|

| | | | | | |
|---|---|---|---|---|---|
| 716 | 170 | 190 | 170 | 195 | 725 |
| 717 | 120 | 165 | 170 | 205 | 660 |
| 718 | 195 | 215 | 205 | 185 | 800 |
| 719 | 410 | 350 | 450 | 415 | 1,625 |
| 720 | 635 | 640 | 650 | 645 | 2,570 |
| 721 | 525 | 555 | 540 | 405 | 2,025 |
| 722 | 300 | 410 | 415 | 410 | 1,535 |
| 723 | 275 | 220 | 225 | 255 | 975 |
| 724 | 430 | 445 | 500 | 410 | 1,785 |
| 725 | 205 | 285 | 200 | 290 | 980 |
| 726 | 100 | 155 | 125 | 210 | 590 |
| 727 | 835 | 835 | 805 | 750 | 3,225 |
| 728 | 270 | 155 | 125 | 135 | 685 |
| 729 | 360 | 430 | 375 | 340 | 1,505 |
| TOTAL | 4,830 | 5,050 | 4,955 | 4,850 | 19,685 |

STATE: CALIFORNIA

| ZIP CODE | QUARTER 1 | QUARTER 2 | QUARTER 3 | QUARTER 4 | TOTAL GRAMS |
|---|---|---|---|---|---|
| 900 | 1,225 | 1,355 | 1,190 | 1,260 | 5,030 |
| 902 | 2,050 | 1,875 | 2,045 | 1,670 | 7,640 |
| 903 | 130 | 120 | 80 | 95 | 425 |
| 904 | 220 | 260 | 205 | 235 | 920 |
| 905 | 505 | 420 | 540 | 405 | 1,870 |
| 906 | 970 | 885 | 880 | 855 | 3,590 |
| 907 | 825 | 835 | 720 | 725 | 3,105 |
| 908 | 670 | 435 | 445 | 390 | 1,940 |

DRUG CODE:2165DRUG NAME:BUTALBITAL
STATE:CALIFORNIA

| ZIP CODE | QUARTER 1 | QUARTER 2 | QUARTER 3 | QUARTER 4 | TOTAL GRAMS |
|---|---|---|---|---|---|
| 910 | 310 | 360 | 340 | 310 | 1,320 |
| 911 | 185 | 225 | 160 | 150 | 720 |
| 912 | 240 | 155 | 205 | 250 | 850 |
| 913 | 2,340 | 2,205 | 2,230 | 2,125 | 8,900 |
| 914 | 505 | 590 | 450 | 330 | 1,875 |
| 915 | 175 | 170 | 185 | 185 | 715 |
| 916 | 150 | 185 | 190 | 160 | 685 |
| 917 | 1,900 | 1,785 | 1,925 | 1,630 | 7,240 |
| 918 | 135 | 120 | 95 | 105 | 455 |
| 919 | 315 | 155 | 150 | 175 | 795 |
| 920 | 1,950 | 2,155 | 2,485 | 1,665 | 8,255 |
| 921 | 770 | 715 | 675 | 710 | 2,870 |
| 922 | 1,130 | 850 | 740 | 800 | 3,520 |
| 923 | 1,500 | 1,345 | 1,435 | 1,255 | 5,535 |
| 924 | 100 | 75 | 75 | 80 | 330 |
| 925 | 1,965 | 1,920 | 2,025 | 1,885 | 7,795 |
| 926 | 2,295 | 2,325 | 2,205 | 2,180 | 9,005 |
| 927 | 440 | 400 | 415 | 425 | 1,680 |
| 928 | 1,635 | 1,565 | 1,450 | 1,180 | 5,830 |
| 930 | 1,740 | 1,680 | 1,545 | 1,545 | 6,510 |
| 931 | 325 | 290 | 310 | 295 | 1,220 |
| 932 | 935 | 875 | 875 | 815 | 3,500 |
| 933 | 750 | 560 | 725 | 765 | 2,800 |
| 934 | 960 | 950 | 845 | 905 | 3,660 |
| 935 | 905 | 780 | 760 | 685 | 3,130 |
| 936 | 540 | 620 | 520 | 520 | 2,200 |
| 937 | 635 | 565 | 565 | 555 | 2,320 |
| 939 | 815 | 830 | 710 | 740 | 3,095 |
| 940 | 700 | 630 | 610 | 595 | 2,535 |
| 941 | 325 | 325 | 275 | 285 | 1,210 |

| | | | | | |
|---|---|---|---|---|---|
| 943 | 65 | 75 | 105 | 50 | 295 |
| 944 | 95 | 80 | 70 | 85 | 330 |
| 945 | 2,955 | 3,030 | 2,670 | 2,570 | 11,225 |
| 946 | 150 | 225 | 175 | 160 | 710 |
| 947 | 85 | 85 | 95 | 60 | 325 |
| 948 | 25 | 70 | 30 | 40 | 165 |
| 949 | 715 | 720 | 670 | 605 | 2,710 |
| 950 | 965 | 890 | 785 | 890 | 3,530 |
| 951 | 750 | 715 | 630 | 620 | 2,715 |
| 952 | 690 | 565 | 655 | 520 | 2,430 |
| 953 | 1,600 | 1,700 | 1,495 | 1,475 | 6,270 |
| 954 | 460 | 435 | 530 | 420 | 1,845 |
| 955 | 200 | 185 | 250 | 210 | 845 |
| 956 | 1,845 | 1,910 | 2,010 | 1,625 | 7,390 |
| 957 | 335 | 350 | 320 | 375 | 1,380 |
| 958 | 680 | 685 | 690 | 545 | 2,600 |
| 959 | 1,200 | 1,155 | 1,195 | 1,090 | 4,640 |
| 960 | 470 | 470 | 330 | 300 | 1,570 |
| 961 | 140 | 95 | 130 | 100 | 465 |
| TOTAL | 46,690 | 45,030 | 44,115 | 40,680 | 176,515 |

STATE: COLORADO

| ZIP CODE | QUARTER 1 | QUARTER 2 | QUARTER 3 | QUARTER 4 | TOTAL GRAMS |
|---|---|---|---|---|---|
| 800 | 495 | 495 | 470 | 525 | 1,985 |
| 801 | 650 | 575 | 690 | 620 | 2,535 |
| 802 | 550 | 610 | 460 | 365 | 1,985 |
| 803 | 35 | 25 | 35 | 20 | 115 |
| 804 | 135 | 245 | 155 | 110 | 645 |
| 805 | 515 | 615 | 500 | 445 | 2,075 |
| 806 | 140 | 170 | 105 | 200 | 615 |
| 807 | 50 | 35 | 55 | 70 | 210 |
| 808 | 55 | 85 | 110 | 100 | 350 |
| 809 | 615 | 550 | 535 | 590 | 2,290 |
| 810 | 350 | 245 | 275 | 300 | 1,170 |
| 811 | 70 | 65 | 60 | 45 | 240 |
| 812 | 195 | 105 | 100 | 140 | 540 |
| 813 | 85 | 20 | 45 | 10 | 160 |

DRUG CODE:2165DRUG NAME:BUTALBITAL
STATE:COLORADO

| ZIP CODE | QUARTER 1 | QUARTER 2 | QUARTER 3 | QUARTER 4 | TOTAL GRAMS |
|---|---|---|---|---|---|
| 814 | 180 | 145 | 130 | 165 | 620 |
| 815 | 155 | 195 | 165 | 195 | 710 |
| 816 | 75 | 60 | 70 | 40 | 245 |
| TOTAL | 4,350 | 4,240 | 3,960 | 3,940 | 16,490 |

STATE: CONNECTICUT

| ZIP CODE | QUARTER 1 | QUARTER 2 | QUARTER 3 | QUARTER 4 | TOTAL GRAMS |
|---|---|---|---|---|---|
| 60 | 1,490 | 1,490 | 1,445 | 1,350 | 5,775 |
| 61 | 260 | 345 | 255 | 320 | 1,180 |
| 62 | 150 | 250 | 165 | 190 | 755 |
| 63 | 490 | 460 | 560 | 425 | 1,935 |
| 64 | 1,490 | 1,470 | 1,350 | 1,385 | 5,695 |
| 65 | 230 | 205 | 240 | 240 | 915 |
| 66 | 195 | 295 | 225 | 185 | 900 |
| 67 | 910 | 765 | 705 | 650 | 3,030 |
| 68 | 1,070 | 870 | 955 | 885 | 3,780 |
| 69 | 180 | 175 | 150 | 190 | 695 |

| | | | | | | |
|---|---|---|---|---|---|---|
| TOTAL | | 6,465 | 6,325 | 6,050 | 5,820 | 24,660 |
| STATE: | DELAWARE | | | | | |
| ZIP CODE | | QUARTER 1 | QUARTER 2 | QUARTER 3 | QUARTER 4 | TOTAL GRAMS |
| | 197 | 405 | 390 | 375 | 465 | 1,635 |
| | 198 | 280 | 230 | 310 | 275 | 1,095 |
| | 199 | 280 | 240 | 310 | 245 | 1,075 |
| TOTAL | | 965 | 860 | 995 | 985 | 3,805 |
| STATE: | DISTRICT OF COLUMBIA | | | | | |
| ZIP CODE | | QUARTER 1 | QUARTER 2 | QUARTER 3 | QUARTER 4 | TOTAL GRAMS |
| | 200 | 645 | 685 | 665 | 670 | 2,665 |
| TOTAL | | 645 | 685 | 665 | 670 | 2,665 |
| STATE: | FLORIDA | | | | | |
| ZIP CODE | | QUARTER 1 | QUARTER 2 | QUARTER 3 | QUARTER 4 | TOTAL GRAMS |
| | 320 | 1,635 | 1,585 | 1,255 | 1,285 | 5,760 |
| | 321 | 1,210 | 1,005 | 1,040 | 985 | 4,240 |
| | 322 | 2,120 | 1,760 | 1,865 | 1,700 | 7,445 |
| | 323 | 705 | 615 | 760 | 685 | 2,765 |
| | 324 | 995 | 825 | 990 | 925 | 3,735 |
| | 325 | 1,025 | 1,005 | 1,055 | 960 | 4,045 |
| | 326 | 850 | 710 | 660 | 605 | 2,825 |
| | 327 | 1,765 | 1,680 | 1,900 | 1,825 | 7,170 |
| | 328 | 1,285 | 1,200 | 1,065 | 1,025 | 4,575 |
| | 329 | 1,685 | 1,920 | 1,590 | 1,645 | 6,840 |
| | 330 | 2,855 | 2,620 | 2,535 | 2,800 | 10,810 |
| | 331 | 1,320 | 1,125 | 910 | 1,190 | 4,545 |
| | 333 | 1,465 | 1,450 | 1,380 | 1,530 | 5,825 |
| | 334 | 4,205 | 3,915 | 3,650 | 3,680 | 15,450 |
| | 335 | 2,010 | 1,995 | 1,735 | 1,910 | 7,650 |
| | 336 | 1,485 | 1,370 | 1,550 | 1,540 | 5,945 |
| | 337 | 1,965 | 1,840 | 1,780 | 1,945 | 7,530 |
| | 338 | 960 | 1,075 | 860 | 1,030 | 3,925 |
| | 339 | 1,980 | 1,815 | 1,715 | 1,865 | 7,375 |
| | 341 | 755 | 725 | 535 | 735 | 2,750 |
| | 342 | 1,700 | 1,375 | 1,355 | 1,600 | 6,030 |
| | 344 | 1,080 | 1,245 | 1,260 | 1,155 | 4,740 |
| | 346 | 6,700 | 1,675 | 1,455 | 1,550 | 11,380 |
| | 347 | 900 | 860 | 815 | 1,015 | 3,590 |
| | 349 | 840 | 875 | 840 | 670 | 3,225 |
| TOTAL | | 43,495 | 36,265 | 34,555 | 35,855 | 150,170 |
| STATE: | GEORGIA | | | | | |
| ZIP CODE | | QUARTER 1 | QUARTER 2 | QUARTER 3 | QUARTER 4 | TOTAL GRAMS |
| | 300 | 2,585 | 2,710 | 2,635 | 2,475 | 10,405 |
| | 301 | 2,375 | 2,360 | 2,250 | 2,445 | 9,430 |
| | 302 | 1,795 | 1,875 | 1,865 | 1,775 | 7,310 |
| | 303 | 950 | 835 | 850 | 925 | 3,560 |
| | 304 | 465 | 440 | 530 | 545 | 1,980 |
| | 305 | 1,360 | 1,270 | 1,170 | 1,190 | 4,990 |
| | 306 | 755 | 810 | 765 | 715 | 3,045 |
| | 307 | 655 | 590 | 575 | 665 | 2,485 |
| | 308 | 250 | 315 | 370 | 275 | 1,210 |

DRUG CODE:2165DRUG NAME:BUTALBITAL
STATE:GEORGIA

| ZIP CODE | QUARTER 1 | QUARTER 2 | QUARTER 3 | QUARTER 4 | TOTAL GRAMS |
|---|---|---|---|---|---|

| | | | | | |
|---|---|---|---|---|---|
| 309 | 470 | 390 | 435 | 440 | 1,735 |
| 310 | 670 | 605 | 580 | 700 | 2,555 |
| 312 | 305 | 265 | 280 | 290 | 1,140 |
| 313 | 220 | 250 | 285 | 330 | 1,085 |
| 314 | 225 | 285 | 275 | 260 | 1,045 |
| 315 | 1,120 | 1,195 | 1,275 | 1,245 | 4,835 |
| 316 | 370 | 400 | 445 | 460 | 1,675 |
| 317 | 680 | 635 | 650 | 625 | 2,590 |
| 318 | 25 | 55 | 25 | 20 | 125 |
| 319 | 450 | 485 | 560 | 500 | 1,995 |
| 398 | 145 | 190 | 170 | 150 | 655 |
| TOTAL | 15,870 | 15,960 | 15,990 | 16,030 | 63,850 |

STATE: GUAM

| ZIP CODE | QUARTER 1 | QUARTER 2 | QUARTER 3 | QUARTER 4 | TOTAL GRAMS |
|---|---|---|---|---|---|
| 969 | 15 | 5 | 15 | 0 | 35 |
| TOTAL | 15 | 5 | 15 | 0 | 35 |

STATE: HAWAII

| ZIP CODE | QUARTER 1 | QUARTER 2 | QUARTER 3 | QUARTER 4 | TOTAL GRAMS |
|---|---|---|---|---|---|
| 967 | 1,270 | 1,220 | 1,220 | 1,260 | 4,970 |
| 968 | 775 | 690 | 815 | 670 | 2,950 |
| TOTAL | 2,045 | 1,910 | 2,035 | 1,930 | 7,920 |

STATE: IDAHO

| ZIP CODE | QUARTER 1 | QUARTER 2 | QUARTER 3 | QUARTER 4 | TOTAL GRAMS |
|---|---|---|---|---|---|
| 832 | 225 | 245 | 230 | 250 | 950 |
| 833 | 195 | 215 | 240 | 215 | 865 |
| 834 | 205 | 235 | 245 | 290 | 975 |
| 835 | 120 | 90 | 135 | 100 | 445 |
| 836 | 575 | 695 | 570 | 585 | 2,425 |
| 837 | 335 | 280 | 320 | 265 | 1,200 |
| 838 | 295 | 345 | 310 | 320 | 1,270 |
| TOTAL | 1,950 | 2,105 | 2,050 | 2,025 | 8,130 |

STATE: ILLINOIS

| ZIP CODE | QUARTER 1 | QUARTER 2 | QUARTER 3 | QUARTER 4 | TOTAL GRAMS |
|---|---|---|---|---|---|
| 600 | 3,450 | 3,755 | 3,755 | 3,780 | 14,740 |
| 601 | 2,320 | 2,040 | 1,915 | 2,310 | 8,585 |
| 602 | 85 | 80 | 70 | 105 | 340 |
| 603 | 45 | 55 | 50 | 85 | 235 |
| 604 | 2,160 | 2,050 | 2,085 | 2,170 | 8,465 |
| 605 | 1,275 | 1,225 | 1,355 | 1,410 | 5,265 |
| 606 | 1,520 | 1,260 | 1,235 | 1,255 | 5,270 |
| 607 | 125 | 170 | 110 | 65 | 470 |
| 608 | 35 | 50 | 55 | 25 | 165 |
| 609 | 140 | 170 | 245 | 225 | 780 |
| 610 | 210 | 260 | 255 | 190 | 915 |
| 611 | 390 | 310 | 380 | 440 | 1,520 |
| 612 | 100 | 60 | 105 | 45 | 310 |
| 613 | 130 | 100 | 155 | 95 | 480 |
| 614 | 275 | 155 | 195 | 205 | 830 |
| 615 | 255 | 240 | 240 | 250 | 985 |
| 616 | 235 | 155 | 190 | 220 | 800 |
| 617 | 205 | 300 | 175 | 190 | 870 |
| 618 | 310 | 400 | 385 | 390 | 1,485 |
| 619 | 30 | 40 | 45 | 25 | 140 |
| 620 | 400 | 505 | 490 | 435 | 1,830 |
| 622 | 355 | 350 | 440 | 455 | 1,600 |
| 623 | 105 | 85 | 135 | 125 | 450 |
| 624 | 110 | 90 | 175 | 115 | 490 |
| 625 | 185 | 265 | 160 | 195 | 805 |

| | | | | | |
|---|---|---|---|---|---|
| 626 | 70 | 110 | 95 | 105 | 380 |
| 627 | 370 | 260 | 265 | 215 | 1,110 |
| 628 | 220 | 250 | 255 | 210 | 935 |
| 629 | 495 | 470 | 475 | 480 | 1,920 |
| TOTAL | 15,605 | 15,260 | 15,490 | 15,815 | 62,170 |

STATE: INDIANA

| ZIP CODE | QUARTER 1 | QUARTER 2 | QUARTER 3 | QUARTER 4 | TOTAL GRAMS |
|---|---|---|---|---|---|
| 460 | 450 | 465 | 480 | 500 | 1,895 |

DRUG CODE:2165DRUG NAME:BUTALBITAL
STATE:INDIANA

| ZIP CODE | QUARTER 1 | QUARTER 2 | QUARTER 3 | QUARTER 4 | TOTAL GRAMS |
|---|---|---|---|---|---|
| 461 | 870 | 850 | 835 | 785 | 3,340 |
| 462 | 1,015 | 1,000 | 990 | 1,025 | 4,030 |
| 463 | 1,230 | 1,160 | 1,215 | 1,050 | 4,655 |
| 464 | 240 | 220 | 180 | 160 | 800 |
| 465 | 300 | 320 | 365 | 315 | 1,300 |
| 466 | 95 | 70 | 55 | 45 | 265 |
| 467 | 320 | 320 | 285 | 260 | 1,185 |
| 468 | 185 | 145 | 195 | 190 | 715 |
| 469 | 305 | 360 | 235 | 320 | 1,220 |
| 470 | 160 | 180 | 210 | 130 | 680 |
| 471 | 450 | 540 | 730 | 430 | 2,150 |
| 472 | 200 | 160 | 165 | 195 | 720 |
| 473 | 250 | 250 | 195 | 255 | 950 |
| 474 | 300 | 260 | 285 | 260 | 1,105 |
| 475 | 260 | 320 | 325 | 295 | 1,200 |
| 476 | 180 | 265 | 305 | 225 | 975 |
| 477 | 355 | 445 | 400 | 395 | 1,595 |
| 478 | 115 | 80 | 110 | 100 | 405 |
| 479 | 290 | 245 | 295 | 305 | 1,135 |
| TOTAL | 7,570 | 7,655 | 7,855 | 7,240 | 30,320 |

STATE: IOWA

| ZIP CODE | QUARTER 1 | QUARTER 2 | QUARTER 3 | QUARTER 4 | TOTAL GRAMS |
|---|---|---|---|---|---|
| 500 | 95 | 130 | 160 | 120 | 505 |
| 501 | 95 | 85 | 85 | 90 | 355 |
| 502 | 155 | 180 | 140 | 185 | 660 |
| 503 | 170 | 175 | 170 | 165 | 680 |
| 504 | 80 | 75 | 40 | 40 | 235 |
| 505 | 85 | 90 | 50 | 80 | 305 |
| 506 | 60 | 55 | 75 | 60 | 250 |
| 507 | 45 | 50 | 80 | 85 | 260 |
| 508 | 30 | 40 | 35 | 45 | 150 |
| 510 | 75 | 85 | 100 | 90 | 350 |
| 511 | 70 | 65 | 50 | 60 | 245 |
| 512 | 15 | 10 | 15 | 10 | 50 |
| 513 | 15 | 30 | 20 | 30 | 95 |
| 514 | 105 | 110 | 70 | 100 | 385 |
| 515 | 95 | 85 | 100 | 115 | 395 |
| 516 | 35 | 55 | 35 | 55 | 180 |
| 520 | 210 | 230 | 200 | 210 | 850 |
| 521 | 30 | 30 | 25 | 35 | 120 |
| 522 | 125 | 70 | 35 | 55 | 285 |
| 523 | 60 | 65 | 45 | 60 | 230 |
| 524 | 95 | 65 | 50 | 45 | 255 |
| 525 | 20 | 55 | 35 | 25 | 135 |

| | | | | | |
|---|---|---|---|---|---|
| 526 | 70 | 160 | 80 | 90 | 400 |
| 527 | 200 | 95 | 150 | 135 | 580 |
| 528 | 175 | 170 | 160 | 160 | 665 |
| TOTAL | 2,210 | 2,260 | 2,005 | 2,145 | 8,620 |

STATE: KANSAS

| ZIP CODE | QUARTER 1 | QUARTER 2 | QUARTER 3 | QUARTER 4 | TOTAL GRAMS |
|---|---|---|---|---|---|
| 660 | 560 | 555 | 630 | 460 | 2,205 |
| 661 | 160 | 220 | 215 | 275 | 870 |
| 662 | 690 | 740 | 1,100 | 1,450 | 3,980 |
| 664 | 45 | 50 | 65 | 80 | 240 |
| 665 | 140 | 145 | 95 | 90 | 470 |
| 666 | 155 | 120 | 85 | 120 | 480 |
| 667 | 175 | 115 | 170 | 230 | 690 |
| 668 | 100 | 85 | 135 | 170 | 490 |
| 669 | 5 | 0 | 5 | 0 | 10 |
| 670 | 185 | 220 | 160 | 215 | 780 |
| 671 | 55 | 55 | 85 | 100 | 295 |
| 672 | 580 | 540 | 480 | 515 | 2,115 |
| 673 | 140 | 115 | 120 | 150 | 525 |
| 674 | 350 | 245 | 300 | 285 | 1,180 |
| 675 | 100 | 85 | 65 | 80 | 330 |
| 676 | 0 | 15 | 10 | 10 | 35 |

DRUG CODE:2165DRUG NAME:BUTALBITAL
STATE:KANSAS

| ZIP CODE | QUARTER 1 | QUARTER 2 | QUARTER 3 | QUARTER 4 | TOTAL GRAMS |
|---|---|---|---|---|---|
| 677 | 30 | 20 | 25 | 0 | 75 |
| 678 | 65 | 55 | 15 | 35 | 170 |
| 679 | 130 | 165 | 130 | 90 | 515 |
| TOTAL | 3,665 | 3,545 | 3,890 | 4,355 | 15,455 |

STATE: KENTUCKY

| ZIP CODE | QUARTER 1 | QUARTER 2 | QUARTER 3 | QUARTER 4 | TOTAL GRAMS |
|---|---|---|---|---|---|
| 400 | 280 | 205 | 215 | 240 | 940 |
| 401 | 50 | 60 | 60 | 90 | 260 |
| 402 | 870 | 925 | 880 | 930 | 3,605 |
| 403 | 290 | 275 | 350 | 315 | 1,230 |
| 404 | 130 | 100 | 75 | 70 | 375 |
| 405 | 210 | 165 | 165 | 180 | 720 |
| 406 | 20 | 20 | 25 | 10 | 75 |
| 407 | 105 | 110 | 130 | 120 | 465 |
| 408 | 105 | 115 | 80 | 75 | 375 |
| 409 | 45 | 50 | 60 | 55 | 210 |
| 410 | 640 | 760 | 585 | 685 | 2,670 |
| 411 | 75 | 65 | 50 | 70 | 260 |
| 412 | 60 | 55 | 45 | 50 | 210 |
| 413 | 5 | 0 | 5 | 20 | 30 |
| 415 | 65 | 100 | 105 | 70 | 340 |
| 416 | 35 | 35 | 60 | 55 | 185 |
| 417 | 45 | 30 | 5 | 45 | 125 |
| 418 | 40 | 30 | 45 | 35 | 150 |
| 420 | 340 | 415 | 415 | 400 | 1,570 |
| 421 | 265 | 240 | 265 | 255 | 1,025 |
| 422 | 140 | 130 | 165 | 105 | 540 |
| 423 | 275 | 265 | 265 | 260 | 1,065 |
| 424 | 230 | 170 | 135 | 140 | 675 |
| 425 | 50 | 35 | 35 | 45 | 165 |
| 426 | 75 | 50 | 50 | 55 | 230 |

| | | | | | |
|---|---|---|---|---|---|
| 427 | 290 | 305 | 325 | 330 | 1,250 |
| TOTAL | 4,735 | 4,710 | 4,595 | 4,705 | 18,745 |

STATE: LOUISIANA

| ZIP CODE | QUARTER 1 | QUARTER 2 | QUARTER 3 | QUARTER 4 | TOTAL GRAMS |
|---|---|---|---|---|---|
| 700 | 2,110 | 2,065 | 2,060 | 2,105 | 8,340 |
| 701 | 520 | 455 | 495 | 465 | 1,935 |
| 703 | 245 | 225 | 205 | 190 | 865 |
| 704 | 1,360 | 1,210 | 1,135 | 1,210 | 4,915 |
| 705 | 840 | 835 | 710 | 800 | 3,185 |
| 706 | 555 | 655 | 605 | 570 | 2,385 |
| 707 | 490 | 465 | 555 | 455 | 1,965 |
| 708 | 350 | 315 | 405 | 370 | 1,440 |
| 710 | 75 | 65 | 85 | 100 | 325 |
| 711 | 295 | 225 | 275 | 355 | 1,150 |
| 712 | 495 | 470 | 485 | 405 | 1,855 |
| 713 | 550 | 480 | 460 | 440 | 1,930 |
| 714 | 540 | 430 | 455 | 470 | 1,895 |
| TOTAL | 8,425 | 7,895 | 7,930 | 7,935 | 32,185 |

STATE: MAINE

| ZIP CODE | QUARTER 1 | QUARTER 2 | QUARTER 3 | QUARTER 4 | TOTAL GRAMS |
|---|---|---|---|---|---|
| 39 | 55 | 35 | 20 | 30 | 140 |
| 40 | 385 | 360 | 375 | 400 | 1,520 |
| 41 | 160 | 115 | 155 | 120 | 550 |
| 42 | 80 | 100 | 65 | 100 | 345 |
| 43 | 60 | 80 | 85 | 60 | 285 |
| 44 | 90 | 80 | 80 | 85 | 335 |
| 45 | 75 | 60 | 25 | 15 | 175 |
| 46 | 75 | 45 | 55 | 55 | 230 |
| 47 | 165 | 145 | 110 | 140 | 560 |
| 48 | 10 | 0 | 5 | 5 | 20 |
| 49 | 245 | 385 | 290 | 305 | 1,225 |
| TOTAL | 1,400 | 1,405 | 1,265 | 1,315 | 5,385 |

STATE: MARYLAND

| ZIP CODE | QUARTER 1 | QUARTER 2 | QUARTER 3 | QUARTER 4 | TOTAL GRAMS |
|---|---|---|---|---|---|
| 206 | 725 | 855 | 720 | 685 | 2,985 |

DRUG CODE:2165DRUG NAME:BUTALBITAL
STATE:MARYLAND

| ZIP CODE | QUARTER 1 | QUARTER 2 | QUARTER 3 | QUARTER 4 | TOTAL GRAMS |
|---|---|---|---|---|---|
| 207 | 680 | 855 | 750 | 660 | 2,945 |
| 208 | 910 | 950 | 920 | 885 | 3,665 |
| 209 | 210 | 245 | 185 | 225 | 865 |
| 210 | 1,175 | 1,265 | 1,280 | 1,310 | 5,030 |
| 211 | 605 | 575 | 550 | 460 | 2,190 |
| 212 | 1,195 | 1,210 | 1,045 | 1,155 | 4,605 |
| 214 | 155 | 195 | 220 | 175 | 745 |
| 215 | 365 | 390 | 390 | 390 | 1,535 |
| 216 | 240 | 235 | 215 | 225 | 915 |
| 217 | 1,305 | 1,230 | 1,215 | 1,260 | 5,010 |
| 218 | 275 | 320 | 345 | 320 | 1,260 |
| 219 | 160 | 155 | 140 | 170 | 625 |
| TOTAL | 8,000 | 8,480 | 7,975 | 7,920 | 32,375 |

STATE: MASSACHUSETTS

| ZIP CODE | QUARTER 1 | QUARTER 2 | QUARTER 3 | QUARTER 4 | TOTAL GRAMS |
|---|---|---|---|---|---|
| 10 | 550 | 645 | 510 | 590 | 2,295 |

| | | | | | |
|---|---|---|---|---|---|
| 11 | 85 | 105 | 115 | 70 | 375 |
| 12 | 115 | 115 | 125 | 125 | 480 |
| 13 | 120 | 65 | 65 | 75 | 325 |
| 14 | 285 | 315 | 270 | 300 | 1,170 |
| 15 | 435 | 430 | 435 | 440 | 1,740 |
| 16 | 260 | 205 | 400 | 240 | 1,105 |
| 17 | 410 | 350 | 335 | 355 | 1,450 |
| 18 | 610 | 575 | 525 | 480 | 2,190 |
| 19 | 400 | 325 | 345 | 250 | 1,320 |
| 20 | 350 | 405 | 325 | 230 | 1,310 |
| 21 | 770 | 785 | 840 | 795 | 3,190 |
| 22 | 25 | 20 | 10 | 20 | 75 |
| 23 | 275 | 200 | 230 | 240 | 945 |
| 24 | 270 | 340 | 295 | 270 | 1,175 |
| 25 | 145 | 115 | 110 | 120 | 490 |
| 26 | 315 | 235 | 215 | 215 | 980 |
| 27 | 520 | 510 | 480 | 365 | 1,875 |
| TOTAL | 5,940 | 5,740 | 5,630 | 5,180 | 22,490 |

STATE: MICHIGAN

| ZIP CODE | QUARTER 1 | QUARTER 2 | QUARTER 3 | QUARTER 4 | TOTAL GRAMS |
|---|---|---|---|---|---|
| 480 | 3,380 | 3,375 | 3,195 | 3,130 | 13,080 |
| 481 | 3,680 | 3,225 | 3,330 | 3,270 | 13,505 |
| 482 | 695 | 605 | 690 | 775 | 2,765 |
| 483 | 2,165 | 2,065 | 2,100 | 2,220 | 8,550 |
| 484 | 820 | 910 | 865 | 1,030 | 3,625 |
| 485 | 490 | 430 | 435 | 310 | 1,665 |
| 486 | 450 | 445 | 480 | 430 | 1,805 |
| 487 | 280 | 255 | 310 | 350 | 1,195 |
| 488 | 1,110 | 935 | 915 | 1,010 | 3,970 |
| 489 | 320 | 390 | 495 | 565 | 1,770 |
| 490 | 795 | 1,025 | 845 | 705 | 3,370 |
| 491 | 70 | 80 | 50 | 25 | 225 |
| 492 | 560 | 550 | 505 | 530 | 2,145 |
| 493 | 230 | 195 | 165 | 315 | 905 |
| 494 | 595 | 655 | 690 | 805 | 2,745 |
| 495 | 360 | 445 | 305 | 390 | 1,500 |
| 496 | 365 | 300 | 325 | 315 | 1,305 |
| 497 | 280 | 280 | 285 | 280 | 1,125 |
| 498 | 190 | 200 | 240 | 195 | 825 |
| 499 | 20 | 25 | 25 | 40 | 110 |
| TOTAL | 16,855 | 16,390 | 16,250 | 16,690 | 66,185 |

STATE: MINNESOTA

| ZIP CODE | QUARTER 1 | QUARTER 2 | QUARTER 3 | QUARTER 4 | TOTAL GRAMS |
|---|---|---|---|---|---|
| 550 | 340 | 375 | 385 | 430 | 1,530 |
| 551 | 625 | 605 | 605 | 595 | 2,430 |
| 553 | 620 | 795 | 675 | 760 | 2,850 |
| 554 | 580 | 535 | 580 | 525 | 2,220 |
| 556 | 5 | 5 | 5 | 10 | 25 |
| 557 | 325 | 260 | 235 | 260 | 1,080 |
| 558 | 60 | 45 | 60 | 60 | 225 |

DRUG CODE:2165DRUG NAME:BUTALBITAL
STATE:MINNESOTA

| ZIP CODE | QUARTER 1 | QUARTER 2 | QUARTER 3 | QUARTER 4 | TOTAL GRAMS |
|---|---|---|---|---|---|
| 559 | 90 | 75 | 40 | 50 | 255 |
| 560 | 140 | 225 | 150 | 160 | 675 |
| 561 | 35 | 70 | 25 | 35 | 165 |
| 562 | 215 | 160 | 120 | 130 | 625 |

| | | | | | |
|---|---|---|---|---|---|
| 563 | 180 | 195 | 215 | 165 | 755 |
| 564 | 95 | 145 | 105 | 90 | 435 |
| 565 | 110 | 160 | 135 | 165 | 570 |
| 566 | 95 | 75 | 100 | 70 | 340 |
| 567 | 100 | 130 | 120 | 45 | 395 |
| TOTAL | 3,615 | 3,855 | 3,555 | 3,550 | 14,575 |

STATE: MISSISSIPPI

| ZIP CODE | QUARTER 1 | QUARTER 2 | QUARTER 3 | QUARTER 4 | TOTAL GRAMS |
|---|---|---|---|---|---|
| 386 | 430 | 450 | 430 | 430 | 1,740 |
| 387 | 25 | 30 | 40 | 35 | 130 |
| 388 | 375 | 295 | 305 | 380 | 1,355 |
| 389 | 35 | 40 | 35 | 70 | 180 |
| 390 | 240 | 290 | 220 | 240 | 990 |
| 391 | 330 | 360 | 220 | 380 | 1,290 |
| 392 | 185 | 190 | 115 | 130 | 620 |
| 393 | 195 | 180 | 195 | 165 | 735 |
| 394 | 650 | 495 | 590 | 485 | 2,220 |
| 395 | 730 | 725 | 730 | 785 | 2,970 |
| 396 | 115 | 90 | 80 | 90 | 375 |
| 397 | 215 | 200 | 220 | 175 | 810 |
| TOTAL | 3,525 | 3,345 | 3,180 | 3,365 | 13,415 |

STATE: MISSOURI

| ZIP CODE | QUARTER 1 | QUARTER 2 | QUARTER 3 | QUARTER 4 | TOTAL GRAMS |
|---|---|---|---|---|---|
| 630 | 1,535 | 1,380 | 1,380 | 1,420 | 5,715 |
| 631 | 2,015 | 2,520 | 2,315 | 2,525 | 9,375 |
| 633 | 1,010 | 1,085 | 955 | 1,015 | 4,065 |
| 634 | 75 | 110 | 85 | 90 | 360 |
| 635 | 65 | 85 | 80 | 35 | 265 |
| 636 | 140 | 255 | 295 | 280 | 970 |
| 637 | 435 | 455 | 445 | 355 | 1,690 |
| 638 | 140 | 170 | 150 | 160 | 620 |
| 639 | 190 | 165 | 135 | 165 | 655 |
| 640 | 1,015 | 965 | 970 | 990 | 3,940 |
| 641 | 1,295 | 1,220 | 1,455 | 1,480 | 5,450 |
| 644 | 90 | 95 | 90 | 90 | 365 |
| 645 | 145 | 115 | 90 | 100 | 450 |
| 646 | 90 | 85 | 135 | 135 | 445 |
| 647 | 140 | 150 | 150 | 170 | 610 |
| 648 | 210 | 200 | 275 | 255 | 940 |
| 650 | 140 | 205 | 200 | 140 | 685 |
| 651 | 105 | 95 | 110 | 85 | 395 |
| 652 | 185 | 105 | 95 | 85 | 470 |
| 653 | 75 | 60 | 80 | 105 | 320 |
| 654 | 75 | 100 | 100 | 125 | 400 |
| 655 | 145 | 185 | 160 | 200 | 690 |
| 656 | 235 | 175 | 175 | 150 | 735 |
| 657 | 325 | 280 | 325 | 200 | 1,130 |
| 658 | 360 | 370 | 355 | 340 | 1,425 |
| TOTAL | 10,235 | 10,630 | 10,605 | 10,695 | 42,165 |

STATE: MONTANA

| ZIP CODE | QUARTER 1 | QUARTER 2 | QUARTER 3 | QUARTER 4 | TOTAL GRAMS |
|---|---|---|---|---|---|
| 590 | 150 | 155 | 120 | 120 | 545 |
| 591 | 245 | 265 | 280 | 375 | 1,165 |
| 592 | 40 | 40 | 50 | 75 | 205 |
| 593 | 150 | 115 | 160 | 115 | 540 |
| 594 | 135 | 105 | 100 | 130 | 470 |
| 595 | 30 | 5 | 30 | 20 | 85 |
| 596 | 65 | 65 | 90 | 55 | 275 |

| | | | | | |
|---|---|---|---|---|---|
| 597 | 290 | 275 | 290 | 235 | 1,090 |
| 598 | 220 | 285 | 320 | 260 | 1,085 |
| 599 | 150 | 290 | 325 | 175 | 940 |
| TOTAL | 1,475 | 1,600 | 1,765 | 1,560 | 6,400 |

DRUG CODE:2165DRUG NAME:BUTALBITAL
STATE: NEBRASKA

| ZIP CODE | QUARTER 1 | QUARTER 2 | QUARTER 3 | QUARTER 4 | TOTAL GRAMS |
|---|---|---|---|---|---|
| 680 | 110 | 125 | 150 | 120 | 505 |
| 681 | 705 | 685 | 615 | 625 | 2,630 |
| 683 | 70 | 105 | 70 | 105 | 350 |
| 684 | 30 | 30 | 40 | 30 | 130 |
| 685 | 510 | 475 | 325 | 370 | 1,680 |
| 686 | 105 | 125 | 105 | 115 | 450 |
| 687 | 70 | 50 | 70 | 60 | 250 |
| 688 | 275 | 255 | 200 | 195 | 925 |
| 689 | 25 | 45 | 0 | 20 | 90 |
| 690 | 5 | 5 | 5 | 10 | 25 |
| 691 | 130 | 110 | 135 | 135 | 510 |
| 692 | 0 | 0 | 5 | 0 | 5 |
| 693 | 195 | 210 | 175 | 160 | 740 |
| TOTAL | 2,230 | 2,220 | 1,895 | 1,945 | 8,290 |

STATE: NEVADA

| ZIP CODE | QUARTER 1 | QUARTER 2 | QUARTER 3 | QUARTER 4 | TOTAL GRAMS |
|---|---|---|---|---|---|
| 890 | 2,290 | 2,060 | 2,225 | 2,155 | 8,730 |
| 891 | 3,845 | 3,680 | 3,625 | 3,760 | 14,910 |
| 893 | 65 | 85 | 50 | 55 | 255 |
| 894 | 1,160 | 1,030 | 925 | 1,010 | 4,125 |
| 895 | 1,375 | 1,315 | 1,280 | 1,150 | 5,120 |
| 897 | 390 | 370 | 255 | 205 | 1,220 |
| 898 | 190 | 140 | 190 | 290 | 810 |
| TOTAL | 9,315 | 8,680 | 8,550 | 8,625 | 35,170 |

STATE: NEW HAMPSHIRE

| ZIP CODE | QUARTER 1 | QUARTER 2 | QUARTER 3 | QUARTER 4 | TOTAL GRAMS |
|---|---|---|---|---|---|
| 30 | 385 | 300 | 395 | 380 | 1,460 |
| 31 | 215 | 235 | 230 | 315 | 995 |
| 32 | 190 | 205 | 155 | 160 | 710 |
| 33 | 70 | 45 | 60 | 25 | 200 |
| 34 | 75 | 90 | 45 | 65 | 275 |
| 35 | 40 | 45 | 25 | 35 | 145 |
| 36 | 10 | 10 | 10 | 10 | 40 |
| 37 | 25 | 25 | 20 | 25 | 95 |
| 38 | 300 | 320 | 330 | 320 | 1,270 |
| TOTAL | 1,310 | 1,275 | 1,270 | 1,335 | 5,190 |

STATE: NEW JERSEY

| ZIP CODE | QUARTER 1 | QUARTER 2 | QUARTER 3 | QUARTER 4 | TOTAL GRAMS |
|---|---|---|---|---|---|
| 70 | 1,785 | 1,710 | 1,600 | 1,550 | 6,645 |
| 71 | 165 | 160 | 125 | 100 | 550 |
| 72 | 90 | 60 | 75 | 60 | 285 |
| 73 | 80 | 90 | 90 | 60 | 320 |
| 74 | 1,155 | 1,110 | 1,165 | 1,200 | 4,630 |
| 75 | 175 | 105 | 110 | 100 | 490 |
| 76 | 750 | 660 | 725 | 695 | 2,830 |
| 77 | 1,470 | 1,540 | 1,395 | 1,265 | 5,670 |
| 78 | 810 | 695 | 715 | 690 | 2,910 |
| 79 | 420 | 430 | 340 | 240 | 1,430 |

| | | | | | |
|---|---|---|---|---|---|
| 80 | 2,425 | 2,210 | 2,040 | 2,100 | 8,775 |
| 81 | 95 | 115 | 80 | 115 | 405 |
| 82 | 545 | 440 | 460 | 360 | 1,805 |
| 83 | 270 | 260 | 255 | 255 | 1,040 |
| 84 | 45 | 10 | 30 | 20 | 105 |
| 85 | 315 | 215 | 245 | 245 | 1,020 |
| 86 | 410 | 400 | 325 | 380 | 1,515 |
| 87 | 615 | 610 | 560 | 500 | 2,285 |
| 88 | 1,245 | 1,280 | 1,180 | 1,050 | 4,755 |
| 89 | 100 | 70 | 90 | 95 | 355 |
| TOTAL | 12,965 | 12,170 | 11,605 | 11,080 | 47,820 |

STATE: NEW MEXICO

| ZIP CODE | QUARTER 1 | QUARTER 2 | QUARTER 3 | QUARTER 4 | TOTAL GRAMS |
|---|---|---|---|---|---|
| 870 | 205 | 180 | 260 | 185 | 830 |
| 871 | 970 | 955 | 920 | 840 | 3,685 |
| 873 | 10 | 5 | 5 | 0 | 20 |
| 874 | 45 | 30 | 30 | 85 | 190 |
| 875 | 170 | 180 | 200 | 145 | 695 |

DRUG CODE:2165DRUG NAME:BUTALBITAL
STATE:NEW MEXICO

| ZIP CODE | QUARTER 1 | QUARTER 2 | QUARTER 3 | QUARTER 4 | TOTAL GRAMS |
|---|---|---|---|---|---|
| 877 | 50 | 60 | 50 | 70 | 230 |
| 879 | 35 | 15 | 40 | 55 | 145 |
| 880 | 335 | 385 | 315 | 255 | 1,290 |
| 881 | 80 | 75 | 45 | 40 | 240 |
| 882 | 240 | 290 | 320 | 315 | 1,165 |
| 883 | 50 | 125 | 50 | 65 | 290 |
| 884 | 15 | 20 | 10 | 10 | 55 |
| TOTAL | 2,205 | 2,320 | 2,245 | 2,065 | 8,835 |

STATE: NEW YORK

| ZIP CODE | QUARTER 1 | QUARTER 2 | QUARTER 3 | QUARTER 4 | TOTAL GRAMS |
|---|---|---|---|---|---|
| 100 | 730 | 770 | 650 | 610 | 2,760 |
| 101 | 100 | 40 | 70 | 110 | 320 |
| 102 | 5 | 0 | 0 | 0 | 5 |
| 103 | 435 | 435 | 405 | 355 | 1,630 |
| 104 | 455 | 395 | 330 | 345 | 1,525 |
| 105 | 520 | 485 | 465 | 430 | 1,900 |
| 106 | 115 | 135 | 125 | 75 | 450 |
| 107 | 130 | 145 | 155 | 205 | 635 |
| 108 | 55 | 80 | 75 | 90 | 300 |
| 109 | 635 | 545 | 565 | 585 | 2,330 |
| 110 | 145 | 90 | 110 | 220 | 565 |
| 111 | 60 | 110 | 105 | 55 | 330 |
| 112 | 730 | 570 | 610 | 675 | 2,585 |
| 113 | 485 | 475 | 335 | 430 | 1,725 |
| 114 | 265 | 275 | 235 | 245 | 1,020 |
| 115 | 760 | 705 | 580 | 580 | 2,625 |
| 116 | 70 | 70 | 80 | 45 | 265 |
| 117 | 1,670 | 1,690 | 1,450 | 1,380 | 6,190 |
| 118 | 190 | 185 | 195 | 55 | 625 |
| 119 | 175 | 195 | 130 | 190 | 690 |
| 120 | 440 | 360 | 315 | 315 | 1,430 |
| 121 | 70 | 130 | 65 | 100 | 365 |
| 122 | 115 | 95 | 95 | 120 | 425 |
| 123 | 220 | 170 | 225 | 205 | 820 |
| 124 | 270 | 260 | 190 | 145 | 865 |
| 125 | 365 | 285 | 250 | 275 | 1,175 |

| | | | | | |
|---|---|---|---|---|---|
| 126 | 170 | 115 | 150 | 120 | 555 |
| 127 | 175 | 135 | 130 | 180 | 620 |
| 128 | 255 | 225 | 250 | 220 | 950 |
| 129 | 135 | 80 | 80 | 55 | 350 |
| 130 | 175 | 285 | 225 | 220 | 905 |
| 131 | 110 | 95 | 120 | 75 | 400 |
| 132 | 150 | 145 | 140 | 110 | 545 |
| 133 | 115 | 120 | 105 | 100 | 440 |
| 134 | 35 | 35 | 30 | 35 | 135 |
| 135 | 30 | 30 | 25 | 40 | 125 |
| 136 | 220 | 175 | 215 | 155 | 765 |
| 137 | 140 | 30 | 40 | 50 | 260 |
| 138 | 80 | 80 | 75 | 105 | 340 |
| 139 | 115 | 155 | 165 | 65 | 500 |
| 140 | 425 | 360 | 325 | 240 | 1,350 |
| 141 | 215 | 115 | 130 | 150 | 610 |
| 142 | 505 | 415 | 415 | 375 | 1,710 |
| 143 | 65 | 15 | 55 | 55 | 190 |
| 144 | 180 | 310 | 280 | 220 | 990 |
| 145 | 155 | 130 | 200 | 155 | 640 |
| 146 | 430 | 460 | 355 | 215 | 1,460 |
| 147 | 105 | 70 | 135 | 95 | 405 |
| 148 | 165 | 165 | 155 | 150 | 635 |
| 149 | 25 | 20 | 10 | 35 | 90 |
| TOTAL | 13,385 | 12,460 | 11,620 | 11,060 | 48,525 |

STATE: NORTH CAROLINA

| ZIP CODE | QUARTER 1 | QUARTER 2 | QUARTER 3 | QUARTER 4 | TOTAL GRAMS |
|---|---|---|---|---|---|
| 270 | 340 | 305 | 310 | 275 | 1,230 |
| 271 | 255 | 295 | 240 | 265 | 1,055 |
| 272 | 445 | 420 | 520 | 460 | 1,845 |

DRUG CODE:2165DRUG NAME:BUTALBITAL
STATE:NORTH CAROLINA

| ZIP CODE | QUARTER 1 | QUARTER 2 | QUARTER 3 | QUARTER 4 | TOTAL GRAMS |
|---|---|---|---|---|---|
| 273 | 230 | 200 | 200 | 225 | 855 |
| 274 | 220 | 215 | 170 | 190 | 795 |
| 275 | 750 | 670 | 670 | 720 | 2,810 |
| 276 | 415 | 430 | 390 | 375 | 1,610 |
| 277 | 165 | 185 | 175 | 110 | 635 |
| 278 | 445 | 355 | 335 | 340 | 1,475 |
| 279 | 365 | 420 | 385 | 310 | 1,480 |
| 280 | 855 | 700 | 745 | 810 | 3,110 |
| 281 | 555 | 680 | 600 | 575 | 2,410 |
| 282 | 595 | 595 | 555 | 640 | 2,385 |
| 283 | 980 | 915 | 820 | 855 | 3,570 |
| 284 | 720 | 575 | 605 | 765 | 2,665 |
| 285 | 615 | 610 | 585 | 705 | 2,515 |
| 286 | 830 | 730 | 680 | 710 | 2,950 |
| 287 | 805 | 835 | 840 | 850 | 3,330 |
| 288 | 240 | 290 | 275 | 250 | 1,055 |
| 289 | 55 | 90 | 120 | 120 | 385 |
| TOTAL | 9,880 | 9,515 | 9,220 | 9,550 | 38,165 |

STATE: NORTH DAKOTA

| ZIP CODE | QUARTER 1 | QUARTER 2 | QUARTER 3 | QUARTER 4 | TOTAL GRAMS |
|---|---|---|---|---|---|
| 580 | 80 | 60 | 70 | 50 | 260 |
| 581 | 40 | 35 | 45 | 40 | 160 |
| 582 | 65 | 85 | 70 | 60 | 280 |
| 583 | 5 | 5 | 10 | 5 | 25 |

| | | | | | |
|---|---|---|---|---|---|
| 584 | 25 | 35 | 25 | 20 | 105 |
| 585 | 110 | 85 | 80 | 80 | 355 |
| 586 | 20 | 60 | 25 | 5 | 110 |
| 587 | 50 | 30 | 35 | 15 | 130 |
| 588 | 40 | 60 | 95 | 70 | 265 |
| TOTAL | 435 | 455 | 455 | 345 | 1,690 |

STATE: OHIO

| ZIP CODE | QUARTER 1 | QUARTER 2 | QUARTER 3 | QUARTER 4 | TOTAL GRAMS |
|---|---|---|---|---|---|
| 430 | 845 | 765 | 765 | 745 | 3,120 |
| 431 | 500 | 555 | 570 | 530 | 2,155 |
| 432 | 2,180 | 2,240 | 1,990 | 1,340 | 7,750 |
| 433 | 100 | 85 | 80 | 80 | 345 |
| 434 | 205 | 235 | 260 | 200 | 900 |
| 435 | 680 | 575 | 655 | 635 | 2,545 |
| 436 | 615 | 790 | 475 | 595 | 2,475 |
| 437 | 165 | 145 | 185 | 175 | 670 |
| 438 | 5 | 30 | 10 | 5 | 50 |
| 439 | 175 | 90 | 75 | 90 | 430 |
| 440 | 1,105 | 935 | 950 | 940 | 3,930 |
| 441 | 1,070 | 770 | 825 | 995 | 3,660 |
| 442 | 550 | 555 | 565 | 445 | 2,115 |
| 443 | 135 | 230 | 185 | 235 | 785 |
| 444 | 215 | 225 | 210 | 190 | 840 |
| 445 | 155 | 70 | 75 | 105 | 405 |
| 446 | 470 | 510 | 475 | 460 | 1,915 |
| 447 | 280 | 295 | 180 | 330 | 1,085 |
| 448 | 135 | 155 | 175 | 135 | 600 |
| 449 | 60 | 150 | 90 | 65 | 365 |
| 450 | 645 | 570 | 575 | 480 | 2,270 |
| 451 | 315 | 285 | 290 | 225 | 1,115 |
| 452 | 845 | 945 | 930 | 815 | 3,535 |
| 453 | 430 | 425 | 455 | 440 | 1,750 |
| 454 | 430 | 425 | 450 | 425 | 1,730 |
| 455 | 105 | 100 | 130 | 75 | 410 |
| 456 | 245 | 365 | 280 | 280 | 1,170 |
| 457 | 85 | 110 | 125 | 110 | 430 |
| 458 | 295 | 220 | 310 | 270 | 1,095 |
| TOTAL | 13,040 | 12,850 | 12,340 | 11,415 | 49,645 |

STATE: OKLAHOMA

| ZIP CODE | QUARTER 1 | QUARTER 2 | QUARTER 3 | QUARTER 4 | TOTAL GRAMS |
|---|---|---|---|---|---|
| 730 | 1,845 | 1,740 | 1,925 | 1,955 | 7,465 |
| 731 | 1,830 | 1,685 | 1,669.95 | 1,780 | 6,964.95 |

DRUG CODE:2165DRUG NAME:BUTALBITAL
STATE:OKLAHOMA

| ZIP CODE | QUARTER 1 | QUARTER 2 | QUARTER 3 | QUARTER 4 | TOTAL GRAMS |
|---|---|---|---|---|---|
| 734 | 180 | 200 | 250 | 135 | 765 |
| 735 | 585 | 570 | 525 | 515 | 2,195 |
| 736 | 95 | 90 | 125 | 160 | 470 |
| 737 | 190 | 220 | 225 | 260 | 895 |
| 738 | 50 | 40 | 45 | 70 | 205 |
| 739 | 60 | 50 | 70 | 45 | 225 |
| 740 | 1,610 | 1,530 | 1,440 | 1,580 | 6,160 |
| 741 | 750 | 695 | 620 | 640 | 2,705 |
| 743 | 320 | 300 | 265 | 225 | 1,110 |
| 744 | 525 | 550 | 525 | 505 | 2,105 |
| 745 | 130 | 170 | 125 | 110 | 535 |

RETAIL DRUG DISTRIBUTION BY ZIP CODE WITHIN STATE BY GRAMS WT

| | | | | | |
|---|---|---|---|---|---|
| 746 | 50 | 25 | 60 | 45 | 180 |
| 747 | 400 | 350 | 310 | 325 | 1,385 |
| 748 | 260 | 230 | 235 | 300 | 1,025 |
| 749 | 190 | 115 | 145 | 160 | 610 |
| TOTAL | 9,070 | 8,560 | 8,559.95 | 8,810 | 34,999.95 |

STATE: OREGON

| ZIP CODE | QUARTER 1 | QUARTER 2 | QUARTER 3 | QUARTER 4 | TOTAL GRAMS |
|---|---|---|---|---|---|
| 970 | 740 | 640 | 615 | 620 | 2,615 |
| 971 | 480 | 380 | 265 | 315 | 1,440 |
| 972 | 500 | 405 | 565 | 480 | 1,950 |
| 973 | 655 | 635 | 655 | 625 | 2,570 |
| 974 | 865 | 865 | 935 | 825 | 3,490 |
| 975 | 435 | 395 | 445 | 435 | 1,710 |
| 976 | 75 | 70 | 60 | 265 | 470 |
| 977 | 300 | 260 | 175 | 220 | 955 |
| 978 | 215 | 190 | 140 | 155 | 700 |
| 979 | 25 | 60 | 20 | 55 | 160 |
| TOTAL | 4,290 | 3,900 | 3,875 | 3,995 | 16,060 |

STATE: PENNSYLVANIA

| ZIP CODE | QUARTER 1 | QUARTER 2 | QUARTER 3 | QUARTER 4 | TOTAL GRAMS |
|---|---|---|---|---|---|
| 150 | 925 | 685 | 630 | 610 | 2,850 |
| 151 | 495 | 260 | 290 | 380 | 1,425 |
| 152 | 900 | 580 | 540 | 665 | 2,685 |
| 153 | 255 | 130 | 145 | 205 | 735 |
| 154 | 265 | 235 | 190 | 310 | 1,000 |
| 155 | 80 | 40 | 60 | 70 | 250 |
| 156 | 320 | 250 | 355 | 415 | 1,340 |
| 157 | 130 | 125 | 175 | 280 | 710 |
| 158 | 160 | 115 | 165 | 130 | 570 |
| 159 | 375 | 240 | 285 | 275 | 1,175 |
| 160 | 190 | 140 | 165 | 220 | 715 |
| 161 | 160 | 100 | 115 | 180 | 555 |
| 162 | 115 | 55 | 85 | 65 | 320 |
| 163 | 85 | 55 | 30 | 80 | 250 |
| 164 | 210 | 185 | 170 | 205 | 770 |
| 165 | 250 | 275 | 260 | 275 | 1,060 |
| 166 | 180 | 90 | 55 | 95 | 420 |
| 167 | 55 | 55 | 60 | 30 | 200 |
| 168 | 90 | 40 | 40 | 70 | 240 |
| 169 | 55 | 80 | 50 | 45 | 230 |
| 170 | 850 | 795 | 710 | 770 | 3,125 |
| 171 | 230 | 205 | 250 | 165 | 850 |
| 172 | 280 | 290 | 250 | 265 | 1,085 |
| 173 | 660 | 575 | 630 | 515 | 2,380 |
| 174 | 365 | 290 | 275 | 255 | 1,185 |
| 175 | 265 | 340 | 330 | 290 | 1,225 |
| 176 | 370 | 260 | 340 | 365 | 1,335 |
| 177 | 270 | 250 | 230 | 225 | 975 |
| 178 | 210 | 195 | 140 | 210 | 755 |
| 179 | 280 | 195 | 190 | 265 | 930 |
| 180 | 625 | 605 | 630 | 605 | 2,465 |
| 181 | 260 | 215 | 300 | 235 | 1,010 |
| 182 | 195 | 190 | 280 | 185 | 850 |
| 183 | 265 | 285 | 200 | 270 | 1,020 |
| 184 | 340 | 380 | 360 | 420 | 1,500 |

RETAIL DRUG DISTRIBUTION BY ZIP CODE WITHIN STATE BY GRAMS WT

| ZIP CODE | QUARTER 1 | QUARTER 2 | QUARTER 3 | QUARTER 4 | TOTAL GRAMS |
|---|---|---|---|---|---|
| 185 | 455 | 425 | 425 | 310 | 1,615 |
| 186 | 385 | 345 | 465 | 275 | 1,470 |
| 187 | 1,515 | 1,200 | 1,260 | 1,330 | 5,305 |
| 188 | 55 | 55 | 55 | 50 | 215 |
| 189 | 565 | 680 | 635 | 515 | 2,395 |
| 190 | 3,145 | 2,975 | 2,730 | 2,930 | 11,780 |
| 191 | 2,025 | 2,185 | 1,930 | 1,875 | 8,015 |
| 193 | 640 | 595 | 510 | 500 | 2,245 |
| 194 | 1,040 | 935 | 885 | 895 | 3,755 |
| 195 | 410 | 330 | 345 | 395 | 1,480 |
| 196 | 715 | 660 | 560 | 600 | 2,535 |
| TOTAL | 21,710 | 19,190 | 18,780 | 19,320 | 79,000 |

STATE: PUERTO RICO

| ZIP CODE | QUARTER 1 | QUARTER 2 | QUARTER 3 | QUARTER 4 | TOTAL GRAMS |
|---|---|---|---|---|---|
| 6 | 55 | 50 | 40 | 45 | 190 |
| 7 | 45 | 60 | 50 | 75 | 230 |
| 9 | 40 | 75 | 80 | 65 | 260 |
| TOTAL | 140 | 185 | 170 | 185 | 680 |

STATE: RHODE ISLAND

| ZIP CODE | QUARTER 1 | QUARTER 2 | QUARTER 3 | QUARTER 4 | TOTAL GRAMS |
|---|---|---|---|---|---|
| 28 | 710 | 565 | 625 | 595 | 2,495 |
| 29 | 485 | 410 | 360 | 440 | 1,695 |
| TOTAL | 1,195 | 975 | 985 | 1,035 | 4,190 |

STATE: SOUTH CAROLINA

| ZIP CODE | QUARTER 1 | QUARTER 2 | QUARTER 3 | QUARTER 4 | TOTAL GRAMS |
|---|---|---|---|---|---|
| 290 | 500 | 445 | 440 | 525 | 1,910 |
| 291 | 370 | 475 | 390 | 355 | 1,590 |
| 292 | 390 | 320 | 335 | 370 | 1,415 |
| 293 | 465 | 585 | 550 | 470 | 2,070 |
| 294 | 1,200 | 1,180 | 1,075 | 1,230 | 4,685 |
| 295 | 1,320 | 1,060 | 995 | 940 | 4,315 |
| 296 | 1,500 | 1,305 | 1,420 | 1,400 | 5,625 |
| 297 | 315 | 290 | 260 | 255 | 1,120 |
| 298 | 160 | 170 | 105 | 145 | 580 |
| 299 | 160 | 185 | 230 | 205 | 780 |
| TOTAL | 6,380 | 6,015 | 5,800 | 5,895 | 24,090 |

STATE: SOUTH DAKOTA

| ZIP CODE | QUARTER 1 | QUARTER 2 | QUARTER 3 | QUARTER 4 | TOTAL GRAMS |
|---|---|---|---|---|---|
| 570 | 135 | 100 | 110 | 60 | 405 |
| 571 | 145 | 190 | 150 | 175 | 660 |
| 572 | 90 | 75 | 105 | 60 | 330 |
| 573 | 70 | 85 | 65 | 110 | 330 |
| 574 | 180 | 200 | 190 | 115 | 685 |
| 575 | 65 | 70 | 55 | 75 | 265 |
| 576 | 100 | 80 | 55 | 95 | 330 |
| 577 | 205 | 305 | 260 | 190 | 960 |
| TOTAL | 990 | 1,105 | 990 | 880 | 3,965 |

STATE: TENNESSEE

| ZIP CODE | QUARTER 1 | QUARTER 2 | QUARTER 3 | QUARTER 4 | TOTAL GRAMS |
|---|---|---|---|---|---|
| 370 | 1,000 | 1,015 | 1,010 | 860 | 3,885 |
| 371 | 885 | 925 | 945 | 945 | 3,700 |
| 372 | 500 | 485 | 490 | 465 | 1,940 |
| 373 | 755 | 935 | 705 | 830 | 3,225 |
| 374 | 370 | 390 | 295 | 325 | 1,380 |
| 376 | 505 | 470 | 465 | 490 | 1,930 |

| | | | | | |
|---|---|---|---|---|---|
| 377 | 300 | 385 | 330 | 435 | 1,450 |
| 378 | 675 | 570 | 620 | 660 | 2,525 |
| 379 | 460 | 480 | 520 | 480 | 1,940 |
| 380 | 385 | 375 | 315 | 305 | 1,380 |
| 381 | 365 | 335 | 360 | 340 | 1,400 |
| 382 | 135 | 160 | 165 | 155 | 615 |
| 383 | 560 | 485 | 525 | 545 | 2,115 |
| 384 | 155 | 140 | 135 | 130 | 560 |
| 385 | 250 | 275 | 265 | 355 | 1,145 |
| TOTAL | 7,300 | 7,425 | 7,145 | 7,320 | 29,190 |

STATE: TEXAS

DRUG CODE:2165DRUG NAME:BUTALBITAL
STATE:TEXAS

| ZIP CODE | QUARTER 1 | QUARTER 2 | QUARTER 3 | QUARTER 4 | TOTAL GRAMS |
|---|---|---|---|---|---|
| 750 | 3,565 | 3,400 | 3,220 | 3,565 | 13,750 |
| 751 | 1,065 | 1,075 | 1,220 | 1,180 | 4,540 |
| 752 | 990 | 755 | 915 | 995 | 3,655 |
| 753 | 0 | 0 | 0 | 15 | 15 |
| 754 | 1,025 | 870 | 965 | 860 | 3,720 |
| 755 | 155 | 230 | 170 | 145 | 700 |
| 756 | 635 | 665 | 545 | 510 | 2,355 |
| 757 | 745 | 780 | 735 | 695 | 2,955 |
| 758 | 400 | 450 | 305 | 370 | 1,525 |
| 759 | 730 | 705 | 705 | 875 | 3,015 |
| 760 | 1,905 | 1,915 | 2,045 | 2,025 | 7,890 |
| 761 | 1,170 | 1,315 | 1,345 | 1,335 | 5,165 |
| 762 | 775 | 835 | 825 | 840 | 3,275 |
| 763 | 410 | 330 | 360 | 340 | 1,440 |
| 764 | 250 | 335 | 310 | 265 | 1,160 |
| 765 | 195 | 185 | 270 | 135 | 785 |
| 766 | 285 | 220 | 325 | 275 | 1,105 |
| 767 | 250 | 240 | 200 | 190 | 880 |
| 768 | 250 | 225 | 255 | 200 | 930 |
| 769 | 200 | 180 | 195 | 105 | 680 |
| 770 | 2,190 | 2,085 | 1,929.95 | 2,259.90 | 8,464.85 |
| 773 | 2,080 | 2,190 | 2,115 | 2,065 | 8,450 |
| 774 | 1,365 | 1,215 | 1,195 | 1,205 | 4,980 |
| 775 | 2,190 | 2,165 | 2,040 | 1,905 | 8,300 |
| 776 | 645 | 545 | 500 | 545 | 2,235 |
| 777 | 330 | 320 | 240 | 325 | 1,215 |
| 778 | 220 | 145 | 275 | 260 | 900 |
| 779 | 130 | 180 | 175 | 155 | 640 |
| 780 | 580 | 625 | 620 | 565 | 2,390 |
| 781 | 545 | 480 | 580 | 625 | 2,230 |
| 782 | 2,040 | 2,060 | 1,785 | 1,630 | 7,515 |
| 783 | 465 | 415 | 445 | 335 | 1,660 |
| 784 | 650 | 630 | 740 | 795 | 2,815 |
| 785 | 585 | 605 | 470 | 495 | 2,155 |
| 786 | 1,085 | 1,110 | 1,175 | 1,100 | 4,470 |
| 787 | 945 | 1,060 | 1,030 | 1,025 | 4,060 |
| 788 | 40 | 30 | 45 | 60 | 175 |
| 789 | 155 | 155 | 145 | 145 | 600 |
| 790 | 285 | 325 | 325 | 335 | 1,270 |
| 791 | 490 | 340 | 390 | 385 | 1,605 |
| 792 | 60 | 40 | 50 | 80 | 230 |
| 793 | 185 | 225 | 125 | 210 | 745 |
| 794 | 445 | 465 | 470 | 365 | 1,745 |
| 795 | 50 | 60 | 85 | 80 | 275 |

| | | | | | |
|---|---|---|---|---|---|
| 796 | 235 | 165 | 265 | 250 | 915 |
| 797 | 540 | 580 | 595 | 535 | 2,250 |
| 798 | 50 | 25 | 25 | 25 | 125 |
| 799 | 525 | 345 | 470 | 325 | 1,665 |
| TOTAL | 34,105 | 33,295 | 33,214.95 | 33,004.90 | 133,619.85 |

STATE: UTAH

| ZIP CODE | QUARTER 1 | QUARTER 2 | QUARTER 3 | QUARTER 4 | TOTAL GRAMS |
|---|---|---|---|---|---|
| 840 | 2,275 | 2,155 | 1,995 | 2,070 | 8,495 |
| 841 | 555 | 535 | 590 | 625 | 2,305 |
| 843 | 335 | 200 | 295 | 295 | 1,125 |
| 844 | 505 | 585 | 535 | 555 | 2,180 |
| 845 | 140 | 155 | 160 | 190 | 645 |
| 846 | 535 | 570 | 550 | 645 | 2,300 |
| 847 | 600 | 560 | 615 | 675 | 2,450 |
| TOTAL | 4,945 | 4,760 | 4,740 | 5,055 | 19,500 |

STATE: VERMONT

| ZIP CODE | QUARTER 1 | QUARTER 2 | QUARTER 3 | QUARTER 4 | TOTAL GRAMS |
|---|---|---|---|---|---|
| 50 | 5 | 15 | 10 | 0 | 30 |
| 51 | 15 | 15 | 5 | 15 | 50 |
| 52 | 65 | 85 | 70 | 30 | 250 |
| 53 | 5 | 0 | 5 | 0 | 10 |
| 54 | 60 | 35 | 75 | 50 | 220 |

DRUG CODE:2165DRUG NAME:BUTALBITAL
STATE:VERMONT

| ZIP CODE | QUARTER 1 | QUARTER 2 | QUARTER 3 | QUARTER 4 | TOTAL GRAMS |
|---|---|---|---|---|---|
| 56 | 5 | 5 | 30 | 15 | 55 |
| 57 | 85 | 110 | 110 | 80 | 385 |
| 58 | 45 | 45 | 30 | 20 | 140 |
| TOTAL | 285 | 310 | 335 | 210 | 1,140 |

STATE: VIRGIN ISLANDS

| ZIP CODE | QUARTER 1 | QUARTER 2 | QUARTER 3 | QUARTER 4 | TOTAL GRAMS |
|---|---|---|---|---|---|
| 8 | 50 | 25 | 20 | 20 | 115 |
| TOTAL | 50 | 25 | 20 | 20 | 115 |

STATE: VIRGINIA

| ZIP CODE | QUARTER 1 | QUARTER 2 | QUARTER 3 | QUARTER 4 | TOTAL GRAMS |
|---|---|---|---|---|---|
| 201 | 1,210 | 1,465 | 1,345 | 1,205 | 5,225 |
| 220 | 450 | 500 | 345 | 310 | 1,605 |
| 221 | 550 | 600 | 510 | 515 | 2,175 |
| 222 | 90 | 65 | 85 | 125 | 365 |
| 223 | 355 | 265 | 295 | 295 | 1,210 |
| 224 | 450 | 420 | 445 | 360 | 1,675 |
| 225 | 210 | 165 | 230 | 155 | 760 |
| 226 | 330 | 235 | 310 | 220 | 1,095 |
| 227 | 50 | 55 | 50 | 50 | 205 |
| 228 | 180 | 110 | 125 | 145 | 560 |
| 229 | 390 | 330 | 295 | 260 | 1,275 |
| 230 | 330 | 300 | 310 | 240 | 1,180 |
| 231 | 555 | 595 | 565 | 485 | 2,200 |
| 232 | 660 | 475 | 570 | 525 | 2,230 |
| 233 | 305 | 275 | 280 | 325 | 1,185 |
| 234 | 690 | 680 | 615 | 615 | 2,600 |
| 235 | 260 | 275 | 305 | 235 | 1,075 |
| 236 | 545 | 530 | 485 | 505 | 2,065 |
| 237 | 105 | 60 | 45 | 80 | 290 |

| | | | | | |
|---|---|---|---|---|---|
| 238 | 460 | 430 | 495 | 525 | 1,910 |
| 239 | 110 | 60 | 55 | 30 | 255 |
| 240 | 360 | 315 | 305 | 350 | 1,330 |
| 241 | 340 | 300 | 310 | 240 | 1,190 |
| 242 | 365 | 360 | 460 | 395 | 1,580 |
| 243 | 255 | 190 | 185 | 265 | 895 |
| 244 | 120 | 125 | 145 | 185 | 575 |
| 245 | 480 | 420 | 520 | 365 | 1,785 |
| 246 | 465 | 530 | 430 | 530 | 1,955 |
| TOTAL | 10,670 | 10,130 | 10,115 | 9,535 | 40,450 |

STATE: WASHINGTON

| ZIP CODE | QUARTER 1 | QUARTER 2 | QUARTER 3 | QUARTER 4 | TOTAL GRAMS |
|---|---|---|---|---|---|
| 980 | 1,295 | 1,300 | 1,385 | 1,395 | 5,375 |
| 981 | 630 | 645 | 735 | 545 | 2,555 |
| 982 | 1,115 | 940 | 900 | 1,020 | 3,975 |
| 983 | 720 | 735 | 860 | 790 | 3,105 |
| 984 | 400 | 440 | 590 | 600 | 2,030 |
| 985 | 685 | 795 | 550 | 600 | 2,630 |
| 986 | 625 | 620 | 575 | 585 | 2,405 |
| 988 | 180 | 180 | 240 | 205 | 805 |
| 989 | 290 | 290 | 240 | 375 | 1,195 |
| 990 | 115 | 225 | 85 | 75 | 500 |
| 991 | 140 | 180 | 125 | 145 | 590 |
| 992 | 480 | 435 | 410 | 470 | 1,795 |
| 993 | 535 | 580 | 610 | 590 | 2,315 |
| 994 | 70 | 65 | 80 | 55 | 270 |
| TOTAL | 7,280 | 7,430 | 7,385 | 7,450 | 29,545 |

STATE: WEST VIRGINIA

| ZIP CODE | QUARTER 1 | QUARTER 2 | QUARTER 3 | QUARTER 4 | TOTAL GRAMS |
|---|---|---|---|---|---|
| 247 | 40 | 40 | 45 | 55 | 180 |
| 248 | 25 | 20 | 0 | 25 | 70 |
| 249 | 25 | 55 | 65 | 40 | 185 |
| 250 | 50 | 20 | 55 | 50 | 175 |
| 251 | 25 | 55 | 55 | 35 | 170 |
| 252 | 85 | 75 | 120 | 170 | 450 |
| 253 | 185 | 130 | 140 | 160 | 615 |
| 254 | 220 | 175 | 185 | 160 | 740 |

DRUG CODE:2165DRUG NAME:BUTALBITAL
STATE:WEST VIRGINIA

| ZIP CODE | QUARTER 1 | QUARTER 2 | QUARTER 3 | QUARTER 4 | TOTAL GRAMS |
|---|---|---|---|---|---|
| 255 | 105 | 130 | 185 | 145 | 565 |
| 256 | 120 | 50 | 45 | 30 | 245 |
| 257 | 90 | 80 | 145 | 140 | 455 |
| 258 | 100 | 85 | 100 | 180 | 465 |
| 259 | 20 | 10 | 0 | 20 | 50 |
| 260 | 205 | 155 | 170 | 125 | 655 |
| 261 | 75 | 80 | 85 | 70 | 310 |
| 262 | 70 | 65 | 45 | 80 | 260 |
| 263 | 105 | 140 | 135 | 105 | 485 |
| 264 | 50 | 35 | 40 | 55 | 180 |
| 265 | 150 | 155 | 145 | 165 | 615 |
| 266 | 5 | 20 | 10 | 20 | 55 |
| 267 | 165 | 105 | 125 | 75 | 470 |
| 268 | 30 | 20 | 5 | 15 | 70 |
| TOTAL | 1,945 | 1,700 | 1,900 | 1,920 | 7,465 |

STATE: WISCONSIN

RETAIL DRUG DISTRIBUTION BY ZIP CODE WITHIN STATE BY GRAMS WT

| ZIP CODE | QUARTER 1 | QUARTER 2 | QUARTER 3 | QUARTER 4 | TOTAL GRAMS |
|---|---|---|---|---|---|
| 530 | 705 | 690 | 570 | 555 | 2,520 |
| 531 | 780 | 820 | 825 | 775 | 3,200 |
| 532 | 590 | 610 | 540 | 475 | 2,215 |
| 534 | 85 | 90 | 150 | 100 | 425 |
| 535 | 370 | 405 | 395 | 335 | 1,505 |
| 537 | 75 | 80 | 55 | 90 | 300 |
| 538 | 15 | 25 | 25 | 35 | 100 |
| 539 | 55 | 50 | 65 | 85 | 255 |
| 540 | 110 | 85 | 40 | 60 | 295 |
| 541 | 140 | 150 | 150 | 150 | 590 |
| 542 | 130 | 110 | 80 | 70 | 390 |
| 543 | 75 | 100 | 90 | 50 | 315 |
| 544 | 155 | 160 | 160 | 175 | 650 |
| 545 | 105 | 110 | 105 | 80 | 400 |
| 546 | 75 | 50 | 35 | 60 | 220 |
| 547 | 350 | 300 | 180 | 305 | 1,135 |
| 548 | 105 | 145 | 105 | 125 | 480 |
| 549 | 440 | 435 | 385 | 335 | 1,595 |
| TOTAL | 4,360 | 4,415 | 3,955 | 3,860 | 16,590 |

STATE: WYOMING

| ZIP CODE | QUARTER 1 | QUARTER 2 | QUARTER 3 | QUARTER 4 | TOTAL GRAMS |
|---|---|---|---|---|---|
| 820 | 70 | 70 | 45 | 70 | 255 |
| 822 | 50 | 65 | 50 | 55 | 220 |
| 823 | 10 | 5 | 0 | 5 | 20 |
| 824 | 110 | 175 | 185 | 215 | 685 |
| 825 | 90 | 75 | 60 | 65 | 290 |
| 826 | 170 | 125 | 125 | 120 | 540 |
| 827 | 20 | 20 | 15 | 15 | 70 |
| 828 | 185 | 215 | 225 | 155 | 780 |
| 829 | 115 | 125 | 135 | 160 | 535 |
| 830 | 0 | 5 | 0 | 0 | 5 |
| 831 | 35 | 25 | 40 | 30 | 130 |
| TOTAL | 855 | 905 | 880 | 890 | 3,530 |

DRUG CODE:2270DRUG NAME:PENTOBARBITAL (SCHEDULE 2)

STATE: ALABAMA

| ZIP CODE | QUARTER 1 | QUARTER 2 | QUARTER 3 | QUARTER 4 | TOTAL GRAMS |
|---|---|---|---|---|---|
| 350 | 799.75 | 1,421.78 | 2,754.16 | 622.03 | 5,597.72 |
| 351 | 2,488.12 | 1,599.51 | 1,155.20 | 799.75 | 6,042.58 |
| 352 | 1,642.33 | 1,954.13 | 1,358.80 | 1,741.77 | 6,697.03 |
| 354 | 88.86 | 1,066.34 | 0 | 0 | 1,155.20 |
| 355 | 0 | 533.17 | 533.17 | 0 | 1,066.34 |
| 356 | 544.56 | 0 | 799.75 | 3,732.18 | 5,076.49 |
| 357 | 1,155.20 | 444.31 | 977.48 | 2,310.40 | 4,887.39 |
| 358 | 18.23 | 266.58 | 266.58 | 0 | 551.39 |
| 359 | 1,066.34 | 2,308.75 | 2,665.85 | 0 | 6,040.94 |
| 360 | 1,065.24 | 1,598.41 | 1,420.68 | 2,042.71 | 6,127.04 |
| 361 | 0 | 2,665.85 | 0 | 2,665.85 | 5,331.70 |
| 362 | 0 | 0 | 177.72 | 888.62 | 1,066.34 |

DRUG CODE:2270DRUG NAME:PENTOBARBITAL (SCHEDULE 2)
STATE:ALABAMA

| ZIP CODE | QUARTER 1 | QUARTER 2 | QUARTER 3 | QUARTER 4 | TOTAL GRAMS |
|---|---|---|---|---|---|
| 363 | 977.48 | 1,421.78 | 710.89 | 583.3 | 3,693.45 |
| 364 | 532.62 | 532.62 | 88.86 | 799.2 | 1,953.30 |
| 365 | 1,954.95 | 1,421.78 | 3,196.81 | 3,199.01 | 9,772.55 |
| 366 | 3,111.96 | 360 | 3,556.27 | 4.56 | 7,032.79 |

| | | | | | | |
|---|---|---|---|---|---|---|
| | 367 | 444.31 | 0 | 355.45 | 88.86 | 888.62 |
| | 368 | 1,599.51 | 622.03 | 533.17 | 977.48 | 3,732.19 |
| TOTAL | | 17,489.46 | 18,217.04 | 20,550.84 | 20,455.72 | 76,713.06 |
| STATE: | ALASKA | | | | | |
| ZIP CODE | QUARTER 1 | | QUARTER 2 | QUARTER 3 | QUARTER 4 | TOTAL GRAMS |
| | 995 | 2,711.42 | 1,800.93 | 3,474.71 | 1,599.51 | 9,586.57 |
| | 996 | 622.03 | 533.17 | 622.03 | 1,332.92 | 3,110.15 |
| | 997 | 1,332.92 | 355.45 | 710.89 | 1,599.51 | 3,998.77 |
| | 998 | 355.45 | 533.17 | 88.86 | 266.58 | 1,244.06 |
| TOTAL | | 5,021.82 | 3,222.72 | 4,896.49 | 4,798.52 | 17,939.55 |
| STATE: | ARIZONA | | | | | |
| ZIP CODE | QUARTER 1 | | QUARTER 2 | QUARTER 3 | QUARTER 4 | TOTAL GRAMS |
| | 850 | 6,653.22 | 7,080.75 | 3,988.74 | 8,378.04 | 26,100.75 |
| | 851 | 3,732.18 | 1,244.06 | 3,640.02 | 177.72 | 8,793.98 |
| | 852 | 12,309.55 | 13,460.09 | 9,155.01 | 10,128.56 | 45,053.21 |
| | 853 | 10,929.96 | 15,728.49 | 8,011.21 | 12,796.06 | 47,465.72 |
| | 855 | 533.17 | 266.58 | 1,604.98 | 0 | 2,404.73 |
| | 856 | 888.62 | 0 | 1,777.23 | 2,579.72 | 5,245.57 |
| | 857 | 5,720.86 | 5,324.40 | 6,965.37 | 10,465.15 | 28,475.78 |
| | 859 | 355.45 | 888.62 | 0 | 1,066.34 | 2,310.41 |
| | 860 | 1,604.06 | 890.42 | 1,066.34 | 182.28 | 3,743.10 |
| | 863 | 3,376.74 | 3,909.91 | 1,244.06 | 2,932.43 | 11,463.14 |
| | 864 | 1,597.31 | 2,043.81 | 444.31 | 533.17 | 4,618.60 |
| TOTAL | | 47,701.12 | 50,837.13 | 37,897.27 | 49,239.47 | 185,674.99 |
| STATE: | ARKANSAS | | | | | |
| ZIP CODE | QUARTER 1 | | QUARTER 2 | QUARTER 3 | QUARTER 4 | TOTAL GRAMS |
| | 717 | 0 | 0 | 533.17 | 177.17 | 710.34 |
| | 718 | 533.17 | 533.17 | 0 | 533.17 | 1,599.51 |
| | 719 | 799.75 | 2,132.68 | 622.03 | 1,599.51 | 5,153.97 |
| | 720 | 1,066.34 | 444.31 | 1,599.51 | 444.31 | 3,554.47 |
| | 721 | 1,779.41 | 2,221.54 | 1,066.34 | 885.87 | 5,953.16 |
| | 722 | 1,084.57 | 0 | 270.32 | 0 | 1,354.89 |
| | 723 | 0 | 0 | 888.62 | 533.17 | 1,421.79 |
| | 724 | 355.45 | 88.86 | 444.31 | 977.48 | 1,866.10 |
| | 726 | 266.03 | 1,334.75 | 0 | 534.44 | 2,135.22 |
| | 727 | 532.89 | 1,154.92 | 1,246.80 | 888.34 | 3,822.95 |
| | 729 | 2,665.85 | 0 | 710.89 | 0 | 3,376.74 |
| TOTAL | | 9,083.46 | 7,910.23 | 7,381.99 | 6,573.46 | 30,949.14 |
| STATE: | CALIFORNIA | | | | | |
| ZIP CODE | QUARTER 1 | | QUARTER 2 | QUARTER 3 | QUARTER 4 | TOTAL GRAMS |
| | 900 | 2,804.83 | 2,727.36 | 3,106.05 | 2,300.37 | 10,938.61 |
| | 902 | 906.84 | 533.17 | 7,119.86 | 1,189.83 | 9,749.70 |
| | 904 | 0 | 36.46 | 16.41 | 11.39 | 64.26 |
| | 905 | 360.91 | 189.57 | 1,765.84 | 34.18 | 2,350.50 |
| | 906 | 1,066.34 | 1,510.65 | 0 | 1,068.62 | 3,645.61 |
| | 907 | 354.35 | 355.45 | 533.17 | 887.52 | 2,130.49 |
| | 908 | 159.5 | 364.56 | 113.93 | 371.4 | 1,009.39 |
| | 910 | 1,444.57 | 498.99 | 1,225.83 | 799.75 | 3,969.14 |
| | 911 | 2,132.68 | 0 | 0 | 0 | 2,132.68 |
| | 912 | 1,510.65 | 0 | 1,510.65 | 0 | 3,021.30 |
| | 913 | 4,002.41 | 7,380.97 | 4,105.86 | 6,056.71 | 21,545.95 |
| | 914 | 0.91 | 355.45 | 444.31 | 446.59 | 1,247.26 |
| | 915 | 0 | 0 | 22.79 | 0 | 22.79 |
| | 916 | 177.72 | 266.58 | 88.86 | 0 | 533.16 |
| | 917 | 1,431.81 | 6,935.75 | 7,393.73 | 1,421.78 | 17,183.07 |
| | 918 | 0 | 177.72 | 0 | 0 | 177.72 |

| | | | | | |
|---|---|---|---|---|---|
| 919 | 1,866.09 | 1,066.34 | 1,421.78 | 2,488.12 | 6,842.33 |
| 920 | 8,620.48 | 7,642.09 | 6,933.48 | 6,600.81 | 29,796.86 |
| 921 | 1,068.16 | 2,981.65 | 3,447.37 | 277.98 | 7,775.16 |
| 922 | 3,110.15 | 8,441.84 | 2,761.54 | 3,732.18 | 18,045.71 |
| 923 | 6,615.66 | 7,748.53 | 4,972.41 | 9,755.99 | 29,092.59 |
| 924 | 2,399.26 | 4,643.58 | 2,132.68 | 4,354.21 | 13,529.73 |

DRUG CODE:2270DRUG NAME:PENTOBARBITAL (SCHEDULE 2)
STATE:CALIFORNIA

| ZIP CODE | QUARTER 1 | QUARTER 2 | QUARTER 3 | QUARTER 4 | TOTAL GRAMS |
|---|---|---|---|---|---|
| 925 | 14,432.02 | 17,003.08 | 14,962.91 | 21,533.19 | 67,931.20 |
| 926 | 177.72 | 1,688.37 | 416.97 | 1,512.47 | 3,795.53 |
| 927 | 419.24 | 268.86 | 36.46 | 152.66 | 877.22 |
| 928 | 3,196.74 | 6,842.34 | 3,137.49 | 8,626.40 | 21,802.97 |
| 930 | 2,044.54 | 1,244.06 | 890.44 | 1,867.91 | 6,046.95 |
| 931 | 444.31 | 291.65 | 533.17 | 202.79 | 1,471.92 |
| 932 | 5,523.08 | 8,531.62 | 9,788.44 | 5,420.55 | 29,263.69 |
| 933 | 6,842.34 | 7,464.37 | 10,399.07 | 13,705.18 | 38,410.96 |
| 934 | 2,488.12 | 1,421.78 | 799.75 | 3,465.60 | 8,175.25 |
| 935 | 10,041.35 | 2,399.26 | 14,128.98 | 2,132.68 | 28,702.27 |
| 936 | 3,110.15 | 2,132.68 | 1,647.81 | 2,510.91 | 9,401.55 |
| 937 | 10,130.21 | 1,689.28 | 8,975.01 | 266.58 | 21,061.08 |
| 939 | 3,385.85 | 1,711.15 | 4,345.10 | 3,411.83 | 12,853.93 |
| 940 | 799.75 | 2,576.98 | 710.89 | 1,244.06 | 5,331.68 |
| 941 | 300.76 | 1,180.72 | 432 | 57.87 | 1,971.35 |
| 943 | 1,080.01 | 3.65 | 48.76 | 444.31 | 1,576.73 |
| 944 | 533.17 | 177.72 | 710.89 | 3,554.46 | 4,976.24 |
| 945 | 2,844.02 | 4,355.58 | 4,001.96 | 5,415.05 | 16,616.61 |
| 946 | 18.23 | 1,085.02 | 888.62 | 18.23 | 2,010.10 |
| 947 | 177.72 | 533.17 | 888.62 | 1,421.78 | 3,021.29 |
| 949 | 2,160.02 | 4,975.97 | 5,153.69 | 3,820.77 | 16,110.45 |
| 950 | 2,932.43 | 3,734.01 | 3,734.01 | 7,385.07 | 17,785.52 |
| 951 | 88.59 | 188.38 | 2,357.97 | 2,348.22 | 4,983.16 |
| 952 | 2,044.73 | 710.89 | 3,021.29 | 1,244.06 | 7,020.97 |
| 953 | 6,665.98 | 6,397.56 | 12,795.59 | 12,441.88 | 38,301.01 |
| 954 | 3,736.28 | 4,176.49 | 4,443.08 | 1,688.37 | 14,044.22 |
| 955 | 710.89 | 3,021.29 | 1,866.09 | 1,066.34 | 6,664.61 |
| 956 | 8,501.81 | 8,097.33 | 9,735.39 | 12,734.54 | 39,069.07 |
| 957 | 177.72 | 0 | 0 | 0 | 177.72 |
| 958 | 73.82 | 4,488.65 | 4,703.74 | 100.25 | 9,366.46 |
| 959 | 9,065.13 | 6,226.23 | 5,152.87 | 3,754.97 | 24,199.20 |
| 960 | 5,509.41 | 3,110.15 | 8,887.52 | 6,593.98 | 24,101.06 |
| 961 | 1,155.20 | 1,244.06 | 2,132.68 | 3,199.01 | 7,730.95 |
| TOTAL | 150,844.66 | 162,829.06 | 190,843.81 | 175,139.40 | 679,656.93 |

STATE: COLORADO

| ZIP CODE | QUARTER 1 | QUARTER 2 | QUARTER 3 | QUARTER 4 | TOTAL GRAMS |
|---|---|---|---|---|---|
| 800 | 3,823.05 | 4,472.70 | 2,273.03 | 1,864.72 | 12,433.50 |
| 801 | 2,576.98 | 5,201.82 | 3,732.18 | 5,082.60 | 16,593.58 |
| 802 | 5,812.45 | 2,148.63 | 2,834.45 | 2,234.30 | 13,029.83 |
| 803 | 462.54 | 1,332.92 | 355.45 | 355.45 | 2,506.36 |
| 804 | 3,199.01 | 1,421.78 | 6,486.89 | 3,110.15 | 14,217.83 |
| 805 | 7,374.40 | 3,376.74 | 8,620.02 | 5,864.31 | 25,235.47 |
| 806 | 2,576.98 | 2,043.81 | 2,221.54 | 1,066.34 | 7,908.67 |
| 807 | 266.58 | 355.45 | 0 | 266.58 | 888.61 |
| 808 | 533.17 | 1,332.92 | 0 | 799.75 | 2,665.84 |
| 809 | 724.56 | 2,932.43 | 2,488.12 | 355.45 | 6,500.56 |
| 810 | 2,399.26 | 1,954.95 | 2,576.98 | 1,511.56 | 8,442.75 |
| 811 | 177.72 | 710.89 | 355.45 | 0 | 1,244.06 |
| 812 | 889.06 | 444.31 | 977.48 | 622.03 | 2,932.88 |

| | | | | | |
|---|---|---|---|---|---|
| 813 | 1,777.23 | 2,043.81 | 622.03 | 1,510.65 | 5,953.72 |
| 814 | 1,333.56 | 1,421.23 | 2,132.68 | 2,843.57 | 7,731.04 |
| 815 | 2,576.98 | 2,665.85 | 710.89 | 2,488.12 | 8,441.84 |
| 816 | 1,954.95 | 1,688.37 | 622.03 | 1,777.23 | 6,042.58 |
| TOTAL | 38,458.48 | 35,548.61 | 37,009.22 | 31,752.81 | 142,769.12 |

STATE: CONNECTICUT

| ZIP CODE | QUARTER 1 | QUARTER 2 | QUARTER 3 | QUARTER 4 | TOTAL GRAMS |
|---|---|---|---|---|---|
| 60 | 977.2 | 2,678.95 | 6,034.33 | 3,640.02 | 13,330.50 |
| 61 | 635.7 | 558.23 | 1,444.57 | 678.99 | 3,317.49 |
| 62 | 710.89 | 2,399.26 | 2,310.40 | 1,332.92 | 6,753.47 |
| 63 | 533.17 | 0 | 1,510.65 | 88.86 | 2,132.68 |
| 64 | 5,420.55 | 5,953.72 | 5,953.72 | 5,152.87 | 22,480.86 |
| 65 | 6.38 | 499.9 | 0.91 | 7.29 | 514.48 |
| 66 | 97.98 | 177.72 | 275.7 | 9.11 | 560.51 |
| 67 | 3,287.60 | 710.34 | 2,488.12 | 3,642.77 | 10,128.83 |
| 68 | 0 | 533.17 | 710.89 | 0 | 1,244.06 |
| 69 | 0 | 0 | 0 | 1,066.34 | 1,066.34 |

DRUG CODE:2270DRUG NAME:PENTOBARBITAL (SCHEDULE 2)
STATE:CONNECTICUT

| ZIP CODE | QUARTER 1 | QUARTER 2 | QUARTER 3 | QUARTER 4 | TOTAL GRAMS |
|---|---|---|---|---|---|
| TOTAL | 11,669.47 | 13,511.29 | 20,729.29 | 15,619.17 | 61,529.22 |

STATE: DELAWARE

| ZIP CODE | QUARTER 1 | QUARTER 2 | QUARTER 3 | QUARTER 4 | TOTAL GRAMS |
|---|---|---|---|---|---|
| 197 | 31.9 | 0 | 0.91 | 31.9 | 64.71 |
| 198 | 0 | 54.68 | 173.17 | 288.46 | 516.31 |
| 199 | 1,420.13 | 2,129.38 | 1,155.20 | 2,042.17 | 6,746.88 |
| TOTAL | 1,452.03 | 2,184.06 | 1,329.28 | 2,362.53 | 7,327.90 |

STATE: DISTRICT OF COLUMBIA

| ZIP CODE | QUARTER 1 | QUARTER 2 | QUARTER 3 | QUARTER 4 | TOTAL GRAMS |
|---|---|---|---|---|---|
| 200 | 152.66 | 246.08 | 209.62 | 284.81 | 893.17 |
| TOTAL | 152.66 | 246.08 | 209.62 | 284.81 | 893.17 |

STATE: FLORIDA

| ZIP CODE | QUARTER 1 | QUARTER 2 | QUARTER 3 | QUARTER 4 | TOTAL GRAMS |
|---|---|---|---|---|---|
| 320 | 1,866.55 | 2,132.68 | 1,688.82 | 1,244.06 | 6,932.11 |
| 321 | 568.71 | 5,321.66 | 1,973.18 | 721.83 | 8,585.38 |
| 322 | 501.27 | 1,222.45 | 1,598.41 | 821.44 | 4,143.57 |
| 323 | 2,043.81 | 2,676.78 | 2,843.57 | 3,116.53 | 10,680.69 |
| 324 | 2,313.13 | 4,354.21 | 799.75 | 3,199.01 | 10,666.10 |
| 325 | 4,976.24 | 5,420.55 | 799.75 | 2,399.26 | 13,595.80 |
| 326 | 624.31 | 2,374.20 | 799.75 | 1,130.14 | 4,928.40 |
| 327 | 3,024.03 | 3,921.47 | 3,019.92 | 2,224.27 | 12,189.69 |
| 328 | 842.04 | 5,112.95 | 3,011.08 | 4,901.05 | 13,867.12 |
| 329 | 3,425.04 | 3,584.08 | 3,375.36 | 3,732.18 | 14,116.66 |
| 330 | 49.22 | 710.89 | 534.99 | 779.25 | 2,074.35 |
| 331 | 185.01 | 953.78 | 139.62 | 8,915.77 | 10,194.18 |
| 333 | 3,950.92 | 6,400.31 | 5,074.22 | 3,221.80 | 18,647.25 |
| 334 | 1,034.44 | 10,307.11 | 514.39 | 1,423.51 | 13,279.45 |
| 335 | 1,953.30 | 977.48 | 1,954.95 | 2,220.71 | 7,106.44 |
| 336 | 4,623.53 | 850.34 | 3,390.41 | 138.99 | 9,003.27 |
| 337 | 856.72 | 7,781.08 | 694.94 | 3,040.42 | 12,373.16 |
| 338 | 800.11 | 9,152.46 | 1,066.34 | 669.6 | 11,688.51 |
| 339 | 2,603.78 | 2,132.68 | 9,685.08 | 2,488.12 | 16,909.66 |
| 341 | 389.62 | 2,314.04 | 488.05 | 88.86 | 3,280.57 |
| 342 | 3,831.98 | 1,688.37 | 177.72 | 4,279.93 | 9,978 |

| | | | | | |
|---|---|---|---|---|---|
| 344 | 14,213.72 | 4,264.53 | 3,286.78 | 5,643.02 | 27,408.05 |
| 346 | 3,020.65 | 2,932.43 | 1,244.06 | 4,619.70 | 11,816.84 |
| 347 | 444.31 | 5,153.42 | 1,687.82 | 2,043.26 | 9,328.81 |
| 349 | 2,489.94 | 7,663.96 | 977.48 | 1,813.69 | 12,945.07 |
| TOTAL | 60,632.38 | 99,403.91 | 50,826.44 | 64,876.40 | 275,739.13 |

STATE: GEORGIA

| ZIP CODE | QUARTER 1 | QUARTER 2 | QUARTER 3 | QUARTER 4 | TOTAL GRAMS |
|---|---|---|---|---|---|
| 300 | 7,295.68 | 7,104.52 | 4,612 | 2,403.82 | 21,416.02 |
| 301 | 1,688.37 | 4,894.67 | 4,086.25 | 2,932.43 | 13,601.72 |
| 302 | 1,952.20 | 177.72 | 1,685.62 | 1,063.59 | 4,879.13 |
| 303 | 422.89 | 959.7 | 713.17 | 2,307.21 | 4,402.97 |
| 304 | 533.17 | 533.17 | 888.62 | 710.89 | 2,665.85 |
| 305 | 443.76 | 3,376.74 | 2,932.43 | 355.45 | 7,108.38 |
| 306 | 3,465.05 | 3,728.33 | 2,840.27 | 2,931.88 | 12,965.53 |
| 307 | 4,618.60 | 622.03 | 4,440.88 | 1,419.58 | 11,101.09 |
| 308 | 0 | 266.58 | 6,398.03 | 0 | 6,664.61 |
| 309 | 2,399.26 | 227.85 | 2,576.98 | 177.72 | 5,381.81 |
| 310 | 1,421.78 | 798.1 | 1,332.10 | 3,109.05 | 6,661.03 |
| 312 | 372.76 | 1,960.42 | 2,492.68 | 15.49 | 4,841.35 |
| 313 | 177.72 | 1,954.95 | 266.58 | 2,310.40 | 4,709.65 |
| 314 | 303.04 | 88.59 | 355.17 | 341.5 | 1,088.30 |
| 315 | 1,686.72 | 2,754.71 | 2,219.89 | 1,332.92 | 7,994.24 |
| 316 | 1,071.81 | 0 | 1,071.81 | 0 | 2,143.62 |
| 317 | 1,599.51 | 5,953.72 | 1,777.23 | 4,887.38 | 14,217.84 |
| 318 | 709.24 | 0 | 531.52 | 0 | 1,240.76 |
| 319 | 546.38 | 546.84 | 799.75 | 1,347.50 | 3,240.47 |
| 398 | 88.86 | 88.86 | 0 | 88.86 | 266.58 |
| TOTAL | 30,796.80 | 36,037.50 | 42,020.98 | 27,735.67 | 136,590.95 |

STATE: GUAM

| ZIP CODE | QUARTER 1 | QUARTER 2 | QUARTER 3 | QUARTER 4 | TOTAL GRAMS |
|---|---|---|---|---|---|
| 969 | 0 | 3,215.42 | 0 | 0 | 3,215.42 |
| TOTAL | 0 | 3,215.42 | 0 | 0 | 3,215.42 |

DRUG CODE:2270DRUG NAME:PENTOBARBITAL (SCHEDULE 2)
STATE: HAWAII

| ZIP CODE | QUARTER 1 | QUARTER 2 | QUARTER 3 | QUARTER 4 | TOTAL GRAMS |
|---|---|---|---|---|---|
| 967 | 6,606.74 | 3,482 | 5,525.36 | 3,654.26 | 19,268.36 |
| 968 | 9,164.13 | 186.84 | 334.94 | 9,176.89 | 18,862.80 |
| TOTAL | 15,770.87 | 3,668.84 | 5,860.30 | 12,831.15 | 38,131.16 |

STATE: IDAHO

| ZIP CODE | QUARTER 1 | QUARTER 2 | QUARTER 3 | QUARTER 4 | TOTAL GRAMS |
|---|---|---|---|---|---|
| 832 | 1,599.51 | 1,602.24 | 802.49 | 3,201.75 | 7,205.99 |
| 833 | 1,066.34 | 2,399.26 | 3,465.60 | 4,087.63 | 11,018.83 |
| 834 | 2,754.71 | 3,821.04 | 4,443.08 | 3,998.77 | 15,017.60 |
| 835 | 533.17 | 1,421.78 | 2,754.71 | 1,866.09 | 6,575.75 |
| 836 | 8,813.69 | 3,734.01 | 7,737.33 | 9,421.49 | 29,706.52 |
| 837 | 838.94 | 2,535.51 | 570.54 | 5,066.47 | 9,011.46 |
| 838 | 4,265.87 | 3,026.76 | 2,665.85 | 3,021.29 | 12,979.77 |
| TOTAL | 19,872.23 | 18,540.60 | 22,439.60 | 30,663.49 | 91,515.92 |

STATE: ILLINOIS

| ZIP CODE | QUARTER 1 | QUARTER 2 | QUARTER 3 | QUARTER 4 | TOTAL GRAMS |
|---|---|---|---|---|---|
| 600 | 10,543.99 | 13,498.98 | 10,945.72 | 12,145.68 | 47,134.37 |
| 601 | 4,002.59 | 5,864.58 | 3,820.49 | 2,907.37 | 16,595.03 |
| 602 | 265.76 | 0 | 0 | 266.58 | 532.34 |
| 603 | 0 | 0 | 531.52 | 0 | 531.52 |

| | | | | | |
|---|---|---|---|---|---|
| 604 | 3,999.03 | 6,075.39 | 7,510.20 | 1,422.70 | 19,007.32 |
| 605 | 4,901.96 | 2,232.20 | 3,200.84 | 2,245.60 | 12,580.60 |
| 606 | 11,114.43 | 5,366.78 | 7,678.09 | 4,560.19 | 28,719.49 |
| 607 | 88.86 | 533.17 | 0 | 177.72 | 799.75 |
| 608 | 0 | 88.86 | 0 | 0 | 88.86 |
| 609 | 888.62 | 886.42 | 1,599.51 | 177.72 | 3,552.27 |
| 610 | 2,221.54 | 3,909.91 | 3,554.46 | 5,951.52 | 15,637.43 |
| 611 | 1,077.73 | 2,662.55 | 5,331.42 | 2,132.68 | 11,204.38 |
| 612 | 1,244.06 | 355.45 | 0 | 533.17 | 2,132.68 |
| 613 | 3,369.59 | 0 | 1,240.21 | 531.52 | 5,141.32 |
| 614 | 3,293.34 | 2,756.07 | 1,421.78 | 1,512.47 | 8,983.66 |
| 615 | 710.89 | 622.03 | 266.58 | 1,155.20 | 2,754.70 |
| 616 | 378.23 | 1,244.06 | 2,233.84 | 387.35 | 4,243.48 |
| 617 | 1,244.06 | 1,066.34 | 1,066.34 | 355.45 | 3,732.19 |
| 618 | 3,826.86 | 2,319.51 | 4,620.80 | 5,509.41 | 16,276.58 |
| 619 | 177.72 | 533.17 | 799.75 | 709.24 | 2,219.88 |
| 620 | 710.89 | 2,488.12 | 1,066.34 | 3,021.29 | 7,286.64 |
| 622 | 3,554.46 | 1,866.09 | 3,199.01 | 3,909.91 | 12,529.47 |
| 623 | 799.75 | 4,265.35 | 88.86 | 710.89 | 5,864.85 |
| 624 | 0 | 177.72 | 1,954.40 | 2,843.57 | 4,975.69 |
| 625 | 888.62 | 977.48 | 355.45 | 622.03 | 2,843.58 |
| 626 | 1,421.23 | 2,042.17 | 1,687.82 | 1,419.58 | 6,570.80 |
| 627 | 9.11 | 2,416.94 | 275.15 | 112.56 | 2,813.76 |
| 628 | 1,244.06 | 887.52 | 2,488.12 | 355.45 | 4,975.15 |
| 629 | 977.48 | 3,821.04 | 533.17 | 1,421.78 | 6,753.47 |
| TOTAL | 62,954.86 | 68,957.90 | 67,469.87 | 57,098.63 | 256,481.26 |

STATE: INDIANA

| ZIP CODE | QUARTER 1 | QUARTER 2 | QUARTER 3 | QUARTER 4 | TOTAL GRAMS |
|---|---|---|---|---|---|
| 460 | 2,140.97 | 3,029.03 | 3,109.33 | 3,286.23 | 11,565.56 |
| 461 | 4,353.94 | 1,601.79 | 2,667.85 | 1,959.51 | 10,583.09 |
| 462 | 879.5 | 1,540.08 | 1,311.50 | 745.53 | 4,476.61 |
| 463 | 2,219.61 | 3,375.64 | 3,195.17 | 3,109.33 | 11,899.75 |
| 464 | 13.67 | 0 | 0 | 888.62 | 902.29 |
| 465 | 8,086.40 | 3,287.88 | 4,887.38 | 5,420.55 | 21,682.21 |
| 466 | 355.45 | 638.44 | 1,355.71 | 177.72 | 2,527.32 |
| 467 | 3,021.29 | 2,665.85 | 3,110.15 | 1,244.06 | 10,041.35 |
| 468 | 2,310.85 | 2,576.98 | 5,798.78 | 1,533.89 | 12,220.50 |
| 469 | 3,640.57 | 2,488.12 | 2,132.68 | 3,462.85 | 11,724.22 |
| 471 | 979.76 | 2,937.90 | 1,421.78 | 4,095.38 | 9,434.82 |
| 472 | 799.48 | 1,332.92 | 2,487.57 | 2,398.71 | 7,018.68 |
| 473 | 888.62 | 3,640.02 | 1,601.79 | 2,129.38 | 8,259.81 |
| 474 | 2,962.68 | 1,864.44 | 2,399.26 | 1,243.24 | 8,469.62 |
| 475 | 177.72 | 355.45 | 533.17 | 1,242.41 | 2,308.75 |
| 476 | 1,506.52 | 1,153.82 | 799.48 | 354.62 | 3,814.44 |
| 477 | 177.72 | 2,843.57 | 0 | 533.17 | 3,554.46 |
| 478 | 2,129.38 | 533.17 | 3,373.44 | 1,064.69 | 7,100.68 |
| 479 | 1,331.27 | 1,602.41 | 265.76 | 2,127.73 | 5,327.17 |

DRUG CODE:2270DRUG NAME:PENTOBARBITAL (SCHEDULE 2)
STATE:INDIANA

| ZIP CODE | QUARTER 1 | QUARTER 2 | QUARTER 3 | QUARTER 4 | TOTAL GRAMS |
|---|---|---|---|---|---|
| TOTAL | 37,975.40 | 37,467.51 | 40,450.80 | 37,017.62 | 152,911.33 |

STATE: IOWA

| ZIP CODE | QUARTER 1 | QUARTER 2 | QUARTER 3 | QUARTER 4 | TOTAL GRAMS |
|---|---|---|---|---|---|
| 500 | 9,063.87 | 533.17 | 6,931.20 | 6,309.17 | 22,837.41 |
| 501 | 535.9 | 710.89 | 1,067.25 | 712.71 | 3,026.75 |
| 502 | 1,066.34 | 179.55 | 177.72 | 622.03 | 2,045.64 |
| 503 | 622.03 | 2,759.26 | 2,133.59 | 905.02 | 6,419.90 |

| | | | | | |
|---|---|---|---|---|---|
| 504 | 179 | 3.65 | 179 | 179 | 540.65 |
| 505 | 444.31 | 0 | 0 | 0 | 444.31 |
| 506 | 1,332.92 | 710.89 | 266.58 | 1,071.81 | 3,382.20 |
| 507 | 1,066.34 | 0 | 0 | 1,066.34 | 2,132.68 |
| 508 | 355.45 | 0 | 0 | 355.45 | 710.9 |
| 510 | 0 | 177.72 | 177.72 | 0 | 355.44 |
| 511 | 0 | 710.89 | 622.94 | 88.86 | 1,422.69 |
| 513 | 355.45 | 533.17 | 888.62 | 0 | 1,777.24 |
| 515 | 0 | 0 | 1,688.37 | 0 | 1,688.37 |
| 516 | 535.9 | 714.54 | 0 | 0 | 1,250.44 |
| 520 | 710.89 | 0 | 1,510.65 | 177.72 | 2,399.26 |
| 521 | 177.72 | 0 | 0 | 0 | 177.72 |
| 522 | 91.14 | 492.16 | 4.56 | 1,029.88 | 1,617.74 |
| 523 | 177.72 | 533.17 | 444.31 | 0 | 1,155.20 |
| 524 | 1,244.06 | 1,777.23 | 1,332.37 | 1,688.37 | 6,042.03 |
| 525 | 2,488.12 | 533.17 | 888.62 | 1,066.34 | 4,976.25 |
| 526 | 533.17 | 1,421.78 | 3,199.01 | 177.72 | 5,331.68 |
| 527 | 533.17 | 0 | 0 | 355.45 | 888.62 |
| 528 | 177.72 | 177.72 | 1,066.34 | 1,332.92 | 2,754.70 |
| TOTAL | 21,691.22 | 11,968.96 | 22,578.85 | 17,138.79 | 73,377.82 |

STATE: KANSAS

| ZIP CODE | QUARTER 1 | QUARTER 2 | QUARTER 3 | QUARTER 4 | TOTAL GRAMS |
|---|---|---|---|---|---|
| 660 | 1,066.34 | 4,443.08 | 1,599.51 | 3,821.04 | 10,929.97 |
| 661 | 0 | 0 | 0 | 533.17 | 533.17 |
| 662 | 282.99 | 277.52 | 888.62 | 544.11 | 1,993.24 |
| 664 | 0 | 1,066.34 | 177.72 | 1,954.95 | 3,199.01 |
| 665 | 8,086.40 | 177.72 | 1,066.34 | 5,509.41 | 14,839.87 |
| 666 | 177.72 | 2,932.43 | 177.72 | 977.48 | 4,265.35 |
| 667 | 0 | 533.17 | 0 | 710.89 | 1,244.06 |
| 668 | 0 | 444.31 | 0 | 0 | 444.31 |
| 669 | 0 | 177.72 | 0 | 0 | 177.72 |
| 670 | 977.48 | 1,244.06 | 5,342.63 | 1,332.92 | 8,897.09 |
| 671 | 888.62 | 2,399.26 | 1,599.51 | 1,688.37 | 6,575.76 |
| 672 | 1,200.77 | 4,759.79 | 4,488.65 | 2,907.37 | 13,356.58 |
| 673 | 177.72 | 355.17 | 0 | 533.17 | 1,066.06 |
| 674 | 2,399.26 | 1,954.95 | 799.75 | 355.45 | 5,509.41 |
| 675 | 3,643.32 | 177.72 | 710.89 | 88.86 | 4,620.79 |
| 676 | 357.27 | 266.58 | 357.27 | 0 | 981.12 |
| 677 | 0 | 0 | 177.72 | 0 | 177.72 |
| 678 | 2,665.85 | 355.45 | 88.86 | 1,954.95 | 5,065.11 |
| 679 | 0 | 266.58 | 0 | 0 | 266.58 |
| TOTAL | 21,923.74 | 21,831.85 | 17,475.19 | 22,912.14 | 84,142.92 |

STATE: KENTUCKY

| ZIP CODE | QUARTER 1 | QUARTER 2 | QUARTER 3 | QUARTER 4 | TOTAL GRAMS |
|---|---|---|---|---|---|
| 400 | 977.48 | 444.31 | 1,155.20 | 977.48 | 3,554.47 |
| 401 | 0 | 177.72 | 177.72 | 1,510.65 | 1,866.09 |
| 402 | 1,063.04 | 3,594.45 | 8,038.24 | 1,063.04 | 13,758.77 |
| 403 | 3,019.64 | 5,245.56 | 2,751.41 | 3,288.96 | 14,305.57 |
| 404 | 2,665.85 | 1,599.51 | 2,843.02 | 1,332.65 | 8,441.03 |
| 405 | 993.78 | 684.73 | 699.22 | 372.85 | 2,750.58 |
| 406 | 177.72 | 802.94 | 177.72 | 177.72 | 1,336.10 |
| 407 | 0 | 1,066.34 | 888.62 | 888.62 | 2,843.58 |
| 408 | 710.89 | 0 | 0 | 0 | 710.89 |
| 409 | 0 | 88.86 | 444.31 | 88.86 | 622.03 |
| 410 | 2,574.78 | 266.58 | 3,642.77 | 1,242.41 | 7,726.54 |
| 420 | 3,732.18 | 4,890.12 | 2,045.18 | 2,754.71 | 13,422.19 |
| 421 | 1,964.07 | 1,954.95 | 2,674.96 | 2,043.81 | 8,637.79 |
| 422 | 1,421.78 | 3,913.10 | 0.46 | 2,135.41 | 7,470.75 |
| 423 | 1,774.21 | 710.89 | 1,154.37 | 2,482.90 | 6,122.37 |

RETAIL DRUG DISTRIBUTION BY ZIP CODE WITHIN STATE BY GRAMS WT

DRUG CODE:2270DRUG NAME:PENTOBARBITAL (SCHEDULE 2)
STATE:KENTUCKY

| ZIP CODE | QUARTER 1 | QUARTER 2 | QUARTER 3 | QUARTER 4 | TOTAL GRAMS |
|---|---|---|---|---|---|
| 424 | 355.45 | 1,332.92 | 1,421.78 | 355.45 | 3,465.60 |
| 425 | 531.52 | 0 | 0 | 354.35 | 885.87 |
| 426 | 177.17 | 88.86 | 0 | 0 | 266.03 |
| 427 | 355.45 | 2,132.68 | 3,552.81 | 5,422.83 | 11,463.77 |
| TOTAL | 22,495.01 | 28,994.52 | 31,667.79 | 26,492.70 | 109,650.02 |

STATE: LOUISIANA

| ZIP CODE | QUARTER 1 | QUARTER 2 | QUARTER 3 | QUARTER 4 | TOTAL GRAMS |
|---|---|---|---|---|---|
| 700 | 1,421.78 | 977.48 | 1,777.23 | 1,066.34 | 5,242.83 |
| 701 | 1,722.55 | 2,221.54 | 2,599.77 | 2,183.26 | 8,727.12 |
| 703 | 1,243.24 | 1,777.23 | 1,777.23 | 3,198.19 | 7,995.89 |
| 704 | 5,597.72 | 1,777.23 | 4,442.80 | 3,997.94 | 15,815.69 |
| 705 | 2,305.45 | 1,523.30 | 2,128.83 | 3,247.65 | 9,205.23 |
| 706 | 4,645.41 | 531.52 | 1,066.34 | 4,443.08 | 10,686.35 |
| 707 | 355.45 | 444.31 | 355.45 | 355.45 | 1,510.66 |
| 708 | 279.43 | 4,202.01 | 2,843.57 | 68.36 | 7,393.37 |
| 710 | 1,329.35 | 0 | 0 | 265.76 | 1,595.11 |
| 711 | 3,177.96 | 709.24 | 3,286.78 | 783.8 | 7,957.78 |
| 712 | 811.69 | 1,889.51 | 1,250.98 | 715.81 | 4,667.99 |
| 713 | 1,332.92 | 1,510.10 | 2,042.44 | 1,332.92 | 6,218.38 |
| 714 | 799.75 | 1,331.27 | 622.03 | 622.03 | 3,375.08 |
| TOTAL | 25,022.70 | 18,894.74 | 24,193.45 | 22,280.59 | 90,391.48 |

STATE: MAINE

| ZIP CODE | QUARTER 1 | QUARTER 2 | QUARTER 3 | QUARTER 4 | TOTAL GRAMS |
|---|---|---|---|---|---|
| 39 | 977.48 | 88.86 | 1,421.78 | 0 | 2,488.12 |
| 40 | 2,044.27 | 4,798.52 | 5,065.11 | 2,576.98 | 14,484.88 |
| 41 | 1,401.28 | 779.25 | 1,490.14 | 799.75 | 4,470.42 |
| 42 | 4,176.49 | 4,359.68 | 622.03 | 4,620.80 | 13,779 |
| 43 | 0 | 888.62 | 177.72 | 355.45 | 1,421.79 |
| 44 | 2,925.59 | 4,887.38 | 3,287.88 | 3,643.32 | 14,744.17 |
| 45 | 266.58 | 177.17 | 0 | 888.07 | 1,331.82 |
| 46 | 1,510.65 | 888.62 | 266.58 | 1,066.34 | 3,732.19 |
| 47 | 266.58 | 710.89 | 266.58 | 620.93 | 1,864.98 |
| 48 | 532.34 | 799.75 | 355.45 | 621.48 | 2,309.02 |
| 49 | 1,599.51 | 1,244.06 | 977.48 | 1,954.95 | 5,776 |
| TOTAL | 15,700.77 | 19,622.80 | 13,930.75 | 17,148.07 | 66,402.39 |

STATE: MARYLAND

| ZIP CODE | QUARTER 1 | QUARTER 2 | QUARTER 3 | QUARTER 4 | TOTAL GRAMS |
|---|---|---|---|---|---|
| 206 | 177.17 | 266.03 | 710.89 | 532.34 | 1,686.43 |
| 207 | 1,920.78 | 3,050.46 | 3,665.65 | 1,777.23 | 10,414.12 |
| 208 | 489.88 | 1,599.51 | 977.48 | 533.17 | 3,600.04 |
| 210 | 2,930.78 | 4,796.87 | 2,220.99 | 3,465.05 | 13,413.69 |
| 211 | 1,954.95 | 444.31 | 2,665.85 | 3,745.85 | 8,810.96 |
| 212 | 3,774.11 | 86.58 | 149.01 | 20.05 | 4,029.75 |
| 215 | 799.75 | 1,066.34 | 355.45 | 266.58 | 2,488.12 |
| 216 | 1,064.69 | 355.45 | 620.38 | 88.86 | 2,129.38 |
| 217 | 2,932.43 | 4,619.24 | 6,395.18 | 4,797.70 | 18,744.55 |
| 218 | 1,421.78 | 709.79 | 710.89 | 1,427.25 | 4,269.71 |
| 219 | 355.45 | 1,332.92 | 88.86 | 710.89 | 2,488.12 |
| TOTAL | 17,821.77 | 18,327.50 | 18,560.63 | 17,364.97 | 72,074.87 |

STATE: MASSACHUSETTS

| ZIP CODE | QUARTER 1 | QUARTER 2 | QUARTER 3 | QUARTER 4 | TOTAL GRAMS |
|---|---|---|---|---|---|
| 10 | 1,421.51 | 2,664.20 | 1,421.51 | 1,242.69 | 6,749.91 |

| | | | | | |
|---|---|---|---|---|---|
| 11 | 1,599.51 | 0 | 888.62 | 0 | 2,488.13 |
| 12 | 799.75 | 1,528.87 | 1,155.20 | 1,066.34 | 4,550.16 |
| 13 | 1,421.78 | 0 | 799.75 | 1,777.23 | 3,998.76 |
| 14 | 1,421.78 | 799.75 | 533.17 | 1,155.20 | 3,909.90 |
| 15 | 2,843.57 | 1,332.65 | 1,244.06 | 1,865.82 | 7,286.10 |
| 16 | 0 | 624.48 | 206.89 | 734.21 | 1,565.58 |
| 17 | 2,843.57 | 888.62 | 1,688.37 | 2,132.68 | 7,553.24 |
| 18 | 2,423.87 | 2,843.29 | 2,071.16 | 4,329.15 | 11,667.47 |
| 19 | 2,221.54 | 1,155.20 | 2,043.81 | 3,998.77 | 9,419.32 |
| 20 | 1,421.78 | 1,866.09 | 1,775.86 | 977.48 | 6,041.21 |
| 21 | 1,203.05 | 5,506.68 | 6,056.25 | 1,558.04 | 14,324.02 |
| 22 | 22.79 | 9.11 | 0 | 31.9 | 63.8 |
| 23 | 2,043.81 | 1,066.34 | 2,488.12 | 710.89 | 6,309.16 |
| 24 | 622.03 | 355.45 | 888.62 | 444.31 | 2,310.41 |

DRUG CODE:2270DRUG NAME:PENTOBARBITAL (SCHEDULE 2)
STATE:MASSACHUSETTS

| ZIP CODE | QUARTER 1 | QUARTER 2 | QUARTER 3 | QUARTER 4 | TOTAL GRAMS |
|---|---|---|---|---|---|
| 25 | 888.62 | 533.17 | 2,399.26 | 2,043.81 | 5,864.86 |
| 26 | 977.48 | 355.45 | 1,866.09 | 710.89 | 3,909.91 |
| 27 | 1,688.37 | 1,688.37 | 1,866.09 | 2,665.85 | 7,908.68 |
| TOTAL | 25,864.81 | 23,217.72 | 29,392.83 | 27,445.26 | 105,920.62 |

STATE: MICHIGAN

| ZIP CODE | QUARTER 1 | QUARTER 2 | QUARTER 3 | QUARTER 4 | TOTAL GRAMS |
|---|---|---|---|---|---|
| 480 | 7,313.07 | 8,908.66 | 10,686.17 | 7,820.72 | 34,728.62 |
| 481 | 15,361.45 | 10,963.95 | 14,850.41 | 11,722.14 | 52,897.95 |
| 482 | 3,059.57 | 7,735.96 | 1,316.52 | 5,448.35 | 17,560.40 |
| 483 | 6,860.01 | 3,656.44 | 5,239.80 | 11,905.52 | 27,661.77 |
| 484 | 4,354.21 | 6,929.36 | 7,372.02 | 6,754.84 | 25,410.43 |
| 485 | 1,599.51 | 4,725.15 | 953.32 | 1,421.78 | 8,699.76 |
| 486 | 712.07 | 1,868.45 | 1,970.16 | 3,733.36 | 8,284.04 |
| 487 | 533.17 | 4,976.24 | 1,421.78 | 1,154.10 | 8,085.29 |
| 488 | 9,066.15 | 9,068.43 | 10,041.07 | 10,574.24 | 38,749.89 |
| 489 | 5,709.01 | 284.81 | 828.01 | 88.86 | 6,910.69 |
| 490 | 11,378.28 | 11,373.72 | 13,506.95 | 15,536.54 | 51,795.49 |
| 491 | 888.62 | 10,219.07 | 1,244.06 | 799.75 | 13,151.50 |
| 492 | 886.14 | 1,510.65 | 2,486.28 | 3,638.92 | 8,521.99 |
| 493 | 2,754.71 | 3,110.15 | 4,976.24 | 3,821.04 | 14,662.14 |
| 494 | 1,514.29 | 5,337.16 | 4,976.24 | 3,648.33 | 15,476.02 |
| 495 | 2,875.47 | 3,176.23 | 2,680.88 | 5,242.83 | 13,975.41 |
| 496 | 3,108.50 | 2,399.26 | 3,554.46 | 1,244.06 | 10,306.28 |
| 497 | 2,221.54 | 799.75 | 2,932.43 | 1,777.23 | 7,730.95 |
| 498 | 533.17 | 2,488.12 | 0 | 533.17 | 3,554.46 |
| 499 | 531.52 | 177.72 | 531.52 | 354.35 | 1,595.11 |
| TOTAL | 81,260.46 | 99,709.28 | 91,568.32 | 97,220.13 | 369,758.19 |

STATE: MINNESOTA

| ZIP CODE | QUARTER 1 | QUARTER 2 | QUARTER 3 | QUARTER 4 | TOTAL GRAMS |
|---|---|---|---|---|---|
| 550 | 2,132.68 | 3,202.66 | 5,065.11 | 3,110.15 | 13,510.60 |
| 551 | 4,763.52 | 5,058 | 2,809.85 | 4,674.38 | 17,305.75 |
| 553 | 4,354.21 | 8,975.01 | 7,020.06 | 8,441.02 | 28,790.30 |
| 554 | 3,860.87 | 2,606.68 | 4,976.32 | 3,757.33 | 15,201.20 |
| 557 | 444.31 | 88.86 | 1,244.06 | 1,421.78 | 3,199.01 |
| 558 | 266.58 | 266.58 | 533.17 | 177.72 | 1,244.05 |
| 559 | 1,748.98 | 1,867 | 2,107.61 | 2,665.85 | 8,389.44 |
| 560 | 1,244.06 | 2,488.12 | 2,488.12 | 2,576.98 | 8,797.28 |
| 561 | 533.17 | 1,066.34 | 266.58 | 266.58 | 2,132.67 |

| | | | | | |
|---|---|---|---|---|---|
| 562 | 0 | 444.31 | 1,244.06 | 0 | 1,688.37 |
| 563 | 1,244.06 | 1,155.20 | 1,095.68 | 2,355.69 | 5,850.63 |
| 564 | 977.48 | 355.45 | 1,066.34 | 977.48 | 3,376.75 |
| 565 | 799.75 | 799.75 | 1,866.09 | 2,310.40 | 5,775.99 |
| 566 | 444.31 | 0 | 1,510.65 | 0 | 1,954.96 |
| 567 | 1,066.34 | 0 | 0 | 442.93 | 1,509.27 |
| TOTAL | 23,880.32 | 28,373.96 | 33,293.70 | 33,178.29 | 118,726.27 |

STATE: MISSISSIPPI

| ZIP CODE | QUARTER 1 | QUARTER 2 | QUARTER 3 | QUARTER 4 | TOTAL GRAMS |
|---|---|---|---|---|---|
| 386 | 2,488.12 | 0 | 1,777.23 | 177.17 | 4,442.52 |
| 387 | 533.17 | 0 | 0 | 177.72 | 710.89 |
| 388 | 0 | 3.65 | 38.28 | 714.54 | 756.47 |
| 389 | 533.17 | 0 | 0 | 533.17 | 1,066.34 |
| 390 | 0 | 177.72 | 0 | 0 | 177.72 |
| 391 | 0 | 710.89 | 0 | 355.45 | 1,066.34 |
| 392 | 2,665.85 | 88.86 | 2,788.88 | 1,790.90 | 7,334.49 |
| 393 | 531.52 | 1,151.90 | 974.73 | 88.86 | 2,747.01 |
| 394 | 0 | 88.86 | 0 | 9.11 | 97.97 |
| 395 | 444.31 | 888.62 | 2,576.98 | 88.86 | 3,998.77 |
| 396 | 0 | 0 | 531.52 | 0 | 531.52 |
| TOTAL | 7,196.14 | 3,110.50 | 8,687.62 | 3,935.78 | 22,930.04 |

STATE: MISSOURI

| ZIP CODE | QUARTER 1 | QUARTER 2 | QUARTER 3 | QUARTER 4 | TOTAL GRAMS |
|---|---|---|---|---|---|
| 630 | 6,308.34 | 1,243.79 | 4,797.97 | 2,840.82 | 15,190.92 |
| 631 | 753.63 | 7,063.35 | 3,507.07 | 10,096.03 | 21,420.08 |
| 633 | 1,775.58 | 1,866.09 | 6,579.84 | 3,908.81 | 14,130.32 |
| 634 | 0 | 888.62 | 1,066.34 | 888.62 | 2,843.58 |
| 635 | 178.63 | 356.36 | 178.63 | 178.63 | 892.25 |

DRUG CODE:2270DRUG NAME:PENTOBARBITAL (SCHEDULE 2)
STATE:MISSOURI

| ZIP CODE | QUARTER 1 | QUARTER 2 | QUARTER 3 | QUARTER 4 | TOTAL GRAMS |
|---|---|---|---|---|---|
| 636 | 2,132.68 | 1,244.06 | 1,421.78 | 2,488.12 | 7,286.64 |
| 637 | 1,603.15 | 466.18 | 1,155.20 | 266.58 | 3,491.11 |
| 638 | 0 | 177.72 | 266.58 | 88.86 | 533.16 |
| 639 | 177.72 | 0 | 177.72 | 177.72 | 533.16 |
| 640 | 6,575.75 | 4,265.35 | 7,197.78 | 3,110.15 | 21,149.03 |
| 641 | 730.94 | 689.47 | 738.23 | 621.57 | 2,780.21 |
| 644 | 1,421.78 | 799.75 | 1,954.95 | 1,332.92 | 5,509.40 |
| 645 | 0 | 0 | 0 | 533.17 | 533.17 |
| 646 | 355.45 | 0 | 533.17 | 355.45 | 1,244.07 |
| 647 | 533.17 | 977.48 | 533.17 | 1,866.09 | 3,909.91 |
| 648 | 444.31 | 1,510.65 | 1,332.92 | 1,244.06 | 4,531.94 |
| 650 | 888.62 | 1,069.07 | 355.45 | 888.62 | 3,201.76 |
| 651 | 266.58 | 1,066.34 | 710.89 | 177.72 | 2,221.53 |
| 652 | 4,974.42 | 2,768.38 | 5,400.05 | 82.03 | 13,224.88 |
| 653 | 0 | 888.62 | 0 | 0 | 888.62 |
| 654 | 533.17 | 533.17 | 0 | 266.58 | 1,332.92 |
| 655 | 177.72 | 798.1 | 710.89 | 2,399.26 | 4,085.97 |
| 656 | 3,376.74 | 886.97 | 799.75 | 1,954.95 | 7,018.41 |
| 657 | 266.58 | 266.58 | 1,777.23 | 3,459 | 5,769.39 |
| 658 | 177.72 | 184.1 | 359.09 | 187.75 | 908.66 |
| TOTAL | 33,652.68 | 30,010.20 | 41,554.70 | 39,413.51 | 144,631.09 |

STATE: MONTANA

| ZIP CODE | QUARTER 1 | QUARTER 2 | QUARTER 3 | QUARTER 4 | TOTAL GRAMS |
|---|---|---|---|---|---|
| 590 | 1,866.09 | 88.86 | 801.58 | 1,244.06 | 4,000.59 |
| 591 | 1,069.07 | 1,693.84 | 551.4 | 3,465.60 | 6,779.91 |

| | | | | | |
|---|---|---|---|---|---|
| 592 | 0 | 177.72 | 0 | 533.17 | 710.89 |
| 593 | 0 | 0 | 0 | 266.58 | 266.58 |
| 594 | 1,335.66 | 1,244.06 | 3,110.15 | 3,287.88 | 8,977.75 |
| 595 | 266.58 | 533.17 | 177.72 | 710.89 | 1,688.36 |
| 596 | 799.75 | 88.59 | 355.45 | 1,066.34 | 2,310.13 |
| 597 | 2,043.81 | 3,554.46 | 2,399.26 | 5,420.55 | 13,418.08 |
| 598 | 3,022.66 | 1,954.95 | 14,401.03 | 3,110.61 | 22,489.25 |
| 599 | 1,244.06 | 1,244.06 | 4,087.63 | 1,954.95 | 8,530.70 |
| TOTAL | 11,647.68 | 10,579.71 | 25,884.22 | 21,060.63 | 69,172.24 |

STATE: NEBRASKA

| ZIP CODE | QUARTER 1 | QUARTER 2 | QUARTER 3 | QUARTER 4 | TOTAL GRAMS |
|---|---|---|---|---|---|
| 680 | 3,643.32 | 1,954.95 | 3,821.04 | 4,531.94 | 13,951.25 |
| 681 | 3,720.79 | 5,067.84 | 2,697.29 | 4,158.72 | 15,644.64 |
| 683 | 355.45 | 1,244.06 | 534.99 | 1,866.09 | 4,000.59 |
| 684 | 622.03 | 0 | 2,310.40 | 2,754.71 | 5,687.14 |
| 685 | 3,111.06 | 3,199.01 | 3,199.93 | 5,153.97 | 14,663.97 |
| 686 | 266.58 | 88.86 | 710.89 | 444.31 | 1,510.64 |
| 687 | 977.48 | 2,665.85 | 0 | 1,421.78 | 5,065.11 |
| 688 | 462.99 | 3,199.01 | 2,957.04 | 1,335.20 | 7,954.24 |
| 689 | 88.86 | 1,244.06 | 0 | 977.48 | 2,310.40 |
| 690 | 0 | 533.17 | 0 | 533.17 | 1,066.34 |
| 691 | 1,423.15 | 355.45 | 1,067.71 | 533.17 | 3,379.48 |
| 693 | 2,310.40 | 888.62 | 2,665.85 | 1,510.65 | 7,375.52 |
| TOTAL | 16,982.11 | 20,440.88 | 19,965.14 | 25,221.19 | 82,609.32 |

STATE: NEVADA

| ZIP CODE | QUARTER 1 | QUARTER 2 | QUARTER 3 | QUARTER 4 | TOTAL GRAMS |
|---|---|---|---|---|---|
| 890 | 1,012.57 | 1,511.56 | 888.62 | 2,044.73 | 5,457.48 |
| 891 | 7,805.32 | 6,357.47 | 8,149.74 | 8,888.43 | 31,200.96 |
| 893 | 0 | 177.72 | 0 | 0 | 177.72 |
| 894 | 4,532.39 | 3,998.77 | 1,688.37 | 3,732.64 | 13,952.17 |
| 895 | 2,577.44 | 764.41 | 1,777.23 | 2,311.31 | 7,430.39 |
| 897 | 177.72 | 444.31 | 446.59 | 1,068.62 | 2,137.24 |
| 898 | 0 | 0 | 799.75 | 0 | 799.75 |
| TOTAL | 16,105.44 | 13,254.24 | 13,750.30 | 18,045.73 | 61,155.71 |

STATE: NEW HAMPSHIRE

| ZIP CODE | QUARTER 1 | QUARTER 2 | QUARTER 3 | QUARTER 4 | TOTAL GRAMS |
|---|---|---|---|---|---|
| 30 | 3,643.32 | 4,102.01 | 6,485.79 | 3,728.33 | 17,959.45 |
| 31 | 0 | 1,244.06 | 88.86 | 97.98 | 1,430.90 |
| 32 | 5,509.41 | 2,932.43 | 4,709.66 | 4,885.73 | 18,037.23 |
| 33 | 0 | 888.62 | 266.58 | 0 | 1,155.20 |
| 34 | 1,244.06 | 1,244.06 | 1,777.23 | 1,688.37 | 5,953.72 |

DRUG CODE:2270DRUG NAME:PENTOBARBITAL (SCHEDULE 2)
STATE:NEW HAMPSHIRE

| ZIP CODE | QUARTER 1 | QUARTER 2 | QUARTER 3 | QUARTER 4 | TOTAL GRAMS |
|---|---|---|---|---|---|
| 35 | 0 | 88.86 | 177.72 | 88.86 | 355.44 |
| 36 | 0 | 0 | 355.45 | 355.45 | 710.9 |
| 37 | 2,174.95 | 710.89 | 2,551.92 | 2,843.57 | 8,281.33 |
| 38 | 1,332.92 | 3,021.75 | 1,866.09 | 2,221.54 | 8,442.30 |
| TOTAL | 13,904.66 | 14,232.68 | 18,279.30 | 15,909.83 | 62,326.47 |

STATE: NEW JERSEY

| ZIP CODE | QUARTER 1 | QUARTER 2 | QUARTER 3 | QUARTER 4 | TOTAL GRAMS |
|---|---|---|---|---|---|
| 70 | 1,066.34 | 1,155.20 | 1,733.94 | 1,244.06 | 5,199.54 |
| 71 | 0 | 63.8 | 2,232.93 | 159.5 | 2,456.23 |
| 73 | 31.9 | 0 | 0 | 0 | 31.9 |
| 74 | 533.17 | 1,421.23 | 620.93 | 622.03 | 3,197.36 |

RETAIL DRUG DISTRIBUTION BY ZIP CODE WITHIN STATE BY GRAMS WT

| | | | | | |
|---|---|---|---|---|---|
| 75 | 2,130.64 | 10.94 | 0 | 0 | 2,141.58 |
| 76 | 1,954.95 | 2,604.33 | 1,711.15 | 4,980.80 | 11,251.23 |
| 77 | 1,155.20 | 0 | 2,221.54 | 533.17 | 3,909.91 |
| 78 | 3,376.74 | 2,488.12 | 4,087.63 | 2,488.12 | 12,440.61 |
| 79 | 444.31 | 1,232.67 | 977.48 | 1,173.43 | 3,827.89 |
| 80 | 2,137.23 | 3,114.71 | 1,958.60 | 3,918.75 | 11,129.29 |
| 81 | 0 | 0 | 18.23 | 0 | 18.23 |
| 82 | 444.03 | 977.48 | 0 | 533.17 | 1,954.68 |
| 83 | 1,599.51 | 3,021.29 | 3,199.01 | 1,244.06 | 9,063.87 |
| 84 | 22.79 | 0 | 0 | 0 | 22.79 |
| 85 | 2,219.89 | 531.52 | 3,638.37 | 1,864.17 | 8,253.95 |
| 86 | 9.11 | 517.22 | 622.03 | 0 | 1,148.36 |
| 87 | 0 | 0 | 1,155.20 | 0 | 1,155.20 |
| 88 | 868.11 | 1,156.29 | 622.03 | 1,510.65 | 4,157.08 |
| 89 | 444.31 | 0 | 587.85 | 266.58 | 1,298.74 |
| TOTAL | 18,438.23 | 18,294.80 | 25,386.92 | 20,538.49 | 82,658.44 |

STATE: NEW MEXICO

| ZIP CODE | QUARTER 1 | QUARTER 2 | QUARTER 3 | QUARTER 4 | TOTAL GRAMS |
|---|---|---|---|---|---|
| 870 | 3,822.41 | 2,844.02 | 6,132.35 | 3,199.47 | 15,998.25 |
| 871 | 2,444.83 | 3,773.20 | 3,287.88 | 7,200.06 | 16,705.97 |
| 874 | 1,599.51 | 1,879.76 | 3,915.37 | 2,843.57 | 10,238.21 |
| 875 | 2,221.54 | 977.48 | 2,843.57 | 2,221.54 | 8,264.13 |
| 877 | 1,066.34 | 444.31 | 266.58 | 710.89 | 2,488.12 |
| 880 | 1,244.06 | 2,665.85 | 1,599.51 | 4,275.38 | 9,784.80 |
| 881 | 533.17 | 0 | 533.17 | 0 | 1,066.34 |
| 882 | 534.54 | 801.12 | 0 | 1,512.01 | 2,847.67 |
| 883 | 444.31 | 0 | 888.62 | 0 | 1,332.93 |
| 884 | 0 | 0 | 0 | 177.72 | 177.72 |
| TOTAL | 13,910.71 | 13,385.74 | 19,467.05 | 22,140.64 | 68,904.14 |

STATE: NEW YORK

| ZIP CODE | QUARTER 1 | QUARTER 2 | QUARTER 3 | QUARTER 4 | TOTAL GRAMS |
|---|---|---|---|---|---|
| 100 | 550.49 | 195.95 | 214.18 | 1,215.81 | 2,176.43 |
| 103 | 0 | 0 | 0 | 177.72 | 177.72 |
| 104 | 12.76 | 225.57 | 4.56 | 59.24 | 302.13 |
| 105 | 1,421.78 | 888.62 | 2,488.12 | 2,889.14 | 7,687.66 |
| 108 | 0 | 0 | 0 | 177.72 | 177.72 |
| 109 | 976.65 | 799.2 | 1,153.55 | 533.17 | 3,462.57 |
| 110 | 1,016.21 | 0 | 31.9 | 54.68 | 1,102.79 |
| 111 | 0 | 355.45 | 0 | 0 | 355.45 |
| 112 | 37.37 | 0 | 1,954.95 | 0 | 1,992.32 |
| 113 | 1.82 | 0 | 5.47 | 1.82 | 9.11 |
| 114 | 0 | 0 | 177.72 | 533.17 | 710.89 |
| 115 | 0 | 0 | 3,375.36 | 0 | 3,375.36 |
| 117 | 1,688.37 | 710.07 | 1,332.65 | 1,244.06 | 4,975.15 |
| 119 | 88.86 | 799.75 | 977.48 | 0 | 1,866.09 |
| 120 | 799.75 | 799.75 | 1,065.79 | 621.48 | 3,286.77 |
| 121 | 4,709.66 | 5,863.21 | 4,530.29 | 6,931.20 | 22,034.36 |
| 122 | 0 | 223.29 | 1,151.90 | 223.29 | 1,598.48 |
| 123 | 88.86 | 143.55 | 89.32 | 0 | 321.73 |
| 124 | 0 | 0 | 799.75 | 88.86 | 888.61 |
| 125 | 6,309.17 | 8,262.47 | 7,284.99 | 9,149.44 | 31,006.07 |
| 126 | 266.58 | 0 | 266.58 | 266.58 | 799.74 |
| 127 | 177.72 | 0 | 355.45 | 0 | 533.17 |
| 128 | 2,043.81 | 3,019.64 | 1,866.09 | 3,287.88 | 10,217.42 |
| 129 | 88.86 | 1,777.23 | 1,332.92 | 1,866.09 | 5,065.10 |

RETAIL DRUG DISTRIBUTION BY ZIP CODE WITHIN STATE BY GRAMS WT

STATE:NEW YORK

| ZIP CODE | QUARTER 1 | QUARTER 2 | QUARTER 3 | QUARTER 4 | TOTAL GRAMS |
|---|---|---|---|---|---|
| 130 | 3,462.30 | 3,548.96 | 4,260.68 | 4,082.13 | 15,354.07 |
| 131 | 0 | 533.17 | 533.17 | 799.75 | 1,866.09 |
| 132 | 1,322.16 | 1,421.33 | 1,299.83 | 762.1 | 4,805.42 |
| 133 | 266.58 | 2,132.68 | 622.03 | 1,777.23 | 4,798.52 |
| 134 | 1,777.23 | 1,866.09 | 2,665.85 | 1,066.34 | 7,375.51 |
| 135 | 0 | 0 | 0 | 4.56 | 4.56 |
| 136 | 1,509 | 355.45 | 798.93 | 1,064.69 | 3,728.07 |
| 137 | 442.93 | 531.52 | 163.32 | 265.76 | 1,403.53 |
| 138 | 2,839.17 | 2,131.58 | 1,330.72 | 3,107.68 | 9,409.15 |
| 139 | 0 | 177.72 | 0 | 177.72 | 355.44 |
| 140 | 6,308.89 | 5,864.58 | 2,754.71 | 6,842.34 | 21,770.52 |
| 141 | 1,954.95 | 5,074.22 | 3,465.60 | 4,627.63 | 15,122.40 |
| 142 | 3,340.91 | 2,582.72 | 5,038.01 | 3,520 | 14,481.64 |
| 143 | 532.62 | 88.59 | 88.59 | 1,243.51 | 1,953.31 |
| 144 | 1,243.51 | 5,694.22 | 3,198.46 | 2,398.71 | 12,534.90 |
| 145 | 5,147.09 | 7,629.99 | 7,453.37 | 10,911.55 | 31,142 |
| 146 | 802.03 | 91.14 | 312.15 | 854.16 | 2,059.48 |
| 147 | 0 | 2,398.16 | 2,131.03 | 355.45 | 4,884.64 |
| 148 | 6,396.38 | 3,998.22 | 6,662.04 | 8,796.74 | 25,853.38 |
| 149 | 0 | 22.79 | 0 | 0 | 22.79 |
| TOTAL | 57,624.47 | 70,206.88 | 73,237.51 | 81,979.40 | 283,048.26 |

STATE: NORTH CAROLINA

| ZIP CODE | QUARTER 1 | QUARTER 2 | QUARTER 3 | QUARTER 4 | TOTAL GRAMS |
|---|---|---|---|---|---|
| 270 | 2,310.40 | 2,928.03 | 1,421.78 | 3,019.64 | 9,679.85 |
| 271 | 5,666.17 | 1,020.77 | 1,311.96 | 710.89 | 8,709.79 |
| 272 | 3,021.29 | 799.75 | 5,331.69 | 6,042.58 | 15,195.31 |
| 273 | 1,865.54 | 621.48 | 6,043.49 | 4,442.80 | 12,973.31 |
| 274 | 1,164.31 | 1,954.95 | 1,187.10 | 355.45 | 4,661.81 |
| 275 | 5,074.85 | 1,845.39 | 5,894.72 | 3,953.91 | 16,768.87 |
| 276 | 209.62 | 177.72 | 4,541.05 | 4,976.24 | 9,904.63 |
| 277 | 2,365.08 | 432.92 | 3,108.78 | 288 | 6,194.78 |
| 278 | 3,909.36 | 1,598.41 | 2,932.43 | 2,131.03 | 10,571.23 |
| 279 | 355.45 | 1,244.06 | 177.72 | 355.45 | 2,132.68 |
| 280 | 1,776.68 | 3,199.01 | 2,931.88 | 4,175.94 | 12,083.51 |
| 281 | 6,836.29 | 7,367.81 | 8,519.16 | 10,208.63 | 32,931.89 |
| 282 | 1,392.16 | 3,944.08 | 3,909.91 | 1,360.26 | 10,606.41 |
| 283 | 7,105.62 | 12,019.72 | 3,908.26 | 5,953.72 | 28,987.32 |
| 284 | 1,386.70 | 2,129.38 | 2,097.59 | 2,392.66 | 8,006.33 |
| 285 | 1,421.78 | 2,663.10 | 1,954.95 | 1,066.34 | 7,106.17 |
| 286 | 3,021.29 | 6,220.31 | 2,310.40 | 7,553.23 | 19,105.23 |
| 287 | 1,332.92 | 4,531.94 | 1,954.95 | 3,821.04 | 11,640.85 |
| 288 | 2,220.99 | 1,698.39 | 2,164.58 | 4,814.01 | 10,897.97 |
| 289 | 88.86 | 266.58 | 88.86 | 88.86 | 533.16 |
| TOTAL | 52,525.36 | 56,663.80 | 61,791.26 | 67,710.68 | 238,691.10 |

STATE: NORTH DAKOTA

| ZIP CODE | QUARTER 1 | QUARTER 2 | QUARTER 3 | QUARTER 4 | TOTAL GRAMS |
|---|---|---|---|---|---|
| 580 | 3,376.74 | 888.62 | 2,310.40 | 1,777.23 | 8,352.99 |
| 581 | 369.12 | 368.21 | 1,632.32 | 38.28 | 2,407.93 |
| 582 | 2,043.81 | 177.72 | 444.31 | 1,066.34 | 3,732.18 |
| 584 | 2,132.68 | 0 | 1,510.65 | 0 | 3,643.33 |
| 585 | 622.03 | 0 | 266.58 | 0 | 888.61 |
| 586 | 0 | 1,066.34 | 0 | 1,599.51 | 2,665.85 |
| 587 | 1,244.06 | 0 | 1,125.58 | 888.62 | 3,258.26 |
| TOTAL | 9,788.44 | 2,500.89 | 7,289.84 | 5,369.98 | 24,949.15 |

STATE: OHIO

| ZIP CODE | QUARTER 1 | QUARTER 2 | QUARTER 3 | QUARTER 4 | TOTAL GRAMS |
|---|---|---|---|---|---|

| | | | | | |
|---|---|---|---|---|---|
| 430 | 5,240.35 | 2,929.95 | 4,623.62 | 3,287.95 | 16,081.87 |
| 431 | 1,420.13 | 1,687.54 | 887.52 | 1,596.21 | 5,591.40 |
| 432 | 2,061.57 | 2,242.67 | 2,643.23 | 2,683.25 | 9,630.72 |
| 433 | 355.45 | 532.89 | 4,643.03 | 4,732.44 | 10,263.81 |
| 434 | 88.86 | 266.03 | 622.03 | 621.21 | 1,598.13 |
| 435 | 1,510.65 | 4,003.32 | 2,845.85 | 2,493.13 | 10,852.95 |
| 436 | 1,759.63 | 7,184.57 | 1,157.48 | 1,000.26 | 11,101.94 |
| 437 | 6,220.31 | 177.72 | 1,954.95 | 2,665.85 | 11,018.83 |
| 438 | 888.62 | 710.89 | 888.62 | 355.45 | 2,843.58 |
| 439 | 622.03 | 977.48 | 355.45 | 1,688.37 | 3,643.33 |

DRUG CODE:2270DRUG NAME:PENTOBARBITAL (SCHEDULE 2)
STATE:OHIO

| ZIP CODE | QUARTER 1 | QUARTER 2 | QUARTER 3 | QUARTER 4 | TOTAL GRAMS |
|---|---|---|---|---|---|
| 440 | 3,998.03 | 3,288.50 | 5,065.47 | 5,415.60 | 17,767.60 |
| 441 | 2,690.91 | 3,978.26 | 6,776.26 | 3,476.99 | 16,922.42 |
| 442 | 5,330.04 | 2,754.71 | 3,559.93 | 2,754.71 | 14,399.39 |
| 443 | 6,630.44 | 2,620.28 | 1,091.40 | 5,281.56 | 15,623.68 |
| 444 | 3,998.77 | 2,665.85 | 2,132.68 | 6,664.61 | 15,461.91 |
| 445 | 1,433.54 | 267.22 | 800.39 | 1,333.56 | 3,834.71 |
| 446 | 1,420.13 | 3,382.21 | 3,737.65 | 3,204.48 | 11,744.47 |
| 447 | 2,219.89 | 1,597.86 | 1,153 | 2,578.62 | 7,549.37 |
| 448 | 2,132.68 | 2,932.43 | 3,109.05 | 3,110.15 | 11,284.31 |
| 449 | 442.93 | 177.72 | 355.45 | 532.07 | 1,508.17 |
| 450 | 5,157.79 | 3,199.01 | 2,758.35 | 2,667.67 | 13,782.82 |
| 451 | 533.17 | 1,243.79 | 1,157.02 | 1,688.09 | 4,622.07 |
| 452 | 1,138.79 | 1,574.44 | 1,800.93 | 191.39 | 4,705.55 |
| 453 | 1,597.86 | 2,751.68 | 5,862.93 | 2,308.75 | 12,521.22 |
| 454 | 223.29 | 2,861.80 | 4,176.49 | 2,305.84 | 9,567.42 |
| 455 | 533.17 | 799.75 | 533.17 | 1,510.65 | 3,376.74 |
| 456 | 888.62 | 355.45 | 977.48 | 0 | 2,221.55 |
| 457 | 444.31 | 1,421.78 | 799.75 | 88.86 | 2,754.70 |
| 458 | 533.17 | 1,068.16 | 1,777.23 | 711.8 | 4,090.36 |
| TOTAL | 61,515.13 | 59,653.96 | 68,246.41 | 66,949.52 | 256,365.02 |

STATE: OKLAHOMA

| ZIP CODE | QUARTER 1 | QUARTER 2 | QUARTER 3 | QUARTER 4 | TOTAL GRAMS |
|---|---|---|---|---|---|
| 730 | 2,221.54 | 2,132.68 | 2,753.06 | 6,136.91 | 13,244.19 |
| 731 | 592.41 | 2,494.96 | 3,183.06 | 5,864.86 | 12,135.29 |
| 734 | 1,066.34 | 1,421.78 | 1,421.78 | 444.31 | 4,354.21 |
| 735 | 0 | 0 | 3,199.01 | 0 | 3,199.01 |
| 737 | 1,066.34 | 0 | 533.17 | 0 | 1,599.51 |
| 738 | 0 | 0 | 2,132.68 | 0 | 2,132.68 |
| 740 | 4,265.35 | 1,244.06 | 2,842.74 | 1,068.62 | 9,420.77 |
| 741 | 7,831.20 | 3,552.81 | 2,861.80 | 1,954.95 | 16,200.76 |
| 743 | 0 | 1,332.37 | 177.17 | 0 | 1,509.54 |
| 744 | 0 | 0 | 533.17 | 1,066.34 | 1,599.51 |
| 745 | 88.59 | 177.17 | 0 | 88.59 | 354.35 |
| 748 | 444.31 | 532.89 | 177.72 | 622.03 | 1,776.95 |
| TOTAL | 17,576.08 | 12,888.72 | 19,815.36 | 17,246.61 | 67,526.77 |

STATE: OREGON

| ZIP CODE | QUARTER 1 | QUARTER 2 | QUARTER 3 | QUARTER 4 | TOTAL GRAMS |
|---|---|---|---|---|---|
| 970 | 6,218.66 | 7,821.63 | 6,309.17 | 6,309.17 | 26,658.63 |
| 971 | 2,488.12 | 2,310.40 | 2,132.68 | 1,332.92 | 8,264.12 |
| 972 | 2,672.68 | 2,936.99 | 2,499.51 | 4,162.82 | 12,272 |
| 973 | 2,520.02 | 5,065.11 | 4,620.80 | 3,110.15 | 15,316.08 |
| 974 | 5,506.11 | 4,354.21 | 3,909.91 | 6,927.90 | 20,698.13 |
| 975 | 3,287.88 | 5,068.75 | 3,199.01 | 3,287.88 | 14,843.52 |

| | | | | | |
|---|---|---|---|---|---|
| 976 | 1,066.34 | 3,021.29 | 533.17 | 533.17 | 5,153.97 |
| 977 | 2,665.85 | 4,095.83 | 2,310.40 | 3,737.65 | 12,809.73 |
| 978 | 1,599.51 | 2,665.85 | 888.62 | 2,665.85 | 7,819.83 |
| 979 | 888.62 | 355.45 | 533.17 | 622.03 | 2,399.27 |
| TOTAL | 28,913.79 | 37,695.51 | 26,936.44 | 32,689.54 | 126,235.28 |

STATE: PENNSYLVANIA

| ZIP CODE | QUARTER 1 | QUARTER 2 | QUARTER 3 | QUARTER 4 | TOTAL GRAMS |
|---|---|---|---|---|---|
| 150 | 1,066.34 | 88.86 | 1,421.78 | 1,332.92 | 3,909.90 |
| 151 | 1,599.51 | 933.64 | 1,598.68 | 621.76 | 4,753.59 |
| 152 | 5,013.61 | 1,124.02 | 2,327.26 | 2,902.81 | 11,367.70 |
| 153 | 0 | 0 | 3,013.04 | 3,467.88 | 6,480.92 |
| 154 | 1,066.34 | 88.86 | 443.21 | 2,310.40 | 3,908.81 |
| 155 | 799.75 | 799.75 | 444.31 | 799.75 | 2,843.56 |
| 156 | 2,843.29 | 2,310.40 | 1,332.65 | 1,866.09 | 8,352.43 |
| 159 | 944.21 | 219.65 | 2,036.98 | 191.39 | 3,392.23 |
| 160 | 177.72 | 531.52 | 177.72 | 0 | 886.96 |
| 161 | 1,332.92 | 0 | 1,777.23 | 0 | 3,110.15 |
| 162 | 0 | 0 | 0 | 1,332.92 | 1,332.92 |
| 163 | 1,599.51 | 1,421.78 | 1,332.92 | 888.62 | 5,242.83 |
| 164 | 1,954.95 | 2,310.40 | 799.75 | 1,688.37 | 6,753.47 |
| 165 | 533.17 | 3,405.90 | 799.75 | 799.75 | 5,538.57 |
| 166 | 1,244.06 | 888.62 | 88.86 | 1,954.95 | 4,176.49 |
| 168 | 3,837.45 | 1,777.23 | 3,199.01 | 444.31 | 9,258 |

DRUG CODE:2270DRUG NAME:PENTOBARBITAL (SCHEDULE 2)
STATE:PENNSYLVANIA

| ZIP CODE | QUARTER 1 | QUARTER 2 | QUARTER 3 | QUARTER 4 | TOTAL GRAMS |
|---|---|---|---|---|---|
| 169 | 266.58 | 1,421.78 | 888.62 | 177.72 | 2,754.70 |
| 170 | 5,064.28 | 6,658.01 | 3,528.57 | 8,899.04 | 24,149.90 |
| 171 | 710.89 | 1,155.20 | 1,332.92 | 194.13 | 3,393.14 |
| 172 | 1,954.95 | 2,665.02 | 3,640.02 | 1,244.06 | 9,504.05 |
| 173 | 1,242.41 | 1,332.92 | 266.58 | 2,575.33 | 5,417.24 |
| 174 | 1,877.48 | 1,777.23 | 100.25 | 1,954.95 | 5,709.91 |
| 175 | 1,421.78 | 0 | 3,554.46 | 1,066.34 | 6,042.58 |
| 176 | 444.31 | 177.72 | 0 | 622.03 | 1,244.06 |
| 177 | 1,421.78 | 0 | 710.89 | 1,599.51 | 3,732.18 |
| 178 | 2,576.98 | 3,290.15 | 2,579.26 | 2,955.21 | 11,401.60 |
| 179 | 355.45 | 355.45 | 533.17 | 177.72 | 1,421.79 |
| 180 | 1,866.09 | 178.63 | 2,399.26 | 1,332.92 | 5,776.90 |
| 181 | 533.17 | 1,066.34 | 339.5 | 177.72 | 2,116.73 |
| 182 | 888.62 | 0 | 355.45 | 355.45 | 1,599.52 |
| 183 | 355.45 | 1,244.06 | 533.17 | 2,399.26 | 4,531.94 |
| 184 | 177.72 | 1,510.65 | 177.72 | 1,954.95 | 3,821.04 |
| 185 | 0 | 0 | 888.62 | 0 | 888.62 |
| 186 | 177.72 | 1,066.34 | 355.45 | 177.72 | 1,777.23 |
| 187 | 622.03 | 0 | 3,215.42 | 622.03 | 4,459.48 |
| 188 | 266.58 | 88.86 | 88.86 | 533.17 | 977.47 |
| 189 | 5,507.76 | 5,152.32 | 5,595.53 | 4,709.66 | 20,965.27 |
| 190 | 2,428.88 | 1,688.37 | 2,313.86 | 1,688.37 | 8,119.48 |
| 191 | 1,327.89 | 788.82 | 537.73 | 945.58 | 3,600.02 |
| 193 | 3,643.32 | 2,222.45 | 3,909.91 | 2,489.03 | 12,264.71 |
| 194 | 2,489.94 | 3,840.64 | 2,247.51 | 4,798.52 | 13,376.61 |
| 195 | 2,310.40 | 444.31 | 1,688.37 | 710.89 | 5,153.97 |
| 196 | 2,757.34 | 2,130.20 | 2,141.15 | 2,046.45 | 9,075.14 |
| TOTAL | 66,702.63 | 56,156.10 | 64,715.40 | 67,009.68 | 254,583.81 |

STATE: PUERTO RICO

| ZIP CODE | QUARTER 1 | QUARTER 2 | QUARTER 3 | QUARTER 4 | TOTAL GRAMS |
|---|---|---|---|---|---|
| 6 | 0 | 1,781.79 | 0 | 0 | 1,781.79 |

| | | | | | |
|---|---|---|---|---|---|
| 7 | 893.17 | 1,066.34 | 0 | 0 | 1,959.51 |
| 9 | 0 | 1,155.20 | 0 | 0 | 1,155.20 |
| TOTAL | 893.17 | 4,003.33 | 0 | 0 | 4,896.50 |

STATE: RHODE ISLAND

| ZIP CODE | QUARTER 1 | QUARTER 2 | QUARTER 3 | QUARTER 4 | TOTAL GRAMS |
|---|---|---|---|---|---|
| 28 | 4,087.63 | 7,553.23 | 5,953.72 | 9,774.77 | 27,369.35 |
| 29 | 532.34 | 1,159.76 | 712.35 | 365.93 | 2,770.38 |
| TOTAL | 4,619.97 | 8,712.99 | 6,666.07 | 10,140.70 | 30,139.73 |

STATE: SOUTH CAROLINA

| ZIP CODE | QUARTER 1 | QUARTER 2 | QUARTER 3 | QUARTER 4 | TOTAL GRAMS |
|---|---|---|---|---|---|
| 290 | 1,688.37 | 2,310.40 | 355.45 | 2,401.54 | 6,755.76 |
| 291 | 1,066.34 | 444.31 | 888.62 | 1,244.06 | 3,643.33 |
| 292 | 952.41 | 1,084.57 | 943.3 | 3,005.34 | 5,985.62 |
| 293 | 2,843.57 | 1,599.51 | 4.56 | 1,080.01 | 5,527.65 |
| 294 | 906.84 | 5,090.97 | 2,776.47 | 2,578.24 | 11,352.52 |
| 295 | 724.56 | 1,444.57 | 1,785.32 | 1,269.12 | 5,223.57 |
| 296 | 2,592.81 | 5,569.91 | 710.89 | 1,980.93 | 10,854.54 |
| 297 | 444.31 | 266.58 | 2,221.54 | 355.45 | 3,287.88 |
| 298 | 1,777.23 | 355.45 | 2,399.26 | 2,399.26 | 6,931.20 |
| 299 | 533.17 | 88.86 | 0 | 444.51 | 1,066.54 |
| TOTAL | 13,529.61 | 18,255.13 | 12,085.41 | 16,758.46 | 60,628.61 |

STATE: SOUTH DAKOTA

| ZIP CODE | QUARTER 1 | QUARTER 2 | QUARTER 3 | QUARTER 4 | TOTAL GRAMS |
|---|---|---|---|---|---|
| 570 | 3,909.91 | 1,066.34 | 533.17 | 2,665.85 | 8,175.27 |
| 571 | 628.87 | 1,155.20 | 1,228.11 | 587.85 | 3,600.03 |
| 572 | 0 | 0 | 533.17 | 0 | 533.17 |
| 573 | 177.72 | 177.72 | 355.45 | 0 | 710.89 |
| 575 | 888.62 | 0 | 0 | 1,421.78 | 2,310.40 |
| 577 | 1,169.33 | 3,287.88 | 2,936.99 | 3,025.85 | 10,420.05 |
| TOTAL | 6,774.45 | 5,687.14 | 5,586.89 | 7,701.33 | 25,749.81 |

STATE: TENNESSEE

| ZIP CODE | QUARTER 1 | QUARTER 2 | QUARTER 3 | QUARTER 4 | TOTAL GRAMS |
|---|---|---|---|---|---|
| 370 | 5,415.60 | 2,751.68 | 8,703.48 | 5,148.19 | 22,018.95 |
| 371 | 3,373.44 | 4,527.26 | 3,371.24 | 1,777.23 | 13,049.17 |
| 372 | 305.32 | 284.81 | 177.72 | 4,837.26 | 5,605.11 |

DRUG CODE:2270DRUG NAME:PENTOBARBITAL (SCHEDULE 2)
STATE:TENNESSEE

| ZIP CODE | QUARTER 1 | QUARTER 2 | QUARTER 3 | QUARTER 4 | TOTAL GRAMS |
|---|---|---|---|---|---|
| 373 | 3,285.40 | 3,131.84 | 4,085.70 | 4,619.70 | 15,122.64 |
| 374 | 2,754.71 | 1,155.20 | 756.46 | 1,845.59 | 6,511.96 |
| 376 | 4,299.25 | 3,765.16 | 6,632.44 | 4,379.08 | 19,075.93 |
| 377 | 2,938.62 | 532.89 | 2,398.16 | 4,629.54 | 10,499.21 |
| 378 | 1,863.07 | 2,753.61 | 5,601.90 | 1,597.86 | 11,816.44 |
| 379 | 2,037.49 | 7,576.08 | 3,370.41 | 2,769.71 | 15,753.69 |
| 380 | 1,155.20 | 710.89 | 887.52 | 1,864.99 | 4,618.60 |
| 381 | 161.5 | 1,332.10 | 1,089.12 | 159.22 | 2,741.94 |
| 382 | 1,066.34 | 533.17 | 355.45 | 1,688.37 | 3,643.33 |
| 383 | 1,243.51 | 1,775.03 | 709.79 | 1,243.51 | 4,971.84 |
| 384 | 2,232.93 | 1,510.37 | 266.31 | 1,512.47 | 5,522.08 |
| 385 | 3,192.42 | 88.86 | 1,066.34 | 888.62 | 5,236.24 |
| TOTAL | 35,324.80 | 32,428.95 | 39,472.04 | 38,961.34 | 146,187.13 |

STATE: TEXAS

| ZIP CODE | QUARTER 1 | QUARTER 2 | QUARTER 3 | QUARTER 4 | TOTAL GRAMS |
|---|---|---|---|---|---|
| 750 | 4,443.08 | 13,862.39 | 5,798.78 | 11,219.33 | 35,323.58 |

| | | | | | |
|---|---|---|---|---|---|
| 751 | 8,257.52 | 3,199.01 | 5,864.86 | 1,862.79 | 19,184.18 |
| 752 | 1,913.94 | 1,845.59 | 28,319.48 | 1,672.42 | 33,751.43 |
| 753 | 46.48 | 36.46 | 0 | 0 | 82.94 |
| 754 | 1,156.11 | 2,044.73 | 3,019.64 | 1,688.37 | 7,908.85 |
| 755 | 533.17 | 177.72 | 888.62 | 533.17 | 2,132.68 |
| 756 | 1,603.33 | 3,732.18 | 15,992.32 | 2,749.76 | 24,077.59 |
| 757 | 1,777.23 | 2,633.95 | 2,131.85 | 1,777.23 | 8,320.26 |
| 758 | 0 | 533.17 | 0 | 0 | 533.17 |
| 759 | 533.17 | 4,708.01 | 2,664.20 | 3,641.67 | 11,547.05 |
| 760 | 9,241.60 | 2,576.98 | 6,042.58 | 10,489.30 | 28,350.46 |
| 761 | 1,244.06 | 15,125.14 | 9,526.41 | 1,542.54 | 27,438.15 |
| 762 | 622.03 | 1,332.92 | 3,199.01 | 1,244.06 | 6,398.02 |
| 763 | 1,866.09 | 3,554.46 | 1,066.34 | 2,665.85 | 9,152.74 |
| 764 | 1,421.78 | 177.72 | 2,488.12 | 1,510.65 | 5,598.27 |
| 765 | 2,674.96 | 2,427.97 | 3,534.86 | 977.48 | 9,615.27 |
| 766 | 88.86 | 177.72 | 266.58 | 177.72 | 710.88 |
| 767 | 888.62 | 0 | 177.72 | 266.58 | 1,332.92 |
| 768 | 0 | 0 | 533.17 | 0 | 533.17 |
| 769 | 2,321.34 | 0 | 0 | 2,143.61 | 4,464.95 |
| 770 | 2,850.86 | 32,880.58 | 1,594.95 | 11,053 | 48,379.39 |
| 773 | 2,044.27 | 622.03 | 1,598.77 | 1,602.41 | 5,867.48 |
| 774 | 355.45 | 1,777.23 | 1,155.20 | 1,777.23 | 5,065.11 |
| 775 | 6,039.28 | 7,297.58 | 6,128.14 | 8,260.82 | 27,725.82 |
| 776 | 444.31 | 4,087.63 | 3,728.88 | 444.31 | 8,705.13 |
| 777 | 0 | 1,954.95 | 0 | 266.58 | 2,221.53 |
| 778 | 1,777.76 | 2,574.78 | 4,621.33 | 3,194.62 | 12,168.49 |
| 779 | 888.62 | 533.17 | 888.62 | 888.62 | 3,199.03 |
| 780 | 4,176.49 | 977.48 | 4,354.21 | 2,932.43 | 12,440.61 |
| 781 | 622.03 | 1,866.09 | 1,066.34 | 2,665.85 | 6,220.31 |
| 782 | 2,319.51 | 5,900.40 | 1,267.30 | 1,815.96 | 11,303.17 |
| 783 | 177.72 | 2,043.81 | 1,421.78 | 533.17 | 4,176.48 |
| 784 | 533.17 | 0 | 538.64 | 5.47 | 1,077.28 |
| 785 | 11,196.55 | 8,352.98 | 12,529.47 | 12,262.89 | 44,341.89 |
| 786 | 9,241.60 | 4,087.63 | 5,687.14 | 5,242.83 | 24,259.20 |
| 787 | 1,013.02 | 2,310.85 | 888.62 | 1,249.53 | 5,462.02 |
| 788 | 1,866.09 | 444.31 | 1,244.06 | 0 | 3,554.46 |
| 789 | 710.89 | 533.17 | 0 | 533.17 | 1,777.23 |
| 790 | 1,155.20 | 444.31 | 4,087.63 | 0 | 5,687.14 |
| 791 | 1,005.73 | 729.12 | 4,901.05 | 658.49 | 7,294.39 |
| 793 | 1,601.33 | 1,066.34 | 888.62 | 177.72 | 3,734.01 |
| 794 | 2,266.20 | 3,561.30 | 2,232.47 | 2,232.93 | 10,292.90 |
| 795 | 355.45 | 1,064.69 | 355.45 | 531.52 | 2,307.11 |
| 796 | 1,332.92 | 3,287.33 | 1,066.34 | 1,066.34 | 6,752.93 |
| 797 | 5,161.43 | 0 | 4,708.01 | 6,549.04 | 16,418.48 |
| 798 | 0 | 0 | 266.58 | 0 | 266.58 |
| 799 | 556.87 | 10,863.89 | 2,152.73 | 29,253.66 | 42,827.15 |
| TOTAL | 100,326.12 | 157,407.77 | 160,886.87 | 141,361.12 | 559,981.88 |

STATE: UTAH

| ZIP CODE | QUARTER 1 | QUARTER 2 | QUARTER 3 | QUARTER 4 | TOTAL GRAMS |
|---|---|---|---|---|---|
| 840 | 8,530.15 | 4,976.68 | 802.49 | 1,954.95 | 16,264.27 |

DRUG CODE:2270DRUG NAME:PENTOBARBITAL (SCHEDULE 2)
STATE:UTAH

| ZIP CODE | QUARTER 1 | QUARTER 2 | QUARTER 3 | QUARTER 4 | TOTAL GRAMS |
|---|---|---|---|---|---|
| 841 | 3,622.82 | 1,822.80 | 3,356.23 | 6,379.34 | 15,181.19 |
| 843 | 0 | 0 | 0 | 88.86 | 88.86 |
| 844 | 0 | 183.19 | 895.91 | 353.62 | 1,432.72 |
| 845 | 88.86 | 0 | 1,335.20 | 533.17 | 1,957.23 |

| | | | | | |
|---|---|---|---|---|---|
| 846 | 1,954.95 | 2,310.40 | 1,066.34 | 1,332.92 | 6,664.61 |
| 847 | 1,244.06 | 478.49 | 1,156.11 | 177.72 | 3,056.38 |
| TOTAL | 15,440.84 | 9,771.56 | 8,612.28 | 10,820.58 | 44,645.26 |

STATE: VERMONT

| ZIP CODE | QUARTER 1 | QUARTER 2 | QUARTER 3 | QUARTER 4 | TOTAL GRAMS |
|---|---|---|---|---|---|
| 50 | 2,221.26 | 710.89 | 1,954.68 | 2,221.26 | 7,108.09 |
| 51 | 355.45 | 0 | 798.65 | 622.03 | 1,776.13 |
| 52 | 799.75 | 977.48 | 1,066.34 | 622.03 | 3,465.60 |
| 53 | 177.72 | 0 | 266.58 | 177.72 | 622.02 |
| 54 | 2,221.71 | 7,103.87 | 3,519.46 | 8,438.76 | 21,283.80 |
| 56 | 1,066.06 | 1,421.78 | 533.17 | 1,332.92 | 4,353.93 |
| 57 | 0 | 2,399.26 | 888.62 | 1,777.23 | 5,065.11 |
| 58 | 0 | 88.86 | 177.72 | 177.72 | 444.3 |
| TOTAL | 6,841.95 | 12,702.14 | 9,205.22 | 15,369.67 | 44,118.98 |

STATE: VIRGINIA

| ZIP CODE | QUARTER 1 | QUARTER 2 | QUARTER 3 | QUARTER 4 | TOTAL GRAMS |
|---|---|---|---|---|---|
| 201 | 2,398.99 | 1,510.65 | 2,310.40 | 1,066.34 | 7,286.38 |
| 220 | 2,144.07 | 2,169.13 | 1,911.66 | 1,779.51 | 8,004.37 |
| 221 | 266.58 | 1,777.23 | 177.72 | 177.72 | 2,399.25 |
| 223 | 355.45 | 1,421.78 | 622.03 | 444.31 | 2,843.57 |
| 224 | 2,665.85 | 88.86 | 620.38 | 533.17 | 3,908.26 |
| 225 | 266.58 | 2,486.47 | 1,064.69 | 1,066.34 | 4,884.08 |
| 226 | 1,688.37 | 2,665.85 | 888.62 | 1,866.09 | 7,108.93 |
| 227 | 443.21 | 1,064.69 | 266.58 | 710.89 | 2,485.37 |
| 228 | 2,221.54 | 1,244.06 | 1,155.20 | 1,155.20 | 5,776 |
| 229 | 3,337.55 | 4,762.07 | 5,364.96 | 5,580.68 | 19,045.26 |
| 230 | 3,376.74 | 3,465.32 | 3,643.32 | 4,531.66 | 15,017.04 |
| 231 | 799.75 | 710.89 | 888.62 | 977.48 | 3,376.74 |
| 232 | 991.15 | 289.37 | 152.66 | 88.86 | 1,522.04 |
| 233 | 444.31 | 2,932.43 | 799.75 | 533.17 | 4,709.66 |
| 234 | 5,680.54 | 4,798.52 | 6,668.04 | 6,398.03 | 23,545.13 |
| 235 | 1,125.58 | 1,371.66 | 2,201.03 | 88.86 | 4,787.13 |
| 236 | 2,310.40 | 1,866.09 | 3,110.61 | 1,955.41 | 9,242.51 |
| 237 | 0 | 0 | 266.58 | 0 | 266.58 |
| 238 | 4,531.94 | 977.48 | 3,110.15 | 977.48 | 9,597.05 |
| 239 | 444.31 | 533.17 | 622.03 | 888.62 | 2,488.13 |
| 240 | 266.31 | 2,262.55 | 3,643.05 | 2,843.29 | 9,015.20 |
| 241 | 444.31 | 716.36 | 1,332.92 | 1,158.39 | 3,651.98 |
| 242 | 0 | 0 | 266.58 | 888.62 | 1,155.20 |
| 243 | 3,554.46 | 708.69 | 1,332.92 | 1,066.34 | 6,662.41 |
| 244 | 1,777.23 | 2,488.12 | 0 | 177.72 | 4,443.07 |
| 245 | 3,377.65 | 2,221.54 | 2,401.08 | 4,360.59 | 12,360.86 |
| 246 | 533.17 | 0 | 533.17 | 177.45 | 1,243.79 |
| TOTAL | 45,446.04 | 44,532.98 | 45,354.75 | 41,492.22 | 176,825.99 |

STATE: WASHINGTON

| ZIP CODE | QUARTER 1 | QUARTER 2 | QUARTER 3 | QUARTER 4 | TOTAL GRAMS |
|---|---|---|---|---|---|
| 980 | 5,242.83 | 3,732.18 | 5,242.83 | 6,927.90 | 21,145.74 |
| 981 | 1,068.62 | 3,194.46 | 777.88 | 1,674.70 | 6,715.66 |
| 982 | 17,163.67 | 6,138.73 | 12,886.74 | 14,845.79 | 51,034.93 |
| 983 | 2,754.71 | 4,265.35 | 2,754.71 | 6,220.31 | 15,995.08 |
| 984 | 1,676.98 | 2,959.77 | 1,688.37 | 1,171.15 | 7,496.27 |
| 985 | 6,398.03 | 7,730.95 | 3,732.18 | 2,576.98 | 20,438.14 |
| 986 | 5,417.25 | 4,970.75 | 5,509.41 | 8,353.44 | 24,250.85 |
| 988 | 1,421.78 | 623.85 | 3,911.73 | 4,003.78 | 9,961.14 |
| 989 | 1,158.85 | 1,783.15 | 1,780.88 | 3,380.38 | 8,103.26 |
| 990 | 6,575.75 | 2,488.12 | 5,065.11 | 4,976.24 | 19,105.22 |
| 991 | 1,066.34 | 1,335.66 | 977.2 | 4,268.09 | 7,647.29 |
| 992 | 2,996.23 | 3,554.46 | 2,119.01 | 2,843.57 | 11,513.27 |

| | | | | | |
|---|---|---|---|---|---|
| 993 | 4,087.63 | 2,488.12 | 710.89 | 1,244.06 | 8,530.70 |
| 994 | 355.45 | 0 | 177.72 | 355.45 | 888.62 |
| TOTAL | 57,384.12 | 45,265.55 | 47,334.66 | 62,841.84 | 212,826.17 |

STATE: WEST VIRGINIA

DRUG CODE:2270DRUG NAME:PENTOBARBITAL (SCHEDULE 2)
STATE:WEST VIRGINIA

| ZIP CODE | QUARTER 1 | QUARTER 2 | QUARTER 3 | QUARTER 4 | TOTAL GRAMS |
|---|---|---|---|---|---|
| 247 | 0 | 0 | 0 | 533.17 | 533.17 |
| 249 | 0 | 0 | 531.52 | 0 | 531.52 |
| 250 | 0 | 0 | 355.45 | 0 | 355.45 |
| 251 | 266.58 | 0 | 266.58 | 0 | 533.16 |
| 252 | 888.62 | 177.72 | 88.86 | 1,066.34 | 2,221.54 |
| 253 | 1,954.95 | 1,777.23 | 888.62 | 1,066.34 | 5,687.14 |
| 254 | 1,332.92 | 1,066.34 | 1,599.51 | 1,421.78 | 5,420.55 |
| 255 | 355.45 | 710.89 | 531.52 | 0 | 1,597.86 |
| 256 | 0 | 355.45 | 0 | 0 | 355.45 |
| 257 | 289.37 | 622.03 | 667.6 | 888.62 | 2,467.62 |
| 258 | 0 | 0 | 533.17 | 0 | 533.17 |
| 259 | 266.58 | 533.17 | 177.72 | 444.31 | 1,421.78 |
| 260 | 2,754.71 | 0 | 2,221.54 | 799.75 | 5,776 |
| 261 | 0 | 799.75 | 1,596.21 | 977.48 | 3,373.44 |
| 262 | 0 | 0 | 355.45 | 533.17 | 888.62 |
| 263 | 0 | 88.86 | 0 | 0 | 88.86 |
| 264 | 0 | 177.72 | 0 | 266.58 | 444.3 |
| 265 | 555.95 | 867.01 | 887.52 | 2,043.81 | 4,354.29 |
| 266 | 0 | 0 | 0 | 885.87 | 885.87 |
| 267 | 444.31 | 533.17 | 533.17 | 1,244.06 | 2,754.71 |
| 268 | 533.17 | 533.17 | 0 | 0 | 1,066.34 |
| TOTAL | 9,642.61 | 8,242.51 | 11,234.44 | 12,171.28 | 41,290.84 |

STATE: WISCONSIN

| ZIP CODE | QUARTER 1 | QUARTER 2 | QUARTER 3 | QUARTER 4 | TOTAL GRAMS |
|---|---|---|---|---|---|
| 530 | 4,798.51 | 976.93 | 5,235.13 | 2,662.27 | 13,672.84 |
| 531 | 4,265.35 | 6,761.68 | 7,017.31 | 3,821.04 | 21,865.38 |
| 532 | 4,743.73 | 4,096.74 | 2,838.74 | 6,140.01 | 17,819.22 |
| 534 | 266.58 | 0 | 444.31 | 622.03 | 1,332.92 |
| 535 | 9,329.91 | 6,397.20 | 6,309.17 | 8,085.85 | 30,122.13 |
| 537 | 683.55 | 3,043.16 | 3,795.53 | 1,739.40 | 9,261.64 |
| 538 | 0 | 1,151.90 | 266.58 | 444.31 | 1,862.79 |
| 539 | 622.03 | 888.62 | 1,690.19 | 1,155.20 | 4,356.04 |
| 540 | 444.31 | 1,954.68 | 1,155.20 | 1,332.92 | 4,887.11 |
| 541 | 355.45 | 1,685.98 | 355.45 | 799.75 | 3,196.63 |
| 542 | 708.69 | 531.52 | 88.86 | 0 | 1,329.07 |
| 543 | 401.02 | 892.98 | 364.56 | 219.65 | 1,878.21 |
| 544 | 2,150.90 | 1,421.23 | 3,640.85 | 2,663.65 | 9,876.63 |
| 545 | 355.45 | 622.03 | 2,399.26 | 444.31 | 3,821.05 |
| 546 | 729.12 | 724.56 | 1,777.23 | 977.48 | 4,208.39 |
| 547 | 2,836.70 | 1,688.37 | 2,129.38 | 1,597.31 | 8,251.76 |
| 548 | 2,043.81 | 2,576.98 | 2,932.43 | 888.62 | 8,441.84 |
| 549 | 5,951.78 | 3,909.36 | 2,598.31 | 4,621.43 | 17,080.88 |
| TOTAL | 40,686.89 | 39,323.92 | 45,038.49 | 38,215.23 | 163,264.53 |

STATE: WYOMING

| ZIP CODE | QUARTER 1 | QUARTER 2 | QUARTER 3 | QUARTER 4 | TOTAL GRAMS |
|---|---|---|---|---|---|
| 820 | 1,332.92 | 1,244.06 | 1,332.92 | 4,887.38 | 8,797.28 |
| 822 | 0 | 0 | 0 | 88.86 | 88.86 |
| 823 | 1,066.34 | 0 | 0 | 1,066.34 | 2,132.68 |

| | | | | | |
|---|---|---|---|---|---|
| 824 | 3,998.77 | 177.72 | 3,909.91 | 177.72 | 8,264.12 |
| 825 | 1.82 | 0.91 | 0 | 0 | 2.73 |
| 826 | 2,404.73 | 1,066.34 | 1,066.34 | 888.62 | 5,426.03 |
| 827 | 1,332.92 | 622.03 | 1,155.20 | 355.45 | 3,465.60 |
| 828 | 1,954.95 | 1,510.65 | 710.89 | 1,954.95 | 6,131.44 |
| 829 | 1,777.23 | 710.89 | 0 | 710.89 | 3,199.01 |
| 831 | 533.17 | 0 | 266.58 | 533.17 | 1,332.92 |
| TOTAL | 14,402.85 | 5,332.60 | 8,441.84 | 10,663.38 | 38,840.67 |

DRUG CODE:2315DRUG NAME:SECOBARBITAL (SCHEDULE 2)
STATE: ARIZONA

| ZIP CODE | QUARTER 1 | QUARTER 2 | QUARTER 3 | QUARTER 4 | TOTAL GRAMS |
|---|---|---|---|---|---|
| 852 | 9.16 | 0 | 0 | 9.16 | 18.32 |
| 853 | 0 | 9.16 | 0 | 0 | 9.16 |
| 857 | 0 | 0 | 0 | 9.16 | 9.16 |
| TOTAL | 9.16 | 9.16 | 0 | 18.32 | 36.64 |

STATE: ARKANSAS

| ZIP CODE | QUARTER 1 | QUARTER 2 | QUARTER 3 | QUARTER 4 | TOTAL GRAMS |
|---|---|---|---|---|---|

DRUG CODE:2315DRUG NAME:SECOBARBITAL (SCHEDULE 2)
STATE:ARKANSAS

| ZIP CODE | QUARTER 1 | QUARTER 2 | QUARTER 3 | QUARTER 4 | TOTAL GRAMS |
|---|---|---|---|---|---|
| 726 | 0 | 0 | 0 | 18.31 | 18.31 |
| TOTAL | 0 | 0 | 0 | 18.31 | 18.31 |

STATE: CALIFORNIA

| ZIP CODE | QUARTER 1 | QUARTER 2 | QUARTER 3 | QUARTER 4 | TOTAL GRAMS |
|---|---|---|---|---|---|
| 900 | 9.16 | 9.16 | 9.16 | 9.16 | 36.64 |
| 904 | 0 | 9.16 | 0 | 0 | 9.16 |
| 906 | 9.16 | 45.78 | 27.47 | 27.47 | 109.88 |
| 908 | 0 | 0 | 0 | 9.16 | 9.16 |
| 911 | 0 | 9.16 | 0 | 0 | 9.16 |
| 914 | 9.16 | 0 | 9.16 | 9.16 | 27.48 |
| 917 | 0 | 9.16 | 9.16 | 0 | 18.32 |
| 920 | 0 | 45.78 | 45.78 | 64.09 | 155.65 |
| 921 | 0 | 0 | 18.31 | 0 | 18.31 |
| 922 | 0 | 9.16 | 0 | 0 | 9.16 |
| 926 | 0 | 9.16 | 9.16 | 9.16 | 27.48 |
| 928 | 0 | 82.4 | 311.27 | 173.95 | 567.62 |
| 931 | 0 | 0 | 0 | 27.47 | 27.47 |
| 934 | 0 | 0 | 0 | 9.16 | 9.16 |
| 939 | 0 | 0 | 0 | 18.31 | 18.31 |
| 940 | 0 | 0 | 0 | 18.31 | 18.31 |
| 941 | 27.47 | 137.33 | 485.22 | 512.68 | 1,162.70 |
| 943 | 0 | 0 | 82.4 | 27.47 | 109.87 |
| 944 | 0 | 45.78 | 18.31 | 18.31 | 82.4 |
| 945 | 0 | 27.47 | 27.47 | 18.31 | 73.25 |
| 947 | 0 | 18.31 | 0 | 0 | 18.31 |
| 949 | 0 | 0 | 18.31 | 9.16 | 27.47 |
| 950 | 0 | 9.16 | 36.62 | 27.47 | 73.25 |
| 952 | 0 | 0 | 0 | 9.16 | 9.16 |
| 953 | 0 | 0 | 9.16 | 9.16 | 18.32 |
| 956 | 0 | 18.31 | 18.31 | 18.31 | 54.93 |
| 957 | 0 | 0 | 9.16 | 0 | 9.16 |
| 958 | 9.16 | 0 | 45.78 | 36.62 | 91.56 |
| 961 | 0 | 0 | 9.16 | 0 | 9.16 |
| TOTAL | 64.11 | 485.28 | 1,199.37 | 1,062.05 | 2,810.81 |

STATE: COLORADO

| ZIP CODE | QUARTER 1 | QUARTER 2 | QUARTER 3 | QUARTER 4 | TOTAL GRAMS |
|---|---|---|---|---|---|

| | | | | | |
|---|---|---|---|---|---|
| 801 | 9.16 | 18.31 | 9.16 | 0 | 36.63 |
| 802 | 9.16 | 0 | 0 | 0 | 9.16 |
| 809 | 0 | 0 | 0 | 9.16 | 9.16 |
| TOTAL | 18.32 | 18.31 | 9.16 | 9.16 | 54.95 |

STATE: CONNECTICUT

| ZIP CODE | QUARTER 1 | QUARTER 2 | QUARTER 3 | QUARTER 4 | TOTAL GRAMS |
|---|---|---|---|---|---|
| 60 | 0 | 9.16 | 0 | 0 | 9.16 |
| 64 | 0 | 18.31 | 9.16 | 9.16 | 36.63 |
| TOTAL | 0 | 27.47 | 9.16 | 9.16 | 45.79 |

STATE: DELAWARE

| ZIP CODE | QUARTER 1 | QUARTER 2 | QUARTER 3 | QUARTER 4 | TOTAL GRAMS |
|---|---|---|---|---|---|
| 197 | 0 | 0 | 0 | 9.16 | 9.16 |
| TOTAL | 0 | 0 | 0 | 9.16 | 9.16 |

STATE: FLORIDA

| ZIP CODE | QUARTER 1 | QUARTER 2 | QUARTER 3 | QUARTER 4 | TOTAL GRAMS |
|---|---|---|---|---|---|
| 330 | 0 | 9.16 | 27.47 | 18.31 | 54.94 |
| 331 | 18.31 | 9.16 | 18.31 | 18.31 | 64.09 |
| 334 | 18.31 | 36.62 | 27.47 | 36.62 | 119.02 |
| 337 | 9.16 | 36.62 | 9.16 | 9.16 | 64.1 |
| 349 | 0 | 0 | 0 | 9.16 | 9.16 |
| TOTAL | 45.78 | 91.56 | 82.41 | 91.56 | 311.31 |

STATE: GEORGIA

| ZIP CODE | QUARTER 1 | QUARTER 2 | QUARTER 3 | QUARTER 4 | TOTAL GRAMS |
|---|---|---|---|---|---|
| 302 | 9.16 | 18.31 | 9.16 | 18.31 | 54.94 |
| 312 | 9.16 | 0 | 0 | 0 | 9.16 |
| TOTAL | 18.32 | 18.31 | 9.16 | 18.31 | 64.1 |

STATE: HAWAII

| ZIP CODE | QUARTER 1 | QUARTER 2 | QUARTER 3 | QUARTER 4 | TOTAL GRAMS |
|---|---|---|---|---|---|
| 967 | 18.31 | 18.31 | 9.16 | 0 | 45.78 |
| 968 | 0 | 9.16 | 0 | 9.16 | 18.32 |

DRUG CODE:2315DRUG NAME:SECOBARBITAL (SCHEDULE 2)
STATE:HAWAII

| ZIP CODE | QUARTER 1 | QUARTER 2 | QUARTER 3 | QUARTER 4 | TOTAL GRAMS |
|---|---|---|---|---|---|
| TOTAL | 18.31 | 27.47 | 9.16 | 9.16 | 64.1 |

STATE: ILLINOIS

| ZIP CODE | QUARTER 1 | QUARTER 2 | QUARTER 3 | QUARTER 4 | TOTAL GRAMS |
|---|---|---|---|---|---|
| 600 | 9.16 | 9.16 | 18.31 | 18.31 | 54.94 |
| 601 | 9.16 | 0 | 0 | 0 | 9.16 |
| 604 | 0 | 0 | 9.16 | 0 | 9.16 |
| 605 | 9.16 | 0 | 0 | 0 | 9.16 |
| 606 | 45.78 | 73.24 | 64.09 | 45.78 | 228.89 |
| 617 | 9.16 | 0 | 0 | 0 | 9.16 |
| TOTAL | 82.42 | 82.4 | 91.56 | 64.09 | 320.47 |

STATE: KANSAS

| ZIP CODE | QUARTER 1 | QUARTER 2 | QUARTER 3 | QUARTER 4 | TOTAL GRAMS |
|---|---|---|---|---|---|
| 664 | 9.16 | 0 | 0 | 9.16 | 18.32 |
| 667 | 9.16 | 27.47 | 9.16 | 0 | 45.79 |
| 672 | 9.16 | 0 | 0 | 0 | 9.16 |
| TOTAL | 27.48 | 27.47 | 9.16 | 9.16 | 73.27 |

RETAIL DRUG DISTRIBUTION BY ZIP CODE WITHIN STATE BY GRAMS WT

STATE: LOUISIANA

| ZIP CODE | QUARTER 1 | QUARTER 2 | QUARTER 3 | QUARTER 4 | TOTAL GRAMS |
|---|---|---|---|---|---|
| 700 | 0 | 9.16 | 0 | 0 | 9.16 |
| 706 | 9.16 | 18.31 | 18.31 | 0 | 45.78 |
| TOTAL | 9.16 | 27.47 | 18.31 | 0 | 54.94 |

STATE: MARYLAND

| ZIP CODE | QUARTER 1 | QUARTER 2 | QUARTER 3 | QUARTER 4 | TOTAL GRAMS |
|---|---|---|---|---|---|
| 207 | 9.16 | 0 | 0 | 0 | 9.16 |
| 209 | 9.16 | 9.16 | 18.31 | 9.16 | 45.79 |
| 211 | 9.16 | 0 | 0 | 0 | 9.16 |
| 217 | 9.16 | 36.62 | 45.78 | 27.47 | 119.03 |
| TOTAL | 36.64 | 45.78 | 64.09 | 36.63 | 183.14 |

STATE: MASSACHUSETTS

| ZIP CODE | QUARTER 1 | QUARTER 2 | QUARTER 3 | QUARTER 4 | TOTAL GRAMS |
|---|---|---|---|---|---|
| 10 | 0 | 0 | 0 | 9.16 | 9.16 |
| 13 | 9.16 | 0 | 0 | 0 | 9.16 |
| 14 | 0 | 9.16 | 0 | 0 | 9.16 |
| 15 | 0 | 18.31 | 9.16 | 0 | 27.47 |
| 16 | 0 | 18.31 | 27.47 | 9.16 | 54.94 |
| 17 | 9.16 | 9.16 | 9.16 | 18.31 | 45.79 |
| 19 | 0 | 0 | 9.16 | 0 | 9.16 |
| 20 | 0 | 0 | 0 | 9.16 | 9.16 |
| 24 | 0 | 27.47 | 18.31 | 0 | 45.78 |
| TOTAL | 18.32 | 82.41 | 73.26 | 45.79 | 219.78 |

STATE: MICHIGAN

| ZIP CODE | QUARTER 1 | QUARTER 2 | QUARTER 3 | QUARTER 4 | TOTAL GRAMS |
|---|---|---|---|---|---|
| 480 | 0 | 0 | 9.16 | 0 | 9.16 |
| 481 | 0 | 0 | 0 | 9.16 | 9.16 |
| TOTAL | 0 | 0 | 9.16 | 9.16 | 18.32 |

STATE: MISSOURI

| ZIP CODE | QUARTER 1 | QUARTER 2 | QUARTER 3 | QUARTER 4 | TOTAL GRAMS |
|---|---|---|---|---|---|
| 637 | 18.31 | 9.16 | 18.31 | 9.16 | 54.94 |
| 641 | 0 | 27.47 | 0 | 9.16 | 36.63 |
| 647 | 18.31 | 0 | 0 | 0 | 18.31 |
| 654 | 9.16 | 9.16 | 9.16 | 9.16 | 36.64 |
| TOTAL | 45.78 | 45.79 | 27.47 | 27.48 | 146.52 |

STATE: NEVADA

| ZIP CODE | QUARTER 1 | QUARTER 2 | QUARTER 3 | QUARTER 4 | TOTAL GRAMS |
|---|---|---|---|---|---|
| 891 | 0 | 0 | 9.16 | 0 | 9.16 |
| TOTAL | 0 | 0 | 9.16 | 0 | 9.16 |

STATE: NEW HAMPSHIRE

| ZIP CODE | QUARTER 1 | QUARTER 2 | QUARTER 3 | QUARTER 4 | TOTAL GRAMS |
|---|---|---|---|---|---|
| 30 | 9.16 | 9.16 | 18.31 | 0 | 36.63 |
| TOTAL | 9.16 | 9.16 | 18.31 | 0 | 36.63 |

STATE: NEW JERSEY

| ZIP CODE | QUARTER 1 | QUARTER 2 | QUARTER 3 | QUARTER 4 | TOTAL GRAMS |
|---|---|---|---|---|---|
| 70 | 18.31 | 45.78 | 45.78 | 73.24 | 183.11 |
| 76 | 0 | 0 | 0 | 18.31 | 18.31 |
| 77 | 0 | 18.31 | 0 | 9.16 | 27.47 |
| 78 | 0 | 9.16 | 0 | 9.16 | 18.32 |

RETAIL DRUG DISTRIBUTION BY ZIP CODE WITHIN STATE BY GRAMS WT

| ZIP CODE | QUARTER 1 | QUARTER 2 | QUARTER 3 | QUARTER 4 | TOTAL GRAMS |
|---|---|---|---|---|---|
| 80 | 0 | 9.16 | 18.31 | 0 | 27.47 |
| 85 | 0 | 0 | 9.16 | 0 | 9.16 |
| 88 | 9.16 | 9.16 | 18.31 | 0 | 36.63 |
| TOTAL | 27.47 | 91.57 | 91.56 | 109.87 | 320.47 |

STATE: NEW MEXICO

| ZIP CODE | QUARTER 1 | QUARTER 2 | QUARTER 3 | QUARTER 4 | TOTAL GRAMS |
|---|---|---|---|---|---|
| 871 | 0 | 9.16 | 18.31 | 9.16 | 36.63 |
| TOTAL | 0 | 9.16 | 18.31 | 9.16 | 36.63 |

STATE: NEW YORK

| ZIP CODE | QUARTER 1 | QUARTER 2 | QUARTER 3 | QUARTER 4 | TOTAL GRAMS |
|---|---|---|---|---|---|
| 100 | 82.4 | 192.26 | 119.02 | 146.48 | 540.16 |
| 101 | 9.16 | 0 | 0 | 9.16 | 18.32 |
| 103 | 9.16 | 27.47 | 36.62 | 9.16 | 82.41 |
| 105 | 9.16 | 36.62 | 18.31 | 0 | 64.09 |
| 106 | 0 | 9.16 | 9.16 | 9.16 | 27.48 |
| 108 | 18.31 | 18.31 | 9.16 | 0 | 45.78 |
| 110 | 27.47 | 27.47 | 18.31 | 9.16 | 82.41 |
| 112 | 54.93 | 164.79 | 173.95 | 155.64 | 549.31 |
| 113 | 9.16 | 0 | 9.16 | 9.16 | 27.48 |
| 114 | 64.09 | 18.31 | 27.47 | 18.31 | 128.18 |
| 116 | 27.47 | 45.78 | 36.62 | 54.93 | 164.8 |
| 117 | 0 | 18.31 | 9.16 | 9.16 | 36.63 |
| 119 | 18.31 | 9.16 | 0 | 9.16 | 36.63 |
| 140 | 0 | 9.16 | 0 | 0 | 9.16 |
| 142 | 0 | 18.31 | 0 | 18.31 | 36.62 |
| 144 | 0 | 0 | 0 | 18.31 | 18.31 |
| TOTAL | 329.62 | 595.11 | 466.94 | 476.1 | 1,867.77 |

STATE: NORTH CAROLINA

| ZIP CODE | QUARTER 1 | QUARTER 2 | QUARTER 3 | QUARTER 4 | TOTAL GRAMS |
|---|---|---|---|---|---|
| 272 | 0 | 0 | 9.16 | 0 | 9.16 |
| 275 | 9.16 | 0 | 9.16 | 0 | 18.32 |
| 280 | 0 | 0 | 9.16 | 0 | 9.16 |
| 283 | 9.16 | 9.16 | 9.16 | 9.16 | 36.64 |
| 288 | 9.16 | 0 | 0 | 0 | 9.16 |
| TOTAL | 27.48 | 9.16 | 36.64 | 9.16 | 82.44 |

STATE: OHIO

| ZIP CODE | QUARTER 1 | QUARTER 2 | QUARTER 3 | QUARTER 4 | TOTAL GRAMS |
|---|---|---|---|---|---|
| 430 | 0 | 9.16 | 9.16 | 0 | 18.32 |
| 440 | 0 | 0 | 9.16 | 0 | 9.16 |
| 441 | 9.16 | 9.16 | 0 | 0 | 18.32 |
| 448 | 9.16 | 27.47 | 27.47 | 27.47 | 91.57 |
| 450 | 18.31 | 9.16 | 9.16 | 9.16 | 45.79 |
| 452 | 9.16 | 0 | 0 | 9.16 | 18.32 |
| TOTAL | 45.79 | 54.95 | 54.95 | 45.79 | 201.48 |

STATE: OREGON

| ZIP CODE | QUARTER 1 | QUARTER 2 | QUARTER 3 | QUARTER 4 | TOTAL GRAMS |
|---|---|---|---|---|---|
| 970 | 9.16 | 27.47 | 9.16 | 27.47 | 73.26 |
| 971 | 9.16 | 0 | 0 | 0 | 9.16 |
| 972 | 91.55 | 201.41 | 73.24 | 82.4 | 448.6 |
| 973 | 27.47 | 146.48 | 54.93 | 36.62 | 265.5 |
| 974 | 18.31 | 82.4 | 36.62 | 18.31 | 155.64 |
| 975 | 9.16 | 36.62 | 18.31 | 0 | 64.09 |
| 977 | 0 | 0 | 0 | 9.16 | 9.16 |

| | | | | | |
|---|---|---|---|---|---|
| 978 | 0 | 0 | 9.16 | 0 | 9.16 |
| TOTAL | 164.81 | 494.38 | 201.42 | 173.96 | 1,034.57 |

STATE: PENNSYLVANIA

| ZIP CODE | QUARTER 1 | QUARTER 2 | QUARTER 3 | QUARTER 4 | TOTAL GRAMS |
|---|---|---|---|---|---|
| 150 | 9.16 | 9.16 | 0 | 0 | 18.32 |
| 156 | 0 | 9.16 | 0 | 0 | 9.16 |
| 173 | 0 | 36.62 | 9.16 | 18.31 | 64.09 |
| 181 | 18.31 | 18.31 | 9.16 | 0 | 45.78 |
| 187 | 27.47 | 18.31 | 9.16 | 9.16 | 64.1 |
| 190 | 0 | 18.31 | 27.47 | 36.62 | 82.4 |
| TOTAL | 54.94 | 109.87 | 54.95 | 64.09 | 283.85 |

STATE: RHODE ISLAND

| ZIP CODE | QUARTER 1 | QUARTER 2 | QUARTER 3 | QUARTER 4 | TOTAL GRAMS |
|---|---|---|---|---|---|

DRUG CODE:2315DRUG NAME:SECOBARBITAL (SCHEDULE 2)
STATE:RHODE ISLAND

| ZIP CODE | QUARTER 1 | QUARTER 2 | QUARTER 3 | QUARTER 4 | TOTAL GRAMS |
|---|---|---|---|---|---|
| 28 | 18.31 | 9.16 | 18.31 | 0 | 45.78 |
| TOTAL | 18.31 | 9.16 | 18.31 | 0 | 45.78 |

STATE: TEXAS

| ZIP CODE | QUARTER 1 | QUARTER 2 | QUARTER 3 | QUARTER 4 | TOTAL GRAMS |
|---|---|---|---|---|---|
| 750 | 0 | 9.16 | 18.31 | 0 | 27.47 |
| 762 | 0 | 9.16 | 0 | 0 | 9.16 |
| 770 | 9.16 | 0 | 0 | 9.16 | 18.32 |
| 778 | 0 | 9.16 | 0 | 0 | 9.16 |
| 786 | 0 | 0 | 9.16 | 0 | 9.16 |
| TOTAL | 9.16 | 27.48 | 27.47 | 9.16 | 73.27 |

STATE: VERMONT

| ZIP CODE | QUARTER 1 | QUARTER 2 | QUARTER 3 | QUARTER 4 | TOTAL GRAMS |
|---|---|---|---|---|---|
| 54 | 0 | 0 | 0 | 9.16 | 9.16 |
| 57 | 0 | 9.16 | 36.62 | 54.93 | 100.71 |
| TOTAL | 0 | 9.16 | 36.62 | 64.09 | 109.87 |

STATE: VIRGINIA

| ZIP CODE | QUARTER 1 | QUARTER 2 | QUARTER 3 | QUARTER 4 | TOTAL GRAMS |
|---|---|---|---|---|---|
| 201 | 0 | 0 | 0 | 9.16 | 9.16 |
| 220 | 9.16 | 0 | 9.16 | 9.16 | 27.48 |
| 235 | 18.31 | 27.47 | 18.31 | 27.47 | 91.56 |
| TOTAL | 27.47 | 27.47 | 27.47 | 45.79 | 128.2 |

STATE: WASHINGTON

| ZIP CODE | QUARTER 1 | QUARTER 2 | QUARTER 3 | QUARTER 4 | TOTAL GRAMS |
|---|---|---|---|---|---|
| 980 | 0 | 0 | 9.16 | 0 | 9.16 |
| 981 | 109.86 | 155.64 | 64.09 | 219.72 | 549.31 |
| 982 | 9.16 | 9.16 | 9.16 | 0 | 27.48 |
| 983 | 9.16 | 9.16 | 18.31 | 18.31 | 54.94 |
| 984 | 45.78 | 18.31 | 0 | 0 | 64.09 |
| 985 | 9.16 | 9.16 | 9.16 | 9.16 | 36.64 |
| 986 | 9.16 | 27.47 | 9.16 | 0 | 45.79 |
| 988 | 0 | 9.16 | 0 | 9.16 | 18.32 |
| 989 | 0 | 0 | 0 | 9.16 | 9.16 |
| TOTAL | 192.28 | 238.06 | 119.04 | 265.51 | 814.89 |

STATE: WEST VIRGINIA

| ZIP CODE | QUARTER 1 | QUARTER 2 | QUARTER 3 | QUARTER 4 | TOTAL GRAMS |
|---|---|---|---|---|---|
| 260 | 0 | 9.16 | 0 | 0 | 9.16 |

| | | | | | |
|---|---|---|---|---|---|
| TOTAL | 0 | 9.16 | 0 | 0 | 9.16 |
| STATE: | WYOMING | | | | |
| ZIP CODE | QUARTER 1 | QUARTER 2 | QUARTER 3 | QUARTER 4 | TOTAL GRAMS |
| 825 | 0 | 9.16 | 9.16 | 0 | 18.32 |
| TOTAL | 0 | 9.16 | 9.16 | 0 | 18.32 |

DRUG CODE: 7379 DRUG NAME: NABILONE

STATE: ARIZONA

| ZIP CODE | QUARTER 1 | QUARTER 2 | QUARTER 3 | QUARTER 4 | TOTAL GRAMS |
|---|---|---|---|---|---|
| 852 | 0 | 0 | 0 | 0.15 | 0.15 |
| TOTAL | 0 | 0 | 0 | 0.15 | 0.15 |

STATE: CALIFORNIA

| ZIP CODE | QUARTER 1 | QUARTER 2 | QUARTER 3 | QUARTER 4 | TOTAL GRAMS |
|---|---|---|---|---|---|
| 910 | 0 | 0 | 0 | 0.05 | 0.05 |
| TOTAL | 0 | 0 | 0 | 0.05 | 0.05 |

STATE: COLORADO

| ZIP CODE | QUARTER 1 | QUARTER 2 | QUARTER 3 | QUARTER 4 | TOTAL GRAMS |
|---|---|---|---|---|---|
| 801 | 0.1 | 0.15 | 0.05 | 0.1 | 0.4 |
| 802 | 0.05 | 0 | 0 | 0 | 0.05 |
| 815 | 0.15 | 0.15 | 0.2 | 0.1 | 0.6 |
| TOTAL | 0.3 | 0.3 | 0.25 | 0.2 | 1.05 |

STATE: FLORIDA

| ZIP CODE | QUARTER 1 | QUARTER 2 | QUARTER 3 | QUARTER 4 | TOTAL GRAMS |
|---|---|---|---|---|---|
| 327 | 0.25 | 0 | 0 | 0 | 0.25 |
| 330 | 0 | 0 | 0.1 | 0.15 | 0.25 |
| 331 | 0 | 0 | 0.1 | 0 | 0.1 |
| 335 | 0.65 | 0.15 | 0.6 | 0.4 | 1.8 |
| 336 | 0 | 0.1 | 0 | 0.1 | 0.2 |
| 337 | 0.1 | 0.05 | 0.1 | 0.1 | 0.35 |
| 338 | 0 | 0 | 0.2 | 0 | 0.2 |
| 339 | 0.1 | 0.05 | 0.1 | 0 | 0.25 |

DRUG CODE:7379DRUG NAME:NABILONE
STATE:FLORIDA

| ZIP CODE | QUARTER 1 | QUARTER 2 | QUARTER 3 | QUARTER 4 | TOTAL GRAMS |
|---|---|---|---|---|---|
| 346 | 0.1 | 0 | 0 | 0 | 0.1 |
| TOTAL | 1.2 | 0.35 | 1.2 | 0.75 | 3.5 |

STATE: HAWAII

| ZIP CODE | QUARTER 1 | QUARTER 2 | QUARTER 3 | QUARTER 4 | TOTAL GRAMS |
|---|---|---|---|---|---|
| 968 | 0 | 0.1 | 0 | 0.1 | 0.2 |
| TOTAL | 0 | 0.1 | 0 | 0.1 | 0.2 |

STATE: ILLINOIS

| ZIP CODE | QUARTER 1 | QUARTER 2 | QUARTER 3 | QUARTER 4 | TOTAL GRAMS |
|---|---|---|---|---|---|
| 600 | 0.1 | 0.05 | 0.05 | 0 | 0.2 |
| 605 | 0 | 0.05 | 0 | 0 | 0.05 |
| 606 | 0.1 | 0 | 0.1 | 0.15 | 0.35 |
| 617 | 0 | 0 | 0.2 | 0 | 0.2 |
| TOTAL | 0.2 | 0.1 | 0.35 | 0.15 | 0.8 |

STATE: INDIANA

| ZIP CODE | QUARTER 1 | QUARTER 2 | QUARTER 3 | QUARTER 4 | TOTAL GRAMS |
|---|---|---|---|---|---|
| 471 | 0 | 0 | 0 | 0.05 | 0.05 |

| | | | | | |
|---|---|---|---|---|---|
| 475 | 0 | 0.1 | 0.15 | 0 | 0.25 |
| TOTAL | 0 | 0.1 | 0.15 | 0.05 | 0.3 |
| STATE: | IOWA | | | | |
| ZIP CODE | QUARTER 1 | QUARTER 2 | QUARTER 3 | QUARTER 4 | TOTAL GRAMS |
| 500 | 0 | 0 | 0.05 | 0 | 0.05 |
| TOTAL | 0 | 0 | 0.05 | 0 | 0.05 |
| STATE: | MAINE | | | | |
| ZIP CODE | QUARTER 1 | QUARTER 2 | QUARTER 3 | QUARTER 4 | TOTAL GRAMS |
| 44 | 0 | 0 | 0 | 0.1 | 0.1 |
| TOTAL | 0 | 0 | 0 | 0.1 | 0.1 |
| STATE: | MARYLAND | | | | |
| ZIP CODE | QUARTER 1 | QUARTER 2 | QUARTER 3 | QUARTER 4 | TOTAL GRAMS |
| 206 | 0.1 | 0 | 0 | 0 | 0.1 |
| 207 | 0.05 | 0.05 | 0 | 0 | 0.1 |
| 208 | 0.9 | 0.8 | 1 | 0.85 | 3.55 |
| 210 | 0.05 | 0 | 0 | 0 | 0.05 |
| 214 | 0.05 | 0.2 | 0 | 0 | 0.25 |
| 216 | 0.5 | 0.3 | 0.2 | 0.3 | 1.3 |
| TOTAL | 1.65 | 1.35 | 1.2 | 1.15 | 5.35 |
| STATE: | MASSACHUSETTS | | | | |
| ZIP CODE | QUARTER 1 | QUARTER 2 | QUARTER 3 | QUARTER 4 | TOTAL GRAMS |
| 16 | 0 | 0 | 0.1 | 0 | 0.1 |
| 24 | 0 | 0.25 | 0.25 | 0.3 | 0.8 |
| TOTAL | 0 | 0.25 | 0.35 | 0.3 | 0.9 |
| STATE: | MINNESOTA | | | | |
| ZIP CODE | QUARTER 1 | QUARTER 2 | QUARTER 3 | QUARTER 4 | TOTAL GRAMS |
| 558 | 0.05 | 0.05 | 0.15 | 0.05 | 0.3 |
| TOTAL | 0.05 | 0.05 | 0.15 | 0.05 | 0.3 |
| STATE: | MISSOURI | | | | |
| ZIP CODE | QUARTER 1 | QUARTER 2 | QUARTER 3 | QUARTER 4 | TOTAL GRAMS |
| 631 | 0 | 0.05 | 0 | 0 | 0.05 |
| 641 | 0 | 0.2 | 0.1 | 0.05 | 0.35 |
| 658 | 0.05 | 0 | 0 | 0 | 0.05 |
| TOTAL | 0.05 | 0.25 | 0.1 | 0.05 | 0.45 |
| STATE: | NEBRASKA | | | | |
| ZIP CODE | QUARTER 1 | QUARTER 2 | QUARTER 3 | QUARTER 4 | TOTAL GRAMS |
| 681 | 0.2 | 0.15 | 0.2 | 0.15 | 0.7 |
| TOTAL | 0.2 | 0.15 | 0.2 | 0.15 | 0.7 |
| STATE: | NEW HAMPSHIRE | | | | |
| ZIP CODE | QUARTER 1 | QUARTER 2 | QUARTER 3 | QUARTER 4 | TOTAL GRAMS |
| 30 | 0 | 0.1 | 0.15 | 0 | 0.25 |
| TOTAL | 0 | 0.1 | 0.15 | 0 | 0.25 |
| STATE: | NEW JERSEY | | | | |
| ZIP CODE | QUARTER 1 | QUARTER 2 | QUARTER 3 | QUARTER 4 | TOTAL GRAMS |
| 81 | 0 | 0 | 0 | 0.05 | 0.05 |
| 87 | 0 | 0 | 0.05 | 0 | 0.05 |
| 88 | 0 | 0 | 0 | 0.05 | 0.05 |
| TOTAL | 0 | 0 | 0.05 | 0.1 | 0.15 |
| STATE: | NEW MEXICO | | | | |
| ZIP CODE | QUARTER 1 | QUARTER 2 | QUARTER 3 | QUARTER 4 | TOTAL GRAMS |

DRUG CODE:7379DRUG NAME:NABILONE
STATE:NEW MEXICO

| ZIP CODE | QUARTER 1 | QUARTER 2 | QUARTER 3 | QUARTER 4 | TOTAL GRAMS |
|---|---|---|---|---|---|
| 871 | 0 | 0.15 | 0 | 0.15 | 0.3 |
| TOTAL | 0 | 0.15 | 0 | 0.15 | 0.3 |

STATE: NEW YORK

| ZIP CODE | QUARTER 1 | QUARTER 2 | QUARTER 3 | QUARTER 4 | TOTAL GRAMS |
|---|---|---|---|---|---|
| 129 | 0 | 0 | 0 | 0.05 | 0.05 |
| 132 | 0.05 | 0.2 | 0.1 | 0.25 | 0.6 |
| 134 | 0 | 0 | 0 | 0.05 | 0.05 |
| 135 | 0 | 0 | 0 | 0.1 | 0.1 |
| 137 | 0 | 0 | 0 | 0.05 | 0.05 |
| 146 | 0.3 | 0 | 0 | 0 | 0.3 |
| TOTAL | 0.35 | 0.2 | 0.1 | 0.5 | 1.15 |

STATE: NORTH CAROLINA

| ZIP CODE | QUARTER 1 | QUARTER 2 | QUARTER 3 | QUARTER 4 | TOTAL GRAMS |
|---|---|---|---|---|---|
| 277 | 0 | 0.15 | 0.25 | 0.15 | 0.55 |
| TOTAL | 0 | 0.15 | 0.25 | 0.15 | 0.55 |

STATE: OHIO

| ZIP CODE | QUARTER 1 | QUARTER 2 | QUARTER 3 | QUARTER 4 | TOTAL GRAMS |
|---|---|---|---|---|---|
| 430 | 0.2 | 0.05 | 0.2 | 0.1 | 0.55 |
| 432 | 0.05 | 0 | 0 | 0 | 0.05 |
| 450 | 0.15 | 0 | 0 | 0 | 0.15 |
| TOTAL | 0.4 | 0.05 | 0.2 | 0.1 | 0.75 |

STATE: PENNSYLVANIA

| ZIP CODE | QUARTER 1 | QUARTER 2 | QUARTER 3 | QUARTER 4 | TOTAL GRAMS |
|---|---|---|---|---|---|
| 157 | 0 | 0.05 | 0 | 0 | 0.05 |
| 165 | 0.1 | 0 | 0 | 0 | 0.1 |
| 187 | 0.15 | 0.3 | 0.2 | 0.2 | 0.85 |
| 191 | 0 | 0 | 0 | 0.05 | 0.05 |
| TOTAL | 0.25 | 0.35 | 0.2 | 0.25 | 1.05 |

STATE: RHODE ISLAND

| ZIP CODE | QUARTER 1 | QUARTER 2 | QUARTER 3 | QUARTER 4 | TOTAL GRAMS |
|---|---|---|---|---|---|
| 28 | 0 | 0.05 | 0 | 0 | 0.05 |
| TOTAL | 0 | 0.05 | 0 | 0 | 0.05 |

STATE: VIRGINIA

| ZIP CODE | QUARTER 1 | QUARTER 2 | QUARTER 3 | QUARTER 4 | TOTAL GRAMS |
|---|---|---|---|---|---|
| 220 | 0 | 0.05 | 0 | 0 | 0.05 |
| 222 | 0.1 | 0.2 | 0.1 | 0.2 | 0.6 |
| 223 | 0 | 0.1 | 0.25 | 0 | 0.35 |
| TOTAL | 0.1 | 0.35 | 0.35 | 0.2 | 1 |

STATE: WISCONSIN

| ZIP CODE | QUARTER 1 | QUARTER 2 | QUARTER 3 | QUARTER 4 | TOTAL GRAMS |
|---|---|---|---|---|---|
| 530 | 0.05 | 0 | 0 | 0 | 0.05 |
| TOTAL | 0.05 | 0 | 0 | 0 | 0.05 |

DRUG CODE:7471DRUG NAME:PHENCYCLIDINE (PCP)
STATE: MINNESOTA

| ZIP CODE | QUARTER 1 | QUARTER 2 | QUARTER 3 | QUARTER 4 | TOTAL GRAMS |
|---|---|---|---|---|---|
| 554 | 0 | 0 | 0 | 0.22 | 0.22 |
| TOTAL | 0 | 0 | 0 | 0.22 | 0.22 |

RETAIL DRUG DISTRIBUTION BY ZIP CODE WITHIN STATE BY GRAMS WT

DRUG CODE:9041LDRUG NAME:COCAINE

STATE: ALABAMA

| ZIP CODE | QUARTER 1 | QUARTER 2 | QUARTER 3 | QUARTER 4 | TOTAL GRAMS |
|---|---|---|---|---|---|
| 350 | 0 | 0.14 | 0.43 | 1 | 1.57 |
| 351 | 0.29 | 0.57 | 0 | 1.86 | 2.72 |
| 352 | 14.14 | 13.71 | 13 | 16.28 | 57.13 |
| 354 | 0.57 | 0.71 | 1 | 1.14 | 3.42 |
| 356 | 1.43 | 1.71 | 3.57 | 2.57 | 9.28 |
| 357 | 1.14 | 0 | 1.14 | 1.71 | 3.99 |
| 358 | 2.14 | 3.29 | 3.14 | 3.29 | 11.86 |
| 359 | 0.29 | 2.43 | 1.86 | 0.86 | 5.44 |
| 360 | 2 | 0.86 | 0.86 | 1.71 | 5.43 |
| 361 | 19.28 | 18.28 | 19.43 | 20.57 | 77.56 |
| 362 | 2.43 | 0 | 3.71 | 0.71 | 6.85 |
| 364 | 0.71 | 0.86 | 0.14 | 1.29 | 3 |
| 365 | 3.14 | 7.14 | 6 | 8.57 | 24.85 |
| 366 | 6.57 | 4.43 | 4.71 | 9.57 | 25.28 |
| 367 | 0 | 0 | 0 | 0.29 | 0.29 |

DRUG CODE:9041LDRUG NAME:COCAINE
STATE:ALABAMA

| ZIP CODE | QUARTER 1 | QUARTER 2 | QUARTER 3 | QUARTER 4 | TOTAL GRAMS |
|---|---|---|---|---|---|
| 368 | 1.29 | 4.43 | 1 | 2.57 | 9.29 |
| TOTAL | 55.42 | 58.56 | 59.99 | 73.99 | 247.96 |

STATE: ALASKA

| ZIP CODE | QUARTER 1 | QUARTER 2 | QUARTER 3 | QUARTER 4 | TOTAL GRAMS |
|---|---|---|---|---|---|
| 995 | 4.29 | 3.29 | 6.14 | 6.43 | 20.15 |
| 996 | 0.14 | 1.14 | 1.86 | 0.43 | 3.57 |
| 997 | 13.21 | 6.43 | 4.29 | 14.5 | 38.43 |
| 998 | 0.14 | 0 | 0 | 0.43 | 0.57 |
| 999 | 0 | 0.14 | 0.29 | 0.14 | 0.57 |
| TOTAL | 17.78 | 11 | 12.58 | 21.93 | 63.29 |

STATE: ARIZONA

| ZIP CODE | QUARTER 1 | QUARTER 2 | QUARTER 3 | QUARTER 4 | TOTAL GRAMS |
|---|---|---|---|---|---|
| 850 | 38.57 | 43.32 | 40 | 36.14 | 158.03 |
| 851 | 4.71 | 1.71 | 0.57 | 2.86 | 9.85 |
| 852 | 54.14 | 42.78 | 36.07 | 46.85 | 179.84 |
| 853 | 11.71 | 9.29 | 10.14 | 15.14 | 46.28 |
| 855 | 0 | 0 | 0 | 0.29 | 0.29 |
| 856 | 2.29 | 3.86 | 2.29 | 2.43 | 10.87 |
| 857 | 12 | 15 | 14 | 14.14 | 55.14 |
| 859 | 0.43 | 0.57 | 1 | 0.86 | 2.86 |
| 860 | 4.43 | 10.14 | 8.14 | 5.71 | 28.42 |
| 863 | 1.29 | 0.14 | 1 | 0.57 | 3 |
| 864 | 1.57 | 1.29 | 2.29 | 2.14 | 7.29 |
| 865 | 0 | 0.57 | 0.14 | 0.86 | 1.57 |
| TOTAL | 131.14 | 128.67 | 115.64 | 127.99 | 503.44 |

STATE: ARKANSAS

| ZIP CODE | QUARTER 1 | QUARTER 2 | QUARTER 3 | QUARTER 4 | TOTAL GRAMS |
|---|---|---|---|---|---|
| 716 | 0 | 0 | 0.29 | 0 | 0.29 |
| 719 | 1.43 | 1.14 | 0 | 0.57 | 3.14 |
| 720 | 25.71 | 7.71 | 7.43 | 10.14 | 50.99 |
| 721 | 1.43 | 2.29 | 1.57 | 1.43 | 6.72 |
| 722 | 1 | 3.86 | 2.43 | 0.86 | 8.15 |
| 723 | 4.61 | 0 | 0.57 | 0 | 5.18 |
| 724 | 12.14 | 16.46 | 17.89 | 12.86 | 59.35 |

| | | | | | |
|---|---|---|---|---|---|
| 725 | 1.43 | 1.71 | 1.86 | 1.43 | 6.43 |
| 726 | 1.57 | 2.14 | 2.43 | 3.86 | 10 |
| 727 | 15.39 | 15.39 | 11.78 | 6.75 | 49.31 |
| 728 | 0.14 | 0 | 0.43 | 0.57 | 1.14 |
| 729 | 2.86 | 3.43 | 3.14 | 5.86 | 15.29 |
| TOTAL | 67.71 | 54.13 | 49.82 | 44.33 | 215.99 |

STATE: CALIFORNIA

| ZIP CODE | QUARTER 1 | QUARTER 2 | QUARTER 3 | QUARTER 4 | TOTAL GRAMS |
|---|---|---|---|---|---|
| 900 | 48.21 | 59.25 | 45.78 | 56.32 | 209.56 |
| 902 | 34.07 | 26.71 | 28.14 | 27.57 | 116.49 |
| 903 | 0.86 | 0.86 | 0.86 | 0.29 | 2.87 |
| 904 | 10.29 | 23.28 | 21.64 | 17 | 72.21 |
| 905 | 15.14 | 12 | 16.14 | 15.57 | 58.85 |
| 906 | 1.43 | 1.14 | 3.29 | 2 | 7.86 |
| 907 | 10.57 | 10 | 16.61 | 7.57 | 44.75 |
| 908 | 20.57 | 16.43 | 16 | 18.86 | 71.86 |
| 910 | 6.86 | 8.14 | 9.71 | 7.29 | 32 |
| 911 | 3.14 | 4.14 | 2.71 | 4.14 | 14.13 |
| 912 | 8.71 | 10.29 | 5.43 | 11.71 | 36.14 |
| 913 | 58.25 | 27.14 | 23.28 | 42.07 | 150.74 |
| 914 | 9.71 | 11.28 | 10.57 | 10.57 | 42.13 |
| 917 | 19.57 | 16.57 | 12.28 | 21.43 | 69.85 |
| 918 | 0.86 | 0 | 0.86 | 0.86 | 2.58 |
| 919 | 12.78 | 17.78 | 15.57 | 7.29 | 53.42 |
| 920 | 39 | 45.32 | 45.93 | 25.71 | 155.96 |
| 921 | 64.42 | 59 | 50.93 | 60.57 | 234.92 |
| 922 | 12.86 | 9.29 | 12.57 | 17.14 | 51.86 |
| 923 | 9.71 | 12.28 | 11 | 14.57 | 47.56 |
| 925 | 13.03 | 7.43 | 10.86 | 8.57 | 39.89 |
| 926 | 31.36 | 27.36 | 24.57 | 30.64 | 113.93 |
| 927 | 9 | 6.14 | 8.43 | 8.57 | 32.14 |
| 928 | 16.46 | 23.21 | 27.86 | 18.28 | 85.81 |

DRUG CODE:9041LDRUG NAME:COCAINE
STATE:CALIFORNIA

| ZIP CODE | QUARTER 1 | QUARTER 2 | QUARTER 3 | QUARTER 4 | TOTAL GRAMS |
|---|---|---|---|---|---|
| 930 | 6.29 | 7.14 | 2.86 | 7 | 23.29 |
| 931 | 15.57 | 8 | 13.14 | 14.57 | 51.28 |
| 932 | 28 | 28.43 | 25.07 | 25.71 | 107.21 |
| 933 | 15.86 | 16.43 | 19.28 | 14.43 | 66 |
| 934 | 5 | 6.29 | 10.29 | 5 | 26.58 |
| 935 | 5.71 | 6.29 | 6.57 | 5.86 | 24.43 |
| 936 | 3.29 | 2.14 | 5.86 | 3.43 | 14.72 |
| 937 | 10.71 | 9.86 | 9.14 | 9.71 | 39.42 |
| 939 | 8.71 | 16.28 | 11.71 | 9 | 45.7 |
| 940 | 18.57 | 27.57 | 30.03 | 24.43 | 100.6 |
| 941 | 55.1 | 62.28 | 57.5 | 65.21 | 240.09 |
| 943 | 13.32 | 19.75 | 20.28 | 17.71 | 71.06 |
| 944 | 3.57 | 2.29 | 7 | 7 | 19.86 |
| 945 | 38.21 | 46.07 | 47.64 | 40.43 | 172.35 |
| 946 | 19.57 | 24.14 | 23.28 | 21.43 | 88.42 |
| 947 | 2.29 | 1.57 | 1.86 | 2.71 | 8.43 |
| 948 | 9.71 | 8.57 | 9.86 | 8.14 | 36.28 |
| 949 | 13.43 | 13.28 | 11.86 | 13.28 | 51.85 |
| 950 | 17.28 | 15.57 | 15.57 | 17.57 | 65.99 |
| 951 | 25.43 | 21 | 24.71 | 23.86 | 95 |
| 952 | 15.53 | 16.39 | 17.53 | 16.53 | 65.98 |
| 953 | 26.5 | 27.57 | 30.78 | 31 | 115.85 |

| | | | | | |
|---|---|---|---|---|---|
| 954 | 7.86 | 9.29 | 9 | 7.57 | 33.72 |
| 955 | 2 | 2.14 | 2.71 | 2.57 | 9.42 |
| 956 | 58.64 | 42.43 | 31.57 | 39.71 | 172.35 |
| 957 | 0.57 | 0 | 0.29 | 0.29 | 1.15 |
| 958 | 33.43 | 37.14 | 30.14 | 38.28 | 138.99 |
| 959 | 18.14 | 14.14 | 19.28 | 19.86 | 71.42 |
| 960 | 12.57 | 10.43 | 10.43 | 13.28 | 46.71 |
| 961 | 2.71 | 3.57 | 3 | 3.86 | 13.14 |
| TOTAL | 950.43 | 941.09 | 929.26 | 944.02 | 3,764.80 |

STATE: COLORADO

| ZIP CODE | QUARTER 1 | QUARTER 2 | QUARTER 3 | QUARTER 4 | TOTAL GRAMS |
|---|---|---|---|---|---|
| 800 | 8.14 | 8.14 | 4.86 | 8.29 | 29.43 |
| 801 | 22.53 | 25.57 | 15.43 | 26.86 | 90.39 |
| 802 | 18.71 | 16.14 | 17.43 | 13.57 | 65.85 |
| 803 | 0.57 | 0.57 | 1.29 | 1.57 | 4 |
| 804 | 0.86 | 0 | 1.43 | 1.57 | 3.86 |
| 805 | 29.14 | 33 | 23.86 | 30.28 | 116.28 |
| 806 | 2.86 | 2.71 | 3.29 | 3.71 | 12.57 |
| 807 | 1.43 | 1.57 | 0 | 0.86 | 3.86 |
| 809 | 1.43 | 1.43 | 3.71 | 2 | 8.57 |
| 810 | 5 | 4.43 | 5.86 | 7.71 | 23 |
| 812 | 0 | 0.29 | 0 | 0 | 0.29 |
| 813 | 0.86 | 0.43 | 0.86 | 0.71 | 2.86 |
| 814 | 2.14 | 1.71 | 2.14 | 1.57 | 7.56 |
| 815 | 0.14 | 0.29 | 0 | 0 | 0.43 |
| 816 | 3.29 | 1.43 | 1.14 | 1.14 | 7 |
| TOTAL | 97.1 | 97.71 | 81.3 | 99.84 | 375.95 |

STATE: CONNECTICUT

| ZIP CODE | QUARTER 1 | QUARTER 2 | QUARTER 3 | QUARTER 4 | TOTAL GRAMS |
|---|---|---|---|---|---|
| 60 | 1.14 | 1.14 | 0.57 | 0.57 | 3.42 |
| 61 | 1.36 | 1.43 | 0.64 | 1.43 | 4.86 |
| 62 | 2.71 | 1.57 | 2.43 | 0.86 | 7.57 |
| 63 | 8.71 | 3.43 | 6 | 5.14 | 23.28 |
| 64 | 3.14 | 3.14 | 4.57 | 2.43 | 13.28 |
| 65 | 11.14 | 10.14 | 9 | 13.28 | 43.56 |
| 66 | 1.43 | 2.43 | 1.43 | 1.43 | 6.72 |
| 67 | 2.86 | 2.14 | 1.86 | 3.14 | 10 |
| 68 | 17.14 | 17.57 | 13.14 | 23.14 | 70.99 |
| TOTAL | 49.63 | 42.99 | 39.64 | 51.42 | 183.68 |

STATE: DELAWARE

| ZIP CODE | QUARTER 1 | QUARTER 2 | QUARTER 3 | QUARTER 4 | TOTAL GRAMS |
|---|---|---|---|---|---|
| 197 | 1.29 | 0.29 | 1.57 | 1.43 | 4.58 |
| 198 | 1.43 | 0.29 | 1.43 | 1.14 | 4.29 |
| 199 | 21.14 | 18.57 | 8 | 18 | 65.71 |

DRUG CODE:9041LDRUG NAME:COCAINE
STATE:DELAWARE

| ZIP CODE | QUARTER 1 | QUARTER 2 | QUARTER 3 | QUARTER 4 | TOTAL GRAMS |
|---|---|---|---|---|---|
| TOTAL | 23.86 | 19.15 | 11 | 20.57 | 74.58 |

STATE: DISTRICT OF COLUMBIA

| ZIP CODE | QUARTER 1 | QUARTER 2 | QUARTER 3 | QUARTER 4 | TOTAL GRAMS |
|---|---|---|---|---|---|
| 200 | 27.43 | 18 | 27.43 | 23.14 | 96 |
| 204 | 0 | 1.43 | 0 | 0 | 1.43 |
| TOTAL | 27.43 | 19.43 | 27.43 | 23.14 | 97.43 |

STATE: FLORIDA

RETAIL DRUG DISTRIBUTION BY ZIP CODE WITHIN STATE BY GRAMS WT

| ZIP CODE | QUARTER 1 | QUARTER 2 | QUARTER 3 | QUARTER 4 | TOTAL GRAMS |
|---|---|---|---|---|---|
| 320 | 5.43 | 5.86 | 3.57 | 5.29 | 20.15 |
| 321 | 1.43 | 0.57 | 1.86 | 1.29 | 5.15 |
| 322 | 27.89 | 34.43 | 36.71 | 34.28 | 133.31 |
| 323 | 4.29 | 3.86 | 1 | 3.57 | 12.72 |
| 324 | 0.14 | 0 | 0.29 | 0.57 | 1 |
| 325 | 4.71 | 8.14 | 7 | 5.43 | 25.28 |
| 326 | 4.43 | 5 | 6.57 | 4.57 | 20.57 |
| 327 | 7.86 | 6 | 4.79 | 8.5 | 27.15 |
| 328 | 4 | 3.29 | 8.29 | 0.86 | 16.44 |
| 329 | 6.71 | 3.86 | 6 | 7.86 | 24.43 |
| 330 | 17 | 21.5 | 26.28 | 19.43 | 84.21 |
| 331 | 9.29 | 9 | 9.43 | 11.75 | 39.47 |
| 333 | 17.28 | 14.43 | 16.93 | 19.86 | 68.5 |
| 334 | 30.46 | 44.21 | 34.32 | 28.14 | 137.13 |
| 335 | 6.86 | 12.75 | 4.43 | 5.86 | 29.9 |
| 336 | 84.92 | 76.96 | 57.14 | 66.07 | 285.09 |
| 337 | 10.43 | 6.43 | 5.71 | 11 | 33.57 |
| 338 | 8.29 | 3 | 7.29 | 5.29 | 23.87 |
| 339 | 13.71 | 11.71 | 11.71 | 12.28 | 49.41 |
| 341 | 8.04 | 11.71 | 16.86 | 10.71 | 47.32 |
| 342 | 28.14 | 20.57 | 21.71 | 16.71 | 87.13 |
| 344 | 1.71 | 1.57 | 3 | 1.29 | 7.57 |
| 346 | 5 | 3.86 | 4.86 | 5 | 18.72 |
| 347 | 0.71 | 1.86 | 2.71 | 0.86 | 6.14 |
| 349 | 10.57 | 8.57 | 7 | 7.57 | 33.71 |
| TOTAL | 319.3 | 319.14 | 305.46 | 294.04 | 1,237.94 |

STATE: GEORGIA

| ZIP CODE | QUARTER 1 | QUARTER 2 | QUARTER 3 | QUARTER 4 | TOTAL GRAMS |
|---|---|---|---|---|---|
| 300 | 51.71 | 52.14 | 55.28 | 91.71 | 250.84 |
| 301 | 21 | 25.43 | 21.43 | 26 | 93.86 |
| 302 | 12.57 | 17.18 | 13.21 | 13.71 | 56.67 |
| 303 | 38.71 | 43.85 | 47.71 | 43.85 | 174.12 |
| 304 | 3 | 2 | 2.86 | 3.86 | 11.72 |
| 305 | 3 | 1 | 3 | 6 | 13 |
| 306 | 2.86 | 3.71 | 4 | 4.14 | 14.71 |
| 307 | 1.14 | 1 | 2.29 | 1.14 | 5.57 |
| 308 | 0 | 0 | 0.57 | 0 | 0.57 |
| 309 | 0.57 | 2.29 | 1.29 | 0.71 | 4.86 |
| 310 | 1.43 | 2.14 | 3.86 | 0.71 | 8.14 |
| 312 | 1.86 | 1.14 | 2.14 | 0.57 | 5.71 |
| 314 | 3 | 1.29 | 2.14 | 1.43 | 7.86 |
| 315 | 2.14 | 2.86 | 0.57 | 1.86 | 7.43 |
| 316 | 4.14 | 3.29 | 1.71 | 5 | 14.14 |
| 317 | 8.29 | 15.61 | 13.75 | 12.46 | 50.11 |
| 319 | 4.43 | 1.43 | 1.43 | 3.57 | 10.86 |
| 398 | 0 | 0 | 0.29 | 0 | 0.29 |
| TOTAL | 159.85 | 176.36 | 177.53 | 216.72 | 730.46 |

STATE: GUAM

| ZIP CODE | QUARTER 1 | QUARTER 2 | QUARTER 3 | QUARTER 4 | TOTAL GRAMS |
|---|---|---|---|---|---|
| 969 | 0 | 0 | 0.71 | 0 | 0.71 |
| TOTAL | 0 | 0 | 0.71 | 0 | 0.71 |

STATE: HAWAII

| ZIP CODE | QUARTER 1 | QUARTER 2 | QUARTER 3 | QUARTER 4 | TOTAL GRAMS |
|---|---|---|---|---|---|
| 967 | 5.71 | 7.29 | 8.71 | 11.57 | 33.28 |
| 968 | 11.43 | 14.14 | 16.28 | 13.28 | 55.13 |
| TOTAL | 17.14 | 21.43 | 24.99 | 24.85 | 88.41 |

STATE: IDAHO

### RETAIL DRUG DISTRIBUTION BY ZIP CODE WITHIN STATE BY GRAMS WT

ZIP CODE QUARTER 1 QUARTER 2 QUARTER 3 QUARTER 4 TOTAL GRAMS

DRUG CODE:9041LDRUG NAME:COCAINE
STATE:IDAHO

| ZIP CODE | QUARTER 1 | QUARTER 2 | QUARTER 3 | QUARTER 4 | TOTAL GRAMS |
|---|---|---|---|---|---|
| 832 | 6.14 | 8 | 7.57 | 5.29 | 27 |
| 833 | 11.93 | 11.36 | 14.96 | 18.57 | 56.82 |
| 834 | 36.78 | 10.86 | 32.61 | 8 | 88.25 |
| 835 | 1.86 | 0.29 | 1.71 | 1.14 | 5 |
| 836 | 10.86 | 11 | 13.86 | 12.57 | 48.29 |
| 837 | 10 | 7.36 | 5.14 | 7.79 | 30.29 |
| 838 | 1.71 | 0.29 | 2.18 | 2 | 6.18 |
| TOTAL | 79.28 | 49.16 | 78.03 | 55.36 | 261.83 |

STATE: ILLINOIS

| ZIP CODE | QUARTER 1 | QUARTER 2 | QUARTER 3 | QUARTER 4 | TOTAL GRAMS |
|---|---|---|---|---|---|
| 600 | 33.43 | 23.71 | 23.14 | 31.71 | 111.99 |
| 601 | 60.14 | 83.49 | 50.89 | 64.85 | 259.37 |
| 602 | 16.86 | 15.14 | 2.71 | 11.86 | 46.57 |
| 603 | 1.14 | 1.86 | 1 | 1 | 5 |
| 604 | 22.57 | 22.71 | 23.14 | 22.57 | 90.99 |
| 605 | 49 | 48.14 | 38.85 | 44.14 | 180.13 |
| 606 | 71.5 | 75.5 | 55.71 | 65 | 267.71 |
| 608 | 0 | 0 | 0 | 0.29 | 0.29 |
| 609 | 28.93 | 23.46 | 26.32 | 10.21 | 88.92 |
| 610 | 13 | 17.71 | 17.14 | 16.14 | 63.99 |
| 611 | 6.43 | 12.57 | 8 | 9 | 36 |
| 612 | 6.14 | 4.86 | 4.86 | 6.14 | 22 |
| 613 | 1.86 | 0.57 | 1.29 | 0.71 | 4.43 |
| 614 | 11.64 | 10.36 | 2 | 19.43 | 43.43 |
| 615 | 0 | 1.43 | 2.86 | 2.86 | 7.15 |
| 616 | 3.29 | 7.43 | 6.14 | 5.29 | 22.15 |
| 617 | 5.14 | 3.86 | 5.57 | 6.43 | 21 |
| 618 | 6.29 | 0.29 | 4 | 3.86 | 14.44 |
| 619 | 0.29 | 0.14 | 0.29 | 0.29 | 1.01 |
| 620 | 4.71 | 5.86 | 7.21 | 7.57 | 25.35 |
| 622 | 6.14 | 6 | 5.86 | 3.86 | 21.86 |
| 623 | 0.57 | 0 | 1.57 | 1.43 | 3.57 |
| 624 | 0.29 | 0.43 | 0.14 | 0.29 | 1.15 |
| 625 | 7.71 | 5.71 | 6.29 | 5.57 | 25.28 |
| 626 | 0.29 | 0 | 0.43 | 0.29 | 1.01 |
| 627 | 2.57 | 1.57 | 5.71 | 2.86 | 12.71 |
| 628 | 5.36 | 2.5 | 5.43 | 9.71 | 23 |
| 629 | 0.29 | 0.57 | 1 | 0.14 | 2 |
| TOTAL | 365.58 | 375.87 | 307.55 | 353.5 | 1,402.50 |

STATE: INDIANA

| ZIP CODE | QUARTER 1 | QUARTER 2 | QUARTER 3 | QUARTER 4 | TOTAL GRAMS |
|---|---|---|---|---|---|
| 460 | 28.78 | 21.46 | 20.5 | 30.36 | 101.1 |
| 461 | 8.43 | 14.5 | 7.29 | 8.93 | 39.15 |
| 462 | 42.18 | 47.85 | 44.57 | 57.1 | 191.7 |
| 463 | 18.86 | 21.71 | 16.14 | 27.14 | 83.85 |
| 464 | 4.79 | 4.57 | 3.86 | 3 | 16.22 |
| 465 | 9.18 | 8.04 | 5.46 | 8.75 | 31.43 |
| 466 | 15.32 | 26 | 7 | 17.21 | 65.53 |
| 467 | 3.71 | 3.14 | 2.71 | 3.57 | 13.13 |
| 468 | 1.57 | 1.14 | 3.86 | 2.43 | 9 |
| 469 | 14 | 15.86 | 9.57 | 10.86 | 50.29 |
| 471 | 12.71 | 7.82 | 9.29 | 12.57 | 42.39 |
| 472 | 11.43 | 10.43 | 8.71 | 7.29 | 37.86 |

| | | | | | |
|---|---|---|---|---|---|
| 473 | 9.29 | 9.86 | 9.29 | 12 | 40.44 |
| 474 | 0.14 | 0 | 0.29 | 0.29 | 0.72 |
| 475 | 6.46 | 6.89 | 1.57 | 1.57 | 16.49 |
| 476 | 1 | 1.43 | 0.86 | 1 | 4.29 |
| 477 | 6.75 | 2.57 | 2.43 | 2 | 13.75 |
| 478 | 2.14 | 1 | 1.43 | 1 | 5.57 |
| 479 | 8.86 | 20.64 | 14.57 | 13.86 | 57.93 |
| TOTAL | 205.6 | 224.91 | 169.4 | 220.93 | 820.84 |

STATE: IOWA

| ZIP CODE | QUARTER 1 | QUARTER 2 | QUARTER 3 | QUARTER 4 | TOTAL GRAMS |
|---|---|---|---|---|---|
| 500 | 0.71 | 0.43 | 0.43 | 0.71 | 2.28 |
| 501 | 0.14 | 0 | 1 | 0 | 1.14 |
| 502 | 2.43 | 3.57 | 2 | 1.86 | 9.86 |

DRUG CODE:9041LDRUG NAME:COCAINE
STATE:IOWA

| ZIP CODE | QUARTER 1 | QUARTER 2 | QUARTER 3 | QUARTER 4 | TOTAL GRAMS |
|---|---|---|---|---|---|
| 503 | 10.86 | 14.14 | 12 | 15.86 | 52.86 |
| 504 | 0 | 0.43 | 0.14 | 0.14 | 0.71 |
| 505 | 2.86 | 2.86 | 2.29 | 1.57 | 9.58 |
| 506 | 0.57 | 1.57 | 1.71 | 1.43 | 5.28 |
| 507 | 23.86 | 25.43 | 26 | 29.86 | 105.15 |
| 508 | 0.14 | 0.29 | 0.14 | 0.57 | 1.14 |
| 510 | 1.14 | 0.43 | 0.14 | 0.57 | 2.28 |
| 511 | 1.43 | 1.86 | 5.43 | 5.14 | 13.86 |
| 512 | 0 | 0 | 0.29 | 0 | 0.29 |
| 513 | 0.14 | 0.14 | 0.14 | 0.29 | 0.71 |
| 514 | 0.29 | 0.43 | 0 | 0.14 | 0.86 |
| 515 | 0.86 | 0.57 | 1 | 1.71 | 4.14 |
| 520 | 6.29 | 2.57 | 1.14 | 3 | 13 |
| 521 | 2 | 1.43 | 0.43 | 0.29 | 4.15 |
| 522 | 10.86 | 12.86 | 11 | 12 | 46.72 |
| 523 | 0 | 0 | 0.29 | 0 | 0.29 |
| 524 | 8.43 | 12.43 | 8.29 | 9.14 | 38.29 |
| 525 | 6.86 | 3.14 | 4 | 8.29 | 22.29 |
| 526 | 2 | 1.71 | 0.29 | 1.86 | 5.86 |
| 527 | 4.14 | 7.43 | 3 | 5.29 | 19.86 |
| 528 | 7.21 | 15.07 | 10.79 | 10 | 43.07 |
| TOTAL | 93.22 | 108.79 | 91.94 | 109.72 | 403.67 |

STATE: KANSAS

| ZIP CODE | QUARTER 1 | QUARTER 2 | QUARTER 3 | QUARTER 4 | TOTAL GRAMS |
|---|---|---|---|---|---|
| 660 | 8.57 | 12 | 8 | 8.14 | 36.71 |
| 661 | 1.86 | 1.71 | 11.64 | 2 | 17.21 |
| 662 | 35.78 | 50.03 | 31.21 | 57.32 | 174.34 |
| 664 | 0.29 | 0 | 0.29 | 0 | 0.58 |
| 665 | 5.43 | 7.29 | 6.57 | 6.14 | 25.43 |
| 666 | 18.78 | 16.71 | 12.93 | 7.5 | 55.92 |
| 667 | 1.29 | 2.57 | 1.86 | 2.43 | 8.15 |
| 669 | 0 | 0.29 | 0 | 0 | 0.29 |
| 670 | 1.14 | 0.57 | 0.71 | 0.71 | 3.13 |
| 671 | 0 | 0.14 | 0.57 | 0 | 0.71 |
| 672 | 8 | 9 | 4.86 | 7.86 | 29.72 |
| 673 | 0.29 | 0.29 | 0.43 | 0 | 1.01 |
| 674 | 1.14 | 0.43 | 0.57 | 1.14 | 3.28 |
| 675 | 0 | 3 | 0 | 0 | 3 |
| 676 | 0.29 | 0.14 | 1.43 | 0 | 1.86 |
| 677 | 0.57 | 0.29 | 0.29 | 0.14 | 1.29 |
| 678 | 0 | 0.29 | 0.71 | 0.57 | 1.57 |

| | | | | | |
|---|---|---|---|---|---|
| 679 | 0.29 | 1.14 | 0.29 | 0.71 | 2.43 |
| TOTAL | 83.72 | 105.89 | 82.36 | 94.66 | 366.63 |

STATE: KENTUCKY

| ZIP CODE | QUARTER 1 | QUARTER 2 | QUARTER 3 | QUARTER 4 | TOTAL GRAMS |
|---|---|---|---|---|---|
| 400 | 1.86 | 2.71 | 2 | 2.14 | 8.71 |
| 402 | 38.43 | 28.14 | 29.57 | 32.86 | 129 |
| 403 | 4.75 | 4.71 | 2.71 | 1.86 | 14.03 |
| 404 | 0.71 | 1.43 | 0 | 1.14 | 3.28 |
| 405 | 11.61 | 32.18 | 6.14 | 10.43 | 60.36 |
| 406 | 0.57 | 0 | 0.57 | 1.14 | 2.28 |
| 407 | 2.29 | 1.86 | 0.71 | 1 | 5.86 |
| 410 | 12.5 | 9.61 | 9.89 | 10.86 | 42.86 |
| 411 | 9.29 | 15.43 | 11.71 | 9 | 45.43 |
| 412 | 0.43 | 0 | 0 | 0.57 | 1 |
| 415 | 0.29 | 0 | 1.29 | 0.29 | 1.87 |
| 416 | 3 | 2.71 | 2.29 | 2.14 | 10.14 |
| 420 | 19.53 | 5.71 | 4.14 | 16.93 | 46.31 |
| 421 | 4.71 | 4.71 | 6.71 | 7.71 | 23.84 |
| 422 | 0.14 | 0.14 | 0.43 | 0 | 0.71 |
| 423 | 2.86 | 3.86 | 2.86 | 4.86 | 14.44 |
| 424 | 1.29 | 0.71 | 1 | 1 | 4 |
| 426 | 0.14 | 0 | 0 | 0.43 | 0.57 |
| 427 | 5.14 | 8.29 | 3.57 | 6.43 | 23.43 |
| TOTAL | 119.54 | 122.2 | 85.59 | 110.79 | 438.12 |

STATE: LOUISIANA

DRUG CODE:9041LDRUG NAME:COCAINE
STATE:LOUISIANA

| ZIP CODE | QUARTER 1 | QUARTER 2 | QUARTER 3 | QUARTER 4 | TOTAL GRAMS |
|---|---|---|---|---|---|
| 700 | 1 | 2.21 | 1.57 | 1.14 | 5.92 |
| 701 | 19.71 | 17 | 16.14 | 17.57 | 70.42 |
| 703 | 3.14 | 3.29 | 4.71 | 3.86 | 15 |
| 704 | 1.57 | 0.71 | 0.57 | 1.86 | 4.71 |
| 705 | 7.29 | 20.28 | 16 | 28.14 | 71.71 |
| 706 | 9.71 | 7.86 | 4.14 | 8.86 | 30.57 |
| 707 | 0.14 | 0 | 0.14 | 0.43 | 0.71 |
| 708 | 1.14 | 1 | 2.29 | 4.43 | 8.86 |
| 710 | 0 | 0 | 0.57 | 0.14 | 0.71 |
| 711 | 41.28 | 31.43 | 35.57 | 34.57 | 142.85 |
| 712 | 0.86 | 0.43 | 0.71 | 2.57 | 4.57 |
| 713 | 5.71 | 4.43 | 3.29 | 5.86 | 19.29 |
| 714 | 2 | 3.43 | 3.86 | 1.29 | 10.58 |
| TOTAL | 93.55 | 92.07 | 89.56 | 110.72 | 385.9 |

STATE: MAINE

| ZIP CODE | QUARTER 1 | QUARTER 2 | QUARTER 3 | QUARTER 4 | TOTAL GRAMS |
|---|---|---|---|---|---|
| 39 | 0 | 1 | 0.43 | 0.43 | 1.86 |
| 40 | 0 | 0.86 | 0.43 | 0.71 | 2 |
| 41 | 8 | 9.14 | 6.14 | 3.86 | 27.14 |
| 42 | 1.64 | 0.71 | 0.29 | 0.43 | 3.07 |
| 43 | 2.43 | 2.86 | 3 | 3.86 | 12.15 |
| 44 | 7 | 5.71 | 10.14 | 8.57 | 31.42 |
| 45 | 0.14 | 0.14 | 0.43 | 0.43 | 1.14 |
| 46 | 1.43 | 0.57 | 1.14 | 1.43 | 4.57 |
| 47 | 0 | 1.57 | 2.57 | 1.71 | 5.85 |
| 48 | 3.71 | 3.43 | 3.86 | 3.71 | 14.71 |
| 49 | 1.29 | 0.57 | 1.29 | 1.43 | 4.58 |

| | | | | | |
|---|---|---|---|---|---|
| TOTAL | 25.64 | 26.56 | 29.72 | 26.57 | 108.49 |

STATE: MARYLAND

| ZIP CODE | QUARTER 1 | QUARTER 2 | QUARTER 3 | QUARTER 4 | TOTAL GRAMS |
|---|---|---|---|---|---|
| 206 | 4 | 1.14 | 3.71 | 0 | 8.85 |
| 207 | 5.57 | 4.86 | 6.57 | 6.43 | 23.43 |
| 208 | 19.57 | 19.14 | 19.86 | 13.64 | 72.21 |
| 209 | 5.86 | 10 | 7 | 5.57 | 28.43 |
| 210 | 7.86 | 10.14 | 6.86 | 8.71 | 33.57 |
| 211 | 13.14 | 8.86 | 7.14 | 7.57 | 36.71 |
| 212 | 42.93 | 48.64 | 43.28 | 37.43 | 172.28 |
| 214 | 37.96 | 31.93 | 22.43 | 29.07 | 121.39 |
| 215 | 3.57 | 2.29 | 1.14 | 0.71 | 7.71 |
| 216 | 1.43 | 2.14 | 1.71 | 3.14 | 8.42 |
| 217 | 5 | 6.57 | 3.43 | 6.43 | 21.43 |
| 218 | 1.29 | 3.29 | 1.86 | 2.43 | 8.87 |
| 219 | 4.29 | 4.29 | 4 | 3.57 | 16.15 |
| TOTAL | 152.47 | 153.29 | 128.99 | 124.7 | 559.45 |

STATE: MASSACHUSETTS

| ZIP CODE | QUARTER 1 | QUARTER 2 | QUARTER 3 | QUARTER 4 | TOTAL GRAMS |
|---|---|---|---|---|---|
| 10 | 8.86 | 7.29 | 6 | 10.43 | 32.58 |
| 11 | 22.14 | 15.86 | 18.86 | 13.57 | 70.43 |
| 12 | 8.29 | 5.57 | 5.71 | 5 | 24.57 |
| 13 | 1.86 | 0.71 | 1.14 | 1.29 | 5 |
| 14 | 12.28 | 11 | 8 | 9.29 | 40.57 |
| 15 | 3.29 | 4.71 | 3.57 | 6 | 17.57 |
| 16 | 22.57 | 24.43 | 25.28 | 23.86 | 96.14 |
| 17 | 8.29 | 6 | 7.86 | 9.29 | 31.44 |
| 18 | 17.57 | 18.57 | 14.71 | 17.28 | 68.13 |
| 19 | 9 | 8 | 9.43 | 8.86 | 35.29 |
| 20 | 0.14 | 0.29 | 0 | 0.14 | 0.57 |
| 21 | 97.64 | 99.99 | 90.14 | 79.49 | 367.26 |
| 22 | 16.43 | 15.57 | 8.43 | 8.86 | 49.29 |
| 23 | 3.71 | 3.43 | 1 | 4.29 | 12.43 |
| 24 | 5.43 | 5.71 | 3 | 1.71 | 15.85 |
| 25 | 3.57 | 0.57 | 4.43 | 4.43 | 13 |
| 26 | 5.14 | 5.71 | 3.71 | 2.29 | 16.85 |
| 27 | 3.14 | 3.57 | 2.71 | 1.86 | 11.28 |
| TOTAL | 249.35 | 236.98 | 213.98 | 207.94 | 908.25 |

STATE: MICHIGAN

DRUG CODE:9041LDRUG NAME:COCAINE
STATE:MICHIGAN

| ZIP CODE | QUARTER 1 | QUARTER 2 | QUARTER 3 | QUARTER 4 | TOTAL GRAMS |
|---|---|---|---|---|---|
| 480 | 92.46 | 65.64 | 58.28 | 90.67 | 307.05 |
| 481 | 25.57 | 44.25 | 31.75 | 40.43 | 142 |
| 482 | 14.14 | 18.71 | 13.14 | 18 | 63.99 |
| 483 | 33.14 | 30.14 | 23.28 | 29 | 115.56 |
| 484 | 7.29 | 5.29 | 4 | 6.14 | 22.72 |
| 485 | 10.57 | 13 | 11.57 | 11.28 | 46.42 |
| 486 | 4.43 | 4.86 | 6.86 | 7.86 | 24.01 |
| 487 | 7.14 | 5.71 | 4.29 | 4.29 | 21.43 |
| 488 | 12.57 | 9.14 | 11 | 13.14 | 45.85 |
| 489 | 9.71 | 12 | 4.14 | 7.71 | 33.56 |
| 490 | 40.71 | 55.71 | 62.75 | 42.14 | 201.31 |
| 491 | 34.1 | 6.75 | 23.46 | 6.61 | 70.92 |
| 492 | 2 | 3.71 | 5.71 | 1.71 | 13.13 |
| 493 | 0.29 | 0 | 0.57 | 0.29 | 1.15 |

| | | | | | |
|---|---|---|---|---|---|
| 494 | 48.57 | 63.14 | 43 | 64.14 | 218.85 |
| 495 | 22 | 32.57 | 21.86 | 35.71 | 112.14 |
| 496 | 13.14 | 15.71 | 11.28 | 14.14 | 54.27 |
| 497 | 25.96 | 14.71 | 8.71 | 14.86 | 64.24 |
| 498 | 4.43 | 5.86 | 3.29 | 6.86 | 20.44 |
| 499 | 1.14 | 1.71 | 0.43 | 0.86 | 4.14 |
| TOTAL | 409.36 | 408.61 | 349.37 | 415.84 | 1,583.18 |

STATE: MINNESOTA

| ZIP CODE | QUARTER 1 | QUARTER 2 | QUARTER 3 | QUARTER 4 | TOTAL GRAMS |
|---|---|---|---|---|---|
| 550 | 15.28 | 13.86 | 10.29 | 13.57 | 53 |
| 551 | 20.14 | 13.14 | 17.36 | 12.86 | 63.5 |
| 553 | 23.14 | 21.14 | 20.71 | 30.86 | 95.85 |
| 554 | 86.21 | 88.49 | 69.39 | 78.99 | 323.08 |
| 557 | 6 | 7.57 | 3.71 | 6.14 | 23.42 |
| 558 | 23.28 | 17.43 | 21.28 | 23.14 | 85.13 |
| 559 | 44.32 | 47.78 | 48.18 | 31 | 171.28 |
| 560 | 8.14 | 5.43 | 7.57 | 6.14 | 27.28 |
| 561 | 0 | 1 | 0.29 | 0 | 1.29 |
| 562 | 2.86 | 4 | 4.71 | 4.86 | 16.43 |
| 563 | 1.57 | 0.29 | 0.43 | 2.43 | 4.72 |
| 564 | 2.14 | 0.57 | 3 | 1.86 | 7.57 |
| 565 | 0.43 | 0.71 | 0 | 0.71 | 1.85 |
| 566 | 1.29 | 2 | 0.71 | 2.14 | 6.14 |
| 567 | 0.43 | 1.71 | 0.57 | 0.71 | 3.42 |
| TOTAL | 235.23 | 225.12 | 208.2 | 215.41 | 883.96 |

STATE: MISSISSIPPI

| ZIP CODE | QUARTER 1 | QUARTER 2 | QUARTER 3 | QUARTER 4 | TOTAL GRAMS |
|---|---|---|---|---|---|
| 386 | 2.29 | 0 | 3.29 | 1.14 | 6.72 |
| 388 | 0.71 | 1 | 0.43 | 0.43 | 2.57 |
| 389 | 8.18 | 3.93 | 3.57 | 1.07 | 16.75 |
| 390 | 1.14 | 0.14 | 0.29 | 0.57 | 2.14 |
| 391 | 0 | 0.14 | 0 | 0.14 | 0.28 |
| 392 | 16.71 | 15.86 | 15.71 | 14.71 | 62.99 |
| 393 | 0 | 0 | 8.93 | 0.14 | 9.07 |
| 394 | 11.78 | 19.43 | 10.79 | 12.5 | 54.5 |
| 395 | 35.6 | 39.18 | 11 | 32.46 | 118.24 |
| 396 | 8.93 | 4.46 | 8.93 | 8.93 | 31.25 |
| 397 | 18.71 | 17.14 | 18.57 | 23.43 | 77.85 |
| TOTAL | 104.05 | 101.28 | 81.51 | 95.52 | 382.36 |

STATE: MISSOURI

| ZIP CODE | QUARTER 1 | QUARTER 2 | QUARTER 3 | QUARTER 4 | TOTAL GRAMS |
|---|---|---|---|---|---|
| 630 | 5 | 6.43 | 4.86 | 3.57 | 19.86 |
| 631 | 27 | 26.14 | 18.14 | 23.14 | 94.42 |
| 633 | 7.71 | 6.57 | 8.86 | 4.43 | 27.57 |
| 634 | 1.43 | 1.29 | 1.57 | 1.43 | 5.72 |
| 635 | 0.14 | 0.86 | 0.86 | 0.57 | 2.43 |
| 636 | 6.43 | 4.14 | 5.43 | 10.43 | 26.43 |
| 637 | 31.82 | 28.21 | 27.21 | 20.71 | 107.95 |
| 638 | 0.43 | 0.43 | 0.14 | 0 | 1 |
| 639 | 0.14 | 0 | 0.57 | 0.14 | 0.85 |
| 640 | 38.25 | 20.53 | 36.82 | 28.75 | 124.35 |
| 641 | 7.43 | 7.86 | 15.36 | 8.93 | 39.58 |

DRUG CODE:9041LDRUG NAME:COCAINE
STATE:MISSOURI

| ZIP CODE | QUARTER 1 | QUARTER 2 | QUARTER 3 | QUARTER 4 | TOTAL GRAMS |
|---|---|---|---|---|---|

| | | | | | |
|---|---|---|---|---|---|
| 644 | 1.71 | 1.29 | 2 | 1.29 | 6.29 |
| 645 | 9.86 | 5.86 | 5.71 | 4.43 | 25.86 |
| 647 | 0.43 | 2.29 | 0.57 | 2.29 | 5.58 |
| 648 | 14.43 | 15.28 | 20.5 | 12.71 | 62.92 |
| 650 | 0 | 0 | 0.14 | 0.14 | 0.28 |
| 651 | 1.57 | 0.86 | 1.29 | 0.86 | 4.58 |
| 652 | 14.61 | 15.61 | 14.03 | 17 | 61.25 |
| 653 | 11.28 | 7.71 | 7.29 | 11.71 | 37.99 |
| 654 | 0 | 1.71 | 0.57 | 0.14 | 2.42 |
| 655 | 0 | 0.36 | 0 | 0.36 | 0.72 |
| 656 | 0.57 | 0.57 | 1.71 | 8 | 10.85 |
| 657 | 0 | 0.43 | 0 | 0.57 | 1 |
| 658 | 8.14 | 10.43 | 11.43 | 11.43 | 41.43 |
| TOTAL | 188.38 | 164.86 | 185.06 | 173.03 | 711.33 |
| STATE: | MONTANA | | | | |
| ZIP CODE | QUARTER 1 | QUARTER 2 | QUARTER 3 | QUARTER 4 | TOTAL GRAMS |
| 590 | 0.14 | 0.43 | 1 | 0.57 | 2.14 |
| 591 | 11.14 | 7.29 | 11.14 | 10.57 | 40.14 |
| 592 | 5 | 1.43 | 3.57 | 5.14 | 15.14 |
| 593 | 0 | 0 | 0.86 | 0 | 0.86 |
| 594 | 4.71 | 3.86 | 6.86 | 6 | 21.43 |
| 596 | 5.57 | 7 | 8 | 9 | 29.57 |
| 597 | 2.14 | 2.71 | 4.43 | 3.71 | 12.99 |
| 598 | 1.86 | 2 | 1.21 | 1.14 | 6.21 |
| 599 | 19 | 23.89 | 18 | 18.71 | 79.6 |
| TOTAL | 49.56 | 48.61 | 55.07 | 54.84 | 208.08 |
| STATE: | NEBRASKA | | | | |
| ZIP CODE | QUARTER 1 | QUARTER 2 | QUARTER 3 | QUARTER 4 | TOTAL GRAMS |
| 680 | 1.86 | 0.57 | 2.71 | 3.43 | 8.57 |
| 681 | 11.43 | 12.86 | 15.43 | 12.57 | 52.29 |
| 683 | 0 | 0.43 | 0.14 | 0.14 | 0.71 |
| 684 | 0 | 0 | 0 | 0.57 | 0.57 |
| 685 | 3.43 | 2.29 | 2.43 | 5.14 | 13.29 |
| 686 | 0.29 | 0.29 | 4.61 | 0.29 | 5.48 |
| 687 | 0.86 | 1.71 | 1.86 | 2 | 6.43 |
| 688 | 7 | 4.14 | 5.86 | 3.43 | 20.43 |
| 689 | 1 | 1.43 | 2.29 | 1.71 | 6.43 |
| 690 | 1.86 | 2.14 | 0 | 0 | 4 |
| 691 | 6.71 | 5.43 | 4.86 | 4.86 | 21.86 |
| 693 | 0.86 | 2.86 | 3.29 | 3.43 | 10.44 |
| TOTAL | 35.3 | 34.15 | 43.48 | 37.57 | 150.5 |
| STATE: | NEVADA | | | | |
| ZIP CODE | QUARTER 1 | QUARTER 2 | QUARTER 3 | QUARTER 4 | TOTAL GRAMS |
| 890 | 2.43 | 2 | 1.57 | 2.57 | 8.57 |
| 891 | 10.71 | 10.75 | 10.43 | 11.71 | 43.6 |
| 894 | 0.43 | 1.14 | 1.57 | 1.71 | 4.85 |
| 895 | 18.86 | 37.25 | 46.64 | 30.78 | 133.53 |
| 897 | 0.29 | 2.71 | 2.43 | 0 | 5.43 |
| 898 | 8 | 3.14 | 6.29 | 3.71 | 21.14 |
| TOTAL | 40.72 | 56.99 | 68.93 | 50.48 | 217.12 |
| STATE: | NEW HAMPSHIRE | | | | |
| ZIP CODE | QUARTER 1 | QUARTER 2 | QUARTER 3 | QUARTER 4 | TOTAL GRAMS |
| 30 | 9.71 | 9.14 | 8.75 | 10.57 | 38.17 |
| 31 | 11.57 | 6.71 | 14.61 | 13.43 | 46.32 |
| 32 | 2.57 | 3.29 | 1.71 | 1.71 | 9.28 |
| 33 | 0.86 | 0.14 | 0.71 | 0.29 | 2 |
| 34 | 0.29 | 1.14 | 0.29 | 0.14 | 1.86 |

| | | | | | |
|---|---|---|---|---|---|
| 35 | 1.71 | 1 | 0.71 | 1.14 | 4.56 |
| 37 | 7.86 | 5.86 | 11 | 8.86 | 33.58 |
| 38 | 23.86 | 20.57 | 14.28 | 16 | 74.71 |
| TOTAL | 58.43 | 47.85 | 52.06 | 52.14 | 210.48 |

STATE: NEW JERSEY

| ZIP CODE | QUARTER 1 | QUARTER 2 | QUARTER 3 | QUARTER 4 | TOTAL GRAMS |
|---|---|---|---|---|---|
| 70 | 25.36 | 22.86 | 22.43 | 22.86 | 93.51 |
| 71 | 10 | 13.86 | 9.29 | 9.71 | 42.86 |
| 72 | 0.57 | 1.14 | 1.29 | 0.86 | 3.86 |

DRUG CODE:9041LDRUG NAME:COCAINE
STATE:NEW JERSEY

| ZIP CODE | QUARTER 1 | QUARTER 2 | QUARTER 3 | QUARTER 4 | TOTAL GRAMS |
|---|---|---|---|---|---|
| 73 | 0.86 | 2 | 0.57 | 0.86 | 4.29 |
| 74 | 14.71 | 27.86 | 39.85 | 33.28 | 115.7 |
| 75 | 7.57 | 7.14 | 0.86 | 1.29 | 16.86 |
| 76 | 10 | 14 | 14.57 | 13.43 | 52 |
| 77 | 9.71 | 7.71 | 12.28 | 10 | 39.7 |
| 78 | 29.71 | 13.57 | 7 | 6.71 | 56.99 |
| 79 | 1.71 | 1.43 | 2.29 | 1.86 | 7.29 |
| 80 | 21.71 | 19.86 | 14.71 | 21 | 77.28 |
| 81 | 0 | 1.43 | 0.71 | 2.14 | 4.28 |
| 82 | 7.46 | 7.75 | 8.75 | 12.93 | 36.89 |
| 83 | 0.71 | 1 | 0.57 | 1.71 | 3.99 |
| 84 | 4 | 3.14 | 3.71 | 2.86 | 13.71 |
| 85 | 4 | 3.71 | 3.71 | 3.86 | 15.28 |
| 86 | 2.86 | 1.86 | 1.71 | 3.14 | 9.57 |
| 87 | 11 | 5.71 | 5.29 | 6 | 28 |
| 88 | 9.36 | 9.71 | 9.71 | 8.71 | 37.49 |
| 89 | 8.29 | 9.29 | 10.29 | 9.86 | 37.73 |
| TOTAL | 179.59 | 175.03 | 169.59 | 173.07 | 697.28 |

STATE: NEW MEXICO

| ZIP CODE | QUARTER 1 | QUARTER 2 | QUARTER 3 | QUARTER 4 | TOTAL GRAMS |
|---|---|---|---|---|---|
| 871 | 74.57 | 69.17 | 61.6 | 61.75 | 267.09 |
| 873 | 0.43 | 0.86 | 1.57 | 0 | 2.86 |
| 874 | 0.29 | 1 | 2.57 | 2 | 5.86 |
| 875 | 2.43 | 1 | 2.14 | 3 | 8.57 |
| 880 | 9.14 | 12.14 | 1.43 | 3.14 | 25.85 |
| 882 | 1.71 | 0 | 0.29 | 0.43 | 2.43 |
| 883 | 0 | 0.14 | 0.14 | 0 | 0.28 |
| TOTAL | 88.57 | 84.31 | 69.74 | 70.32 | 312.94 |

STATE: NEW YORK

| ZIP CODE | QUARTER 1 | QUARTER 2 | QUARTER 3 | QUARTER 4 | TOTAL GRAMS |
|---|---|---|---|---|---|
| 100 | 141.49 | 104.85 | 129.17 | 119.81 | 495.32 |
| 101 | 0.43 | 0.86 | 0.86 | 0.43 | 2.58 |
| 103 | 0.86 | 0.86 | 0.43 | 1.29 | 3.44 |
| 104 | 3 | 7.71 | 6.86 | 6 | 23.57 |
| 105 | 3.14 | 4.14 | 4.57 | 5.14 | 16.99 |
| 106 | 1 | 2.57 | 0.86 | 0.71 | 5.14 |
| 107 | 3.43 | 1.43 | 0.71 | 3.29 | 8.86 |
| 108 | 0.86 | 0.71 | 0 | 0.29 | 1.86 |
| 109 | 4.14 | 7.43 | 5 | 5.57 | 22.14 |
| 110 | 9.86 | 17.71 | 15.14 | 9.57 | 52.28 |
| 111 | 0.43 | 1.18 | 0.86 | 0 | 2.47 |
| 112 | 21.18 | 15.89 | 17.46 | 11.57 | 66.1 |
| 113 | 3.57 | 2.14 | 6.14 | 4 | 15.85 |
| 115 | 5.57 | 6.86 | 7.57 | 10.71 | 30.71 |

| | | | | | |
|---|---|---|---|---|---|
| 116 | 1.57 | 0.86 | 2.14 | 0.86 | 5.43 |
| 117 | 5.57 | 6.29 | 5.86 | 6.71 | 24.43 |
| 118 | 3.14 | 2.71 | 0.86 | 3.43 | 10.14 |
| 119 | 2.14 | 1 | 2.64 | 1.29 | 7.07 |
| 120 | 0 | 0 | 0.14 | 0 | 0.14 |
| 121 | 0.14 | 0 | 1.43 | 0.43 | 2 |
| 122 | 3.86 | 2.14 | 2.29 | 1.86 | 10.15 |
| 123 | 0.86 | 1.14 | 1.14 | 0.86 | 4 |
| 125 | 0 | 2.93 | 2.29 | 0.29 | 5.51 |
| 126 | 6.93 | 8.14 | 7.14 | 7.43 | 29.64 |
| 127 | 1.43 | 0 | 1.43 | 1.43 | 4.29 |
| 128 | 14.21 | 3.43 | 10.57 | 9 | 37.21 |
| 129 | 1.57 | 0.71 | 0.14 | 2.29 | 4.71 |
| 130 | 4.71 | 8.14 | 2.36 | 0.14 | 15.35 |
| 131 | 3.71 | 2.57 | 2.43 | 3.43 | 12.14 |
| 132 | 12.5 | 8.71 | 14.71 | 9.5 | 45.42 |
| 133 | 0 | 0.71 | 0.71 | 0 | 1.42 |
| 134 | 3.71 | 3.71 | 2.14 | 3.43 | 12.99 |
| 135 | 2.43 | 0.86 | 0 | 2.14 | 5.43 |
| 136 | 2.57 | 3.71 | 3 | 3.57 | 12.85 |
| 137 | 0.86 | 0 | 0.86 | 0.43 | 2.15 |
| 138 | 0.86 | 0.29 | 1.71 | 1.14 | 4 |

DRUG CODE:9041LDRUG NAME:COCAINE
STATE:NEW YORK

| ZIP CODE | QUARTER 1 | QUARTER 2 | QUARTER 3 | QUARTER 4 | TOTAL GRAMS |
|---|---|---|---|---|---|
| 139 | 2.57 | 1.71 | 2.86 | 0.86 | 8 |
| 140 | 1.14 | 2.43 | 2.43 | 2 | 8 |
| 141 | 1.57 | 0.86 | 2.71 | 0.71 | 5.85 |
| 142 | 48.35 | 62.46 | 64.32 | 47.71 | 222.84 |
| 144 | 7.29 | 4.71 | 5.57 | 3.57 | 21.14 |
| 145 | 1 | 0.86 | 1.43 | 0.57 | 3.86 |
| 146 | 38.14 | 37.93 | 39.89 | 40.32 | 156.28 |
| 148 | 6.43 | 5.86 | 5.86 | 11.43 | 29.58 |
| 149 | 3.43 | 4.71 | 3.57 | 3 | 14.71 |
| TOTAL | 381.65 | 353.92 | 390.26 | 348.21 | 1,474.04 |

STATE: NORTH CAROLINA

| ZIP CODE | QUARTER 1 | QUARTER 2 | QUARTER 3 | QUARTER 4 | TOTAL GRAMS |
|---|---|---|---|---|---|
| 270 | 0.14 | 0.14 | 1 | 0.86 | 2.14 |
| 271 | 18.71 | 17.43 | 18.28 | 21.28 | 75.7 |
| 272 | 3.14 | 1.57 | 2.43 | 1.14 | 8.28 |
| 273 | 2.43 | 0.29 | 0.71 | 0.29 | 3.72 |
| 274 | 1 | 2.29 | 0.14 | 2.14 | 5.57 |
| 275 | 18.03 | 13.32 | 13.75 | 20.5 | 65.6 |
| 276 | 0.71 | 4.71 | 1.43 | 2.29 | 9.14 |
| 277 | 0.29 | 1.29 | 1.86 | 1.71 | 5.15 |
| 278 | 17.36 | 23.82 | 38.5 | 11.28 | 90.96 |
| 279 | 3.14 | 3.14 | 2.43 | 1.29 | 10 |
| 280 | 16 | 16.14 | 17.71 | 15.14 | 64.99 |
| 281 | 2 | 2 | 2.14 | 2.57 | 8.71 |
| 282 | 6.43 | 7.61 | 6.86 | 11.18 | 32.08 |
| 283 | 2.57 | 1.57 | 1.29 | 2.71 | 8.14 |
| 284 | 10.86 | 7.86 | 9.14 | 11.28 | 39.14 |
| 285 | 1.71 | 8.43 | 1.5 | 2.43 | 14.07 |
| 286 | 31.64 | 24.28 | 47.53 | 28.14 | 131.59 |
| 287 | 11 | 12.86 | 8.57 | 15 | 47.43 |
| 288 | 1.86 | 1.29 | 1 | 3.29 | 7.44 |
| TOTAL | 149.02 | 150.04 | 176.27 | 154.52 | 629.85 |

RETAIL DRUG DISTRIBUTION BY ZIP CODE WITHIN STATE BY GRAMS WT

STATE: NORTH DAKOTA

| ZIP CODE | QUARTER 1 | QUARTER 2 | QUARTER 3 | QUARTER 4 | TOTAL GRAMS |
|---|---|---|---|---|---|
| 581 | 0.86 | 1.71 | 1.29 | 4 | 7.86 |
| 582 | 7.86 | 5.71 | 9.57 | 10.43 | 33.57 |
| 583 | 0 | 0 | 0 | 0.29 | 0.29 |
| 584 | 1.14 | 0 | 0 | 0.43 | 1.57 |
| 585 | 25.14 | 81.89 | 6.71 | 2.29 | 116.03 |
| 586 | 1.43 | 0 | 0.71 | 1.43 | 3.57 |
| 587 | 6.29 | 4.57 | 3.57 | 6.14 | 20.57 |
| 588 | 0 | 0 | 0.14 | 0.29 | 0.43 |
| TOTAL | 42.72 | 93.88 | 21.99 | 25.3 | 183.89 |

STATE: OHIO

| ZIP CODE | QUARTER 1 | QUARTER 2 | QUARTER 3 | QUARTER 4 | TOTAL GRAMS |
|---|---|---|---|---|---|
| 430 | 2.29 | 2.14 | 1.86 | 2.86 | 9.15 |
| 431 | 1.29 | 0 | 0.71 | 0.14 | 2.14 |
| 432 | 14.14 | 9.18 | 10.29 | 16.96 | 50.57 |
| 433 | 2.86 | 5.57 | 4.43 | 4.29 | 17.15 |
| 434 | 0 | 0.14 | 0 | 0 | 0.14 |
| 435 | 6 | 6.86 | 6 | 7 | 25.86 |
| 436 | 8.32 | 3 | 3.43 | 4.57 | 19.32 |
| 437 | 1.71 | 1 | 0.86 | 2 | 5.57 |
| 438 | 0 | 0 | 0.14 | 0.57 | 0.71 |
| 439 | 6.29 | 4.71 | 2.57 | 6.57 | 20.14 |
| 440 | 20.14 | 15.86 | 13.43 | 21.57 | 71 |
| 441 | 18.5 | 21.14 | 27.03 | 24.14 | 90.81 |
| 442 | 9.57 | 15.28 | 10.14 | 16.07 | 51.06 |
| 443 | 4.14 | 4.43 | 8.57 | 4.43 | 21.57 |
| 444 | 1 | 1.86 | 1 | 0.86 | 4.72 |
| 445 | 13.43 | 15.86 | 11.14 | 15 | 55.43 |
| 446 | 3.86 | 0.71 | 2.29 | 3.86 | 10.72 |
| 447 | 1.86 | 1.71 | 1.71 | 1.14 | 6.42 |
| 448 | 6.86 | 1.71 | 9.57 | 6.14 | 24.28 |
| 449 | 0.71 | 0 | 1.14 | 0.57 | 2.42 |
| 450 | 7.86 | 12.28 | 11.14 | 11 | 42.28 |

DRUG CODE:9041LDRUG NAME:COCAINE
STATE:OHIO

| ZIP CODE | QUARTER 1 | QUARTER 2 | QUARTER 3 | QUARTER 4 | TOTAL GRAMS |
|---|---|---|---|---|---|
| 451 | 1.29 | 0.86 | 0.43 | 0.57 | 3.15 |
| 452 | 20.18 | 12.43 | 17.36 | 22.03 | 72 |
| 453 | 2.86 | 6.14 | 6.57 | 6.14 | 21.71 |
| 454 | 7.57 | 9.43 | 13.32 | 7.29 | 37.61 |
| 455 | 4.71 | 4.86 | 3.57 | 3.86 | 17 |
| 456 | 8 | 5.43 | 4.57 | 7.43 | 25.43 |
| 457 | 0.57 | 0 | 2.07 | 0 | 2.64 |
| 458 | 4 | 2.71 | 3 | 2.29 | 12 |
| TOTAL | 180.01 | 165.3 | 178.34 | 199.35 | 723 |

STATE: OKLAHOMA

| ZIP CODE | QUARTER 1 | QUARTER 2 | QUARTER 3 | QUARTER 4 | TOTAL GRAMS |
|---|---|---|---|---|---|
| 730 | 2.14 | 2.71 | 1.14 | 3.86 | 9.85 |
| 731 | 14.71 | 17.78 | 10.43 | 8.29 | 51.21 |
| 735 | 1.71 | 0.71 | 0.57 | 0.71 | 3.7 |
| 737 | 0.71 | 0.29 | 0.57 | 0 | 1.57 |
| 740 | 8.43 | 4.29 | 4.43 | 6.29 | 23.44 |
| 741 | 17.43 | 8.86 | 13.43 | 18.86 | 58.58 |
| 743 | 0.29 | 0 | 0 | 0.43 | 0.72 |
| 744 | 1.14 | 1.14 | 0.43 | 2.43 | 5.14 |

| | | | | | |
|---|---|---|---|---|---|
| TOTAL | 46.56 | 35.78 | 31 | 40.87 | 154.21 |

STATE: OREGON

| ZIP CODE | QUARTER 1 | QUARTER 2 | QUARTER 3 | QUARTER 4 | TOTAL GRAMS |
|---|---|---|---|---|---|
| 970 | 8.29 | 6.29 | 8.14 | 9.43 | 32.15 |
| 971 | 6.43 | 6.86 | 5.57 | 5.14 | 24 |
| 972 | 51 | 37.03 | 42.5 | 34.89 | 165.42 |
| 973 | 11.14 | 12.43 | 6.29 | 7.57 | 37.43 |
| 974 | 8.86 | 7.43 | 9.14 | 8 | 33.43 |
| 975 | 45.6 | 7 | 30.93 | 10.61 | 94.14 |
| 977 | 0.86 | 3.86 | 5.71 | 3.29 | 13.72 |
| 978 | 3.29 | 3.14 | 2 | 2.29 | 10.72 |
| 979 | 1.43 | 0.43 | 1.43 | 1.71 | 5 |
| TOTAL | 136.9 | 84.47 | 111.71 | 82.93 | 416.01 |

STATE: PENNSYLVANIA

| ZIP CODE | QUARTER 1 | QUARTER 2 | QUARTER 3 | QUARTER 4 | TOTAL GRAMS |
|---|---|---|---|---|---|
| 150 | 4.57 | 8.43 | 5.57 | 5.57 | 24.14 |
| 151 | 1.43 | 1.29 | 4 | 3 | 9.72 |
| 152 | 11.86 | 14.46 | 11.71 | 15.03 | 53.06 |
| 154 | 1 | 0.71 | 1.43 | 1.29 | 4.43 |
| 155 | 0.29 | 0.57 | 0 | 1.43 | 2.29 |
| 156 | 7.29 | 9.43 | 5.43 | 6.43 | 28.58 |
| 157 | 2.14 | 3.29 | 3.29 | 2.29 | 11.01 |
| 161 | 9.43 | 7.29 | 7.71 | 0.71 | 25.14 |
| 162 | 0.57 | 0.71 | 1.86 | 0.57 | 3.71 |
| 163 | 0 | 0 | 0.14 | 0 | 0.14 |
| 165 | 3 | 3 | 3.14 | 3.71 | 12.85 |
| 166 | 0 | 0 | 0.29 | 0.14 | 0.43 |
| 167 | 0.57 | 0.71 | 0.43 | 0.86 | 2.57 |
| 168 | 10.64 | 10.21 | 5.75 | 10.79 | 37.39 |
| 169 | 0.57 | 3 | 1.14 | 1.29 | 6 |
| 170 | 8.14 | 6.57 | 9.71 | 9.86 | 34.28 |
| 171 | 3 | 1.29 | 4 | 2.43 | 10.72 |
| 172 | 1.14 | 1.86 | 2.29 | 0.43 | 5.72 |
| 173 | 0.29 | 0 | 0 | 0 | 0.29 |
| 174 | 14.36 | 10 | 7.14 | 10.32 | 41.82 |
| 175 | 2.29 | 2 | 1.14 | 1.86 | 7.29 |
| 176 | 18.14 | 18.28 | 17.71 | 15.71 | 69.84 |
| 177 | 14.57 | 17.57 | 17.14 | 19.57 | 68.85 |
| 178 | 17.21 | 4.14 | 8.14 | 9 | 38.49 |
| 179 | 10.57 | 11.14 | 9.43 | 9.57 | 40.71 |
| 180 | 2.14 | 7.43 | 5.71 | 2.86 | 18.14 |
| 181 | 21.46 | 19.11 | 15.07 | 15.71 | 71.35 |
| 182 | 0 | 0.86 | 0 | 0.57 | 1.43 |
| 183 | 5.71 | 4.29 | 3.57 | 0 | 13.57 |
| 184 | 0.86 | 0 | 0 | 0.29 | 1.15 |
| 185 | 2.29 | 4.43 | 5.93 | 2 | 14.65 |
| 186 | 1.71 | 3.57 | 1.43 | 2.86 | 9.57 |

DRUG CODE:9041LDRUG NAME:COCAINE
STATE:PENNSYLVANIA

| ZIP CODE | QUARTER 1 | QUARTER 2 | QUARTER 3 | QUARTER 4 | TOTAL GRAMS |
|---|---|---|---|---|---|
| 187 | 6.43 | 8.14 | 2.86 | 6.5 | 23.93 |
| 188 | 1.43 | 0.86 | 1.14 | 3.57 | 7 |
| 189 | 17.28 | 15.86 | 14.71 | 16.14 | 63.99 |
| 190 | 70.71 | 74.64 | 73.85 | 49.32 | 268.52 |
| 191 | 103.85 | 110.21 | 103.99 | 125.74 | 443.79 |
| 193 | 23.57 | 21.71 | 22 | 26.71 | 93.99 |

| | | | | | |
|---|---|---|---|---|---|
| 194 | 45.14 | 48.78 | 31.43 | 33.85 | 159.2 |
| 196 | 0.29 | 0.43 | 3.71 | 3 | 7.43 |
| TOTAL | 445.94 | 456.27 | 413.99 | 420.98 | 1,737.18 |
| STATE: | RHODE ISLAND | | | | |
| ZIP CODE | QUARTER 1 | QUARTER 2 | QUARTER 3 | QUARTER 4 | TOTAL GRAMS |
| 28 | 21.32 | 17.71 | 11.86 | 13.43 | 64.32 |
| 29 | 6.86 | 5.29 | 8.86 | 10.93 | 31.94 |
| TOTAL | 28.18 | 23 | 20.72 | 24.36 | 96.26 |
| STATE: | SOUTH CAROLINA | | | | |
| ZIP CODE | QUARTER 1 | QUARTER 2 | QUARTER 3 | QUARTER 4 | TOTAL GRAMS |
| 290 | 1.14 | 1.71 | 1.14 | 0 | 3.99 |
| 291 | 10.93 | 16.21 | 2.86 | 3 | 33 |
| 292 | 4.54 | 2.86 | 2.86 | 5.57 | 15.83 |
| 293 | 1 | 0.71 | 0 | 0.86 | 2.57 |
| 294 | 5.86 | 3.71 | 6.57 | 7.14 | 23.28 |
| 295 | 6.71 | 4.71 | 4.57 | 6.86 | 22.85 |
| 296 | 10 | 4.71 | 11.71 | 9.57 | 35.99 |
| 297 | 2.14 | 0.43 | 0.29 | 1.14 | 4 |
| 298 | 0 | 0.29 | 0 | 0 | 0.29 |
| TOTAL | 42.32 | 35.34 | 30 | 34.14 | 141.8 |
| STATE: | SOUTH DAKOTA | | | | |
| ZIP CODE | QUARTER 1 | QUARTER 2 | QUARTER 3 | QUARTER 4 | TOTAL GRAMS |
| 570 | 5.82 | 9.14 | 11.18 | 5.54 | 31.68 |
| 571 | 9.18 | 31.25 | 15.96 | 3 | 59.39 |
| 572 | 0.57 | 0.43 | 1 | 1.57 | 3.57 |
| 573 | 0.43 | 0.71 | 0.43 | 0.14 | 1.71 |
| 574 | 0 | 0.14 | 0.57 | 0.29 | 1 |
| 575 | 0 | 0.57 | 1.14 | 0 | 1.71 |
| 577 | 8.71 | 14.71 | 11.57 | 12.43 | 47.42 |
| TOTAL | 24.71 | 56.95 | 41.85 | 22.97 | 146.48 |
| STATE: | TENNESSEE | | | | |
| ZIP CODE | QUARTER 1 | QUARTER 2 | QUARTER 3 | QUARTER 4 | TOTAL GRAMS |
| 370 | 28.14 | 28.14 | 23.93 | 23.86 | 104.07 |
| 371 | 26.57 | 16.86 | 19.43 | 14.71 | 77.57 |
| 372 | 24.75 | 24.14 | 25.5 | 25.86 | 100.25 |
| 373 | 3.71 | 9.57 | 9.14 | 6.57 | 28.99 |
| 374 | 4.71 | 7.29 | 6.89 | 7.71 | 26.6 |
| 376 | 16.89 | 20.61 | 17 | 17 | 71.5 |
| 377 | 2.43 | 2.57 | 3.57 | 7.14 | 15.71 |
| 378 | 7.29 | 6 | 6.43 | 4.71 | 24.43 |
| 379 | 34.85 | 43 | 35.71 | 39.28 | 152.84 |
| 380 | 0.86 | 0.71 | 2 | 1 | 4.57 |
| 381 | 24.57 | 25.46 | 25.46 | 28.93 | 104.42 |
| 382 | 0.43 | 0.57 | 0 | 0.29 | 1.29 |
| 383 | 9.43 | 8 | 7.14 | 11.43 | 36 |
| 384 | 1.57 | 5.29 | 1.14 | 2 | 10 |
| 385 | 1.29 | 0 | 1.57 | 2 | 4.86 |
| TOTAL | 187.49 | 198.21 | 184.91 | 192.49 | 763.1 |
| STATE: | TEXAS | | | | |
| ZIP CODE | QUARTER 1 | QUARTER 2 | QUARTER 3 | QUARTER 4 | TOTAL GRAMS |
| 750 | 13.5 | 16.14 | 11.71 | 21.28 | 62.63 |
| 751 | 1.43 | 0.14 | 1.43 | 2.14 | 5.14 |
| 752 | 19.86 | 16.86 | 17 | 18.86 | 72.58 |
| 753 | 1.86 | 1.29 | 2.29 | 3 | 8.44 |
| 754 | 1.14 | 0 | 0 | 1 | 2.14 |
| 756 | 2.29 | 1.14 | 2 | 4 | 9.43 |

| | | | | | |
|---|---|---|---|---|---|
| 757 | 1.29 | 0.86 | 2.29 | 1.71 | 6.15 |
| 758 | 0.29 | 0.14 | 0.29 | 0 | 0.72 |
| 759 | 3.57 | 4 | 2.71 | 3.86 | 14.14 |
| 760 | 24.28 | 33.14 | 28.21 | 35.71 | 121.34 |

DRUG CODE:9041LDRUG NAME:COCAINE
STATE:TEXAS

| ZIP CODE | QUARTER 1 | QUARTER 2 | QUARTER 3 | QUARTER 4 | TOTAL GRAMS |
|---|---|---|---|---|---|
| 761 | 34.78 | 51.57 | 43.5 | 59.14 | 188.99 |
| 762 | 17.53 | 18.96 | 3.57 | 10.46 | 50.52 |
| 763 | 0.29 | 0.14 | 0.29 | 0 | 0.72 |
| 764 | 0.29 | 0 | 0 | 0.14 | 0.43 |
| 765 | 2.71 | 2.43 | 2.57 | 2.14 | 9.85 |
| 767 | 0 | 0 | 0 | 0.14 | 0.14 |
| 769 | 7.71 | 5.43 | 7.14 | 7.43 | 27.71 |
| 770 | 55.28 | 57.28 | 60.21 | 61.57 | 234.34 |
| 773 | 16.57 | 15.43 | 11.28 | 19.14 | 62.42 |
| 774 | 9 | 3.29 | 3.43 | 8 | 23.72 |
| 775 | 13.86 | 12.14 | 11.14 | 12.57 | 49.71 |
| 776 | 7.43 | 5.57 | 4 | 7.14 | 24.14 |
| 777 | 7.04 | 0 | 0.86 | 5.46 | 13.36 |
| 779 | 4.43 | 3.86 | 3.14 | 6.43 | 17.86 |
| 780 | 1.14 | 1.43 | 1 | 1.86 | 5.43 |
| 781 | 1.29 | 0 | 0.71 | 3.14 | 5.14 |
| 782 | 14.57 | 24.28 | 15.71 | 22.57 | 77.13 |
| 784 | 0.71 | 1.43 | 4.57 | 3.14 | 9.85 |
| 785 | 6.57 | 4 | 7.86 | 6.86 | 25.29 |
| 786 | 15.71 | 13.71 | 17.57 | 17.43 | 64.42 |
| 787 | 43.14 | 40.43 | 40.21 | 52.64 | 176.42 |
| 788 | 0.29 | 0 | 0.29 | 0.14 | 0.72 |
| 789 | 0.14 | 0 | 0.14 | 0.14 | 0.42 |
| 790 | 0.71 | 0 | 0 | 0.29 | 1 |
| 791 | 3.71 | 3 | 3.43 | 5.71 | 15.85 |
| 794 | 0.43 | 0.14 | 1 | 0.29 | 1.86 |
| 795 | 0.57 | 0.43 | 0 | 0.43 | 1.43 |
| 796 | 1 | 0.86 | 1 | 0 | 2.86 |
| 797 | 7.29 | 8.57 | 6.86 | 9.57 | 32.29 |
| 799 | 30 | 29.14 | 31 | 31.43 | 121.57 |
| TOTAL | 373.7 | 377.23 | 350.41 | 446.96 | 1,548.30 |

STATE: UTAH

| ZIP CODE | QUARTER 1 | QUARTER 2 | QUARTER 3 | QUARTER 4 | TOTAL GRAMS |
|---|---|---|---|---|---|
| 840 | 23.86 | 28.28 | 23.11 | 34.71 | 109.96 |
| 841 | 26.28 | 25.57 | 23.86 | 31.14 | 106.85 |
| 843 | 7 | 12.71 | 7.36 | 10.86 | 37.93 |
| 844 | 45.5 | 57.71 | 37.28 | 46.35 | 186.84 |
| 845 | 0.14 | 0.57 | 0.14 | 0.43 | 1.28 |
| 846 | 4 | 3.57 | 1 | 3.29 | 11.86 |
| 847 | 2.14 | 1.86 | 1.43 | 1.43 | 6.86 |
| TOTAL | 108.92 | 130.27 | 94.18 | 128.21 | 461.58 |

STATE: VERMONT

| ZIP CODE | QUARTER 1 | QUARTER 2 | QUARTER 3 | QUARTER 4 | TOTAL GRAMS |
|---|---|---|---|---|---|
| 50 | 0.57 | 1.43 | 0.57 | 0.57 | 3.14 |
| 52 | 1.14 | 0.71 | 0 | 0.71 | 2.56 |
| 53 | 0.14 | 0.29 | 0 | 0 | 0.43 |
| 54 | 6.71 | 5.71 | 0 | 5.86 | 18.28 |
| 56 | 1.43 | 0 | 3.43 | 1.29 | 6.15 |
| 58 | 0 | 0.57 | 0 | 0 | 0.57 |

| | | | | | |
|---|---|---|---|---|---|
| TOTAL | 9.99 | 8.71 | 4 | 8.43 | 31.13 |

STATE: VIRGINIA

| ZIP CODE | QUARTER 1 | QUARTER 2 | QUARTER 3 | QUARTER 4 | TOTAL GRAMS |
|---|---|---|---|---|---|
| 201 | 13.43 | 15.71 | 13.71 | 18.57 | 61.42 |
| 220 | 27.86 | 40.14 | 29.28 | 38 | 135.28 |
| 221 | 5.86 | 12.86 | 6.57 | 13.28 | 38.57 |
| 222 | 0.57 | 2 | 0.86 | 0.57 | 4 |
| 223 | 2.71 | 4.43 | 3 | 3.71 | 13.85 |
| 224 | 5.57 | 6.86 | 5.29 | 6.29 | 24.01 |
| 225 | 0.86 | 1.71 | 0.29 | 1 | 3.86 |
| 226 | 0.86 | 1.29 | 0.86 | 1.43 | 4.44 |
| 227 | 0 | 0 | 0.57 | 0 | 0.57 |
| 228 | 0.89 | 0 | 0.57 | 0.14 | 1.6 |
| 229 | 1.86 | 2 | 1.86 | 0.86 | 6.58 |
| 230 | 0.14 | 0.14 | 0.43 | 0 | 0.71 |
| 231 | 10.71 | 7 | 6.57 | 11.86 | 36.14 |
| 232 | 27.71 | 31.14 | 17.78 | 21.43 | 98.06 |

DRUG CODE:9041LDRUG NAME:COCAINE
STATE:VIRGINIA

| ZIP CODE | QUARTER 1 | QUARTER 2 | QUARTER 3 | QUARTER 4 | TOTAL GRAMS |
|---|---|---|---|---|---|
| 233 | 2.14 | 0.71 | 1.43 | 1.43 | 5.71 |
| 234 | 16.32 | 3.43 | 6.71 | 9.86 | 36.32 |
| 235 | 10.57 | 16 | 8.14 | 8.71 | 43.42 |
| 236 | 27.43 | 19.75 | 19.71 | 20.43 | 87.32 |
| 237 | 1.29 | 1.43 | 1.71 | 1.86 | 6.29 |
| 238 | 2.14 | 0.71 | 1 | 0 | 3.85 |
| 239 | 3.29 | 2.43 | 2.43 | 2.57 | 10.72 |
| 240 | 3.86 | 2.29 | 2.86 | 2.43 | 11.44 |
| 241 | 15.11 | 5.75 | 10.5 | 10.93 | 42.29 |
| 242 | 3.86 | 1.57 | 2.71 | 2.43 | 10.57 |
| 243 | 1.43 | 0.14 | 0.57 | 1.14 | 3.28 |
| 244 | 1.14 | 1.43 | 1.57 | 0.86 | 5 |
| 245 | 1.86 | 3 | 0.71 | 2.43 | 8 |
| 246 | 0 | 0.71 | 0.29 | 0.71 | 1.71 |
| TOTAL | 189.47 | 184.63 | 147.98 | 182.93 | 705.01 |

STATE: WASHINGTON

| ZIP CODE | QUARTER 1 | QUARTER 2 | QUARTER 3 | QUARTER 4 | TOTAL GRAMS |
|---|---|---|---|---|---|
| 980 | 19.14 | 12.71 | 12.86 | 17.43 | 62.14 |
| 981 | 94.71 | 60.42 | 57.14 | 75.75 | 288.02 |
| 982 | 4.57 | 5 | 4.86 | 5 | 19.43 |
| 983 | 4.29 | 5.43 | 4.86 | 2.71 | 17.29 |
| 984 | 9.43 | 9.86 | 12.86 | 13.57 | 45.72 |
| 985 | 3.14 | 2.86 | 6.14 | 3.14 | 15.28 |
| 986 | 4.14 | 2.29 | 3.57 | 5.46 | 15.46 |
| 988 | 0.57 | 0.86 | 1.71 | 0.43 | 3.57 |
| 989 | 0.57 | 0.86 | 0.29 | 2.86 | 4.58 |
| 991 | 0.43 | 0.14 | 0.14 | 0.29 | 1 |
| 992 | 1.14 | 0.29 | 1.43 | 1.14 | 4 |
| 993 | 16.86 | 16.86 | 18.14 | 11.71 | 63.57 |
| 994 | 1.86 | 0.86 | 0.71 | 2.29 | 5.72 |
| TOTAL | 160.85 | 118.44 | 124.71 | 141.78 | 545.78 |

STATE: WEST VIRGINIA

| ZIP CODE | QUARTER 1 | QUARTER 2 | QUARTER 3 | QUARTER 4 | TOTAL GRAMS |
|---|---|---|---|---|---|
| 247 | 2 | 2 | 2.86 | 1.71 | 8.57 |
| 249 | 0.14 | 0.29 | 0 | 0 | 0.43 |
| 252 | 0.29 | 0 | 0 | 0.43 | 0.72 |

| | | | | | |
|---|---|---|---|---|---|
| 253 | 10 | 5.86 | 18.86 | 8.86 | 43.58 |
| 254 | 4.14 | 3.57 | 3.57 | 4.71 | 15.99 |
| 255 | 0.14 | 0.57 | 0 | 1.14 | 1.85 |
| 256 | 1.43 | 1.14 | 1.57 | 1.14 | 5.28 |
| 257 | 3 | 3.71 | 3.86 | 2.43 | 13 |
| 258 | 2.71 | 3.29 | 2.14 | 1.86 | 10 |
| 260 | 4.86 | 6 | 6.14 | 4.57 | 21.57 |
| 261 | 0.14 | 0.14 | 0.43 | 1.14 | 1.85 |
| 262 | 0.43 | 0 | 0 | 0.43 | 0.86 |
| 263 | 10 | 8.71 | 10 | 7.29 | 36 |
| 264 | 1 | 1.29 | 0.71 | 0.86 | 3.86 |
| 265 | 10 | 8.57 | 8 | 7 | 33.57 |
| TOTAL | 50.28 | 45.14 | 58.14 | 43.57 | 197.13 |

STATE: WISCONSIN

| ZIP CODE | QUARTER 1 | QUARTER 2 | QUARTER 3 | QUARTER 4 | TOTAL GRAMS |
|---|---|---|---|---|---|
| 530 | 10.71 | 13.86 | 15.71 | 19.14 | 59.42 |
| 531 | 15.14 | 16.86 | 17.86 | 12.86 | 62.72 |
| 532 | 24.64 | 27.21 | 23.14 | 31 | 105.99 |
| 534 | 4.71 | 6.29 | 2.86 | 3.71 | 17.57 |
| 535 | 29.71 | 26 | 24.36 | 25.28 | 105.35 |
| 537 | 22.43 | 23.43 | 20.28 | 23.86 | 90 |
| 538 | 1.36 | 1.29 | 1.07 | 1.21 | 4.93 |
| 539 | 4.29 | 5.71 | 1.14 | 4.29 | 15.43 |
| 540 | 2 | 1.29 | 1.57 | 1.57 | 6.43 |
| 541 | 0.43 | 1.29 | 0.71 | 1 | 3.43 |
| 542 | 1 | 1.43 | 0.43 | 1.71 | 4.57 |
| 543 | 5.86 | 5.57 | 5.29 | 5.29 | 22.01 |
| 544 | 2 | 2.43 | 1.64 | 2.71 | 8.78 |
| 545 | 1.29 | 3.29 | 1.86 | 1.57 | 8.01 |
| 546 | 3.29 | 8.71 | 3.14 | 6.14 | 21.28 |

DRUG CODE:9041LDRUG NAME:COCAINE
STATE:WISCONSIN

| ZIP CODE | QUARTER 1 | QUARTER 2 | QUARTER 3 | QUARTER 4 | TOTAL GRAMS |
|---|---|---|---|---|---|
| 547 | 4 | 2.57 | 1 | 1.71 | 9.28 |
| 548 | 1 | 3.43 | 2 | 3.86 | 10.29 |
| 549 | 21.57 | 20.43 | 12.14 | 19.57 | 73.71 |
| TOTAL | 155.43 | 171.09 | 136.2 | 166.48 | 629.2 |

STATE: WYOMING

| ZIP CODE | QUARTER 1 | QUARTER 2 | QUARTER 3 | QUARTER 4 | TOTAL GRAMS |
|---|---|---|---|---|---|
| 820 | 1.43 | 2 | 2.71 | 1.14 | 7.28 |
| 822 | 0.14 | 0.14 | 0.57 | 0.57 | 1.42 |
| 823 | 0.29 | 0.71 | 0 | 0.43 | 1.43 |
| 824 | 0.43 | 1.86 | 6.18 | 1.29 | 9.76 |
| 825 | 0.43 | 0.29 | 2.43 | 0.71 | 3.86 |
| 826 | 2.14 | 2 | 3.14 | 3 | 10.28 |
| 827 | 1.14 | 0.14 | 0.71 | 0.86 | 2.85 |
| 829 | 10.29 | 6.57 | 7.14 | 12.86 | 36.86 |
| 830 | 0.57 | 0 | 1.14 | 0 | 1.71 |
| 831 | 0.43 | 1.57 | 1.29 | 0.29 | 3.58 |
| TOTAL | 17.29 | 15.28 | 25.31 | 21.15 | 79.03 |

DRUG CODE: 9050 DRUG NAME: CODEINE
STATE: ALABAMA

| ZIP CODE | QUARTER 1 | QUARTER 2 | QUARTER 3 | QUARTER 4 | TOTAL GRAMS |
|---|---|---|---|---|---|
| 350 | 6,378.32 | 6,366.85 | 6,240.16 | 6,152.06 | 25,137.39 |

| | | | | | |
|---|---|---|---|---|---|
| 351 | 3,488.29 | 3,620.94 | 3,797.90 | 3,827.96 | 14,735.09 |
| 352 | 6,150.46 | 6,623.22 | 6,077.48 | 5,985.38 | 24,836.54 |
| 354 | 1,465.24 | 1,584.60 | 1,522.61 | 1,593.35 | 6,165.80 |
| 355 | 2,160.92 | 1,763.09 | 1,915.59 | 1,826.59 | 7,666.19 |
| 356 | 4,531.99 | 4,431.61 | 4,507.97 | 5,909.40 | 19,380.97 |
| 357 | 973.96 | 933.54 | 819.03 | 877.47 | 3,604 |
| 358 | 1,152.40 | 1,056.65 | 1,111.70 | 1,087.99 | 4,408.74 |
| 359 | 4,078.50 | 2,274.75 | 2,260.69 | 2,226.65 | 10,840.59 |
| 360 | 3,969.92 | 3,503.51 | 3,360.40 | 2,927.24 | 13,761.07 |
| 361 | 3,056.55 | 3,024.81 | 2,600.78 | 2,548.52 | 11,230.66 |
| 362 | 1,552.67 | 1,441.17 | 1,549.46 | 1,289.40 | 5,832.70 |
| 363 | 1,709.56 | 1,573.01 | 1,823.23 | 1,533.74 | 6,639.54 |
| 364 | 2,384.32 | 2,512.45 | 2,543.48 | 2,247.02 | 9,687.27 |
| 365 | 4,569.80 | 4,191.30 | 4,230.66 | 4,005.17 | 16,996.93 |
| 366 | 3,928.72 | 4,003.16 | 3,806.49 | 3,666.38 | 15,404.75 |
| 367 | 2,445.11 | 2,722.44 | 2,531.23 | 2,431.73 | 10,130.51 |
| 368 | 2,506.34 | 2,318.50 | 2,266.83 | 2,438.32 | 9,529.99 |
| 369 | 412.85 | 362.46 | 396.71 | 432.07 | 1,604.09 |
| TOTAL | 56,915.92 | 54,308.06 | 53,362.40 | 53,006.44 | 217,592.82 |

STATE: ALASKA

| ZIP CODE | QUARTER 1 | QUARTER 2 | QUARTER 3 | QUARTER 4 | TOTAL GRAMS |
|---|---|---|---|---|---|
| 995 | 3,380.03 | 2,951.46 | 3,332.28 | 2,635.16 | 12,298.93 |
| 996 | 1,773.46 | 1,506.98 | 1,690.54 | 1,661.30 | 6,632.28 |
| 997 | 1,816.57 | 2,003.73 | 1,703.06 | 1,887.67 | 7,411.03 |
| 998 | 599.28 | 649.74 | 575.33 | 548.44 | 2,372.79 |
| 999 | 296.34 | 247.71 | 219.16 | 336.3 | 1,099.51 |
| TOTAL | 7,865.68 | 7,359.62 | 7,520.37 | 7,068.87 | 29,814.54 |

STATE: AMERICAN SAMOA

| ZIP CODE | QUARTER 1 | QUARTER 2 | QUARTER 3 | QUARTER 4 | TOTAL GRAMS |
|---|---|---|---|---|---|
| 967 | 764.69 | 503.9 | 291.73 | 590.1 | 2,150.42 |
| TOTAL | 764.69 | 503.9 | 291.73 | 590.1 | 2,150.42 |

STATE: ARIZONA

| ZIP CODE | QUARTER 1 | QUARTER 2 | QUARTER 3 | QUARTER 4 | TOTAL GRAMS |
|---|---|---|---|---|---|
| 850 | 20,457.97 | 20,654.13 | 20,379.46 | 20,345.31 | 81,836.87 |
| 851 | 3,341.83 | 2,928.54 | 2,936.22 | 3,151.26 | 12,357.85 |
| 852 | 22,446.41 | 21,036.05 | 21,509.09 | 22,010.83 | 87,002.38 |
| 853 | 12,566.18 | 10,927.37 | 11,144.46 | 11,559.61 | 46,197.62 |
| 855 | 1,265.34 | 1,049.80 | 1,190.14 | 905.76 | 4,411.04 |
| 856 | 2,970.19 | 2,281.10 | 2,642.51 | 2,388.95 | 10,282.75 |
| 857 | 10,528.31 | 9,784.05 | 9,997.89 | 9,672.62 | 39,982.87 |
| 859 | 833.01 | 821.56 | 965.1 | 666.36 | 3,286.03 |
| 860 | 1,897.55 | 1,707.62 | 1,869.59 | 1,602.62 | 7,077.38 |
| 863 | 2,261.59 | 2,150.18 | 2,003.59 | 2,329.23 | 8,744.59 |
| 864 | 3,229.09 | 2,904.79 | 2,914.41 | 2,871.63 | 11,919.92 |
| 865 | 1,969.54 | 1,899.75 | 1,638.93 | 1,558.30 | 7,066.52 |

DRUG CODE:9050DRUG NAME:CODEINE
STATE:ARIZONA

| ZIP CODE | QUARTER 1 | QUARTER 2 | QUARTER 3 | QUARTER 4 | TOTAL GRAMS |
|---|---|---|---|---|---|
| TOTAL | 83,767.01 | 78,144.94 | 79,191.39 | 79,062.48 | 320,165.82 |

STATE: ARKANSAS

| ZIP CODE | QUARTER 1 | QUARTER 2 | QUARTER 3 | QUARTER 4 | TOTAL GRAMS |
|---|---|---|---|---|---|
| 716 | 3,968 | 4,118.48 | 4,073.23 | 3,837.68 | 15,997.39 |
| 717 | 3,565.47 | 4,045.62 | 3,921.52 | 3,812.02 | 15,344.63 |
| 718 | 6,725.28 | 6,354.61 | 6,152.24 | 5,858.22 | 25,090.35 |
| 719 | 6,453.08 | 6,468.62 | 5,925.66 | 5,863.40 | 24,710.76 |

| | | | | | |
|---|---|---|---|---|---|
| 720 | 7,722.22 | 7,891.57 | 7,875.48 | 7,367.60 | 30,856.87 |
| 721 | 7,374.95 | 7,396.24 | 7,832.77 | 7,448.13 | 30,052.09 |
| 722 | 3,230.62 | 3,281.34 | 3,074.96 | 3,492.34 | 13,079.26 |
| 723 | 8,671.70 | 8,747.92 | 9,167.49 | 9,512.27 | 36,099.38 |
| 724 | 8,952.98 | 8,609.24 | 8,612.92 | 8,285.28 | 34,460.42 |
| 725 | 4,612.02 | 4,594.04 | 4,422.54 | 4,619.58 | 18,248.18 |
| 726 | 3,599.24 | 3,380.83 | 2,822.83 | 2,642.27 | 12,445.17 |
| 727 | 4,578.55 | 4,330.33 | 4,127.55 | 4,120.41 | 17,156.84 |
| 728 | 3,112.42 | 2,907.93 | 2,750.31 | 2,913.08 | 11,683.74 |
| 729 | 5,540.41 | 6,112.57 | 6,660.32 | 5,931.91 | 24,245.21 |
| TOTAL | 78,106.94 | 78,239.34 | 77,419.82 | 75,704.19 | 309,470.29 |

STATE: CALIFORNIA

| ZIP CODE | QUARTER 1 | QUARTER 2 | QUARTER 3 | QUARTER 4 | TOTAL GRAMS |
|---|---|---|---|---|---|
| 900 | 33,329.28 | 33,587.80 | 31,534.17 | 29,846.96 | 128,298.21 |
| 902 | 42,148.39 | 27,388.17 | 26,803.69 | 26,041.26 | 122,381.51 |
| 903 | 3,691.01 | 3,967.64 | 4,125.58 | 4,305.05 | 16,089.28 |
| 904 | 1,280.66 | 1,283.80 | 1,504.69 | 1,350.51 | 5,419.66 |
| 905 | 2,439.88 | 2,565.11 | 2,715.02 | 2,530.94 | 10,250.95 |
| 906 | 8,912.87 | 8,270.94 | 8,321.70 | 7,758.51 | 33,264.02 |
| 907 | 10,589.67 | 10,506.65 | 10,613.42 | 10,405.56 | 42,115.30 |
| 908 | 8,993.21 | 9,082.69 | 8,615.30 | 7,976.90 | 34,668.10 |
| 910 | 4,318.59 | 4,573.04 | 4,021 | 3,717.72 | 16,630.35 |
| 911 | 2,418.04 | 2,667.57 | 2,788.98 | 2,431.29 | 10,305.88 |
| 912 | 2,948.78 | 2,981.90 | 2,989.93 | 3,338.48 | 12,259.09 |
| 913 | 16,244.78 | 15,886.98 | 16,166.35 | 15,368.26 | 63,666.37 |
| 914 | 6,187.53 | 6,187.16 | 6,248.73 | 5,518.42 | 24,141.84 |
| 915 | 1,702.33 | 1,697.28 | 1,802.51 | 1,622.84 | 6,824.96 |
| 916 | 1,529.25 | 1,440.17 | 1,430.16 | 1,364.92 | 5,764.50 |
| 917 | 24,526.72 | 25,586.20 | 25,327.37 | 24,240.50 | 99,680.79 |
| 918 | 1,304.10 | 1,185.72 | 1,057.09 | 1,042.69 | 4,589.60 |
| 919 | 4,931.16 | 5,054.94 | 4,684.59 | 4,658.05 | 19,328.74 |
| 920 | 16,933.50 | 18,823.35 | 18,714.02 | 17,516.27 | 71,987.14 |
| 921 | 12,591.12 | 12,218.11 | 12,698.34 | 11,558.38 | 49,065.95 |
| 922 | 13,645.84 | 12,418.77 | 11,407.59 | 10,829.32 | 48,301.52 |
| 923 | 21,262.26 | 22,610.02 | 21,508.42 | 21,762.31 | 87,143.01 |
| 924 | 3,956.39 | 4,194.55 | 4,026.36 | 4,062.75 | 16,240.05 |
| 925 | 21,033.75 | 20,849.14 | 20,128.69 | 19,676.96 | 81,688.54 |
| 926 | 16,912.40 | 18,410.13 | 16,639.48 | 12,077.09 | 64,039.10 |
| 927 | 4,315.92 | 3,959.39 | 3,937.31 | 3,710.99 | 15,923.61 |
| 928 | 14,446.60 | 15,194.34 | 13,401.78 | 14,228.76 | 57,271.48 |
| 930 | 8,309.03 | 8,256.12 | 8,163.25 | 8,008.59 | 32,736.99 |
| 931 | 2,197.38 | 1,835.21 | 2,047.76 | 2,124.65 | 8,205 |
| 932 | 14,363.33 | 13,376.66 | 13,006.92 | 12,092.19 | 52,839.10 |
| 933 | 8,980.90 | 9,759.84 | 9,354.47 | 8,731.97 | 36,827.18 |
| 934 | 7,065.15 | 6,777.15 | 6,229.84 | 6,337.71 | 26,409.85 |
| 935 | 12,048.04 | 12,361.87 | 11,098.40 | 11,393.83 | 46,902.14 |
| 936 | 10,113.56 | 9,983.93 | 9,181.51 | 9,078.25 | 38,357.25 |
| 937 | 11,469.57 | 11,432.20 | 11,116.99 | 10,638.20 | 44,656.96 |
| 939 | 4,738.62 | 4,533.01 | 4,532.73 | 4,069.42 | 17,873.78 |
| 940 | 7,598.57 | 6,774.72 | 6,947.95 | 6,318.59 | 27,639.83 |
| 941 | 10,395.37 | 9,998.80 | 9,606.21 | 9,126.22 | 39,126.60 |
| 943 | 737.71 | 834.23 | 902.97 | 743.13 | 3,218.04 |
| 944 | 1,086.64 | 1,043.18 | 971.7 | 1,034.22 | 4,135.74 |
| 945 | 40,466.45 | 41,951.61 | 39,843.98 | 40,381.77 | 162,643.81 |
| 946 | 5,217.94 | 5,141.20 | 4,690.15 | 4,572.99 | 19,622.28 |
| 947 | 1,080.66 | 1,133.42 | 1,063.19 | 1,145.76 | 4,423.03 |
| 948 | 1,841.18 | 1,688.93 | 1,859.23 | 1,565.80 | 6,955.14 |
| 949 | 4,321.18 | 4,165.06 | 4,309.87 | 4,003.56 | 16,799.67 |
| 950 | 8,536.14 | 7,177.02 | 7,608.07 | 7,442.83 | 30,764.06 |

RETAIL DRUG DISTRIBUTION BY ZIP CODE WITHIN STATE BY GRAMS WT

DRUG CODE:9050DRUG NAME:CODEINE
STATE:CALIFORNIA

| ZIP CODE | QUARTER 1 | QUARTER 2 | QUARTER 3 | QUARTER 4 | TOTAL GRAMS |
|---|---|---|---|---|---|
| 951 | 8,381.32 | 7,790.11 | 7,757.46 | 7,727.32 | 31,656.21 |
| 952 | 13,037.63 | 12,183.88 | 12,688.85 | 11,761.98 | 49,672.34 |
| 953 | 19,137.81 | 18,831.38 | 17,542.22 | 17,342.82 | 72,854.23 |
| 954 | 6,086.94 | 6,326.21 | 6,276.61 | 5,882.71 | 24,572.47 |
| 955 | 4,108.75 | 3,995.21 | 3,695.46 | 3,571.94 | 15,371.36 |
| 956 | 18,754.51 | 18,670.38 | 18,318 | 17,721.29 | 73,464.18 |
| 957 | 2,778.90 | 2,556.89 | 2,636.22 | 2,676.67 | 10,648.68 |
| 958 | 9,658.29 | 9,947.59 | 10,182.58 | 9,207.10 | 38,995.56 |
| 959 | 9,019.57 | 9,203.21 | 9,211.69 | 8,687.59 | 36,122.06 |
| 960 | 3,884.23 | 4,302.97 | 3,949.85 | 3,898.72 | 16,035.77 |
| 961 | 1,506.63 | 1,527.22 | 1,664.21 | 1,821.21 | 6,519.27 |
| TOTAL | 559,516.03 | 546,150.77 | 529,670.61 | 508,050.72 | 2,143,388.13 |

STATE: COLORADO

| ZIP CODE | QUARTER 1 | QUARTER 2 | QUARTER 3 | QUARTER 4 | TOTAL GRAMS |
|---|---|---|---|---|---|
| 800 | 4,704.71 | 4,358.61 | 4,561.42 | 4,390.01 | 18,014.75 |
| 801 | 3,454.33 | 3,047.79 | 3,208.25 | 3,302.24 | 13,012.61 |
| 802 | 5,507.81 | 5,180.13 | 5,300.14 | 4,993.03 | 20,981.11 |
| 803 | 340.12 | 408.64 | 391.41 | 379.94 | 1,520.11 |
| 804 | 758.18 | 841.8 | 760.21 | 812.44 | 3,172.63 |
| 805 | 2,622.53 | 2,363.94 | 2,377.68 | 2,537.60 | 9,901.75 |
| 806 | 1,199.98 | 1,110.68 | 983.17 | 1,014 | 4,307.83 |
| 807 | 545.47 | 408.77 | 341.07 | 449.93 | 1,745.24 |
| 808 | 644.69 | 621.85 | 707.39 | 685.7 | 2,659.63 |
| 809 | 3,072.32 | 2,901.42 | 2,728.14 | 2,812.09 | 11,513.97 |
| 810 | 2,307.83 | 2,064.75 | 1,956.12 | 1,985.54 | 8,314.24 |
| 811 | 408.17 | 441.24 | 480.88 | 450.78 | 1,781.07 |
| 812 | 867.78 | 749.42 | 744.56 | 774.03 | 3,135.79 |
| 813 | 493.22 | 367.06 | 335.47 | 303.15 | 1,498.90 |
| 814 | 740.32 | 712.73 | 648.81 | 653.49 | 2,755.35 |
| 815 | 986.55 | 1,228.74 | 1,103.70 | 1,069.04 | 4,388.03 |
| 816 | 634.88 | 380.31 | 500.12 | 450.24 | 1,965.55 |
| TOTAL | 29,288.89 | 27,187.88 | 27,128.54 | 27,063.25 | 110,668.56 |

STATE: CONNECTICUT

| ZIP CODE | QUARTER 1 | QUARTER 2 | QUARTER 3 | QUARTER 4 | TOTAL GRAMS |
|---|---|---|---|---|---|
| 60 | 6,185.50 | 5,786.28 | 5,843.45 | 5,749.61 | 23,564.84 |
| 61 | 1,782.53 | 1,772.28 | 1,772.62 | 1,753.49 | 7,080.92 |
| 62 | 1,059.35 | 851.42 | 948.12 | 872.05 | 3,730.94 |
| 63 | 1,884.07 | 1,714.66 | 1,771.11 | 1,876.04 | 7,245.88 |
| 64 | 6,543.90 | 6,290.97 | 6,241.83 | 6,187.94 | 25,264.64 |
| 65 | 2,081.73 | 2,139.93 | 1,945.78 | 1,890.79 | 8,058.23 |
| 66 | 1,779.66 | 1,854.69 | 1,689.79 | 1,673.65 | 6,997.79 |
| 67 | 3,239.79 | 3,076.90 | 2,896.01 | 2,814.79 | 12,027.49 |
| 68 | 3,721.45 | 3,778.14 | 3,554.84 | 3,354.88 | 14,409.31 |
| 69 | 764.12 | 892.23 | 665.5 | 761.38 | 3,083.23 |
| TOTAL | 29,042.10 | 28,157.50 | 27,329.05 | 26,934.62 | 111,463.27 |

STATE: DELAWARE

| ZIP CODE | QUARTER 1 | QUARTER 2 | QUARTER 3 | QUARTER 4 | TOTAL GRAMS |
|---|---|---|---|---|---|
| 197 | 4,302.79 | 4,189.16 | 4,164.97 | 3,858.77 | 16,515.69 |
| 198 | 2,409.36 | 2,872.54 | 2,668.07 | 2,370.88 | 10,320.85 |
| 199 | 4,264.59 | 4,053.18 | 4,137.46 | 4,295.43 | 16,750.66 |
| TOTAL | 10,976.74 | 11,114.88 | 10,970.50 | 10,525.08 | 43,587.20 |

STATE: DISTRICT OF COLUMBIA

| ZIP CODE | QUARTER 1 | QUARTER 2 | QUARTER 3 | QUARTER 4 | TOTAL GRAMS |
|---|---|---|---|---|---|
| 200 | 13,137.26 | 13,713.47 | 13,015.70 | 12,892.08 | 52,758.51 |

| | | | | | |
|---|---|---|---|---|---|
| 203 | 50.81 | 0 | 33.15 | 0 | 83.96 |
| 204 | 284.35 | 265.76 | 370.9 | 164.64 | 1,085.65 |
| 205 | 0 | 0 | 2.21 | 0 | 2.21 |
| TOTAL | 13,472.42 | 13,979.23 | 13,421.96 | 13,056.72 | 53,930.33 |

STATE: FLORIDA

| ZIP CODE | QUARTER 1 | QUARTER 2 | QUARTER 3 | QUARTER 4 | TOTAL GRAMS |
|---|---|---|---|---|---|
| 320 | 5,174.91 | 4,561.66 | 4,498.24 | 4,679.98 | 18,914.79 |
| 321 | 6,809.15 | 6,101.57 | 5,601.55 | 5,722.29 | 24,234.56 |
| 322 | 5,867.80 | 5,653.33 | 6,003.67 | 6,213.90 | 23,738.70 |
| 323 | 4,083.12 | 3,697.11 | 3,792.30 | 3,656.69 | 15,229.22 |
| 324 | 4,941.86 | 4,645.80 | 3,894.10 | 3,734.13 | 17,215.89 |
| 325 | 4,718.06 | 4,284.59 | 4,039.02 | 4,033.12 | 17,074.79 |

DRUG CODE:9050DRUG NAME:CODEINE
STATE:FLORIDA

| ZIP CODE | QUARTER 1 | QUARTER 2 | QUARTER 3 | QUARTER 4 | TOTAL GRAMS |
|---|---|---|---|---|---|
| 326 | 2,969.67 | 2,498.58 | 2,628.97 | 2,421.85 | 10,519.07 |
| 327 | 8,777.98 | 8,138.79 | 7,593.21 | 7,999.77 | 32,509.75 |
| 328 | 6,569.74 | 5,797.87 | 5,963.19 | 5,954.96 | 24,285.76 |
| 329 | 5,166.34 | 5,273.77 | 4,990.98 | 5,160.51 | 20,591.60 |
| 330 | 15,073.76 | 13,618.86 | 13,443.60 | 13,419.19 | 55,555.41 |
| 331 | 17,486.76 | 16,886.47 | 15,984.19 | 16,813.24 | 67,170.66 |
| 333 | 7,826.37 | 7,275.28 | 6,748.74 | 7,255.87 | 29,106.26 |
| 334 | 14,169.69 | 12,281.68 | 12,245.81 | 13,326.93 | 52,024.11 |
| 335 | 8,687.78 | 8,323.48 | 7,726.60 | 8,107.35 | 32,845.21 |
| 336 | 8,991.94 | 9,258.53 | 8,388 | 8,909.49 | 35,547.96 |
| 337 | 8,508.88 | 8,246.13 | 7,923.23 | 9,659.48 | 34,337.72 |
| 338 | 9,253.70 | 9,032.75 | 7,910.13 | 8,527.88 | 34,724.46 |
| 339 | 9,373.91 | 7,746.96 | 6,966.80 | 7,125.51 | 31,213.18 |
| 341 | 3,155.65 | 2,114.66 | 1,667.60 | 2,796.29 | 9,734.20 |
| 342 | 7,612.65 | 6,663.24 | 5,653.37 | 6,442.80 | 26,372.06 |
| 344 | 6,493.69 | 5,857.43 | 5,229.02 | 5,034.66 | 22,614.80 |
| 346 | 9,495.20 | 7,291.93 | 6,383.84 | 6,689.43 | 29,860.40 |
| 347 | 5,097.03 | 5,012.42 | 4,745.13 | 4,920.74 | 19,775.32 |
| 349 | 3,212.85 | 3,362.54 | 3,081.50 | 2,888.87 | 12,545.76 |
| TOTAL | 189,518.49 | 173,625.43 | 163,102.79 | 171,494.93 | 697,741.64 |

STATE: GEORGIA

| ZIP CODE | QUARTER 1 | QUARTER 2 | QUARTER 3 | QUARTER 4 | TOTAL GRAMS |
|---|---|---|---|---|---|
| 300 | 16,057.22 | 15,971.57 | 15,850.95 | 16,387 | 64,266.74 |
| 301 | 9,191.38 | 9,447.95 | 9,221 | 9,482.11 | 37,342.44 |
| 302 | 12,395.03 | 11,993.58 | 12,283.83 | 12,192.65 | 48,865.09 |
| 303 | 9,146.49 | 8,797.07 | 8,413.35 | 8,618.58 | 34,975.49 |
| 304 | 2,642.11 | 2,868.66 | 2,892.96 | 3,541.05 | 11,944.78 |
| 305 | 7,470.77 | 7,734.75 | 7,040.80 | 7,404.27 | 29,650.59 |
| 306 | 5,944.78 | 5,817.32 | 6,282.85 | 6,048.55 | 24,093.50 |
| 307 | 2,751.52 | 2,803.14 | 2,616.05 | 2,865.04 | 11,035.75 |
| 308 | 1,847.33 | 1,975.09 | 2,143.99 | 2,043.10 | 8,009.51 |
| 309 | 3,933.47 | 3,856.19 | 4,069.39 | 4,097.09 | 15,956.14 |
| 310 | 6,025.16 | 6,084.67 | 5,977.75 | 5,596.93 | 23,684.51 |
| 312 | 2,095.39 | 1,958.67 | 2,260.59 | 2,016.18 | 8,330.83 |
| 313 | 1,621.03 | 1,563.75 | 1,895.62 | 1,716.92 | 6,797.32 |
| 314 | 1,674.83 | 1,635.46 | 1,783.05 | 1,739.87 | 6,833.21 |
| 315 | 6,662.15 | 7,124.69 | 6,411.72 | 6,238.97 | 26,437.53 |
| 316 | 2,768.19 | 2,897.25 | 2,555.56 | 2,599.99 | 10,820.99 |
| 317 | 7,633.55 | 7,156.52 | 7,281.99 | 6,708.16 | 28,780.22 |
| 318 | 633.73 | 656.55 | 571.38 | 518.26 | 2,379.92 |
| 319 | 3,647.22 | 3,578.33 | 3,375.27 | 3,277.51 | 13,878.33 |

| | | | | | | |
|---|---|---|---|---|---|---|
| | 398 | 2,284.14 | 2,064.23 | 1,990.20 | 2,023.35 | 8,361.92 |
| TOTAL | | 106,425.49 | 105,985.44 | 104,918.30 | 105,115.58 | 422,444.81 |
| STATE: | GUAM | | | | | |
| ZIP CODE | QUARTER 1 | | QUARTER 2 | QUARTER 3 | QUARTER 4 | TOTAL GRAMS |
| | 969 | 1,384.25 | 1,306.60 | 1,732.30 | 1,007.81 | 5,430.96 |
| TOTAL | | 1,384.25 | 1,306.60 | 1,732.30 | 1,007.81 | 5,430.96 |
| STATE: | HAWAII | | | | | |
| ZIP CODE | QUARTER 1 | | QUARTER 2 | QUARTER 3 | QUARTER 4 | TOTAL GRAMS |
| | 967 | 10,105.63 | 9,547.28 | 8,991.23 | 8,540.97 | 37,185.11 |
| | 968 | 5,748.99 | 5,174.80 | 5,521.16 | 4,691.66 | 21,136.61 |
| TOTAL | | 15,854.62 | 14,722.08 | 14,512.39 | 13,232.63 | 58,321.72 |
| STATE: | IDAHO | | | | | |
| ZIP CODE | QUARTER 1 | | QUARTER 2 | QUARTER 3 | QUARTER 4 | TOTAL GRAMS |
| | 832 | 1,747.64 | 1,692.44 | 1,663.23 | 1,781.97 | 6,885.28 |
| | 833 | 1,560.85 | 1,514.10 | 1,610.19 | 1,404.38 | 6,089.52 |
| | 834 | 2,091.29 | 1,893.86 | 1,891.26 | 1,678.92 | 7,555.33 |
| | 835 | 1,170.33 | 1,207.50 | 1,276.89 | 1,164.37 | 4,819.09 |
| | 836 | 3,893.71 | 3,703.30 | 3,515.20 | 3,222 | 14,334.21 |
| | 837 | 2,086.90 | 2,324.90 | 2,166.93 | 1,927.17 | 8,505.90 |
| | 838 | 2,870.34 | 2,888.76 | 2,701.08 | 2,528.32 | 10,988.50 |
| TOTAL | | 15,421.06 | 15,224.86 | 14,824.78 | 13,707.13 | 59,177.83 |
| STATE: | ILLINOIS | | | | | |
| ZIP CODE | QUARTER 1 | | QUARTER 2 | QUARTER 3 | QUARTER 4 | TOTAL GRAMS |
| | 600 | 22,467.46 | 22,975.17 | 21,125.34 | 23,385.60 | 89,953.57 |
| | 601 | 14,881.30 | 15,212.24 | 14,743.76 | 15,313.19 | 60,150.49 |

DRUG CODE:9050DRUG NAME:CODEINE
STATE:ILLINOIS

| ZIP CODE | QUARTER 1 | | QUARTER 2 | QUARTER 3 | QUARTER 4 | TOTAL GRAMS |
|---|---|---|---|---|---|---|
| | 602 | 601.15 | 606.95 | 398.84 | 606.67 | 2,213.61 |
| | 603 | 1,186.92 | 1,241.70 | 1,081.47 | 1,163.57 | 4,673.66 |
| | 604 | 21,181.02 | 20,484.27 | 20,185.84 | 20,672.28 | 82,523.41 |
| | 605 | 10,055.32 | 10,007.52 | 9,910.83 | 10,332.34 | 40,306.01 |
| | 606 | 38,258.22 | 37,726.14 | 34,820.93 | 35,312.35 | 146,117.64 |
| | 607 | 1,524.02 | 1,569.61 | 1,502.28 | 1,693.12 | 6,289.03 |
| | 608 | 2,133.11 | 1,819.09 | 1,771.67 | 2,125.36 | 7,849.23 |
| | 609 | 2,401 | 2,737.21 | 2,623.85 | 2,746.28 | 10,508.34 |
| | 610 | 3,152.18 | 3,158.74 | 3,455.64 | 3,223.98 | 12,990.54 |
| | 611 | 4,284.52 | 4,488.35 | 5,068.93 | 5,305.71 | 19,147.51 |
| | 612 | 2,043.86 | 2,188.36 | 2,234.07 | 2,055.73 | 8,522.02 |
| | 613 | 1,609.64 | 1,834.14 | 1,747.37 | 1,715.73 | 6,906.88 |
| | 614 | 1,879.08 | 2,151.12 | 2,186.87 | 1,986.76 | 8,203.83 |
| | 615 | 2,564.90 | 2,518.94 | 2,486.35 | 2,448.66 | 10,018.85 |
| | 616 | 3,661.57 | 3,125.01 | 3,471.54 | 3,425.50 | 13,683.62 |
| | 617 | 3,288.21 | 3,692.38 | 3,298.81 | 3,308.21 | 13,587.61 |
| | 618 | 4,360.35 | 4,487.36 | 4,378.85 | 4,451.82 | 17,678.38 |
| | 619 | 1,365.44 | 1,470.06 | 1,648.59 | 1,483.32 | 5,967.41 |
| | 620 | 5,311.53 | 5,433.03 | 5,394.34 | 5,158.65 | 21,297.55 |
| | 622 | 6,798.38 | 6,891.95 | 6,545.86 | 6,475.70 | 26,711.89 |
| | 623 | 1,227.89 | 1,440.95 | 1,373.63 | 1,536.20 | 5,578.67 |
| | 624 | 1,564.81 | 1,532.51 | 1,736.75 | 1,732.42 | 6,566.49 |
| | 625 | 2,807.75 | 2,741.86 | 2,575.50 | 2,653.11 | 10,778.22 |
| | 626 | 2,301.96 | 2,442.94 | 2,279.34 | 2,362.30 | 9,386.54 |
| | 627 | 3,749.49 | 3,624.11 | 3,392.43 | 3,665.84 | 14,431.87 |
| | 628 | 2,716.73 | 2,682.85 | 3,013.81 | 2,653.73 | 11,067.12 |

| | | | | | |
|---|---|---|---|---|---|
| 629 | 4,852.05 | 5,218.96 | 4,777.14 | 4,830.77 | 19,678.92 |
| TOTAL | 174,229.86 | 175,503.52 | 169,230.63 | 173,824.90 | 692,788.91 |

STATE: INDIANA

| ZIP CODE | QUARTER 1 | QUARTER 2 | QUARTER 3 | QUARTER 4 | TOTAL GRAMS |
|---|---|---|---|---|---|
| 460 | 3,270.87 | 3,054.53 | 2,843.11 | 2,869.28 | 12,037.79 |
| 461 | 4,009.03 | 4,178.65 | 3,842.96 | 4,304.78 | 16,335.42 |
| 462 | 7,800.41 | 8,230.64 | 7,422.98 | 7,736.41 | 31,190.44 |
| 463 | 9,870.67 | 9,728.61 | 9,373.16 | 9,404.78 | 38,377.22 |
| 464 | 2,602.89 | 2,651.12 | 2,469.65 | 2,593.31 | 10,316.97 |
| 465 | 3,424.70 | 3,427.63 | 3,306.89 | 3,063.79 | 13,223.01 |
| 466 | 1,135.82 | 999.33 | 1,178.07 | 951.81 | 4,265.03 |
| 467 | 1,791.97 | 3,094.29 | 2,319.09 | 1,986.88 | 9,192.23 |
| 468 | 2,383.80 | 2,302.02 | 2,166.73 | 2,168.41 | 9,020.96 |
| 469 | 3,323.97 | 3,398.22 | 3,049.83 | 3,059.10 | 12,831.12 |
| 470 | 703.25 | 786.42 | 625.43 | 660.73 | 2,775.83 |
| 471 | 3,438.82 | 4,255.14 | 4,622.29 | 2,255.37 | 14,571.62 |
| 472 | 1,379.34 | 1,417.37 | 1,324.13 | 1,465.64 | 5,586.48 |
| 473 | 1,645.31 | 1,803.20 | 1,380.96 | 1,677.76 | 6,507.23 |
| 474 | 1,651.66 | 1,947.54 | 2,052.53 | 1,672.96 | 7,324.69 |
| 475 | 1,161 | 1,336.97 | 1,353.43 | 1,158.67 | 5,010.07 |
| 476 | 989.79 | 962.66 | 1,027.97 | 982.47 | 3,962.89 |
| 477 | 3,876.90 | 3,144.41 | 2,970.65 | 3,092.67 | 13,084.63 |
| 478 | 2,546.20 | 2,434.84 | 2,407.22 | 2,286.86 | 9,675.12 |
| 479 | 2,008.93 | 2,244.37 | 2,212.74 | 2,326.90 | 8,792.94 |
| TOTAL | 59,015.33 | 61,397.96 | 57,949.82 | 55,718.58 | 234,081.69 |

STATE: IOWA

| ZIP CODE | QUARTER 1 | QUARTER 2 | QUARTER 3 | QUARTER 4 | TOTAL GRAMS |
|---|---|---|---|---|---|
| 500 | 1,082.61 | 1,240.07 | 1,396.20 | 1,339.67 | 5,058.55 |
| 501 | 953.53 | 1,240.71 | 1,260.72 | 1,279.05 | 4,734.01 |
| 502 | 918.09 | 1,106.61 | 1,202.18 | 1,108.19 | 4,335.07 |
| 503 | 2,212.42 | 2,542.71 | 2,356.32 | 2,274.33 | 9,385.78 |
| 504 | 491.37 | 506.1 | 483.09 | 571.31 | 2,051.87 |
| 505 | 565.16 | 974.32 | 708.25 | 767.86 | 3,015.59 |
| 506 | 777.88 | 848.91 | 911.77 | 931.26 | 3,469.82 |
| 507 | 730.41 | 582.72 | 652.89 | 686.76 | 2,652.78 |
| 508 | 173.67 | 203.33 | 246.79 | 253.6 | 877.39 |
| 510 | 465.29 | 508.77 | 530.23 | 475.99 | 1,980.28 |
| 511 | 600.63 | 747.08 | 669.45 | 684.64 | 2,701.80 |
| 512 | 174.6 | 194.49 | 163.55 | 176.81 | 709.45 |
| 513 | 302.41 | 488.97 | 437.22 | 450.44 | 1,679.04 |

DRUG CODE:9050DRUG NAME:CODEINE
STATE:IOWA

| ZIP CODE | QUARTER 1 | QUARTER 2 | QUARTER 3 | QUARTER 4 | TOTAL GRAMS |
|---|---|---|---|---|---|
| 514 | 314.74 | 381.53 | 319.01 | 284.51 | 1,299.79 |
| 515 | 1,137.83 | 1,100.57 | 1,134.17 | 1,158.56 | 4,531.13 |
| 516 | 227.19 | 345.02 | 262.75 | 256.38 | 1,091.34 |
| 520 | 1,294.32 | 1,439.78 | 1,259.81 | 1,309.91 | 5,303.82 |
| 521 | 412.73 | 337.21 | 305.54 | 347.35 | 1,402.83 |
| 522 | 1,294.47 | 1,395.90 | 1,263.12 | 1,321.29 | 5,274.78 |
| 523 | 823.8 | 856.81 | 733.73 | 653.06 | 3,067.40 |
| 524 | 635.68 | 1,031.10 | 852.35 | 842.13 | 3,361.26 |
| 525 | 728.14 | 1,263.59 | 1,094.79 | 1,013.49 | 4,100.01 |
| 526 | 874.09 | 1,052.23 | 964.86 | 1,190.16 | 4,081.34 |
| 527 | 1,160.71 | 1,429.74 | 1,245.37 | 1,378.35 | 5,214.17 |
| 528 | 1,051.08 | 1,152.47 | 1,156.39 | 1,104.79 | 4,464.73 |
| TOTAL | 19,402.85 | 22,970.74 | 21,610.55 | 21,859.89 | 85,844.03 |

RETAIL DRUG DISTRIBUTION BY ZIP CODE WITHIN STATE BY GRAMS WT

STATE: KANSAS

| ZIP CODE | QUARTER 1 | QUARTER 2 | QUARTER 3 | QUARTER 4 | TOTAL GRAMS |
|---|---|---|---|---|---|
| 660 | 3,084.77 | 2,909.09 | 2,957.47 | 3,150.12 | 12,101.45 |
| 661 | 1,161.12 | 1,137.11 | 1,091.49 | 1,160.59 | 4,550.31 |
| 662 | 9,541.76 | 9,123.65 | 7,892.54 | 8,895.61 | 35,453.56 |
| 664 | 564.93 | 561.46 | 587.11 | 514.48 | 2,227.98 |
| 665 | 973.26 | 1,097.94 | 768.19 | 886.47 | 3,725.86 |
| 666 | 1,139.89 | 1,149.57 | 1,193.28 | 1,067.90 | 4,550.64 |
| 667 | 1,202.45 | 1,066.91 | 1,104.48 | 1,147.21 | 4,521.05 |
| 668 | 385.84 | 329.52 | 348.84 | 383.04 | 1,447.24 |
| 669 | 300.57 | 273.4 | 262.72 | 327.82 | 1,164.51 |
| 670 | 1,796.95 | 1,757.55 | 1,802.69 | 1,698.22 | 7,055.41 |
| 671 | 1,028.62 | 963.4 | 1,129.89 | 919.31 | 4,041.22 |
| 672 | 4,224.21 | 4,068.99 | 3,780.58 | 3,990.99 | 16,064.77 |
| 673 | 754.63 | 1,002.44 | 896.57 | 901.25 | 3,554.89 |
| 674 | 1,557.42 | 1,644.34 | 1,664.47 | 1,521.63 | 6,387.86 |
| 675 | 847.57 | 792.71 | 831.88 | 995.37 | 3,467.53 |
| 676 | 453.06 | 444.32 | 365.39 | 400.86 | 1,663.63 |
| 677 | 230.96 | 203.62 | 256.46 | 254.34 | 945.38 |
| 678 | 1,048.01 | 1,040.74 | 912.19 | 955.02 | 3,955.96 |
| 679 | 732.74 | 710.64 | 740.38 | 729.33 | 2,913.09 |
| TOTAL | 31,028.76 | 30,277.40 | 28,586.62 | 29,899.56 | 119,792.34 |

STATE: KENTUCKY

| ZIP CODE | QUARTER 1 | QUARTER 2 | QUARTER 3 | QUARTER 4 | TOTAL GRAMS |
|---|---|---|---|---|---|
| 400 | 2,004.83 | 2,090.59 | 2,123.04 | 1,760.68 | 7,979.14 |
| 401 | 1,148.55 | 1,004.30 | 1,174.27 | 1,048.15 | 4,375.27 |
| 402 | 7,209.97 | 6,896.04 | 6,490.16 | 7,106.73 | 27,702.90 |
| 403 | 2,793.98 | 2,723.31 | 2,941.93 | 2,897.86 | 11,357.08 |
| 404 | 1,460.42 | 1,373.93 | 1,351.40 | 1,374.86 | 5,560.61 |
| 405 | 1,605.79 | 1,522.15 | 1,581.56 | 1,515.67 | 6,225.17 |
| 406 | 143.84 | 250.77 | 199.36 | 168.15 | 762.12 |
| 407 | 2,066.55 | 2,276.01 | 2,024.82 | 2,192.60 | 8,559.98 |
| 408 | 939.03 | 1,012.59 | 941.5 | 895.27 | 3,788.39 |
| 409 | 3,101.97 | 3,134.01 | 2,826.72 | 2,806.83 | 11,869.53 |
| 410 | 3,985.62 | 4,219.37 | 4,005.23 | 3,845.36 | 16,055.58 |
| 411 | 1,152.19 | 1,542.05 | 1,301.07 | 1,297.29 | 5,292.60 |
| 412 | 2,210.10 | 2,361.49 | 2,163.69 | 2,045.54 | 8,780.82 |
| 413 | 1,618.90 | 1,824.44 | 1,629.95 | 1,634.37 | 6,707.66 |
| 414 | 498.3 | 548.03 | 438.11 | 484.45 | 1,968.89 |
| 415 | 2,343.64 | 2,552.67 | 2,292.98 | 2,268.59 | 9,457.88 |
| 416 | 737.29 | 1,249.15 | 861.94 | 1,068.36 | 3,916.74 |
| 417 | 719.71 | 655.08 | 656 | 685.83 | 2,716.62 |
| 418 | 490.64 | 558.12 | 654.55 | 720.49 | 2,423.80 |
| 420 | 2,295.59 | 2,184.96 | 2,447.87 | 2,261.02 | 9,189.44 |
| 421 | 2,717.24 | 3,050.56 | 2,692.20 | 2,690.49 | 11,150.49 |
| 422 | 1,331.01 | 1,650.94 | 1,548.72 | 1,617.60 | 6,148.27 |
| 423 | 2,205.36 | 2,307.50 | 2,216.12 | 2,270.45 | 8,999.43 |
| 424 | 1,927.62 | 1,809.82 | 1,849.44 | 1,997.62 | 7,584.50 |
| 425 | 898.53 | 866.36 | 842.05 | 570.21 | 3,177.15 |
| 426 | 1,529.84 | 1,600.11 | 1,697.44 | 1,447.46 | 6,274.85 |
| 427 | 3,413 | 3,641.73 | 3,573.26 | 3,005.45 | 13,633.44 |
| TOTAL | 52,549.51 | 54,906.08 | 52,525.38 | 51,677.38 | 211,658.35 |

STATE: LOUISIANA

DRUG CODE:9050DRUG NAME:CODEINE
STATE:LOUISIANA

| ZIP CODE | QUARTER 1 | QUARTER 2 | QUARTER 3 | QUARTER 4 | TOTAL GRAMS |
|---|---|---|---|---|---|
| 700 | 5,478.69 | 5,188.99 | 5,213.42 | 5,077.20 | 20,958.30 |

| | | | | | |
|---|---|---|---|---|---|
| 701 | 3,022.64 | 2,754.36 | 2,766.92 | 2,588.89 | 11,132.81 |
| 703 | 2,160.47 | 1,981.54 | 2,218.95 | 1,909.05 | 8,270.01 |
| 704 | 3,501.62 | 3,195.65 | 3,408.64 | 3,279.95 | 13,385.86 |
| 705 | 8,859.80 | 9,244.28 | 8,562.64 | 9,169.45 | 35,836.17 |
| 706 | 3,388.21 | 3,270.34 | 3,408.34 | 3,107.93 | 13,174.82 |
| 707 | 1,892.90 | 2,013.72 | 2,208.59 | 2,044.85 | 8,160.06 |
| 708 | 2,208.78 | 2,114.62 | 2,371.15 | 2,116.36 | 8,810.91 |
| 710 | 1,592.74 | 1,292.35 | 1,681.13 | 1,678.71 | 6,244.93 |
| 711 | 3,249.47 | 3,286.37 | 3,378.84 | 3,971.28 | 13,885.96 |
| 712 | 5,239.86 | 5,424.46 | 5,332.75 | 5,065.95 | 21,063.02 |
| 713 | 3,310.50 | 3,419.07 | 3,435.56 | 3,320.04 | 13,485.17 |
| 714 | 1,889.65 | 1,849.03 | 1,845.98 | 2,100.28 | 7,684.94 |
| TOTAL | 45,795.33 | 45,034.78 | 45,832.91 | 45,429.94 | 182,092.96 |

STATE: MAINE

| ZIP CODE | QUARTER 1 | QUARTER 2 | QUARTER 3 | QUARTER 4 | TOTAL GRAMS |
|---|---|---|---|---|---|
| 39 | 181.23 | 294.49 | 243.2 | 212.17 | 931.09 |
| 40 | 2,448.67 | 2,626.41 | 2,490.71 | 2,328.46 | 9,894.25 |
| 41 | 675.07 | 536.3 | 557.75 | 589.05 | 2,358.17 |
| 42 | 1,436.13 | 1,900.68 | 1,792.97 | 1,933.13 | 7,062.91 |
| 43 | 963.07 | 1,186.92 | 867.39 | 968.8 | 3,986.18 |
| 44 | 1,810.71 | 1,695.79 | 1,812.73 | 1,831.50 | 7,150.73 |
| 45 | 340.34 | 329.67 | 355.63 | 307.67 | 1,333.31 |
| 46 | 858.33 | 980.79 | 954.56 | 885.76 | 3,679.44 |
| 47 | 1,469.59 | 1,463.23 | 1,502 | 1,394.07 | 5,828.89 |
| 48 | 332.33 | 343.5 | 370.33 | 324.65 | 1,370.81 |
| 49 | 1,659.98 | 1,589.44 | 1,526.95 | 1,400.13 | 6,176.50 |
| TOTAL | 12,175.45 | 12,947.22 | 12,474.22 | 12,175.39 | 49,772.28 |

STATE: MARYLAND

| ZIP CODE | QUARTER 1 | QUARTER 2 | QUARTER 3 | QUARTER 4 | TOTAL GRAMS |
|---|---|---|---|---|---|
| 206 | 4,649.09 | 4,933.98 | 4,259.01 | 4,578.28 | 18,420.36 |
| 207 | 16,072.26 | 16,169.05 | 15,459.67 | 15,659.97 | 63,360.95 |
| 208 | 7,175.10 | 7,052.76 | 6,623.81 | 6,991.81 | 27,843.48 |
| 209 | 2,656.50 | 2,852.39 | 2,879.04 | 2,847.13 | 11,235.06 |
| 210 | 7,832.70 | 7,587.91 | 7,855.01 | 7,125.92 | 30,401.54 |
| 211 | 3,693.89 | 3,907.08 | 3,718.92 | 3,644.69 | 14,964.58 |
| 212 | 12,141.53 | 12,088.39 | 12,027.89 | 11,492.23 | 47,750.04 |
| 214 | 1,177.96 | 1,186.82 | 1,270.72 | 1,089.74 | 4,725.24 |
| 215 | 1,214.63 | 1,135.49 | 997.59 | 1,110.24 | 4,457.95 |
| 216 | 2,047.42 | 1,930.76 | 1,655.08 | 1,719.03 | 7,352.29 |
| 217 | 5,357.91 | 5,175.30 | 5,246.76 | 5,490.39 | 21,270.36 |
| 218 | 1,852.12 | 1,614.78 | 1,950.12 | 1,602.73 | 7,019.75 |
| 219 | 1,215.84 | 1,117.83 | 1,137.44 | 1,121.01 | 4,592.12 |
| TOTAL | 67,086.95 | 66,752.54 | 65,081.06 | 64,473.17 | 263,393.72 |

STATE: MASSACHUSETTS

| ZIP CODE | QUARTER 1 | QUARTER 2 | QUARTER 3 | QUARTER 4 | TOTAL GRAMS |
|---|---|---|---|---|---|
| 10 | 3,735.50 | 3,783.75 | 3,494.28 | 3,516.39 | 14,529.92 |
| 11 | 1,691.86 | 1,349.02 | 1,488.06 | 1,421.77 | 5,950.71 |
| 12 | 722.91 | 551.84 | 599.29 | 606.96 | 2,481 |
| 13 | 722.9 | 692.95 | 718.16 | 598.12 | 2,732.13 |
| 14 | 2,285.99 | 2,257.19 | 2,158.48 | 2,147.07 | 8,848.73 |
| 15 | 3,538.20 | 3,241.97 | 3,159.84 | 3,150.08 | 13,090.09 |
| 16 | 2,339.30 | 2,281.44 | 2,376.97 | 2,153.47 | 9,151.18 |
| 17 | 3,045.47 | 3,054.50 | 3,079.70 | 2,578.19 | 11,757.86 |
| 18 | 9,920.46 | 9,007.08 | 8,333.72 | 8,425.80 | 35,687.06 |
| 19 | 3,617.63 | 3,521.58 | 3,751.76 | 3,124.88 | 14,015.85 |
| 20 | 2,443.33 | 2,579.48 | 2,487.46 | 2,195.15 | 9,705.42 |
| 21 | 9,665.51 | 9,271.91 | 8,715.65 | 8,636.75 | 36,289.82 |
| 22 | 293.43 | 313.86 | 278.79 | 225.39 | 1,111.47 |
| 23 | 3,463.74 | 3,491.43 | 2,947.29 | 3,057.49 | 12,959.95 |

| | | | | | |
|---|---|---|---|---|---|
| 24 | 2,047.28 | 2,164.88 | 1,743.57 | 1,932.53 | 7,888.26 |
| 25 | 903.06 | 1,040.60 | 996.46 | 803.74 | 3,743.86 |
| 26 | 1,210.82 | 1,350.23 | 1,250.87 | 1,151.59 | 4,963.51 |
| 27 | 4,830.50 | 4,495.07 | 4,434.28 | 4,032.60 | 17,792.45 |
| TOTAL | 56,477.89 | 54,448.78 | 52,014.63 | 49,757.97 | 212,699.27 |

STATE: MICHIGAN

DRUG CODE:9050DRUG NAME:CODEINE
STATE:MICHIGAN

| ZIP CODE | QUARTER 1 | QUARTER 2 | QUARTER 3 | QUARTER 4 | TOTAL GRAMS |
|---|---|---|---|---|---|
| 480 | 36,745.73 | 37,245.39 | 35,797.93 | 37,664.83 | 147,453.88 |
| 481 | 54,696.73 | 57,251.81 | 54,988.44 | 56,326.27 | 223,263.25 |
| 482 | 56,142.16 | 57,406.11 | 55,408.43 | 54,760.67 | 223,717.37 |
| 483 | 18,026.01 | 18,800.38 | 18,162.35 | 19,828.48 | 74,817.22 |
| 484 | 9,627.17 | 10,376.30 | 10,432.56 | 10,573.76 | 41,009.79 |
| 485 | 11,356.30 | 10,877.09 | 11,020.46 | 10,660.38 | 43,914.23 |
| 486 | 15,813.16 | 15,850.57 | 15,162.38 | 13,445.13 | 60,271.24 |
| 487 | 6,416.27 | 7,125.90 | 6,728.90 | 6,740.60 | 27,011.67 |
| 488 | 10,971.76 | 11,522.41 | 9,401.11 | 11,190.41 | 43,085.69 |
| 489 | 3,910.15 | 3,834.54 | 3,261.90 | 4,043.34 | 15,049.93 |
| 490 | 11,296.57 | 11,897.74 | 12,201.34 | 13,870.88 | 49,266.53 |
| 491 | 1,157.98 | 1,156.61 | 1,091.23 | 1,183.86 | 4,589.68 |
| 492 | 6,804.21 | 6,853.07 | 6,140.92 | 6,766.81 | 26,565.01 |
| 493 | 3,261.51 | 3,256.70 | 2,686.92 | 3,278.79 | 12,483.92 |
| 494 | 9,090.29 | 9,294.07 | 8,503.01 | 9,258.59 | 36,145.96 |
| 495 | 4,900.13 | 4,723.33 | 3,801.74 | 4,977.10 | 18,402.30 |
| 496 | 4,406.54 | 4,271.71 | 4,131.49 | 4,222.81 | 17,032.55 |
| 497 | 3,853.47 | 3,984.15 | 3,968.32 | 4,216.51 | 16,022.45 |
| 498 | 3,334.66 | 3,225.66 | 3,469.23 | 3,189.54 | 13,219.09 |
| 499 | 1,287.15 | 1,294.94 | 1,451.83 | 1,322.03 | 5,355.95 |
| TOTAL | 273,097.95 | 280,248.48 | 267,810.49 | 277,520.79 | 1,098,677.71 |

STATE: MINNESOTA

| ZIP CODE | QUARTER 1 | QUARTER 2 | QUARTER 3 | QUARTER 4 | TOTAL GRAMS |
|---|---|---|---|---|---|
| 550 | 4,482.73 | 4,267.19 | 4,008.48 | 4,555.76 | 17,314.16 |
| 551 | 7,190.47 | 7,143.03 | 6,505.59 | 6,716.51 | 27,555.60 |
| 553 | 6,522.25 | 6,241.81 | 6,162.77 | 6,313.12 | 25,239.95 |
| 554 | 8,852.85 | 8,660.76 | 8,175.56 | 7,853.87 | 33,543.04 |
| 556 | 176.53 | 153 | 118.4 | 88.55 | 536.48 |
| 557 | 2,280.49 | 2,009.61 | 1,970.43 | 1,532.40 | 7,792.93 |
| 558 | 1,045.13 | 1,075.69 | 908.3 | 916.39 | 3,945.51 |
| 559 | 1,924.77 | 1,648.23 | 1,729.56 | 1,656.77 | 6,959.33 |
| 560 | 1,488.58 | 1,497.85 | 1,602.25 | 1,467.84 | 6,056.52 |
| 561 | 742.56 | 680.49 | 831.97 | 631.67 | 2,886.69 |
| 562 | 1,784.84 | 2,012.48 | 1,841.39 | 1,742 | 7,380.71 |
| 563 | 3,036.19 | 2,965.20 | 3,009.48 | 2,621.82 | 11,632.69 |
| 564 | 1,389.19 | 1,604.20 | 1,530.82 | 1,258.95 | 5,783.16 |
| 565 | 1,578.17 | 1,486.68 | 1,535.30 | 1,412.07 | 6,012.22 |
| 566 | 900.46 | 945.78 | 940.93 | 854.41 | 3,641.58 |
| 567 | 925.53 | 847.13 | 876.3 | 850.45 | 3,499.41 |
| TOTAL | 44,320.74 | 43,239.13 | 41,747.53 | 40,472.58 | 169,779.98 |

STATE: MISSISSIPPI

| ZIP CODE | QUARTER 1 | QUARTER 2 | QUARTER 3 | QUARTER 4 | TOTAL GRAMS |
|---|---|---|---|---|---|
| 386 | 3,340.50 | 3,304.98 | 3,043.84 | 3,257.67 | 12,946.99 |
| 387 | 1,422.21 | 1,650.94 | 1,782.81 | 1,557.20 | 6,413.16 |
| 388 | 3,138.43 | 3,330.14 | 2,806.26 | 3,235.74 | 12,510.57 |
| 389 | 1,065.58 | 1,235 | 1,159.42 | 1,259.98 | 4,719.98 |

| | | | | | |
|---|---|---|---|---|---|
| 390 | 2,074.46 | 2,080.93 | 2,293.73 | 2,271.80 | 8,720.92 |
| 391 | 3,025.45 | 2,959.21 | 3,250.08 | 3,266.75 | 12,501.49 |
| 392 | 1,778.03 | 1,614.85 | 1,713 | 1,770.52 | 6,876.40 |
| 393 | 2,489.38 | 2,337.58 | 2,472.14 | 2,513.91 | 9,813.01 |
| 394 | 3,613.50 | 3,630.82 | 3,757.78 | 3,589.88 | 14,591.98 |
| 395 | 3,411.73 | 2,936.90 | 2,778.80 | 2,766.86 | 11,894.29 |
| 396 | 1,121.53 | 1,253.36 | 1,031.59 | 1,115.66 | 4,522.14 |
| 397 | 1,708.33 | 2,025.26 | 1,977.10 | 1,996.62 | 7,707.31 |
| TOTAL | 28,189.13 | 28,359.97 | 28,066.55 | 28,602.59 | 113,218.24 |

STATE: MISSOURI

| ZIP CODE | QUARTER 1 | QUARTER 2 | QUARTER 3 | QUARTER 4 | TOTAL GRAMS |
|---|---|---|---|---|---|
| 630 | 7,608.66 | 7,601.36 | 7,232.05 | 7,592.45 | 30,034.52 |
| 631 | 20,212.98 | 19,845.97 | 20,425.79 | 20,274.27 | 80,759.01 |
| 633 | 3,885.61 | 3,930.49 | 3,922.79 | 3,823.15 | 15,562.04 |
| 634 | 842.57 | 870.77 | 792.67 | 644.23 | 3,150.24 |
| 635 | 768.01 | 955.13 | 889.66 | 771.51 | 3,384.31 |
| 636 | 2,123.67 | 2,183.66 | 2,129.81 | 2,204.65 | 8,641.79 |
| 637 | 1,135.08 | 1,095.68 | 1,289.81 | 1,099.33 | 4,619.90 |
| 638 | 2,190.90 | 2,218.93 | 2,636.09 | 2,323.36 | 9,369.28 |
| 639 | 1,638.02 | 1,648.40 | 1,446.22 | 1,319.42 | 6,052.06 |

DRUG CODE:9050DRUG NAME:CODEINE
STATE:MISSOURI

| ZIP CODE | QUARTER 1 | QUARTER 2 | QUARTER 3 | QUARTER 4 | TOTAL GRAMS |
|---|---|---|---|---|---|
| 640 | 6,745.87 | 6,520.82 | 6,327.27 | 6,239.74 | 25,833.70 |
| 641 | 6,346.17 | 6,462.52 | 6,332.28 | 5,835.71 | 24,976.68 |
| 644 | 609.06 | 646.74 | 722.07 | 607.08 | 2,584.95 |
| 645 | 794.7 | 736.28 | 717.68 | 699.08 | 2,947.74 |
| 646 | 679.59 | 703.68 | 841.44 | 738.18 | 2,962.89 |
| 647 | 1,770.73 | 1,657.91 | 1,493.46 | 1,662.80 | 6,584.90 |
| 648 | 2,119.82 | 1,999.87 | 2,010.50 | 1,855.02 | 7,985.21 |
| 650 | 2,870.62 | 2,963.91 | 2,329.52 | 1,372.64 | 9,536.69 |
| 651 | 761.21 | 918.59 | 750.03 | 770.22 | 3,200.05 |
| 652 | 2,403.63 | 2,367.65 | 2,162.70 | 2,100.59 | 9,034.57 |
| 653 | 1,214.06 | 1,332.30 | 1,196.74 | 1,233.76 | 4,976.86 |
| 654 | 754.71 | 896.72 | 823.35 | 834.95 | 3,309.73 |
| 655 | 1,541.87 | 1,377.86 | 1,190.62 | 1,301.98 | 5,412.33 |
| 656 | 1,769.13 | 1,453.74 | 1,407.98 | 1,522.75 | 6,153.60 |
| 657 | 2,886.90 | 2,969.23 | 2,960.47 | 2,761.29 | 11,577.89 |
| 658 | 2,222.02 | 2,295.37 | 2,318.22 | 2,115.89 | 8,951.50 |
| TOTAL | 75,895.59 | 75,653.58 | 74,349.22 | 71,704.05 | 297,602.44 |

STATE: MONTANA

| ZIP CODE | QUARTER 1 | QUARTER 2 | QUARTER 3 | QUARTER 4 | TOTAL GRAMS |
|---|---|---|---|---|---|
| 590 | 784.91 | 787.72 | 844.43 | 733.18 | 3,150.24 |
| 591 | 1,286.86 | 1,290.16 | 1,333.61 | 1,319.55 | 5,230.18 |
| 592 | 622.84 | 494.81 | 449.83 | 384.48 | 1,951.96 |
| 593 | 353.62 | 428.94 | 475.17 | 393.4 | 1,651.13 |
| 594 | 1,646.09 | 1,548.31 | 1,604.03 | 1,333.65 | 6,132.08 |
| 595 | 279.58 | 353.62 | 247.53 | 249.74 | 1,130.47 |
| 596 | 1,145.76 | 975.23 | 931.63 | 996.53 | 4,049.15 |
| 597 | 1,625.62 | 1,540.39 | 1,689.03 | 1,638.18 | 6,493.22 |
| 598 | 2,485.90 | 2,273.09 | 2,426.79 | 2,379.18 | 9,564.96 |
| 599 | 1,604.77 | 1,586.16 | 1,617.55 | 1,565.99 | 6,374.47 |
| TOTAL | 11,835.95 | 11,278.43 | 11,619.60 | 10,993.88 | 45,727.86 |

STATE: NEBRASKA

| ZIP CODE | QUARTER 1 | QUARTER 2 | QUARTER 3 | QUARTER 4 | TOTAL GRAMS |
|---|---|---|---|---|---|

| | | | | | |
|---|---|---|---|---|---|
| 680 | 1,146.84 | 1,234.11 | 1,226.26 | 1,229.28 | 4,836.49 |
| 681 | 4,905.64 | 5,119.38 | 4,787.40 | 4,577.30 | 19,389.72 |
| 683 | 259.7 | 290.44 | 260.51 | 267.4 | 1,078.05 |
| 684 | 331.66 | 294.27 | 281.32 | 325.69 | 1,232.94 |
| 685 | 2,361.01 | 2,345.80 | 2,445.70 | 2,378.50 | 9,531.01 |
| 686 | 201.12 | 242.28 | 211.08 | 168.7 | 823.18 |
| 687 | 714.3 | 901.17 | 916.86 | 974.25 | 3,506.58 |
| 688 | 1,125.51 | 1,321.63 | 1,158.28 | 1,296.30 | 4,901.72 |
| 689 | 418.22 | 435.1 | 436.86 | 495.7 | 1,785.88 |
| 690 | 204.21 | 207.31 | 173.27 | 156.92 | 741.71 |
| 691 | 1,079.17 | 1,130.51 | 1,125.84 | 823.24 | 4,158.76 |
| 692 | 39.78 | 66.3 | 67.41 | 54.15 | 227.64 |
| 693 | 585.48 | 562.34 | 576.63 | 579.4 | 2,303.85 |
| TOTAL | 13,372.64 | 14,150.64 | 13,667.42 | 13,326.83 | 54,517.53 |

STATE: NEVADA

| ZIP CODE | QUARTER 1 | QUARTER 2 | QUARTER 3 | QUARTER 4 | TOTAL GRAMS |
|---|---|---|---|---|---|
| 890 | 8,177.01 | 7,267.96 | 6,846.34 | 7,085.74 | 29,377.05 |
| 891 | 17,030.63 | 16,087.93 | 14,813.65 | 15,419.66 | 63,351.87 |
| 893 | 181.23 | 130.4 | 132.61 | 70.72 | 514.96 |
| 894 | 2,963.83 | 2,774.98 | 2,473.95 | 2,654.41 | 10,867.17 |
| 895 | 2,620.81 | 2,539.11 | 2,381.19 | 2,159.47 | 9,700.58 |
| 897 | 629.83 | 730.21 | 651.67 | 711.76 | 2,723.47 |
| 898 | 365.77 | 335.94 | 374.61 | 382.35 | 1,458.67 |
| TOTAL | 31,969.11 | 29,866.53 | 27,674.02 | 28,484.11 | 117,993.77 |

STATE: NEW HAMPSHIRE

| ZIP CODE | QUARTER 1 | QUARTER 2 | QUARTER 3 | QUARTER 4 | TOTAL GRAMS |
|---|---|---|---|---|---|
| 30 | 2,470.09 | 2,376.89 | 2,377.33 | 2,173.27 | 9,397.58 |
| 31 | 1,496.70 | 1,489.77 | 1,595.15 | 1,396.50 | 5,978.12 |
| 32 | 1,046.64 | 1,159.45 | 1,018.62 | 990.41 | 4,215.12 |
| 33 | 713.79 | 657.22 | 644.13 | 787.11 | 2,802.25 |
| 34 | 473.78 | 486.57 | 607.75 | 563 | 2,131.10 |
| 35 | 380.2 | 391.27 | 364 | 448.08 | 1,583.55 |
| 36 | 26.52 | 48.62 | 30.94 | 26.52 | 132.6 |
| 37 | 705.54 | 406.44 | 542 | 672.03 | 2,326.01 |

DRUG CODE:9050DRUG NAME:CODEINE
STATE:NEW HAMPSHIRE

| ZIP CODE | QUARTER 1 | QUARTER 2 | QUARTER 3 | QUARTER 4 | TOTAL GRAMS |
|---|---|---|---|---|---|
| 38 | 1,833.72 | 1,758.69 | 1,583.26 | 1,698.18 | 6,873.85 |
| TOTAL | 9,146.98 | 8,774.92 | 8,763.18 | 8,755.10 | 35,440.18 |

STATE: NEW JERSEY

| ZIP CODE | QUARTER 1 | QUARTER 2 | QUARTER 3 | QUARTER 4 | TOTAL GRAMS |
|---|---|---|---|---|---|
| 70 | 14,542.37 | 14,428.18 | 14,328.12 | 14,059.27 | 57,357.94 |
| 71 | 3,018.07 | 3,218.59 | 2,986.27 | 2,963.07 | 12,186 |
| 72 | 1,677.58 | 1,724.17 | 1,404.42 | 1,130.64 | 5,936.81 |
| 73 | 1,982.91 | 2,136.55 | 1,982.63 | 2,236.82 | 8,338.91 |
| 74 | 5,022.83 | 4,893.60 | 4,660.58 | 5,350.96 | 19,927.97 |
| 75 | 1,596.80 | 1,595.12 | 1,614.59 | 1,884.80 | 6,691.31 |
| 76 | 3,988.57 | 3,899.85 | 4,107.82 | 4,737.23 | 16,733.47 |
| 77 | 6,512.65 | 6,493 | 6,621.36 | 6,049.47 | 25,676.48 |
| 78 | 3,551.20 | 3,551.24 | 3,341.19 | 3,320.68 | 13,764.31 |
| 79 | 1,872.52 | 2,167.82 | 1,826.46 | 1,846.08 | 7,712.88 |
| 80 | 15,576.77 | 15,920.97 | 15,412.75 | 15,500.53 | 62,411.02 |
| 81 | 2,637.17 | 3,096.93 | 2,600.37 | 2,784.72 | 11,119.19 |
| 82 | 2,698.91 | 2,956.71 | 2,686.64 | 2,751.75 | 11,094.01 |
| 83 | 3,912.33 | 3,953.75 | 3,494.41 | 3,396.10 | 14,756.59 |

| | | | | | |
|---|---|---|---|---|---|
| 84 | 547.36 | 486.5 | 591.57 | 414.39 | 2,039.82 |
| 85 | 2,063.55 | 2,188.32 | 2,041.97 | 2,242.27 | 8,536.11 |
| 86 | 3,240.31 | 3,176.96 | 3,185.28 | 3,180.88 | 12,783.43 |
| 87 | 4,566.77 | 4,889.66 | 4,825.59 | 4,716.10 | 18,998.12 |
| 88 | 7,740.03 | 8,183.43 | 8,104.98 | 8,138.91 | 32,167.35 |
| 89 | 669.47 | 690.74 | 659.63 | 627.79 | 2,647.63 |
| TOTAL | 87,418.17 | 89,652.09 | 86,476.63 | 87,332.46 | 350,879.35 |

STATE: NEW MEXICO

| ZIP CODE | QUARTER 1 | QUARTER 2 | QUARTER 3 | QUARTER 4 | TOTAL GRAMS |
|---|---|---|---|---|---|
| 870 | 2,045.81 | 2,134.76 | 2,213.92 | 2,023.71 | 8,418.20 |
| 871 | 7,901.58 | 6,961.27 | 6,961.13 | 6,540.35 | 28,364.33 |
| 873 | 2,098.21 | 1,798.93 | 1,851.25 | 2,041.49 | 7,789.88 |
| 874 | 1,285.90 | 1,087.84 | 1,082.41 | 1,155.88 | 4,612.03 |
| 875 | 1,316.60 | 1,348.83 | 1,267.08 | 1,204.93 | 5,137.44 |
| 877 | 573.61 | 324.88 | 445.7 | 373.31 | 1,717.50 |
| 878 | 39.78 | 66.3 | 100 | 79.56 | 285.64 |
| 879 | 209.96 | 165.76 | 204.43 | 212.72 | 792.87 |
| 880 | 1,880.72 | 1,795.81 | 1,807.39 | 2,073.64 | 7,557.56 |
| 881 | 968.15 | 1,035.83 | 1,062.50 | 1,017.41 | 4,083.89 |
| 882 | 2,830.70 | 2,690.01 | 3,054.95 | 2,890.36 | 11,466.02 |
| 883 | 853.71 | 963.65 | 891.99 | 900.46 | 3,609.81 |
| 884 | 112.42 | 81.92 | 184.62 | 109.91 | 488.87 |
| TOTAL | 22,117.15 | 20,455.79 | 21,127.37 | 20,623.73 | 84,324.04 |

STATE: NEW YORK

| ZIP CODE | QUARTER 1 | QUARTER 2 | QUARTER 3 | QUARTER 4 | TOTAL GRAMS |
|---|---|---|---|---|---|
| 63 | 0 | 0 | 0 | 2.21 | 2.21 |
| 100 | 19,739.18 | 17,934.73 | 15,830.40 | 16,065.96 | 69,570.27 |
| 101 | 581.97 | 696.07 | 569.17 | 534.52 | 2,381.73 |
| 102 | 34.71 | 22.1 | 23.21 | 60.04 | 140.06 |
| 103 | 3,521.01 | 3,605.11 | 3,331.55 | 3,358.01 | 13,815.68 |
| 104 | 16,455.50 | 14,017.97 | 12,886.21 | 12,886.38 | 56,246.06 |
| 105 | 4,366.75 | 3,807.87 | 3,812.66 | 4,068.73 | 16,056.01 |
| 106 | 801.96 | 869.38 | 666.42 | 703.52 | 3,041.28 |
| 107 | 2,200.60 | 1,768.26 | 1,899.58 | 1,737.28 | 7,605.72 |
| 108 | 797.64 | 556.4 | 620.84 | 479.4 | 2,454.28 |
| 109 | 4,163.79 | 4,004.15 | 3,972.20 | 4,464.49 | 16,604.63 |
| 110 | 1,759 | 1,729.04 | 1,589 | 1,664.46 | 6,741.50 |
| 111 | 1,643.77 | 1,844.62 | 1,533.61 | 1,700.11 | 6,722.11 |
| 112 | 23,286.17 | 21,540.71 | 20,268.42 | 19,799.15 | 84,894.45 |
| 113 | 9,071.91 | 8,514.23 | 8,388.66 | 7,702.03 | 33,676.83 |
| 114 | 4,586.32 | 4,633.51 | 4,053.63 | 4,341.63 | 17,615.09 |
| 115 | 5,721.87 | 5,069.09 | 4,729.01 | 4,738.42 | 20,258.39 |
| 116 | 1,088.07 | 1,187.31 | 993.74 | 954.76 | 4,223.88 |
| 117 | 10,200.27 | 9,833.44 | 9,360.13 | 9,232.28 | 38,626.12 |
| 118 | 756.47 | 736.52 | 483.12 | 496.3 | 2,472.41 |
| 119 | 1,456.28 | 1,515.43 | 1,429.95 | 1,498.71 | 5,900.37 |
| 120 | 1,709.23 | 1,694.46 | 1,731.16 | 1,801.83 | 6,936.68 |
| 121 | 899.77 | 771.3 | 730.96 | 800.77 | 3,202.80 |

DRUG CODE:9050DRUG NAME:CODEINE
STATE:NEW YORK

| ZIP CODE | QUARTER 1 | QUARTER 2 | QUARTER 3 | QUARTER 4 | TOTAL GRAMS |
|---|---|---|---|---|---|
| 122 | 846.37 | 701.32 | 644.31 | 691.27 | 2,883.27 |
| 123 | 988.64 | 764.88 | 802.62 | 889.93 | 3,446.07 |
| 124 | 1,229.12 | 1,239.22 | 1,237.44 | 1,116.23 | 4,822.01 |
| 125 | 2,652.29 | 2,900.90 | 2,784.35 | 2,729.72 | 11,067.26 |
| 126 | 665.89 | 584.19 | 565.68 | 483.27 | 2,299.03 |
| 127 | 1,144.62 | 1,114.89 | 1,146.93 | 1,197.30 | 4,603.74 |

| | | | | | |
|---|---|---|---|---|---|
| 128 | 1,821.07 | 1,651.02 | 1,815.61 | 1,677.18 | 6,964.88 |
| 129 | 1,608.36 | 1,441.59 | 1,481.37 | 1,630.65 | 6,161.97 |
| 130 | 3,410.70 | 3,383.10 | 3,374.78 | 3,401.14 | 13,569.72 |
| 131 | 1,308.16 | 1,331.77 | 1,334.88 | 1,241.56 | 5,216.37 |
| 132 | 2,536.65 | 2,407.65 | 2,384.99 | 2,407.34 | 9,736.63 |
| 133 | 981.45 | 970.4 | 901.89 | 1,021.48 | 3,875.22 |
| 134 | 1,428.88 | 1,169.57 | 1,110.46 | 1,215.79 | 4,924.70 |
| 135 | 621.88 | 589.17 | 557.67 | 534.47 | 2,303.19 |
| 136 | 3,992.56 | 3,857.43 | 3,672.71 | 3,547.88 | 15,070.58 |
| 137 | 1,122.63 | 997.96 | 898.41 | 956.41 | 3,975.41 |
| 138 | 1,579.99 | 1,391.36 | 1,419.94 | 1,394.90 | 5,786.19 |
| 139 | 1,145.16 | 1,136.77 | 1,019.07 | 1,019.94 | 4,320.94 |
| 140 | 5,751.27 | 5,310.93 | 5,216.66 | 5,470.44 | 21,749.30 |
| 141 | 3,136.70 | 2,651.78 | 2,765.42 | 2,969.10 | 11,523 |
| 142 | 11,746.42 | 11,128.57 | 10,325.42 | 10,189.94 | 43,390.35 |
| 143 | 1,018.45 | 815.83 | 902.5 | 876.63 | 3,613.41 |
| 144 | 3,016.66 | 3,039.66 | 2,798.90 | 2,835.34 | 11,690.56 |
| 145 | 2,276.53 | 2,102.50 | 2,244.31 | 2,257.96 | 8,881.30 |
| 146 | 4,759 | 4,675.29 | 4,599.94 | 4,314.31 | 18,348.54 |
| 147 | 1,393.34 | 1,344.51 | 1,325.39 | 1,274.46 | 5,337.70 |
| 148 | 2,662.97 | 2,357.09 | 2,379.08 | 2,405.02 | 9,804.16 |
| 149 | 491.36 | 447.9 | 483.82 | 455.07 | 1,878.15 |
| TOTAL | 180,179.36 | 167,858.95 | 159,098.18 | 159,295.72 | 666,432.21 |

STATE: NORTH CAROLINA

| ZIP CODE | QUARTER 1 | QUARTER 2 | QUARTER 3 | QUARTER 4 | TOTAL GRAMS |
|---|---|---|---|---|---|
| 270 | 1,128.57 | 1,263.49 | 1,166.28 | 1,126.64 | 4,684.98 |
| 271 | 1,304.58 | 1,307.78 | 1,212.15 | 1,210.48 | 5,034.99 |
| 272 | 2,703.07 | 2,195.83 | 2,397.10 | 2,278.11 | 9,574.11 |
| 273 | 1,850.90 | 1,780.02 | 1,721.23 | 1,914.56 | 7,266.71 |
| 274 | 2,085.94 | 1,756.17 | 1,814.68 | 1,946.60 | 7,603.39 |
| 275 | 5,778.53 | 5,907.74 | 5,243.24 | 5,839.86 | 22,769.37 |
| 276 | 2,226.96 | 2,085.68 | 2,333.35 | 2,325.68 | 8,971.67 |
| 277 | 2,001.81 | 1,606.30 | 1,570.76 | 1,751.22 | 6,930.09 |
| 278 | 2,834.83 | 3,227.20 | 3,103.24 | 3,322.57 | 12,487.84 |
| 279 | 1,363.34 | 1,576.70 | 1,450.73 | 1,578.61 | 5,969.38 |
| 280 | 2,922.27 | 2,893.56 | 2,972.29 | 2,917.31 | 11,705.43 |
| 281 | 2,450.38 | 2,377.09 | 2,122.56 | 2,293.83 | 9,243.86 |
| 282 | 2,727.45 | 2,561.23 | 2,487.53 | 2,800.80 | 10,577.01 |
| 283 | 8,464.67 | 8,088.61 | 7,557.50 | 7,725.69 | 31,836.47 |
| 284 | 2,750.93 | 2,222.70 | 2,507.14 | 2,273.36 | 9,754.13 |
| 285 | 3,701.65 | 3,410.05 | 3,524.62 | 3,663.41 | 14,299.73 |
| 286 | 3,446.08 | 3,146.87 | 3,131.77 | 3,188.79 | 12,913.51 |
| 287 | 2,344.03 | 2,362.24 | 2,392.04 | 2,224.91 | 9,323.22 |
| 288 | 937.44 | 1,013.49 | 929.99 | 922.15 | 3,803.07 |
| 289 | 847.09 | 730.14 | 860.26 | 988.08 | 3,425.57 |
| TOTAL | 53,870.52 | 51,512.89 | 50,498.46 | 52,292.66 | 208,174.53 |

STATE: NORTH DAKOTA

| ZIP CODE | QUARTER 1 | QUARTER 2 | QUARTER 3 | QUARTER 4 | TOTAL GRAMS |
|---|---|---|---|---|---|
| 580 | 478.66 | 466.85 | 519.35 | 413.09 | 1,877.95 |
| 581 | 1,280.24 | 973.55 | 979.16 | 919.05 | 4,152 |
| 582 | 945.61 | 1,114.87 | 1,055.09 | 686.39 | 3,801.96 |
| 583 | 703.18 | 714.23 | 668.56 | 480.7 | 2,566.67 |
| 584 | 401.08 | 269.78 | 272.31 | 258.93 | 1,202.10 |
| 585 | 1,453.17 | 1,402.03 | 1,286.57 | 983.81 | 5,125.58 |
| 586 | 293.94 | 282.33 | 337.76 | 241.53 | 1,155.56 |
| 587 | 773.9 | 841.37 | 633.74 | 465.76 | 2,714.77 |
| 588 | 603.98 | 589.1 | 633.7 | 666.86 | 2,493.64 |
| TOTAL | 6,933.76 | 6,654.11 | 6,386.24 | 5,116.12 | 25,090.23 |

STATE: OHIO

RETAIL DRUG DISTRIBUTION BY ZIP CODE WITHIN STATE BY GRAMS WT

DRUG CODE:9050DRUG NAME:CODEINE
STATE:OHIO

| ZIP CODE | QUARTER 1 | QUARTER 2 | QUARTER 3 | QUARTER 4 | TOTAL GRAMS |
|---|---|---|---|---|---|
| 430 | 5,684.61 | 5,247.28 | 5,335.21 | 4,888.69 | 21,155.79 |
| 431 | 3,679.35 | 3,622.86 | 3,663.53 | 3,829.68 | 14,795.42 |
| 432 | 17,208.16 | 16,873.73 | 16,946.14 | 14,895.95 | 65,923.98 |
| 433 | 1,120.43 | 1,141.40 | 1,124.72 | 1,018.86 | 4,405.41 |
| 434 | 1,823.44 | 1,865.66 | 1,635.28 | 1,624.95 | 6,949.33 |
| 435 | 3,834.12 | 3,763.05 | 3,582.73 | 3,542.02 | 14,721.92 |
| 436 | 7,094.30 | 7,210.47 | 6,770.82 | 7,195.70 | 28,271.29 |
| 437 | 1,905.86 | 1,776.07 | 2,011.15 | 2,012.42 | 7,705.50 |
| 438 | 327.46 | 373.87 | 448.01 | 382.04 | 1,531.38 |
| 439 | 2,151.09 | 2,086.14 | 2,178.54 | 2,027.35 | 8,443.12 |
| 440 | 6,970.68 | 5,779.21 | 5,520.83 | 5,746.27 | 24,016.99 |
| 441 | 12,082.84 | 10,540.97 | 10,789.73 | 11,428.40 | 44,841.94 |
| 442 | 3,393.90 | 2,710.57 | 2,840.88 | 3,099.89 | 12,045.24 |
| 443 | 1,893.26 | 1,602.06 | 1,518.80 | 1,519.36 | 6,533.48 |
| 444 | 2,648.43 | 2,443.32 | 2,624.14 | 2,861.85 | 10,577.74 |
| 445 | 2,032.93 | 1,897.82 | 1,877.35 | 2,122.31 | 7,930.41 |
| 446 | 3,135.94 | 2,555.28 | 2,769.72 | 2,779.07 | 11,240.01 |
| 447 | 2,211.98 | 2,043.50 | 1,897.95 | 2,313.14 | 8,466.57 |
| 448 | 3,323.67 | 3,058.92 | 2,865.87 | 2,843.50 | 12,091.96 |
| 449 | 1,391.43 | 1,519.60 | 1,320.69 | 1,276.93 | 5,508.65 |
| 450 | 4,991.43 | 5,136.57 | 4,991.98 | 4,757.61 | 19,877.59 |
| 451 | 2,587.73 | 2,434.50 | 2,209.69 | 2,049.47 | 9,281.39 |
| 452 | 7,501.68 | 7,567.32 | 7,239.76 | 7,204.32 | 29,513.08 |
| 453 | 3,996.56 | 4,146.10 | 3,905.20 | 3,792.15 | 15,840.01 |
| 454 | 6,443.79 | 6,304.53 | 5,701.45 | 6,245.11 | 24,694.88 |
| 455 | 883.63 | 945.16 | 881.72 | 827.35 | 3,537.86 |
| 456 | 4,316.57 | 4,485.66 | 4,165.35 | 4,234.78 | 17,202.36 |
| 457 | 1,412.79 | 1,487 | 1,557.75 | 1,809.70 | 6,267.24 |
| 458 | 3,695.32 | 3,401.24 | 3,274.67 | 3,353.95 | 13,725.18 |
| TOTAL | 119,743.38 | 114,019.86 | 111,649.66 | 111,682.82 | 457,095.72 |

STATE: OKLAHOMA

| ZIP CODE | QUARTER 1 | QUARTER 2 | QUARTER 3 | QUARTER 4 | TOTAL GRAMS |
|---|---|---|---|---|---|
| 730 | 14,569.14 | 14,055.82 | 13,745.45 | 14,604.02 | 56,974.43 |
| 731 | 13,149.71 | 12,936.71 | 12,962.55 | 13,227.17 | 52,276.14 |
| 734 | 3,497.96 | 3,875.37 | 3,790.34 | 3,709.51 | 14,873.18 |
| 735 | 9,727.13 | 9,404.85 | 9,492.48 | 9,075.28 | 37,699.74 |
| 736 | 1,341.15 | 1,333.18 | 1,368.49 | 1,862.30 | 5,905.12 |
| 737 | 1,890.44 | 1,863.26 | 1,809.40 | 1,988.76 | 7,551.86 |
| 738 | 685.48 | 593.4 | 520.66 | 544.35 | 2,343.89 |
| 739 | 422.49 | 535.21 | 577.02 | 542.94 | 2,077.66 |
| 740 | 12,847.44 | 12,807.80 | 13,215.80 | 13,116.66 | 51,987.70 |
| 741 | 4,848.13 | 5,078.08 | 5,261.88 | 4,699.81 | 19,887.90 |
| 743 | 3,715.27 | 3,994.38 | 4,122.09 | 3,790.75 | 15,622.49 |
| 744 | 6,865.20 | 7,049.86 | 6,620.63 | 6,475.45 | 27,011.14 |
| 745 | 1,977.82 | 2,023.36 | 2,061.80 | 2,042.49 | 8,105.47 |
| 746 | 1,942.46 | 1,898.25 | 1,901.13 | 1,836.59 | 7,578.43 |
| 747 | 5,350.09 | 5,325.01 | 6,418.13 | 6,222.49 | 23,315.72 |
| 748 | 8,693.44 | 8,806.39 | 8,838.24 | 8,853.14 | 35,191.21 |
| 749 | 3,452.18 | 3,432.29 | 3,149.57 | 3,750.54 | 13,784.58 |
| TOTAL | 94,975.53 | 95,013.22 | 95,855.66 | 96,342.25 | 382,186.66 |

STATE: OREGON

| ZIP CODE | QUARTER 1 | QUARTER 2 | QUARTER 3 | QUARTER 4 | TOTAL GRAMS |
|---|---|---|---|---|---|

| | | | | | |
|---|---|---|---|---|---|
| 970 | 7,108.58 | 6,353.36 | 6,819.66 | 6,852.84 | 27,134.44 |
| 971 | 2,580.69 | 2,533.76 | 2,560.82 | 2,444.24 | 10,119.51 |
| 972 | 9,221.17 | 8,920.34 | 8,829.39 | 8,520.06 | 35,490.96 |
| 973 | 5,286.93 | 5,150.68 | 4,920.17 | 4,723.42 | 20,081.20 |
| 974 | 5,820.69 | 5,738.07 | 5,505.79 | 5,532.31 | 22,596.86 |
| 975 | 3,448.91 | 3,966.12 | 3,644.61 | 3,433.71 | 14,493.35 |
| 976 | 722.27 | 555.56 | 546.62 | 668.72 | 2,493.17 |
| 977 | 1,996.68 | 1,846.55 | 1,627.55 | 1,673.77 | 7,144.55 |
| 978 | 1,879.68 | 1,902.14 | 1,637.25 | 1,731.86 | 7,150.93 |
| 979 | 340.92 | 392.02 | 343.36 | 424.5 | 1,500.80 |
| TOTAL | 38,406.52 | 37,358.60 | 36,435.22 | 36,005.43 | 148,205.77 |

STATE: PENNSYLVANIA

| ZIP CODE | QUARTER 1 | QUARTER 2 | QUARTER 3 | QUARTER 4 | TOTAL GRAMS |
|---|---|---|---|---|---|
| 150 | 4,439.60 | 3,054.42 | 3,300.85 | 4,115.94 | 14,910.81 |

DRUG CODE:9050DRUG NAME:CODEINE
STATE:PENNSYLVANIA

| ZIP CODE | QUARTER 1 | QUARTER 2 | QUARTER 3 | QUARTER 4 | TOTAL GRAMS |
|---|---|---|---|---|---|
| 151 | 3,628.90 | 2,372.77 | 2,394.10 | 3,288.12 | 11,683.89 |
| 152 | 6,547.21 | 4,803.43 | 4,950.58 | 5,791.08 | 22,092.30 |
| 153 | 1,565.75 | 1,335.78 | 1,358.46 | 1,527.57 | 5,787.56 |
| 154 | 1,213.45 | 1,203.76 | 1,123.43 | 1,041.63 | 4,582.27 |
| 155 | 2,606.69 | 2,772.42 | 2,741.79 | 2,621.72 | 10,742.62 |
| 156 | 2,392.12 | 1,873.66 | 1,845.74 | 2,066.29 | 8,177.81 |
| 157 | 2,245.90 | 2,020.06 | 1,576.82 | 1,840.59 | 7,683.37 |
| 158 | 781.01 | 790.11 | 892.88 | 856.43 | 3,320.43 |
| 159 | 1,449.67 | 1,453.54 | 1,254.91 | 1,502.42 | 5,660.54 |
| 160 | 1,936.59 | 1,364.94 | 1,561.08 | 1,610.13 | 6,472.74 |
| 161 | 2,053.32 | 1,748.93 | 1,916.59 | 2,070.40 | 7,789.24 |
| 162 | 1,795.42 | 1,679.45 | 1,509.90 | 1,481.35 | 6,466.12 |
| 163 | 1,676.79 | 1,549.06 | 1,604.03 | 1,508.87 | 6,338.75 |
| 164 | 807.1 | 496.05 | 655.72 | 760.33 | 2,719.20 |
| 165 | 2,057.30 | 1,714.40 | 1,752.16 | 1,623.91 | 7,147.77 |
| 166 | 1,963.78 | 1,888.23 | 1,850.22 | 1,788.84 | 7,491.07 |
| 167 | 757.32 | 753.64 | 770.22 | 702.81 | 2,983.99 |
| 168 | 988.01 | 1,020.28 | 929.95 | 972.67 | 3,910.91 |
| 169 | 443.45 | 422.37 | 386.91 | 495.06 | 1,747.79 |
| 170 | 5,828.63 | 5,989.88 | 5,529.11 | 5,718.22 | 23,065.84 |
| 171 | 1,307.63 | 1,616.81 | 1,353.76 | 1,527.64 | 5,805.84 |
| 172 | 1,808.87 | 1,959.17 | 1,711.64 | 1,648.83 | 7,128.51 |
| 173 | 2,938.49 | 2,805.91 | 2,765.76 | 2,869.31 | 11,379.47 |
| 174 | 1,954.15 | 1,787.80 | 1,680.75 | 1,783.71 | 7,206.41 |
| 175 | 1,790.55 | 1,843.02 | 1,794.33 | 1,808.10 | 7,236 |
| 176 | 1,407.12 | 1,383.19 | 1,437.52 | 1,478.21 | 5,706.04 |
| 177 | 1,534 | 1,435.09 | 1,332.26 | 1,297.24 | 5,598.59 |
| 178 | 2,401.09 | 2,573.46 | 2,133.07 | 2,185.45 | 9,293.07 |
| 179 | 2,008.02 | 1,906.54 | 1,840.94 | 1,799.39 | 7,554.89 |
| 180 | 3,678.40 | 3,990.27 | 3,763 | 3,258.96 | 14,690.63 |
| 181 | 2,065.38 | 1,901.13 | 1,868.15 | 1,719.53 | 7,554.19 |
| 182 | 1,864.85 | 1,722.96 | 1,548.91 | 1,536.47 | 6,673.19 |
| 183 | 1,996.30 | 2,279.15 | 1,906 | 1,943.28 | 8,124.73 |
| 184 | 2,115.49 | 2,414.28 | 2,322.05 | 1,975.61 | 8,827.43 |
| 185 | 2,082.75 | 2,047.45 | 2,093.36 | 1,787.29 | 8,010.85 |
| 186 | 2,827.87 | 2,968.05 | 2,547.08 | 2,667.06 | 11,010.06 |
| 187 | 5,795.98 | 5,756.10 | 5,376.90 | 5,768.16 | 22,697.14 |
| 188 | 875.12 | 883.17 | 979.26 | 965.25 | 3,702.80 |
| 189 | 3,625.36 | 3,670.69 | 3,510.29 | 3,540.84 | 14,347.18 |

| | | | | | |
|---|---|---|---|---|---|
| 190 | 17,124.43 | 16,499.75 | 16,361.06 | 15,827.49 | 65,812.73 |
| 191 | 36,352.03 | 37,039.55 | 34,649.60 | 32,505.74 | 140,546.92 |
| 193 | 3,600.52 | 3,742.37 | 3,715.79 | 3,610.89 | 14,669.57 |
| 194 | 6,965.21 | 6,890.19 | 6,531.74 | 6,407.08 | 26,794.22 |
| 195 | 2,102.39 | 2,078.48 | 1,975.87 | 1,889.38 | 8,046.12 |
| 196 | 3,190.97 | 3,380.93 | 2,908.84 | 2,855.77 | 12,336.51 |
| TOTAL | 160,590.98 | 154,882.69 | 148,013.38 | 148,041.06 | 611,528.11 |

STATE: PUERTO RICO

| ZIP CODE | QUARTER 1 | QUARTER 2 | QUARTER 3 | QUARTER 4 | TOTAL GRAMS |
|---|---|---|---|---|---|
| 6 | 4,996.11 | 5,144.15 | 5,153.69 | 5,243.24 | 20,537.19 |
| 7 | 4,313.89 | 4,818.68 | 4,801.88 | 5,079.25 | 19,013.70 |
| 9 | 3,044.41 | 3,669.65 | 3,422.34 | 3,493.06 | 13,629.46 |
| TOTAL | 12,354.41 | 13,632.48 | 13,377.91 | 13,815.55 | 53,180.35 |

STATE: RHODE ISLAND

| ZIP CODE | QUARTER 1 | QUARTER 2 | QUARTER 3 | QUARTER 4 | TOTAL GRAMS |
|---|---|---|---|---|---|
| 28 | 5,483.46 | 5,342.06 | 5,202.95 | 5,091.93 | 21,120.40 |
| 29 | 3,622.96 | 3,204.76 | 3,481.71 | 2,986.89 | 13,296.32 |
| TOTAL | 9,106.42 | 8,546.82 | 8,684.66 | 8,078.82 | 34,416.72 |

STATE: SOUTH CAROLINA

| ZIP CODE | QUARTER 1 | QUARTER 2 | QUARTER 3 | QUARTER 4 | TOTAL GRAMS |
|---|---|---|---|---|---|
| 290 | 3,227.64 | 3,389.53 | 2,981 | 3,166.95 | 12,765.12 |
| 291 | 3,282.94 | 3,189.33 | 3,095.81 | 3,052.13 | 12,620.21 |
| 292 | 4,578.60 | 4,293.64 | 3,921.47 | 3,918.12 | 16,711.83 |
| 293 | 4,352.87 | 4,164 | 4,182.19 | 4,012.07 | 16,711.13 |
| 294 | 47,030.80 | 44,100.38 | 45,191.43 | 35,898.61 | 172,221.22 |
| 295 | 9,797.90 | 8,794.12 | 8,692.65 | 8,390.78 | 35,675.45 |
| 296 | 8,270.04 | 7,961.01 | 7,785.74 | 7,184.79 | 31,201.58 |

DRUG CODE:9050DRUG NAME:CODEINE
STATE:SOUTH CAROLINA

| ZIP CODE | QUARTER 1 | QUARTER 2 | QUARTER 3 | QUARTER 4 | TOTAL GRAMS |
|---|---|---|---|---|---|
| 297 | 1,957.89 | 2,255.52 | 1,719.75 | 1,714.90 | 7,648.06 |
| 298 | 2,323.27 | 2,353.75 | 2,252.23 | 2,338.40 | 9,267.65 |
| 299 | 1,406.26 | 1,141.91 | 1,191.24 | 1,173.46 | 4,912.87 |
| TOTAL | 86,228.21 | 81,643.19 | 81,013.51 | 70,850.21 | 319,735.12 |

STATE: SOUTH DAKOTA

| ZIP CODE | QUARTER 1 | QUARTER 2 | QUARTER 3 | QUARTER 4 | TOTAL GRAMS |
|---|---|---|---|---|---|
| 570 | 954.2 | 1,151.63 | 1,060.29 | 1,067.74 | 4,233.86 |
| 571 | 2,435.31 | 2,638.07 | 2,349.43 | 2,517.03 | 9,939.84 |
| 572 | 594.91 | 557.01 | 607.39 | 599.85 | 2,359.16 |
| 573 | 647.62 | 743.07 | 709.12 | 692.78 | 2,792.59 |
| 574 | 670.19 | 544.57 | 597.79 | 556.06 | 2,368.61 |
| 575 | 500.37 | 479.99 | 363.74 | 370.92 | 1,715.02 |
| 576 | 335.68 | 358.48 | 297.92 | 236.48 | 1,228.56 |
| 577 | 2,186.55 | 1,940.82 | 1,962.78 | 1,837.45 | 7,927.60 |
| TOTAL | 8,324.83 | 8,413.64 | 7,948.46 | 7,878.31 | 32,565.24 |

STATE: TENNESSEE

| ZIP CODE | QUARTER 1 | QUARTER 2 | QUARTER 3 | QUARTER 4 | TOTAL GRAMS |
|---|---|---|---|---|---|
| 370 | 5,131.02 | 4,877.39 | 5,617.56 | 4,916.38 | 20,542.35 |
| 371 | 56,745.80 | 61,350.64 | 62,179.58 | 58,476.57 | 238,752.59 |
| 372 | 2,975.65 | 2,350.93 | 2,711.97 | 2,682.48 | 10,721.03 |
| 373 | 4,152.14 | 4,154.42 | 4,004.18 | 3,888.05 | 16,198.79 |
| 374 | 1,457.94 | 1,474.20 | 1,254.43 | 1,369.25 | 5,555.82 |
| 376 | 3,234.46 | 2,973.31 | 2,846.22 | 2,833.44 | 11,887.43 |

| | | | | | |
|---|---|---|---|---|---|
| 377 | 2,819.99 | 2,573 | 2,732.86 | 2,894.40 | 11,020.25 |
| 378 | 4,427.35 | 4,105.52 | 4,658.39 | 4,166.92 | 17,358.18 |
| 379 | 2,630.73 | 2,452.97 | 2,478.77 | 2,623.53 | 10,186 |
| 380 | 2,875.02 | 2,834.35 | 2,935.83 | 2,773.05 | 11,418.25 |
| 381 | 6,275.23 | 5,454 | 5,594.06 | 5,530.90 | 22,854.19 |
| 382 | 953.91 | 1,023.34 | 1,032.85 | 972.94 | 3,983.04 |
| 383 | 4,283.76 | 3,832.31 | 3,843.36 | 4,058.85 | 16,018.28 |
| 384 | 1,239.94 | 1,272.64 | 1,266.94 | 1,231.39 | 5,010.91 |
| 385 | 2,452.13 | 2,908.86 | 2,839.78 | 2,816.40 | 11,017.17 |
| TOTAL | 101,655.07 | 103,637.88 | 105,996.78 | 101,234.55 | 412,524.28 |

STATE: TEXAS

| ZIP CODE | QUARTER 1 | QUARTER 2 | QUARTER 3 | QUARTER 4 | TOTAL GRAMS |
|---|---|---|---|---|---|
| 750 | 64,608.24 | 66,439.48 | 66,547.13 | 70,884.79 | 268,479.64 |
| 751 | 28,725.26 | 30,213.19 | 32,465.28 | 32,459.82 | 123,863.55 |
| 752 | 38,229.48 | 39,735.89 | 40,886.38 | 40,347.46 | 159,199.21 |
| 753 | 287.86 | 362.46 | 369.27 | 471.95 | 1,491.54 |
| 754 | 20,979.93 | 21,317.27 | 22,210.04 | 22,412.08 | 86,919.32 |
| 755 | 9,245.68 | 8,698.95 | 9,392.43 | 8,857.56 | 36,194.62 |
| 756 | 23,840.86 | 25,128.35 | 25,105.57 | 25,196.15 | 99,270.93 |
| 757 | 25,321.19 | 26,526.09 | 25,797.28 | 26,828.43 | 104,472.99 |
| 758 | 5,195.28 | 5,314.19 | 5,159.48 | 5,501.31 | 21,170.26 |
| 759 | 20,818.75 | 19,631.45 | 19,691.19 | 19,444.17 | 79,585.56 |
| 760 | 49,032.07 | 49,412.49 | 49,760.45 | 51,407.65 | 199,612.66 |
| 761 | 40,248.41 | 41,115.57 | 39,859.38 | 41,342.34 | 162,565.70 |
| 762 | 17,563.03 | 17,920.13 | 18,334.81 | 18,929.24 | 72,747.21 |
| 763 | 10,352.89 | 11,358.11 | 10,986.84 | 11,509.06 | 44,206.90 |
| 764 | 6,854.46 | 6,901.04 | 6,838.05 | 7,408.45 | 28,002 |
| 765 | 16,707.17 | 17,337.94 | 17,526.05 | 17,059.97 | 68,631.13 |
| 766 | 6,321.17 | 6,392.99 | 6,144.61 | 6,094.63 | 24,953.40 |
| 767 | 10,487.91 | 10,634.53 | 10,526.34 | 10,194.80 | 41,843.58 |
| 768 | 6,235.80 | 6,929.77 | 7,052.43 | 7,080.42 | 27,298.42 |
| 769 | 6,683.52 | 6,943.03 | 7,609.56 | 7,787.65 | 29,023.76 |
| 770 | 92,656.18 | 94,076.74 | 94,434 | 93,427.34 | 374,594.26 |
| 772 | 22.1 | 0 | 0 | 0.43 | 22.53 |
| 773 | 42,731.81 | 44,018.41 | 44,679.51 | 46,345.68 | 177,775.41 |
| 774 | 34,368.90 | 34,814.62 | 35,042.01 | 35,671.79 | 139,897.32 |
| 775 | 49,433.26 | 50,351.31 | 51,513.84 | 50,842.20 | 202,140.61 |
| 776 | 16,119.18 | 17,086.55 | 17,827.22 | 17,399 | 68,431.95 |
| 777 | 8,729.40 | 9,196.73 | 8,959.85 | 9,343.66 | 36,229.64 |
| 778 | 12,382.22 | 12,618.29 | 12,555.27 | 13,085.68 | 50,641.46 |
| 779 | 4,987.16 | 5,558.51 | 5,791.57 | 5,826.94 | 22,164.18 |
| 780 | 16,023.40 | 15,862.86 | 16,134.06 | 16,244.87 | 64,265.19 |
| 781 | 13,865.91 | 13,569.77 | 13,825.28 | 13,817.04 | 55,078 |

DRUG CODE:9050DRUG NAME:CODEINE
STATE:TEXAS

| ZIP CODE | QUARTER 1 | QUARTER 2 | QUARTER 3 | QUARTER 4 | TOTAL GRAMS |
|---|---|---|---|---|---|
| 782 | 77,819.43 | 77,543.49 | 79,686.09 | 77,878.30 | 312,927.31 |
| 783 | 14,142.46 | 13,990.48 | 14,447.65 | 14,441.63 | 57,022.22 |
| 784 | 18,183.47 | 18,386.87 | 18,108.38 | 17,942.37 | 72,621.09 |
| 785 | 32,294.28 | 32,333.38 | 32,806.43 | 32,348.38 | 129,782.47 |
| 786 | 23,630.13 | 23,641.80 | 23,965.75 | 23,862.07 | 95,099.75 |
| 787 | 21,759.18 | 21,168.11 | 21,448.69 | 21,389.04 | 85,765.02 |
| 788 | 5,922.06 | 5,739.78 | 5,723.24 | 5,599.13 | 22,984.21 |
| 789 | 1,920.58 | 1,765.79 | 1,841.01 | 1,833.28 | 7,360.66 |
| 790 | 8,835.38 | 8,769.88 | 8,055.31 | 8,320.88 | 33,981.45 |
| 791 | 16,384.81 | 16,662.31 | 16,708.76 | 15,543.98 | 65,299.86 |
| 792 | 1,783.91 | 1,720.97 | 1,716.29 | 1,559.81 | 6,780.98 |
| 793 | 6,343.21 | 6,492.31 | 6,363.54 | 6,314.92 | 25,513.98 |

| | | | | | |
|---|---|---|---|---|---|
| 794 | 15,137.58 | 14,964.63 | 15,116.57 | 14,569.42 | 59,788.20 |
| 795 | 3,459.01 | 3,545.22 | 3,421.23 | 3,463.37 | 13,888.83 |
| 796 | 8,937.49 | 8,829.15 | 9,171.60 | 9,604.56 | 36,542.80 |
| 797 | 20,692.52 | 21,024.38 | 21,706 | 21,720.01 | 85,142.91 |
| 798 | 1,402.31 | 1,435.46 | 1,392.36 | 1,256.36 | 5,486.49 |
| 799 | 29,258.92 | 28,193.80 | 29,421.75 | 29,935.34 | 116,809.81 |
| TOTAL | 1,006,965.21 | 1,021,674.52 | 1,034,125.83 | 1,040,805.41 | |
| 4,103,570.97 | | | | | |

STATE: UTAH

| ZIP CODE | QUARTER 1 | QUARTER 2 | QUARTER 3 | QUARTER 4 | TOTAL GRAMS |
|---|---|---|---|---|---|
| 840 | 7,670.30 | 7,279.14 | 7,582.86 | 7,178 | 29,710.30 |
| 841 | 3,764.89 | 3,680.55 | 3,893.42 | 4,071.42 | 15,410.28 |
| 843 | 825.28 | 861.36 | 887.95 | 758.68 | 3,333.27 |
| 844 | 975.83 | 1,025.18 | 1,015.05 | 1,042.18 | 4,058.24 |
| 845 | 647.92 | 633.36 | 563.76 | 613.29 | 2,458.33 |
| 846 | 2,086.82 | 2,018.62 | 2,170.28 | 2,022.76 | 8,298.48 |
| 847 | 2,166.60 | 2,003.05 | 1,685.27 | 1,803.87 | 7,658.79 |
| TOTAL | 18,137.64 | 17,501.26 | 17,798.59 | 17,490.20 | 70,927.69 |

STATE: VERMONT

| ZIP CODE | QUARTER 1 | QUARTER 2 | QUARTER 3 | QUARTER 4 | TOTAL GRAMS |
|---|---|---|---|---|---|
| 50 | 352.81 | 415.11 | 338.41 | 270.86 | 1,377.19 |
| 51 | 172.01 | 98.8 | 132.97 | 139.5 | 543.28 |
| 52 | 338.1 | 295.14 | 278.53 | 120 | 1,031.77 |
| 53 | 240.6 | 174.77 | 197.43 | 241.07 | 853.87 |
| 54 | 1,449.61 | 1,290.77 | 1,359.32 | 1,265.42 | 5,365.12 |
| 56 | 680.44 | 691.14 | 621.07 | 627.33 | 2,619.98 |
| 57 | 369.23 | 434 | 396.61 | 474.25 | 1,674.09 |
| 58 | 472.37 | 469.67 | 399.63 | 489.96 | 1,831.63 |
| TOTAL | 4,075.17 | 3,869.40 | 3,723.97 | 3,628.39 | 15,296.93 |

STATE: VIRGIN ISLANDS

| ZIP CODE | QUARTER 1 | QUARTER 2 | QUARTER 3 | QUARTER 4 | TOTAL GRAMS |
|---|---|---|---|---|---|
| 8 | 710.94 | 759.24 | 727.27 | 612.2 | 2,809.65 |
| TOTAL | 710.94 | 759.24 | 727.27 | 612.2 | 2,809.65 |

STATE: VIRGINIA

| ZIP CODE | QUARTER 1 | QUARTER 2 | QUARTER 3 | QUARTER 4 | TOTAL GRAMS |
|---|---|---|---|---|---|
| 201 | 6,692.06 | 6,270.89 | 6,122.49 | 6,426 | 25,511.44 |
| 220 | 4,470.43 | 4,215.12 | 4,366.11 | 4,175.90 | 17,227.56 |
| 221 | 3,727.25 | 3,905.25 | 3,587.05 | 3,608.25 | 14,827.80 |
| 222 | 1,539.62 | 1,416 | 1,409.40 | 1,435.21 | 5,800.23 |
| 223 | 2,322.07 | 2,329.88 | 2,152.84 | 2,143.76 | 8,948.55 |
| 224 | 2,614.48 | 2,484.85 | 2,448.86 | 2,434.76 | 9,982.95 |
| 225 | 1,117.23 | 1,187.76 | 1,013.30 | 1,106.01 | 4,424.30 |
| 226 | 1,236.15 | 1,243.34 | 1,309.73 | 1,251.09 | 5,040.31 |
| 227 | 376.26 | 361.25 | 450.11 | 323.95 | 1,511.57 |
| 228 | 1,145.18 | 1,259.13 | 1,249.59 | 1,222.31 | 4,876.21 |
| 229 | 1,595.45 | 1,414.10 | 1,499.21 | 1,409.32 | 5,918.08 |
| 230 | 1,364.46 | 1,121.98 | 1,294.86 | 1,357.43 | 5,138.73 |
| 231 | 2,413.66 | 2,380.75 | 2,204.59 | 2,180.72 | 9,179.72 |
| 232 | 5,146.24 | 4,779.98 | 5,056.29 | 4,771.79 | 19,754.30 |
| 233 | 1,583.16 | 1,549.22 | 1,592.72 | 1,391.62 | 6,116.72 |
| 234 | 4,405.79 | 4,641.92 | 4,329.18 | 3,984.65 | 17,361.54 |
| 235 | 1,600.62 | 1,830.95 | 1,560.49 | 1,661.46 | 6,653.52 |
| 236 | 3,680.60 | 3,745.04 | 3,548.97 | 3,090.28 | 14,064.89 |
| 237 | 1,081 | 1,000.34 | 1,157.09 | 837.36 | 4,075.79 |
| 238 | 3,035.40 | 2,907.17 | 2,842.04 | 2,655.15 | 11,439.76 |

RETAIL DRUG DISTRIBUTION BY ZIP CODE WITHIN STATE BY GRAMS WT

DRUG CODE:9050DRUG NAME:CODEINE
STATE:VIRGINIA

| ZIP CODE | QUARTER 1 | QUARTER 2 | QUARTER 3 | QUARTER 4 | TOTAL GRAMS |
|---|---|---|---|---|---|
| 239 | 723.08 | 731.91 | 705.56 | 682.08 | 2,842.63 |
| 240 | 1,811.06 | 1,580.08 | 1,783.44 | 1,527.82 | 6,702.40 |
| 241 | 1,503.39 | 1,206.05 | 1,311.72 | 1,132.22 | 5,153.38 |
| 242 | 1,996.19 | 1,764.67 | 1,991.93 | 1,706.66 | 7,459.45 |
| 243 | 1,644.57 | 1,648.15 | 1,623.02 | 1,619.21 | 6,534.95 |
| 244 | 680.14 | 576.77 | 762.76 | 671.79 | 2,691.46 |
| 245 | 2,154.57 | 2,303.23 | 2,025.85 | 2,150.56 | 8,634.21 |
| 246 | 1,599.01 | 1,708.41 | 1,985.85 | 1,836.89 | 7,130.16 |
| TOTAL | 63,259.12 | 61,564.19 | 61,385.05 | 58,794.25 | 245,002.61 |

STATE: WASHINGTON

| ZIP CODE | QUARTER 1 | QUARTER 2 | QUARTER 3 | QUARTER 4 | TOTAL GRAMS |
|---|---|---|---|---|---|
| 980 | 11,761.72 | 11,459.57 | 11,702.74 | 12,034.31 | 46,958.34 |
| 981 | 9,033.23 | 9,086.35 | 8,331.42 | 7,838.50 | 34,289.50 |
| 982 | 9,077.98 | 8,821.33 | 8,241.65 | 8,457.02 | 34,597.98 |
| 983 | 7,786.66 | 7,500.55 | 7,445.54 | 6,935.39 | 29,668.14 |
| 984 | 4,418.30 | 4,587.77 | 4,515.68 | 4,256.58 | 17,778.33 |
| 985 | 4,797.43 | 4,406.53 | 4,493.73 | 4,448.96 | 18,146.65 |
| 986 | 4,849.76 | 4,882.26 | 4,836.44 | 4,956.91 | 19,525.37 |
| 988 | 1,863.30 | 1,744.42 | 1,539.25 | 1,585.73 | 6,732.70 |
| 989 | 2,660.73 | 2,678.57 | 2,416.33 | 2,477.09 | 10,232.72 |
| 990 | 1,367.33 | 1,653.11 | 1,104.74 | 1,232.33 | 5,357.51 |
| 991 | 1,133.57 | 1,425.13 | 1,456.99 | 1,319.44 | 5,335.13 |
| 992 | 6,179.24 | 6,550.58 | 6,058.26 | 5,998.94 | 24,787.02 |
| 993 | 2,743.31 | 3,216.30 | 3,057.30 | 2,894.52 | 11,911.43 |
| 994 | 651.23 | 570.01 | 560.07 | 541.64 | 2,322.95 |
| TOTAL | 68,323.79 | 68,582.48 | 65,760.14 | 64,977.36 | 267,643.77 |

STATE: WEST VIRGINIA

| ZIP CODE | QUARTER 1 | QUARTER 2 | QUARTER 3 | QUARTER 4 | TOTAL GRAMS |
|---|---|---|---|---|---|
| 247 | 763.53 | 828.79 | 878.7 | 1,055.09 | 3,526.11 |
| 248 | 1,677.47 | 1,515.02 | 1,733.91 | 1,484.17 | 6,410.57 |
| 249 | 587.91 | 551.41 | 502.93 | 506.29 | 2,148.54 |
| 250 | 1,022.31 | 999.28 | 885.15 | 977.73 | 3,884.47 |
| 251 | 882.49 | 845.94 | 934.79 | 908.41 | 3,571.63 |
| 252 | 998.14 | 956.89 | 908.35 | 905.43 | 3,768.81 |
| 253 | 2,209.28 | 2,574.84 | 2,414 | 2,496.19 | 9,694.31 |
| 254 | 1,429.17 | 1,646.52 | 1,350.32 | 1,669.39 | 6,095.40 |
| 255 | 2,139.95 | 2,277.11 | 2,294.54 | 1,994.40 | 8,706 |
| 256 | 3,676.12 | 3,309.54 | 3,879.75 | 3,617.48 | 14,482.89 |
| 257 | 1,474.36 | 1,411.51 | 1,343.89 | 1,534.92 | 5,764.68 |
| 258 | 2,265.59 | 2,162.28 | 2,539.65 | 2,320.84 | 9,288.36 |
| 259 | 732.64 | 724.52 | 818.09 | 720.46 | 2,995.71 |
| 260 | 1,513.72 | 1,457.15 | 1,419.79 | 1,494.82 | 5,885.48 |
| 261 | 1,788.86 | 1,929.18 | 1,813.09 | 1,744.38 | 7,275.51 |
| 262 | 1,783.17 | 1,630.50 | 1,733.63 | 1,801.22 | 6,948.52 |
| 263 | 1,365.66 | 1,331.52 | 1,307.97 | 1,422.66 | 5,427.81 |
| 264 | 449.85 | 470.93 | 507.31 | 508.51 | 1,936.60 |
| 265 | 2,779.78 | 2,535.66 | 2,566.10 | 2,621.36 | 10,502.90 |
| 266 | 882.93 | 1,012.23 | 1,034.33 | 934.8 | 3,864.29 |
| 267 | 491.72 | 555.2 | 500.93 | 607.01 | 2,154.86 |
| 268 | 352.29 | 432.3 | 406.97 | 345.43 | 1,536.99 |
| TOTAL | 31,266.94 | 31,158.32 | 31,774.19 | 31,670.99 | 125,870.44 |

STATE: WISCONSIN

| ZIP CODE | QUARTER 1 | QUARTER 2 | QUARTER 3 | QUARTER 4 | TOTAL GRAMS |
|---|---|---|---|---|---|
| 530 | 6,019.84 | 5,669.79 | 5,413.18 | 5,656.12 | 22,758.93 |
| 531 | 6,149.98 | 6,249.99 | 6,369.70 | 6,200.20 | 24,969.87 |

| | | | | | |
|---|---|---|---|---|---|
| 532 | 11,279.63 | 10,806.50 | 10,414.81 | 10,483.35 | 42,984.29 |
| 534 | 1,815.74 | 1,874.40 | 1,737.82 | 1,796.05 | 7,224.01 |
| 535 | 4,946.26 | 4,686 | 4,617.34 | 4,408.78 | 18,658.38 |
| 537 | 2,619.45 | 2,704.95 | 2,353.14 | 2,405.69 | 10,083.23 |
| 538 | 649.75 | 497.5 | 444.09 | 546.58 | 2,137.92 |
| 539 | 2,481.32 | 2,376.64 | 2,275.25 | 2,136.79 | 9,270 |
| 540 | 1,129.34 | 1,051.33 | 926.55 | 924.85 | 4,032.07 |
| 541 | 1,798.05 | 1,504.69 | 1,707.62 | 1,640.99 | 6,651.35 |
| 542 | 1,241.09 | 1,232.85 | 1,231.12 | 1,088.38 | 4,793.44 |
| 543 | 2,400.49 | 2,250.61 | 2,221.66 | 2,264.22 | 9,136.98 |
| 544 | 3,785.45 | 3,694.91 | 3,825.88 | 3,365.55 | 14,671.79 |

DRUG CODE:9050DRUG NAME:CODEINE
STATE:WISCONSIN

| ZIP CODE | QUARTER 1 | QUARTER 2 | QUARTER 3 | QUARTER 4 | TOTAL GRAMS |
|---|---|---|---|---|---|
| 545 | 1,166.05 | 1,315.70 | 1,065.56 | 1,134.68 | 4,681.99 |
| 546 | 1,508.75 | 1,399.84 | 1,607.38 | 1,356.72 | 5,872.69 |
| 547 | 2,009.38 | 2,058.38 | 1,988.39 | 1,944.56 | 8,000.71 |
| 548 | 1,783.98 | 1,964.29 | 2,090.87 | 1,842.67 | 7,681.81 |
| 549 | 4,963.88 | 4,428.38 | 4,509.07 | 4,190.69 | 18,092.02 |
| TOTAL | 57,748.43 | 55,766.75 | 54,799.43 | 53,386.87 | 221,701.48 |

STATE: WYOMING

| ZIP CODE | QUARTER 1 | QUARTER 2 | QUARTER 3 | QUARTER 4 | TOTAL GRAMS |
|---|---|---|---|---|---|
| 820 | 1,033.39 | 1,143.95 | 852.19 | 1,060.76 | 4,090.29 |
| 822 | 185.65 | 205.54 | 148.08 | 164.65 | 703.92 |
| 823 | 129.51 | 114.93 | 83.98 | 61.88 | 390.3 |
| 824 | 604.24 | 679.57 | 688.23 | 733.44 | 2,705.48 |
| 825 | 617.35 | 526.19 | 534.84 | 459.42 | 2,137.80 |
| 826 | 671.24 | 502.7 | 642.22 | 608.87 | 2,425.03 |
| 827 | 483.08 | 408.87 | 361.89 | 379.93 | 1,633.77 |
| 828 | 531.18 | 457.26 | 477.38 | 284.68 | 1,750.50 |
| 829 | 548.1 | 547 | 590.1 | 554.74 | 2,239.94 |
| 830 | 86.19 | 110.51 | 78.9 | 143.66 | 419.26 |
| 831 | 143.66 | 61.88 | 132.61 | 75.14 | 413.29 |
| TOTAL | 5,033.59 | 4,758.40 | 4,590.42 | 4,527.17 | 18,909.58 |

DRUG CODE: 9064 DRUG NAME: BUPRENORPHINE
STATE: ALABAMA

| ZIP CODE | QUARTER 1 | QUARTER 2 | QUARTER 3 | QUARTER 4 | TOTAL GRAMS |
|---|---|---|---|---|---|
| 350 | 3,518.48 | 3,564.91 | 3,573.56 | 3,509.48 | 14,166.43 |
| 351 | 2,070.09 | 2,052.86 | 1,989.24 | 2,100.70 | 8,212.89 |
| 352 | 2,878.57 | 2,865.89 | 3,014.55 | 2,991.91 | 11,750.92 |
| 354 | 331.16 | 342.2 | 360.73 | 353.29 | 1,387.38 |
| 355 | 1,339.86 | 1,396.88 | 1,522.90 | 1,525.91 | 5,785.55 |
| 356 | 2,925.19 | 2,505.04 | 2,521.41 | 2,594.75 | 10,546.39 |
| 357 | 641.6 | 655.2 | 730.5 | 802.14 | 2,829.44 |
| 358 | 1,031.22 | 1,062.38 | 1,115.12 | 1,144.29 | 4,353.01 |
| 359 | 1,952.46 | 1,799.42 | 2,011.34 | 2,239.43 | 8,002.65 |
| 360 | 512.3 | 525.19 | 481.84 | 452.33 | 1,971.66 |
| 361 | 298.07 | 298.8 | 298.55 | 324.44 | 1,219.86 |
| 362 | 766.87 | 790.91 | 823.89 | 829.81 | 3,211.48 |
| 363 | 742.07 | 725.21 | 664.67 | 661.94 | 2,793.89 |
| 364 | 328.48 | 353.89 | 359.72 | 360.79 | 1,402.88 |
| 365 | 1,479.12 | 1,563.50 | 1,600.63 | 1,625.77 | 6,269.02 |
| 366 | 982.48 | 1,015.18 | 965.8 | 999.2 | 3,962.66 |
| 367 | 168.78 | 177.18 | 178.62 | 186.09 | 710.67 |
| 368 | 516.6 | 537.63 | 527.78 | 532.56 | 2,114.57 |
| 369 | 35.6 | 33.96 | 32.23 | 37.38 | 139.17 |

RETAIL DRUG DISTRIBUTION BY ZIP CODE WITHIN STATE BY GRAMS WT

| | | | | | |
|---|---|---|---|---|---|
| TOTAL | 22,519 | 22,266.23 | 22,773.08 | 23,272.21 | 90,830.52 |

STATE: ALASKA

| ZIP CODE | QUARTER 1 | QUARTER 2 | QUARTER 3 | QUARTER 4 | TOTAL GRAMS |
|---|---|---|---|---|---|
| 995 | 858.73 | 844.97 | 906.24 | 953.54 | 3,563.48 |
| 996 | 494.22 | 530.32 | 537.86 | 576.7 | 2,139.10 |
| 997 | 151.11 | 161.13 | 164.94 | 153.43 | 630.61 |
| 998 | 199.51 | 206.87 | 217.2 | 201.61 | 825.19 |
| 999 | 145.5 | 137.16 | 117.3 | 132.56 | 532.52 |
| TOTAL | 1,849.07 | 1,880.45 | 1,943.54 | 2,017.84 | 7,690.90 |

STATE: AMERICAN SAMOA

| ZIP CODE | QUARTER 1 | QUARTER 2 | QUARTER 3 | QUARTER 4 | TOTAL GRAMS |
|---|---|---|---|---|---|
| 967 | 0 | 0.01 | 0 | 0 | 0.01 |
| TOTAL | 0 | 0.01 | 0 | 0 | 0.01 |

STATE: ARIZONA

| ZIP CODE | QUARTER 1 | QUARTER 2 | QUARTER 3 | QUARTER 4 | TOTAL GRAMS |
|---|---|---|---|---|---|
| 850 | 1,823.24 | 1,900.91 | 1,970.02 | 2,004.98 | 7,699.15 |
| 851 | 398.26 | 441.56 | 485.95 | 519.79 | 1,845.56 |
| 852 | 2,472.32 | 2,498.53 | 2,562.42 | 2,552.74 | 10,086.01 |
| 853 | 1,190.68 | 1,259.97 | 1,289.12 | 1,329.35 | 5,069.12 |
| 855 | 80.18 | 95.12 | 79.14 | 85.53 | 339.97 |
| 856 | 79.44 | 93.64 | 93.32 | 85.17 | 351.57 |
| 857 | 968.54 | 1,063.67 | 1,130.69 | 1,183.96 | 4,346.86 |
| 859 | 97.37 | 104.43 | 118.59 | 115.13 | 435.52 |
| 860 | 116.04 | 102.71 | 119.8 | 116.4 | 454.95 |

DRUG CODE:9064DRUG NAME:BUPRENORPHINE
STATE:ARIZONA

| ZIP CODE | QUARTER 1 | QUARTER 2 | QUARTER 3 | QUARTER 4 | TOTAL GRAMS |
|---|---|---|---|---|---|
| 863 | 422.11 | 442.04 | 486.88 | 528.12 | 1,879.15 |
| 864 | 148.79 | 147.51 | 138.29 | 155.15 | 589.74 |
| 865 | 8.28 | 5.4 | 8.64 | 7.74 | 30.06 |
| TOTAL | 7,805.25 | 8,155.49 | 8,482.86 | 8,684.06 | 33,127.66 |

STATE: ARKANSAS

| ZIP CODE | QUARTER 1 | QUARTER 2 | QUARTER 3 | QUARTER 4 | TOTAL GRAMS |
|---|---|---|---|---|---|
| 716 | 86.84 | 87.41 | 85.31 | 89.12 | 348.68 |
| 717 | 111.43 | 106.68 | 103.47 | 118.69 | 440.27 |
| 718 | 149.12 | 149.89 | 162.56 | 168.45 | 630.02 |
| 719 | 748.29 | 743.15 | 769.37 | 827.58 | 3,088.39 |
| 720 | 578.38 | 578.54 | 551.61 | 566.7 | 2,275.23 |
| 721 | 943.06 | 981.22 | 962.48 | 975.41 | 3,862.17 |
| 722 | 501.72 | 527.67 | 520.51 | 525.66 | 2,075.56 |
| 723 | 131.78 | 137.3 | 139.99 | 150.09 | 559.16 |
| 724 | 490.61 | 508.07 | 496.51 | 586.34 | 2,081.53 |
| 725 | 126.06 | 135.39 | 126.8 | 145.16 | 533.41 |
| 726 | 166.5 | 178.48 | 170.86 | 198.09 | 713.93 |
| 727 | 378.07 | 440.07 | 516.49 | 620.55 | 1,955.18 |
| 728 | 114.74 | 106.56 | 115.03 | 106.12 | 442.45 |
| 729 | 218.13 | 262.05 | 279.99 | 293.03 | 1,053.20 |
| TOTAL | 4,744.73 | 4,942.48 | 5,000.98 | 5,370.99 | 20,059.18 |

STATE: CALIFORNIA

| ZIP CODE | QUARTER 1 | QUARTER 2 | QUARTER 3 | QUARTER 4 | TOTAL GRAMS |
|---|---|---|---|---|---|
| 900 | 733.61 | 785.32 | 783.31 | 862.81 | 3,165.05 |
| 902 | 840.17 | 803.23 | 864.05 | 903.52 | 3,410.97 |

| | | | | | |
|---|---|---|---|---|---|
| 903 | 30.6 | 19.58 | 24.6 | 21.76 | 96.54 |
| 904 | 169.05 | 206.19 | 192.04 | 197.68 | 764.96 |
| 905 | 173.05 | 183.94 | 185.91 | 175.95 | 718.85 |
| 906 | 312.46 | 326.98 | 284.53 | 244.13 | 1,168.10 |
| 907 | 611.81 | 610.96 | 635.02 | 597.8 | 2,455.59 |
| 908 | 320.8 | 330.62 | 342.75 | 336.49 | 1,330.66 |
| 910 | 179.09 | 124.91 | 117.33 | 114.66 | 535.99 |
| 911 | 122.74 | 141.53 | 131.19 | 154.25 | 549.71 |
| 912 | 137.96 | 184.36 | 202.94 | 224.41 | 749.67 |
| 913 | 1,109.27 | 1,133 | 1,102.23 | 1,155.03 | 4,499.53 |
| 914 | 411.35 | 413.39 | 426.06 | 435.21 | 1,686.01 |
| 915 | 169.38 | 174.73 | 177.18 | 188.85 | 710.14 |
| 916 | 133.65 | 140.95 | 126.45 | 102.79 | 503.84 |
| 917 | 790.05 | 823.05 | 848.78 | 819.02 | 3,280.90 |
| 918 | 7.73 | 8.86 | 11.37 | 8.18 | 36.14 |
| 919 | 227.67 | 220.15 | 225.69 | 221.28 | 894.79 |
| 920 | 1,321.07 | 1,273.76 | 1,350.91 | 1,304.28 | 5,250.02 |
| 921 | 1,059.96 | 1,153.88 | 1,168.21 | 1,168.43 | 4,550.48 |
| 922 | 268.25 | 257.97 | 254.32 | 323.85 | 1,104.39 |
| 923 | 344.48 | 370.04 | 339.21 | 382.51 | 1,436.24 |
| 924 | 72.28 | 62.02 | 57.43 | 62.07 | 253.8 |
| 925 | 1,165.01 | 1,288.82 | 1,290.11 | 1,305.93 | 5,049.87 |
| 926 | 1,634.60 | 1,701.23 | 1,756.08 | 1,808.80 | 6,900.71 |
| 927 | 301.75 | 312.51 | 306.86 | 312.02 | 1,233.14 |
| 928 | 638.45 | 699.64 | 702.97 | 694.07 | 2,735.13 |
| 930 | 631.51 | 624.16 | 635.28 | 630.69 | 2,521.64 |
| 931 | 424.08 | 431.84 | 442.96 | 426.29 | 1,725.17 |
| 932 | 267.65 | 266.08 | 284.23 | 306.03 | 1,123.99 |
| 933 | 293.71 | 299.96 | 308.16 | 314.88 | 1,216.71 |
| 934 | 755.82 | 749.63 | 785.2 | 789.3 | 3,079.95 |
| 935 | 341.24 | 385.35 | 377.25 | 399.18 | 1,503.02 |
| 936 | 196.55 | 194.2 | 204.07 | 196.46 | 791.28 |
| 937 | 232.3 | 240.38 | 268.65 | 281.04 | 1,022.37 |
| 939 | 562.15 | 586.29 | 610 | 628.82 | 2,387.26 |
| 940 | 634.69 | 651.23 | 688.26 | 657.51 | 2,631.69 |
| 941 | 951.37 | 1,145.23 | 916.72 | 1,055.93 | 4,069.25 |
| 943 | 76.38 | 95.59 | 90.82 | 88.04 | 350.83 |
| 944 | 101.71 | 102.19 | 117.96 | 121.39 | 443.25 |
| 945 | 2,379.69 | 2,434.76 | 2,457.60 | 2,466.99 | 9,739.04 |
| 946 | 299.58 | 290.53 | 313.14 | 315.99 | 1,219.24 |
| 947 | 103.17 | 106.15 | 96.79 | 85.07 | 391.18 |

DRUG CODE:9064DRUG NAME:BUPRENORPHINE
STATE:CALIFORNIA

| ZIP CODE | QUARTER 1 | QUARTER 2 | QUARTER 3 | QUARTER 4 | TOTAL GRAMS |
|---|---|---|---|---|---|
| 948 | 75.37 | 84.3 | 95.49 | 96.09 | 351.25 |
| 949 | 655.52 | 707.67 | 748.06 | 775.77 | 2,887.02 |
| 950 | 835.94 | 837.02 | 842.39 | 902.56 | 3,417.91 |
| 951 | 546.21 | 639.97 | 622.59 | 630.7 | 2,439.47 |
| 952 | 309.92 | 330.58 | 331.17 | 310.82 | 1,282.49 |
| 953 | 917.1 | 986.16 | 937.52 | 971.5 | 3,812.28 |
| 954 | 1,658.76 | 1,760.04 | 1,855.97 | 1,928.85 | 7,203.62 |
| 955 | 894.97 | 1,031.42 | 1,033.86 | 1,116.19 | 4,076.44 |
| 956 | 1,428.04 | 1,451.70 | 1,579.80 | 1,685.04 | 6,144.58 |
| 957 | 248.2 | 265.45 | 277.69 | 306.48 | 1,097.82 |
| 958 | 1,815.01 | 1,878.35 | 1,961.33 | 2,016.78 | 7,671.47 |
| 959 | 1,055.25 | 1,131.07 | 1,249.64 | 1,258.74 | 4,694.70 |
| 960 | 671.22 | 716.02 | 695.87 | 761.63 | 2,844.74 |
| 961 | 84.99 | 89.31 | 96.19 | 103.62 | 374.11 |
| TOTAL | 32,734.39 | 34,264.25 | 34,832.19 | 35,754.16 | 137,584.99 |

RETAIL DRUG DISTRIBUTION BY ZIP CODE WITHIN STATE BY GRAMS WT

STATE: COLORADO

| ZIP CODE | QUARTER 1 | QUARTER 2 | QUARTER 3 | QUARTER 4 | TOTAL GRAMS |
|---|---|---|---|---|---|
| 800 | 1,043.41 | 1,112.30 | 1,099.30 | 1,170.45 | 4,425.46 |
| 801 | 724.2 | 731.27 | 781.24 | 777.19 | 3,013.90 |
| 802 | 1,445.45 | 1,535.93 | 1,515.35 | 1,502.82 | 5,999.55 |
| 803 | 123.49 | 126.77 | 121.18 | 119.81 | 491.25 |
| 804 | 134.64 | 132.77 | 154.29 | 151.36 | 573.06 |
| 805 | 759.69 | 789.53 | 837.79 | 903.07 | 3,290.08 |
| 806 | 349.43 | 328.92 | 339.53 | 361.27 | 1,379.15 |
| 807 | 54.53 | 43.6 | 46.45 | 55.53 | 200.11 |
| 808 | 64.8 | 66.02 | 74.14 | 72.35 | 277.31 |
| 809 | 634.87 | 659.91 | 658.5 | 635.73 | 2,589.01 |
| 810 | 437.6 | 474.68 | 457.13 | 475.25 | 1,844.66 |
| 811 | 87.88 | 97.4 | 87.91 | 83 | 356.19 |
| 812 | 107.24 | 128.52 | 108.44 | 101.55 | 445.75 |
| 813 | 126.13 | 128.42 | 158.44 | 113 | 525.99 |
| 814 | 82.27 | 76.84 | 75.38 | 62.26 | 296.75 |
| 815 | 179.78 | 172.86 | 173.04 | 217.66 | 743.34 |
| 816 | 117.61 | 111.47 | 118.61 | 137.82 | 485.51 |
| TOTAL | 6,473.02 | 6,717.21 | 6,806.72 | 6,940.12 | 26,937.07 |

STATE: CONNECTICUT

| ZIP CODE | QUARTER 1 | QUARTER 2 | QUARTER 3 | QUARTER 4 | TOTAL GRAMS |
|---|---|---|---|---|---|
| 60 | 2,683.91 | 2,783.68 | 2,789.79 | 2,928.79 | 11,186.17 |
| 61 | 1,054.99 | 1,110.93 | 1,069.20 | 1,173.92 | 4,409.04 |
| 62 | 641.47 | 697.47 | 710.29 | 726.57 | 2,775.80 |
| 63 | 1,099.66 | 1,209.16 | 1,264.84 | 1,250.85 | 4,824.51 |
| 64 | 3,007.54 | 3,078.90 | 3,142.29 | 3,295.51 | 12,524.24 |
| 65 | 2,162.20 | 2,209.84 | 2,255.44 | 2,238.13 | 8,865.61 |
| 66 | 299.58 | 316.36 | 313.97 | 295.01 | 1,224.92 |
| 67 | 1,291.55 | 1,365.11 | 1,427.74 | 1,382.58 | 5,466.98 |
| 68 | 660.71 | 690.44 | 645.64 | 643.76 | 2,640.55 |
| 69 | 186.14 | 190.15 | 176.51 | 190.82 | 743.62 |
| TOTAL | 13,087.75 | 13,652.04 | 13,795.71 | 14,125.94 | 54,661.44 |

STATE: DELAWARE

| ZIP CODE | QUARTER 1 | QUARTER 2 | QUARTER 3 | QUARTER 4 | TOTAL GRAMS |
|---|---|---|---|---|---|
| 197 | 979.88 | 962.89 | 979.87 | 973.36 | 3,896 |
| 198 | 748.56 | 796.37 | 748.78 | 832.78 | 3,126.49 |
| 199 | 1,056.93 | 1,141.82 | 1,270.52 | 1,174.79 | 4,644.06 |
| TOTAL | 2,785.37 | 2,901.08 | 2,999.17 | 2,980.93 | 11,666.55 |

STATE: DISTRICT OF COLUMBIA

| ZIP CODE | QUARTER 1 | QUARTER 2 | QUARTER 3 | QUARTER 4 | TOTAL GRAMS |
|---|---|---|---|---|---|
| 200 | 1,628.23 | 1,790.78 | 1,909.19 | 1,992.54 | 7,320.74 |
| 204 | 10.94 | 9.98 | 18.72 | 21.8 | 61.44 |
| TOTAL | 1,639.17 | 1,800.76 | 1,927.91 | 2,014.34 | 7,382.18 |

STATE: FLORIDA

| ZIP CODE | QUARTER 1 | QUARTER 2 | QUARTER 3 | QUARTER 4 | TOTAL GRAMS |
|---|---|---|---|---|---|
| 320 | 2,548.78 | 2,682.43 | 2,701.08 | 2,841.42 | 10,773.71 |
| 321 | 2,040.15 | 2,098.79 | 2,111.51 | 2,093.87 | 8,344.32 |
| 322 | 3,276.43 | 3,411.68 | 3,477.64 | 3,619.44 | 13,785.19 |
| 323 | 567.32 | 584.88 | 595.58 | 638.98 | 2,386.76 |
| 324 | 708.58 | 758.77 | 720.04 | 762.22 | 2,949.61 |

DRUG CODE:9064DRUG NAME:BUPRENORPHINE
STATE:FLORIDA

| ZIP CODE | QUARTER 1 | QUARTER 2 | QUARTER 3 | QUARTER 4 | TOTAL GRAMS |
|---|---|---|---|---|---|

| | | | | | |
|---|---|---|---|---|---|
| 325 | 1,694.63 | 1,709.64 | 1,707.50 | 1,799.69 | 6,911.46 |
| 326 | 1,909.78 | 2,095.88 | 2,164.69 | 2,200.81 | 8,371.16 |
| 327 | 2,169.74 | 2,290.98 | 2,299.25 | 2,481.32 | 9,241.29 |
| 328 | 1,037.23 | 1,043.42 | 1,067.65 | 1,063.64 | 4,211.94 |
| 329 | 2,562.39 | 2,542.41 | 2,610.30 | 2,677.88 | 10,392.98 |
| 330 | 1,425.57 | 1,470.73 | 1,462.32 | 1,440.95 | 5,799.57 |
| 331 | 763.54 | 785.77 | 789.86 | 832.51 | 3,171.68 |
| 333 | 1,088.43 | 1,155.60 | 1,113.30 | 1,062.80 | 4,420.13 |
| 334 | 3,568.69 | 3,639.05 | 3,573.11 | 3,670.39 | 14,451.24 |
| 335 | 1,255.21 | 1,278.36 | 1,302.40 | 1,376.48 | 5,212.45 |
| 336 | 1,237.41 | 1,197.16 | 1,209.23 | 1,235.50 | 4,879.30 |
| 337 | 2,070.51 | 2,279.70 | 2,170.34 | 2,293.30 | 8,813.85 |
| 338 | 694.03 | 742.92 | 753.99 | 761.32 | 2,952.26 |
| 339 | 2,193.53 | 2,150.51 | 2,214.21 | 2,335.07 | 8,893.32 |
| 341 | 882.46 | 852.23 | 855.67 | 896.28 | 3,486.64 |
| 342 | 3,125.77 | 3,224.40 | 3,067.09 | 3,231.47 | 12,648.73 |
| 344 | 1,091.21 | 1,154.97 | 1,178.28 | 1,259.03 | 4,683.49 |
| 346 | 1,690.19 | 1,730.76 | 1,721.53 | 1,753.68 | 6,896.16 |
| 347 | 913.69 | 950.44 | 972.54 | 1,002.77 | 3,839.44 |
| 349 | 1,214.88 | 1,253.88 | 1,201.92 | 1,249.04 | 4,919.72 |
| TOTAL | 41,730.15 | 43,085.36 | 43,041.03 | 44,579.86 | 172,436.40 |

STATE: GEORGIA

| ZIP CODE | QUARTER 1 | QUARTER 2 | QUARTER 3 | QUARTER 4 | TOTAL GRAMS |
|---|---|---|---|---|---|
| 300 | 2,289.96 | 2,349.68 | 2,370.61 | 2,414.40 | 9,424.65 |
| 301 | 1,914.84 | 1,952.45 | 1,947.27 | 2,006.15 | 7,820.71 |
| 302 | 1,222.11 | 1,239.37 | 1,300.02 | 1,298.35 | 5,059.85 |
| 303 | 698.28 | 673.81 | 673.16 | 674.57 | 2,719.82 |
| 304 | 799.59 | 770.76 | 783.97 | 771.19 | 3,125.51 |
| 305 | 790.95 | 807.79 | 815.33 | 832.95 | 3,247.02 |
| 306 | 663.19 | 692.85 | 720.03 | 716.64 | 2,792.71 |
| 307 | 390.41 | 420.4 | 456.15 | 496.74 | 1,763.70 |
| 308 | 108.18 | 123.48 | 111.28 | 120.11 | 463.05 |
| 309 | 250.97 | 244.24 | 247.65 | 257.1 | 999.96 |
| 310 | 933.01 | 955.37 | 962.74 | 990.67 | 3,841.79 |
| 312 | 396.29 | 405.9 | 397.97 | 380.92 | 1,581.08 |
| 313 | 670.89 | 673.17 | 645.86 | 706.07 | 2,695.99 |
| 314 | 829.23 | 840.52 | 784.57 | 786.93 | 3,241.25 |
| 315 | 1,412.41 | 1,425.12 | 1,472.38 | 1,478.70 | 5,788.61 |
| 316 | 122.8 | 127.43 | 129.03 | 134.6 | 513.86 |
| 317 | 265.09 | 289.16 | 302.05 | 315.26 | 1,171.56 |
| 318 | 12.16 | 10.53 | 11.67 | 14.91 | 49.27 |
| 319 | 236.53 | 258.66 | 253.28 | 266.59 | 1,015.06 |
| 398 | 60.65 | 55.61 | 51.45 | 53.75 | 221.46 |
| TOTAL | 14,067.54 | 14,316.30 | 14,436.47 | 14,716.60 | 57,536.91 |

STATE: GUAM

| ZIP CODE | QUARTER 1 | QUARTER 2 | QUARTER 3 | QUARTER 4 | TOTAL GRAMS |
|---|---|---|---|---|---|
| 969 | 3.56 | 0.49 | 0.49 | 0.02 | 4.56 |
| TOTAL | 3.56 | 0.49 | 0.49 | 0.02 | 4.56 |

STATE: HAWAII

| ZIP CODE | QUARTER 1 | QUARTER 2 | QUARTER 3 | QUARTER 4 | TOTAL GRAMS |
|---|---|---|---|---|---|
| 967 | 1,323 | 1,350.35 | 1,436.70 | 1,368.13 | 5,478.18 |
| 968 | 386.18 | 443.92 | 442.96 | 458.37 | 1,731.43 |
| TOTAL | 1,709.18 | 1,794.27 | 1,879.66 | 1,826.50 | 7,209.61 |

STATE: IDAHO

| ZIP CODE | QUARTER 1 | QUARTER 2 | QUARTER 3 | QUARTER 4 | TOTAL GRAMS |
|---|---|---|---|---|---|
| 832 | 192.37 | 214.91 | 250.1 | 277.52 | 934.9 |
| 833 | 152.58 | 166.7 | 178.24 | 186.52 | 684.04 |

| | | | | | |
|---|---|---|---|---|---|
| 834 | 179.18 | 193.76 | 203.34 | 208.85 | 785.13 |
| 835 | 218.95 | 232.48 | 233.77 | 255.78 | 940.98 |
| 836 | 444.55 | 474.29 | 448.67 | 429.87 | 1,797.38 |
| 837 | 421.81 | 459.06 | 502.66 | 521.24 | 1,904.77 |
| 838 | 1,225.20 | 1,286.86 | 1,306.94 | 1,279.25 | 5,098.25 |
| TOTAL | 2,834.64 | 3,028.06 | 3,123.72 | 3,159.03 | 12,145.45 |

STATE: ILLINOIS

| ZIP CODE | QUARTER 1 | QUARTER 2 | QUARTER 3 | QUARTER 4 | TOTAL GRAMS |
|---|---|---|---|---|---|
| 600 | 1,753.73 | 1,900.75 | 1,851.35 | 1,974.99 | 7,480.82 |

DRUG CODE:9064DRUG NAME:BUPRENORPHINE
STATE:ILLINOIS

| ZIP CODE | QUARTER 1 | QUARTER 2 | QUARTER 3 | QUARTER 4 | TOTAL GRAMS |
|---|---|---|---|---|---|
| 601 | 981.95 | 1,032.66 | 1,001.34 | 1,050.93 | 4,066.88 |
| 602 | 35.24 | 30.54 | 37.64 | 31.75 | 135.17 |
| 603 | 29.28 | 24.5 | 50.9 | 93.02 | 197.7 |
| 604 | 1,578.90 | 1,704 | 1,684.39 | 1,766.65 | 6,733.94 |
| 605 | 751.03 | 802.88 | 813.7 | 851.45 | 3,219.06 |
| 606 | 1,343.02 | 1,452.76 | 1,525.73 | 1,584.07 | 5,905.58 |
| 607 | 97.46 | 97.39 | 110.72 | 101.17 | 406.74 |
| 608 | 66.15 | 70.63 | 59.87 | 66.91 | 263.56 |
| 609 | 111.43 | 129.08 | 133.58 | 150.06 | 524.15 |
| 610 | 142.6 | 153.15 | 174.45 | 162.19 | 632.39 |
| 611 | 440.76 | 462.45 | 485.27 | 485 | 1,873.48 |
| 612 | 142.42 | 165.36 | 152.16 | 162.05 | 621.99 |
| 613 | 205.01 | 216.62 | 228.05 | 233.64 | 883.32 |
| 614 | 36.79 | 37.78 | 38.35 | 39.84 | 152.76 |
| 615 | 131.63 | 141.94 | 163.95 | 160.54 | 598.06 |
| 616 | 271.47 | 288.74 | 285.99 | 283.33 | 1,129.53 |
| 617 | 93.85 | 94.63 | 95.7 | 101.47 | 385.65 |
| 618 | 569.81 | 641.93 | 653.34 | 712.38 | 2,577.46 |
| 619 | 59.22 | 73.88 | 76.81 | 76.11 | 286.02 |
| 620 | 247.59 | 263.99 | 285.1 | 304.52 | 1,101.20 |
| 622 | 333.52 | 301.07 | 257.35 | 269.7 | 1,161.64 |
| 623 | 30.18 | 36.3 | 45.8 | 57.74 | 170.02 |
| 624 | 90.22 | 102.14 | 101.67 | 120.48 | 414.51 |
| 625 | 243.98 | 252.72 | 259.99 | 258.19 | 1,014.88 |
| 626 | 116.67 | 123.05 | 144.72 | 142.27 | 526.71 |
| 627 | 225.34 | 246.85 | 260 | 256.61 | 988.8 |
| 628 | 223.47 | 243.51 | 246.93 | 293.5 | 1,007.41 |
| 629 | 716.36 | 776.02 | 788.75 | 871.98 | 3,153.11 |
| TOTAL | 11,069.08 | 11,867.32 | 12,013.60 | 12,662.54 | 47,612.54 |

STATE: INDIANA

| ZIP CODE | QUARTER 1 | QUARTER 2 | QUARTER 3 | QUARTER 4 | TOTAL GRAMS |
|---|---|---|---|---|---|
| 460 | 1,694.32 | 1,658.74 | 1,606.21 | 1,685.60 | 6,644.87 |
| 461 | 1,749.12 | 1,793.38 | 1,918.12 | 1,954.06 | 7,414.68 |
| 462 | 2,811.53 | 2,922.09 | 2,979.32 | 3,025.07 | 11,738.01 |
| 463 | 1,786.99 | 1,873.90 | 1,883.52 | 1,999.56 | 7,543.97 |
| 464 | 150.29 | 190.2 | 261.2 | 152.95 | 754.64 |
| 465 | 354.23 | 379.48 | 384.96 | 418.5 | 1,537.17 |
| 466 | 121.83 | 121.89 | 147.9 | 141.84 | 533.46 |
| 467 | 92.28 | 101.22 | 108.02 | 108.58 | 410.1 |
| 468 | 209.14 | 222.12 | 216.8 | 242.27 | 890.33 |
| 469 | 559.78 | 562.9 | 566.35 | 592.43 | 2,281.46 |
| 470 | 540.84 | 531 | 535.24 | 609.44 | 2,216.52 |
| 471 | 1,289.43 | 1,451.29 | 1,407.85 | 1,477.50 | 5,626.07 |
| 472 | 779.81 | 798.74 | 787.72 | 885.53 | 3,251.80 |

| | | | | | |
|---|---|---|---|---|---|
| 473 | 1,372.56 | 1,411.33 | 1,470.76 | 1,485.97 | 5,740.62 |
| 474 | 642.15 | 605.63 | 638.07 | 678.75 | 2,564.60 |
| 475 | 177.66 | 182.75 | 178.37 | 180.69 | 719.47 |
| 476 | 29.37 | 34.32 | 36.63 | 50.05 | 150.37 |
| 477 | 262.25 | 283.34 | 259.95 | 327.67 | 1,133.21 |
| 478 | 252.63 | 281.77 | 297.37 | 348.12 | 1,179.89 |
| 479 | 368.04 | 309.71 | 326.18 | 353.63 | 1,357.56 |
| TOTAL | 15,244.25 | 15,715.80 | 16,010.54 | 16,718.21 | 63,688.80 |

STATE: IOWA

| ZIP CODE | QUARTER 1 | QUARTER 2 | QUARTER 3 | QUARTER 4 | TOTAL GRAMS |
|---|---|---|---|---|---|
| 500 | 44.03 | 49.05 | 59.9 | 63.74 | 216.72 |
| 501 | 31.65 | 41.34 | 64.34 | 52.7 | 190.03 |
| 502 | 30.41 | 31.77 | 39.19 | 34.45 | 135.82 |
| 503 | 254.2 | 277.42 | 303.65 | 268.75 | 1,104.02 |
| 504 | 130.03 | 139.03 | 141.4 | 136.21 | 546.67 |
| 505 | 34.57 | 38.93 | 39.65 | 38.31 | 151.46 |
| 506 | 15.07 | 15.76 | 13.73 | 19.21 | 63.77 |
| 507 | 26.81 | 29.78 | 24.55 | 29.37 | 110.51 |
| 508 | 6.73 | 9.11 | 9.45 | 6.13 | 31.42 |
| 510 | 91.18 | 106.39 | 94.18 | 75.4 | 367.15 |
| 511 | 73.48 | 83.5 | 91.27 | 95.94 | 344.19 |
| 512 | 13.68 | 20.92 | 22.27 | 23.81 | 80.68 |

DRUG CODE:9064DRUG NAME:BUPRENORPHINE
STATE:IOWA

| ZIP CODE | QUARTER 1 | QUARTER 2 | QUARTER 3 | QUARTER 4 | TOTAL GRAMS |
|---|---|---|---|---|---|
| 513 | 12.4 | 13.81 | 12.1 | 12.78 | 51.09 |
| 514 | 6.91 | 6.34 | 6.57 | 7.46 | 27.28 |
| 515 | 64.54 | 57.72 | 55.09 | 56.04 | 233.39 |
| 516 | 9.24 | 9.72 | 10.35 | 5.51 | 34.82 |
| 520 | 29.04 | 53.6 | 61.71 | 68.28 | 212.63 |
| 521 | 4.97 | 3.89 | 5.03 | 3.68 | 17.57 |
| 522 | 90.04 | 94.29 | 97.28 | 110.07 | 391.68 |
| 523 | 32.77 | 26.45 | 27.33 | 26.57 | 113.12 |
| 524 | 19.7 | 26.52 | 15.92 | 25.27 | 87.41 |
| 525 | 38.07 | 34.55 | 37.43 | 39.2 | 149.25 |
| 526 | 33.97 | 41.43 | 41.26 | 40.1 | 156.76 |
| 527 | 168.23 | 172.6 | 163.65 | 180.54 | 685.02 |
| 528 | 92.42 | 87.04 | 81.46 | 74.14 | 335.06 |
| TOTAL | 1,354.14 | 1,470.96 | 1,518.76 | 1,493.66 | 5,837.52 |

STATE: KANSAS

| ZIP CODE | QUARTER 1 | QUARTER 2 | QUARTER 3 | QUARTER 4 | TOTAL GRAMS |
|---|---|---|---|---|---|
| 660 | 474.3 | 469.96 | 526.49 | 486.85 | 1,957.60 |
| 661 | 78.18 | 74.77 | 76.93 | 81.46 | 311.34 |
| 662 | 610.83 | 622.74 | 600.49 | 624.95 | 2,459.01 |
| 664 | 27.02 | 31.94 | 37.38 | 40.08 | 136.42 |
| 665 | 43.43 | 35.97 | 35.05 | 40.65 | 155.1 |
| 666 | 142.51 | 137.68 | 156 | 130.22 | 566.41 |
| 667 | 136.09 | 125.59 | 117.91 | 121.07 | 500.66 |
| 668 | 9.66 | 6.4 | 12.35 | 14.86 | 43.27 |
| 669 | 4.65 | 3.18 | 4.8 | 4.42 | 17.05 |
| 670 | 69.92 | 61.75 | 76.46 | 92.36 | 300.49 |
| 671 | 45.02 | 44.2 | 53.66 | 55.77 | 198.65 |
| 672 | 391.44 | 399.54 | 443.46 | 453.58 | 1,688.02 |
| 673 | 20.44 | 15.86 | 26.82 | 23.04 | 86.16 |
| 674 | 57.14 | 58.71 | 68.36 | 53.44 | 237.65 |
| 675 | 48.23 | 51.55 | 58.15 | 59.1 | 217.03 |
| 676 | 53.61 | 51.9 | 51.9 | 58.3 | 215.71 |

| | | | | | |
|---|---|---|---|---|---|
| 677 | 16.7 | 21.15 | 16.62 | 16.07 | 70.54 |
| 678 | 60.39 | 57.08 | 51.19 | 47.09 | 215.75 |
| 679 | 9.9 | 13.34 | 15.08 | 12.4 | 50.72 |
| TOTAL | 2,299.46 | 2,283.31 | 2,429.10 | 2,415.71 | 9,427.58 |

STATE: KENTUCKY

| ZIP CODE | QUARTER 1 | QUARTER 2 | QUARTER 3 | QUARTER 4 | TOTAL GRAMS |
|---|---|---|---|---|---|
| 400 | 631.37 | 662.03 | 615.96 | 614.07 | 2,523.43 |
| 401 | 151.71 | 150.33 | 140.63 | 147.23 | 589.9 |
| 402 | 2,491.62 | 2,476.77 | 2,559.60 | 2,466.39 | 9,994.38 |
| 403 | 3,592.50 | 3,628.14 | 3,698.78 | 3,772.47 | 14,691.89 |
| 404 | 1,981.15 | 2,037.15 | 1,994.58 | 2,045.82 | 8,058.70 |
| 405 | 2,278.88 | 2,283.48 | 2,486.72 | 2,442.05 | 9,491.13 |
| 406 | 371.38 | 374.78 | 394.73 | 396.6 | 1,537.49 |
| 407 | 2,320.78 | 2,120.70 | 1,820.01 | 1,701.51 | 7,963 |
| 408 | 295.05 | 299.44 | 261.87 | 308.46 | 1,164.82 |
| 409 | 1,684.79 | 1,491.57 | 1,335.78 | 1,401.57 | 5,913.71 |
| 410 | 1,712.52 | 1,773.47 | 1,775.20 | 1,853.28 | 7,114.47 |
| 411 | 1,602.82 | 1,688.72 | 1,621.35 | 1,635.74 | 6,548.63 |
| 412 | 989.67 | 1,005.43 | 1,059.96 | 1,114.64 | 4,169.70 |
| 413 | 631.9 | 632.15 | 673.25 | 742.6 | 2,679.90 |
| 414 | 360.91 | 437.85 | 469.11 | 546.55 | 1,814.42 |
| 415 | 2,521.67 | 2,507.09 | 2,380.75 | 2,498.08 | 9,907.59 |
| 416 | 1,154.84 | 1,238.13 | 1,324.73 | 1,539.04 | 5,256.74 |
| 417 | 759.89 | 840.61 | 942.29 | 1,037.17 | 3,579.96 |
| 418 | 325.28 | 344.68 | 384.81 | 399.81 | 1,454.58 |
| 420 | 721.34 | 832.31 | 867.8 | 847.67 | 3,269.12 |
| 421 | 663.09 | 727.44 | 738.92 | 812.27 | 2,941.72 |
| 422 | 224.32 | 248.2 | 246.51 | 243.09 | 962.12 |
| 423 | 543.51 | 580.26 | 568.27 | 593.67 | 2,285.71 |
| 424 | 219.03 | 253.78 | 280.64 | 297.8 | 1,051.25 |
| 425 | 591.76 | 615.72 | 617.96 | 627.38 | 2,452.82 |
| 426 | 591.65 | 614.22 | 731.65 | 787.51 | 2,725.03 |
| 427 | 1,103.15 | 1,070.67 | 1,031.31 | 1,047.19 | 4,252.32 |
| TOTAL | 30,516.58 | 30,935.12 | 31,023.17 | 31,919.66 | 124,394.53 |

DRUG CODE:9064DRUG NAME:BUPRENORPHINE
STATE: LOUISIANA

| ZIP CODE | QUARTER 1 | QUARTER 2 | QUARTER 3 | QUARTER 4 | TOTAL GRAMS |
|---|---|---|---|---|---|
| 700 | 3,524.60 | 3,513.13 | 3,671.97 | 3,778.48 | 14,488.18 |
| 701 | 797.25 | 827.89 | 879.85 | 835.38 | 3,340.37 |
| 703 | 707.13 | 704.71 | 1,033.42 | 1,012.96 | 3,458.22 |
| 704 | 2,302.45 | 2,338.37 | 2,994.47 | 3,044.05 | 10,679.34 |
| 705 | 1,280.46 | 1,286.51 | 2,388.28 | 2,398.93 | 7,354.18 |
| 706 | 766.25 | 792.08 | 1,057.64 | 1,073.91 | 3,689.88 |
| 707 | 934.22 | 969.87 | 1,610.03 | 1,653.42 | 5,167.54 |
| 708 | 680.12 | 651.22 | 781.42 | 797.55 | 2,910.31 |
| 710 | 243.66 | 272.79 | 335.95 | 391.74 | 1,244.14 |
| 711 | 1,583.01 | 1,471.73 | 1,070.77 | 1,061.83 | 5,187.34 |
| 712 | 848.94 | 902.27 | 949.82 | 1,022.87 | 3,723.90 |
| 713 | 291.83 | 293.35 | 375.95 | 402.14 | 1,363.27 |
| 714 | 237.28 | 266.66 | 305.1 | 307.5 | 1,116.54 |
| TOTAL | 14,197.20 | 14,290.58 | 17,454.67 | 17,780.76 | 63,723.21 |

STATE: MAINE

| ZIP CODE | QUARTER 1 | QUARTER 2 | QUARTER 3 | QUARTER 4 | TOTAL GRAMS |
|---|---|---|---|---|---|
| 39 | 48.11 | 51.67 | 61.67 | 62.97 | 224.42 |
| 40 | 1,807.28 | 1,842.95 | 1,906.99 | 1,944.41 | 7,501.63 |
| 41 | 883.2 | 872.81 | 945.41 | 946.6 | 3,648.02 |

| | | | | | |
|---|---|---|---|---|---|
| 42 | 1,020.83 | 1,091.09 | 1,145.55 | 1,226.16 | 4,483.63 |
| 43 | 626.36 | 647.25 | 649.82 | 666.82 | 2,590.25 |
| 44 | 831.24 | 879.99 | 916.2 | 958.66 | 3,586.09 |
| 45 | 286.89 | 294.58 | 306.23 | 307.18 | 1,194.88 |
| 46 | 362.11 | 404.36 | 451.66 | 465.31 | 1,683.44 |
| 47 | 238.85 | 275.35 | 311.34 | 417.76 | 1,243.30 |
| 48 | 441.09 | 483.14 | 572.74 | 646.89 | 2,143.86 |
| 49 | 832.05 | 833.82 | 910.02 | 954.03 | 3,529.92 |
| TOTAL | 7,378.01 | 7,677.01 | 8,177.63 | 8,596.79 | 31,829.44 |

STATE: MARYLAND

| ZIP CODE | QUARTER 1 | QUARTER 2 | QUARTER 3 | QUARTER 4 | TOTAL GRAMS |
|---|---|---|---|---|---|
| 206 | 1,188.64 | 1,334.44 | 1,318.90 | 1,402.32 | 5,244.30 |
| 207 | 914.39 | 932.19 | 1,016.95 | 1,048.29 | 3,911.82 |
| 208 | 978.63 | 1,031.85 | 1,057.07 | 1,085.47 | 4,153.02 |
| 209 | 292.34 | 374.76 | 394.41 | 403.96 | 1,465.47 |
| 210 | 2,962.02 | 3,113.70 | 3,080.22 | 3,080.52 | 12,236.46 |
| 211 | 1,535.86 | 1,620.47 | 1,581.91 | 1,562.19 | 6,300.43 |
| 212 | 8,978.86 | 9,501.53 | 9,202.06 | 9,623.52 | 37,305.97 |
| 214 | 273.59 | 310.7 | 254.4 | 271.77 | 1,110.46 |
| 215 | 941.43 | 962.12 | 1,025.09 | 950.43 | 3,879.07 |
| 216 | 700.63 | 686.72 | 710.1 | 676.11 | 2,773.56 |
| 217 | 1,497.83 | 1,625.43 | 1,670.53 | 1,724.13 | 6,517.92 |
| 218 | 620.93 | 659.32 | 721.75 | 682.41 | 2,684.41 |
| 219 | 883.85 | 936.22 | 946.27 | 1,082.22 | 3,848.56 |
| TOTAL | 21,769 | 23,089.45 | 22,979.66 | 23,593.34 | 91,431.45 |

STATE: MASSACHUSETTS

| ZIP CODE | QUARTER 1 | QUARTER 2 | QUARTER 3 | QUARTER 4 | TOTAL GRAMS |
|---|---|---|---|---|---|
| 10 | 3,178.77 | 3,274.80 | 3,323.18 | 3,408.07 | 13,184.82 |
| 11 | 1,112.95 | 1,145.12 | 1,244.50 | 1,218.56 | 4,721.13 |
| 12 | 1,107.65 | 1,105.84 | 1,236.90 | 1,165.39 | 4,615.78 |
| 13 | 805.41 | 761.81 | 862.32 | 824.05 | 3,253.59 |
| 14 | 897.37 | 931.58 | 986.72 | 957.9 | 3,773.57 |
| 15 | 1,194.65 | 1,266.73 | 1,338.96 | 1,301.51 | 5,101.85 |
| 16 | 2,194.01 | 2,135.66 | 2,302.14 | 2,167.90 | 8,799.71 |
| 17 | 1,403.86 | 1,407.79 | 1,420.15 | 1,366.82 | 5,598.62 |
| 18 | 2,918.97 | 3,005.54 | 3,120.58 | 3,084.85 | 12,129.94 |
| 19 | 2,349.48 | 2,412.61 | 2,596.03 | 2,597.85 | 9,955.97 |
| 20 | 1,512.55 | 1,530.46 | 1,589.97 | 1,555.33 | 6,188.31 |
| 21 | 7,510.70 | 7,743 | 8,114.42 | 8,117.52 | 31,485.64 |
| 22 | 81.26 | 83.65 | 73.72 | 94.71 | 333.34 |
| 23 | 2,472.94 | 2,518.15 | 2,583.27 | 2,695.66 | 10,270.02 |
| 24 | 957.95 | 1,016.12 | 1,041.22 | 1,050.11 | 4,065.40 |
| 25 | 935.58 | 976.5 | 949.83 | 977.54 | 3,839.45 |
| 26 | 1,178.79 | 1,251.83 | 1,280.33 | 1,256.81 | 4,967.76 |
| 27 | 4,791.25 | 4,890.81 | 4,952.93 | 4,978.60 | 19,613.59 |
| TOTAL | 36,604.14 | 37,458 | 39,017.17 | 38,819.18 | 151,898.49 |

STATE: MICHIGAN

DRUG CODE:9064DRUG NAME:BUPRENORPHINE
STATE:MICHIGAN

| ZIP CODE | QUARTER 1 | QUARTER 2 | QUARTER 3 | QUARTER 4 | TOTAL GRAMS |
|---|---|---|---|---|---|
| 480 | 3,484.44 | 3,685.33 | 3,660.70 | 3,739.03 | 14,569.50 |
| 481 | 7,047.03 | 7,255.91 | 7,290.82 | 7,317.73 | 28,911.49 |
| 482 | 821.99 | 888.44 | 918.44 | 894.79 | 3,523.66 |
| 483 | 2,684.42 | 2,683.79 | 2,710.67 | 2,820.08 | 10,898.96 |
| 484 | 769.97 | 815.16 | 831.1 | 840.47 | 3,256.70 |
| 485 | 492.62 | 566.96 | 548.31 | 534.18 | 2,142.07 |

| | | | | | |
|---|---|---|---|---|---|
| 486 | 715.81 | 764.73 | 791.62 | 843.96 | 3,116.12 |
| 487 | 509.05 | 477.43 | 510.86 | 544.98 | 2,042.32 |
| 488 | 970.4 | 937.96 | 934.32 | 958.92 | 3,801.60 |
| 489 | 198.34 | 200.76 | 214.99 | 218.35 | 832.44 |
| 490 | 973.7 | 1,023.87 | 1,047.41 | 1,118.32 | 4,163.30 |
| 491 | 61.85 | 71.06 | 76.72 | 76.44 | 286.07 |
| 492 | 684.56 | 714.43 | 679.25 | 651.57 | 2,729.81 |
| 493 | 203.74 | 204.41 | 221.79 | 266.57 | 896.51 |
| 494 | 597.87 | 622.01 | 593.89 | 639.4 | 2,453.17 |
| 495 | 675.87 | 675.68 | 750.9 | 775.14 | 2,877.59 |
| 496 | 913.13 | 1,014.04 | 977.42 | 1,062.19 | 3,966.78 |
| 497 | 867.58 | 912.45 | 967.76 | 1,014.97 | 3,762.76 |
| 498 | 583.64 | 593.77 | 675.28 | 638.08 | 2,490.77 |
| 499 | 120 | 103.55 | 104.38 | 84.62 | 412.55 |
| TOTAL | 23,376.01 | 24,211.74 | 24,506.63 | 25,039.79 | 97,134.17 |

STATE: MINNESOTA

| ZIP CODE | QUARTER 1 | QUARTER 2 | QUARTER 3 | QUARTER 4 | TOTAL GRAMS |
|---|---|---|---|---|---|
| 550 | 742.73 | 745.76 | 782.37 | 772.49 | 3,043.35 |
| 551 | 1,029.99 | 1,059.12 | 1,123.72 | 1,214.20 | 4,427.03 |
| 553 | 668.01 | 707.66 | 776.67 | 866.04 | 3,018.38 |
| 554 | 1,534.19 | 1,647.08 | 1,651.78 | 1,765.16 | 6,598.21 |
| 556 | 12.48 | 15.15 | 11.94 | 14.79 | 54.36 |
| 557 | 160.1 | 159.52 | 160.9 | 159.27 | 639.79 |
| 558 | 268.54 | 263.85 | 271.51 | 282.35 | 1,086.25 |
| 559 | 206.5 | 207.6 | 215.58 | 241.7 | 871.38 |
| 560 | 144.17 | 153.88 | 162.99 | 183.38 | 644.42 |
| 561 | 5.45 | 5.88 | 4.23 | 9.32 | 24.88 |
| 562 | 39.92 | 44.11 | 49.56 | 54.67 | 188.26 |
| 563 | 279.19 | 297.84 | 258.81 | 246.19 | 1,082.03 |
| 564 | 99.45 | 97.35 | 93.76 | 105.63 | 396.19 |
| 565 | 132.03 | 169.82 | 215.37 | 247.13 | 764.35 |
| 566 | 367.8 | 411.41 | 430.77 | 489.56 | 1,699.54 |
| 567 | 12.56 | 16.54 | 39.08 | 64.24 | 132.42 |
| TOTAL | 5,703.11 | 6,002.57 | 6,249.04 | 6,716.12 | 24,670.84 |

STATE: MISSISSIPPI

| ZIP CODE | QUARTER 1 | QUARTER 2 | QUARTER 3 | QUARTER 4 | TOTAL GRAMS |
|---|---|---|---|---|---|
| 386 | 1,155.11 | 1,188.39 | 1,254.23 | 1,166.75 | 4,764.48 |
| 387 | 66.38 | 77.14 | 83.99 | 69.43 | 296.94 |
| 388 | 1,371.91 | 1,413.68 | 1,417.13 | 1,504.56 | 5,707.28 |
| 389 | 105.04 | 109.52 | 128.76 | 130.09 | 473.41 |
| 390 | 595.58 | 607.25 | 618.49 | 624.69 | 2,446.01 |
| 391 | 728.55 | 739.82 | 716.25 | 726.53 | 2,911.15 |
| 392 | 508.13 | 507.57 | 526.47 | 529.5 | 2,071.67 |
| 393 | 415.32 | 449.6 | 427.79 | 449.97 | 1,742.68 |
| 394 | 1,587.66 | 1,672.36 | 1,628.41 | 1,641.02 | 6,529.45 |
| 395 | 2,001.33 | 2,065.93 | 2,076.33 | 2,178.73 | 8,322.32 |
| 396 | 263.5 | 252.86 | 274.61 | 270.89 | 1,061.86 |
| 397 | 185.16 | 194.77 | 203.45 | 174.58 | 757.96 |
| TOTAL | 8,983.67 | 9,278.89 | 9,355.91 | 9,466.74 | 37,085.21 |

STATE: MISSOURI

| ZIP CODE | QUARTER 1 | QUARTER 2 | QUARTER 3 | QUARTER 4 | TOTAL GRAMS |
|---|---|---|---|---|---|
| 630 | 911.93 | 903.18 | 962.29 | 969.31 | 3,746.71 |
| 631 | 1,370.11 | 1,494.80 | 1,505.86 | 1,449.29 | 5,820.06 |
| 633 | 470.4 | 507.7 | 503.6 | 503.41 | 1,985.11 |
| 634 | 36.03 | 34.74 | 49.79 | 66.75 | 187.31 |
| 635 | 18.38 | 24.13 | 22.4 | 20.61 | 85.52 |
| 636 | 386.14 | 436.18 | 421.27 | 409.95 | 1,653.54 |
| 637 | 124.83 | 106.24 | 104.46 | 138.34 | 473.87 |

| | | | | | |
|---|---|---|---|---|---|
| 638 | 147.78 | 150.01 | 155.27 | 164.5 | 617.56 |
| 639 | 109.56 | 123.5 | 125.02 | 131.06 | 489.14 |

DRUG CODE:9064DRUG NAME:BUPRENORPHINE
STATE:MISSOURI

| ZIP CODE | QUARTER 1 | QUARTER 2 | QUARTER 3 | QUARTER 4 | TOTAL GRAMS |
|---|---|---|---|---|---|
| 640 | 632.69 | 683.94 | 680.7 | 708.64 | 2,705.97 |
| 641 | 514.67 | 548.09 | 589.32 | 634.12 | 2,286.20 |
| 644 | 54.66 | 55.93 | 55.63 | 60.03 | 226.25 |
| 645 | 206.38 | 212.82 | 225.37 | 227.22 | 871.79 |
| 646 | 17.4 | 22.94 | 18.1 | 17.65 | 76.09 |
| 647 | 79.76 | 80.41 | 85.57 | 93.11 | 338.85 |
| 648 | 146 | 167.05 | 151.74 | 175.95 | 640.74 |
| 650 | 127.71 | 134.87 | 133.4 | 143.9 | 539.88 |
| 651 | 103.76 | 106.8 | 118.17 | 110.55 | 439.28 |
| 652 | 386.72 | 398.16 | 420.51 | 424.7 | 1,630.09 |
| 653 | 63.74 | 65.92 | 70.83 | 73.62 | 274.11 |
| 654 | 124.21 | 129.32 | 128.71 | 122.09 | 504.33 |
| 655 | 119 | 128.79 | 127.35 | 155.34 | 530.48 |
| 656 | 202.97 | 195.69 | 206.72 | 215.51 | 820.89 |
| 657 | 304.12 | 294.68 | 287.89 | 296.14 | 1,182.83 |
| 658 | 1,067.83 | 1,110.53 | 1,202.01 | 1,316.16 | 4,696.53 |
| TOTAL | 7,726.78 | 8,116.42 | 8,351.98 | 8,627.95 | 32,823.13 |

STATE: MONTANA

| ZIP CODE | QUARTER 1 | QUARTER 2 | QUARTER 3 | QUARTER 4 | TOTAL GRAMS |
|---|---|---|---|---|---|
| 80 | 0.02 | 0.05 | 0 | 0 | 0.07 |
| 590 | 26.8 | 30.77 | 38.95 | 40.12 | 136.64 |
| 591 | 155.65 | 187.6 | 189.8 | 245.47 | 778.52 |
| 592 | 35.88 | 42.61 | 42.49 | 62.6 | 183.58 |
| 593 | 33.16 | 34.8 | 34.55 | 26.86 | 129.37 |
| 594 | 295.62 | 320.38 | 402.28 | 546.19 | 1,564.47 |
| 595 | 14.82 | 15.71 | 19.83 | 21.74 | 72.1 |
| 596 | 239.14 | 239.07 | 235.4 | 236.56 | 950.17 |
| 597 | 190.92 | 196.58 | 241.2 | 232.75 | 861.45 |
| 598 | 340.65 | 434.4 | 440.2 | 453.26 | 1,668.51 |
| 599 | 214.1 | 236.19 | 223.97 | 207.92 | 882.18 |
| TOTAL | 1,546.76 | 1,738.16 | 1,868.67 | 2,073.47 | 7,227.06 |

STATE: NEBRASKA

| ZIP CODE | QUARTER 1 | QUARTER 2 | QUARTER 3 | QUARTER 4 | TOTAL GRAMS |
|---|---|---|---|---|---|
| 680 | 51.9 | 59.64 | 65.85 | 73.04 | 250.43 |
| 681 | 434.91 | 483.35 | 501.23 | 529.67 | 1,949.16 |
| 683 | 23.07 | 32.44 | 33.2 | 32.69 | 121.4 |
| 684 | 20.41 | 26.98 | 19.63 | 19.91 | 86.93 |
| 685 | 233.74 | 241.44 | 267.54 | 261.42 | 1,004.14 |
| 686 | 19.02 | 16.04 | 27.19 | 21.66 | 83.91 |
| 687 | 35.41 | 49.24 | 51.39 | 62.26 | 198.3 |
| 688 | 53.13 | 52.4 | 58.84 | 57.33 | 221.7 |
| 689 | 20.2 | 13.4 | 17.64 | 26.19 | 77.43 |
| 690 | 4.11 | 5.44 | 1.34 | 0.83 | 11.72 |
| 691 | 149.08 | 137.94 | 141.99 | 130.93 | 559.94 |
| 692 | 0.14 | 0.16 | 0.76 | 0.06 | 1.12 |
| 693 | 23.39 | 23.16 | 20.15 | 26.63 | 93.33 |
| TOTAL | 1,068.51 | 1,141.63 | 1,206.75 | 1,242.62 | 4,659.51 |

STATE: NEVADA

| ZIP CODE | QUARTER 1 | QUARTER 2 | QUARTER 3 | QUARTER 4 | TOTAL GRAMS |
|---|---|---|---|---|---|
| 890 | 647.29 | 632.79 | 639.59 | 706.58 | 2,626.25 |

| | | | | | |
|---|---|---|---|---|---|
| 891 | 1,453.48 | 1,481.25 | 1,464.44 | 1,444.32 | 5,843.49 |
| 893 | 11.72 | 9.32 | 11.16 | 11.5 | 43.7 |
| 894 | 269.24 | 276.63 | 284.44 | 285.39 | 1,115.70 |
| 895 | 461.83 | 517.26 | 540.19 | 578.55 | 2,097.83 |
| 897 | 89.96 | 96.54 | 90.14 | 99.41 | 376.05 |
| 898 | 59.46 | 53.81 | 54.06 | 54.93 | 222.26 |
| TOTAL | 2,992.98 | 3,067.60 | 3,084.02 | 3,180.68 | 12,325.28 |

STATE: NEW HAMPSHIRE

| ZIP CODE | QUARTER 1 | QUARTER 2 | QUARTER 3 | QUARTER 4 | TOTAL GRAMS |
|---|---|---|---|---|---|
| 30 | 1,150.58 | 1,184.67 | 1,239.87 | 1,186.19 | 4,761.31 |
| 31 | 992.19 | 1,080.87 | 1,154.66 | 1,141.01 | 4,368.73 |
| 32 | 899.48 | 924.3 | 1,004.75 | 1,071.35 | 3,899.88 |
| 33 | 642.94 | 699.03 | 764.44 | 745.5 | 2,851.91 |
| 34 | 206.06 | 228.59 | 311.42 | 284.31 | 1,030.38 |
| 35 | 225.82 | 239.1 | 279.89 | 312.41 | 1,057.22 |
| 36 | 8.64 | 11.16 | 9.84 | 9.24 | 38.88 |

DRUG CODE:9064DRUG NAME:BUPRENORPHINE
STATE:NEW HAMPSHIRE

| ZIP CODE | QUARTER 1 | QUARTER 2 | QUARTER 3 | QUARTER 4 | TOTAL GRAMS |
|---|---|---|---|---|---|
| 37 | 980.31 | 927.13 | 1,058.89 | 1,154.13 | 4,120.46 |
| 38 | 1,414.82 | 1,493.28 | 1,616.28 | 1,634.32 | 6,158.70 |
| TOTAL | 6,520.84 | 6,788.13 | 7,440.04 | 7,538.46 | 28,287.47 |

STATE: NEW JERSEY

| ZIP CODE | QUARTER 1 | QUARTER 2 | QUARTER 3 | QUARTER 4 | TOTAL GRAMS |
|---|---|---|---|---|---|
| 70 | 2,768.44 | 2,822.20 | 2,912.77 | 2,864.02 | 11,367.43 |
| 71 | 835.63 | 898.8 | 917.4 | 943.88 | 3,595.71 |
| 72 | 242.23 | 249.52 | 228.72 | 241.07 | 961.54 |
| 73 | 184.89 | 175.65 | 180.7 | 174.5 | 715.74 |
| 74 | 1,197.70 | 1,223.10 | 1,171.81 | 1,198.42 | 4,791.03 |
| 75 | 284.14 | 252.4 | 242.74 | 233.96 | 1,013.24 |
| 76 | 757.63 | 765.61 | 748.54 | 803.83 | 3,075.61 |
| 77 | 1,862.93 | 1,958.70 | 1,975.39 | 2,001.44 | 7,798.46 |
| 78 | 740.28 | 767.93 | 804.93 | 809.94 | 3,123.08 |
| 79 | 329.69 | 354.06 | 339.74 | 375.15 | 1,398.64 |
| 80 | 6,599.32 | 6,848.70 | 6,782.26 | 6,798.67 | 27,028.95 |
| 81 | 602 | 655.37 | 667.61 | 686.54 | 2,611.52 |
| 82 | 1,681.94 | 1,719.57 | 1,775.55 | 1,735.22 | 6,912.28 |
| 83 | 882.77 | 875.74 | 871.29 | 929.5 | 3,559.30 |
| 84 | 248.78 | 269.49 | 250.04 | 215.61 | 983.92 |
| 85 | 521.58 | 518.07 | 503.4 | 551.56 | 2,094.61 |
| 86 | 653.77 | 668.74 | 654.51 | 671.35 | 2,648.37 |
| 87 | 2,240.39 | 2,342.30 | 2,396.46 | 2,381.03 | 9,360.18 |
| 88 | 1,659.69 | 1,763.93 | 1,783.75 | 1,733.25 | 6,940.62 |
| 89 | 163.99 | 186.27 | 166.79 | 168.02 | 685.07 |
| TOTAL | 24,457.79 | 25,316.15 | 25,374.40 | 25,516.96 | 100,665.30 |

STATE: NEW MEXICO

| ZIP CODE | QUARTER 1 | QUARTER 2 | QUARTER 3 | QUARTER 4 | TOTAL GRAMS |
|---|---|---|---|---|---|
| 870 | 297.94 | 329.27 | 349.29 | 352.17 | 1,328.67 |
| 871 | 3,086.08 | 3,163.78 | 3,346.13 | 3,187.72 | 12,783.71 |
| 873 | 103.52 | 106.74 | 118.42 | 111.26 | 439.94 |
| 874 | 294.86 | 288.23 | 274.69 | 282.04 | 1,139.82 |
| 875 | 1,326.40 | 1,438.56 | 1,573.41 | 1,672.92 | 6,011.29 |
| 877 | 135.51 | 127.31 | 132.54 | 145.81 | 541.17 |
| 878 | 58.1 | 36.27 | 42.37 | 41.53 | 178.27 |
| 879 | 10.84 | 12.12 | 12.28 | 11.18 | 46.42 |
| 880 | 306.93 | 314.92 | 317.34 | 309.2 | 1,248.39 |

| | | | | | |
|---|---|---|---|---|---|
| 881 | 19.35 | 18.45 | 15.8 | 24.54 | 78.14 |
| 882 | 321.89 | 326.18 | 321.99 | 351.01 | 1,321.07 |
| 883 | 65.22 | 74.04 | 70.32 | 77.63 | 287.21 |
| 884 | 4.32 | 4.42 | 5.82 | 7.57 | 22.13 |
| TOTAL | 6,030.96 | 6,240.29 | 6,580.40 | 6,574.58 | 25,426.23 |

STATE: NEW YORK

| ZIP CODE | QUARTER 1 | QUARTER 2 | QUARTER 3 | QUARTER 4 | TOTAL GRAMS |
|---|---|---|---|---|---|
| 100 | 2,758.54 | 2,911.83 | 2,807.44 | 2,658.76 | 11,136.57 |
| 101 | 85.01 | 77.92 | 67.44 | 66.66 | 297.03 |
| 102 | 3.07 | 1.92 | 1.09 | 1.22 | 7.3 |
| 103 | 1,576.11 | 1,729.83 | 1,757.32 | 1,697.02 | 6,760.28 |
| 104 | 2,528.50 | 2,903.76 | 2,961.33 | 2,495.53 | 10,889.12 |
| 105 | 1,030.57 | 1,008.18 | 1,054.50 | 1,007.25 | 4,100.50 |
| 106 | 243.13 | 251.5 | 258.47 | 241.01 | 994.11 |
| 107 | 342.79 | 357.51 | 341.82 | 349.62 | 1,391.74 |
| 108 | 94.17 | 94.5 | 89.31 | 101.19 | 379.17 |
| 109 | 1,271.25 | 1,328.88 | 1,355.81 | 1,375.95 | 5,331.89 |
| 110 | 409.46 | 448.6 | 479.25 | 488.74 | 1,826.05 |
| 111 | 191.2 | 218.96 | 201.68 | 221.57 | 833.41 |
| 112 | 2,687.87 | 2,612.06 | 2,701.77 | 2,780.25 | 10,781.95 |
| 113 | 1,014.51 | 1,012.57 | 1,098.18 | 1,054.42 | 4,179.68 |
| 114 | 430.68 | 455.73 | 442.36 | 487.75 | 1,816.52 |
| 115 | 1,214.58 | 1,226.62 | 1,237.44 | 1,253.79 | 4,932.43 |
| 116 | 91.32 | 91.87 | 96.02 | 85.09 | 364.3 |
| 117 | 5,548.44 | 5,857.71 | 5,797.68 | 5,821.69 | 23,025.52 |
| 118 | 535.02 | 527.13 | 484.52 | 452.8 | 1,999.47 |
| 119 | 990.89 | 1,056.78 | 1,067.77 | 1,071.16 | 4,186.60 |
| 120 | 1,321.63 | 1,372 | 1,436.33 | 1,477.41 | 5,607.37 |
| 121 | 769.64 | 774.52 | 797.71 | 806.67 | 3,148.54 |

DRUG CODE:9064DRUG NAME:BUPRENORPHINE
STATE:NEW YORK

| ZIP CODE | QUARTER 1 | QUARTER 2 | QUARTER 3 | QUARTER 4 | TOTAL GRAMS |
|---|---|---|---|---|---|
| 122 | 600.72 | 592.04 | 584.16 | 593.28 | 2,370.20 |
| 123 | 560.18 | 589.11 | 607.92 | 618.25 | 2,375.46 |
| 124 | 691.5 | 708.96 | 742.29 | 750.42 | 2,893.17 |
| 125 | 1,116.54 | 1,189.52 | 1,164.05 | 1,268.75 | 4,738.86 |
| 126 | 292.44 | 255.8 | 277.43 | 258.98 | 1,084.65 |
| 127 | 649.67 | 708.49 | 723.4 | 745.17 | 2,826.73 |
| 128 | 1,136.71 | 1,128.49 | 1,217.58 | 1,195.54 | 4,678.32 |
| 129 | 684.4 | 752.77 | 790.2 | 853.29 | 3,080.66 |
| 130 | 598.2 | 696.38 | 717.91 | 844.3 | 2,856.79 |
| 131 | 221.29 | 264.27 | 299.92 | 301.97 | 1,087.45 |
| 132 | 661.32 | 710.99 | 755.01 | 806.49 | 2,933.81 |
| 133 | 257.77 | 289.69 | 289.07 | 306.13 | 1,142.66 |
| 134 | 564.4 | 589.51 | 584.51 | 602.33 | 2,340.75 |
| 135 | 307.72 | 353.36 | 343.57 | 321.49 | 1,326.14 |
| 136 | 642.7 | 698.13 | 745.66 | 750.73 | 2,837.22 |
| 137 | 232.26 | 254.34 | 245.91 | 262.76 | 995.27 |
| 138 | 578.94 | 599.21 | 593.98 | 622.68 | 2,394.81 |
| 139 | 280.69 | 284.7 | 287.23 | 341.9 | 1,194.52 |
| 140 | 1,946.92 | 1,985.90 | 2,073.63 | 2,155.46 | 8,161.91 |
| 141 | 921.9 | 919.43 | 945.08 | 951.03 | 3,737.44 |
| 142 | 3,065.76 | 2,991.62 | 3,055.52 | 3,066.85 | 12,179.75 |
| 143 | 659.82 | 645.15 | 659.42 | 632.18 | 2,596.57 |
| 144 | 785.66 | 774.91 | 788.63 | 818.85 | 3,168.05 |
| 145 | 594.68 | 622.95 | 649.61 | 616.92 | 2,484.16 |
| 146 | 1,956.11 | 1,945.69 | 2,032.80 | 2,053.17 | 7,987.77 |
| 147 | 596.31 | 688.69 | 711.06 | 742.7 | 2,738.76 |

| | | | | | |
|---|---|---|---|---|---|
| 148 | 699.19 | 731.4 | 744.92 | 727.19 | 2,902.70 |
| 149 | 196.07 | 189.34 | 198.44 | 218.34 | 802.19 |
| TOTAL | 46,638.25 | 48,481.22 | 49,364.15 | 49,422.70 | 193,906.32 |

STATE: NORTH CAROLINA

| ZIP CODE | QUARTER 1 | QUARTER 2 | QUARTER 3 | QUARTER 4 | TOTAL GRAMS |
|---|---|---|---|---|---|
| 270 | 493.82 | 505.12 | 481.67 | 495.49 | 1,976.10 |
| 271 | 496.66 | 506.19 | 483.71 | 513.74 | 2,000.30 |
| 272 | 978.72 | 987.21 | 996.62 | 1,151.07 | 4,113.62 |
| 273 | 704.96 | 785.84 | 816.47 | 805.48 | 3,112.75 |
| 274 | 446.05 | 457.05 | 539.44 | 582.92 | 2,025.46 |
| 275 | 1,828.38 | 1,901.21 | 1,884.66 | 1,950.70 | 7,564.95 |
| 276 | 802.5 | 893.58 | 927.97 | 1,004.85 | 3,628.90 |
| 277 | 1,507.60 | 1,517.16 | 1,514.86 | 1,527.92 | 6,067.54 |
| 278 | 790.5 | 775.69 | 761.51 | 813.51 | 3,141.21 |
| 279 | 380.54 | 424.12 | 477.18 | 455.82 | 1,737.66 |
| 280 | 2,290.25 | 2,461.03 | 2,372.21 | 2,579.51 | 9,703 |
| 281 | 1,258.18 | 1,376.53 | 1,450.85 | 1,635.53 | 5,721.09 |
| 282 | 1,114.48 | 1,188.54 | 1,243.19 | 1,370.42 | 4,916.63 |
| 283 | 1,692.88 | 1,827.30 | 1,896.48 | 2,034.38 | 7,451.04 |
| 284 | 2,397.92 | 2,619.45 | 2,697.02 | 2,910.36 | 10,624.75 |
| 285 | 1,125.78 | 1,123.87 | 1,143.78 | 1,129.68 | 4,523.11 |
| 286 | 3,721.31 | 4,073.54 | 4,050.41 | 4,620.05 | 16,465.31 |
| 287 | 1,979.51 | 2,063.25 | 2,081.02 | 2,176.47 | 8,300.25 |
| 288 | 928.7 | 995.38 | 1,075.13 | 1,190.05 | 4,189.26 |
| 289 | 140.47 | 146.28 | 146.06 | 155.73 | 588.54 |
| TOTAL | 25,079.21 | 26,628.34 | 27,040.24 | 29,103.68 | 107,851.47 |

STATE: NORTH DAKOTA

| ZIP CODE | QUARTER 1 | QUARTER 2 | QUARTER 3 | QUARTER 4 | TOTAL GRAMS |
|---|---|---|---|---|---|
| 580 | 5.84 | 3.38 | 4.8 | 4.35 | 18.37 |
| 581 | 223.79 | 266.73 | 261.63 | 251.15 | 1,003.30 |
| 582 | 25.93 | 23.81 | 29.92 | 38.35 | 118.01 |
| 583 | 11.85 | 31.82 | 40.39 | 36.54 | 120.6 |
| 584 | 15.1 | 14.52 | 18.5 | 17.85 | 65.97 |
| 585 | 320.25 | 350.25 | 358.5 | 348.21 | 1,377.21 |
| 586 | 54.27 | 52.68 | 48.15 | 47.61 | 202.71 |
| 587 | 32.03 | 37.36 | 39.35 | 41.49 | 150.23 |
| 588 | 50.76 | 59.38 | 62.76 | 55.87 | 228.77 |
| TOTAL | 739.82 | 839.93 | 864 | 841.42 | 3,285.17 |

STATE: OHIO

| ZIP CODE | QUARTER 1 | QUARTER 2 | QUARTER 3 | QUARTER 4 | TOTAL GRAMS |
|---|---|---|---|---|---|

DRUG CODE:9064DRUG NAME:BUPRENORPHINE
STATE:OHIO

| ZIP CODE | QUARTER 1 | QUARTER 2 | QUARTER 3 | QUARTER 4 | TOTAL GRAMS |
|---|---|---|---|---|---|
| 430 | 2,903.61 | 3,066.32 | 3,215.30 | 3,151.28 | 12,336.51 |
| 431 | 2,268.84 | 2,257.05 | 2,360.33 | 2,388.80 | 9,275.02 |
| 432 | 4,499.81 | 4,592.73 | 4,650.46 | 4,843.40 | 18,586.40 |
| 433 | 706.49 | 798.82 | 765.45 | 841.93 | 3,112.69 |
| 434 | 244.52 | 294.1 | 274.53 | 268.81 | 1,081.96 |
| 435 | 656.31 | 684.9 | 678.99 | 692.66 | 2,712.86 |
| 436 | 1,728.89 | 1,825.19 | 1,781.71 | 1,852.71 | 7,188.50 |
| 437 | 985.52 | 1,103.45 | 1,230.02 | 1,280.03 | 4,599.02 |
| 438 | 62.2 | 74.99 | 70.8 | 63.54 | 271.53 |
| 439 | 1,445.16 | 1,494.96 | 1,586.87 | 1,755.71 | 6,282.70 |
| 440 | 2,135.74 | 2,176.70 | 2,201.33 | 2,149.27 | 8,663.04 |
| 441 | 2,597 | 2,723.55 | 2,894.16 | 2,898.95 | 11,113.66 |
| 442 | 1,397.73 | 1,395.90 | 1,426.28 | 1,415.76 | 5,635.67 |

| | | | | | |
|---|---|---|---|---|---|
| 443 | 687.83 | 727.82 | 737.93 | 774.03 | 2,927.61 |
| 444 | 1,150.01 | 1,206.15 | 1,208.84 | 1,232.15 | 4,797.15 |
| 445 | 1,254.26 | 1,246.21 | 1,323.11 | 1,323.14 | 5,146.72 |
| 446 | 784.79 | 825.76 | 831.57 | 795.6 | 3,237.72 |
| 447 | 606.58 | 637.15 | 712.99 | 743.92 | 2,700.64 |
| 448 | 1,000.87 | 972.69 | 997.19 | 1,048.72 | 4,019.47 |
| 449 | 867.17 | 826.52 | 814.32 | 845.66 | 3,353.67 |
| 450 | 2,686 | 2,763.21 | 2,819.64 | 2,789.54 | 11,058.39 |
| 451 | 1,874.81 | 2,035.26 | 2,125.26 | 2,102.40 | 8,137.73 |
| 452 | 4,049.15 | 4,188.54 | 4,388.44 | 4,479.65 | 17,105.78 |
| 453 | 912.06 | 976.97 | 958.47 | 992.31 | 3,839.81 |
| 454 | 1,754.36 | 1,804.86 | 1,879.87 | 2,002 | 7,441.09 |
| 455 | 861.66 | 983.62 | 820.73 | 796.47 | 3,462.48 |
| 456 | 3,512.17 | 3,692.38 | 3,952.07 | 4,206.33 | 15,362.95 |
| 457 | 966.92 | 1,049.55 | 1,072.37 | 1,095.31 | 4,184.15 |
| 458 | 435.73 | 459.38 | 471.92 | 475.76 | 1,842.79 |
| TOTAL | 45,036.19 | 46,884.73 | 48,250.95 | 49,305.84 | 189,477.71 |

STATE: OKLAHOMA

| ZIP CODE | QUARTER 1 | QUARTER 2 | QUARTER 3 | QUARTER 4 | TOTAL GRAMS |
|---|---|---|---|---|---|
| 730 | 1,500.32 | 1,567.50 | 1,602.46 | 1,624.22 | 6,294.50 |
| 731 | 2,118.91 | 2,166.74 | 2,165.74 | 2,272.94 | 8,724.33 |
| 734 | 94.21 | 91.31 | 90.64 | 91.82 | 367.98 |
| 735 | 454.98 | 465.89 | 522.36 | 533.06 | 1,976.29 |
| 736 | 90.62 | 81.71 | 82.08 | 77.63 | 332.04 |
| 737 | 249.58 | 233.58 | 262.77 | 262.46 | 1,008.39 |
| 738 | 50.6 | 49.08 | 49.15 | 47.81 | 196.64 |
| 739 | 4.92 | 12.41 | 13.29 | 14.25 | 44.87 |
| 740 | 1,430.44 | 1,565.63 | 1,599.80 | 1,637.15 | 6,233.02 |
| 741 | 1,431.62 | 1,443.83 | 1,408.92 | 1,398.91 | 5,683.28 |
| 743 | 172.71 | 179.72 | 173.16 | 183.66 | 709.25 |
| 744 | 425.16 | 456.65 | 460.02 | 553.09 | 1,894.92 |
| 745 | 170.27 | 169.42 | 174.25 | 180.95 | 694.89 |
| 746 | 186.61 | 184.24 | 190.77 | 187.83 | 749.45 |
| 747 | 71.9 | 72.86 | 71.75 | 86.58 | 303.09 |
| 748 | 496.73 | 527.49 | 548.79 | 599.53 | 2,172.54 |
| 749 | 122.75 | 140.13 | 146.64 | 164.99 | 574.51 |
| TOTAL | 9,072.33 | 9,408.19 | 9,562.59 | 9,916.88 | 37,959.99 |

STATE: OREGON

| ZIP CODE | QUARTER 1 | QUARTER 2 | QUARTER 3 | QUARTER 4 | TOTAL GRAMS |
|---|---|---|---|---|---|
| 970 | 1,441.46 | 1,539.78 | 1,626.35 | 1,632.57 | 6,240.16 |
| 971 | 507.96 | 544.63 | 577.59 | 586.63 | 2,216.81 |
| 972 | 2,374.61 | 2,398.34 | 2,584.92 | 2,492.95 | 9,850.82 |
| 973 | 1,204.93 | 1,254.56 | 1,286.15 | 1,272.88 | 5,018.52 |
| 974 | 1,107.26 | 1,146.83 | 1,179.67 | 1,208.46 | 4,642.22 |
| 975 | 798.66 | 826.73 | 841.64 | 825.21 | 3,292.24 |
| 976 | 30.71 | 35.58 | 29.3 | 31.78 | 127.37 |
| 977 | 333.68 | 381.5 | 350.94 | 364.76 | 1,430.88 |
| 978 | 374.01 | 410.48 | 440.12 | 496.99 | 1,721.60 |
| 979 | 21.89 | 26.33 | 21.02 | 29.14 | 98.38 |
| TOTAL | 8,195.17 | 8,564.76 | 8,937.70 | 8,941.37 | 34,639 |

STATE: PENNSYLVANIA

| ZIP CODE | QUARTER 1 | QUARTER 2 | QUARTER 3 | QUARTER 4 | TOTAL GRAMS |
|---|---|---|---|---|---|
| 150 | 3,076.49 | 3,064.08 | 3,126.93 | 3,232.96 | 12,500.46 |

RETAIL DRUG DISTRIBUTION BY ZIP CODE WITHIN STATE BY GRAMS WT

STATE:PENNSYLVANIA

| ZIP CODE | QUARTER 1 | QUARTER 2 | QUARTER 3 | QUARTER 4 | TOTAL GRAMS |
|---|---|---|---|---|---|
| 151 | 1,882.26 | 1,963.39 | 2,065.40 | 2,195.12 | 8,106.17 |
| 152 | 5,464.68 | 5,481.95 | 5,564.83 | 5,759.18 | 22,270.64 |
| 153 | 2,027.11 | 2,180.64 | 2,245.50 | 2,211.33 | 8,664.58 |
| 154 | 1,123.19 | 1,143.30 | 1,166.97 | 1,196.73 | 4,630.19 |
| 155 | 267.89 | 266.28 | 272.7 | 335.6 | 1,142.47 |
| 156 | 2,081.19 | 2,104.39 | 2,180.70 | 2,327.57 | 8,693.85 |
| 157 | 489.04 | 547.71 | 582.77 | 571.52 | 2,191.04 |
| 158 | 578.83 | 604.96 | 626.32 | 639.85 | 2,449.96 |
| 159 | 917.9 | 865.65 | 903.41 | 992.32 | 3,679.28 |
| 160 | 1,118.74 | 1,140.87 | 1,206.93 | 1,276.32 | 4,742.86 |
| 161 | 925.16 | 899.55 | 868.55 | 937.06 | 3,630.32 |
| 162 | 721.07 | 759.07 | 738.25 | 735.8 | 2,954.19 |
| 163 | 572.44 | 573.2 | 573.68 | 613.53 | 2,332.85 |
| 164 | 177.66 | 186.54 | 188.42 | 228.22 | 780.84 |
| 165 | 1,382.24 | 1,462.74 | 1,650.59 | 1,657.26 | 6,152.83 |
| 166 | 1,308.79 | 1,381.43 | 1,459.62 | 1,541.06 | 5,690.90 |
| 167 | 153.28 | 165.7 | 109.38 | 118.04 | 546.4 |
| 168 | 550.18 | 575.23 | 543.3 | 496.2 | 2,164.91 |
| 169 | 92.08 | 94.55 | 105.39 | 113.57 | 405.59 |
| 170 | 1,434.82 | 1,427.34 | 1,481.39 | 1,533.67 | 5,877.22 |
| 171 | 309.67 | 328.28 | 325.61 | 349.4 | 1,312.96 |
| 172 | 680.41 | 662.28 | 703.52 | 745.09 | 2,791.30 |
| 173 | 855.61 | 846.93 | 914.01 | 947.16 | 3,563.71 |
| 174 | 416.9 | 406.19 | 425.15 | 454.61 | 1,702.85 |
| 175 | 323.05 | 326.07 | 347.24 | 368.13 | 1,364.49 |
| 176 | 442.82 | 444.82 | 411.79 | 429.57 | 1,729 |
| 177 | 1,078.25 | 1,165.57 | 1,305.94 | 1,356.82 | 4,906.58 |
| 178 | 622.23 | 646.5 | 643.5 | 713.41 | 2,625.64 |
| 179 | 368.93 | 394.58 | 379.46 | 393.64 | 1,536.61 |
| 180 | 690.33 | 690.77 | 697.8 | 761.59 | 2,840.49 |
| 181 | 300.79 | 328.68 | 318.45 | 357.39 | 1,305.31 |
| 182 | 360.6 | 380.58 | 348.12 | 340.03 | 1,429.33 |
| 183 | 394.23 | 429.39 | 406.29 | 442.04 | 1,671.95 |
| 184 | 576.25 | 632.92 | 776.01 | 816.09 | 2,801.27 |
| 185 | 1,883.61 | 1,985.98 | 1,904.19 | 1,844.84 | 7,618.62 |
| 186 | 604.94 | 646.35 | 730.01 | 768.3 | 2,749.60 |
| 187 | 1,828.41 | 1,916.73 | 1,911.50 | 1,975.40 | 7,632.04 |
| 188 | 211.71 | 261.1 | 254.33 | 274.04 | 1,001.18 |
| 189 | 1,015.93 | 1,102.39 | 1,084.67 | 1,085.75 | 4,288.74 |
| 190 | 4,705.95 | 5,027.50 | 5,160.05 | 5,006.81 | 19,900.31 |
| 191 | 6,178.23 | 6,499.09 | 6,529.75 | 6,687.67 | 25,894.74 |
| 193 | 871.35 | 914.03 | 914.41 | 868.43 | 3,568.22 |
| 194 | 1,695.62 | 1,791.97 | 1,783.34 | 2,046.21 | 7,317.14 |
| 195 | 243.65 | 272.66 | 279.09 | 268.95 | 1,064.35 |
| 196 | 458.71 | 477.98 | 460.71 | 531.71 | 1,929.11 |
| TOTAL | 53,463.22 | 55,467.91 | 56,675.97 | 58,545.99 | 224,153.09 |

STATE: PUERTO RICO

| ZIP CODE | QUARTER 1 | QUARTER 2 | QUARTER 3 | QUARTER 4 | TOTAL GRAMS |
|---|---|---|---|---|---|
| 6 | 578.73 | 577.86 | 578.92 | 576.78 | 2,312.29 |
| 7 | 1,193.87 | 1,295.58 | 1,419.06 | 1,401.18 | 5,309.69 |
| 9 | 587 | 590.91 | 592.16 | 576.03 | 2,346.10 |
| TOTAL | 2,359.60 | 2,464.35 | 2,590.14 | 2,553.99 | 9,968.08 |

STATE: RHODE ISLAND

| ZIP CODE | QUARTER 1 | QUARTER 2 | QUARTER 3 | QUARTER 4 | TOTAL GRAMS |
|---|---|---|---|---|---|
| 28 | 3,694.74 | 3,784.55 | 3,751.65 | 3,869.02 | 15,099.96 |
| 29 | 2,935.67 | 3,431.56 | 3,008.57 | 3,186.95 | 12,562.75 |
| TOTAL | 6,630.41 | 7,216.11 | 6,760.22 | 7,055.97 | 27,662.71 |

RETAIL DRUG DISTRIBUTION BY ZIP CODE WITHIN STATE BY GRAMS WT

STATE: SOUTH CAROLINA

| ZIP CODE | QUARTER 1 | QUARTER 2 | QUARTER 3 | QUARTER 4 | TOTAL GRAMS |
|---|---|---|---|---|---|
| 290 | 287.46 | 306.27 | 285.33 | 288.47 | 1,167.53 |
| 291 | 349.87 | 376.25 | 372.15 | 360.93 | 1,459.20 |
| 292 | 428.18 | 393.29 | 444.26 | 426.23 | 1,691.96 |
| 293 | 1,204.36 | 1,281.63 | 1,266.97 | 1,299.28 | 5,052.24 |
| 294 | 1,653.25 | 1,687.58 | 1,660.65 | 1,668.50 | 6,669.98 |
| 295 | 1,330.02 | 1,419.83 | 1,449.72 | 1,422.32 | 5,621.89 |
| 296 | 2,789.30 | 2,998.64 | 2,915.61 | 2,988.81 | 11,692.36 |

DRUG CODE:9064DRUG NAME:BUPRENORPHINE
STATE:SOUTH CAROLINA

| ZIP CODE | QUARTER 1 | QUARTER 2 | QUARTER 3 | QUARTER 4 | TOTAL GRAMS |
|---|---|---|---|---|---|
| 297 | 459.3 | 534.62 | 545.38 | 559.14 | 2,098.44 |
| 298 | 203.19 | 196.3 | 181.16 | 184.23 | 764.88 |
| 299 | 446.52 | 456.91 | 464.33 | 451.62 | 1,819.38 |
| TOTAL | 9,151.45 | 9,651.32 | 9,585.56 | 9,649.53 | 38,037.86 |

STATE: SOUTH DAKOTA

| ZIP CODE | QUARTER 1 | QUARTER 2 | QUARTER 3 | QUARTER 4 | TOTAL GRAMS |
|---|---|---|---|---|---|
| 570 | 42.45 | 42.97 | 41.14 | 37.97 | 164.53 |
| 571 | 111.62 | 125.22 | 127.65 | 127.83 | 492.32 |
| 572 | 24.4 | 30.41 | 37.07 | 42.89 | 134.77 |
| 573 | 11.45 | 18.05 | 19.67 | 17.9 | 67.07 |
| 574 | 53.2 | 63.52 | 63.41 | 74.48 | 254.61 |
| 575 | 6.27 | 6.31 | 5.9 | 7.25 | 25.73 |
| 576 | 4.46 | 5.04 | 5.08 | 4.55 | 19.13 |
| 577 | 100.41 | 94.02 | 87.21 | 80.89 | 362.53 |
| TOTAL | 354.26 | 385.54 | 387.13 | 393.76 | 1,520.69 |

STATE: TENNESSEE

| ZIP CODE | QUARTER 1 | QUARTER 2 | QUARTER 3 | QUARTER 4 | TOTAL GRAMS |
|---|---|---|---|---|---|
| 370 | 2,614.21 | 2,677.06 | 2,702.06 | 2,732.80 | 10,726.13 |
| 371 | 2,862.61 | 2,952.61 | 3,067.64 | 3,126.68 | 12,009.54 |
| 372 | 1,733.40 | 1,815.21 | 1,925.17 | 1,983.62 | 7,457.40 |
| 373 | 1,246.26 | 1,225.97 | 1,249.62 | 1,262.86 | 4,984.71 |
| 374 | 619.07 | 642.28 | 639.15 | 609.4 | 2,509.90 |
| 376 | 7,144.80 | 7,149.26 | 7,046.74 | 6,996.34 | 28,337.14 |
| 377 | 2,841.91 | 2,853.98 | 2,807.20 | 2,874.57 | 11,377.66 |
| 378 | 4,298.61 | 4,371.13 | 4,332.63 | 4,458.82 | 17,461.19 |
| 379 | 2,133.25 | 2,193.40 | 2,168.19 | 2,174.65 | 8,669.49 |
| 380 | 672.37 | 706.19 | 706.28 | 718.65 | 2,803.49 |
| 381 | 1,114.38 | 1,101.47 | 1,108.85 | 1,140.13 | 4,464.83 |
| 382 | 399.47 | 438.17 | 440.87 | 458.27 | 1,736.78 |
| 383 | 869.74 | 916.73 | 968.82 | 1,022.09 | 3,777.38 |
| 384 | 651.71 | 716.66 | 782.73 | 800.14 | 2,951.24 |
| 385 | 1,547.95 | 1,504.91 | 1,504.95 | 1,596.96 | 6,154.77 |
| TOTAL | 30,749.74 | 31,265.03 | 31,450.90 | 31,955.98 | 125,421.65 |

STATE: TEXAS

| ZIP CODE | QUARTER 1 | QUARTER 2 | QUARTER 3 | QUARTER 4 | TOTAL GRAMS |
|---|---|---|---|---|---|
| 750 | 2,479.27 | 2,534.14 | 2,558.63 | 2,589.79 | 10,161.83 |
| 751 | 718.95 | 724.57 | 767.9 | 774.97 | 2,986.39 |
| 752 | 992.01 | 1,040.11 | 1,024.93 | 1,060.54 | 4,117.59 |
| 753 | 1.92 | 1.1 | 1.27 | 0.98 | 5.27 |
| 754 | 408.86 | 434.01 | 416.52 | 413.58 | 1,672.97 |
| 755 | 127.6 | 133.63 | 112.45 | 122.91 | 496.59 |
| 756 | 566.85 | 558.27 | 543.44 | 529.54 | 2,198.10 |

| | | | | | |
|---|---|---|---|---|---|
| 757 | 403.47 | 419.15 | 381.06 | 423.31 | 1,626.99 |
| 758 | 68.06 | 63.04 | 65.38 | 92.35 | 288.83 |
| 759 | 427.91 | 412.12 | 387.31 | 405.97 | 1,633.31 |
| 760 | 1,773.68 | 1,833.83 | 1,836.52 | 1,907.43 | 7,351.46 |
| 761 | 958.02 | 998.58 | 968.05 | 1,016.70 | 3,941.35 |
| 762 | 518.82 | 539.95 | 540.84 | 556.9 | 2,156.51 |
| 763 | 284.99 | 282.22 | 289.71 | 290.33 | 1,147.25 |
| 764 | 90.63 | 94.44 | 95.22 | 92.46 | 372.75 |
| 765 | 251.34 | 289.15 | 267.62 | 291.52 | 1,099.63 |
| 766 | 71.1 | 69.63 | 67.08 | 63.09 | 270.9 |
| 767 | 108.84 | 125.15 | 94.69 | 118.22 | 446.9 |
| 768 | 34.26 | 36.23 | 34.35 | 37.39 | 142.23 |
| 769 | 63.87 | 61.42 | 54.46 | 61.25 | 241 |
| 770 | 2,213.35 | 2,277.39 | 2,269.53 | 2,434.79 | 9,195.06 |
| 772 | 0 | 0 | 0 | 0 | 0 |
| 773 | 2,036.29 | 2,106.78 | 2,096.75 | 2,122.97 | 8,362.79 |
| 774 | 965.56 | 972.4 | 953.13 | 924.85 | 3,815.94 |
| 775 | 2,273.56 | 2,316.76 | 2,299.98 | 2,254.10 | 9,144.40 |
| 776 | 877.35 | 902.93 | 864.84 | 891.21 | 3,536.33 |
| 777 | 314.77 | 305.3 | 319.2 | 321.21 | 1,260.48 |
| 778 | 279.67 | 292.41 | 280.17 | 298.44 | 1,150.69 |
| 779 | 151.55 | 148.52 | 135.68 | 131.7 | 567.45 |
| 780 | 315.17 | 307.73 | 339.84 | 333.65 | 1,296.39 |
| 781 | 318.48 | 309.79 | 303.21 | 305.77 | 1,237.25 |

DRUG CODE:9064DRUG NAME:BUPRENORPHINE
STATE:TEXAS

| ZIP CODE | QUARTER 1 | QUARTER 2 | QUARTER 3 | QUARTER 4 | TOTAL GRAMS |
|---|---|---|---|---|---|
| 782 | 943.39 | 902.52 | 926.76 | 897.95 | 3,670.62 |
| 783 | 141.79 | 149.55 | 141.1 | 153.01 | 585.45 |
| 784 | 334.84 | 360.84 | 351.95 | 314.61 | 1,362.24 |
| 785 | 108.92 | 111.46 | 110.62 | 107.25 | 438.25 |
| 786 | 842.56 | 874.37 | 872.49 | 891.18 | 3,480.60 |
| 787 | 1,251.98 | 1,350.55 | 1,243.06 | 1,300.34 | 5,145.93 |
| 788 | 28.39 | 26.14 | 25.56 | 26.14 | 106.23 |
| 789 | 29.32 | 35.98 | 30.35 | 26.88 | 122.53 |
| 790 | 32.05 | 30.19 | 30.32 | 40.15 | 132.71 |
| 791 | 84.6 | 96.24 | 90.7 | 90.59 | 362.13 |
| 792 | 2.01 | 1.82 | 2.52 | 3.92 | 10.27 |
| 793 | 34.47 | 36.67 | 33.93 | 31.88 | 136.95 |
| 794 | 158.65 | 147.83 | 150.75 | 149.53 | 606.76 |
| 795 | 36.48 | 36.19 | 32.11 | 26.84 | 131.62 |
| 796 | 160.78 | 151.37 | 162.8 | 195.59 | 670.54 |
| 797 | 408.38 | 386.82 | 388.89 | 427.99 | 1,612.08 |
| 798 | 8.81 | 8.16 | 8.28 | 11.07 | 36.32 |
| 799 | 108.62 | 133.85 | 123.51 | 131.25 | 497.23 |
| TOTAL | 24,812.24 | 25,431.30 | 25,095.46 | 25,694.09 | 101,033.09 |

STATE: UTAH

| ZIP CODE | QUARTER 1 | QUARTER 2 | QUARTER 3 | QUARTER 4 | TOTAL GRAMS |
|---|---|---|---|---|---|
| 840 | 3,943.09 | 4,016.74 | 4,148.80 | 4,284.05 | 16,392.68 |
| 841 | 2,187.54 | 2,214.42 | 2,293.78 | 2,277.76 | 8,973.50 |
| 843 | 319.89 | 327.48 | 343.37 | 359.63 | 1,350.37 |
| 844 | 745.51 | 765.36 | 803.99 | 768.77 | 3,083.63 |
| 845 | 187.36 | 197.97 | 194.22 | 202.95 | 782.5 |
| 846 | 933.43 | 964.35 | 968.13 | 1,024.46 | 3,890.37 |
| 847 | 245.45 | 257.47 | 298.44 | 287.86 | 1,089.22 |
| TOTAL | 8,562.27 | 8,743.79 | 9,050.73 | 9,205.48 | 35,562.27 |

STATE: VERMONT

| ZIP CODE | QUARTER 1 | QUARTER 2 | QUARTER 3 | QUARTER 4 | TOTAL GRAMS |
|---|---|---|---|---|---|
| 50 | 570.24 | 609.06 | 621.28 | 613.82 | 2,414.40 |
| 51 | 214.56 | 227.85 | 234.29 | 251.03 | 927.73 |
| 52 | 297.92 | 317.72 | 346.19 | 347.54 | 1,309.37 |
| 53 | 307.81 | 359.3 | 344.02 | 353.57 | 1,364.70 |
| 54 | 1,744.17 | 1,871.05 | 1,978.34 | 2,047.19 | 7,640.75 |
| 56 | 637.58 | 727.57 | 743.93 | 746.32 | 2,855.40 |
| 57 | 863.88 | 867.59 | 892.91 | 948.18 | 3,572.56 |
| 58 | 354.71 | 346.23 | 370 | 382.37 | 1,453.31 |
| TOTAL | 4,990.87 | 5,326.37 | 5,530.96 | 5,690.02 | 21,538.22 |

STATE: VIRGIN ISLANDS

| ZIP CODE | QUARTER 1 | QUARTER 2 | QUARTER 3 | QUARTER 4 | TOTAL GRAMS |
|---|---|---|---|---|---|
| 8 | 24.39 | 25.31 | 18.9 | 20.37 | 88.97 |
| TOTAL | 24.39 | 25.31 | 18.9 | 20.37 | 88.97 |

STATE: VIRGINIA

| ZIP CODE | QUARTER 1 | QUARTER 2 | QUARTER 3 | QUARTER 4 | TOTAL GRAMS |
|---|---|---|---|---|---|
| 201 | 1,391.23 | 1,465.73 | 1,451.04 | 1,418.55 | 5,726.55 |
| 220 | 810.5 | 800.96 | 849.32 | 845.75 | 3,306.53 |
| 221 | 521.84 | 540.86 | 552.04 | 568.62 | 2,183.36 |
| 222 | 164.22 | 158.99 | 193.53 | 330.16 | 846.9 |
| 223 | 221.46 | 226.07 | 205.57 | 209.24 | 862.34 |
| 224 | 440.21 | 449.98 | 453.62 | 452.69 | 1,796.50 |
| 225 | 195.96 | 199.11 | 186.11 | 203.83 | 785.01 |
| 226 | 1,001.68 | 1,025.12 | 1,110.93 | 1,097.97 | 4,235.70 |
| 227 | 129.09 | 140.47 | 148.9 | 151.01 | 569.47 |
| 228 | 176.11 | 186.34 | 205.8 | 210.86 | 779.11 |
| 229 | 356.77 | 373.49 | 384.77 | 366.71 | 1,481.74 |
| 230 | 297.42 | 318.19 | 304.69 | 318.63 | 1,238.93 |
| 231 | 924.83 | 936.59 | 901.17 | 938.44 | 3,701.03 |
| 232 | 919.46 | 1,002.58 | 1,067.25 | 1,018.20 | 4,007.49 |
| 233 | 313.66 | 356.81 | 328.46 | 302.35 | 1,301.28 |
| 234 | 601.87 | 632.72 | 674.84 | 668.56 | 2,577.99 |
| 235 | 129.19 | 130.81 | 113.86 | 124.21 | 498.07 |
| 236 | 574.87 | 635.86 | 613.88 | 637.04 | 2,461.65 |
| 237 | 128.32 | 102.98 | 107.51 | 88.57 | 427.38 |
| 238 | 401.42 | 407.79 | 421.12 | 396.95 | 1,627.28 |

DRUG CODE:9064DRUG NAME:BUPRENORPHINE
STATE:VIRGINIA

| ZIP CODE | QUARTER 1 | QUARTER 2 | QUARTER 3 | QUARTER 4 | TOTAL GRAMS |
|---|---|---|---|---|---|
| 239 | 119.08 | 130.12 | 114.07 | 121.64 | 484.91 |
| 240 | 1,656.63 | 1,801.07 | 1,876.72 | 1,837.22 | 7,171.64 |
| 241 | 746.61 | 760.5 | 775.52 | 746 | 3,028.63 |
| 242 | 3,657.48 | 3,661.69 | 3,755.71 | 3,809.18 | 14,884.06 |
| 243 | 936.35 | 960.93 | 844.59 | 896.72 | 3,638.59 |
| 244 | 157.79 | 163.39 | 153.16 | 161.66 | 636 |
| 245 | 616.54 | 732.99 | 717.81 | 650.44 | 2,717.78 |
| 246 | 917.88 | 930.61 | 952.86 | 994.48 | 3,795.83 |
| TOTAL | 18,508.47 | 19,232.75 | 19,464.85 | 19,565.68 | 76,771.75 |

STATE: WASHINGTON

| ZIP CODE | QUARTER 1 | QUARTER 2 | QUARTER 3 | QUARTER 4 | TOTAL GRAMS |
|---|---|---|---|---|---|
| 980 | 3,153.61 | 3,199.21 | 3,270.40 | 3,249.54 | 12,872.76 |
| 981 | 1,723.52 | 1,914.93 | 1,988.64 | 1,971.90 | 7,598.99 |
| 982 | 3,015.80 | 3,041.17 | 3,247.75 | 3,327.55 | 12,632.27 |
| 983 | 1,366 | 1,476.55 | 1,520.30 | 1,570.01 | 5,932.86 |
| 984 | 999.44 | 1,053.90 | 1,101.39 | 1,172.98 | 4,327.71 |

| | | | | | |
|---|---|---|---|---|---|
| 985 | 1,125.46 | 1,208.46 | 1,372.26 | 1,419.21 | 5,125.39 |
| 986 | 926.57 | 1,057.67 | 1,081.26 | 1,159.34 | 4,224.84 |
| 988 | 334.47 | 357.8 | 377.21 | 441.1 | 1,510.58 |
| 989 | 440.68 | 457.02 | 508.1 | 544.85 | 1,950.65 |
| 990 | 69.69 | 86.63 | 91.94 | 91 | 339.26 |
| 991 | 229.88 | 257.18 | 296.13 | 306.47 | 1,089.66 |
| 992 | 1,253.66 | 1,206.27 | 1,275.72 | 1,427.39 | 5,163.04 |
| 993 | 660.23 | 660.87 | 671.52 | 716.41 | 2,709.03 |
| 994 | 117.86 | 145.21 | 152.21 | 140.62 | 555.9 |
| TOTAL | 15,416.87 | 16,122.87 | 16,954.83 | 17,538.37 | 66,032.94 |

STATE: WEST VIRGINIA

| ZIP CODE | QUARTER 1 | QUARTER 2 | QUARTER 3 | QUARTER 4 | TOTAL GRAMS |
|---|---|---|---|---|---|
| 247 | 251.74 | 226.76 | 196.13 | 195.46 | 870.09 |
| 248 | 389.55 | 393.17 | 415.24 | 448.23 | 1,646.19 |
| 249 | 183.02 | 170.73 | 132.48 | 143.25 | 629.48 |
| 250 | 645.34 | 780.05 | 650.18 | 671.45 | 2,747.02 |
| 251 | 331.49 | 309.2 | 312.92 | 311.36 | 1,264.97 |
| 252 | 298.05 | 318.81 | 313.61 | 311.55 | 1,242.02 |
| 253 | 1,137.57 | 1,214.20 | 1,192.20 | 1,329.59 | 4,873.56 |
| 254 | 1,001.01 | 981.08 | 1,024.81 | 933.37 | 3,940.27 |
| 255 | 1,434.78 | 1,447 | 1,416.41 | 1,443.36 | 5,741.55 |
| 256 | 1,047.09 | 1,099.16 | 1,211.90 | 1,246.67 | 4,604.82 |
| 257 | 1,089.64 | 1,092.87 | 1,075.74 | 1,008.89 | 4,267.14 |
| 258 | 983.67 | 1,119.85 | 1,232.98 | 1,502.24 | 4,838.74 |
| 259 | 316.83 | 309.16 | 316.7 | 329.03 | 1,271.72 |
| 260 | 1,777.70 | 1,749.45 | 1,695.28 | 1,600.22 | 6,822.65 |
| 261 | 463.88 | 478.48 | 523.9 | 526.09 | 1,992.35 |
| 262 | 489.71 | 477.06 | 470.08 | 496.6 | 1,933.45 |
| 263 | 572.87 | 611.41 | 668.8 | 727.43 | 2,580.51 |
| 264 | 107.61 | 117.83 | 110.65 | 125.26 | 461.35 |
| 265 | 1,496.64 | 1,395.38 | 1,409.01 | 1,311.70 | 5,612.73 |
| 266 | 93.6 | 87.44 | 93.87 | 100.93 | 375.84 |
| 267 | 101.79 | 114.57 | 95.38 | 100.86 | 412.6 |
| 268 | 20.42 | 22.93 | 19.21 | 17.6 | 80.16 |
| TOTAL | 14,234 | 14,516.59 | 14,577.48 | 14,881.14 | 58,209.21 |

STATE: WISCONSIN

| ZIP CODE | QUARTER 1 | QUARTER 2 | QUARTER 3 | QUARTER 4 | TOTAL GRAMS |
|---|---|---|---|---|---|
| 530 | 1,057.38 | 1,064.59 | 1,108.09 | 1,102.25 | 4,332.31 |
| 531 | 1,169.73 | 1,161.37 | 1,200.56 | 1,179.17 | 4,710.83 |
| 532 | 2,055.30 | 2,124.67 | 2,263.63 | 2,329.10 | 8,772.70 |
| 534 | 225.47 | 205.16 | 197.34 | 206.69 | 834.66 |
| 535 | 748.63 | 774.51 | 765.24 | 775.27 | 3,063.65 |
| 537 | 634.06 | 656.55 | 684.4 | 663.9 | 2,638.91 |
| 538 | 20.12 | 26.12 | 23.72 | 23.1 | 93.06 |
| 539 | 324.26 | 328.87 | 320.83 | 284.2 | 1,258.16 |
| 540 | 70.58 | 84.69 | 78.34 | 83.34 | 316.95 |
| 541 | 207.12 | 212.99 | 215.36 | 236.91 | 872.38 |
| 542 | 109.27 | 113.89 | 112.29 | 97.96 | 433.41 |
| 543 | 173.19 | 206.16 | 206.42 | 216.49 | 802.26 |
| 544 | 450.5 | 461.47 | 419.65 | 420.12 | 1,751.74 |

DRUG CODE:9064DRUG NAME:BUPRENORPHINE
STATE:WISCONSIN

| ZIP CODE | QUARTER 1 | QUARTER 2 | QUARTER 3 | QUARTER 4 | TOTAL GRAMS |
|---|---|---|---|---|---|
| 545 | 199.84 | 217.04 | 228.29 | 208.4 | 853.57 |
| 546 | 383.44 | 384.35 | 391.2 | 389.18 | 1,548.17 |
| 547 | 202.67 | 201.11 | 202.92 | 198.27 | 804.97 |
| 548 | 177.5 | 192.64 | 186.53 | 176.39 | 733.06 |

| | | | | | |
|---|---|---|---|---|---|
| 549 | 913.68 | 945.45 | 953.23 | 916.6 | 3,728.96 |
| TOTAL | 9,122.74 | 9,361.63 | 9,558.04 | 9,507.34 | 37,549.75 |

STATE: WYOMING

| ZIP CODE | QUARTER 1 | QUARTER 2 | QUARTER 3 | QUARTER 4 | TOTAL GRAMS |
|---|---|---|---|---|---|
| 820 | 153.05 | 279.46 | 238.53 | 283.02 | 954.06 |
| 822 | 4.06 | 5.66 | 4.39 | 6.05 | 20.16 |
| 823 | 16.32 | 19.86 | 18.38 | 13.08 | 67.64 |
| 824 | 90.27 | 90.37 | 80.66 | 102.59 | 363.89 |
| 825 | 42.41 | 53.93 | 59.99 | 71.96 | 228.29 |
| 826 | 249.98 | 284.21 | 284.78 | 299.84 | 1,118.81 |
| 827 | 82.13 | 76.98 | 76.59 | 75.94 | 311.64 |
| 828 | 60.86 | 60.91 | 40.93 | 40.9 | 203.6 |
| 829 | 134.92 | 114.49 | 128.6 | 145.05 | 523.06 |
| 830 | 28.75 | 40.96 | 43.55 | 37.66 | 150.92 |
| 831 | 18.26 | 18.61 | 20.07 | 17.38 | 74.32 |
| TOTAL | 881.01 | 1,045.44 | 996.47 | 1,093.47 | 4,016.39 |

DRUG CODE:9120DRUG NAME:DIHYDROCODEINE

STATE: ALABAMA

| ZIP CODE | QUARTER 1 | QUARTER 2 | QUARTER 3 | QUARTER 4 | TOTAL GRAMS |
|---|---|---|---|---|---|
| 350 | 118.01 | 101.9 | 138.75 | 139.02 | 497.68 |
| 351 | 24.78 | 26.12 | 32.26 | 34.07 | 117.23 |
| 352 | 43.97 | 44.18 | 40.86 | 48.49 | 177.5 |
| 354 | 103.13 | 121.88 | 133.15 | 146.07 | 504.23 |
| 355 | 33.86 | 33.75 | 33.22 | 45.5 | 146.33 |
| 356 | 119.05 | 83.42 | 169.41 | 129.89 | 501.77 |
| 357 | 14.42 | 30.23 | 27.45 | 35.14 | 107.24 |
| 358 | 12.55 | 12.18 | 21.15 | 17.25 | 63.13 |
| 359 | 25.69 | 26.76 | 41.6 | 32.53 | 126.58 |
| 360 | 13.99 | 11.75 | 11.26 | 5.34 | 42.34 |
| 361 | 20.3 | 12.18 | 15.69 | 20.8 | 68.97 |
| 362 | 20.67 | 15.38 | 22.54 | 20.19 | 78.78 |
| 363 | 23.18 | 14.95 | 9.93 | 12.92 | 60.98 |
| 364 | 0.11 | 0 | 2.35 | 3.2 | 5.66 |
| 365 | 66.3 | 45.74 | 37.65 | 27.98 | 177.67 |
| 366 | 16.44 | 20.51 | 21.63 | 24.12 | 82.7 |
| 367 | 5.34 | 11.75 | 7.48 | 16.02 | 40.59 |
| 368 | 7.8 | 8.55 | 5.87 | 10.04 | 32.26 |
| TOTAL | 669.59 | 621.23 | 772.25 | 768.57 | 2,831.64 |

STATE: ALASKA

| ZIP CODE | QUARTER 1 | QUARTER 2 | QUARTER 3 | QUARTER 4 | TOTAL GRAMS |
|---|---|---|---|---|---|
| 996 | 0 | 0 | 1.07 | 0 | 1.07 |
| TOTAL | 0 | 0 | 1.07 | 0 | 1.07 |

STATE: ARIZONA

| ZIP CODE | QUARTER 1 | QUARTER 2 | QUARTER 3 | QUARTER 4 | TOTAL GRAMS |
|---|---|---|---|---|---|
| 850 | 4.27 | 4.27 | 21.58 | 25.21 | 55.33 |
| 852 | 43.79 | 33.11 | 39.52 | 44.76 | 161.18 |
| 857 | 1.07 | 0 | 1.07 | 0 | 2.14 |
| 864 | 0 | 2.24 | 0 | 1.07 | 3.31 |
| TOTAL | 49.13 | 39.62 | 62.17 | 71.04 | 221.96 |

STATE: ARKANSAS

| ZIP CODE | QUARTER 1 | QUARTER 2 | QUARTER 3 | QUARTER 4 | TOTAL GRAMS |
|---|---|---|---|---|---|
| 716 | 0 | 0 | 0 | 1.17 | 1.17 |
| 717 | 0 | 2.14 | 2.14 | 1.07 | 5.35 |
| 718 | 17.09 | 11.84 | 5.34 | 18.37 | 52.64 |

| | | | | | |
|---|---|---|---|---|---|
| 719 | 1.07 | 2.14 | 4.27 | 15.86 | 23.34 |
| 720 | 7.48 | 6.41 | 4.27 | 12.28 | 30.44 |
| 721 | 2.14 | 0 | 0 | 7.58 | 9.72 |
| 722 | 0 | 0 | 0 | 0.69 | 0.69 |
| 723 | 20.41 | 12.76 | 7.05 | 5.39 | 45.61 |
| 724 | 24.09 | 9.5 | 21.52 | 10.84 | 65.95 |
| 727 | 8.82 | 7.48 | 8.12 | 5.34 | 29.76 |
| 728 | 0 | 1.07 | 1.07 | 0 | 2.14 |

DRUG CODE:9120DRUG NAME:DIHYDROCODEINE
STATE:ARKANSAS

| ZIP CODE | QUARTER 1 | QUARTER 2 | QUARTER 3 | QUARTER 4 | TOTAL GRAMS |
|---|---|---|---|---|---|
| 729 | 23.8 | 44.21 | 30.34 | 24.57 | 122.92 |
| TOTAL | 104.9 | 97.55 | 84.12 | 103.16 | 389.73 |

STATE: CALIFORNIA

| ZIP CODE | QUARTER 1 | QUARTER 2 | QUARTER 3 | QUARTER 4 | TOTAL GRAMS |
|---|---|---|---|---|---|
| 900 | 5.34 | 50.84 | 16.66 | 19.23 | 92.07 |
| 902 | 44.22 | 34.72 | 67.29 | 41.66 | 187.89 |
| 906 | 0 | 0 | 1.07 | 0 | 1.07 |
| 907 | 0 | 1.28 | 8.01 | 0 | 9.29 |
| 908 | 0 | 0 | 0 | 9.61 | 9.61 |
| 910 | 0 | 7.48 | 0 | 0 | 7.48 |
| 913 | 0 | 1,090.80 | 96.78 | 28.84 | 1,216.42 |
| 914 | 0 | 28.84 | 7.69 | 0 | 36.53 |
| 917 | 108.95 | 138.86 | 100.41 | 125.19 | 473.41 |
| 920 | 14.95 | 6.41 | 6.41 | 18.16 | 45.93 |
| 921 | 2.14 | 1.07 | 9.19 | 9.72 | 22.12 |
| 922 | 0 | 4.49 | 0 | 0 | 4.49 |
| 923 | 0 | 6.41 | 1.6 | 3.52 | 11.53 |
| 924 | 0 | 19.23 | 6.41 | 9.61 | 35.25 |
| 925 | 1.07 | 1.07 | 2.03 | 2.14 | 6.31 |
| 926 | 1.07 | 21.36 | 7.48 | 5.77 | 35.68 |
| 927 | 8.01 | 21.9 | 4.81 | 17.52 | 52.24 |
| 928 | 145.8 | 178.38 | 117.5 | 108.53 | 550.21 |
| 930 | 73.7 | 80.11 | 139.07 | 70.5 | 363.38 |
| 931 | 83.32 | 57.68 | 32.04 | 6.41 | 179.45 |
| 933 | 122.84 | 182.12 | 143.67 | 119.85 | 568.48 |
| 934 | 132.45 | 131.38 | 85.45 | 55.44 | 404.72 |
| 935 | 1.07 | 12.82 | 6.41 | 14.42 | 34.72 |
| 936 | 0 | 0 | 1.07 | 1.07 | 2.14 |
| 937 | 0 | 9.61 | 0 | 1.07 | 10.68 |
| 940 | 1.07 | 0 | 0 | 5.13 | 6.2 |
| 941 | 12.82 | 13.89 | 14.95 | 9.61 | 51.27 |
| 944 | 0 | 0 | 8.97 | 13.46 | 22.43 |
| 945 | 1.07 | 4.81 | 5.87 | 3.2 | 14.95 |
| 946 | 0 | 0 | 1.07 | 2.14 | 3.21 |
| 950 | 0 | 12.82 | 17.3 | 19.23 | 49.35 |
| 951 | 0 | 0 | 0 | 1.07 | 1.07 |
| 952 | 0 | 0 | 1.07 | 2.14 | 3.21 |
| 953 | 10.25 | 22.43 | 10.9 | 9.61 | 53.19 |
| 954 | 1.07 | 0 | 1.07 | 3.2 | 5.34 |
| 956 | 3.2 | 1.07 | 2.14 | 1.07 | 7.48 |
| 957 | 0 | 0 | 1.07 | 1.07 | 2.14 |
| 958 | 10.68 | 20.3 | 19.01 | 20.51 | 70.5 |
| 959 | 0 | 0 | 1.07 | 0 | 1.07 |
| TOTAL | 785.09 | 2,162.18 | 945.54 | 759.7 | 4,652.51 |

STATE: COLORADO

| ZIP CODE | QUARTER 1 | QUARTER 2 | QUARTER 3 | QUARTER 4 | TOTAL GRAMS |
|---|---|---|---|---|---|

| | | | | | |
|---|---|---|---|---|---|
| 806 | 0 | 0 | 1.07 | 0 | 1.07 |
| 811 | 0 | 1.07 | 0 | 0 | 1.07 |
| 812 | 0 | 1.07 | 0 | 0 | 1.07 |
| TOTAL | 0 | 2.14 | 1.07 | 0 | 3.21 |

STATE: CONNECTICUT

| ZIP CODE | QUARTER 1 | QUARTER 2 | QUARTER 3 | QUARTER 4 | TOTAL GRAMS |
|---|---|---|---|---|---|
| 68 | 0 | 0 | 0 | 3.2 | 3.2 |
| TOTAL | 0 | 0 | 0 | 3.2 | 3.2 |

STATE: DELAWARE

| ZIP CODE | QUARTER 1 | QUARTER 2 | QUARTER 3 | QUARTER 4 | TOTAL GRAMS |
|---|---|---|---|---|---|
| 198 | 0 | 1.07 | 0 | 0 | 1.07 |
| 199 | 0 | 0.32 | 1.07 | 0 | 1.39 |
| TOTAL | 0 | 1.39 | 1.07 | 0 | 2.46 |

STATE: FLORIDA

| ZIP CODE | QUARTER 1 | QUARTER 2 | QUARTER 3 | QUARTER 4 | TOTAL GRAMS |
|---|---|---|---|---|---|
| 321 | 0 | 0 | 17.3 | 6.41 | 23.71 |
| 323 | 0 | 0 | 0 | 2.14 | 2.14 |
| 324 | 3.74 | 6.41 | 1.07 | 2.78 | 14 |
| 325 | 8.71 | 9.73 | 16.45 | 26.81 | 61.7 |
| 326 | 1.07 | 0 | 0 | 0 | 1.07 |

DRUG CODE:9120DRUG NAME:DIHYDROCODEINE
STATE:FLORIDA

| ZIP CODE | QUARTER 1 | QUARTER 2 | QUARTER 3 | QUARTER 4 | TOTAL GRAMS |
|---|---|---|---|---|---|
| 327 | 1.07 | 1.28 | 2.14 | 0 | 4.49 |
| 328 | 24.57 | 43.79 | 22.43 | 18.16 | 108.95 |
| 330 | 2.14 | 1.07 | 0 | 6.68 | 9.89 |
| 331 | 0 | 14.95 | 10.68 | 19.01 | 44.64 |
| 333 | 0 | 3.2 | 0 | 2.24 | 5.44 |
| 334 | 0.21 | 1.07 | 1.07 | 1.07 | 3.42 |
| 335 | 2.14 | 2.14 | 0 | 1.07 | 5.35 |
| 336 | 13.89 | 5.34 | 13.35 | 14.95 | 47.53 |
| 337 | 1.07 | 1.07 | 1.07 | 0 | 3.21 |
| 338 | 2.14 | 0 | 0 | 0 | 2.14 |
| 339 | 1.07 | 0 | 2.14 | 1.07 | 4.28 |
| 341 | 2.14 | 3.2 | 0 | 1.07 | 6.41 |
| 342 | 3.2 | 5.34 | 0 | 0 | 8.54 |
| 347 | 0 | 0 | 0 | 1.07 | 1.07 |
| 349 | 0 | 0.53 | 0 | 5.98 | 6.51 |
| TOTAL | 67.16 | 99.12 | 87.7 | 110.51 | 364.49 |

STATE: GEORGIA

| ZIP CODE | QUARTER 1 | QUARTER 2 | QUARTER 3 | QUARTER 4 | TOTAL GRAMS |
|---|---|---|---|---|---|
| 300 | 45.4 | 48.49 | 33.01 | 35.52 | 162.42 |
| 301 | 11.75 | 3.74 | 3.2 | 6.94 | 25.63 |
| 302 | 13.14 | 112.9 | 95.07 | 85.4 | 306.51 |
| 303 | 18.05 | 13.02 | 16.08 | 9.51 | 56.66 |
| 304 | 9.08 | 1.82 | 8.76 | 81.71 | 101.37 |
| 305 | 58.8 | 35.84 | 34.45 | 35.25 | 164.34 |
| 306 | 23.4 | 15.43 | 8.55 | 13.89 | 61.27 |
| 307 | 3.2 | 3.2 | 2.14 | 3.2 | 11.74 |
| 309 | 0 | 0 | 0.21 | 1.07 | 1.28 |
| 310 | 38.35 | 32.08 | 31.78 | 20.67 | 122.88 |
| 312 | 39.2 | 21.58 | 20.2 | 18.53 | 99.51 |
| 313 | 0 | 1.28 | 1.17 | 0.64 | 3.09 |
| 314 | 3.42 | 13.89 | 17.84 | 19.92 | 55.07 |

| | | | | | |
|---|---|---|---|---|---|
| 315 | 4.27 | 2.14 | 8.76 | 6.41 | 21.58 |
| 316 | 2.61 | 2.14 | 0 | 1.07 | 5.82 |
| 317 | 106.28 | 91.48 | 96.56 | 69.38 | 363.7 |
| 318 | 0 | 0 | 0 | 6.41 | 6.41 |
| 319 | 0.43 | 0 | 1.07 | 1.07 | 2.57 |
| 398 | 9.53 | 18.21 | 9.29 | 5.34 | 42.37 |
| TOTAL | 386.91 | 417.24 | 388.14 | 421.93 | 1,614.22 |

STATE: HAWAII

| ZIP CODE | QUARTER 1 | QUARTER 2 | QUARTER 3 | QUARTER 4 | TOTAL GRAMS |
|---|---|---|---|---|---|
| 967 | 1.71 | 1.92 | 6.09 | 0.32 | 10.04 |
| 968 | 5.34 | 2.78 | 3.85 | 2.56 | 14.53 |
| TOTAL | 7.05 | 4.7 | 9.94 | 2.88 | 24.57 |

STATE: IDAHO

| ZIP CODE | QUARTER 1 | QUARTER 2 | QUARTER 3 | QUARTER 4 | TOTAL GRAMS |
|---|---|---|---|---|---|
| 836 | 15.49 | 10.68 | 19.23 | 51.27 | 96.67 |
| 837 | 11.75 | 94 | 10.68 | 0 | 116.43 |
| TOTAL | 27.24 | 104.68 | 29.91 | 51.27 | 213.1 |

STATE: ILLINOIS

| ZIP CODE | QUARTER 1 | QUARTER 2 | QUARTER 3 | QUARTER 4 | TOTAL GRAMS |
|---|---|---|---|---|---|
| 600 | 8.55 | 10.68 | 4.27 | 3.2 | 26.7 |
| 601 | 0 | 1.07 | 0 | 6.41 | 7.48 |
| 603 | 0 | 0 | 0 | 28.84 | 28.84 |
| 604 | 1.07 | 7.48 | 22.43 | 44.22 | 75.2 |
| 606 | 0 | 1.07 | 2.14 | 51.91 | 55.12 |
| 608 | 0 | 1.07 | 0 | 0 | 1.07 |
| 617 | 1.07 | 0 | 0 | 0 | 1.07 |
| 618 | 0 | 1.07 | 0 | 1.07 | 2.14 |
| 619 | 0 | 1.07 | 0 | 0 | 1.07 |
| 620 | 0 | 0 | 1.07 | 0 | 1.07 |
| 622 | 0.28 | 0 | 0 | 0 | 0.28 |
| 628 | 3.2 | 0 | 0 | 0 | 3.2 |
| 629 | 4.27 | 1.07 | 1.07 | 4.27 | 10.68 |
| TOTAL | 18.44 | 24.58 | 30.98 | 139.92 | 213.92 |

STATE: INDIANA

| ZIP CODE | QUARTER 1 | QUARTER 2 | QUARTER 3 | QUARTER 4 | TOTAL GRAMS |
|---|---|---|---|---|---|

DRUG CODE:9120DRUG NAME:DIHYDROCODEINE
STATE:INDIANA

| ZIP CODE | QUARTER 1 | QUARTER 2 | QUARTER 3 | QUARTER 4 | TOTAL GRAMS |
|---|---|---|---|---|---|
| 460 | 0 | 4.27 | 0 | 1.07 | 5.34 |
| 463 | 12.82 | 1.07 | 1.07 | 0 | 14.96 |
| 467 | 0 | 0 | 1.07 | 0 | 1.07 |
| 476 | 0 | 4.27 | 2.14 | 0 | 6.41 |
| 477 | 4.81 | 24.99 | 9.61 | 1.07 | 40.48 |
| TOTAL | 17.63 | 34.6 | 13.89 | 2.14 | 68.26 |

STATE: KANSAS

| ZIP CODE | QUARTER 1 | QUARTER 2 | QUARTER 3 | QUARTER 4 | TOTAL GRAMS |
|---|---|---|---|---|---|
| 660 | 1.07 | 1.07 | 0 | 0 | 2.14 |
| 662 | 0 | 0 | 5.34 | 0 | 5.34 |
| TOTAL | 1.07 | 1.07 | 5.34 | 0 | 7.48 |

STATE: KENTUCKY

| ZIP CODE | QUARTER 1 | QUARTER 2 | QUARTER 3 | QUARTER 4 | TOTAL GRAMS |
|---|---|---|---|---|---|
| 400 | 5.34 | 6.41 | 0 | 0 | 11.75 |

| | | | | | |
|---|---|---|---|---|---|
| 401 | 0 | 0 | 0 | 8.55 | 8.55 |
| 402 | 4.27 | 2.14 | 0.21 | 2.14 | 8.76 |
| 405 | 0 | 0 | 0 | 1.07 | 1.07 |
| 406 | 0 | 0 | 0 | 1.07 | 1.07 |
| 409 | 0 | 4.27 | 0 | 5.34 | 9.61 |
| 410 | 4.27 | 2.14 | 2.14 | 0 | 8.55 |
| 420 | 1.07 | 1.07 | 1.07 | 0 | 3.21 |
| 421 | 4.27 | 1.07 | 0 | 0 | 5.34 |
| 422 | 1.07 | 1.07 | 1.07 | 5.34 | 8.55 |
| 424 | 0 | 0 | 1.07 | 1.07 | 2.14 |
| 426 | 0 | 0 | 2.14 | 2.14 | 4.28 |
| 427 | 13.89 | 14.95 | 11.75 | 7.48 | 48.07 |
| TOTAL | 34.18 | 33.12 | 19.45 | 34.2 | 120.95 |

STATE: LOUISIANA

| ZIP CODE | QUARTER 1 | QUARTER 2 | QUARTER 3 | QUARTER 4 | TOTAL GRAMS |
|---|---|---|---|---|---|
| 700 | 21.05 | 28.05 | 101.74 | 67.4 | 218.24 |
| 701 | 7.16 | 6.62 | 2.08 | 1.98 | 17.84 |
| 703 | 58 | 45.82 | 49.78 | 72.9 | 226.5 |
| 704 | 24.99 | 35.19 | 35.2 | 54.74 | 150.12 |
| 705 | 129.94 | 343.31 | 411.88 | 458.72 | 1,343.85 |
| 706 | 8.55 | 8.6 | 10.68 | 7.48 | 35.31 |
| 707 | 15.11 | 15.33 | 20.56 | 2.51 | 53.51 |
| 708 | 27.08 | 30.07 | 12.76 | 13.67 | 83.58 |
| 710 | 3.31 | 7.8 | 8.55 | 4.27 | 23.93 |
| 711 | 26.69 | 26.3 | 23.83 | 20.4 | 97.22 |
| 712 | 24.62 | 13.35 | 17.62 | 13.03 | 68.62 |
| 713 | 2.14 | 3.2 | 1.07 | 2.14 | 8.55 |
| 714 | 1.07 | 2.14 | 0 | 0 | 3.21 |
| TOTAL | 349.71 | 565.78 | 695.75 | 719.24 | 2,330.48 |

STATE: MAINE

| ZIP CODE | QUARTER 1 | QUARTER 2 | QUARTER 3 | QUARTER 4 | TOTAL GRAMS |
|---|---|---|---|---|---|
| 40 | 0 | 0 | 1.07 | 0 | 1.07 |
| TOTAL | 0 | 0 | 1.07 | 0 | 1.07 |

STATE: MARYLAND

| ZIP CODE | QUARTER 1 | QUARTER 2 | QUARTER 3 | QUARTER 4 | TOTAL GRAMS |
|---|---|---|---|---|---|
| 207 | 1.07 | 0 | 0 | 0 | 1.07 |
| 208 | 0 | 0 | 3.2 | 0 | 3.2 |
| 209 | 0 | 0 | 1.07 | 0 | 1.07 |
| 210 | 0 | 1.07 | 2.14 | 1.07 | 4.28 |
| 211 | 0 | 0 | 3.2 | 2.14 | 5.34 |
| 212 | 0 | 0 | 6.62 | 3.2 | 9.82 |
| TOTAL | 1.07 | 1.07 | 16.23 | 6.41 | 24.78 |

STATE: MASSACHUSETTS

| ZIP CODE | QUARTER 1 | QUARTER 2 | QUARTER 3 | QUARTER 4 | TOTAL GRAMS |
|---|---|---|---|---|---|
| 18 | 19.23 | 12.82 | 9.61 | 9.61 | 51.27 |
| TOTAL | 19.23 | 12.82 | 9.61 | 9.61 | 51.27 |

STATE: MICHIGAN

| ZIP CODE | QUARTER 1 | QUARTER 2 | QUARTER 3 | QUARTER 4 | TOTAL GRAMS |
|---|---|---|---|---|---|
| 480 | 91.86 | 1.07 | 0 | 2.14 | 95.07 |
| 481 | 1.07 | 0 | 0 | 0 | 1.07 |
| 482 | 1.07 | 0 | 0 | 0 | 1.07 |
| 483 | 2.14 | 0 | 0 | 0 | 2.14 |

RETAIL DRUG DISTRIBUTION BY ZIP CODE WITHIN STATE BY GRAMS WT

DRUG CODE:9120DRUG NAME:DIHYDROCODEINE
STATE:MICHIGAN

| ZIP CODE | QUARTER 1 | QUARTER 2 | QUARTER 3 | QUARTER 4 | TOTAL GRAMS |
|---|---|---|---|---|---|
| 488 | 0 | 0 | 1.07 | 6.41 | 7.48 |
| 496 | 0 | 0 | 2.14 | 0 | 2.14 |
| TOTAL | 96.14 | 1.07 | 3.21 | 8.55 | 108.97 |

STATE: MINNESOTA

| ZIP CODE | QUARTER 1 | QUARTER 2 | QUARTER 3 | QUARTER 4 | TOTAL GRAMS |
|---|---|---|---|---|---|
| 551 | 0 | 0 | 1.07 | 0 | 1.07 |
| 554 | 0 | 0 | 2.14 | 0 | 2.14 |
| TOTAL | 0 | 0 | 3.21 | 0 | 3.21 |

STATE: MISSISSIPPI

| ZIP CODE | QUARTER 1 | QUARTER 2 | QUARTER 3 | QUARTER 4 | TOTAL GRAMS |
|---|---|---|---|---|---|
| 386 | 29.03 | 27.67 | 31.67 | 36.48 | 124.85 |
| 387 | 4.27 | 4.27 | 12.6 | 19.23 | 40.37 |
| 388 | 21.31 | 26.01 | 19.49 | 20.19 | 87 |
| 389 | 3.95 | 2.14 | 13.15 | 20.56 | 39.8 |
| 390 | 36.42 | 41.77 | 67.51 | 55.01 | 200.71 |
| 391 | 24.57 | 35.6 | 44.44 | 31.99 | 136.6 |
| 392 | 37.07 | 46.14 | 64.61 | 69.32 | 217.14 |
| 393 | 13.67 | 18.27 | 28.84 | 11.96 | 72.74 |
| 394 | 20.94 | 13.03 | 32.31 | 17.09 | 83.37 |
| 395 | 25.42 | 19.23 | 26.44 | 15.97 | 87.06 |
| 396 | 4.27 | 10.68 | 6.41 | 6.41 | 27.77 |
| 397 | 1.07 | 2.14 | 16.45 | 16.66 | 36.32 |
| TOTAL | 221.99 | 246.95 | 363.92 | 320.87 | 1,153.73 |

STATE: MISSOURI

| ZIP CODE | QUARTER 1 | QUARTER 2 | QUARTER 3 | QUARTER 4 | TOTAL GRAMS |
|---|---|---|---|---|---|
| 631 | 13.89 | 8.55 | 4.27 | 10.68 | 37.39 |
| 633 | 4.27 | 3.2 | 8.55 | 3.2 | 19.22 |
| 636 | 2.14 | 0 | 0 | 0 | 2.14 |
| 641 | 0 | 0 | 5.34 | 0 | 5.34 |
| 646 | 0 | 3.2 | 1.07 | 2.14 | 6.41 |
| 658 | 0 | 1.07 | 0 | 0 | 1.07 |
| TOTAL | 20.3 | 16.02 | 19.23 | 16.02 | 71.57 |

STATE: NEBRASKA

| ZIP CODE | QUARTER 1 | QUARTER 2 | QUARTER 3 | QUARTER 4 | TOTAL GRAMS |
|---|---|---|---|---|---|
| 685 | 2.14 | 1.07 | 3.2 | 0 | 6.41 |
| TOTAL | 2.14 | 1.07 | 3.2 | 0 | 6.41 |

STATE: NEVADA

| ZIP CODE | QUARTER 1 | QUARTER 2 | QUARTER 3 | QUARTER 4 | TOTAL GRAMS |
|---|---|---|---|---|---|
| 890 | 1.6 | 4.86 | 0 | 0 | 6.46 |
| 891 | 35.84 | 75.84 | 67.29 | 16.13 | 195.1 |
| 895 | 0 | 0 | 0 | 1.07 | 1.07 |
| TOTAL | 37.44 | 80.7 | 67.29 | 17.2 | 202.63 |

STATE: NEW HAMPSHIRE

| ZIP CODE | QUARTER 1 | QUARTER 2 | QUARTER 3 | QUARTER 4 | TOTAL GRAMS |
|---|---|---|---|---|---|
| 31 | 0 | 0 | 1.07 | 0 | 1.07 |
| TOTAL | 0 | 0 | 1.07 | 0 | 1.07 |

STATE: NEW JERSEY

| ZIP CODE | QUARTER 1 | QUARTER 2 | QUARTER 3 | QUARTER 4 | TOTAL GRAMS |
|---|---|---|---|---|---|
| 70 | 0 | 0 | 13.86 | 24.18 | 38.04 |
| 71 | 0 | 0 | 0 | 123.91 | 123.91 |
| 72 | 0 | 0 | 0.09 | 0 | 0.09 |

| | | | | | |
|---|---|---|---|---|---|
| 73 | 0 | 0 | 0.68 | 2.09 | 2.77 |
| 74 | 0 | 0 | 2.03 | 5.3 | 7.33 |
| 76 | 0 | 2.14 | 4.27 | 1.92 | 8.33 |
| 77 | 0 | 5.23 | 1.19 | 7.37 | 13.79 |
| 78 | 0 | 0 | 0.51 | 0 | 0.51 |
| 79 | 0 | 0 | 0 | 2.82 | 2.82 |
| 80 | 2.67 | 34.5 | 11.75 | 13.9 | 62.82 |
| 81 | 0 | 0 | 0 | 1.07 | 1.07 |
| 82 | 1.39 | 7.48 | 5.77 | 2.03 | 16.67 |
| 83 | 1.07 | 2.24 | 2.14 | 1.2 | 6.65 |
| 84 | 0.16 | 0 | 0 | 0 | 0.16 |
| 85 | 0 | 0 | 0.21 | 2.24 | 2.45 |
| 86 | 0 | 22.43 | 17.09 | 178.62 | 218.14 |
| 87 | 0 | 2.76 | 0.92 | 2.67 | 6.35 |
| 88 | 0 | 0 | 0 | 5.08 | 5.08 |

DRUG CODE:9120DRUG NAME:DIHYDROCODEINE
STATE:NEW JERSEY

| ZIP CODE | QUARTER 1 | QUARTER 2 | QUARTER 3 | QUARTER 4 | TOTAL GRAMS |
|---|---|---|---|---|---|
| TOTAL | 5.29 | 76.78 | 60.51 | 374.4 | 516.98 |

STATE: NEW MEXICO

| ZIP CODE | QUARTER 1 | QUARTER 2 | QUARTER 3 | QUARTER 4 | TOTAL GRAMS |
|---|---|---|---|---|---|
| 870 | 0 | 0 | 0 | 9.61 | 9.61 |
| 871 | 26.7 | 12.82 | 19.23 | 22.43 | 81.18 |
| 875 | 13.89 | 29.91 | 10.68 | 12.82 | 67.3 |
| 877 | 0 | 0 | 12.82 | 2.14 | 14.96 |
| 878 | 6.41 | 0 | 0 | 0 | 6.41 |
| 880 | 103.61 | 100.41 | 115.36 | 115.36 | 434.74 |
| 883 | 0 | 0 | 2.14 | 0 | 2.14 |
| TOTAL | 150.61 | 143.14 | 160.23 | 162.36 | 616.34 |

STATE: NEW YORK

| ZIP CODE | QUARTER 1 | QUARTER 2 | QUARTER 3 | QUARTER 4 | TOTAL GRAMS |
|---|---|---|---|---|---|
| 100 | 0 | 0 | 14.9 | 17.09 | 31.99 |
| 103 | 0 | 0 | 0.21 | 1.07 | 1.28 |
| 104 | 0 | 0 | 0 | 2.14 | 2.14 |
| 110 | 0 | 0 | 3.2 | 1.07 | 4.27 |
| 112 | 0 | 0 | 4.27 | 2.14 | 6.41 |
| 113 | 0 | 12.82 | 1.34 | 3.2 | 17.36 |
| 117 | 4.27 | 4.27 | 44.12 | 66.25 | 118.91 |
| 125 | 1.07 | 0 | 1.07 | 0 | 2.14 |
| 127 | 0 | 0 | 1.07 | 0 | 1.07 |
| 130 | 0 | 0 | 2.14 | 0 | 2.14 |
| 132 | 4.27 | 0 | 0 | 0 | 4.27 |
| 136 | 0 | 0 | 0 | 5.34 | 5.34 |
| 141 | 0 | 0 | 0 | 1.07 | 1.07 |
| 142 | 0 | 0 | 0 | 1.07 | 1.07 |
| 143 | 0 | 0 | 1.07 | 0 | 1.07 |
| 146 | 0 | 1.07 | 0 | 0 | 1.07 |
| TOTAL | 9.61 | 18.16 | 73.39 | 100.44 | 201.6 |

STATE: NORTH CAROLINA

| ZIP CODE | QUARTER 1 | QUARTER 2 | QUARTER 3 | QUARTER 4 | TOTAL GRAMS |
|---|---|---|---|---|---|
| 274 | 0 | 0 | 1.07 | 0 | 1.07 |
| 275 | 1.07 | 0 | 1.07 | 3.2 | 5.34 |
| 276 | 1.07 | 0 | 1.07 | 0 | 2.14 |
| 278 | 4.27 | 9.61 | 0 | 0 | 13.88 |
| 280 | 1.07 | 1.07 | 0 | 0 | 2.14 |

| | | | | | |
|---|---|---|---|---|---|
| 281 | 1.28 | 0 | 1.07 | 0 | 2.35 |
| 282 | 0 | 0 | 1.07 | 0 | 1.07 |
| 283 | 1.07 | 0 | 0 | 0 | 1.07 |
| 285 | 0 | 0 | 4.27 | 0 | 4.27 |
| 286 | 0 | 2.14 | 0 | 1.07 | 3.21 |
| TOTAL | 9.83 | 12.82 | 9.62 | 4.27 | 36.54 |

STATE: NORTH DAKOTA

| ZIP CODE | QUARTER 1 | QUARTER 2 | QUARTER 3 | QUARTER 4 | TOTAL GRAMS |
|---|---|---|---|---|---|
| 585 | 0 | 0 | 0 | 2.14 | 2.14 |
| TOTAL | 0 | 0 | 0 | 2.14 | 2.14 |

STATE: OHIO

| ZIP CODE | QUARTER 1 | QUARTER 2 | QUARTER 3 | QUARTER 4 | TOTAL GRAMS |
|---|---|---|---|---|---|
| 430 | 4.27 | 9.61 | 5.34 | 6.41 | 25.63 |
| 431 | 0.21 | 1.07 | 0 | 0 | 1.28 |
| 432 | 19.23 | 14.95 | 33.11 | 21.36 | 88.65 |
| 452 | 0 | 0 | 1.07 | 0 | 1.07 |
| 453 | 3.2 | 0 | 0 | 0 | 3.2 |
| 455 | 1.07 | 0 | 0 | 1.07 | 2.14 |
| 456 | 4.27 | 3.2 | 3.2 | 2.14 | 12.81 |
| TOTAL | 32.25 | 28.83 | 42.72 | 30.98 | 134.78 |

STATE: OKLAHOMA

| ZIP CODE | QUARTER 1 | QUARTER 2 | QUARTER 3 | QUARTER 4 | TOTAL GRAMS |
|---|---|---|---|---|---|
| 730 | 56.92 | 44.51 | 62.59 | 62.91 | 226.93 |
| 731 | 37.78 | 66.25 | 102.48 | 83 | 289.51 |
| 734 | 16.49 | 32.46 | 26.83 | 27.13 | 102.91 |
| 735 | 21.95 | 19.44 | 24.99 | 16.02 | 82.4 |
| 736 | 0 | 0 | 0 | 1.07 | 1.07 |
| 737 | 1.07 | 2.14 | 2.14 | 1.07 | 6.42 |
| 738 | 2.14 | 3.2 | 1.07 | 2.14 | 8.55 |

DRUG CODE:9120DRUG NAME:DIHYDROCODEINE
STATE:OKLAHOMA

| ZIP CODE | QUARTER 1 | QUARTER 2 | QUARTER 3 | QUARTER 4 | TOTAL GRAMS |
|---|---|---|---|---|---|
| 740 | 13.51 | 13.18 | 12.28 | 17.09 | 56.06 |
| 741 | 36.88 | 28.37 | 6.41 | 16.29 | 87.95 |
| 743 | 15.02 | 12.82 | 11.75 | 12.82 | 52.41 |
| 744 | 18.82 | 9.72 | 20.94 | 23.5 | 72.98 |
| 745 | 0.09 | 1.07 | 0 | 0 | 1.16 |
| 747 | 0 | 1.07 | 0 | 2.14 | 3.21 |
| 748 | 6.41 | 12.82 | 20.3 | 8.55 | 48.08 |
| 749 | 6.41 | 2.46 | 1.28 | 1.07 | 11.22 |
| TOTAL | 233.49 | 249.51 | 293.06 | 274.8 | 1,050.86 |

STATE: PENNSYLVANIA

| ZIP CODE | QUARTER 1 | QUARTER 2 | QUARTER 3 | QUARTER 4 | TOTAL GRAMS |
|---|---|---|---|---|---|
| 157 | 1.07 | 3.2 | 1.07 | 30.44 | 35.78 |
| 170 | 3.2 | 3.2 | 1.07 | 0 | 7.47 |
| 187 | 0 | 4.27 | 5.34 | 7.48 | 17.09 |
| 189 | 0 | 3.76 | 1.5 | 4.46 | 9.72 |
| 190 | 99.02 | 117.92 | 87.06 | 23.22 | 327.22 |
| 191 | 108.1 | 146.46 | 167.59 | 250.95 | 673.1 |
| 193 | 3.74 | 17.41 | 10.9 | 12.16 | 44.21 |
| 194 | 2.35 | 1.89 | 0 | 10.68 | 14.92 |
| 196 | 0 | 0 | 0 | 2.14 | 2.14 |
| TOTAL | 217.48 | 298.11 | 274.53 | 341.53 | 1,131.65 |

RETAIL DRUG DISTRIBUTION BY ZIP CODE WITHIN STATE BY GRAMS WT

STATE: RHODE ISLAND

| ZIP CODE | QUARTER 1 | QUARTER 2 | QUARTER 3 | QUARTER 4 | TOTAL GRAMS |
|---|---|---|---|---|---|
| 28 | 0 | 1.07 | 5.34 | 0 | 6.41 |
| TOTAL | 0 | 1.07 | 5.34 | 0 | 6.41 |

STATE: SOUTH CAROLINA

| ZIP CODE | QUARTER 1 | QUARTER 2 | QUARTER 3 | QUARTER 4 | TOTAL GRAMS |
|---|---|---|---|---|---|
| 290 | 7.48 | 9.61 | 0 | 0 | 17.09 |
| 291 | 0 | 0 | 2.14 | 0 | 2.14 |
| 293 | 2.14 | 0 | 1.07 | 0 | 3.21 |
| 294 | 0 | 3.2 | 1.07 | 1.07 | 5.34 |
| 295 | 1.28 | 0 | 1.28 | 2.14 | 4.7 |
| 296 | 5.55 | 4.27 | 2.14 | 5.34 | 17.3 |
| 297 | 1.07 | 3.2 | 0 | 2.14 | 6.41 |
| 298 | 0 | 0 | 1.07 | 1.07 | 2.14 |
| 299 | 1.07 | 0 | 1.07 | 1.07 | 3.21 |
| TOTAL | 18.59 | 20.28 | 9.84 | 12.83 | 61.54 |

STATE: TENNESSEE

| ZIP CODE | QUARTER 1 | QUARTER 2 | QUARTER 3 | QUARTER 4 | TOTAL GRAMS |
|---|---|---|---|---|---|
| 370 | 447.08 | 640.27 | 743.97 | 608.92 | 2,440.24 |
| 371 | 17.77 | 10.77 | 14.4 | 16.79 | 59.73 |
| 372 | 116.97 | 236.04 | 343.21 | 298.2 | 994.42 |
| 373 | 11.75 | 17.09 | 18.16 | 14.95 | 61.95 |
| 374 | 0.32 | 2.62 | 2.35 | 6.94 | 12.23 |
| 377 | 14.31 | 13.03 | 8.83 | 7.26 | 43.43 |
| 378 | 41.87 | 64.32 | 92 | 24.89 | 223.08 |
| 379 | 43.63 | 50.47 | 49.98 | 35.25 | 179.33 |
| 380 | 13.62 | 12.21 | 23.98 | 15.49 | 65.3 |
| 381 | 12.67 | 10.95 | 2.51 | 11.32 | 37.45 |
| 382 | 1.17 | 0.6 | 0.66 | 1.83 | 4.26 |
| 383 | 23.58 | 14.9 | 28.04 | 20.99 | 87.51 |
| 384 | 7.69 | 10.68 | 2.35 | 7.48 | 28.2 |
| 385 | 0 | 0 | 4.27 | 2.14 | 6.41 |
| TOTAL | 752.43 | 1,083.95 | 1,334.71 | 1,072.45 | 4,243.54 |

STATE: TEXAS

| ZIP CODE | QUARTER 1 | QUARTER 2 | QUARTER 3 | QUARTER 4 | TOTAL GRAMS |
|---|---|---|---|---|---|
| 750 | 322.42 | 528.25 | 473.13 | 363.49 | 1,687.29 |
| 751 | 72.34 | 78.51 | 112.24 | 75.54 | 338.63 |
| 752 | 437.34 | 186.9 | 94.87 | 84.33 | 803.44 |
| 753 | 0 | 0.11 | 0 | 1.07 | 1.18 |
| 754 | 31.7 | 24.59 | 56.74 | 61.97 | 175 |
| 755 | 32.21 | 29.35 | 29.05 | 46.36 | 136.97 |
| 756 | 16.36 | 24.67 | 13.59 | 23.93 | 78.55 |
| 757 | 9.61 | 4.27 | 4.27 | 6.41 | 24.56 |
| 758 | 0 | 0 | 1.07 | 0 | 1.07 |
| 759 | 19.07 | 24.94 | 25.66 | 45.56 | 115.23 |

DRUG CODE:9120DRUG NAME:DIHYDROCODEINE
STATE:TEXAS

| ZIP CODE | QUARTER 1 | QUARTER 2 | QUARTER 3 | QUARTER 4 | TOTAL GRAMS |
|---|---|---|---|---|---|
| 760 | 91.88 | 242.62 | 574.67 | 546.47 | 1,455.64 |
| 761 | 120.64 | 122.14 | 122.74 | 97.9 | 463.42 |
| 762 | 38.27 | 90.92 | 94.76 | 57.47 | 281.42 |
| 763 | 106.91 | 104.97 | 107.67 | 123.11 | 442.66 |
| 764 | 11.75 | 18.16 | 18.16 | 10.68 | 58.75 |
| 765 | 0 | 0 | 0 | 20.3 | 20.3 |

| | | | | | |
|---|---|---|---|---|---|
| 766 | 0 | 2.14 | 0 | 2.14 | 4.28 |
| 767 | 1.07 | 3.2 | 5.34 | 19.23 | 28.84 |
| 769 | 2.56 | 2.03 | 1.07 | 0 | 5.66 |
| 770 | 101.51 | 114.47 | 103.71 | 104.34 | 424.03 |
| 773 | 75.41 | 82.96 | 93.68 | 97.3 | 349.35 |
| 774 | 34.19 | 62.46 | 27.18 | 69.65 | 193.48 |
| 775 | 127.03 | 114.35 | 133.73 | 124.71 | 499.82 |
| 776 | 29.04 | 18.53 | 20.08 | 22.64 | 90.29 |
| 777 | 8.33 | 12.07 | 14.74 | 14.53 | 49.67 |
| 778 | 2.14 | 0 | 5.34 | 5.34 | 12.82 |
| 779 | 11.11 | 12.82 | 8.97 | 4.51 | 37.41 |
| 780 | 22.86 | 71.25 | 39.52 | 31.08 | 164.71 |
| 781 | 34.29 | 20.46 | 24.03 | 61.42 | 140.2 |
| 782 | 120.09 | 96.88 | 113.76 | 152.96 | 483.69 |
| 783 | 71.14 | 69.22 | 95.28 | 78.19 | 313.83 |
| 784 | 117.07 | 94.32 | 185.01 | 272.87 | 669.27 |
| 785 | 80.53 | 87.59 | 206.58 | 161.12 | 535.82 |
| 786 | 2.35 | 3.2 | 6.41 | 12.82 | 24.78 |
| 787 | 5.55 | 2.14 | 0.91 | 24.46 | 33.06 |
| 788 | 1.07 | 0 | 0 | 0 | 1.07 |
| 789 | 0 | 0 | 0 | 1.07 | 1.07 |
| 790 | 1.07 | 1.07 | 0 | 1.07 | 3.21 |
| 791 | 2.14 | 2.14 | 0 | 0 | 4.28 |
| 793 | 1.07 | 1.07 | 0 | 0 | 2.14 |
| 794 | 5.34 | 3.2 | 9.61 | 7.48 | 25.63 |
| 796 | 0 | 0 | 0 | 2.14 | 2.14 |
| 797 | 16.02 | 8.55 | 8.55 | 23.5 | 56.62 |
| 799 | 8.97 | 2.14 | 2.14 | 1.07 | 14.32 |
| TOTAL | 2,192.45 | 2,368.66 | 2,834.26 | 2,860.23 | 10,255.60 |

STATE: VIRGINIA

| ZIP CODE | QUARTER 1 | QUARTER 2 | QUARTER 3 | QUARTER 4 | TOTAL GRAMS |
|---|---|---|---|---|---|
| 223 | 2.14 | 0 | 0 | 0 | 2.14 |
| 224 | 0 | 1.07 | 0 | 0 | 1.07 |
| 226 | 1.07 | 2.14 | 3.2 | 3.2 | 9.61 |
| 227 | 4.27 | 1.07 | 0 | 0 | 5.34 |
| 229 | 0 | 0 | 1.07 | 0 | 1.07 |
| 231 | 0 | 0 | 1.07 | 0 | 1.07 |
| 232 | 0 | 2.14 | 2.14 | 2.14 | 6.42 |
| 234 | 1.07 | 0 | 1.07 | 0 | 2.14 |
| 241 | 0 | 0 | 1.07 | 0 | 1.07 |
| 242 | 0 | 1.07 | 0 | 0 | 1.07 |
| TOTAL | 8.55 | 7.49 | 9.62 | 5.34 | 31 |

STATE: WASHINGTON

| ZIP CODE | QUARTER 1 | QUARTER 2 | QUARTER 3 | QUARTER 4 | TOTAL GRAMS |
|---|---|---|---|---|---|
| 980 | 0 | 1.07 | 0 | 0 | 1.07 |
| TOTAL | 0 | 1.07 | 0 | 0 | 1.07 |

STATE: WEST VIRGINIA

| ZIP CODE | QUARTER 1 | QUARTER 2 | QUARTER 3 | QUARTER 4 | TOTAL GRAMS |
|---|---|---|---|---|---|
| 255 | 0 | 0 | 0.21 | 0 | 0.21 |
| 256 | 0 | 0 | 0 | 12.82 | 12.82 |
| 258 | 0.21 | 0 | 0 | 0 | 0.21 |
| 260 | 6.41 | 6.41 | 1.07 | 4.27 | 18.16 |
| TOTAL | 6.62 | 6.41 | 1.28 | 17.09 | 31.4 |

STATE: WISCONSIN

| ZIP CODE | QUARTER 1 | QUARTER 2 | QUARTER 3 | QUARTER 4 | TOTAL GRAMS |
|---|---|---|---|---|---|
| 531 | 9.61 | 0 | 1.07 | 4.27 | 14.95 |
| 532 | 0 | 1.07 | 2.14 | 0 | 3.21 |
| 534 | 0 | 2.14 | 0 | 0 | 2.14 |

| | | | | | |
|---|---|---|---|---|---|
| 535 | 0 | 0 | 0 | 1.07 | 1.07 |
| TOTAL | 9.61 | 3.21 | 3.21 | 5.34 | 21.37 |

DRUG CODE:9143DRUG NAME:OXYCODONE
STATE: ALABAMA

| ZIP CODE | QUARTER 1 | QUARTER 2 | QUARTER 3 | QUARTER 4 | TOTAL GRAMS |
|---|---|---|---|---|---|
| 350 | 32,032.14 | 31,872.77 | 31,959.53 | 30,728.56 | 126,593 |
| 351 | 18,006.80 | 17,588.84 | 17,785.15 | 16,818.12 | 70,198.91 |
| 352 | 26,407.65 | 25,080.13 | 25,741.60 | 24,384.50 | 101,613.88 |
| 354 | 9,271.36 | 9,368.49 | 9,024.52 | 9,091.57 | 36,755.94 |
| 355 | 14,958.12 | 14,677.48 | 14,709.07 | 14,011.47 | 58,356.14 |
| 356 | 27,955.09 | 26,596.22 | 25,702.97 | 24,043.42 | 104,297.70 |
| 357 | 11,862.11 | 11,741.70 | 11,794.69 | 11,673.02 | 47,071.52 |
| 358 | 17,607.51 | 17,102.40 | 17,429.96 | 16,303.49 | 68,443.36 |
| 359 | 26,154.80 | 24,320.63 | 22,484.16 | 21,972.21 | 94,931.80 |
| 360 | 9,232.05 | 9,482.05 | 9,097.22 | 9,189.60 | 37,000.92 |
| 361 | 6,017.64 | 5,999.01 | 6,365.97 | 6,155.19 | 24,537.81 |
| 362 | 13,031.91 | 12,572.89 | 12,373.69 | 11,868.33 | 49,846.82 |
| 363 | 9,930.25 | 9,818.78 | 10,267.42 | 9,930.72 | 39,947.17 |
| 364 | 5,275.80 | 5,224.96 | 5,352.54 | 5,128.59 | 20,981.89 |
| 365 | 22,238.55 | 21,743.35 | 22,038.63 | 19,999.48 | 86,020.01 |
| 366 | 14,957.72 | 14,729.85 | 14,395.51 | 13,682.60 | 57,765.68 |
| 367 | 2,892.10 | 2,789.28 | 2,898.28 | 2,646.81 | 11,226.47 |
| 368 | 6,319.39 | 6,107.34 | 6,152.05 | 6,408.03 | 24,986.81 |
| 369 | 615.67 | 554.57 | 576.81 | 623.74 | 2,370.79 |
| TOTAL | 274,766.66 | 267,370.74 | 266,149.77 | 254,659.45 | 1,062,946.62 |

STATE: ALASKA

| ZIP CODE | QUARTER 1 | QUARTER 2 | QUARTER 3 | QUARTER 4 | TOTAL GRAMS |
|---|---|---|---|---|---|
| 995 | 21,612.28 | 20,788.97 | 19,836.23 | 20,325.24 | 82,562.72 |
| 996 | 11,207.64 | 10,804.56 | 11,109.30 | 10,671.29 | 43,792.79 |
| 997 | 4,253.85 | 3,870.13 | 3,872.93 | 3,657.12 | 15,654.03 |
| 998 | 1,624.16 | 1,395.46 | 1,388.89 | 1,386.16 | 5,794.67 |
| 999 | 626.99 | 654.89 | 604.47 | 640.37 | 2,526.72 |
| TOTAL | 39,324.92 | 37,514.01 | 36,811.82 | 36,680.18 | 150,330.93 |

STATE: AMERICAN SAMOA

| ZIP CODE | QUARTER 1 | QUARTER 2 | QUARTER 3 | QUARTER 4 | TOTAL GRAMS |
|---|---|---|---|---|---|
| 967 | 25.55 | 43.03 | 32.27 | 32.27 | 133.12 |
| TOTAL | 25.55 | 43.03 | 32.27 | 32.27 | 133.12 |

STATE: ARIZONA

| ZIP CODE | QUARTER 1 | QUARTER 2 | QUARTER 3 | QUARTER 4 | TOTAL GRAMS |
|---|---|---|---|---|---|
| 850 | 112,562.29 | 105,879.72 | 103,686.46 | 100,016.68 | 422,145.15 |
| 851 | 24,180.52 | 22,476.46 | 22,280.35 | 22,216.03 | 91,153.36 |
| 852 | 127,662.89 | 120,232.06 | 117,258.96 | 117,103.28 | 482,257.19 |
| 853 | 93,666.32 | 88,288.58 | 86,631.14 | 87,579.63 | 356,165.67 |
| 855 | 11,481.24 | 10,667.33 | 10,150.35 | 8,899.08 | 41,198 |
| 856 | 16,889.03 | 15,554.41 | 14,837.94 | 14,246.43 | 61,527.81 |
| 857 | 76,679.57 | 73,217.23 | 73,765.92 | 70,890.40 | 294,553.12 |
| 859 | 7,032.13 | 6,835.43 | 7,399.43 | 7,020.45 | 28,287.44 |
| 860 | 8,004.39 | 8,046.61 | 7,776.41 | 7,628.72 | 31,456.13 |
| 863 | 15,840.23 | 15,257.70 | 15,168.40 | 14,832.93 | 61,099.26 |
| 864 | 24,380.28 | 23,026.78 | 23,304.95 | 23,712.75 | 94,424.76 |
| 865 | 416.42 | 367.57 | 346.05 | 329.02 | 1,459.06 |
| TOTAL | 518,795.31 | 489,849.88 | 482,606.36 | 474,475.40 | 1,965,726.95 |

STATE: ARKANSAS

| ZIP CODE | QUARTER 1 | QUARTER 2 | QUARTER 3 | QUARTER 4 | TOTAL GRAMS |
|---|---|---|---|---|---|

| | | | | | |
|---|---|---|---|---|---|
| 716 | 4,931.02 | 4,805.30 | 4,722.92 | 4,751.91 | 19,211.15 |
| 717 | 3,564.34 | 3,519.17 | 3,155.47 | 3,252.04 | 13,491.02 |
| 718 | 4,028.41 | 3,482.36 | 3,698.76 | 3,577.10 | 14,786.63 |
| 719 | 7,345.55 | 6,876.14 | 6,676.92 | 6,440.72 | 27,339.33 |
| 720 | 19,769.74 | 19,293.65 | 18,395.84 | 17,994.76 | 75,453.99 |
| 721 | 19,116.53 | 18,649.19 | 17,909.53 | 17,046.50 | 72,721.75 |
| 722 | 14,737.43 | 14,435.45 | 13,398.16 | 13,129.89 | 55,700.93 |
| 723 | 4,416.03 | 4,404.24 | 4,600.14 | 4,605.16 | 18,025.57 |
| 724 | 7,005.95 | 6,789.69 | 6,986.98 | 6,912.71 | 27,695.33 |
| 725 | 6,063.46 | 6,122.80 | 6,088.47 | 6,101.15 | 24,375.88 |
| 726 | 9,403.75 | 8,887.38 | 8,694.27 | 8,264.33 | 35,249.73 |
| 727 | 37,273.08 | 36,488.64 | 35,506.35 | 34,986.10 | 144,254.17 |
| 728 | 6,180.25 | 6,032.83 | 5,789.28 | 5,956.37 | 23,958.73 |
| 729 | 22,664.40 | 19,139.47 | 18,979.68 | 20,712.37 | 81,495.92 |
| TOTAL | 166,499.94 | 158,926.31 | 154,602.77 | 153,731.11 | 633,760.13 |

STATE: CALIFORNIA

ZIP CODE QUARTER 1 QUARTER 2 QUARTER 3 QUARTER 4 TOTAL GRAMS

DRUG CODE:9143DRUG NAME:OXYCODONE
STATE:CALIFORNIA

| ZIP CODE | QUARTER 1 | QUARTER 2 | QUARTER 3 | QUARTER 4 | TOTAL GRAMS |
|---|---|---|---|---|---|
| 900 | 29,382.82 | 28,120.11 | 27,398.76 | 27,450.63 | 112,352.32 |
| 902 | 19,179.80 | 18,464.49 | 18,194.56 | 17,148.06 | 72,986.91 |
| 903 | 1,321.04 | 1,298.22 | 1,486.74 | 1,286.07 | 5,392.07 |
| 904 | 6,946.26 | 6,215.69 | 5,165.84 | 5,431.11 | 23,758.90 |
| 905 | 4,280.18 | 4,160.93 | 4,034.08 | 4,253.55 | 16,728.74 |
| 906 | 9,883.73 | 9,866.17 | 9,066.53 | 8,836.73 | 37,653.16 |
| 907 | 9,300.93 | 9,582.12 | 9,166.76 | 9,232.06 | 37,281.87 |
| 908 | 7,964.91 | 7,761.92 | 7,676.13 | 7,343.62 | 30,746.58 |
| 910 | 6,072.05 | 5,712.19 | 5,429.38 | 4,925.22 | 22,138.84 |
| 911 | 3,317.56 | 3,032.07 | 3,024.21 | 3,051.87 | 12,425.71 |
| 912 | 3,682.08 | 3,175.44 | 3,401.43 | 3,748.92 | 14,007.87 |
| 913 | 35,126.97 | 35,584.42 | 34,274.43 | 34,572.49 | 139,558.31 |
| 914 | 10,150.50 | 8,965.02 | 8,325.13 | 9,059.20 | 36,499.85 |
| 915 | 4,397.51 | 4,193.38 | 4,166.34 | 3,928.37 | 16,685.60 |
| 916 | 3,448.16 | 3,326.88 | 2,726.81 | 2,408.64 | 11,910.49 |
| 917 | 29,130.11 | 28,527.90 | 28,793.19 | 28,971.20 | 115,422.40 |
| 918 | 734.23 | 692.32 | 493.3 | 539.02 | 2,458.87 |
| 919 | 14,024.64 | 13,862.79 | 13,798.24 | 13,441.13 | 55,126.80 |
| 920 | 64,238.85 | 62,220.46 | 61,129.20 | 59,439.57 | 247,028.08 |
| 921 | 41,979.09 | 39,463.44 | 39,486.38 | 38,163.79 | 159,092.70 |
| 922 | 32,186.33 | 31,146.82 | 30,305.35 | 30,220.67 | 123,859.17 |
| 923 | 35,635.04 | 34,708.18 | 33,914.74 | 33,970.29 | 138,228.25 |
| 924 | 5,705.69 | 5,053.45 | 5,191.84 | 5,276.04 | 21,227.02 |
| 925 | 34,939.87 | 33,703.94 | 33,673.82 | 33,490.62 | 135,808.25 |
| 926 | 34,874.45 | 32,205.67 | 31,128.38 | 31,605.41 | 129,813.91 |
| 927 | 8,277.37 | 7,706.47 | 7,129.36 | 6,636.84 | 29,750.04 |
| 928 | 28,596.79 | 27,938.11 | 26,762.80 | 27,063.21 | 110,360.91 |
| 930 | 20,426.46 | 20,080.53 | 18,861.86 | 19,628.17 | 78,997.02 |
| 931 | 3,932.87 | 3,826.16 | 3,472.76 | 3,200.46 | 14,432.25 |
| 932 | 11,825.48 | 11,593.37 | 11,013.75 | 11,405.75 | 45,838.35 |
| 933 | 15,580.50 | 14,712.42 | 14,337.61 | 14,571.86 | 59,202.39 |
| 934 | 19,340.44 | 18,692.54 | 17,074.76 | 17,865.97 | 72,973.71 |
| 935 | 22,527.69 | 22,237.62 | 22,128.68 | 22,671.57 | 89,565.56 |
| 936 | 13,726.29 | 13,897.19 | 13,320.29 | 13,369.08 | 54,312.85 |
| 937 | 15,459.10 | 14,958.94 | 14,524.64 | 14,793.48 | 59,736.16 |
| 939 | 10,382.11 | 9,631.63 | 8,604.29 | 8,461.46 | 37,079.49 |
| 940 | 16,865.87 | 15,216.17 | 15,299.78 | 15,181.70 | 62,563.52 |

| | | | | | |
|---|---|---|---|---|---|
| 941 | 29,437.22 | 27,614.38 | 27,429.59 | 26,127.23 | 110,608.42 |
| 943 | 4,865.35 | 4,653.74 | 4,465.32 | 4,537.99 | 18,522.40 |
| 944 | 2,776.80 | 2,656.94 | 2,624.96 | 2,680.17 | 10,738.87 |
| 945 | 68,958.98 | 66,419.57 | 64,357.56 | 62,446.95 | 262,183.06 |
| 946 | 8,496.30 | 8,079.31 | 7,340.81 | 7,147.34 | 31,063.76 |
| 947 | 1,815.76 | 1,793.84 | 1,691.47 | 1,676.50 | 6,977.57 |
| 948 | 2,469.19 | 2,102.92 | 1,810.29 | 1,874.65 | 8,257.05 |
| 949 | 13,401.53 | 13,029.10 | 12,406.89 | 11,536.29 | 50,373.81 |
| 950 | 18,246.68 | 17,449.57 | 16,757.39 | 16,197.13 | 68,650.77 |
| 951 | 10,699.81 | 10,752.30 | 10,613.87 | 10,557.73 | 42,623.71 |
| 952 | 13,326.67 | 12,170.06 | 12,002.61 | 11,859.97 | 49,359.31 |
| 953 | 46,636.55 | 44,752.34 | 42,751.63 | 42,600.60 | 176,741.12 |
| 954 | 32,453.95 | 29,966.33 | 29,055.03 | 27,833.87 | 119,309.18 |
| 955 | 7,780.80 | 7,942.16 | 7,445.54 | 7,379 | 30,547.50 |
| 956 | 42,386.31 | 42,193.44 | 39,802.73 | 39,223.33 | 163,605.81 |
| 957 | 7,470.24 | 7,347.94 | 7,139.21 | 6,998.88 | 28,956.27 |
| 958 | 26,841.26 | 26,213.93 | 26,066.27 | 25,684.24 | 104,805.70 |
| 959 | 29,821.45 | 29,138.63 | 27,919.13 | 27,471.97 | 114,351.18 |
| 960 | 16,434.34 | 15,649.17 | 15,109.66 | 14,048.98 | 61,242.15 |
| 961 | 4,327.22 | 4,082.97 | 4,112.72 | 3,622.32 | 16,145.23 |
| TOTAL | 1,023,490.18 | 985,543.81 | 954,884.83 | 944,148.97 | 3,908,067.79 |

STATE: COLORADO

| ZIP CODE | QUARTER 1 | QUARTER 2 | QUARTER 3 | QUARTER 4 | TOTAL GRAMS |
|---|---|---|---|---|---|
| 800 | 43,215.68 | 41,497.01 | 40,830.97 | 39,555.52 | 165,099.18 |
| 801 | 31,042.66 | 30,863.64 | 28,955.42 | 27,940.84 | 118,802.56 |
| 802 | 51,206.02 | 49,900.16 | 49,916.33 | 47,945.10 | 198,967.61 |
| 803 | 5,227.49 | 4,985.90 | 4,659.22 | 4,871.47 | 19,744.08 |
| 804 | 7,533.83 | 7,261.91 | 7,427.30 | 6,943.72 | 29,166.76 |
| 805 | 26,806.70 | 24,318.33 | 23,820.75 | 22,430.77 | 97,376.55 |

DRUG CODE:9143DRUG NAME:OXYCODONE
STATE:COLORADO

| ZIP CODE | QUARTER 1 | QUARTER 2 | QUARTER 3 | QUARTER 4 | TOTAL GRAMS |
|---|---|---|---|---|---|
| 806 | 12,415.12 | 11,433.89 | 11,464.48 | 10,983.23 | 46,296.72 |
| 807 | 2,511.41 | 2,583.79 | 2,545.07 | 2,437.27 | 10,077.54 |
| 808 | 6,108.92 | 5,743.53 | 5,631.28 | 5,542 | 23,025.73 |
| 809 | 32,280.17 | 30,151.41 | 30,488.60 | 30,622.19 | 123,542.37 |
| 810 | 21,324.66 | 20,319.40 | 21,006.72 | 20,455.09 | 83,105.87 |
| 811 | 3,982.35 | 3,713.23 | 3,676.10 | 3,665.83 | 15,037.51 |
| 812 | 3,624.98 | 3,625.01 | 3,759.58 | 3,747.75 | 14,757.32 |
| 813 | 2,306.78 | 2,135.42 | 2,115.58 | 1,822.52 | 8,380.30 |
| 814 | 2,820.72 | 2,824.36 | 2,864.65 | 2,600.31 | 11,110.04 |
| 815 | 6,271.32 | 6,212.69 | 6,121.20 | 6,087.20 | 24,692.41 |
| 816 | 4,494.87 | 4,187.22 | 4,070.40 | 4,218.11 | 16,970.60 |
| TOTAL | 263,173.68 | 251,756.90 | 249,353.65 | 241,868.92 | 1,006,153.15 |

STATE: CONNECTICUT

| ZIP CODE | QUARTER 1 | QUARTER 2 | QUARTER 3 | QUARTER 4 | TOTAL GRAMS |
|---|---|---|---|---|---|
| 60 | 37,418.08 | 35,887.27 | 34,886.47 | 33,338.68 | 141,530.50 |
| 61 | 10,583.05 | 10,684.06 | 9,947.28 | 9,378.45 | 40,592.84 |
| 62 | 6,722.48 | 6,124.71 | 5,794.13 | 5,642.07 | 24,283.39 |
| 63 | 18,123.04 | 17,435.04 | 17,165.97 | 16,092.51 | 68,816.56 |
| 64 | 56,394.59 | 54,008.60 | 51,227.89 | 49,629.75 | 211,260.83 |
| 65 | 20,878.40 | 19,868.74 | 18,941.59 | 18,174.50 | 77,863.23 |
| 66 | 12,033.09 | 11,520.87 | 11,226.77 | 10,854.72 | 45,635.45 |
| 67 | 23,158.75 | 22,289.32 | 20,855.53 | 20,346.32 | 86,649.92 |
| 68 | 21,326.39 | 20,554.25 | 19,567.93 | 18,565 | 80,013.57 |
| 69 | 3,434.43 | 3,478.07 | 2,870.42 | 2,859.95 | 12,642.87 |
| TOTAL | 210,072.30 | 201,850.93 | 192,483.98 | 184,881.95 | 789,289.16 |

RETAIL DRUG DISTRIBUTION BY ZIP CODE WITHIN STATE BY GRAMS WT

STATE: DELAWARE

| ZIP CODE | QUARTER 1 | QUARTER 2 | QUARTER 3 | QUARTER 4 | TOTAL GRAMS |
|---|---|---|---|---|---|
| 197 | 28,532.95 | 27,153.41 | 26,880.45 | 26,501.33 | 109,068.14 |
| 198 | 17,520.47 | 16,963.07 | 16,472.73 | 16,354.72 | 67,310.99 |
| 199 | 32,372.02 | 30,872.26 | 31,906.62 | 31,298.74 | 126,449.64 |
| TOTAL | 78,425.44 | 74,988.74 | 75,259.80 | 74,154.79 | 302,828.77 |

STATE: DISTRICT OF COLUMBIA

| ZIP CODE | QUARTER 1 | QUARTER 2 | QUARTER 3 | QUARTER 4 | TOTAL GRAMS |
|---|---|---|---|---|---|
| 200 | 16,037.76 | 15,020.43 | 15,417.98 | 14,652.56 | 61,128.73 |
| 203 | 146.13 | 85.62 | 41.69 | 59.62 | 333.06 |
| 204 | 1,441.84 | 1,326.46 | 1,530.15 | 1,184.90 | 5,483.35 |
| 205 | 0 | 0.9 | 2.24 | 1.34 | 4.48 |
| TOTAL | 17,625.73 | 16,433.41 | 16,992.06 | 15,898.42 | 66,949.62 |

STATE: FLORIDA

| ZIP CODE | QUARTER 1 | QUARTER 2 | QUARTER 3 | QUARTER 4 | TOTAL GRAMS |
|---|---|---|---|---|---|
| 320 | 34,665.71 | 34,564.35 | 34,190.96 | 34,632.67 | 138,053.69 |
| 321 | 31,531.82 | 30,505.25 | 29,584.01 | 30,203.40 | 121,824.48 |
| 322 | 66,226.45 | 64,802.93 | 66,248.55 | 63,480.86 | 260,758.79 |
| 323 | 12,844.23 | 12,687.57 | 12,581.40 | 13,000.17 | 51,113.37 |
| 324 | 15,452.20 | 15,519.81 | 16,161.17 | 16,025.89 | 63,159.07 |
| 325 | 31,291.97 | 31,254.54 | 31,796.85 | 32,058.80 | 126,402.16 |
| 326 | 25,290.12 | 24,940.98 | 24,518.71 | 24,048.39 | 98,798.20 |
| 327 | 44,497.03 | 44,758.23 | 45,371.88 | 46,043.72 | 180,670.86 |
| 328 | 30,955.73 | 30,435.92 | 30,585.37 | 29,629.57 | 121,606.59 |
| 329 | 54,514.25 | 54,573.92 | 54,271.22 | 53,817.92 | 217,177.31 |
| 330 | 63,313.97 | 64,112.51 | 64,816.52 | 65,288.97 | 257,531.97 |
| 331 | 51,154.68 | 51,783.89 | 52,541.69 | 53,660.64 | 209,140.90 |
| 333 | 44,159.73 | 45,597.75 | 46,763.49 | 45,930.90 | 182,451.87 |
| 334 | 81,178.09 | 80,457.18 | 79,474.27 | 81,807.96 | 322,917.50 |
| 335 | 40,194.68 | 40,271.24 | 40,721.73 | 41,313.92 | 162,501.57 |
| 336 | 56,364.78 | 56,577.30 | 58,122.89 | 58,902.99 | 229,967.96 |
| 337 | 54,038.82 | 53,208.06 | 53,103.60 | 53,332.76 | 213,683.24 |
| 338 | 32,538.23 | 32,436.52 | 32,735.46 | 33,120.73 | 130,830.94 |
| 339 | 46,259.37 | 44,665.44 | 43,609 | 45,496.45 | 180,030.26 |
| 341 | 19,205.63 | 17,278.70 | 17,065.49 | 18,014.94 | 71,564.76 |
| 342 | 51,409.87 | 49,593.98 | 49,800.53 | 49,865.24 | 200,669.62 |
| 344 | 31,489.77 | 31,372.74 | 31,094.29 | 32,056.83 | 126,013.63 |
| 346 | 49,945.09 | 50,363.86 | 48,879.47 | 50,712.50 | 199,900.92 |
| 347 | 27,128.12 | 26,100.98 | 26,115.10 | 26,073.39 | 105,417.59 |
| 349 | 28,070.08 | 27,656.46 | 28,342.06 | 29,126.94 | 113,195.54 |
| TOTAL | 1,023,720.42 | 1,015,520.11 | 1,018,495.71 | 1,027,646.55 | 4,085,382.79 |

DRUG CODE:9143DRUG NAME:OXYCODONE

STATE: GEORGIA

| ZIP CODE | QUARTER 1 | QUARTER 2 | QUARTER 3 | QUARTER 4 | TOTAL GRAMS |
|---|---|---|---|---|---|
| 300 | 81,820.18 | 81,498.08 | 81,717.72 | 81,209.27 | 326,245.25 |
| 301 | 71,353.86 | 69,594.23 | 70,824.70 | 70,418.57 | 282,191.36 |
| 302 | 41,252.02 | 40,257.57 | 40,550.36 | 41,256.49 | 163,316.44 |
| 303 | 28,616.29 | 28,080.95 | 28,227.68 | 28,808.99 | 113,733.91 |
| 304 | 10,359.09 | 10,015.39 | 10,311.94 | 10,368.46 | 41,054.88 |
| 305 | 38,896.29 | 39,066 | 39,087.68 | 39,894.14 | 156,944.11 |
| 306 | 26,749.22 | 26,994.73 | 27,180.59 | 28,160.13 | 109,084.67 |
| 307 | 18,639.39 | 18,624.38 | 19,905.45 | 19,881.71 | 77,050.93 |
| 308 | 8,788.91 | 8,859.64 | 8,620 | 8,382.44 | 34,650.99 |
| 309 | 19,279.13 | 18,222.43 | 18,589.52 | 18,206.93 | 74,298.01 |

| | | | | | | |
|---|---|---|---|---|---|---|
| | 310 | 26,043.03 | 25,673.85 | 26,513.97 | 26,001.09 | 104,231.94 |
| | 312 | 13,152.91 | 12,761.93 | 13,100.33 | 12,694.06 | 51,709.23 |
| | 313 | 11,646.72 | 12,059.01 | 12,244.15 | 12,298.62 | 48,248.50 |
| | 314 | 11,879.61 | 11,819.15 | 11,717.09 | 11,009.26 | 46,425.11 |
| | 315 | 19,226.06 | 19,279.12 | 20,386.52 | 19,769.64 | 78,661.34 |
| | 316 | 6,504.47 | 6,540.79 | 6,537.53 | 6,424.64 | 26,007.43 |
| | 317 | 13,885.95 | 13,530.88 | 13,560.73 | 13,221.55 | 54,199.11 |
| | 318 | 903.49 | 866.54 | 969.39 | 1,013.16 | 3,752.58 |
| | 319 | 7,837.34 | 8,016.58 | 8,623.45 | 7,233.42 | 31,710.79 |
| | 398 | 2,664.02 | 2,738.77 | 2,619.31 | 2,767.06 | 10,789.16 |
| TOTAL | | 459,497.98 | 454,500.02 | 461,288.11 | 459,019.63 | 1,834,305.74 |
| STATE: | GUAM | | | | | |
| ZIP CODE | QUARTER 1 | | QUARTER 2 | QUARTER 3 | QUARTER 4 | TOTAL GRAMS |
| | 969 | 890.2 | 1,005.84 | 988.84 | 1,080.03 | 3,964.91 |
| TOTAL | | 890.2 | 1,005.84 | 988.84 | 1,080.03 | 3,964.91 |
| STATE: | HAWAII | | | | | |
| ZIP CODE | QUARTER 1 | | QUARTER 2 | QUARTER 3 | QUARTER 4 | TOTAL GRAMS |
| | 967 | 35,411.52 | 33,713.82 | 32,273.13 | 32,115.92 | 133,514.39 |
| | 968 | 15,892.26 | 15,729.35 | 15,654.92 | 14,203.36 | 61,479.89 |
| TOTAL | | 51,303.78 | 49,443.17 | 47,928.05 | 46,319.28 | 194,994.28 |
| STATE: | IDAHO | | | | | |
| ZIP CODE | QUARTER 1 | | QUARTER 2 | QUARTER 3 | QUARTER 4 | TOTAL GRAMS |
| | 832 | 6,925.39 | 6,475.85 | 6,517.12 | 6,809.19 | 26,727.55 |
| | 833 | 7,581.25 | 7,480.44 | 7,524.55 | 7,412.75 | 29,998.99 |
| | 834 | 6,440.11 | 6,393.51 | 5,952.36 | 6,177.97 | 24,963.95 |
| | 835 | 2,090.62 | 2,038.27 | 2,187.61 | 2,153.50 | 8,470 |
| | 836 | 18,643.17 | 18,692.67 | 18,054.18 | 18,374.37 | 73,764.39 |
| | 837 | 14,502.69 | 14,240.13 | 13,802.71 | 14,135.03 | 56,680.56 |
| | 838 | 8,610.99 | 8,629.26 | 8,568.35 | 8,319.87 | 34,128.47 |
| TOTAL | | 64,794.22 | 63,950.13 | 62,606.88 | 63,382.68 | 254,733.91 |
| STATE: | ILLINOIS | | | | | |
| ZIP CODE | QUARTER 1 | | QUARTER 2 | QUARTER 3 | QUARTER 4 | TOTAL GRAMS |
| | 600 | 27,399.08 | 27,877.38 | 27,874.01 | 27,502.97 | 110,653.44 |
| | 601 | 17,379.64 | 16,503.46 | 16,997.20 | 16,852.63 | 67,732.93 |
| | 602 | 641.94 | 580.89 | 525.91 | 606.03 | 2,354.77 |
| | 603 | 617.69 | 666.5 | 609.47 | 666.91 | 2,560.57 |
| | 604 | 24,579.31 | 24,287.29 | 24,435.03 | 23,914.27 | 97,215.90 |
| | 605 | 12,358.70 | 12,451.09 | 12,604.37 | 11,746.25 | 49,160.41 |
| | 606 | 14,875 | 14,429.74 | 14,300.75 | 14,204.97 | 57,810.46 |
| | 607 | 1,266.04 | 1,347.82 | 1,524.45 | 1,497.58 | 5,635.89 |
| | 608 | 891.26 | 769.69 | 894.17 | 827.74 | 3,382.86 |
| | 609 | 3,165.68 | 3,200.59 | 3,245.37 | 3,254.67 | 12,866.31 |
| | 610 | 3,907.71 | 3,545.36 | 3,410.55 | 3,443.71 | 14,307.33 |
| | 611 | 4,042.07 | 3,882.65 | 3,839.01 | 3,955.33 | 15,719.06 |
| | 612 | 1,743.02 | 1,771.93 | 1,739.61 | 1,903.54 | 7,158.10 |
| | 613 | 1,902.09 | 1,930.37 | 1,907.65 | 1,904.21 | 7,644.32 |
| | 614 | 1,275.48 | 1,345.80 | 1,387.84 | 1,438.25 | 5,447.37 |
| | 615 | 2,857.95 | 2,662.93 | 2,655.61 | 2,814.18 | 10,990.67 |
| | 616 | 3,615.90 | 3,401.44 | 3,169.46 | 3,233.91 | 13,420.71 |
| | 617 | 2,568.19 | 2,540.83 | 2,532.49 | 2,516.49 | 10,158 |
| | 618 | 3,849.45 | 4,035.95 | 3,736.20 | 3,876.96 | 15,498.56 |
| | 619 | 1,083.47 | 1,002.56 | 921.29 | 1,044.73 | 4,052.05 |
| | 620 | 9,317.19 | 9,077.09 | 9,231.10 | 9,455.03 | 37,080.41 |
| | 622 | 10,725.72 | 10,216.02 | 10,090.94 | 10,463.12 | 41,495.80 |
| | 623 | 2,160.57 | 2,113.52 | 2,098.80 | 2,055.85 | 8,428.74 |
| | 624 | 2,394.06 | 2,313.28 | 2,239.19 | 2,322.91 | 9,269.44 |

RETAIL DRUG DISTRIBUTION BY ZIP CODE WITHIN STATE BY GRAMS WT

DRUG CODE:9143DRUG NAME:OXYCODONE
STATE:ILLINOIS

| ZIP CODE | QUARTER 1 | QUARTER 2 | QUARTER 3 | QUARTER 4 | TOTAL GRAMS |
|---|---|---|---|---|---|
| 625 | 2,071.60 | 2,081.09 | 1,983.98 | 1,983.47 | 8,120.14 |
| 626 | 2,165.27 | 2,149.88 | 2,160.73 | 2,238.52 | 8,714.40 |
| 627 | 2,231.02 | 2,064.88 | 2,082.33 | 1,983.42 | 8,361.65 |
| 628 | 5,020.17 | 5,227.91 | 5,049.45 | 5,215.81 | 20,513.34 |
| 629 | 9,354.18 | 9,424.71 | 9,339.90 | 9,168.17 | 37,286.96 |
| TOTAL | 175,459.45 | 172,902.65 | 172,586.86 | 172,091.63 | 693,040.59 |

STATE: INDIANA

| ZIP CODE | QUARTER 1 | QUARTER 2 | QUARTER 3 | QUARTER 4 | TOTAL GRAMS |
|---|---|---|---|---|---|
| 460 | 16,256.69 | 15,563.22 | 14,613.24 | 14,134.61 | 60,567.76 |
| 461 | 21,724.29 | 21,247.84 | 20,634.50 | 20,299.85 | 83,906.48 |
| 462 | 45,409.92 | 44,243.46 | 43,456.58 | 45,575.21 | 178,685.17 |
| 463 | 21,061.14 | 20,455.73 | 20,379.28 | 19,560.15 | 81,456.30 |
| 464 | 2,343.59 | 2,179.97 | 2,197.43 | 2,095.05 | 8,816.04 |
| 465 | 21,232.91 | 19,978.99 | 19,383.78 | 19,405.38 | 80,001.06 |
| 466 | 7,267.42 | 7,523.91 | 7,096.37 | 7,513.76 | 29,401.46 |
| 467 | 9,843.21 | 8,886.66 | 8,921.29 | 9,002.94 | 36,654.10 |
| 468 | 14,769.69 | 11,563.61 | 12,105.03 | 12,128.77 | 50,567.10 |
| 469 | 12,154.15 | 11,234.92 | 10,875.95 | 10,469.20 | 44,734.22 |
| 470 | 7,541.32 | 7,508.90 | 7,941.46 | 7,535.12 | 30,526.80 |
| 471 | 22,444.16 | 21,813.04 | 19,934.31 | 18,866.78 | 83,058.29 |
| 472 | 9,226.55 | 8,500.81 | 8,524.28 | 8,265.57 | 34,517.21 |
| 473 | 14,592.10 | 13,953.85 | 13,257.58 | 13,483.53 | 55,287.06 |
| 474 | 10,289.49 | 9,853.98 | 9,867.92 | 9,509.10 | 39,520.49 |
| 475 | 5,698.90 | 5,798.78 | 5,669.08 | 5,908.04 | 23,074.80 |
| 476 | 4,430.70 | 4,526.51 | 4,307.01 | 4,518.59 | 17,782.81 |
| 477 | 13,210.66 | 12,872.66 | 12,503.25 | 12,145.65 | 50,732.22 |
| 478 | 3,986.60 | 3,801.81 | 3,672.63 | 3,619.63 | 15,080.67 |
| 479 | 9,742.79 | 9,732.35 | 9,259.98 | 9,560.12 | 38,295.24 |
| TOTAL | 273,226.28 | 261,241 | 254,600.95 | 253,597.05 | 1,042,665.28 |

STATE: IOWA

| ZIP CODE | QUARTER 1 | QUARTER 2 | QUARTER 3 | QUARTER 4 | TOTAL GRAMS |
|---|---|---|---|---|---|
| 500 | 4,786.84 | 4,829.63 | 4,759.43 | 4,845.45 | 19,221.35 |
| 501 | 3,888.79 | 3,562.21 | 3,654.29 | 3,539.15 | 14,644.44 |
| 502 | 4,558.91 | 4,390.93 | 4,232.95 | 3,936.08 | 17,118.87 |
| 503 | 12,766.63 | 12,351.37 | 11,686.44 | 11,452.86 | 48,257.30 |
| 504 | 2,103.09 | 2,209.47 | 2,132.73 | 2,055.26 | 8,500.55 |
| 505 | 2,027.12 | 1,911.79 | 1,702.21 | 1,806.54 | 7,447.66 |
| 506 | 1,876.46 | 1,784.61 | 1,647.14 | 1,711.34 | 7,019.55 |
| 507 | 2,050.09 | 1,971.28 | 2,008.40 | 1,974.36 | 8,004.13 |
| 508 | 551.03 | 482.99 | 485.77 | 495.67 | 2,015.46 |
| 510 | 708.34 | 681.3 | 731.43 | 731.62 | 2,852.69 |
| 511 | 1,522.44 | 1,460.50 | 1,416.68 | 1,357.23 | 5,756.85 |
| 512 | 496.18 | 447.81 | 496.97 | 463.65 | 1,904.61 |
| 513 | 537.99 | 535.91 | 510.98 | 465.64 | 2,050.52 |
| 514 | 916.61 | 866.56 | 776.92 | 798.92 | 3,359.01 |
| 515 | 4,249.68 | 4,050.07 | 3,911.65 | 3,985.39 | 16,196.79 |
| 516 | 594.83 | 566.05 | 481.2 | 464.66 | 2,106.74 |
| 520 | 1,522.67 | 1,488.31 | 1,499.82 | 1,413.35 | 5,924.15 |
| 521 | 671.39 | 608.97 | 585.24 | 591.91 | 2,457.51 |
| 522 | 3,266.41 | 3,148.29 | 3,118.12 | 2,864.50 | 12,397.32 |
| 523 | 1,457.40 | 1,520.74 | 1,261.05 | 1,336.54 | 5,575.73 |
| 524 | 3,067.31 | 2,981.78 | 2,871.34 | 2,858.61 | 11,779.04 |
| 525 | 2,060.64 | 2,037.19 | 2,240.48 | 2,055.47 | 8,393.78 |
| 526 | 2,621.72 | 2,374.80 | 2,306.19 | 2,278.62 | 9,581.33 |

| | | | | | | |
|---|---|---|---|---|---|---|
| | 527 | 2,249.04 | 2,098.48 | 2,155.79 | 2,188 | 8,691.31 |
| | 528 | 2,568.70 | 2,439.22 | 2,553.46 | 2,566.63 | 10,128.01 |
| TOTAL | | 63,120.31 | 60,800.26 | 59,226.68 | 58,237.45 | 241,384.70 |
| STATE: | KANSAS | | | | | |
| ZIP CODE | QUARTER 1 | | QUARTER 2 | QUARTER 3 | QUARTER 4 | TOTAL GRAMS |
| | 660 | 19,502.64 | 18,830.82 | 18,726.60 | 18,381.47 | 75,441.53 |
| | 661 | 9,858.38 | 9,683.77 | 9,127.08 | 9,287.77 | 37,957 |
| | 662 | 17,034.56 | 16,534.80 | 17,246.74 | 15,910.45 | 66,726.55 |
| | 664 | 2,962.58 | 2,893.45 | 2,834.83 | 2,775.57 | 11,466.43 |
| | 665 | 3,167.63 | 3,123.53 | 3,132.64 | 2,981.43 | 12,405.23 |
| | 666 | 7,809.75 | 7,158.46 | 8,010.74 | 6,576.69 | 29,555.64 |
| | 667 | 5,426.62 | 5,346.28 | 5,169.91 | 5,482.10 | 21,424.91 |

DRUG CODE:9143DRUG NAME:OXYCODONE
STATE:KANSAS

| ZIP CODE | QUARTER 1 | | QUARTER 2 | QUARTER 3 | QUARTER 4 | TOTAL GRAMS |
|---|---|---|---|---|---|---|
| | 668 | 2,119.48 | 2,132.32 | 1,963.32 | 2,068.96 | 8,284.08 |
| | 669 | 730.34 | 749.47 | 719.4 | 692.41 | 2,891.62 |
| | 670 | 9,314.93 | 9,145.33 | 8,843.01 | 9,035.17 | 36,338.44 |
| | 671 | 4,064.63 | 3,975.65 | 4,038.04 | 3,975.83 | 16,054.15 |
| | 672 | 27,296.87 | 27,021.02 | 27,202.91 | 26,730.75 | 108,251.55 |
| | 673 | 2,503.70 | 2,602.27 | 2,581.29 | 2,641.71 | 10,328.97 |
| | 674 | 5,206.24 | 5,364.91 | 5,568.76 | 5,491.15 | 21,631.06 |
| | 675 | 6,751.84 | 6,759.07 | 6,785.31 | 6,323.03 | 26,619.25 |
| | 676 | 2,661.84 | 2,630.90 | 2,615.25 | 2,683.21 | 10,591.20 |
| | 677 | 1,390.29 | 1,261.83 | 1,397.56 | 1,506.18 | 5,555.86 |
| | 678 | 4,832.99 | 4,825.89 | 4,896.85 | 4,692.98 | 19,248.71 |
| | 679 | 1,237.62 | 1,203.23 | 1,269.94 | 1,273.11 | 4,983.90 |
| TOTAL | | 133,872.93 | 131,243 | 132,130.18 | 128,509.97 | 525,756.08 |
| STATE: | KENTUCKY | | | | | |
| ZIP CODE | QUARTER 1 | | QUARTER 2 | QUARTER 3 | QUARTER 4 | TOTAL GRAMS |
| | 400 | 8,888.95 | 8,634.30 | 8,627.12 | 8,776.31 | 34,926.68 |
| | 401 | 4,488.69 | 4,548.01 | 4,554.77 | 4,395 | 17,986.47 |
| | 402 | 38,618.74 | 37,231.26 | 36,997.13 | 36,617.46 | 149,464.59 |
| | 403 | 16,171.14 | 15,734.70 | 15,027.42 | 14,539.31 | 61,472.57 |
| | 404 | 8,658.77 | 8,263.92 | 7,959.61 | 8,207.64 | 33,089.94 |
| | 405 | 13,583.78 | 13,275.65 | 13,297.20 | 12,735.56 | 52,892.19 |
| | 406 | 1,430.28 | 1,461.38 | 1,501.10 | 1,381.07 | 5,773.83 |
| | 407 | 7,531.60 | 7,359.13 | 6,331.40 | 6,147.31 | 27,369.44 |
| | 408 | 1,320.19 | 1,287.78 | 1,257.88 | 1,252.86 | 5,118.71 |
| | 409 | 4,047.48 | 4,044.12 | 3,864.59 | 3,743.23 | 15,699.42 |
| | 410 | 32,450.60 | 32,154.36 | 31,567.18 | 31,190.83 | 127,362.97 |
| | 411 | 6,609.94 | 6,785.89 | 6,666.99 | 6,219.10 | 26,281.92 |
| | 412 | 2,256 | 2,299.84 | 2,370.26 | 2,141.20 | 9,067.30 |
| | 413 | 1,740.82 | 1,648.22 | 1,674.06 | 1,684.05 | 6,747.15 |
| | 414 | 988.26 | 901.65 | 859.52 | 818.15 | 3,567.58 |
| | 415 | 3,994.77 | 3,813.80 | 3,998.79 | 3,611.95 | 15,419.31 |
| | 416 | 3,079.39 | 3,053.86 | 3,006.10 | 3,024.21 | 12,163.56 |
| | 417 | 5,202.30 | 5,174.43 | 5,155.86 | 4,459.04 | 19,991.63 |
| | 418 | 2,040.75 | 1,949.22 | 1,854.41 | 1,754.14 | 7,598.52 |
| | 420 | 12,431.10 | 12,506.72 | 12,637.36 | 12,461.35 | 50,036.53 |
| | 421 | 10,392.45 | 10,472.22 | 10,503.59 | 10,788.36 | 42,156.62 |
| | 422 | 6,214.31 | 6,361.35 | 6,350.06 | 6,155.62 | 25,081.34 |
| | 423 | 7,857.15 | 7,771.27 | 7,760.07 | 7,630.88 | 31,019.37 |
| | 424 | 8,818.85 | 9,043.22 | 8,890.14 | 8,910.27 | 35,662.48 |
| | 425 | 3,366.51 | 3,241.54 | 3,045.59 | 2,623.97 | 12,277.61 |
| | 426 | 3,641.54 | 3,697.12 | 3,581.65 | 3,598.95 | 14,519.26 |
| | 427 | 7,539.80 | 7,289.72 | 7,566.25 | 7,498.18 | 29,893.95 |

| | | | | | |
|---|---|---|---|---|---|
| TOTAL | 223,364.16 | 220,004.68 | 216,906.10 | 212,366 | 872,640.94 |
| STATE: | LOUISIANA | | | | |
| ZIP CODE | QUARTER 1 | QUARTER 2 | QUARTER 3 | QUARTER 4 | TOTAL GRAMS |
| 700 | 40,179.72 | 40,092.12 | 39,856.09 | 39,584.46 | 159,712.39 |
| 701 | 15,144.67 | 15,241.47 | 14,838.60 | 15,172.19 | 60,396.93 |
| 703 | 9,913.92 | 9,875.10 | 9,656.08 | 9,906.48 | 39,351.58 |
| 704 | 33,175.73 | 33,070.13 | 32,288.03 | 30,817.81 | 129,351.70 |
| 705 | 23,395.13 | 23,305.39 | 23,097.91 | 23,186.52 | 92,984.95 |
| 706 | 7,968.17 | 7,753.41 | 6,917.36 | 6,399.47 | 29,038.41 |
| 707 | 23,995.33 | 24,202.78 | 23,836.99 | 22,944.17 | 94,979.27 |
| 708 | 16,147.69 | 15,754.74 | 15,698 | 15,466 | 63,066.43 |
| 710 | 3,778.60 | 3,757.99 | 3,690.77 | 3,850.72 | 15,078.08 |
| 711 | 14,640.14 | 15,021.48 | 15,443.10 | 15,157.29 | 60,262.01 |
| 712 | 7,058.11 | 7,081.91 | 7,246.45 | 7,251.36 | 28,637.83 |
| 713 | 13,374.79 | 13,101.37 | 13,108.20 | 12,072.30 | 51,656.66 |
| 714 | 7,156.94 | 6,649.77 | 6,709.91 | 6,213.70 | 26,730.32 |
| TOTAL | 215,928.94 | 214,907.66 | 212,387.49 | 208,022.47 | 851,246.56 |
| STATE: | MAINE | | | | |
| ZIP CODE | QUARTER 1 | QUARTER 2 | QUARTER 3 | QUARTER 4 | TOTAL GRAMS |
| 39 | 1,641.81 | 1,591.06 | 1,292.27 | 1,119.37 | 5,644.51 |
| 40 | 20,954.22 | 20,608.29 | 19,098.94 | 17,701.40 | 78,362.85 |
| 41 | 4,973.91 | 4,720.09 | 4,595.45 | 4,365.14 | 18,654.59 |
| 42 | 8,796.21 | 8,290.78 | 7,467.57 | 7,218.30 | 31,772.86 |
| 43 | 9,355.08 | 9,004.18 | 8,635.34 | 8,010.37 | 35,004.97 |

DRUG CODE:9143DRUG NAME:OXYCODONE
STATE:MAINE

| ZIP CODE | QUARTER 1 | QUARTER 2 | QUARTER 3 | QUARTER 4 | TOTAL GRAMS |
|---|---|---|---|---|---|
| 44 | 9,108.14 | 8,347.62 | 7,356.39 | 6,858.15 | 31,670.30 |
| 45 | 2,481.60 | 2,268.10 | 2,017.26 | 1,850.18 | 8,617.14 |
| 46 | 3,907.27 | 3,592.96 | 3,770.50 | 3,163.82 | 14,434.55 |
| 47 | 2,793.32 | 2,537.58 | 2,436.10 | 2,228.68 | 9,995.68 |
| 48 | 1,969.05 | 1,887.28 | 1,609.64 | 1,421.66 | 6,887.63 |
| 49 | 11,220.22 | 10,541.81 | 9,967.50 | 8,666.55 | 40,396.08 |
| TOTAL | 77,200.83 | 73,389.75 | 68,246.96 | 62,603.62 | 281,441.16 |
| STATE: | MARYLAND | | | | |
| ZIP CODE | QUARTER 1 | QUARTER 2 | QUARTER 3 | QUARTER 4 | TOTAL GRAMS |
| 206 | 26,983.66 | 26,731.93 | 27,123.60 | 26,696.14 | 107,535.33 |
| 207 | 30,592.76 | 30,542.23 | 29,811.81 | 29,179.55 | 120,126.35 |
| 208 | 27,762.42 | 25,531.05 | 26,599.14 | 25,051.42 | 104,944.03 |
| 209 | 5,028.26 | 4,726.35 | 4,991.55 | 4,751.02 | 19,497.18 |
| 210 | 60,427.71 | 59,719.07 | 58,341.94 | 56,500.57 | 234,989.29 |
| 211 | 28,272.09 | 28,366.56 | 28,164.92 | 27,367.55 | 112,171.12 |
| 212 | 113,925.50 | 109,291.65 | 110,481.23 | 108,130.84 | 441,829.22 |
| 214 | 5,153.25 | 5,160.78 | 4,986.46 | 4,727.42 | 20,027.91 |
| 215 | 9,793.95 | 9,477.91 | 9,408.52 | 9,254.60 | 37,934.98 |
| 216 | 10,636.56 | 10,513.77 | 10,689.06 | 10,923.68 | 42,763.07 |
| 217 | 30,692.02 | 29,926.01 | 30,661.29 | 30,422.06 | 121,701.38 |
| 218 | 10,896.08 | 10,545.97 | 10,603.35 | 10,350.23 | 42,395.63 |
| 219 | 9,932.28 | 9,042.82 | 8,946.51 | 8,541.38 | 36,462.99 |
| TOTAL | 370,096.54 | 359,576.10 | 360,809.38 | 351,896.46 | 1,442,378.48 |
| STATE: | MASSACHUSETTS | | | | |
| ZIP CODE | QUARTER 1 | QUARTER 2 | QUARTER 3 | QUARTER 4 | TOTAL GRAMS |
| 10 | 32,054.41 | 30,235.30 | 28,728.91 | 27,124.72 | 118,143.34 |

| | | | | | |
|---|---|---|---|---|---|
| 11 | 12,705.95 | 11,863.62 | 11,312.38 | 10,779.84 | 46,661.79 |
| 12 | 5,335.92 | 5,003.98 | 4,860.57 | 4,472.65 | 19,673.12 |
| 13 | 4,331.04 | 4,250.31 | 4,139.56 | 3,863.35 | 16,584.26 |
| 14 | 9,414.99 | 9,279.57 | 8,532.75 | 8,152.11 | 35,379.42 |
| 15 | 13,412.54 | 13,258.12 | 12,674.99 | 12,019.18 | 51,364.83 |
| 16 | 11,762.78 | 11,389.88 | 10,971.31 | 10,392.50 | 44,516.47 |
| 17 | 11,835.90 | 10,663.49 | 9,733.69 | 9,929.50 | 42,162.58 |
| 18 | 38,071.79 | 36,849.16 | 34,749.24 | 33,196.81 | 142,867 |
| 19 | 19,683.21 | 19,226.38 | 17,925.82 | 17,074.16 | 73,909.57 |
| 20 | 13,578.50 | 13,257 | 12,782.23 | 12,179.49 | 51,797.22 |
| 21 | 44,027.01 | 41,721.48 | 40,593.62 | 39,066.80 | 165,408.91 |
| 22 | 3,374.17 | 3,241.83 | 2,853.62 | 2,836.36 | 12,305.98 |
| 23 | 24,286.63 | 22,804.18 | 21,125.51 | 20,505.53 | 88,721.85 |
| 24 | 7,232.31 | 6,845.82 | 6,468.15 | 6,373.21 | 26,919.49 |
| 25 | 6,145.25 | 5,870.59 | 5,659.68 | 5,234.84 | 22,910.36 |
| 26 | 9,049.27 | 7,736.39 | 7,776.22 | 7,046.23 | 31,608.11 |
| 27 | 35,206.03 | 32,718.09 | 31,489.75 | 30,520.04 | 129,933.91 |
| TOTAL | 301,507.70 | 286,215.19 | 272,378 | 260,767.32 | 1,120,868.21 |
| STATE: | MICHIGAN | | | | |
| ZIP CODE | QUARTER 1 | QUARTER 2 | QUARTER 3 | QUARTER 4 | TOTAL GRAMS |
| 480 | 61,293.03 | 59,083.86 | 55,610.26 | 54,798.93 | 230,786.08 |
| 481 | 67,219.28 | 63,846.63 | 61,810.58 | 60,452.24 | 253,328.73 |
| 482 | 47,589.90 | 45,239.62 | 45,584.89 | 44,427.01 | 182,841.42 |
| 483 | 42,073.17 | 39,856.67 | 39,753.63 | 39,549.39 | 161,232.86 |
| 484 | 9,526.93 | 9,351.67 | 9,053.23 | 9,075.62 | 37,007.45 |
| 485 | 5,418.58 | 5,233.55 | 4,970.39 | 4,960.75 | 20,583.27 |
| 486 | 11,417.90 | 11,901.06 | 11,188.16 | 11,021.80 | 45,528.92 |
| 487 | 6,471.45 | 6,427.98 | 6,369.04 | 5,764.48 | 25,032.95 |
| 488 | 12,607.62 | 11,872.38 | 10,976.87 | 11,696.52 | 47,153.39 |
| 489 | 6,279.63 | 5,537.99 | 5,101.31 | 5,371.10 | 22,290.03 |
| 490 | 22,037.39 | 20,952.15 | 19,942.70 | 20,019.14 | 82,951.38 |
| 491 | 2,357.56 | 2,274.17 | 2,350.79 | 2,283.13 | 9,265.65 |
| 492 | 12,852.14 | 12,350.52 | 11,132.74 | 11,399.24 | 47,734.64 |
| 493 | 3,989.34 | 3,726.54 | 3,264.48 | 3,505.83 | 14,486.19 |
| 494 | 8,650.35 | 8,176.89 | 7,618.29 | 7,911.95 | 32,357.48 |
| 495 | 8,656.90 | 8,312.10 | 7,692.54 | 8,185.45 | 32,846.99 |
| 496 | 10,279.59 | 10,174.53 | 9,928.31 | 9,379.66 | 39,762.09 |
| 497 | 9,771.68 | 9,584.87 | 9,133.45 | 9,021.54 | 37,511.54 |
| 498 | 5,132.76 | 4,835.45 | 5,182.10 | 4,600.93 | 19,751.24 |
| 499 | 1,795.64 | 1,743.56 | 1,664.40 | 1,637.18 | 6,840.78 |

DRUG CODE:9143DRUG NAME:OXYCODONE
STATE:MICHIGAN

| ZIP CODE | QUARTER 1 | QUARTER 2 | QUARTER 3 | QUARTER 4 | TOTAL GRAMS |
|---|---|---|---|---|---|
| TOTAL | 355,420.84 | 340,482.19 | 328,328.16 | 325,061.89 | 1,349,293.08 |
| STATE: | MINNESOTA | | | | |
| ZIP CODE | QUARTER 1 | QUARTER 2 | QUARTER 3 | QUARTER 4 | TOTAL GRAMS |
| 550 | 18,928.32 | 17,937.23 | 17,814.03 | 17,652.45 | 72,332.03 |
| 551 | 29,235.82 | 28,755.29 | 27,919.90 | 27,910.71 | 113,821.72 |
| 553 | 25,880.52 | 25,042.19 | 25,111.51 | 25,123.51 | 101,157.73 |
| 554 | 45,716.48 | 44,344.27 | 43,995.36 | 44,391.70 | 178,447.81 |
| 556 | 245.19 | 287.91 | 239.05 | 222.66 | 994.81 |
| 557 | 6,482.18 | 5,719.20 | 5,375.97 | 4,859 | 22,436.35 |
| 558 | 3,855.49 | 3,446.77 | 3,247.80 | 2,930.32 | 13,480.38 |
| 559 | 9,198.38 | 9,002.63 | 8,609.17 | 8,225.54 | 35,035.72 |
| 560 | 5,058.97 | 4,701.19 | 4,834.87 | 4,792.95 | 19,387.98 |
| 561 | 1,255.12 | 1,267.92 | 1,233.21 | 1,200.75 | 4,957 |
| 562 | 2,811.19 | 2,706.68 | 2,663 | 2,549.26 | 10,730.13 |

| | | | | | |
|---|---|---|---|---|---|
| 563 | 8,947.80 | 8,641.20 | 8,405.20 | 8,019.57 | 34,013.77 |
| 564 | 4,048.80 | 4,120.41 | 3,988.62 | 3,646.18 | 15,804.01 |
| 565 | 3,769.64 | 3,611.67 | 3,534.15 | 3,126.65 | 14,042.11 |
| 566 | 1,572.43 | 1,603.93 | 1,680.24 | 1,614.11 | 6,470.71 |
| 567 | 1,800.81 | 1,697.93 | 1,618.70 | 1,616.51 | 6,733.95 |
| TOTAL | 168,807.14 | 162,886.42 | 160,270.78 | 157,881.87 | 649,846.21 |

STATE: MISSISSIPPI

| ZIP CODE | QUARTER 1 | QUARTER 2 | QUARTER 3 | QUARTER 4 | TOTAL GRAMS |
|---|---|---|---|---|---|
| 386 | 14,261.65 | 14,319.54 | 14,305.93 | 14,340.24 | 57,227.36 |
| 387 | 1,866.32 | 1,876.73 | 1,990.72 | 1,993.55 | 7,727.32 |
| 388 | 8,849.59 | 9,041.67 | 9,398.28 | 9,674.39 | 36,963.93 |
| 389 | 1,976.22 | 1,981.88 | 2,030.02 | 2,098.94 | 8,087.06 |
| 390 | 6,064.54 | 6,172.90 | 6,239.54 | 6,237.60 | 24,714.58 |
| 391 | 5,660.99 | 5,588.55 | 5,640.94 | 6,110.66 | 23,001.14 |
| 392 | 6,738.02 | 6,387.73 | 6,400.46 | 6,717.89 | 26,244.10 |
| 393 | 7,373.59 | 7,345.24 | 7,471.52 | 7,266.61 | 29,456.96 |
| 394 | 21,440.04 | 21,439.36 | 21,651.58 | 21,844.45 | 86,375.43 |
| 395 | 27,477.24 | 26,560.72 | 26,113.94 | 25,476.70 | 105,628.60 |
| 396 | 5,937.21 | 5,992.74 | 5,819.94 | 6,250.73 | 24,000.62 |
| 397 | 3,668.23 | 3,820.75 | 4,064.52 | 3,994.18 | 15,547.68 |
| TOTAL | 111,313.64 | 110,527.81 | 111,127.39 | 112,005.94 | 444,974.78 |

STATE: MISSOURI

| ZIP CODE | QUARTER 1 | QUARTER 2 | QUARTER 3 | QUARTER 4 | TOTAL GRAMS |
|---|---|---|---|---|---|
| 630 | 40,892.62 | 38,840.10 | 39,632.56 | 38,133.04 | 157,498.32 |
| 631 | 69,455.15 | 69,367.03 | 71,114.45 | 62,595.21 | 272,531.84 |
| 633 | 20,856.86 | 19,319.91 | 19,594.01 | 19,845.77 | 79,616.55 |
| 634 | 1,710.55 | 1,656.01 | 1,718.12 | 1,525.73 | 6,610.41 |
| 635 | 2,780.13 | 2,664.76 | 2,727.90 | 2,591.98 | 10,764.77 |
| 636 | 11,492.76 | 11,461.19 | 11,520.88 | 11,473.26 | 45,948.09 |
| 637 | 6,428.01 | 6,309.59 | 6,077.50 | 6,134.46 | 24,949.56 |
| 638 | 6,293.21 | 6,163.38 | 6,148.89 | 6,295.17 | 24,900.65 |
| 639 | 3,352.13 | 2,981.66 | 3,085.06 | 2,928.87 | 12,347.72 |
| 640 | 29,033.91 | 28,356.11 | 28,474.99 | 27,650.41 | 113,515.42 |
| 641 | 28,025.89 | 26,581.53 | 25,776.36 | 25,812.44 | 106,196.22 |
| 644 | 2,834.44 | 2,652.51 | 2,534.18 | 2,763.25 | 10,784.38 |
| 645 | 6,275.59 | 6,184.71 | 5,940.03 | 5,824.38 | 24,224.71 |
| 646 | 2,739.75 | 2,556.45 | 2,496.67 | 2,940.21 | 10,733.08 |
| 647 | 7,705.95 | 7,610.50 | 7,428.20 | 7,302.22 | 30,046.87 |
| 648 | 11,875.22 | 12,367.20 | 12,341.70 | 12,003.10 | 48,587.22 |
| 650 | 5,213.99 | 5,142.77 | 5,146.66 | 4,862.86 | 20,366.28 |
| 651 | 2,613.97 | 2,433.43 | 2,289.84 | 2,250.51 | 9,587.75 |
| 652 | 12,577.36 | 12,622.84 | 12,455.41 | 12,322.38 | 49,977.99 |
| 653 | 3,928.39 | 3,812.84 | 3,856.09 | 4,012.88 | 15,610.20 |
| 654 | 7,249.08 | 7,065.54 | 7,049.33 | 6,715.90 | 28,079.85 |
| 655 | 8,067.52 | 8,282.32 | 8,389.52 | 8,382.47 | 33,121.83 |
| 656 | 12,618.29 | 12,474.57 | 12,712.27 | 12,112.88 | 49,918.01 |
| 657 | 17,621.39 | 17,047.88 | 16,829.14 | 16,321.58 | 67,819.99 |
| 658 | 18,651.63 | 18,368.98 | 18,654.57 | 17,879.15 | 73,554.33 |
| TOTAL | 340,293.79 | 332,323.81 | 333,994.33 | 320,680.11 | 1,327,292.04 |

STATE: MONTANA

| ZIP CODE | QUARTER 1 | QUARTER 2 | QUARTER 3 | QUARTER 4 | TOTAL GRAMS |
|---|---|---|---|---|---|
| 590 | 3,139.31 | 3,158.98 | 2,822.63 | 2,786.06 | 11,906.98 |

DRUG CODE:9143DRUG NAME:OXYCODONE
STATE:MONTANA

| ZIP CODE | QUARTER 1 | QUARTER 2 | QUARTER 3 | QUARTER 4 | TOTAL GRAMS |
|---|---|---|---|---|---|

| | | | | | |
|---|---|---|---|---|---|
| 591 | 11,249.49 | 10,453.28 | 9,959.91 | 9,790.13 | 41,452.81 |
| 592 | 1,535.93 | 1,517.84 | 1,434.13 | 1,295 | 5,782.90 |
| 593 | 1,971.97 | 1,807.48 | 1,931.17 | 1,733.88 | 7,444.50 |
| 594 | 7,995.60 | 7,193.84 | 7,011.58 | 6,406.71 | 28,607.73 |
| 595 | 853.71 | 869.3 | 980.33 | 850.17 | 3,553.51 |
| 596 | 3,845.08 | 3,813.03 | 3,505.23 | 3,429.98 | 14,593.32 |
| 597 | 5,915.26 | 5,503.28 | 5,510.84 | 5,270.23 | 22,199.61 |
| 598 | 6,852.58 | 6,796.50 | 6,410.46 | 6,233.13 | 26,292.67 |
| 599 | 5,424.23 | 5,254.13 | 5,364.29 | 5,126.82 | 21,169.47 |
| TOTAL | 48,783.16 | 46,367.66 | 44,930.57 | 42,922.11 | 183,003.50 |
| STATE: | NEBRASKA | | | | |
| ZIP CODE | QUARTER 1 | QUARTER 2 | QUARTER 3 | QUARTER 4 | TOTAL GRAMS |
| 680 | 4,460.30 | 4,124.29 | 4,124.42 | 4,096.36 | 16,805.37 |
| 681 | 20,020.21 | 19,762.25 | 20,624.31 | 20,592.46 | 80,999.23 |
| 683 | 1,371.93 | 1,355.08 | 1,290.60 | 1,200.68 | 5,218.29 |
| 684 | 1,093.90 | 1,032.27 | 1,092.70 | 1,037.03 | 4,255.90 |
| 685 | 5,959.93 | 5,748.02 | 5,804.41 | 5,600.62 | 23,112.98 |
| 686 | 644.98 | 617.96 | 634.78 | 602.36 | 2,500.08 |
| 687 | 1,598.64 | 1,513.52 | 1,556.41 | 1,493.39 | 6,161.96 |
| 688 | 3,258.31 | 3,260.42 | 3,273.35 | 3,325.24 | 13,117.32 |
| 689 | 1,534.09 | 1,349.92 | 1,358.35 | 1,411.89 | 5,654.25 |
| 690 | 795.82 | 707.22 | 752.43 | 716.93 | 2,972.40 |
| 691 | 2,079.49 | 2,084.90 | 2,115.79 | 1,896.92 | 8,177.10 |
| 692 | 250.12 | 228.61 | 236.68 | 270.34 | 985.75 |
| 693 | 2,097.32 | 2,090.06 | 2,005.02 | 2,052 | 8,244.40 |
| TOTAL | 45,165.04 | 43,874.52 | 44,869.25 | 44,296.22 | 178,205.03 |
| STATE: | NEVADA | | | | |
| ZIP CODE | QUARTER 1 | QUARTER 2 | QUARTER 3 | QUARTER 4 | TOTAL GRAMS |
| 890 | 44,361.38 | 43,016.33 | 43,653.59 | 44,099.24 | 175,130.54 |
| 891 | 108,289.24 | 102,759.63 | 102,122.47 | 103,870.03 | 417,041.37 |
| 893 | 934.29 | 889.01 | 835.99 | 751.49 | 3,410.78 |
| 894 | 18,327.39 | 16,792.23 | 15,956.61 | 15,570.84 | 66,647.07 |
| 895 | 23,671.74 | 21,904.43 | 20,365.82 | 19,650.47 | 85,592.46 |
| 897 | 6,163.20 | 6,095.08 | 6,071.85 | 6,001.98 | 24,332.11 |
| 898 | 2,358.44 | 2,054.55 | 1,921.89 | 1,904.48 | 8,239.36 |
| TOTAL | 204,105.68 | 193,511.26 | 190,928.22 | 191,848.53 | 780,393.69 |
| STATE: | NEW HAMPSHIRE | | | | |
| ZIP CODE | QUARTER 1 | QUARTER 2 | QUARTER 3 | QUARTER 4 | TOTAL GRAMS |
| 30 | 16,965.71 | 15,823.27 | 15,552.93 | 14,839.29 | 63,181.20 |
| 31 | 10,319.28 | 9,803.27 | 9,359.73 | 9,134.09 | 38,616.37 |
| 32 | 8,883.68 | 8,550.92 | 8,462.90 | 7,744.97 | 33,642.47 |
| 33 | 5,747.83 | 5,359.15 | 5,482.04 | 5,011.55 | 21,600.57 |
| 34 | 3,173.75 | 3,221.82 | 2,999.95 | 2,952.23 | 12,347.75 |
| 35 | 3,720.12 | 3,243.45 | 3,334.13 | 3,005.49 | 13,303.19 |
| 36 | 279.93 | 309.07 | 284.41 | 280.6 | 1,154.01 |
| 37 | 5,205.66 | 4,936.95 | 4,724.21 | 4,351.52 | 19,218.34 |
| 38 | 23,833.62 | 22,332.24 | 21,456 | 20,665.58 | 88,287.44 |
| TOTAL | 78,129.58 | 73,580.14 | 71,656.30 | 67,985.32 | 291,351.34 |
| STATE: | NEW JERSEY | | | | |
| ZIP CODE | QUARTER 1 | QUARTER 2 | QUARTER 3 | QUARTER 4 | TOTAL GRAMS |
| 70 | 89,935.61 | 85,973.21 | 83,697.66 | 80,759.74 | 340,366.22 |
| 71 | 20,749.71 | 20,613.80 | 19,966.92 | 19,557.40 | 80,887.83 |
| 72 | 11,119.48 | 10,521.15 | 8,101.54 | 6,837.51 | 36,579.68 |
| 73 | 11,784.79 | 11,840.19 | 11,285.28 | 11,046.07 | 45,956.33 |
| 74 | 27,666.51 | 26,270.77 | 25,454.20 | 24,669.25 | 104,060.73 |
| 75 | 11,706.23 | 10,541.38 | 10,032.74 | 10,008.01 | 42,288.36 |
| 76 | 24,771.14 | 24,316.55 | 26,383.24 | 27,553.21 | 103,024.14 |
| 77 | 41,038 | 40,178.42 | 39,758.90 | 37,812.02 | 158,787.34 |

| | | | | | |
|---|---|---|---|---|---|
| 78 | 20,036.61 | 19,387.43 | 18,755.64 | 17,172.40 | 75,352.08 |
| 79 | 8,589.48 | 8,155.45 | 7,858.40 | 7,577.07 | 32,180.40 |
| 80 | 143,991.13 | 140,474.78 | 135,995.38 | 130,526.86 | 550,988.15 |
| 81 | 17,246.32 | 17,269.32 | 17,663.45 | 17,611.63 | 69,790.72 |
| 82 | 27,003.62 | 27,323.64 | 27,663.61 | 26,123.78 | 108,114.65 |
| 83 | 20,494.99 | 18,980.49 | 18,098.18 | 17,468.30 | 75,041.96 |
| 84 | 5,953.43 | 5,887.41 | 5,590.65 | 4,903.02 | 22,334.51 |
| 85 | 15,899.63 | 15,441.99 | 15,243 | 15,755.93 | 62,340.55 |

DRUG CODE:9143DRUG NAME:OXYCODONE
STATE:NEW JERSEY

| ZIP CODE | QUARTER 1 | QUARTER 2 | QUARTER 3 | QUARTER 4 | TOTAL GRAMS |
|---|---|---|---|---|---|
| 86 | 18,502.13 | 18,493.44 | 18,172.25 | 17,299.20 | 72,467.02 |
| 87 | 33,987.91 | 32,195.83 | 31,973.26 | 30,866.67 | 129,023.67 |
| 88 | 57,536.52 | 56,677.19 | 55,687.88 | 55,024.19 | 224,925.78 |
| 89 | 5,712.22 | 5,385.18 | 5,446.92 | 4,907.61 | 21,451.93 |
| TOTAL | 613,725.46 | 595,927.62 | 582,829.10 | 563,479.87 | 2,355,962.05 |

STATE: NEW MEXICO

| ZIP CODE | QUARTER 1 | QUARTER 2 | QUARTER 3 | QUARTER 4 | TOTAL GRAMS |
|---|---|---|---|---|---|
| 870 | 10,532.17 | 10,204.46 | 10,123.58 | 10,105.61 | 40,965.82 |
| 871 | 57,932.89 | 55,091.82 | 54,734.93 | 52,647.57 | 220,407.21 |
| 873 | 1,346.81 | 1,291.40 | 1,237.26 | 1,271.69 | 5,147.16 |
| 874 | 3,161.21 | 3,257.39 | 3,106.69 | 2,979.97 | 12,505.26 |
| 875 | 16,055.03 | 15,519.19 | 15,530.52 | 15,645.60 | 62,750.34 |
| 877 | 1,811.60 | 1,694.82 | 1,784.98 | 1,933.26 | 7,224.66 |
| 878 | 686.94 | 652.19 | 615.75 | 617.55 | 2,572.43 |
| 879 | 820.66 | 658.7 | 569.05 | 579.9 | 2,628.31 |
| 880 | 8,964.24 | 8,287.52 | 7,443.20 | 7,373.46 | 32,068.42 |
| 881 | 2,424.75 | 2,342.11 | 1,748.53 | 1,355.78 | 7,871.17 |
| 882 | 8,899.96 | 8,334.22 | 8,066 | 8,129.33 | 33,429.51 |
| 883 | 3,870.05 | 3,609.85 | 3,329.94 | 3,427.41 | 14,237.25 |
| 884 | 415.3 | 316.69 | 312.85 | 348.51 | 1,393.35 |
| TOTAL | 116,921.61 | 111,260.36 | 108,603.28 | 106,415.64 | 443,200.89 |

STATE: NEW YORK

| ZIP CODE | QUARTER 1 | QUARTER 2 | QUARTER 3 | QUARTER 4 | TOTAL GRAMS |
|---|---|---|---|---|---|
| 63 | 0 | 0 | 0.45 | 0 | 0.45 |
| 100 | 89,632.32 | 86,296.45 | 82,469.48 | 82,016.94 | 340,415.19 |
| 101 | 2,118.46 | 1,967.53 | 1,766.43 | 1,756.36 | 7,608.78 |
| 102 | 72.29 | 86.41 | 84.36 | 100.68 | 343.74 |
| 103 | 34,769.11 | 34,020.46 | 33,736.20 | 33,269.78 | 135,795.55 |
| 104 | 75,617.66 | 71,378.28 | 68,892.20 | 65,602.78 | 281,490.92 |
| 105 | 22,141.14 | 21,372.22 | 20,863.41 | 20,802.68 | 85,179.45 |
| 106 | 4,213.56 | 3,921.58 | 3,691.68 | 3,728.05 | 15,554.87 |
| 107 | 9,026.56 | 8,883.42 | 8,491.17 | 8,506.88 | 34,908.03 |
| 108 | 3,017.24 | 3,035.73 | 2,909.20 | 2,816.60 | 11,778.77 |
| 109 | 33,908.26 | 32,522.92 | 31,729.54 | 30,657.79 | 128,818.51 |
| 110 | 13,903.16 | 13,572.32 | 13,352.89 | 13,151.94 | 53,980.31 |
| 111 | 10,992.10 | 10,479.72 | 9,677.46 | 9,526.02 | 40,675.30 |
| 112 | 84,390.22 | 82,446.13 | 82,220.65 | 80,719.25 | 329,776.25 |
| 113 | 35,278.82 | 32,907.44 | 31,621.30 | 31,364.46 | 131,172.02 |
| 114 | 18,436.54 | 17,270.75 | 16,676.60 | 17,110.83 | 69,494.72 |
| 115 | 35,262.70 | 33,851.81 | 33,562.39 | 32,214.60 | 134,891.50 |
| 116 | 4,124.28 | 4,162.63 | 4,091.80 | 4,061.95 | 16,440.66 |
| 117 | 107,656.99 | 106,797.45 | 104,905.01 | 107,244.64 | 426,604.09 |
| 118 | 4,362.95 | 4,209.39 | 3,837.94 | 3,625.20 | 16,035.48 |
| 119 | 10,995 | 10,976.23 | 10,771.80 | 10,343.50 | 43,086.53 |
| 120 | 11,524.63 | 11,277.50 | 10,822.65 | 10,493.14 | 44,117.92 |
| 121 | 6,404.72 | 6,432.19 | 6,055.63 | 5,931.40 | 24,823.94 |

| | | | | | |
|---|---|---|---|---|---|
| 122 | 6,695.07 | 6,259.48 | 6,016.46 | 6,102.06 | 25,073.07 |
| 123 | 3,429.02 | 3,234.05 | 3,308.35 | 3,271.75 | 13,243.17 |
| 124 | 15,989.51 | 14,979.04 | 15,386.17 | 14,667.34 | 61,022.06 |
| 125 | 26,416.88 | 25,463.91 | 25,259.20 | 24,851.02 | 101,991.01 |
| 126 | 8,064.23 | 7,470.97 | 7,398.81 | 7,024.05 | 29,958.06 |
| 127 | 13,493.42 | 13,179.98 | 12,569.09 | 12,335.67 | 51,578.16 |
| 128 | 10,147.97 | 10,104.59 | 9,929.61 | 9,608.40 | 39,790.57 |
| 129 | 6,996.40 | 7,053.73 | 6,778.91 | 6,593.04 | 27,422.08 |
| 130 | 15,645.96 | 14,169.75 | 13,282.96 | 12,922.44 | 56,021.11 |
| 131 | 4,650.06 | 4,212.74 | 4,188.66 | 3,952.54 | 17,004 |
| 132 | 11,722.30 | 10,127.42 | 9,665.96 | 8,572.73 | 40,088.41 |
| 133 | 5,008.94 | 4,753.82 | 4,582.75 | 4,416.12 | 18,761.63 |
| 134 | 6,087.98 | 5,574.18 | 5,458.64 | 5,090.68 | 22,211.48 |
| 135 | 2,445.05 | 2,508.24 | 2,375.10 | 2,197.92 | 9,526.31 |
| 136 | 7,758.68 | 7,667.43 | 7,702.35 | 7,194.49 | 30,322.95 |
| 137 | 5,579.64 | 5,184.17 | 4,963.72 | 5,010.86 | 20,738.39 |
| 138 | 9,977.68 | 9,620.11 | 9,740.48 | 9,497.98 | 38,836.25 |
| 139 | 6,294.05 | 5,935.54 | 5,904.83 | 5,523.30 | 23,657.72 |
| 140 | 18,253.26 | 16,144.19 | 15,500.01 | 14,702.88 | 64,600.34 |
| 141 | 8,882.60 | 7,795.92 | 7,027.39 | 6,861.17 | 30,567.08 |

DRUG CODE:9143DRUG NAME:OXYCODONE
STATE:NEW YORK

| ZIP CODE | QUARTER 1 | QUARTER 2 | QUARTER 3 | QUARTER 4 | TOTAL GRAMS |
|---|---|---|---|---|---|
| 142 | 24,657.49 | 21,471.87 | 20,521.77 | 19,761.67 | 86,412.80 |
| 143 | 3,311.72 | 2,942.85 | 2,706.34 | 2,588.93 | 11,549.84 |
| 144 | 10,158.96 | 9,685.92 | 9,411.66 | 9,281.67 | 38,538.21 |
| 145 | 9,418.55 | 9,173.75 | 8,969.46 | 8,453.52 | 36,015.28 |
| 146 | 23,370.47 | 22,409.05 | 21,931.73 | 21,197.80 | 88,909.05 |
| 147 | 5,694.61 | 4,960.58 | 4,604.30 | 4,428.32 | 19,687.81 |
| 148 | 13,636.98 | 12,986.09 | 12,967.65 | 12,424.60 | 52,015.32 |
| 149 | 5,157.20 | 4,425.29 | 4,138.39 | 3,829.55 | 17,550.43 |
| TOTAL | 906,793.39 | 867,363.22 | 844,520.99 | 827,407.97 | 3,446,085.57 |

STATE: NORTH CAROLINA

| ZIP CODE | QUARTER 1 | QUARTER 2 | QUARTER 3 | QUARTER 4 | TOTAL GRAMS |
|---|---|---|---|---|---|
| 270 | 21,472.44 | 21,328.05 | 20,655.21 | 19,787.70 | 83,243.40 |
| 271 | 16,494.89 | 15,922.99 | 15,191.22 | 14,301.44 | 61,910.54 |
| 272 | 38,387.52 | 37,470.58 | 36,732.43 | 36,941.64 | 149,532.17 |
| 273 | 26,191.26 | 25,178.97 | 24,986.13 | 24,607.66 | 100,964.02 |
| 274 | 14,462.19 | 13,924.32 | 13,756.39 | 13,522.96 | 55,665.86 |
| 275 | 58,379.51 | 55,737.87 | 53,345.35 | 53,049.79 | 220,512.52 |
| 276 | 16,307.41 | 15,667.11 | 15,662.61 | 16,625.14 | 64,262.27 |
| 277 | 18,356.59 | 17,007.84 | 17,177.56 | 16,280.35 | 68,822.34 |
| 278 | 33,674.70 | 32,879.87 | 32,809.89 | 32,738.14 | 132,102.60 |
| 279 | 13,617.03 | 13,647.60 | 13,520.41 | 13,256.10 | 54,041.14 |
| 280 | 42,819.22 | 41,771.73 | 41,174.74 | 39,916.76 | 165,682.45 |
| 281 | 33,738.05 | 32,457.64 | 31,722.84 | 31,127.85 | 129,046.38 |
| 282 | 25,568.98 | 24,474.83 | 23,966.68 | 24,398.21 | 98,408.70 |
| 283 | 84,280.40 | 79,612.48 | 78,556.80 | 77,593.06 | 320,042.74 |
| 284 | 53,725.13 | 50,555.50 | 49,698.92 | 47,677.14 | 201,656.69 |
| 285 | 40,707.10 | 39,899.30 | 39,132.01 | 37,821.29 | 157,559.70 |
| 286 | 69,473.60 | 66,592.07 | 64,923.55 | 62,792.18 | 263,781.40 |
| 287 | 35,256.26 | 33,749.11 | 33,054.52 | 31,963.32 | 134,023.21 |
| 288 | 15,044.75 | 14,234.64 | 13,446.60 | 13,603.63 | 56,329.62 |
| 289 | 3,433.51 | 3,318.14 | 3,400.76 | 3,358.42 | 13,510.83 |
| TOTAL | 661,390.54 | 635,430.64 | 622,914.62 | 611,362.78 | 2,531,098.58 |

STATE: NORTH DAKOTA

| ZIP CODE | QUARTER 1 | QUARTER 2 | QUARTER 3 | QUARTER 4 | TOTAL GRAMS |
|---|---|---|---|---|---|

| | | | | | |
|---|---|---|---|---|---|
| 580 | 947.25 | 866.87 | 759.11 | 702.41 | 3,275.64 |
| 581 | 3,709.55 | 3,376.02 | 3,210.63 | 2,789.68 | 13,085.88 |
| 582 | 3,273.42 | 2,956.19 | 2,638.06 | 2,313.91 | 11,181.58 |
| 583 | 1,670.45 | 1,601.96 | 1,581.43 | 1,378.32 | 6,232.16 |
| 584 | 1,012.10 | 958.76 | 830.96 | 767.72 | 3,569.54 |
| 585 | 4,912.73 | 5,086.83 | 4,524.41 | 4,106.16 | 18,630.13 |
| 586 | 1,109.08 | 1,053.94 | 1,000.45 | 858.27 | 4,021.74 |
| 587 | 1,546.66 | 1,430.24 | 1,507.71 | 1,270.50 | 5,755.11 |
| 588 | 942.4 | 940.07 | 935.95 | 802.3 | 3,620.72 |
| TOTAL | 19,123.64 | 18,270.88 | 16,988.71 | 14,989.27 | 69,372.50 |

STATE: OHIO

| ZIP CODE | QUARTER 1 | QUARTER 2 | QUARTER 3 | QUARTER 4 | TOTAL GRAMS |
|---|---|---|---|---|---|
| 430 | 33,550.37 | 32,544.02 | 31,603.63 | 31,706.04 | 129,404.06 |
| 431 | 26,166.90 | 25,390.93 | 24,306.53 | 24,187.27 | 100,051.63 |
| 432 | 59,350.56 | 59,647.22 | 59,563 | 54,268.18 | 232,828.96 |
| 433 | 7,587.76 | 7,447.58 | 7,315.84 | 7,238.74 | 29,589.92 |
| 434 | 6,341.22 | 6,053.17 | 6,046.69 | 5,999.02 | 24,440.10 |
| 435 | 10,208.83 | 10,047.51 | 9,548.90 | 9,528.07 | 39,333.31 |
| 436 | 26,174.05 | 25,619.11 | 24,995.11 | 24,205.52 | 100,993.79 |
| 437 | 12,291.85 | 12,224.61 | 11,999.36 | 11,221.69 | 47,737.51 |
| 438 | 1,972.57 | 1,909.50 | 1,878.44 | 1,699.45 | 7,459.96 |
| 439 | 12,164.75 | 12,009.73 | 11,948.33 | 11,158.85 | 47,281.66 |
| 440 | 31,456.03 | 29,510.73 | 29,380.07 | 29,049.62 | 119,396.45 |
| 441 | 45,033.72 | 43,163.35 | 43,530.06 | 42,154.77 | 173,881.90 |
| 442 | 23,332.87 | 21,282.02 | 20,758.05 | 20,671.70 | 86,044.64 |
| 443 | 15,598.34 | 14,194.15 | 14,610.49 | 14,129.73 | 58,532.71 |
| 444 | 21,364.42 | 20,223.12 | 20,575.56 | 19,718.15 | 81,881.25 |
| 445 | 11,198.38 | 10,553.44 | 10,619.21 | 10,356.73 | 42,727.76 |
| 446 | 15,036.80 | 13,696.79 | 13,830.98 | 13,539.62 | 56,104.19 |
| 447 | 9,574.35 | 8,524.39 | 8,574.31 | 8,473.16 | 35,146.21 |
| 448 | 14,645.97 | 14,296.73 | 13,319.04 | 13,520.35 | 55,782.09 |
| 449 | 4,555.39 | 4,288.87 | 4,136.08 | 4,025.43 | 17,005.77 |

DRUG CODE:9143DRUG NAME:OXYCODONE
STATE:OHIO

| ZIP CODE | QUARTER 1 | QUARTER 2 | QUARTER 3 | QUARTER 4 | TOTAL GRAMS |
|---|---|---|---|---|---|
| 450 | 37,582.73 | 36,237.38 | 36,006.39 | 36,074.63 | 145,901.13 |
| 451 | 15,800.51 | 15,347.65 | 15,126 | 14,961.83 | 61,235.99 |
| 452 | 47,752.76 | 46,840.14 | 47,014.81 | 45,923.61 | 187,531.32 |
| 453 | 15,982.97 | 15,045.50 | 14,984.04 | 14,965.34 | 60,977.85 |
| 454 | 22,581.66 | 21,436.19 | 20,995.16 | 21,327.72 | 86,340.73 |
| 455 | 6,271.85 | 6,333.04 | 6,289.93 | 6,137.39 | 25,032.21 |
| 456 | 25,803.28 | 25,191.20 | 24,958.27 | 24,190.91 | 100,143.66 |
| 457 | 8,392.08 | 8,354.55 | 8,190.82 | 8,023.93 | 32,961.38 |
| 458 | 7,951.78 | 7,788.84 | 7,557.67 | 7,805.87 | 31,104.16 |
| TOTAL | 575,724.75 | 555,201.46 | 549,662.77 | 536,263.32 | 2,216,852.30 |

STATE: OKLAHOMA

| ZIP CODE | QUARTER 1 | QUARTER 2 | QUARTER 3 | QUARTER 4 | TOTAL GRAMS |
|---|---|---|---|---|---|
| 730 | 41,117.16 | 40,268.50 | 39,964.14 | 39,768.39 | 161,118.19 |
| 731 | 49,395.92 | 47,765.17 | 48,847.64 | 47,127.03 | 193,135.76 |
| 734 | 5,030.89 | 4,918.89 | 5,054.12 | 4,660.17 | 19,664.07 |
| 735 | 12,599.99 | 12,726.53 | 12,919.28 | 12,086.11 | 50,331.91 |
| 736 | 5,668.38 | 5,706.79 | 6,049.24 | 5,667.33 | 23,091.74 |
| 737 | 3,789.88 | 3,956.50 | 3,810.53 | 4,027.66 | 15,584.57 |
| 738 | 930.51 | 956.12 | 882.96 | 949.3 | 3,718.89 |
| 739 | 319.12 | 182.86 | 182.59 | 192.39 | 876.96 |
| 740 | 38,386.22 | 37,460.68 | 37,714.86 | 37,590.12 | 151,151.88 |
| 741 | 30,339.26 | 29,156.52 | 28,111.38 | 27,685.84 | 115,293 |

| | | | | | |
|---|---|---|---|---|---|
| 743 | 6,600.93 | 6,446.55 | 6,336.97 | 6,583.99 | 25,968.44 |
| 744 | 18,545.08 | 17,743.32 | 17,810.62 | 17,651.17 | 71,750.19 |
| 745 | 4,732.79 | 4,573.78 | 4,499.10 | 4,568.01 | 18,373.68 |
| 746 | 2,868.53 | 2,837.28 | 2,735.13 | 2,616.50 | 11,057.44 |
| 747 | 4,274.51 | 4,425.08 | 4,468.47 | 4,223.86 | 17,391.92 |
| 748 | 11,263.23 | 11,019.02 | 10,952.16 | 11,381.79 | 44,616.20 |
| 749 | 10,527.55 | 7,826.71 | 7,909.55 | 8,672.27 | 34,936.08 |
| TOTAL | 246,389.95 | 237,970.30 | 238,248.74 | 235,451.93 | 958,060.92 |

STATE: OREGON

| ZIP CODE | QUARTER 1 | QUARTER 2 | QUARTER 3 | QUARTER 4 | TOTAL GRAMS |
|---|---|---|---|---|---|
| 970 | 35,443.37 | 33,709.13 | 32,999.44 | 31,406.34 | 133,558.28 |
| 971 | 15,769.87 | 14,821.97 | 14,889.29 | 14,022.19 | 59,503.32 |
| 972 | 45,095.36 | 42,775.54 | 41,747.25 | 38,927.74 | 168,545.89 |
| 973 | 30,717.18 | 28,944.67 | 28,494.79 | 26,923.48 | 115,080.12 |
| 974 | 41,413.88 | 39,457.71 | 38,462.19 | 36,380.91 | 155,714.69 |
| 975 | 25,651.66 | 24,764.14 | 23,954.38 | 23,004.25 | 97,374.43 |
| 976 | 3,196.30 | 3,108.70 | 3,061.05 | 3,119.28 | 12,485.33 |
| 977 | 8,764.57 | 8,653.94 | 8,841.23 | 8,197.50 | 34,457.24 |
| 978 | 8,533.29 | 8,377.37 | 7,986.35 | 7,922.95 | 32,819.96 |
| 979 | 1,926.72 | 1,791.77 | 1,851.21 | 1,909.11 | 7,478.81 |
| TOTAL | 216,512.20 | 206,404.94 | 202,287.18 | 191,813.75 | 817,018.07 |

STATE: PENNSYLVANIA

| ZIP CODE | QUARTER 1 | QUARTER 2 | QUARTER 3 | QUARTER 4 | TOTAL GRAMS |
|---|---|---|---|---|---|
| 150 | 27,932.64 | 26,906.11 | 26,071.70 | 26,684.90 | 107,595.35 |
| 151 | 15,961.23 | 14,248.06 | 14,061.41 | 14,055.41 | 58,326.11 |
| 152 | 42,638.16 | 39,226.08 | 39,014.93 | 38,290.55 | 159,169.72 |
| 153 | 8,564.97 | 8,067.14 | 7,532.23 | 7,586.38 | 31,750.72 |
| 154 | 10,671.24 | 10,501.14 | 10,897.68 | 10,688.51 | 42,758.57 |
| 155 | 4,623.38 | 4,355.42 | 4,203.39 | 4,202.12 | 17,384.31 |
| 156 | 17,102.92 | 16,381.49 | 16,505.67 | 16,431.08 | 66,421.16 |
| 157 | 6,678.10 | 6,398.67 | 6,241.08 | 6,349.40 | 25,667.25 |
| 158 | 5,403.29 | 5,424.64 | 5,511.78 | 5,038.22 | 21,377.93 |
| 159 | 15,618.82 | 14,766.07 | 14,815.32 | 15,231.77 | 60,431.98 |
| 160 | 8,873.99 | 8,315.41 | 8,218.84 | 8,292.24 | 33,700.48 |
| 161 | 14,078.99 | 12,863.52 | 12,213.16 | 12,255.90 | 51,411.57 |
| 162 | 4,602.57 | 4,627.34 | 4,571.61 | 4,298.57 | 18,100.09 |
| 163 | 9,393.69 | 8,945.64 | 8,739.96 | 7,926.06 | 35,005.35 |
| 164 | 3,369.85 | 3,168.48 | 3,155.19 | 3,007.67 | 12,701.19 |
| 165 | 7,798.81 | 7,267.94 | 6,916.06 | 6,370.79 | 28,353.60 |
| 166 | 12,943.89 | 12,685.67 | 12,552.90 | 12,139.13 | 50,321.59 |
| 167 | 1,867.69 | 1,975.18 | 2,015.75 | 1,914.04 | 7,772.66 |
| 168 | 5,931.65 | 5,847.72 | 5,659.81 | 5,224.50 | 22,663.68 |
| 169 | 2,712.05 | 2,770.66 | 2,527.25 | 2,407.65 | 10,417.61 |
| 170 | 37,249.32 | 36,320.78 | 34,846.56 | 33,716.81 | 142,133.47 |

DRUG CODE:9143DRUG NAME:OXYCODONE
STATE:PENNSYLVANIA

| ZIP CODE | QUARTER 1 | QUARTER 2 | QUARTER 3 | QUARTER 4 | TOTAL GRAMS |
|---|---|---|---|---|---|
| 171 | 11,715.78 | 11,187.45 | 10,539.59 | 9,672.91 | 43,115.73 |
| 172 | 11,484.96 | 11,090.78 | 10,683.17 | 10,182.73 | 43,441.64 |
| 173 | 17,053.06 | 16,213.79 | 15,823.53 | 15,174.73 | 64,265.11 |
| 174 | 9,450.56 | 9,400.98 | 8,916.69 | 8,651.39 | 36,419.62 |
| 175 | 14,349.78 | 13,852.37 | 13,635.93 | 12,593.38 | 54,431.46 |
| 176 | 12,102.87 | 11,783.09 | 10,978.81 | 10,592.91 | 45,457.68 |
| 177 | 9,509.63 | 9,377.62 | 9,025.84 | 8,657.81 | 36,570.90 |
| 178 | 13,611.81 | 12,594.95 | 12,826.93 | 12,651.57 | 51,685.26 |

| | | | | | |
|---|---|---|---|---|---|
| 179 | 10,354.09 | 10,334.21 | 9,716.48 | 8,416.08 | 38,820.86 |
| 180 | 21,968.18 | 20,545.35 | 20,411.46 | 19,402.61 | 82,327.60 |
| 181 | 13,222.33 | 12,869.19 | 12,507.33 | 11,623.77 | 50,222.62 |
| 182 | 8,525.66 | 8,516.73 | 8,556.39 | 8,658.90 | 34,257.68 |
| 183 | 14,217.64 | 13,674.34 | 12,904.01 | 12,034.72 | 52,830.71 |
| 184 | 13,864.60 | 13,356.47 | 13,208.30 | 13,108.85 | 53,538.22 |
| 185 | 18,576.18 | 18,262.08 | 17,599.84 | 17,518.09 | 71,956.19 |
| 186 | 11,226.67 | 11,286.39 | 10,922.52 | 10,505.41 | 43,940.99 |
| 187 | 40,566.86 | 45,843.40 | 42,860.68 | 36,957.09 | 166,228.03 |
| 188 | 4,137.75 | 4,153.85 | 4,090.49 | 3,844.48 | 16,226.57 |
| 189 | 20,731.67 | 19,957.83 | 19,306.90 | 18,636.30 | 78,632.70 |
| 190 | 122,042.78 | 122,145.16 | 116,176.26 | 112,574.99 | 472,939.19 |
| 191 | 161,776.90 | 159,240.01 | 152,825.84 | 144,293.22 | 618,135.97 |
| 193 | 22,505.70 | 22,341.85 | 21,679.53 | 20,745.08 | 87,272.16 |
| 194 | 37,043.43 | 35,465.27 | 34,098.93 | 31,913.24 | 138,520.87 |
| 195 | 7,804.57 | 7,658.93 | 7,327.95 | 7,160.88 | 29,952.33 |
| 196 | 11,184.93 | 10,501.19 | 9,916.65 | 9,820.14 | 41,422.91 |
| TOTAL | 903,045.64 | 882,716.44 | 852,812.33 | 817,502.98 | 3,456,077.39 |

STATE: PUERTO RICO

| ZIP CODE | QUARTER 1 | QUARTER 2 | QUARTER 3 | QUARTER 4 | TOTAL GRAMS |
|---|---|---|---|---|---|
| 6 | 4,422.84 | 4,540.95 | 4,605.68 | 4,774.94 | 18,344.41 |
| 7 | 6,063.93 | 6,117.45 | 6,591.74 | 6,297.02 | 25,070.14 |
| 9 | 7,099.74 | 7,518.54 | 7,380.17 | 7,646.74 | 29,645.19 |
| TOTAL | 17,586.51 | 18,176.94 | 18,577.59 | 18,718.70 | 73,059.74 |

STATE: RHODE ISLAND

| ZIP CODE | QUARTER 1 | QUARTER 2 | QUARTER 3 | QUARTER 4 | TOTAL GRAMS |
|---|---|---|---|---|---|
| 28 | 30,085.94 | 28,601.57 | 28,515.15 | 27,334.13 | 114,536.79 |
| 29 | 21,520.28 | 20,794.28 | 19,763.37 | 19,524.43 | 81,602.36 |
| TOTAL | 51,606.22 | 49,395.85 | 48,278.52 | 46,858.56 | 196,139.15 |

STATE: SOUTH CAROLINA

| ZIP CODE | QUARTER 1 | QUARTER 2 | QUARTER 3 | QUARTER 4 | TOTAL GRAMS |
|---|---|---|---|---|---|
| 290 | 13,366 | 13,087.11 | 13,383.35 | 13,640.79 | 53,477.25 |
| 291 | 16,985.07 | 16,487.42 | 16,757.75 | 16,592.69 | 66,822.93 |
| 292 | 11,584.03 | 10,970.83 | 11,236.83 | 11,893.47 | 45,685.16 |
| 293 | 28,095.36 | 26,985.94 | 27,512.07 | 27,001.94 | 109,595.31 |
| 294 | 45,800.05 | 44,135.21 | 44,170.53 | 43,370.03 | 177,475.82 |
| 295 | 63,869.20 | 62,314.12 | 62,394.64 | 61,961.20 | 250,539.16 |
| 296 | 52,622.45 | 51,380.16 | 50,879.81 | 48,738.48 | 203,620.90 |
| 297 | 12,333.46 | 12,092.97 | 11,861.75 | 11,712.82 | 48,001 |
| 298 | 11,567.69 | 11,159.71 | 11,212.56 | 11,247.95 | 45,187.91 |
| 299 | 7,850.88 | 7,786.13 | 7,688.61 | 7,523.22 | 30,848.84 |
| TOTAL | 264,074.19 | 256,399.60 | 257,097.90 | 253,682.59 | 1,031,254.28 |

STATE: SOUTH DAKOTA

| ZIP CODE | QUARTER 1 | QUARTER 2 | QUARTER 3 | QUARTER 4 | TOTAL GRAMS |
|---|---|---|---|---|---|
| 570 | 3,237.03 | 3,025.68 | 2,939.53 | 2,921.62 | 12,123.86 |
| 571 | 9,085.08 | 7,448.34 | 7,439.90 | 6,844.17 | 30,817.49 |
| 572 | 918.28 | 901.34 | 837.2 | 836.21 | 3,493.03 |
| 573 | 1,031.19 | 1,001.30 | 924 | 901.03 | 3,857.52 |
| 574 | 1,215.88 | 1,150.12 | 1,190.46 | 1,046.44 | 4,602.90 |
| 575 | 1,106.77 | 1,032.91 | 1,115.31 | 1,035.78 | 4,290.77 |
| 576 | 247.34 | 206.2 | 186.65 | 153.26 | 793.45 |
| 577 | 6,565.27 | 6,269.40 | 6,405.85 | 5,641.58 | 24,882.10 |
| TOTAL | 23,406.84 | 21,035.29 | 21,038.90 | 19,380.09 | 84,861.12 |

STATE: TENNESSEE

| ZIP CODE | QUARTER 1 | QUARTER 2 | QUARTER 3 | QUARTER 4 | TOTAL GRAMS |
|---|---|---|---|---|---|

| | | | | | |
|---|---|---|---|---|---|
| 370 | 70,618.11 | 69,265.41 | 68,181.18 | 66,754.95 | 274,819.65 |
| 371 | 54,336.85 | 53,531.39 | 52,524.03 | 51,600.20 | 211,992.47 |
| 372 | 37,774.54 | 37,371.75 | 36,497.18 | 35,604.37 | 147,247.84 |

DRUG CODE:9143DRUG NAME:OXYCODONE
STATE:TENNESSEE

| ZIP CODE | QUARTER 1 | QUARTER 2 | QUARTER 3 | QUARTER 4 | TOTAL GRAMS |
|---|---|---|---|---|---|
| 373 | 51,479.78 | 48,806.94 | 47,546.43 | 47,208.03 | 195,041.18 |
| 374 | 20,782.70 | 19,771.07 | 19,140.33 | 18,493.18 | 78,187.28 |
| 376 | 46,478.13 | 44,739.80 | 43,154.49 | 40,660.11 | 175,032.53 |
| 377 | 41,215.52 | 38,165.08 | 36,903.59 | 37,152.30 | 153,436.49 |
| 378 | 60,573.97 | 56,156.43 | 52,475.07 | 50,753.37 | 219,958.84 |
| 379 | 31,117.06 | 29,116.11 | 27,081.09 | 26,972.31 | 114,286.57 |
| 380 | 15,487.55 | 14,996.33 | 15,490.88 | 15,459.34 | 61,434.10 |
| 381 | 31,445.87 | 32,716.36 | 31,735.36 | 31,232.14 | 127,129.73 |
| 382 | 9,407.34 | 9,292.82 | 9,174.97 | 8,890.41 | 36,765.54 |
| 383 | 17,580.78 | 16,770.44 | 16,271.88 | 16,579.24 | 67,202.34 |
| 384 | 15,857.76 | 15,257.55 | 15,068.92 | 14,619.69 | 60,803.92 |
| 385 | 33,536.22 | 32,780.11 | 31,913.48 | 31,915.90 | 130,145.71 |
| TOTAL | 537,692.18 | 518,737.59 | 503,158.88 | 493,895.54 | 2,053,484.19 |

STATE: TEXAS

| ZIP CODE | QUARTER 1 | QUARTER 2 | QUARTER 3 | QUARTER 4 | TOTAL GRAMS |
|---|---|---|---|---|---|
| 750 | 37,204.35 | 38,532.40 | 38,605.22 | 39,542.45 | 153,884.42 |
| 751 | 8,955.05 | 8,564.77 | 8,834.37 | 8,342.08 | 34,696.27 |
| 752 | 13,686.34 | 12,579.18 | 12,463.25 | 12,596.21 | 51,324.98 |
| 753 | 160.47 | 120.36 | 102.96 | 145.86 | 529.65 |
| 754 | 5,603.09 | 6,053.99 | 5,964.90 | 6,005.70 | 23,627.68 |
| 755 | 2,007.76 | 1,969.40 | 1,935.63 | 1,701.43 | 7,614.22 |
| 756 | 2,990.94 | 2,982.49 | 2,710.15 | 2,831.45 | 11,515.03 |
| 757 | 4,709.02 | 4,700.19 | 4,776.52 | 4,746.29 | 18,932.02 |
| 758 | 955.43 | 843.65 | 826.08 | 854.21 | 3,479.37 |
| 759 | 2,427.99 | 2,207.99 | 2,253.40 | 2,007.32 | 8,896.70 |
| 760 | 27,606.47 | 27,968.51 | 26,971.46 | 27,223.58 | 109,770.02 |
| 761 | 20,322.04 | 20,633.67 | 20,937.72 | 20,639.85 | 82,533.28 |
| 762 | 7,992.31 | 8,233.02 | 8,259.16 | 8,468.71 | 32,953.20 |
| 763 | 3,572.19 | 3,458.47 | 3,398.97 | 3,322.29 | 13,751.92 |
| 764 | 1,455.60 | 1,626.73 | 1,674.08 | 1,533.51 | 6,289.92 |
| 765 | 6,863.65 | 7,246.68 | 6,650.33 | 6,505.03 | 27,265.69 |
| 766 | 1,485.19 | 1,392.87 | 1,555.44 | 1,574.93 | 6,008.43 |
| 767 | 2,029.14 | 2,075.15 | 2,048.81 | 1,966.25 | 8,119.35 |
| 768 | 1,214.31 | 1,170.43 | 1,014.79 | 1,200.28 | 4,599.81 |
| 769 | 1,487.16 | 1,375.01 | 1,371.88 | 1,327.32 | 5,561.37 |
| 770 | 33,332.29 | 33,794.28 | 36,827.86 | 35,982.34 | 139,936.77 |
| 773 | 13,310.81 | 13,248.54 | 13,418.35 | 13,441.88 | 53,419.58 |
| 774 | 13,384.16 | 12,667.04 | 13,337.24 | 13,087.68 | 52,476.12 |
| 775 | 13,128.33 | 12,335.56 | 12,198.10 | 12,337.44 | 49,999.43 |
| 776 | 2,836.09 | 2,828.22 | 2,705.67 | 2,603.51 | 10,973.49 |
| 777 | 1,008.02 | 1,049.62 | 1,094.85 | 1,087.90 | 4,240.39 |
| 778 | 1,648.44 | 1,561.66 | 1,537.46 | 1,590.11 | 6,337.67 |
| 779 | 3,107.79 | 3,188 | 3,067.35 | 3,285.12 | 12,648.26 |
| 780 | 3,579.72 | 3,325.39 | 3,319.44 | 3,474.88 | 13,699.43 |
| 781 | 3,953.10 | 3,898.80 | 3,757.76 | 3,783.39 | 15,393.05 |
| 782 | 25,705.15 | 24,388.75 | 23,958.99 | 23,226.82 | 97,279.71 |
| 783 | 1,305.66 | 1,251.28 | 1,311.67 | 1,260.59 | 5,129.20 |
| 784 | 2,117.15 | 2,305.49 | 2,114.12 | 2,219.45 | 8,756.21 |
| 785 | 3,726.25 | 3,503.16 | 3,601.72 | 3,803.37 | 14,634.50 |
| 786 | 19,675.25 | 19,929.31 | 19,889.75 | 20,257.63 | 79,751.94 |
| 787 | 19,094.58 | 19,540.09 | 18,408.04 | 19,779.50 | 76,822.21 |
| 788 | 626.56 | 624.29 | 612.23 | 588.42 | 2,451.50 |

| | | | | | |
|---|---|---|---|---|---|
| 789 | 794.12 | 667.25 | 647.91 | 619.03 | 2,728.31 |
| 790 | 1,515.63 | 1,557.84 | 1,548.33 | 1,569.79 | 6,191.59 |
| 791 | 2,522.66 | 2,962.13 | 2,959.93 | 2,989.29 | 11,434.01 |
| 792 | 335.25 | 332.74 | 310.05 | 348.2 | 1,326.24 |
| 793 | 590.03 | 578.51 | 625.71 | 693.89 | 2,488.14 |
| 794 | 2,393.74 | 2,393.98 | 2,555.84 | 2,611.87 | 9,955.43 |
| 795 | 793.22 | 716.8 | 732.49 | 757.09 | 2,999.60 |
| 796 | 2,237.21 | 2,374.07 | 2,238.11 | 2,096.62 | 8,946.01 |
| 797 | 4,379.15 | 4,324.61 | 4,305.59 | 4,050.75 | 17,060.10 |
| 798 | 226.14 | 143.89 | 125.85 | 120 | 615.88 |
| 799 | 5,560.39 | 5,247.43 | 5,536.21 | 4,678.52 | 21,022.55 |
| TOTAL | 335,615.39 | 334,473.69 | 335,101.74 | 334,879.83 | 1,340,070.65 |

STATE: UTAH

| ZIP CODE | QUARTER 1 | QUARTER 2 | QUARTER 3 | QUARTER 4 | TOTAL GRAMS |
|---|---|---|---|---|---|

DRUG CODE:9143DRUG NAME:OXYCODONE
STATE:UTAH

| ZIP CODE | QUARTER 1 | QUARTER 2 | QUARTER 3 | QUARTER 4 | TOTAL GRAMS |
|---|---|---|---|---|---|
| 840 | 74,755.81 | 71,750.63 | 73,720.27 | 70,797.01 | 291,023.72 |
| 841 | 55,017.84 | 52,410.10 | 51,543.76 | 49,560.98 | 208,532.68 |
| 843 | 6,528.32 | 6,083.49 | 5,887.92 | 5,860.53 | 24,360.26 |
| 844 | 15,478.06 | 14,905.97 | 15,133.40 | 14,259.30 | 59,776.73 |
| 845 | 6,521.72 | 6,309.75 | 6,473.90 | 6,145.96 | 25,451.33 |
| 846 | 15,813.67 | 15,314.48 | 14,804.50 | 14,259.92 | 60,192.57 |
| 847 | 20,350.70 | 19,743.51 | 19,792.93 | 19,348.78 | 79,235.92 |
| TOTAL | 194,466.12 | 186,517.93 | 187,356.68 | 180,232.48 | 748,573.21 |

STATE: VERMONT

| ZIP CODE | QUARTER 1 | QUARTER 2 | QUARTER 3 | QUARTER 4 | TOTAL GRAMS |
|---|---|---|---|---|---|
| 50 | 3,085.11 | 3,083.38 | 2,964.26 | 2,510.33 | 11,643.08 |
| 51 | 1,963.78 | 1,842.06 | 1,694.74 | 1,753.15 | 7,253.73 |
| 52 | 1,563.92 | 1,486.77 | 1,441.04 | 1,364.43 | 5,856.16 |
| 53 | 2,115.88 | 2,154.78 | 2,078.09 | 1,954.08 | 8,302.83 |
| 54 | 10,637.76 | 10,282.77 | 10,569.10 | 10,244.93 | 41,734.56 |
| 56 | 2,933.15 | 2,642.89 | 2,593.41 | 2,391.95 | 10,561.40 |
| 57 | 2,691.40 | 2,721.71 | 2,619.38 | 2,426.91 | 10,459.40 |
| 58 | 2,107.08 | 2,028.24 | 1,918.63 | 2,056.78 | 8,110.73 |
| TOTAL | 27,098.08 | 26,242.60 | 25,878.65 | 24,702.56 | 103,921.89 |

STATE: VIRGIN ISLANDS

| ZIP CODE | QUARTER 1 | QUARTER 2 | QUARTER 3 | QUARTER 4 | TOTAL GRAMS |
|---|---|---|---|---|---|
| 8 | 742.3 | 677.08 | 587.66 | 607.78 | 2,614.82 |
| TOTAL | 742.3 | 677.08 | 587.66 | 607.78 | 2,614.82 |

STATE: VIRGINIA

| ZIP CODE | QUARTER 1 | QUARTER 2 | QUARTER 3 | QUARTER 4 | TOTAL GRAMS |
|---|---|---|---|---|---|
| 201 | 24,673.27 | 24,280.21 | 22,895.83 | 22,732.31 | 94,581.62 |
| 220 | 14,720.59 | 14,269.44 | 14,553.57 | 13,608.50 | 57,152.10 |
| 221 | 12,925.85 | 13,040.78 | 12,668.40 | 12,276.15 | 50,911.18 |
| 222 | 3,466.38 | 3,329.72 | 3,126.31 | 3,257.43 | 13,179.84 |
| 223 | 8,109.67 | 7,548.46 | 7,332.27 | 7,329.96 | 30,320.36 |
| 224 | 12,120.23 | 11,554.30 | 11,539.46 | 10,938.93 | 46,152.92 |
| 225 | 4,131.33 | 4,094.85 | 3,964.89 | 4,006.09 | 16,197.16 |
| 226 | 11,194.24 | 10,858.90 | 11,214.17 | 10,805.22 | 44,072.53 |
| 227 | 2,588.09 | 2,454.29 | 2,409 | 2,374.08 | 9,825.46 |
| 228 | 6,606.11 | 6,477.10 | 6,359.51 | 6,542.04 | 25,984.76 |
| 229 | 10,606.10 | 10,468.27 | 10,389.78 | 9,985.25 | 41,449.40 |
| 230 | 9,293.91 | 8,582.69 | 8,478.69 | 8,102.85 | 34,458.14 |
| 231 | 15,752.09 | 15,162.79 | 15,217.12 | 14,248.95 | 60,380.95 |

| | | | | | |
|---|---|---|---|---|---|
| 232 | 32,759.41 | 31,611.50 | 30,042.76 | 29,498.93 | 123,912.60 |
| 233 | 10,663.60 | 10,371.72 | 10,410.79 | 9,865.31 | 41,311.42 |
| 234 | 23,240.35 | 22,571.95 | 22,452.21 | 22,528.92 | 90,793.43 |
| 235 | 9,854.71 | 9,425.05 | 9,125.87 | 9,215.52 | 37,621.15 |
| 236 | 15,447.75 | 14,365.63 | 14,313.61 | 13,654.80 | 57,781.79 |
| 237 | 7,358.63 | 7,130.82 | 6,683.32 | 6,876.79 | 28,049.56 |
| 238 | 19,159 | 18,010.77 | 17,682.69 | 17,258.05 | 72,110.51 |
| 239 | 3,403.52 | 3,283.88 | 3,319.49 | 3,032 | 13,038.89 |
| 240 | 19,418.64 | 19,504.86 | 19,523.93 | 19,464.52 | 77,911.95 |
| 241 | 19,319.38 | 18,707.94 | 18,453.81 | 17,891.81 | 74,372.94 |
| 242 | 23,992.69 | 23,768.49 | 23,997.40 | 23,425.32 | 95,183.90 |
| 243 | 9,790.57 | 10,087.13 | 10,187.14 | 10,337.38 | 40,402.22 |
| 244 | 5,774.93 | 5,543.81 | 5,367.91 | 5,015.59 | 21,702.24 |
| 245 | 16,804.78 | 15,949.75 | 15,274.89 | 15,847.97 | 63,877.39 |
| 246 | 8,591.67 | 8,515.81 | 7,692.33 | 6,863.56 | 31,663.37 |
| TOTAL | 361,767.49 | 350,970.91 | 344,677.15 | 336,984.23 | 1,394,399.78 |

STATE: WASHINGTON

| ZIP CODE | QUARTER 1 | QUARTER 2 | QUARTER 3 | QUARTER 4 | TOTAL GRAMS |
|---|---|---|---|---|---|
| 980 | 59,203.59 | 56,427.63 | 56,656.73 | 54,611.13 | 226,899.08 |
| 981 | 39,035.09 | 37,582.77 | 36,856.02 | 35,032.60 | 148,506.48 |
| 982 | 46,310.24 | 44,617.20 | 43,455.59 | 41,356.32 | 175,739.35 |
| 983 | 37,796.20 | 37,449.17 | 36,617.70 | 35,779.46 | 147,642.53 |
| 984 | 26,972.90 | 26,295.23 | 25,688.03 | 25,782.27 | 104,738.43 |
| 985 | 24,779.42 | 23,335.96 | 23,351.96 | 22,222.06 | 93,689.40 |
| 986 | 32,070.20 | 30,260.46 | 30,874.94 | 31,219.08 | 124,424.68 |
| 988 | 10,503.55 | 10,535.38 | 10,422.58 | 10,230.30 | 41,691.81 |
| 989 | 13,056.68 | 13,279.18 | 13,072.38 | 12,529.30 | 51,937.54 |
| 990 | 3,334.97 | 3,153.44 | 3,373.92 | 3,182.61 | 13,044.94 |

DRUG CODE:9143DRUG NAME:OXYCODONE
STATE:WASHINGTON

| ZIP CODE | QUARTER 1 | QUARTER 2 | QUARTER 3 | QUARTER 4 | TOTAL GRAMS |
|---|---|---|---|---|---|
| 991 | 5,350.29 | 5,244.29 | 5,496.94 | 5,145.95 | 21,237.47 |
| 992 | 19,968.45 | 19,771.17 | 19,477.84 | 19,083.50 | 78,300.96 |
| 993 | 19,751.87 | 19,055.81 | 18,700.38 | 18,058.40 | 75,566.46 |
| 994 | 1,342.19 | 1,326.60 | 1,175.71 | 1,279.98 | 5,124.48 |
| TOTAL | 339,475.64 | 328,334.29 | 325,220.72 | 315,512.96 | 1,308,543.61 |

STATE: WEST VIRGINIA

| ZIP CODE | QUARTER 1 | QUARTER 2 | QUARTER 3 | QUARTER 4 | TOTAL GRAMS |
|---|---|---|---|---|---|
| 247 | 3,550.26 | 3,347.35 | 3,159.17 | 3,057.17 | 13,113.95 |
| 248 | 3,240.76 | 2,796.77 | 2,468.60 | 2,419.32 | 10,925.45 |
| 249 | 3,362.67 | 3,210.40 | 2,881.74 | 2,569.15 | 12,023.96 |
| 250 | 4,002.43 | 3,957.35 | 3,620.45 | 3,092.76 | 14,672.99 |
| 251 | 3,020.62 | 2,936.68 | 2,708.07 | 2,479.26 | 11,144.63 |
| 252 | 2,723.51 | 2,332.56 | 2,160.09 | 2,039.97 | 9,256.13 |
| 253 | 8,950.24 | 8,414.46 | 8,475.98 | 7,877.34 | 33,718.02 |
| 254 | 16,275.94 | 15,677.87 | 15,278.73 | 15,504.54 | 62,737.08 |
| 255 | 9,049.36 | 8,372.81 | 7,913.58 | 6,973.56 | 32,309.31 |
| 256 | 3,104.45 | 2,790.80 | 2,772.96 | 2,376.05 | 11,044.26 |
| 257 | 6,599.25 | 6,698.38 | 6,040.37 | 5,463.52 | 24,801.52 |
| 258 | 7,684.84 | 6,106.55 | 6,013.45 | 5,041.56 | 24,846.40 |
| 259 | 4,002.29 | 2,965.17 | 3,293 | 2,729.90 | 12,990.36 |
| 260 | 12,799.27 | 11,623.36 | 10,993.40 | 10,715.06 | 46,131.09 |
| 261 | 4,363.25 | 3,976.92 | 3,738.99 | 3,744.56 | 15,823.72 |
| 262 | 2,887.63 | 2,695.42 | 2,540.05 | 2,473.95 | 10,597.05 |
| 263 | 5,940.70 | 5,464.37 | 5,145.60 | 5,143.53 | 21,694.20 |
| 264 | 1,644.29 | 1,681.16 | 1,658.44 | 1,512.35 | 6,496.24 |
| 265 | 8,228.61 | 8,069.61 | 8,076.14 | 7,601.91 | 31,976.27 |

| | | | | | |
|---|---|---|---|---|---|
| 266 | 1,965.67 | 1,805.86 | 1,868.49 | 1,629.38 | 7,269.40 |
| 267 | 2,686.45 | 2,624.46 | 2,755.45 | 2,699.97 | 10,766.33 |
| 268 | 1,006.59 | 1,002.78 | 855.89 | 843.61 | 3,708.87 |
| TOTAL | 117,089.08 | 108,551.09 | 104,418.64 | 97,988.42 | 428,047.23 |

STATE: WISCONSIN

| ZIP CODE | QUARTER 1 | QUARTER 2 | QUARTER 3 | QUARTER 4 | TOTAL GRAMS |
|---|---|---|---|---|---|
| 530 | 31,148.14 | 29,465.89 | 28,872.89 | 27,477.03 | 116,963.95 |
| 531 | 37,721.49 | 35,876.74 | 33,774.04 | 32,871.12 | 140,243.39 |
| 532 | 92,942.91 | 86,486.97 | 86,712.68 | 82,171.57 | 348,314.13 |
| 534 | 8,312.81 | 7,568.16 | 7,049.90 | 6,727.69 | 29,658.56 |
| 535 | 21,258.95 | 20,468.70 | 19,897.09 | 18,655.55 | 80,280.29 |
| 537 | 17,380.06 | 16,575.94 | 16,272.51 | 15,200.48 | 65,428.99 |
| 538 | 1,331.71 | 1,322.54 | 1,194.56 | 1,106.76 | 4,955.57 |
| 539 | 8,996.98 | 8,446.55 | 8,130.56 | 7,408.58 | 32,982.67 |
| 540 | 3,920.99 | 3,586.47 | 3,671 | 3,599.51 | 14,777.97 |
| 541 | 4,172.63 | 3,994.15 | 3,823.06 | 3,766.24 | 15,756.08 |
| 542 | 3,859.66 | 3,181.24 | 2,842.51 | 2,617.20 | 12,500.61 |
| 543 | 5,017.63 | 4,590.34 | 4,474.98 | 4,513.18 | 18,596.13 |
| 544 | 9,949.79 | 9,356.58 | 8,875.24 | 8,671.23 | 36,852.84 |
| 545 | 3,437.94 | 3,491.16 | 3,366.81 | 3,186.80 | 13,482.71 |
| 546 | 8,685.31 | 8,103.71 | 8,034.39 | 7,901.18 | 32,724.59 |
| 547 | 6,120.89 | 5,840.91 | 5,565.35 | 5,679.37 | 23,206.52 |
| 548 | 5,730.47 | 5,484.19 | 5,524.53 | 5,117.17 | 21,856.36 |
| 549 | 15,775.54 | 14,379.16 | 14,365.89 | 13,890.58 | 58,411.17 |
| TOTAL | 285,763.90 | 268,219.40 | 262,447.99 | 250,561.24 | 1,066,992.53 |

STATE: WYOMING

| ZIP CODE | QUARTER 1 | QUARTER 2 | QUARTER 3 | QUARTER 4 | TOTAL GRAMS |
|---|---|---|---|---|---|
| 820 | 6,717.19 | 6,398.37 | 6,058.50 | 6,031.16 | 25,205.22 |
| 822 | 655.14 | 697.01 | 685.29 | 663.64 | 2,701.08 |
| 823 | 534.89 | 693.63 | 673.48 | 640.92 | 2,542.92 |
| 824 | 4,470.37 | 4,237.34 | 4,001.88 | 4,076.48 | 16,786.07 |
| 825 | 2,365.36 | 2,189.26 | 2,514.50 | 2,224.08 | 9,293.20 |
| 826 | 5,869.26 | 5,275.05 | 5,505.83 | 5,151.34 | 21,801.48 |
| 827 | 1,617.15 | 1,398.72 | 1,328.86 | 1,345.02 | 5,689.75 |
| 828 | 1,875.48 | 1,832.67 | 1,753.78 | 1,590.53 | 7,052.46 |
| 829 | 5,051.79 | 5,029.32 | 5,217.12 | 5,184.01 | 20,482.24 |
| 830 | 367.47 | 308.31 | 294.59 | 274.33 | 1,244.70 |
| 831 | 1,046.80 | 1,045.95 | 984.44 | 975.39 | 4,052.58 |
| TOTAL | 30,570.90 | 29,105.63 | 29,018.27 | 28,156.90 | 116,851.70 |

DRUG CODE:9150DRUG NAME:HYDROMORPHONE
STATE: ALABAMA

| ZIP CODE | QUARTER 1 | QUARTER 2 | QUARTER 3 | QUARTER 4 | TOTAL GRAMS |
|---|---|---|---|---|---|
| 350 | 252.09 | 297.91 | 285.23 | 269.45 | 1,104.68 |
| 351 | 132.29 | 109.83 | 112.91 | 101.63 | 456.66 |
| 352 | 605.12 | 538.66 | 545.44 | 491.3 | 2,180.52 |
| 354 | 147.84 | 154.52 | 136.68 | 150.66 | 589.7 |
| 355 | 46.37 | 48.71 | 44.35 | 39.68 | 179.11 |
| 356 | 247.18 | 277.38 | 275.44 | 223.08 | 1,023.08 |
| 357 | 115.11 | 130.92 | 120.18 | 116.57 | 482.78 |
| 358 | 313.73 | 316.27 | 288.72 | 286.1 | 1,204.82 |
| 359 | 299.22 | 281.66 | 206.61 | 141.81 | 929.3 |
| 360 | 237.92 | 240.02 | 242.36 | 203.28 | 923.58 |
| 361 | 266.45 | 204.05 | 243.06 | 221.22 | 934.78 |
| 362 | 113.04 | 131.68 | 107.29 | 149.93 | 501.94 |
| 363 | 218.28 | 274.09 | 267.53 | 285.24 | 1,045.14 |
| 364 | 101.57 | 92.21 | 91.95 | 84.58 | 370.31 |

| | | | | | | |
|---|---|---|---|---|---|---|
| | 365 | 396.93 | 402.14 | 402.68 | 397.27 | 1,599.02 |
| | 366 | 305.88 | 350.75 | 309.73 | 293.43 | 1,259.79 |
| | 367 | 28.17 | 30.71 | 28.14 | 31.1 | 118.12 |
| | 368 | 158.75 | 153.42 | 115.31 | 119.67 | 547.15 |
| | 369 | 14.36 | 11.92 | 15.38 | 15.56 | 57.22 |
| TOTAL | | 4,000.30 | 4,046.85 | 3,838.99 | 3,621.56 | 15,507.70 |
| STATE: | ALASKA | | | | | |
| ZIP CODE | QUARTER 1 | | QUARTER 2 | QUARTER 3 | QUARTER 4 | TOTAL GRAMS |
| | 995 | 673.96 | 600.43 | 594.59 | 638.78 | 2,507.76 |
| | 996 | 348.69 | 354.51 | 353.01 | 344.41 | 1,400.62 |
| | 997 | 45.03 | 49.19 | 40.71 | 41.14 | 176.07 |
| | 998 | 44.13 | 43.36 | 35.93 | 34.75 | 158.17 |
| | 999 | 8.7 | 15.31 | 13.94 | 26.2 | 64.15 |
| TOTAL | | 1,120.51 | 1,062.80 | 1,038.18 | 1,085.28 | 4,306.77 |
| STATE: | AMERICAN SAMOA | | | | | |
| ZIP CODE | QUARTER 1 | | QUARTER 2 | QUARTER 3 | QUARTER 4 | TOTAL GRAMS |
| | 967 | 0.61 | 1.95 | 0.06 | 0.53 | 3.15 |
| TOTAL | | 0.61 | 1.95 | 0.06 | 0.53 | 3.15 |
| STATE: | ARIZONA | | | | | |
| ZIP CODE | QUARTER 1 | | QUARTER 2 | QUARTER 3 | QUARTER 4 | TOTAL GRAMS |
| | 850 | 1,428.66 | 1,390.71 | 1,399.90 | 1,416.57 | 5,635.84 |
| | 851 | 214.71 | 204.1 | 193.83 | 205.91 | 818.55 |
| | 852 | 2,227.54 | 2,090.31 | 1,971.79 | 2,083.28 | 8,372.92 |
| | 853 | 1,294.54 | 1,339.34 | 1,304.22 | 1,223.79 | 5,161.89 |
| | 855 | 118.05 | 113.95 | 106.75 | 115.46 | 454.21 |
| | 856 | 273.67 | 256.74 | 230.91 | 216.26 | 977.58 |
| | 857 | 1,262.10 | 1,194.66 | 1,219.61 | 1,168.74 | 4,845.11 |
| | 859 | 158.83 | 121.73 | 135.93 | 132.8 | 549.29 |
| | 860 | 169.75 | 184.39 | 177.08 | 141.22 | 672.44 |
| | 863 | 255.41 | 300.62 | 247.82 | 269.01 | 1,072.86 |
| | 864 | 881.56 | 767.56 | 782.6 | 705.09 | 3,136.81 |
| | 865 | 4.48 | 0.49 | 3.37 | 3.06 | 11.4 |
| TOTAL | | 8,289.30 | 7,964.60 | 7,773.81 | 7,681.19 | 31,708.90 |
| STATE: | ARKANSAS | | | | | |
| ZIP CODE | QUARTER 1 | | QUARTER 2 | QUARTER 3 | QUARTER 4 | TOTAL GRAMS |
| | 716 | 85.92 | 102.67 | 97.46 | 81.44 | 367.49 |
| | 717 | 66.42 | 60.94 | 43.73 | 51.15 | 222.24 |
| | 718 | 138.88 | 137.52 | 115.37 | 86.49 | 478.26 |
| | 719 | 229.21 | 194.01 | 183.62 | 163.02 | 769.86 |
| | 720 | 403.98 | 455.31 | 301.09 | 288.04 | 1,448.42 |
| | 721 | 396.24 | 392.36 | 337.12 | 272.6 | 1,398.32 |
| | 722 | 645.28 | 583.48 | 648.96 | 607.55 | 2,485.27 |
| | 723 | 55.44 | 54.32 | 57.11 | 72.16 | 239.03 |
| | 724 | 276.79 | 304.88 | 264.45 | 211.51 | 1,057.63 |
| | 725 | 160.37 | 171.62 | 152.64 | 143.78 | 628.41 |
| | 726 | 147.74 | 179.32 | 171.42 | 179.99 | 678.47 |
| | 727 | 855.76 | 884.97 | 914.85 | 870.18 | 3,525.76 |
| | 728 | 115.26 | 104.32 | 105.52 | 90.2 | 415.3 |
| | 729 | 425.4 | 401.97 | 429.51 | 386.28 | 1,643.16 |
| TOTAL | | 4,002.69 | 4,027.69 | 3,822.85 | 3,504.39 | 15,357.62 |
| STATE: | CALIFORNIA | | | | | |
| ZIP CODE | QUARTER 1 | | QUARTER 2 | QUARTER 3 | QUARTER 4 | TOTAL GRAMS |

DRUG CODE:9150DRUG NAME:HYDROMORPHONE
STATE:CALIFORNIA

RETAIL DRUG DISTRIBUTION BY ZIP CODE WITHIN STATE BY GRAMS WT

| ZIP CODE | QUARTER 1 | QUARTER 2 | QUARTER 3 | QUARTER 4 | TOTAL GRAMS |
|---|---|---|---|---|---|
| 900 | 1,754.20 | 1,719.13 | 1,533.98 | 1,666.14 | 6,673.45 |
| 902 | 798.05 | 716.18 | 692.83 | 605.6 | 2,812.66 |
| 903 | 52.16 | 47.45 | 56.52 | 40.73 | 196.86 |
| 904 | 268.44 | 275.26 | 233.47 | 249.18 | 1,026.35 |
| 905 | 512.93 | 434.27 | 427.04 | 441.1 | 1,815.34 |
| 906 | 433.95 | 452.2 | 416.65 | 435.86 | 1,738.66 |
| 907 | 462.98 | 490.55 | 428.94 | 449.62 | 1,832.09 |
| 908 | 2,139.08 | 1,228.89 | 812.52 | 1,085.62 | 5,266.11 |
| 910 | 433.5 | 471.62 | 434.63 | 396.96 | 1,736.71 |
| 911 | 174.7 | 189.47 | 183.24 | 202.1 | 749.51 |
| 912 | 170.18 | 169.92 | 174.51 | 208.62 | 723.23 |
| 913 | 1,544.42 | 1,524.49 | 1,460.28 | 1,443.37 | 5,972.56 |
| 914 | 445.05 | 408.81 | 339.5 | 394.07 | 1,587.43 |
| 915 | 346.51 | 237.27 | 202.18 | 283.96 | 1,069.92 |
| 916 | 156.64 | 130.94 | 116.82 | 84.12 | 488.52 |
| 917 | 2,134.51 | 2,094.20 | 1,868.59 | 1,842.90 | 7,940.20 |
| 918 | 43.71 | 40 | 17.62 | 17.58 | 118.91 |
| 919 | 498.75 | 470.36 | 560.03 | 445.14 | 1,974.28 |
| 920 | 2,335.66 | 2,244.28 | 2,169.45 | 2,197.81 | 8,947.20 |
| 921 | 1,698.08 | 1,632.91 | 1,515.81 | 1,572.26 | 6,419.06 |
| 922 | 1,035.05 | 1,035.69 | 996.71 | 1,030.02 | 4,097.47 |
| 923 | 1,760.42 | 1,730.72 | 1,587.80 | 1,601.07 | 6,680.01 |
| 924 | 209.54 | 209.07 | 197.62 | 211.71 | 827.94 |
| 925 | 1,289.51 | 1,306.41 | 1,199.76 | 1,317.60 | 5,113.28 |
| 926 | 1,072.97 | 1,001.49 | 1,262.68 | 896.98 | 4,234.12 |
| 927 | 320.38 | 312.97 | 315.83 | 292.25 | 1,241.43 |
| 928 | 1,677.31 | 1,633.14 | 1,544.41 | 1,629.24 | 6,484.10 |
| 930 | 815.12 | 763.79 | 737.78 | 723.38 | 3,040.07 |
| 931 | 221.95 | 181.26 | 178.73 | 179.35 | 761.29 |
| 932 | 685.97 | 664.44 | 693.07 | 738.59 | 2,782.07 |
| 933 | 979.76 | 1,016.72 | 913.73 | 919.11 | 3,829.32 |
| 934 | 1,083.50 | 1,048.88 | 1,010.85 | 1,012.45 | 4,155.68 |
| 935 | 1,252.05 | 1,321.51 | 1,220.75 | 1,323.03 | 5,117.34 |
| 936 | 314.05 | 302.5 | 315.18 | 309.08 | 1,240.81 |
| 937 | 433.35 | 434.53 | 400.39 | 387.51 | 1,655.78 |
| 939 | 411.33 | 407.01 | 360.82 | 295.93 | 1,475.09 |
| 940 | 724.11 | 656.42 | 668.95 | 618.88 | 2,668.36 |
| 941 | 1,167.35 | 1,119.31 | 1,125.72 | 1,005.75 | 4,418.13 |
| 943 | 301.4 | 347.25 | 277.98 | 354.55 | 1,281.18 |
| 944 | 122.92 | 105.66 | 109.39 | 95.21 | 433.18 |
| 945 | 3,469.25 | 3,375.10 | 3,263.84 | 3,175.94 | 13,284.13 |
| 946 | 537.14 | 545.82 | 465.81 | 491.54 | 2,040.31 |
| 947 | 156.23 | 123.37 | 223.42 | 203.8 | 706.82 |
| 948 | 144.97 | 109.3 | 112.82 | 105.7 | 472.79 |
| 949 | 582.85 | 649.25 | 555.88 | 560.54 | 2,348.52 |
| 950 | 631.96 | 595.38 | 568.27 | 545.18 | 2,340.79 |
| 951 | 484.58 | 467.46 | 433.4 | 394.96 | 1,780.40 |
| 952 | 646.19 | 599.57 | 542.45 | 555.07 | 2,343.28 |
| 953 | 1,237 | 1,138.46 | 1,132.64 | 1,145.07 | 4,653.17 |
| 954 | 1,127.19 | 1,097.72 | 1,055.04 | 1,041.54 | 4,321.49 |
| 955 | 265.14 | 315.89 | 270.27 | 273.72 | 1,125.02 |
| 956 | 2,036.56 | 2,025.78 | 1,886.99 | 1,891.85 | 7,841.18 |
| 957 | 232.31 | 225.58 | 231.69 | 205.08 | 894.66 |
| 958 | 1,236.48 | 1,192.97 | 1,181.36 | 1,232.74 | 4,843.55 |
| 959 | 1,346.12 | 1,279.47 | 1,246.96 | 1,193.01 | 5,065.56 |
| 960 | 604.76 | 607.26 | 601.67 | 571.7 | 2,385.39 |
| 961 | 110.4 | 110.75 | 100.8 | 104.12 | 426.07 |
| TOTAL | 47,130.67 | 45,036.10 | 42,636.07 | 42,695.99 | 177,498.83 |

STATE: COLORADO

RETAIL DRUG DISTRIBUTION BY ZIP CODE WITHIN STATE BY GRAMS WT

| ZIP CODE | QUARTER 1 | QUARTER 2 | QUARTER 3 | QUARTER 4 | TOTAL GRAMS |
|---|---|---|---|---|---|
| 800 | 1,445.18 | 1,480 | 1,412.44 | 1,394.19 | 5,731.81 |
| 801 | 1,415.77 | 1,488.15 | 1,670.40 | 1,337.55 | 5,911.87 |
| 802 | 1,388.61 | 1,279.09 | 1,280.06 | 1,236.08 | 5,183.84 |
| 803 | 144.24 | 151.63 | 153.62 | 159.33 | 608.82 |
| 804 | 285.54 | 324.38 | 283.17 | 262.93 | 1,156.02 |
| 805 | 865.59 | 763.39 | 717.85 | 702.06 | 3,048.89 |

DRUG CODE:9150DRUG NAME:HYDROMORPHONE
STATE:COLORADO

| ZIP CODE | QUARTER 1 | QUARTER 2 | QUARTER 3 | QUARTER 4 | TOTAL GRAMS |
|---|---|---|---|---|---|
| 806 | 387.57 | 363.41 | 339.09 | 349.34 | 1,439.41 |
| 807 | 71.32 | 64.69 | 70.25 | 59.65 | 265.91 |
| 808 | 151.72 | 124.98 | 135.58 | 159.89 | 572.17 |
| 809 | 924.26 | 881.44 | 830.02 | 838.04 | 3,473.76 |
| 810 | 431.37 | 410.68 | 415.3 | 416.22 | 1,673.57 |
| 811 | 65.12 | 63.86 | 54.79 | 54.41 | 238.18 |
| 812 | 116.07 | 104.31 | 130.91 | 112.22 | 463.51 |
| 813 | 89.65 | 74.88 | 58.93 | 65.86 | 289.32 |
| 814 | 80.9 | 75.3 | 66.18 | 70.14 | 292.52 |
| 815 | 200.5 | 176.58 | 136.84 | 173.42 | 687.34 |
| 816 | 110.97 | 98.34 | 99.3 | 110.08 | 418.69 |
| TOTAL | 8,174.38 | 7,925.11 | 7,854.73 | 7,501.41 | 31,455.63 |

STATE: CONNECTICUT

| ZIP CODE | QUARTER 1 | QUARTER 2 | QUARTER 3 | QUARTER 4 | TOTAL GRAMS |
|---|---|---|---|---|---|
| 60 | 1,462.35 | 1,381.74 | 1,342.55 | 1,418.01 | 5,604.65 |
| 61 | 678.78 | 631.69 | 574.41 | 579.69 | 2,464.57 |
| 62 | 207.33 | 193.41 | 171.99 | 195.32 | 768.05 |
| 63 | 432.69 | 426.45 | 367.45 | 373.48 | 1,600.07 |
| 64 | 1,827.89 | 1,786.65 | 1,616.87 | 1,886.70 | 7,118.11 |
| 65 | 522.57 | 551.34 | 523.13 | 524.71 | 2,121.75 |
| 66 | 196.96 | 168.18 | 166.93 | 155.19 | 687.26 |
| 67 | 514.23 | 508.41 | 490.08 | 475.09 | 1,987.81 |
| 68 | 1,520.85 | 1,494.49 | 1,474.58 | 1,457.88 | 5,947.80 |
| 69 | 112.94 | 97.82 | 95.39 | 92.66 | 398.81 |
| TOTAL | 7,476.59 | 7,240.18 | 6,823.38 | 7,158.73 | 28,698.88 |

STATE: DELAWARE

| ZIP CODE | QUARTER 1 | QUARTER 2 | QUARTER 3 | QUARTER 4 | TOTAL GRAMS |
|---|---|---|---|---|---|
| 197 | 1,060.81 | 1,037.82 | 1,037.08 | 967.67 | 4,103.38 |
| 198 | 421.05 | 458.58 | 439.86 | 398.54 | 1,718.03 |
| 199 | 976.43 | 966.47 | 985.51 | 944.79 | 3,873.20 |
| TOTAL | 2,458.29 | 2,462.87 | 2,462.45 | 2,311 | 9,694.61 |

STATE: DISTRICT OF COLUMBIA

| ZIP CODE | QUARTER 1 | QUARTER 2 | QUARTER 3 | QUARTER 4 | TOTAL GRAMS |
|---|---|---|---|---|---|
| 200 | 623.82 | 579.33 | 599.7 | 613.57 | 2,416.42 |
| 203 | 2.84 | 0 | 0 | 0 | 2.84 |
| 204 | 19.42 | 14.23 | 21.59 | 9.49 | 64.73 |
| TOTAL | 646.08 | 593.56 | 621.29 | 623.06 | 2,483.99 |

STATE: FLORIDA

| ZIP CODE | QUARTER 1 | QUARTER 2 | QUARTER 3 | QUARTER 4 | TOTAL GRAMS |
|---|---|---|---|---|---|
| 320 | 828.16 | 841.23 | 827.66 | 803.08 | 3,300.13 |
| 321 | 1,209.92 | 1,155.84 | 1,071.74 | 1,173.11 | 4,610.61 |
| 322 | 3,246.37 | 2,620.19 | 2,426.74 | 2,317.38 | 10,610.68 |
| 323 | 410.92 | 427.34 | 388.53 | 393.35 | 1,620.14 |
| 324 | 413.61 | 470.39 | 483.41 | 398.12 | 1,765.53 |

| | | | | | |
|---|---|---|---|---|---|
| 325 | 1,074.85 | 1,124.15 | 1,200.54 | 1,069.27 | 4,468.81 |
| 326 | 549.34 | 525.9 | 523.84 | 564.26 | 2,163.34 |
| 327 | 3,069.97 | 3,185.10 | 3,127.90 | 3,116.83 | 12,499.80 |
| 328 | 2,339.62 | 2,240.62 | 2,174.46 | 2,059.57 | 8,814.27 |
| 329 | 2,646.89 | 2,111.56 | 2,365.95 | 2,229.52 | 9,353.92 |
| 330 | 2,338.96 | 2,342.94 | 2,209.07 | 2,068.41 | 8,959.38 |
| 331 | 1,910.52 | 2,042.40 | 1,885.92 | 1,749.86 | 7,588.70 |
| 333 | 1,501.05 | 1,551.32 | 1,599.80 | 1,495.08 | 6,147.25 |
| 334 | 2,890.77 | 2,847.29 | 2,623.68 | 2,419.90 | 10,781.64 |
| 335 | 2,583.70 | 2,493.92 | 2,355.36 | 2,428.89 | 9,861.87 |
| 336 | 10,375.01 | 8,927.29 | 6,834.72 | 8,530.78 | 34,667.80 |
| 337 | 3,775.96 | 3,690.24 | 3,776.79 | 3,587.64 | 14,830.63 |
| 338 | 1,318.02 | 1,431.67 | 1,307.61 | 1,305.30 | 5,362.60 |
| 339 | 2,281.10 | 2,014.99 | 2,191.74 | 2,295.85 | 8,783.68 |
| 341 | 768.57 | 707.19 | 694.12 | 715.01 | 2,884.89 |
| 342 | 2,665.18 | 2,648.83 | 2,697.76 | 2,623.80 | 10,635.57 |
| 344 | 1,050.59 | 1,151.16 | 1,188.08 | 1,161.34 | 4,551.17 |
| 346 | 3,546.09 | 3,470.96 | 3,465.21 | 3,517.61 | 13,999.87 |
| 347 | 1,892.90 | 1,682.35 | 1,675.50 | 1,733.07 | 6,983.82 |
| 349 | 1,237.88 | 1,234.80 | 1,261.99 | 1,262.50 | 4,997.17 |
| TOTAL | 55,925.95 | 52,939.67 | 50,358.12 | 51,019.53 | 210,243.27 |

STATE: GEORGIA

DRUG CODE:9150DRUG NAME:HYDROMORPHONE
STATE:GEORGIA

| ZIP CODE | QUARTER 1 | QUARTER 2 | QUARTER 3 | QUARTER 4 | TOTAL GRAMS |
|---|---|---|---|---|---|
| 300 | 1,979.33 | 2,128.53 | 1,926.83 | 2,089.79 | 8,124.48 |
| 301 | 1,075.35 | 1,067 | 990.98 | 992.67 | 4,126 |
| 302 | 711.98 | 674.18 | 676.8 | 700.77 | 2,763.73 |
| 303 | 917.47 | 789.83 | 900.95 | 877.2 | 3,485.45 |
| 304 | 185.04 | 164.93 | 161.72 | 152.71 | 664.4 |
| 305 | 597.58 | 622.86 | 608.61 | 598.78 | 2,427.83 |
| 306 | 511.74 | 499.29 | 490.66 | 496.63 | 1,998.32 |
| 307 | 219.66 | 243.05 | 243.21 | 224.48 | 930.4 |
| 308 | 117.84 | 131.18 | 143.61 | 154.38 | 547.01 |
| 309 | 326.93 | 306.67 | 292.76 | 284.63 | 1,210.99 |
| 310 | 315.92 | 312.96 | 302.92 | 311.93 | 1,243.73 |
| 312 | 106.9 | 110.98 | 141.2 | 163.29 | 522.37 |
| 313 | 263.58 | 324.42 | 358.99 | 366.57 | 1,313.56 |
| 314 | 269.17 | 290.94 | 286.87 | 236.47 | 1,083.45 |
| 315 | 549.8 | 532.26 | 480.58 | 496.45 | 2,059.09 |
| 316 | 144.47 | 144.34 | 155.78 | 182.73 | 627.32 |
| 317 | 465.08 | 469.63 | 494.94 | 424.11 | 1,853.76 |
| 318 | 15.43 | 11.13 | 12.15 | 11.53 | 50.24 |
| 319 | 115 | 107.54 | 104.5 | 98.64 | 425.68 |
| 398 | 70.35 | 73.8 | 84.16 | 77.19 | 305.5 |
| TOTAL | 8,958.62 | 9,005.52 | 8,858.22 | 8,940.95 | 35,763.31 |

STATE: GUAM

| ZIP CODE | QUARTER 1 | QUARTER 2 | QUARTER 3 | QUARTER 4 | TOTAL GRAMS |
|---|---|---|---|---|---|
| 969 | 18.14 | 37.35 | 36 | 34.63 | 126.12 |
| TOTAL | 18.14 | 37.35 | 36 | 34.63 | 126.12 |

STATE: HAWAII

| ZIP CODE | QUARTER 1 | QUARTER 2 | QUARTER 3 | QUARTER 4 | TOTAL GRAMS |
|---|---|---|---|---|---|
| 967 | 440.56 | 410.91 | 400.21 | 393.94 | 1,645.62 |
| 968 | 404.85 | 360.9 | 370.46 | 333.46 | 1,469.67 |
| TOTAL | 845.41 | 771.81 | 770.67 | 727.4 | 3,115.29 |

RETAIL DRUG DISTRIBUTION BY ZIP CODE WITHIN STATE BY GRAMS WT

STATE: IDAHO

| ZIP CODE | QUARTER 1 | QUARTER 2 | QUARTER 3 | QUARTER 4 | TOTAL GRAMS |
|---|---|---|---|---|---|
| 832 | 107.95 | 113.28 | 100.23 | 130.49 | 451.95 |
| 833 | 231.04 | 220.24 | 228.48 | 202.8 | 882.56 |
| 834 | 86.24 | 92.25 | 89.3 | 117.63 | 385.42 |
| 835 | 135.87 | 128.44 | 113.75 | 116.06 | 494.12 |
| 836 | 647.88 | 671.98 | 555.72 | 613.83 | 2,489.41 |
| 837 | 631.11 | 650.04 | 498.67 | 555.94 | 2,335.76 |
| 838 | 291.41 | 304.15 | 309.48 | 301.98 | 1,207.02 |
| TOTAL | 2,131.50 | 2,180.38 | 1,895.63 | 2,038.73 | 8,246.24 |

STATE: ILLINOIS

| ZIP CODE | QUARTER 1 | QUARTER 2 | QUARTER 3 | QUARTER 4 | TOTAL GRAMS |
|---|---|---|---|---|---|
| 600 | 1,821.22 | 1,971.05 | 1,856.51 | 1,909.75 | 7,558.53 |
| 601 | 1,379.47 | 1,429.51 | 1,275.20 | 1,238.66 | 5,322.84 |
| 602 | 70.66 | 115.74 | 158.54 | 89.07 | 434.01 |
| 603 | 96.44 | 113.29 | 88.94 | 80.2 | 378.87 |
| 604 | 1,592.46 | 1,551.49 | 1,497.45 | 1,485.22 | 6,126.62 |
| 605 | 828.86 | 845.9 | 853.98 | 839.41 | 3,368.15 |
| 606 | 1,813.50 | 1,761.76 | 1,796.35 | 1,703.88 | 7,075.49 |
| 607 | 63.24 | 70.36 | 76.36 | 54.31 | 264.27 |
| 608 | 171.69 | 157.7 | 143.34 | 146.87 | 619.6 |
| 609 | 173.28 | 193.93 | 174.74 | 188.78 | 730.73 |
| 610 | 107.03 | 105.34 | 89.54 | 91 | 392.91 |
| 611 | 210.82 | 219.25 | 256.62 | 227.28 | 913.97 |
| 612 | 75.55 | 84.98 | 87.93 | 79.93 | 328.39 |
| 613 | 90.67 | 86.14 | 76.28 | 84.63 | 337.72 |
| 614 | 50.98 | 49.03 | 55.46 | 45.56 | 201.03 |
| 615 | 107.85 | 111.57 | 96.75 | 90.74 | 406.91 |
| 616 | 223.01 | 204.99 | 229.14 | 193.52 | 850.66 |
| 617 | 190.48 | 193.16 | 216.01 | 220.32 | 819.97 |
| 618 | 315.77 | 272.72 | 292.28 | 286.21 | 1,166.98 |
| 619 | 54.25 | 43.11 | 33.9 | 99.75 | 231.01 |
| 620 | 207.52 | 213.17 | 226.49 | 206.37 | 853.55 |
| 622 | 320.57 | 282.4 | 286.71 | 256.43 | 1,146.11 |
| 623 | 75.88 | 78.09 | 71.13 | 71.63 | 296.73 |
| 624 | 102.73 | 110.11 | 99.34 | 107.45 | 419.63 |

DRUG CODE:9150DRUG NAME:HYDROMORPHONE
STATE:ILLINOIS

| ZIP CODE | QUARTER 1 | QUARTER 2 | QUARTER 3 | QUARTER 4 | TOTAL GRAMS |
|---|---|---|---|---|---|
| 625 | 256.79 | 233.37 | 251.57 | 244.48 | 986.21 |
| 626 | 101.88 | 112.58 | 113.29 | 92 | 419.75 |
| 627 | 160.24 | 194.64 | 197.21 | 170.56 | 722.65 |
| 628 | 105.03 | 85.65 | 88.84 | 75.88 | 355.4 |
| 629 | 183.72 | 156.11 | 179.16 | 184.42 | 703.41 |
| TOTAL | 10,951.59 | 11,047.14 | 10,869.06 | 10,564.31 | 43,432.10 |

STATE: INDIANA

| ZIP CODE | QUARTER 1 | QUARTER 2 | QUARTER 3 | QUARTER 4 | TOTAL GRAMS |
|---|---|---|---|---|---|
| 460 | 462.44 | 407.32 | 406.75 | 442.98 | 1,719.49 |
| 461 | 397.09 | 391.88 | 359.98 | 372.98 | 1,521.93 |
| 462 | 1,621.59 | 1,559.16 | 1,815.63 | 1,594.62 | 6,591 |
| 463 | 737.57 | 752.12 | 676.72 | 593.48 | 2,759.89 |
| 464 | 53.25 | 52.27 | 47.35 | 41.48 | 194.35 |
| 465 | 648.94 | 504 | 499.33 | 488.58 | 2,140.85 |
| 466 | 105.39 | 125.84 | 117.23 | 91.97 | 440.43 |
| 467 | 301.83 | 343.46 | 296.8 | 292.98 | 1,235.07 |

| | | | | | |
|---|---|---|---|---|---|
| 468 | 526.52 | 410.78 | 408.02 | 439.87 | 1,785.19 |
| 469 | 221.66 | 182.73 | 196.36 | 180.7 | 781.45 |
| 470 | 52 | 54.04 | 56.31 | 35.4 | 197.75 |
| 471 | 456.99 | 353.98 | 312.07 | 262.58 | 1,385.62 |
| 472 | 245.04 | 254.98 | 211.73 | 197.24 | 908.99 |
| 473 | 420.42 | 343.56 | 306.52 | 298.74 | 1,369.24 |
| 474 | 294.94 | 228.29 | 197.48 | 221.46 | 942.17 |
| 475 | 314.76 | 339.58 | 236.03 | 289.38 | 1,179.75 |
| 476 | 104.04 | 95.97 | 95.81 | 89.29 | 385.11 |
| 477 | 314.94 | 372.81 | 329.21 | 277.15 | 1,294.11 |
| 478 | 169.25 | 195.65 | 182.59 | 149.2 | 696.69 |
| 479 | 208.46 | 212.48 | 210 | 246.33 | 877.27 |
| TOTAL | 7,657.12 | 7,180.90 | 6,961.92 | 6,606.41 | 28,406.35 |

STATE: IOWA

| ZIP CODE | QUARTER 1 | QUARTER 2 | QUARTER 3 | QUARTER 4 | TOTAL GRAMS |
|---|---|---|---|---|---|
| 500 | 307.67 | 339.04 | 316.65 | 313.34 | 1,276.70 |
| 501 | 268.88 | 227.91 | 229.9 | 224.61 | 951.3 |
| 502 | 227.38 | 216.66 | 171.82 | 203.67 | 819.53 |
| 503 | 930.59 | 854.04 | 852.3 | 737.23 | 3,374.16 |
| 504 | 62.29 | 53.67 | 64.36 | 78.84 | 259.16 |
| 505 | 62.81 | 59.18 | 54.32 | 55.86 | 232.17 |
| 506 | 102.68 | 72.57 | 80.16 | 85.86 | 341.27 |
| 507 | 96.96 | 87.22 | 70.6 | 76.9 | 331.68 |
| 508 | 23.49 | 25.76 | 22.68 | 22.75 | 94.68 |
| 510 | 11.99 | 13.89 | 12.55 | 13.94 | 52.37 |
| 511 | 59.93 | 48.75 | 65.87 | 57.84 | 232.39 |
| 512 | 11.63 | 6.49 | 9.68 | 8.26 | 36.06 |
| 513 | 25.53 | 30.16 | 26 | 27.72 | 109.41 |
| 514 | 30.24 | 20.94 | 25.59 | 27.12 | 103.89 |
| 515 | 71.78 | 66.97 | 62.18 | 69.02 | 269.95 |
| 516 | 9.51 | 9.07 | 7.8 | 10.55 | 36.93 |
| 520 | 85.96 | 86.25 | 76.01 | 74.37 | 322.59 |
| 521 | 18.9 | 15.37 | 24.8 | 21.19 | 80.26 |
| 522 | 451.77 | 440.43 | 380.35 | 397.67 | 1,670.22 |
| 523 | 94.56 | 90.49 | 91.2 | 77.8 | 354.05 |
| 524 | 192.89 | 152.1 | 153.27 | 169.82 | 668.08 |
| 525 | 208.74 | 236.48 | 259.48 | 237.17 | 941.87 |
| 526 | 122.89 | 114.26 | 117.87 | 108.22 | 463.24 |
| 527 | 114.5 | 116.85 | 120.11 | 130.38 | 481.84 |
| 528 | 126.66 | 116.36 | 179.82 | 131.88 | 554.72 |
| TOTAL | 3,720.23 | 3,500.91 | 3,475.37 | 3,362.01 | 14,058.52 |

STATE: KANSAS

| ZIP CODE | QUARTER 1 | QUARTER 2 | QUARTER 3 | QUARTER 4 | TOTAL GRAMS |
|---|---|---|---|---|---|
| 660 | 491.84 | 478.6 | 456.46 | 429.95 | 1,856.85 |
| 661 | 203.53 | 213.91 | 218.05 | 231.97 | 867.46 |
| 662 | 904.13 | 1,108.14 | 1,031.19 | 907.38 | 3,950.84 |
| 664 | 136.52 | 131.03 | 149.08 | 109.66 | 526.29 |
| 665 | 169.02 | 152.23 | 120.19 | 141.27 | 582.71 |
| 666 | 141.76 | 141.33 | 151.01 | 126.06 | 560.16 |
| 667 | 105.42 | 117.97 | 130.18 | 140.97 | 494.54 |

DRUG CODE:9150DRUG NAME:HYDROMORPHONE
STATE:KANSAS

| ZIP CODE | QUARTER 1 | QUARTER 2 | QUARTER 3 | QUARTER 4 | TOTAL GRAMS |
|---|---|---|---|---|---|
| 668 | 46.92 | 41.25 | 40.96 | 39.53 | 168.66 |
| 669 | 19.78 | 25.6 | 15.86 | 17.05 | 78.29 |
| 670 | 175.47 | 167.27 | 173.03 | 160.84 | 676.61 |
| 671 | 135.38 | 119.83 | 123.96 | 124 | 503.17 |

| | | | | | |
|---|---|---|---|---|---|
| 672 | 788.74 | 732.61 | 690.28 | 753.27 | 2,964.90 |
| 673 | 33.79 | 33.99 | 32.8 | 26.16 | 126.74 |
| 674 | 143.45 | 165.06 | 169.15 | 189.93 | 667.59 |
| 675 | 242.14 | 219.53 | 237.55 | 249.85 | 949.07 |
| 676 | 65.01 | 85.57 | 70.64 | 68.08 | 289.3 |
| 677 | 20.55 | 27.67 | 31.74 | 27.02 | 106.98 |
| 678 | 69.52 | 78.87 | 75.05 | 82.54 | 305.98 |
| 679 | 18.31 | 22.09 | 27.79 | 22.45 | 90.64 |
| TOTAL | 3,911.28 | 4,062.55 | 3,944.97 | 3,847.98 | 15,766.78 |

STATE: KENTUCKY

| ZIP CODE | QUARTER 1 | QUARTER 2 | QUARTER 3 | QUARTER 4 | TOTAL GRAMS |
|---|---|---|---|---|---|
| 400 | 134.95 | 126.68 | 120.14 | 121.21 | 502.98 |
| 401 | 53.02 | 39.4 | 43.27 | 45.07 | 180.76 |
| 402 | 815.8 | 931.02 | 896.11 | 870.68 | 3,513.61 |
| 403 | 103.43 | 108.37 | 76.34 | 90.22 | 378.36 |
| 404 | 99.12 | 37.07 | 41.98 | 33.28 | 211.45 |
| 405 | 263.25 | 314.72 | 322.5 | 358.84 | 1,259.31 |
| 406 | 32.22 | 29.72 | 36.88 | 34.74 | 133.56 |
| 407 | 32.03 | 27.36 | 17.73 | 26.15 | 103.27 |
| 408 | 8.66 | 10.26 | 6.51 | 6.69 | 32.12 |
| 409 | 15.25 | 17.2 | 19.13 | 18.17 | 69.75 |
| 410 | 179.79 | 159.61 | 148.99 | 150.88 | 639.27 |
| 411 | 70.77 | 94.56 | 66.01 | 114.92 | 346.26 |
| 412 | 11.28 | 12.39 | 8.01 | 6.94 | 38.62 |
| 413 | 4.14 | 13.34 | 8.11 | 7.49 | 33.08 |
| 414 | 8.51 | 4.83 | 5.36 | 1.1 | 19.8 |
| 415 | 49.9 | 54.24 | 44.32 | 43.94 | 192.4 |
| 416 | 8.49 | 8.1 | 7.03 | 6.9 | 30.52 |
| 417 | 62.34 | 82.52 | 75.69 | 48.4 | 268.95 |
| 418 | 3.1 | 1.13 | 4.4 | 2.03 | 10.66 |
| 420 | 264.74 | 241.76 | 292.36 | 307.57 | 1,106.43 |
| 421 | 177.35 | 168.86 | 175.83 | 202.94 | 724.98 |
| 422 | 136.92 | 155.61 | 164.34 | 157.74 | 614.61 |
| 423 | 188.03 | 240.22 | 170.65 | 185.63 | 784.53 |
| 424 | 166.34 | 205.94 | 186.55 | 167.05 | 725.88 |
| 425 | 28.02 | 42.99 | 65.84 | 95.13 | 231.98 |
| 426 | 10.46 | 9.01 | 14.79 | 13.02 | 47.28 |
| 427 | 107.51 | 103.34 | 98.23 | 97.6 | 406.68 |
| TOTAL | 3,035.42 | 3,240.25 | 3,117.10 | 3,214.33 | 12,607.10 |

STATE: LOUISIANA

| ZIP CODE | QUARTER 1 | QUARTER 2 | QUARTER 3 | QUARTER 4 | TOTAL GRAMS |
|---|---|---|---|---|---|
| 700 | 419.59 | 400.78 | 423.51 | 365.13 | 1,609.01 |
| 701 | 232.75 | 236.33 | 237.65 | 234.56 | 941.29 |
| 703 | 124.12 | 131.54 | 103.32 | 123.91 | 482.89 |
| 704 | 644.97 | 582.52 | 584.04 | 570.74 | 2,382.27 |
| 705 | 1,108.58 | 953.66 | 855.25 | 819.48 | 3,736.97 |
| 706 | 241.14 | 218.33 | 172.5 | 168.26 | 800.23 |
| 707 | 333.6 | 302.79 | 285.96 | 316.1 | 1,238.45 |
| 708 | 351.3 | 355.04 | 334.63 | 348.16 | 1,389.13 |
| 710 | 35.73 | 39.76 | 37.92 | 43.44 | 156.85 |
| 711 | 174.81 | 290.53 | 186.64 | 218.83 | 870.81 |
| 712 | 114.72 | 114.59 | 138.15 | 121.56 | 489.02 |
| 713 | 190 | 230.36 | 207.53 | 181.45 | 809.34 |
| 714 | 110.3 | 83.04 | 79.09 | 77.88 | 350.31 |
| TOTAL | 4,081.61 | 3,939.27 | 3,646.19 | 3,589.50 | 15,256.57 |

STATE: MAINE

| ZIP CODE | QUARTER 1 | QUARTER 2 | QUARTER 3 | QUARTER 4 | TOTAL GRAMS |
|---|---|---|---|---|---|
| 39 | 44.49 | 40.83 | 31.49 | 27.93 | 144.74 |

| | | | | | |
|---|---|---|---|---|---|
| 40 | 442.76 | 466.72 | 483.8 | 391.65 | 1,784.93 |
| 41 | 476.19 | 417.56 | 303.23 | 295.74 | 1,492.72 |
| 42 | 159.02 | 149.92 | 117.74 | 124.45 | 551.13 |
| 43 | 308.32 | 238.39 | 151.44 | 533.05 | 1,231.20 |

DRUG CODE:9150DRUG NAME:HYDROMORPHONE
STATE:MAINE

| ZIP CODE | QUARTER 1 | QUARTER 2 | QUARTER 3 | QUARTER 4 | TOTAL GRAMS |
|---|---|---|---|---|---|
| 44 | 231.53 | 234.04 | 232.2 | 238.73 | 936.5 |
| 45 | 69.74 | 60.25 | 63.23 | 60.93 | 254.15 |
| 46 | 106.52 | 87.24 | 79.19 | 55.41 | 328.36 |
| 47 | 42.47 | 36.56 | 36.73 | 68.05 | 183.81 |
| 48 | 48.6 | 46.62 | 43.13 | 38.53 | 176.88 |
| 49 | 171.67 | 149.58 | 157.64 | 140.63 | 619.52 |
| TOTAL | 2,101.31 | 1,927.71 | 1,699.82 | 1,975.10 | 7,703.94 |

STATE: MARYLAND

| ZIP CODE | QUARTER 1 | QUARTER 2 | QUARTER 3 | QUARTER 4 | TOTAL GRAMS |
|---|---|---|---|---|---|
| 206 | 532.38 | 527.79 | 495 | 519.37 | 2,074.54 |
| 207 | 861.25 | 830.77 | 790.25 | 748.29 | 3,230.56 |
| 208 | 1,284.29 | 1,269.36 | 1,299.43 | 1,170.29 | 5,023.37 |
| 209 | 260.25 | 247.87 | 244.02 | 244.54 | 996.68 |
| 210 | 1,838.35 | 1,835.41 | 1,648.29 | 1,582.04 | 6,904.09 |
| 211 | 641.89 | 629.36 | 712.13 | 610.79 | 2,594.17 |
| 212 | 2,586.34 | 2,592.41 | 2,437.30 | 2,338.17 | 9,954.22 |
| 214 | 157.76 | 165.96 | 132.17 | 147.82 | 603.71 |
| 215 | 148.15 | 142.87 | 189.36 | 85.77 | 566.15 |
| 216 | 183.28 | 180.38 | 196.39 | 165.79 | 725.84 |
| 217 | 884.63 | 859.45 | 855.42 | 854.8 | 3,454.30 |
| 218 | 287.4 | 245.29 | 284.01 | 290.81 | 1,107.51 |
| 219 | 477.89 | 498.94 | 456.21 | 455.87 | 1,888.91 |
| TOTAL | 10,143.86 | 10,025.86 | 9,739.98 | 9,214.35 | 39,124.05 |

STATE: MASSACHUSETTS

| ZIP CODE | QUARTER 1 | QUARTER 2 | QUARTER 3 | QUARTER 4 | TOTAL GRAMS |
|---|---|---|---|---|---|
| 10 | 587.37 | 641.12 | 545.3 | 557.13 | 2,330.92 |
| 11 | 382.04 | 388.12 | 390.93 | 412.27 | 1,573.36 |
| 12 | 134.67 | 129.86 | 110.36 | 109.28 | 484.17 |
| 13 | 69.6 | 65.29 | 64.39 | 68.54 | 267.82 |
| 14 | 102.89 | 101.17 | 96 | 93.82 | 393.88 |
| 15 | 214.51 | 188.75 | 196.33 | 173.93 | 773.52 |
| 16 | 246.29 | 242.17 | 228.65 | 214.54 | 931.65 |
| 17 | 408.73 | 376.57 | 393.27 | 404.12 | 1,582.69 |
| 18 | 1,612.91 | 1,202.52 | 1,453.96 | 1,387.76 | 5,657.15 |
| 19 | 545.1 | 504.57 | 470.85 | 454.44 | 1,974.96 |
| 20 | 381.86 | 365.84 | 415.73 | 393.27 | 1,556.70 |
| 21 | 1,721.03 | 1,717.44 | 1,642.48 | 1,614.52 | 6,695.47 |
| 22 | 352.45 | 353.43 | 333.85 | 311.79 | 1,351.52 |
| 23 | 531.64 | 517.55 | 491.58 | 516.19 | 2,056.96 |
| 24 | 253.22 | 272.04 | 246.29 | 279.19 | 1,050.74 |
| 25 | 141.73 | 136.17 | 123.89 | 117.79 | 519.58 |
| 26 | 219.92 | 176.84 | 143.56 | 148.4 | 688.72 |
| 27 | 578.81 | 522.75 | 546.61 | 545.05 | 2,193.22 |
| TOTAL | 8,484.77 | 7,902.20 | 7,894.03 | 7,802.03 | 32,083.03 |

STATE: MICHIGAN

| ZIP CODE | QUARTER 1 | QUARTER 2 | QUARTER 3 | QUARTER 4 | TOTAL GRAMS |
|---|---|---|---|---|---|
| 480 | 1,744.62 | 1,714.84 | 1,504.39 | 1,475.79 | 6,439.64 |
| 481 | 1,506.80 | 1,378.46 | 1,261.39 | 1,503.85 | 5,650.50 |

| | | | | | |
|---|---|---|---|---|---|
| 482 | 974.79 | 915.47 | 962.59 | 902.4 | 3,755.25 |
| 483 | 1,581.79 | 1,855.15 | 1,174.48 | 1,590.51 | 6,201.93 |
| 484 | 480.32 | 543.62 | 414.77 | 421.22 | 1,859.93 |
| 485 | 284.93 | 245.64 | 275.6 | 263.35 | 1,069.52 |
| 486 | 591.83 | 518.86 | 514.52 | 488.67 | 2,113.88 |
| 487 | 166.75 | 191.24 | 181.55 | 147.95 | 687.49 |
| 488 | 445.9 | 414.37 | 411.38 | 377.05 | 1,648.70 |
| 489 | 248.05 | 213.73 | 194.83 | 207.28 | 863.89 |
| 490 | 635.95 | 704.39 | 614.17 | 597.15 | 2,551.66 |
| 491 | 24.56 | 40.25 | 32.01 | 42.29 | 139.11 |
| 492 | 213.06 | 220.14 | 206 | 160.77 | 799.97 |
| 493 | 172.62 | 173.24 | 184.96 | 166.91 | 697.73 |
| 494 | 433.23 | 375.94 | 376.66 | 363.05 | 1,548.88 |
| 495 | 577.76 | 555.27 | 559.94 | 502.68 | 2,195.65 |
| 496 | 485.97 | 514.03 | 446.73 | 423.15 | 1,869.88 |
| 497 | 333.21 | 338.24 | 318.37 | 323.75 | 1,313.57 |
| 498 | 251.3 | 222.16 | 246.08 | 224.76 | 944.3 |
| 499 | 64.89 | 55.74 | 51.51 | 63.94 | 236.08 |

DRUG CODE:9150DRUG NAME:HYDROMORPHONE
STATE:MICHIGAN

| ZIP CODE | QUARTER 1 | QUARTER 2 | QUARTER 3 | QUARTER 4 | TOTAL GRAMS |
|---|---|---|---|---|---|
| TOTAL | 11,218.33 | 11,190.78 | 9,931.93 | 10,246.52 | 42,587.56 |

STATE: MINNESOTA

| ZIP CODE | QUARTER 1 | QUARTER 2 | QUARTER 3 | QUARTER 4 | TOTAL GRAMS |
|---|---|---|---|---|---|
| 550 | 554.77 | 534.31 | 550.78 | 545 | 2,184.86 |
| 551 | 1,097.71 | 1,038.18 | 978.82 | 1,521.03 | 4,635.74 |
| 553 | 700.16 | 651.26 | 713.46 | 741.6 | 2,806.48 |
| 554 | 2,141.56 | 2,077.70 | 2,077.79 | 1,999.42 | 8,296.47 |
| 556 | 7.05 | 6.99 | 7.23 | 4.87 | 26.14 |
| 557 | 175.78 | 171.11 | 151.32 | 137.53 | 635.74 |
| 558 | 225.05 | 265.2 | 212.69 | 186.84 | 889.78 |
| 559 | 563.33 | 574.65 | 523.24 | 596.91 | 2,258.13 |
| 560 | 144.58 | 135.76 | 133.79 | 136.41 | 550.54 |
| 561 | 17.12 | 12.74 | 14.92 | 11.09 | 55.87 |
| 562 | 84.72 | 80.6 | 78.12 | 83.41 | 326.85 |
| 563 | 214.61 | 232.59 | 203.17 | 170.82 | 821.19 |
| 564 | 160.71 | 161.53 | 149.56 | 143.13 | 614.93 |
| 565 | 168.12 | 165.62 | 176.82 | 175.54 | 686.1 |
| 566 | 62.41 | 70.13 | 62.73 | 61.25 | 256.52 |
| 567 | 18 | 15.8 | 16.39 | 26.33 | 76.52 |
| TOTAL | 6,335.68 | 6,194.17 | 6,050.83 | 6,541.18 | 25,121.86 |

STATE: MISSISSIPPI

| ZIP CODE | QUARTER 1 | QUARTER 2 | QUARTER 3 | QUARTER 4 | TOTAL GRAMS |
|---|---|---|---|---|---|
| 386 | 149.4 | 144.53 | 163.9 | 191.79 | 649.62 |
| 387 | 14.69 | 21.62 | 10.81 | 13.02 | 60.14 |
| 388 | 174.92 | 209.73 | 235.94 | 178.33 | 798.92 |
| 389 | 23.86 | 26.89 | 27.08 | 30.49 | 108.32 |
| 390 | 129.83 | 108.47 | 110.7 | 109.03 | 458.03 |
| 391 | 3,305.19 | 3,210.46 | 3,061.19 | 3,388.55 | 12,965.39 |
| 392 | 413.05 | 556.82 | 395.47 | 298.56 | 1,663.90 |
| 393 | 109.33 | 105.93 | 90.76 | 91.51 | 397.53 |
| 394 | 420.83 | 439.06 | 444.32 | 370.43 | 1,674.64 |
| 395 | 382.42 | 329.15 | 351.39 | 359.57 | 1,422.53 |
| 396 | 105.68 | 69.17 | 91.43 | 112.89 | 379.17 |
| 397 | 31.64 | 36.01 | 43.87 | 46.47 | 157.99 |
| TOTAL | 5,260.84 | 5,257.84 | 5,026.86 | 5,190.64 | 20,736.18 |

STATE: MISSOURI

RETAIL DRUG DISTRIBUTION BY ZIP CODE WITHIN STATE BY GRAMS WT

| ZIP CODE | QUARTER 1 | QUARTER 2 | QUARTER 3 | QUARTER 4 | TOTAL GRAMS |
|---|---|---|---|---|---|
| 630 | 753.92 | 826.94 | 720.38 | 961.83 | 3,263.07 |
| 631 | 1,345.18 | 1,192.01 | 1,206.87 | 1,181.45 | 4,925.51 |
| 633 | 335.03 | 333.16 | 318.57 | 303.59 | 1,290.35 |
| 634 | 51.65 | 36.87 | 47.81 | 36.03 | 172.36 |
| 635 | 97.22 | 82.44 | 79.77 | 76.66 | 336.09 |
| 636 | 244.92 | 230.05 | 203.64 | 198.82 | 877.43 |
| 637 | 120.76 | 118.53 | 111.51 | 87.16 | 437.96 |
| 638 | 82.79 | 72.12 | 78.07 | 64.79 | 297.77 |
| 639 | 86.69 | 61.34 | 60.94 | 66.23 | 275.2 |
| 640 | 551.88 | 585.2 | 517.83 | 569.64 | 2,224.55 |
| 641 | 579.08 | 539.46 | 517.02 | 492.5 | 2,128.06 |
| 644 | 131.84 | 148.46 | 114.62 | 124.54 | 519.46 |
| 645 | 658.5 | 602.57 | 592.82 | 580.18 | 2,434.07 |
| 646 | 128.56 | 110.92 | 109.14 | 141.76 | 490.38 |
| 647 | 125.05 | 146.95 | 121.91 | 135.74 | 529.65 |
| 648 | 339.62 | 330.92 | 331.94 | 310.84 | 1,313.32 |
| 650 | 78.74 | 85.24 | 90.6 | 79.07 | 333.65 |
| 651 | 59.05 | 56.53 | 52.29 | 36.32 | 204.19 |
| 652 | 422.79 | 389.13 | 342.14 | 343.04 | 1,497.10 |
| 653 | 53.19 | 62.36 | 52.31 | 47.09 | 214.95 |
| 654 | 50.74 | 40.78 | 41.99 | 38.63 | 172.14 |
| 655 | 83.77 | 89.85 | 72.79 | 89.72 | 336.13 |
| 656 | 337.45 | 352.57 | 353.83 | 354.52 | 1,398.37 |
| 657 | 634.47 | 613.39 | 638.12 | 562.12 | 2,448.10 |
| 658 | 614.13 | 537.28 | 690.78 | 465.07 | 2,307.26 |
| TOTAL | 7,967.02 | 7,645.07 | 7,467.69 | 7,347.34 | 30,427.12 |

STATE: MONTANA

| ZIP CODE | QUARTER 1 | QUARTER 2 | QUARTER 3 | QUARTER 4 | TOTAL GRAMS |
|---|---|---|---|---|---|
| 590 | 110.66 | 100.58 | 104.87 | 102.85 | 418.96 |

DRUG CODE:9150DRUG NAME:HYDROMORPHONE
STATE:MONTANA

| ZIP CODE | QUARTER 1 | QUARTER 2 | QUARTER 3 | QUARTER 4 | TOTAL GRAMS |
|---|---|---|---|---|---|
| 591 | 607.34 | 595.74 | 584.59 | 546.69 | 2,334.36 |
| 592 | 46.17 | 43.82 | 51.81 | 37.44 | 179.24 |
| 593 | 68.14 | 60.24 | 60.95 | 57.77 | 247.1 |
| 594 | 140.92 | 156.31 | 167.27 | 196.21 | 660.71 |
| 595 | 15.21 | 14.2 | 16.91 | 15.69 | 62.01 |
| 596 | 107.33 | 147.23 | 140.3 | 111.38 | 506.24 |
| 597 | 230.36 | 224.4 | 177.48 | 250.82 | 883.06 |
| 598 | 321.31 | 311.88 | 315.86 | 319.16 | 1,268.21 |
| 599 | 138.98 | 132.09 | 196.07 | 199.22 | 666.36 |
| TOTAL | 1,786.42 | 1,786.49 | 1,816.11 | 1,837.23 | 7,226.25 |

STATE: NEBRASKA

| ZIP CODE | QUARTER 1 | QUARTER 2 | QUARTER 3 | QUARTER 4 | TOTAL GRAMS |
|---|---|---|---|---|---|
| 680 | 211.17 | 198.65 | 212.92 | 200.5 | 823.24 |
| 681 | 727.37 | 628.25 | 622.65 | 629.38 | 2,607.65 |
| 683 | 22.23 | 33.23 | 35.52 | 27.74 | 118.72 |
| 684 | 26.46 | 20.05 | 16.01 | 29.88 | 92.4 |
| 685 | 163.36 | 193.28 | 189.16 | 171.44 | 717.24 |
| 686 | 22.61 | 21.62 | 23.79 | 25.82 | 93.84 |
| 687 | 47.32 | 49.13 | 53.15 | 48.25 | 197.85 |
| 688 | 125.93 | 137.63 | 142.32 | 120.08 | 525.96 |
| 689 | 36.99 | 51.05 | 50.9 | 21.5 | 160.44 |
| 690 | 24.76 | 24.14 | 28.61 | 20.55 | 98.06 |

| | | | | | |
|---|---|---|---|---|---|
| 691 | 48.07 | 43.05 | 47 | 42.52 | 180.64 |
| 692 | 3.78 | 4.3 | 4.24 | 2.74 | 15.06 |
| 693 | 54.08 | 50.15 | 61.44 | 56.66 | 222.33 |
| TOTAL | 1,514.13 | 1,454.53 | 1,487.71 | 1,397.06 | 5,853.43 |

STATE: NEVADA

| ZIP CODE | QUARTER 1 | QUARTER 2 | QUARTER 3 | QUARTER 4 | TOTAL GRAMS |
|---|---|---|---|---|---|
| 890 | 960.73 | 824.76 | 776.16 | 777.79 | 3,339.44 |
| 891 | 2,141.74 | 1,909.89 | 1,883 | 2,014.92 | 7,949.55 |
| 893 | 7.04 | 4.02 | 2.84 | 1.23 | 15.13 |
| 894 | 263.12 | 257.28 | 203.98 | 213.87 | 938.25 |
| 895 | 397.96 | 352.66 | 334.95 | 325.11 | 1,410.68 |
| 897 | 146.05 | 161.87 | 129.77 | 123.3 | 560.99 |
| 898 | 30.78 | 28.49 | 34.1 | 30.07 | 123.44 |
| TOTAL | 3,947.42 | 3,538.97 | 3,364.80 | 3,486.29 | 14,337.48 |

STATE: NEW HAMPSHIRE

| ZIP CODE | QUARTER 1 | QUARTER 2 | QUARTER 3 | QUARTER 4 | TOTAL GRAMS |
|---|---|---|---|---|---|
| 30 | 426.05 | 446.58 | 398.36 | 423.66 | 1,694.65 |
| 31 | 286.69 | 265.71 | 259.66 | 315.35 | 1,127.41 |
| 32 | 206.23 | 186.84 | 190.31 | 193.1 | 776.48 |
| 33 | 164.51 | 203.45 | 202.49 | 264.07 | 834.52 |
| 34 | 112.82 | 97.16 | 109.07 | 100.49 | 419.54 |
| 35 | 102.54 | 109.7 | 94.42 | 97.54 | 404.2 |
| 36 | 9.58 | 8.01 | 10.89 | 8.23 | 36.71 |
| 37 | 334.82 | 302.39 | 294.97 | 301.99 | 1,234.17 |
| 38 | 490.48 | 442.75 | 451.18 | 473.34 | 1,857.75 |
| TOTAL | 2,133.72 | 2,062.59 | 2,011.35 | 2,177.77 | 8,385.43 |

STATE: NEW JERSEY

| ZIP CODE | QUARTER 1 | QUARTER 2 | QUARTER 3 | QUARTER 4 | TOTAL GRAMS |
|---|---|---|---|---|---|
| 70 | 1,873.68 | 1,776.85 | 1,768.24 | 1,657.39 | 7,076.16 |
| 71 | 248.9 | 239.91 | 255.77 | 266 | 1,010.58 |
| 72 | 279.06 | 238.1 | 151.92 | 105.69 | 774.77 |
| 73 | 90.12 | 87.29 | 77.28 | 73.35 | 328.04 |
| 74 | 878.22 | 1,076.39 | 934.31 | 684.32 | 3,573.24 |
| 75 | 146.81 | 215.18 | 140.08 | 143.6 | 645.67 |
| 76 | 817.63 | 817.96 | 914.43 | 988.42 | 3,538.44 |
| 77 | 789.5 | 802.73 | 836.24 | 823.02 | 3,251.49 |
| 78 | 417.8 | 419.65 | 421.85 | 448.16 | 1,707.46 |
| 79 | 457.62 | 479.93 | 491.57 | 549.91 | 1,979.03 |
| 80 | 1,631.30 | 1,626.07 | 1,520.20 | 1,463.90 | 6,241.47 |
| 81 | 179.54 | 198.99 | 188.61 | 198.43 | 765.57 |
| 82 | 261.66 | 278 | 312.43 | 222.37 | 1,074.46 |
| 83 | 204.32 | 165.25 | 166.35 | 155.62 | 691.54 |
| 84 | 23.03 | 26.91 | 30.22 | 23.58 | 103.74 |
| 85 | 328.09 | 296.27 | 251.44 | 244.3 | 1,120.10 |

DRUG CODE:9150DRUG NAME:HYDROMORPHONE
STATE:NEW JERSEY

| ZIP CODE | QUARTER 1 | QUARTER 2 | QUARTER 3 | QUARTER 4 | TOTAL GRAMS |
|---|---|---|---|---|---|
| 86 | 199.7 | 189.87 | 170.39 | 171.43 | 731.39 |
| 87 | 481.17 | 450.63 | 430.34 | 417.7 | 1,779.84 |
| 88 | 1,208.29 | 1,146.06 | 1,100.18 | 1,154.68 | 4,609.21 |
| 89 | 180.97 | 192.69 | 182.21 | 167.33 | 723.2 |
| TOTAL | 10,697.41 | 10,724.73 | 10,344.06 | 9,959.20 | 41,725.40 |

STATE: NEW MEXICO

| ZIP CODE | QUARTER 1 | QUARTER 2 | QUARTER 3 | QUARTER 4 | TOTAL GRAMS |
|---|---|---|---|---|---|
| 870 | 88.29 | 93.88 | 91.52 | 93.94 | 367.63 |

| | | | | | |
|---|---|---|---|---|---|
| 871 | 960 | 949.35 | 882.77 | 821.9 | 3,614.02 |
| 873 | 13.83 | 12.46 | 8.1 | 7.9 | 42.29 |
| 874 | 89.11 | 86.82 | 93.96 | 81.95 | 351.84 |
| 875 | 170.51 | 150.62 | 145.12 | 145.37 | 611.62 |
| 877 | 35.65 | 27.48 | 27.19 | 30.89 | 121.21 |
| 878 | 3.04 | 3.81 | 2.31 | 2.84 | 12 |
| 879 | 3.55 | 5.62 | 8.6 | 6.9 | 24.67 |
| 880 | 412.03 | 319.22 | 289.49 | 262.22 | 1,282.96 |
| 881 | 30.49 | 27.55 | 30.28 | 21.37 | 109.69 |
| 882 | 129.53 | 140.71 | 145 | 131.4 | 546.64 |
| 883 | 58.5 | 60.01 | 49.29 | 35.03 | 202.83 |
| 884 | 8.03 | 5.07 | 6.96 | 1.8 | 21.86 |
| TOTAL | 2,002.56 | 1,882.60 | 1,780.59 | 1,643.51 | 7,309.26 |

STATE: NEW YORK

| ZIP CODE | QUARTER 1 | QUARTER 2 | QUARTER 3 | QUARTER 4 | TOTAL GRAMS |
|---|---|---|---|---|---|
| 100 | 3,726.06 | 3,642.52 | 3,583.22 | 3,425.79 | 14,377.59 |
| 101 | 54.69 | 38.9 | 41.52 | 40.53 | 175.64 |
| 102 | 3.37 | 3.72 | 5.32 | 4.26 | 16.67 |
| 103 | 600.16 | 679.92 | 681.36 | 767.99 | 2,729.43 |
| 104 | 885.4 | 777.97 | 811.59 | 905.07 | 3,380.03 |
| 105 | 611.43 | 562.59 | 559.61 | 549.18 | 2,282.81 |
| 106 | 160.87 | 151.15 | 129.01 | 144.25 | 585.28 |
| 107 | 182.31 | 180.22 | 183.77 | 182.02 | 728.32 |
| 108 | 93.78 | 72.03 | 93.33 | 77.78 | 336.92 |
| 109 | 928.19 | 846.14 | 794.06 | 739.77 | 3,308.16 |
| 110 | 1,028.97 | 987.85 | 1,106.66 | 1,331.25 | 4,454.73 |
| 111 | 216.56 | 221.34 | 211.16 | 214.21 | 863.27 |
| 112 | 1,682.95 | 1,605.12 | 1,650.35 | 1,556.12 | 6,494.54 |
| 113 | 744.2 | 714.09 | 655.16 | 628.47 | 2,741.92 |
| 114 | 360.05 | 335.62 | 327.2 | 390.84 | 1,413.71 |
| 115 | 982.03 | 908.81 | 947.31 | 1,036.45 | 3,874.60 |
| 116 | 62.2 | 68.92 | 50.18 | 42.11 | 223.41 |
| 117 | 2,868.79 | 2,868.47 | 2,830.40 | 3,011.22 | 11,578.88 |
| 118 | 243.89 | 228.27 | 175.88 | 161.04 | 809.08 |
| 119 | 207.08 | 236.41 | 209.48 | 250.41 | 903.38 |
| 120 | 277.57 | 280.8 | 231.99 | 222.38 | 1,012.74 |
| 121 | 120.46 | 103.21 | 112.19 | 96.28 | 432.14 |
| 122 | 287.66 | 279.51 | 244.83 | 273.06 | 1,085.06 |
| 123 | 66.04 | 77.09 | 113.43 | 79.05 | 335.61 |
| 124 | 296.23 | 264.09 | 241.55 | 246.91 | 1,048.78 |
| 125 | 451.72 | 406.54 | 426.9 | 369.07 | 1,654.23 |
| 126 | 139.31 | 161.76 | 198.09 | 146.25 | 645.41 |
| 127 | 174.23 | 200.7 | 135.19 | 207.79 | 717.91 |
| 128 | 257.25 | 244.28 | 278.52 | 290.13 | 1,070.18 |
| 129 | 362.51 | 374.55 | 366.36 | 364.29 | 1,467.71 |
| 130 | 268.99 | 238.36 | 216.35 | 193.87 | 917.57 |
| 131 | 128.2 | 114.45 | 108.23 | 108.3 | 459.18 |
| 132 | 241.28 | 212.59 | 192.64 | 172.92 | 819.43 |
| 133 | 162.28 | 135.39 | 122.61 | 94.44 | 514.72 |
| 134 | 149.96 | 140.32 | 134.57 | 95.8 | 520.65 |
| 135 | 104.88 | 102.09 | 100.31 | 74.06 | 381.34 |
| 136 | 168.55 | 168.1 | 156.46 | 151.69 | 644.8 |
| 137 | 78.24 | 79.89 | 76.94 | 82.58 | 317.65 |
| 138 | 148.72 | 136.04 | 133.35 | 130.35 | 548.46 |
| 139 | 112.6 | 97.09 | 89.44 | 86.03 | 385.16 |
| 140 | 401.16 | 366.22 | 328.74 | 329.59 | 1,425.71 |
| 141 | 173.47 | 165.97 | 164.58 | 158.06 | 662.08 |
| 142 | 816.82 | 725.91 | 706.08 | 647.12 | 2,895.93 |

RETAIL DRUG DISTRIBUTION BY ZIP CODE WITHIN STATE BY GRAMS WT

STATE:NEW YORK

| ZIP CODE | QUARTER 1 | QUARTER 2 | QUARTER 3 | QUARTER 4 | TOTAL GRAMS |
|---|---|---|---|---|---|
| 143 | 76.95 | 62.03 | 65.42 | 61.35 | 265.75 |
| 144 | 416.41 | 407.48 | 358.02 | 368.35 | 1,550.26 |
| 145 | 327.3 | 314.53 | 310.24 | 306.99 | 1,259.06 |
| 146 | 1,168.53 | 1,151.38 | 1,048.35 | 1,181.31 | 4,549.57 |
| 147 | 107.62 | 95.67 | 91.74 | 91.08 | 386.11 |
| 148 | 319.33 | 296.62 | 291.03 | 274.07 | 1,181.05 |
| 149 | 63.54 | 51.41 | 49.69 | 48.54 | 213.18 |
| TOTAL | 23,510.79 | 22,584.13 | 22,140.41 | 22,410.47 | 90,645.80 |

STATE: NORTH CAROLINA

| ZIP CODE | QUARTER 1 | QUARTER 2 | QUARTER 3 | QUARTER 4 | TOTAL GRAMS |
|---|---|---|---|---|---|
| 270 | 356.99 | 334.8 | 320.56 | 325.55 | 1,337.90 |
| 271 | 757.96 | 699.26 | 663.39 | 690.58 | 2,811.19 |
| 272 | 739.01 | 847.7 | 798.86 | 807.08 | 3,192.65 |
| 273 | 443.42 | 428.44 | 393.1 | 416.37 | 1,681.33 |
| 274 | 352.15 | 333.26 | 301.63 | 313.4 | 1,300.44 |
| 275 | 1,380.48 | 1,614.06 | 1,223.92 | 1,378.32 | 5,596.78 |
| 276 | 533.19 | 502.23 | 519.75 | 505.09 | 2,060.26 |
| 277 | 607.88 | 778.46 | 1,078.09 | 1,084.53 | 3,548.96 |
| 278 | 537.68 | 538.95 | 567.53 | 513.99 | 2,158.15 |
| 279 | 310.62 | 303.68 | 277.49 | 305.78 | 1,197.57 |
| 280 | 868.8 | 826.93 | 739.16 | 755.64 | 3,190.53 |
| 281 | 552.36 | 515.78 | 507.46 | 494.96 | 2,070.56 |
| 282 | 751.51 | 858.29 | 1,029.39 | 938.33 | 3,577.52 |
| 283 | 959.81 | 898.99 | 840.5 | 801.5 | 3,500.80 |
| 284 | 746.21 | 725.73 | 696.55 | 598.84 | 2,767.33 |
| 285 | 567 | 578.45 | 551.24 | 555.2 | 2,251.89 |
| 286 | 833.18 | 867.62 | 869 | 1,021.51 | 3,591.31 |
| 287 | 529.59 | 491.71 | 454 | 450.16 | 1,925.46 |
| 288 | 504.67 | 452.06 | 329.48 | 263.45 | 1,549.66 |
| 289 | 69.88 | 83.77 | 78.38 | 79.32 | 311.35 |
| TOTAL | 12,402.39 | 12,680.17 | 12,239.48 | 12,299.60 | 49,621.64 |

STATE: NORTH DAKOTA

| ZIP CODE | QUARTER 1 | QUARTER 2 | QUARTER 3 | QUARTER 4 | TOTAL GRAMS |
|---|---|---|---|---|---|
| 580 | 37.11 | 40.61 | 34.99 | 28.58 | 141.29 |
| 581 | 245.14 | 193.14 | 207.78 | 192.86 | 838.92 |
| 582 | 70.23 | 66.52 | 65.82 | 59.1 | 261.67 |
| 583 | 34.55 | 28.17 | 27.01 | 19.52 | 109.25 |
| 584 | 32.18 | 29.13 | 28.96 | 21.18 | 111.45 |
| 585 | 321.74 | 347.14 | 318.97 | 281.77 | 1,269.62 |
| 586 | 23.5 | 32.58 | 27.01 | 22.78 | 105.87 |
| 587 | 55.52 | 58.56 | 57.77 | 62.1 | 233.95 |
| 588 | 26.44 | 30.77 | 34.7 | 31 | 122.91 |
| TOTAL | 846.41 | 826.62 | 803.01 | 718.89 | 3,194.93 |

STATE: OHIO

| ZIP CODE | QUARTER 1 | QUARTER 2 | QUARTER 3 | QUARTER 4 | TOTAL GRAMS |
|---|---|---|---|---|---|
| 430 | 859.84 | 773.95 | 959.5 | 892.08 | 3,485.37 |
| 431 | 355.79 | 331.96 | 319.32 | 298.87 | 1,305.94 |
| 432 | 1,434.14 | 1,575.09 | 1,292.67 | 1,447.35 | 5,749.25 |
| 433 | 133.88 | 125.31 | 137.74 | 132.47 | 529.4 |
| 434 | 74.42 | 74.93 | 76.83 | 80.01 | 306.19 |
| 435 | 286.51 | 447.37 | 298.02 | 416.97 | 1,448.87 |
| 436 | 355.95 | 338.59 | 297.59 | 327.85 | 1,319.98 |
| 437 | 199.83 | 196.93 | 179.29 | 176.16 | 752.21 |
| 438 | 31.7 | 30.9 | 27.13 | 34.84 | 124.57 |
| 439 | 116.44 | 128.92 | 115.31 | 124.54 | 485.21 |

| | | | | | |
|---|---|---|---|---|---|
| 440 | 369.61 | 355.63 | 342.42 | 352.55 | 1,420.21 |
| 441 | 1,216.99 | 1,085.21 | 1,125.92 | 1,099.40 | 4,527.52 |
| 442 | 457.16 | 424.16 | 431.18 | 418.71 | 1,731.21 |
| 443 | 364.7 | 357.58 | 412.73 | 357.65 | 1,492.66 |
| 444 | 144.36 | 127.77 | 127.89 | 146.38 | 546.4 |
| 445 | 94.57 | 122.69 | 106.74 | 104.48 | 428.48 |
| 446 | 501.83 | 462.58 | 504.78 | 452.54 | 1,921.73 |
| 447 | 431.24 | 380.93 | 389.31 | 377.53 | 1,579.01 |
| 448 | 225.83 | 222.11 | 214.03 | 222.38 | 884.35 |
| 449 | 48.83 | 46.16 | 43.25 | 53.44 | 191.68 |
| 450 | 783.6 | 741.86 | 624.83 | 626.92 | 2,777.21 |

DRUG CODE:9150DRUG NAME:HYDROMORPHONE
STATE:OHIO

| ZIP CODE | QUARTER 1 | QUARTER 2 | QUARTER 3 | QUARTER 4 | TOTAL GRAMS |
|---|---|---|---|---|---|
| 451 | 165.83 | 164.22 | 168.24 | 163.7 | 661.99 |
| 452 | 653.36 | 902.34 | 646.45 | 633.35 | 2,835.50 |
| 453 | 238.07 | 212.69 | 185.97 | 176.58 | 813.31 |
| 454 | 315.28 | 354.15 | 329.29 | 347.51 | 1,346.23 |
| 455 | 66.77 | 75.85 | 62.3 | 62.14 | 267.06 |
| 456 | 200.35 | 181.3 | 200.39 | 255.79 | 837.83 |
| 457 | 112.25 | 128.79 | 103.94 | 78.22 | 423.2 |
| 458 | 284.94 | 257.36 | 207.84 | 243.8 | 993.94 |
| TOTAL | 10,524.07 | 10,627.33 | 9,930.90 | 10,104.21 | 41,186.51 |

STATE: OKLAHOMA

| ZIP CODE | QUARTER 1 | QUARTER 2 | QUARTER 3 | QUARTER 4 | TOTAL GRAMS |
|---|---|---|---|---|---|
| 730 | 738.87 | 818.55 | 613.75 | 677.41 | 2,848.58 |
| 731 | 1,097.93 | 1,015.20 | 971.82 | 1,011.65 | 4,096.60 |
| 734 | 84.01 | 88.48 | 72.45 | 76.44 | 321.38 |
| 735 | 254.21 | 246.32 | 254.32 | 219.7 | 974.55 |
| 736 | 37.34 | 39.79 | 26.19 | 21.87 | 125.19 |
| 737 | 60.96 | 78.78 | 75.34 | 68.56 | 283.64 |
| 738 | 15.79 | 17.59 | 19.58 | 14.77 | 67.73 |
| 739 | 8.87 | 10.69 | 11.05 | 9.71 | 40.32 |
| 740 | 695.84 | 516.64 | 437.15 | 432.19 | 2,081.82 |
| 741 | 659.65 | 588.39 | 633.92 | 703.08 | 2,585.04 |
| 743 | 91.6 | 93.45 | 83.37 | 78.87 | 347.29 |
| 744 | 164.49 | 166.04 | 178.64 | 174.85 | 684.02 |
| 745 | 85.09 | 106.11 | 105.57 | 115.57 | 412.34 |
| 746 | 17.78 | 19.51 | 30.92 | 23.94 | 92.15 |
| 747 | 132.25 | 158.77 | 153.8 | 166.43 | 611.25 |
| 748 | 128.79 | 120.3 | 113.39 | 124.79 | 487.27 |
| 749 | 92.71 | 100.98 | 93.6 | 96.96 | 384.25 |
| TOTAL | 4,366.18 | 4,185.59 | 3,874.86 | 4,016.79 | 16,443.42 |

STATE: OREGON

| ZIP CODE | QUARTER 1 | QUARTER 2 | QUARTER 3 | QUARTER 4 | TOTAL GRAMS |
|---|---|---|---|---|---|
| 970 | 1,000.10 | 899.39 | 889.68 | 838.01 | 3,627.18 |
| 971 | 390.92 | 368.56 | 382.28 | 366.45 | 1,508.21 |
| 972 | 1,483.15 | 1,603.54 | 1,418.90 | 1,421.85 | 5,927.44 |
| 973 | 835.87 | 806.96 | 834.64 | 826.93 | 3,304.40 |
| 974 | 1,083.35 | 1,005.03 | 946.48 | 913.93 | 3,948.79 |
| 975 | 515.09 | 450.45 | 475.58 | 494.13 | 1,935.25 |
| 976 | 54.8 | 61.36 | 60.94 | 58.61 | 235.71 |
| 977 | 259.58 | 237.8 | 238.46 | 164.44 | 900.28 |
| 978 | 198.9 | 239.81 | 196.69 | 168.87 | 804.27 |
| 979 | 28.5 | 30.21 | 27.29 | 34.83 | 120.83 |
| TOTAL | 5,850.26 | 5,703.11 | 5,470.94 | 5,288.05 | 22,312.36 |

RETAIL DRUG DISTRIBUTION BY ZIP CODE WITHIN STATE BY GRAMS WT

STATE: PENNSYLVANIA

| ZIP CODE | QUARTER 1 | QUARTER 2 | QUARTER 3 | QUARTER 4 | TOTAL GRAMS |
|---|---|---|---|---|---|
| 150 | 335.09 | 358.54 | 360.4 | 332.61 | 1,386.64 |
| 151 | 182.21 | 144.48 | 153.76 | 160.59 | 641.04 |
| 152 | 1,069.23 | 1,152.36 | 1,060.31 | 961.91 | 4,243.81 |
| 153 | 132.01 | 119.62 | 130.91 | 112.36 | 494.9 |
| 154 | 143.86 | 121.12 | 103.13 | 87.24 | 455.35 |
| 155 | 82 | 101.17 | 80.63 | 81.86 | 345.66 |
| 156 | 308.02 | 305.73 | 198.66 | 191.02 | 1,003.43 |
| 157 | 70.64 | 52.22 | 55.17 | 73.92 | 251.95 |
| 158 | 102.05 | 82.83 | 81.63 | 77.51 | 344.02 |
| 159 | 142.76 | 135.82 | 141.61 | 186.44 | 606.63 |
| 160 | 94.09 | 99.8 | 126.32 | 93.89 | 414.1 |
| 161 | 197.59 | 180.94 | 196.58 | 175.66 | 750.77 |
| 162 | 42.11 | 39.48 | 46.58 | 42.18 | 170.35 |
| 163 | 418.9 | 347.19 | 321.9 | 324.79 | 1,412.78 |
| 164 | 107.2 | 102.49 | 98.55 | 84.43 | 392.67 |
| 165 | 148.67 | 115.46 | 118.48 | 147.57 | 530.18 |
| 166 | 120.72 | 93.31 | 101.1 | 123.48 | 438.61 |
| 167 | 28.83 | 28.22 | 24.18 | 25 | 106.23 |
| 168 | 79.04 | 80.18 | 77.88 | 81.85 | 318.95 |
| 169 | 38.94 | 36.48 | 34.78 | 30.99 | 141.19 |
| 170 | 667.12 | 776.91 | 719.86 | 716.24 | 2,880.13 |
| 171 | 230.8 | 212.96 | 211.82 | 179.74 | 835.32 |

DRUG CODE:9150DRUG NAME:HYDROMORPHONE
STATE:PENNSYLVANIA

| ZIP CODE | QUARTER 1 | QUARTER 2 | QUARTER 3 | QUARTER 4 | TOTAL GRAMS |
|---|---|---|---|---|---|
| 172 | 180.48 | 183.03 | 148.43 | 155.99 | 667.93 |
| 173 | 244.74 | 256.22 | 274.04 | 265.84 | 1,040.84 |
| 174 | 160.01 | 209.87 | 184.2 | 151.03 | 705.11 |
| 175 | 227.18 | 236.01 | 255.26 | 250.4 | 968.85 |
| 176 | 415.18 | 336.78 | 315.79 | 318.98 | 1,386.73 |
| 177 | 198.56 | 172.29 | 162.22 | 170.39 | 703.46 |
| 178 | 394.23 | 335.7 | 365.46 | 359.35 | 1,454.74 |
| 179 | 108.95 | 76.82 | 62.77 | 27.06 | 275.6 |
| 180 | 578.86 | 619.36 | 490.96 | 625.58 | 2,314.76 |
| 181 | 338.51 | 340 | 385.34 | 338.38 | 1,402.23 |
| 182 | 69.28 | 69.2 | 66.95 | 77.04 | 282.47 |
| 183 | 175.82 | 185.11 | 207.57 | 189.33 | 757.83 |
| 184 | 213.68 | 192.49 | 175.04 | 198.71 | 779.92 |
| 185 | 268.62 | 219.99 | 274.05 | 365.72 | 1,128.38 |
| 186 | 78.25 | 86.51 | 81.01 | 94.29 | 340.06 |
| 187 | 975.23 | 1,019.60 | 1,024.61 | 852.23 | 3,871.67 |
| 188 | 55.31 | 53.64 | 60.79 | 71.41 | 241.15 |
| 189 | 424.21 | 447.83 | 422.57 | 401.43 | 1,696.04 |
| 190 | 3,666.80 | 3,848.65 | 3,880.78 | 4,435.75 | 15,831.98 |
| 191 | 1,942.24 | 1,843.14 | 1,769.30 | 1,743.26 | 7,297.94 |
| 193 | 665.67 | 643.6 | 644.45 | 479.63 | 2,433.35 |
| 194 | 948.36 | 1,014.97 | 954.89 | 765 | 3,683.22 |
| 195 | 98.06 | 103.26 | 79.18 | 81.95 | 362.45 |
| 196 | 183.87 | 191.53 | 171.31 | 186.19 | 732.9 |
| TOTAL | 17,353.98 | 17,372.91 | 16,901.21 | 16,896.22 | 68,524.32 |

STATE: PUERTO RICO

| ZIP CODE | QUARTER 1 | QUARTER 2 | QUARTER 3 | QUARTER 4 | TOTAL GRAMS |
|---|---|---|---|---|---|
| 6 | 12.41 | 14.54 | 16.31 | 9.22 | 52.48 |
| 7 | 0.18 | 2.84 | 2.48 | 5.14 | 10.64 |
| 9 | 53.75 | 57.27 | 62.22 | 50.57 | 223.81 |
| TOTAL | 66.34 | 74.65 | 81.01 | 64.93 | 286.93 |

RETAIL DRUG DISTRIBUTION BY ZIP CODE WITHIN STATE BY GRAMS WT

STATE: RHODE ISLAND

| ZIP CODE | QUARTER 1 | QUARTER 2 | QUARTER 3 | QUARTER 4 | TOTAL GRAMS |
|---|---|---|---|---|---|
| 28 | 460.23 | 431.79 | 431.27 | 447.96 | 1,771.25 |
| 29 | 385.46 | 449.54 | 457.94 | 476.15 | 1,769.09 |
| TOTAL | 845.69 | 881.33 | 889.21 | 924.11 | 3,540.34 |

STATE: SOUTH CAROLINA

| ZIP CODE | QUARTER 1 | QUARTER 2 | QUARTER 3 | QUARTER 4 | TOTAL GRAMS |
|---|---|---|---|---|---|
| 290 | 372.5 | 378.02 | 409.25 | 439.91 | 1,599.68 |
| 291 | 737.69 | 675.27 | 615.93 | 618.89 | 2,647.78 |
| 292 | 650.89 | 625.46 | 674.24 | 666.01 | 2,616.60 |
| 293 | 306.92 | 304.1 | 296.17 | 306.92 | 1,214.11 |
| 294 | 1,062.38 | 1,005.89 | 1,028.18 | 1,009.03 | 4,105.48 |
| 295 | 1,119.53 | 1,142.53 | 1,141.34 | 1,126.50 | 4,529.90 |
| 296 | 1,061.92 | 1,001.26 | 1,038.52 | 965 | 4,066.70 |
| 297 | 287.12 | 236.23 | 227.99 | 229.63 | 980.97 |
| 298 | 295.4 | 252.45 | 229.12 | 282.13 | 1,059.10 |
| 299 | 419.61 | 503.95 | 407.83 | 402.32 | 1,733.71 |
| TOTAL | 6,313.96 | 6,125.16 | 6,068.57 | 6,046.34 | 24,554.03 |

STATE: SOUTH DAKOTA

| ZIP CODE | QUARTER 1 | QUARTER 2 | QUARTER 3 | QUARTER 4 | TOTAL GRAMS |
|---|---|---|---|---|---|
| 570 | 99.19 | 89.88 | 75.73 | 81.01 | 345.81 |
| 571 | 432.27 | 362.37 | 355.9 | 313.46 | 1,464 |
| 572 | 20.14 | 21.12 | 17.42 | 23.67 | 82.35 |
| 573 | 18.77 | 17.6 | 23.24 | 20.96 | 80.57 |
| 574 | 29.79 | 25.59 | 20.35 | 23.03 | 98.76 |
| 575 | 18.8 | 22.4 | 20.56 | 20.79 | 82.55 |
| 576 | 3.21 | 7.13 | 2.88 | 3.42 | 16.64 |
| 577 | 205.64 | 140.41 | 115.24 | 101.72 | 563.01 |
| TOTAL | 827.81 | 686.5 | 631.32 | 588.06 | 2,733.69 |

STATE: TENNESSEE

| ZIP CODE | QUARTER 1 | QUARTER 2 | QUARTER 3 | QUARTER 4 | TOTAL GRAMS |
|---|---|---|---|---|---|
| 370 | 797.8 | 712.09 | 650.43 | 717.13 | 2,877.45 |
| 371 | 602.48 | 537.45 | 448.86 | 408.98 | 1,997.77 |
| 372 | 764.94 | 812.15 | 1,225.74 | 1,014.02 | 3,816.85 |
| 373 | 700.41 | 602.03 | 512.23 | 533.71 | 2,348.38 |

DRUG CODE:9150DRUG NAME:HYDROMORPHONE
STATE:TENNESSEE

| ZIP CODE | QUARTER 1 | QUARTER 2 | QUARTER 3 | QUARTER 4 | TOTAL GRAMS |
|---|---|---|---|---|---|
| 374 | 380.48 | 409.34 | 378.34 | 390.56 | 1,558.72 |
| 376 | 454.74 | 435.21 | 387.29 | 363.76 | 1,641 |
| 377 | 156.55 | 143.54 | 117.16 | 103.11 | 520.36 |
| 378 | 315.22 | 295.64 | 249.77 | 246.82 | 1,107.45 |
| 379 | 433.28 | 460.22 | 503.4 | 364.16 | 1,761.06 |
| 380 | 202.15 | 165.89 | 186.36 | 194.54 | 748.94 |
| 381 | 2,342.44 | 1,750.83 | 2,015.36 | 2,348.79 | 8,457.42 |
| 382 | 61.38 | 68.16 | 72.37 | 62.38 | 264.29 |
| 383 | 134.31 | 151.03 | 146.42 | 150.8 | 582.56 |
| 384 | 145.84 | 101.97 | 129.51 | 170.92 | 548.24 |
| 385 | 198.69 | 177.59 | 185.36 | 192.49 | 754.13 |
| TOTAL | 7,690.71 | 6,823.14 | 7,208.60 | 7,262.17 | 28,984.62 |

STATE: TEXAS

| ZIP CODE | QUARTER 1 | QUARTER 2 | QUARTER 3 | QUARTER 4 | TOTAL GRAMS |
|---|---|---|---|---|---|
| 750 | 2,588.54 | 2,683.91 | 2,756.03 | 2,718.80 | 10,747.28 |

| | | | | | |
|---|---|---|---|---|---|
| 751 | 508.53 | 504.27 | 505.48 | 536.68 | 2,054.96 |
| 752 | 1,547.14 | 1,449.25 | 1,645.15 | 1,763.19 | 6,404.73 |
| 753 | 109.8 | 99.76 | 108.28 | 96.83 | 414.67 |
| 754 | 581.74 | 590.32 | 573.61 | 597.97 | 2,343.64 |
| 755 | 82.19 | 73.98 | 78.93 | 88.75 | 323.85 |
| 756 | 625.27 | 619.06 | 784.76 | 351.41 | 2,380.50 |
| 757 | 332.17 | 310.73 | 319.45 | 385.88 | 1,348.23 |
| 758 | 48.46 | 43.71 | 58.36 | 32.19 | 182.72 |
| 759 | 247.07 | 242.83 | 213.45 | 195.5 | 898.85 |
| 760 | 1,338.22 | 1,355.97 | 1,283.69 | 1,221.93 | 5,199.81 |
| 761 | 1,417.24 | 1,391.81 | 1,435.20 | 1,239.15 | 5,483.40 |
| 762 | 785.74 | 696.69 | 821.67 | 737.93 | 3,042.03 |
| 763 | 330.65 | 311.66 | 289.05 | 343.93 | 1,275.29 |
| 764 | 126.48 | 124.6 | 130.82 | 124.81 | 506.71 |
| 765 | 450.41 | 487.41 | 437.48 | 471.4 | 1,846.70 |
| 766 | 58.88 | 62.29 | 56.09 | 56.76 | 234.02 |
| 767 | 96.12 | 85.94 | 96.31 | 102.45 | 380.82 |
| 768 | 40.91 | 34.31 | 61.69 | 45.99 | 182.9 |
| 769 | 81 | 83.41 | 76.22 | 74.13 | 314.76 |
| 770 | 2,887.43 | 2,686.01 | 2,977.13 | 3,171.82 | 11,722.39 |
| 773 | 1,074.59 | 1,064.51 | 1,080.05 | 1,092.98 | 4,312.13 |
| 774 | 785.29 | 766.97 | 796.28 | 753.18 | 3,101.72 |
| 775 | 1,284.93 | 1,258.72 | 1,496.75 | 1,315.22 | 5,355.62 |
| 776 | 623.14 | 608.66 | 517.55 | 498.76 | 2,248.11 |
| 777 | 276.74 | 309.93 | 278.5 | 274.08 | 1,139.25 |
| 778 | 104.68 | 120.55 | 111.63 | 123.61 | 460.47 |
| 779 | 432.82 | 411.14 | 414.67 | 422.31 | 1,680.94 |
| 780 | 255.05 | 250.09 | 251.61 | 252.07 | 1,008.82 |
| 781 | 307.41 | 348.53 | 298.69 | 315.28 | 1,269.91 |
| 782 | 2,379.55 | 2,061.46 | 1,985.25 | 2,068.29 | 8,494.55 |
| 783 | 138.63 | 128.41 | 126.21 | 125.66 | 518.91 |
| 784 | 316.81 | 291.85 | 306.38 | 319.68 | 1,234.72 |
| 785 | 423.92 | 330.53 | 362.86 | 370.99 | 1,488.30 |
| 786 | 904.81 | 944.47 | 939.57 | 955.39 | 3,744.24 |
| 787 | 1,276.11 | 1,187.11 | 1,186.01 | 1,224.82 | 4,874.05 |
| 788 | 30.47 | 39.33 | 54.04 | 41.16 | 165 |
| 789 | 51.09 | 48.35 | 45.44 | 48.28 | 193.16 |
| 790 | 50.15 | 57.41 | 48.91 | 62.79 | 219.26 |
| 791 | 171.79 | 141.2 | 154.42 | 175.17 | 642.58 |
| 792 | 2.84 | 5.1 | 4.21 | 4.03 | 16.18 |
| 793 | 89.13 | 85 | 84.98 | 73.67 | 332.78 |
| 794 | 356.46 | 352.39 | 370.52 | 405.57 | 1,484.94 |
| 795 | 40.01 | 49.8 | 41.76 | 44.13 | 175.7 |
| 796 | 103.14 | 122.97 | 76.55 | 88.32 | 390.98 |
| 797 | 253.85 | 253.96 | 227.31 | 207.62 | 942.74 |
| 798 | 21.46 | 11.53 | 13.52 | 10.42 | 56.93 |
| 799 | 529.73 | 468.68 | 468.88 | 478.43 | 1,945.72 |
| TOTAL | 26,568.59 | 25,656.57 | 26,451.40 | 26,109.41 | 104,785.97 |

STATE: UTAH

| ZIP CODE | QUARTER 1 | QUARTER 2 | QUARTER 3 | QUARTER 4 | TOTAL GRAMS |
|---|---|---|---|---|---|
| 840 | 1,103.20 | 1,049.21 | 995.69 | 1,218.16 | 4,366.26 |

DRUG CODE:9150DRUG NAME:HYDROMORPHONE
STATE:UTAH

| ZIP CODE | QUARTER 1 | QUARTER 2 | QUARTER 3 | QUARTER 4 | TOTAL GRAMS |
|---|---|---|---|---|---|
| 841 | 1,011.03 | 1,024.15 | 1,057.24 | 1,055.67 | 4,148.09 |
| 843 | 157.76 | 138.84 | 145.94 | 143.25 | 585.79 |
| 844 | 223.65 | 200.59 | 191.66 | 197.93 | 813.83 |

| | | | | | |
|---|---|---|---|---|---|
| 845 | 38.14 | 30.27 | 36.75 | 27.37 | 132.53 |
| 846 | 203.9 | 226.23 | 196.94 | 227.52 | 854.59 |
| 847 | 293.89 | 259.15 | 236.63 | 272.8 | 1,062.47 |
| TOTAL | 3,031.57 | 2,928.44 | 2,860.85 | 3,142.70 | 11,963.56 |

STATE: VERMONT

| ZIP CODE | QUARTER 1 | QUARTER 2 | QUARTER 3 | QUARTER 4 | TOTAL GRAMS |
|---|---|---|---|---|---|
| 50 | 85.11 | 90.97 | 67.67 | 61.29 | 305.04 |
| 51 | 87.71 | 72.46 | 63.87 | 51.82 | 275.86 |
| 52 | 40.95 | 42.66 | 39.67 | 37.14 | 160.42 |
| 53 | 35.23 | 40.77 | 51.09 | 33.58 | 160.67 |
| 54 | 782.72 | 712.37 | 783.26 | 724.61 | 3,002.96 |
| 56 | 137.35 | 131.26 | 122.72 | 112.24 | 503.57 |
| 57 | 150.77 | 155.21 | 184.38 | 244.66 | 735.02 |
| 58 | 72.93 | 77.74 | 64.73 | 67.42 | 282.82 |
| TOTAL | 1,392.77 | 1,323.44 | 1,377.39 | 1,332.76 | 5,426.36 |

STATE: VIRGIN ISLANDS

| ZIP CODE | QUARTER 1 | QUARTER 2 | QUARTER 3 | QUARTER 4 | TOTAL GRAMS |
|---|---|---|---|---|---|
| 8 | 29.97 | 31.21 | 27.27 | 13.5 | 101.95 |
| TOTAL | 29.97 | 31.21 | 27.27 | 13.5 | 101.95 |

STATE: VIRGINIA

| ZIP CODE | QUARTER 1 | QUARTER 2 | QUARTER 3 | QUARTER 4 | TOTAL GRAMS |
|---|---|---|---|---|---|
| 201 | 1,176.85 | 1,221.67 | 1,226.27 | 1,225.25 | 4,850.04 |
| 220 | 830.68 | 829.73 | 730.75 | 779.05 | 3,170.21 |
| 221 | 505.72 | 493.94 | 479.33 | 484.04 | 1,963.03 |
| 222 | 202.13 | 241.61 | 213.32 | 183.4 | 840.46 |
| 223 | 282.87 | 263.8 | 239.35 | 281.29 | 1,067.31 |
| 224 | 518.34 | 454.42 | 459.34 | 433.95 | 1,866.05 |
| 225 | 175.69 | 143.28 | 157.4 | 138.75 | 615.12 |
| 226 | 610.38 | 578.18 | 581.22 | 582.16 | 2,351.94 |
| 227 | 83.06 | 71.68 | 73.74 | 82.13 | 310.61 |
| 228 | 379.15 | 328.13 | 346.15 | 335.79 | 1,389.22 |
| 229 | 727.66 | 685.5 | 641.52 | 640.89 | 2,695.57 |
| 230 | 451.06 | 397.1 | 461.51 | 434.08 | 1,743.75 |
| 231 | 718.19 | 666.21 | 715.06 | 615.99 | 2,715.45 |
| 232 | 1,529.91 | 1,464.82 | 1,387.46 | 1,444.71 | 5,826.90 |
| 233 | 568.25 | 495.63 | 587.99 | 647.83 | 2,299.70 |
| 234 | 1,128.50 | 1,097.07 | 1,098.23 | 1,111.80 | 4,435.60 |
| 235 | 682.13 | 728.82 | 668.31 | 653.85 | 2,733.11 |
| 236 | 770.74 | 658.73 | 743.89 | 622.78 | 2,796.14 |
| 237 | 301.27 | 294.93 | 310.74 | 298.32 | 1,205.26 |
| 238 | 417.91 | 353.92 | 377.03 | 360.69 | 1,509.55 |
| 239 | 100.17 | 96.79 | 76.24 | 82.36 | 355.56 |
| 240 | 616.85 | 542.44 | 597.22 | 492.31 | 2,248.82 |
| 241 | 446.55 | 402.01 | 401.97 | 386.3 | 1,636.83 |
| 242 | 185.64 | 199.88 | 205.97 | 159.22 | 750.71 |
| 243 | 134.79 | 138.08 | 151.28 | 144.89 | 569.04 |
| 244 | 216.18 | 210.72 | 205.64 | 188.18 | 820.72 |
| 245 | 454.14 | 387.33 | 375.68 | 331.51 | 1,548.66 |
| 246 | 67.87 | 66.19 | 64.62 | 53.94 | 252.62 |
| TOTAL | 14,282.68 | 13,512.61 | 13,577.23 | 13,195.46 | 54,567.98 |

STATE: WASHINGTON

| ZIP CODE | QUARTER 1 | QUARTER 2 | QUARTER 3 | QUARTER 4 | TOTAL GRAMS |
|---|---|---|---|---|---|
| 980 | 2,176.72 | 2,243.17 | 2,083.73 | 1,855.31 | 8,358.93 |
| 981 | 2,037.67 | 2,058.90 | 1,955.51 | 1,874.24 | 7,926.32 |
| 982 | 1,728.36 | 1,770.37 | 1,616.61 | 1,567.80 | 6,683.14 |
| 983 | 997.57 | 1,022.45 | 988.54 | 927.39 | 3,935.95 |

| | | | | | |
|---|---|---|---|---|---|
| 984 | 649.04 | 711.59 | 646.66 | 549.58 | 2,556.87 |
| 985 | 1,300.19 | 1,241.66 | 1,071 | 864.63 | 4,477.48 |
| 986 | 1,207.51 | 1,194.54 | 1,209.28 | 1,218.04 | 4,829.37 |
| 988 | 232.47 | 271.93 | 205.19 | 187.84 | 897.43 |
| 989 | 347.08 | 401.54 | 475.14 | 346.51 | 1,570.27 |
| 990 | 94.46 | 88.8 | 95.43 | 74.05 | 352.74 |
| 991 | 140.75 | 144.81 | 137.18 | 137.87 | 560.61 |

DRUG CODE:9150DRUG NAME:HYDROMORPHONE
STATE:WASHINGTON

| ZIP CODE | QUARTER 1 | QUARTER 2 | QUARTER 3 | QUARTER 4 | TOTAL GRAMS |
|---|---|---|---|---|---|
| 992 | 916.73 | 820.35 | 917.34 | 931.59 | 3,586.01 |
| 993 | 773.55 | 708.61 | 695.37 | 644.85 | 2,822.38 |
| 994 | 117.92 | 182.97 | 80.09 | 82.28 | 463.26 |
| TOTAL | 12,720.02 | 12,861.69 | 12,177.07 | 11,261.98 | 49,020.76 |

STATE: WEST VIRGINIA

| ZIP CODE | QUARTER 1 | QUARTER 2 | QUARTER 3 | QUARTER 4 | TOTAL GRAMS |
|---|---|---|---|---|---|
| 247 | 42.29 | 45.49 | 34.83 | 49.31 | 171.92 |
| 248 | 18.97 | 13.65 | 5.14 | 4.79 | 42.55 |
| 249 | 43.75 | 37.11 | 36.03 | 38.16 | 155.05 |
| 250 | 13.96 | 20.39 | 9.78 | 1.6 | 45.73 |
| 251 | 16.96 | 101.56 | 9.69 | 10.02 | 138.23 |
| 252 | 11.22 | 5.63 | 6.44 | 6.28 | 29.57 |
| 253 | 135.1 | 148.59 | 200.86 | 199.48 | 684.03 |
| 254 | 439.41 | 432.2 | 414.26 | 457.49 | 1,743.36 |
| 255 | 65.34 | 67.59 | 52.5 | 57.23 | 242.66 |
| 256 | 13.11 | 22.75 | 20.58 | 20.63 | 77.07 |
| 257 | 90.96 | 122.47 | 88.89 | 133.57 | 435.89 |
| 258 | 155.63 | 170.16 | 164.87 | 127.52 | 618.18 |
| 259 | 15.96 | 18.62 | 21.46 | 16.49 | 72.53 |
| 260 | 113.47 | 118.47 | 109.49 | 97.87 | 439.3 |
| 261 | 73.12 | 58.27 | 46.67 | 73.21 | 251.27 |
| 262 | 28.07 | 27.34 | 24.08 | 23.4 | 102.89 |
| 263 | 104.57 | 127.24 | 101.59 | 104.98 | 438.38 |
| 264 | 24.48 | 19.7 | 14.92 | 20.32 | 79.42 |
| 265 | 284.68 | 358.34 | 249.67 | 215.48 | 1,108.17 |
| 266 | 22.62 | 17.5 | 13.52 | 14.65 | 68.29 |
| 267 | 55.53 | 58.21 | 54.05 | 55.9 | 223.69 |
| 268 | 30.61 | 28.46 | 29.29 | 32.72 | 121.08 |
| TOTAL | 1,799.81 | 2,019.74 | 1,708.61 | 1,761.10 | 7,289.26 |

STATE: WISCONSIN

| ZIP CODE | QUARTER 1 | QUARTER 2 | QUARTER 3 | QUARTER 4 | TOTAL GRAMS |
|---|---|---|---|---|---|
| 530 | 293.83 | 312.58 | 282.29 | 298.02 | 1,186.72 |
| 531 | 797.5 | 631.54 | 556.48 | 584.62 | 2,570.14 |
| 532 | 773.88 | 874.74 | 638.62 | 764.85 | 3,052.09 |
| 534 | 104.17 | 84.95 | 70.25 | 76.98 | 336.35 |
| 535 | 341.29 | 350.26 | 373.76 | 304.44 | 1,369.75 |
| 537 | 364.5 | 342.13 | 298.66 | 319.8 | 1,325.09 |
| 538 | 48.35 | 32.2 | 47.19 | 34.89 | 162.63 |
| 539 | 91.78 | 87.24 | 77.39 | 65.85 | 322.26 |
| 540 | 88.2 | 94.12 | 87.16 | 86.13 | 355.61 |
| 541 | 55.42 | 51.29 | 54.84 | 51.58 | 213.13 |
| 542 | 32.09 | 31.02 | 32.64 | 29.52 | 125.27 |
| 543 | 143.74 | 129.83 | 139.25 | 175.87 | 588.69 |
| 544 | 206.23 | 296.16 | 240.24 | 230.98 | 973.61 |
| 545 | 64.3 | 53.52 | 65.56 | 51.77 | 235.15 |
| 546 | 209.11 | 208.59 | 231.59 | 310.09 | 959.38 |

| | | | | | |
|---|---|---|---|---|---|
| 547 | 217.31 | 237.81 | 258.9 | 243.08 | 957.1 |
| 548 | 113.19 | 122.36 | 130.72 | 150.42 | 516.69 |
| 549 | 428.93 | 345.47 | 406.91 | 420.08 | 1,601.39 |
| TOTAL | 4,373.82 | 4,285.81 | 3,992.45 | 4,198.97 | 16,851.05 |

STATE: WYOMING

| ZIP CODE | QUARTER 1 | QUARTER 2 | QUARTER 3 | QUARTER 4 | TOTAL GRAMS |
|---|---|---|---|---|---|
| 820 | 185.17 | 179.97 | 198.47 | 151.24 | 714.85 |
| 822 | 6.08 | 3.9 | 4.72 | 11.99 | 26.69 |
| 823 | 9.49 | 9.5 | 8.32 | 7.47 | 34.78 |
| 824 | 126.7 | 137.24 | 130.47 | 123.7 | 518.11 |
| 825 | 40.09 | 36.25 | 45.06 | 46.82 | 168.22 |
| 826 | 110.27 | 132.96 | 119.66 | 113.25 | 476.14 |
| 827 | 60.64 | 44.2 | 49.66 | 91.89 | 246.39 |
| 828 | 44.69 | 39.77 | 39.82 | 41.86 | 166.14 |
| 829 | 129.48 | 117.23 | 106.07 | 95.03 | 447.81 |
| 830 | 3.98 | 7.37 | 5.78 | 9.52 | 26.65 |
| 831 | 18.1 | 26 | 23.4 | 24.2 | 91.7 |
| TOTAL | 734.69 | 734.39 | 731.43 | 716.97 | 2,917.48 |

DRUG CODE:9193DRUG NAME:HYDROCODONE

DRUG CODE:9193DRUG NAME:HYDROCODONE
STATE: ALABAMA

| ZIP CODE | QUARTER 1 | QUARTER 2 | QUARTER 3 | QUARTER 4 | TOTAL GRAMS |
|---|---|---|---|---|---|
| 350 | 32,785.45 | 31,175.26 | 31,327.32 | 30,341.91 | 125,629.94 |
| 351 | 16,218.41 | 15,315.98 | 15,481.92 | 15,324.67 | 62,340.98 |
| 352 | 29,034.93 | 27,051.69 | 27,484.66 | 27,158.86 | 110,730.14 |
| 354 | 12,469.83 | 12,333.52 | 11,925.44 | 12,037.39 | 48,766.18 |
| 355 | 13,609.37 | 13,191.41 | 12,850.31 | 13,114.76 | 52,765.85 |
| 356 | 26,809.15 | 26,514.71 | 27,002.85 | 25,680.80 | 106,007.51 |
| 357 | 10,746.98 | 10,405.24 | 10,441.26 | 10,287.23 | 41,880.71 |
| 358 | 12,421.50 | 12,058.29 | 12,229.12 | 11,960.39 | 48,669.30 |
| 359 | 22,994.71 | 22,342.83 | 21,617.73 | 21,836.44 | 88,791.71 |
| 360 | 14,175.24 | 13,627.07 | 13,306.61 | 13,024.24 | 54,133.16 |
| 361 | 10,703.24 | 10,317.52 | 10,885.02 | 10,169.14 | 42,074.92 |
| 362 | 10,631.31 | 10,536.36 | 10,242.09 | 10,429.60 | 41,839.36 |
| 363 | 10,612.09 | 10,252.06 | 10,451.06 | 10,106.02 | 41,421.23 |
| 364 | 6,675.69 | 6,508 | 6,404.05 | 6,448.48 | 26,036.22 |
| 365 | 22,873.96 | 22,011.62 | 21,963 | 21,124.72 | 87,973.30 |
| 366 | 18,631.50 | 17,622.15 | 17,415.67 | 16,396.45 | 70,065.77 |
| 367 | 3,971.94 | 3,762.95 | 3,730.50 | 3,759.68 | 15,225.07 |
| 368 | 8,300.87 | 7,722.70 | 7,563.93 | 7,721.31 | 31,308.81 |
| 369 | 1,273.50 | 1,198.19 | 1,180.65 | 1,137.50 | 4,789.84 |
| TOTAL | 284,939.67 | 273,947.55 | 273,503.19 | 268,059.59 | 1,100,450 |

STATE: ALASKA

| ZIP CODE | QUARTER 1 | QUARTER 2 | QUARTER 3 | QUARTER 4 | TOTAL GRAMS |
|---|---|---|---|---|---|
| 995 | 6,524.65 | 6,137.51 | 6,047.17 | 5,843.86 | 24,553.19 |
| 996 | 4,401.01 | 4,251.54 | 4,196.81 | 4,002.41 | 16,851.77 |
| 997 | 1,458.91 | 1,428.02 | 1,399.32 | 1,266.49 | 5,552.74 |
| 998 | 927.2 | 833.16 | 788.28 | 732.37 | 3,281.01 |
| 999 | 668.33 | 684.31 | 634.47 | 605.97 | 2,593.08 |
| TOTAL | 13,980.10 | 13,334.54 | 13,066.05 | 12,451.10 | 52,831.79 |

STATE: AMERICAN SAMOA

| ZIP CODE | QUARTER 1 | QUARTER 2 | QUARTER 3 | QUARTER 4 | TOTAL GRAMS |
|---|---|---|---|---|---|
| 967 | 7.26 | 14.53 | 7.26 | 3.63 | 32.68 |

| | | | | | |
|---|---|---|---|---|---|
| TOTAL | 7.26 | 14.53 | 7.26 | 3.63 | 32.68 |

STATE: ARIZONA

| ZIP CODE | QUARTER 1 | QUARTER 2 | QUARTER 3 | QUARTER 4 | TOTAL GRAMS |
|---|---|---|---|---|---|
| 850 | 20,073.21 | 18,355.91 | 17,832.37 | 17,199.81 | 73,461.30 |
| 851 | 6,402.42 | 5,489.38 | 5,473.99 | 5,605.68 | 22,971.47 |
| 852 | 31,289.03 | 28,575.69 | 27,841.20 | 28,491.87 | 116,197.79 |
| 853 | 21,936.90 | 19,381.92 | 18,939.14 | 20,158.11 | 80,416.07 |
| 855 | 2,357.79 | 2,365.65 | 2,288.60 | 2,180.87 | 9,192.91 |
| 856 | 4,571.23 | 4,183.73 | 4,053.98 | 4,042.69 | 16,851.63 |
| 857 | 15,851.42 | 14,664.11 | 14,364.24 | 14,279.43 | 59,159.20 |
| 859 | 2,499.60 | 2,327.47 | 2,810.97 | 2,466.48 | 10,104.52 |
| 860 | 2,627.28 | 2,514.22 | 2,529.23 | 2,495.39 | 10,166.12 |
| 863 | 8,023.99 | 7,806.06 | 7,552.58 | 7,598.69 | 30,981.32 |
| 864 | 9,867.17 | 9,318.08 | 9,304.07 | 9,649.58 | 38,138.90 |
| 865 | 171.61 | 164.65 | 130.88 | 124.64 | 591.78 |
| TOTAL | 125,671.65 | 115,146.87 | 113,121.25 | 114,293.24 | 468,233.01 |

STATE: ARKANSAS

| ZIP CODE | QUARTER 1 | QUARTER 2 | QUARTER 3 | QUARTER 4 | TOTAL GRAMS |
|---|---|---|---|---|---|
| 716 | 6,812.89 | 6,533.76 | 6,452.72 | 6,481.02 | 26,280.39 |
| 717 | 5,830.20 | 5,676.46 | 5,702.18 | 5,513.51 | 22,722.35 |
| 718 | 8,058.16 | 7,458.83 | 7,841.40 | 7,558.54 | 30,916.93 |
| 719 | 11,986.86 | 11,559.86 | 11,268.64 | 11,170.37 | 45,985.73 |
| 720 | 13,696 | 13,293.20 | 12,889.56 | 13,359.55 | 53,238.31 |
| 721 | 17,502.31 | 17,076.85 | 16,625.56 | 16,647.42 | 67,852.14 |
| 722 | 8,652.17 | 8,342.68 | 8,289 | 8,434.42 | 33,718.27 |
| 723 | 8,035.50 | 7,708.84 | 8,105.23 | 8,087.12 | 31,936.69 |
| 724 | 12,503.36 | 12,415.90 | 12,697.46 | 13,355.63 | 50,972.35 |
| 725 | 7,098.17 | 6,876.17 | 6,975.29 | 7,095.03 | 28,044.66 |
| 726 | 7,184.43 | 7,085.69 | 7,097.63 | 7,054.89 | 28,422.64 |
| 727 | 19,210.49 | 18,273.74 | 18,539.05 | 18,134.51 | 74,157.79 |
| 728 | 5,466.63 | 5,348.91 | 4,959.46 | 4,916.94 | 20,691.94 |
| 729 | 15,223.77 | 14,813.14 | 13,976.93 | 13,796.66 | 57,810.50 |
| TOTAL | 147,260.94 | 142,464.03 | 141,420.11 | 141,605.61 | 572,750.69 |

STATE: CALIFORNIA

| ZIP CODE | QUARTER 1 | QUARTER 2 | QUARTER 3 | QUARTER 4 | TOTAL GRAMS |
|---|---|---|---|---|---|

DRUG CODE:9193DRUG NAME:HYDROCODONE
STATE:CALIFORNIA

| ZIP CODE | QUARTER 1 | QUARTER 2 | QUARTER 3 | QUARTER 4 | TOTAL GRAMS |
|---|---|---|---|---|---|
| 900 | 24,246.02 | 23,104.64 | 22,327.19 | 21,900.08 | 91,577.93 |
| 902 | 19,507.14 | 18,481.67 | 17,918.93 | 17,862.64 | 73,770.38 |
| 903 | 2,067.36 | 1,867.27 | 2,145.51 | 1,978.77 | 8,058.91 |
| 904 | 2,272.79 | 2,086.71 | 2,002.40 | 2,139.65 | 8,501.55 |
| 905 | 5,174.98 | 4,784.01 | 4,723.89 | 4,688.63 | 19,371.51 |
| 906 | 14,579.78 | 13,933.80 | 13,748.22 | 13,461.49 | 55,723.29 |
| 907 | 13,802.62 | 13,258.17 | 13,241.54 | 12,865.21 | 53,167.54 |
| 908 | 9,666.64 | 9,286.26 | 9,302.18 | 8,927.56 | 37,182.64 |
| 910 | 6,963.78 | 6,350.15 | 5,482.09 | 5,492.75 | 24,288.77 |
| 911 | 2,952.69 | 2,850.90 | 2,851.59 | 2,739.44 | 11,394.62 |
| 912 | 3,031.89 | 3,018.72 | 3,212.47 | 3,352.04 | 12,615.12 |
| 913 | 27,680.37 | 27,053.12 | 26,284.17 | 26,311.33 | 107,328.99 |
| 914 | 7,204.09 | 6,992.34 | 6,575.63 | 6,654.20 | 27,426.26 |
| 915 | 2,387.97 | 2,388.40 | 2,347.82 | 2,385.44 | 9,509.63 |
| 916 | 1,989.47 | 1,939.17 | 1,847.48 | 1,749.30 | 7,525.42 |
| 917 | 36,117.59 | 35,197.78 | 35,823.51 | 35,521.03 | 142,659.91 |
| 918 | 1,186.50 | 1,076.69 | 1,025.84 | 1,045.10 | 4,334.13 |
| 919 | 10,097.75 | 9,663.54 | 9,880.47 | 9,532.41 | 39,174.17 |

| | | | | | |
|---|---|---|---|---|---|
| 920 | 34,170.63 | 33,172.71 | 32,328.45 | 31,736.09 | 131,407.88 |
| 921 | 26,295.10 | 24,511.28 | 23,780.43 | 23,063.73 | 97,650.54 |
| 922 | 23,855.91 | 22,577.92 | 21,890.07 | 22,315.37 | 90,639.27 |
| 923 | 53,324.31 | 50,717.05 | 49,874.47 | 49,841.92 | 203,757.75 |
| 924 | 9,034.78 | 8,453.61 | 7,714.87 | 8,680.08 | 33,883.34 |
| 925 | 45,385.16 | 44,006.65 | 44,137.07 | 42,852.40 | 176,381.28 |
| 926 | 28,440.14 | 27,205.23 | 26,082.70 | 26,110.27 | 107,838.34 |
| 927 | 8,167.45 | 8,030.39 | 7,687.69 | 7,549.07 | 31,434.60 |
| 928 | 28,866.37 | 27,787.05 | 27,353.90 | 26,859.10 | 110,866.42 |
| 930 | 19,016.95 | 18,607.98 | 18,437.57 | 18,500.01 | 74,562.51 |
| 931 | 3,621.63 | 3,612.67 | 3,447.99 | 3,343.59 | 14,025.88 |
| 932 | 27,443.50 | 26,429.01 | 26,182.10 | 25,510.22 | 105,564.83 |
| 933 | 22,496.69 | 21,338.06 | 20,643.33 | 21,109.11 | 85,587.19 |
| 934 | 17,754.81 | 17,149.61 | 16,874.15 | 16,478.21 | 68,256.78 |
| 935 | 19,500.66 | 18,419.88 | 18,478.58 | 18,785.47 | 75,184.59 |
| 936 | 21,071.61 | 20,490.18 | 19,685.12 | 19,507.63 | 80,754.54 |
| 937 | 21,472.92 | 21,030.02 | 20,743.39 | 20,534.48 | 83,780.81 |
| 939 | 10,482 | 10,044.39 | 9,671.42 | 9,408.74 | 39,606.55 |
| 940 | 12,218.97 | 11,119.08 | 11,252.03 | 10,912.17 | 45,502.25 |
| 941 | 11,479.02 | 10,051.80 | 10,551.79 | 10,089.53 | 42,172.14 |
| 943 | 2,560.67 | 2,434.57 | 2,482 | 2,285.50 | 9,762.74 |
| 944 | 1,947.92 | 1,914.24 | 1,813.56 | 1,771.48 | 7,447.20 |
| 945 | 73,654.03 | 71,834.17 | 70,367.71 | 68,320.43 | 284,176.34 |
| 946 | 11,579.27 | 11,052.95 | 10,468.90 | 10,280.44 | 43,381.56 |
| 947 | 1,736.36 | 1,525.25 | 1,546.68 | 1,542.87 | 6,351.16 |
| 948 | 4,241.60 | 3,622.47 | 3,600.53 | 3,659.83 | 15,124.43 |
| 949 | 10,255.82 | 9,783.28 | 9,414.14 | 9,323.22 | 38,776.46 |
| 950 | 16,551.56 | 15,752.77 | 15,675.14 | 15,166.76 | 63,146.23 |
| 951 | 10,655.31 | 10,332.90 | 10,284.17 | 9,847.48 | 41,119.86 |
| 952 | 22,672.93 | 21,630.30 | 21,160.60 | 20,589.47 | 86,053.30 |
| 953 | 57,127.73 | 54,661.58 | 53,361.70 | 53,460.60 | 218,611.61 |
| 954 | 21,739.22 | 20,839.92 | 20,622.07 | 19,616.38 | 82,817.59 |
| 955 | 10,351.91 | 10,015.56 | 9,360.23 | 9,150.81 | 38,878.51 |
| 956 | 63,019.22 | 61,305.39 | 59,774.75 | 58,042.80 | 242,142.16 |
| 957 | 10,986 | 10,358.70 | 10,750.63 | 10,846.76 | 42,942.09 |
| 958 | 39,995.34 | 38,309.61 | 37,113.37 | 36,219.80 | 151,638.12 |
| 959 | 38,539.20 | 36,686.90 | 36,671.33 | 35,860.22 | 147,757.65 |
| 960 | 26,620.60 | 25,609.83 | 24,022.98 | 24,096 | 100,349.41 |
| 961 | 4,351.03 | 3,976.09 | 3,923.65 | 3,537.56 | 15,788.33 |
| TOTAL | 1,063,613.76 | 1,019,734.39 | 999,972.19 | 985,412.67 | 4,068,733.01 |

STATE: COLORADO

| ZIP CODE | QUARTER 1 | QUARTER 2 | QUARTER 3 | QUARTER 4 | TOTAL GRAMS |
|---|---|---|---|---|---|
| 800 | 11,114.52 | 10,459.60 | 10,462.20 | 10,119.76 | 42,156.08 |
| 801 | 8,612.66 | 7,993.11 | 7,785.62 | 8,193 | 32,584.39 |
| 802 | 12,578.96 | 11,688.10 | 11,495.71 | 11,359.82 | 47,122.59 |
| 803 | 1,132.81 | 1,044.98 | 1,024.46 | 1,060.31 | 4,262.56 |
| 804 | 2,578.19 | 2,396.34 | 2,490.58 | 2,396.32 | 9,861.43 |
| 805 | 7,876.72 | 7,309.62 | 7,525.17 | 7,390.66 | 30,102.17 |

DRUG CODE:9193DRUG NAME:HYDROCODONE
STATE:COLORADO

| ZIP CODE | QUARTER 1 | QUARTER 2 | QUARTER 3 | QUARTER 4 | TOTAL GRAMS |
|---|---|---|---|---|---|
| 806 | 3,342.56 | 3,172.10 | 3,178.55 | 3,042.58 | 12,735.79 |
| 807 | 1,118.31 | 1,026.52 | 1,001.42 | 981.53 | 4,127.78 |
| 808 | 1,993.51 | 1,846.22 | 1,882.94 | 1,857.75 | 7,580.42 |
| 809 | 8,913.40 | 8,470.55 | 8,477.29 | 8,259.13 | 34,120.37 |
| 810 | 6,471.66 | 6,071.74 | 6,122.79 | 6,012.38 | 24,678.57 |
| 811 | 970.67 | 935.23 | 968.8 | 912.34 | 3,787.04 |
| 812 | 1,690.32 | 1,610.48 | 1,641.10 | 1,513.40 | 6,455.30 |

| | | | | | |
|---|---|---|---|---|---|
| 813 | 1,529.98 | 1,339.41 | 1,436.51 | 1,347.15 | 5,653.05 |
| 814 | 1,403.69 | 1,358.99 | 1,292.51 | 1,232.37 | 5,287.56 |
| 815 | 3,184.23 | 2,932.70 | 3,002.15 | 2,968.29 | 12,087.37 |
| 816 | 1,893.76 | 1,690.94 | 1,666.40 | 1,699.83 | 6,950.93 |
| TOTAL | 76,405.95 | 71,346.63 | 71,454.20 | 70,346.62 | 289,553.40 |

STATE: CONNECTICUT

| ZIP CODE | QUARTER 1 | QUARTER 2 | QUARTER 3 | QUARTER 4 | TOTAL GRAMS |
|---|---|---|---|---|---|
| 60 | 5,758.46 | 5,330.73 | 5,021.49 | 5,010.82 | 21,121.50 |
| 61 | 1,570.15 | 1,390.02 | 1,350.78 | 1,413.28 | 5,724.23 |
| 62 | 1,363.23 | 1,228 | 1,214.46 | 1,140.47 | 4,946.16 |
| 63 | 2,940.42 | 2,787.50 | 2,623.35 | 2,589.68 | 10,940.95 |
| 64 | 6,029.94 | 5,652.40 | 5,438.23 | 5,275.90 | 22,396.47 |
| 65 | 1,668.18 | 1,540.76 | 1,520.85 | 1,387.75 | 6,117.54 |
| 66 | 1,065.41 | 1,009.70 | 993.3 | 1,008.41 | 4,076.82 |
| 67 | 3,230.37 | 3,073.29 | 2,963.32 | 2,871.25 | 12,138.23 |
| 68 | 2,201.75 | 1,911.91 | 1,898.92 | 1,830.27 | 7,842.85 |
| 69 | 414.04 | 318.13 | 299.74 | 327.1 | 1,359.01 |
| TOTAL | 26,241.95 | 24,242.44 | 23,324.44 | 22,854.93 | 96,663.76 |

STATE: DELAWARE

| ZIP CODE | QUARTER 1 | QUARTER 2 | QUARTER 3 | QUARTER 4 | TOTAL GRAMS |
|---|---|---|---|---|---|
| 197 | 2,139.62 | 2,038.28 | 1,959.57 | 1,936.99 | 8,074.46 |
| 198 | 1,499.01 | 1,356.92 | 1,341.86 | 1,337.48 | 5,535.27 |
| 199 | 3,679.18 | 3,489.25 | 3,633.32 | 3,478.93 | 14,280.68 |
| TOTAL | 7,317.81 | 6,884.45 | 6,934.75 | 6,753.40 | 27,890.41 |

STATE: DISTRICT OF COLUMBIA

| ZIP CODE | QUARTER 1 | QUARTER 2 | QUARTER 3 | QUARTER 4 | TOTAL GRAMS |
|---|---|---|---|---|---|
| 200 | 1,633.84 | 1,683.45 | 1,550.38 | 1,474.04 | 6,341.71 |
| 203 | 7.57 | 7.26 | 6.96 | 6.51 | 28.3 |
| 204 | 137.12 | 137.12 | 128.95 | 99.29 | 502.48 |
| 205 | 0 | 3.33 | 0 | 0 | 3.33 |
| TOTAL | 1,778.53 | 1,831.16 | 1,686.29 | 1,579.84 | 6,875.82 |

STATE: FLORIDA

| ZIP CODE | QUARTER 1 | QUARTER 2 | QUARTER 3 | QUARTER 4 | TOTAL GRAMS |
|---|---|---|---|---|---|
| 320 | 21,040.66 | 20,524.97 | 20,453.77 | 20,402.08 | 82,421.48 |
| 321 | 22,898.32 | 22,045.90 | 21,508.92 | 21,998.25 | 88,451.39 |
| 322 | 40,649.71 | 37,294.63 | 39,038.68 | 37,110.08 | 154,093.10 |
| 323 | 8,462.44 | 8,066.88 | 7,840.56 | 8,203.67 | 32,573.55 |
| 324 | 18,573.50 | 18,253.47 | 18,375.49 | 18,029.92 | 73,232.38 |
| 325 | 30,061.70 | 28,848.79 | 28,912.52 | 28,840.22 | 116,663.23 |
| 326 | 8,270.09 | 7,904.12 | 8,034.31 | 7,073.11 | 31,281.63 |
| 327 | 22,184.30 | 20,490.68 | 20,368.76 | 21,003.19 | 84,046.93 |
| 328 | 10,467.11 | 9,828.77 | 9,539.26 | 9,653.98 | 39,489.12 |
| 329 | 18,343.17 | 17,210.28 | 16,835.45 | 17,029.36 | 69,418.26 |
| 330 | 11,897.63 | 10,881.87 | 10,297.77 | 10,513.43 | 43,590.70 |
| 331 | 6,582.10 | 5,706.42 | 5,507 | 5,390.73 | 23,186.25 |
| 333 | 7,751.30 | 7,323.12 | 7,089.52 | 7,066.46 | 29,230.40 |
| 334 | 17,150.82 | 15,685.73 | 14,801.89 | 15,350.68 | 62,989.12 |
| 335 | 14,543.46 | 13,769.81 | 13,227.31 | 13,906.90 | 55,447.48 |
| 336 | 15,117.27 | 13,990.09 | 14,097.44 | 14,133.98 | 57,338.78 |
| 337 | 23,581.52 | 21,789.96 | 20,833.88 | 20,496.90 | 86,702.26 |
| 338 | 14,842.10 | 13,926.23 | 13,595.82 | 13,672.36 | 56,036.51 |
| 339 | 17,339.03 | 15,498.79 | 14,954.28 | 15,523.55 | 63,315.65 |
| 341 | 5,627.91 | 4,421.23 | 4,010.03 | 4,460.41 | 18,519.58 |
| 342 | 20,986.40 | 18,896.24 | 18,183.60 | 18,911.70 | 76,977.94 |
| 344 | 15,033.60 | 14,386.25 | 14,179.82 | 14,560.65 | 58,160.32 |
| 346 | 22,862.56 | 21,582.98 | 21,136.58 | 21,356.40 | 86,938.52 |
| 347 | 9,896.27 | 9,301.32 | 9,238.56 | 9,213.47 | 37,649.62 |
| 349 | 8,142.65 | 7,572.25 | 7,324.11 | 7,693.53 | 30,732.54 |

| | | | | | |
|---|---|---|---|---|---|
| TOTAL | 412,305.62 | 385,200.78 | 379,385.33 | 381,595.01 | 1,558,486.74 |

DRUG CODE:9193DRUG NAME:HYDROCODONE
STATE: GEORGIA

| ZIP CODE | QUARTER 1 | QUARTER 2 | QUARTER 3 | QUARTER 4 | TOTAL GRAMS |
|---|---|---|---|---|---|
| 300 | 36,754.60 | 34,540.25 | 33,873.39 | 34,453.30 | 139,621.54 |
| 301 | 38,711.28 | 37,096.49 | 36,806.24 | 36,741.76 | 149,355.77 |
| 302 | 22,941.98 | 21,868.72 | 21,366.11 | 21,501.44 | 87,678.25 |
| 303 | 9,129.39 | 8,391.82 | 8,220.52 | 8,419.76 | 34,161.49 |
| 304 | 7,615.98 | 7,344.96 | 7,427.61 | 7,190.53 | 29,579.08 |
| 305 | 23,101.94 | 22,657.34 | 22,601.85 | 22,537.24 | 90,898.37 |
| 306 | 16,860.43 | 15,926.30 | 15,948.36 | 15,715.96 | 64,451.05 |
| 307 | 14,577.80 | 14,083.67 | 14,221.34 | 14,136.44 | 57,019.25 |
| 308 | 3,891.48 | 3,783.42 | 3,437.73 | 3,491.75 | 14,604.38 |
| 309 | 8,020.89 | 7,558.57 | 7,425.29 | 7,001.05 | 30,005.80 |
| 310 | 25,588.05 | 23,868.14 | 24,192.51 | 23,693.71 | 97,342.41 |
| 312 | 9,145.16 | 8,589.08 | 8,632.11 | 8,681.43 | 35,047.78 |
| 313 | 5,728.73 | 5,563.65 | 5,671.18 | 5,469 | 22,432.56 |
| 314 | 6,258.41 | 5,707.38 | 5,555.80 | 5,564.18 | 23,085.77 |
| 315 | 17,956.55 | 17,571.82 | 17,851.18 | 17,521.85 | 70,901.40 |
| 316 | 9,261.04 | 9,263.29 | 9,059.17 | 9,165.43 | 36,748.93 |
| 317 | 12,859.03 | 12,416.15 | 12,385.88 | 12,298.62 | 49,959.68 |
| 318 | 1,382.06 | 1,340.52 | 1,342.49 | 1,320.62 | 5,385.69 |
| 319 | 7,226.56 | 7,038.70 | 6,997.52 | 6,610.37 | 27,873.15 |
| 398 | 3,744.19 | 3,700.65 | 3,673.70 | 3,733.12 | 14,851.66 |
| TOTAL | 280,755.55 | 268,310.92 | 266,689.98 | 265,247.56 | 1,081,004.01 |

STATE: GUAM

| ZIP CODE | QUARTER 1 | QUARTER 2 | QUARTER 3 | QUARTER 4 | TOTAL GRAMS |
|---|---|---|---|---|---|
| 969 | 215.9 | 207.3 | 223.58 | 173.35 | 820.13 |
| TOTAL | 215.9 | 207.3 | 223.58 | 173.35 | 820.13 |

STATE: HAWAII

| ZIP CODE | QUARTER 1 | QUARTER 2 | QUARTER 3 | QUARTER 4 | TOTAL GRAMS |
|---|---|---|---|---|---|
| 967 | 15,174.92 | 14,287.08 | 13,854.09 | 13,809.71 | 57,125.80 |
| 968 | 5,459.34 | 5,289.59 | 5,042.40 | 4,833.69 | 20,625.02 |
| TOTAL | 20,634.26 | 19,576.67 | 18,896.49 | 18,643.40 | 77,750.82 |

STATE: IDAHO

| ZIP CODE | QUARTER 1 | QUARTER 2 | QUARTER 3 | QUARTER 4 | TOTAL GRAMS |
|---|---|---|---|---|---|
| 832 | 6,485.63 | 6,007.22 | 6,236 | 6,027.38 | 24,756.23 |
| 833 | 7,511.30 | 7,127.77 | 7,179.51 | 6,896.62 | 28,715.20 |
| 834 | 5,490.50 | 5,131.61 | 5,113.20 | 5,123.82 | 20,859.13 |
| 835 | 4,201.19 | 3,941.13 | 3,819.56 | 3,841.36 | 15,803.24 |
| 836 | 17,629.96 | 17,002.67 | 16,913.22 | 17,070.09 | 68,615.94 |
| 837 | 11,559.41 | 11,345.01 | 11,047.51 | 10,852.73 | 44,804.66 |
| 838 | 10,586.52 | 10,171.34 | 10,375.11 | 10,169.31 | 41,302.28 |
| TOTAL | 63,464.51 | 60,726.75 | 60,684.11 | 59,981.31 | 244,856.68 |

STATE: ILLINOIS

| ZIP CODE | QUARTER 1 | QUARTER 2 | QUARTER 3 | QUARTER 4 | TOTAL GRAMS |
|---|---|---|---|---|---|
| 600 | 40,400.61 | 39,760.49 | 38,723.77 | 40,077.13 | 158,962 |
| 601 | 36,073.42 | 34,633.62 | 34,368.85 | 34,758.86 | 139,834.75 |
| 602 | 1,421.39 | 1,389.61 | 1,294.38 | 1,295.13 | 5,400.51 |
| 603 | 1,676.33 | 1,611.60 | 1,687.99 | 1,616.29 | 6,592.21 |
| 604 | 44,592.71 | 42,313.16 | 42,436.31 | 43,014.57 | 172,356.75 |
| 605 | 21,255.35 | 20,408.50 | 20,662.98 | 20,819.35 | 83,146.18 |

| | | | | | |
|---|---|---|---|---|---|
| 606 | 39,398.72 | 37,363.33 | 37,199.76 | 36,328.76 | 150,290.57 |
| 607 | 3,608.23 | 3,571.80 | 3,495.27 | 3,654.10 | 14,329.40 |
| 608 | 2,801.09 | 2,477.89 | 2,429.54 | 2,494.18 | 10,202.70 |
| 609 | 6,864.97 | 6,740.09 | 6,475.77 | 6,577.79 | 26,658.62 |
| 610 | 7,603.04 | 7,352.51 | 7,249.79 | 7,289.23 | 29,494.57 |
| 611 | 11,971.73 | 11,519.39 | 10,773.16 | 11,166.29 | 45,430.57 |
| 612 | 4,072.68 | 3,849.98 | 3,815.64 | 3,647.95 | 15,386.25 |
| 613 | 4,659.54 | 4,675.85 | 4,624.17 | 4,597.33 | 18,556.89 |
| 614 | 4,774.72 | 4,453.98 | 4,693.01 | 4,596.19 | 18,517.90 |
| 615 | 6,481.69 | 6,370.71 | 6,139.21 | 6,270.08 | 25,261.69 |
| 616 | 9,242.07 | 9,061.01 | 8,957.35 | 8,575.53 | 35,835.96 |
| 617 | 5,643.51 | 5,488.72 | 5,319.62 | 5,399.05 | 21,850.90 |
| 618 | 10,168.40 | 9,684.21 | 9,074.48 | 9,569.08 | 38,496.17 |
| 619 | 3,382.08 | 3,047.82 | 3,090.82 | 2,937.92 | 12,458.64 |
| 620 | 13,485.35 | 13,248.40 | 13,171.54 | 13,299.89 | 53,205.18 |
| 622 | 13,444.83 | 12,576.77 | 12,611.06 | 12,473.09 | 51,105.75 |
| 623 | 4,520.89 | 4,446.65 | 4,260.80 | 4,322.46 | 17,550.80 |
| 624 | 5,139.93 | 5,084.14 | 4,934.37 | 5,042.91 | 20,201.35 |

DRUG CODE:9193DRUG NAME:HYDROCODONE
STATE:ILLINOIS

| ZIP CODE | QUARTER 1 | QUARTER 2 | QUARTER 3 | QUARTER 4 | TOTAL GRAMS |
|---|---|---|---|---|---|
| 625 | 5,898.60 | 5,630.26 | 5,865.73 | 5,732.60 | 23,127.19 |
| 626 | 5,462.53 | 5,132.52 | 5,267.51 | 5,336.42 | 21,198.98 |
| 627 | 7,880.80 | 7,491.99 | 7,368.35 | 7,300.31 | 30,041.45 |
| 628 | 9,340.12 | 9,299.16 | 8,533.39 | 8,888.41 | 36,061.08 |
| 629 | 19,787.56 | 19,438.99 | 19,122.31 | 18,887.39 | 77,236.25 |
| TOTAL | 351,052.89 | 338,123.15 | 333,646.93 | 335,968.29 | 1,358,791.26 |

STATE: INDIANA

| ZIP CODE | QUARTER 1 | QUARTER 2 | QUARTER 3 | QUARTER 4 | TOTAL GRAMS |
|---|---|---|---|---|---|
| 460 | 17,519.73 | 16,969.03 | 16,143.40 | 15,468.11 | 66,100.27 |
| 461 | 21,511.79 | 20,572.66 | 19,678.62 | 20,598.64 | 82,361.71 |
| 462 | 34,909.71 | 33,812.61 | 32,632.66 | 33,875.92 | 135,230.90 |
| 463 | 30,670.93 | 30,288.04 | 29,837.37 | 30,280.01 | 121,076.35 |
| 464 | 4,729.40 | 4,541.07 | 4,224.49 | 4,221.01 | 17,715.97 |
| 465 | 11,898.86 | 11,821.48 | 10,965.79 | 11,621.46 | 46,307.59 |
| 466 | 4,072.74 | 4,083.75 | 3,904.62 | 4,126.15 | 16,187.26 |
| 467 | 7,610.13 | 6,750.41 | 6,621.64 | 6,647.09 | 27,629.27 |
| 468 | 9,957.11 | 8,983.90 | 8,642.93 | 9,237.67 | 36,821.61 |
| 469 | 16,041.18 | 14,873.70 | 14,562.33 | 13,842.97 | 59,320.18 |
| 470 | 3,898.02 | 3,804.40 | 3,933.48 | 3,826.18 | 15,462.08 |
| 471 | 17,908.63 | 17,066.23 | 16,596.15 | 16,766.68 | 68,337.69 |
| 472 | 8,877.54 | 8,484.67 | 8,267.64 | 8,263.29 | 33,893.14 |
| 473 | 16,827.07 | 16,170.96 | 15,332.27 | 15,258.25 | 63,588.55 |
| 474 | 11,432.12 | 11,152.09 | 11,099.46 | 10,777.98 | 44,461.65 |
| 475 | 8,250.24 | 8,289.21 | 7,829.36 | 7,945.59 | 32,314.40 |
| 476 | 5,809.89 | 5,781.25 | 5,518.44 | 5,590.04 | 22,699.62 |
| 477 | 13,428.26 | 13,123.38 | 12,938.34 | 12,933.05 | 52,423.03 |
| 478 | 6,539.51 | 6,383.85 | 6,085.21 | 6,092.45 | 25,101.02 |
| 479 | 10,919.45 | 10,671.85 | 10,324.17 | 10,426.42 | 42,341.89 |
| TOTAL | 262,812.31 | 253,624.54 | 245,138.37 | 247,798.96 | 1,009,374.18 |

STATE: IOWA

| ZIP CODE | QUARTER 1 | QUARTER 2 | QUARTER 3 | QUARTER 4 | TOTAL GRAMS |
|---|---|---|---|---|---|
| 500 | 4,721.89 | 4,779.54 | 4,733.79 | 4,720.31 | 18,955.53 |
| 501 | 4,478.53 | 4,255.44 | 4,387.58 | 4,156.26 | 17,277.81 |
| 502 | 4,505.27 | 4,312.88 | 4,271.07 | 4,370.81 | 17,460.03 |
| 503 | 11,024.40 | 10,767.02 | 10,448.57 | 10,205.45 | 42,445.44 |
| 504 | 2,476.23 | 2,328.22 | 2,294.54 | 2,303.54 | 9,402.53 |

| | | | | | |
|---|---|---|---|---|---|
| 505 | 2,313.54 | 2,250.86 | 2,245.21 | 2,252.79 | 9,062.40 |
| 506 | 2,166.75 | 2,017.27 | 2,049.51 | 1,958.90 | 8,192.43 |
| 507 | 1,958.80 | 1,978.79 | 1,807.95 | 1,856.81 | 7,602.35 |
| 508 | 835.55 | 838.53 | 799.96 | 788.17 | 3,262.21 |
| 510 | 1,106.18 | 1,038.27 | 1,052.15 | 1,013.19 | 4,209.79 |
| 511 | 2,449.33 | 2,375.19 | 2,269.01 | 2,198.68 | 9,292.21 |
| 512 | 646.81 | 561.4 | 571.8 | 528.66 | 2,308.67 |
| 513 | 911.67 | 826.87 | 871.79 | 869.88 | 3,480.21 |
| 514 | 963.12 | 872.3 | 876.54 | 833.23 | 3,545.19 |
| 515 | 3,177.15 | 3,137.93 | 3,111.45 | 3,039.43 | 12,465.96 |
| 516 | 612.66 | 571.61 | 579.01 | 580.54 | 2,343.82 |
| 520 | 2,284.69 | 2,162.74 | 2,026.76 | 2,070.51 | 8,544.70 |
| 521 | 721.55 | 716.71 | 620.33 | 639.96 | 2,698.55 |
| 522 | 3,394.55 | 3,275.76 | 3,001.72 | 2,962.56 | 12,634.59 |
| 523 | 1,757.47 | 1,769.83 | 1,668.73 | 1,631.87 | 6,827.90 |
| 524 | 3,184.92 | 3,095.89 | 2,836.01 | 2,840.13 | 11,956.95 |
| 525 | 3,540.05 | 3,541.89 | 3,512 | 3,270.38 | 13,864.32 |
| 526 | 2,767.63 | 2,570.77 | 2,522.98 | 2,489.01 | 10,350.39 |
| 527 | 2,587.23 | 2,428.93 | 2,354.56 | 2,348.52 | 9,719.24 |
| 528 | 3,306.55 | 3,157.03 | 3,122.01 | 3,097.63 | 12,683.22 |
| TOTAL | 67,892.52 | 65,631.67 | 64,035.03 | 63,027.22 | 260,586.44 |

STATE: KANSAS

| ZIP CODE | QUARTER 1 | QUARTER 2 | QUARTER 3 | QUARTER 4 | TOTAL GRAMS |
|---|---|---|---|---|---|
| 660 | 11,967.21 | 11,257.84 | 11,540.12 | 11,559.91 | 46,325.08 |
| 661 | 3,789.69 | 3,593.58 | 3,664.72 | 3,654.48 | 14,702.47 |
| 662 | 9,396.56 | 9,023.69 | 8,998.76 | 8,771.87 | 36,190.88 |
| 664 | 1,644.81 | 1,639.90 | 1,599.60 | 1,617.84 | 6,502.15 |
| 665 | 2,110.14 | 2,066.71 | 1,996.39 | 2,042.87 | 8,216.11 |
| 666 | 5,799.27 | 5,373.42 | 5,404.01 | 5,123.64 | 21,700.34 |
| 667 | 5,490.08 | 5,309.51 | 5,188.33 | 5,268.29 | 21,256.21 |

DRUG CODE:9193DRUG NAME:HYDROCODONE
STATE:KANSAS

| ZIP CODE | QUARTER 1 | QUARTER 2 | QUARTER 3 | QUARTER 4 | TOTAL GRAMS |
|---|---|---|---|---|---|
| 668 | 1,433.86 | 1,429.33 | 1,399.22 | 1,355.30 | 5,617.71 |
| 669 | 721.08 | 713.8 | 688.53 | 683.13 | 2,806.54 |
| 670 | 7,054.20 | 6,740.57 | 6,656.78 | 6,724.62 | 27,176.17 |
| 671 | 3,735.33 | 3,714.02 | 3,737.53 | 3,629.06 | 14,815.94 |
| 672 | 20,777.57 | 19,817.09 | 19,714.28 | 19,362.24 | 79,671.18 |
| 673 | 3,052.55 | 2,874.06 | 2,792.58 | 2,821.99 | 11,541.18 |
| 674 | 4,809.12 | 4,702.41 | 4,750.23 | 4,593.98 | 18,855.74 |
| 675 | 4,208.34 | 4,031.41 | 3,956.57 | 3,843.34 | 16,039.66 |
| 676 | 1,682.08 | 1,564.74 | 1,597.07 | 1,618.51 | 6,462.40 |
| 677 | 751.25 | 764.62 | 724.09 | 744.81 | 2,984.77 |
| 678 | 2,474.72 | 2,490.95 | 2,470.07 | 2,450.62 | 9,886.36 |
| 679 | 1,112.47 | 1,006.01 | 980.14 | 994.09 | 4,092.71 |
| TOTAL | 92,010.33 | 88,113.66 | 87,859.02 | 86,860.59 | 354,843.60 |

STATE: KENTUCKY

| ZIP CODE | QUARTER 1 | QUARTER 2 | QUARTER 3 | QUARTER 4 | TOTAL GRAMS |
|---|---|---|---|---|---|
| 400 | 8,892.46 | 8,588.47 | 8,655.74 | 8,531.03 | 34,667.70 |
| 401 | 4,744.85 | 4,649.56 | 4,776.46 | 4,506.44 | 18,677.31 |
| 402 | 40,688.67 | 38,636.76 | 38,099.37 | 37,830.27 | 155,255.07 |
| 403 | 12,758.97 | 12,431.87 | 12,256.21 | 11,801.39 | 49,248.44 |
| 404 | 8,299.13 | 8,086.54 | 7,722.40 | 7,915.88 | 32,023.95 |
| 405 | 8,896.99 | 8,610.63 | 8,374.57 | 8,319.89 | 34,202.08 |
| 406 | 2,167.59 | 2,154.64 | 2,242.16 | 2,136.23 | 8,700.62 |
| 407 | 6,874.37 | 6,740.95 | 6,724.78 | 6,327.37 | 26,667.47 |
| 408 | 1,970.69 | 2,039.22 | 1,821.55 | 1,852.03 | 7,683.49 |

| | | | | | |
|---|---|---|---|---|---|
| 409 | 8,506.12 | 8,544.95 | 8,689.76 | 7,926.17 | 33,667 |
| 410 | 11,377.35 | 11,314.10 | 10,936.16 | 10,885.04 | 44,512.65 |
| 411 | 4,331.44 | 4,293.72 | 4,374.43 | 4,236.02 | 17,235.61 |
| 412 | 3,382.87 | 3,437.06 | 3,327.19 | 3,016.18 | 13,163.30 |
| 413 | 3,334.61 | 3,367.30 | 3,221.55 | 3,198.57 | 13,122.03 |
| 414 | 1,343.28 | 1,355.89 | 1,410.38 | 1,360.25 | 5,469.80 |
| 415 | 8,367.05 | 8,184.15 | 7,826.34 | 7,171.07 | 31,548.61 |
| 416 | 5,166.77 | 5,093.04 | 4,748.87 | 4,455.49 | 19,464.17 |
| 417 | 5,439.88 | 5,439.35 | 5,136.51 | 4,774.28 | 20,790.02 |
| 418 | 2,418.12 | 2,338.07 | 2,061.46 | 2,037.65 | 8,855.30 |
| 420 | 16,050.47 | 15,945.88 | 16,019.12 | 15,546.44 | 63,561.91 |
| 421 | 13,763.74 | 13,756.85 | 13,624.35 | 13,623.55 | 54,768.49 |
| 422 | 7,017.58 | 6,719.78 | 7,069.35 | 6,792.94 | 27,599.65 |
| 423 | 10,494.10 | 10,414.24 | 10,615.45 | 10,449.70 | 41,973.49 |
| 424 | 11,168.35 | 11,087.01 | 10,829.64 | 10,950.76 | 44,035.76 |
| 425 | 3,874.68 | 3,766.10 | 3,404.23 | 3,019.73 | 14,064.74 |
| 426 | 4,338.39 | 4,248.93 | 4,156.70 | 3,903.16 | 16,647.18 |
| 427 | 9,503.60 | 9,134.39 | 9,357.92 | 9,081.33 | 37,077.24 |
| TOTAL | 225,172.12 | 220,379.45 | 217,482.65 | 211,648.86 | 874,683.08 |

STATE: LOUISIANA

| ZIP CODE | QUARTER 1 | QUARTER 2 | QUARTER 3 | QUARTER 4 | TOTAL GRAMS |
|---|---|---|---|---|---|
| 700 | 20,621.53 | 19,916.60 | 19,789.41 | 19,325.37 | 79,652.91 |
| 701 | 9,417.94 | 8,957.04 | 8,756.01 | 8,279.29 | 35,410.28 |
| 703 | 12,705.71 | 11,944.31 | 12,165.20 | 12,078.03 | 48,893.25 |
| 704 | 18,441.32 | 18,084 | 18,160.41 | 17,952.84 | 72,638.57 |
| 705 | 31,613.46 | 30,666.85 | 30,922.12 | 30,846.65 | 124,049.08 |
| 706 | 9,162.19 | 8,571.74 | 8,338.09 | 8,098.59 | 34,170.61 |
| 707 | 16,199.27 | 15,753.25 | 15,592.07 | 15,460.04 | 63,004.63 |
| 708 | 14,090.98 | 13,556.34 | 13,394.66 | 13,465.85 | 54,507.83 |
| 710 | 4,896.69 | 4,756.16 | 4,883.87 | 5,038.25 | 19,574.97 |
| 711 | 15,897.62 | 15,751.82 | 15,954.87 | 15,836.70 | 63,441.01 |
| 712 | 12,643.02 | 12,406.25 | 12,446.49 | 12,599.13 | 50,094.89 |
| 713 | 15,545.98 | 15,100.03 | 15,690.39 | 14,771.97 | 61,108.37 |
| 714 | 6,029.08 | 5,919.79 | 5,977.91 | 5,932.69 | 23,859.47 |
| TOTAL | 187,264.79 | 181,384.18 | 182,071.50 | 179,685.40 | 730,405.87 |

STATE: MAINE

| ZIP CODE | QUARTER 1 | QUARTER 2 | QUARTER 3 | QUARTER 4 | TOTAL GRAMS |
|---|---|---|---|---|---|
| 39 | 279.77 | 268.99 | 242.04 | 231.1 | 1,021.90 |
| 40 | 5,517.17 | 5,165.77 | 5,052.79 | 4,865.06 | 20,600.79 |
| 41 | 1,332.51 | 1,227.34 | 1,107.08 | 1,069.12 | 4,736.05 |
| 42 | 3,029.96 | 2,720.28 | 2,533.73 | 2,413.92 | 10,697.89 |
| 43 | 2,499.35 | 2,370.09 | 2,302.18 | 2,189.08 | 9,360.70 |

DRUG CODE:9193DRUG NAME:HYDROCODONE
STATE:MAINE

| ZIP CODE | QUARTER 1 | QUARTER 2 | QUARTER 3 | QUARTER 4 | TOTAL GRAMS |
|---|---|---|---|---|---|
| 44 | 2,834.70 | 2,642.63 | 2,408.58 | 2,447.03 | 10,332.94 |
| 45 | 648.1 | 610.87 | 581.72 | 588.28 | 2,428.97 |
| 46 | 1,503.78 | 1,356.43 | 1,412.33 | 1,282.45 | 5,554.99 |
| 47 | 1,534.37 | 1,412.24 | 1,329.23 | 1,302.40 | 5,578.24 |
| 48 | 711.53 | 656.23 | 594.95 | 558.22 | 2,520.93 |
| 49 | 2,897.77 | 2,729.91 | 2,573.11 | 2,399.22 | 10,600.01 |
| TOTAL | 22,789.01 | 21,160.78 | 20,137.74 | 19,345.88 | 83,433.41 |

STATE: MARYLAND

| ZIP CODE | QUARTER 1 | QUARTER 2 | QUARTER 3 | QUARTER 4 | TOTAL GRAMS |
|---|---|---|---|---|---|

| | | | | | |
|---|---|---|---|---|---|
| 206 | 3,205.13 | 2,991.22 | 2,933.74 | 2,856.59 | 11,986.68 |
| 207 | 3,800.68 | 3,531.58 | 3,400.84 | 3,438.37 | 14,171.47 |
| 208 | 3,070.80 | 3,020 | 2,897.70 | 2,956.63 | 11,945.13 |
| 209 | 814.02 | 697.19 | 725.93 | 682.36 | 2,919.50 |
| 210 | 7,152.24 | 6,701.46 | 6,615.43 | 6,521.89 | 26,991.02 |
| 211 | 3,466.48 | 3,316.10 | 3,325.33 | 3,223.49 | 13,331.40 |
| 212 | 7,960.54 | 7,389.92 | 7,368.98 | 7,205.36 | 29,924.80 |
| 214 | 836.94 | 893.69 | 813.87 | 760.01 | 3,304.51 |
| 215 | 3,036.89 | 2,909.05 | 2,914.38 | 2,909.52 | 11,769.84 |
| 216 | 1,687.34 | 1,607.14 | 1,619.52 | 1,624.13 | 6,538.13 |
| 217 | 4,605.31 | 4,363.03 | 4,265.25 | 4,163.07 | 17,396.66 |
| 218 | 1,845.41 | 1,842 | 1,771.72 | 1,742.58 | 7,201.71 |
| 219 | 850.77 | 781.98 | 823.19 | 776.8 | 3,232.74 |
| TOTAL | 42,332.55 | 40,044.36 | 39,475.88 | 38,860.80 | 160,713.59 |

STATE: MASSACHUSETTS

| ZIP CODE | QUARTER 1 | QUARTER 2 | QUARTER 3 | QUARTER 4 | TOTAL GRAMS |
|---|---|---|---|---|---|
| 10 | 2,751.14 | 2,649.69 | 2,656.25 | 2,463.76 | 10,520.84 |
| 11 | 595.34 | 596.6 | 569.02 | 481.34 | 2,242.30 |
| 12 | 770.65 | 681.41 | 691.22 | 666.03 | 2,809.31 |
| 13 | 381.35 | 381.83 | 378.56 | 383.02 | 1,524.76 |
| 14 | 1,629.80 | 1,542.93 | 1,453.05 | 1,399.92 | 6,025.70 |
| 15 | 2,470.10 | 2,432.02 | 2,317.57 | 2,239.17 | 9,458.86 |
| 16 | 1,607.78 | 1,517.69 | 1,385.44 | 1,313.76 | 5,824.67 |
| 17 | 1,839.71 | 1,697.08 | 1,613.88 | 1,605.09 | 6,755.76 |
| 18 | 6,580.10 | 6,084.76 | 5,824.98 | 5,572.44 | 24,062.28 |
| 19 | 3,294.34 | 3,115.48 | 3,054.67 | 2,890.28 | 12,354.77 |
| 20 | 1,822.74 | 1,663.55 | 1,617.71 | 1,566.27 | 6,670.27 |
| 21 | 4,348.66 | 3,999.12 | 3,745.79 | 3,713.36 | 15,806.93 |
| 22 | 80.82 | 73.12 | 77.46 | 76.39 | 307.79 |
| 23 | 2,934.21 | 2,685.08 | 2,607.40 | 2,473.10 | 10,699.79 |
| 24 | 1,094.12 | 1,123.94 | 951.19 | 954.25 | 4,123.50 |
| 25 | 621.8 | 602.56 | 615.05 | 577.15 | 2,416.56 |
| 26 | 1,171.89 | 1,113.68 | 1,071.87 | 1,008.37 | 4,365.81 |
| 27 | 3,970.57 | 3,742.03 | 3,598.37 | 3,418.57 | 14,729.54 |
| TOTAL | 37,965.12 | 35,702.57 | 34,229.48 | 32,802.27 | 140,699.44 |

STATE: MICHIGAN

| ZIP CODE | QUARTER 1 | QUARTER 2 | QUARTER 3 | QUARTER 4 | TOTAL GRAMS |
|---|---|---|---|---|---|
| 480 | 62,223.36 | 60,161.94 | 56,513.89 | 58,365.67 | 237,264.86 |
| 481 | 80,071.52 | 76,096.02 | 71,578.93 | 74,150.53 | 301,897 |
| 482 | 37,041.39 | 35,011.35 | 34,937.89 | 34,584.27 | 141,574.90 |
| 483 | 40,150.78 | 38,748.23 | 35,703.69 | 37,940.69 | 152,543.39 |
| 484 | 22,476.37 | 21,572.92 | 19,984.03 | 21,153.55 | 85,186.87 |
| 485 | 14,296.57 | 13,765.16 | 12,561.53 | 13,357 | 53,980.26 |
| 486 | 25,017.30 | 24,420.40 | 23,394.01 | 23,783 | 96,614.71 |
| 487 | 13,380.30 | 13,272.25 | 12,498.80 | 12,621.56 | 51,772.91 |
| 488 | 23,770.94 | 22,863.74 | 18,297.56 | 22,435.38 | 87,367.62 |
| 489 | 9,346.56 | 8,892.53 | 7,954.38 | 8,962.68 | 35,156.15 |
| 490 | 35,379.01 | 33,740.67 | 28,924.44 | 31,740.41 | 129,784.53 |
| 491 | 3,317.65 | 3,100.20 | 2,782.76 | 2,901.50 | 12,102.11 |
| 492 | 14,274.14 | 13,846.02 | 12,003.10 | 13,527.46 | 53,650.72 |
| 493 | 10,185.44 | 10,047.89 | 7,915.24 | 9,678.09 | 37,826.66 |
| 494 | 27,435.15 | 25,373.62 | 21,416.36 | 24,598.54 | 98,823.67 |
| 495 | 19,249.96 | 17,928.15 | 15,200.92 | 17,956.66 | 70,335.69 |
| 496 | 12,495.98 | 12,497.87 | 11,171.92 | 12,107.41 | 48,273.18 |
| 497 | 10,473.44 | 10,227.72 | 9,894.38 | 9,672.29 | 40,267.83 |
| 498 | 9,096.73 | 8,658.36 | 8,907.04 | 8,295.08 | 34,957.21 |
| 499 | 2,697.95 | 2,688.14 | 2,778.24 | 2,595.87 | 10,760.20 |

RETAIL DRUG DISTRIBUTION BY ZIP CODE WITHIN STATE BY GRAMS WT

STATE:MICHIGAN

| ZIP CODE | QUARTER 1 | QUARTER 2 | QUARTER 3 | QUARTER 4 | TOTAL GRAMS |
|---|---|---|---|---|---|
| TOTAL | 472,380.54 | 452,913.18 | 414,419.11 | 440,427.64 | 1,780,140.47 |

STATE: MINNESOTA

| ZIP CODE | QUARTER 1 | QUARTER 2 | QUARTER 3 | QUARTER 4 | TOTAL GRAMS |
|---|---|---|---|---|---|
| 550 | 5,629.87 | 5,351.07 | 5,225.90 | 5,007.91 | 21,214.75 |
| 551 | 6,213.10 | 5,908.55 | 5,734.32 | 5,745.26 | 23,601.23 |
| 553 | 7,411.80 | 7,064.33 | 6,908.74 | 6,895.01 | 28,279.88 |
| 554 | 9,760.85 | 9,359.93 | 9,268.24 | 8,862.74 | 37,251.76 |
| 556 | 279.45 | 284 | 246.52 | 242.06 | 1,052.03 |
| 557 | 3,989.52 | 3,770.98 | 3,744.47 | 3,417.72 | 14,922.69 |
| 558 | 2,994.05 | 2,861.24 | 2,731.41 | 2,668.71 | 11,255.41 |
| 559 | 2,549.70 | 2,575.58 | 2,438.45 | 2,242.89 | 9,806.62 |
| 560 | 2,306.96 | 2,154.79 | 2,108.55 | 2,121.99 | 8,692.29 |
| 561 | 1,009.12 | 960.66 | 916.07 | 864.45 | 3,750.30 |
| 562 | 1,866.10 | 1,737.37 | 1,733.92 | 1,668.14 | 7,005.53 |
| 563 | 4,622.05 | 4,298.52 | 4,243.98 | 4,058.68 | 17,223.23 |
| 564 | 2,234.90 | 2,215.55 | 2,146 | 1,977.50 | 8,573.95 |
| 565 | 2,622.70 | 2,511.66 | 2,372.35 | 2,217.02 | 9,723.73 |
| 566 | 1,548 | 1,497.51 | 1,496.72 | 1,364.69 | 5,906.92 |
| 567 | 1,116.95 | 1,055.72 | 1,006.34 | 960.7 | 4,139.71 |
| TOTAL | 56,155.12 | 53,607.46 | 52,321.98 | 50,315.47 | 212,400.03 |

STATE: MISSISSIPPI

| ZIP CODE | QUARTER 1 | QUARTER 2 | QUARTER 3 | QUARTER 4 | TOTAL GRAMS |
|---|---|---|---|---|---|
| 386 | 12,319.13 | 11,751.52 | 11,667.55 | 11,665.38 | 47,403.58 |
| 387 | 3,641.96 | 3,659.82 | 3,636.36 | 3,646.97 | 14,585.11 |
| 388 | 15,013.40 | 14,757.47 | 14,387.88 | 14,538.82 | 58,697.57 |
| 389 | 4,123.38 | 4,059.24 | 4,075.86 | 3,952.87 | 16,211.35 |
| 390 | 9,383.60 | 9,176.97 | 9,063.49 | 9,004.71 | 36,628.77 |
| 391 | 10,143.21 | 9,358.08 | 9,492.37 | 9,697.61 | 38,691.27 |
| 392 | 10,494.77 | 9,675.85 | 9,413.63 | 9,502.55 | 39,086.80 |
| 393 | 13,527.66 | 13,167.24 | 13,163.60 | 13,089.85 | 52,948.35 |
| 394 | 22,936.21 | 22,192.14 | 22,306.21 | 22,530.06 | 89,964.62 |
| 395 | 23,688.22 | 22,711.54 | 22,609.58 | 21,585.73 | 90,595.07 |
| 396 | 6,286.81 | 6,147.79 | 5,927.86 | 6,368.08 | 24,730.54 |
| 397 | 6,610.26 | 6,478.10 | 6,441.47 | 6,120.61 | 25,650.44 |
| TOTAL | 138,168.61 | 133,135.76 | 132,185.86 | 131,703.24 | 535,193.47 |

STATE: MISSOURI

| ZIP CODE | QUARTER 1 | QUARTER 2 | QUARTER 3 | QUARTER 4 | TOTAL GRAMS |
|---|---|---|---|---|---|
| 630 | 19,003.28 | 18,123.13 | 18,193.84 | 18,043.03 | 73,363.28 |
| 631 | 26,609.19 | 25,505.32 | 24,904.40 | 22,582.42 | 99,601.33 |
| 633 | 11,176.23 | 10,725.92 | 10,481.51 | 10,605.44 | 42,989.10 |
| 634 | 2,203.47 | 2,158.97 | 2,075.86 | 2,049.02 | 8,487.32 |
| 635 | 1,918.40 | 1,917.83 | 1,873.35 | 1,936.72 | 7,646.30 |
| 636 | 7,652.37 | 7,631.75 | 7,428.92 | 7,427.32 | 30,140.36 |
| 637 | 6,014.82 | 6,050.23 | 5,753.77 | 5,742.69 | 23,561.51 |
| 638 | 10,582.77 | 10,411.90 | 10,744.81 | 10,521.86 | 42,261.34 |
| 639 | 7,787.53 | 7,197.63 | 7,226.92 | 7,166.02 | 29,378.10 |
| 640 | 18,970.90 | 18,425.56 | 18,344 | 18,217.51 | 73,957.97 |
| 641 | 15,432.21 | 14,553.37 | 14,603.38 | 14,217.70 | 58,806.66 |
| 644 | 2,584.12 | 2,420.94 | 2,362.45 | 2,347.25 | 9,714.76 |
| 645 | 4,833.14 | 4,840.17 | 4,742.11 | 4,642.82 | 19,058.24 |
| 646 | 1,627.62 | 1,578.62 | 1,526.16 | 1,700.81 | 6,433.21 |
| 647 | 5,085.68 | 5,006.81 | 4,946.63 | 4,930.08 | 19,969.20 |
| 648 | 8,680.61 | 8,555.18 | 8,389.68 | 8,250.98 | 33,876.45 |
| 650 | 3,947.14 | 3,678.94 | 3,573.45 | 3,535.06 | 14,734.59 |
| 651 | 2,059.97 | 1,976.01 | 1,883.81 | 1,864.90 | 7,784.69 |
| 652 | 5,680.08 | 5,431.65 | 5,299.63 | 5,242.58 | 21,653.94 |

| | | | | | |
|---|---|---|---|---|---|
| 653 | 2,540.27 | 2,485.60 | 2,557.95 | 2,540.92 | 10,124.74 |
| 654 | 2,665.14 | 2,626.63 | 2,550.65 | 2,557.10 | 10,399.52 |
| 655 | 4,443.39 | 4,351.22 | 4,264.88 | 4,331.55 | 17,391.04 |
| 656 | 8,110.67 | 7,873.95 | 7,831.83 | 7,688.51 | 31,504.96 |
| 657 | 12,130.74 | 11,827.54 | 11,868.68 | 11,648.58 | 47,475.54 |
| 658 | 11,601.83 | 10,871.23 | 11,021.71 | 10,986.03 | 44,480.80 |
| TOTAL | 203,341.57 | 196,226.10 | 194,450.38 | 190,776.90 | 784,794.95 |

STATE: MONTANA

| ZIP CODE | QUARTER 1 | QUARTER 2 | QUARTER 3 | QUARTER 4 | TOTAL GRAMS |
|---|---|---|---|---|---|
| 590 | 1,301.13 | 1,259.88 | 1,259.95 | 1,153.88 | 4,974.84 |

DRUG CODE:9193DRUG NAME:HYDROCODONE
STATE:MONTANA

| ZIP CODE | QUARTER 1 | QUARTER 2 | QUARTER 3 | QUARTER 4 | TOTAL GRAMS |
|---|---|---|---|---|---|
| 591 | 3,910.83 | 3,697.84 | 3,838.01 | 3,922.97 | 15,369.65 |
| 592 | 666.44 | 637.71 | 638.28 | 571.59 | 2,514.02 |
| 593 | 798.49 | 758.43 | 826.16 | 751.02 | 3,134.10 |
| 594 | 3,931.94 | 3,740.78 | 3,630.84 | 3,577.05 | 14,880.61 |
| 595 | 810.35 | 777.63 | 771.45 | 717.86 | 3,077.29 |
| 596 | 3,343.60 | 3,392.48 | 3,249.72 | 3,169.61 | 13,155.41 |
| 597 | 3,558.21 | 3,413.94 | 3,418.96 | 3,356.07 | 13,747.18 |
| 598 | 6,704.08 | 6,748.39 | 6,740.94 | 6,579.46 | 26,772.87 |
| 599 | 4,346.83 | 4,187.54 | 4,372.99 | 4,122.33 | 17,029.69 |
| TOTAL | 29,371.90 | 28,614.62 | 28,747.30 | 27,921.84 | 114,655.66 |

STATE: NEBRASKA

| ZIP CODE | QUARTER 1 | QUARTER 2 | QUARTER 3 | QUARTER 4 | TOTAL GRAMS |
|---|---|---|---|---|---|
| 680 | 4,053.33 | 3,944.09 | 3,957.04 | 3,890.72 | 15,845.18 |
| 681 | 13,018.83 | 12,510.89 | 12,710.74 | 12,522.97 | 50,763.43 |
| 683 | 1,672.53 | 1,627.11 | 1,570.71 | 1,536.86 | 6,407.21 |
| 684 | 1,201.31 | 1,129.25 | 1,048.05 | 1,023.02 | 4,401.63 |
| 685 | 6,006.06 | 5,837.54 | 5,677.59 | 5,484.44 | 23,005.63 |
| 686 | 1,013.27 | 941.59 | 978.56 | 918.96 | 3,852.38 |
| 687 | 2,138.52 | 2,023.09 | 1,966.50 | 1,957.55 | 8,085.66 |
| 688 | 3,470.25 | 3,387.56 | 3,373.21 | 3,339.18 | 13,570.20 |
| 689 | 1,397.70 | 1,315.54 | 1,269.21 | 1,334.50 | 5,316.95 |
| 690 | 510.31 | 511.38 | 490.35 | 434.67 | 1,946.71 |
| 691 | 1,806.89 | 1,782.06 | 1,686.61 | 1,692.49 | 6,968.05 |
| 692 | 221.82 | 226.81 | 222.82 | 227.13 | 898.58 |
| 693 | 1,342.85 | 1,322.19 | 1,320.65 | 1,272.30 | 5,257.99 |
| TOTAL | 37,853.67 | 36,559.10 | 36,272.04 | 35,634.79 | 146,319.60 |

STATE: NEVADA

| ZIP CODE | QUARTER 1 | QUARTER 2 | QUARTER 3 | QUARTER 4 | TOTAL GRAMS |
|---|---|---|---|---|---|
| 890 | 26,906.27 | 26,362.03 | 26,054.23 | 25,798.76 | 105,121.29 |
| 891 | 57,368 | 52,285.94 | 52,223.55 | 51,812.88 | 213,690.37 |
| 893 | 603.49 | 590.94 | 558.61 | 541.75 | 2,294.79 |
| 894 | 10,450.21 | 9,620.02 | 9,055.96 | 9,026.37 | 38,152.56 |
| 895 | 11,841.85 | 11,253.93 | 10,480.27 | 10,414.13 | 43,990.18 |
| 897 | 2,902.56 | 2,832.75 | 2,860.44 | 2,845.67 | 11,441.42 |
| 898 | 1,434.25 | 1,301.79 | 1,242.91 | 1,249.62 | 5,228.57 |
| TOTAL | 111,506.63 | 104,247.40 | 102,475.97 | 101,689.18 | 419,919.18 |

STATE: NEW HAMPSHIRE

| ZIP CODE | QUARTER 1 | QUARTER 2 | QUARTER 3 | QUARTER 4 | TOTAL GRAMS |
|---|---|---|---|---|---|
| 30 | 2,615.44 | 2,445.02 | 2,398.48 | 2,366.10 | 9,825.04 |
| 31 | 1,583.26 | 1,512.54 | 1,427.73 | 1,461.57 | 5,985.10 |
| 32 | 1,285.53 | 1,226.70 | 1,230.62 | 1,212.33 | 4,955.18 |

| | | | | | |
|---|---|---|---|---|---|
| 33 | 862.16 | 745.11 | 741.96 | 723.51 | 3,072.74 |
| 34 | 662.51 | 630.45 | 600.28 | 596.63 | 2,489.87 |
| 35 | 429.27 | 429.1 | 450.1 | 376.46 | 1,684.93 |
| 36 | 42.44 | 39.89 | 42.23 | 34.88 | 159.44 |
| 37 | 547.25 | 524.36 | 531.7 | 526.52 | 2,129.83 |
| 38 | 2,911.30 | 2,681.15 | 2,722.62 | 2,753.67 | 11,068.74 |
| TOTAL | 10,939.16 | 10,234.32 | 10,145.72 | 10,051.67 | 41,370.87 |

STATE: NEW JERSEY

| ZIP CODE | QUARTER 1 | QUARTER 2 | QUARTER 3 | QUARTER 4 | TOTAL GRAMS |
|---|---|---|---|---|---|
| 70 | 5,288.26 | 4,845.78 | 4,625.27 | 4,702.68 | 19,461.99 |
| 71 | 510.73 | 459.41 | 396.66 | 418.35 | 1,785.15 |
| 72 | 424.39 | 384.44 | 311.3 | 262.23 | 1,382.36 |
| 73 | 482.43 | 508.86 | 464.77 | 464.35 | 1,920.41 |
| 74 | 2,664.81 | 2,315.47 | 2,303.80 | 2,438.44 | 9,722.52 |
| 75 | 374.56 | 340.7 | 318.15 | 311.68 | 1,345.09 |
| 76 | 1,603.86 | 1,439.37 | 1,430.21 | 1,590.18 | 6,063.62 |
| 77 | 3,803.76 | 3,624.59 | 3,454.65 | 3,519 | 14,402 |
| 78 | 2,453.46 | 2,301.21 | 2,301.47 | 2,324.46 | 9,380.60 |
| 79 | 997.22 | 857.59 | 814.27 | 899.77 | 3,568.85 |
| 80 | 10,129.39 | 9,804.29 | 9,529.64 | 9,379.20 | 38,842.52 |
| 81 | 716.56 | 705.66 | 647.61 | 652.16 | 2,721.99 |
| 82 | 2,580.88 | 2,578.09 | 2,589.30 | 2,453.67 | 10,201.94 |
| 83 | 2,262.96 | 2,184.17 | 2,095.12 | 2,104.10 | 8,646.35 |
| 84 | 354.65 | 338 | 355.16 | 316.1 | 1,363.91 |
| 85 | 1,206.23 | 1,099.91 | 1,118.72 | 1,113.72 | 4,538.58 |

DRUG CODE:9193DRUG NAME:HYDROCODONE
STATE:NEW JERSEY

| ZIP CODE | QUARTER 1 | QUARTER 2 | QUARTER 3 | QUARTER 4 | TOTAL GRAMS |
|---|---|---|---|---|---|
| 86 | 1,378.22 | 1,381.41 | 1,275.76 | 1,311.31 | 5,346.70 |
| 87 | 3,020.48 | 2,827.22 | 2,854.74 | 2,856.73 | 11,559.17 |
| 88 | 3,767.05 | 3,149.32 | 3,180.94 | 3,310.41 | 13,407.72 |
| 89 | 268.08 | 225.54 | 209.13 | 222.31 | 925.06 |
| TOTAL | 44,287.98 | 41,371.03 | 40,276.67 | 40,650.85 | 166,586.53 |

STATE: NEW MEXICO

| ZIP CODE | QUARTER 1 | QUARTER 2 | QUARTER 3 | QUARTER 4 | TOTAL GRAMS |
|---|---|---|---|---|---|
| 870 | 3,091.02 | 2,927.95 | 2,893.14 | 2,853.22 | 11,765.33 |
| 871 | 16,020.69 | 14,778.89 | 14,951.26 | 14,426.97 | 60,177.81 |
| 873 | 673.97 | 674.64 | 676.85 | 615.22 | 2,640.68 |
| 874 | 2,362.51 | 2,130.23 | 2,088.89 | 2,074.35 | 8,655.98 |
| 875 | 4,894.59 | 4,406.75 | 4,488.75 | 4,367.80 | 18,157.89 |
| 877 | 1,128.64 | 1,007.02 | 985.59 | 968.74 | 4,089.99 |
| 878 | 261.35 | 220.62 | 214.42 | 227.85 | 924.24 |
| 879 | 371.28 | 333.53 | 317.49 | 307.5 | 1,329.80 |
| 880 | 3,457.05 | 3,152.46 | 3,109.47 | 3,126.12 | 12,845.10 |
| 881 | 1,976.09 | 1,807.03 | 1,552.51 | 1,479 | 6,814.63 |
| 882 | 6,905.02 | 6,330.67 | 6,330.39 | 6,336.52 | 25,902.60 |
| 883 | 1,924.79 | 1,817.38 | 1,771.24 | 1,747.18 | 7,260.59 |
| 884 | 387.57 | 321.51 | 329.08 | 321.6 | 1,359.76 |
| TOTAL | 43,454.57 | 39,908.68 | 39,709.08 | 38,852.07 | 161,924.40 |

STATE: NEW YORK

| ZIP CODE | QUARTER 1 | QUARTER 2 | QUARTER 3 | QUARTER 4 | TOTAL GRAMS |
|---|---|---|---|---|---|
| 63 | 0.3 | 0.3 | 0 | 0.3 | 0.9 |
| 100 | 5,866.25 | 5,059.82 | 4,689.65 | 4,904.32 | 20,520.04 |
| 101 | 255.67 | 295.7 | 214.1 | 194.51 | 959.98 |
| 102 | 24.21 | 21.9 | 24.32 | 19.68 | 90.11 |
| 103 | 3,159.52 | 2,966.99 | 2,795.23 | 2,894.79 | 11,816.53 |

| | | | | | |
|---|---|---|---|---|---|
| 104 | 2,339.94 | 2,110.49 | 1,907.51 | 1,917.58 | 8,275.52 |
| 105 | 3,221.66 | 2,988.69 | 2,819.80 | 2,845.64 | 11,875.79 |
| 106 | 432.46 | 400.91 | 394.21 | 385.41 | 1,612.99 |
| 107 | 1,082.93 | 1,023.39 | 986.64 | 967.98 | 4,060.94 |
| 108 | 342.02 | 321.95 | 272.26 | 262.12 | 1,198.35 |
| 109 | 3,430.14 | 3,152.74 | 2,971.81 | 3,120.41 | 12,675.10 |
| 110 | 1,251.71 | 1,111.56 | 1,080.74 | 1,080.41 | 4,524.42 |
| 111 | 685.22 | 688.71 | 616.3 | 627.69 | 2,617.92 |
| 112 | 5,072.95 | 4,813.91 | 4,394.78 | 4,458.36 | 18,740 |
| 113 | 2,742.53 | 2,521.17 | 2,350 | 2,370.81 | 9,984.51 |
| 114 | 1,244.72 | 1,240.34 | 1,134.38 | 1,165.84 | 4,785.28 |
| 115 | 3,826.79 | 3,461 | 3,291.71 | 3,183.70 | 13,763.20 |
| 116 | 354.82 | 355.32 | 327.49 | 334.1 | 1,371.73 |
| 117 | 15,810.09 | 14,647.31 | 14,334.62 | 14,536.16 | 59,328.18 |
| 118 | 576.14 | 483.35 | 399.31 | 463.32 | 1,922.12 |
| 119 | 2,560.82 | 2,381.33 | 2,395.06 | 2,331.40 | 9,668.61 |
| 120 | 8,199.39 | 7,696.83 | 7,377.88 | 7,484.81 | 30,758.91 |
| 121 | 4,502.94 | 4,197.98 | 4,022.70 | 4,043.40 | 16,767.02 |
| 122 | 3,732.12 | 3,421.69 | 3,303.25 | 3,473.24 | 13,930.30 |
| 123 | 3,423.40 | 3,140.03 | 3,023.63 | 3,028.13 | 12,615.19 |
| 124 | 3,140.53 | 2,925.80 | 2,964.96 | 2,834.46 | 11,865.75 |
| 125 | 5,347.13 | 5,134.42 | 4,874.57 | 5,003.58 | 20,359.70 |
| 126 | 895.18 | 866.93 | 748.38 | 809.19 | 3,319.68 |
| 127 | 1,207.44 | 1,212.10 | 1,156.60 | 1,122.80 | 4,698.94 |
| 128 | 5,525.48 | 5,376.41 | 5,313.45 | 5,107.60 | 21,322.94 |
| 129 | 3,275.69 | 3,222.10 | 3,220.02 | 3,090.62 | 12,808.43 |
| 130 | 6,961.29 | 6,505.13 | 6,040.09 | 6,015.37 | 25,521.88 |
| 131 | 2,347.87 | 2,208.67 | 2,182.19 | 2,121.19 | 8,859.92 |
| 132 | 5,607.75 | 5,196.28 | 4,755.30 | 4,539.89 | 20,099.22 |
| 133 | 3,258.88 | 3,154.72 | 3,007.95 | 2,766.14 | 12,187.69 |
| 134 | 4,705.42 | 4,566.76 | 4,406.78 | 4,414.76 | 18,093.72 |
| 135 | 2,230.85 | 2,176.89 | 2,102.14 | 1,986.94 | 8,496.82 |
| 136 | 4,009.75 | 4,077.15 | 4,041.31 | 3,900.20 | 16,028.41 |
| 137 | 2,045.64 | 1,956.32 | 1,904.45 | 1,883.58 | 7,789.99 |
| 138 | 4,327.12 | 4,218.74 | 4,246.48 | 3,984.72 | 16,777.06 |
| 139 | 2,051.52 | 1,935.03 | 1,821.27 | 1,767.21 | 7,575.03 |
| 140 | 14,459.19 | 13,524.50 | 13,198.91 | 13,037.23 | 54,219.83 |
| 141 | 7,238.67 | 6,948.50 | 6,681.63 | 6,445.14 | 27,313.94 |

DRUG CODE:9193DRUG NAME:HYDROCODONE
STATE:NEW YORK

| ZIP CODE | QUARTER 1 | QUARTER 2 | QUARTER 3 | QUARTER 4 | TOTAL GRAMS |
|---|---|---|---|---|---|
| 142 | 26,862.80 | 25,280.87 | 24,547.66 | 23,569.22 | 100,260.55 |
| 143 | 3,338.15 | 3,094.35 | 2,976.81 | 2,895.55 | 12,304.86 |
| 144 | 5,084.18 | 4,896.34 | 4,744.68 | 4,700.84 | 19,426.04 |
| 145 | 4,162.06 | 4,028.02 | 3,907.51 | 3,839.60 | 15,937.19 |
| 146 | 7,599.65 | 7,168.13 | 6,913.45 | 6,686.80 | 28,368.03 |
| 147 | 5,863.18 | 5,714.09 | 5,252.91 | 5,142.53 | 21,972.71 |
| 148 | 5,007.57 | 4,789.75 | 4,687.72 | 4,559.17 | 19,044.21 |
| 149 | 1,291.80 | 1,184.25 | 1,076.09 | 1,038.40 | 4,590.54 |
| TOTAL | 211,983.53 | 199,865.66 | 191,900.69 | 189,356.84 | 793,106.72 |

STATE: NORTH CAROLINA

| ZIP CODE | QUARTER 1 | QUARTER 2 | QUARTER 3 | QUARTER 4 | TOTAL GRAMS |
|---|---|---|---|---|---|
| 270 | 8,667.26 | 8,302.97 | 8,167.61 | 8,210.36 | 33,348.20 |
| 271 | 7,372.18 | 6,701.46 | 6,459.60 | 6,456.80 | 26,990.04 |
| 272 | 14,410.04 | 14,177.80 | 13,947.97 | 14,537.33 | 57,073.14 |
| 273 | 8,632.68 | 8,151.34 | 8,041.38 | 7,994.59 | 32,819.99 |
| 274 | 5,818.17 | 5,254.55 | 5,229.76 | 5,294.17 | 21,596.65 |
| 275 | 17,441.68 | 16,226.83 | 15,373.82 | 15,877.54 | 64,919.87 |

| | | | | | |
|---|---|---|---|---|---|
| 276 | 5,715.95 | 5,065.42 | 5,122.33 | 5,443.72 | 21,347.42 |
| 277 | 4,750.67 | 4,157.05 | 4,353.75 | 4,137.60 | 17,399.07 |
| 278 | 10,513.69 | 10,075.51 | 10,125.78 | 10,055.85 | 40,770.83 |
| 279 | 3,499.54 | 3,287.22 | 3,181.19 | 3,092.95 | 13,060.90 |
| 280 | 25,168.80 | 23,601.97 | 23,244.92 | 23,179.42 | 95,195.11 |
| 281 | 17,662.74 | 16,574.39 | 16,267.71 | 16,367.35 | 66,872.19 |
| 282 | 13,547.53 | 12,161.38 | 11,720.98 | 12,193.52 | 49,623.41 |
| 283 | 20,414.78 | 19,134.98 | 19,006.89 | 18,638.15 | 77,194.80 |
| 284 | 13,750.06 | 12,824.01 | 12,425.49 | 12,428.21 | 51,427.77 |
| 285 | 9,977.77 | 9,514.18 | 9,070.15 | 8,863.07 | 37,425.17 |
| 286 | 22,002.38 | 21,307.52 | 20,843.79 | 20,611.25 | 84,764.94 |
| 287 | 12,622.64 | 12,124.85 | 11,961.30 | 12,055.32 | 48,764.11 |
| 288 | 6,809.83 | 6,544.22 | 6,427.71 | 6,303.01 | 26,084.77 |
| 289 | 1,607.23 | 1,510.84 | 1,558.76 | 1,504.08 | 6,180.91 |
| TOTAL | 230,385.62 | 216,698.49 | 212,530.89 | 213,244.29 | 872,859.29 |

STATE: NORTH DAKOTA

| ZIP CODE | QUARTER 1 | QUARTER 2 | QUARTER 3 | QUARTER 4 | TOTAL GRAMS |
|---|---|---|---|---|---|
| 580 | 827.66 | 753.15 | 693.34 | 597.15 | 2,871.30 |
| 581 | 2,768.39 | 2,442.91 | 2,286.71 | 2,093.36 | 9,591.37 |
| 582 | 1,648.15 | 1,592.24 | 1,384.80 | 1,276.98 | 5,902.17 |
| 583 | 765.74 | 797.72 | 732.35 | 694 | 2,989.81 |
| 584 | 681.72 | 653.61 | 670.88 | 572.76 | 2,578.97 |
| 585 | 4,242.88 | 4,069.28 | 3,759.91 | 3,541.57 | 15,613.64 |
| 586 | 721.99 | 738.06 | 677.62 | 635.05 | 2,772.72 |
| 587 | 996.22 | 949.2 | 893.24 | 778.61 | 3,617.27 |
| 588 | 754.29 | 712.58 | 686.48 | 568.52 | 2,721.87 |
| TOTAL | 13,407.04 | 12,708.75 | 11,785.33 | 10,758 | 48,659.12 |

STATE: OHIO

| ZIP CODE | QUARTER 1 | QUARTER 2 | QUARTER 3 | QUARTER 4 | TOTAL GRAMS |
|---|---|---|---|---|---|
| 430 | 11,839.94 | 11,446.04 | 10,873.95 | 11,194.63 | 45,354.56 |
| 431 | 8,885.80 | 8,615.03 | 8,196.95 | 8,218.44 | 33,916.22 |
| 432 | 18,816.33 | 20,161.48 | 19,437.51 | 16,335.86 | 74,751.18 |
| 433 | 3,363.28 | 3,320.51 | 3,207.61 | 3,188.35 | 13,079.75 |
| 434 | 2,105.74 | 2,042.12 | 1,920.20 | 2,024.68 | 8,092.74 |
| 435 | 4,432.29 | 4,369.99 | 4,187.48 | 4,340.25 | 17,330.01 |
| 436 | 8,415.90 | 8,504.99 | 7,898.33 | 7,755.35 | 32,574.57 |
| 437 | 4,737.35 | 4,657 | 4,502.74 | 4,394.46 | 18,291.55 |
| 438 | 702.41 | 699.45 | 693.63 | 665.57 | 2,761.06 |
| 439 | 4,381.39 | 4,313.44 | 4,194.09 | 4,011.47 | 16,900.39 |
| 440 | 11,353.60 | 10,944.99 | 10,636.01 | 10,191.11 | 43,125.71 |
| 441 | 12,749.22 | 12,266.14 | 11,964.42 | 11,754.12 | 48,733.90 |
| 442 | 8,521.94 | 8,282.02 | 8,125.39 | 7,814.70 | 32,744.05 |
| 443 | 4,378.62 | 4,233 | 4,056.46 | 3,868.24 | 16,536.32 |
| 444 | 10,020.72 | 9,435.48 | 9,112.71 | 8,718.55 | 37,287.46 |
| 445 | 6,116.69 | 5,773.92 | 5,677.93 | 5,481.95 | 23,050.49 |
| 446 | 7,665.80 | 7,274.51 | 7,232.62 | 6,962.14 | 29,135.07 |
| 447 | 4,254.92 | 4,124.53 | 3,971.69 | 3,940.34 | 16,291.48 |
| 448 | 4,548.99 | 4,375.27 | 4,097.11 | 3,994.28 | 17,015.65 |
| 449 | 1,126.39 | 1,044.61 | 1,012.14 | 1,069.81 | 4,252.95 |

DRUG CODE:9193DRUG NAME:HYDROCODONE
STATE:OHIO

| ZIP CODE | QUARTER 1 | QUARTER 2 | QUARTER 3 | QUARTER 4 | TOTAL GRAMS |
|---|---|---|---|---|---|
| 450 | 18,980.40 | 18,242.36 | 17,279.56 | 17,487.51 | 71,989.83 |
| 451 | 6,364.46 | 6,089.05 | 5,864.24 | 5,833.17 | 24,150.92 |
| 452 | 14,959.54 | 14,235.02 | 13,583.44 | 13,823.23 | 56,601.23 |
| 453 | 11,383.58 | 10,841.50 | 10,248.89 | 10,510.30 | 42,984.27 |
| 454 | 13,501.93 | 12,977.62 | 12,283.73 | 12,751.03 | 51,514.31 |

| | | | | | |
|---|---|---|---|---|---|
| 455 | 2,809.01 | 2,833.90 | 2,722.69 | 2,683.08 | 11,048.68 |
| 456 | 10,579.87 | 10,248.83 | 10,240.16 | 9,852.08 | 40,920.94 |
| 457 | 4,196.10 | 3,976.66 | 3,978.16 | 3,826.46 | 15,977.38 |
| 458 | 5,487.74 | 5,408.15 | 5,138.79 | 5,291.12 | 21,325.80 |
| TOTAL | 226,679.95 | 220,737.61 | 212,338.63 | 207,982.28 | 867,738.47 |

STATE: OKLAHOMA

| ZIP CODE | QUARTER 1 | QUARTER 2 | QUARTER 3 | QUARTER 4 | TOTAL GRAMS |
|---|---|---|---|---|---|
| 730 | 28,783.18 | 27,224.45 | 26,994 | 27,222.13 | 110,223.76 |
| 731 | 39,573.52 | 37,859 | 38,584.85 | 37,834.79 | 153,852.16 |
| 734 | 5,964.53 | 5,805.45 | 5,695.70 | 5,738.37 | 23,204.05 |
| 735 | 10,804.33 | 10,349.53 | 10,905.72 | 10,341.71 | 42,401.29 |
| 736 | 2,692.72 | 2,521.97 | 2,454.84 | 2,266.19 | 9,935.72 |
| 737 | 4,408.68 | 4,228.95 | 4,398.31 | 4,468.87 | 17,504.81 |
| 738 | 811.83 | 777.86 | 785.82 | 740.68 | 3,116.19 |
| 739 | 674.93 | 454.81 | 412.68 | 378.09 | 1,920.51 |
| 740 | 32,137.86 | 30,624.40 | 30,782.67 | 30,879.81 | 124,424.74 |
| 741 | 23,765.84 | 22,819.93 | 22,473.30 | 21,573.05 | 90,632.12 |
| 743 | 5,595.06 | 5,468.13 | 5,435.70 | 5,415.37 | 21,914.26 |
| 744 | 16,293.03 | 15,748.07 | 15,638.87 | 15,823.96 | 63,503.93 |
| 745 | 5,432.60 | 5,273.73 | 5,086.15 | 4,749.56 | 20,542.04 |
| 746 | 2,082.25 | 2,060.08 | 2,103.64 | 2,034.38 | 8,280.35 |
| 747 | 7,521.32 | 7,250.09 | 7,313.72 | 7,049.94 | 29,135.07 |
| 748 | 10,189.28 | 9,684.34 | 9,606.89 | 9,448.10 | 38,928.61 |
| 749 | 5,896.08 | 5,382.86 | 5,425.54 | 5,567.42 | 22,271.90 |
| TOTAL | 202,627.04 | 193,533.65 | 194,098.40 | 191,532.42 | 781,791.51 |

STATE: OREGON

| ZIP CODE | QUARTER 1 | QUARTER 2 | QUARTER 3 | QUARTER 4 | TOTAL GRAMS |
|---|---|---|---|---|---|
| 970 | 17,218.83 | 16,500.80 | 16,156.31 | 15,526.91 | 65,402.85 |
| 971 | 7,473.90 | 7,052.58 | 7,024.09 | 6,720.96 | 28,271.53 |
| 972 | 21,459.71 | 21,075.92 | 19,805.43 | 19,098.52 | 81,439.58 |
| 973 | 14,566.21 | 13,871.40 | 13,735.99 | 13,183.02 | 55,356.62 |
| 974 | 22,389.02 | 21,779.21 | 21,141.16 | 20,413.52 | 85,722.91 |
| 975 | 13,600.79 | 13,215.28 | 12,507.82 | 12,102.88 | 51,426.77 |
| 976 | 3,255.72 | 3,154.28 | 2,898.19 | 2,921.74 | 12,229.93 |
| 977 | 6,081.06 | 5,965.04 | 5,906.73 | 5,621.71 | 23,574.54 |
| 978 | 5,282.06 | 4,980.27 | 4,998.37 | 4,893.35 | 20,154.05 |
| 979 | 2,130.53 | 2,063.95 | 2,006.82 | 1,925.37 | 8,126.67 |
| TOTAL | 113,457.83 | 109,658.73 | 106,180.91 | 102,407.98 | 431,705.45 |

STATE: PENNSYLVANIA

| ZIP CODE | QUARTER 1 | QUARTER 2 | QUARTER 3 | QUARTER 4 | TOTAL GRAMS |
|---|---|---|---|---|---|
| 150 | 11,644.74 | 10,298.52 | 10,559.53 | 10,177.10 | 42,679.89 |
| 151 | 7,514.38 | 7,132.50 | 6,946.22 | 6,878.14 | 28,471.24 |
| 152 | 15,473.75 | 15,038.32 | 14,175.45 | 13,561.71 | 58,249.23 |
| 153 | 4,633.90 | 4,350.24 | 4,342.03 | 3,703.49 | 17,029.66 |
| 154 | 5,705.35 | 5,481.98 | 5,294.45 | 5,207.10 | 21,688.88 |
| 155 | 1,878.43 | 1,861.18 | 1,814.29 | 1,758.83 | 7,312.73 |
| 156 | 7,041.64 | 6,817.24 | 6,691.23 | 6,490.42 | 27,040.53 |
| 157 | 2,836.86 | 2,712.01 | 2,692.04 | 2,690.21 | 10,931.12 |
| 158 | 2,538.19 | 2,551.73 | 2,412.35 | 2,315.17 | 9,817.44 |
| 159 | 3,050.80 | 2,965.32 | 2,913.75 | 2,880.11 | 11,809.98 |
| 160 | 3,384.99 | 3,409.07 | 3,209.60 | 3,110.74 | 13,114.40 |
| 161 | 5,074.82 | 4,659.37 | 4,569.52 | 4,538.45 | 18,842.16 |
| 162 | 1,803.52 | 1,909.91 | 1,808.83 | 1,732.92 | 7,255.18 |
| 163 | 4,120.45 | 3,946.09 | 3,920.25 | 3,760.41 | 15,747.20 |
| 164 | 2,687.85 | 2,611.91 | 2,568.51 | 2,399.51 | 10,267.78 |
| 165 | 9,754.69 | 9,487.53 | 8,924.94 | 8,300.03 | 36,467.19 |
| 166 | 6,595.19 | 6,508.26 | 6,330.92 | 6,021.02 | 25,455.39 |

| | | | | | |
|---|---|---|---|---|---|
| 167 | 1,456.78 | 1,425.76 | 1,365.52 | 1,403.66 | 5,651.72 |
| 168 | 2,072.70 | 1,917.97 | 1,843.95 | 1,753.54 | 7,588.16 |
| 169 | 1,121.79 | 1,127.18 | 1,032.55 | 1,033.84 | 4,315.36 |
| 170 | 10,152.44 | 9,934.60 | 9,463.99 | 9,079.59 | 38,630.62 |

DRUG CODE:9193DRUG NAME:HYDROCODONE
STATE:PENNSYLVANIA

| ZIP CODE | QUARTER 1 | QUARTER 2 | QUARTER 3 | QUARTER 4 | TOTAL GRAMS |
|---|---|---|---|---|---|
| 171 | 2,621.31 | 2,526.43 | 2,410.83 | 2,247.16 | 9,805.73 |
| 172 | 1,935.41 | 1,795.20 | 1,700.28 | 1,670.59 | 7,101.48 |
| 173 | 3,348.49 | 3,140.92 | 3,036.91 | 2,924.51 | 12,450.83 |
| 174 | 1,959.02 | 1,910.12 | 1,830.82 | 1,770.65 | 7,470.61 |
| 175 | 3,406.50 | 3,273.18 | 3,216.31 | 3,059.58 | 12,955.57 |
| 176 | 2,705.44 | 2,574.47 | 2,454.96 | 2,403.74 | 10,138.61 |
| 177 | 3,748.23 | 3,595.58 | 3,450.94 | 3,349.69 | 14,144.44 |
| 178 | 2,533.89 | 2,330.25 | 2,270.26 | 2,287.95 | 9,422.35 |
| 179 | 2,283.70 | 2,209.50 | 2,112.24 | 1,953.49 | 8,558.93 |
| 180 | 3,969.41 | 3,701.88 | 3,589.33 | 3,504.19 | 14,764.81 |
| 181 | 1,742.59 | 1,660.35 | 1,541.64 | 1,587.71 | 6,532.29 |
| 182 | 2,661.70 | 2,555.06 | 2,519.87 | 2,439.40 | 10,176.03 |
| 183 | 1,996.56 | 1,980.40 | 1,814.43 | 1,847.74 | 7,639.13 |
| 184 | 3,491.37 | 3,542.97 | 3,355.25 | 3,292.27 | 13,681.86 |
| 185 | 5,046.08 | 4,827.61 | 4,693.29 | 4,559.18 | 19,126.16 |
| 186 | 3,806.39 | 3,669.57 | 3,560.62 | 3,409.76 | 14,446.34 |
| 187 | 10,413.36 | 9,970.57 | 9,562.87 | 9,178.88 | 39,125.68 |
| 188 | 1,342.47 | 1,354.36 | 1,258.67 | 1,254.02 | 5,209.52 |
| 189 | 3,004.80 | 2,911.02 | 2,788.92 | 2,776.92 | 11,481.66 |
| 190 | 12,497.99 | 11,845.10 | 12,040.62 | 11,370.04 | 47,753.75 |
| 191 | 9,425.88 | 9,009.46 | 8,427.35 | 8,253.29 | 35,115.98 |
| 193 | 3,284.42 | 3,135.87 | 3,080.36 | 2,995.79 | 12,496.44 |
| 194 | 5,120.72 | 4,934.71 | 4,805.67 | 4,622.86 | 19,483.96 |
| 195 | 2,039.55 | 2,042.05 | 1,960.80 | 1,930.66 | 7,973.06 |
| 196 | 2,543.21 | 2,427.12 | 2,320.44 | 2,172.16 | 9,462.93 |
| TOTAL | 207,471.75 | 199,070.44 | 192,683.60 | 185,658.22 | 784,884.01 |

STATE: PUERTO RICO

| ZIP CODE | QUARTER 1 | QUARTER 2 | QUARTER 3 | QUARTER 4 | TOTAL GRAMS |
|---|---|---|---|---|---|
| 6 | 520.22 | 410.65 | 380.29 | 409.6 | 1,720.76 |
| 7 | 413.13 | 363.04 | 314.77 | 326.37 | 1,417.31 |
| 9 | 380.38 | 304.38 | 295.82 | 292.36 | 1,272.94 |
| TOTAL | 1,313.73 | 1,078.07 | 990.88 | 1,028.33 | 4,411.01 |

STATE: RHODE ISLAND

| ZIP CODE | QUARTER 1 | QUARTER 2 | QUARTER 3 | QUARTER 4 | TOTAL GRAMS |
|---|---|---|---|---|---|
| 28 | 7,584.31 | 7,351.56 | 7,283.32 | 7,025.47 | 29,244.66 |
| 29 | 5,617.73 | 5,233.42 | 5,144.85 | 5,041.53 | 21,037.53 |
| TOTAL | 13,202.04 | 12,584.98 | 12,428.17 | 12,067 | 50,282.19 |

STATE: SOUTH CAROLINA

| ZIP CODE | QUARTER 1 | QUARTER 2 | QUARTER 3 | QUARTER 4 | TOTAL GRAMS |
|---|---|---|---|---|---|
| 290 | 8,833.85 | 8,094.82 | 8,115.84 | 8,188.92 | 33,233.43 |
| 291 | 9,802.01 | 9,253.57 | 9,049.55 | 9,096.11 | 37,201.24 |
| 292 | 7,280.32 | 6,835.47 | 6,631.38 | 6,851.04 | 27,598.21 |
| 293 | 25,748.51 | 24,108.24 | 24,581.87 | 24,141.70 | 98,580.32 |
| 294 | 22,801.19 | 21,549.21 | 20,937.59 | 21,109.68 | 86,397.67 |
| 295 | 26,973.34 | 25,954.72 | 25,140.10 | 25,132.11 | 103,200.27 |
| 296 | 45,204.70 | 42,437.20 | 42,195.66 | 41,650.26 | 171,487.82 |
| 297 | 8,161.95 | 7,649.95 | 7,381.91 | 7,562.18 | 30,755.99 |
| 298 | 4,452.97 | 3,975.28 | 4,065.87 | 4,050.20 | 16,544.32 |

| | | | | | | |
|---|---|---|---|---|---|---|
| | 299 | 3,413.81 | 3,117.71 | 3,024.35 | 3,131.38 | 12,687.25 |
| TOTAL | | 162,672.65 | 152,976.17 | 151,124.12 | 150,913.58 | 617,686.52 |

STATE: SOUTH DAKOTA

| ZIP CODE | | QUARTER 1 | QUARTER 2 | QUARTER 3 | QUARTER 4 | TOTAL GRAMS |
|---|---|---|---|---|---|---|
| | 570 | 2,661.21 | 2,599.55 | 2,500.50 | 2,490.28 | 10,251.54 |
| | 571 | 6,430.39 | 5,976.16 | 5,789.48 | 5,678.04 | 23,874.07 |
| | 572 | 1,137.51 | 1,133.83 | 1,090.88 | 1,107.51 | 4,469.73 |
| | 573 | 1,539.88 | 1,496.62 | 1,442.88 | 1,388.31 | 5,867.69 |
| | 574 | 1,093.78 | 1,087.58 | 1,012 | 1,003.16 | 4,196.52 |
| | 575 | 1,517.27 | 1,359.44 | 1,336.09 | 1,309.91 | 5,522.71 |
| | 576 | 305.11 | 288.4 | 287.57 | 214.15 | 1,095.23 |
| | 577 | 3,813.45 | 3,647.67 | 3,631.97 | 3,334.06 | 14,427.15 |
| TOTAL | | 18,498.60 | 17,589.25 | 17,091.37 | 16,525.42 | 69,704.64 |

STATE: TENNESSEE

| ZIP CODE | | QUARTER 1 | QUARTER 2 | QUARTER 3 | QUARTER 4 | TOTAL GRAMS |
|---|---|---|---|---|---|---|
| | 370 | 34,040.22 | 32,644.52 | 32,186.26 | 31,797.69 | 130,668.69 |
| | 371 | 31,176.05 | 29,827.96 | 29,476.63 | 29,128.60 | 119,609.24 |
| | 372 | 19,990.11 | 18,680.29 | 18,062.25 | 17,664.53 | 74,397.18 |

DRUG CODE:9193DRUG NAME:HYDROCODONE
STATE:TENNESSEE

| ZIP CODE | | QUARTER 1 | QUARTER 2 | QUARTER 3 | QUARTER 4 | TOTAL GRAMS |
|---|---|---|---|---|---|---|
| | 373 | 31,389.98 | 30,157.93 | 29,820.67 | 29,155.24 | 120,523.82 |
| | 374 | 11,555.39 | 11,114.73 | 11,009.99 | 10,617.18 | 44,297.29 |
| | 376 | 22,683.77 | 21,876.58 | 21,776.26 | 19,899.70 | 86,236.31 |
| | 377 | 15,374.56 | 14,898.94 | 15,029.82 | 14,684.88 | 59,988.20 |
| | 378 | 25,692.50 | 24,484.55 | 23,772.93 | 23,420.41 | 97,370.39 |
| | 379 | 12,734.26 | 11,944.27 | 11,825.88 | 11,759.38 | 48,263.79 |
| | 380 | 15,168.15 | 14,069.86 | 13,822.94 | 14,008.20 | 57,069.15 |
| | 381 | 20,918.58 | 19,690.51 | 19,633.66 | 17,872.64 | 78,115.39 |
| | 382 | 8,737.28 | 8,659.05 | 8,531.15 | 8,151.34 | 34,078.82 |
| | 383 | 18,286.83 | 17,621.81 | 17,135.57 | 17,107.79 | 70,152 |
| | 384 | 10,338.32 | 9,912.76 | 9,722.09 | 9,396.23 | 39,369.40 |
| | 385 | 9,399.50 | 9,120.66 | 8,926.05 | 9,215.55 | 36,661.76 |
| TOTAL | | 287,485.50 | 274,704.42 | 270,732.15 | 263,879.36 | 1,096,801.43 |

STATE: TEXAS

| ZIP CODE | | QUARTER 1 | QUARTER 2 | QUARTER 3 | QUARTER 4 | TOTAL GRAMS |
|---|---|---|---|---|---|---|
| | 750 | 59,939.84 | 58,127.87 | 58,029.48 | 58,160.14 | 234,257.33 |
| | 751 | 24,795.14 | 23,843.78 | 23,973.56 | 23,739.52 | 96,352 |
| | 752 | 32,810.92 | 31,422.44 | 31,976.69 | 30,504 | 126,714.05 |
| | 753 | 245.2 | 264.63 | 251.96 | 297.8 | 1,059.59 |
| | 754 | 17,333.86 | 17,524.91 | 17,133.71 | 16,665.60 | 68,658.08 |
| | 755 | 5,820.57 | 5,696.23 | 5,606.28 | 5,732.66 | 22,855.74 |
| | 756 | 12,263.99 | 11,935.37 | 11,879.73 | 11,715.24 | 47,794.33 |
| | 757 | 16,294.76 | 15,990.83 | 15,594.65 | 15,976.37 | 63,856.61 |
| | 758 | 3,392.03 | 3,188.70 | 3,044.44 | 3,040.96 | 12,666.13 |
| | 759 | 12,093.80 | 10,743.60 | 10,132.54 | 10,147.44 | 43,117.38 |
| | 760 | 42,865.45 | 41,525.01 | 41,677.83 | 41,851.45 | 167,919.74 |
| | 761 | 36,945.55 | 36,499.30 | 37,066.92 | 35,663.33 | 146,175.10 |
| | 762 | 15,292.46 | 14,883.64 | 15,274.69 | 15,141.06 | 60,591.85 |
| | 763 | 7,898.32 | 7,995.34 | 7,751.26 | 7,607.11 | 31,252.03 |
| | 764 | 5,118.89 | 5,079.53 | 5,021.31 | 4,885.74 | 20,105.47 |
| | 765 | 16,341.70 | 15,920.64 | 15,649.52 | 15,172.86 | 63,084.72 |
| | 766 | 3,713.55 | 3,613.08 | 3,602.54 | 3,626.49 | 14,555.66 |
| | 767 | 11,343.77 | 11,296.15 | 10,740.49 | 10,293.66 | 43,674.07 |
| | 768 | 3,528.97 | 3,559.67 | 3,660.53 | 3,692.25 | 14,441.42 |

| | | | | | |
|---|---|---|---|---|---|
| 769 | 4,569.09 | 4,220.43 | 4,305.68 | 4,125.09 | 17,220.29 |
| 770 | 62,998.74 | 58,140.47 | 57,622.90 | 55,110.83 | 233,872.94 |
| 772 | 2.72 | 4.51 | 5.6 | 0 | 12.83 |
| 773 | 34,955.01 | 33,316.74 | 33,440.43 | 32,675.65 | 134,387.83 |
| 774 | 22,741.14 | 20,823.12 | 20,714.96 | 20,562.08 | 84,841.30 |
| 775 | 37,439.48 | 35,660.44 | 35,573.73 | 34,645.91 | 143,319.56 |
| 776 | 10,670.30 | 10,191.60 | 10,162.23 | 10,230.54 | 41,254.67 |
| 777 | 5,121.41 | 4,812.90 | 4,875.21 | 4,891.95 | 19,701.47 |
| 778 | 7,801.92 | 7,540.85 | 7,570.40 | 7,190.97 | 30,104.14 |
| 779 | 8,271.88 | 8,158.83 | 8,309.48 | 8,050.35 | 32,790.54 |
| 780 | 10,459.21 | 10,091.86 | 10,016.18 | 9,730.19 | 40,297.44 |
| 781 | 11,689.19 | 10,938.79 | 11,063.84 | 10,947.44 | 44,639.26 |
| 782 | 50,303.70 | 48,802.84 | 48,597.14 | 48,951.55 | 196,655.23 |
| 783 | 6,716.58 | 6,583.63 | 6,738.17 | 6,756.46 | 26,794.84 |
| 784 | 13,994.42 | 13,669.72 | 13,710.23 | 13,024.49 | 54,398.86 |
| 785 | 9,945.29 | 9,116.55 | 8,998.36 | 8,925.05 | 36,985.25 |
| 786 | 26,111.77 | 25,142.44 | 25,131.16 | 25,397.33 | 101,782.70 |
| 787 | 24,391.27 | 23,919.49 | 23,790.80 | 23,872.68 | 95,974.24 |
| 788 | 3,156.70 | 3,018.06 | 3,039.92 | 2,977.06 | 12,191.74 |
| 789 | 1,778.74 | 1,680.26 | 1,790.74 | 1,749.04 | 6,998.78 |
| 790 | 4,795.16 | 4,722.25 | 4,774.15 | 4,686.43 | 18,977.99 |
| 791 | 9,270.98 | 9,093.59 | 8,999.89 | 8,610.13 | 35,974.59 |
| 792 | 1,028.58 | 992.44 | 1,025.81 | 1,066.78 | 4,113.61 |
| 793 | 4,008.54 | 3,603.25 | 3,809.85 | 3,818.22 | 15,239.86 |
| 794 | 11,485.89 | 11,080.98 | 11,277.63 | 11,480.16 | 45,324.66 |
| 795 | 3,384.27 | 3,230.63 | 3,139.53 | 3,162.66 | 12,917.09 |
| 796 | 6,756.80 | 6,335.73 | 6,455.60 | 6,372.87 | 25,921 |
| 797 | 14,059.24 | 13,551.79 | 12,875.06 | 12,714.48 | 53,200.57 |
| 798 | 356.4 | 355.39 | 409.21 | 368.62 | 1,489.62 |
| 799 | 10,127.52 | 9,443.99 | 9,547.46 | 9,039.03 | 38,158 |
| TOTAL | 746,430.71 | 717,354.29 | 715,839.48 | 705,047.72 | 2,884,672.20 |

DRUG CODE:9193DRUG NAME:HYDROCODONE
STATE: UTAH

| ZIP CODE | QUARTER 1 | QUARTER 2 | QUARTER 3 | QUARTER 4 | TOTAL GRAMS |
|---|---|---|---|---|---|
| 840 | 24,811.63 | 23,608.25 | 23,911.59 | 23,276.82 | 95,608.29 |
| 841 | 18,666.94 | 17,868.29 | 17,157.76 | 16,232.59 | 69,925.58 |
| 843 | 3,266.98 | 3,067.78 | 3,051.82 | 3,131.71 | 12,518.29 |
| 844 | 5,139.52 | 4,769.85 | 4,802.35 | 4,608.46 | 19,320.18 |
| 845 | 2,005.19 | 2,097.59 | 1,918.03 | 1,861.84 | 7,882.65 |
| 846 | 6,136.18 | 5,791.30 | 5,788.96 | 5,667.56 | 23,384 |
| 847 | 8,319.40 | 8,030.96 | 8,053.27 | 7,980.50 | 32,384.13 |
| TOTAL | 68,345.84 | 65,234.02 | 64,683.78 | 62,759.48 | 261,023.12 |

STATE: VERMONT

| ZIP CODE | QUARTER 1 | QUARTER 2 | QUARTER 3 | QUARTER 4 | TOTAL GRAMS |
|---|---|---|---|---|---|
| 50 | 448.41 | 446.42 | 430.19 | 400.42 | 1,725.44 |
| 51 | 269.74 | 230.56 | 210.81 | 244.67 | 955.78 |
| 52 | 326.27 | 333.24 | 306.38 | 285.7 | 1,251.59 |
| 53 | 381.57 | 350.5 | 359.06 | 363.47 | 1,454.60 |
| 54 | 2,376.60 | 2,297.14 | 2,256.02 | 2,229.30 | 9,159.06 |
| 56 | 1,094.72 | 1,076.44 | 984.52 | 937.11 | 4,092.79 |
| 57 | 558.05 | 546.49 | 511.35 | 533.93 | 2,149.82 |
| 58 | 675.39 | 654.38 | 617.15 | 603 | 2,549.92 |
| TOTAL | 6,130.75 | 5,935.17 | 5,675.48 | 5,597.60 | 23,339 |

STATE: VIRGIN ISLANDS

| ZIP CODE | QUARTER 1 | QUARTER 2 | QUARTER 3 | QUARTER 4 | TOTAL GRAMS |
|---|---|---|---|---|---|
| 8 | 436.02 | 363.59 | 313.07 | 275.43 | 1,388.11 |

| | | | | | |
|---|---|---|---|---|---|
| TOTAL | 436.02 | 363.59 | 313.07 | 275.43 | 1,388.11 |

STATE: VIRGINIA

| ZIP CODE | QUARTER 1 | QUARTER 2 | QUARTER 3 | QUARTER 4 | TOTAL GRAMS |
|---|---|---|---|---|---|
| 201 | 5,488.18 | 4,916.85 | 4,821.94 | 5,116.88 | 20,343.85 |
| 220 | 2,435.10 | 2,292.47 | 2,154.19 | 2,189.55 | 9,071.31 |
| 221 | 2,079.38 | 1,998.40 | 1,891.07 | 1,870.58 | 7,839.43 |
| 222 | 717.35 | 686.44 | 610.49 | 631.93 | 2,646.21 |
| 223 | 1,308.02 | 1,146.04 | 1,059.44 | 1,148.84 | 4,662.34 |
| 224 | 3,334.22 | 3,114.05 | 2,917.08 | 3,073.52 | 12,438.87 |
| 225 | 1,452.62 | 1,347.45 | 1,260.19 | 1,286.29 | 5,346.55 |
| 226 | 3,324.36 | 3,114.84 | 2,945.71 | 3,023.79 | 12,408.70 |
| 227 | 703.39 | 659.37 | 589.32 | 580.9 | 2,532.98 |
| 228 | 2,684.28 | 2,508.50 | 2,472.40 | 2,539.41 | 10,204.59 |
| 229 | 3,221.75 | 3,089.62 | 2,971.49 | 2,874.90 | 12,157.76 |
| 230 | 3,022.21 | 2,822.70 | 2,845.14 | 2,704.36 | 11,394.41 |
| 231 | 5,415.48 | 4,922.77 | 4,863.12 | 4,821.81 | 20,023.18 |
| 232 | 8,592.09 | 8,095.74 | 7,940.75 | 7,806.42 | 32,435 |
| 233 | 3,585.15 | 3,297.69 | 3,239.20 | 3,238.56 | 13,360.60 |
| 234 | 7,931.95 | 7,241.28 | 7,121.87 | 7,182.15 | 29,477.25 |
| 235 | 3,027.29 | 2,906.35 | 2,676.34 | 2,723.45 | 11,333.43 |
| 236 | 5,635.05 | 5,015.80 | 4,814.96 | 4,811.61 | 20,277.42 |
| 237 | 3,001.84 | 2,709.74 | 2,640.35 | 2,662 | 11,013.93 |
| 238 | 4,335.67 | 4,017.68 | 3,845.27 | 3,929.56 | 16,128.18 |
| 239 | 1,661.08 | 1,594.45 | 1,533.58 | 1,457.65 | 6,246.76 |
| 240 | 10,456.75 | 9,774.25 | 9,691.85 | 9,710.67 | 39,633.52 |
| 241 | 11,341.37 | 10,814.49 | 10,549.13 | 10,065.59 | 42,770.58 |
| 242 | 15,261.29 | 14,972.52 | 15,091.84 | 14,620.97 | 59,946.62 |
| 243 | 6,077.37 | 6,067.22 | 5,694.58 | 5,442.35 | 23,281.52 |
| 244 | 3,252.32 | 3,108.68 | 2,959.42 | 2,996.68 | 12,317.10 |
| 245 | 8,939.38 | 8,457.85 | 8,223.86 | 7,951.89 | 33,572.98 |
| 246 | 4,900.93 | 4,928.29 | 4,745.47 | 4,873.55 | 19,448.24 |
| TOTAL | 133,185.87 | 125,621.53 | 122,170.05 | 121,335.86 | 502,313.31 |

STATE: WASHINGTON

| ZIP CODE | QUARTER 1 | QUARTER 2 | QUARTER 3 | QUARTER 4 | TOTAL GRAMS |
|---|---|---|---|---|---|
| 980 | 19,643.02 | 18,729.87 | 18,507.37 | 18,406.69 | 75,286.95 |
| 981 | 10,858.58 | 10,177.14 | 10,001.45 | 9,044.47 | 40,081.64 |
| 982 | 18,138.29 | 17,586.67 | 16,839.59 | 16,551.50 | 69,116.05 |
| 983 | 14,106.38 | 13,946.29 | 13,945.54 | 13,611.70 | 55,609.91 |
| 984 | 7,908.76 | 7,733.48 | 7,570.55 | 7,320.57 | 30,533.36 |
| 985 | 10,794.95 | 10,259.75 | 10,091.79 | 9,348.37 | 40,494.86 |
| 986 | 14,176.34 | 13,473.70 | 13,230.22 | 13,409.87 | 54,290.13 |
| 988 | 5,235.65 | 5,138.25 | 5,054.48 | 4,956.96 | 20,385.34 |
| 989 | 6,768.62 | 6,640.62 | 6,506.95 | 6,135.64 | 26,051.83 |
| 990 | 3,856.98 | 3,880.52 | 3,772.06 | 3,564.01 | 15,073.57 |

DRUG CODE:9193DRUG NAME:HYDROCODONE
STATE:WASHINGTON

| ZIP CODE | QUARTER 1 | QUARTER 2 | QUARTER 3 | QUARTER 4 | TOTAL GRAMS |
|---|---|---|---|---|---|
| 991 | 5,223.39 | 5,156.77 | 5,110.52 | 4,818.77 | 20,309.45 |
| 992 | 21,277.47 | 21,312.63 | 20,883.09 | 20,346.75 | 83,819.94 |
| 993 | 13,006.95 | 12,768.56 | 12,193.38 | 11,880.45 | 49,849.34 |
| 994 | 2,277.62 | 2,270.58 | 2,158.52 | 2,253.18 | 8,959.90 |
| TOTAL | 153,273 | 149,074.83 | 145,865.51 | 141,648.93 | 589,862.27 |

STATE: WEST VIRGINIA

| ZIP CODE | QUARTER 1 | QUARTER 2 | QUARTER 3 | QUARTER 4 | TOTAL GRAMS |
|---|---|---|---|---|---|
| 247 | 2,689.37 | 2,573.32 | 2,401.52 | 2,338.22 | 10,002.43 |
| 248 | 1,976.78 | 1,875.55 | 1,744.93 | 1,685.94 | 7,283.20 |
| 249 | 1,475.45 | 1,449.68 | 1,358.69 | 1,386.58 | 5,670.40 |

| | | | | | |
|---|---|---|---|---|---|
| 250 | 2,981.59 | 2,945.04 | 2,838.08 | 2,736.94 | 11,501.65 |
| 251 | 2,491.04 | 2,435.24 | 2,394.28 | 2,284.97 | 9,605.53 |
| 252 | 1,942.57 | 1,865.70 | 1,735.75 | 1,674.10 | 7,218.12 |
| 253 | 7,049.76 | 6,641.24 | 6,789.59 | 6,337.16 | 26,817.75 |
| 254 | 2,883.37 | 2,837.40 | 2,649.81 | 2,636.71 | 11,007.29 |
| 255 | 6,246.06 | 5,977.64 | 5,845.25 | 5,619.55 | 23,688.50 |
| 256 | 3,705.09 | 3,685.31 | 3,503.19 | 3,354.94 | 14,248.53 |
| 257 | 6,087.02 | 5,826.92 | 5,482.75 | 5,316.36 | 22,713.05 |
| 258 | 6,250.10 | 5,258.68 | 5,208.99 | 4,619.26 | 21,337.03 |
| 259 | 1,937.33 | 1,884 | 1,773.19 | 1,674.10 | 7,268.62 |
| 260 | 4,310.79 | 4,079.18 | 3,992.61 | 4,058.86 | 16,441.44 |
| 261 | 5,101.01 | 4,688.40 | 4,486.80 | 4,283.07 | 18,559.28 |
| 262 | 2,742.19 | 2,671.98 | 2,547.94 | 2,491.66 | 10,453.77 |
| 263 | 6,240.46 | 6,017.20 | 5,550.52 | 5,709.21 | 23,517.39 |
| 264 | 1,548.19 | 1,492.25 | 1,469.88 | 1,371.79 | 5,882.11 |
| 265 | 5,680.23 | 5,198.83 | 4,953.79 | 4,721.02 | 20,553.87 |
| 266 | 1,468.46 | 1,361.02 | 1,373.22 | 1,297.66 | 5,500.36 |
| 267 | 1,057.93 | 1,059.30 | 998.71 | 975.97 | 4,091.91 |
| 268 | 554.61 | 531.28 | 469.9 | 448.02 | 2,003.81 |
| TOTAL | 76,419.40 | 72,355.16 | 69,569.39 | 67,022.09 | 285,366.04 |

STATE: WISCONSIN

| ZIP CODE | QUARTER 1 | QUARTER 2 | QUARTER 3 | QUARTER 4 | TOTAL GRAMS |
|---|---|---|---|---|---|
| 530 | 8,523.26 | 8,087.40 | 7,997.40 | 8,132.72 | 32,740.78 |
| 531 | 12,240.04 | 11,560.21 | 11,332.11 | 11,237.95 | 46,370.31 |
| 532 | 12,870.35 | 12,216.57 | 12,258.87 | 12,140.17 | 49,485.96 |
| 534 | 3,298.21 | 2,987.30 | 2,903.22 | 2,825.57 | 12,014.30 |
| 535 | 9,588.28 | 9,126.29 | 8,578.85 | 8,265.65 | 35,559.07 |
| 537 | 4,929.16 | 4,584.26 | 4,627.71 | 4,375.08 | 18,516.21 |
| 538 | 857.3 | 872.47 | 828.53 | 813.8 | 3,372.10 |
| 539 | 4,627.23 | 4,526.06 | 4,392.22 | 4,053.75 | 17,599.26 |
| 540 | 1,075.89 | 1,033.73 | 997.07 | 982.74 | 4,089.43 |
| 541 | 3,129.51 | 2,993.39 | 2,902.18 | 2,793.59 | 11,818.67 |
| 542 | 1,758.40 | 1,668.63 | 1,646.77 | 1,589.42 | 6,663.22 |
| 543 | 3,296.74 | 3,150 | 3,077.79 | 3,062.59 | 12,587.12 |
| 544 | 6,481.25 | 6,261.86 | 6,011.56 | 6,099.76 | 24,854.43 |
| 545 | 1,654.26 | 1,669.84 | 1,784.51 | 1,591.62 | 6,700.23 |
| 546 | 5,549.02 | 5,340.79 | 5,426.03 | 5,137.51 | 21,453.35 |
| 547 | 3,668.11 | 3,584.21 | 3,569.82 | 3,532.75 | 14,354.89 |
| 548 | 3,791.16 | 3,674.47 | 3,678.26 | 3,465.66 | 14,609.55 |
| 549 | 7,923.30 | 7,773.89 | 7,585.56 | 7,301.79 | 30,584.54 |
| TOTAL | 95,261.47 | 91,111.37 | 89,598.46 | 87,402.12 | 363,373.42 |

STATE: WYOMING

| ZIP CODE | QUARTER 1 | QUARTER 2 | QUARTER 3 | QUARTER 4 | TOTAL GRAMS |
|---|---|---|---|---|---|
| 820 | 2,807.48 | 2,621.19 | 2,625.84 | 2,628.47 | 10,682.98 |
| 822 | 503.45 | 501.75 | 488.61 | 471.45 | 1,965.26 |
| 823 | 575.17 | 656.76 | 652.91 | 602.81 | 2,487.65 |
| 824 | 1,499.31 | 1,501.55 | 1,357.54 | 1,349.15 | 5,707.55 |
| 825 | 911.43 | 927.79 | 854.64 | 822.64 | 3,516.50 |
| 826 | 2,655.30 | 2,333.77 | 2,293.16 | 2,408.09 | 9,690.32 |
| 827 | 965.55 | 889.91 | 876.09 | 868.52 | 3,600.07 |
| 828 | 1,517.81 | 1,393.01 | 1,365.32 | 1,253.53 | 5,529.67 |
| 829 | 1,938.25 | 1,886.29 | 1,888.08 | 1,832.10 | 7,544.72 |
| 830 | 312.7 | 272.81 | 283.34 | 241.02 | 1,109.87 |
| 831 | 495.31 | 527.9 | 493.22 | 496.96 | 2,013.39 |
| TOTAL | 14,181.76 | 13,512.73 | 13,178.75 | 12,974.74 | 53,847.98 |

DRUG CODE:9220LDRUG NAME:LEVORPHANOL
STATE: ALABAMA

| ZIP CODE | QUARTER 1 | QUARTER 2 | QUARTER 3 | QUARTER 4 | TOTAL GRAMS |
|---|---|---|---|---|---|
| 350 | 0 | 0 | 0.46 | 0.81 | 1.27 |
| 352 | 0.23 | 0.35 | 0.46 | 0.7 | 1.74 |
| 354 | 1.74 | 1.63 | 1.04 | 1.04 | 5.45 |
| 360 | 0.12 | 0 | 0 | 0 | 0.12 |
| 361 | 3.37 | 1.74 | 2.09 | 0.81 | 8.01 |
| 363 | 0.35 | 0 | 0 | 0 | 0.35 |
| 364 | 0.81 | 0.7 | 0.58 | 0.7 | 2.79 |
| 368 | 0.12 | 0 | 0 | 0 | 0.12 |
| TOTAL | 6.74 | 4.42 | 4.63 | 4.06 | 19.85 |

STATE: ARIZONA

| ZIP CODE | QUARTER 1 | QUARTER 2 | QUARTER 3 | QUARTER 4 | TOTAL GRAMS |
|---|---|---|---|---|---|
| 850 | 0.46 | 1.28 | 3.37 | 15.09 | 20.2 |
| 851 | 0 | 0 | 0 | 0.23 | 0.23 |
| 852 | 0.7 | 3.37 | 6.15 | 6.96 | 17.18 |
| 853 | 0.23 | 0 | 0 | 0 | 0.23 |
| 855 | 0 | 0.12 | 0 | 0 | 0.12 |
| 857 | 0 | 0.35 | 0.46 | 0.46 | 1.27 |
| 860 | 0.35 | 0 | 0 | 0 | 0.35 |
| TOTAL | 1.74 | 5.12 | 9.98 | 22.74 | 39.58 |

STATE: ARKANSAS

| ZIP CODE | QUARTER 1 | QUARTER 2 | QUARTER 3 | QUARTER 4 | TOTAL GRAMS |
|---|---|---|---|---|---|
| 719 | 0.46 | 1.97 | 2.67 | 2.44 | 7.54 |
| 722 | 0.12 | 0 | 0 | 0 | 0.12 |
| 726 | 0 | 0 | 0.23 | 0 | 0.23 |
| TOTAL | 0.58 | 1.97 | 2.9 | 2.44 | 7.89 |

STATE: CALIFORNIA

| ZIP CODE | QUARTER 1 | QUARTER 2 | QUARTER 3 | QUARTER 4 | TOTAL GRAMS |
|---|---|---|---|---|---|
| 900 | 9.52 | 6.5 | 2.9 | 1.28 | 20.2 |
| 902 | 0.46 | 0.35 | 0.7 | 1.51 | 3.02 |
| 904 | 5.34 | 13.12 | 35.64 | 11.96 | 66.06 |
| 905 | 0.35 | 0 | 0 | 0 | 0.35 |
| 906 | 0 | 0.12 | 0 | 0 | 0.12 |
| 907 | 0 | 0.23 | 0 | 0 | 0.23 |
| 908 | 0 | 0 | 0.23 | 0 | 0.23 |
| 911 | 0.46 | 0.7 | 0.58 | 0.93 | 2.67 |
| 913 | 3.37 | 3.83 | 5.34 | 3.02 | 15.56 |
| 914 | 0.12 | 0.12 | 0.58 | 0.46 | 1.28 |
| 915 | 0.58 | 0 | 0 | 0 | 0.58 |
| 917 | 18.8 | 34.59 | 32.27 | 40.74 | 126.4 |
| 918 | 0 | 0 | 0.12 | 0 | 0.12 |
| 919 | 0.58 | 0.58 | 0.58 | 1.63 | 3.37 |
| 920 | 3.02 | 2.44 | 1.74 | 4.53 | 11.73 |
| 921 | 4.64 | 2.32 | 3.48 | 4.88 | 15.32 |
| 923 | 0.23 | 0.81 | 1.04 | 3.13 | 5.21 |
| 924 | 3.71 | 11.26 | 10.91 | 1.74 | 27.62 |
| 925 | 0.35 | 0.23 | 0.46 | 0 | 1.04 |
| 926 | 0.35 | 0.58 | 0.46 | 1.39 | 2.78 |
| 927 | 0 | 0.12 | 0 | 0 | 0.12 |
| 928 | 0.23 | 0.23 | 0 | 0 | 0.46 |
| 930 | 0.12 | 0.7 | 1.28 | 0 | 2.1 |
| 931 | 0.58 | 0 | 0.58 | 0.58 | 1.74 |
| 932 | 0 | 0.12 | 0 | 0 | 0.12 |
| 933 | 0.12 | 0 | 0.12 | 0.23 | 0.47 |
| 934 | 0.23 | 0.46 | 0.58 | 0 | 1.27 |
| 935 | 0.23 | 0.12 | 0.81 | 0.46 | 1.62 |
| 936 | 1.51 | 0.93 | 0.35 | 0.81 | 3.6 |
| 937 | 2.09 | 0.35 | 0.46 | 0.93 | 3.83 |

| | | | | | |
|---|---|---|---|---|---|
| 939 | 0.58 | 0.7 | 0.35 | 0.46 | 2.09 |
| 940 | 1.97 | 0.81 | 0.93 | 0.7 | 4.41 |
| 941 | 2.44 | 2.67 | 1.51 | 1.04 | 7.66 |
| 943 | 0 | 0.12 | 0 | 0.23 | 0.35 |
| 945 | 4.18 | 2.79 | 3.13 | 1.86 | 11.96 |
| 949 | 0.81 | 1.16 | 1.28 | 0.93 | 4.18 |
| 950 | 7.31 | 1.97 | 5.92 | 2.32 | 17.52 |
| 951 | 0.46 | 0.23 | 0.23 | 0 | 0.92 |
| 953 | 0.81 | 0.35 | 0.46 | 0.35 | 1.97 |

DRUG CODE:9220LDRUG NAME:LEVORPHANOL
STATE:CALIFORNIA

| ZIP CODE | QUARTER 1 | QUARTER 2 | QUARTER 3 | QUARTER 4 | TOTAL GRAMS |
|---|---|---|---|---|---|
| 954 | 2.09 | 1.74 | 1.63 | 1.63 | 7.09 |
| 955 | 0.35 | 0 | 0 | 0 | 0.35 |
| 956 | 2.9 | 0 | 3.6 | 0.7 | 7.2 |
| 958 | 1.63 | 2.44 | 1.74 | 5.57 | 11.38 |
| 961 | 0 | 0 | 0 | 0.23 | 0.23 |
| TOTAL | 82.52 | 95.79 | 121.99 | 96.23 | 396.53 |

STATE: COLORADO

| ZIP CODE | QUARTER 1 | QUARTER 2 | QUARTER 3 | QUARTER 4 | TOTAL GRAMS |
|---|---|---|---|---|---|
| 801 | 0.58 | 0.46 | 0 | 0 | 1.04 |
| 802 | 1.28 | 4.99 | 5.22 | 5.11 | 16.6 |
| 803 | 0 | 0 | 0 | 0.12 | 0.12 |
| 805 | 0 | 0.23 | 0 | 0.7 | 0.93 |
| 815 | 0.35 | 0.46 | 0 | 0 | 0.81 |
| TOTAL | 2.21 | 6.14 | 5.22 | 5.93 | 19.5 |

STATE: CONNECTICUT

| ZIP CODE | QUARTER 1 | QUARTER 2 | QUARTER 3 | QUARTER 4 | TOTAL GRAMS |
|---|---|---|---|---|---|
| 60 | 0.23 | 0.35 | 0.81 | 0 | 1.39 |
| 61 | 0 | 1.16 | 0.58 | 0 | 1.74 |
| 64 | 0.12 | 0.35 | 0 | 0 | 0.47 |
| 65 | 0 | 0 | 0 | 0.12 | 0.12 |
| 67 | 0 | 0.23 | 0.46 | 0.12 | 0.81 |
| 68 | 0 | 0.12 | 0 | 0.7 | 0.82 |
| TOTAL | 0.35 | 2.21 | 1.85 | 0.94 | 5.35 |

STATE: FLORIDA

| ZIP CODE | QUARTER 1 | QUARTER 2 | QUARTER 3 | QUARTER 4 | TOTAL GRAMS |
|---|---|---|---|---|---|
| 327 | 6.04 | 18.57 | 23.56 | 18.22 | 66.39 |
| 328 | 0 | 0 | 0 | 0.12 | 0.12 |
| 330 | 0 | 0.46 | 0.23 | 0.12 | 0.81 |
| 331 | 0.12 | 0 | 0 | 0 | 0.12 |
| 334 | 1.16 | 1.74 | 0.23 | 0 | 3.13 |
| 335 | 0 | 0 | 0.46 | 0.46 | 0.92 |
| 337 | 0 | 0.12 | 0 | 0 | 0.12 |
| 338 | 0.12 | 0.12 | 0.12 | 0.58 | 0.94 |
| 339 | 0 | 0 | 0.23 | 0.35 | 0.58 |
| 342 | 0.46 | 0.93 | 2.21 | 2.32 | 5.92 |
| 346 | 0.12 | 0 | 0 | 0 | 0.12 |
| TOTAL | 8.02 | 21.94 | 27.04 | 22.17 | 79.17 |

STATE: GEORGIA

| ZIP CODE | QUARTER 1 | QUARTER 2 | QUARTER 3 | QUARTER 4 | TOTAL GRAMS |
|---|---|---|---|---|---|
| 300 | 0 | 0 | 0.12 | 0 | 0.12 |
| 301 | 0.12 | 0.35 | 0.35 | 0.35 | 1.17 |

| ZIP CODE | QUARTER 1 | QUARTER 2 | QUARTER 3 | QUARTER 4 | TOTAL GRAMS |
|---|---|---|---|---|---|
| 303 | 0.81 | 0.81 | 0.7 | 0.81 | 3.13 |
| 317 | 0 | 0 | 0.12 | 0 | 0.12 |
| TOTAL | 0.93 | 1.16 | 1.29 | 1.16 | 4.54 |

STATE: ILLINOIS

| ZIP CODE | QUARTER 1 | QUARTER 2 | QUARTER 3 | QUARTER 4 | TOTAL GRAMS |
|---|---|---|---|---|---|
| 600 | 0 | 0.58 | 0.46 | 0.35 | 1.39 |
| 601 | 0.81 | 0.81 | 1.28 | 1.97 | 4.87 |
| 604 | 0.46 | 0.81 | 0.23 | 0.81 | 2.31 |
| 605 | 1.74 | 1.28 | 1.04 | 1.16 | 5.22 |
| 606 | 1.28 | 0.23 | 0 | 0 | 1.51 |
| 620 | 2.09 | 2.21 | 2.9 | 0.12 | 7.32 |
| 622 | 0 | 0 | 0 | 3.48 | 3.48 |
| 627 | 0.58 | 0.35 | 0.46 | 0.58 | 1.97 |
| 628 | 0.23 | 0.12 | 0.35 | 0.23 | 0.93 |
| TOTAL | 7.19 | 6.39 | 6.72 | 8.7 | 29 |

STATE: INDIANA

| ZIP CODE | QUARTER 1 | QUARTER 2 | QUARTER 3 | QUARTER 4 | TOTAL GRAMS |
|---|---|---|---|---|---|
| 460 | 1.04 | 0.35 | 0.7 | 0 | 2.09 |
| 461 | 0 | 0.12 | 0 | 0.12 | 0.24 |
| 462 | 1.63 | 1.04 | 3.48 | 0.7 | 6.85 |
| 465 | 0 | 0 | 0 | 0.12 | 0.12 |
| 467 | 10.91 | 11.96 | 12.54 | 10.68 | 46.09 |
| 468 | 1.51 | 1.39 | 1.51 | 1.97 | 6.38 |
| 469 | 5.57 | 4.99 | 6.04 | 5.22 | 21.82 |
| 471 | 0 | 0 | 0 | 1.28 | 1.28 |

DRUG CODE:9220LDRUG NAME:LEVORPHANOL
STATE:INDIANA

| ZIP CODE | QUARTER 1 | QUARTER 2 | QUARTER 3 | QUARTER 4 | TOTAL GRAMS |
|---|---|---|---|---|---|
| 472 | 0.35 | 0.23 | 0.23 | 0.46 | 1.27 |
| 473 | 3.71 | 3.37 | 1.51 | 2.55 | 11.14 |
| 479 | 0.93 | 0.93 | 1.16 | 0.93 | 3.95 |
| TOTAL | 25.65 | 24.38 | 27.17 | 24.03 | 101.23 |

STATE: IOWA

| ZIP CODE | QUARTER 1 | QUARTER 2 | QUARTER 3 | QUARTER 4 | TOTAL GRAMS |
|---|---|---|---|---|---|
| 500 | 0 | 0 | 0.12 | 0 | 0.12 |
| 501 | 0 | 0 | 0.12 | 0 | 0.12 |
| 502 | 0.46 | 0.12 | 0 | 0 | 0.58 |
| 503 | 0.12 | 0 | 0.12 | 0.12 | 0.36 |
| 506 | 0.23 | 0.35 | 0.7 | 0.7 | 1.98 |
| 507 | 3.48 | 2.79 | 1.86 | 1.28 | 9.41 |
| 520 | 1.04 | 0.93 | 0.93 | 1.39 | 4.29 |
| 524 | 0.35 | 0.7 | 0.35 | 0.81 | 2.21 |
| TOTAL | 5.68 | 4.89 | 4.2 | 4.3 | 19.07 |

STATE: KANSAS

| ZIP CODE | QUARTER 1 | QUARTER 2 | QUARTER 3 | QUARTER 4 | TOTAL GRAMS |
|---|---|---|---|---|---|
| 660 | 1.39 | 0.81 | 1.04 | 2.44 | 5.68 |
| 662 | 4.88 | 6.38 | 7.66 | 7.31 | 26.23 |
| 670 | 0.23 | 0 | 0 | 0 | 0.23 |
| 672 | 0.12 | 0.23 | 0 | 0 | 0.35 |
| 675 | 0 | 0.23 | 0 | 0 | 0.23 |
| TOTAL | 6.62 | 7.65 | 8.7 | 9.75 | 32.72 |

STATE: LOUISIANA

| ZIP CODE | QUARTER 1 | QUARTER 2 | QUARTER 3 | QUARTER 4 | TOTAL GRAMS |
|---|---|---|---|---|---|

| | | | | | |
|---|---|---|---|---|---|
| 701 | 0.12 | 0 | 0 | 0 | 0.12 |
| 703 | 0.93 | 1.16 | 0.46 | 0.93 | 3.48 |
| 704 | 1.39 | 0.7 | 0.7 | 0.23 | 3.02 |
| 705 | 1.74 | 6.38 | 11.61 | 13.35 | 33.08 |
| 706 | 0.12 | 0 | 0 | 0 | 0.12 |
| 707 | 0.12 | 0.12 | 0.23 | 0 | 0.47 |
| 708 | 0.35 | 0.23 | 0.12 | 0 | 0.7 |
| 711 | 0 | 0 | 0.12 | 0.12 | 0.24 |
| TOTAL | 4.77 | 8.59 | 13.24 | 14.63 | 41.23 |

STATE: MAINE

| ZIP CODE | QUARTER 1 | QUARTER 2 | QUARTER 3 | QUARTER 4 | TOTAL GRAMS |
|---|---|---|---|---|---|
| 39 | 0 | 0 | 0 | 0.23 | 0.23 |
| TOTAL | 0 | 0 | 0 | 0.23 | 0.23 |

STATE: MARYLAND

| ZIP CODE | QUARTER 1 | QUARTER 2 | QUARTER 3 | QUARTER 4 | TOTAL GRAMS |
|---|---|---|---|---|---|
| 208 | 0 | 0 | 0 | 0.12 | 0.12 |
| 210 | 0 | 0.35 | 0 | 1.04 | 1.39 |
| 211 | 0 | 0 | 0 | 1.16 | 1.16 |
| 212 | 0 | 0 | 0.35 | 0.58 | 0.93 |
| 214 | 0 | 0 | 0 | 0.12 | 0.12 |
| 217 | 0.12 | 0.12 | 0 | 0.12 | 0.36 |
| TOTAL | 0.12 | 0.47 | 0.35 | 3.14 | 4.08 |

STATE: MASSACHUSETTS

| ZIP CODE | QUARTER 1 | QUARTER 2 | QUARTER 3 | QUARTER 4 | TOTAL GRAMS |
|---|---|---|---|---|---|
| 10 | 10.56 | 10.8 | 8.71 | 9.63 | 39.7 |
| 11 | 5.46 | 1.39 | 0.58 | 0.7 | 8.13 |
| 12 | 0.23 | 0 | 0 | 0 | 0.23 |
| 13 | 0.81 | 0.35 | 0.58 | 0 | 1.74 |
| 16 | 0.46 | 0 | 0 | 0 | 0.46 |
| 17 | 0 | 0.12 | 0 | 0.12 | 0.24 |
| 18 | 3.48 | 4.41 | 3.83 | 4.53 | 16.25 |
| 20 | 0 | 0.12 | 0 | 0.58 | 0.7 |
| 21 | 0.23 | 0.7 | 0.81 | 1.04 | 2.78 |
| 23 | 0.23 | 0.58 | 1.04 | 1.86 | 3.71 |
| 26 | 0.35 | 0.46 | 0 | 0 | 0.81 |
| 27 | 0 | 0 | 0.35 | 0.23 | 0.58 |
| TOTAL | 21.81 | 18.93 | 15.9 | 18.69 | 75.33 |

STATE: MICHIGAN

| ZIP CODE | QUARTER 1 | QUARTER 2 | QUARTER 3 | QUARTER 4 | TOTAL GRAMS |
|---|---|---|---|---|---|
| 495 | 0.12 | 0.12 | 0 | 0 | 0.24 |
| 497 | 0 | 1.04 | 0.93 | 1.28 | 3.25 |

DRUG CODE:9220LDRUG NAME:LEVORPHANOL
STATE:MICHIGAN

| ZIP CODE | QUARTER 1 | QUARTER 2 | QUARTER 3 | QUARTER 4 | TOTAL GRAMS |
|---|---|---|---|---|---|
| 498 | 0.12 | 0 | 0 | 0 | 0.12 |
| TOTAL | 0.24 | 1.16 | 0.93 | 1.28 | 3.61 |

STATE: MINNESOTA

| ZIP CODE | QUARTER 1 | QUARTER 2 | QUARTER 3 | QUARTER 4 | TOTAL GRAMS |
|---|---|---|---|---|---|
| 550 | 0.23 | 0 | 0.23 | 0 | 0.46 |
| 551 | 0.12 | 0.12 | 1.39 | 0.46 | 2.09 |
| TOTAL | 0.35 | 0.12 | 1.62 | 0.46 | 2.55 |

STATE: MISSISSIPPI

RETAIL DRUG DISTRIBUTION BY ZIP CODE WITHIN STATE BY GRAMS WT

| ZIP CODE | QUARTER 1 | QUARTER 2 | QUARTER 3 | QUARTER 4 | TOTAL GRAMS |
|---|---|---|---|---|---|
| 394 | 0 | 0.12 | 0 | 0.35 | 0.47 |
| 395 | 0.46 | 0.58 | 0.23 | 0 | 1.27 |
| 396 | 0.58 | 0.7 | 0 | 0 | 1.28 |
| TOTAL | 1.04 | 1.4 | 0.23 | 0.35 | 3.02 |

STATE: MISSOURI

| ZIP CODE | QUARTER 1 | QUARTER 2 | QUARTER 3 | QUARTER 4 | TOTAL GRAMS |
|---|---|---|---|---|---|
| 630 | 1.04 | 0.12 | 0 | 0.35 | 1.51 |
| 631 | 1.16 | 3.25 | 6.73 | 8.71 | 19.85 |
| 633 | 2.21 | 1.74 | 1.63 | 2.09 | 7.67 |
| 636 | 0 | 0 | 0.12 | 0 | 0.12 |
| 640 | 0.12 | 0.12 | 0.35 | 0.12 | 0.71 |
| 645 | 0 | 0 | 0.12 | 0 | 0.12 |
| 652 | 0.35 | 0 | 0 | 0 | 0.35 |
| 658 | 0 | 0.12 | 0.23 | 0.23 | 0.58 |
| TOTAL | 4.88 | 5.35 | 9.18 | 11.5 | 30.91 |

STATE: NEBRASKA

| ZIP CODE | QUARTER 1 | QUARTER 2 | QUARTER 3 | QUARTER 4 | TOTAL GRAMS |
|---|---|---|---|---|---|
| 680 | 0.35 | 0 | 0.12 | 0.46 | 0.93 |
| 681 | 0 | 0.35 | 0.93 | 1.39 | 2.67 |
| TOTAL | 0.35 | 0.35 | 1.05 | 1.85 | 3.6 |

STATE: NEVADA

| ZIP CODE | QUARTER 1 | QUARTER 2 | QUARTER 3 | QUARTER 4 | TOTAL GRAMS |
|---|---|---|---|---|---|
| 891 | 0 | 3.25 | 1.39 | 1.63 | 6.27 |
| 894 | 0.58 | 0.23 | 0.7 | 0.58 | 2.09 |
| 895 | 0 | 0 | 0 | 0.12 | 0.12 |
| TOTAL | 0.58 | 3.48 | 2.09 | 2.33 | 8.48 |

STATE: NEW HAMPSHIRE

| ZIP CODE | QUARTER 1 | QUARTER 2 | QUARTER 3 | QUARTER 4 | TOTAL GRAMS |
|---|---|---|---|---|---|
| 30 | 0.23 | 0.58 | 0 | 0.23 | 1.04 |
| 31 | 0.23 | 0 | 0 | 0 | 0.23 |
| 32 | 0.12 | 0 | 0 | 0 | 0.12 |
| 33 | 0 | 0.12 | 0 | 0 | 0.12 |
| 38 | 0.7 | 0.23 | 0.35 | 0.58 | 1.86 |
| TOTAL | 1.28 | 0.93 | 0.35 | 0.81 | 3.37 |

STATE: NEW JERSEY

| ZIP CODE | QUARTER 1 | QUARTER 2 | QUARTER 3 | QUARTER 4 | TOTAL GRAMS |
|---|---|---|---|---|---|
| 70 | 42.49 | 95.88 | 183.52 | 266.98 | 588.87 |
| 71 | 0.23 | 0 | 0 | 0 | 0.23 |
| 74 | 0 | 0.12 | 0 | 0 | 0.12 |
| 77 | 0 | 0.12 | 0.58 | 0.81 | 1.51 |
| 79 | 0.35 | 0.35 | 0.23 | 0.23 | 1.16 |
| 80 | 0.46 | 0.7 | 0.81 | 0.35 | 2.32 |
| 81 | 0 | 0 | 0.12 | 0 | 0.12 |
| 84 | 0 | 0 | 0 | 0.12 | 0.12 |
| 88 | 0 | 0.23 | 0 | 0 | 0.23 |
| TOTAL | 43.53 | 97.4 | 185.26 | 268.49 | 594.68 |

STATE: NEW MEXICO

| ZIP CODE | QUARTER 1 | QUARTER 2 | QUARTER 3 | QUARTER 4 | TOTAL GRAMS |
|---|---|---|---|---|---|
| 883 | 0 | 0 | 0 | 0.12 | 0.12 |
| TOTAL | 0 | 0 | 0 | 0.12 | 0.12 |

STATE: NEW YORK

| ZIP CODE | QUARTER 1 | QUARTER 2 | QUARTER 3 | QUARTER 4 | TOTAL GRAMS |
|---|---|---|---|---|---|
| 100 | 3.95 | 2.44 | 0.93 | 2.09 | 9.41 |

| | | | | | |
|---|---|---|---|---|---|
| 110 | 0 | 0.23 | 0.35 | 0.12 | 0.7 |
| 112 | 0 | 0 | 0 | 0.35 | 0.35 |
| 113 | 0 | 0 | 0.12 | 0.35 | 0.47 |
| 115 | 0 | 0 | 0.23 | 0.46 | 0.69 |

DRUG CODE:9220LDRUG NAME:LEVORPHANOL
STATE:NEW YORK

| ZIP CODE | QUARTER 1 | QUARTER 2 | QUARTER 3 | QUARTER 4 | TOTAL GRAMS |
|---|---|---|---|---|---|
| 117 | 28.44 | 11.26 | 15.44 | 30.3 | 85.44 |
| 118 | 0 | 0 | 0.58 | 0.58 | 1.16 |
| 119 | 0 | 0 | 0 | 0.23 | 0.23 |
| 120 | 0 | 0 | 0.12 | 0.46 | 0.58 |
| 121 | 0.23 | 0.58 | 0.23 | 0.23 | 1.27 |
| 122 | 0 | 1.28 | 2.09 | 2.79 | 6.16 |
| 125 | 0.58 | 0 | 0 | 0 | 0.58 |
| 126 | 0 | 0 | 0.12 | 0.7 | 0.82 |
| 128 | 0.46 | 0.46 | 0.23 | 0 | 1.15 |
| 130 | 0.81 | 1.04 | 0.35 | 0.35 | 2.55 |
| 132 | 0 | 0.12 | 0 | 0 | 0.12 |
| 133 | 0.23 | 0.35 | 0.35 | 0.46 | 1.39 |
| 134 | 0 | 0 | 0.23 | 0.7 | 0.93 |
| 135 | 0.12 | 0.35 | 0.35 | 0.23 | 1.05 |
| 137 | 0 | 0 | 0.46 | 1.16 | 1.62 |
| 138 | 0 | 0 | 0.81 | 1.97 | 2.78 |
| 139 | 0 | 0 | 1.04 | 2.32 | 3.36 |
| 140 | 0 | 0.7 | 0.58 | 0.12 | 1.4 |
| 141 | 0 | 0.46 | 0.12 | 0.35 | 0.93 |
| 142 | 0.93 | 2.79 | 2.21 | 2.9 | 8.83 |
| 143 | 0 | 0 | 0.46 | 0.23 | 0.69 |
| 146 | 0 | 0 | 0 | 0.35 | 0.35 |
| 148 | 0 | 0.35 | 0.12 | 0.46 | 0.93 |
| TOTAL | 35.75 | 22.41 | 27.52 | 50.26 | 135.94 |

STATE: NORTH CAROLINA

| ZIP CODE | QUARTER 1 | QUARTER 2 | QUARTER 3 | QUARTER 4 | TOTAL GRAMS |
|---|---|---|---|---|---|
| 270 | 0.81 | 1.04 | 0.46 | 0.46 | 2.77 |
| 271 | 0.93 | 0.58 | 1.16 | 0.12 | 2.79 |
| 272 | 0.46 | 0.35 | 0.46 | 0 | 1.27 |
| 273 | 0.7 | 0.35 | 0.93 | 0.23 | 2.21 |
| 275 | 5.46 | 5.34 | 8.71 | 10.45 | 29.96 |
| 276 | 1.04 | 0.7 | 2.55 | 1.51 | 5.8 |
| 277 | 1.04 | 0.12 | 0.23 | 0 | 1.39 |
| 278 | 0.81 | 0.46 | 1.86 | 5.92 | 9.05 |
| 280 | 0 | 0.35 | 0.58 | 1.97 | 2.9 |
| 281 | 0 | 0 | 0.35 | 0.35 | 0.7 |
| 282 | 0.7 | 0.12 | 1.74 | 0.81 | 3.37 |
| 283 | 1.97 | 1.74 | 6.38 | 8.47 | 18.56 |
| 284 | 1.74 | 2.44 | 3.25 | 3.37 | 10.8 |
| 285 | 2.44 | 2.44 | 3.95 | 4.99 | 13.82 |
| 286 | 1.39 | 0.7 | 1.86 | 1.39 | 5.34 |
| 287 | 0 | 0 | 0.12 | 0.12 | 0.24 |
| 288 | 0.35 | 0.35 | 0.46 | 0.35 | 1.51 |
| TOTAL | 19.84 | 17.08 | 35.05 | 40.51 | 112.48 |

STATE: OHIO

| ZIP CODE | QUARTER 1 | QUARTER 2 | QUARTER 3 | QUARTER 4 | TOTAL GRAMS |
|---|---|---|---|---|---|
| 430 | 0 | 0 | 0 | 0.12 | 0.12 |
| 432 | 1.16 | 2.21 | 7.78 | 11.96 | 23.11 |
| 433 | 0.12 | 0 | 0 | 0 | 0.12 |
| 436 | 0 | 0.35 | 0 | 0 | 0.35 |

| | | | | | |
|---|---|---|---|---|---|
| 439 | 0 | 0 | 0.46 | 0.35 | 0.81 |
| 445 | 0 | 0 | 0 | 0.12 | 0.12 |
| TOTAL | 1.28 | 2.56 | 8.24 | 12.55 | 24.63 |

STATE: OKLAHOMA

| ZIP CODE | QUARTER 1 | QUARTER 2 | QUARTER 3 | QUARTER 4 | TOTAL GRAMS |
|---|---|---|---|---|---|
| 730 | 0.35 | 0.46 | 0.35 | 0.35 | 1.51 |
| 731 | 0.12 | 0.23 | 0 | 0 | 0.35 |
| TOTAL | 0.47 | 0.69 | 0.35 | 0.35 | 1.86 |

STATE: OREGON

| ZIP CODE | QUARTER 1 | QUARTER 2 | QUARTER 3 | QUARTER 4 | TOTAL GRAMS |
|---|---|---|---|---|---|
| 970 | 0.46 | 0.35 | 0.58 | 0.35 | 1.74 |
| 972 | 0.93 | 0.46 | 0.58 | 0.58 | 2.55 |
| 974 | 4.29 | 3.02 | 2.32 | 2.09 | 11.72 |
| 978 | 0.93 | 0.81 | 0.58 | 0.81 | 3.13 |
| TOTAL | 6.61 | 4.64 | 4.06 | 3.83 | 19.14 |

STATE: PENNSYLVANIA

DRUG CODE:9220LDRUG NAME:LEVORPHANOL
STATE:PENNSYLVANIA

| ZIP CODE | QUARTER 1 | QUARTER 2 | QUARTER 3 | QUARTER 4 | TOTAL GRAMS |
|---|---|---|---|---|---|
| 153 | 0 | 0 | 0.12 | 0.35 | 0.47 |
| 159 | 0 | 0.12 | 0.12 | 0.23 | 0.47 |
| 163 | 0 | 0 | 0 | 0.12 | 0.12 |
| 170 | 0.35 | 0.46 | 0.23 | 0.46 | 1.5 |
| 173 | 0 | 0 | 0 | 0.12 | 0.12 |
| 177 | 1.39 | 1.16 | 1.04 | 0.46 | 4.05 |
| 178 | 3.13 | 3.25 | 3.37 | 3.83 | 13.58 |
| 179 | 0.23 | 0.23 | 0 | 0.12 | 0.58 |
| 183 | 0.23 | 0.35 | 0.35 | 0.35 | 1.28 |
| 186 | 0.12 | 0 | 0 | 0 | 0.12 |
| 188 | 0.12 | 0.12 | 0.23 | 0.23 | 0.7 |
| 189 | 0.23 | 0.58 | 0.46 | 0.35 | 1.62 |
| 190 | 1.86 | 2.32 | 1.16 | 1.39 | 6.73 |
| 191 | 2.21 | 1.86 | 1.28 | 1.97 | 7.32 |
| 193 | 0 | 0 | 0 | 0.12 | 0.12 |
| TOTAL | 9.87 | 10.45 | 8.36 | 10.1 | 38.78 |

STATE: SOUTH CAROLINA

| ZIP CODE | QUARTER 1 | QUARTER 2 | QUARTER 3 | QUARTER 4 | TOTAL GRAMS |
|---|---|---|---|---|---|
| 292 | 0.12 | 0 | 0 | 0.23 | 0.35 |
| 294 | 0.58 | 0 | 0.12 | 0.23 | 0.93 |
| 295 | 0 | 0 | 0.12 | 0.23 | 0.35 |
| 296 | 0 | 0 | 0.23 | 0 | 0.23 |
| TOTAL | 0.7 | 0 | 0.47 | 0.69 | 1.86 |

STATE: TENNESSEE

| ZIP CODE | QUARTER 1 | QUARTER 2 | QUARTER 3 | QUARTER 4 | TOTAL GRAMS |
|---|---|---|---|---|---|
| 372 | 0.23 | 0 | 0 | 0 | 0.23 |
| 376 | 0 | 0.23 | 0.12 | 0 | 0.35 |
| 378 | 0.12 | 0.12 | 0 | 0.12 | 0.36 |
| TOTAL | 0.35 | 0.35 | 0.12 | 0.12 | 0.94 |

STATE: TEXAS

| ZIP CODE | QUARTER 1 | QUARTER 2 | QUARTER 3 | QUARTER 4 | TOTAL GRAMS |
|---|---|---|---|---|---|
| 750 | 8.82 | 9.05 | 22.52 | 29.6 | 69.99 |
| 751 | 1.74 | 0 | 1.39 | 0.35 | 3.48 |

| | | | | | |
|---|---|---|---|---|---|
| 752 | 1.04 | 1.51 | 1.16 | 1.97 | 5.68 |
| 754 | 0.12 | 0.12 | 0 | 0.35 | 0.59 |
| 757 | 0.23 | 0 | 0 | 0.12 | 0.35 |
| 760 | 3.71 | 4.29 | 3.95 | 4.41 | 16.36 |
| 761 | 0.7 | 1.97 | 0.58 | 0.23 | 3.48 |
| 762 | 1.16 | 1.28 | 0.93 | 1.63 | 5 |
| 765 | 1.51 | 2.44 | 1.86 | 1.51 | 7.32 |
| 770 | 0.81 | 1.04 | 1.16 | 1.39 | 4.4 |
| 773 | 0 | 0 | 0.23 | 0.46 | 0.69 |
| 774 | 0 | 0.58 | 1.04 | 0.12 | 1.74 |
| 775 | 0.23 | 0.23 | 0.58 | 0.23 | 1.27 |
| 786 | 1.39 | 0.7 | 0.23 | 0.58 | 2.9 |
| 787 | 0.93 | 0.35 | 0.35 | 0.46 | 2.09 |
| 799 | 0 | 0 | 0.81 | 0 | 0.81 |
| TOTAL | 22.39 | 23.56 | 36.79 | 43.41 | 126.15 |

STATE: UTAH

| ZIP CODE | QUARTER 1 | QUARTER 2 | QUARTER 3 | QUARTER 4 | TOTAL GRAMS |
|---|---|---|---|---|---|
| 843 | 0.23 | 0 | 0 | 0 | 0.23 |
| 844 | 0.12 | 0 | 0 | 0 | 0.12 |
| TOTAL | 0.35 | 0 | 0 | 0 | 0.35 |

STATE: VERMONT

| ZIP CODE | QUARTER 1 | QUARTER 2 | QUARTER 3 | QUARTER 4 | TOTAL GRAMS |
|---|---|---|---|---|---|
| 54 | 0.58 | 0.35 | 0.46 | 0.58 | 1.97 |
| TOTAL | 0.58 | 0.35 | 0.46 | 0.58 | 1.97 |

STATE: VIRGINIA

| ZIP CODE | QUARTER 1 | QUARTER 2 | QUARTER 3 | QUARTER 4 | TOTAL GRAMS |
|---|---|---|---|---|---|
| 201 | 0.81 | 0 | 0 | 0 | 0.81 |
| 220 | 0.12 | 0 | 0 | 0 | 0.12 |
| 230 | 0.12 | 0.81 | 0.12 | 0.12 | 1.17 |
| 231 | 1.63 | 1.04 | 0.7 | 0.81 | 4.18 |
| 232 | 1.97 | 0.12 | 1.04 | 1.04 | 4.17 |
| 233 | 0 | 0.46 | 0.35 | 0.58 | 1.39 |
| 234 | 0.81 | 1.63 | 1.86 | 0.23 | 4.53 |

DRUG CODE:9220LDRUG NAME:LEVORPHANOL
STATE:VIRGINIA

| ZIP CODE | QUARTER 1 | QUARTER 2 | QUARTER 3 | QUARTER 4 | TOTAL GRAMS |
|---|---|---|---|---|---|
| 236 | 0.35 | 0.23 | 0.58 | 1.28 | 2.44 |
| 237 | 0.35 | 0 | 0 | 0 | 0.35 |
| 238 | 0.93 | 0 | 0.23 | 1.04 | 2.2 |
| 239 | 0.35 | 0.12 | 0 | 0 | 0.47 |
| 243 | 0 | 0.23 | 0.23 | 0.12 | 0.58 |
| 244 | 0 | 0 | 0.35 | 0.58 | 0.93 |
| TOTAL | 7.44 | 4.64 | 5.46 | 5.8 | 23.34 |

STATE: WASHINGTON

| ZIP CODE | QUARTER 1 | QUARTER 2 | QUARTER 3 | QUARTER 4 | TOTAL GRAMS |
|---|---|---|---|---|---|
| 980 | 5.57 | 2.79 | 1.74 | 2.44 | 12.54 |
| 981 | 1.28 | 0.23 | 0.35 | 0.23 | 2.09 |
| 982 | 5.92 | 7.55 | 8.36 | 9.87 | 31.7 |
| 983 | 0 | 0 | 0.12 | 0.58 | 0.7 |
| 984 | 0.35 | 1.86 | 1.63 | 0.12 | 3.96 |
| 985 | 0.23 | 0.35 | 0.81 | 1.97 | 3.36 |
| 986 | 0 | 0 | 0 | 0.23 | 0.23 |
| 992 | 0 | 0.23 | 0 | 0 | 0.23 |

| | | | | | | |
|---|---|---|---|---|---|---|
| TOTAL | | 13.35 | 13.01 | 13.01 | 15.44 | 54.81 |
| STATE: | WEST VIRGINIA | | | | | |
| ZIP CODE | QUARTER 1 | | QUARTER 2 | QUARTER 3 | QUARTER 4 | TOTAL GRAMS |
| 247 | | 0.12 | 0.35 | 0.35 | 0.12 | 0.94 |
| 254 | | 0 | 0 | 0 | 0.12 | 0.12 |
| TOTAL | | 0.12 | 0.35 | 0.35 | 0.24 | 1.06 |
| STATE: | WISCONSIN | | | | | |
| ZIP CODE | QUARTER 1 | | QUARTER 2 | QUARTER 3 | QUARTER 4 | TOTAL GRAMS |
| 531 | | 0.7 | 0 | 0 | 0 | 0.7 |
| 532 | | 0.12 | 0 | 0 | 0 | 0.12 |
| 535 | | 1.51 | 1.51 | 1.04 | 1.39 | 5.45 |
| 537 | | 0.23 | 0.58 | 0.81 | 0.81 | 2.43 |
| 539 | | 0 | 0 | 0 | 0.12 | 0.12 |
| 543 | | 0 | 0 | 0.12 | 0.12 | 0.24 |
| 544 | | 0 | 0.58 | 0.58 | 0.93 | 2.09 |
| 549 | | 0.58 | 0.81 | 0.35 | 0 | 1.74 |
| TOTAL | | 3.14 | 3.48 | 2.9 | 3.37 | 12.89 |
| DRUG CODE: | | 9230 | DRUG NAME: | MEPERIDINE (PETHIDINE) | | |
| STATE: | ALABAMA | | | | | |
| ZIP CODE | QUARTER 1 | | QUARTER 2 | QUARTER 3 | QUARTER 4 | TOTAL GRAMS |
| 350 | | 777.62 | 720.43 | 702.65 | 730.08 | 2,930.78 |
| 351 | | 316.79 | 359.28 | 289.99 | 301.97 | 1,268.03 |
| 352 | | 975.58 | 898.41 | 795.03 | 817.38 | 3,486.40 |
| 354 | | 601.55 | 584.12 | 492.62 | 499.15 | 2,177.44 |
| 355 | | 292.24 | 293.8 | 218.09 | 233.13 | 1,037.26 |
| 356 | | 773.43 | 730.27 | 664.04 | 542.66 | 2,710.40 |
| 357 | | 389.6 | 368.75 | 389.34 | 341.24 | 1,488.93 |
| 358 | | 659.94 | 628.13 | 629.33 | 573.23 | 2,490.63 |
| 359 | | 927.69 | 694.61 | 531.62 | 540.92 | 2,694.84 |
| 360 | | 343.55 | 362.7 | 336.77 | 319.23 | 1,362.25 |
| 361 | | 359.6 | 377.25 | 494.79 | 294.02 | 1,525.66 |
| 362 | | 574.97 | 566.04 | 571.49 | 10,094.04 | 11,806.54 |
| 363 | | 574.38 | 539.39 | 512.68 | 497.13 | 2,123.58 |
| 364 | | 260.27 | 299.01 | 283.78 | 314.39 | 1,157.45 |
| 365 | | 624.1 | 685.65 | 610.49 | 595.34 | 2,515.58 |
| 366 | | 762.56 | 742.58 | 773.3 | 610.42 | 2,888.86 |
| 367 | | 167.87 | 175.28 | 202.41 | 165.69 | 711.25 |
| 368 | | 459.43 | 458.08 | 337.97 | 436.62 | 1,692.10 |
| 369 | | 22.33 | 7.63 | 17.43 | 7.08 | 54.47 |
| TOTAL | | 9,863.50 | 9,491.41 | 8,853.82 | 17,913.72 | 46,122.45 |
| STATE: | ALASKA | | | | | |
| ZIP CODE | QUARTER 1 | | QUARTER 2 | QUARTER 3 | QUARTER 4 | TOTAL GRAMS |
| 995 | | 510.37 | 615.93 | 479.65 | 496.32 | 2,102.27 |
| 996 | | 171.47 | 174.95 | 199.46 | 198.59 | 744.47 |
| 997 | | 94.99 | 88.02 | 77.78 | 109.37 | 370.16 |
| 998 | | 9.8 | 26.36 | 21.57 | 16.12 | 73.85 |
| 999 | | 8.93 | 8.28 | 0.65 | 0.98 | 18.84 |
| TOTAL | | 795.56 | 913.54 | 779.11 | 821.38 | 3,309.59 |

DRUG CODE:9230DRUG NAME:MEPERIDINE (PETHIDINE)

| | | | | | | |
|---|---|---|---|---|---|---|
| STATE: | AMERICAN SAMOA | | | | | |
| ZIP CODE | QUARTER 1 | | QUARTER 2 | QUARTER 3 | QUARTER 4 | TOTAL GRAMS |
| 967 | | 6.1 | 0 | 0 | 0 | 6.1 |
| TOTAL | | 6.1 | 0 | 0 | 0 | 6.1 |
| STATE: | ARIZONA | | | | | |

RETAIL DRUG DISTRIBUTION BY ZIP CODE WITHIN STATE BY GRAMS WT

| ZIP CODE | QUARTER 1 | QUARTER 2 | QUARTER 3 | QUARTER 4 | TOTAL GRAMS |
|---|---|---|---|---|---|
| 850 | 1,099.05 | 1,176.61 | 1,016.34 | 1,087.52 | 4,379.52 |
| 851 | 229.86 | 215.81 | 163.52 | 159.92 | 769.11 |
| 852 | 1,905.53 | 1,898.13 | 2,085.83 | 1,807.58 | 7,697.07 |
| 853 | 747.09 | 951.9 | 936.54 | 792.3 | 3,427.83 |
| 855 | 165.59 | 143.14 | 112.86 | 170.16 | 591.75 |
| 856 | 415.49 | 368.86 | 381.87 | 284.87 | 1,451.09 |
| 857 | 1,219.88 | 1,266.83 | 1,149.73 | 972.59 | 4,609.03 |
| 859 | 15.9 | 23.53 | 17.32 | 8.28 | 65.03 |
| 860 | 161.01 | 93.9 | 92.38 | 82.36 | 429.65 |
| 863 | 135.08 | 132.25 | 123.97 | 98.92 | 490.22 |
| 864 | 208.62 | 245.44 | 165.04 | 237.81 | 856.91 |
| 865 | 0.22 | 1.31 | 0.44 | 0.22 | 2.19 |
| TOTAL | 6,303.32 | 6,517.71 | 6,245.84 | 5,702.53 | 24,769.40 |

STATE: ARKANSAS

| ZIP CODE | QUARTER 1 | QUARTER 2 | QUARTER 3 | QUARTER 4 | TOTAL GRAMS |
|---|---|---|---|---|---|
| 716 | 605.69 | 644.98 | 667.81 | 602.9 | 2,521.38 |
| 717 | 411.98 | 273.3 | 350.15 | 181.05 | 1,216.48 |
| 718 | 106.98 | 113.38 | 78.17 | 70.42 | 368.95 |
| 719 | 575.95 | 529.22 | 487.17 | 560.48 | 2,152.82 |
| 720 | 1,466.87 | 1,375.42 | 1,616.33 | 1,512.51 | 5,971.13 |
| 721 | 1,654.11 | 1,646.94 | 1,568.90 | 1,603.71 | 6,473.66 |
| 722 | 1,309.84 | 1,169.36 | 1,027.61 | 1,096.41 | 4,603.22 |
| 723 | 603.3 | 433.79 | 557.11 | 565.06 | 2,159.26 |
| 724 | 643.62 | 701.71 | 634.56 | 653.63 | 2,633.52 |
| 725 | 282.63 | 339.78 | 293.26 | 278.01 | 1,193.68 |
| 726 | 222.34 | 239.99 | 306.66 | 215.91 | 984.9 |
| 727 | 1,214.65 | 1,264.11 | 1,191.78 | 1,138.40 | 4,808.94 |
| 728 | 263.08 | 272.21 | 277.79 | 189.88 | 1,002.96 |
| 729 | 582.75 | 508.08 | 605.47 | 409.17 | 2,105.47 |
| TOTAL | 9,943.79 | 9,512.27 | 9,662.77 | 9,077.54 | 38,196.37 |

STATE: CALIFORNIA

| ZIP CODE | QUARTER 1 | QUARTER 2 | QUARTER 3 | QUARTER 4 | TOTAL GRAMS |
|---|---|---|---|---|---|
| 900 | 974.66 | 1,006.80 | 841.65 | 872.81 | 3,695.92 |
| 902 | 429.32 | 527.58 | 430.74 | 496.86 | 1,884.50 |
| 903 | 1.63 | 3.81 | 14.16 | 8.82 | 28.42 |
| 904 | 158.07 | 170.81 | 122.66 | 101.64 | 553.18 |
| 905 | 222.23 | 261.78 | 263.74 | 258.4 | 1,006.15 |
| 906 | 151.64 | 158.5 | 237.59 | 140.97 | 688.7 |
| 907 | 102.29 | 128.11 | 80.94 | 64.38 | 375.72 |
| 908 | 154.58 | 85.41 | 108.07 | 101.2 | 449.26 |
| 910 | 61.01 | 83.88 | 56.65 | 71.25 | 272.79 |
| 911 | 150.66 | 156.76 | 93.36 | 118.96 | 519.74 |
| 912 | 106.11 | 148.7 | 137.7 | 148.81 | 541.32 |
| 913 | 1,045.04 | 858.65 | 826.84 | 1,008.33 | 3,738.86 |
| 914 | 197.98 | 274.09 | 230.4 | 211.56 | 914.03 |
| 915 | 115.47 | 89.55 | 81.92 | 125.06 | 412 |
| 916 | 6.54 | 7.63 | 2.72 | 7.63 | 24.52 |
| 917 | 1,087.09 | 1,400.83 | 1,026.19 | 1,105.61 | 4,619.72 |
| 918 | 44.12 | 59.04 | 51.64 | 48.8 | 203.6 |
| 919 | 62.09 | 30.94 | 48.15 | 37.91 | 179.09 |
| 920 | 900.37 | 913.01 | 1,006.15 | 1,133.82 | 3,953.35 |
| 921 | 752.32 | 750.14 | 738.16 | 715.17 | 2,955.79 |
| 922 | 393.05 | 268.31 | 271.36 | 308.62 | 1,241.34 |
| 923 | 465.27 | 278.55 | 379.21 | 464.73 | 1,587.76 |
| 924 | 37.47 | 32.25 | 33.12 | 36.71 | 139.55 |
| 925 | 806.79 | 989.15 | 739.79 | 828.69 | 3,364.42 |
| 926 | 658.53 | 763.43 | 632.82 | 758.86 | 2,813.64 |
| 927 | 211.88 | 298.27 | 242.06 | 222.34 | 974.55 |

| | | | | | |
|---|---|---|---|---|---|
| 928 | 683.47 | 653.84 | 528.56 | 655.8 | 2,521.67 |
| 930 | 205.78 | 241.84 | 119.07 | 131.81 | 698.5 |
| 931 | 196.63 | 247.83 | 196.31 | 217.44 | 858.21 |
| 932 | 411.11 | 442.5 | 390.21 | 265.92 | 1,509.74 |

DRUG CODE:9230DRUG NAME:MEPERIDINE (PETHIDINE)
STATE:CALIFORNIA

| ZIP CODE | QUARTER 1 | QUARTER 2 | QUARTER 3 | QUARTER 4 | TOTAL GRAMS |
|---|---|---|---|---|---|
| 933 | 749.6 | 666.92 | 759.4 | 710.05 | 2,885.97 |
| 934 | 256.77 | 311.56 | 277.25 | 296.09 | 1,141.67 |
| 935 | 273.43 | 222.45 | 212.86 | 152.84 | 861.58 |
| 936 | 125.74 | 220.64 | 160.84 | 137.57 | 644.79 |
| 937 | 337.27 | 483.14 | 343.48 | 441.52 | 1,605.41 |
| 939 | 174.52 | 156 | 187.81 | 219.29 | 737.62 |
| 940 | 170.38 | 244.24 | 207.63 | 246.42 | 868.67 |
| 941 | 567.56 | 346.2 | 534.01 | 646.65 | 2,094.42 |
| 943 | 160.14 | 164.28 | 144.23 | 185.63 | 654.28 |
| 944 | 9.15 | 11.98 | 10.89 | 17.65 | 49.67 |
| 945 | 1,102.77 | 1,049.29 | 922.26 | 916.93 | 3,991.25 |
| 946 | 31.7 | 23.09 | 18.41 | 15.8 | 89 |
| 947 | 3.05 | 9.26 | 6.54 | 3.05 | 21.9 |
| 948 | 4.36 | 4.58 | 2.18 | 2.18 | 13.3 |
| 949 | 47.71 | 53.38 | 20.48 | 45.43 | 167 |
| 950 | 197.07 | 208.4 | 170.16 | 133.23 | 708.86 |
| 951 | 118.31 | 112.86 | 135.95 | 118.09 | 485.21 |
| 952 | 214.93 | 133.34 | 196.31 | 210.58 | 755.16 |
| 953 | 842.11 | 931.96 | 839.69 | 808.1 | 3,421.86 |
| 954 | 141.95 | 203.71 | 185.74 | 178.66 | 710.06 |
| 955 | 123.45 | 93.9 | 54.25 | 78.87 | 350.47 |
| 956 | 470.61 | 701.34 | 543.82 | 572.79 | 2,288.56 |
| 957 | 43.58 | 26.58 | 28.32 | 33.23 | 131.71 |
| 958 | 404.48 | 423.77 | 331.28 | 294.02 | 1,453.55 |
| 959 | 343.65 | 382.15 | 340.76 | 212.97 | 1,279.53 |
| 960 | 96.41 | 118.92 | 102.53 | 99.44 | 417.3 |
| 961 | 68.19 | 55.56 | 50.98 | 44.23 | 218.96 |
| TOTAL | 17,872.09 | 18,691.49 | 16,720 | 17,490.22 | 70,773.80 |

STATE: COLORADO

| ZIP CODE | QUARTER 1 | QUARTER 2 | QUARTER 3 | QUARTER 4 | TOTAL GRAMS |
|---|---|---|---|---|---|
| 800 | 212.54 | 243.8 | 205.02 | 202.41 | 863.77 |
| 801 | 458.41 | 429.65 | 454.05 | 358.19 | 1,700.30 |
| 802 | 282.91 | 335.85 | 319.19 | 297.29 | 1,235.24 |
| 803 | 64.71 | 42.27 | 25.93 | 51.53 | 184.44 |
| 804 | 118.52 | 77.13 | 106.11 | 66.45 | 368.21 |
| 805 | 232.91 | 241.51 | 182.25 | 180.73 | 837.4 |
| 806 | 179.31 | 157.09 | 165.8 | 162.1 | 664.3 |
| 807 | 71.25 | 41.4 | 26.15 | 52.29 | 191.09 |
| 808 | 63.4 | 89.76 | 83.88 | 79.52 | 316.56 |
| 809 | 218.09 | 196.41 | 222.67 | 181.93 | 819.1 |
| 810 | 174.19 | 138.9 | 177.79 | 143.03 | 633.91 |
| 811 | 10.78 | 5.88 | 13.62 | 14.16 | 44.44 |
| 812 | 64.16 | 34.75 | 35.4 | 32.14 | 166.45 |
| 813 | 65.58 | 51.85 | 87.8 | 92.81 | 298.04 |
| 814 | 70.81 | 89.98 | 87.15 | 90.96 | 338.9 |
| 815 | 118.96 | 87.37 | 105.89 | 103.27 | 415.49 |
| 816 | 134.97 | 123.64 | 111.88 | 149.24 | 519.73 |
| TOTAL | 2,541.50 | 2,387.24 | 2,410.58 | 2,258.05 | 9,597.37 |

STATE: CONNECTICUT

| ZIP CODE | QUARTER 1 | QUARTER 2 | QUARTER 3 | QUARTER 4 | TOTAL GRAMS |
|---|---|---|---|---|---|

| | | | | | |
|---|---|---|---|---|---|
| 60 | 121.36 | 100.55 | 117.11 | 116.89 | 455.91 |
| 61 | 88.78 | 78.33 | 58.72 | 87.69 | 313.52 |
| 62 | 17.65 | 23.97 | 11.55 | 11.33 | 64.5 |
| 63 | 93.03 | 51.42 | 84.75 | 102.51 | 331.71 |
| 64 | 491.09 | 419.08 | 429.21 | 363.96 | 1,703.34 |
| 65 | 108.72 | 86.93 | 108.61 | 94.56 | 398.82 |
| 66 | 97.06 | 114.28 | 118.96 | 97.39 | 427.69 |
| 67 | 132.9 | 118.09 | 106.11 | 173.86 | 530.96 |
| 68 | 277.68 | 276.27 | 233.45 | 268.75 | 1,056.15 |
| 69 | 33.33 | 10.02 | 11.98 | 18.85 | 74.18 |
| TOTAL | 1,461.60 | 1,278.94 | 1,280.45 | 1,335.79 | 5,356.78 |

STATE: DELAWARE

| ZIP CODE | QUARTER 1 | QUARTER 2 | QUARTER 3 | QUARTER 4 | TOTAL GRAMS |
|---|---|---|---|---|---|
| 197 | 83.12 | 68.41 | 82.68 | 76.47 | 310.68 |
| 198 | 22.44 | 11.55 | 25.93 | 5.99 | 65.91 |
| 199 | 130.53 | 161.53 | 124.54 | 141.01 | 557.61 |

DRUG CODE:9230DRUG NAME:MEPERIDINE (PETHIDINE)
STATE:DELAWARE

| ZIP CODE | QUARTER 1 | QUARTER 2 | QUARTER 3 | QUARTER 4 | TOTAL GRAMS |
|---|---|---|---|---|---|
| TOTAL | 236.09 | 241.49 | 233.15 | 223.47 | 934.2 |

STATE: DISTRICT OF COLUMBIA

| ZIP CODE | QUARTER 1 | QUARTER 2 | QUARTER 3 | QUARTER 4 | TOTAL GRAMS |
|---|---|---|---|---|---|
| 200 | 183.67 | 159.38 | 207.85 | 245.33 | 796.23 |
| 204 | 0 | 0 | 0.44 | 0 | 0.44 |
| 205 | 0 | 0.44 | 0 | 0 | 0.44 |
| TOTAL | 183.67 | 159.82 | 208.29 | 245.33 | 797.11 |

STATE: FLORIDA

| ZIP CODE | QUARTER 1 | QUARTER 2 | QUARTER 3 | QUARTER 4 | TOTAL GRAMS |
|---|---|---|---|---|---|
| 320 | 557.11 | 459.93 | 337.38 | 414.94 | 1,769.36 |
| 321 | 329.86 | 328.23 | 310.04 | 291.84 | 1,259.97 |
| 322 | 816.81 | 909.95 | 776.83 | 809.51 | 3,313.10 |
| 323 | 290.43 | 404.48 | 346.75 | 381.17 | 1,422.83 |
| 324 | 213.74 | 173.54 | 163.95 | 148.37 | 699.6 |
| 325 | 888.93 | 829.45 | 815.62 | 748.29 | 3,282.29 |
| 326 | 230.62 | 233.13 | 186.28 | 206.98 | 857.01 |
| 327 | 454.16 | 407.21 | 387.38 | 287.92 | 1,536.67 |
| 328 | 178.77 | 153.38 | 125.06 | 150.44 | 607.65 |
| 329 | 660.14 | 523.55 | 504.71 | 558.09 | 2,246.49 |
| 330 | 665.28 | 745.13 | 829.67 | 724.43 | 2,964.51 |
| 331 | 886.97 | 762.78 | 764.85 | 622.03 | 3,036.63 |
| 333 | 463.09 | 455.79 | 452.42 | 535.75 | 1,907.05 |
| 334 | 589.68 | 679.77 | 500.68 | 676.61 | 2,446.74 |
| 335 | 343.15 | 387.93 | 454.71 | 415.16 | 1,600.95 |
| 336 | 403.5 | 464.18 | 455.58 | 470.72 | 1,793.98 |
| 337 | 314.39 | 340.43 | 312.32 | 271.47 | 1,238.61 |
| 338 | 450.46 | 458.3 | 379.76 | 477.91 | 1,766.43 |
| 339 | 446.86 | 443.48 | 481.18 | 553.29 | 1,924.81 |
| 341 | 86.93 | 80.83 | 109.81 | 86.5 | 364.07 |
| 342 | 296.75 | 301.76 | 245.44 | 220.05 | 1,064 |
| 344 | 449.26 | 393.48 | 362.65 | 387.6 | 1,592.99 |
| 346 | 358.4 | 371.8 | 328.12 | 358.3 | 1,416.62 |
| 347 | 189.77 | 139.11 | 144.12 | 147.5 | 620.5 |
| 349 | 304.26 | 364.5 | 350.78 | 310.15 | 1,329.69 |
| TOTAL | 10,869.32 | 10,812.12 | 10,126.09 | 10,255.02 | 42,062.55 |

STATE: GEORGIA

| ZIP CODE | QUARTER 1 | QUARTER 2 | QUARTER 3 | QUARTER 4 | TOTAL GRAMS |
|---|---|---|---|---|---|
| 300 | 1,155.17 | 1,215.52 | 1,119.55 | 1,250.82 | 4,741.06 |
| 301 | 1,336.34 | 1,373.81 | 1,196.90 | 1,103.97 | 5,011.02 |
| 302 | 863 | 795.46 | 797.97 | 1,000.70 | 3,457.13 |
| 303 | 850.26 | 646.76 | 741.43 | 724.54 | 2,962.99 |
| 304 | 345.85 | 325.53 | 347.51 | 285.68 | 1,304.57 |
| 305 | 680.21 | 696.87 | 653.06 | 662.06 | 2,692.20 |
| 306 | 597.24 | 513.64 | 476.82 | 579.33 | 2,167.03 |
| 307 | 809.51 | 917.19 | 944.16 | 800.39 | 3,471.25 |
| 308 | 445.88 | 257.96 | 278.53 | 269.84 | 1,252.21 |
| 309 | 693.5 | 537.17 | 632.06 | 534.67 | 2,397.40 |
| 310 | 893.46 | 886.82 | 813.48 | 854.33 | 3,448.09 |
| 312 | 391.3 | 387.27 | 328.99 | 416.9 | 1,524.46 |
| 313 | 307.2 | 282.15 | 337.16 | 324.31 | 1,250.82 |
| 314 | 383.46 | 318.1 | 313.2 | 359.49 | 1,374.25 |
| 315 | 748.42 | 666.48 | 679.07 | 699.25 | 2,793.22 |
| 316 | 1,010.46 | 842.39 | 984.64 | 840.95 | 3,678.44 |
| 317 | 909.21 | 917.97 | 781.52 | 815.83 | 3,424.53 |
| 318 | 104.04 | 99.79 | 55.56 | 105.12 | 364.51 |
| 319 | 512.57 | 480.5 | 434.01 | 416.56 | 1,843.64 |
| 398 | 200.01 | 216.33 | 150.86 | 206.04 | 773.24 |
| TOTAL | 13,237.09 | 12,377.71 | 12,066.48 | 12,250.78 | 49,932.06 |

STATE: GUAM

| ZIP CODE | QUARTER 1 | QUARTER 2 | QUARTER 3 | QUARTER 4 | TOTAL GRAMS |
|---|---|---|---|---|---|
| 969 | 34.32 | 70.37 | 9.59 | 43.58 | 157.86 |
| TOTAL | 34.32 | 70.37 | 9.59 | 43.58 | 157.86 |

STATE: HAWAII

| ZIP CODE | QUARTER 1 | QUARTER 2 | QUARTER 3 | QUARTER 4 | TOTAL GRAMS |
|---|---|---|---|---|---|
| 967 | 294.24 | 304.26 | 239.88 | 198.27 | 1,036.65 |
| 968 | 201.32 | 192.71 | 149.35 | 186.39 | 729.77 |

DRUG CODE:9230DRUG NAME:MEPERIDINE (PETHIDINE)
STATE:HAWAII

| ZIP CODE | QUARTER 1 | QUARTER 2 | QUARTER 3 | QUARTER 4 | TOTAL GRAMS |
|---|---|---|---|---|---|
| TOTAL | 495.56 | 496.97 | 389.23 | 384.66 | 1,766.42 |

STATE: IDAHO

| ZIP CODE | QUARTER 1 | QUARTER 2 | QUARTER 3 | QUARTER 4 | TOTAL GRAMS |
|---|---|---|---|---|---|
| 832 | 155.89 | 113.84 | 134.76 | 112.21 | 516.7 |
| 833 | 100.96 | 95.91 | 131.16 | 62.57 | 390.6 |
| 834 | 151.97 | 90.53 | 139.22 | 129.2 | 510.92 |
| 835 | 50.66 | 36.06 | 56.87 | 64.71 | 208.3 |
| 836 | 210.03 | 259.49 | 178.77 | 212.43 | 860.72 |
| 837 | 215.91 | 203.39 | 219.84 | 170.16 | 809.3 |
| 838 | 82.57 | 122.6 | 141.18 | 84.1 | 430.45 |
| TOTAL | 967.99 | 921.82 | 1,001.80 | 835.38 | 3,726.99 |

STATE: ILLINOIS

| ZIP CODE | QUARTER 1 | QUARTER 2 | QUARTER 3 | QUARTER 4 | TOTAL GRAMS |
|---|---|---|---|---|---|
| 600 | 446.1 | 357.53 | 392.18 | 349.04 | 1,544.85 |
| 601 | 481.05 | 456.67 | 399.19 | 499.26 | 1,836.17 |
| 602 | 0.7 | 0.44 | 10.46 | 2.18 | 13.78 |
| 603 | 3.92 | 19.61 | 2.61 | 14.16 | 40.3 |
| 604 | 351.65 | 300.23 | 380.74 | 280.19 | 1,312.81 |
| 605 | 224.08 | 204.27 | 247.07 | 168.09 | 843.51 |
| 606 | 212.43 | 257.86 | 208.07 | 189.88 | 868.24 |
| 607 | 0 | 4.36 | 4.36 | 0.65 | 9.37 |

| | | | | | |
|---|---|---|---|---|---|
| 608 | 63.18 | 52.29 | 43.58 | 69.72 | 228.77 |
| 609 | 14.21 | 1.53 | 3.16 | 0.22 | 19.12 |
| 610 | 101.75 | 92.38 | 111.33 | 130.07 | 435.53 |
| 611 | 98.48 | 135.08 | 62.75 | 68.63 | 364.94 |
| 612 | 10.94 | 35.73 | 13.29 | 9.8 | 69.76 |
| 613 | 90.07 | 63.36 | 66.32 | 57.52 | 277.27 |
| 614 | 94.43 | 77.82 | 86.93 | 75.17 | 334.35 |
| 615 | 80.07 | 55.99 | 82.57 | 76.15 | 294.78 |
| 616 | 175.17 | 162.64 | 182.8 | 191.51 | 712.12 |
| 617 | 59.52 | 34.42 | 68.63 | 39.65 | 202.22 |
| 618 | 171.47 | 206.55 | 167.87 | 159.07 | 704.96 |
| 619 | 44.56 | 53.05 | 50.44 | 58.72 | 206.77 |
| 620 | 61.55 | 84.75 | 35.08 | 55.34 | 236.72 |
| 622 | 376.05 | 363.63 | 328.66 | 247.72 | 1,316.06 |
| 623 | 35.67 | 17.97 | 14.05 | 23.97 | 91.66 |
| 624 | 33.6 | 34.53 | 25.93 | 21.68 | 115.74 |
| 625 | 11.33 | 32.25 | 22.01 | 15.47 | 81.06 |
| 626 | 244.28 | 171.03 | 196.96 | 231.17 | 843.44 |
| 627 | 102.62 | 131.05 | 72.12 | 82.36 | 388.15 |
| 628 | 118.28 | 120.53 | 96.3 | 98.92 | 434.03 |
| 629 | 241.27 | 288.62 | 240.32 | 175.61 | 945.82 |
| TOTAL | 3,948.43 | 3,816.17 | 3,615.78 | 3,391.92 | 14,772.30 |

STATE: INDIANA

| ZIP CODE | QUARTER 1 | QUARTER 2 | QUARTER 3 | QUARTER 4 | TOTAL GRAMS |
|---|---|---|---|---|---|
| 460 | 275.61 | 270.17 | 271.58 | 293.26 | 1,110.62 |
| 461 | 562.34 | 514.45 | 422.9 | 506.67 | 2,006.36 |
| 462 | 1,060.18 | 746.55 | 733.48 | 1,062.03 | 3,602.24 |
| 463 | 261.45 | 320.6 | 261.23 | 270.38 | 1,113.66 |
| 464 | 54.47 | 75.38 | 64.93 | 55.56 | 250.34 |
| 465 | 43.14 | 52.62 | 78.33 | 59.15 | 233.24 |
| 466 | 41.4 | 30.5 | 37.04 | 19.5 | 128.44 |
| 467 | 145.37 | 81.49 | 80.18 | 78.7 | 385.74 |
| 468 | 210.95 | 146.56 | 176.04 | 158.94 | 692.49 |
| 469 | 176.39 | 146.74 | 102.4 | 101.36 | 526.89 |
| 470 | 150.9 | 112.45 | 100.09 | 108.2 | 471.64 |
| 471 | 281.84 | 211.8 | 227.16 | 224.08 | 944.88 |
| 472 | 136.72 | 121.9 | 156 | 158.18 | 572.8 |
| 473 | 154.47 | 168.85 | 220.05 | 177.89 | 721.26 |
| 474 | 115.8 | 81.16 | 108.17 | 74.73 | 379.86 |
| 475 | 72.49 | 60.35 | 74.84 | 42.16 | 249.84 |
| 476 | 65.36 | 69.5 | 51.53 | 71.68 | 258.07 |
| 477 | 137.91 | 130.29 | 150.12 | 163.84 | 582.16 |
| 478 | 117.87 | 118.96 | 90.85 | 97.83 | 425.51 |
| 479 | 88.89 | 97.61 | 96.52 | 88.57 | 371.59 |
| TOTAL | 4,153.55 | 3,557.93 | 3,503.44 | 3,812.71 | 15,027.63 |

DRUG CODE:9230DRUG NAME:MEPERIDINE (PETHIDINE)
STATE: IOWA

| ZIP CODE | QUARTER 1 | QUARTER 2 | QUARTER 3 | QUARTER 4 | TOTAL GRAMS |
|---|---|---|---|---|---|
| 500 | 63.6 | 98.26 | 69.39 | 90.42 | 321.67 |
| 501 | 17.43 | 19.39 | 23.31 | 15.69 | 75.82 |
| 502 | 160.57 | 217.66 | 190.2 | 194.78 | 763.21 |
| 503 | 471.92 | 539.68 | 456.67 | 389.34 | 1,857.61 |
| 504 | 28.69 | 6.97 | 2.61 | 7.41 | 45.68 |
| 505 | 102.01 | 66.67 | 74.51 | 65.36 | 308.55 |
| 506 | 53.82 | 37.26 | 56.21 | 47.06 | 194.35 |
| 507 | 211.34 | 251.21 | 257.53 | 266.9 | 986.98 |
| 508 | 3.68 | 2.83 | 11.77 | 24.08 | 42.36 |

| | | | | | |
|---|---|---|---|---|---|
| 510 | 8.28 | 13.07 | 15.47 | 14.16 | 50.98 |
| 511 | 45.21 | 46.84 | 88.78 | 105.02 | 285.85 |
| 512 | 3.05 | 9.8 | 4.79 | 18.74 | 36.38 |
| 513 | 28.76 | 23.97 | 22.66 | 11.55 | 86.94 |
| 514 | 16.12 | 6.32 | 6.75 | 9.37 | 38.56 |
| 515 | 31.83 | 34.1 | 35.6 | 35.95 | 137.48 |
| 516 | 1.74 | 4.58 | 6.32 | 0 | 12.64 |
| 520 | 47.06 | 91.29 | 44.01 | 58.83 | 241.19 |
| 521 | 0.87 | 1.53 | 0.87 | 0 | 3.27 |
| 522 | 109.5 | 99.35 | 78.22 | 119.83 | 406.9 |
| 523 | 69.81 | 66.02 | 41.18 | 46.73 | 223.74 |
| 524 | 177.89 | 211.77 | 203.5 | 198.27 | 791.43 |
| 525 | 135.63 | 82.55 | 95.8 | 103.03 | 417.01 |
| 526 | 36.65 | 22.05 | 31.59 | 29.2 | 119.49 |
| 527 | 34.86 | 47.28 | 50.22 | 112.42 | 244.78 |
| 528 | 24.18 | 56.21 | 24.08 | 32.46 | 136.93 |
| TOTAL | 1,884.50 | 2,056.66 | 1,892.04 | 1,996.60 | 7,829.80 |

STATE: KANSAS

| ZIP CODE | QUARTER 1 | QUARTER 2 | QUARTER 3 | QUARTER 4 | TOTAL GRAMS |
|---|---|---|---|---|---|
| 660 | 205.17 | 238.03 | 275 | 288.44 | 1,006.64 |
| 661 | 120.48 | 75.17 | 84.97 | 76.91 | 357.53 |
| 662 | 221.62 | 257.53 | 235.31 | 193.91 | 908.37 |
| 664 | 21.68 | 57.78 | 30.07 | 35.23 | 144.76 |
| 665 | 47.32 | 75.6 | 81.7 | 70.37 | 274.99 |
| 666 | 63.84 | 77.45 | 62.2 | 56.87 | 260.36 |
| 667 | 141.03 | 133.64 | 126.37 | 107.3 | 508.34 |
| 668 | 55.71 | 29.89 | 23.75 | 60.35 | 169.7 |
| 669 | 21.61 | 17.87 | 9.15 | 20.48 | 69.11 |
| 670 | 104.99 | 90.2 | 149.24 | 110.79 | 455.22 |
| 671 | 91.73 | 64.6 | 40.74 | 47.06 | 244.13 |
| 672 | 272.52 | 334.11 | 282.26 | 313.13 | 1,202.02 |
| 673 | 30.5 | 43.14 | 53.38 | 64.71 | 191.73 |
| 674 | 97.65 | 141.84 | 109.81 | 115.8 | 465.1 |
| 675 | 151.68 | 156.22 | 166.46 | 174.19 | 648.55 |
| 676 | 116.82 | 99.02 | 148.81 | 63.73 | 428.38 |
| 677 | 13.99 | 31.16 | 12.85 | 12.64 | 70.64 |
| 678 | 90.85 | 76.45 | 64.16 | 89.66 | 321.12 |
| 679 | 48.46 | 75.17 | 62.09 | 76.26 | 261.98 |
| TOTAL | 1,917.65 | 2,074.87 | 2,018.32 | 1,977.83 | 7,988.67 |

STATE: KENTUCKY

| ZIP CODE | QUARTER 1 | QUARTER 2 | QUARTER 3 | QUARTER 4 | TOTAL GRAMS |
|---|---|---|---|---|---|
| 400 | 111.16 | 68.85 | 49.89 | 71.46 | 301.36 |
| 401 | 67.76 | 86.28 | 41.4 | 48.59 | 244.03 |
| 402 | 343.09 | 329.03 | 420.54 | 457.86 | 1,550.52 |
| 403 | 69.79 | 80.18 | 91.62 | 93.14 | 334.73 |
| 404 | 64.49 | 67.41 | 62.97 | 73.42 | 268.29 |
| 405 | 118.81 | 91.83 | 104.38 | 96.85 | 411.87 |
| 406 | 13.51 | 16.34 | 4.36 | 6.54 | 40.75 |
| 407 | 43.79 | 59.04 | 54.25 | 48.37 | 205.45 |
| 408 | 56.87 | 38.56 | 39 | 42.7 | 177.13 |
| 409 | 92.53 | 110.57 | 159.59 | 69.39 | 432.08 |
| 410 | 132.51 | 161.12 | 144.45 | 161.66 | 599.74 |
| 411 | 76.26 | 34.86 | 48.19 | 51.2 | 210.51 |
| 412 | 0 | 4.36 | 0 | 0 | 4.36 |
| 413 | 17.43 | 15.25 | 15.8 | 17.32 | 65.8 |
| 414 | 0 | 0 | 0.44 | 0 | 0.44 |
| 415 | 31.92 | 41.29 | 23.31 | 29.74 | 126.26 |

RETAIL DRUG DISTRIBUTION BY ZIP CODE WITHIN STATE BY GRAMS WT

STATE:KENTUCKY

| ZIP CODE | QUARTER 1 | QUARTER 2 | QUARTER 3 | QUARTER 4 | TOTAL GRAMS |
|---|---|---|---|---|---|
| 416 | 7.41 | 12.2 | 16.34 | 7.19 | 43.14 |
| 417 | 4.03 | 8.28 | 3.7 | 23.31 | 39.32 |
| 418 | 67.21 | 44.66 | 72.99 | 53.38 | 238.24 |
| 420 | 442.63 | 418.89 | 305.87 | 279.45 | 1,446.84 |
| 421 | 421.76 | 468.74 | 341.98 | 395.31 | 1,627.79 |
| 422 | 236.07 | 153.28 | 255.35 | 148.48 | 793.18 |
| 423 | 169.92 | 206.48 | 179.31 | 151.25 | 706.96 |
| 424 | 205.24 | 224.72 | 206.22 | 179.49 | 815.67 |
| 425 | 110.24 | 69.07 | 88.89 | 68.52 | 336.72 |
| 426 | 19.65 | 28.11 | 58.39 | 51.42 | 157.57 |
| 427 | 123.4 | 135.69 | 117.43 | 153.19 | 529.71 |
| TOTAL | 3,047.48 | 2,975.09 | 2,906.66 | 2,779.23 | 11,708.46 |

STATE: LOUISIANA

| ZIP CODE | QUARTER 1 | QUARTER 2 | QUARTER 3 | QUARTER 4 | TOTAL GRAMS |
|---|---|---|---|---|---|
| 700 | 1,039.05 | 1,053.75 | 1,035.78 | 846.44 | 3,975.02 |
| 701 | 245.55 | 176.48 | 211.56 | 189.99 | 823.58 |
| 703 | 623.47 | 434.57 | 550.83 | 492.72 | 2,101.59 |
| 704 | 954.68 | 1,012.36 | 756.03 | 910.83 | 3,633.90 |
| 705 | 2,277.71 | 2,028.11 | 1,913.40 | 1,769.04 | 7,988.26 |
| 706 | 938.3 | 710.34 | 775.13 | 670.1 | 3,093.87 |
| 707 | 624.76 | 549.05 | 622.03 | 479.33 | 2,275.17 |
| 708 | 640.71 | 666.68 | 634.67 | 615.82 | 2,557.88 |
| 710 | 374.13 | 303.28 | 250.23 | 389.23 | 1,316.87 |
| 711 | 1,096.02 | 994.05 | 990.46 | 868.67 | 3,949.20 |
| 712 | 735.68 | 560.7 | 675.52 | 587.98 | 2,559.88 |
| 713 | 532.92 | 554.17 | 448.82 | 495.56 | 2,031.47 |
| 714 | 322.91 | 194.43 | 235.52 | 265.26 | 1,018.12 |
| TOTAL | 10,405.89 | 9,237.97 | 9,099.98 | 8,580.97 | 37,324.81 |

STATE: MAINE

| ZIP CODE | QUARTER 1 | QUARTER 2 | QUARTER 3 | QUARTER 4 | TOTAL GRAMS |
|---|---|---|---|---|---|
| 39 | 74.73 | 49.24 | 32.9 | 11.11 | 167.98 |
| 40 | 116.56 | 105.67 | 136.06 | 133.78 | 492.07 |
| 41 | 37.26 | 7.52 | 8.28 | 30.5 | 83.56 |
| 42 | 18.52 | 7.84 | 24.18 | 15.69 | 66.23 |
| 43 | 0 | 8.28 | 10.46 | 8.72 | 27.46 |
| 44 | 154.69 | 115.8 | 118.52 | 158.4 | 547.41 |
| 45 | 17.43 | 6.1 | 0 | 0 | 23.53 |
| 46 | 81.92 | 40.52 | 54.03 | 33.99 | 210.46 |
| 47 | 49.68 | 38.35 | 34.21 | 39 | 161.24 |
| 48 | 4.36 | 4.36 | 4.36 | 5.45 | 18.53 |
| 49 | 39.22 | 97.28 | 32.03 | 41.83 | 210.36 |
| TOTAL | 594.37 | 480.96 | 455.03 | 478.47 | 2,008.83 |

STATE: MARYLAND

| ZIP CODE | QUARTER 1 | QUARTER 2 | QUARTER 3 | QUARTER 4 | TOTAL GRAMS |
|---|---|---|---|---|---|
| 206 | 203.5 | 142.38 | 181.71 | 149.35 | 676.94 |
| 207 | 302.41 | 320.82 | 318.64 | 285.2 | 1,227.07 |
| 208 | 290.97 | 291.41 | 330.73 | 337.27 | 1,250.38 |
| 209 | 56.65 | 19.83 | 83.23 | 32.9 | 192.61 |
| 210 | 641.42 | 723.13 | 640.23 | 580.75 | 2,585.53 |
| 211 | 292.39 | 320.49 | 161.77 | 145.54 | 920.19 |
| 212 | 531.07 | 545.78 | 505.14 | 554.82 | 2,136.81 |
| 214 | 135.41 | 144.12 | 124.52 | 176.3 | 580.35 |
| 215 | 80.72 | 133.99 | 115.69 | 91.51 | 421.91 |
| 216 | 142.32 | 89.11 | 130.29 | 128.55 | 490.27 |
| 217 | 129.42 | 122.99 | 153.38 | 125.93 | 531.72 |
| 218 | 99.57 | 185.41 | 101.97 | 113.73 | 500.68 |

| | | | | | |
|---|---|---|---|---|---|
| 219 | 17.87 | 16.56 | 10.89 | 9.59 | 54.91 |
| TOTAL | 2,923.72 | 3,056.02 | 2,858.19 | 2,731.44 | 11,569.37 |

STATE: MASSACHUSETTS

| ZIP CODE | QUARTER 1 | QUARTER 2 | QUARTER 3 | QUARTER 4 | TOTAL GRAMS |
|---|---|---|---|---|---|
| 10 | 432.7 | 381.5 | 393.92 | 385.64 | 1,593.76 |
| 11 | 34.42 | 51.42 | 25.71 | 51.75 | 163.3 |
| 12 | 13.73 | 23.53 | 15.03 | 23.09 | 75.38 |
| 13 | 21.79 | 22.33 | 17.87 | 28.54 | 90.53 |
| 14 | 128.98 | 121.79 | 146.96 | 109.59 | 507.32 |
| 15 | 86.06 | 54.9 | 77.13 | 50.98 | 269.07 |

DRUG CODE:9230DRUG NAME:MEPERIDINE (PETHIDINE)
STATE:MASSACHUSETTS

| ZIP CODE | QUARTER 1 | QUARTER 2 | QUARTER 3 | QUARTER 4 | TOTAL GRAMS |
|---|---|---|---|---|---|
| 16 | 40.09 | 32.57 | 24.18 | 18.63 | 115.47 |
| 17 | 143.47 | 163.52 | 119.07 | 146.19 | 572.25 |
| 18 | 145.1 | 144.67 | 184.32 | 220.49 | 694.58 |
| 19 | 251.86 | 247.51 | 247.51 | 144.02 | 890.9 |
| 20 | 266.9 | 300.56 | 215.91 | 154.04 | 937.41 |
| 21 | 371.85 | 524.86 | 502.29 | 404.48 | 1,803.48 |
| 22 | 48.59 | 54.9 | 52.07 | 32.9 | 188.46 |
| 23 | 276.27 | 319.62 | 202.84 | 321.15 | 1,119.88 |
| 24 | 127.78 | 134.21 | 129.42 | 162.21 | 553.62 |
| 25 | 135.95 | 114.6 | 83.23 | 134.65 | 468.43 |
| 26 | 49.68 | 21.57 | 26.36 | 21.13 | 118.74 |
| 27 | 168.2 | 169.29 | 177.79 | 188.03 | 703.31 |
| TOTAL | 2,743.42 | 2,883.35 | 2,641.61 | 2,597.51 | 10,865.89 |

STATE: MICHIGAN

| ZIP CODE | QUARTER 1 | QUARTER 2 | QUARTER 3 | QUARTER 4 | TOTAL GRAMS |
|---|---|---|---|---|---|
| 480 | 528.13 | 436.73 | 383.35 | 398.6 | 1,746.81 |
| 481 | 362 | 368.86 | 388.69 | 340.43 | 1,459.98 |
| 482 | 77.78 | 67.54 | 93.03 | 52.07 | 290.42 |
| 483 | 147.72 | 226.26 | 79.31 | 98.04 | 551.33 |
| 484 | 146.19 | 153.21 | 146.41 | 128.11 | 573.92 |
| 485 | 71.03 | 32.79 | 54.47 | 72.66 | 230.95 |
| 486 | 415.1 | 453.62 | 380.76 | 410.48 | 1,659.96 |
| 487 | 15.95 | 36.82 | 39 | 35.95 | 127.72 |
| 488 | 259.69 | 219.4 | 252.52 | 171.14 | 902.75 |
| 489 | 301.65 | 126.59 | 133.67 | 215.81 | 777.72 |
| 490 | 384.88 | 407.21 | 387.05 | 384.77 | 1,563.91 |
| 492 | 168.85 | 133.78 | 211.12 | 210.9 | 724.65 |
| 493 | 73.64 | 87.8 | 86.71 | 60.57 | 308.72 |
| 494 | 120.38 | 128.76 | 127.78 | 150.88 | 527.8 |
| 495 | 272.13 | 332.7 | 225.5 | 241.84 | 1,072.17 |
| 496 | 76.26 | 76.15 | 80.83 | 99.35 | 332.59 |
| 497 | 257.96 | 280.08 | 268.31 | 235.96 | 1,042.31 |
| 498 | 154.69 | 135.21 | 102.18 | 49.89 | 441.97 |
| 499 | 29.63 | 51.2 | 42.92 | 52.73 | 176.48 |
| TOTAL | 3,863.66 | 3,754.71 | 3,483.61 | 3,410.18 | 14,512.16 |

STATE: MINNESOTA

| ZIP CODE | QUARTER 1 | QUARTER 2 | QUARTER 3 | QUARTER 4 | TOTAL GRAMS |
|---|---|---|---|---|---|
| 550 | 71.68 | 57.3 | 91.94 | 96.3 | 317.22 |
| 551 | 35.73 | 48.15 | 27.67 | 48.8 | 160.35 |
| 553 | 125.82 | 83.01 | 93.58 | 90.74 | 393.15 |
| 554 | 105.23 | 110.14 | 71.68 | 100.44 | 387.49 |
| 556 | 0 | 5.66 | 8.72 | 4.36 | 18.74 |
| 557 | 31.81 | 49.46 | 41.83 | 60.79 | 183.89 |

| | | | | | |
|---|---|---|---|---|---|
| 558 | 113.08 | 129.64 | 123.32 | 97.39 | 463.43 |
| 559 | 111.99 | 143.36 | 87.37 | 128.55 | 471.27 |
| 560 | 26.15 | 30.72 | 16.12 | 25.93 | 98.92 |
| 561 | 5.66 | 3.27 | 5.01 | 1.31 | 15.25 |
| 562 | 36.6 | 18.41 | 31.37 | 29.41 | 115.79 |
| 563 | 46.84 | 39.22 | 43.03 | 47.93 | 177.02 |
| 564 | 24.18 | 47.41 | 54.51 | 48.19 | 174.29 |
| 565 | 55.99 | 34.21 | 40.09 | 34.21 | 164.5 |
| 566 | 11.77 | 6.54 | 12.85 | 8.72 | 39.88 |
| 567 | 36.82 | 37.47 | 38.35 | 34.64 | 147.28 |
| TOTAL | 839.35 | 843.97 | 787.44 | 857.71 | 3,328.47 |

STATE: MISSISSIPPI

| ZIP CODE | QUARTER 1 | QUARTER 2 | QUARTER 3 | QUARTER 4 | TOTAL GRAMS |
|---|---|---|---|---|---|
| 386 | 738.05 | 628.44 | 654.5 | 642.08 | 2,663.07 |
| 387 | 466.43 | 412.33 | 408.08 | 406.18 | 1,693.02 |
| 388 | 496.19 | 454.4 | 453.62 | 398.28 | 1,802.49 |
| 389 | 343.33 | 264.7 | 350.5 | 289.01 | 1,247.54 |
| 390 | 443.27 | 472.79 | 412.55 | 400.56 | 1,729.17 |
| 391 | 588.07 | 559.15 | 471.15 | 371.04 | 1,989.41 |
| 392 | 525.84 | 613.6 | 475.19 | 401.11 | 2,015.74 |
| 393 | 445.6 | 461.02 | 427.78 | 437.84 | 1,772.24 |
| 394 | 520.2 | 579.59 | 545.04 | 489.96 | 2,134.79 |
| 395 | 839.15 | 1,047.11 | 967.84 | 973.79 | 3,827.89 |

DRUG CODE:9230DRUG NAME:MEPERIDINE (PETHIDINE)
STATE:MISSISSIPPI

| ZIP CODE | QUARTER 1 | QUARTER 2 | QUARTER 3 | QUARTER 4 | TOTAL GRAMS |
|---|---|---|---|---|---|
| 396 | 225.65 | 242.71 | 182.8 | 231.06 | 882.22 |
| 397 | 252.65 | 292.63 | 184.95 | 239.58 | 969.81 |
| TOTAL | 5,884.43 | 6,028.47 | 5,534 | 5,280.49 | 22,727.39 |

STATE: MISSOURI

| ZIP CODE | QUARTER 1 | QUARTER 2 | QUARTER 3 | QUARTER 4 | TOTAL GRAMS |
|---|---|---|---|---|---|
| 630 | 240.51 | 245.76 | 245.44 | 220.05 | 951.76 |
| 631 | 1,248.58 | 1,254.20 | 1,346.58 | 1,043.84 | 4,893.20 |
| 633 | 129.09 | 80.5 | 120.92 | 47.39 | 377.9 |
| 634 | 45.97 | 50.33 | 24.18 | 38.13 | 158.61 |
| 635 | 117.43 | 108.11 | 107.52 | 156.87 | 489.93 |
| 636 | 112.53 | 90.74 | 96.95 | 86.93 | 387.15 |
| 637 | 81.09 | 63.18 | 55.34 | 68.63 | 268.24 |
| 638 | 107.41 | 141.23 | 110.03 | 119.53 | 478.2 |
| 639 | 103.49 | 80.94 | 120.92 | 112.21 | 417.56 |
| 640 | 439.72 | 297.49 | 423.18 | 318.58 | 1,478.97 |
| 641 | 439.13 | 323.44 | 297.18 | 264.5 | 1,324.25 |
| 644 | 37.47 | 36.6 | 42.27 | 29.41 | 145.75 |
| 645 | 25.53 | 23.42 | 54.58 | 21.4 | 124.93 |
| 646 | 61.22 | 37.26 | 76.69 | 47.06 | 222.23 |
| 647 | 62.29 | 80.61 | 70.85 | 66.45 | 280.2 |
| 648 | 262.32 | 195.22 | 204.8 | 195.43 | 857.77 |
| 650 | 40.74 | 110.9 | 92.16 | 66.02 | 309.82 |
| 651 | 178.66 | 173.43 | 86.5 | 105.23 | 543.82 |
| 652 | 465.05 | 476.93 | 430.74 | 417.78 | 1,790.50 |
| 653 | 107.63 | 111.6 | 73.64 | 86.02 | 378.89 |
| 654 | 134.21 | 72.12 | 21.35 | 77.89 | 305.57 |
| 655 | 199.57 | 190.42 | 218.53 | 213.08 | 821.6 |
| 656 | 96.95 | 150.99 | 149.24 | 134.21 | 531.39 |
| 657 | 285.85 | 294.57 | 235.74 | 251.65 | 1,067.81 |
| 658 | 233.93 | 197.56 | 259.7 | 203.86 | 895.05 |
| TOTAL | 5,256.37 | 4,887.55 | 4,965.03 | 4,392.15 | 19,501.10 |

RETAIL DRUG DISTRIBUTION BY ZIP CODE WITHIN STATE BY GRAMS WT

STATE: MONTANA

| ZIP CODE | QUARTER 1 | QUARTER 2 | QUARTER 3 | QUARTER 4 | TOTAL GRAMS |
|---|---|---|---|---|---|
| 590 | 29.89 | 33.12 | 13.29 | 30.07 | 106.37 |
| 591 | 87.59 | 80.18 | 110.24 | 86.06 | 364.07 |
| 592 | 17.87 | 30.72 | 33.55 | 27.45 | 109.59 |
| 593 | 32.46 | 34.86 | 46.41 | 31.81 | 145.54 |
| 594 | 179.62 | 281.06 | 169.29 | 191.51 | 821.48 |
| 595 | 1.74 | 10.02 | 9.37 | 8.28 | 29.41 |
| 596 | 86.54 | 91.51 | 116.13 | 108.5 | 402.68 |
| 597 | 52.94 | 60.57 | 59.26 | 56.43 | 229.2 |
| 598 | 112.9 | 121.14 | 122.01 | 162.53 | 518.58 |
| 599 | 104.58 | 183.67 | 149.46 | 104.36 | 542.07 |
| TOTAL | 706.13 | 926.85 | 829.01 | 807 | 3,268.99 |

STATE: NEBRASKA

| ZIP CODE | QUARTER 1 | QUARTER 2 | QUARTER 3 | QUARTER 4 | TOTAL GRAMS |
|---|---|---|---|---|---|
| 680 | 88.68 | 95.43 | 59.15 | 89.22 | 332.48 |
| 681 | 207.42 | 285.42 | 236.5 | 171.79 | 901.13 |
| 683 | 13.42 | 18.08 | 17.43 | 13.29 | 62.22 |
| 684 | 31.55 | 36.6 | 14.38 | 11.2 | 93.73 |
| 685 | 153.93 | 178.35 | 111.14 | 192.25 | 635.67 |
| 686 | 23.31 | 22.01 | 13.94 | 15.25 | 74.51 |
| 687 | 26.8 | 54.69 | 64.06 | 45.97 | 191.52 |
| 688 | 116.96 | 164.06 | 100.99 | 107.46 | 489.47 |
| 689 | 58.17 | 63.84 | 56.97 | 56.87 | 235.85 |
| 690 | 17.65 | 17.65 | 13.73 | 13.51 | 62.54 |
| 691 | 26.36 | 45.86 | 32.57 | 46.19 | 150.98 |
| 692 | 7.23 | 8.58 | 15.73 | 12.72 | 44.26 |
| 693 | 38.78 | 84.54 | 22.66 | 41.18 | 187.16 |
| TOTAL | 810.26 | 1,075.11 | 759.25 | 816.9 | 3,461.52 |

STATE: NEVADA

| ZIP CODE | QUARTER 1 | QUARTER 2 | QUARTER 3 | QUARTER 4 | TOTAL GRAMS |
|---|---|---|---|---|---|
| 890 | 285.63 | 460.15 | 326.7 | 270.6 | 1,343.08 |
| 891 | 1,008.33 | 911.04 | 984.9 | 908.1 | 3,812.37 |
| 893 | 14.97 | 9.8 | 9.59 | 4.14 | 38.5 |
| 894 | 125.71 | 93.58 | 112.21 | 88.02 | 419.52 |

DRUG CODE:9230DRUG NAME:MEPERIDINE (PETHIDINE)
STATE:NEVADA

| ZIP CODE | QUARTER 1 | QUARTER 2 | QUARTER 3 | QUARTER 4 | TOTAL GRAMS |
|---|---|---|---|---|---|
| 895 | 278.36 | 230.51 | 224.3 | 160.57 | 893.74 |
| 897 | 78.48 | 88.37 | 55.56 | 102.84 | 325.25 |
| 898 | 65.91 | 39.44 | 77.89 | 32.14 | 215.38 |
| TOTAL | 1,857.39 | 1,832.89 | 1,791.15 | 1,566.41 | 7,047.84 |

STATE: NEW HAMPSHIRE

| ZIP CODE | QUARTER 1 | QUARTER 2 | QUARTER 3 | QUARTER 4 | TOTAL GRAMS |
|---|---|---|---|---|---|
| 30 | 24.84 | 59.48 | 48.8 | 52.07 | 185.19 |
| 31 | 101.31 | 78.65 | 87.37 | 78.65 | 345.98 |
| 32 | 124.41 | 129.53 | 145 | 120.27 | 519.21 |
| 33 | 192.6 | 225.17 | 181.93 | 223.98 | 823.68 |
| 34 | 4.36 | 13.94 | 30.94 | 18.52 | 67.76 |
| 35 | 41.61 | 24.84 | 37.69 | 17.21 | 121.35 |
| 37 | 37.47 | 29.85 | 38.13 | 27.45 | 132.9 |
| 38 | 214.82 | 194.89 | 160.36 | 134.65 | 704.72 |
| TOTAL | 741.42 | 756.35 | 730.22 | 672.8 | 2,900.79 |

RETAIL DRUG DISTRIBUTION BY ZIP CODE WITHIN STATE BY GRAMS WT

STATE: NEW JERSEY

| ZIP CODE | QUARTER 1 | QUARTER 2 | QUARTER 3 | QUARTER 4 | TOTAL GRAMS |
|---|---|---|---|---|---|
| 70 | 331.17 | 332.15 | 252.84 | 288.79 | 1,204.95 |
| 71 | 32.03 | 47.28 | 62.2 | 33.88 | 175.39 |
| 72 | 22.88 | 1.09 | 7.63 | 10.89 | 42.49 |
| 73 | 70.16 | 85.52 | 31.05 | 113.84 | 300.57 |
| 74 | 158.07 | 123.21 | 145.98 | 126.04 | 553.3 |
| 75 | 30.72 | 42.92 | 32.46 | 38.78 | 144.88 |
| 76 | 146.63 | 188.46 | 185.85 | 202.41 | 723.35 |
| 77 | 217.44 | 212.97 | 239.34 | 200.55 | 870.3 |
| 78 | 127.46 | 108.5 | 132.25 | 118.96 | 487.17 |
| 79 | 106.11 | 89.33 | 45.21 | 88.46 | 329.11 |
| 80 | 392.83 | 339.78 | 293.7 | 293.26 | 1,319.57 |
| 81 | 1.31 | 1.31 | 8.06 | 7.19 | 17.87 |
| 82 | 40.31 | 39.44 | 52.73 | 98.04 | 230.52 |
| 83 | 29.85 | 43.79 | 47.5 | 32.35 | 153.49 |
| 84 | 4.14 | 1.09 | 2.18 | 1.53 | 8.94 |
| 85 | 31.59 | 33.33 | 20.04 | 21.13 | 106.09 |
| 86 | 67 | 77.35 | 78.44 | 15.9 | 238.69 |
| 87 | 106 | 92.27 | 66.02 | 129.96 | 394.25 |
| 88 | 575.84 | 458.84 | 499.91 | 471.26 | 2,005.85 |
| 89 | 66.45 | 44.45 | 68.19 | 64.71 | 243.8 |
| TOTAL | 2,557.99 | 2,363.08 | 2,271.58 | 2,357.93 | 9,550.58 |

STATE: NEW MEXICO

| ZIP CODE | QUARTER 1 | QUARTER 2 | QUARTER 3 | QUARTER 4 | TOTAL GRAMS |
|---|---|---|---|---|---|
| 870 | 201.75 | 188.68 | 191.73 | 107.85 | 690.01 |
| 871 | 829.01 | 751.45 | 759.4 | 618.11 | 2,957.97 |
| 873 | 6.54 | 9.15 | 0.87 | 7.19 | 23.75 |
| 874 | 47.5 | 63.07 | 62.53 | 65.36 | 238.46 |
| 875 | 303.06 | 244.02 | 293.91 | 231.82 | 1,072.81 |
| 877 | 21.79 | 45.32 | 26.15 | 29.85 | 123.11 |
| 878 | 13.94 | 5.23 | 16.12 | 14.82 | 50.11 |
| 879 | 8.72 | 17.43 | 0 | 2.18 | 28.33 |
| 880 | 198.92 | 338.69 | 323.98 | 266.35 | 1,127.94 |
| 881 | 85.12 | 40.57 | 63.4 | 85.3 | 274.39 |
| 882 | 171.03 | 234.43 | 181.27 | 164.82 | 751.55 |
| 883 | 49.46 | 16.34 | 10.02 | 39.44 | 115.26 |
| 884 | 8.72 | 13.94 | 2.18 | 2.18 | 27.02 |
| TOTAL | 1,945.56 | 1,968.32 | 1,931.56 | 1,635.27 | 7,480.71 |

STATE: NEW YORK

| ZIP CODE | QUARTER 1 | QUARTER 2 | QUARTER 3 | QUARTER 4 | TOTAL GRAMS |
|---|---|---|---|---|---|
| 100 | 822.8 | 735.65 | 700.58 | 819.1 | 3,078.13 |
| 101 | 33.99 | 29.85 | 37.26 | 59.26 | 160.36 |
| 103 | 125.6 | 170.16 | 170.38 | 161.01 | 627.15 |
| 104 | 110.68 | 205.46 | 148.16 | 113.73 | 578.03 |
| 105 | 367.23 | 256.66 | 189.01 | 266.79 | 1,079.69 |
| 106 | 2.4 | 30.94 | 19.61 | 4.36 | 57.31 |
| 107 | 60.68 | 55.12 | 78.44 | 54.03 | 248.27 |
| 108 | 24.18 | 10.89 | 8.5 | 1.74 | 45.31 |
| 109 | 99.13 | 115.91 | 110.03 | 111.44 | 436.51 |
| 110 | 80.83 | 128.55 | 165.59 | 136.72 | 511.69 |

DRUG CODE:9230DRUG NAME:MEPERIDINE (PETHIDINE)
STATE:NEW YORK

| ZIP CODE | QUARTER 1 | QUARTER 2 | QUARTER 3 | QUARTER 4 | TOTAL GRAMS |
|---|---|---|---|---|---|
| 111 | 13.07 | 3.27 | 54.58 | 18.52 | 89.44 |
| 112 | 255.35 | 258.62 | 277.35 | 245.33 | 1,036.65 |

RETAIL DRUG DISTRIBUTION BY ZIP CODE WITHIN STATE BY GRAMS WT

| | | | | | |
|---|---|---|---|---|---|
| 113 | 155.02 | 134.43 | 112.75 | 217.44 | 619.64 |
| 114 | 71.68 | 76.47 | 69.72 | 48.37 | 266.24 |
| 115 | 128 | 77.13 | 111.12 | 113.73 | 429.98 |
| 116 | 8.72 | 5.45 | 8.72 | 9.8 | 32.69 |
| 117 | 427.47 | 376.38 | 406.99 | 409.17 | 1,620.01 |
| 118 | 78.44 | 52.29 | 91.51 | 87.15 | 309.39 |
| 119 | 19.06 | 15.58 | 14.71 | 19.17 | 68.52 |
| 120 | 174.08 | 151.42 | 140.31 | 126.15 | 591.96 |
| 121 | 90.85 | 65.58 | 63.84 | 37.04 | 257.31 |
| 122 | 32.68 | 47.28 | 59.92 | 32.9 | 172.78 |
| 123 | 68.74 | 68.41 | 116.13 | 75.17 | 328.45 |
| 124 | 130.07 | 144.02 | 125.82 | 139 | 538.91 |
| 125 | 81.49 | 96.85 | 75.71 | 64.06 | 318.11 |
| 126 | 60.46 | 84.32 | 107.85 | 55.56 | 308.19 |
| 127 | 50.66 | 104.58 | 69.18 | 33.77 | 258.19 |
| 128 | 139.88 | 119.61 | 98.7 | 85.19 | 443.38 |
| 129 | 69.72 | 135.95 | 60.24 | 22.99 | 288.9 |
| 130 | 288.9 | 278.12 | 265.15 | 181.71 | 1,013.88 |
| 131 | 52.29 | 38.13 | 17.43 | 50.76 | 158.61 |
| 132 | 110.46 | 80.94 | 134.97 | 94.45 | 420.82 |
| 133 | 63.62 | 99.57 | 71.81 | 63.01 | 298.01 |
| 134 | 70.64 | 60.55 | 64.29 | 78.78 | 274.26 |
| 135 | 52.83 | 105.89 | 67.76 | 58.72 | 285.2 |
| 136 | 30.44 | 27.34 | 15.69 | 29.74 | 103.21 |
| 137 | 3.7 | 8.93 | 10.24 | 8.28 | 31.15 |
| 138 | 145.15 | 137.74 | 179.14 | 136.83 | 598.86 |
| 139 | 4.9 | 13.94 | 15.03 | 36.17 | 70.04 |
| 140 | 98.26 | 117.11 | 106.21 | 81.27 | 402.85 |
| 141 | 50.11 | 66.02 | 63.73 | 59.92 | 239.78 |
| 142 | 372.13 | 351.98 | 355.68 | 224.74 | 1,304.53 |
| 143 | 87.69 | 59.92 | 126.37 | 62.64 | 336.62 |
| 144 | 42.16 | 25.82 | 17.1 | 15.9 | 100.98 |
| 145 | 108.5 | 100.66 | 50.11 | 39.87 | 299.14 |
| 146 | 142.82 | 78.04 | 98.48 | 75.49 | 394.83 |
| 147 | 60.96 | 50.98 | 68.85 | 57.3 | 238.09 |
| 148 | 128.33 | 164.71 | 179.53 | 135.08 | 607.65 |
| 149 | 176.04 | 70.37 | 36.17 | 18.52 | 301.1 |
| TOTAL | 5,872.89 | 5,693.59 | 5,636.45 | 5,077.87 | 22,280.80 |

STATE: NORTH CAROLINA

| ZIP CODE | QUARTER 1 | QUARTER 2 | QUARTER 3 | QUARTER 4 | TOTAL GRAMS |
|---|---|---|---|---|---|
| 270 | 331.39 | 335.09 | 346.42 | 342.5 | 1,355.40 |
| 271 | 321.58 | 301.1 | 218.31 | 238.25 | 1,079.24 |
| 272 | 418.54 | 372.35 | 420.5 | 241.19 | 1,452.58 |
| 273 | 253.82 | 345.55 | 244.46 | 162.75 | 1,006.58 |
| 274 | 231.82 | 268.64 | 264.39 | 161.45 | 926.3 |
| 275 | 802.87 | 758.1 | 737.51 | 648.29 | 2,946.77 |
| 276 | 255.79 | 300.23 | 208.07 | 209.27 | 973.36 |
| 277 | 92.38 | 30.94 | 48.37 | 77.56 | 249.25 |
| 278 | 498.76 | 493.7 | 431.17 | 518.54 | 1,942.17 |
| 279 | 252.74 | 285.42 | 248.27 | 192.6 | 979.03 |
| 280 | 782.5 | 730.32 | 698.29 | 565.06 | 2,776.17 |
| 281 | 669.2 | 556.02 | 532.81 | 705.15 | 2,463.18 |
| 282 | 716.7 | 685.54 | 696 | 623.67 | 2,721.91 |
| 283 | 392.37 | 369.19 | 325.83 | 317.99 | 1,405.38 |
| 284 | 325.72 | 291.3 | 337.49 | 306.88 | 1,261.39 |
| 285 | 318.32 | 432.59 | 401.54 | 253.39 | 1,405.84 |
| 286 | 786.96 | 710.71 | 763.22 | 526.6 | 2,787.49 |
| 287 | 295.87 | 353.5 | 326.59 | 334.76 | 1,310.72 |
| 288 | 118.63 | 102.95 | 71.03 | 47.61 | 340.22 |
| 289 | 20.7 | 17.43 | 57.74 | 26.15 | 122.02 |

| | | | | | |
|---|---|---|---|---|---|
| TOTAL | 7,886.66 | 7,740.67 | 7,378.01 | 6,499.66 | 29,505 |

STATE: NORTH DAKOTA

| ZIP CODE | QUARTER 1 | QUARTER 2 | QUARTER 3 | QUARTER 4 | TOTAL GRAMS |
|---|---|---|---|---|---|
| 580 | 25.27 | 37.69 | 44.66 | 14.6 | 122.22 |

DRUG CODE:9230DRUG NAME:MEPERIDINE (PETHIDINE)
STATE:NORTH DAKOTA

| ZIP CODE | QUARTER 1 | QUARTER 2 | QUARTER 3 | QUARTER 4 | TOTAL GRAMS |
|---|---|---|---|---|---|
| 581 | 61.88 | 55.12 | 19.83 | 45.32 | 182.15 |
| 582 | 29.63 | 44.56 | 32.46 | 9.37 | 116.02 |
| 583 | 24.18 | 34.21 | 22.01 | 22.01 | 102.41 |
| 584 | 19.61 | 10.68 | 15.47 | 10.24 | 56 |
| 585 | 80.18 | 79.09 | 75.6 | 84.32 | 319.19 |
| 586 | 0.87 | 10.68 | 8.06 | 9.59 | 29.2 |
| 587 | 59.26 | 40.09 | 28.32 | 60.13 | 187.8 |
| 588 | 1.42 | 7.63 | 4.79 | 5.45 | 19.29 |
| TOTAL | 302.3 | 319.75 | 251.2 | 261.03 | 1,134.28 |

STATE: OHIO

| ZIP CODE | QUARTER 1 | QUARTER 2 | QUARTER 3 | QUARTER 4 | TOTAL GRAMS |
|---|---|---|---|---|---|
| 430 | 181.32 | 195.11 | 214.5 | 237.37 | 828.3 |
| 431 | 244.02 | 197.87 | 222.23 | 235.96 | 900.08 |
| 432 | 237.48 | 290.43 | 686.52 | 244.56 | 1,458.99 |
| 433 | 30.28 | 44.23 | 25.71 | 35.73 | 135.95 |
| 434 | 163.67 | 192.82 | 126.63 | 113.3 | 596.42 |
| 435 | 143.84 | 83.99 | 114.82 | 66.34 | 408.99 |
| 436 | 102.62 | 113.51 | 84.32 | 114.6 | 415.05 |
| 437 | 178.68 | 201.86 | 157.31 | 197.83 | 735.68 |
| 438 | 3.22 | 3.27 | 0 | 0 | 6.49 |
| 439 | 95.65 | 110.14 | 57.08 | 60.9 | 323.77 |
| 440 | 276.81 | 411.54 | 268.86 | 328.66 | 1,285.87 |
| 441 | 1,068.57 | 1,020.31 | 1,093.62 | 808.64 | 3,991.14 |
| 442 | 191.95 | 161.01 | 155.78 | 123.86 | 632.6 |
| 443 | 99.46 | 85.19 | 156.22 | 85.95 | 426.82 |
| 444 | 68.98 | 52.51 | 65.82 | 94.49 | 281.8 |
| 445 | 92.6 | 113.51 | 78 | 88.35 | 372.46 |
| 446 | 203.58 | 160.86 | 150.55 | 143.62 | 658.61 |
| 447 | 71.57 | 82.9 | 79.09 | 96.08 | 329.64 |
| 448 | 214.17 | 247.64 | 211.86 | 231.36 | 905.03 |
| 449 | 22.88 | 30.28 | 36.71 | 49.13 | 139 |
| 450 | 379.97 | 325.61 | 308.12 | 265.81 | 1,279.51 |
| 451 | 91.51 | 93.69 | 162.75 | 43.68 | 391.63 |
| 452 | 737.4 | 777.81 | 716.59 | 703.2 | 2,935 |
| 453 | 225.59 | 196.63 | 162.21 | 208.62 | 793.05 |
| 454 | 232.58 | 235.96 | 320.71 | 307.86 | 1,097.11 |
| 455 | 43.79 | 28.76 | 41.4 | 21.79 | 135.74 |
| 456 | 35.43 | 55.56 | 57.3 | 54.36 | 202.65 |
| 457 | 33.55 | 33.55 | 24.51 | 28.54 | 120.15 |
| 458 | 196.57 | 184.98 | 198.57 | 191.29 | 771.41 |
| TOTAL | 5,667.74 | 5,731.53 | 5,977.79 | 5,181.88 | 22,558.94 |

STATE: OKLAHOMA

| ZIP CODE | QUARTER 1 | QUARTER 2 | QUARTER 3 | QUARTER 4 | TOTAL GRAMS |
|---|---|---|---|---|---|
| 730 | 1,640.25 | 1,514.43 | 1,590.36 | 1,485.56 | 6,230.60 |
| 731 | 1,743.46 | 1,916.12 | 1,957.41 | 1,879.98 | 7,496.97 |
| 734 | 147.72 | 135.52 | 67 | 59.26 | 409.5 |
| 735 | 906.34 | 880.24 | 814.94 | 781.93 | 3,383.45 |
| 736 | 157.41 | 150.12 | 132.9 | 90.96 | 531.39 |

| | | | | | |
|---|---|---|---|---|---|
| 737 | 144.76 | 261.45 | 218.96 | 301.1 | 926.27 |
| 738 | 42.49 | 92.49 | 46.41 | 9.8 | 191.19 |
| 739 | 182.14 | 79.96 | 21.57 | 22.44 | 306.11 |
| 740 | 1,577.20 | 1,428.52 | 1,437.32 | 1,274.90 | 5,717.94 |
| 741 | 783.59 | 685.54 | 845.9 | 786.96 | 3,101.99 |
| 743 | 207.02 | 196.09 | 191.95 | 209.6 | 804.66 |
| 744 | 436.25 | 519.96 | 449.8 | 474.53 | 1,880.54 |
| 745 | 279.03 | 323.44 | 304.74 | 226.05 | 1,133.26 |
| 746 | 67.76 | 69.72 | 24.29 | 48.91 | 210.68 |
| 747 | 283.56 | 208.51 | 308.73 | 173.32 | 974.12 |
| 748 | 474.86 | 434.01 | 327.68 | 362.76 | 1,599.31 |
| 749 | 251.45 | 196.52 | 212.75 | 242.82 | 903.54 |
| TOTAL | 9,325.29 | 9,092.64 | 8,952.71 | 8,430.88 | 35,801.52 |

STATE: OREGON

| ZIP CODE | QUARTER 1 | QUARTER 2 | QUARTER 3 | QUARTER 4 | TOTAL GRAMS |
|---|---|---|---|---|---|
| 970 | 338.9 | 378.78 | 397.95 | 430.85 | 1,546.48 |
| 971 | 208.72 | 139.66 | 160.03 | 133.88 | 642.29 |
| 972 | 264.28 | 265.59 | 328.77 | 247.29 | 1,105.93 |

DRUG CODE:9230DRUG NAME:MEPERIDINE (PETHIDINE)
STATE:OREGON

| ZIP CODE | QUARTER 1 | QUARTER 2 | QUARTER 3 | QUARTER 4 | TOTAL GRAMS |
|---|---|---|---|---|---|
| 973 | 244.46 | 306.99 | 325.07 | 313.3 | 1,189.82 |
| 974 | 344.46 | 302.13 | 239.77 | 255.13 | 1,141.49 |
| 975 | 204.15 | 220.49 | 143.25 | 169.51 | 737.4 |
| 976 | 52.51 | 69.5 | 65.36 | 65.36 | 252.73 |
| 977 | 54.69 | 65.36 | 57.3 | 64.93 | 242.28 |
| 978 | 79.31 | 71.51 | 102.73 | 74.95 | 328.5 |
| 979 | 21.24 | 26.04 | 6.75 | 21.24 | 75.27 |
| TOTAL | 1,812.72 | 1,846.05 | 1,826.98 | 1,776.44 | 7,262.19 |

STATE: PENNSYLVANIA

| ZIP CODE | QUARTER 1 | QUARTER 2 | QUARTER 3 | QUARTER 4 | TOTAL GRAMS |
|---|---|---|---|---|---|
| 150 | 73.97 | 74.19 | 102.18 | 145.98 | 396.32 |
| 151 | 91.29 | 87.69 | 110.9 | 67.76 | 357.64 |
| 152 | 232.58 | 207.63 | 224.85 | 211.56 | 876.62 |
| 153 | 39.87 | 29.41 | 53.6 | 30.72 | 153.6 |
| 154 | 28.11 | 27.45 | 10.02 | 23.75 | 89.33 |
| 155 | 11.94 | 16.34 | 22.88 | 8.28 | 59.44 |
| 156 | 42.7 | 32.9 | 35.51 | 30.94 | 142.05 |
| 157 | 22.75 | 25.71 | 59.37 | 10.02 | 117.85 |
| 158 | 129.2 | 81.05 | 147.5 | 71.25 | 429 |
| 159 | 119.29 | 79.79 | 84.21 | 119.61 | 402.9 |
| 160 | 101.53 | 78.87 | 48.59 | 54.47 | 283.46 |
| 161 | 131.05 | 28.11 | 63.62 | 40.96 | 263.74 |
| 162 | 12.2 | 10.89 | 15.47 | 0.87 | 39.43 |
| 163 | 27.45 | 74.51 | 43.79 | 47.93 | 193.68 |
| 164 | 88.24 | 9.8 | 14.16 | 4.36 | 116.56 |
| 165 | 39.22 | 50.87 | 56.21 | 40.31 | 186.61 |
| 166 | 128.59 | 108.07 | 140.75 | 111.12 | 488.53 |
| 167 | 0 | 0 | 0 | 1.09 | 1.09 |
| 168 | 52.94 | 53.16 | 41.18 | 46.41 | 193.69 |
| 169 | 5.66 | 0.44 | 19.83 | 4.14 | 30.07 |
| 170 | 166.67 | 176.04 | 228.01 | 171.25 | 741.97 |
| 171 | 43.9 | 44.23 | 88.68 | 39.76 | 216.57 |
| 172 | 50.98 | 61.22 | 50.55 | 9.59 | 172.34 |
| 173 | 204.15 | 143.36 | 227.79 | 184.54 | 759.84 |
| 174 | 89.66 | 99.13 | 145.98 | 129.09 | 463.86 |
| 175 | 93.03 | 83.88 | 84.64 | 72.12 | 333.67 |

| | | | | | |
|---|---|---|---|---|---|
| 176 | 40.74 | 35.51 | 81.05 | 39.54 | 196.84 |
| 177 | 47.06 | 67.32 | 52.73 | 86.93 | 254.04 |
| 178 | 92.81 | 74.73 | 102.18 | 71.25 | 340.97 |
| 179 | 68.63 | 41.61 | 27.02 | 41.83 | 179.09 |
| 180 | 76.26 | 71.57 | 72.55 | 55.78 | 276.16 |
| 181 | 79.2 | 124.84 | 158.83 | 146.52 | 509.39 |
| 182 | 32.46 | 31.59 | 38.89 | 41.51 | 144.45 |
| 183 | 21.79 | 30.5 | 17.43 | 8.72 | 78.44 |
| 184 | 73.25 | 25.93 | 81.49 | 50.98 | 231.65 |
| 185 | 59.48 | 92.81 | 89.11 | 76.47 | 317.87 |
| 186 | 64.49 | 63.62 | 52.83 | 79.09 | 260.03 |
| 187 | 416.14 | 342.5 | 357.97 | 343.81 | 1,460.42 |
| 188 | 86.89 | 108.85 | 111.12 | 88.24 | 395.1 |
| 189 | 47.61 | 36.06 | 50.98 | 55.34 | 189.99 |
| 190 | 591.64 | 418.76 | 522.68 | 584.12 | 2,117.20 |
| 191 | 325.83 | 362.87 | 393.92 | 336.29 | 1,418.91 |
| 193 | 290.54 | 249.79 | 170.38 | 217.55 | 928.26 |
| 194 | 133.12 | 153.49 | 154.8 | 131.71 | 573.12 |
| 195 | 36.17 | 36.71 | 31.81 | 37.47 | 142.16 |
| 196 | 94.56 | 52.29 | 28.98 | 57.3 | 233.13 |
| TOTAL | 4,705.64 | 4,106.09 | 4,717.02 | 4,228.33 | 17,757.08 |

STATE: PUERTO RICO

| ZIP CODE | QUARTER 1 | QUARTER 2 | QUARTER 3 | QUARTER 4 | TOTAL GRAMS |
|---|---|---|---|---|---|
| 6 | 1,942.03 | 2,027.54 | 1,754.33 | 1,681.34 | 7,405.24 |
| 7 | 2,139.10 | 2,085.28 | 2,083.21 | 1,570.66 | 7,878.25 |
| 9 | 2,479.96 | 2,581.06 | 2,659.93 | 2,236.92 | 9,957.87 |
| TOTAL | 6,561.09 | 6,693.88 | 6,497.47 | 5,488.92 | 25,241.36 |

STATE: RHODE ISLAND

| ZIP CODE | QUARTER 1 | QUARTER 2 | QUARTER 3 | QUARTER 4 | TOTAL GRAMS |
|---|---|---|---|---|---|
| 28 | 94.45 | 86.06 | 128.66 | 131.92 | 441.09 |

DRUG CODE:9230DRUG NAME:MEPERIDINE (PETHIDINE)
STATE:RHODE ISLAND

| ZIP CODE | QUARTER 1 | QUARTER 2 | QUARTER 3 | QUARTER 4 | TOTAL GRAMS |
|---|---|---|---|---|---|
| 29 | 141.84 | 134.65 | 123.1 | 110.14 | 509.73 |
| TOTAL | 236.29 | 220.71 | 251.76 | 242.06 | 950.82 |

STATE: SOUTH CAROLINA

| ZIP CODE | QUARTER 1 | QUARTER 2 | QUARTER 3 | QUARTER 4 | TOTAL GRAMS |
|---|---|---|---|---|---|
| 290 | 940.35 | 915.73 | 882.61 | 832.83 | 3,571.52 |
| 291 | 807.77 | 774.33 | 697.42 | 585.87 | 2,865.39 |
| 292 | 1,358.67 | 1,066.72 | 1,082.40 | 1,063.34 | 4,571.13 |
| 293 | 602.53 | 555.25 | 442.29 | 518.98 | 2,119.05 |
| 294 | 1,182.19 | 1,013.45 | 1,090.25 | 975.54 | 4,261.43 |
| 295 | 980.98 | 952.66 | 892.42 | 783.37 | 3,609.43 |
| 296 | 1,611.45 | 1,426.10 | 1,216.29 | 1,152.99 | 5,406.83 |
| 297 | 331.28 | 400.45 | 334 | 279.75 | 1,345.48 |
| 298 | 561.25 | 435.97 | 401.98 | 446.86 | 1,846.06 |
| 299 | 269.08 | 181.05 | 250.67 | 193.26 | 894.06 |
| TOTAL | 8,645.55 | 7,721.71 | 7,290.33 | 6,832.79 | 30,490.38 |

STATE: SOUTH DAKOTA

| ZIP CODE | QUARTER 1 | QUARTER 2 | QUARTER 3 | QUARTER 4 | TOTAL GRAMS |
|---|---|---|---|---|---|
| 570 | 189.99 | 182.14 | 185.85 | 144.89 | 702.87 |
| 571 | 187.15 | 205.67 | 169.94 | 162.75 | 725.51 |
| 572 | 38.13 | 33.12 | 39.44 | 27.45 | 138.14 |
| 573 | 84.97 | 57.63 | 39.22 | 31.48 | 213.3 |

RETAIL DRUG DISTRIBUTION BY ZIP CODE WITHIN STATE BY GRAMS WT

| | | | | | |
|---|---|---|---|---|---|
| 574 | 58.39 | 21.35 | 42.16 | 40.09 | 161.99 |
| 575 | 9.59 | 15.9 | 12.42 | 22.88 | 60.79 |
| 576 | 8.72 | 2.18 | 0 | 5.23 | 16.13 |
| 577 | 133.67 | 157.09 | 142.71 | 105.02 | 538.49 |
| TOTAL | 710.61 | 675.08 | 631.74 | 539.79 | 2,557.22 |

STATE: TENNESSEE

| ZIP CODE | QUARTER 1 | QUARTER 2 | QUARTER 3 | QUARTER 4 | TOTAL GRAMS |
|---|---|---|---|---|---|
| 370 | 2,062.51 | 1,649.86 | 1,952.70 | 1,381.98 | 7,047.05 |
| 371 | 1,417.49 | 1,397.34 | 1,166.28 | 1,318.58 | 5,299.69 |
| 372 | 940.89 | 697.64 | 880.76 | 757.88 | 3,277.17 |
| 373 | 1,205.79 | 1,100.49 | 935.56 | 958.8 | 4,200.64 |
| 374 | 506.12 | 380.24 | 417.23 | 420.17 | 1,723.76 |
| 376 | 298.82 | 310.8 | 261.89 | 228.44 | 1,099.95 |
| 377 | 305.77 | 225.5 | 270.82 | 287.27 | 1,089.36 |
| 378 | 462.64 | 418.91 | 366.14 | 394.35 | 1,642.04 |
| 379 | 311.74 | 297.51 | 291.93 | 299.51 | 1,200.69 |
| 380 | 578.81 | 506.89 | 461.68 | 432.26 | 1,979.64 |
| 381 | 812.87 | 754.09 | 845.03 | 826.29 | 3,238.28 |
| 382 | 298.88 | 265.94 | 301.06 | 201.1 | 1,066.98 |
| 383 | 602.42 | 553.95 | 568.92 | 509.61 | 2,234.90 |
| 384 | 379.87 | 338.8 | 289.23 | 328.12 | 1,336.02 |
| 385 | 355.94 | 317.99 | 345.11 | 172.34 | 1,191.38 |
| TOTAL | 10,540.56 | 9,215.95 | 9,354.34 | 8,516.70 | 37,627.55 |

STATE: TEXAS

| ZIP CODE | QUARTER 1 | QUARTER 2 | QUARTER 3 | QUARTER 4 | TOTAL GRAMS |
|---|---|---|---|---|---|
| 750 | 889.08 | 944.58 | 906.64 | 768.1 | 3,508.40 |
| 751 | 242.87 | 192.17 | 142.6 | 145.87 | 723.51 |
| 752 | 534.9 | 542.81 | 450.7 | 459.32 | 1,987.73 |
| 753 | 6.86 | 16.89 | 16.67 | 8.93 | 49.35 |
| 754 | 210.82 | 149.51 | 140.25 | 242.84 | 743.42 |
| 755 | 179.96 | 106.76 | 121.9 | 104.04 | 512.66 |
| 756 | 694.56 | 359.97 | 372.5 | 266.07 | 1,693.10 |
| 757 | 209.55 | 243.58 | 200.38 | 192.67 | 846.18 |
| 758 | 54.34 | 26.15 | 18.17 | 13.12 | 111.78 |
| 759 | 274.06 | 167.74 | 197.66 | 193.8 | 833.26 |
| 760 | 489.7 | 488.17 | 572.64 | 466.8 | 2,017.31 |
| 761 | 595.45 | 561.35 | 603.4 | 504.27 | 2,264.47 |
| 762 | 260.14 | 224.85 | 266.13 | 269.18 | 1,020.30 |
| 763 | 138.22 | 153.3 | 164.5 | 98.59 | 554.61 |
| 764 | 75.58 | 61.38 | 67.28 | 46.56 | 250.8 |
| 765 | 193.43 | 231.06 | 215.26 | 86.82 | 726.57 |
| 766 | 61.31 | 61.01 | 49.57 | 57.52 | 229.41 |
| 767 | 125.06 | 84.1 | 112.86 | 72.12 | 394.14 |
| 768 | 67.04 | 52.83 | 78.87 | 43.58 | 242.32 |
| 769 | 100.33 | 207.74 | 108.61 | 99.68 | 516.36 |

DRUG CODE:9230DRUG NAME:MEPERIDINE (PETHIDINE)
STATE:TEXAS

| ZIP CODE | QUARTER 1 | QUARTER 2 | QUARTER 3 | QUARTER 4 | TOTAL GRAMS |
|---|---|---|---|---|---|
| 770 | 2,026.28 | 2,468.20 | 2,470.35 | 2,317.21 | 9,282.04 |
| 773 | 445.12 | 381.32 | 460.48 | 481.77 | 1,768.69 |
| 774 | 257.46 | 297.73 | 335.85 | 324.42 | 1,215.46 |
| 775 | 651.66 | 537.82 | 571.53 | 551.44 | 2,312.45 |
| 776 | 154.8 | 171.58 | 179.75 | 166.35 | 672.48 |
| 777 | 147.83 | 175.06 | 128.11 | 133.45 | 584.45 |
| 778 | 92.05 | 118.2 | 157.52 | 132.25 | 500.02 |
| 779 | 76.37 | 89.22 | 101.9 | 75.93 | 343.42 |
| 780 | 210.29 | 238.07 | 197.29 | 219.4 | 865.05 |

| | | | | | |
|---|---|---|---|---|---|
| 781 | 71.66 | 97.61 | 76.47 | 90.09 | 335.83 |
| 782 | 949.19 | 1,002.05 | 916.93 | 833.96 | 3,702.13 |
| 783 | 71.9 | 97.83 | 59.92 | 64.27 | 293.92 |
| 784 | 516.84 | 450.28 | 460.48 | 407.25 | 1,834.85 |
| 785 | 987.71 | 1,191.28 | 1,059.09 | 1,011.46 | 4,249.54 |
| 786 | 288.36 | 268.29 | 222.12 | 249.68 | 1,028.45 |
| 787 | 320.49 | 300.45 | 310.25 | 297.94 | 1,229.13 |
| 788 | 103.71 | 110.07 | 73.31 | 44.45 | 331.54 |
| 789 | 34.42 | 42.49 | 32.51 | 30.94 | 140.36 |
| 790 | 83.25 | 29.06 | 42.38 | 49.89 | 204.58 |
| 791 | 354.05 | 279.53 | 303.06 | 155.78 | 1,092.42 |
| 792 | 67.11 | 35.95 | 48.48 | 41.94 | 193.48 |
| 793 | 103.71 | 75.99 | 72.55 | 84.86 | 337.11 |
| 794 | 168.24 | 214.5 | 197.98 | 183.23 | 763.95 |
| 795 | 145.1 | 122.66 | 80.4 | 102.62 | 450.78 |
| 796 | 178.44 | 138.79 | 128.22 | 116.13 | 561.58 |
| 797 | 558.3 | 560.81 | 443.27 | 443.92 | 2,006.30 |
| 798 | 2.18 | 15.25 | 0 | 13.07 | 30.5 |
| 799 | 290.86 | 359.06 | 193.04 | 212.86 | 1,055.82 |
| TOTAL | 14,760.64 | 14,745.10 | 14,129.83 | 12,976.44 | 56,612.01 |

STATE: UTAH

| ZIP CODE | QUARTER 1 | QUARTER 2 | QUARTER 3 | QUARTER 4 | TOTAL GRAMS |
|---|---|---|---|---|---|
| 840 | 961.79 | 970.5 | 854.38 | 936.71 | 3,723.38 |
| 841 | 468.65 | 424.68 | 581.4 | 367.77 | 1,842.50 |
| 843 | 189.55 | 95.65 | 103.27 | 129.2 | 517.67 |
| 844 | 220.05 | 151.42 | 146.85 | 226.37 | 744.69 |
| 845 | 129.96 | 132.36 | 132.36 | 94.34 | 489.02 |
| 846 | 180.4 | 202.23 | 240.97 | 202.95 | 826.55 |
| 847 | 227.44 | 236.81 | 260.58 | 234.22 | 959.05 |
| TOTAL | 2,377.84 | 2,213.65 | 2,319.81 | 2,191.56 | 9,102.86 |

STATE: VERMONT

| ZIP CODE | QUARTER 1 | QUARTER 2 | QUARTER 3 | QUARTER 4 | TOTAL GRAMS |
|---|---|---|---|---|---|
| 50 | 2.18 | 0 | 1.09 | 0 | 3.27 |
| 51 | 0.87 | 0 | 5.66 | 1.74 | 8.27 |
| 52 | 0 | 0.22 | 0.87 | 1.31 | 2.4 |
| 53 | 7.41 | 7.41 | 13.51 | 7.41 | 35.74 |
| 54 | 286.51 | 260.25 | 235.85 | 236.5 | 1,019.11 |
| 56 | 27.56 | 37.91 | 34.86 | 29.2 | 129.53 |
| 57 | 60.57 | 62.75 | 56.65 | 49.24 | 229.21 |
| 58 | 0.44 | 0.44 | 0.87 | 2.4 | 4.15 |
| TOTAL | 385.54 | 368.98 | 349.36 | 327.8 | 1,431.68 |

STATE: VIRGIN ISLANDS

| ZIP CODE | QUARTER 1 | QUARTER 2 | QUARTER 3 | QUARTER 4 | TOTAL GRAMS |
|---|---|---|---|---|---|
| 8 | 41.4 | 79.42 | 77.35 | 39.22 | 237.39 |
| TOTAL | 41.4 | 79.42 | 77.35 | 39.22 | 237.39 |

STATE: VIRGINIA

| ZIP CODE | QUARTER 1 | QUARTER 2 | QUARTER 3 | QUARTER 4 | TOTAL GRAMS |
|---|---|---|---|---|---|
| 201 | 314.39 | 338.58 | 405.79 | 364.18 | 1,422.94 |
| 220 | 243.04 | 260.25 | 259.38 | 205.35 | 968.02 |
| 221 | 168.64 | 127.89 | 118.85 | 137.91 | 553.29 |
| 222 | 45.1 | 52.73 | 27.02 | 70.81 | 195.66 |
| 223 | 229.64 | 216.35 | 140.53 | 221.58 | 808.1 |
| 224 | 366.68 | 273.65 | 285.42 | 239.44 | 1,165.19 |
| 225 | 196.31 | 178.66 | 138.35 | 106.21 | 619.53 |
| 226 | 131.2 | 202.23 | 192.17 | 172.77 | 698.37 |
| 227 | 22.44 | 23.09 | 15.69 | 23.09 | 84.31 |
| 228 | 172.34 | 131.6 | 102.18 | 94.99 | 501.11 |

RETAIL DRUG DISTRIBUTION BY ZIP CODE WITHIN STATE BY GRAMS WT

DRUG CODE:9230DRUG NAME:MEPERIDINE (PETHIDINE)
STATE:VIRGINIA

| ZIP CODE | QUARTER 1 | QUARTER 2 | QUARTER 3 | QUARTER 4 | TOTAL GRAMS |
|---|---|---|---|---|---|
| 229 | 318.1 | 416.14 | 325.61 | 361.67 | 1,421.52 |
| 230 | 324.85 | 356.88 | 252.52 | 266.46 | 1,200.71 |
| 231 | 721.49 | 639.35 | 733.69 | 673.39 | 2,767.92 |
| 232 | 901.17 | 918.78 | 960.83 | 990.46 | 3,771.24 |
| 233 | 340.97 | 324.42 | 277.79 | 290.21 | 1,233.39 |
| 234 | 826.73 | 742.19 | 975.21 | 802.22 | 3,346.35 |
| 235 | 255.57 | 321.91 | 256.66 | 222.67 | 1,056.81 |
| 236 | 419.19 | 525.73 | 341.63 | 352.52 | 1,639.07 |
| 237 | 108.07 | 128 | 113.73 | 49.02 | 398.82 |
| 238 | 387.6 | 375.29 | 309.82 | 236.18 | 1,308.89 |
| 239 | 93.08 | 127.46 | 86.06 | 61.01 | 367.61 |
| 240 | 221.69 | 150.77 | 237.81 | 168.2 | 778.47 |
| 241 | 290.32 | 191.62 | 165.59 | 204.15 | 851.68 |
| 242 | 272.13 | 250.56 | 265.92 | 261.45 | 1,050.06 |
| 243 | 158.5 | 143.03 | 192.47 | 78.65 | 572.65 |
| 244 | 68.52 | 109.81 | 57.52 | 70.16 | 306.01 |
| 245 | 177.48 | 205.76 | 184.21 | 172.34 | 739.79 |
| 246 | 17.43 | 17.87 | 5.23 | 4.36 | 44.89 |
| TOTAL | 7,792.67 | 7,750.60 | 7,427.68 | 6,901.45 | 29,872.40 |

STATE: WASHINGTON

| ZIP CODE | QUARTER 1 | QUARTER 2 | QUARTER 3 | QUARTER 4 | TOTAL GRAMS |
|---|---|---|---|---|---|
| 980 | 775.64 | 808.86 | 611.47 | 591.64 | 2,787.61 |
| 981 | 474.75 | 687.18 | 533.47 | 479.11 | 2,174.51 |
| 982 | 617.46 | 637.61 | 570.72 | 497.95 | 2,323.74 |
| 983 | 540.44 | 487.93 | 462.88 | 502.86 | 1,994.11 |
| 984 | 198.59 | 197.18 | 187.81 | 160.14 | 743.72 |
| 985 | 319.4 | 304.15 | 228.77 | 324.63 | 1,176.95 |
| 986 | 320.93 | 264.06 | 250.77 | 183.89 | 1,019.65 |
| 988 | 308.08 | 347.29 | 289.77 | 271.47 | 1,216.61 |
| 989 | 147.61 | 132.25 | 156.22 | 153.71 | 589.79 |
| 990 | 75.17 | 5.45 | 15.25 | 36.6 | 132.47 |
| 991 | 169.62 | 173.32 | 157.2 | 192.17 | 692.31 |
| 992 | 281.49 | 346.75 | 199.68 | 281.39 | 1,109.31 |
| 993 | 224.19 | 226.52 | 179.96 | 212.84 | 843.51 |
| 994 | 52.29 | 20.7 | 28.32 | 23.53 | 124.84 |
| TOTAL | 4,505.66 | 4,639.25 | 3,872.29 | 3,911.93 | 16,929.13 |

STATE: WEST VIRGINIA

| ZIP CODE | QUARTER 1 | QUARTER 2 | QUARTER 3 | QUARTER 4 | TOTAL GRAMS |
|---|---|---|---|---|---|
| 247 | 59.7 | 66.45 | 43.79 | 32.25 | 202.19 |
| 248 | 18.74 | 20.7 | 8.72 | 32.68 | 80.84 |
| 249 | 17.43 | 45.1 | 34.86 | 58.83 | 156.22 |
| 250 | 9.8 | 17.43 | 0 | 4.36 | 31.59 |
| 251 | 2.61 | 2.61 | 5.23 | 1.31 | 11.76 |
| 252 | 7.63 | 10.02 | 1.74 | 1.31 | 20.7 |
| 253 | 78.04 | 87.59 | 76.69 | 84.54 | 326.86 |
| 254 | 73.42 | 97.17 | 87.15 | 70.81 | 328.55 |
| 255 | 37.04 | 45.32 | 25.71 | 22.88 | 130.95 |
| 256 | 21.79 | 20.7 | 11.98 | 29.63 | 84.1 |
| 257 | 50.55 | 68.19 | 63.29 | 81.27 | 263.3 |
| 258 | 27.23 | 20.7 | 18.74 | 10.89 | 77.56 |
| 259 | 13.07 | 4.36 | 8.72 | 16.89 | 43.04 |
| 260 | 94.27 | 96.74 | 72.27 | 78 | 341.28 |
| 261 | 94.78 | 133.23 | 128.22 | 141.73 | 497.96 |
| 262 | 3.05 | 2.4 | 2.4 | 0.22 | 8.07 |

| | | | | | |
|---|---|---|---|---|---|
| 263 | 65.8 | 53.6 | 60.79 | 105.45 | 285.64 |
| 264 | 22.77 | 13.07 | 3.81 | 4.25 | 43.9 |
| 265 | 56.91 | 54.47 | 54.14 | 33.99 | 199.51 |
| 266 | 17.65 | 1.35 | 2.61 | 5.66 | 27.27 |
| 267 | 3.27 | 8.93 | 8.28 | 9.8 | 30.28 |
| 268 | 7.71 | 6.97 | 13.73 | 9.37 | 37.78 |
| TOTAL | 783.26 | 877.1 | 732.87 | 836.12 | 3,229.35 |

STATE: WISCONSIN

| ZIP CODE | QUARTER 1 | QUARTER 2 | QUARTER 3 | QUARTER 4 | TOTAL GRAMS |
|---|---|---|---|---|---|
| 530 | 300.67 | 362.76 | 397.4 | 272.02 | 1,332.85 |
| 531 | 331.93 | 419.08 | 310.8 | 315.05 | 1,376.86 |
| 532 | 231.71 | 185.08 | 156.65 | 148.16 | 721.6 |

DRUG CODE:9230DRUG NAME:MEPERIDINE (PETHIDINE)
STATE:WISCONSIN

| ZIP CODE | QUARTER 1 | QUARTER 2 | QUARTER 3 | QUARTER 4 | TOTAL GRAMS |
|---|---|---|---|---|---|
| 534 | 21.57 | 76.26 | 70.81 | 45.75 | 214.39 |
| 535 | 122.23 | 85.84 | 85.84 | 79.31 | 373.22 |
| 537 | 119.61 | 104.8 | 101.75 | 108.07 | 434.23 |
| 538 | 4.36 | 1.31 | 3.27 | 5.01 | 13.95 |
| 539 | 2.51 | 39.54 | 39.22 | 6.43 | 87.7 |
| 540 | 6.32 | 10.46 | 16.56 | 13.94 | 47.28 |
| 541 | 1.74 | 33.33 | 20.26 | 41.61 | 96.94 |
| 542 | 79.31 | 71.35 | 61.22 | 83.45 | 295.33 |
| 543 | 179.31 | 164.06 | 188.03 | 215.04 | 746.44 |
| 544 | 130.4 | 101.75 | 106.65 | 137.37 | 476.17 |
| 545 | 3.92 | 12.2 | 8.28 | 5.23 | 29.63 |
| 546 | 66.89 | 45.1 | 38.56 | 38.35 | 188.9 |
| 547 | 49.78 | 33.99 | 29.63 | 61.77 | 175.17 |
| 548 | 20.26 | 27.93 | 22.01 | 35.3 | 105.5 |
| 549 | 266.57 | 268.86 | 265.81 | 152.51 | 953.75 |
| TOTAL | 1,939.09 | 2,043.70 | 1,922.75 | 1,764.37 | 7,669.91 |

STATE: WYOMING

| ZIP CODE | QUARTER 1 | QUARTER 2 | QUARTER 3 | QUARTER 4 | TOTAL GRAMS |
|---|---|---|---|---|---|
| 820 | 69.81 | 70.03 | 102.23 | 51.64 | 293.71 |
| 822 | 61.22 | 35.08 | 48.59 | 121.79 | 266.68 |
| 823 | 69.72 | 52.29 | 8.72 | 0 | 130.73 |
| 824 | 63.01 | 66.5 | 46.63 | 43.14 | 219.28 |
| 825 | 59.92 | 34.64 | 37.37 | 22.66 | 154.59 |
| 826 | 96.3 | 42.92 | 81.92 | 42.05 | 263.19 |
| 827 | 31.59 | 17.43 | 25.27 | 29.2 | 103.49 |
| 828 | 49.72 | 58.39 | 67.11 | 59.7 | 234.92 |
| 829 | 39.76 | 40.31 | 5.23 | 66.89 | 152.19 |
| 830 | 2.18 | 2.18 | 1.09 | 1.09 | 6.54 |
| 831 | 21.79 | 54.69 | 55.34 | 43.79 | 175.61 |
| TOTAL | 565.02 | 474.46 | 479.5 | 481.95 | 2,000.93 |

DRUG CODE:9250BDRUG NAME:METHADONE
STATE: ALABAMA

| ZIP CODE | QUARTER 1 | QUARTER 2 | QUARTER 3 | QUARTER 4 | TOTAL GRAMS |
|---|---|---|---|---|---|
| 350 | 13,369.49 | 11,983.62 | 11,738.28 | 12,061.43 | 49,152.82 |
| 351 | 1,771.90 | 1,725.03 | 1,602.76 | 1,509.21 | 6,608.90 |
| 352 | 13,759.77 | 14,399.62 | 11,879.32 | 15,725.68 | 55,764.39 |
| 354 | 4,739.34 | 5,135.11 | 4,953.74 | 3,807.37 | 18,635.56 |
| 355 | 6,615.25 | 6,909.69 | 6,235.76 | 5,529.18 | 25,289.88 |
| 356 | 3,615.43 | 2,859.04 | 2,618.80 | 1,070.42 | 10,163.69 |
| 357 | 1,014.70 | 971.77 | 939.57 | 1,285.43 | 4,211.47 |

| | | | | | |
|---|---|---|---|---|---|
| 358 | 12,732.68 | 14,333.48 | 11,904.73 | 13,935.20 | 52,906.09 |
| 359 | 4,488.81 | 4,631.65 | 2,344.67 | 1,493.20 | 12,958.33 |
| 360 | 825.71 | 834.03 | 767.04 | 745.84 | 3,172.62 |
| 361 | 1,735.49 | 2,240.92 | 2,190.39 | 1,336.14 | 7,502.94 |
| 362 | 3,625.90 | 3,656.31 | 3,328.06 | 3,010.10 | 13,620.37 |
| 363 | 2,375.08 | 2,396.54 | 2,361.22 | 1,873.14 | 9,005.98 |
| 364 | 512.04 | 538.43 | 480.29 | 466.43 | 1,997.19 |
| 365 | 4,404.38 | 4,090.36 | 4,228.28 | 3,924.63 | 16,647.65 |
| 366 | 12,782.76 | 9,322.96 | 12,365.80 | 7,090.45 | 41,561.97 |
| 367 | 666.33 | 684.22 | 655.6 | 579.12 | 2,585.27 |
| 368 | 1,082.94 | 1,100.29 | 970.96 | 972.39 | 4,126.58 |
| 369 | 32.65 | 26.83 | 33.09 | 21.47 | 114.04 |
| TOTAL | 90,150.65 | 87,839.90 | 81,598.36 | 76,436.83 | 336,025.74 |

STATE: ALASKA

| ZIP CODE | QUARTER 1 | QUARTER 2 | QUARTER 3 | QUARTER 4 | TOTAL GRAMS |
|---|---|---|---|---|---|
| 995 | 5,610.93 | 5,756.63 | 4,486.85 | 4,625.84 | 20,480.25 |
| 996 | 1,812.77 | 1,713.31 | 1,893.53 | 1,980.20 | 7,399.81 |
| 997 | 850.57 | 802.63 | 805.68 | 795.39 | 3,254.27 |
| 998 | 243.28 | 220.47 | 209.74 | 178.43 | 851.92 |
| 999 | 63.5 | 54.11 | 55.01 | 46.96 | 219.58 |
| TOTAL | 8,581.05 | 8,547.15 | 7,450.81 | 7,626.82 | 32,205.83 |

STATE: ARIZONA

| ZIP CODE | QUARTER 1 | QUARTER 2 | QUARTER 3 | QUARTER 4 | TOTAL GRAMS |
|---|---|---|---|---|---|
| 850 | 26,993.35 | 27,858.95 | 28,264.47 | 20,821.72 | 103,938.49 |
| 851 | 741.1 | 719.1 | 646.47 | 639.67 | 2,746.34 |

DRUG CODE:9250BDRUG NAME:METHADONE
STATE:ARIZONA

| ZIP CODE | QUARTER 1 | QUARTER 2 | QUARTER 3 | QUARTER 4 | TOTAL GRAMS |
|---|---|---|---|---|---|
| 852 | 18,004.99 | 17,930.66 | 16,744.42 | 15,343.70 | 68,023.77 |
| 853 | 7,704.67 | 8,089.52 | 5,758.43 | 7,121.16 | 28,673.78 |
| 855 | 845.21 | 715.07 | 708.36 | 675.72 | 2,944.36 |
| 856 | 758 | 648.53 | 627.6 | 552.11 | 2,586.24 |
| 857 | 26,830.48 | 25,482.80 | 28,289.34 | 23,180.43 | 103,783.05 |
| 859 | 503.19 | 435.57 | 409.37 | 412.59 | 1,760.72 |
| 860 | 888.59 | 899.95 | 1,103.51 | 1,072.92 | 3,964.97 |
| 863 | 1,947.56 | 2,289.13 | 1,873.32 | 1,734.24 | 7,844.25 |
| 864 | 1,750.79 | 2,554.85 | 1,596.95 | 1,569.67 | 7,472.26 |
| 865 | 15.2 | 8.5 | 4.47 | 8.5 | 36.67 |
| TOTAL | 86,983.13 | 87,632.63 | 86,026.71 | 73,132.43 | 333,774.90 |

STATE: ARKANSAS

| ZIP CODE | QUARTER 1 | QUARTER 2 | QUARTER 3 | QUARTER 4 | TOTAL GRAMS |
|---|---|---|---|---|---|
| 716 | 308.84 | 329.59 | 324.04 | 291.66 | 1,254.13 |
| 717 | 583.6 | 562.4 | 490.31 | 485.03 | 2,121.34 |
| 718 | 1,953.46 | 2,187.97 | 2,123.75 | 1,709.02 | 7,974.20 |
| 719 | 1,021.14 | 891.45 | 828.93 | 744.59 | 3,486.11 |
| 720 | 1,064.96 | 1,010.49 | 911.39 | 861.75 | 3,848.59 |
| 721 | 2,629.36 | 3,365.45 | 3,045.88 | 2,404.59 | 11,445.28 |
| 722 | 1,404.66 | 1,693.65 | 1,123.19 | 1,090.81 | 5,312.31 |
| 723 | 276.82 | 311.25 | 244.17 | 253.12 | 1,085.36 |
| 724 | 3,079.87 | 2,757.44 | 2,643.85 | 2,603.15 | 11,084.31 |
| 725 | 355.52 | 323.77 | 323.86 | 322.43 | 1,325.58 |
| 726 | 720.35 | 648.71 | 615.35 | 602.38 | 2,586.79 |
| 727 | 5,033.06 | 4,660.18 | 4,878.24 | 4,023.82 | 18,595.30 |
| 728 | 336.29 | 324.22 | 298.73 | 263.85 | 1,223.09 |
| 729 | 1,744.97 | 1,587.56 | 1,481.13 | 1,430.24 | 6,243.90 |

RETAIL DRUG DISTRIBUTION BY ZIP CODE WITHIN STATE BY GRAMS WT

| | | | | | |
|---|---|---|---|---|---|
| TOTAL | 20,512.90 | 20,654.13 | 19,332.82 | 17,086.44 | 77,586.29 |

STATE: CALIFORNIA

| ZIP CODE | QUARTER 1 | QUARTER 2 | QUARTER 3 | QUARTER 4 | TOTAL GRAMS |
|---|---|---|---|---|---|
| 900 | 16,809.93 | 19,187.04 | 17,756.01 | 14,897.31 | 68,650.29 |
| 902 | 4,658.75 | 3,344.97 | 3,665.79 | 2,350.57 | 14,020.08 |
| 903 | 1,418.07 | 1,272.73 | 1,573.70 | 909.16 | 5,173.66 |
| 904 | 245.69 | 282.18 | 251.15 | 298.19 | 1,077.21 |
| 905 | 551.76 | 516.07 | 536.19 | 481.54 | 2,085.56 |
| 906 | 7,129.44 | 6,653.89 | 6,842.88 | 4,558.31 | 25,184.52 |
| 907 | 4,913.48 | 4,186.87 | 4,751.68 | 3,581.62 | 17,433.65 |
| 908 | 3,596.38 | 1,794.52 | 3,234.78 | 1,124.62 | 9,750.30 |
| 910 | 602.47 | 614.72 | 563.38 | 566.78 | 2,347.35 |
| 911 | 669.01 | 986.43 | 575.73 | 1,016.84 | 3,248.01 |
| 912 | 1,600.71 | 1,543.64 | 1,296.43 | 965.06 | 5,405.84 |
| 913 | 6,829.37 | 6,530.82 | 6,243.09 | 5,543.22 | 25,146.50 |
| 914 | 3,954.32 | 3,338.80 | 3,064.30 | 2,790.98 | 13,148.40 |
| 915 | 306.96 | 327.8 | 340.23 | 307.32 | 1,282.31 |
| 916 | 1,293.30 | 1,038.40 | 941.8 | 711.5 | 3,985 |
| 917 | 8,267.65 | 8,148.16 | 8,446.89 | 6,990.63 | 31,853.33 |
| 918 | 62.61 | 56.79 | 34.43 | 32.2 | 186.03 |
| 919 | 2,589.56 | 2,360.05 | 2,755.74 | 1,599.90 | 9,305.25 |
| 920 | 15,490.20 | 14,349.84 | 13,590.86 | 10,931.27 | 54,362.17 |
| 921 | 13,514.21 | 11,553.77 | 12,373.22 | 7,877.70 | 45,318.90 |
| 922 | 6,319.38 | 7,620.02 | 6,159.91 | 4,007.99 | 24,107.30 |
| 923 | 7,541.56 | 7,703.63 | 9,158.42 | 5,952.22 | 30,355.83 |
| 924 | 307.58 | 263.4 | 265.19 | 249.45 | 1,085.62 |
| 925 | 6,988.75 | 7,862.85 | 7,330.23 | 5,990.33 | 28,172.16 |
| 926 | 2,994.34 | 2,758.80 | 2,711.66 | 2,604.51 | 11,069.31 |
| 927 | 4,143.67 | 3,686.72 | 3,592 | 2,618.18 | 14,040.57 |
| 928 | 6,274.71 | 5,445.09 | 5,614.61 | 4,890.58 | 22,224.99 |
| 930 | 9,807.45 | 9,675.98 | 10,208.41 | 6,496.56 | 36,188.40 |
| 931 | 1,955.87 | 2,789.19 | 2,763.25 | 1,934.32 | 9,442.63 |
| 932 | 8,786.76 | 9,818.90 | 9,941.43 | 6,044.80 | 34,591.89 |
| 933 | 9,407.39 | 9,207.30 | 9,973.70 | 7,212.71 | 35,801.10 |
| 934 | 5,642.68 | 7,102.61 | 5,930.86 | 5,446.54 | 24,122.69 |
| 935 | 4,778.87 | 4,167.64 | 4,383.45 | 3,452.74 | 16,782.70 |
| 936 | 2,721.84 | 2,606.01 | 2,434.91 | 2,445.02 | 10,207.78 |
| 937 | 17,880.84 | 18,746.09 | 15,919.43 | 12,293.53 | 64,839.89 |
| 939 | 3,157.23 | 2,961.09 | 2,691.25 | 2,026.08 | 10,835.65 |

DRUG CODE:9250BDRUG NAME:METHADONE
STATE:CALIFORNIA

| ZIP CODE | QUARTER 1 | QUARTER 2 | QUARTER 3 | QUARTER 4 | TOTAL GRAMS |
|---|---|---|---|---|---|
| 940 | 5,095.31 | 5,232.15 | 4,781.02 | 3,722.94 | 18,831.42 |
| 941 | 23,119.08 | 24,260.86 | 23,176.11 | 19,114.03 | 89,670.08 |
| 943 | 677.24 | 646.41 | 592.75 | 623.68 | 2,540.08 |
| 944 | 423.23 | 416.25 | 395.68 | 376.18 | 1,611.34 |
| 945 | 27,100.14 | 25,698.53 | 25,084.97 | 20,662.43 | 98,546.07 |
| 946 | 10,269.23 | 10,375.93 | 10,503.39 | 7,362.43 | 38,510.98 |
| 947 | 1,845.15 | 1,517.62 | 1,496.42 | 1,108.43 | 5,967.62 |
| 948 | 3,059.74 | 2,898.93 | 2,935.78 | 2,858.59 | 11,753.04 |
| 949 | 3,929.17 | 3,885.99 | 3,582.52 | 2,628.12 | 14,025.80 |
| 950 | 8,252.63 | 7,171.48 | 7,878.51 | 5,781.77 | 29,084.39 |
| 951 | 3,878.74 | 3,792.52 | 2,934.97 | 2,946.60 | 13,552.83 |
| 952 | 16,089.63 | 16,681.54 | 16,561.34 | 11,507.53 | 60,840.04 |
| 953 | 18,675.43 | 18,421.60 | 17,269.70 | 13,643.45 | 68,010.18 |
| 954 | 8,190.83 | 7,906.50 | 7,366.01 | 7,781.46 | 31,244.80 |
| 955 | 1,734.87 | 1,671.10 | 1,520.39 | 1,473.61 | 6,399.97 |
| 956 | 15,628.39 | 15,699.85 | 15,471.42 | 12,411.23 | 59,210.89 |

| | | | | | |
|---|---|---|---|---|---|
| 957 | 780.36 | 747.72 | 822.4 | 789.67 | 3,140.15 |
| 958 | 19,310.66 | 19,660.17 | 19,530.45 | 15,171.64 | 73,672.92 |
| 959 | 12,627.14 | 11,850.35 | 11,664.68 | 8,777.82 | 44,919.99 |
| 960 | 4,083.11 | 3,783.13 | 3,749.06 | 3,383.78 | 14,999.08 |
| 961 | 459.9 | 444.25 | 421.26 | 393.54 | 1,718.95 |
| TOTAL | 378,442.77 | 373,265.69 | 365,655.82 | 283,719.21 | 1,401,083.49 |

STATE: COLORADO

| ZIP CODE | QUARTER 1 | QUARTER 2 | QUARTER 3 | QUARTER 4 | TOTAL GRAMS |
|---|---|---|---|---|---|
| 800 | 7,200.29 | 6,236.08 | 6,650.27 | 7,200.88 | 27,287.52 |
| 801 | 1,577.27 | 1,871.53 | 1,927.61 | 1,470.75 | 6,847.16 |
| 802 | 17,159.33 | 21,527.05 | 20,478.45 | 15,952.52 | 75,117.35 |
| 803 | 302.75 | 280.39 | 248.64 | 234.33 | 1,066.11 |
| 804 | 312.32 | 307.41 | 277.26 | 339.69 | 1,236.68 |
| 805 | 3,036.22 | 3,243.81 | 3,050.44 | 4,008.16 | 13,338.63 |
| 806 | 589.86 | 559.72 | 495.5 | 436.29 | 2,081.37 |
| 807 | 115.82 | 104.91 | 96.15 | 71.1 | 387.98 |
| 808 | 347.03 | 320.2 | 324.67 | 296.05 | 1,287.95 |
| 809 | 3,779.20 | 3,498.98 | 3,660.42 | 2,764.77 | 13,703.37 |
| 810 | 1,970.63 | 1,924.75 | 1,748.82 | 1,979.66 | 7,623.86 |
| 811 | 245.07 | 225.39 | 359.1 | 268.32 | 1,097.88 |
| 812 | 229.86 | 196.77 | 191.4 | 188.27 | 806.3 |
| 813 | 267.87 | 372.07 | 504.89 | 139.53 | 1,284.36 |
| 814 | 172.17 | 139.08 | 134.61 | 116.27 | 562.13 |
| 815 | 897.08 | 1,158.52 | 878.39 | 1,274.97 | 4,208.96 |
| 816 | 205.89 | 151.6 | 145.79 | 152.85 | 656.13 |
| TOTAL | 38,408.66 | 42,118.26 | 41,172.41 | 36,894.41 | 158,593.74 |

STATE: CONNECTICUT

| ZIP CODE | QUARTER 1 | QUARTER 2 | QUARTER 3 | QUARTER 4 | TOTAL GRAMS |
|---|---|---|---|---|---|
| 60 | 14,272.48 | 12,097.12 | 14,195.20 | 12,035.14 | 52,599.94 |
| 61 | 14,870.12 | 10,343.74 | 17,150.57 | 11,509.50 | 53,873.93 |
| 62 | 5,837.75 | 3,657.20 | 3,676.88 | 3,481.45 | 16,653.28 |
| 63 | 6,989.02 | 9,172.07 | 7,017.02 | 4,852.21 | 28,030.32 |
| 64 | 8,945.88 | 15,606.48 | 8,801.25 | 8,720.31 | 42,073.92 |
| 65 | 28,235.22 | 23,724.50 | 19,991.63 | 24,096.84 | 96,048.19 |
| 66 | 16,896.65 | 19,616.88 | 10,462.69 | 19,566.65 | 66,542.87 |
| 67 | 9,468.83 | 9,456.40 | 9,458.01 | 9,446.92 | 37,830.16 |
| 68 | 6,103.56 | 5,987.02 | 5,248.70 | 6,652.28 | 23,991.56 |
| 69 | 1,921.26 | 1,906.95 | 1,928.77 | 1,925.02 | 7,682 |
| TOTAL | 113,540.77 | 111,568.36 | 97,930.72 | 102,286.32 | 425,326.17 |

STATE: DELAWARE

| ZIP CODE | QUARTER 1 | QUARTER 2 | QUARTER 3 | QUARTER 4 | TOTAL GRAMS |
|---|---|---|---|---|---|
| 197 | 13,134.17 | 12,463.46 | 11,092.71 | 10,004.67 | 46,695.01 |
| 198 | 6,431.63 | 6,404.80 | 6,664.17 | 3,830.36 | 23,330.96 |
| 199 | 7,430.41 | 8,876.47 | 9,712.74 | 5,137.43 | 31,157.05 |
| TOTAL | 26,996.21 | 27,744.73 | 27,469.62 | 18,972.46 | 101,183.02 |

STATE: DISTRICT OF COLUMBIA

| ZIP CODE | QUARTER 1 | QUARTER 2 | QUARTER 3 | QUARTER 4 | TOTAL GRAMS |
|---|---|---|---|---|---|
| 200 | 10,126.47 | 10,842.77 | 10,205.77 | 6,500.82 | 37,675.83 |
| 204 | 721.33 | 678.85 | 719.1 | 651.12 | 2,770.40 |
| TOTAL | 10,847.80 | 11,521.62 | 10,924.87 | 7,151.94 | 40,446.23 |

DRUG CODE:9250BDRUG NAME:METHADONE

STATE: FLORIDA

| ZIP CODE | QUARTER 1 | QUARTER 2 | QUARTER 3 | QUARTER 4 | TOTAL GRAMS |
|---|---|---|---|---|---|
| 320 | 7,473.52 | 8,230.63 | 7,392.57 | 6,905.22 | 30,001.94 |

| | | | | | | |
|---|---|---|---|---|---|---|
| | 321 | 10,779.76 | 10,717.42 | 8,174.73 | 10,964.99 | 40,636.90 |
| | 322 | 19,733.24 | 19,396.05 | 22,574.79 | 17,429.29 | 79,133.37 |
| | 323 | 5,940.34 | 4,196.97 | 4,450.09 | 4,152.70 | 18,740.10 |
| | 324 | 5,663.70 | 6,316.34 | 4,487.12 | 899.86 | 17,367.02 |
| | 325 | 13,821.16 | 15,177.61 | 12,065.55 | 15,898.76 | 56,963.08 |
| | 326 | 5,248.97 | 5,086.54 | 3,831.07 | 2,412.82 | 16,579.40 |
| | 327 | 2,735.43 | 2,733.11 | 2,636.60 | 2,529.72 | 10,634.86 |
| | 328 | 12,459.17 | 10,614.56 | 12,896.17 | 9,513.20 | 45,483.10 |
| | 329 | 7,840.40 | 9,058.04 | 5,672.55 | 4,204.13 | 26,775.12 |
| | 330 | 7,183.19 | 6,016.47 | 7,996.94 | 4,838.82 | 26,035.42 |
| | 331 | 2,315.51 | 2,495.60 | 2,273.61 | 1,855.52 | 8,940.24 |
| | 333 | 3,099.45 | 3,070.03 | 2,486.70 | 2,933.18 | 11,589.36 |
| | 334 | 8,595.54 | 8,494.56 | 9,181.19 | 10,212.08 | 36,483.37 |
| | 335 | 3,144.17 | 2,996.96 | 2,847.05 | 2,716.47 | 11,704.65 |
| | 336 | 12,506.13 | 13,760.79 | 13,606.42 | 11,855.16 | 51,728.50 |
| | 337 | 17,467.18 | 20,403.28 | 20,093.10 | 18,589.57 | 76,553.13 |
| | 338 | 2,874.42 | 1,783.08 | 2,438.85 | 2,392.25 | 9,488.60 |
| | 339 | 17,635.51 | 14,842.84 | 16,872.86 | 15,542.26 | 64,893.47 |
| | 341 | 3,561.32 | 4,324.87 | 3,468.39 | 4,396.96 | 15,751.54 |
| | 342 | 16,199.82 | 16,061.19 | 16,335.95 | 16,707.39 | 65,304.35 |
| | 344 | 6,939.38 | 11,479 | 9,105.26 | 10,182.30 | 37,705.94 |
| | 346 | 14,034.66 | 16,246.33 | 16,173.70 | 13,883.77 | 60,338.46 |
| | 347 | 5,012.66 | 4,878.06 | 3,279.41 | 4,698.37 | 17,868.50 |
| | 349 | 2,706.10 | 2,254.78 | 2,325.89 | 2,460.94 | 9,747.71 |
| TOTAL | | 214,970.73 | 220,635.11 | 212,666.56 | 198,175.73 | 846,448.13 |
| STATE: | GEORGIA | | | | | |
| ZIP CODE | QUARTER 1 | | QUARTER 2 | QUARTER 3 | QUARTER 4 | TOTAL GRAMS |
| | 300 | 18,517.94 | 17,660.65 | 15,193.63 | 17,665.30 | 69,037.52 |
| | 301 | 13,752.03 | 14,547.95 | 14,480.60 | 12,639.03 | 55,419.61 |
| | 302 | 8,622.11 | 8,183.67 | 8,483.56 | 6,886.34 | 32,175.68 |
| | 303 | 5,503.96 | 5,075.41 | 4,204.31 | 4,806.49 | 19,590.17 |
| | 304 | 1,966.25 | 1,425.41 | 2,143.43 | 2,050.77 | 7,585.86 |
| | 305 | 13,135.29 | 12,824.22 | 13,622.74 | 12,882.09 | 52,464.34 |
| | 306 | 5,041.91 | 5,162.03 | 5,187.16 | 4,423.43 | 19,814.53 |
| | 307 | 19,130.77 | 23,669.94 | 26,822.43 | 25,028.26 | 94,651.40 |
| | 308 | 1,663.58 | 1,067.91 | 1,020.96 | 445.86 | 4,198.31 |
| | 309 | 3,274.09 | 2,397.67 | 3,037.70 | 3,058.12 | 11,767.58 |
| | 310 | 4,812.05 | 4,113.35 | 6,152.40 | 4,019.34 | 19,097.14 |
| | 312 | 2,151.48 | 2,485.90 | 2,074.65 | 2,130.46 | 8,842.49 |
| | 313 | 477.7 | 514.46 | 461.42 | 476.18 | 1,929.76 |
| | 314 | 4,171.48 | 4,204.48 | 4,318.25 | 3,753.17 | 16,447.38 |
| | 315 | 3,582.07 | 3,006.53 | 3,040.07 | 1,914.91 | 11,543.58 |
| | 316 | 2,023.40 | 2,074.02 | 1,506.17 | 454.53 | 6,058.12 |
| | 317 | 3,551.77 | 3,418.62 | 3,320.57 | 3,527.60 | 13,818.56 |
| | 318 | 123.43 | 134.61 | 130.49 | 122.62 | 511.15 |
| | 319 | 3,655.50 | 3,624.02 | 2,615.32 | 3,080.49 | 12,975.33 |
| | 398 | 1,229.80 | 971.77 | 1,013.36 | 1,021.23 | 4,236.16 |
| TOTAL | | 116,386.61 | 116,562.62 | 118,829.22 | 110,386.22 | 462,164.67 |
| STATE: | GUAM | | | | | |
| ZIP CODE | QUARTER 1 | | QUARTER 2 | QUARTER 3 | QUARTER 4 | TOTAL GRAMS |
| | 969 | 51.88 | 39.35 | 47.4 | 25.94 | 164.57 |
| TOTAL | | 51.88 | 39.35 | 47.4 | 25.94 | 164.57 |
| STATE: | HAWAII | | | | | |
| ZIP CODE | QUARTER 1 | | QUARTER 2 | QUARTER 3 | QUARTER 4 | TOTAL GRAMS |
| | 967 | 4,946.66 | 4,474.68 | 4,977.43 | 3,454.89 | 17,853.66 |
| | 968 | 4,544.98 | 3,497.28 | 4,254.75 | 3,454.44 | 15,751.45 |
| TOTAL | | 9,491.64 | 7,971.96 | 9,232.18 | 6,909.33 | 33,605.11 |

RETAIL DRUG DISTRIBUTION BY ZIP CODE WITHIN STATE BY GRAMS WT

STATE: IDAHO

| ZIP CODE | QUARTER 1 | QUARTER 2 | QUARTER 3 | QUARTER 4 | TOTAL GRAMS |
|---|---|---|---|---|---|
| 832 | 1,252.43 | 1,136.34 | 1,073.73 | 1,017.56 | 4,480.06 |
| 833 | 622.95 | 640.39 | 607.3 | 531.27 | 2,401.91 |
| 834 | 768.29 | 688.69 | 614.54 | 559.89 | 2,631.41 |
| 835 | 509.36 | 478.06 | 482.98 | 403.37 | 1,873.77 |
| 836 | 2,908.68 | 2,950.09 | 2,832.74 | 2,260.06 | 10,951.57 |
| 837 | 6,115.19 | 6,231.64 | 5,804.30 | 4,285.96 | 22,437.09 |

DRUG CODE:9250BDRUG NAME:METHADONE
STATE:IDAHO

| ZIP CODE | QUARTER 1 | QUARTER 2 | QUARTER 3 | QUARTER 4 | TOTAL GRAMS |
|---|---|---|---|---|---|
| 838 | 1,612.60 | 1,640.87 | 1,621.64 | 1,445.44 | 6,320.55 |
| TOTAL | 13,789.50 | 13,766.08 | 13,037.23 | 10,503.55 | 51,096.36 |

STATE: ILLINOIS

| ZIP CODE | QUARTER 1 | QUARTER 2 | QUARTER 3 | QUARTER 4 | TOTAL GRAMS |
|---|---|---|---|---|---|
| 600 | 9,965.05 | 8,222.31 | 8,510.84 | 6,697.71 | 33,395.91 |
| 601 | 8,246.73 | 7,848.27 | 8,391.98 | 6,733.04 | 31,220.02 |
| 602 | 4,511.53 | 4,709.28 | 4,766.35 | 3,192.65 | 17,179.81 |
| 603 | 48.74 | 63.95 | 47.4 | 49.19 | 209.28 |
| 604 | 7,733.97 | 7,076.05 | 6,431 | 6,396.66 | 27,637.68 |
| 605 | 6,791.63 | 6,575.45 | 6,697.09 | 5,096.11 | 25,160.28 |
| 606 | 54,919.83 | 56,807.99 | 54,538.28 | 44,380.13 | 210,646.23 |
| 607 | 1,189.28 | 718.56 | 1,720.11 | 551.13 | 4,179.08 |
| 608 | 3,936.70 | 3,867.39 | 4,011.38 | 3,622.77 | 15,438.24 |
| 609 | 955.85 | 681.35 | 829.11 | 698.35 | 3,164.66 |
| 610 | 1,206.55 | 939.57 | 1,306.27 | 736.54 | 4,188.93 |
| 611 | 2,252.73 | 3,013.32 | 2,156.49 | 3,839.84 | 11,262.38 |
| 612 | 1,610.46 | 1,319.69 | 1,246.79 | 1,148.86 | 5,325.80 |
| 613 | 364.29 | 630.82 | 327.35 | 583.15 | 1,905.61 |
| 614 | 208.84 | 214.21 | 233.44 | 223.15 | 879.64 |
| 615 | 420.64 | 357.76 | 340.77 | 319.75 | 1,438.92 |
| 616 | 8,618.25 | 9,180.12 | 8,386.45 | 7,692.70 | 33,877.52 |
| 617 | 1,302.96 | 1,812.95 | 1,580.85 | 1,848.10 | 6,544.86 |
| 618 | 1,983.87 | 2,293.87 | 2,344.40 | 2,535.18 | 9,157.32 |
| 619 | 84.07 | 82.73 | 81.39 | 86.31 | 334.5 |
| 620 | 2,360.59 | 1,761.34 | 2,336.98 | 1,765.64 | 8,224.55 |
| 622 | 1,715.64 | 1,676.73 | 1,655.80 | 1,101.72 | 6,149.89 |
| 623 | 485.03 | 453.19 | 432.53 | 439.15 | 1,809.90 |
| 624 | 226.28 | 220.02 | 208.4 | 185.14 | 839.84 |
| 625 | 1,674.59 | 2,065.17 | 1,668.95 | 1,500.45 | 6,909.16 |
| 626 | 217.79 | 237.02 | 200.79 | 220.92 | 876.52 |
| 627 | 1,383.19 | 1,149.30 | 1,312.98 | 913.18 | 4,758.65 |
| 628 | 278.25 | 267.87 | 249.54 | 192.74 | 988.4 |
| 629 | 845.21 | 717.58 | 749.6 | 686.63 | 2,999.02 |
| TOTAL | 125,538.54 | 124,963.86 | 122,763.31 | 103,436.89 | 476,702.60 |

STATE: INDIANA

| ZIP CODE | QUARTER 1 | QUARTER 2 | QUARTER 3 | QUARTER 4 | TOTAL GRAMS |
|---|---|---|---|---|---|
| 460 | 891.09 | 804.87 | 828.39 | 711.85 | 3,236.20 |
| 461 | 1,840.05 | 1,742.56 | 1,639.17 | 1,593.11 | 6,814.89 |
| 462 | 18,551.67 | 18,575.99 | 18,727.62 | 14,568 | 70,423.28 |
| 463 | 2,122.05 | 2,176.34 | 2,094.35 | 2,316.85 | 8,709.59 |
| 464 | 5,751.89 | 4,937.98 | 6,098.91 | 5,112.39 | 21,901.17 |
| 465 | 3,145.25 | 3,083.89 | 2,653.06 | 2,437.42 | 11,319.62 |
| 466 | 3,315.09 | 3,351.76 | 2,872.54 | 2,368.37 | 11,907.76 |
| 467 | 1,732.10 | 1,371.38 | 1,378.99 | 1,229.71 | 5,712.18 |
| 468 | 6,562.21 | 6,147.57 | 6,093.46 | 6,706.66 | 25,509.90 |
| 469 | 8,748.75 | 8,396.98 | 8,007.47 | 8,105.86 | 33,259.06 |

| | | | | | |
|---|---|---|---|---|---|
| 470 | 8,188.68 | 8,111.31 | 8,215.96 | 4,892.82 | 29,408.77 |
| 471 | 15,679.01 | 16,123.53 | 15,384.31 | 9,350.50 | 56,537.35 |
| 472 | 855.05 | 683.77 | 676.61 | 637.26 | 2,852.69 |
| 473 | 7,227.74 | 8,130.01 | 7,058.43 | 5,092.98 | 27,509.16 |
| 474 | 1,186.87 | 1,143.49 | 1,099.22 | 963.27 | 4,392.85 |
| 475 | 337.64 | 351.95 | 349.26 | 323.33 | 1,362.18 |
| 476 | 340.32 | 332.63 | 294.26 | 316.62 | 1,283.83 |
| 477 | 4,004.68 | 4,583.80 | 4,173.45 | 2,375.79 | 15,137.72 |
| 478 | 428.78 | 422.51 | 376.99 | 367.15 | 1,595.43 |
| 479 | 1,242.14 | 1,304.75 | 1,220.14 | 1,206.81 | 4,973.84 |
| TOTAL | 92,151.06 | 91,777.07 | 89,242.59 | 70,676.75 | 343,847.47 |

STATE: IOWA

| ZIP CODE | QUARTER 1 | QUARTER 2 | QUARTER 3 | QUARTER 4 | TOTAL GRAMS |
|---|---|---|---|---|---|
| 500 | 284.96 | 278.61 | 263.4 | 266.8 | 1,093.77 |
| 501 | 254.01 | 239.97 | 286.21 | 235.05 | 1,015.24 |
| 502 | 300.97 | 308.12 | 300.07 | 258.03 | 1,167.19 |
| 503 | 4,795.68 | 5,747.77 | 4,480.05 | 4,744.52 | 19,768.02 |
| 504 | 199.45 | 160.1 | 160.54 | 144 | 664.09 |
| 505 | 222.71 | 214.66 | 199.45 | 228.52 | 865.34 |
| 506 | 459.72 | 800.04 | 457.49 | 466.88 | 2,184.13 |

DRUG CODE:9250BDRUG NAME:METHADONE
STATE:IOWA

| ZIP CODE | QUARTER 1 | QUARTER 2 | QUARTER 3 | QUARTER 4 | TOTAL GRAMS |
|---|---|---|---|---|---|
| 507 | 112.25 | 120.03 | 99.73 | 95.7 | 427.71 |
| 508 | 122.09 | 127.9 | 109.56 | 94.81 | 454.36 |
| 510 | 213.76 | 209.74 | 209.74 | 187.82 | 821.06 |
| 511 | 427.97 | 483.42 | 854.15 | 407.49 | 2,173.03 |
| 512 | 33.54 | 24.6 | 28.17 | 19.23 | 105.54 |
| 513 | 37.12 | 39.8 | 35.33 | 42.93 | 155.18 |
| 514 | 89.44 | 77.37 | 72.45 | 63.95 | 303.21 |
| 515 | 448.54 | 459.27 | 403.82 | 406.5 | 1,718.13 |
| 516 | 48.3 | 39.35 | 46.06 | 46.96 | 180.67 |
| 520 | 166.9 | 158.67 | 146.86 | 135.77 | 608.2 |
| 521 | 23.7 | 23.16 | 29.07 | 27.28 | 103.21 |
| 522 | 314.02 | 272.88 | 262.77 | 267.69 | 1,117.36 |
| 523 | 1,156.10 | 1,165.22 | 1,151.99 | 807.02 | 4,280.33 |
| 524 | 1,666.89 | 1,619.22 | 1,235.61 | 829.73 | 5,351.45 |
| 525 | 253.56 | 226.55 | 208.84 | 203.48 | 892.43 |
| 526 | 177.47 | 180.94 | 204.73 | 189.52 | 752.66 |
| 527 | 214.75 | 200.79 | 168.15 | 173.51 | 757.2 |
| 528 | 1,826.99 | 1,827.26 | 1,831.11 | 1,007 | 6,492.36 |
| TOTAL | 13,850.89 | 15,005.44 | 13,245.35 | 11,350.19 | 53,451.87 |

STATE: KANSAS

| ZIP CODE | QUARTER 1 | QUARTER 2 | QUARTER 3 | QUARTER 4 | TOTAL GRAMS |
|---|---|---|---|---|---|
| 660 | 2,237.34 | 2,551.54 | 1,714.65 | 2,368.73 | 8,872.26 |
| 661 | 3,454.62 | 3,908.35 | 3,765.42 | 3,691.19 | 14,819.58 |
| 662 | 2,137.53 | 2,326.24 | 2,095.22 | 1,466.10 | 8,025.09 |
| 664 | 97.94 | 125.66 | 109.12 | 121.64 | 454.36 |
| 665 | 222.08 | 220.74 | 171.99 | 151.15 | 765.96 |
| 666 | 2,171.42 | 2,086.81 | 2,082.97 | 1,894.70 | 8,235.90 |
| 667 | 469.11 | 432.8 | 431.55 | 406.95 | 1,740.41 |
| 668 | 171.72 | 161.89 | 151.15 | 145.34 | 630.1 |
| 669 | 88.55 | 91.68 | 103.75 | 110.91 | 394.89 |
| 670 | 739.04 | 782.15 | 776.43 | 734.3 | 3,031.92 |
| 671 | 320.91 | 281.65 | 293.63 | 275.92 | 1,172.11 |
| 672 | 11,308.97 | 10,974.02 | 10,673.32 | 9,773.20 | 42,729.51 |
| 673 | 186.48 | 174.41 | 164.57 | 162.33 | 687.79 |

| | | | | | |
|---|---|---|---|---|---|
| 674 | 388.62 | 408.29 | 375.56 | 371.8 | 1,544.27 |
| 675 | 312.15 | 309.91 | 306.33 | 282.63 | 1,211.02 |
| 676 | 82.73 | 72.89 | 75.58 | 102.41 | 333.61 |
| 677 | 60.82 | 54.56 | 42.04 | 47.4 | 204.82 |
| 678 | 210.63 | 218.68 | 199 | 195.87 | 824.18 |
| 679 | 38.91 | 33.09 | 38.46 | 33.09 | 143.55 |
| TOTAL | 24,699.57 | 25,215.36 | 23,570.74 | 22,335.66 | 95,821.33 |

STATE: KENTUCKY

| ZIP CODE | QUARTER 1 | QUARTER 2 | QUARTER 3 | QUARTER 4 | TOTAL GRAMS |
|---|---|---|---|---|---|
| 400 | 524.57 | 461.51 | 471.8 | 456.23 | 1,914.11 |
| 401 | 245.51 | 225.39 | 215.55 | 196.32 | 882.77 |
| 402 | 10,273.70 | 8,304.86 | 10,270.40 | 10,510.99 | 39,359.95 |
| 403 | 1,830.03 | 2,374.45 | 2,435.99 | 2,026.08 | 8,666.55 |
| 404 | 925.7 | 884.02 | 721.33 | 733.5 | 3,264.55 |
| 405 | 5,431.14 | 6,004.88 | 5,808.61 | 4,501.86 | 21,746.49 |
| 406 | 1,387.66 | 1,942.82 | 1,921.08 | 1,363.07 | 6,614.63 |
| 407 | 1,960.08 | 1,543.29 | 1,881.82 | 1,921.62 | 7,306.81 |
| 408 | 80.94 | 113.14 | 91.23 | 87.65 | 372.96 |
| 409 | 212.42 | 218.68 | 190.95 | 179.77 | 801.82 |
| 410 | 10,027.12 | 10,653.65 | 10,844.15 | 10,495.78 | 42,020.70 |
| 411 | 3,717.13 | 3,734.57 | 3,733.94 | 3,719.81 | 14,905.45 |
| 412 | 922.84 | 1,302.25 | 914.08 | 1,255.74 | 4,394.91 |
| 413 | 253.56 | 216.44 | 230.31 | 207.95 | 908.26 |
| 414 | 96.6 | 112.69 | 123.43 | 139.08 | 471.8 |
| 415 | 904.6 | 1,633.62 | 919.53 | 1,279.44 | 4,737.19 |
| 416 | 249.09 | 210.63 | 201.24 | 193.01 | 853.97 |
| 417 | 1,981.54 | 1,959.18 | 1,958.29 | 1,894.79 | 7,793.80 |
| 418 | 51.43 | 68.87 | 55.45 | 50.09 | 225.84 |
| 420 | 2,061.59 | 2,105.86 | 1,839.42 | 1,817.60 | 7,824.47 |
| 421 | 2,608.96 | 2,546.36 | 2,706.90 | 2,432.77 | 10,294.99 |
| 422 | 2,733.29 | 1,493.65 | 1,810.27 | 1,761.52 | 7,798.73 |
| 423 | 589.41 | 820.16 | 1,228.01 | 1,205.65 | 3,843.23 |

DRUG CODE:9250BDRUG NAME:METHADONE
STATE:KENTUCKY

| ZIP CODE | QUARTER 1 | QUARTER 2 | QUARTER 3 | QUARTER 4 | TOTAL GRAMS |
|---|---|---|---|---|---|
| 424 | 495.32 | 511.15 | 448.54 | 440.94 | 1,895.95 |
| 425 | 200.79 | 201.24 | 185.14 | 180.67 | 767.84 |
| 426 | 158.31 | 149.36 | 135.95 | 101.96 | 545.58 |
| 427 | 1,761.97 | 1,759.28 | 1,739.61 | 1,739.61 | 7,000.47 |
| TOTAL | 51,685.30 | 51,552 | 53,083.02 | 50,893.50 | 207,213.82 |

STATE: LOUISIANA

| ZIP CODE | QUARTER 1 | QUARTER 2 | QUARTER 3 | QUARTER 4 | TOTAL GRAMS |
|---|---|---|---|---|---|
| 700 | 5,235.73 | 6,897.26 | 6,976.32 | 6,693.51 | 25,802.82 |
| 701 | 3,978.11 | 5,121.24 | 4,098.95 | 4,984.40 | 18,182.70 |
| 703 | 266.53 | 267.43 | 240.59 | 218.14 | 992.69 |
| 704 | 3,117.25 | 4,184.45 | 4,219.87 | 4,137.94 | 15,659.51 |
| 705 | 5,179.47 | 5,159.53 | 4,161.11 | 5,895.88 | 20,395.99 |
| 706 | 1,626.02 | 1,993.62 | 1,479.34 | 1,908.02 | 7,007 |
| 707 | 741.9 | 670.35 | 636.81 | 565.71 | 2,614.77 |
| 708 | 4,714.83 | 4,239.46 | 4,202.43 | 2,520.06 | 15,676.78 |
| 710 | 613.11 | 545.14 | 545.58 | 593.79 | 2,297.62 |
| 711 | 5,199.68 | 5,080.55 | 4,964.90 | 4,796.13 | 20,041.26 |
| 712 | 1,106.19 | 1,756.51 | 474.21 | 1,051.37 | 4,388.28 |
| 713 | 4,839.60 | 4,072.20 | 5,133.86 | 3,980.08 | 18,025.74 |
| 714 | 556.32 | 520.54 | 558.55 | 518.75 | 2,154.16 |
| TOTAL | 37,174.74 | 40,508.28 | 37,692.52 | 37,863.78 | 153,239.32 |

STATE: MAINE

RETAIL DRUG DISTRIBUTION BY ZIP CODE WITHIN STATE BY GRAMS WT

| ZIP CODE | QUARTER 1 | QUARTER 2 | QUARTER 3 | QUARTER 4 | TOTAL GRAMS |
|---|---|---|---|---|---|
| 39 | 145.79 | 133.27 | 124.77 | 84.97 | 488.8 |
| 40 | 6,450.68 | 6,315.90 | 5,820.58 | 4,412.70 | 22,999.86 |
| 41 | 6,203.02 | 6,207.49 | 5,239.48 | 4,734.86 | 22,384.85 |
| 42 | 2,372.40 | 2,358.44 | 2,837.57 | 1,617.79 | 9,186.20 |
| 43 | 1,933.69 | 1,982.62 | 1,678.97 | 1,282.84 | 6,878.12 |
| 44 | 11,272.75 | 10,343.91 | 10,070.14 | 7,100.82 | 38,787.62 |
| 45 | 273.69 | 243.72 | 247.3 | 194.53 | 959.24 |
| 46 | 1,615.29 | 1,667.16 | 1,313.87 | 1,206.10 | 5,802.42 |
| 47 | 261.16 | 205.26 | 149.81 | 131.92 | 748.15 |
| 48 | 1,754.01 | 1,390.43 | 1,543.38 | 1,278.54 | 5,966.36 |
| 49 | 6,567.58 | 8,056.49 | 8,055.59 | 4,534.88 | 27,214.54 |
| TOTAL | 38,850.06 | 38,904.69 | 37,081.46 | 26,579.95 | 141,416.16 |

STATE: MARYLAND

| ZIP CODE | QUARTER 1 | QUARTER 2 | QUARTER 3 | QUARTER 4 | TOTAL GRAMS |
|---|---|---|---|---|---|
| 206 | 4,328.72 | 4,358.41 | 4,325.14 | 4,597.48 | 17,609.75 |
| 207 | 3,962.73 | 3,860.41 | 3,562.75 | 3,827.05 | 15,212.94 |
| 208 | 3,340.76 | 3,825.89 | 3,501.39 | 2,780.59 | 13,448.63 |
| 209 | 1,067.65 | 905.67 | 761.58 | 887.69 | 3,622.59 |
| 210 | 37,478.22 | 40,829.45 | 36,851.61 | 26,099.22 | 141,258.50 |
| 211 | 6,580.28 | 6,645.93 | 5,467.02 | 6,356.86 | 25,050.09 |
| 212 | 98,029.50 | 93,579.17 | 97,950.64 | 81,459.19 | 371,018.50 |
| 214 | 1,237.94 | 1,251.09 | 1,800.87 | 1,030.97 | 5,320.87 |
| 215 | 4,252.42 | 3,721.15 | 3,183.44 | 3,104.10 | 14,261.11 |
| 216 | 2,963.95 | 4,064.51 | 4,829.94 | 2,810.83 | 14,669.23 |
| 217 | 12,831.51 | 15,265.80 | 9,001.96 | 12,598.61 | 49,697.88 |
| 218 | 3,045.88 | 2,592.78 | 2,600.02 | 2,030.20 | 10,268.88 |
| 219 | 8,531.68 | 7,635.40 | 7,599.54 | 6,664.17 | 30,430.79 |
| TOTAL | 187,651.24 | 188,535.66 | 181,435.90 | 154,246.96 | 711,869.76 |

STATE: MASSACHUSETTS

| ZIP CODE | QUARTER 1 | QUARTER 2 | QUARTER 3 | QUARTER 4 | TOTAL GRAMS |
|---|---|---|---|---|---|
| 10 | 11,354.68 | 11,459.68 | 11,792.84 | 9,135.76 | 43,742.96 |
| 11 | 6,200.16 | 7,047.60 | 6,961.56 | 4,632.54 | 24,841.86 |
| 12 | 4,657.14 | 4,853.46 | 4,949.25 | 3,694.68 | 18,154.53 |
| 13 | 1,253.95 | 1,264.14 | 1,378.72 | 1,037.50 | 4,934.31 |
| 14 | 5,495.19 | 5,991.59 | 6,527.33 | 4,792.64 | 22,806.75 |
| 15 | 3,017.53 | 3,175.84 | 3,551.48 | 2,260.77 | 12,005.62 |
| 16 | 10,669.19 | 9,490.19 | 10,611.62 | 7,281.30 | 38,052.30 |
| 17 | 3,946.72 | 3,853.61 | 3,822.13 | 3,375.82 | 14,998.28 |
| 18 | 18,855.65 | 17,398.58 | 15,968.08 | 12,968.71 | 65,191.02 |
| 19 | 14,759.30 | 16,476.64 | 14,720.12 | 11,558.69 | 57,514.75 |
| 20 | 710.6 | 680.19 | 666.06 | 646.29 | 2,703.14 |
| 21 | 24,372.71 | 26,778.78 | 24,790.43 | 17,876.72 | 93,818.64 |
| 22 | 278.61 | 353.47 | 299.62 | 294.44 | 1,226.14 |

DRUG CODE:9250BDRUG NAME:METHADONE
STATE:MASSACHUSETTS

| ZIP CODE | QUARTER 1 | QUARTER 2 | QUARTER 3 | QUARTER 4 | TOTAL GRAMS |
|---|---|---|---|---|---|
| 23 | 7,406.26 | 9,981.77 | 9,980.25 | 5,717.27 | 33,085.55 |
| 24 | 1,510.91 | 1,428.27 | 1,802.04 | 1,006.74 | 5,747.96 |
| 25 | 3,178.34 | 3,617.67 | 3,149.36 | 2,235.64 | 12,181.01 |
| 26 | 3,416.88 | 4,115.31 | 3,430.47 | 2,689.73 | 13,652.39 |
| 27 | 21,010.53 | 20,222.65 | 20,288.75 | 15,165.27 | 76,687.20 |
| TOTAL | 142,094.35 | 148,189.44 | 144,690.11 | 106,370.51 | 541,344.41 |

STATE: MICHIGAN

| ZIP CODE | QUARTER 1 | QUARTER 2 | QUARTER 3 | QUARTER 4 | TOTAL GRAMS |
|---|---|---|---|---|---|

| | | | | | | |
|---|---|---|---|---|---|---|
| | 480 | 15,533.93 | 17,122.11 | 10,658.73 | 15,354.97 | 58,669.74 |
| | 481 | 14,086.53 | 14,575.05 | 14,242.96 | 11,807.06 | 54,711.60 |
| | 482 | 11,509.48 | 15,975.49 | 14,587.20 | 13,579.75 | 55,651.92 |
| | 483 | 10,382.46 | 13,175.14 | 10,430.76 | 11,127.23 | 45,115.59 |
| | 484 | 2,812.35 | 2,742.68 | 1,580.40 | 2,632.22 | 9,767.65 |
| | 485 | 8,553.59 | 8,793.29 | 12,521.60 | 8,820.04 | 38,688.52 |
| | 486 | 4,844.96 | 5,512.63 | 5,467.02 | 3,793.33 | 19,617.94 |
| | 487 | 928.39 | 927.49 | 877.41 | 833.58 | 3,566.87 |
| | 488 | 5,915.56 | 5,885.51 | 5,845.26 | 7,278.45 | 24,924.78 |
| | 489 | 7,504.02 | 8,487.71 | 7,340.45 | 4,200.08 | 27,532.26 |
| | 490 | 12,411.14 | 10,654.99 | 11,811.63 | 8,633.91 | 43,511.67 |
| | 491 | 571.97 | 594.33 | 538.34 | 447.65 | 2,152.29 |
| | 492 | 5,225.35 | 5,132.78 | 4,753.47 | 4,178.82 | 19,290.42 |
| | 493 | 1,513.95 | 1,501.43 | 1,243.93 | 1,367.27 | 5,626.58 |
| | 494 | 8,315.77 | 8,049.51 | 7,567.43 | 7,008.43 | 30,941.14 |
| | 495 | 9,592.45 | 10,294.58 | 9,803.01 | 7,902.57 | 37,592.61 |
| | 496 | 3,484.85 | 3,318.31 | 3,252.13 | 3,051.96 | 13,107.25 |
| | 497 | 4,157.35 | 4,947.73 | 4,523.43 | 4,162.98 | 17,791.49 |
| | 498 | 994.13 | 949.05 | 969.08 | 812.47 | 3,724.73 |
| | 499 | 228.52 | 258.48 | 253.56 | 227.18 | 967.74 |
| TOTAL | | 128,566.75 | 138,898.29 | 128,267.80 | 117,219.95 | 512,952.79 |
| STATE: | MINNESOTA | | | | | |
| ZIP CODE | QUARTER 1 | | QUARTER 2 | QUARTER 3 | QUARTER 4 | TOTAL GRAMS |
| | 550 | 1,327.38 | 1,253.14 | 1,242.14 | 1,183.74 | 5,006.40 |
| | 551 | 14,328.04 | 14,289.74 | 11,140.74 | 11,131.12 | 50,889.64 |
| | 553 | 2,572.92 | 2,594.21 | 2,536.79 | 2,899.82 | 10,603.74 |
| | 554 | 28,138.81 | 28,602.46 | 26,464.85 | 23,300.48 | 106,506.60 |
| | 556 | 35.33 | 30.41 | 26.38 | 18.34 | 110.46 |
| | 557 | 581.09 | 492.81 | 442.37 | 430.56 | 1,946.83 |
| | 558 | 2,679.71 | 3,182.90 | 2,685.70 | 3,332.53 | 11,880.84 |
| | 559 | 1,430.24 | 1,149.48 | 1,471.65 | 1,833.25 | 5,884.62 |
| | 560 | 177.36 | 182.99 | 197.39 | 172.98 | 730.72 |
| | 561 | 37.12 | 38.01 | 39.8 | 37.12 | 152.05 |
| | 562 | 177 | 171.64 | 176.47 | 144.18 | 669.29 |
| | 563 | 1,658.31 | 1,988.52 | 1,711.17 | 2,848.04 | 8,206.04 |
| | 564 | 4,930.38 | 3,129.06 | 3,992.42 | 3,980.08 | 16,031.94 |
| | 565 | 332.63 | 269.21 | 245.16 | 251.33 | 1,098.33 |
| | 566 | 170.03 | 289.79 | 169.4 | 155.54 | 784.76 |
| | 567 | 126.2 | 138.54 | 108.04 | 136.4 | 509.18 |
| TOTAL | | 58,702.55 | 57,802.91 | 52,650.47 | 51,855.51 | 221,011.44 |
| STATE: | MISSISSIPPI | | | | | |
| ZIP CODE | QUARTER 1 | | QUARTER 2 | QUARTER 3 | QUARTER 4 | TOTAL GRAMS |
| | 386 | 783.49 | 820.43 | 767.4 | 634.58 | 3,005.90 |
| | 387 | 490.58 | 435.13 | 397.11 | 427.52 | 1,750.34 |
| | 388 | 364.92 | 359.01 | 326.55 | 217.79 | 1,268.27 |
| | 389 | 154.28 | 150.71 | 139.97 | 109.12 | 554.08 |
| | 390 | 266.08 | 258.48 | 267.87 | 251.33 | 1,043.76 |
| | 391 | 266.08 | 279.5 | 203.92 | 227.62 | 977.12 |
| | 392 | 1,546.42 | 1,591.50 | 1,509.75 | 2,001.85 | 6,649.52 |
| | 393 | 603.72 | 616.24 | 615.35 | 508.91 | 2,344.22 |
| | 394 | 760.24 | 758.45 | 736.54 | 677.87 | 2,933.10 |
| | 395 | 1,494.54 | 1,437.30 | 1,291.25 | 1,041.17 | 5,264.26 |
| | 396 | 216.89 | 211.53 | 177.54 | 190.06 | 796.02 |
| | 397 | 177.9 | 165.2 | 172.8 | 171.28 | 687.18 |
| TOTAL | | 7,125.14 | 7,083.48 | 6,606.05 | 6,459.10 | 27,273.77 |
| STATE: | MISSOURI | | | | | |
| ZIP CODE | QUARTER 1 | | QUARTER 2 | QUARTER 3 | QUARTER 4 | TOTAL GRAMS |
| | 630 | 5,278.48 | 5,388.40 | 5,388.94 | 3,576.97 | 19,632.79 |

RETAIL DRUG DISTRIBUTION BY ZIP CODE WITHIN STATE BY GRAMS WT

DRUG CODE:9250BDRUG NAME:METHADONE
STATE:MISSOURI

| ZIP CODE | QUARTER 1 | QUARTER 2 | QUARTER 3 | QUARTER 4 | TOTAL GRAMS |
|---|---|---|---|---|---|
| 631 | 12,230.17 | 12,907.04 | 12,597.95 | 9,306.12 | 47,041.28 |
| 633 | 1,481.22 | 1,435.96 | 1,405.10 | 1,333.55 | 5,655.83 |
| 634 | 264.74 | 257.14 | 260.27 | 280.39 | 1,062.54 |
| 635 | 91.68 | 89.71 | 87.2 | 77.81 | 346.4 |
| 636 | 360.17 | 328.42 | 333.07 | 351.68 | 1,373.34 |
| 637 | 1,742.47 | 1,989.86 | 2,072.50 | 1,112.99 | 6,917.82 |
| 638 | 161.44 | 158.31 | 145.79 | 141.14 | 606.68 |
| 639 | 870.7 | 957.01 | 884.56 | 469.11 | 3,181.38 |
| 640 | 1,281.85 | 1,313.34 | 1,228.37 | 1,267.72 | 5,091.28 |
| 641 | 4,758.74 | 4,952.47 | 4,525.57 | 4,796.31 | 19,033.09 |
| 644 | 167.7 | 166.81 | 134.16 | 131.92 | 600.59 |
| 645 | 2,057.57 | 2,259.70 | 2,410.86 | 2,989.53 | 9,717.66 |
| 646 | 133.27 | 146.68 | 117.61 | 171.72 | 569.28 |
| 647 | 233.53 | 224.94 | 248.2 | 224.49 | 931.16 |
| 648 | 1,691.31 | 2,237.79 | 1,701.95 | 2,218.11 | 7,849.16 |
| 650 | 165.91 | 183.8 | 165.91 | 176.2 | 691.82 |
| 651 | 176.64 | 174.86 | 166.36 | 146.23 | 664.09 |
| 652 | 1,705.62 | 1,697.12 | 1,339.81 | 1,638.54 | 6,381.09 |
| 653 | 84.97 | 92.12 | 89.71 | 93.91 | 360.71 |
| 654 | 167.25 | 186.21 | 173.51 | 160.1 | 687.07 |
| 655 | 372.07 | 335.85 | 299.62 | 271.9 | 1,279.44 |
| 656 | 666.24 | 652.73 | 689.58 | 661.23 | 2,669.78 |
| 657 | 1,638.18 | 1,868.04 | 1,879.04 | 1,385.96 | 6,771.22 |
| 658 | 4,866.79 | 4,308.68 | 3,863.72 | 4,499.28 | 17,538.47 |
| TOTAL | 42,648.71 | 44,312.99 | 42,209.36 | 37,482.91 | 166,653.97 |

STATE: MONTANA

| ZIP CODE | QUARTER 1 | QUARTER 2 | QUARTER 3 | QUARTER 4 | TOTAL GRAMS |
|---|---|---|---|---|---|
| 590 | 83.63 | 80.5 | 88.1 | 84.97 | 337.2 |
| 591 | 1,521.46 | 1,130.97 | 1,287.31 | 1,269.96 | 5,209.70 |
| 592 | 25.49 | 35.33 | 16.1 | 16.1 | 93.02 |
| 593 | 39.35 | 49.64 | 54.56 | 47.4 | 190.95 |
| 594 | 355.52 | 329.85 | 331.91 | 309.02 | 1,326.30 |
| 595 | 47.85 | 51.43 | 53.22 | 53.66 | 206.16 |
| 596 | 435.13 | 430.21 | 450.33 | 385.04 | 1,700.71 |
| 597 | 631.89 | 626.26 | 867.3 | 497.73 | 2,623.18 |
| 598 | 2,417.12 | 2,472.39 | 2,322.40 | 2,322.76 | 9,534.67 |
| 599 | 2,486.52 | 2,048 | 1,991.11 | 1,969.74 | 8,495.37 |
| TOTAL | 8,043.96 | 7,254.58 | 7,462.34 | 6,956.38 | 29,717.26 |

STATE: NEBRASKA

| ZIP CODE | QUARTER 1 | QUARTER 2 | QUARTER 3 | QUARTER 4 | TOTAL GRAMS |
|---|---|---|---|---|---|
| 680 | 504.35 | 513.3 | 520.45 | 481.46 | 2,019.56 |
| 681 | 5,662.98 | 5,519.70 | 5,119.37 | 5,188.24 | 21,490.29 |
| 683 | 113.59 | 111.71 | 119.4 | 98.83 | 443.53 |
| 684 | 54.11 | 59.57 | 43.47 | 42.04 | 199.19 |
| 685 | 1,759.87 | 1,409.26 | 2,112.40 | 1,667.25 | 6,948.78 |
| 686 | 78.26 | 79.6 | 80.94 | 66.63 | 305.43 |
| 687 | 163.23 | 165.02 | 162.78 | 177.54 | 668.57 |
| 688 | 343.36 | 327.98 | 271.72 | 290.5 | 1,233.56 |
| 689 | 57.24 | 47.67 | 48.3 | 45.97 | 199.18 |
| 690 | 48.3 | 46.51 | 41.14 | 42.04 | 177.99 |
| 691 | 157.24 | 185.14 | 168.86 | 145.79 | 657.03 |
| 692 | 33.09 | 18.78 | 23.7 | 27.28 | 102.85 |
| 693 | 140.69 | 154.55 | 133.27 | 129.06 | 557.57 |
| TOTAL | 9,116.31 | 8,638.79 | 8,845.80 | 8,402.63 | 35,003.53 |

RETAIL DRUG DISTRIBUTION BY ZIP CODE WITHIN STATE BY GRAMS WT

STATE: NEVADA

| ZIP CODE | QUARTER 1 | QUARTER 2 | QUARTER 3 | QUARTER 4 | TOTAL GRAMS |
|---|---|---|---|---|---|
| 890 | 13,669.29 | 12,709.60 | 9,716.67 | 12,398.08 | 48,493.64 |
| 891 | 16,630.47 | 16,676.34 | 16,345.88 | 15,957.04 | 65,609.73 |
| 893 | 46.51 | 42.93 | 35.78 | 38.46 | 163.68 |
| 894 | 4,411.90 | 4,589.79 | 4,519.58 | 4,200.91 | 17,722.18 |
| 895 | 5,014.01 | 3,969.35 | 4,749.71 | 3,729.92 | 17,462.99 |
| 897 | 531.9 | 1,083.21 | 1,101.27 | 1,617.61 | 4,333.99 |
| 898 | 98.83 | 96.15 | 75.13 | 84.97 | 355.08 |
| TOTAL | 40,402.91 | 39,167.37 | 36,544.02 | 38,026.99 | 154,141.29 |

STATE: NEW HAMPSHIRE

| ZIP CODE | QUARTER 1 | QUARTER 2 | QUARTER 3 | QUARTER 4 | TOTAL GRAMS |
|---|---|---|---|---|---|

DRUG CODE:9250BDRUG NAME:METHADONE

STATE:NEW HAMPSHIRE

| ZIP CODE | QUARTER 1 | QUARTER 2 | QUARTER 3 | QUARTER 4 | TOTAL GRAMS |
|---|---|---|---|---|---|
| 30 | 3,872.22 | 4,018.81 | 4,183.02 | 2,948.84 | 15,022.89 |
| 31 | 9,527.86 | 9,732.15 | 9,588.06 | 6,794.04 | 35,642.11 |
| 32 | 731.89 | 742.35 | 696.47 | 647.1 | 2,817.81 |
| 33 | 4,672.17 | 6,728.66 | 3,714 | 5,081.98 | 20,196.81 |
| 34 | 2,879.97 | 1,979.75 | 3,746.19 | 2,008.82 | 10,614.73 |
| 35 | 279.5 | 337.19 | 300.97 | 254.46 | 1,172.12 |
| 36 | 54.11 | 42.48 | 40.7 | 33.99 | 171.28 |
| 37 | 2,630.52 | 2,667.64 | 2,558.43 | 2,009.90 | 9,866.49 |
| 38 | 6,320.72 | 7,067.10 | 6,553.09 | 5,167.13 | 25,108.04 |
| TOTAL | 30,968.96 | 33,316.13 | 31,380.93 | 24,946.26 | 120,612.28 |

STATE: NEW JERSEY

| ZIP CODE | QUARTER 1 | QUARTER 2 | QUARTER 3 | QUARTER 4 | TOTAL GRAMS |
|---|---|---|---|---|---|
| 70 | 9,916.48 | 8,638.47 | 7,912.49 | 7,776.63 | 34,244.07 |
| 71 | 11,570.07 | 11,613.35 | 12,437.23 | 13,563.65 | 49,184.30 |
| 72 | 2,113.20 | 2,944.45 | 1,926.81 | 968.19 | 7,952.65 |
| 73 | 5,433.66 | 5,152.19 | 4,543.55 | 3,809.70 | 18,939.10 |
| 74 | 848.88 | 818.82 | 817.48 | 794.32 | 3,279.50 |
| 75 | 2,873.53 | 4,672.17 | 2,867.36 | 2,860.11 | 13,273.17 |
| 76 | 2,042.81 | 2,551.81 | 2,600.47 | 2,288.77 | 9,483.86 |
| 77 | 6,907.72 | 6,895.73 | 7,870.90 | 8,293.59 | 29,967.94 |
| 78 | 563.56 | 2,694.83 | 536.64 | 2,595.64 | 6,390.67 |
| 79 | 262.42 | 344.25 | 215.19 | 256.25 | 1,078.11 |
| 80 | 5,460.85 | 4,763.93 | 3,877.76 | 3,462.58 | 17,565.12 |
| 81 | 20,967.96 | 21,391.72 | 26,025.97 | 16,879.92 | 85,265.57 |
| 82 | 6,091.76 | 6,595.40 | 8,418.36 | 7,379.87 | 28,485.39 |
| 83 | 740.12 | 968.19 | 2,600.11 | 4,009.60 | 8,318.02 |
| 84 | 7,207.16 | 4,279.35 | 695.93 | 140.51 | 12,322.95 |
| 85 | 454.36 | 432.62 | 392.19 | 445.86 | 1,725.03 |
| 86 | 5,653.32 | 5,846.16 | 5,512.90 | 4,812.32 | 21,824.70 |
| 87 | 9,212.32 | 10,453.30 | 8,611.01 | 9,027.98 | 37,304.61 |
| 88 | 9,953.42 | 9,596.55 | 8,857.96 | 9,428.23 | 37,836.16 |
| 89 | 2,980.32 | 3,583.50 | 2,283.76 | 2,896.60 | 11,744.18 |
| TOTAL | 111,253.92 | 114,236.79 | 109,004.07 | 101,690.32 | 436,185.10 |

STATE: NEW MEXICO

| ZIP CODE | QUARTER 1 | QUARTER 2 | QUARTER 3 | QUARTER 4 | TOTAL GRAMS |
|---|---|---|---|---|---|
| 870 | 1,321.03 | 1,306.27 | 1,302.69 | 728.94 | 4,658.93 |
| 871 | 30,829.61 | 31,426.80 | 30,516.48 | 23,723.51 | 116,496.40 |
| 873 | 70.21 | 67.97 | 82.28 | 75.13 | 295.59 |
| 874 | 509.36 | 155.45 | 1,036.61 | 135.05 | 1,836.47 |
| 875 | 3,165.10 | 4,195 | 5,236.71 | 3,995.46 | 16,592.27 |

| | | | | | |
|---|---|---|---|---|---|
| 877 | 61.71 | 62.61 | 68.87 | 69.32 | 262.51 |
| 878 | 7.16 | 3.58 | 2.68 | 5.81 | 19.23 |
| 879 | 45.17 | 46.96 | 29.52 | 15.2 | 136.85 |
| 880 | 2,057.84 | 1,911.60 | 2,096.03 | 1,699.81 | 7,765.28 |
| 881 | 46.96 | 39.8 | 33.99 | 46.06 | 166.81 |
| 882 | 704.88 | 1,081.96 | 1,006.20 | 1,041.26 | 3,834.30 |
| 883 | 276.28 | 221.36 | 212.6 | 174.41 | 884.65 |
| 884 | 17.44 | 21.47 | 16.99 | 14.31 | 70.21 |
| TOTAL | 39,112.75 | 40,540.83 | 41,641.65 | 31,724.27 | 153,019.50 |

STATE: NEW YORK

| ZIP CODE | QUARTER 1 | QUARTER 2 | QUARTER 3 | QUARTER 4 | TOTAL GRAMS |
|---|---|---|---|---|---|
| 100 | 84,854.32 | 92,199.61 | 89,341.85 | 75,958.81 | 342,354.59 |
| 101 | 1,337.49 | 1,202.52 | 1,330.42 | 1,340.08 | 5,210.51 |
| 102 | 15.2 | 16.99 | 15.2 | 13.42 | 60.81 |
| 103 | 13,204.65 | 13,157.97 | 11,750.98 | 8,922.18 | 47,035.78 |
| 104 | 55,913.06 | 60,768.49 | 62,033.88 | 47,258.98 | 225,974.41 |
| 105 | 4,131.39 | 4,752.39 | 4,487.29 | 3,645.04 | 17,016.11 |
| 106 | 164.12 | 121.37 | 152.41 | 122.8 | 560.7 |
| 107 | 6,566.24 | 6,689.13 | 4,832.98 | 4,855.43 | 22,943.78 |
| 108 | 1,313.87 | 2,035.30 | 1,323.26 | 2,011.51 | 6,683.94 |
| 109 | 3,256.42 | 3,552.29 | 2,609.05 | 3,180.22 | 12,597.98 |
| 110 | 3,136.03 | 3,444.48 | 3,046.42 | 3,219.61 | 12,846.54 |
| 111 | 254.46 | 260.27 | 229.41 | 218.05 | 962.19 |
| 112 | 31,725.17 | 36,604.75 | 37,819.79 | 24,155.24 | 130,304.95 |
| 113 | 4,523.25 | 4,389.89 | 4,874.57 | 3,702.64 | 17,490.35 |
| 114 | 7,205.73 | 7,024.62 | 7,631.29 | 4,933.78 | 26,795.42 |

DRUG CODE:9250BDRUG NAME:METHADONE
STATE:NEW YORK

| ZIP CODE | QUARTER 1 | QUARTER 2 | QUARTER 3 | QUARTER 4 | TOTAL GRAMS |
|---|---|---|---|---|---|
| 115 | 3,022.80 | 4,704.72 | 4,895.91 | 4,632.37 | 17,255.80 |
| 116 | 161.44 | 149.81 | 162.07 | 143.28 | 616.6 |
| 117 | 10,424.23 | 11,049.86 | 9,335.30 | 9,443.70 | 40,253.09 |
| 118 | 208.84 | 226.01 | 166.36 | 158.31 | 759.52 |
| 119 | 2,586.60 | 2,611.20 | 1,915.27 | 2,574.08 | 9,687.15 |
| 120 | 1,384.53 | 1,415.48 | 1,348.76 | 1,134.10 | 5,282.87 |
| 121 | 441.12 | 388.35 | 354.63 | 416.25 | 1,600.35 |
| 122 | 4,550.26 | 4,961.33 | 4,891.47 | 4,332.65 | 18,735.71 |
| 123 | 409.72 | 425.91 | 288.89 | 265.64 | 1,390.16 |
| 124 | 3,111.71 | 2,806.09 | 2,756.72 | 2,980.32 | 11,654.84 |
| 125 | 4,564.30 | 3,947.88 | 5,105.15 | 4,530.14 | 18,147.47 |
| 126 | 2,089.32 | 2,159.98 | 2,120.62 | 1,962.58 | 8,332.50 |
| 127 | 546.03 | 574.74 | 556.76 | 535.3 | 2,212.83 |
| 128 | 796.91 | 791.99 | 800.04 | 758.9 | 3,147.84 |
| 129 | 907.01 | 1,038.85 | 891.54 | 851.38 | 3,688.78 |
| 130 | 747.45 | 966.13 | 1,077.75 | 1,177.48 | 3,968.81 |
| 131 | 263.4 | 262.86 | 249.09 | 281.29 | 1,056.64 |
| 132 | 5,671.66 | 6,283.88 | 5,161.58 | 5,018.12 | 22,135.24 |
| 133 | 474.03 | 537.27 | 529.22 | 292.74 | 1,833.26 |
| 134 | 543.53 | 616.78 | 542.01 | 444.52 | 2,146.84 |
| 135 | 452.48 | 411.16 | 396.4 | 254.46 | 1,514.50 |
| 136 | 355.08 | 342.11 | 401.14 | 379.67 | 1,478 |
| 137 | 210.63 | 224.49 | 235.23 | 241.49 | 911.84 |
| 138 | 532.97 | 565.08 | 479.49 | 532.62 | 2,110.16 |
| 139 | 1,131.95 | 1,094.12 | 1,130.52 | 1,203.86 | 4,560.45 |
| 140 | 2,079.66 | 1,944.60 | 1,808.83 | 1,745.42 | 7,578.51 |
| 141 | 818.91 | 752.46 | 721.96 | 688.06 | 2,981.39 |
| 142 | 14,278.24 | 12,555.26 | 13,130.15 | 10,742.24 | 50,705.89 |
| 143 | 869.27 | 846.28 | 1,467.26 | 798.25 | 3,981.06 |

| | | | | | |
|---|---|---|---|---|---|
| 144 | 745.48 | 708.28 | 729.11 | 684.39 | 2,867.26 |
| 145 | 505.78 | 412.77 | 421.98 | 392.64 | 1,733.17 |
| 146 | 5,903.04 | 6,600.49 | 6,125.66 | 4,657.77 | 23,286.96 |
| 147 | 806.3 | 765.96 | 672.59 | 677.06 | 2,921.91 |
| 148 | 1,038.58 | 841.9 | 815.07 | 820.97 | 3,516.52 |
| 149 | 393.54 | 349.26 | 321.18 | 308.57 | 1,372.55 |
| TOTAL | 290,628.20 | 310,553.41 | 303,484.51 | 249,598.41 | 1,154,264.53 |

STATE: NORTH CAROLINA

| ZIP CODE | QUARTER 1 | QUARTER 2 | QUARTER 3 | QUARTER 4 | TOTAL GRAMS |
|---|---|---|---|---|---|
| 270 | 686 | 703 | 692.71 | 635.29 | 2,717 |
| 271 | 3,653.09 | 3,570.53 | 4,319.77 | 3,015.74 | 14,559.13 |
| 272 | 6,919.44 | 6,972.46 | 6,968.72 | 5,247.44 | 26,108.06 |
| 273 | 2,874.15 | 4,209.94 | 2,814.23 | 4,107.08 | 14,005.40 |
| 274 | 8,566.92 | 5,515.14 | 7,968.75 | 6,170.56 | 28,221.37 |
| 275 | 7,234.68 | 7,190.01 | 7,063.89 | 6,660.51 | 28,149.09 |
| 276 | 5,512.01 | 5,205.68 | 5,144.32 | 5,171.78 | 21,033.79 |
| 277 | 8,006.94 | 7,252.78 | 7,064.87 | 7,527.72 | 29,852.31 |
| 278 | 5,813.33 | 6,025.57 | 6,210.18 | 5,810.65 | 23,859.73 |
| 279 | 606.85 | 628.32 | 604.61 | 628.32 | 2,468.10 |
| 280 | 9,125.21 | 8,969.76 | 9,705.58 | 8,615.67 | 36,416.22 |
| 281 | 6,501.48 | 6,436.91 | 6,730.90 | 5,326.15 | 24,995.44 |
| 282 | 7,280.51 | 7,812.94 | 6,792.34 | 7,630.66 | 29,516.45 |
| 283 | 10,389.53 | 11,028.94 | 11,488.39 | 8,828.98 | 41,735.84 |
| 284 | 7,021.30 | 6,622.41 | 6,098.63 | 6,261.43 | 26,003.77 |
| 285 | 6,511.59 | 6,016.36 | 6,077 | 5,267.39 | 23,872.34 |
| 286 | 15,610.50 | 13,329.42 | 14,462 | 14,547.95 | 57,949.87 |
| 287 | 9,205.88 | 10,246.60 | 8,197.44 | 9,014.93 | 36,664.85 |
| 288 | 14,318.45 | 16,856.31 | 15,169.29 | 9,901.99 | 56,246.04 |
| 289 | 229.86 | 224.05 | 245.07 | 202.13 | 901.11 |
| TOTAL | 136,067.72 | 134,817.13 | 133,818.69 | 120,572.37 | 525,275.91 |

STATE: NORTH DAKOTA

| ZIP CODE | QUARTER 1 | QUARTER 2 | QUARTER 3 | QUARTER 4 | TOTAL GRAMS |
|---|---|---|---|---|---|
| 580 | 95.25 | 102.23 | 72.45 | 69.14 | 339.07 |
| 581 | 407.67 | 314.74 | 263.94 | 215.73 | 1,202.08 |
| 582 | 314.38 | 290.5 | 284.87 | 201.51 | 1,091.26 |
| 583 | 40.7 | 42.04 | 42.04 | 34.88 | 159.66 |
| 584 | 92.75 | 73.16 | 62.61 | 42.04 | 270.56 |

DRUG CODE:9250BDRUG NAME:METHADONE
STATE:NORTH DAKOTA

| ZIP CODE | QUARTER 1 | QUARTER 2 | QUARTER 3 | QUARTER 4 | TOTAL GRAMS |
|---|---|---|---|---|---|
| 585 | 307.23 | 257.59 | 270.11 | 284.69 | 1,119.62 |
| 586 | 46.96 | 48.74 | 41.59 | 37.65 | 174.94 |
| 587 | 48.3 | 52.32 | 332.27 | 51.88 | 484.77 |
| 588 | 72.45 | 65.29 | 81.84 | 55.45 | 275.03 |
| TOTAL | 1,425.69 | 1,246.61 | 1,451.72 | 992.97 | 5,116.99 |

STATE: OHIO

| ZIP CODE | QUARTER 1 | QUARTER 2 | QUARTER 3 | QUARTER 4 | TOTAL GRAMS |
|---|---|---|---|---|---|
| 430 | 1,975.55 | 1,929.58 | 1,755.89 | 1,776.64 | 7,437.66 |
| 431 | 1,055.66 | 1,024.98 | 953.79 | 931.61 | 3,966.04 |
| 432 | 15,594.04 | 15,306.20 | 15,994.86 | 15,361.43 | 62,256.53 |
| 433 | 838.05 | 823.21 | 775.8 | 698.08 | 3,135.14 |
| 434 | 334.51 | 337.28 | 306.33 | 300.07 | 1,278.19 |
| 435 | 458.2 | 481.9 | 458.29 | 447.29 | 1,845.68 |
| 436 | 3,667.49 | 4,526.74 | 4,455.10 | 4,275.41 | 16,924.74 |
| 437 | 459.81 | 443.8 | 408.51 | 401.94 | 1,714.06 |
| 438 | 119.31 | 115.91 | 95.7 | 108.13 | 439.05 |

RETAIL DRUG DISTRIBUTION BY ZIP CODE WITHIN STATE BY GRAMS WT

| | | | | | |
|---|---|---|---|---|---|
| 439 | 673.21 | 646.83 | 585.65 | 609.27 | 2,514.96 |
| 440 | 1,823.50 | 1,720.83 | 1,544.36 | 1,603.93 | 6,692.62 |
| 441 | 11,038.77 | 10,438.54 | 11,069.09 | 10,145.09 | 42,691.49 |
| 442 | 996.9 | 955.49 | 1,017.83 | 957.28 | 3,927.50 |
| 443 | 4,251.17 | 4,316.64 | 4,167.55 | 4,323.35 | 17,058.71 |
| 444 | 477.16 | 442.73 | 1,174.35 | 1,785.22 | 3,879.46 |
| 445 | 4,724.67 | 5,079.74 | 4,666.71 | 4,160.84 | 18,631.96 |
| 446 | 683.05 | 667.49 | 621.7 | 667.85 | 2,640.09 |
| 447 | 2,170.71 | 2,165.79 | 2,114.90 | 1,883.87 | 8,335.27 |
| 448 | 921.32 | 859.25 | 810.95 | 803.89 | 3,395.41 |
| 449 | 203.03 | 162.87 | 173.96 | 155.63 | 695.49 |
| 450 | 2,717.72 | 2,905.82 | 3,105.63 | 3,418.40 | 12,147.57 |
| 451 | 1,087.14 | 1,012.01 | 987.86 | 962.37 | 4,049.38 |
| 452 | 12,310.91 | 12,683.32 | 12,377.72 | 12,859.78 | 50,231.73 |
| 453 | 1,126.41 | 1,043.41 | 997.43 | 994.75 | 4,162 |
| 454 | 5,285.46 | 5,377.76 | 5,150.13 | 5,145.93 | 20,959.28 |
| 455 | 177.99 | 197.22 | 182.46 | 173.51 | 731.18 |
| 456 | 868.55 | 842.88 | 836.98 | 922.57 | 3,470.98 |
| 457 | 523.94 | 524.92 | 506.23 | 495.14 | 2,050.23 |
| 458 | 1,166.92 | 730.1 | 946.81 | 858.18 | 3,702.01 |
| TOTAL | 77,731.15 | 77,763.24 | 78,242.57 | 77,227.45 | 310,964.41 |

STATE: OKLAHOMA

| ZIP CODE | QUARTER 1 | QUARTER 2 | QUARTER 3 | QUARTER 4 | TOTAL GRAMS |
|---|---|---|---|---|---|
| 730 | 1,378.63 | 1,374.60 | 1,214.68 | 1,194.11 | 5,162.02 |
| 731 | 12,624.81 | 11,210.59 | 11,381.06 | 8,097.27 | 43,313.73 |
| 734 | 3,214.92 | 2,231.08 | 2,581.15 | 1,223.63 | 9,250.78 |
| 735 | 1,847.83 | 1,407.79 | 1,368.88 | 474.93 | 5,099.43 |
| 736 | 66.19 | 55.9 | 65.29 | 84.07 | 271.45 |
| 737 | 151.78 | 122.53 | 143.1 | 137.56 | 554.97 |
| 738 | 82.73 | 78.71 | 88.99 | 84.07 | 334.5 |
| 739 | 24.6 | 24.15 | 22.81 | 23.7 | 95.26 |
| 740 | 3,542.90 | 3,640.12 | 2,915.74 | 2,785.43 | 12,884.19 |
| 741 | 13,732.35 | 13,166.28 | 11,184.29 | 9,268.04 | 47,350.96 |
| 743 | 888.59 | 1,387.21 | 1,380.95 | 817.03 | 4,473.78 |
| 744 | 903.34 | 814.8 | 826.87 | 795.12 | 3,340.13 |
| 745 | 262.06 | 265.19 | 259.38 | 266.53 | 1,053.16 |
| 746 | 145.79 | 158.76 | 448.54 | 135.5 | 888.59 |
| 747 | 266.53 | 276.37 | 298.73 | 285.76 | 1,127.39 |
| 748 | 394.79 | 381.1 | 328.24 | 367.6 | 1,471.73 |
| 749 | 4,298.93 | 5,059.17 | 3,829.82 | 3,300.78 | 16,488.70 |
| TOTAL | 43,826.77 | 41,654.35 | 38,338.52 | 29,341.13 | 153,160.77 |

STATE: OREGON

| ZIP CODE | QUARTER 1 | QUARTER 2 | QUARTER 3 | QUARTER 4 | TOTAL GRAMS |
|---|---|---|---|---|---|
| 970 | 5,423.73 | 3,233.97 | 2,896.42 | 2,674.08 | 14,228.20 |
| 971 | 1,822.88 | 1,745.87 | 1,698.47 | 1,518.60 | 6,785.82 |
| 972 | 33,719.05 | 37,209.87 | 34,679.29 | 29,768.52 | 135,376.73 |
| 973 | 9,802.18 | 7,644.62 | 9,423.49 | 7,154.22 | 34,024.51 |
| 974 | 8,164.98 | 8,948.29 | 9,075.39 | 6,812.38 | 33,001.04 |
| 975 | 7,319.23 | 7,743.27 | 7,243.57 | 5,388.22 | 27,694.29 |
| 976 | 466.52 | 444.7 | 385.93 | 328.24 | 1,625.39 |

DRUG CODE:9250BDRUG NAME:METHADONE
STATE:OREGON

| ZIP CODE | QUARTER 1 | QUARTER 2 | QUARTER 3 | QUARTER 4 | TOTAL GRAMS |
|---|---|---|---|---|---|
| 977 | 1,500.27 | 1,395.89 | 1,440.88 | 1,341.33 | 5,678.37 |
| 978 | 733.32 | 682.25 | 621.61 | 580.91 | 2,618.09 |
| 979 | 233.44 | 241.04 | 211.97 | 190.33 | 876.78 |

RETAIL DRUG DISTRIBUTION BY ZIP CODE WITHIN STATE BY GRAMS WT

| | | | | | |
|---|---|---|---|---|---|
| TOTAL | 69,185.60 | 69,289.77 | 67,677.02 | 55,756.83 | 261,909.22 |
| STATE: | PENNSYLVANIA | | | | |
| ZIP CODE | QUARTER 1 | QUARTER 2 | QUARTER 3 | QUARTER 4 | TOTAL GRAMS |
| 150 | 3,868.82 | 3,157.14 | 3,750.49 | 3,637.44 | 14,413.89 |
| 151 | 811.04 | 652.55 | 649.96 | 667.58 | 2,781.13 |
| 152 | 20,994.07 | 20,246.09 | 22,096.87 | 19,529.76 | 82,866.79 |
| 153 | 2,499.76 | 3,031.30 | 2,875.05 | 2,052.74 | 10,458.85 |
| 154 | 4,844.96 | 4,613.32 | 3,561.32 | 2,219.45 | 15,239.05 |
| 155 | 88.1 | 73.16 | 61.71 | 105.54 | 328.51 |
| 156 | 7,979.84 | 8,835.87 | 8,462.37 | 7,097.33 | 32,375.41 |
| 157 | 1,058.97 | 1,510.82 | 884.56 | 901.11 | 4,355.46 |
| 158 | 160.1 | 143.73 | 141.05 | 112.96 | 557.84 |
| 159 | 3,948.33 | 3,160.63 | 3,170.11 | 3,939.21 | 14,218.28 |
| 160 | 5,131.71 | 3,775.71 | 4,249.20 | 4,213.97 | 17,370.59 |
| 161 | 6,529.03 | 7,743.45 | 5,341.71 | 7,701.95 | 27,316.14 |
| 162 | 239.25 | 221.63 | 222.71 | 220.47 | 904.06 |
| 163 | 2,102.11 | 2,678.82 | 2,297.27 | 2,218.92 | 9,297.12 |
| 164 | 183.35 | 138.63 | 128.79 | 119.4 | 570.17 |
| 165 | 3,124.41 | 3,331.73 | 2,944.99 | 2,731.14 | 12,132.27 |
| 166 | 6,208.48 | 7,072.02 | 5,464.52 | 5,760.29 | 24,505.31 |
| 167 | 259.82 | 254.01 | 225.39 | 213.14 | 952.36 |
| 168 | 3,913.89 | 3,198.37 | 3,159.92 | 2,629.09 | 12,901.27 |
| 169 | 136.4 | 129.69 | 131.48 | 126.56 | 524.13 |
| 170 | 5,546.04 | 6,456.11 | 5,506.43 | 5,355.63 | 22,864.21 |
| 171 | 3,979.19 | 3,935.81 | 4,796.22 | 3,024.41 | 15,735.63 |
| 172 | 740.12 | 686 | 655.6 | 612.22 | 2,693.94 |
| 173 | 1,356.80 | 1,210.21 | 1,251.71 | 1,113.80 | 4,932.52 |
| 174 | 3,774.37 | 3,745.93 | 3,703.62 | 2,941.68 | 14,165.60 |
| 175 | 509.36 | 537.53 | 507.12 | 464.64 | 2,018.65 |
| 176 | 3,574.65 | 4,625.03 | 4,627.27 | 4,577.27 | 17,404.22 |
| 177 | 4,442.48 | 4,209.23 | 3,716.14 | 3,833.49 | 16,201.34 |
| 178 | 1,666.98 | 1,698.29 | 1,552.95 | 1,589.17 | 6,507.39 |
| 179 | 185.59 | 178.88 | 183.98 | 165.02 | 713.47 |
| 180 | 6,042.03 | 5,267.93 | 5,281.25 | 5,303.26 | 21,894.47 |
| 181 | 2,050.68 | 2,253.71 | 2,353.17 | 1,735.23 | 8,392.79 |
| 182 | 225.84 | 260.72 | 235.67 | 201.69 | 923.92 |
| 183 | 3,506.50 | 3,560.61 | 3,479.66 | 3,409.90 | 13,956.67 |
| 184 | 470.54 | 404.89 | 353.82 | 360.62 | 1,589.87 |
| 185 | 2,506.47 | 2,472.39 | 2,623.28 | 1,833.88 | 9,436.02 |
| 186 | 588.43 | 601.75 | 594.42 | 625.63 | 2,410.23 |
| 187 | 6,093.28 | 6,266.08 | 6,107.14 | 4,371.56 | 22,838.06 |
| 188 | 212.87 | 216.44 | 209.74 | 212.87 | 851.92 |
| 189 | 5,396.18 | 4,855.25 | 4,987 | 4,589.08 | 19,827.51 |
| 190 | 15,374.74 | 14,422.29 | 13,931.89 | 11,007.02 | 54,735.94 |
| 191 | 40,510.51 | 37,997.91 | 36,444.16 | 27,098.18 | 142,050.76 |
| 193 | 4,061.92 | 4,953.54 | 3,958.88 | 3,910.14 | 16,884.48 |
| 194 | 11,735.15 | 10,110.48 | 12,064.74 | 15,304.97 | 49,215.34 |
| 195 | 211.08 | 206.16 | 208.4 | 173.42 | 799.06 |
| 196 | 2,562.01 | 3,657.11 | 2,550.83 | 2,548.15 | 11,318.10 |
| TOTAL | 201,406.25 | 198,758.95 | 191,704.56 | 172,560.98 | 764,430.74 |
| STATE: | PUERTO RICO | | | | |
| ZIP CODE | QUARTER 1 | QUARTER 2 | QUARTER 3 | QUARTER 4 | TOTAL GRAMS |
| 6 | 1,336.68 | 1,293.75 | 1,079.99 | 1,294.20 | 5,004.62 |
| 7 | 2,691.70 | 6,275.02 | 4,477.37 | 5,378.65 | 18,822.74 |
| 9 | 13,718.75 | 13,813.56 | 13,896.29 | 9,426.08 | 50,854.68 |
| TOTAL | 17,747.13 | 21,382.33 | 19,453.65 | 16,098.93 | 74,682.04 |
| STATE: | RHODE ISLAND | | | | |
| ZIP CODE | QUARTER 1 | QUARTER 2 | QUARTER 3 | QUARTER 4 | TOTAL GRAMS |

| | | | | | | |
|---|---|---|---|---|---|---|
| | 28 | 23,874.76 | 23,016.22 | 21,626.32 | 20,784.51 | 89,301.81 |
| | 29 | 20,698.83 | 17,476.84 | 22,147.40 | 20,177.13 | 80,500.20 |
| TOTAL | | 44,573.59 | 40,493.06 | 43,773.72 | 40,961.64 | 169,802.01 |

STATE: SOUTH CAROLINA

| ZIP CODE | QUARTER 1 | QUARTER 2 | QUARTER 3 | QUARTER 4 | TOTAL GRAMS |
|---|---|---|---|---|---|

DRUG CODE:9250BDRUG NAME:METHADONE
STATE:SOUTH CAROLINA

| ZIP CODE | QUARTER 1 | QUARTER 2 | QUARTER 3 | QUARTER 4 | TOTAL GRAMS |
|---|---|---|---|---|---|
| 290 | 1,024.09 | 986.08 | 1,059.69 | 992.78 | 4,062.64 |
| 291 | 3,300.34 | 2,662.90 | 2,981.93 | 3,168.86 | 12,114.03 |
| 292 | 3,291.57 | 3,429.58 | 3,309.10 | 2,127.96 | 12,158.21 |
| 293 | 6,358.02 | 6,524.38 | 5,788.65 | 4,781.28 | 23,452.33 |
| 294 | 8,697.15 | 8,224.63 | 7,225.32 | 4,824.48 | 28,971.58 |
| 295 | 16,510.71 | 15,946.17 | 16,320.56 | 13,753.55 | 62,530.99 |
| 296 | 13,621 | 13,250.71 | 12,315.17 | 11,419.16 | 50,606.04 |
| 297 | 5,112.75 | 3,324.57 | 4,645.51 | 4,606.16 | 17,688.99 |
| 298 | 1,671.19 | 1,942.19 | 1,731.56 | 1,216.38 | 6,561.32 |
| 299 | 1,137.23 | 1,112.63 | 953.88 | 796.91 | 4,000.65 |
| TOTAL | 60,724.05 | 57,403.84 | 56,331.37 | 47,687.52 | 222,146.78 |

STATE: SOUTH DAKOTA

| ZIP CODE | QUARTER 1 | QUARTER 2 | QUARTER 3 | QUARTER 4 | TOTAL GRAMS |
|---|---|---|---|---|---|
| 570 | 505.78 | 488.79 | 479.4 | 502.3 | 1,976.27 |
| 571 | 1,422.81 | 1,471.29 | 1,260.66 | 1,204.76 | 5,359.52 |
| 572 | 24.6 | 25.49 | 29.07 | 25.49 | 104.65 |
| 573 | 128.35 | 112.69 | 124.32 | 140.69 | 506.05 |
| 574 | 46.06 | 40.52 | 41.14 | 28.17 | 155.89 |
| 575 | 46.51 | 62.61 | 55.01 | 32.2 | 196.33 |
| 576 | 12.07 | 14.31 | 9.39 | 6.26 | 42.03 |
| 577 | 371.53 | 405.07 | 362.41 | 343 | 1,482.01 |
| TOTAL | 2,557.71 | 2,620.77 | 2,361.40 | 2,282.87 | 9,822.75 |

STATE: TENNESSEE

| ZIP CODE | QUARTER 1 | QUARTER 2 | QUARTER 3 | QUARTER 4 | TOTAL GRAMS |
|---|---|---|---|---|---|
| 370 | 2,256.03 | 2,097.55 | 2,037.17 | 2,002.92 | 8,393.67 |
| 371 | 1,697.57 | 1,579.42 | 1,605.72 | 1,498.39 | 6,381.10 |
| 372 | 5,046.38 | 5,211.76 | 5,133.05 | 5,016.60 | 20,407.79 |
| 373 | 1,464.13 | 1,338.56 | 1,236.51 | 1,136.07 | 5,175.27 |
| 374 | 8,890.78 | 8,746.34 | 7,766.70 | 7,735.67 | 33,139.49 |
| 376 | 944.04 | 943.59 | 876.06 | 784.75 | 3,548.44 |
| 377 | 460.17 | 340.05 | 250.25 | 230.31 | 1,280.78 |
| 378 | 538.88 | 339.6 | 255.62 | 284.42 | 1,418.52 |
| 379 | 10,287.48 | 11,866.81 | 10,336.13 | 9,997.74 | 42,488.16 |
| 380 | 3,602.64 | 3,576.71 | 3,498.54 | 2,443.23 | 13,121.12 |
| 381 | 8,755.82 | 9,570.17 | 9,584.30 | 7,794.25 | 35,704.54 |
| 382 | 2,957.78 | 3,269.03 | 3,509.63 | 3,118.33 | 12,854.77 |
| 383 | 4,455.72 | 3,812.83 | 3,193.01 | 2,669.69 | 14,131.25 |
| 384 | 1,154.58 | 1,045.82 | 1,493.65 | 1,066.75 | 4,760.80 |
| 385 | 383.25 | 368.94 | 314.83 | 337.19 | 1,404.21 |
| TOTAL | 52,895.25 | 54,107.18 | 51,091.17 | 46,116.31 | 204,209.91 |

STATE: TEXAS

| ZIP CODE | QUARTER 1 | QUARTER 2 | QUARTER 3 | QUARTER 4 | TOTAL GRAMS |
|---|---|---|---|---|---|
| 750 | 7,576.46 | 9,188.53 | 7,254.30 | 7,474.41 | 31,493.70 |
| 751 | 1,334.44 | 1,328.45 | 1,397.05 | 1,253.05 | 5,312.99 |
| 752 | 8,847.58 | 9,742.12 | 9,106.11 | 7,785 | 35,480.81 |
| 753 | 19.32 | 34.43 | 17.62 | 20.75 | 92.12 |

| | | | | | |
|---|---|---|---|---|---|
| 754 | 1,793 | 1,898.54 | 1,770.64 | 1,665.64 | 7,127.82 |
| 755 | 318.41 | 325.56 | 318.41 | 309.46 | 1,271.84 |
| 756 | 3,864.61 | 3,016.81 | 3,406.14 | 3,883.48 | 14,171.04 |
| 757 | 5,434.28 | 5,343.15 | 4,711.70 | 4,513.50 | 20,002.63 |
| 758 | 342.64 | 336.74 | 326.9 | 256.69 | 1,262.97 |
| 759 | 1,969.92 | 2,274.01 | 1,187.05 | 1,267.36 | 6,698.34 |
| 760 | 3,674.20 | 2,940.88 | 3,577.69 | 2,801.44 | 12,994.21 |
| 761 | 10,222.28 | 10,922.95 | 10,829.75 | 9,117.60 | 41,092.58 |
| 762 | 2,396.72 | 2,381.79 | 2,333.67 | 1,878.69 | 8,990.87 |
| 763 | 858.36 | 914.08 | 806.57 | 721.6 | 3,300.61 |
| 764 | 386.92 | 331.82 | 314.47 | 296.14 | 1,329.35 |
| 765 | 633.95 | 1,047.97 | 958.44 | 553.63 | 3,193.99 |
| 766 | 290.23 | 263.85 | 279.5 | 251.33 | 1,084.91 |
| 767 | 2,383.58 | 1,788.80 | 2,347.35 | 1,715.82 | 8,235.55 |
| 768 | 355.97 | 391.03 | 371.53 | 346.58 | 1,465.11 |
| 769 | 1,352.24 | 1,401.35 | 1,037.68 | 1,050.74 | 4,842.01 |
| 770 | 14,058.90 | 13,203.56 | 13,689.44 | 11,101.87 | 52,053.77 |
| 772 | 429.31 | 429.31 | 0 | 429.31 | 1,287.93 |
| 773 | 4,202.07 | 4,379.97 | 4,472.89 | 3,537.89 | 16,592.82 |
| 774 | 1,072.74 | 1,092.06 | 1,083.03 | 1,088.84 | 4,336.67 |

DRUG CODE:9250BDRUG NAME:METHADONE
STATE:TEXAS

| ZIP CODE | QUARTER 1 | QUARTER 2 | QUARTER 3 | QUARTER 4 | TOTAL GRAMS |
|---|---|---|---|---|---|
| 775 | 5,045.94 | 6,232.36 | 5,316.67 | 4,723.06 | 21,318.03 |
| 776 | 1,741.22 | 1,730.04 | 2,681.68 | 1,658.22 | 7,811.16 |
| 777 | 1,038.22 | 1,074.71 | 778.75 | 244.62 | 3,136.30 |
| 778 | 265.19 | 263.4 | 262.42 | 276.55 | 1,067.56 |
| 779 | 1,509.30 | 1,305.38 | 1,709.65 | 826.87 | 5,351.20 |
| 780 | 2,206.22 | 2,582.31 | 2,202.91 | 2,138.78 | 9,130.22 |
| 781 | 1,785.85 | 1,931.37 | 2,047.73 | 1,487.03 | 7,251.98 |
| 782 | 29,460.82 | 27,945.26 | 26,515.02 | 25,687.35 | 109,608.45 |
| 783 | 355.35 | 336.74 | 326.01 | 317.51 | 1,335.61 |
| 784 | 6,083.08 | 7,170.40 | 6,316.70 | 6,201.05 | 25,771.23 |
| 785 | 1,782.45 | 1,295.63 | 2,571.22 | 476.18 | 6,125.48 |
| 786 | 2,058.82 | 2,586.34 | 2,302.99 | 2,763.52 | 9,711.67 |
| 787 | 7,626.37 | 8,015.70 | 8,224.99 | 7,121.84 | 30,988.90 |
| 788 | 131.92 | 141.76 | 122.09 | 123.87 | 519.64 |
| 789 | 68.87 | 58.58 | 65.11 | 42.48 | 235.04 |
| 790 | 254.46 | 269.66 | 258.3 | 243.55 | 1,025.97 |
| 791 | 1,310.74 | 1,557.15 | 1,299.83 | 1,011.57 | 5,179.29 |
| 792 | 63.77 | 59.48 | 43.38 | 53.66 | 220.29 |
| 793 | 168.59 | 141.76 | 161.89 | 136.84 | 609.08 |
| 794 | 1,594.45 | 2,014.90 | 1,981.01 | 850.75 | 6,441.11 |
| 795 | 193.64 | 215.55 | 245.07 | 231.2 | 885.46 |
| 796 | 348.37 | 680.19 | 408.2 | 465.54 | 1,902.30 |
| 797 | 2,796.07 | 2,518.99 | 2,388.76 | 1,980.74 | 9,684.56 |
| 798 | 23.7 | 11.63 | 13.42 | 7.6 | 56.35 |
| 799 | 2,766.74 | 2,782.21 | 2,074.11 | 1,818.32 | 9,441.38 |
| TOTAL | 144,498.28 | 147,899.26 | 141,915.84 | 124,209.52 | 558,522.90 |

STATE: UTAH

| ZIP CODE | QUARTER 1 | QUARTER 2 | QUARTER 3 | QUARTER 4 | TOTAL GRAMS |
|---|---|---|---|---|---|
| 840 | 7,811.06 | 8,426.32 | 7,769.03 | 6,251.41 | 30,257.82 |
| 841 | 11,958.31 | 16,445.78 | 11,097 | 11,045.57 | 50,546.66 |
| 843 | 524.74 | 381.73 | 378.78 | 344.79 | 1,630.04 |
| 844 | 2,334.92 | 2,350.30 | 2,345.92 | 2,311.40 | 9,342.54 |
| 845 | 282.27 | 219.13 | 220.29 | 229.41 | 951.1 |
| 846 | 2,050.95 | 1,538.19 | 2,014.90 | 1,519.50 | 7,123.54 |
| 847 | 2,856.18 | 2,816.20 | 2,898.93 | 2,153.63 | 10,724.94 |

| | | | | | |
|---|---|---|---|---|---|
| TOTAL | 27,818.43 | 32,177.65 | 26,724.85 | 23,855.71 | 110,576.64 |
| STATE: | VERMONT | | | | |
| ZIP CODE | QUARTER 1 | QUARTER 2 | QUARTER 3 | QUARTER 4 | TOTAL GRAMS |
| 50 | 710.6 | 662.75 | 630.55 | 526.8 | 2,530.70 |
| 51 | 262.06 | 285.76 | 277.71 | 260.27 | 1,085.80 |
| 52 | 202.13 | 192.3 | 186.04 | 182.01 | 762.48 |
| 53 | 1,821.45 | 1,816.08 | 2,002.56 | 1,191.79 | 6,831.88 |
| 54 | 5,971.10 | 7,526.64 | 4,865.36 | 5,423.19 | 23,786.29 |
| 56 | 2,384.92 | 1,914.11 | 2,534.73 | 1,476.21 | 8,309.97 |
| 57 | 2,383.40 | 2,769.69 | 2,750.55 | 3,071.01 | 10,974.65 |
| 58 | 3,242.38 | 4,131.77 | 3,657.02 | 2,867.45 | 13,898.62 |
| TOTAL | 16,978.04 | 19,299.10 | 16,904.52 | 14,998.73 | 68,180.39 |
| STATE: | VIRGIN ISLANDS | | | | |
| ZIP CODE | QUARTER 1 | QUARTER 2 | QUARTER 3 | QUARTER 4 | TOTAL GRAMS |
| 8 | 36.22 | 44.27 | 47.4 | 37.12 | 165.01 |
| TOTAL | 36.22 | 44.27 | 47.4 | 37.12 | 165.01 |
| STATE: | VIRGINIA | | | | |
| ZIP CODE | QUARTER 1 | QUARTER 2 | QUARTER 3 | QUARTER 4 | TOTAL GRAMS |
| 201 | 2,684.72 | 3,435.75 | 2,232.87 | 3,064.30 | 11,417.64 |
| 220 | 1,664.12 | 1,848.90 | 2,160.24 | 1,068.27 | 6,741.53 |
| 221 | 649.33 | 656.22 | 563.02 | 582.25 | 2,450.82 |
| 222 | 170.38 | 133.62 | 136.22 | 133.98 | 574.2 |
| 223 | 960.59 | 925.7 | 469.56 | 917.21 | 3,273.06 |
| 224 | 4,254.03 | 3,530.91 | 4,208.96 | 2,775.86 | 14,769.76 |
| 225 | 266.98 | 246.85 | 234.33 | 191.4 | 939.56 |
| 226 | 2,988.01 | 2,896.96 | 2,941.59 | 3,130.58 | 11,957.14 |
| 227 | 259.38 | 244.62 | 261.61 | 263.85 | 1,029.46 |
| 228 | 3,319.48 | 2,738.65 | 3,143.46 | 3,673.84 | 12,875.43 |
| 229 | 4,366.19 | 4,434.52 | 5,014.54 | 3,514.28 | 17,329.53 |
| 230 | 579.12 | 1,144.38 | 1,088.04 | 1,568.60 | 4,380.14 |
| 231 | 1,303.77 | 1,072.30 | 1,224.17 | 1,441.68 | 5,041.92 |

DRUG CODE:9250BDRUG NAME:METHADONE
STATE:VIRGINIA

| ZIP CODE | QUARTER 1 | QUARTER 2 | QUARTER 3 | QUARTER 4 | TOTAL GRAMS |
|---|---|---|---|---|---|
| 232 | 15,394.95 | 17,163.27 | 14,807.06 | 12,386.10 | 59,751.38 |
| 233 | 2,962.07 | 3,306.78 | 2,869.77 | 3,301.77 | 12,440.39 |
| 234 | 4,711.34 | 3,981.15 | 4,945.94 | 3,914.07 | 17,552.50 |
| 235 | 2,115.57 | 1,531.43 | 2,091.16 | 1,499.01 | 7,237.17 |
| 236 | 5,058.55 | 4,681.47 | 4,651.15 | 6,072.62 | 20,463.79 |
| 237 | 880.89 | 885.19 | 871.86 | 872.58 | 3,510.52 |
| 238 | 581.36 | 559 | 516.07 | 474.03 | 2,130.46 |
| 239 | 213.94 | 236.12 | 182.01 | 164.12 | 796.19 |
| 240 | 3,794.49 | 4,861.42 | 4,887.72 | 3,355.79 | 16,899.42 |
| 241 | 1,459.03 | 1,405.55 | 1,239.55 | 1,185.62 | 5,289.75 |
| 242 | 812.12 | 753.08 | 720.89 | 714.36 | 3,000.45 |
| 243 | 4,934.40 | 4,082.94 | 5,037.71 | 3,558.82 | 17,613.87 |
| 244 | 586.73 | 602.83 | 564.37 | 556.59 | 2,310.52 |
| 245 | 4,029.81 | 4,112.09 | 3,815.06 | 3,338.62 | 15,295.58 |
| 246 | 3,971.58 | 3,895.11 | 4,283.28 | 2,377.58 | 14,527.55 |
| TOTAL | 74,972.93 | 75,366.81 | 75,162.21 | 66,097.78 | 291,599.73 |
| STATE: | WASHINGTON | | | | |
| ZIP CODE | QUARTER 1 | QUARTER 2 | QUARTER 3 | QUARTER 4 | TOTAL GRAMS |
| 980 | 15,052.66 | 16,331.92 | 14,433.29 | 12,446.11 | 58,263.98 |
| 981 | 25,760.24 | 25,975.25 | 25,670.08 | 14,526.58 | 91,932.15 |
| 982 | 15,904.85 | 15,570.25 | 16,531.64 | 11,662.80 | 59,669.54 |

| | | | | | |
|---|---|---|---|---|---|
| 983 | 4,864.10 | 4,624.05 | 4,367.27 | 4,208.60 | 18,064.02 |
| 984 | 13,292.57 | 12,312.94 | 12,507.20 | 8,931.66 | 47,044.37 |
| 985 | 8,263.99 | 7,589.52 | 7,770.01 | 1,565.02 | 25,188.54 |
| 986 | 7,680.57 | 8,321.59 | 7,847.38 | 6,324.48 | 30,174.02 |
| 988 | 999.58 | 939.12 | 883.04 | 832.78 | 3,654.52 |
| 989 | 2,563.35 | 2,167.58 | 2,073.40 | 1,993.53 | 8,797.86 |
| 990 | 374.31 | 350.6 | 287.55 | 329.59 | 1,342.05 |
| 991 | 784.84 | 772.31 | 717.76 | 687.97 | 2,962.88 |
| 992 | 11,043.16 | 11,609.13 | 11,755.55 | 9,003.03 | 43,410.87 |
| 993 | 1,690.42 | 1,604.11 | 1,488.28 | 1,392.76 | 6,175.57 |
| 994 | 348.37 | 360 | 257.14 | 313.49 | 1,279 |
| TOTAL | 108,623.01 | 108,528.37 | 106,589.59 | 74,218.40 | 397,959.37 |

STATE: WEST VIRGINIA

| ZIP CODE | QUARTER 1 | QUARTER 2 | QUARTER 3 | QUARTER 4 | TOTAL GRAMS |
|---|---|---|---|---|---|
| 247 | 67.08 | 63.5 | 59.48 | 56.35 | 246.41 |
| 248 | 24.6 | 34.88 | 27.28 | 16.1 | 102.86 |
| 249 | 72 | 70.66 | 57.24 | 50.98 | 250.88 |
| 250 | 102.41 | 87.03 | 56.35 | 38.01 | 283.8 |
| 251 | 76.47 | 70.66 | 67.53 | 67.53 | 282.19 |
| 252 | 66.63 | 58.14 | 50.53 | 51.88 | 227.18 |
| 253 | 2,297.45 | 3,064.48 | 1,654.55 | 1,618.77 | 8,635.25 |
| 254 | 4,323.53 | 4,231.85 | 4,218.88 | 3,501.75 | 16,276.01 |
| 255 | 96.77 | 114.57 | 120.03 | 105.09 | 436.46 |
| 256 | 3,287.81 | 2,674.26 | 2,592.87 | 1,956.05 | 10,510.99 |
| 257 | 5,418.90 | 6,291.48 | 5,420.78 | 4,594.98 | 21,726.14 |
| 258 | 3,844.58 | 3,830.72 | 3,526.17 | 2,193.07 | 13,394.54 |
| 259 | 76.92 | 78.71 | 74.24 | 72.45 | 302.32 |
| 260 | 1,932.80 | 1,446.51 | 1,427.28 | 1,435.69 | 6,242.28 |
| 261 | 1,612.60 | 1,643.01 | 1,623.96 | 1,156.91 | 6,036.48 |
| 262 | 132.73 | 125.66 | 117.17 | 134.61 | 510.17 |
| 263 | 1,955.16 | 1,824.13 | 2,115.26 | 1,145.10 | 7,039.65 |
| 264 | 103.3 | 92.57 | 84.97 | 80.05 | 360.89 |
| 265 | 1,768.41 | 1,679.50 | 2,093.43 | 1,176.40 | 6,717.74 |
| 266 | 25.49 | 23.88 | 18.78 | 17.89 | 86.04 |
| 267 | 129.78 | 146.23 | 151.6 | 165.02 | 592.63 |
| 268 | 66.63 | 80.05 | 64.84 | 66.63 | 278.15 |
| TOTAL | 27,482.05 | 27,732.48 | 25,623.22 | 19,701.31 | 100,539.06 |

STATE: WISCONSIN

| ZIP CODE | QUARTER 1 | QUARTER 2 | QUARTER 3 | QUARTER 4 | TOTAL GRAMS |
|---|---|---|---|---|---|
| 530 | 3,862.73 | 4,038.66 | 3,974.27 | 2,790.53 | 14,666.19 |
| 531 | 6,001.87 | 5,602.43 | 5,370.42 | 3,799.86 | 20,774.58 |
| 532 | 20,000.53 | 19,447.21 | 17,899.85 | 11,429.05 | 68,776.64 |
| 534 | 3,630.10 | 3,141.04 | 3,333.61 | 1,894.25 | 11,999 |
| 535 | 4,755.08 | 4,551.78 | 4,153.86 | 2,873.26 | 16,333.98 |
| 537 | 9,900.83 | 10,148.40 | 10,484.34 | 6,517.58 | 37,051.15 |

DRUG CODE:9250BDRUG NAME:METHADONE
STATE:WISCONSIN

| ZIP CODE | QUARTER 1 | QUARTER 2 | QUARTER 3 | QUARTER 4 | TOTAL GRAMS |
|---|---|---|---|---|---|
| 538 | 161.44 | 144.89 | 132.37 | 114.57 | 553.27 |
| 539 | 1,175.69 | 1,104.49 | 940.91 | 745.93 | 3,967.02 |
| 540 | 332.27 | 353.82 | 340.59 | 273.15 | 1,299.83 |
| 541 | 352.39 | 380.57 | 311.25 | 299.18 | 1,343.39 |
| 542 | 201.24 | 176.2 | 138.18 | 132.82 | 648.44 |
| 543 | 4,240.98 | 4,769.48 | 4,905.96 | 3,185.58 | 17,102 |
| 544 | 2,325.62 | 2,486.61 | 2,581.69 | 2,144.95 | 9,538.87 |
| 545 | 118.51 | 101.96 | 94.81 | 58.58 | 373.86 |
| 546 | 3,477.96 | 3,507.11 | 3,447.82 | 3,424.93 | 13,857.82 |

RETAIL DRUG DISTRIBUTION BY ZIP CODE WITHIN STATE BY GRAMS WT

| | | | | | |
|---|---|---|---|---|---|
| 547 | 2,031.36 | 981.07 | 2,172.77 | 1,500.18 | 6,685.38 |
| 548 | 438.7 | 387.54 | 369.3 | 426.9 | 1,622.44 |
| 549 | 7,020.06 | 7,422.98 | 7,672.79 | 7,012.99 | 29,128.82 |
| TOTAL | 70,027.36 | 68,746.24 | 68,324.79 | 48,624.29 | 255,722.68 |

STATE: WYOMING

| ZIP CODE | QUARTER 1 | QUARTER 2 | QUARTER 3 | QUARTER 4 | TOTAL GRAMS |
|---|---|---|---|---|---|
| 820 | 308.57 | 289.79 | 295.15 | 268.77 | 1,162.28 |
| 822 | 54.56 | 64.84 | 59.48 | 57.69 | 236.57 |
| 823 | 11.18 | 17.89 | 16.99 | 12.97 | 59.03 |
| 824 | 114.48 | 113.59 | 62.61 | 69.76 | 360.44 |
| 825 | 127.63 | 132.64 | 125.31 | 134.52 | 520.1 |
| 826 | 361.78 | 275.92 | 312.15 | 295.15 | 1,245 |
| 827 | 51.43 | 52.77 | 45.17 | 46.06 | 195.43 |
| 828 | 94.81 | 89.44 | 77.81 | 105.54 | 367.6 |
| 829 | 309.46 | 308.12 | 294.26 | 246.41 | 1,158.25 |
| 830 | 59.48 | 50.98 | 54.56 | 35.78 | 200.8 |
| 831 | 68.87 | 69.76 | 76.47 | 67.88 | 282.98 |
| TOTAL | 1,562.25 | 1,465.74 | 1,419.96 | 1,340.53 | 5,788.48 |

DRUG CODE: 9300 DRUG NAME: MORPHINE

STATE: ALABAMA

| ZIP CODE | QUARTER 1 | QUARTER 2 | QUARTER 3 | QUARTER 4 | TOTAL GRAMS |
|---|---|---|---|---|---|
| 350 | 9,234.35 | 9,063.03 | 9,081.13 | 8,636.86 | 36,015.37 |
| 351 | 4,400.93 | 4,685.65 | 4,397.07 | 4,204.99 | 17,688.64 |
| 352 | 8,269.68 | 7,842.60 | 8,051.01 | 7,317.58 | 31,480.87 |
| 354 | 5,104.77 | 4,963.27 | 4,852.42 | 4,730.75 | 19,651.21 |
| 355 | 2,721.93 | 2,831.57 | 2,690.64 | 2,439.68 | 10,683.82 |
| 356 | 6,637.76 | 6,614.86 | 6,362.16 | 5,881.56 | 25,496.34 |
| 357 | 3,582.67 | 3,241.36 | 3,302.67 | 3,049.76 | 13,176.46 |
| 358 | 5,625.47 | 5,233.46 | 5,139.95 | 5,070.73 | 21,069.61 |
| 359 | 7,983.57 | 7,328.42 | 6,762.90 | 6,712.59 | 28,787.48 |
| 360 | 4,400.28 | 4,629.14 | 4,382.56 | 4,390.02 | 17,802 |
| 361 | 3,239.51 | 3,231.87 | 3,727.47 | 3,622.95 | 13,821.80 |
| 362 | 3,483.55 | 3,403.13 | 3,054.69 | 4,511.03 | 14,452.40 |
| 363 | 2,116.42 | 2,481.98 | 2,414.27 | 2,360.65 | 9,373.32 |
| 364 | 1,386.67 | 1,317.04 | 1,367.38 | 1,176.76 | 5,247.85 |
| 365 | 7,348.10 | 7,267.65 | 7,400 | 6,907.53 | 28,923.28 |
| 366 | 6,468.84 | 6,119.04 | 5,700.89 | 5,075.21 | 23,363.98 |
| 367 | 1,149.45 | 1,122.48 | 1,211.07 | 1,130.40 | 4,613.40 |
| 368 | 4,213.68 | 4,155.54 | 4,129.80 | 3,962.67 | 16,461.69 |
| 369 | 192.35 | 189.89 | 164.27 | 213.88 | 760.39 |
| TOTAL | 87,559.98 | 85,721.98 | 84,192.35 | 81,395.60 | 338,869.91 |

STATE: ALASKA

| ZIP CODE | QUARTER 1 | QUARTER 2 | QUARTER 3 | QUARTER 4 | TOTAL GRAMS |
|---|---|---|---|---|---|
| 995 | 6,686.39 | 6,144.57 | 5,965.72 | 5,450.11 | 24,246.79 |
| 996 | 5,168.20 | 4,857.60 | 4,578.81 | 4,443.27 | 19,047.88 |
| 997 | 1,920.64 | 1,718.38 | 1,678.25 | 1,602.44 | 6,919.71 |
| 998 | 548.46 | 573.8 | 452.6 | 511.99 | 2,086.85 |
| 999 | 251.99 | 320.79 | 280.97 | 253.26 | 1,107.01 |
| TOTAL | 14,575.68 | 13,615.14 | 12,956.35 | 12,261.07 | 53,408.24 |

STATE: AMERICAN SAMOA

| ZIP CODE | QUARTER 1 | QUARTER 2 | QUARTER 3 | QUARTER 4 | TOTAL GRAMS |
|---|---|---|---|---|---|
| 967 | 30.46 | 41.36 | 62.79 | 14.29 | 148.9 |
| TOTAL | 30.46 | 41.36 | 62.79 | 14.29 | 148.9 |

STATE: ARIZONA

| ZIP CODE | QUARTER 1 | QUARTER 2 | QUARTER 3 | QUARTER 4 | TOTAL GRAMS |
|---|---|---|---|---|---|
| 850 | 40,863.12 | 37,870.80 | 35,461.81 | 34,846.76 | 149,042.49 |

| | | | | | |
|---|---|---|---|---|---|
| 851 | 7,489.92 | 7,146.93 | 6,587.75 | 6,631.54 | 27,856.14 |

DRUG CODE:9300DRUG NAME:MORPHINE
STATE:ARIZONA

| ZIP CODE | QUARTER 1 | QUARTER 2 | QUARTER 3 | QUARTER 4 | TOTAL GRAMS |
|---|---|---|---|---|---|
| 852 | 42,908.03 | 40,894.82 | 39,226.21 | 37,205.45 | 160,234.51 |
| 853 | 30,357.82 | 29,382.70 | 28,302.34 | 27,918.96 | 115,961.82 |
| 855 | 5,596.95 | 5,191.80 | 5,155.31 | 4,772 | 20,716.06 |
| 856 | 8,967.53 | 7,951.64 | 8,204.38 | 7,399.50 | 32,523.05 |
| 857 | 44,532.35 | 41,495.99 | 43,233.07 | 41,042.91 | 170,304.32 |
| 859 | 3,707.57 | 3,894.49 | 4,077.57 | 3,875.41 | 15,555.04 |
| 860 | 4,329.68 | 4,163.52 | 4,309.47 | 4,221.40 | 17,024.07 |
| 863 | 10,056.99 | 10,041.96 | 10,038.06 | 9,101.21 | 39,238.22 |
| 864 | 13,451.41 | 12,487.14 | 12,076.90 | 12,217.38 | 50,232.83 |
| 865 | 468.79 | 454.69 | 421.32 | 413.45 | 1,758.25 |
| TOTAL | 212,730.16 | 200,976.48 | 197,094.19 | 189,645.97 | 800,446.80 |

STATE: ARKANSAS

| ZIP CODE | QUARTER 1 | QUARTER 2 | QUARTER 3 | QUARTER 4 | TOTAL GRAMS |
|---|---|---|---|---|---|
| 716 | 1,995.99 | 1,990.59 | 1,768.98 | 1,795.39 | 7,550.95 |
| 717 | 1,471.76 | 1,328.16 | 1,580.70 | 1,519.90 | 5,900.52 |
| 718 | 1,469.12 | 1,415.96 | 1,544.59 | 1,297.64 | 5,727.31 |
| 719 | 3,404.56 | 3,394.41 | 3,192.77 | 2,950.99 | 12,942.73 |
| 720 | 5,420.42 | 5,465.59 | 5,638.70 | 5,500.01 | 22,024.72 |
| 721 | 7,141.63 | 6,742.33 | 6,705.50 | 6,389.81 | 26,979.27 |
| 722 | 3,800.70 | 3,714.08 | 3,541.60 | 3,043.42 | 14,099.80 |
| 723 | 1,552.90 | 1,299.81 | 1,439.85 | 1,313.94 | 5,606.50 |
| 724 | 3,408.30 | 3,448.31 | 3,597.58 | 3,547.56 | 14,001.75 |
| 725 | 2,052.26 | 2,032.63 | 2,188.41 | 2,167.26 | 8,440.56 |
| 726 | 3,702.39 | 3,453.58 | 3,358.86 | 3,356.06 | 13,870.89 |
| 727 | 10,568.52 | 10,300.37 | 10,468.86 | 9,633.05 | 40,970.80 |
| 728 | 2,244.36 | 1,906.29 | 2,203.76 | 2,121.58 | 8,475.99 |
| 729 | 5,057.67 | 4,944.59 | 4,989.14 | 4,816.09 | 19,807.49 |
| TOTAL | 53,290.58 | 51,436.70 | 52,219.30 | 49,452.70 | 206,399.28 |

STATE: CALIFORNIA

| ZIP CODE | QUARTER 1 | QUARTER 2 | QUARTER 3 | QUARTER 4 | TOTAL GRAMS |
|---|---|---|---|---|---|
| 900 | 9,779.15 | 9,036.64 | 9,230.61 | 8,495.04 | 36,541.44 |
| 902 | 6,618.15 | 6,571.49 | 5,928.25 | 5,577.80 | 24,695.69 |
| 903 | 276.74 | 273.2 | 336.3 | 316.67 | 1,202.91 |
| 904 | 1,494.24 | 1,374.21 | 1,321.54 | 1,547.19 | 5,737.18 |
| 905 | 2,716.94 | 2,362.90 | 2,634.58 | 2,947.34 | 10,661.76 |
| 906 | 6,230.90 | 6,231.17 | 5,856.60 | 5,556.65 | 23,875.32 |
| 907 | 5,243.67 | 5,345.20 | 5,074.43 | 4,533.42 | 20,196.72 |
| 908 | 7,036.27 | 5,820.69 | 5,821.15 | 5,656.26 | 24,334.37 |
| 910 | 2,904.02 | 3,116.75 | 3,071.74 | 2,843.72 | 11,936.23 |
| 911 | 1,693.87 | 1,736.05 | 1,633.88 | 1,489.79 | 6,553.59 |
| 912 | 2,174.09 | 2,375.95 | 2,540.48 | 2,676.04 | 9,766.56 |
| 913 | 13,890.18 | 13,093.37 | 13,765.46 | 13,072.32 | 53,821.33 |
| 914 | 4,837.95 | 4,613.17 | 4,625.26 | 4,318.45 | 18,394.83 |
| 915 | 1,066.70 | 1,066.92 | 1,194.29 | 1,071.61 | 4,399.52 |
| 916 | 876.12 | 832.5 | 785 | 815.63 | 3,309.25 |
| 917 | 18,706.96 | 18,454.50 | 17,473.24 | 18,350.70 | 72,985.40 |
| 918 | 311.64 | 319.63 | 154.66 | 189.28 | 975.21 |
| 919 | 6,300.23 | 6,264.80 | 6,028.53 | 5,338.67 | 23,932.23 |
| 920 | 28,956.08 | 26,734.49 | 26,294.33 | 24,995.18 | 106,980.08 |
| 921 | 19,552.51 | 18,008.66 | 17,044.72 | 15,229.13 | 69,835.02 |
| 922 | 11,064.35 | 10,292.83 | 10,797.43 | 10,537.73 | 42,692.34 |
| 923 | 23,477.06 | 22,642.83 | 21,253.87 | 21,583.88 | 88,957.64 |
| 924 | 2,809.67 | 2,586.33 | 2,613.81 | 2,516.45 | 10,526.26 |

| | | | | | |
|---|---|---|---|---|---|
| 925 | 16,561.61 | 16,871 | 15,664.98 | 15,327.25 | 64,424.84 |
| 926 | 13,475.85 | 12,946.78 | 12,195.77 | 12,199.65 | 50,818.05 |
| 927 | 3,951.07 | 4,070.92 | 3,719.53 | 3,368.02 | 15,109.54 |
| 928 | 15,438.58 | 14,758.32 | 13,614.84 | 13,415.53 | 57,227.27 |
| 930 | 9,327.95 | 9,262.27 | 8,808.90 | 8,813.36 | 36,212.48 |
| 931 | 1,364.33 | 1,207.18 | 1,158.76 | 1,182.78 | 4,913.05 |
| 932 | 7,295.67 | 7,390.29 | 7,229.73 | 7,202.14 | 29,117.83 |
| 933 | 10,403.82 | 10,216.37 | 9,981.87 | 9,740.91 | 40,342.97 |
| 934 | 10,220.80 | 10,118.69 | 9,483.40 | 9,794.67 | 39,617.56 |
| 935 | 11,640.34 | 11,689.55 | 11,234.44 | 10,656.82 | 45,221.15 |
| 936 | 8,223.08 | 8,620.12 | 8,205.01 | 8,000.57 | 33,048.78 |
| 937 | 11,034.24 | 10,388.82 | 10,246.02 | 9,722.78 | 41,391.86 |
| 939 | 5,795.86 | 5,382.55 | 4,765.32 | 4,376.87 | 20,320.60 |

DRUG CODE:9300DRUG NAME:MORPHINE
STATE:CALIFORNIA

| ZIP CODE | QUARTER 1 | QUARTER 2 | QUARTER 3 | QUARTER 4 | TOTAL GRAMS |
|---|---|---|---|---|---|
| 940 | 8,980.80 | 8,091.17 | 7,998.60 | 7,857.46 | 32,928.03 |
| 941 | 17,738 | 16,763.90 | 16,074.86 | 15,333.26 | 65,910.02 |
| 943 | 2,324.48 | 1,957.71 | 2,411.20 | 1,998.41 | 8,691.80 |
| 944 | 1,270.79 | 1,312.30 | 1,183.13 | 1,253.18 | 5,019.40 |
| 945 | 44,531.17 | 42,894.84 | 41,543.68 | 40,299.49 | 169,269.18 |
| 946 | 5,468.83 | 5,456.19 | 4,679.38 | 4,723.63 | 20,328.03 |
| 947 | 966.62 | 890.95 | 1,015.06 | 811.34 | 3,683.97 |
| 948 | 1,861.58 | 1,507.98 | 1,337.84 | 1,373.86 | 6,081.26 |
| 949 | 7,068.96 | 7,006.56 | 6,953.94 | 6,850.50 | 27,879.96 |
| 950 | 7,696.38 | 7,131.47 | 7,204.33 | 7,015.82 | 29,048 |
| 951 | 3,957.85 | 3,944.47 | 4,106.21 | 3,654.02 | 15,662.55 |
| 952 | 10,522.02 | 9,099.83 | 9,272.49 | 9,198.21 | 38,092.55 |
| 953 | 25,766.61 | 24,975.74 | 23,728.20 | 24,864.99 | 99,335.54 |
| 954 | 17,018 | 15,171.30 | 15,877.54 | 15,531.42 | 63,598.26 |
| 955 | 6,846.91 | 6,591.99 | 6,253.86 | 5,920.89 | 25,613.65 |
| 956 | 31,848.56 | 31,985.95 | 30,315.43 | 28,154.79 | 122,304.73 |
| 957 | 3,968.74 | 3,765.76 | 3,726.67 | 3,252.46 | 14,713.63 |
| 958 | 17,191.42 | 17,385.04 | 16,046.30 | 16,223.12 | 66,845.88 |
| 959 | 15,553.18 | 14,807.91 | 13,853.91 | 13,636.63 | 57,851.63 |
| 960 | 11,176.53 | 10,766.22 | 10,387.89 | 9,923.64 | 42,254.28 |
| 961 | 2,351.11 | 2,221.37 | 2,140.65 | 2,139.40 | 8,852.53 |
| TOTAL | 546,859.23 | 525,805.79 | 507,899.90 | 493,476.82 | 2,074,041.74 |

STATE: COLORADO

| ZIP CODE | QUARTER 1 | QUARTER 2 | QUARTER 3 | QUARTER 4 | TOTAL GRAMS |
|---|---|---|---|---|---|
| 800 | 16,079.60 | 15,244.20 | 14,873.74 | 14,076.64 | 60,274.18 |
| 801 | 11,303.47 | 11,433.30 | 10,602.20 | 9,466.24 | 42,805.21 |
| 802 | 19,866.14 | 18,524.74 | 18,115.55 | 17,304.33 | 73,810.76 |
| 803 | 1,541.45 | 1,374.40 | 1,318.22 | 1,382.33 | 5,616.40 |
| 804 | 3,528.40 | 3,543.78 | 3,605.78 | 3,344.11 | 14,022.07 |
| 805 | 10,250.39 | 9,828.24 | 9,172.20 | 8,757.62 | 38,008.45 |
| 806 | 4,133.67 | 4,295.66 | 3,649.04 | 3,694.11 | 15,772.48 |
| 807 | 1,499.80 | 1,419.22 | 1,300.42 | 1,223.38 | 5,442.82 |
| 808 | 2,150.73 | 2,121.80 | 2,059.04 | 1,949.27 | 8,280.84 |
| 809 | 10,321.30 | 9,966.22 | 9,890.90 | 10,105.04 | 40,283.46 |
| 810 | 6,208.21 | 5,357.99 | 5,321 | 5,107.71 | 21,994.91 |
| 811 | 765.81 | 746.32 | 708.14 | 750.95 | 2,971.22 |
| 812 | 1,690.51 | 1,470.76 | 1,649.88 | 1,374.01 | 6,185.16 |
| 813 | 886.84 | 772.02 | 773.81 | 754.98 | 3,187.65 |
| 814 | 842.01 | 896.3 | 812.66 | 705.18 | 3,256.15 |
| 815 | 2,957.40 | 2,893.89 | 2,726.27 | 2,510.26 | 11,087.82 |
| 816 | 1,415.49 | 1,378.72 | 1,289.98 | 1,283.80 | 5,367.99 |
| TOTAL | 95,441.22 | 91,267.56 | 87,868.83 | 83,789.96 | 358,367.57 |

STATE: CONNECTICUT

| ZIP CODE | QUARTER 1 | QUARTER 2 | QUARTER 3 | QUARTER 4 | TOTAL GRAMS |
|---|---|---|---|---|---|
| 60 | 7,907.80 | 7,632 | 7,500.06 | 8,176.75 | 31,216.61 |
| 61 | 2,323.31 | 2,111.94 | 2,068.06 | 1,895.10 | 8,398.41 |
| 62 | 1,979.58 | 1,929.50 | 1,946.49 | 1,794.99 | 7,650.56 |
| 63 | 2,967.80 | 2,740.56 | 2,720.53 | 2,817.18 | 11,246.07 |
| 64 | 12,430.52 | 11,755.79 | 11,626.53 | 11,135.54 | 46,948.38 |
| 65 | 4,687.98 | 4,541.67 | 4,540.49 | 4,316.58 | 18,086.72 |
| 66 | 1,981.48 | 1,648.80 | 1,619.29 | 1,727.94 | 6,977.51 |
| 67 | 3,941.92 | 3,691.09 | 3,206.13 | 3,214.23 | 14,053.37 |
| 68 | 4,517.19 | 4,823.04 | 4,685.84 | 4,257.03 | 18,283.10 |
| 69 | 424.58 | 448.43 | 373.05 | 337.93 | 1,583.99 |
| TOTAL | 43,162.16 | 41,322.82 | 40,286.47 | 39,673.27 | 164,444.72 |

STATE: DELAWARE

| ZIP CODE | QUARTER 1 | QUARTER 2 | QUARTER 3 | QUARTER 4 | TOTAL GRAMS |
|---|---|---|---|---|---|
| 197 | 8,466.30 | 8,400.36 | 8,213.62 | 8,449.01 | 33,529.29 |
| 198 | 5,394.63 | 5,491.45 | 5,000 | 5,152.95 | 21,039.03 |
| 199 | 9,411.15 | 9,298.07 | 9,231.44 | 8,890.65 | 36,831.31 |
| TOTAL | 23,272.08 | 23,189.88 | 22,445.06 | 22,492.61 | 91,399.63 |

STATE: DISTRICT OF COLUMBIA

| ZIP CODE | QUARTER 1 | QUARTER 2 | QUARTER 3 | QUARTER 4 | TOTAL GRAMS |
|---|---|---|---|---|---|
| 200 | 3,314.22 | 2,869.61 | 3,082.68 | 2,957.25 | 12,223.76 |
| 203 | 22.61 | 0.12 | 6.77 | 22.56 | 52.06 |
| 204 | 1,470.01 | 1,151.24 | 1,540.89 | 1,071.41 | 5,233.55 |

DRUG CODE:9300DRUG NAME:MORPHINE
STATE:DISTRICT OF COLUMBIA

| ZIP CODE | QUARTER 1 | QUARTER 2 | QUARTER 3 | QUARTER 4 | TOTAL GRAMS |
|---|---|---|---|---|---|
| 205 | 0 | 0.56 | 0.38 | 0 | 0.94 |
| TOTAL | 4,806.84 | 4,021.53 | 4,630.72 | 4,051.22 | 17,510.31 |

STATE: FLORIDA

| ZIP CODE | QUARTER 1 | QUARTER 2 | QUARTER 3 | QUARTER 4 | TOTAL GRAMS |
|---|---|---|---|---|---|
| 320 | 14,134.49 | 13,932.21 | 13,572.62 | 13,357.76 | 54,997.08 |
| 321 | 17,468.54 | 17,123.91 | 17,050.27 | 16,402.58 | 68,045.30 |
| 322 | 27,313 | 25,641.29 | 25,200.09 | 24,451.79 | 102,606.17 |
| 323 | 4,715.55 | 4,748.61 | 4,900.85 | 4,755.84 | 19,120.85 |
| 324 | 4,733.41 | 4,966.14 | 4,722.70 | 4,644.18 | 19,066.43 |
| 325 | 10,719.44 | 9,753.36 | 9,310.61 | 9,426.64 | 39,210.05 |
| 326 | 11,845.97 | 11,129.81 | 10,598.18 | 10,106.15 | 43,680.11 |
| 327 | 22,522.98 | 21,792.43 | 21,734.16 | 21,478.20 | 87,527.77 |
| 328 | 14,804.39 | 13,626.49 | 13,220.91 | 12,298.30 | 53,950.09 |
| 329 | 22,437.95 | 22,673.87 | 21,517.38 | 20,047.95 | 86,677.15 |
| 330 | 26,076.45 | 24,168.61 | 24,272.20 | 22,744.60 | 97,261.86 |
| 331 | 9,917.56 | 9,603.93 | 10,065.07 | 9,222.60 | 38,809.16 |
| 333 | 17,443.43 | 17,995.13 | 17,939.89 | 17,311.94 | 70,690.39 |
| 334 | 32,279.78 | 31,448.90 | 29,423.65 | 29,598.78 | 122,751.11 |
| 335 | 17,992.22 | 17,193.63 | 17,030.48 | 17,308.18 | 69,524.51 |
| 336 | 30,847.49 | 29,612.63 | 28,182.96 | 28,528.40 | 117,171.48 |
| 337 | 30,376.18 | 29,025.88 | 28,453.87 | 28,216.11 | 116,072.04 |
| 338 | 9,412.69 | 9,809.42 | 9,151.31 | 9,136.17 | 37,509.59 |
| 339 | 24,325.20 | 22,951.82 | 23,381.88 | 23,064.32 | 93,723.22 |
| 341 | 7,600.68 | 7,233.05 | 7,157.65 | 6,938.21 | 28,929.59 |
| 342 | 23,315.22 | 22,227.49 | 21,945.50 | 22,253.89 | 89,742.10 |
| 344 | 14,660 | 14,631.83 | 14,501.44 | 14,759.85 | 58,553.12 |
| 346 | 30,350.71 | 30,039.53 | 29,448.53 | 29,319.43 | 119,158.20 |
| 347 | 12,212.02 | 11,696.16 | 11,047.86 | 11,122.08 | 46,078.12 |

| | | | | | |
|---|---|---|---|---|---|
| 349 | 15,053.42 | 14,351.56 | 14,364.76 | 14,027.78 | 57,797.52 |
| TOTAL | 452,558.77 | 437,377.69 | 428,194.82 | 420,521.73 | 1,738,653.01 |

STATE: GEORGIA

| ZIP CODE | QUARTER 1 | QUARTER 2 | QUARTER 3 | QUARTER 4 | TOTAL GRAMS |
|---|---|---|---|---|---|
| 300 | 25,160.35 | 24,829.71 | 24,301.15 | 23,766.99 | 98,058.20 |
| 301 | 18,185.84 | 17,270.06 | 17,080.23 | 17,064.63 | 69,600.76 |
| 302 | 11,478.97 | 10,716.71 | 11,018.11 | 10,797.25 | 44,011.04 |
| 303 | 6,419.98 | 6,593.60 | 6,643.25 | 6,639.55 | 26,296.38 |
| 304 | 2,756.05 | 2,611.84 | 2,857.03 | 2,875.69 | 11,100.61 |
| 305 | 8,536.29 | 8,572.75 | 8,709.82 | 8,846.12 | 34,664.98 |
| 306 | 4,657.92 | 4,482.26 | 4,600.69 | 4,653.40 | 18,394.27 |
| 307 | 4,356.46 | 4,291.54 | 4,382.56 | 4,159.32 | 17,189.88 |
| 308 | 2,236.61 | 2,308.50 | 2,292.63 | 2,192.04 | 9,029.78 |
| 309 | 6,223.91 | 5,435.30 | 5,406.96 | 5,069.28 | 22,135.45 |
| 310 | 8,107.44 | 7,549.85 | 7,508.95 | 7,721.79 | 30,888.03 |
| 312 | 3,200.31 | 2,985.53 | 2,888.82 | 2,798.98 | 11,873.64 |
| 313 | 2,850.45 | 2,879.32 | 2,804.52 | 2,955.67 | 11,489.96 |
| 314 | 3,069.30 | 2,911.98 | 2,847.57 | 2,754.92 | 11,583.77 |
| 315 | 4,957.64 | 4,783.15 | 4,743.66 | 4,746.23 | 19,230.68 |
| 316 | 2,567.39 | 2,689.38 | 3,020.61 | 3,246.94 | 11,524.32 |
| 317 | 3,361.22 | 3,295.96 | 3,257.43 | 3,659.52 | 13,574.13 |
| 318 | 470.86 | 431.57 | 487.3 | 529.3 | 1,919.03 |
| 319 | 4,213.11 | 4,463.48 | 4,550.74 | 3,892.77 | 17,120.10 |
| 398 | 992.28 | 880.06 | 990.91 | 1,062.88 | 3,926.13 |
| TOTAL | 123,802.38 | 119,982.55 | 120,392.94 | 119,433.27 | 483,611.14 |

STATE: GUAM

| ZIP CODE | QUARTER 1 | QUARTER 2 | QUARTER 3 | QUARTER 4 | TOTAL GRAMS |
|---|---|---|---|---|---|
| 969 | 1,003.37 | 1,290.67 | 1,276.39 | 1,225.44 | 4,795.87 |
| TOTAL | 1,003.37 | 1,290.67 | 1,276.39 | 1,225.44 | 4,795.87 |

STATE: HAWAII

| ZIP CODE | QUARTER 1 | QUARTER 2 | QUARTER 3 | QUARTER 4 | TOTAL GRAMS |
|---|---|---|---|---|---|
| 967 | 18,175.27 | 17,305.48 | 16,670.78 | 16,378.20 | 68,529.73 |
| 968 | 8,867.89 | 8,840.72 | 8,896.34 | 8,407.41 | 35,012.36 |
| TOTAL | 27,043.16 | 26,146.20 | 25,567.12 | 24,785.61 | 103,542.09 |

STATE: IDAHO

| ZIP CODE | QUARTER 1 | QUARTER 2 | QUARTER 3 | QUARTER 4 | TOTAL GRAMS |
|---|---|---|---|---|---|
| 832 | 3,493.38 | 3,153.05 | 3,141.45 | 3,093.84 | 12,881.72 |
| 833 | 3,798.48 | 3,546.12 | 3,640.30 | 3,500.25 | 14,485.15 |

DRUG CODE:9300DRUG NAME:MORPHINE
STATE:IDAHO

| ZIP CODE | QUARTER 1 | QUARTER 2 | QUARTER 3 | QUARTER 4 | TOTAL GRAMS |
|---|---|---|---|---|---|
| 834 | 2,291.70 | 2,221.26 | 2,347.38 | 2,216.24 | 9,076.58 |
| 835 | 2,297.76 | 2,019.63 | 2,073.91 | 1,899.27 | 8,290.57 |
| 836 | 8,129.46 | 8,569.59 | 7,658.66 | 7,753.08 | 32,110.79 |
| 837 | 6,835.36 | 7,045.47 | 6,558.51 | 6,361.55 | 26,800.89 |
| 838 | 5,548.30 | 5,487.76 | 5,195.95 | 5,215.09 | 21,447.10 |
| TOTAL | 32,394.44 | 32,042.88 | 30,616.16 | 30,039.32 | 125,092.80 |

STATE: ILLINOIS

| ZIP CODE | QUARTER 1 | QUARTER 2 | QUARTER 3 | QUARTER 4 | TOTAL GRAMS |
|---|---|---|---|---|---|
| 600 | 19,345.72 | 19,756.34 | 19,376.79 | 18,231.90 | 76,710.75 |
| 601 | 11,027.16 | 10,942.32 | 11,216.73 | 11,057.46 | 44,243.67 |
| 602 | 527.04 | 476.93 | 456.49 | 336.31 | 1,796.77 |

| | | | | | |
|---|---|---|---|---|---|
| 603 | 586.02 | 525.83 | 647.96 | 575.16 | 2,334.97 |
| 604 | 12,815.50 | 12,613.33 | 12,465.16 | 12,156.03 | 50,050.02 |
| 605 | 6,219.79 | 6,141.96 | 6,454.94 | 5,940.74 | 24,757.43 |
| 606 | 12,503.57 | 12,357.62 | 12,052.08 | 11,540.18 | 48,453.45 |
| 607 | 676.44 | 656.08 | 686.66 | 649.68 | 2,668.86 |
| 608 | 928.05 | 794.4 | 750.25 | 1,045.53 | 3,518.23 |
| 609 | 1,643.32 | 1,905.40 | 1,958.17 | 1,853.41 | 7,360.30 |
| 610 | 3,039.26 | 3,005.39 | 2,884.84 | 3,261.09 | 12,190.58 |
| 611 | 4,335.87 | 4,205.91 | 4,316.55 | 4,257.69 | 17,116.02 |
| 612 | 1,060.18 | 1,281.66 | 1,203.33 | 1,242.05 | 4,787.22 |
| 613 | 1,691.06 | 1,630.74 | 1,739.77 | 1,358.65 | 6,420.22 |
| 614 | 1,063.03 | 1,164.63 | 1,054.06 | 1,070.57 | 4,352.29 |
| 615 | 2,019.20 | 2,004.30 | 2,052.78 | 1,879.73 | 7,956.01 |
| 616 | 3,554.64 | 3,661.21 | 3,611.74 | 3,374.91 | 14,202.50 |
| 617 | 1,674.35 | 1,574.73 | 1,526.21 | 1,575.98 | 6,351.27 |
| 618 | 3,059.66 | 2,922.14 | 2,897.87 | 3,146.11 | 12,025.78 |
| 619 | 1,136.04 | 1,124.76 | 1,226.26 | 1,261.40 | 4,748.46 |
| 620 | 4,209.96 | 3,851.84 | 4,146.40 | 3,894.56 | 16,102.76 |
| 622 | 4,874.31 | 4,930.71 | 4,932.47 | 4,710.48 | 19,447.97 |
| 623 | 1,285.40 | 1,208.39 | 1,456.48 | 1,408.24 | 5,358.51 |
| 624 | 1,418.03 | 1,303.61 | 1,314.11 | 1,321.30 | 5,357.05 |
| 625 | 2,495.29 | 2,185.24 | 2,408.60 | 2,223.29 | 9,312.42 |
| 626 | 1,578 | 1,829.99 | 1,678.38 | 1,946.35 | 7,032.72 |
| 627 | 1,935.12 | 1,932.84 | 1,662.96 | 1,846.52 | 7,377.44 |
| 628 | 2,250.88 | 2,381.63 | 2,420.62 | 2,238.87 | 9,292 |
| 629 | 5,175.79 | 4,908.37 | 4,617.94 | 4,747.62 | 19,449.72 |
| TOTAL | 114,128.68 | 113,278.30 | 113,216.60 | 110,151.81 | 450,775.39 |

STATE: INDIANA

| ZIP CODE | QUARTER 1 | QUARTER 2 | QUARTER 3 | QUARTER 4 | TOTAL GRAMS |
|---|---|---|---|---|---|
| 460 | 7,083.99 | 6,678.66 | 6,309.97 | 5,893.33 | 25,965.95 |
| 461 | 8,963.76 | 8,840.03 | 8,749.11 | 8,058.91 | 34,611.81 |
| 462 | 19,288.17 | 18,955.81 | 17,420.60 | 19,221.78 | 74,886.36 |
| 463 | 9,957.14 | 9,224.02 | 8,783.81 | 8,065.18 | 36,030.15 |
| 464 | 994.49 | 834.89 | 798.42 | 751.7 | 3,379.50 |
| 465 | 7,538.77 | 7,226.10 | 7,137.62 | 6,865.13 | 28,767.62 |
| 466 | 2,466.94 | 2,398.76 | 2,695.48 | 2,492.64 | 10,053.82 |
| 467 | 4,703.92 | 3,632.06 | 3,746.80 | 3,538.59 | 15,621.37 |
| 468 | 7,149.59 | 5,768.17 | 5,564.36 | 4,946.52 | 23,428.64 |
| 469 | 5,912.97 | 5,569.92 | 5,571.36 | 4,742.23 | 21,796.48 |
| 470 | 1,963.79 | 1,741.54 | 2,080.08 | 1,902.43 | 7,687.84 |
| 471 | 6,884.55 | 6,314.44 | 6,166.92 | 5,537.52 | 24,903.43 |
| 472 | 3,273.49 | 3,188.96 | 2,927.19 | 2,784.62 | 12,174.26 |
| 473 | 6,889.02 | 6,336.13 | 6,289.58 | 6,068.26 | 25,582.99 |
| 474 | 2,528.64 | 2,243.87 | 2,434.17 | 2,181.24 | 9,387.92 |
| 475 | 2,130.44 | 1,951.56 | 1,978.40 | 2,073.89 | 8,134.29 |
| 476 | 1,347.44 | 1,374.14 | 1,288.04 | 1,276.38 | 5,286 |
| 477 | 4,474.21 | 4,596.03 | 4,567.54 | 3,817.28 | 17,455.06 |
| 478 | 2,780.40 | 2,701.23 | 2,404.27 | 2,341.55 | 10,227.45 |
| 479 | 3,708.11 | 4,007.82 | 3,530.45 | 3,559.82 | 14,806.20 |
| TOTAL | 110,039.83 | 103,584.14 | 100,444.17 | 96,119 | 410,187.14 |

STATE: IOWA

| ZIP CODE | QUARTER 1 | QUARTER 2 | QUARTER 3 | QUARTER 4 | TOTAL GRAMS |
|---|---|---|---|---|---|
| 500 | 2,713.57 | 2,956.21 | 2,912.80 | 2,529.16 | 11,111.74 |
| 501 | 2,584.28 | 2,625.84 | 2,576.88 | 2,495.54 | 10,282.54 |
| 502 | 2,653.73 | 2,772.72 | 2,708.46 | 2,424.55 | 10,559.46 |

DRUG CODE:9300DRUG NAME:MORPHINE
STATE:IOWA

RETAIL DRUG DISTRIBUTION BY ZIP CODE WITHIN STATE BY GRAMS WT

| ZIP CODE | QUARTER 1 | QUARTER 2 | QUARTER 3 | QUARTER 4 | TOTAL GRAMS |
|---|---|---|---|---|---|
| 503 | 7,080.13 | 6,372.03 | 6,059.56 | 5,973.80 | 25,485.52 |
| 504 | 1,515.87 | 1,543.82 | 1,488.96 | 1,554.62 | 6,103.27 |
| 505 | 2,017.48 | 1,812.19 | 1,703.59 | 1,780.84 | 7,314.10 |
| 506 | 1,361.85 | 1,243.93 | 1,364.17 | 1,089.17 | 5,059.12 |
| 507 | 916.99 | 876.31 | 970.5 | 892.51 | 3,656.31 |
| 508 | 547.84 | 541.49 | 502.73 | 448.63 | 2,040.69 |
| 510 | 658.53 | 700.99 | 658.03 | 564.74 | 2,582.29 |
| 511 | 1,017.46 | 1,089.87 | 1,128.53 | 1,034.40 | 4,270.26 |
| 512 | 371.23 | 216.4 | 280.94 | 293.19 | 1,161.76 |
| 513 | 440.81 | 454.75 | 476.88 | 375.16 | 1,747.60 |
| 514 | 664.27 | 599.46 | 655.01 | 629 | 2,547.74 |
| 515 | 1,712.15 | 1,525.52 | 1,365.02 | 1,286.93 | 5,889.62 |
| 516 | 243.79 | 219.24 | 167.94 | 223.95 | 854.92 |
| 520 | 1,644.08 | 1,521.25 | 1,506.68 | 1,467.88 | 6,139.89 |
| 521 | 401.71 | 427.71 | 460.92 | 419.05 | 1,709.39 |
| 522 | 2,447.64 | 2,572.47 | 2,328.80 | 2,366.76 | 9,715.67 |
| 523 | 997.01 | 910.05 | 881.22 | 826.22 | 3,614.50 |
| 524 | 1,572.48 | 1,536.38 | 1,440.81 | 1,485.01 | 6,034.68 |
| 525 | 1,982.73 | 1,953.24 | 1,914.21 | 1,736.94 | 7,587.12 |
| 526 | 1,373.18 | 1,362.53 | 1,404.69 | 1,275.06 | 5,415.46 |
| 527 | 1,270.41 | 1,221.14 | 1,256.83 | 1,298.28 | 5,046.66 |
| 528 | 2,076.31 | 1,685.78 | 1,816.97 | 1,736.11 | 7,315.17 |
| TOTAL | 40,265.53 | 38,741.32 | 38,031.13 | 36,207.50 | 153,245.48 |

STATE: KANSAS

| ZIP CODE | QUARTER 1 | QUARTER 2 | QUARTER 3 | QUARTER 4 | TOTAL GRAMS |
|---|---|---|---|---|---|
| 660 | 7,767.53 | 7,542.71 | 7,772.37 | 7,282.88 | 30,365.49 |
| 661 | 2,268.70 | 2,365.71 | 1,938.96 | 1,806.75 | 8,380.12 |
| 662 | 7,932.45 | 7,748.44 | 7,456.39 | 6,767.04 | 29,904.32 |
| 664 | 907.24 | 1,029.58 | 1,146.02 | 986.24 | 4,069.08 |
| 665 | 1,204.77 | 1,302.08 | 1,486.82 | 1,192.87 | 5,186.54 |
| 666 | 3,903.71 | 3,906.96 | 4,657.11 | 2,786.51 | 15,254.29 |
| 667 | 3,201.33 | 2,917.17 | 3,213.83 | 3,143.67 | 12,476 |
| 668 | 890.75 | 805.14 | 791.93 | 833.41 | 3,321.23 |
| 669 | 710.83 | 631.2 | 644.9 | 705.99 | 2,692.92 |
| 670 | 2,870.32 | 2,792.79 | 2,783.24 | 2,661.65 | 11,108 |
| 671 | 1,851.15 | 1,865.31 | 1,823.89 | 1,679.62 | 7,219.97 |
| 672 | 8,966.33 | 9,020.43 | 8,935.83 | 8,796.82 | 35,719.41 |
| 673 | 1,924.50 | 1,775.42 | 1,787.28 | 1,506.11 | 6,993.31 |
| 674 | 2,548.68 | 2,518.14 | 2,405.92 | 2,299.88 | 9,772.62 |
| 675 | 1,637.89 | 1,547.13 | 1,497.71 | 1,371.97 | 6,054.70 |
| 676 | 1,097.74 | 1,163.67 | 1,210.81 | 1,087.73 | 4,559.95 |
| 677 | 689.67 | 640.59 | 802.2 | 729.68 | 2,862.14 |
| 678 | 869.66 | 921.37 | 1,033.17 | 1,100.01 | 3,924.21 |
| 679 | 425.26 | 380.14 | 396.83 | 390.29 | 1,592.52 |
| TOTAL | 51,668.51 | 50,873.98 | 51,785.21 | 47,129.12 | 201,456.82 |

STATE: KENTUCKY

| ZIP CODE | QUARTER 1 | QUARTER 2 | QUARTER 3 | QUARTER 4 | TOTAL GRAMS |
|---|---|---|---|---|---|
| 400 | 1,951.78 | 1,939.05 | 1,719.15 | 1,767.75 | 7,377.73 |
| 401 | 1,452.33 | 1,439.57 | 1,410.50 | 1,341.50 | 5,643.90 |
| 402 | 8,934.97 | 8,769.71 | 8,770.89 | 7,773.35 | 34,248.92 |
| 403 | 3,797.55 | 3,638.24 | 3,409.09 | 3,522.40 | 14,367.28 |
| 404 | 1,986.66 | 1,887.61 | 1,713.42 | 1,679.33 | 7,267.02 |
| 405 | 4,702.86 | 4,518.65 | 4,763.35 | 4,037.05 | 18,021.91 |
| 406 | 304.31 | 348.98 | 300.14 | 303.45 | 1,256.88 |
| 407 | 1,437.85 | 1,531.19 | 1,539.15 | 1,553.14 | 6,061.33 |
| 408 | 361.96 | 334.9 | 374.48 | 369.65 | 1,440.99 |
| 409 | 791.7 | 761.35 | 699.72 | 682.13 | 2,934.90 |
| 410 | 6,682.87 | 6,218.73 | 6,410.85 | 6,090.95 | 25,403.40 |
| 411 | 1,416.53 | 1,301.19 | 1,420.75 | 1,351.49 | 5,489.96 |

| | | | | | |
|---|---|---|---|---|---|
| 412 | 369.72 | 384.24 | 317.16 | 290.09 | 1,361.21 |
| 413 | 446.78 | 366.43 | 430 | 359.25 | 1,602.46 |
| 414 | 289.44 | 228.08 | 280.14 | 194.92 | 992.58 |
| 415 | 620.32 | 532.95 | 509.68 | 545.46 | 2,208.41 |
| 416 | 528.93 | 518.03 | 371.42 | 347.83 | 1,766.21 |
| 417 | 1,402.58 | 1,209.97 | 1,451.31 | 1,207.13 | 5,270.99 |
| 418 | 199.36 | 179.65 | 164.07 | 153.28 | 696.36 |

DRUG CODE:9300DRUG NAME:MORPHINE
STATE:KENTUCKY

| ZIP CODE | QUARTER 1 | QUARTER 2 | QUARTER 3 | QUARTER 4 | TOTAL GRAMS |
|---|---|---|---|---|---|
| 420 | 2,653.12 | 2,838.68 | 2,990.69 | 2,894.40 | 11,376.89 |
| 421 | 3,812.81 | 3,736.46 | 3,752.72 | 3,467.52 | 14,769.51 |
| 422 | 1,870.09 | 1,788.93 | 1,771.77 | 1,690.42 | 7,121.21 |
| 423 | 2,371.26 | 2,177.87 | 2,433.61 | 2,507.40 | 9,490.14 |
| 424 | 2,701.71 | 2,719.59 | 2,447.27 | 2,482.77 | 10,351.34 |
| 425 | 952.06 | 831.9 | 906.45 | 861.63 | 3,552.04 |
| 426 | 829.61 | 693.52 | 697.34 | 725.55 | 2,946.02 |
| 427 | 2,741.71 | 2,461.29 | 2,395.42 | 2,348.13 | 9,946.55 |
| TOTAL | 55,610.87 | 53,356.76 | 53,450.54 | 50,547.97 | 212,966.14 |

STATE: LOUISIANA

| ZIP CODE | QUARTER 1 | QUARTER 2 | QUARTER 3 | QUARTER 4 | TOTAL GRAMS |
|---|---|---|---|---|---|
| 700 | 9,922.68 | 9,138.31 | 8,781.20 | 8,090.07 | 35,932.26 |
| 701 | 4,585.70 | 4,326.16 | 4,172.96 | 3,808.85 | 16,893.67 |
| 703 | 2,259.96 | 2,193.33 | 1,870.13 | 2,271.45 | 8,594.87 |
| 704 | 8,255.48 | 8,034.90 | 8,018.06 | 7,255.18 | 31,563.62 |
| 705 | 6,700.97 | 6,665.10 | 6,380 | 6,178.57 | 25,924.64 |
| 706 | 2,010.99 | 1,964.71 | 1,675.71 | 1,599.32 | 7,250.73 |
| 707 | 7,369.68 | 7,221.89 | 7,248.58 | 6,192.03 | 28,032.18 |
| 708 | 5,428.93 | 6,153.80 | 5,834.33 | 5,257.45 | 22,674.51 |
| 710 | 1,993.24 | 1,899.35 | 1,771.65 | 1,754.22 | 7,418.46 |
| 711 | 6,833.60 | 6,489.54 | 6,354.83 | 6,431.47 | 26,109.44 |
| 712 | 2,145.29 | 2,192.07 | 2,040.98 | 2,101.31 | 8,479.65 |
| 713 | 5,991.49 | 5,675.22 | 6,058.10 | 5,257.50 | 22,982.31 |
| 714 | 2,501.73 | 2,482.78 | 2,465.41 | 2,431.90 | 9,881.82 |
| TOTAL | 65,999.74 | 64,437.16 | 62,671.94 | 58,629.32 | 251,738.16 |

STATE: MAINE

| ZIP CODE | QUARTER 1 | QUARTER 2 | QUARTER 3 | QUARTER 4 | TOTAL GRAMS |
|---|---|---|---|---|---|
| 39 | 450.15 | 463.11 | 403.88 | 325.15 | 1,642.29 |
| 40 | 5,961.47 | 5,988.96 | 5,297.57 | 4,789.75 | 22,037.75 |
| 41 | 1,811.05 | 1,408.28 | 1,183.18 | 1,164.85 | 5,567.36 |
| 42 | 2,721.49 | 2,808.70 | 2,565.85 | 2,090.56 | 10,186.60 |
| 43 | 3,875.28 | 4,058.32 | 3,708.80 | 3,303.79 | 14,946.19 |
| 44 | 1,974.45 | 2,096.98 | 1,988.41 | 1,897.01 | 7,956.85 |
| 45 | 595.17 | 489.74 | 463.94 | 449.97 | 1,998.82 |
| 46 | 994.25 | 942.92 | 966.24 | 887.02 | 3,790.43 |
| 47 | 890.93 | 926.98 | 756.31 | 633 | 3,207.22 |
| 48 | 761.36 | 802.46 | 814.23 | 716.86 | 3,094.91 |
| 49 | 3,911.19 | 3,809.03 | 3,633.07 | 3,197.04 | 14,550.33 |
| TOTAL | 23,946.79 | 23,795.48 | 21,781.48 | 19,455 | 88,978.75 |

STATE: MARYLAND

| ZIP CODE | QUARTER 1 | QUARTER 2 | QUARTER 3 | QUARTER 4 | TOTAL GRAMS |
|---|---|---|---|---|---|
| 206 | 6,764.65 | 6,496.02 | 6,341.13 | 6,183.25 | 25,785.05 |
| 207 | 6,725.76 | 7,164.88 | 6,856.17 | 6,749.63 | 27,496.44 |
| 208 | 6,587.09 | 6,489.68 | 6,824.85 | 6,633.76 | 26,535.38 |
| 209 | 1,268.04 | 1,149.77 | 1,236.95 | 1,102.66 | 4,757.42 |
| 210 | 15,241.04 | 15,479.92 | 14,427.83 | 14,401.14 | 59,549.93 |

| | | | | | |
|---|---|---|---|---|---|
| 211 | 7,400.22 | 7,236.77 | 7,172.95 | 6,612.11 | 28,422.05 |
| 212 | 22,647.86 | 22,743 | 22,141.52 | 21,522.02 | 89,054.40 |
| 214 | 1,446.73 | 1,427.68 | 1,504.89 | 1,257.14 | 5,636.44 |
| 215 | 2,812.17 | 2,660.33 | 2,626.53 | 2,370.90 | 10,469.93 |
| 216 | 2,957.31 | 2,829.52 | 2,749.78 | 2,821.42 | 11,358.03 |
| 217 | 9,579.12 | 9,293.69 | 9,318.57 | 9,285.23 | 37,476.61 |
| 218 | 2,944.23 | 2,761.16 | 2,772.79 | 2,668.82 | 11,147 |
| 219 | 4,755.38 | 4,466.03 | 4,391.60 | 4,052.52 | 17,665.53 |
| TOTAL | 91,129.60 | 90,198.45 | 88,365.56 | 85,660.60 | 355,354.21 |

STATE: MASSACHUSETTS

| ZIP CODE | QUARTER 1 | QUARTER 2 | QUARTER 3 | QUARTER 4 | TOTAL GRAMS |
|---|---|---|---|---|---|
| 10 | 9,472.59 | 8,835.97 | 8,595.85 | 8,216.88 | 35,121.29 |
| 11 | 3,954.58 | 3,859.61 | 3,456.99 | 3,414.58 | 14,685.76 |
| 12 | 1,497.96 | 1,440.92 | 1,238.89 | 1,279.74 | 5,457.51 |
| 13 | 1,775.18 | 1,717.43 | 1,576.77 | 1,625.28 | 6,694.66 |
| 14 | 2,810.77 | 2,883.67 | 2,587.37 | 2,713.59 | 10,995.40 |
| 15 | 3,947.88 | 3,875.45 | 3,639.42 | 3,552.47 | 15,015.22 |
| 16 | 2,607.99 | 2,597.99 | 2,631.69 | 2,429.87 | 10,267.54 |
| 17 | 3,882.05 | 3,527.35 | 3,721.30 | 3,610.18 | 14,740.88 |
| 18 | 10,166.14 | 9,962.54 | 9,666.22 | 9,843.10 | 39,638 |

DRUG CODE:9300DRUG NAME:MORPHINE
STATE:MASSACHUSETTS

| ZIP CODE | QUARTER 1 | QUARTER 2 | QUARTER 3 | QUARTER 4 | TOTAL GRAMS |
|---|---|---|---|---|---|
| 19 | 5,007.65 | 4,616.59 | 4,397.70 | 4,298.39 | 18,320.33 |
| 20 | 2,693.09 | 2,623.84 | 2,361.40 | 2,491.85 | 10,170.18 |
| 21 | 9,026.41 | 9,023.58 | 8,491.77 | 8,201.94 | 34,743.70 |
| 22 | 926.77 | 892.12 | 1,088.08 | 853.29 | 3,760.26 |
| 23 | 6,086.13 | 5,559.26 | 5,553.69 | 5,286.63 | 22,485.71 |
| 24 | 1,633.44 | 1,325.91 | 1,553.99 | 1,557.33 | 6,070.67 |
| 25 | 1,335.14 | 1,432.74 | 1,228.62 | 1,141.68 | 5,138.18 |
| 26 | 2,128.75 | 2,108.23 | 2,051.89 | 1,856.21 | 8,145.08 |
| 27 | 9,846.69 | 9,303.58 | 8,957.88 | 9,319.67 | 37,427.82 |
| TOTAL | 78,799.21 | 75,586.78 | 72,799.52 | 71,692.68 | 298,878.19 |

STATE: MICHIGAN

| ZIP CODE | QUARTER 1 | QUARTER 2 | QUARTER 3 | QUARTER 4 | TOTAL GRAMS |
|---|---|---|---|---|---|
| 480 | 27,242.94 | 25,923.86 | 25,253.70 | 24,465.66 | 102,886.16 |
| 481 | 26,701.08 | 25,780.69 | 24,904.89 | 24,526.23 | 101,912.89 |
| 482 | 15,509.74 | 14,118.85 | 14,471.38 | 13,444.46 | 57,544.43 |
| 483 | 21,416.46 | 20,456.04 | 20,602.44 | 20,290.39 | 82,765.33 |
| 484 | 8,960.43 | 8,514.18 | 8,468.96 | 8,184.11 | 34,127.68 |
| 485 | 5,410.28 | 5,217.46 | 5,606.07 | 5,634.70 | 21,868.51 |
| 486 | 16,451.47 | 16,087.64 | 16,114.04 | 14,935.98 | 63,589.13 |
| 487 | 7,952.31 | 7,837.38 | 8,233.63 | 7,509.81 | 31,533.13 |
| 488 | 11,014.17 | 10,839.91 | 11,119.77 | 10,603.15 | 43,577 |
| 489 | 5,294.04 | 4,782.65 | 4,640.90 | 4,683.90 | 19,401.49 |
| 490 | 21,124.35 | 20,818.12 | 19,601.33 | 18,755.95 | 80,299.75 |
| 491 | 1,659.36 | 1,691.70 | 1,637.78 | 1,509.53 | 6,498.37 |
| 492 | 7,904.30 | 7,455.13 | 7,182.30 | 6,978.27 | 29,520 |
| 493 | 4,319.40 | 4,086.85 | 4,306.10 | 4,337.76 | 17,050.11 |
| 494 | 12,113.82 | 12,382.98 | 12,053.87 | 11,375.23 | 47,925.90 |
| 495 | 9,280.84 | 9,621.97 | 9,077.46 | 9,133.22 | 37,113.49 |
| 496 | 11,105.76 | 11,108.90 | 10,574.97 | 10,070.32 | 42,859.95 |
| 497 | 10,066.34 | 9,846.58 | 9,823.68 | 9,030.85 | 38,767.45 |
| 498 | 5,722.77 | 5,126.03 | 5,514.31 | 4,946.82 | 21,309.93 |
| 499 | 1,627.13 | 1,639.32 | 1,583.68 | 1,483.07 | 6,333.20 |
| TOTAL | 230,876.99 | 223,336.24 | 220,771.26 | 211,899.41 | 886,883.90 |

RETAIL DRUG DISTRIBUTION BY ZIP CODE WITHIN STATE BY GRAMS WT

STATE: MINNESOTA

| ZIP CODE | QUARTER 1 | QUARTER 2 | QUARTER 3 | QUARTER 4 | TOTAL GRAMS |
|---|---|---|---|---|---|
| 550 | 5,706.31 | 5,635.14 | 5,557.66 | 5,361.55 | 22,260.66 |
| 551 | 7,567.56 | 7,472.05 | 8,692.32 | 8,488.34 | 32,220.27 |
| 553 | 6,911.03 | 6,971.80 | 6,974.73 | 6,759.68 | 27,617.24 |
| 554 | 13,300.56 | 13,711.16 | 12,510.94 | 12,340.31 | 51,862.97 |
| 556 | 231.93 | 233.38 | 241.39 | 204.49 | 911.19 |
| 557 | 2,400.21 | 2,111.46 | 1,967.82 | 1,675.19 | 8,154.68 |
| 558 | 1,948.57 | 1,943.69 | 1,967.13 | 1,612.23 | 7,471.62 |
| 559 | 2,435.55 | 2,308.81 | 2,481.69 | 2,198.82 | 9,424.87 |
| 560 | 2,212.73 | 2,036.23 | 1,933.21 | 2,072.67 | 8,254.84 |
| 561 | 811.89 | 792.24 | 801.74 | 834.45 | 3,240.32 |
| 562 | 1,386.36 | 1,280 | 1,165.88 | 1,132.66 | 4,964.90 |
| 563 | 4,519.14 | 4,507.50 | 4,157.53 | 3,841.81 | 17,025.98 |
| 564 | 1,827.47 | 1,874.69 | 1,709.19 | 1,636.69 | 7,048.04 |
| 565 | 1,909.05 | 1,858.24 | 1,763.29 | 1,500.50 | 7,031.08 |
| 566 | 1,029.91 | 940.03 | 945.38 | 823.09 | 3,738.41 |
| 567 | 863.24 | 876.26 | 841.52 | 798.19 | 3,379.21 |
| TOTAL | 55,061.51 | 54,552.68 | 53,711.42 | 51,280.67 | 214,606.28 |

STATE: MISSISSIPPI

| ZIP CODE | QUARTER 1 | QUARTER 2 | QUARTER 3 | QUARTER 4 | TOTAL GRAMS |
|---|---|---|---|---|---|
| 386 | 4,267.64 | 4,367.65 | 4,145.21 | 4,211.95 | 16,992.45 |
| 387 | 546.55 | 615.04 | 477.42 | 527.09 | 2,166.10 |
| 388 | 3,985.74 | 4,248.40 | 4,453.74 | 4,296.41 | 16,984.29 |
| 389 | 872.22 | 777.12 | 814.39 | 869.39 | 3,333.12 |
| 390 | 3,054.45 | 3,005.14 | 3,331.58 | 3,169.24 | 12,560.41 |
| 391 | 5,123.37 | 5,213.84 | 5,456.75 | 5,766.64 | 21,560.60 |
| 392 | 3,861.18 | 3,625.06 | 3,235.37 | 3,624.53 | 14,346.14 |
| 393 | 3,807.57 | 3,962.29 | 3,975.99 | 4,024.34 | 15,770.19 |
| 394 | 6,879.81 | 6,305.44 | 6,146.77 | 6,035.96 | 25,367.98 |
| 395 | 7,555.02 | 7,432.70 | 7,432.48 | 6,618.62 | 29,038.82 |
| 396 | 2,082.66 | 2,190.46 | 2,439.09 | 2,611.14 | 9,323.35 |
| 397 | 1,662.97 | 1,645.23 | 1,396.70 | 1,429.13 | 6,134.03 |

DRUG CODE:9300DRUG NAME:MORPHINE
STATE:MISSISSIPPI

| ZIP CODE | QUARTER 1 | QUARTER 2 | QUARTER 3 | QUARTER 4 | TOTAL GRAMS |
|---|---|---|---|---|---|
| TOTAL | 43,699.18 | 43,388.37 | 43,305.49 | 43,184.44 | 173,577.48 |

STATE: MISSOURI

| ZIP CODE | QUARTER 1 | QUARTER 2 | QUARTER 3 | QUARTER 4 | TOTAL GRAMS |
|---|---|---|---|---|---|
| 630 | 11,735.08 | 12,121.76 | 11,839.38 | 11,182.64 | 46,878.86 |
| 631 | 20,166.11 | 19,311.08 | 18,295.74 | 16,893.39 | 74,666.32 |
| 633 | 6,471.97 | 6,692.31 | 6,636.17 | 6,395.51 | 26,195.96 |
| 634 | 694.96 | 653.91 | 739.23 | 678.41 | 2,766.51 |
| 635 | 1,204.80 | 1,291.07 | 1,304.02 | 1,453.22 | 5,253.11 |
| 636 | 3,503.29 | 3,480.94 | 3,325.50 | 3,151.85 | 13,461.58 |
| 637 | 1,771.03 | 1,584.61 | 1,511.21 | 1,483.51 | 6,350.36 |
| 638 | 1,561.35 | 1,545.21 | 1,725.89 | 1,710.05 | 6,542.50 |
| 639 | 1,747.83 | 1,692.17 | 1,739.42 | 1,733.60 | 6,913.02 |
| 640 | 8,485.66 | 7,940.43 | 7,814.23 | 7,786.23 | 32,026.55 |
| 641 | 8,231.85 | 8,723.74 | 7,562.10 | 7,275.19 | 31,792.88 |
| 644 | 989.27 | 974.39 | 875.77 | 889.53 | 3,728.96 |
| 645 | 1,803.74 | 1,764.28 | 1,590.92 | 1,606.21 | 6,765.15 |
| 646 | 1,051.37 | 969.55 | 908.38 | 1,008.49 | 3,937.79 |
| 647 | 3,009.97 | 2,992.21 | 2,867.45 | 2,974.93 | 11,844.56 |
| 648 | 3,889 | 4,137.01 | 4,156.08 | 4,115.80 | 16,297.89 |
| 650 | 1,952.96 | 2,053.92 | 2,097.56 | 1,872.19 | 7,976.63 |
| 651 | 1,321.87 | 1,474.66 | 1,391.55 | 1,262.09 | 5,450.17 |

| | | | | | |
|---|---|---|---|---|---|
| 652 | 5,109.96 | 4,814.87 | 5,279.59 | 5,069.29 | 20,273.71 |
| 653 | 1,210.14 | 1,132.71 | 1,193.51 | 1,144.32 | 4,680.68 |
| 654 | 1,692.20 | 1,725.05 | 1,635.55 | 1,522.70 | 6,575.50 |
| 655 | 2,670.01 | 2,720.28 | 2,712.73 | 2,676.13 | 10,779.15 |
| 656 | 4,823.84 | 4,801.46 | 4,729.08 | 4,717.41 | 19,071.79 |
| 657 | 8,082.64 | 8,109.86 | 7,785.83 | 7,630.29 | 31,608.62 |
| 658 | 9,522.36 | 9,235.33 | 9,183.50 | 8,499.68 | 36,440.87 |
| TOTAL | 112,703.26 | 111,942.81 | 108,900.39 | 104,732.66 | 438,279.12 |

STATE: MONTANA

| ZIP CODE | QUARTER 1 | QUARTER 2 | QUARTER 3 | QUARTER 4 | TOTAL GRAMS |
|---|---|---|---|---|---|
| 590 | 1,561.54 | 1,644.38 | 1,383.45 | 1,287.24 | 5,876.61 |
| 591 | 4,332.14 | 4,000.72 | 3,816.95 | 3,662.95 | 15,812.76 |
| 592 | 699.84 | 658.74 | 657.24 | 580.49 | 2,596.31 |
| 593 | 892.7 | 794.77 | 847.04 | 756.56 | 3,291.07 |
| 594 | 3,442.07 | 3,265.41 | 3,352.08 | 2,926.50 | 12,986.06 |
| 595 | 463.98 | 507.1 | 389.51 | 375.64 | 1,736.23 |
| 596 | 2,758.85 | 2,679.47 | 2,437.58 | 2,450.99 | 10,326.89 |
| 597 | 2,339.30 | 2,384.98 | 2,326.18 | 2,084.37 | 9,134.83 |
| 598 | 3,356.22 | 3,357.02 | 3,248.53 | 3,092.30 | 13,054.07 |
| 599 | 2,890.50 | 2,850.88 | 2,669.66 | 2,624.79 | 11,035.83 |
| TOTAL | 22,737.14 | 22,143.47 | 21,128.22 | 19,841.83 | 85,850.66 |

STATE: NEBRASKA

| ZIP CODE | QUARTER 1 | QUARTER 2 | QUARTER 3 | QUARTER 4 | TOTAL GRAMS |
|---|---|---|---|---|---|
| 680 | 2,073.62 | 2,122.11 | 1,996.94 | 2,077.99 | 8,270.66 |
| 681 | 6,889.09 | 6,913.26 | 7,389.42 | 7,060.77 | 28,252.54 |
| 683 | 575.81 | 640.97 | 519.27 | 656.64 | 2,392.69 |
| 684 | 597.67 | 625.47 | 558.77 | 604.02 | 2,385.93 |
| 685 | 3,257.51 | 3,218.27 | 3,234.34 | 2,994.41 | 12,704.53 |
| 686 | 515.7 | 408.67 | 440.33 | 446.34 | 1,811.04 |
| 687 | 1,053.93 | 1,063.36 | 998.31 | 946.96 | 4,062.56 |
| 688 | 2,065.34 | 1,955.35 | 1,970.96 | 2,017.67 | 8,009.32 |
| 689 | 734.7 | 657.51 | 666.95 | 664.5 | 2,723.66 |
| 690 | 370.37 | 393.97 | 380.47 | 381.33 | 1,526.14 |
| 691 | 914.49 | 888.84 | 892.64 | 847.93 | 3,543.90 |
| 692 | 54.37 | 62.69 | 64.86 | 57.29 | 239.21 |
| 693 | 1,176.91 | 1,120.40 | 1,096.58 | 1,057.86 | 4,451.75 |
| TOTAL | 20,279.51 | 20,070.87 | 20,209.84 | 19,813.71 | 80,373.93 |

STATE: NEVADA

| ZIP CODE | QUARTER 1 | QUARTER 2 | QUARTER 3 | QUARTER 4 | TOTAL GRAMS |
|---|---|---|---|---|---|
| 890 | 18,558.05 | 17,668.30 | 17,958.98 | 17,177.96 | 71,363.29 |
| 891 | 39,754.70 | 36,820.22 | 35,458.63 | 33,763.99 | 145,797.54 |
| 893 | 227.77 | 168.6 | 166.57 | 128.08 | 691.02 |
| 894 | 5,888.65 | 5,373.48 | 5,223.11 | 5,141.02 | 21,626.26 |
| 895 | 9,653.05 | 10,033.01 | 8,049.50 | 6,881.77 | 34,617.33 |
| 897 | 2,053.52 | 2,081.59 | 2,128.31 | 1,982.02 | 8,245.44 |

DRUG CODE:9300DRUG NAME:MORPHINE
STATE:NEVADA

| ZIP CODE | QUARTER 1 | QUARTER 2 | QUARTER 3 | QUARTER 4 | TOTAL GRAMS |
|---|---|---|---|---|---|
| 898 | 657.2 | 662.55 | 567.75 | 594.69 | 2,482.19 |
| TOTAL | 76,792.94 | 72,807.75 | 69,552.85 | 65,669.53 | 284,823.07 |

STATE: NEW HAMPSHIRE

| ZIP CODE | QUARTER 1 | QUARTER 2 | QUARTER 3 | QUARTER 4 | TOTAL GRAMS |
|---|---|---|---|---|---|
| 30 | 4,974.83 | 4,848.53 | 4,940.37 | 4,695.73 | 19,459.46 |
| 31 | 3,733.06 | 3,347.54 | 2,890.69 | 3,084.90 | 13,056.19 |
| 32 | 2,099.24 | 2,046.98 | 2,131.03 | 1,892.31 | 8,169.56 |

| | | | | | |
|---|---|---|---|---|---|
| 33 | 1,998.93 | 2,282.16 | 1,978 | 2,024.28 | 8,283.37 |
| 34 | 1,123.96 | 1,070.38 | 1,053.33 | 1,069.99 | 4,317.66 |
| 35 | 979.26 | 835.77 | 903.4 | 759.11 | 3,477.54 |
| 36 | 159.12 | 155.06 | 140.25 | 132.28 | 586.71 |
| 37 | 1,122.31 | 1,071.22 | 1,200.04 | 1,055.07 | 4,448.64 |
| 38 | 4,637.20 | 4,428.18 | 4,610.14 | 4,242.78 | 17,918.30 |
| TOTAL | 20,827.91 | 20,085.82 | 19,847.25 | 18,956.45 | 79,717.43 |

STATE: NEW JERSEY

| ZIP CODE | QUARTER 1 | QUARTER 2 | QUARTER 3 | QUARTER 4 | TOTAL GRAMS |
|---|---|---|---|---|---|
| 70 | 14,868.19 | 13,762.06 | 15,266.13 | 14,943.46 | 58,839.84 |
| 71 | 2,137.29 | 2,073.56 | 1,958.64 | 1,816.11 | 7,985.60 |
| 72 | 1,961.97 | 1,826.34 | 1,115.19 | 791.29 | 5,694.79 |
| 73 | 1,165.22 | 1,119.99 | 1,069.48 | 1,128.80 | 4,483.49 |
| 74 | 5,096.49 | 5,101.80 | 5,299.30 | 5,368.39 | 20,865.98 |
| 75 | 1,195.07 | 947.07 | 932.71 | 875.24 | 3,950.09 |
| 76 | 5,432.08 | 5,180.13 | 6,344.87 | 7,040.15 | 23,997.23 |
| 77 | 6,615.05 | 6,283.55 | 6,133.79 | 6,379.89 | 25,412.28 |
| 78 | 4,047.81 | 4,050.29 | 3,839.12 | 3,509.53 | 15,446.75 |
| 79 | 2,119.32 | 2,644.67 | 2,047.33 | 2,220.80 | 9,032.12 |
| 80 | 23,454.29 | 23,723.61 | 22,229.60 | 21,930.52 | 91,338.02 |
| 81 | 1,713.91 | 1,756.24 | 1,747.84 | 2,826.87 | 8,044.86 |
| 82 | 4,139.40 | 4,208.01 | 4,147.96 | 3,944.82 | 16,440.19 |
| 83 | 2,860.58 | 2,565.65 | 2,395.13 | 2,386.61 | 10,207.97 |
| 84 | 510.08 | 463.93 | 478.69 | 393.62 | 1,846.32 |
| 85 | 3,192.70 | 2,928.25 | 2,922.25 | 2,938.07 | 11,981.27 |
| 86 | 2,530.39 | 2,591.63 | 2,777.70 | 2,383.89 | 10,283.61 |
| 87 | 6,613.48 | 6,285.89 | 6,130.90 | 6,413.62 | 25,443.89 |
| 88 | 10,200.13 | 10,508.91 | 10,485.31 | 10,572.91 | 41,767.26 |
| 89 | 779.43 | 736.24 | 900.01 | 784.54 | 3,200.22 |
| TOTAL | 100,632.88 | 98,757.82 | 98,221.95 | 98,649.13 | 396,261.78 |

STATE: NEW MEXICO

| ZIP CODE | QUARTER 1 | QUARTER 2 | QUARTER 3 | QUARTER 4 | TOTAL GRAMS |
|---|---|---|---|---|---|
| 870 | 2,570.36 | 2,449.95 | 2,342.43 | 2,303.32 | 9,666.06 |
| 871 | 17,888.26 | 17,308.79 | 16,269.58 | 15,296.95 | 66,763.58 |
| 873 | 1,126.04 | 1,082.33 | 1,103.71 | 1,052.68 | 4,364.76 |
| 874 | 1,183.01 | 1,088.36 | 1,049.48 | 961.96 | 4,282.81 |
| 875 | 3,382.48 | 3,135.59 | 3,083.20 | 3,134.58 | 12,735.85 |
| 877 | 389.5 | 411.94 | 396.92 | 479.65 | 1,678.01 |
| 878 | 138.07 | 134.31 | 126.5 | 154.16 | 553.04 |
| 879 | 291.48 | 321.86 | 245.98 | 287.4 | 1,146.72 |
| 880 | 4,360.82 | 3,532.19 | 3,189.92 | 2,728.83 | 13,811.76 |
| 881 | 746.56 | 760.74 | 718.46 | 614.99 | 2,840.75 |
| 882 | 3,198.80 | 3,108.47 | 2,906.16 | 2,832.72 | 12,046.15 |
| 883 | 923.82 | 791.68 | 775.15 | 737.4 | 3,228.05 |
| 884 | 108.45 | 92.21 | 93.76 | 118.02 | 412.44 |
| TOTAL | 36,307.65 | 34,218.42 | 32,301.25 | 30,702.66 | 133,529.98 |

STATE: NEW YORK

| ZIP CODE | QUARTER 1 | QUARTER 2 | QUARTER 3 | QUARTER 4 | TOTAL GRAMS |
|---|---|---|---|---|---|
| 63 | 0.03 | 0.03 | 0 | 0 | 0.06 |
| 100 | 16,096.51 | 15,850.18 | 15,812.26 | 15,059.95 | 62,818.90 |
| 101 | 309.35 | 259.75 | 230.82 | 227.77 | 1,027.69 |
| 102 | 9.93 | 5.64 | 18.05 | 5.64 | 39.26 |
| 103 | 7,315.42 | 7,272.69 | 6,950.51 | 7,019.20 | 28,557.82 |
| 104 | 9,172.93 | 9,376.98 | 7,583.04 | 7,296.37 | 33,429.32 |
| 105 | 3,139.83 | 3,003.76 | 2,895.21 | 2,814.59 | 11,853.39 |
| 106 | 551.42 | 507.3 | 467.68 | 464.16 | 1,990.56 |

| | | | | | |
|---|---|---|---|---|---|
| 107 | 1,088.75 | 1,148.91 | 1,016.69 | 1,047.16 | 4,301.51 |
| 108 | 314.02 | 336.81 | 320.19 | 331.8 | 1,302.82 |
| 109 | 4,287.02 | 4,066.16 | 4,283.51 | 3,937.78 | 16,574.47 |

DRUG CODE:9300DRUG NAME:MORPHINE
STATE:NEW YORK

| ZIP CODE | QUARTER 1 | QUARTER 2 | QUARTER 3 | QUARTER 4 | TOTAL GRAMS |
|---|---|---|---|---|---|
| 110 | 3,797.18 | 3,461.22 | 3,253.86 | 3,612.13 | 14,124.39 |
| 111 | 1,141.91 | 1,216.44 | 1,088.77 | 991.87 | 4,438.99 |
| 112 | 9,979.38 | 11,082.91 | 10,236.90 | 9,656.41 | 40,955.60 |
| 113 | 4,244.43 | 4,040.12 | 3,771.19 | 3,650.18 | 15,705.92 |
| 114 | 2,233.99 | 2,391.86 | 2,001.82 | 2,102.91 | 8,730.58 |
| 115 | 5,631 | 5,559.88 | 5,408.25 | 5,235.92 | 21,835.05 |
| 116 | 710.09 | 632.14 | 679.31 | 564.93 | 2,586.47 |
| 117 | 21,973.21 | 22,106.83 | 21,930.34 | 22,762.03 | 88,772.41 |
| 118 | 735.51 | 715.22 | 739.31 | 654.33 | 2,844.37 |
| 119 | 2,205.98 | 2,086.30 | 2,089.37 | 1,932.88 | 8,314.53 |
| 120 | 3,804.47 | 4,053.66 | 3,879.73 | 3,909.60 | 15,647.46 |
| 121 | 2,129.97 | 1,926.27 | 1,992.94 | 1,830.91 | 7,880.09 |
| 122 | 3,445.61 | 3,148.75 | 3,304.65 | 3,264.42 | 13,163.43 |
| 123 | 1,118.28 | 983.07 | 987.13 | 1,032.12 | 4,120.60 |
| 124 | 3,236.43 | 3,133.30 | 3,022.33 | 2,875.93 | 12,267.99 |
| 125 | 4,596.87 | 4,343.81 | 4,066.39 | 3,842.34 | 16,849.41 |
| 126 | 945.27 | 993.57 | 852.51 | 956.44 | 3,747.79 |
| 127 | 2,264.42 | 2,202.42 | 2,137.52 | 2,170.75 | 8,775.11 |
| 128 | 3,509.09 | 3,343.74 | 3,478.08 | 3,210.75 | 13,541.66 |
| 129 | 2,732.88 | 2,596.14 | 2,637.57 | 2,677.29 | 10,643.88 |
| 130 | 5,072.26 | 4,839.65 | 4,562.37 | 4,473.32 | 18,947.60 |
| 131 | 1,571.95 | 1,510.13 | 1,422.75 | 1,386.14 | 5,890.97 |
| 132 | 5,204.17 | 4,663.22 | 4,591.14 | 4,415.07 | 18,873.60 |
| 133 | 1,814.64 | 1,815.06 | 1,671.46 | 1,591.47 | 6,892.63 |
| 134 | 2,445.03 | 2,360.90 | 2,125.86 | 1,981.64 | 8,913.43 |
| 135 | 812.76 | 748.15 | 838.57 | 744.31 | 3,143.79 |
| 136 | 2,705.44 | 2,644.70 | 2,623.67 | 2,513.49 | 10,487.30 |
| 137 | 1,062.49 | 1,113.13 | 948.44 | 948.85 | 4,072.91 |
| 138 | 3,384.70 | 3,607.97 | 3,474.94 | 3,347.22 | 13,814.83 |
| 139 | 961.89 | 939.96 | 859.85 | 909.77 | 3,671.47 |
| 140 | 6,493 | 6,146.86 | 5,550.21 | 5,448.34 | 23,638.41 |
| 141 | 3,231.83 | 2,934.58 | 2,740.75 | 2,626.64 | 11,533.80 |
| 142 | 9,291.97 | 8,665.41 | 7,708.88 | 7,319.93 | 32,986.19 |
| 143 | 1,463.28 | 1,280.37 | 1,183.60 | 1,031.29 | 4,958.54 |
| 144 | 3,815.60 | 3,654.02 | 3,625.41 | 3,468.98 | 14,564.01 |
| 145 | 3,380.63 | 3,240.22 | 2,982.94 | 2,990.94 | 12,594.73 |
| 146 | 7,237.78 | 6,786.88 | 6,728.42 | 6,336.40 | 27,089.48 |
| 147 | 2,657.62 | 2,660.98 | 2,532.61 | 2,311.53 | 10,162.74 |
| 148 | 4,507.88 | 4,385.84 | 4,496.55 | 4,069.41 | 17,459.68 |
| 149 | 1,083.44 | 1,088.14 | 928.34 | 918.73 | 4,018.65 |
| TOTAL | 190,919.54 | 186,932.03 | 178,732.69 | 173,972.03 | 730,556.29 |

STATE: NORTH CAROLINA

| ZIP CODE | QUARTER 1 | QUARTER 2 | QUARTER 3 | QUARTER 4 | TOTAL GRAMS |
|---|---|---|---|---|---|
| 270 | 4,854.79 | 4,711.64 | 4,453.79 | 4,093.30 | 18,113.52 |
| 271 | 5,139.98 | 4,491.15 | 4,821.11 | 4,066.02 | 18,518.26 |
| 272 | 8,165.71 | 8,335.16 | 8,110.55 | 7,862.02 | 32,473.44 |
| 273 | 5,261.92 | 4,845.88 | 4,925.80 | 4,683.05 | 19,716.65 |
| 274 | 3,300.64 | 3,315.15 | 3,315.76 | 3,338.12 | 13,269.67 |
| 275 | 11,994.11 | 12,320.49 | 11,254.18 | 11,421.49 | 46,990.27 |
| 276 | 4,159.43 | 4,386.58 | 4,180.05 | 4,611.77 | 17,337.83 |
| 277 | 6,693.79 | 6,613.10 | 6,954.12 | 5,958.35 | 26,219.36 |
| 278 | 7,576.17 | 7,803.61 | 7,490.91 | 7,368.68 | 30,239.37 |

| | | | | | |
|---|---|---|---|---|---|
| 279 | 2,619.12 | 2,464.24 | 2,525.83 | 2,608.12 | 10,217.31 |
| 280 | 11,670.76 | 10,808.25 | 10,928.49 | 10,256.83 | 43,664.33 |
| 281 | 9,371.13 | 8,579.27 | 8,698.77 | 8,027.82 | 34,676.99 |
| 282 | 7,064.54 | 6,265.67 | 6,415.31 | 6,142.79 | 25,888.31 |
| 283 | 11,328.15 | 10,659.88 | 10,972.05 | 9,740.29 | 42,700.37 |
| 284 | 9,103.63 | 9,069.02 | 8,705.62 | 7,526.60 | 34,404.87 |
| 285 | 7,203.82 | 7,169.49 | 7,170.96 | 6,956.52 | 28,500.79 |
| 286 | 12,484.95 | 12,749.14 | 12,179.31 | 12,234.05 | 49,647.45 |
| 287 | 9,529.47 | 9,156.10 | 8,881.61 | 8,604.28 | 36,171.46 |
| 288 | 5,978.68 | 4,860.50 | 4,406.57 | 4,194.40 | 19,440.15 |
| 289 | 1,155.39 | 1,030.29 | 1,031.56 | 1,083.67 | 4,300.91 |
| TOTAL | 144,656.18 | 139,634.61 | 137,422.35 | 130,778.17 | 552,491.31 |

STATE: NORTH DAKOTA

| ZIP CODE | QUARTER 1 | QUARTER 2 | QUARTER 3 | QUARTER 4 | TOTAL GRAMS |
|---|---|---|---|---|---|

DRUG CODE:9300DRUG NAME:MORPHINE
STATE:NORTH DAKOTA

| ZIP CODE | QUARTER 1 | QUARTER 2 | QUARTER 3 | QUARTER 4 | TOTAL GRAMS |
|---|---|---|---|---|---|
| 580 | 510.88 | 536 | 472.38 | 417.39 | 1,936.65 |
| 581 | 2,596.68 | 2,521.58 | 1,918.31 | 1,682.34 | 8,718.91 |
| 582 | 1,117.90 | 1,050.07 | 992.77 | 878.01 | 4,038.75 |
| 583 | 370.8 | 429.4 | 332.21 | 311.86 | 1,444.27 |
| 584 | 365.05 | 335.85 | 360.07 | 315.75 | 1,376.72 |
| 585 | 2,381.74 | 2,326.89 | 2,177.53 | 1,970.03 | 8,856.19 |
| 586 | 371.48 | 338.31 | 277.19 | 330.01 | 1,316.99 |
| 587 | 465.06 | 441.96 | 413.49 | 362.3 | 1,682.81 |
| 588 | 354.86 | 297.34 | 254.64 | 239.76 | 1,146.60 |
| TOTAL | 8,534.45 | 8,277.40 | 7,198.59 | 6,507.45 | 30,517.89 |

STATE: OHIO

| ZIP CODE | QUARTER 1 | QUARTER 2 | QUARTER 3 | QUARTER 4 | TOTAL GRAMS |
|---|---|---|---|---|---|
| 430 | 7,744.25 | 7,380.87 | 8,039.99 | 7,296.06 | 30,461.17 |
| 431 | 4,457.88 | 4,545.48 | 4,362.77 | 4,333.13 | 17,699.26 |
| 432 | 16,441.93 | 15,400.15 | 15,503.09 | 12,901.96 | 60,247.13 |
| 433 | 1,642.58 | 1,642.07 | 1,494.59 | 1,393.15 | 6,172.39 |
| 434 | 1,122.53 | 1,140.45 | 1,177.34 | 1,065.52 | 4,505.84 |
| 435 | 3,616.09 | 3,788.52 | 3,488.61 | 3,441.58 | 14,334.80 |
| 436 | 6,284.03 | 6,396.72 | 6,113.73 | 5,949.53 | 24,744.01 |
| 437 | 2,419.77 | 2,302.42 | 2,322.37 | 2,301.24 | 9,345.80 |
| 438 | 398.47 | 395.66 | 397.13 | 401.42 | 1,592.68 |
| 439 | 2,844.66 | 2,800.91 | 2,778.67 | 2,550.08 | 10,974.32 |
| 440 | 6,578.74 | 6,663.47 | 6,301.15 | 5,836.77 | 25,380.13 |
| 441 | 10,431.52 | 10,136.02 | 10,098.49 | 9,586.08 | 40,252.11 |
| 442 | 6,702.92 | 6,781.70 | 6,457.65 | 6,143.97 | 26,086.24 |
| 443 | 3,463.80 | 3,205.41 | 3,217.41 | 2,971.61 | 12,858.23 |
| 444 | 4,461.63 | 4,407.84 | 4,696.75 | 4,369.31 | 17,935.53 |
| 445 | 2,973.61 | 2,955.54 | 2,911.81 | 2,989.21 | 11,830.17 |
| 446 | 4,045.09 | 3,892.13 | 3,878.89 | 3,726.55 | 15,542.66 |
| 447 | 2,789.93 | 2,762.91 | 2,706.84 | 2,688.94 | 10,948.62 |
| 448 | 2,899.31 | 2,756.84 | 2,833 | 2,810.21 | 11,299.36 |
| 449 | 754.67 | 810.09 | 957.37 | 928.57 | 3,450.70 |
| 450 | 10,540.07 | 10,416.30 | 10,764.83 | 10,220.01 | 41,941.21 |
| 451 | 3,416.33 | 3,351.38 | 3,375.13 | 3,151.52 | 13,294.36 |
| 452 | 10,370.36 | 10,713.93 | 9,771.43 | 10,006.69 | 40,862.41 |
| 453 | 4,973.19 | 4,912.03 | 4,882.60 | 4,657.38 | 19,425.20 |
| 454 | 7,112 | 7,010.95 | 7,107.50 | 6,472.20 | 27,702.65 |
| 455 | 1,055.58 | 1,035.27 | 1,073.99 | 915.43 | 4,080.27 |
| 456 | 6,145.48 | 6,170.03 | 5,586.72 | 5,735.38 | 23,637.61 |
| 457 | 1,515.64 | 1,315.18 | 1,310.80 | 1,404.07 | 5,545.69 |

| | | | | | | |
|---|---|---|---|---|---|---|
| | 458 | 3,073.80 | 2,829.28 | 2,786.63 | 2,906.83 | 11,596.54 |
| TOTAL | | 140,275.86 | 137,919.55 | 136,397.28 | 129,154.40 | 543,747.09 |

STATE: OKLAHOMA

| ZIP CODE | | QUARTER 1 | QUARTER 2 | QUARTER 3 | QUARTER 4 | TOTAL GRAMS |
|---|---|---|---|---|---|---|
| | 730 | 19,256.59 | 18,497.45 | 17,944.70 | 17,715.68 | 73,414.42 |
| | 731 | 23,717.72 | 22,867.46 | 22,397.32 | 21,228.36 | 90,210.86 |
| | 734 | 2,601.90 | 2,610.48 | 2,421.46 | 2,389.16 | 10,023 |
| | 735 | 6,649.61 | 6,645.28 | 6,401.88 | 6,196.67 | 25,893.44 |
| | 736 | 1,865.17 | 1,871.29 | 1,933.70 | 1,807.88 | 7,478.04 |
| | 737 | 1,675.39 | 1,638.63 | 1,689.26 | 1,576.76 | 6,580.04 |
| | 738 | 490.44 | 443.55 | 462.66 | 460.1 | 1,856.75 |
| | 739 | 177.92 | 110.15 | 142.56 | 105.02 | 535.65 |
| | 740 | 18,524.13 | 17,315.28 | 17,876.97 | 17,595.36 | 71,311.74 |
| | 741 | 12,689.28 | 12,378.33 | 11,664.73 | 10,828.07 | 47,560.41 |
| | 743 | 2,934.74 | 2,732.89 | 2,628.93 | 2,557.32 | 10,853.88 |
| | 744 | 6,765.79 | 6,723.22 | 6,236.92 | 6,379.61 | 26,105.54 |
| | 745 | 1,989.02 | 2,007.05 | 2,090.09 | 2,049.97 | 8,136.13 |
| | 746 | 1,193.87 | 1,241.33 | 1,372.01 | 1,204.30 | 5,011.51 |
| | 747 | 1,548.33 | 1,516.05 | 1,520.54 | 1,450.92 | 6,035.84 |
| | 748 | 4,592.87 | 4,579.18 | 4,488.61 | 4,180.69 | 17,841.35 |
| | 749 | 1,957.69 | 1,769.97 | 1,553.90 | 1,608.42 | 6,889.98 |
| TOTAL | | 108,630.46 | 104,947.59 | 102,826.24 | 99,334.29 | 415,738.58 |

STATE: OREGON

| ZIP CODE | | QUARTER 1 | QUARTER 2 | QUARTER 3 | QUARTER 4 | TOTAL GRAMS |
|---|---|---|---|---|---|---|
| | 970 | 14,786.22 | 14,589.23 | 13,824.87 | 12,620.58 | 55,820.90 |
| | 971 | 7,499.45 | 7,411.65 | 7,206.35 | 6,783.99 | 28,901.44 |

DRUG CODE:9300DRUG NAME:MORPHINE
STATE:OREGON

| ZIP CODE | | QUARTER 1 | QUARTER 2 | QUARTER 3 | QUARTER 4 | TOTAL GRAMS |
|---|---|---|---|---|---|---|
| | 972 | 23,350.55 | 21,942.04 | 19,884.04 | 19,108.29 | 84,284.92 |
| | 973 | 12,017.03 | 11,043.69 | 10,555.02 | 9,796.15 | 43,411.89 |
| | 974 | 20,349.45 | 19,375.25 | 18,484.05 | 16,314.26 | 74,523.01 |
| | 975 | 10,206.43 | 10,098.16 | 9,769.97 | 8,847.50 | 38,922.06 |
| | 976 | 1,751.45 | 1,671.71 | 1,526.08 | 1,561.79 | 6,511.03 |
| | 977 | 3,348.14 | 3,400.09 | 3,268.33 | 3,176.35 | 13,192.91 |
| | 978 | 3,408.85 | 3,185.28 | 3,118.29 | 3,004.79 | 12,717.21 |
| | 979 | 548.4 | 541.68 | 516.65 | 540.69 | 2,147.42 |
| TOTAL | | 97,265.97 | 93,258.78 | 88,153.65 | 81,754.39 | 360,432.79 |

STATE: PENNSYLVANIA

| ZIP CODE | | QUARTER 1 | QUARTER 2 | QUARTER 3 | QUARTER 4 | TOTAL GRAMS |
|---|---|---|---|---|---|---|
| | 150 | 13,589.43 | 13,635.79 | 12,881.38 | 13,074.68 | 53,181.28 |
| | 151 | 3,440.22 | 3,586.74 | 3,266.99 | 3,318.90 | 13,612.85 |
| | 152 | 10,846.13 | 10,976.19 | 11,287.51 | 10,129.11 | 43,238.94 |
| | 153 | 2,571.55 | 2,620.33 | 2,590.19 | 2,442.01 | 10,224.08 |
| | 154 | 2,636.31 | 2,404.55 | 2,575.15 | 2,382.67 | 9,998.68 |
| | 155 | 1,607.66 | 1,668.59 | 1,589.79 | 1,618.29 | 6,484.33 |
| | 156 | 5,576.54 | 5,579.03 | 5,830.10 | 5,447.83 | 22,433.50 |
| | 157 | 2,810.27 | 3,015.26 | 2,966.03 | 3,071.38 | 11,862.94 |
| | 158 | 2,343.83 | 2,402.20 | 2,346.21 | 2,183.94 | 9,276.18 |
| | 159 | 3,161.91 | 3,198.15 | 3,057.77 | 3,166.68 | 12,584.51 |
| | 160 | 3,667.98 | 3,568.06 | 3,448.22 | 3,451.82 | 14,136.08 |
| | 161 | 5,558.86 | 5,153.67 | 4,857.70 | 4,788.93 | 20,359.16 |
| | 162 | 1,548.80 | 1,642.31 | 1,582.21 | 1,567.07 | 6,340.39 |
| | 163 | 5,580.74 | 5,560.33 | 5,053.16 | 4,901.51 | 21,095.74 |
| | 164 | 1,844.75 | 1,899.78 | 1,831.51 | 1,665.29 | 7,241.33 |
| | 165 | 5,761.72 | 5,531.42 | 5,444.72 | 4,816.36 | 21,554.22 |

| | | | | | |
|---|---|---|---|---|---|
| 166 | 5,373.77 | 5,475.51 | 5,325.49 | 5,106.69 | 21,281.46 |
| 167 | 1,938.90 | 1,996.49 | 2,055.90 | 2,061.27 | 8,052.56 |
| 168 | 1,580.91 | 1,600.72 | 1,503.73 | 1,510.26 | 6,195.62 |
| 169 | 927.21 | 936.07 | 868.21 | 844.1 | 3,575.59 |
| 170 | 13,195.65 | 13,633.25 | 12,792.21 | 12,218.39 | 51,839.50 |
| 171 | 3,112.15 | 3,164.86 | 3,209.93 | 3,075.32 | 12,562.26 |
| 172 | 3,027.35 | 2,850.36 | 2,785.67 | 2,846.76 | 11,510.14 |
| 173 | 5,229.15 | 5,329.85 | 4,941.03 | 4,771.15 | 20,271.18 |
| 174 | 3,515.03 | 3,677.90 | 3,540.67 | 3,157.39 | 13,890.99 |
| 175 | 4,237.53 | 4,047.20 | 4,228.46 | 4,198.34 | 16,711.53 |
| 176 | 4,099.32 | 3,941.77 | 3,522.91 | 3,850.95 | 15,414.95 |
| 177 | 2,751.73 | 2,562.45 | 2,537.34 | 2,395.30 | 10,246.82 |
| 178 | 3,796.33 | 3,548.72 | 3,664.41 | 3,523.09 | 14,532.55 |
| 179 | 1,710.31 | 1,594.62 | 1,469.26 | 1,584.85 | 6,359.04 |
| 180 | 5,168.79 | 4,802.51 | 4,763.37 | 4,782.49 | 19,517.16 |
| 181 | 3,860.79 | 4,071.41 | 4,487.68 | 4,191.35 | 16,611.23 |
| 182 | 2,599.80 | 2,713.04 | 2,468.20 | 2,875.29 | 10,656.33 |
| 183 | 3,069.44 | 3,174.17 | 3,006.88 | 3,147.98 | 12,398.47 |
| 184 | 3,787.62 | 3,605.92 | 3,280.87 | 3,611.91 | 14,286.32 |
| 185 | 4,634.35 | 4,742.27 | 4,450.72 | 4,099.02 | 17,926.36 |
| 186 | 2,891.29 | 2,602.73 | 2,791.12 | 2,905.49 | 11,190.63 |
| 187 | 13,875.44 | 13,406.01 | 12,121.44 | 11,420.43 | 50,823.32 |
| 188 | 939 | 979.32 | 968.13 | 1,077.80 | 3,964.25 |
| 189 | 4,541.26 | 4,567.50 | 4,543.43 | 4,382.27 | 18,034.46 |
| 190 | 52,948.31 | 54,232.89 | 48,756.38 | 45,705.58 | 201,643.16 |
| 191 | 21,248.31 | 20,071.61 | 18,591.14 | 17,937.13 | 77,848.19 |
| 193 | 6,220.61 | 6,038.62 | 6,241.40 | 5,993.43 | 24,494.06 |
| 194 | 10,828.43 | 10,350.37 | 9,822.82 | 9,672.87 | 40,674.49 |
| 195 | 1,958.36 | 2,002.24 | 2,039.73 | 1,839.07 | 7,839.40 |
| 196 | 2,905.13 | 2,629.12 | 2,421.03 | 2,445.81 | 10,401.09 |
| TOTAL | 268,518.97 | 266,791.90 | 253,808.20 | 245,258.25 | 1,034,377.32 |

STATE: PUERTO RICO

| ZIP CODE | QUARTER 1 | QUARTER 2 | QUARTER 3 | QUARTER 4 | TOTAL GRAMS |
|---|---|---|---|---|---|
| 6 | 615.06 | 546 | 558.71 | 523.44 | 2,243.21 |
| 7 | 676.48 | 690.35 | 699.74 | 729.48 | 2,796.05 |
| 9 | 1,584.83 | 1,983.81 | 1,667.30 | 1,644.41 | 6,880.35 |
| TOTAL | 2,876.37 | 3,220.16 | 2,925.75 | 2,897.33 | 11,919.61 |

STATE: RHODE ISLAND

| ZIP CODE | QUARTER 1 | QUARTER 2 | QUARTER 3 | QUARTER 4 | TOTAL GRAMS |
|---|---|---|---|---|---|

DRUG CODE:9300DRUG NAME:MORPHINE
STATE:RHODE ISLAND

| ZIP CODE | QUARTER 1 | QUARTER 2 | QUARTER 3 | QUARTER 4 | TOTAL GRAMS |
|---|---|---|---|---|---|
| 28 | 7,665.48 | 7,993.11 | 7,795.04 | 8,076.55 | 31,530.18 |
| 29 | 5,227.26 | 4,782.95 | 5,144.63 | 5,258.20 | 20,413.04 |
| TOTAL | 12,892.74 | 12,776.06 | 12,939.67 | 13,334.75 | 51,943.22 |

STATE: SOUTH CAROLINA

| ZIP CODE | QUARTER 1 | QUARTER 2 | QUARTER 3 | QUARTER 4 | TOTAL GRAMS |
|---|---|---|---|---|---|
| 290 | 5,895.31 | 5,683.04 | 5,713.33 | 5,765.49 | 23,057.17 |
| 291 | 4,958.92 | 4,681.06 | 4,934.02 | 4,900.56 | 19,474.56 |
| 292 | 5,502.53 | 5,621.08 | 5,473.13 | 5,496.30 | 22,093.04 |
| 293 | 7,479.88 | 7,553.68 | 7,166.60 | 7,130.25 | 29,330.41 |
| 294 | 14,437.87 | 13,846 | 14,314.57 | 14,033.43 | 56,631.87 |
| 295 | 10,790.92 | 10,407.40 | 10,732.30 | 10,393.31 | 42,323.93 |
| 296 | 16,215.86 | 16,094.90 | 15,463.57 | 15,334.17 | 63,108.50 |
| 297 | 3,203.58 | 3,099.67 | 3,036.54 | 2,935.81 | 12,275.60 |
| 298 | 2,574.31 | 2,461.75 | 2,241.12 | 2,462.82 | 9,740 |

| | | | | | |
|---|---|---|---|---|---|
| 299 | 2,959.10 | 2,750.49 | 2,654.59 | 2,745.97 | 11,110.15 |
| TOTAL | 74,018.28 | 72,199.07 | 71,729.77 | 71,198.11 | 289,145.23 |

STATE: SOUTH DAKOTA

| ZIP CODE | QUARTER 1 | QUARTER 2 | QUARTER 3 | QUARTER 4 | TOTAL GRAMS |
|---|---|---|---|---|---|
| 570 | 1,621.02 | 1,587.68 | 1,608.57 | 1,407.98 | 6,225.25 |
| 571 | 4,712.90 | 3,934.27 | 3,735.86 | 3,422.86 | 15,805.89 |
| 572 | 451.21 | 432.3 | 480.61 | 477.62 | 1,841.74 |
| 573 | 564.05 | 623.82 | 552.56 | 484.8 | 2,225.23 |
| 574 | 665.56 | 640.03 | 679.34 | 543.29 | 2,528.22 |
| 575 | 565.5 | 629.3 | 608.44 | 560.13 | 2,363.37 |
| 576 | 129.39 | 101.29 | 91.26 | 50.91 | 372.85 |
| 577 | 3,239.87 | 3,076.05 | 3,268.66 | 2,714.17 | 12,298.75 |
| TOTAL | 11,949.50 | 11,024.74 | 11,025.30 | 9,661.76 | 43,661.30 |

STATE: TENNESSEE

| ZIP CODE | QUARTER 1 | QUARTER 2 | QUARTER 3 | QUARTER 4 | TOTAL GRAMS |
|---|---|---|---|---|---|
| 370 | 22,459.51 | 21,834.32 | 21,306.41 | 20,350.03 | 85,950.27 |
| 371 | 20,342.28 | 19,965.42 | 19,109.79 | 18,609.54 | 78,027.03 |
| 372 | 13,822.61 | 12,952.04 | 11,843.87 | 11,199.44 | 49,817.96 |
| 373 | 16,373.73 | 15,438.64 | 15,275.91 | 14,650.82 | 61,739.10 |
| 374 | 6,316.81 | 5,820.79 | 5,480.82 | 5,517.11 | 23,135.53 |
| 376 | 13,400.54 | 13,007.26 | 12,268.83 | 11,361.41 | 50,038.04 |
| 377 | 11,289.56 | 10,150.92 | 9,726.21 | 8,899.79 | 40,066.48 |
| 378 | 19,284.74 | 17,171.19 | 16,151.79 | 14,795.78 | 67,403.50 |
| 379 | 11,559.75 | 10,125.15 | 9,320.17 | 9,010.27 | 40,015.34 |
| 380 | 5,492.46 | 5,127.45 | 4,940.56 | 4,963.20 | 20,523.67 |
| 381 | 34,717.15 | 35,703.86 | 33,426.78 | 28,283.93 | 132,131.72 |
| 382 | 4,153.27 | 3,843.67 | 3,850.60 | 3,374.96 | 15,222.50 |
| 383 | 9,892.81 | 9,486.20 | 8,848.19 | 8,347.96 | 36,575.16 |
| 384 | 5,758.30 | 5,387.65 | 4,981.67 | 5,055.88 | 21,183.50 |
| 385 | 11,364.37 | 10,574.92 | 10,286.96 | 9,903.02 | 42,129.27 |
| TOTAL | 206,227.89 | 196,589.48 | 186,818.56 | 174,323.14 | 763,959.07 |

STATE: TEXAS

| ZIP CODE | QUARTER 1 | QUARTER 2 | QUARTER 3 | QUARTER 4 | TOTAL GRAMS |
|---|---|---|---|---|---|
| 750 | 18,220.03 | 18,444.53 | 18,964.17 | 18,604.84 | 74,233.57 |
| 751 | 5,821.79 | 5,372.79 | 5,462.11 | 5,360.08 | 22,016.77 |
| 752 | 10,316.35 | 9,314.81 | 8,946.66 | 14,041.38 | 42,619.20 |
| 753 | 204.15 | 186.04 | 159.11 | 161.95 | 711.25 |
| 754 | 5,255.32 | 5,604.86 | 5,661.87 | 5,505.95 | 22,028 |
| 755 | 1,261.81 | 1,467.93 | 1,140.45 | 1,286.58 | 5,156.77 |
| 756 | 3,907.32 | 3,483.44 | 4,188.87 | 3,239.58 | 14,819.21 |
| 757 | 4,187.18 | 4,089.89 | 3,676.45 | 3,826.78 | 15,780.30 |
| 758 | 988.53 | 888.44 | 885.92 | 915.58 | 3,678.47 |
| 759 | 2,621.75 | 2,356.57 | 2,556.81 | 2,499.56 | 10,034.69 |
| 760 | 12,712.93 | 13,287.28 | 12,732.51 | 12,184.23 | 50,916.95 |
| 761 | 15,238.44 | 15,475.18 | 15,185.43 | 13,610.21 | 59,509.26 |
| 762 | 4,265.49 | 4,501.81 | 4,563.34 | 3,976.96 | 17,307.60 |
| 763 | 3,392.75 | 3,240.88 | 3,421.93 | 3,247.61 | 13,303.17 |
| 764 | 1,609.55 | 1,725.68 | 1,791.07 | 1,575.30 | 6,701.60 |
| 765 | 4,351.51 | 4,565.11 | 4,111.49 | 4,190.02 | 17,218.13 |
| 766 | 1,307.18 | 1,118.04 | 1,200.14 | 1,086.71 | 4,712.07 |
| 767 | 3,821.05 | 3,924.55 | 3,711.85 | 3,522.94 | 14,980.39 |
| 768 | 1,196.48 | 1,174.94 | 1,315.94 | 1,241.60 | 4,928.96 |

DRUG CODE:9300DRUG NAME:MORPHINE
STATE:TEXAS

| ZIP CODE | QUARTER 1 | QUARTER 2 | QUARTER 3 | QUARTER 4 | TOTAL GRAMS |
|---|---|---|---|---|---|

| | | | | | |
|---|---|---|---|---|---|
| 769 | 1,631.56 | 1,412.92 | 1,971.61 | 1,908.72 | 6,924.81 |
| 770 | 15,100.32 | 13,897.58 | 13,880.60 | 15,028.11 | 57,906.61 |
| 773 | 9,034.94 | 9,449.89 | 8,840.14 | 8,379.36 | 35,704.33 |
| 774 | 7,080.52 | 6,667.89 | 7,111 | 6,220.15 | 27,079.56 |
| 775 | 12,698.15 | 11,690.80 | 12,530.17 | 11,862.99 | 48,782.11 |
| 776 | 3,735.90 | 2,946.51 | 2,650.12 | 3,430.54 | 12,763.07 |
| 777 | 1,448.16 | 1,462.68 | 1,842.21 | 1,970.28 | 6,723.33 |
| 778 | 2,411.35 | 2,465.66 | 2,321.56 | 2,167.40 | 9,365.97 |
| 779 | 2,570.22 | 2,483.42 | 2,596.37 | 2,331.52 | 9,981.53 |
| 780 | 2,922.50 | 2,958.05 | 2,780.86 | 2,829.01 | 11,490.42 |
| 781 | 2,928.60 | 3,052.53 | 2,980.09 | 3,281.19 | 12,242.41 |
| 782 | 18,159.95 | 17,638.38 | 16,692.65 | 16,359.96 | 68,850.94 |
| 783 | 1,586.94 | 1,570.83 | 1,686.01 | 1,825.43 | 6,669.21 |
| 784 | 3,726.74 | 3,427.65 | 3,417.30 | 3,257.84 | 13,829.53 |
| 785 | 2,542.47 | 2,427.53 | 2,791.81 | 2,776.05 | 10,537.86 |
| 786 | 11,060.31 | 10,447.54 | 10,607.28 | 10,293.78 | 42,408.91 |
| 787 | 10,390.01 | 10,512.69 | 10,031.64 | 9,641.80 | 40,576.14 |
| 788 | 625.42 | 517.1 | 606.71 | 568.33 | 2,317.56 |
| 789 | 581.18 | 518.72 | 492.46 | 562.1 | 2,154.46 |
| 790 | 1,646.38 | 1,602.20 | 1,755.55 | 1,719.19 | 6,723.32 |
| 791 | 4,219.82 | 4,783.88 | 4,710.51 | 3,679.33 | 17,393.54 |
| 792 | 426.46 | 397.41 | 444.17 | 407.78 | 1,675.82 |
| 793 | 873.74 | 827.32 | 842.03 | 746.43 | 3,289.52 |
| 794 | 3,936.06 | 3,573.97 | 3,810.24 | 5,213.85 | 16,534.12 |
| 795 | 918.62 | 864.01 | 983.83 | 948.34 | 3,714.80 |
| 796 | 2,794.88 | 2,755.27 | 2,692.83 | 2,744.11 | 10,987.09 |
| 797 | 3,694.78 | 4,057.11 | 3,162.96 | 3,414.72 | 14,329.57 |
| 798 | 115.36 | 84.71 | 72.15 | 127.95 | 400.17 |
| 799 | 3,997.31 | 3,754.83 | 3,559.83 | 4,860.20 | 16,172.17 |
| TOTAL | 233,538.26 | 228,471.85 | 227,540.81 | 228,634.32 | 918,185.24 |

STATE: UTAH

| ZIP CODE | QUARTER 1 | QUARTER 2 | QUARTER 3 | QUARTER 4 | TOTAL GRAMS |
|---|---|---|---|---|---|
| 840 | 21,202.47 | 20,948.93 | 21,226.63 | 20,326.77 | 83,704.80 |
| 841 | 17,990.02 | 17,505.22 | 16,906.41 | 15,821.78 | 68,223.43 |
| 843 | 2,377.31 | 2,181.56 | 2,309.49 | 2,048.04 | 8,916.40 |
| 844 | 5,287.14 | 4,943.10 | 4,911.22 | 4,601.39 | 19,742.85 |
| 845 | 1,596.78 | 1,388.41 | 1,315.12 | 1,352.55 | 5,652.86 |
| 846 | 3,928.32 | 3,845.29 | 3,679.89 | 3,410.99 | 14,864.49 |
| 847 | 5,747.12 | 5,561.41 | 5,544.53 | 5,602.94 | 22,456 |
| TOTAL | 58,129.16 | 56,373.92 | 55,893.29 | 53,164.46 | 223,560.83 |

STATE: VERMONT

| ZIP CODE | QUARTER 1 | QUARTER 2 | QUARTER 3 | QUARTER 4 | TOTAL GRAMS |
|---|---|---|---|---|---|
| 50 | 1,483.72 | 1,629.66 | 1,358.82 | 1,385.65 | 5,857.85 |
| 51 | 620.71 | 577.31 | 547.02 | 538.74 | 2,283.78 |
| 52 | 649.28 | 633.5 | 620.09 | 562.64 | 2,465.51 |
| 53 | 837.01 | 699.79 | 707.94 | 712.72 | 2,957.46 |
| 54 | 4,715.42 | 4,471.13 | 4,333.80 | 4,525.19 | 18,045.54 |
| 56 | 1,174.55 | 1,081.75 | 1,039.59 | 897.16 | 4,193.05 |
| 57 | 1,547.37 | 1,603.67 | 1,484.01 | 1,469.09 | 6,104.14 |
| 58 | 842.97 | 801.89 | 791.39 | 824.23 | 3,260.48 |
| TOTAL | 11,871.03 | 11,498.70 | 10,882.66 | 10,915.42 | 45,167.81 |

STATE: VIRGIN ISLANDS

| ZIP CODE | QUARTER 1 | QUARTER 2 | QUARTER 3 | QUARTER 4 | TOTAL GRAMS |
|---|---|---|---|---|---|
| 8 | 133.25 | 93.11 | 103.7 | 86.71 | 416.77 |
| TOTAL | 133.25 | 93.11 | 103.7 | 86.71 | 416.77 |

STATE: VIRGINIA

| ZIP CODE | QUARTER 1 | QUARTER 2 | QUARTER 3 | QUARTER 4 | TOTAL GRAMS |
|---|---|---|---|---|---|
| 201 | 6,641.30 | 6,369.32 | 6,794.96 | 6,923.86 | 26,729.44 |
| 220 | 4,112.21 | 3,759.74 | 3,530.47 | 3,453.07 | 14,855.49 |
| 221 | 3,437.48 | 3,556.64 | 3,249.59 | 3,228.68 | 13,472.39 |
| 222 | 919.59 | 785.99 | 720.76 | 754.23 | 3,180.57 |
| 223 | 1,582.49 | 1,665.52 | 1,473.36 | 1,489.12 | 6,210.49 |
| 224 | 3,308.04 | 2,892.26 | 2,950.47 | 3,055.08 | 12,205.85 |
| 225 | 1,033.03 | 1,040.60 | 907.92 | 900.45 | 3,882 |
| 226 | 2,981.98 | 3,185.38 | 3,205.37 | 2,882.90 | 12,255.63 |
| 227 | 623.07 | 655.48 | 618.22 | 554.74 | 2,451.51 |

DRUG CODE:9300DRUG NAME:MORPHINE
STATE:VIRGINIA

| ZIP CODE | QUARTER 1 | QUARTER 2 | QUARTER 3 | QUARTER 4 | TOTAL GRAMS |
|---|---|---|---|---|---|
| 228 | 2,321.83 | 2,149.45 | 2,390.71 | 2,173.89 | 9,035.88 |
| 229 | 2,752.86 | 2,867.41 | 2,717.66 | 2,753.13 | 11,091.06 |
| 230 | 2,557.59 | 2,268.39 | 2,469.86 | 2,135.72 | 9,431.56 |
| 231 | 3,720.15 | 3,706.90 | 3,774.66 | 3,306.79 | 14,508.50 |
| 232 | 9,980.59 | 9,597.95 | 9,116.05 | 9,315.43 | 38,010.02 |
| 233 | 2,736.16 | 2,471.92 | 2,676.27 | 2,671.62 | 10,555.97 |
| 234 | 6,406.41 | 6,277.45 | 6,193.21 | 6,042.20 | 24,919.27 |
| 235 | 2,587.90 | 2,388.66 | 2,572.29 | 2,396.14 | 9,944.99 |
| 236 | 4,826.26 | 4,354.71 | 4,782.94 | 4,621.35 | 18,585.26 |
| 237 | 2,028.11 | 1,911.18 | 2,024.90 | 2,015.11 | 7,979.30 |
| 238 | 3,821.44 | 3,688.20 | 3,608.34 | 3,797.11 | 14,915.09 |
| 239 | 1,048.74 | 897.62 | 808.17 | 833.22 | 3,587.75 |
| 240 | 4,725.39 | 4,366.05 | 4,340.46 | 4,098.38 | 17,530.28 |
| 241 | 6,101.46 | 5,786.49 | 5,260.88 | 5,187.79 | 22,336.62 |
| 242 | 5,070.40 | 4,651.24 | 4,823.03 | 4,562.52 | 19,107.19 |
| 243 | 2,896.52 | 2,811.38 | 2,679.33 | 2,723.40 | 11,110.63 |
| 244 | 1,445.97 | 1,502.98 | 1,433.48 | 1,359.73 | 5,742.16 |
| 245 | 3,849.36 | 3,645.55 | 3,501.33 | 3,683.35 | 14,679.59 |
| 246 | 1,337.82 | 1,307.81 | 1,256.92 | 1,144.52 | 5,047.07 |
| TOTAL | 94,854.15 | 90,562.27 | 89,881.61 | 88,063.53 | 363,361.56 |

STATE: WASHINGTON

| ZIP CODE | QUARTER 1 | QUARTER 2 | QUARTER 3 | QUARTER 4 | TOTAL GRAMS |
|---|---|---|---|---|---|
| 980 | 18,211.97 | 17,351.78 | 16,971.33 | 16,442.37 | 68,977.45 |
| 981 | 15,199.60 | 13,933.96 | 13,735.23 | 12,818.57 | 55,687.36 |
| 982 | 15,577.59 | 15,157.99 | 15,474.93 | 14,345.78 | 60,556.29 |
| 983 | 13,618.50 | 13,762.81 | 13,479.30 | 12,655.29 | 53,515.90 |
| 984 | 9,823.77 | 9,409.89 | 9,098.12 | 9,015.81 | 37,347.59 |
| 985 | 9,332.88 | 8,575.40 | 8,505.79 | 7,431.45 | 33,845.52 |
| 986 | 16,728.81 | 16,170.52 | 15,262.06 | 15,129.65 | 63,291.04 |
| 988 | 3,647.28 | 3,725.98 | 3,400 | 3,521.77 | 14,295.03 |
| 989 | 4,573.55 | 4,624.57 | 4,287.64 | 3,842.09 | 17,327.85 |
| 990 | 1,693.25 | 1,771.03 | 1,855.96 | 1,766.57 | 7,086.81 |
| 991 | 2,863.31 | 2,684.58 | 2,884.44 | 2,634.67 | 11,067 |
| 992 | 12,425.05 | 12,601.39 | 12,421.31 | 12,086.19 | 49,533.94 |
| 993 | 8,809.03 | 8,651.40 | 7,961.51 | 7,346.99 | 32,768.93 |
| 994 | 1,278.25 | 1,161.84 | 1,154.17 | 1,190.68 | 4,784.94 |
| TOTAL | 133,782.84 | 129,583.14 | 126,491.79 | 120,227.88 | 510,085.65 |

STATE: WEST VIRGINIA

| ZIP CODE | QUARTER 1 | QUARTER 2 | QUARTER 3 | QUARTER 4 | TOTAL GRAMS |
|---|---|---|---|---|---|
| 247 | 791.83 | 843.26 | 837.53 | 1,000.89 | 3,473.51 |
| 248 | 456.16 | 356.85 | 412.25 | 298.24 | 1,523.50 |
| 249 | 751.19 | 679.36 | 717.24 | 680.06 | 2,827.85 |
| 250 | 796.52 | 714.55 | 636.91 | 635.14 | 2,783.12 |

| | | | | | |
|---|---|---|---|---|---|
| 251 | 686.28 | 671.69 | 547.64 | 567.76 | 2,473.37 |
| 252 | 640.94 | 673.42 | 562.63 | 468.93 | 2,345.92 |
| 253 | 1,910.12 | 1,885.62 | 1,982.08 | 1,621.70 | 7,399.52 |
| 254 | 5,063.64 | 4,849.62 | 4,340.40 | 4,684.99 | 18,938.65 |
| 255 | 1,569.72 | 1,503.21 | 1,504.99 | 1,307.42 | 5,885.34 |
| 256 | 630.14 | 628.23 | 570.64 | 495.44 | 2,324.45 |
| 257 | 2,267.21 | 2,296.47 | 2,069.39 | 1,776.56 | 8,409.63 |
| 258 | 1,637.50 | 1,740.05 | 1,662.45 | 1,334.63 | 6,374.63 |
| 259 | 540.95 | 488.56 | 487.63 | 457.25 | 1,974.39 |
| 260 | 2,808.15 | 2,686.82 | 2,546.55 | 2,561.83 | 10,603.35 |
| 261 | 1,083.22 | 1,066.50 | 1,043 | 897.8 | 4,090.52 |
| 262 | 1,091.64 | 1,056.68 | 985.24 | 895.04 | 4,028.60 |
| 263 | 2,077.95 | 2,107.62 | 2,032.84 | 2,030.58 | 8,248.99 |
| 264 | 438.61 | 525.48 | 508.19 | 485.51 | 1,957.79 |
| 265 | 2,123.97 | 2,072.23 | 2,107.53 | 1,811.80 | 8,115.53 |
| 266 | 568.9 | 565.51 | 518.87 | 520.75 | 2,174.03 |
| 267 | 782.27 | 889.31 | 818.57 | 934.9 | 3,425.05 |
| 268 | 393.14 | 357.49 | 374.47 | 387.45 | 1,512.55 |
| TOTAL | 29,110.05 | 28,658.53 | 27,267.04 | 25,854.67 | 110,890.29 |

STATE: WISCONSIN

| ZIP CODE | QUARTER 1 | QUARTER 2 | QUARTER 3 | QUARTER 4 | TOTAL GRAMS |
|---|---|---|---|---|---|
| 530 | 10,476.65 | 10,085.40 | 10,108 | 9,363.09 | 40,033.14 |
| 531 | 11,917.34 | 11,914.58 | 10,965.76 | 9,993.10 | 44,790.78 |

DRUG CODE:9300DRUG NAME:MORPHINE
STATE:WISCONSIN

| ZIP CODE | QUARTER 1 | QUARTER 2 | QUARTER 3 | QUARTER 4 | TOTAL GRAMS |
|---|---|---|---|---|---|
| 532 | 21,451.81 | 20,286.29 | 19,080.65 | 17,577.45 | 78,396.20 |
| 534 | 2,389.56 | 1,994.95 | 1,908.23 | 1,782.41 | 8,075.15 |
| 535 | 9,942.03 | 9,974.90 | 9,280.73 | 8,572.81 | 37,770.47 |
| 537 | 8,805.49 | 8,917.66 | 8,803.19 | 7,696.53 | 34,222.87 |
| 538 | 845.08 | 882.98 | 835.74 | 738.54 | 3,302.34 |
| 539 | 5,232.39 | 5,006.67 | 5,123 | 4,351.79 | 19,713.85 |
| 540 | 1,338.73 | 1,113.68 | 1,191.69 | 1,093.08 | 4,737.18 |
| 541 | 1,870.28 | 1,943.34 | 1,875.04 | 1,887.63 | 7,576.29 |
| 542 | 1,944.47 | 1,379.26 | 1,243.33 | 1,260.67 | 5,827.73 |
| 543 | 2,814.45 | 2,683.16 | 2,836.52 | 2,398.79 | 10,732.92 |
| 544 | 4,495.25 | 4,170.70 | 4,110.60 | 3,886.95 | 16,663.50 |
| 545 | 1,514.55 | 1,566.82 | 1,526.92 | 1,260.51 | 5,868.80 |
| 546 | 4,271.99 | 4,145.74 | 4,062.08 | 3,922.87 | 16,402.68 |
| 547 | 3,578.31 | 3,538.83 | 3,502.95 | 3,568.99 | 14,189.08 |
| 548 | 2,428.36 | 2,417.23 | 2,590.35 | 2,647.10 | 10,083.04 |
| 549 | 7,135.09 | 6,999.06 | 6,561.17 | 6,110.72 | 26,806.04 |
| TOTAL | 102,451.83 | 99,021.25 | 95,605.95 | 88,113.03 | 385,192.06 |

STATE: WYOMING

| ZIP CODE | QUARTER 1 | QUARTER 2 | QUARTER 3 | QUARTER 4 | TOTAL GRAMS |
|---|---|---|---|---|---|
| 820 | 2,431.32 | 2,080.43 | 2,260.78 | 2,105.21 | 8,877.74 |
| 822 | 260.91 | 250.3 | 309.73 | 291.02 | 1,111.96 |
| 823 | 166.58 | 162.29 | 216.5 | 187.12 | 732.49 |
| 824 | 1,117.28 | 1,088.97 | 1,050.17 | 1,102.63 | 4,359.05 |
| 825 | 840.47 | 901.05 | 737.19 | 791.7 | 3,270.41 |
| 826 | 1,935.71 | 1,502.71 | 1,571.35 | 1,381.91 | 6,391.68 |
| 827 | 524.92 | 466.98 | 408.45 | 353.33 | 1,753.68 |
| 828 | 1,593.76 | 1,481.45 | 1,657.35 | 908.07 | 5,640.63 |
| 829 | 1,891.31 | 1,850.17 | 1,805.46 | 1,837.74 | 7,384.68 |
| 830 | 45.52 | 55.54 | 59.57 | 76.24 | 236.87 |
| 831 | 347.81 | 399.81 | 368.26 | 298.33 | 1,414.21 |

| | | | | | |
|---|---|---|---|---|---|
| TOTAL | 11,155.59 | 10,239.70 | 10,444.81 | 9,333.30 | 41,173.40 |

DRUG CODE:9630DRUG NAME:OPIUM TINCTURE (LAUDANUM)

STATE: ARIZONA

| ZIP CODE | QUARTER 1 | QUARTER 2 | QUARTER 3 | QUARTER 4 | TOTAL GRAMS |
|---|---|---|---|---|---|
| 857 | 0 | 0 | 0 | 9.46 | 9.46 |
| TOTAL | 0 | 0 | 0 | 9.46 | 9.46 |

STATE: CALIFORNIA

| ZIP CODE | QUARTER 1 | QUARTER 2 | QUARTER 3 | QUARTER 4 | TOTAL GRAMS |
|---|---|---|---|---|---|
| 900 | 0 | 1.18 | 0 | 0 | 1.18 |
| 930 | 0 | 0 | 1.18 | 0 | 1.18 |
| 934 | 0 | 0 | 1.18 | 0 | 1.18 |
| 941 | 0 | 0 | 9.46 | 0 | 9.46 |
| 945 | 0 | 14.19 | 0 | 0 | 14.19 |
| 950 | 0 | 9.46 | 0 | 0 | 9.46 |
| 958 | 4.73 | 4.73 | 0 | 0 | 9.46 |
| TOTAL | 4.73 | 29.56 | 11.82 | 0 | 46.11 |

STATE: COLORADO

| ZIP CODE | QUARTER 1 | QUARTER 2 | QUARTER 3 | QUARTER 4 | TOTAL GRAMS |
|---|---|---|---|---|---|
| 804 | 0 | 0 | 1.18 | 0 | 1.18 |
| TOTAL | 0 | 0 | 1.18 | 0 | 1.18 |

STATE: DISTRICT OF COLUMBIA

| ZIP CODE | QUARTER 1 | QUARTER 2 | QUARTER 3 | QUARTER 4 | TOTAL GRAMS |
|---|---|---|---|---|---|
| 200 | 0 | 0 | 1.18 | 0 | 1.18 |
| TOTAL | 0 | 0 | 1.18 | 0 | 1.18 |

STATE: FLORIDA

| ZIP CODE | QUARTER 1 | QUARTER 2 | QUARTER 3 | QUARTER 4 | TOTAL GRAMS |
|---|---|---|---|---|---|
| 321 | 14.19 | 4.73 | 0 | 9.46 | 28.38 |
| 329 | 0 | 0 | 0 | 4.73 | 4.73 |
| 331 | 0 | 0 | 4.73 | 0 | 4.73 |
| TOTAL | 14.19 | 4.73 | 4.73 | 14.19 | 37.84 |

STATE: IDAHO

| ZIP CODE | QUARTER 1 | QUARTER 2 | QUARTER 3 | QUARTER 4 | TOTAL GRAMS |
|---|---|---|---|---|---|
| 838 | 0 | 1.18 | 0 | 0 | 1.18 |
| TOTAL | 0 | 1.18 | 0 | 0 | 1.18 |

STATE: ILLINOIS

| ZIP CODE | QUARTER 1 | QUARTER 2 | QUARTER 3 | QUARTER 4 | TOTAL GRAMS |
|---|---|---|---|---|---|

DRUG CODE:9630DRUG NAME:OPIUM TINCTURE (LAUDANUM)

STATE:ILLINOIS

| ZIP CODE | QUARTER 1 | QUARTER 2 | QUARTER 3 | QUARTER 4 | TOTAL GRAMS |
|---|---|---|---|---|---|
| 600 | 4.73 | 0 | 0 | 7.09 | 11.82 |
| 601 | 0 | 2.36 | 0 | 0 | 2.36 |
| 605 | 0 | 1.18 | 0 | 0 | 1.18 |
| 606 | 0 | 1.18 | 0 | 0 | 1.18 |
| 617 | 1.18 | 1.18 | 0 | 4.73 | 7.09 |
| 618 | 0 | 0 | 0 | 1.18 | 1.18 |
| 626 | 4.73 | 0 | 0 | 4.73 | 9.46 |
| TOTAL | 10.64 | 5.9 | 0 | 17.73 | 34.27 |

STATE: INDIANA

| ZIP CODE | QUARTER 1 | QUARTER 2 | QUARTER 3 | QUARTER 4 | TOTAL GRAMS |
|---|---|---|---|---|---|
| 461 | 2.36 | 11.82 | 14.19 | 4.73 | 33.1 |

| | | | | | | |
|---|---|---|---|---|---|---|
| | 462 | 8.27 | 0 | 0 | 5.91 | 14.18 |
| | 463 | 0 | 18.92 | 0 | 1.18 | 20.1 |
| | 468 | 4.73 | 4.73 | 0 | 0 | 9.46 |
| | 479 | 0 | 0 | 0 | 4.73 | 4.73 |
| TOTAL | | 15.36 | 35.47 | 14.19 | 16.55 | 81.57 |

STATE: KANSAS

| ZIP CODE | | QUARTER 1 | QUARTER 2 | QUARTER 3 | QUARTER 4 | TOTAL GRAMS |
|---|---|---|---|---|---|---|
| | 660 | 0 | 0 | 0 | 4.73 | 4.73 |
| | 672 | 0 | 0 | 0 | 1.18 | 1.18 |
| TOTAL | | 0 | 0 | 0 | 5.91 | 5.91 |

STATE: MAINE

| ZIP CODE | | QUARTER 1 | QUARTER 2 | QUARTER 3 | QUARTER 4 | TOTAL GRAMS |
|---|---|---|---|---|---|---|
| | 46 | 0 | 0 | 4.73 | 0 | 4.73 |
| | 47 | 0 | 1.18 | 0 | 0 | 1.18 |
| TOTAL | | 0 | 1.18 | 4.73 | 0 | 5.91 |

STATE: MARYLAND

| ZIP CODE | | QUARTER 1 | QUARTER 2 | QUARTER 3 | QUARTER 4 | TOTAL GRAMS |
|---|---|---|---|---|---|---|
| | 207 | 0 | 0 | 0 | 1.18 | 1.18 |
| | 212 | 1.18 | 4.72 | 0 | 0 | 5.9 |
| | 216 | 1.18 | 2.36 | 0 | 0 | 3.54 |
| TOTAL | | 2.36 | 7.08 | 0 | 1.18 | 10.62 |

STATE: MASSACHUSETTS

| ZIP CODE | | QUARTER 1 | QUARTER 2 | QUARTER 3 | QUARTER 4 | TOTAL GRAMS |
|---|---|---|---|---|---|---|
| | 10 | 0 | 0 | 0 | 4.73 | 4.73 |
| | 17 | 0 | 0 | 4.73 | 0 | 4.73 |
| | 19 | 0 | 33.11 | 47.3 | 42.57 | 122.98 |
| | 20 | 5.91 | 4.73 | 0 | 4.73 | 15.37 |
| | 21 | 0 | 0 | 1.18 | 1.18 | 2.36 |
| TOTAL | | 5.91 | 37.84 | 53.21 | 53.21 | 150.17 |

STATE: MICHIGAN

| ZIP CODE | | QUARTER 1 | QUARTER 2 | QUARTER 3 | QUARTER 4 | TOTAL GRAMS |
|---|---|---|---|---|---|---|
| | 480 | 0 | 9.46 | 0 | 14.19 | 23.65 |
| | 481 | 0 | 1.18 | 2.36 | 10.64 | 14.18 |
| | 489 | 0 | 9.46 | 0 | 0 | 9.46 |
| TOTAL | | 0 | 20.1 | 2.36 | 24.83 | 47.29 |

STATE: MINNESOTA

| ZIP CODE | | QUARTER 1 | QUARTER 2 | QUARTER 3 | QUARTER 4 | TOTAL GRAMS |
|---|---|---|---|---|---|---|
| | 559 | 0 | 0 | 1.18 | 0 | 1.18 |
| TOTAL | | 0 | 0 | 1.18 | 0 | 1.18 |

STATE: MISSOURI

| ZIP CODE | | QUARTER 1 | QUARTER 2 | QUARTER 3 | QUARTER 4 | TOTAL GRAMS |
|---|---|---|---|---|---|---|
| | 630 | 0 | 0 | 4.73 | 15.37 | 20.1 |
| | 648 | 0 | 0 | 0 | 1.18 | 1.18 |
| | 657 | 0 | 0 | 0 | 4.73 | 4.73 |
| TOTAL | | 0 | 0 | 4.73 | 21.28 | 26.01 |

STATE: NEBRASKA

| ZIP CODE | | QUARTER 1 | QUARTER 2 | QUARTER 3 | QUARTER 4 | TOTAL GRAMS |
|---|---|---|---|---|---|---|
| | 688 | 0 | 0 | 14.19 | 0 | 14.19 |
| TOTAL | | 0 | 0 | 14.19 | 0 | 14.19 |

STATE: NEW HAMPSHIRE

| ZIP CODE | | QUARTER 1 | QUARTER 2 | QUARTER 3 | QUARTER 4 | TOTAL GRAMS |
|---|---|---|---|---|---|---|

| | | | | | |
|---|---|---|---|---|---|
| 32 | 0 | 1.18 | 0 | 0 | 1.18 |
| TOTAL | 0 | 1.18 | 0 | 0 | 1.18 |

STATE: NEW JERSEY

| ZIP CODE | QUARTER 1 | QUARTER 2 | QUARTER 3 | QUARTER 4 | TOTAL GRAMS |
|---|---|---|---|---|---|

DRUG CODE:9630DRUG NAME:OPIUM TINCTURE (LAUDANUM)
STATE:NEW JERSEY

| ZIP CODE | QUARTER 1 | QUARTER 2 | QUARTER 3 | QUARTER 4 | TOTAL GRAMS |
|---|---|---|---|---|---|
| 70 | 0 | 0 | 0 | 1.18 | 1.18 |
| 77 | 2.36 | 0 | 0 | 0 | 2.36 |
| 79 | 0 | 1.18 | 0 | 1.18 | 2.36 |
| 80 | 0 | 1.18 | 0 | 0 | 1.18 |
| 88 | 0 | 0 | 1.18 | 1.18 | 2.36 |
| TOTAL | 2.36 | 2.36 | 1.18 | 3.54 | 9.44 |

STATE: NEW MEXICO

| ZIP CODE | QUARTER 1 | QUARTER 2 | QUARTER 3 | QUARTER 4 | TOTAL GRAMS |
|---|---|---|---|---|---|
| 871 | 0 | 0 | 0 | 4.73 | 4.73 |
| TOTAL | 0 | 0 | 0 | 4.73 | 4.73 |

STATE: NEW YORK

| ZIP CODE | QUARTER 1 | QUARTER 2 | QUARTER 3 | QUARTER 4 | TOTAL GRAMS |
|---|---|---|---|---|---|
| 117 | 1.18 | 0 | 0 | 0 | 1.18 |
| 130 | 1.18 | 0 | 0 | 0 | 1.18 |
| 140 | 0 | 0 | 1.18 | 0 | 1.18 |
| TOTAL | 2.36 | 0 | 1.18 | 0 | 3.54 |

STATE: OHIO

| ZIP CODE | QUARTER 1 | QUARTER 2 | QUARTER 3 | QUARTER 4 | TOTAL GRAMS |
|---|---|---|---|---|---|
| 430 | 4.73 | 0 | 0 | 3.54 | 8.27 |
| 431 | 0 | 0 | 0 | 2.36 | 2.36 |
| 432 | 0 | 9.46 | 0 | 1.18 | 10.64 |
| 436 | 0 | 0 | 4.73 | 0 | 4.73 |
| 440 | 0 | 1.18 | 1.18 | 3.54 | 5.9 |
| 450 | 0 | 4.73 | 0 | 0 | 4.73 |
| 452 | 1.18 | 0 | 0 | 1.18 | 2.36 |
| 453 | 4.73 | 4.73 | 4.73 | 2.36 | 16.55 |
| 454 | 0 | 4.73 | 4.73 | 7.09 | 16.55 |
| 456 | 4.73 | 9.46 | 9.46 | 0 | 23.65 |
| 457 | 0 | 0 | 1.18 | 0 | 1.18 |
| TOTAL | 15.37 | 34.29 | 26.01 | 21.25 | 96.92 |

STATE: OREGON

| ZIP CODE | QUARTER 1 | QUARTER 2 | QUARTER 3 | QUARTER 4 | TOTAL GRAMS |
|---|---|---|---|---|---|
| 972 | 0 | 0 | 0 | 3.54 | 3.54 |
| 973 | 0 | 0 | 0 | 1.18 | 1.18 |
| TOTAL | 0 | 0 | 0 | 4.72 | 4.72 |

STATE: PENNSYLVANIA

| ZIP CODE | QUARTER 1 | QUARTER 2 | QUARTER 3 | QUARTER 4 | TOTAL GRAMS |
|---|---|---|---|---|---|
| 152 | 0 | 0 | 0 | 1.18 | 1.18 |
| 170 | 4.73 | 0 | 0 | 0 | 4.73 |
| 190 | 1.18 | 1.18 | 0 | 4.72 | 7.08 |
| TOTAL | 5.91 | 1.18 | 0 | 5.9 | 12.99 |

STATE: TEXAS

| ZIP CODE | QUARTER 1 | QUARTER 2 | QUARTER 3 | QUARTER 4 | TOTAL GRAMS |
|---|---|---|---|---|---|
| 750 | 1.18 | 1.18 | 0 | 0 | 2.36 |

| | | | | | |
|---|---|---|---|---|---|
| 760 | 0 | 0 | 1.18 | 1.18 | 2.36 |
| 761 | 0 | 0 | 7.08 | 4.72 | 11.8 |
| 770 | 1.18 | 0 | 1.18 | 0 | 2.36 |
| 790 | 0 | 0 | 0 | 2.36 | 2.36 |
| TOTAL | 2.36 | 1.18 | 9.44 | 8.26 | 21.24 |

STATE: VERMONT

| ZIP CODE | QUARTER 1 | QUARTER 2 | QUARTER 3 | QUARTER 4 | TOTAL GRAMS |
|---|---|---|---|---|---|
| 56 | 0 | 0 | 2.36 | 0 | 2.36 |
| TOTAL | 0 | 0 | 2.36 | 0 | 2.36 |

STATE: VIRGINIA

| ZIP CODE | QUARTER 1 | QUARTER 2 | QUARTER 3 | QUARTER 4 | TOTAL GRAMS |
|---|---|---|---|---|---|
| 201 | 0 | 3.54 | 3.54 | 0 | 7.08 |
| 220 | 0 | 0 | 2.36 | 2.36 | 4.72 |
| 221 | 0 | 1.18 | 0 | 0 | 1.18 |
| 223 | 0 | 4.72 | 0 | 0 | 4.72 |
| TOTAL | 0 | 9.44 | 5.9 | 2.36 | 17.7 |

STATE: WASHINGTON

| ZIP CODE | QUARTER 1 | QUARTER 2 | QUARTER 3 | QUARTER 4 | TOTAL GRAMS |
|---|---|---|---|---|---|
| 980 | 2.36 | 0 | 0 | 1.18 | 3.54 |
| 981 | 0 | 0 | 2.36 | 0 | 2.36 |
| 984 | 1.18 | 2.36 | 0 | 1.18 | 4.72 |
| 985 | 0 | 2.36 | 0 | 0 | 2.36 |

DRUG CODE:9630DRUG NAME:OPIUM TINCTURE (LAUDANUM)
STATE:WASHINGTON

| ZIP CODE | QUARTER 1 | QUARTER 2 | QUARTER 3 | QUARTER 4 | TOTAL GRAMS |
|---|---|---|---|---|---|
| 986 | 0 | 0 | 3.54 | 3.54 | 7.08 |
| TOTAL | 3.54 | 4.72 | 5.9 | 5.9 | 20.06 |

STATE: WEST VIRGINIA

| ZIP CODE | QUARTER 1 | QUARTER 2 | QUARTER 3 | QUARTER 4 | TOTAL GRAMS |
|---|---|---|---|---|---|
| 253 | 1.18 | 0 | 0 | 2.36 | 3.54 |
| 258 | 4.73 | 4.73 | 0 | 4.73 | 14.19 |
| 263 | 0 | 0 | 0 | 1.18 | 1.18 |
| 265 | 0 | 0 | 0 | 1.18 | 1.18 |
| TOTAL | 5.91 | 4.73 | 0 | 9.45 | 20.09 |

STATE: WISCONSIN

| ZIP CODE | QUARTER 1 | QUARTER 2 | QUARTER 3 | QUARTER 4 | TOTAL GRAMS |
|---|---|---|---|---|---|
| 537 | 0 | 0 | 0 | 1.18 | 1.18 |
| TOTAL | 0 | 0 | 0 | 1.18 | 1.18 |

DRUG CODE:9639DRUG NAME:OPIUM POWDERED
STATE: ALABAMA

| ZIP CODE | QUARTER 1 | QUARTER 2 | QUARTER 3 | QUARTER 4 | TOTAL GRAMS |
|---|---|---|---|---|---|
| 350 | 5.04 | 11.52 | 6.12 | 7.92 | 30.6 |
| 351 | 2.88 | 2.88 | 2.16 | 3.6 | 11.52 |
| 352 | 29.16 | 29.16 | 35.28 | 42.12 | 135.72 |
| 354 | 6.12 | 4.68 | 3.6 | 3.24 | 17.64 |
| 355 | 0.72 | 0.36 | 0.72 | 1.44 | 3.24 |
| 356 | 9.36 | 10.08 | 9.72 | 6.48 | 35.64 |
| 357 | 0.72 | 2.88 | 1.8 | 1.08 | 6.48 |
| 358 | 14.04 | 5.76 | 14.04 | 15.12 | 48.96 |
| 359 | 4.68 | 3.6 | 2.52 | 1.8 | 12.6 |
| 360 | 0 | 0 | 0 | 0.36 | 0.36 |

| | | | | | |
|---|---|---|---|---|---|
| 361 | 4.68 | 5.4 | 1.44 | 3.96 | 15.48 |
| 362 | 0 | 0.72 | 1.08 | 0.36 | 2.16 |
| 363 | 0 | 1.8 | 2.16 | 0.36 | 4.32 |
| 364 | 0 | 0.36 | 0 | 0 | 0.36 |
| 365 | 5.4 | 4.68 | 7.2 | 6.84 | 24.12 |
| 366 | 6.84 | 5.4 | 8.28 | 7.56 | 28.08 |
| 367 | 0.72 | 0.36 | 0.72 | 0 | 1.8 |
| TOTAL | 90.36 | 89.64 | 96.84 | 102.24 | 379.08 |

STATE: ALASKA

| ZIP CODE | QUARTER 1 | QUARTER 2 | QUARTER 3 | QUARTER 4 | TOTAL GRAMS |
|---|---|---|---|---|---|
| 995 | 5.76 | 10.8 | 6.12 | 14.76 | 37.44 |
| 996 | 4.32 | 5.4 | 4.32 | 5.4 | 19.44 |
| 997 | 3.6 | 1.44 | 3.6 | 2.16 | 10.8 |
| 998 | 0.72 | 1.44 | 2.16 | 2.16 | 6.48 |
| 999 | 0 | 2.16 | 0.72 | 0.72 | 3.6 |
| TOTAL | 14.4 | 21.24 | 16.92 | 25.2 | 77.76 |

STATE: ARIZONA

| ZIP CODE | QUARTER 1 | QUARTER 2 | QUARTER 3 | QUARTER 4 | TOTAL GRAMS |
|---|---|---|---|---|---|
| 850 | 96.84 | 90.72 | 136.08 | 105.12 | 428.76 |
| 851 | 12.96 | 9.72 | 3.24 | 0.72 | 26.64 |
| 852 | 59.4 | 80.28 | 103.68 | 117.36 | 360.72 |
| 853 | 18.36 | 29.52 | 26.64 | 26.64 | 101.16 |
| 855 | 0 | 0.36 | 0 | 0 | 0.36 |
| 856 | 0.36 | 1.08 | 0.72 | 0.72 | 2.88 |
| 857 | 28.8 | 19.08 | 20.16 | 29.16 | 97.2 |
| 859 | 1.08 | 6.84 | 1.44 | 0.36 | 9.72 |
| 860 | 0 | 0.72 | 1.08 | 0 | 1.8 |
| 863 | 17.64 | 7.92 | 13.68 | 24.12 | 63.36 |
| 864 | 5.04 | 4.68 | 2.16 | 3.96 | 15.84 |
| TOTAL | 240.48 | 250.92 | 308.88 | 308.16 | 1,108.44 |

STATE: ARKANSAS

| ZIP CODE | QUARTER 1 | QUARTER 2 | QUARTER 3 | QUARTER 4 | TOTAL GRAMS |
|---|---|---|---|---|---|
| 716 | 6.12 | 2.88 | 3.96 | 2.88 | 15.84 |
| 717 | 0 | 0 | 1.08 | 0 | 1.08 |
| 718 | 1.44 | 1.08 | 0 | 0.36 | 2.88 |
| 719 | 2.16 | 0.72 | 2.16 | 4.32 | 9.36 |
| 720 | 1.8 | 0.72 | 2.88 | 0.36 | 5.76 |
| 721 | 25.2 | 30.96 | 28.8 | 27.36 | 112.32 |
| 722 | 25.92 | 30.24 | 29.88 | 29.88 | 115.92 |
| 723 | 0.36 | 1.08 | 0 | 0 | 1.44 |

DRUG CODE:9639DRUG NAME:OPIUM POWDERED
STATE:ARKANSAS

| ZIP CODE | QUARTER 1 | QUARTER 2 | QUARTER 3 | QUARTER 4 | TOTAL GRAMS |
|---|---|---|---|---|---|
| 724 | 13.68 | 14.4 | 10.8 | 12.24 | 51.12 |
| 725 | 2.88 | 2.52 | 3.24 | 1.8 | 10.44 |
| 726 | 2.16 | 0.36 | 4.32 | 1.44 | 8.28 |
| 727 | 7.56 | 18.72 | 14.4 | 11.52 | 52.2 |
| 728 | 0 | 0 | 0 | 0.36 | 0.36 |
| 729 | 1.08 | 1.08 | 0 | 2.16 | 4.32 |
| TOTAL | 90.36 | 104.76 | 101.52 | 94.68 | 391.32 |

STATE: CALIFORNIA

| ZIP CODE | QUARTER 1 | QUARTER 2 | QUARTER 3 | QUARTER 4 | TOTAL GRAMS |
|---|---|---|---|---|---|
| 900 | 15.12 | 23.76 | 21.6 | 10.44 | 70.92 |

| | | | | | |
|---|---|---|---|---|---|
| 902 | 5.4 | 7.56 | 10.08 | 3.6 | 26.64 |
| 903 | 0 | 0 | 0.36 | 0 | 0.36 |
| 904 | 3.96 | 7.2 | 10.44 | 9 | 30.6 |
| 905 | 0 | 1.44 | 0.72 | 2.52 | 4.68 |
| 906 | 1.08 | 1.8 | 6.12 | 1.08 | 10.08 |
| 907 | 12.24 | 14.76 | 10.44 | 6.12 | 43.56 |
| 908 | 2.88 | 2.88 | 1.44 | 7.2 | 14.4 |
| 910 | 5.04 | 6.12 | 3.24 | 8.64 | 23.04 |
| 911 | 2.88 | 5.04 | 3.6 | 3.6 | 15.12 |
| 912 | 1.44 | 0.72 | 2.16 | 3.6 | 7.92 |
| 913 | 9.36 | 8.28 | 10.08 | 12.24 | 39.96 |
| 914 | 0.36 | 0.36 | 0.72 | 0.36 | 1.8 |
| 915 | 1.08 | 1.44 | 0 | 0.72 | 3.24 |
| 917 | 14.4 | 15.12 | 15.12 | 15.12 | 59.76 |
| 919 | 6.48 | 0 | 7.56 | 10.44 | 24.48 |
| 920 | 36.36 | 18.36 | 22.68 | 23.76 | 101.16 |
| 921 | 36.36 | 35.28 | 38.52 | 23.04 | 133.2 |
| 922 | 17.64 | 21.96 | 25.2 | 26.64 | 91.44 |
| 923 | 5.4 | 8.64 | 7.92 | 4.32 | 26.28 |
| 925 | 7.2 | 5.4 | 4.32 | 8.64 | 25.56 |
| 926 | 15.12 | 17.28 | 11.52 | 14.04 | 57.96 |
| 927 | 9.36 | 11.16 | 7.2 | 5.4 | 33.12 |
| 928 | 8.64 | 8.64 | 10.08 | 6.48 | 33.84 |
| 930 | 3.24 | 7.2 | 4.68 | 3.24 | 18.36 |
| 931 | 6.12 | 2.88 | 3.24 | 1.44 | 13.68 |
| 932 | 9 | 5.76 | 6.12 | 11.52 | 32.4 |
| 933 | 5.76 | 8.28 | 3.96 | 6.12 | 24.12 |
| 934 | 11.16 | 7.56 | 10.8 | 5.4 | 34.92 |
| 935 | 1.8 | 3.96 | 2.88 | 3.96 | 12.6 |
| 936 | 0.72 | 0 | 1.08 | 1.8 | 3.6 |
| 937 | 10.8 | 8.28 | 8.28 | 6.84 | 34.2 |
| 939 | 4.68 | 2.88 | 1.8 | 3.6 | 12.96 |
| 940 | 7.56 | 5.4 | 5.76 | 3.24 | 21.96 |
| 941 | 12.6 | 24.48 | 27.72 | 17.28 | 82.08 |
| 943 | 9.72 | 5.04 | 4.68 | 10.8 | 30.24 |
| 944 | 0 | 0 | 0 | 0.36 | 0.36 |
| 945 | 28.44 | 37.08 | 32.4 | 31.68 | 129.6 |
| 946 | 8.64 | 3.96 | 8.64 | 2.52 | 23.76 |
| 947 | 0.72 | 2.16 | 0.36 | 0 | 3.24 |
| 948 | 2.16 | 3.96 | 3.24 | 3.6 | 12.96 |
| 949 | 6.84 | 6.48 | 1.8 | 3.6 | 18.72 |
| 950 | 16.56 | 7.2 | 13.32 | 7.92 | 45 |
| 951 | 2.88 | 5.76 | 4.32 | 3.6 | 16.56 |
| 952 | 14.76 | 18.36 | 9.36 | 6.48 | 48.96 |
| 953 | 19.8 | 17.64 | 13.68 | 9.36 | 60.48 |
| 954 | 15.48 | 10.44 | 10.44 | 6.12 | 42.48 |
| 955 | 1.44 | 5.04 | 1.08 | 2.16 | 9.72 |
| 956 | 18.36 | 40.32 | 18.36 | 42.84 | 119.88 |
| 957 | 1.8 | 0.72 | 1.44 | 0.72 | 4.68 |
| 958 | 7.92 | 20.52 | 27.72 | 29.16 | 85.32 |
| 959 | 19.44 | 18.36 | 17.28 | 16.2 | 71.28 |
| 960 | 2.88 | 2.52 | 3.96 | 5.4 | 14.76 |
| 961 | 1.08 | 1.8 | 0 | 1.44 | 4.32 |
| TOTAL | 470.16 | 507.24 | 479.52 | 455.4 | 1,912.32 |

STATE: COLORADO

| ZIP CODE | QUARTER 1 | QUARTER 2 | QUARTER 3 | QUARTER 4 | TOTAL GRAMS |
|---|---|---|---|---|---|

DRUG CODE:9639DRUG NAME:OPIUM POWDERED
STATE:COLORADO

| ZIP CODE | QUARTER 1 | QUARTER 2 | QUARTER 3 | QUARTER 4 | TOTAL GRAMS |
|---|---|---|---|---|---|
| 800 | 40.68 | 26.28 | 24.12 | 32.4 | 123.48 |
| 801 | 28.44 | 29.16 | 24.48 | 20.88 | 102.96 |
| 802 | 59.76 | 40.32 | 44.64 | 45.36 | 190.08 |
| 803 | 1.08 | 0 | 0 | 0.72 | 1.8 |
| 804 | 3.6 | 6.12 | 7.56 | 4.32 | 21.6 |
| 805 | 51.48 | 46.08 | 47.52 | 54 | 199.08 |
| 806 | 4.68 | 3.24 | 4.68 | 1.8 | 14.4 |
| 807 | 2.52 | 1.44 | 1.44 | 1.8 | 7.2 |
| 808 | 0 | 0.36 | 0 | 1.44 | 1.8 |
| 809 | 46.44 | 38.52 | 35.28 | 31.68 | 151.92 |
| 810 | 0.36 | 6.84 | 2.52 | 4.68 | 14.4 |
| 812 | 0.36 | 0 | 0.72 | 0 | 1.08 |
| 813 | 0.36 | 1.44 | 1.44 | 2.16 | 5.4 |
| 814 | 5.04 | 5.4 | 6.12 | 5.04 | 21.6 |
| 815 | 9 | 8.28 | 13.68 | 9 | 39.96 |
| 816 | 2.88 | 0.72 | 2.88 | 0.36 | 6.84 |
| TOTAL | 256.68 | 214.2 | 217.08 | 215.64 | 903.6 |

STATE: CONNECTICUT

| ZIP CODE | QUARTER 1 | QUARTER 2 | QUARTER 3 | QUARTER 4 | TOTAL GRAMS |
|---|---|---|---|---|---|
| 60 | 37.08 | 15.12 | 21.96 | 18.72 | 92.88 |
| 61 | 8.28 | 11.16 | 8.64 | 6.84 | 34.92 |
| 62 | 0.36 | 0 | 1.08 | 1.8 | 3.24 |
| 63 | 7.92 | 5.76 | 13.68 | 8.28 | 35.64 |
| 64 | 28.8 | 29.52 | 14.04 | 40.68 | 113.04 |
| 65 | 4.32 | 10.8 | 8.28 | 2.88 | 26.28 |
| 66 | 3.24 | 9 | 2.16 | 1.08 | 15.48 |
| 67 | 12.24 | 2.88 | 10.08 | 5.76 | 30.96 |
| 68 | 4.68 | 1.44 | 2.16 | 2.88 | 11.16 |
| 69 | 0.36 | 0 | 0.36 | 0 | 0.72 |
| TOTAL | 107.28 | 85.68 | 82.44 | 88.92 | 364.32 |

STATE: DELAWARE

| ZIP CODE | QUARTER 1 | QUARTER 2 | QUARTER 3 | QUARTER 4 | TOTAL GRAMS |
|---|---|---|---|---|---|
| 197 | 9 | 6.12 | 5.04 | 3.24 | 23.4 |
| 198 | 2.16 | 9.36 | 2.16 | 1.8 | 15.48 |
| 199 | 13.32 | 8.64 | 10.44 | 6.84 | 39.24 |
| TOTAL | 24.48 | 24.12 | 17.64 | 11.88 | 78.12 |

STATE: DISTRICT OF COLUMBIA

| ZIP CODE | QUARTER 1 | QUARTER 2 | QUARTER 3 | QUARTER 4 | TOTAL GRAMS |
|---|---|---|---|---|---|
| 200 | 12.24 | 10.8 | 6.12 | 17.28 | 46.44 |
| TOTAL | 12.24 | 10.8 | 6.12 | 17.28 | 46.44 |

STATE: FLORIDA

| ZIP CODE | QUARTER 1 | QUARTER 2 | QUARTER 3 | QUARTER 4 | TOTAL GRAMS |
|---|---|---|---|---|---|
| 320 | 10.44 | 11.52 | 8.28 | 5.76 | 36 |
| 321 | 19.8 | 12.24 | 19.44 | 25.92 | 77.4 |
| 322 | 44.64 | 54.72 | 36 | 53.64 | 189 |
| 323 | 6.84 | 3.6 | 7.56 | 7.56 | 25.56 |
| 324 | 5.04 | 12.24 | 10.44 | 3.96 | 31.68 |
| 325 | 34.92 | 26.64 | 23.04 | 36.36 | 120.96 |
| 326 | 6.48 | 0 | 4.68 | 4.68 | 15.84 |
| 327 | 36 | 34.2 | 25.2 | 34.2 | 129.6 |
| 328 | 25.92 | 27.36 | 29.52 | 25.2 | 108 |
| 329 | 26.28 | 39.6 | 27.72 | 26.64 | 120.24 |
| 330 | 13.68 | 13.32 | 7.56 | 7.92 | 42.48 |
| 331 | 12.96 | 5.76 | 13.68 | 16.2 | 48.6 |

| | | | | | |
|---|---|---|---|---|---|
| 333 | 13.32 | 12.24 | 12.6 | 12.96 | 51.12 |
| 334 | 30.96 | 61.2 | 45 | 48.6 | 185.76 |
| 335 | 7.92 | 9 | 7.2 | 9 | 33.12 |
| 336 | 24.48 | 18.36 | 30.6 | 28.08 | 101.52 |
| 337 | 24.48 | 15.12 | 20.88 | 16.92 | 77.4 |
| 338 | 6.12 | 7.2 | 5.76 | 6.84 | 25.92 |
| 339 | 35.64 | 26.28 | 24.84 | 23.04 | 109.8 |
| 341 | 27 | 19.08 | 11.52 | 30.6 | 88.2 |
| 342 | 15.12 | 11.16 | 9.36 | 18.72 | 54.36 |
| 344 | 0.36 | 1.44 | 0.72 | 2.52 | 5.04 |
| 346 | 24.84 | 12.96 | 15.84 | 14.04 | 67.68 |
| 347 | 28.08 | 22.32 | 22.32 | 30.96 | 103.68 |

DRUG CODE:9639DRUG NAME:OPIUM POWDERED
STATE:FLORIDA

| ZIP CODE | QUARTER 1 | QUARTER 2 | QUARTER 3 | QUARTER 4 | TOTAL GRAMS |
|---|---|---|---|---|---|
| 349 | 1.08 | 6.12 | 4.68 | 11.52 | 23.4 |
| TOTAL | 482.4 | 463.68 | 424.44 | 501.84 | 1,872.36 |

STATE: GEORGIA

| ZIP CODE | QUARTER 1 | QUARTER 2 | QUARTER 3 | QUARTER 4 | TOTAL GRAMS |
|---|---|---|---|---|---|
| 300 | 17.28 | 13.68 | 19.44 | 28.44 | 78.84 |
| 301 | 33.12 | 38.16 | 33.84 | 39.24 | 144.36 |
| 302 | 21.96 | 24.48 | 17.64 | 17.28 | 81.36 |
| 303 | 28.08 | 24.84 | 27.36 | 26.64 | 106.92 |
| 304 | 3.6 | 0.72 | 2.88 | 1.08 | 8.28 |
| 305 | 15.84 | 17.64 | 10.44 | 7.92 | 51.84 |
| 306 | 18 | 11.52 | 11.16 | 14.4 | 55.08 |
| 307 | 9.72 | 8.28 | 10.08 | 8.64 | 36.72 |
| 309 | 9.36 | 7.92 | 20.16 | 22.68 | 60.12 |
| 310 | 2.88 | 3.24 | 11.52 | 3.24 | 20.88 |
| 312 | 8.64 | 9.72 | 11.16 | 7.56 | 37.08 |
| 313 | 0 | 1.44 | 2.52 | 0 | 3.96 |
| 314 | 5.4 | 4.68 | 8.28 | 5.76 | 24.12 |
| 315 | 3.6 | 7.92 | 7.56 | 4.68 | 23.76 |
| 316 | 5.76 | 1.8 | 2.88 | 4.32 | 14.76 |
| 317 | 5.04 | 9.36 | 6.48 | 3.24 | 24.12 |
| 319 | 1.8 | 2.16 | 1.8 | 0 | 5.76 |
| 398 | 0 | 0.72 | 0.72 | 0.72 | 2.16 |
| TOTAL | 190.08 | 188.28 | 205.92 | 195.84 | 780.12 |

STATE: GUAM

| ZIP CODE | QUARTER 1 | QUARTER 2 | QUARTER 3 | QUARTER 4 | TOTAL GRAMS |
|---|---|---|---|---|---|
| 969 | 0.72 | 0.72 | 1.44 | 0 | 2.88 |
| TOTAL | 0.72 | 0.72 | 1.44 | 0 | 2.88 |

STATE: HAWAII

| ZIP CODE | QUARTER 1 | QUARTER 2 | QUARTER 3 | QUARTER 4 | TOTAL GRAMS |
|---|---|---|---|---|---|
| 967 | 8.28 | 25.2 | 5.76 | 5.4 | 44.64 |
| 968 | 17.64 | 12.24 | 15.12 | 11.16 | 56.16 |
| TOTAL | 25.92 | 37.44 | 20.88 | 16.56 | 100.8 |

STATE: IDAHO

| ZIP CODE | QUARTER 1 | QUARTER 2 | QUARTER 3 | QUARTER 4 | TOTAL GRAMS |
|---|---|---|---|---|---|
| 832 | 5.04 | 1.44 | 1.08 | 2.16 | 9.72 |
| 833 | 2.16 | 3.6 | 2.88 | 3.24 | 11.88 |
| 834 | 5.76 | 3.96 | 5.4 | 1.8 | 16.92 |
| 835 | 2.16 | 2.16 | 1.08 | 1.8 | 7.2 |
| 836 | 16.56 | 16.2 | 14.4 | 13.68 | 60.84 |

| | | | | | |
|---|---|---|---|---|---|
| 837 | 6.48 | 6.48 | 3.6 | 5.04 | 21.6 |
| 838 | 15.48 | 11.88 | 10.44 | 15.48 | 53.28 |
| TOTAL | 53.64 | 45.72 | 38.88 | 43.2 | 181.44 |

STATE: ILLINOIS

| ZIP CODE | QUARTER 1 | QUARTER 2 | QUARTER 3 | QUARTER 4 | TOTAL GRAMS |
|---|---|---|---|---|---|
| 600 | 30.6 | 29.16 | 41.04 | 24.48 | 125.28 |
| 601 | 32.04 | 20.52 | 33.48 | 42.12 | 128.16 |
| 602 | 1.44 | 1.8 | 1.44 | 0 | 4.68 |
| 603 | 3.96 | 0.36 | 1.8 | 0 | 6.12 |
| 604 | 25.56 | 25.92 | 25.92 | 30.24 | 107.64 |
| 605 | 22.68 | 27.36 | 12.6 | 10.44 | 73.08 |
| 606 | 17.28 | 12.24 | 21.6 | 24.84 | 75.96 |
| 607 | 0 | 0.72 | 0.36 | 0 | 1.08 |
| 608 | 2.16 | 0 | 3.6 | 0.72 | 6.48 |
| 609 | 3.6 | 1.44 | 1.8 | 1.44 | 8.28 |
| 610 | 0.72 | 3.24 | 3.6 | 1.44 | 9 |
| 611 | 14.4 | 9.72 | 11.88 | 11.16 | 47.16 |
| 612 | 0.36 | 2.52 | 5.4 | 0.36 | 8.64 |
| 613 | 3.96 | 2.88 | 4.68 | 0.72 | 12.24 |
| 614 | 3.24 | 2.16 | 1.44 | 2.16 | 9 |
| 615 | 4.32 | 1.8 | 0.72 | 1.8 | 8.64 |
| 616 | 13.32 | 14.04 | 12.6 | 15.84 | 55.8 |
| 617 | 5.4 | 5.04 | 5.04 | 8.64 | 24.12 |
| 618 | 1.44 | 4.32 | 1.44 | 0 | 7.2 |
| 619 | 0.72 | 0.72 | 0.36 | 0.36 | 2.16 |
| 620 | 5.4 | 1.44 | 1.08 | 3.6 | 11.52 |
| 622 | 2.88 | 2.16 | 0.36 | 3.24 | 8.64 |

DRUG CODE:9639DRUG NAME:OPIUM POWDERED
STATE:ILLINOIS

| ZIP CODE | QUARTER 1 | QUARTER 2 | QUARTER 3 | QUARTER 4 | TOTAL GRAMS |
|---|---|---|---|---|---|
| 623 | 1.44 | 2.88 | 2.16 | 1.44 | 7.92 |
| 624 | 0 | 0.72 | 3.24 | 2.52 | 6.48 |
| 625 | 0.72 | 2.52 | 2.88 | 0.72 | 6.84 |
| 626 | 2.88 | 0.72 | 2.16 | 0.36 | 6.12 |
| 627 | 2.52 | 3.6 | 7.56 | 3.6 | 17.28 |
| 628 | 0.72 | 0.72 | 1.44 | 1.08 | 3.96 |
| 629 | 1.8 | 7.92 | 2.16 | 5.04 | 16.92 |
| TOTAL | 205.56 | 188.64 | 213.84 | 198.36 | 806.4 |

STATE: INDIANA

| ZIP CODE | QUARTER 1 | QUARTER 2 | QUARTER 3 | QUARTER 4 | TOTAL GRAMS |
|---|---|---|---|---|---|
| 460 | 15.48 | 18.72 | 24.12 | 24.84 | 83.16 |
| 461 | 40.32 | 33.12 | 41.76 | 16.2 | 131.4 |
| 462 | 83.52 | 102.6 | 99.72 | 95.04 | 380.88 |
| 463 | 9.36 | 9.36 | 5.76 | 7.56 | 32.04 |
| 464 | 1.08 | 3.24 | 0 | 4.32 | 8.64 |
| 465 | 16.2 | 10.08 | 16.92 | 12.96 | 56.16 |
| 466 | 7.2 | 5.4 | 6.48 | 4.32 | 23.4 |
| 467 | 0 | 0.72 | 0 | 0.72 | 1.44 |
| 468 | 16.56 | 7.2 | 10.08 | 14.4 | 48.24 |
| 469 | 2.16 | 5.76 | 8.64 | 3.6 | 20.16 |
| 470 | 1.8 | 1.08 | 1.08 | 0.72 | 4.68 |
| 471 | 11.16 | 24.84 | 11.88 | 10.8 | 58.68 |
| 472 | 5.04 | 8.64 | 9.36 | 10.8 | 33.84 |
| 473 | 5.4 | 9.72 | 9.36 | 10.44 | 34.92 |
| 474 | 2.52 | 8.64 | 3.6 | 4.68 | 19.44 |
| 475 | 6.12 | 8.28 | 7.92 | 6.48 | 28.8 |
| 476 | 0.72 | 2.16 | 0 | 2.16 | 5.04 |

| | | | | | |
|---|---|---|---|---|---|
| 477 | 11.16 | 7.92 | 10.8 | 6.84 | 36.72 |
| 478 | 1.44 | 0.72 | 1.44 | 0.72 | 4.32 |
| 479 | 4.32 | 10.8 | 2.52 | 1.8 | 19.44 |
| TOTAL | 241.56 | 279 | 271.44 | 239.4 | 1,031.40 |

STATE: IOWA

| ZIP CODE | QUARTER 1 | QUARTER 2 | QUARTER 3 | QUARTER 4 | TOTAL GRAMS |
|---|---|---|---|---|---|
| 500 | 5.04 | 5.76 | 3.96 | 8.64 | 23.4 |
| 501 | 16.56 | 16.2 | 17.64 | 17.28 | 67.68 |
| 502 | 3.24 | 5.4 | 3.24 | 3.24 | 15.12 |
| 503 | 4.68 | 5.76 | 4.68 | 10.44 | 25.56 |
| 504 | 0 | 0.72 | 3.6 | 2.16 | 6.48 |
| 505 | 2.52 | 1.44 | 0.72 | 0.36 | 5.04 |
| 506 | 3.24 | 1.44 | 0.72 | 1.8 | 7.2 |
| 507 | 7.56 | 6.12 | 15.48 | 9 | 38.16 |
| 510 | 2.16 | 1.44 | 0.72 | 1.08 | 5.4 |
| 511 | 0.72 | 3.6 | 5.04 | 5.76 | 15.12 |
| 512 | 0.72 | 2.52 | 1.8 | 0.72 | 5.76 |
| 513 | 2.88 | 2.52 | 2.16 | 1.08 | 8.64 |
| 514 | 0.36 | 0 | 2.16 | 0 | 2.52 |
| 515 | 4.68 | 4.32 | 6.48 | 2.16 | 17.64 |
| 520 | 1.08 | 2.52 | 1.8 | 2.16 | 7.56 |
| 521 | 0.72 | 1.44 | 0 | 0 | 2.16 |
| 522 | 10.8 | 23.76 | 11.52 | 20.88 | 66.96 |
| 523 | 0 | 0 | 0.36 | 0 | 0.36 |
| 524 | 5.04 | 5.76 | 3.96 | 5.04 | 19.8 |
| 525 | 1.44 | 0.72 | 1.44 | 1.8 | 5.4 |
| 526 | 0 | 1.8 | 1.44 | 3.24 | 6.48 |
| 527 | 0.36 | 1.8 | 0 | 2.52 | 4.68 |
| 528 | 7.56 | 12.24 | 16.2 | 17.28 | 53.28 |
| TOTAL | 81.36 | 107.28 | 105.12 | 116.64 | 410.4 |

STATE: KANSAS

| ZIP CODE | QUARTER 1 | QUARTER 2 | QUARTER 3 | QUARTER 4 | TOTAL GRAMS |
|---|---|---|---|---|---|
| 660 | 18.36 | 16.92 | 22.32 | 28.08 | 85.68 |
| 661 | 12.96 | 17.64 | 20.52 | 24.84 | 75.96 |
| 662 | 74.52 | 81.36 | 79.56 | 70.56 | 306 |
| 664 | 1.08 | 0.36 | 1.08 | 1.44 | 3.96 |
| 665 | 14.4 | 15.84 | 15.12 | 18 | 63.36 |
| 666 | 2.16 | 5.76 | 6.84 | 5.04 | 19.8 |
| 667 | 1.44 | 2.16 | 1.44 | 0.36 | 5.4 |

DRUG CODE:9639DRUG NAME:OPIUM POWDERED
STATE:KANSAS

| ZIP CODE | QUARTER 1 | QUARTER 2 | QUARTER 3 | QUARTER 4 | TOTAL GRAMS |
|---|---|---|---|---|---|
| 668 | 0 | 1.44 | 1.08 | 0 | 2.52 |
| 669 | 0 | 0.72 | 0 | 0 | 0.72 |
| 670 | 2.88 | 3.6 | 5.4 | 5.04 | 16.92 |
| 671 | 2.88 | 0.72 | 0.72 | 1.44 | 5.76 |
| 672 | 21.6 | 21.6 | 28.8 | 18 | 90 |
| 673 | 0 | 1.08 | 0 | 2.88 | 3.96 |
| 674 | 7.92 | 9 | 9.36 | 5.76 | 32.04 |
| 675 | 3.96 | 2.16 | 1.08 | 1.44 | 8.64 |
| 676 | 3.6 | 3.6 | 2.88 | 3.6 | 13.68 |
| 677 | 1.08 | 1.08 | 0.36 | 0 | 2.52 |
| 678 | 2.16 | 1.44 | 2.52 | 2.16 | 8.28 |
| 679 | 1.44 | 2.52 | 0.72 | 0 | 4.68 |
| TOTAL | 172.44 | 189 | 199.8 | 188.64 | 749.88 |

STATE: KENTUCKY

RETAIL DRUG DISTRIBUTION BY ZIP CODE WITHIN STATE BY GRAMS WT

| ZIP CODE | QUARTER 1 | QUARTER 2 | QUARTER 3 | QUARTER 4 | TOTAL GRAMS |
|---|---|---|---|---|---|
| 400 | 1.8 | 4.68 | 1.44 | 0.36 | 8.28 |
| 402 | 50.76 | 41.04 | 46.44 | 53.64 | 191.88 |
| 403 | 9.36 | 13.32 | 9 | 2.16 | 33.84 |
| 404 | 5.4 | 13.68 | 6.84 | 8.64 | 34.56 |
| 405 | 48.6 | 44.64 | 42.84 | 54.72 | 190.8 |
| 406 | 0 | 0 | 0 | 2.52 | 2.52 |
| 407 | 2.16 | 1.08 | 3.6 | 23.76 | 30.6 |
| 408 | 0 | 0 | 0 | 0.72 | 0.72 |
| 409 | 0 | 0 | 0.36 | 0 | 0.36 |
| 410 | 10.44 | 7.2 | 4.32 | 5.4 | 27.36 |
| 411 | 2.52 | 4.32 | 5.04 | 2.88 | 14.76 |
| 412 | 1.08 | 0.72 | 0 | 1.44 | 3.24 |
| 413 | 0.36 | 0 | 0.72 | 0 | 1.08 |
| 415 | 4.68 | 4.68 | 0.72 | 1.08 | 11.16 |
| 416 | 0.72 | 0.36 | 1.08 | 0.36 | 2.52 |
| 417 | 1.08 | 3.96 | 0.72 | 1.44 | 7.2 |
| 420 | 8.28 | 8.28 | 18 | 5.4 | 39.96 |
| 421 | 8.28 | 9 | 7.92 | 11.52 | 36.72 |
| 422 | 1.44 | 0 | 4.32 | 0.72 | 6.48 |
| 423 | 0.72 | 0.36 | 2.52 | 0.72 | 4.32 |
| 424 | 6.12 | 10.44 | 13.68 | 15.12 | 45.36 |
| 425 | 0 | 0.72 | 0.36 | 2.16 | 3.24 |
| 426 | 0 | 0 | 1.8 | 0 | 1.8 |
| 427 | 1.8 | 2.16 | 1.44 | 0.72 | 6.12 |
| TOTAL | 165.6 | 170.64 | 173.16 | 195.48 | 704.88 |

STATE: LOUISIANA

| ZIP CODE | QUARTER 1 | QUARTER 2 | QUARTER 3 | QUARTER 4 | TOTAL GRAMS |
|---|---|---|---|---|---|
| 700 | 37.44 | 24.12 | 23.04 | 14.04 | 98.64 |
| 701 | 3.96 | 6.12 | 6.48 | 7.56 | 24.12 |
| 703 | 7.92 | 11.16 | 2.16 | 5.04 | 26.28 |
| 704 | 9 | 8.28 | 8.64 | 5.76 | 31.68 |
| 705 | 18.36 | 29.88 | 16.56 | 24.12 | 88.92 |
| 706 | 7.92 | 8.64 | 6.84 | 7.2 | 30.6 |
| 707 | 2.16 | 0.72 | 5.4 | 10.8 | 19.08 |
| 708 | 4.68 | 6.84 | 6.84 | 2.88 | 21.24 |
| 710 | 0 | 0.72 | 0 | 3.24 | 3.96 |
| 711 | 9 | 11.88 | 11.16 | 14.4 | 46.44 |
| 712 | 1.44 | 2.88 | 3.6 | 1.44 | 9.36 |
| 713 | 10.08 | 10.8 | 2.16 | 4.32 | 27.36 |
| 714 | 0 | 1.44 | 0 | 0 | 1.44 |
| TOTAL | 111.96 | 123.48 | 92.88 | 100.8 | 429.12 |

STATE: MAINE

| ZIP CODE | QUARTER 1 | QUARTER 2 | QUARTER 3 | QUARTER 4 | TOTAL GRAMS |
|---|---|---|---|---|---|
| 39 | 0.36 | 0.72 | 0 | 0 | 1.08 |
| 40 | 2.88 | 3.96 | 9.36 | 1.8 | 18 |
| 41 | 8.64 | 5.76 | 4.32 | 4.32 | 23.04 |
| 42 | 16.56 | 12.96 | 12.96 | 10.08 | 52.56 |
| 43 | 4.68 | 2.88 | 3.24 | 5.76 | 16.56 |
| 44 | 6.48 | 7.56 | 10.8 | 6.48 | 31.32 |
| 45 | 0 | 0 | 2.52 | 0 | 2.52 |
| 46 | 2.52 | 2.16 | 1.8 | 1.8 | 8.28 |

DRUG CODE:9639DRUG NAME:OPIUM POWDERED
STATE:MAINE

| ZIP CODE | QUARTER 1 | QUARTER 2 | QUARTER 3 | QUARTER 4 | TOTAL GRAMS |
|---|---|---|---|---|---|
| 47 | 2.52 | 3.24 | 1.44 | 0.72 | 7.92 |

| | | | | | |
|---|---|---|---|---|---|
| 48 | 1.44 | 0 | 0 | 0 | 1.44 |
| 49 | 0.72 | 0.72 | 0.36 | 0.36 | 2.16 |
| TOTAL | 46.8 | 39.96 | 46.8 | 31.32 | 164.88 |

STATE: MARYLAND

| ZIP CODE | QUARTER 1 | QUARTER 2 | QUARTER 3 | QUARTER 4 | TOTAL GRAMS |
|---|---|---|---|---|---|
| 206 | 3.6 | 3.6 | 3.24 | 3.24 | 13.68 |
| 207 | 1.44 | 3.96 | 9.36 | 5.4 | 20.16 |
| 208 | 14.4 | 27.72 | 14.04 | 12.6 | 68.76 |
| 209 | 1.08 | 0 | 0.36 | 0.36 | 1.8 |
| 210 | 11.52 | 4.68 | 10.44 | 13.68 | 40.32 |
| 211 | 2.88 | 5.04 | 4.32 | 5.04 | 17.28 |
| 212 | 17.64 | 8.64 | 48.6 | 13.68 | 88.56 |
| 214 | 1.08 | 1.44 | 0.72 | 1.08 | 4.32 |
| 215 | 0.36 | 0.36 | 4.32 | 0 | 5.04 |
| 216 | 3.6 | 5.76 | 4.32 | 5.4 | 19.08 |
| 217 | 0.72 | 0.36 | 2.88 | 0 | 3.96 |
| 218 | 1.8 | 2.16 | 1.8 | 1.8 | 7.56 |
| 219 | 0 | 0 | 3.6 | 2.52 | 6.12 |
| TOTAL | 60.12 | 63.72 | 108 | 64.8 | 296.64 |

STATE: MASSACHUSETTS

| ZIP CODE | QUARTER 1 | QUARTER 2 | QUARTER 3 | QUARTER 4 | TOTAL GRAMS |
|---|---|---|---|---|---|
| 10 | 28.08 | 5.04 | 4.68 | 20.52 | 58.32 |
| 11 | 30.96 | 20.52 | 37.44 | 34.56 | 123.48 |
| 13 | 1.08 | 0 | 0.72 | 0 | 1.8 |
| 14 | 1.8 | 2.16 | 1.44 | 1.08 | 6.48 |
| 15 | 2.52 | 2.16 | 5.76 | 10.44 | 20.88 |
| 16 | 2.88 | 3.96 | 1.08 | 0 | 7.92 |
| 17 | 11.52 | 7.2 | 11.16 | 7.56 | 37.44 |
| 18 | 7.2 | 3.24 | 7.92 | 15.12 | 33.48 |
| 19 | 13.68 | 7.92 | 24.12 | 7.92 | 53.64 |
| 20 | 2.88 | 5.76 | 7.2 | 5.4 | 21.24 |
| 21 | 36.72 | 46.44 | 35.28 | 54.36 | 172.8 |
| 22 | 8.28 | 2.16 | 1.8 | 2.88 | 15.12 |
| 23 | 5.4 | 7.92 | 7.56 | 6.12 | 27 |
| 24 | 12.6 | 19.44 | 17.64 | 13.32 | 63 |
| 25 | 3.24 | 0.72 | 0.72 | 1.44 | 6.12 |
| 26 | 1.08 | 3.24 | 1.44 | 2.16 | 7.92 |
| 27 | 6.12 | 5.4 | 9 | 20.16 | 40.68 |
| TOTAL | 176.04 | 143.28 | 174.96 | 203.04 | 697.32 |

STATE: MICHIGAN

| ZIP CODE | QUARTER 1 | QUARTER 2 | QUARTER 3 | QUARTER 4 | TOTAL GRAMS |
|---|---|---|---|---|---|
| 480 | 32.04 | 44.28 | 34.92 | 41.4 | 152.64 |
| 481 | 50.4 | 48.24 | 54 | 56.88 | 209.52 |
| 482 | 20.16 | 29.88 | 15.84 | 32.04 | 97.92 |
| 483 | 36.72 | 27.72 | 60.48 | 46.08 | 171 |
| 484 | 3.96 | 2.88 | 3.24 | 5.76 | 15.84 |
| 485 | 3.96 | 6.12 | 3.96 | 5.76 | 19.8 |
| 486 | 6.48 | 19.44 | 13.68 | 18.36 | 57.96 |
| 487 | 4.32 | 5.76 | 5.76 | 5.4 | 21.24 |
| 488 | 12.24 | 9.72 | 14.4 | 1.8 | 38.16 |
| 489 | 14.04 | 14.76 | 5.4 | 9.72 | 43.92 |
| 490 | 24.84 | 59.04 | 33.12 | 35.28 | 152.28 |
| 491 | 3.24 | 1.44 | 2.16 | 6.48 | 13.32 |
| 492 | 19.44 | 25.56 | 27.72 | 19.44 | 92.16 |
| 493 | 0 | 1.44 | 0.72 | 1.08 | 3.24 |
| 494 | 13.32 | 6.84 | 13.32 | 11.52 | 45 |

| | | | | | |
|---|---|---|---|---|---|
| 495 | 28.8 | 38.88 | 33.84 | 43.92 | 145.44 |
| 496 | 21.24 | 24.48 | 29.88 | 16.92 | 92.52 |
| 497 | 12.6 | 16.56 | 11.88 | 16.2 | 57.24 |
| 498 | 11.88 | 6.12 | 5.04 | 8.28 | 31.32 |
| 499 | 7.2 | 0 | 0 | 0 | 7.2 |
| TOTAL | 326.88 | 389.16 | 369.36 | 382.32 | 1,467.72 |

STATE: MINNESOTA

| ZIP CODE | QUARTER 1 | QUARTER 2 | QUARTER 3 | QUARTER 4 | TOTAL GRAMS |
|---|---|---|---|---|---|
| 550 | 38.52 | 28.44 | 41.04 | 35.64 | 143.64 |

DRUG CODE:9639DRUG NAME:OPIUM POWDERED
STATE:MINNESOTA

| ZIP CODE | QUARTER 1 | QUARTER 2 | QUARTER 3 | QUARTER 4 | TOTAL GRAMS |
|---|---|---|---|---|---|
| 551 | 60.12 | 48.6 | 60.84 | 58.68 | 228.24 |
| 553 | 57.96 | 53.64 | 53.64 | 53.64 | 218.88 |
| 554 | 132.48 | 155.52 | 140.04 | 123.48 | 551.52 |
| 556 | 0 | 0 | 0.72 | 1.44 | 2.16 |
| 557 | 0.72 | 0.36 | 3.24 | 2.16 | 6.48 |
| 558 | 5.04 | 7.2 | 9.36 | 8.64 | 30.24 |
| 559 | 47.16 | 48.24 | 60.48 | 66.96 | 222.84 |
| 560 | 9 | 8.64 | 8.64 | 6.84 | 33.12 |
| 561 | 2.16 | 0.36 | 0.36 | 1.8 | 4.68 |
| 562 | 5.04 | 10.08 | 16.2 | 20.16 | 51.48 |
| 563 | 28.8 | 30.96 | 33.12 | 23.4 | 116.28 |
| 564 | 13.32 | 16.2 | 15.12 | 14.4 | 59.04 |
| 565 | 1.8 | 5.04 | 6.48 | 1.08 | 14.4 |
| 566 | 2.16 | 0 | 1.44 | 1.44 | 5.04 |
| 567 | 0 | 0.36 | 3.24 | 1.44 | 5.04 |
| TOTAL | 404.28 | 413.64 | 453.96 | 421.2 | 1,693.08 |

STATE: MISSISSIPPI

| ZIP CODE | QUARTER 1 | QUARTER 2 | QUARTER 3 | QUARTER 4 | TOTAL GRAMS |
|---|---|---|---|---|---|
| 386 | 8.64 | 4.68 | 6.12 | 4.68 | 24.12 |
| 387 | 3.96 | 2.16 | 0.36 | 3.6 | 10.08 |
| 388 | 6.84 | 5.4 | 15.12 | 5.04 | 32.4 |
| 389 | 0.36 | 0 | 1.44 | 0 | 1.8 |
| 390 | 0 | 0.36 | 0 | 0.36 | 0.72 |
| 391 | 2.16 | 3.24 | 1.08 | 0 | 6.48 |
| 392 | 11.16 | 4.68 | 7.92 | 6.48 | 30.24 |
| 393 | 1.08 | 0.72 | 0 | 0 | 1.8 |
| 394 | 2.52 | 3.24 | 7.2 | 0.36 | 13.32 |
| 395 | 11.16 | 2.16 | 3.6 | 4.32 | 21.24 |
| 396 | 0 | 0.72 | 0.36 | 0.36 | 1.44 |
| 397 | 0 | 2.88 | 1.44 | 2.88 | 7.2 |
| TOTAL | 47.88 | 30.24 | 44.64 | 28.08 | 150.84 |

STATE: MISSOURI

| ZIP CODE | QUARTER 1 | QUARTER 2 | QUARTER 3 | QUARTER 4 | TOTAL GRAMS |
|---|---|---|---|---|---|
| 630 | 5.76 | 8.64 | 7.2 | 7.2 | 28.8 |
| 631 | 29.16 | 50.04 | 70.92 | 48.96 | 199.08 |
| 633 | 5.76 | 6.12 | 6.84 | 4.32 | 23.04 |
| 634 | 1.44 | 0 | 1.44 | 2.52 | 5.4 |
| 635 | 0 | 1.8 | 0 | 3.24 | 5.04 |
| 636 | 3.6 | 2.52 | 3.6 | 2.88 | 12.6 |
| 637 | 4.32 | 6.48 | 7.56 | 2.16 | 20.52 |
| 638 | 1.8 | 1.08 | 1.08 | 0.72 | 4.68 |
| 639 | 0.72 | 1.44 | 0 | 1.44 | 3.6 |
| 640 | 24.48 | 28.8 | 32.76 | 31.68 | 117.72 |

| | | | | | |
|---|---|---|---|---|---|
| 641 | 37.08 | 40.68 | 25.92 | 28.08 | 131.76 |
| 644 | 0.36 | 1.44 | 0.72 | 1.08 | 3.6 |
| 645 | 1.44 | 4.32 | 6.12 | 6.12 | 18 |
| 646 | 0 | 0.72 | 0.36 | 0 | 1.08 |
| 647 | 12.24 | 13.68 | 11.88 | 12.24 | 50.04 |
| 648 | 0.72 | 1.8 | 3.24 | 1.08 | 6.84 |
| 650 | 0.72 | 2.16 | 0.36 | 0 | 3.24 |
| 651 | 0.72 | 1.44 | 1.08 | 2.16 | 5.4 |
| 652 | 6.12 | 5.04 | 15.84 | 16.2 | 43.2 |
| 653 | 1.8 | 2.52 | 1.44 | 2.88 | 8.64 |
| 654 | 2.16 | 1.08 | 15.48 | 7.92 | 26.64 |
| 655 | 7.56 | 3.6 | 7.2 | 7.56 | 25.92 |
| 656 | 0 | 0 | 0.72 | 0 | 0.72 |
| 657 | 0.36 | 1.08 | 0 | 0 | 1.44 |
| 658 | 10.8 | 15.84 | 12.24 | 14.76 | 53.64 |
| TOTAL | 159.12 | 202.32 | 234 | 205.2 | 800.64 |

STATE: MONTANA

| ZIP CODE | QUARTER 1 | QUARTER 2 | QUARTER 3 | QUARTER 4 | TOTAL GRAMS |
|---|---|---|---|---|---|
| 590 | 0 | 0 | 0.36 | 0.36 | 0.72 |
| 591 | 19.8 | 18.72 | 23.76 | 17.28 | 79.56 |
| 592 | 0.72 | 2.88 | 0 | 1.44 | 5.04 |
| 593 | 0.36 | 0 | 0 | 0 | 0.36 |
| 594 | 9.36 | 6.48 | 6.12 | 10.8 | 32.76 |

DRUG CODE:9639DRUG NAME:OPIUM POWDERED
STATE:MONTANA

| ZIP CODE | QUARTER 1 | QUARTER 2 | QUARTER 3 | QUARTER 4 | TOTAL GRAMS |
|---|---|---|---|---|---|
| 595 | 1.44 | 1.8 | 2.16 | 2.16 | 7.56 |
| 596 | 1.44 | 0.72 | 3.96 | 3.6 | 9.72 |
| 597 | 1.44 | 2.88 | 2.52 | 2.16 | 9 |
| 598 | 10.8 | 15.12 | 11.52 | 14.76 | 52.2 |
| 599 | 5.4 | 5.76 | 5.04 | 3.6 | 19.8 |
| TOTAL | 50.76 | 54.36 | 55.44 | 56.16 | 216.72 |

STATE: NEBRASKA

| ZIP CODE | QUARTER 1 | QUARTER 2 | QUARTER 3 | QUARTER 4 | TOTAL GRAMS |
|---|---|---|---|---|---|
| 680 | 15.12 | 20.52 | 23.4 | 11.52 | 70.56 |
| 681 | 39.6 | 49.32 | 41.76 | 45 | 175.68 |
| 683 | 0.72 | 3.96 | 6.12 | 5.04 | 15.84 |
| 684 | 1.08 | 0.36 | 0.72 | 0 | 2.16 |
| 685 | 10.08 | 2.88 | 7.56 | 7.2 | 27.72 |
| 686 | 0.72 | 0 | 1.44 | 0.36 | 2.52 |
| 687 | 1.8 | 3.6 | 1.08 | 2.16 | 8.64 |
| 688 | 17.28 | 27.36 | 13.32 | 19.44 | 77.4 |
| 689 | 1.08 | 5.04 | 3.24 | 4.68 | 14.04 |
| 690 | 0 | 0 | 0 | 1.08 | 1.08 |
| 691 | 2.16 | 2.16 | 3.24 | 3.24 | 10.8 |
| 693 | 2.16 | 2.16 | 1.08 | 1.8 | 7.2 |
| TOTAL | 91.8 | 117.36 | 102.96 | 101.52 | 413.64 |

STATE: NEVADA

| ZIP CODE | QUARTER 1 | QUARTER 2 | QUARTER 3 | QUARTER 4 | TOTAL GRAMS |
|---|---|---|---|---|---|
| 890 | 9.72 | 3.24 | 3.6 | 6.12 | 22.68 |
| 891 | 16.56 | 21.96 | 9.36 | 6.84 | 54.72 |
| 894 | 0 | 3.24 | 0.72 | 1.08 | 5.04 |
| 895 | 10.44 | 8.28 | 14.76 | 5.4 | 38.88 |
| 897 | 0 | 2.16 | 2.16 | 2.16 | 6.48 |
| 898 | 1.08 | 0 | 0.36 | 0 | 1.44 |

| | | | | | |
|---|---|---|---|---|---|
| TOTAL | 37.8 | 38.88 | 30.96 | 21.6 | 129.24 |

STATE: NEW HAMPSHIRE

| ZIP CODE | QUARTER 1 | QUARTER 2 | QUARTER 3 | QUARTER 4 | TOTAL GRAMS |
|---|---|---|---|---|---|
| 30 | 7.56 | 13.32 | 9.72 | 15.84 | 46.44 |
| 31 | 0.72 | 5.76 | 1.44 | 3.24 | 11.16 |
| 32 | 0 | 0.36 | 0 | 0 | 0.36 |
| 33 | 9.36 | 7.92 | 9.36 | 8.64 | 35.28 |
| 34 | 0 | 0 | 0 | 0.36 | 0.36 |
| 35 | 0.72 | 1.44 | 1.44 | 1.44 | 5.04 |
| 37 | 5.04 | 10.8 | 11.16 | 4.32 | 31.32 |
| 38 | 3.96 | 9.36 | 3.96 | 2.52 | 19.8 |
| TOTAL | 27.36 | 48.96 | 37.08 | 36.36 | 149.76 |

STATE: NEW JERSEY

| ZIP CODE | QUARTER 1 | QUARTER 2 | QUARTER 3 | QUARTER 4 | TOTAL GRAMS |
|---|---|---|---|---|---|
| 70 | 33.48 | 41.76 | 38.16 | 21.24 | 134.64 |
| 72 | 0 | 1.08 | 0 | 0 | 1.08 |
| 73 | 1.8 | 0 | 0 | 0 | 1.8 |
| 74 | 3.24 | 2.52 | 6.84 | 7.92 | 20.52 |
| 76 | 11.88 | 10.08 | 7.2 | 4.68 | 33.84 |
| 77 | 4.68 | 5.76 | 5.04 | 6.48 | 21.96 |
| 78 | 1.44 | 8.28 | 4.32 | 1.44 | 15.48 |
| 79 | 0.72 | 0.36 | 1.08 | 3.6 | 5.76 |
| 80 | 41.4 | 49.32 | 49.68 | 64.08 | 204.48 |
| 81 | 2.16 | 3.24 | 2.88 | 1.44 | 9.72 |
| 82 | 0 | 1.08 | 1.8 | 1.08 | 3.96 |
| 83 | 2.52 | 3.6 | 4.68 | 2.88 | 13.68 |
| 84 | 0 | 0 | 0 | 0.36 | 0.36 |
| 85 | 1.08 | 4.68 | 1.8 | 3.96 | 11.52 |
| 86 | 0.36 | 0.72 | 1.8 | 1.08 | 3.96 |
| 87 | 1.08 | 1.44 | 1.44 | 2.52 | 6.48 |
| 88 | 12.6 | 10.08 | 10.8 | 15.48 | 48.96 |
| 89 | 6.48 | 8.64 | 5.76 | 7.2 | 28.08 |
| TOTAL | 124.92 | 152.64 | 143.28 | 145.44 | 566.28 |

STATE: NEW MEXICO

| ZIP CODE | QUARTER 1 | QUARTER 2 | QUARTER 3 | QUARTER 4 | TOTAL GRAMS |
|---|---|---|---|---|---|
| 871 | 25.92 | 27.36 | 34.56 | 31.32 | 119.16 |
| 874 | 0 | 3.6 | 0 | 1.44 | 5.04 |

DRUG CODE:9639DRUG NAME:OPIUM POWDERED
STATE:NEW MEXICO

| ZIP CODE | QUARTER 1 | QUARTER 2 | QUARTER 3 | QUARTER 4 | TOTAL GRAMS |
|---|---|---|---|---|---|
| 875 | 10.08 | 7.2 | 14.04 | 10.08 | 41.4 |
| 878 | 0 | 0 | 0.36 | 0 | 0.36 |
| 880 | 7.2 | 14.76 | 10.8 | 5.04 | 37.8 |
| 881 | 0 | 0 | 0 | 0.36 | 0.36 |
| 882 | 6.48 | 5.4 | 7.2 | 2.16 | 21.24 |
| 883 | 0.36 | 1.08 | 0 | 0 | 1.44 |
| TOTAL | 50.04 | 59.4 | 66.96 | 50.4 | 226.8 |

STATE: NEW YORK

| ZIP CODE | QUARTER 1 | QUARTER 2 | QUARTER 3 | QUARTER 4 | TOTAL GRAMS |
|---|---|---|---|---|---|
| 100 | 36.36 | 32.4 | 30.24 | 34.2 | 133.2 |
| 101 | 0.72 | 0 | 0 | 0 | 0.72 |
| 103 | 0 | 0.72 | 1.08 | 0 | 1.8 |
| 104 | 1.44 | 0.72 | 0.72 | 1.08 | 3.96 |
| 105 | 3.6 | 2.88 | 5.04 | 3.24 | 14.76 |

| | | | | | |
|---|---|---|---|---|---|
| 106 | 0.36 | 1.08 | 0.36 | 0 | 1.8 |
| 109 | 1.44 | 5.04 | 0 | 0.36 | 6.84 |
| 110 | 5.4 | 5.76 | 15.12 | 8.28 | 34.56 |
| 112 | 2.16 | 3.24 | 4.68 | 4.68 | 14.76 |
| 113 | 0 | 0 | 0 | 1.8 | 1.8 |
| 114 | 0.72 | 1.08 | 0 | 0.36 | 2.16 |
| 115 | 2.52 | 1.44 | 1.8 | 1.8 | 7.56 |
| 117 | 18 | 20.16 | 27 | 23.4 | 88.56 |
| 118 | 0 | 0.36 | 1.44 | 0 | 1.8 |
| 119 | 0 | 5.76 | 1.8 | 0.72 | 8.28 |
| 120 | 0.36 | 0.72 | 0.36 | 1.8 | 3.24 |
| 121 | 1.8 | 9.36 | 1.44 | 3.6 | 16.2 |
| 122 | 4.32 | 9.36 | 9.36 | 4.68 | 27.72 |
| 123 | 4.32 | 5.4 | 10.44 | 7.56 | 27.72 |
| 125 | 0 | 0 | 0 | 0.72 | 0.72 |
| 128 | 0 | 5.04 | 2.16 | 4.32 | 11.52 |
| 129 | 9 | 6.84 | 21.6 | 5.76 | 43.2 |
| 130 | 3.96 | 12.24 | 0.36 | 1.44 | 18 |
| 131 | 1.44 | 0.36 | 0.36 | 1.44 | 3.6 |
| 132 | 1.8 | 15.48 | 15.48 | 11.88 | 44.64 |
| 133 | 2.16 | 0 | 5.04 | 2.16 | 9.36 |
| 134 | 5.04 | 5.76 | 5.04 | 4.32 | 20.16 |
| 135 | 3.96 | 4.32 | 2.16 | 2.52 | 12.96 |
| 136 | 5.4 | 3.96 | 7.2 | 3.24 | 19.8 |
| 137 | 0.36 | 0.72 | 0 | 0.72 | 1.8 |
| 138 | 1.44 | 0 | 0 | 0 | 1.44 |
| 139 | 3.6 | 1.8 | 2.16 | 6.48 | 14.04 |
| 140 | 3.24 | 6.48 | 6.12 | 1.44 | 17.28 |
| 141 | 1.08 | 1.08 | 3.24 | 0 | 5.4 |
| 142 | 33.48 | 38.16 | 44.64 | 50.4 | 166.68 |
| 143 | 2.16 | 0.72 | 2.52 | 1.08 | 6.48 |
| 144 | 0.72 | 1.08 | 4.32 | 3.24 | 9.36 |
| 145 | 4.32 | 10.8 | 6.12 | 4.32 | 25.56 |
| 146 | 9.72 | 7.2 | 19.08 | 7.92 | 43.92 |
| 147 | 1.44 | 0.36 | 1.44 | 1.8 | 5.04 |
| 148 | 1.44 | 5.4 | 2.52 | 1.8 | 11.16 |
| 149 | 2.52 | 0.72 | 2.88 | 0.72 | 6.84 |
| TOTAL | 181.8 | 234 | 265.32 | 215.28 | 896.4 |

STATE: NORTH CAROLINA

| ZIP CODE | QUARTER 1 | QUARTER 2 | QUARTER 3 | QUARTER 4 | TOTAL GRAMS |
|---|---|---|---|---|---|
| 270 | 1.44 | 2.52 | 1.44 | 1.08 | 6.48 |
| 271 | 11.52 | 16.56 | 10.08 | 13.68 | 51.84 |
| 272 | 34.56 | 30.6 | 26.64 | 20.88 | 112.68 |
| 273 | 8.64 | 3.96 | 10.08 | 6.84 | 29.52 |
| 274 | 7.2 | 8.64 | 10.08 | 15.48 | 41.4 |
| 275 | 31.68 | 22.68 | 16.92 | 28.8 | 100.08 |
| 276 | 12.6 | 15.12 | 14.76 | 10.8 | 53.28 |
| 277 | 6.48 | 6.48 | 18.36 | 13.32 | 44.64 |
| 278 | 10.08 | 7.92 | 8.28 | 6.84 | 33.12 |
| 279 | 2.16 | 7.56 | 0 | 3.6 | 13.32 |
| 280 | 6.84 | 5.4 | 7.2 | 6.48 | 25.92 |
| 281 | 18 | 16.56 | 21.6 | 27.36 | 83.52 |

DRUG CODE:9639DRUG NAME:OPIUM POWDERED
STATE:NORTH CAROLINA

| ZIP CODE | QUARTER 1 | QUARTER 2 | QUARTER 3 | QUARTER 4 | TOTAL GRAMS |
|---|---|---|---|---|---|
| 282 | 23.76 | 19.44 | 23.76 | 24.48 | 91.44 |
| 283 | 12.96 | 14.76 | 6.48 | 9 | 43.2 |
| 284 | 19.44 | 25.56 | 25.2 | 19.08 | 89.28 |

| | | | | | |
|---|---|---|---|---|---|
| 285 | 6.12 | 5.76 | 6.12 | 5.04 | 23.04 |
| 286 | 10.08 | 10.8 | 9.72 | 8.28 | 38.88 |
| 287 | 4.32 | 9.36 | 9.72 | 12.24 | 35.64 |
| 288 | 2.16 | 2.52 | 3.6 | 1.08 | 9.36 |
| 289 | 0 | 0.72 | 0.72 | 0 | 1.44 |
| TOTAL | 230.04 | 232.92 | 230.76 | 234.36 | 928.08 |

STATE: NORTH DAKOTA

| ZIP CODE | QUARTER 1 | QUARTER 2 | QUARTER 3 | QUARTER 4 | TOTAL GRAMS |
|---|---|---|---|---|---|
| 581 | 19.08 | 11.16 | 9 | 11.88 | 51.12 |
| 582 | 17.28 | 7.92 | 10.08 | 0.72 | 36 |
| 583 | 0 | 0.72 | 0.72 | 0 | 1.44 |
| 584 | 2.16 | 0.72 | 0.72 | 1.44 | 5.04 |
| 585 | 14.76 | 10.08 | 12.96 | 14.76 | 52.56 |
| 586 | 1.08 | 0.36 | 0 | 0.36 | 1.8 |
| 587 | 0 | 0 | 3.96 | 0 | 3.96 |
| 588 | 0.72 | 0.72 | 0.36 | 0.36 | 2.16 |
| TOTAL | 55.08 | 31.68 | 37.8 | 29.52 | 154.08 |

STATE: OHIO

| ZIP CODE | QUARTER 1 | QUARTER 2 | QUARTER 3 | QUARTER 4 | TOTAL GRAMS |
|---|---|---|---|---|---|
| 430 | 12.6 | 9 | 11.88 | 13.68 | 47.16 |
| 431 | 2.88 | 5.4 | 5.76 | 3.96 | 18 |
| 432 | 46.44 | 44.28 | 32.4 | 46.44 | 169.56 |
| 433 | 3.24 | 4.32 | 3.24 | 2.16 | 12.96 |
| 434 | 1.08 | 1.8 | 0.72 | 1.8 | 5.4 |
| 435 | 7.2 | 9.36 | 3.96 | 12.6 | 33.12 |
| 436 | 23.76 | 21.96 | 27 | 27.72 | 100.44 |
| 437 | 1.8 | 2.52 | 1.44 | 4.32 | 10.08 |
| 439 | 1.44 | 0.72 | 2.16 | 2.88 | 7.2 |
| 440 | 12.24 | 5.76 | 12.24 | 13.32 | 43.56 |
| 441 | 83.16 | 79.56 | 91.08 | 64.08 | 317.88 |
| 442 | 23.04 | 14.4 | 12.24 | 10.8 | 60.48 |
| 443 | 20.88 | 23.76 | 21.6 | 18.72 | 84.96 |
| 444 | 5.04 | 19.8 | 2.16 | 1.44 | 28.44 |
| 445 | 18.36 | 12.96 | 10.08 | 9.36 | 50.76 |
| 446 | 3.6 | 9 | 4.68 | 3.24 | 20.52 |
| 447 | 11.88 | 6.48 | 9 | 14.4 | 41.76 |
| 448 | 4.32 | 9 | 6.12 | 8.64 | 28.08 |
| 449 | 0.72 | 3.96 | 0 | 2.16 | 6.84 |
| 450 | 14.04 | 42.84 | 40.32 | 29.88 | 127.08 |
| 451 | 7.56 | 12.24 | 37.08 | 33.12 | 90 |
| 452 | 55.8 | 60.12 | 60.48 | 41.76 | 218.16 |
| 453 | 4.68 | 9.72 | 9.36 | 8.64 | 32.4 |
| 454 | 28.08 | 19.08 | 33.48 | 32.04 | 112.68 |
| 455 | 1.08 | 2.52 | 3.6 | 1.44 | 8.64 |
| 456 | 7.2 | 8.28 | 9 | 2.16 | 26.64 |
| 457 | 4.68 | 1.08 | 1.8 | 1.8 | 9.36 |
| 458 | 2.88 | 4.68 | 4.68 | 3.24 | 15.48 |
| TOTAL | 409.68 | 444.6 | 457.56 | 415.8 | 1,727.64 |

STATE: OKLAHOMA

| ZIP CODE | QUARTER 1 | QUARTER 2 | QUARTER 3 | QUARTER 4 | TOTAL GRAMS |
|---|---|---|---|---|---|
| 730 | 2.88 | 1.08 | 3.96 | 2.88 | 10.8 |
| 731 | 20.16 | 21.96 | 29.88 | 25.92 | 97.92 |
| 735 | 3.24 | 2.16 | 5.76 | 5.04 | 16.2 |
| 736 | 0.36 | 0.72 | 0.72 | 0.36 | 2.16 |
| 737 | 0 | 0 | 1.44 | 1.44 | 2.88 |
| 738 | 2.52 | 3.6 | 1.8 | 4.32 | 12.24 |
| 740 | 3.24 | 2.16 | 3.6 | 3.96 | 12.96 |
| 741 | 22.32 | 18.36 | 27.36 | 25.56 | 93.6 |

| | | | | | |
|---|---|---|---|---|---|
| 743 | 11.16 | 5.76 | 4.32 | 7.2 | 28.44 |
| 744 | 0 | 2.16 | 1.44 | 6.84 | 10.44 |
| 745 | 1.8 | 2.52 | 2.16 | 2.88 | 9.36 |
| 746 | 0 | 0.72 | 0 | 0 | 0.72 |
| 747 | 1.44 | 2.16 | 2.16 | 2.88 | 8.64 |

DRUG CODE:9639DRUG NAME:OPIUM POWDERED
STATE:OKLAHOMA

| ZIP CODE | QUARTER 1 | QUARTER 2 | QUARTER 3 | QUARTER 4 | TOTAL GRAMS |
|---|---|---|---|---|---|
| 748 | 0 | 0.72 | 0 | 1.44 | 2.16 |
| TOTAL | 69.12 | 64.08 | 84.6 | 90.72 | 308.52 |

STATE: OREGON

| ZIP CODE | QUARTER 1 | QUARTER 2 | QUARTER 3 | QUARTER 4 | TOTAL GRAMS |
|---|---|---|---|---|---|
| 970 | 19.08 | 18 | 22.68 | 19.8 | 79.56 |
| 971 | 12.24 | 10.8 | 14.4 | 11.88 | 49.32 |
| 972 | 64.8 | 52.56 | 47.16 | 73.8 | 238.32 |
| 973 | 33.84 | 35.28 | 25.92 | 15.48 | 110.52 |
| 974 | 38.52 | 33.84 | 36.36 | 38.88 | 147.6 |
| 975 | 37.8 | 37.44 | 34.2 | 55.44 | 164.88 |
| 976 | 1.08 | 1.8 | 1.8 | 2.88 | 7.56 |
| 977 | 2.52 | 8.64 | 3.96 | 4.32 | 19.44 |
| 978 | 0.72 | 3.96 | 2.52 | 2.52 | 9.72 |
| 979 | 0.36 | 0 | 0 | 0.36 | 0.72 |
| TOTAL | 210.96 | 202.32 | 189 | 225.36 | 827.64 |

STATE: PENNSYLVANIA

| ZIP CODE | QUARTER 1 | QUARTER 2 | QUARTER 3 | QUARTER 4 | TOTAL GRAMS |
|---|---|---|---|---|---|
| 150 | 8.64 | 3.96 | 4.32 | 3.24 | 20.16 |
| 151 | 1.08 | 1.44 | 3.6 | 1.08 | 7.2 |
| 152 | 13.68 | 11.52 | 6.12 | 10.08 | 41.4 |
| 153 | 5.4 | 1.8 | 1.08 | 3.96 | 12.24 |
| 154 | 0 | 0.36 | 0 | 0 | 0.36 |
| 155 | 0.36 | 0.72 | 0.72 | 0.72 | 2.52 |
| 156 | 5.76 | 4.68 | 2.88 | 2.16 | 15.48 |
| 157 | 2.88 | 1.8 | 6.12 | 3.24 | 14.04 |
| 158 | 3.24 | 1.8 | 3.24 | 3.96 | 12.24 |
| 159 | 2.88 | 0 | 2.16 | 0 | 5.04 |
| 160 | 0 | 0 | 2.52 | 1.44 | 3.96 |
| 161 | 0.36 | 8.64 | 5.4 | 3.96 | 18.36 |
| 162 | 0.72 | 1.08 | 0.72 | 1.08 | 3.6 |
| 163 | 3.24 | 1.8 | 3.96 | 1.44 | 10.44 |
| 164 | 0.72 | 0 | 0 | 1.44 | 2.16 |
| 165 | 3.6 | 4.68 | 4.68 | 3.6 | 16.56 |
| 166 | 6.84 | 9.72 | 1.8 | 2.52 | 20.88 |
| 167 | 0.36 | 0.36 | 0.36 | 0.72 | 1.8 |
| 168 | 5.76 | 9 | 10.08 | 9.72 | 34.56 |
| 169 | 0 | 2.16 | 0.36 | 0 | 2.52 |
| 170 | 16.56 | 16.2 | 15.84 | 18 | 66.6 |
| 171 | 2.52 | 4.32 | 6.12 | 7.92 | 20.88 |
| 172 | 19.44 | 9.36 | 16.56 | 14.04 | 59.4 |
| 173 | 3.6 | 5.04 | 7.92 | 6.48 | 23.04 |
| 174 | 5.76 | 0.72 | 4.32 | 2.88 | 13.68 |
| 175 | 8.64 | 8.64 | 1.44 | 1.44 | 20.16 |
| 176 | 11.88 | 10.44 | 5.76 | 1.08 | 29.16 |
| 177 | 1.44 | 1.44 | 1.8 | 0.72 | 5.4 |
| 178 | 5.04 | 6.84 | 7.92 | 7.56 | 27.36 |
| 179 | 7.56 | 7.56 | 7.56 | 10.08 | 32.76 |
| 180 | 17.64 | 23.76 | 28.08 | 24.84 | 94.32 |
| 181 | 3.24 | 21.6 | 7.56 | 2.88 | 35.28 |

| | | | | | |
|---|---|---|---|---|---|
| 182 | 1.08 | 2.52 | 1.44 | 4.32 | 9.36 |
| 183 | 0 | 0.72 | 3.6 | 1.08 | 5.4 |
| 184 | 0.72 | 1.8 | 1.44 | 0.72 | 4.68 |
| 185 | 11.52 | 10.8 | 13.32 | 11.52 | 47.16 |
| 186 | 3.24 | 2.16 | 9 | 3.96 | 18.36 |
| 187 | 27 | 21.6 | 39.24 | 11.88 | 99.72 |
| 188 | 1.44 | 2.16 | 1.44 | 1.44 | 6.48 |
| 189 | 3.24 | 1.8 | 3.6 | 0 | 8.64 |
| 190 | 55.44 | 50.04 | 58.32 | 77.4 | 241.2 |
| 191 | 54.72 | 66.6 | 47.52 | 47.16 | 216 |
| 193 | 21.6 | 14.04 | 19.8 | 11.52 | 66.96 |
| 194 | 24.48 | 39.6 | 5.4 | 2.52 | 72 |
| 195 | 0 | 1.8 | 0 | 0 | 1.8 |
| TOTAL | 373.32 | 397.08 | 375.12 | 325.8 | 1,471.32 |

STATE: RHODE ISLAND

| ZIP CODE | QUARTER 1 | QUARTER 2 | QUARTER 3 | QUARTER 4 | TOTAL GRAMS |
|---|---|---|---|---|---|
| 28 | 10.8 | 11.16 | 15.84 | 12.6 | 50.4 |

DRUG CODE:9639DRUG NAME:OPIUM POWDERED
STATE:RHODE ISLAND

| ZIP CODE | QUARTER 1 | QUARTER 2 | QUARTER 3 | QUARTER 4 | TOTAL GRAMS |
|---|---|---|---|---|---|
| 29 | 35.64 | 12.24 | 16.92 | 17.28 | 82.08 |
| TOTAL | 46.44 | 23.4 | 32.76 | 29.88 | 132.48 |

STATE: SOUTH CAROLINA

| ZIP CODE | QUARTER 1 | QUARTER 2 | QUARTER 3 | QUARTER 4 | TOTAL GRAMS |
|---|---|---|---|---|---|
| 290 | 2.16 | 2.16 | 2.88 | 3.6 | 10.8 |
| 291 | 3.24 | 10.08 | 6.48 | 10.44 | 30.24 |
| 292 | 9.36 | 6.84 | 9.36 | 10.44 | 36 |
| 293 | 6.12 | 13.32 | 7.56 | 11.52 | 38.52 |
| 294 | 17.28 | 23.76 | 36.36 | 30.24 | 107.64 |
| 295 | 20.88 | 26.64 | 27.72 | 28.44 | 103.68 |
| 296 | 31.68 | 34.92 | 25.56 | 23.76 | 115.92 |
| 297 | 18.36 | 8.64 | 5.76 | 6.84 | 39.6 |
| 298 | 9.36 | 9.36 | 7.2 | 10.08 | 36 |
| 299 | 17.64 | 16.2 | 18 | 14.4 | 66.24 |
| TOTAL | 136.08 | 151.92 | 146.88 | 149.76 | 584.64 |

STATE: SOUTH DAKOTA

| ZIP CODE | QUARTER 1 | QUARTER 2 | QUARTER 3 | QUARTER 4 | TOTAL GRAMS |
|---|---|---|---|---|---|
| 570 | 3.6 | 2.52 | 2.52 | 3.6 | 12.24 |
| 571 | 11.16 | 9.36 | 16.2 | 19.08 | 55.8 |
| 572 | 2.16 | 1.44 | 2.52 | 2.52 | 8.64 |
| 573 | 0.72 | 0.72 | 0.36 | 1.08 | 2.88 |
| 574 | 0.72 | 2.16 | 0 | 0 | 2.88 |
| 575 | 0.72 | 0.36 | 1.08 | 0.36 | 2.52 |
| 577 | 2.88 | 0 | 1.08 | 1.44 | 5.4 |
| TOTAL | 21.96 | 16.56 | 23.76 | 28.08 | 90.36 |

STATE: TENNESSEE

| ZIP CODE | QUARTER 1 | QUARTER 2 | QUARTER 3 | QUARTER 4 | TOTAL GRAMS |
|---|---|---|---|---|---|
| 370 | 5.04 | 12.6 | 15.12 | 12.6 | 45.36 |
| 371 | 5.4 | 3.24 | 10.8 | 2.88 | 22.32 |
| 372 | 28.8 | 31.32 | 24.48 | 29.16 | 113.76 |
| 373 | 3.6 | 7.2 | 9.36 | 6.12 | 26.28 |
| 374 | 6.84 | 9.36 | 7.92 | 9 | 33.12 |
| 376 | 19.8 | 29.88 | 33.12 | 24.84 | 107.64 |
| 377 | 7.2 | 5.76 | 9.36 | 12.6 | 34.92 |

| | | | | | |
|---|---|---|---|---|---|
| 378 | 9.36 | 12.24 | 12.96 | 9.36 | 43.92 |
| 379 | 26.64 | 37.44 | 39.6 | 24.48 | 128.16 |
| 380 | 1.8 | 1.08 | 2.16 | 3.24 | 8.28 |
| 381 | 49.68 | 74.16 | 86.76 | 100.08 | 310.68 |
| 382 | 2.52 | 1.8 | 1.08 | 2.16 | 7.56 |
| 383 | 15.84 | 10.44 | 14.4 | 14.4 | 55.08 |
| 384 | 3.24 | 1.44 | 3.24 | 2.16 | 10.08 |
| 385 | 2.88 | 1.08 | 5.76 | 1.8 | 11.52 |
| TOTAL | 188.64 | 239.04 | 276.12 | 254.88 | 958.68 |

STATE: TEXAS

| ZIP CODE | QUARTER 1 | QUARTER 2 | QUARTER 3 | QUARTER 4 | TOTAL GRAMS |
|---|---|---|---|---|---|
| 750 | 33.84 | 37.8 | 45.36 | 44.64 | 161.64 |
| 751 | 1.08 | 4.32 | 5.04 | 7.2 | 17.64 |
| 752 | 61.2 | 65.52 | 69.12 | 65.88 | 261.72 |
| 753 | 9.36 | 8.64 | 1.44 | 8.64 | 28.08 |
| 754 | 3.6 | 3.96 | 1.44 | 3.96 | 12.96 |
| 755 | 6.84 | 4.32 | 6.48 | 5.4 | 23.04 |
| 756 | 7.56 | 11.16 | 8.64 | 8.64 | 36 |
| 757 | 30.6 | 33.12 | 18 | 27.72 | 109.44 |
| 759 | 15.48 | 16.92 | 17.64 | 17.28 | 67.32 |
| 760 | 19.44 | 31.68 | 43.56 | 36.72 | 131.4 |
| 761 | 30.96 | 20.16 | 36.36 | 19.44 | 106.92 |
| 762 | 8.64 | 3.96 | 4.68 | 5.76 | 23.04 |
| 763 | 3.6 | 4.68 | 2.52 | 2.88 | 13.68 |
| 764 | 2.88 | 2.52 | 0 | 0.36 | 5.76 |
| 765 | 2.52 | 5.04 | 9.72 | 14.04 | 31.32 |
| 766 | 0 | 0.72 | 0.72 | 1.08 | 2.52 |
| 767 | 7.56 | 3.96 | 3.24 | 3.96 | 18.72 |
| 768 | 1.8 | 0.72 | 0.72 | 0.72 | 3.96 |
| 769 | 2.16 | 3.6 | 1.08 | 1.44 | 8.28 |
| 770 | 57.24 | 57.6 | 54.36 | 47.52 | 216.72 |
| 773 | 3.24 | 3.96 | 7.2 | 2.16 | 16.56 |

DRUG CODE:9639DRUG NAME:OPIUM POWDERED
STATE:TEXAS

| ZIP CODE | QUARTER 1 | QUARTER 2 | QUARTER 3 | QUARTER 4 | TOTAL GRAMS |
|---|---|---|---|---|---|
| 774 | 7.2 | 5.4 | 2.16 | 5.76 | 20.52 |
| 775 | 20.88 | 21.24 | 22.32 | 19.44 | 83.88 |
| 776 | 0 | 0.72 | 0 | 0 | 0.72 |
| 777 | 2.52 | 1.44 | 1.08 | 1.44 | 6.48 |
| 778 | 7.2 | 7.2 | 6.84 | 7.2 | 28.44 |
| 779 | 3.96 | 0.72 | 1.8 | 5.4 | 11.88 |
| 780 | 1.44 | 0.36 | 0.72 | 0.72 | 3.24 |
| 781 | 4.32 | 9.36 | 2.52 | 2.52 | 18.72 |
| 782 | 57.24 | 50.76 | 42.12 | 45 | 195.12 |
| 784 | 2.88 | 2.16 | 1.8 | 2.16 | 9 |
| 785 | 2.88 | 1.8 | 3.96 | 2.88 | 11.52 |
| 786 | 29.88 | 18.36 | 26.28 | 28.08 | 102.6 |
| 787 | 24.84 | 27.72 | 40.32 | 37.8 | 130.68 |
| 788 | 4.68 | 0.72 | 1.44 | 1.44 | 8.28 |
| 789 | 0 | 1.08 | 0 | 0 | 1.08 |
| 790 | 4.32 | 3.24 | 4.32 | 2.88 | 14.76 |
| 791 | 9.72 | 10.44 | 17.28 | 12.96 | 50.4 |
| 792 | 0.72 | 0 | 0.72 | 0 | 1.44 |
| 794 | 3.24 | 15.12 | 11.88 | 6.48 | 36.72 |
| 796 | 1.8 | 6.48 | 1.44 | 1.44 | 11.16 |
| 797 | 0.36 | 4.32 | 2.88 | 4.32 | 11.88 |
| 799 | 7.56 | 6.84 | 18 | 1.44 | 33.84 |

| | | | | | |
|---|---|---|---|---|---|
| TOTAL | 507.24 | 519.84 | 547.2 | 514.8 | 2,089.08 |
| STATE: | UTAH | | | | |
| ZIP CODE | QUARTER 1 | QUARTER 2 | QUARTER 3 | QUARTER 4 | TOTAL GRAMS |
| 840 | 23.04 | 28.44 | 23.4 | 25.2 | 100.08 |
| 841 | 23.04 | 26.64 | 23.76 | 24.48 | 97.92 |
| 843 | 1.44 | 2.88 | 3.6 | 2.52 | 10.44 |
| 844 | 1.8 | 3.6 | 1.08 | 1.8 | 8.28 |
| 845 | 0 | 0 | 0.72 | 0 | 0.72 |
| 846 | 0.36 | 5.04 | 3.96 | 6.12 | 15.48 |
| 847 | 3.6 | 3.6 | 3.6 | 3.6 | 14.4 |
| TOTAL | 53.28 | 70.2 | 60.12 | 63.72 | 247.32 |
| STATE: | VERMONT | | | | |
| ZIP CODE | QUARTER 1 | QUARTER 2 | QUARTER 3 | QUARTER 4 | TOTAL GRAMS |
| 50 | 15.12 | 9.72 | 2.16 | 2.52 | 29.52 |
| 51 | 0 | 0.36 | 0 | 0 | 0.36 |
| 53 | 0.36 | 0 | 0 | 0 | 0.36 |
| 54 | 30.24 | 13.68 | 14.04 | 16.92 | 74.88 |
| 56 | 2.16 | 4.32 | 4.68 | 5.76 | 16.92 |
| 57 | 8.28 | 7.2 | 7.92 | 5.04 | 28.44 |
| 58 | 0 | 0.72 | 1.44 | 0.36 | 2.52 |
| TOTAL | 56.16 | 36 | 30.24 | 30.6 | 153 |
| STATE: | VIRGIN ISLANDS | | | | |
| ZIP CODE | QUARTER 1 | QUARTER 2 | QUARTER 3 | QUARTER 4 | TOTAL GRAMS |
| 8 | 0 | 0.72 | 0.72 | 0.36 | 1.8 |
| TOTAL | 0 | 0.72 | 0.72 | 0.36 | 1.8 |
| STATE: | VIRGINIA | | | | |
| ZIP CODE | QUARTER 1 | QUARTER 2 | QUARTER 3 | QUARTER 4 | TOTAL GRAMS |
| 201 | 16.56 | 19.44 | 12.6 | 13.68 | 62.28 |
| 220 | 16.56 | 9.72 | 12.96 | 9.72 | 48.96 |
| 221 | 1.8 | 5.4 | 4.32 | 11.16 | 22.68 |
| 222 | 4.32 | 3.6 | 0 | 1.08 | 9 |
| 223 | 0.36 | 0 | 2.88 | 2.16 | 5.4 |
| 224 | 2.16 | 0.72 | 4.68 | 2.88 | 10.44 |
| 225 | 0 | 0.72 | 0 | 0 | 0.72 |
| 226 | 10.44 | 7.92 | 2.88 | 3.6 | 24.84 |
| 228 | 4.32 | 1.44 | 1.44 | 2.88 | 10.08 |
| 229 | 8.28 | 23.76 | 23.4 | 38.88 | 94.32 |
| 230 | 2.16 | 0.72 | 0 | 3.6 | 6.48 |
| 231 | 12.24 | 9.72 | 15.84 | 13.32 | 51.12 |
| 232 | 15.12 | 31.68 | 15.84 | 23.76 | 86.4 |
| 233 | 4.68 | 6.12 | 1.44 | 5.04 | 17.28 |
| 234 | 9 | 10.44 | 16.2 | 11.88 | 47.52 |
| 235 | 11.16 | 4.68 | 9.72 | 7.2 | 32.76 |
| 236 | 4.68 | 5.4 | 7.92 | 3.24 | 21.24 |

DRUG CODE:9639DRUG NAME:OPIUM POWDERED
STATE:VIRGINIA

| ZIP CODE | QUARTER 1 | QUARTER 2 | QUARTER 3 | QUARTER 4 | TOTAL GRAMS |
|---|---|---|---|---|---|
| 237 | 21.96 | 9 | 12.24 | 14.04 | 57.24 |
| 238 | 4.32 | 5.04 | 1.44 | 0 | 10.8 |
| 239 | 1.8 | 0 | 0.36 | 0.36 | 2.52 |
| 240 | 4.68 | 4.32 | 4.32 | 8.64 | 21.96 |
| 241 | 2.52 | 2.16 | 6.12 | 2.52 | 13.32 |
| 242 | 3.6 | 5.4 | 3.6 | 0.72 | 13.32 |
| 243 | 0 | 0.72 | 1.08 | 1.44 | 3.24 |
| 244 | 1.44 | 0.72 | 1.8 | 0.72 | 4.68 |

| | | | | | | |
|---|---|---|---|---|---|---|
| | 245 | 7.56 | 9 | 6.12 | 5.04 | 27.72 |
| | 246 | 4.32 | 0 | 3.6 | 2.16 | 10.08 |
| TOTAL | | 176.04 | 177.84 | 172.8 | 189.72 | 716.4 |
| STATE: | WASHINGTON | | | | | |
| ZIP CODE | QUARTER 1 | | QUARTER 2 | QUARTER 3 | QUARTER 4 | TOTAL GRAMS |
| | 980 | 61.2 | 76.32 | 57.96 | 52.92 | 248.4 |
| | 981 | 76.32 | 66.96 | 52.92 | 69.84 | 266.04 |
| | 982 | 28.44 | 34.2 | 37.8 | 33.12 | 133.56 |
| | 983 | 26.28 | 31.32 | 36.36 | 28.8 | 122.76 |
| | 984 | 23.4 | 24.84 | 27.72 | 18.36 | 94.32 |
| | 985 | 20.88 | 11.88 | 11.16 | 20.16 | 64.08 |
| | 986 | 30.6 | 29.16 | 25.92 | 21.24 | 106.92 |
| | 988 | 30.6 | 55.44 | 41.04 | 38.52 | 165.6 |
| | 989 | 0 | 4.32 | 0.72 | 0.72 | 5.76 |
| | 991 | 2.16 | 4.32 | 2.88 | 2.16 | 11.52 |
| | 992 | 12.96 | 18 | 11.88 | 10.08 | 52.92 |
| | 993 | 4.32 | 10.8 | 10.08 | 9.36 | 34.56 |
| | 994 | 0.72 | 0.72 | 0.72 | 1.08 | 3.24 |
| TOTAL | | 317.88 | 368.28 | 317.16 | 306.36 | 1,309.68 |
| STATE: | WEST VIRGINIA | | | | | |
| ZIP CODE | QUARTER 1 | | QUARTER 2 | QUARTER 3 | QUARTER 4 | TOTAL GRAMS |
| | 247 | 2.88 | 0.72 | 0 | 1.44 | 5.04 |
| | 249 | 6.48 | 9.36 | 16.56 | 6.48 | 38.88 |
| | 250 | 0.72 | 0.72 | 0.72 | 1.08 | 3.24 |
| | 251 | 0 | 0.36 | 0 | 0 | 0.36 |
| | 252 | 1.8 | 2.88 | 3.24 | 4.32 | 12.24 |
| | 253 | 27.36 | 33.12 | 22.32 | 29.88 | 112.68 |
| | 254 | 2.16 | 2.16 | 3.24 | 0 | 7.56 |
| | 255 | 3.6 | 3.6 | 5.76 | 3.24 | 16.2 |
| | 256 | 0 | 0 | 0.72 | 0 | 0.72 |
| | 257 | 14.76 | 11.52 | 13.32 | 12.24 | 51.84 |
| | 258 | 8.64 | 11.88 | 8.64 | 6.84 | 36 |
| | 259 | 5.04 | 7.2 | 10.8 | 10.8 | 33.84 |
| | 260 | 2.52 | 2.52 | 2.16 | 2.88 | 10.08 |
| | 261 | 1.08 | 0 | 0.36 | 0 | 1.44 |
| | 262 | 4.32 | 4.68 | 2.16 | 1.08 | 12.24 |
| | 263 | 3.6 | 1.44 | 6.48 | 2.52 | 14.04 |
| | 264 | 0 | 0 | 0 | 1.08 | 1.08 |
| | 265 | 6.12 | 5.4 | 8.64 | 5.76 | 25.92 |
| | 266 | 0 | 7.2 | 10.8 | 10.8 | 28.8 |
| | 267 | 0 | 2.16 | 0 | 0 | 2.16 |
| | 268 | 0 | 0 | 0.36 | 0 | 0.36 |
| TOTAL | | 91.08 | 106.92 | 116.28 | 100.44 | 414.72 |
| STATE: | WISCONSIN | | | | | |
| ZIP CODE | QUARTER 1 | | QUARTER 2 | QUARTER 3 | QUARTER 4 | TOTAL GRAMS |
| | 530 | 28.8 | 19.8 | 20.88 | 21.24 | 90.72 |
| | 531 | 7.92 | 11.88 | 18.72 | 8.28 | 46.8 |
| | 532 | 40.68 | 47.16 | 35.28 | 30.96 | 154.08 |
| | 534 | 3.6 | 1.44 | 1.08 | 2.16 | 8.28 |
| | 535 | 6.48 | 12.6 | 5.04 | 11.88 | 36 |
| | 537 | 30.96 | 29.52 | 32.4 | 33.12 | 126 |
| | 538 | 2.88 | 0.72 | 1.08 | 0 | 4.68 |
| | 539 | 6.12 | 3.24 | 11.52 | 6.12 | 27 |
| | 540 | 3.96 | 3.6 | 5.04 | 3.6 | 16.2 |
| | 541 | 2.16 | 0.72 | 0 | 0 | 2.88 |
| | 542 | 1.08 | 2.88 | 4.32 | 1.44 | 9.72 |
| | 543 | 10.8 | 7.2 | 6.84 | 5.04 | 29.88 |
| | 544 | 5.4 | 6.84 | 10.8 | 13.32 | 36.36 |

DRUG CODE:9639DRUG NAME:OPIUM POWDERED
STATE:WISCONSIN

| ZIP CODE | QUARTER 1 | QUARTER 2 | QUARTER 3 | QUARTER 4 | TOTAL GRAMS |
|---|---|---|---|---|---|
| 545 | 2.88 | 4.68 | 3.6 | 4.32 | 15.48 |
| 546 | 12.24 | 11.52 | 18.36 | 11.88 | 54 |
| 547 | 8.28 | 14.04 | 22.68 | 9.36 | 54.36 |
| 548 | 3.96 | 0 | 2.16 | 2.16 | 8.28 |
| 549 | 22.32 | 7.92 | 12.96 | 20.88 | 64.08 |
| TOTAL | 200.52 | 185.76 | 212.76 | 185.76 | 784.8 |

STATE: WYOMING

| ZIP CODE | QUARTER 1 | QUARTER 2 | QUARTER 3 | QUARTER 4 | TOTAL GRAMS |
|---|---|---|---|---|---|
| 820 | 24.48 | 14.76 | 8.28 | 19.08 | 66.6 |
| 822 | 0 | 0 | 0 | 1.08 | 1.08 |
| 824 | 0.72 | 0 | 5.04 | 0.36 | 6.12 |
| 825 | 0 | 0 | 0.72 | 0 | 0.72 |
| 826 | 7.92 | 10.08 | 12.24 | 12.24 | 42.48 |
| 827 | 0.36 | 1.44 | 1.44 | 1.44 | 4.68 |
| 828 | 0.72 | 2.16 | 0.72 | 0.72 | 4.32 |
| 829 | 0 | 0 | 0 | 3.6 | 3.6 |
| 830 | 0.72 | 0.72 | 0 | 0.72 | 2.16 |
| TOTAL | 34.92 | 29.16 | 28.44 | 39.24 | 131.76 |

DRUG CODE:9652DRUG NAME:OXYMORPHONE
STATE: ALABAMA

| ZIP CODE | QUARTER 1 | QUARTER 2 | QUARTER 3 | QUARTER 4 | TOTAL GRAMS |
|---|---|---|---|---|---|
| 350 | 851.55 | 848.97 | 873.05 | 743.48 | 3,317.05 |
| 351 | 590.75 | 530.93 | 506.85 | 425.31 | 2,053.84 |
| 352 | 415.63 | 450.6 | 393.55 | 397.65 | 1,657.43 |
| 354 | 226.95 | 227.44 | 195.55 | 189.48 | 839.42 |
| 355 | 285.78 | 337.66 | 367.59 | 280.92 | 1,271.95 |
| 356 | 906.95 | 938.27 | 810.43 | 600.61 | 3,256.26 |
| 357 | 443.46 | 537.36 | 475.49 | 464.92 | 1,921.23 |
| 358 | 922.39 | 937.24 | 957.45 | 850.62 | 3,667.70 |
| 359 | 1,594.41 | 1,653.95 | 1,229.27 | 1,122.44 | 5,600.07 |
| 360 | 333.38 | 318.03 | 258.04 | 222.13 | 1,131.58 |
| 361 | 131.09 | 122.53 | 123.6 | 123.07 | 500.29 |
| 362 | 929.26 | 907 | 691.11 | 633.7 | 3,161.07 |
| 363 | 147.64 | 143.18 | 175.3 | 164.19 | 630.31 |
| 364 | 34.57 | 32.56 | 30.69 | 33.01 | 130.83 |
| 365 | 962.17 | 950.84 | 917.03 | 807.9 | 3,637.94 |
| 366 | 933.18 | 957.67 | 848.12 | 712.79 | 3,451.76 |
| 367 | 91.44 | 131.81 | 93.4 | 124.45 | 441.1 |
| 368 | 261.65 | 271.38 | 270.22 | 283.46 | 1,086.71 |
| 369 | 39.07 | 29.44 | 38.27 | 23.28 | 130.06 |
| TOTAL | 10,101.32 | 10,326.86 | 9,255.01 | 8,203.41 | 37,886.60 |

STATE: ALASKA

| ZIP CODE | QUARTER 1 | QUARTER 2 | QUARTER 3 | QUARTER 4 | TOTAL GRAMS |
|---|---|---|---|---|---|
| 995 | 148 | 124.63 | 108.21 | 113.74 | 494.58 |
| 996 | 176.9 | 176.73 | 177.97 | 169.63 | 701.23 |
| 997 | 17.57 | 22.48 | 18.2 | 18.02 | 76.27 |
| 998 | 0.54 | 0.27 | 0.27 | 0 | 1.08 |
| 999 | 4.28 | 4.28 | 3.75 | 4.28 | 16.59 |
| TOTAL | 347.29 | 328.39 | 308.4 | 305.67 | 1,289.75 |

STATE: ARIZONA

| ZIP CODE | QUARTER 1 | QUARTER 2 | QUARTER 3 | QUARTER 4 | TOTAL GRAMS |
|---|---|---|---|---|---|
| 850 | 1,787.47 | 1,705.94 | 1,643.35 | 1,646.19 | 6,782.95 |

| | | | | | |
|---|---|---|---|---|---|
| 851 | 426.29 | 388.11 | 392.26 | 466.84 | 1,673.50 |
| 852 | 3,038.56 | 3,016.89 | 2,859.99 | 3,050.17 | 11,965.61 |
| 853 | 1,705.92 | 1,717.87 | 1,648.82 | 1,621.39 | 6,694 |
| 855 | 162.09 | 188.5 | 191.36 | 190.91 | 732.86 |
| 856 | 290.65 | 273.74 | 266.16 | 263.04 | 1,093.59 |
| 857 | 1,519.43 | 1,415.10 | 1,266.22 | 1,275.82 | 5,476.57 |
| 859 | 53.35 | 84.66 | 91.31 | 89.61 | 318.93 |
| 860 | 84.13 | 89.83 | 105.85 | 97.82 | 377.63 |
| 863 | 257.68 | 321.25 | 296.53 | 344.26 | 1,219.72 |
| 864 | 512.96 | 482.8 | 455.64 | 442.84 | 1,894.24 |
| TOTAL | 9,838.53 | 9,684.69 | 9,217.49 | 9,488.89 | 38,229.60 |

STATE: ARKANSAS

| ZIP CODE | QUARTER 1 | QUARTER 2 | QUARTER 3 | QUARTER 4 | TOTAL GRAMS |
|---|---|---|---|---|---|
| 716 | 50.94 | 56.11 | 48.35 | 64.41 | 219.81 |

DRUG CODE:9652DRUG NAME:OXYMORPHONE
STATE:ARKANSAS

| ZIP CODE | QUARTER 1 | QUARTER 2 | QUARTER 3 | QUARTER 4 | TOTAL GRAMS |
|---|---|---|---|---|---|
| 717 | 52.19 | 58.74 | 63.96 | 46.7 | 221.59 |
| 718 | 58.88 | 54.2 | 56.92 | 52.81 | 222.81 |
| 719 | 183.59 | 169.5 | 161.83 | 129 | 643.92 |
| 720 | 392.12 | 387.57 | 346.54 | 365.94 | 1,492.17 |
| 721 | 330.26 | 301.44 | 308.04 | 262.99 | 1,202.73 |
| 722 | 236.63 | 232.39 | 211.7 | 150.68 | 831.4 |
| 723 | 48.8 | 66.06 | 51.47 | 57.63 | 223.96 |
| 724 | 62.63 | 71.81 | 94.7 | 95.9 | 325.04 |
| 725 | 79.66 | 77.92 | 85.02 | 67.31 | 309.91 |
| 726 | 55 | 71.59 | 58.61 | 79.58 | 264.78 |
| 727 | 452.92 | 460.86 | 450.56 | 467.28 | 1,831.62 |
| 728 | 104.42 | 73.46 | 81.23 | 63.79 | 322.9 |
| 729 | 537.89 | 479.33 | 488.83 | 464.38 | 1,970.43 |
| TOTAL | 2,645.93 | 2,560.98 | 2,507.76 | 2,368.40 | 10,083.07 |

STATE: CALIFORNIA

| ZIP CODE | QUARTER 1 | QUARTER 2 | QUARTER 3 | QUARTER 4 | TOTAL GRAMS |
|---|---|---|---|---|---|
| 900 | 626.7 | 639.81 | 622.69 | 627.59 | 2,516.79 |
| 902 | 542.04 | 546.05 | 589.54 | 448.19 | 2,125.82 |
| 903 | 176.9 | 179.85 | 138.9 | 129.35 | 625 |
| 904 | 82.07 | 86.36 | 62.98 | 70.3 | 301.71 |
| 905 | 35.51 | 27.92 | 13.83 | 16.59 | 93.85 |
| 906 | 105.31 | 114.37 | 110.09 | 100.5 | 430.27 |
| 907 | 205.23 | 208.75 | 168.92 | 180.29 | 763.19 |
| 908 | 137.74 | 130.16 | 123.56 | 104.82 | 496.28 |
| 910 | 113.25 | 106.07 | 101.88 | 118.56 | 439.76 |
| 911 | 133.91 | 136.85 | 145.23 | 150.23 | 566.22 |
| 912 | 58.34 | 36.75 | 50.23 | 53.35 | 198.67 |
| 913 | 737.7 | 714.55 | 737.97 | 606.25 | 2,796.47 |
| 914 | 222.94 | 154.96 | 108.84 | 150.14 | 636.88 |
| 915 | 30.87 | 14.01 | 27.3 | 38.98 | 111.16 |
| 916 | 37.83 | 26.23 | 35.59 | 30.47 | 130.12 |
| 917 | 727.55 | 715.69 | 705.47 | 838.54 | 2,987.25 |
| 918 | 16.41 | 19.8 | 1.61 | 15.52 | 53.34 |
| 919 | 274.01 | 250.95 | 271.04 | 249.66 | 1,045.66 |
| 920 | 2,181.14 | 1,972.40 | 1,910.44 | 1,826.02 | 7,890 |
| 921 | 880.01 | 792.01 | 723 | 741.91 | 3,136.93 |
| 922 | 518.22 | 427.05 | 435.26 | 397.07 | 1,777.60 |
| 923 | 450.47 | 491.01 | 449.48 | 383.87 | 1,774.83 |
| 924 | 224.36 | 152.82 | 165.26 | 136.76 | 679.2 |

| | | | | | |
|---|---|---|---|---|---|
| 925 | 390.96 | 315.05 | 346.71 | 340.52 | 1,393.24 |
| 926 | 903.15 | 750.47 | 751.47 | 798.59 | 3,203.68 |
| 927 | 146.77 | 125.71 | 115.53 | 123.56 | 511.57 |
| 928 | 748.65 | 723.98 | 688.14 | 563.64 | 2,724.41 |
| 930 | 386.41 | 360.02 | 405.64 | 461.44 | 1,613.51 |
| 931 | 29.35 | 20.88 | 20.16 | 31.05 | 101.44 |
| 932 | 179.9 | 126.28 | 107.37 | 104.42 | 517.97 |
| 933 | 125.08 | 134.37 | 108.66 | 114.08 | 482.19 |
| 934 | 279.83 | 296 | 217.32 | 230.34 | 1,023.49 |
| 935 | 489.18 | 465.59 | 456.26 | 475.13 | 1,886.16 |
| 936 | 553.19 | 544.85 | 550.51 | 454.3 | 2,102.85 |
| 937 | 662.21 | 575.94 | 574.42 | 563.54 | 2,376.11 |
| 939 | 169.68 | 167.09 | 134.26 | 153.23 | 624.26 |
| 940 | 551.15 | 586.07 | 495.71 | 463.12 | 2,096.05 |
| 941 | 339.63 | 299.22 | 284.58 | 266.93 | 1,190.36 |
| 943 | 37.29 | 20.59 | 28.73 | 23.64 | 110.25 |
| 944 | 109.59 | 151.22 | 114.55 | 83.88 | 459.24 |
| 945 | 2,695.56 | 2,577.55 | 2,361.61 | 2,194.03 | 9,828.75 |
| 946 | 45.81 | 47.28 | 32.2 | 45.54 | 170.83 |
| 947 | 37.47 | 50.34 | 41.58 | 12.07 | 141.46 |
| 948 | 52.63 | 44.52 | 38.98 | 33.77 | 169.9 |
| 949 | 271.72 | 202.31 | 156.28 | 172.88 | 803.19 |
| 950 | 302.53 | 323.9 | 331.4 | 280.18 | 1,238.01 |
| 951 | 106.7 | 130.07 | 100.96 | 115.54 | 453.27 |
| 952 | 220.84 | 200.45 | 191.85 | 176.73 | 789.87 |
| 953 | 784.31 | 736.5 | 665.7 | 594.85 | 2,781.36 |
| 954 | 427.72 | 394.03 | 439.8 | 343.27 | 1,604.82 |

DRUG CODE:9652DRUG NAME:OXYMORPHONE
STATE:CALIFORNIA

| ZIP CODE | QUARTER 1 | QUARTER 2 | QUARTER 3 | QUARTER 4 | TOTAL GRAMS |
|---|---|---|---|---|---|
| 955 | 42.28 | 59.95 | 46.75 | 52.08 | 201.06 |
| 956 | 814.38 | 811.54 | 745.78 | 680.35 | 3,052.05 |
| 957 | 117.94 | 129.98 | 131.76 | 112.23 | 491.91 |
| 958 | 525.02 | 506.75 | 455.35 | 414.46 | 1,901.58 |
| 959 | 535.97 | 472.55 | 462.38 | 449.31 | 1,920.21 |
| 960 | 350.43 | 291.24 | 340.46 | 340.69 | 1,322.82 |
| 961 | 69.32 | 84.57 | 82.88 | 82.52 | 319.29 |
| TOTAL | 22,021.16 | 20,671.28 | 19,724.85 | 18,766.87 | 81,184.16 |

STATE: COLORADO

| ZIP CODE | QUARTER 1 | QUARTER 2 | QUARTER 3 | QUARTER 4 | TOTAL GRAMS |
|---|---|---|---|---|---|
| 800 | 820.06 | 715.33 | 649.18 | 688.92 | 2,873.49 |
| 801 | 813.09 | 868.33 | 885.01 | 712.73 | 3,279.16 |
| 802 | 961.34 | 869.29 | 880.15 | 765.4 | 3,476.18 |
| 803 | 179.89 | 138.65 | 132.66 | 135.45 | 586.65 |
| 804 | 115.26 | 148.8 | 146.39 | 118.16 | 528.61 |
| 805 | 793.84 | 670.84 | 668.37 | 610.51 | 2,743.56 |
| 806 | 218.97 | 191.44 | 190.28 | 188.59 | 789.28 |
| 807 | 49.33 | 56.92 | 59.5 | 69.58 | 235.33 |
| 808 | 164.42 | 176.01 | 167.55 | 144.48 | 652.46 |
| 809 | 638.61 | 574.33 | 510.39 | 506.47 | 2,229.80 |
| 810 | 257.28 | 239.26 | 205.57 | 198.72 | 900.83 |
| 811 | 14.56 | 30.11 | 20.52 | 16.24 | 81.43 |
| 812 | 74.13 | 70.21 | 51.52 | 47.55 | 243.41 |
| 813 | 11.51 | 5 | 5.62 | 4.28 | 26.41 |
| 814 | 7.32 | 10.17 | 6.69 | 3.21 | 27.39 |
| 815 | 93.94 | 73.42 | 81.63 | 79.75 | 328.74 |
| 816 | 36.13 | 26.85 | 25.6 | 29.8 | 118.38 |
| TOTAL | 5,249.68 | 4,864.96 | 4,686.63 | 4,319.84 | 19,121.11 |

RETAIL DRUG DISTRIBUTION BY ZIP CODE WITHIN STATE BY GRAMS WT

STATE: CONNECTICUT

| ZIP CODE | QUARTER 1 | QUARTER 2 | QUARTER 3 | QUARTER 4 | TOTAL GRAMS |
|---|---|---|---|---|---|
| 60 | 659.73 | 548.03 | 483.65 | 490 | 2,181.41 |
| 61 | 127.97 | 94.61 | 93.09 | 110.09 | 425.76 |
| 62 | 98.4 | 84.66 | 78.33 | 81.45 | 342.84 |
| 63 | 229.18 | 272.09 | 209.02 | 297.6 | 1,007.89 |
| 64 | 1,384.69 | 1,361.75 | 1,367.38 | 1,248.63 | 5,362.45 |
| 65 | 505.2 | 510.77 | 478.12 | 458.49 | 1,952.58 |
| 66 | 207.1 | 225.17 | 236.58 | 233.42 | 902.27 |
| 67 | 391.9 | 384 | 388.46 | 318.66 | 1,483.02 |
| 68 | 561.67 | 591.19 | 437.45 | 408.19 | 1,998.50 |
| 69 | 53.08 | 38.54 | 53.53 | 65.57 | 210.72 |
| TOTAL | 4,218.92 | 4,110.81 | 3,825.61 | 3,712.10 | 15,867.44 |

STATE: DELAWARE

| ZIP CODE | QUARTER 1 | QUARTER 2 | QUARTER 3 | QUARTER 4 | TOTAL GRAMS |
|---|---|---|---|---|---|
| 197 | 870.81 | 861.3 | 819.67 | 817.97 | 3,369.75 |
| 198 | 469.74 | 492.97 | 474.86 | 465.23 | 1,902.80 |
| 199 | 918.42 | 869.31 | 928.27 | 857.15 | 3,573.15 |
| TOTAL | 2,258.97 | 2,223.58 | 2,222.80 | 2,140.35 | 8,845.70 |

STATE: DISTRICT OF COLUMBIA

| ZIP CODE | QUARTER 1 | QUARTER 2 | QUARTER 3 | QUARTER 4 | TOTAL GRAMS |
|---|---|---|---|---|---|
| 200 | 152.03 | 135.79 | 171.81 | 139.39 | 599.02 |
| 204 | 0 | 0 | 0 | 0.54 | 0.54 |
| TOTAL | 152.03 | 135.79 | 171.81 | 139.93 | 599.56 |

STATE: FLORIDA

| ZIP CODE | QUARTER 1 | QUARTER 2 | QUARTER 3 | QUARTER 4 | TOTAL GRAMS |
|---|---|---|---|---|---|
| 320 | 1,131.76 | 1,067.40 | 1,030.46 | 1,024.26 | 4,253.88 |
| 321 | 510.5 | 551.01 | 549.89 | 510.64 | 2,122.04 |
| 322 | 2,145.10 | 2,121.19 | 2,235.42 | 2,100.54 | 8,602.25 |
| 323 | 425.71 | 376.21 | 380.12 | 383.02 | 1,565.06 |
| 324 | 554.44 | 657.7 | 608.23 | 543.07 | 2,363.44 |
| 325 | 352.11 | 361.52 | 333.65 | 350.19 | 1,397.47 |
| 326 | 709.31 | 610.6 | 617.11 | 616.53 | 2,553.55 |
| 327 | 1,564.52 | 1,622.46 | 1,634.82 | 1,549.85 | 6,371.65 |
| 328 | 772.42 | 724.39 | 746.24 | 679.87 | 2,922.92 |
| 329 | 2,170.70 | 2,046.97 | 2,020.07 | 2,052.86 | 8,290.60 |
| 330 | 1,509.70 | 1,600.56 | 1,609.13 | 1,730.55 | 6,449.94 |
| 331 | 694.77 | 830.41 | 749.45 | 690.67 | 2,965.30 |

DRUG CODE:9652DRUG NAME:OXYMORPHONE
STATE:FLORIDA

| ZIP CODE | QUARTER 1 | QUARTER 2 | QUARTER 3 | QUARTER 4 | TOTAL GRAMS |
|---|---|---|---|---|---|
| 333 | 1,068.51 | 970.43 | 886.52 | 857.4 | 3,782.86 |
| 334 | 2,200.05 | 2,136.96 | 2,095.86 | 2,207.68 | 8,640.55 |
| 335 | 1,165.17 | 1,193.99 | 1,182.39 | 1,199.34 | 4,740.89 |
| 336 | 1,299.25 | 1,371.38 | 1,299.97 | 1,310.88 | 5,281.48 |
| 337 | 1,843.88 | 1,647.89 | 1,666.22 | 1,629.61 | 6,787.60 |
| 338 | 424.51 | 385.79 | 388.15 | 390.83 | 1,589.28 |
| 339 | 1,239.95 | 1,155.81 | 1,150.20 | 1,204.20 | 4,750.16 |
| 341 | 497.88 | 523.31 | 497.35 | 493.96 | 2,012.50 |
| 342 | 1,942.99 | 1,910.79 | 1,840.67 | 1,763.59 | 7,458.04 |
| 344 | 552.34 | 569.47 | 531.74 | 558.45 | 2,212 |
| 346 | 2,620.28 | 2,752.62 | 2,733.08 | 2,836.92 | 10,942.90 |
| 347 | 879.92 | 832.95 | 677.86 | 710.16 | 3,100.89 |
| 349 | 1,042.15 | 1,077.12 | 1,073.46 | 1,075.25 | 4,267.98 |
| TOTAL | 29,317.92 | 29,098.93 | 28,538.06 | 28,470.32 | 115,425.23 |

RETAIL DRUG DISTRIBUTION BY ZIP CODE WITHIN STATE BY GRAMS WT

STATE: GEORGIA

| ZIP CODE | QUARTER 1 | QUARTER 2 | QUARTER 3 | QUARTER 4 | TOTAL GRAMS |
|---|---|---|---|---|---|
| 300 | 1,456.92 | 1,455.36 | 1,505.40 | 1,335.40 | 5,753.08 |
| 301 | 1,526.52 | 1,507.38 | 1,491.24 | 1,429.01 | 5,954.15 |
| 302 | 801.94 | 704.18 | 705.12 | 678.11 | 2,889.35 |
| 303 | 376.3 | 366.79 | 342.42 | 359.87 | 1,445.38 |
| 304 | 153.13 | 142.11 | 180.47 | 170.03 | 645.74 |
| 305 | 611.67 | 588.38 | 565.19 | 575.67 | 2,340.91 |
| 306 | 444.58 | 428.65 | 395.6 | 363.95 | 1,632.78 |
| 307 | 449.8 | 402.83 | 404.3 | 370.22 | 1,627.15 |
| 308 | 53.79 | 74.31 | 99.51 | 101.92 | 329.53 |
| 309 | 132.48 | 105.18 | 136 | 118.65 | 492.31 |
| 310 | 563.99 | 559.03 | 578.62 | 535.93 | 2,237.57 |
| 312 | 236.32 | 218.56 | 192.96 | 171.37 | 819.21 |
| 313 | 409.47 | 385.61 | 360.5 | 388.15 | 1,543.73 |
| 314 | 291.9 | 332.35 | 335.43 | 354.12 | 1,313.80 |
| 315 | 271.06 | 272.45 | 268.43 | 256.79 | 1,068.73 |
| 316 | 647.93 | 620.72 | 591.73 | 578.57 | 2,438.95 |
| 317 | 231.86 | 227.84 | 205.99 | 239.8 | 905.49 |
| 318 | 14.99 | 13.47 | 18.29 | 16.77 | 63.52 |
| 319 | 262.9 | 296.44 | 279.41 | 251.39 | 1,090.14 |
| 398 | 54.24 | 49.42 | 46.52 | 68.91 | 219.09 |
| TOTAL | 8,991.79 | 8,751.06 | 8,703.13 | 8,364.63 | 34,810.61 |

STATE: HAWAII

| ZIP CODE | QUARTER 1 | QUARTER 2 | QUARTER 3 | QUARTER 4 | TOTAL GRAMS |
|---|---|---|---|---|---|
| 967 | 500.11 | 553.8 | 486.34 | 466.38 | 2,006.63 |
| 968 | 221.95 | 210.62 | 215.53 | 220.97 | 869.07 |
| TOTAL | 722.06 | 764.42 | 701.87 | 687.35 | 2,875.70 |

STATE: IDAHO

| ZIP CODE | QUARTER 1 | QUARTER 2 | QUARTER 3 | QUARTER 4 | TOTAL GRAMS |
|---|---|---|---|---|---|
| 832 | 124.58 | 113.34 | 97.68 | 90.01 | 425.61 |
| 833 | 182.61 | 211.43 | 204.51 | 224.01 | 822.56 |
| 834 | 85.82 | 66.28 | 72.44 | 74.58 | 299.12 |
| 835 | 21.95 | 19.63 | 24.09 | 20.7 | 86.37 |
| 836 | 691.78 | 751.91 | 647.31 | 608.64 | 2,699.64 |
| 837 | 498.62 | 471.38 | 513.05 | 442.84 | 1,925.89 |
| 838 | 151.05 | 137.45 | 134.89 | 102.36 | 525.75 |
| TOTAL | 1,756.41 | 1,771.42 | 1,693.97 | 1,563.14 | 6,784.94 |

STATE: ILLINOIS

| ZIP CODE | QUARTER 1 | QUARTER 2 | QUARTER 3 | QUARTER 4 | TOTAL GRAMS |
|---|---|---|---|---|---|
| 600 | 1,592.68 | 1,597.31 | 1,613.23 | 1,440.92 | 6,244.14 |
| 601 | 622.28 | 548.11 | 595.92 | 624.92 | 2,391.23 |
| 602 | 60.22 | 69.94 | 66.95 | 53.79 | 250.9 |
| 603 | 35.06 | 33.1 | 25.87 | 34.17 | 128.2 |
| 604 | 886.75 | 918.33 | 843.75 | 833.54 | 3,482.37 |
| 605 | 655.78 | 620.9 | 631.61 | 641.33 | 2,549.62 |
| 606 | 622.42 | 636.96 | 557.83 | 532.27 | 2,349.48 |
| 607 | 40.23 | 21.77 | 25.51 | 16.41 | 103.92 |
| 608 | 21.59 | 15.66 | 23.55 | 31.22 | 92.02 |
| 609 | 83.14 | 85.91 | 71.81 | 63.61 | 304.47 |
| 610 | 100.41 | 68.62 | 85.41 | 69.91 | 324.35 |
| 611 | 135.6 | 118.02 | 120.43 | 122.22 | 496.27 |

DRUG CODE:9652DRUG NAME:OXYMORPHONE
STATE:ILLINOIS

| ZIP CODE | QUARTER 1 | QUARTER 2 | QUARTER 3 | QUARTER 4 | TOTAL GRAMS |
|---|---|---|---|---|---|

| | | | | | |
|---|---|---|---|---|---|
| 612 | 54.2 | 77.08 | 59.28 | 71.19 | 261.75 |
| 613 | 60.84 | 69.67 | 50.05 | 67.35 | 247.91 |
| 614 | 24.39 | 29.74 | 12.91 | 20.4 | 87.44 |
| 615 | 17.97 | 13.61 | 11.91 | 15.92 | 59.41 |
| 616 | 49.65 | 61.08 | 54.19 | 47.94 | 212.86 |
| 617 | 141.75 | 141.04 | 117.94 | 95.81 | 496.54 |
| 618 | 315.58 | 314.82 | 335.07 | 286.63 | 1,252.10 |
| 619 | 48.17 | 53.08 | 66.91 | 58.34 | 226.5 |
| 620 | 186.49 | 172.44 | 190.42 | 189.39 | 738.74 |
| 622 | 124.14 | 134.89 | 158.21 | 161.51 | 578.75 |
| 623 | 21.32 | 20.61 | 28.59 | 31.76 | 102.28 |
| 624 | 23.15 | 18.69 | 21.9 | 43.09 | 106.83 |
| 625 | 83.59 | 80.74 | 85.2 | 70.57 | 320.1 |
| 626 | 57.63 | 56.83 | 53.97 | 55.62 | 224.05 |
| 627 | 11.06 | 7.94 | 12.85 | 14.01 | 45.86 |
| 628 | 37.56 | 39.34 | 28.73 | 13.65 | 119.28 |
| 629 | 122.62 | 127.26 | 118.6 | 102.9 | 471.38 |
| TOTAL | 6,236.27 | 6,153.49 | 6,068.60 | 5,810.39 | 24,268.75 |

STATE: INDIANA

| ZIP CODE | QUARTER 1 | QUARTER 2 | QUARTER 3 | QUARTER 4 | TOTAL GRAMS |
|---|---|---|---|---|---|
| 460 | 511.12 | 467.5 | 525.13 | 436.45 | 1,940.20 |
| 461 | 799.63 | 820.2 | 770.86 | 723.72 | 3,114.41 |
| 462 | 1,067.25 | 1,069.22 | 991.63 | 1,296.47 | 4,424.57 |
| 463 | 718.58 | 657.75 | 641.11 | 573.84 | 2,591.28 |
| 464 | 12.67 | 7.05 | 3.57 | 4.82 | 28.11 |
| 465 | 646.64 | 639.1 | 610.73 | 564.65 | 2,461.12 |
| 466 | 99.38 | 93.27 | 105.85 | 101.7 | 400.2 |
| 467 | 309.29 | 230.38 | 246.13 | 216.16 | 1,001.96 |
| 468 | 360.27 | 317.5 | 321.29 | 267.81 | 1,266.87 |
| 469 | 411.48 | 359.56 | 357.91 | 342.7 | 1,471.65 |
| 470 | 152.01 | 138.72 | 159.33 | 149.78 | 599.84 |
| 471 | 941.83 | 794.28 | 679.02 | 548.42 | 2,963.55 |
| 472 | 265.04 | 238.99 | 195.82 | 196.35 | 896.2 |
| 473 | 339.67 | 321.29 | 321.56 | 314.64 | 1,297.16 |
| 474 | 315.89 | 266.83 | 298.76 | 268.61 | 1,150.09 |
| 475 | 178.33 | 151.03 | 161.11 | 177.71 | 668.18 |
| 476 | 140.42 | 154.02 | 134.57 | 129.89 | 558.9 |
| 477 | 570.9 | 623.22 | 528.48 | 476.56 | 2,199.16 |
| 478 | 447.03 | 466.75 | 397.88 | 458 | 1,769.66 |
| 479 | 181.11 | 198.65 | 190.53 | 203.22 | 773.51 |
| TOTAL | 8,468.54 | 8,015.31 | 7,641.27 | 7,451.50 | 31,576.62 |

STATE: IOWA

| ZIP CODE | QUARTER 1 | QUARTER 2 | QUARTER 3 | QUARTER 4 | TOTAL GRAMS |
|---|---|---|---|---|---|
| 500 | 164.37 | 157.63 | 146.41 | 142.26 | 610.67 |
| 501 | 126.05 | 134.97 | 141.13 | 107.01 | 509.16 |
| 502 | 214.68 | 218.12 | 208.44 | 205.23 | 846.47 |
| 503 | 681.43 | 551.14 | 504.84 | 487.49 | 2,224.90 |
| 504 | 75.96 | 57.45 | 50.85 | 44.34 | 228.6 |
| 505 | 17.93 | 11.78 | 20.61 | 22.48 | 72.8 |
| 506 | 19.27 | 23.19 | 16.06 | 27.57 | 86.09 |
| 507 | 28.81 | 21.32 | 35.91 | 19.8 | 105.84 |
| 508 | 4.64 | 5.17 | 9.19 | 11.42 | 30.42 |
| 510 | 17.13 | 17.93 | 19.18 | 21.68 | 75.92 |
| 511 | 19.8 | 17.57 | 15.43 | 17.75 | 70.55 |
| 512 | 0.8 | 1.34 | 1.61 | 1.07 | 4.82 |
| 513 | 0 | 0 | 2.05 | 1.87 | 3.92 |
| 514 | 6.42 | 4.55 | 9.37 | 6.82 | 27.16 |
| 515 | 110.53 | 116.6 | 119.32 | 129.31 | 475.76 |
| 516 | 5.35 | 3.48 | 5.08 | 5.62 | 19.53 |
| 520 | 28.64 | 21.68 | 38 | 18.64 | 106.96 |

| | | | | | |
|---|---|---|---|---|---|
| 522 | 21.81 | 19.09 | 28.95 | 11.73 | 81.58 |
| 523 | 38.9 | 38.9 | 33.99 | 32.2 | 143.99 |
| 524 | 43.27 | 29.44 | 19.27 | 19.18 | 111.16 |
| 525 | 97.15 | 61.38 | 50.14 | 59.28 | 267.95 |
| 526 | 39.34 | 24.62 | 20.83 | 19.72 | 104.51 |
| 527 | 53.95 | 31.85 | 34.93 | 31.76 | 152.49 |

DRUG CODE:9652DRUG NAME:OXYMORPHONE
STATE:IOWA

| ZIP CODE | QUARTER 1 | QUARTER 2 | QUARTER 3 | QUARTER 4 | TOTAL GRAMS |
|---|---|---|---|---|---|
| 528 | 63.65 | 58.3 | 65.08 | 60.57 | 247.6 |
| TOTAL | 1,879.88 | 1,627.50 | 1,596.67 | 1,504.80 | 6,608.85 |

STATE: KANSAS

| ZIP CODE | QUARTER 1 | QUARTER 2 | QUARTER 3 | QUARTER 4 | TOTAL GRAMS |
|---|---|---|---|---|---|
| 660 | 796.11 | 823.32 | 767.43 | 748.07 | 3,134.93 |
| 661 | 221.95 | 210.89 | 145.01 | 139.52 | 717.37 |
| 662 | 818.9 | 755.65 | 698.78 | 627.99 | 2,901.32 |
| 664 | 53.88 | 35.86 | 46.39 | 25.96 | 162.09 |
| 665 | 19.09 | 29.35 | 36.75 | 27.08 | 112.27 |
| 666 | 165.22 | 180.29 | 176.19 | 164.15 | 685.85 |
| 667 | 96.44 | 64.01 | 58.08 | 58.16 | 276.69 |
| 668 | 52.86 | 31 | 23.73 | 21.59 | 129.18 |
| 669 | 16.24 | 20.43 | 14.63 | 13.38 | 64.68 |
| 670 | 172.26 | 169.86 | 166.73 | 173.91 | 682.76 |
| 671 | 68.33 | 58.16 | 54.73 | 50.54 | 231.76 |
| 672 | 451.05 | 472.9 | 424.68 | 410.1 | 1,758.73 |
| 673 | 34.12 | 26.18 | 34.39 | 45.05 | 139.74 |
| 674 | 33.63 | 51.21 | 54.86 | 47.37 | 187.07 |
| 675 | 147.64 | 152.73 | 141.67 | 104.91 | 546.95 |
| 676 | 50.05 | 37.74 | 31.85 | 33.99 | 153.63 |
| 677 | 9.01 | 14.45 | 11.51 | 9.72 | 44.69 |
| 678 | 55.13 | 59.95 | 62.54 | 53.62 | 231.24 |
| 679 | 15.79 | 21.41 | 17.66 | 13.65 | 68.51 |
| TOTAL | 3,277.70 | 3,215.39 | 2,967.61 | 2,768.76 | 12,229.46 |

STATE: KENTUCKY

| ZIP CODE | QUARTER 1 | QUARTER 2 | QUARTER 3 | QUARTER 4 | TOTAL GRAMS |
|---|---|---|---|---|---|
| 400 | 100.81 | 84.57 | 108.84 | 125.07 | 419.29 |
| 401 | 100 | 103.66 | 93.05 | 108.03 | 404.74 |
| 402 | 587.72 | 574.82 | 615.19 | 569.2 | 2,346.93 |
| 403 | 462.24 | 467.51 | 390.25 | 387.13 | 1,707.13 |
| 404 | 376.51 | 362.73 | 320.93 | 264.73 | 1,324.90 |
| 405 | 264.78 | 255.72 | 252.29 | 261.43 | 1,034.22 |
| 406 | 38.45 | 28.5 | 26.85 | 24.18 | 117.98 |
| 407 | 389.36 | 462.29 | 250.86 | 294.57 | 1,397.08 |
| 408 | 37.65 | 45.72 | 40.06 | 38.23 | 161.66 |
| 409 | 191.31 | 206.52 | 178.42 | 174.49 | 750.74 |
| 410 | 641.6 | 609.79 | 637.99 | 611.04 | 2,500.42 |
| 411 | 276.42 | 276.15 | 240.38 | 173.11 | 966.06 |
| 412 | 24.58 | 25.69 | 9.23 | 5.93 | 65.43 |
| 413 | 27.39 | 33.45 | 31.8 | 26.9 | 119.54 |
| 414 | 10.13 | 9.37 | 12.58 | 4.82 | 36.9 |
| 415 | 252.29 | 244.79 | 291.05 | 261.16 | 1,049.29 |
| 416 | 89.79 | 82.39 | 75.11 | 72.44 | 319.73 |
| 417 | 84.08 | 81.31 | 78.59 | 62.8 | 306.78 |
| 418 | 21.95 | 23.37 | 9.46 | 3.39 | 58.17 |
| 420 | 260.23 | 227.57 | 232.04 | 220.35 | 940.19 |
| 421 | 261.83 | 248.72 | 241.22 | 246.09 | 997.86 |
| 422 | 211.92 | 192.78 | 191.67 | 204.02 | 800.39 |

| | | | | | |
|---|---|---|---|---|---|
| 423 | 174.05 | 172.89 | 169.28 | 126.32 | 642.54 |
| 424 | 129.18 | 123.11 | 123.47 | 123.56 | 499.32 |
| 425 | 87.25 | 80.69 | 70.57 | 93.89 | 332.4 |
| 426 | 43.8 | 42.2 | 40.86 | 45.85 | 172.71 |
| 427 | 232.12 | 242.47 | 248.27 | 224.1 | 946.96 |
| TOTAL | 5,377.44 | 5,308.78 | 4,980.31 | 4,752.83 | 20,419.36 |

STATE: LOUISIANA

| ZIP CODE | QUARTER 1 | QUARTER 2 | QUARTER 3 | QUARTER 4 | TOTAL GRAMS |
|---|---|---|---|---|---|
| 700 | 1,392.30 | 1,159.60 | 1,308.80 | 1,233.37 | 5,094.07 |
| 701 | 197.24 | 180.03 | 180.34 | 176.64 | 734.25 |
| 703 | 374.82 | 380.61 | 397.61 | 351.49 | 1,504.53 |
| 704 | 1,665.06 | 1,763.24 | 1,632.28 | 1,447.08 | 6,507.66 |
| 705 | 647.4 | 627.64 | 662.74 | 669.48 | 2,607.26 |
| 706 | 113.56 | 101.25 | 80.11 | 68.33 | 363.25 |
| 707 | 2,433.51 | 2,288.64 | 2,204.91 | 2,009.72 | 8,936.78 |
| 708 | 1,132.43 | 1,145.23 | 1,344.22 | 1,225.66 | 4,847.54 |
| 710 | 257.86 | 263.62 | 268.7 | 243.23 | 1,033.41 |
| 711 | 888.53 | 880.55 | 818.01 | 885.41 | 3,472.50 |

DRUG CODE:9652DRUG NAME:OXYMORPHONE
STATE:LOUISIANA

| ZIP CODE | QUARTER 1 | QUARTER 2 | QUARTER 3 | QUARTER 4 | TOTAL GRAMS |
|---|---|---|---|---|---|
| 712 | 63.43 | 69.85 | 71.01 | 72.26 | 276.55 |
| 713 | 241.13 | 234.89 | 221.51 | 175.21 | 872.74 |
| 714 | 127.53 | 106.38 | 128.28 | 97.11 | 459.3 |
| TOTAL | 9,534.80 | 9,201.53 | 9,318.52 | 8,654.99 | 36,709.84 |

STATE: MAINE

| ZIP CODE | QUARTER 1 | QUARTER 2 | QUARTER 3 | QUARTER 4 | TOTAL GRAMS |
|---|---|---|---|---|---|
| 39 | 33.63 | 10.71 | 17.84 | 8.03 | 70.21 |
| 40 | 352.56 | 329.85 | 295.11 | 286.68 | 1,264.20 |
| 41 | 43.46 | 36.67 | 29.01 | 28.14 | 137.28 |
| 42 | 342.34 | 365.76 | 329.41 | 281.1 | 1,318.61 |
| 43 | 457.11 | 436.33 | 425.04 | 326.6 | 1,645.08 |
| 44 | 133.37 | 112.72 | 90.06 | 62.4 | 398.55 |
| 45 | 73.82 | 77.3 | 74.67 | 79.58 | 305.37 |
| 46 | 8.65 | 11.6 | 8.12 | 2.54 | 30.91 |
| 47 | 44.61 | 61.91 | 27.03 | 28.28 | 161.83 |
| 48 | 40.32 | 36.75 | 15.17 | 17.89 | 110.13 |
| 49 | 416.57 | 397.03 | 319.73 | 297.34 | 1,430.67 |
| TOTAL | 1,946.44 | 1,876.63 | 1,631.19 | 1,418.58 | 6,872.84 |

STATE: MARYLAND

| ZIP CODE | QUARTER 1 | QUARTER 2 | QUARTER 3 | QUARTER 4 | TOTAL GRAMS |
|---|---|---|---|---|---|
| 206 | 1,071.99 | 1,064.77 | 1,081.18 | 1,230.70 | 4,448.64 |
| 207 | 688.92 | 723.18 | 760.84 | 759.44 | 2,932.38 |
| 208 | 829.61 | 841.07 | 888.8 | 791.87 | 3,351.35 |
| 209 | 113.7 | 112.67 | 152.82 | 146.71 | 525.9 |
| 210 | 2,136.27 | 2,320.04 | 2,340.58 | 2,240.51 | 9,037.40 |
| 211 | 821.8 | 823.23 | 931.44 | 957.85 | 3,534.32 |
| 212 | 1,311.43 | 1,343.77 | 1,333.69 | 1,241.37 | 5,230.26 |
| 214 | 224.76 | 229.05 | 260.67 | 260.41 | 974.89 |
| 215 | 360.5 | 396.27 | 432.36 | 426.25 | 1,615.38 |
| 216 | 168.7 | 158.75 | 242.65 | 289.49 | 859.59 |
| 217 | 1,820.73 | 1,845.93 | 1,998.88 | 2,130.69 | 7,796.23 |
| 218 | 200.45 | 213.12 | 227.98 | 222.76 | 864.31 |
| 219 | 344.26 | 314.33 | 306.79 | 323.21 | 1,288.59 |
| TOTAL | 10,093.12 | 10,386.18 | 10,958.68 | 11,021.26 | 42,459.24 |

RETAIL DRUG DISTRIBUTION BY ZIP CODE WITHIN STATE BY GRAMS WT

STATE: MASSACHUSETTS

| ZIP CODE | QUARTER 1 | QUARTER 2 | QUARTER 3 | QUARTER 4 | TOTAL GRAMS |
|---|---|---|---|---|---|
| 10 | 138.19 | 128.37 | 130.43 | 127.66 | 524.65 |
| 11 | 18.02 | 17.75 | 23.46 | 13.92 | 73.15 |
| 12 | 32.12 | 23.82 | 24.53 | 33.19 | 113.66 |
| 13 | 41.17 | 33.68 | 32.43 | 19.4 | 126.68 |
| 14 | 30.33 | 52.19 | 28.64 | 23.82 | 134.98 |
| 15 | 130.8 | 161.94 | 141.94 | 150.52 | 585.2 |
| 16 | 73.6 | 55.96 | 36.49 | 37 | 203.05 |
| 17 | 238.25 | 181.28 | 87.21 | 88.17 | 594.91 |
| 18 | 1,253.98 | 1,266.75 | 1,167.99 | 1,072.17 | 4,760.89 |
| 19 | 83.01 | 105.62 | 76.23 | 107.85 | 372.71 |
| 20 | 89.42 | 82.6 | 81.52 | 64.85 | 318.39 |
| 21 | 148.29 | 125.8 | 125.16 | 130.9 | 530.15 |
| 22 | 86.58 | 79.66 | 76.63 | 67.26 | 310.13 |
| 23 | 96.61 | 83.14 | 80.16 | 71.5 | 331.41 |
| 24 | 121 | 113.5 | 124.8 | 134.8 | 494.1 |
| 25 | 29.82 | 20.34 | 10.38 | 7.4 | 67.94 |
| 26 | 46.61 | 46.66 | 45.9 | 41.75 | 180.92 |
| 27 | 583.92 | 558.75 | 521.67 | 565.94 | 2,230.28 |
| TOTAL | 3,241.72 | 3,137.81 | 2,815.57 | 2,758.10 | 11,953.20 |

STATE: MICHIGAN

| ZIP CODE | QUARTER 1 | QUARTER 2 | QUARTER 3 | QUARTER 4 | TOTAL GRAMS |
|---|---|---|---|---|---|
| 480 | 4,121.50 | 3,769.75 | 4,145.28 | 4,198.58 | 16,235.11 |
| 481 | 3,421.20 | 3,437.66 | 3,338.24 | 3,343.77 | 13,540.87 |
| 482 | 7,403.36 | 7,253.44 | 7,601.23 | 7,740.66 | 29,998.69 |
| 483 | 2,053.44 | 1,908.51 | 2,189.75 | 2,344.08 | 8,495.78 |
| 484 | 223.47 | 212.59 | 240.38 | 279.76 | 956.2 |
| 485 | 399.13 | 377.18 | 351.04 | 359.78 | 1,487.13 |
| 486 | 224.23 | 251.93 | 265.13 | 238.9 | 980.19 |
| 487 | 227.4 | 217.4 | 213.3 | 184.13 | 842.23 |
| 488 | 304.2 | 226.9 | 245.73 | 268.84 | 1,045.67 |

DRUG CODE:9652DRUG NAME:OXYMORPHONE
STATE:MICHIGAN

| ZIP CODE | QUARTER 1 | QUARTER 2 | QUARTER 3 | QUARTER 4 | TOTAL GRAMS |
|---|---|---|---|---|---|
| 489 | 177.08 | 177.97 | 130.69 | 144.79 | 630.53 |
| 490 | 477.59 | 536.2 | 474.73 | 454.08 | 1,942.60 |
| 491 | 76.01 | 52.01 | 45.23 | 38.9 | 212.15 |
| 492 | 190.62 | 200.14 | 218.14 | 171.24 | 780.14 |
| 493 | 92.87 | 77.52 | 75.92 | 57.01 | 303.32 |
| 494 | 243.77 | 332.84 | 303.09 | 255.19 | 1,134.89 |
| 495 | 176.46 | 157.19 | 141.4 | 165.84 | 640.89 |
| 496 | 247.87 | 219.1 | 268.52 | 199.56 | 935.05 |
| 497 | 272.36 | 281.64 | 301.89 | 312.5 | 1,168.39 |
| 498 | 30.42 | 30.24 | 32.74 | 33.72 | 127.12 |
| 499 | 4.01 | 2.41 | 1.87 | 2.14 | 10.43 |
| TOTAL | 20,366.99 | 19,722.62 | 20,584.30 | 20,793.47 | 81,467.38 |

STATE: MINNESOTA

| ZIP CODE | QUARTER 1 | QUARTER 2 | QUARTER 3 | QUARTER 4 | TOTAL GRAMS |
|---|---|---|---|---|---|
| 550 | 81.36 | 60.86 | 99.38 | 78.09 | 319.69 |
| 551 | 138.05 | 143.4 | 116.46 | 67.03 | 464.94 |
| 553 | 136.41 | 114.1 | 110.09 | 108.75 | 469.35 |
| 554 | 102.78 | 104.2 | 68.87 | 60.49 | 336.34 |
| 557 | 28.99 | 22.75 | 33.01 | 25.07 | 109.82 |
| 558 | 16.59 | 19 | 20.07 | 17.13 | 72.79 |
| 559 | 12.05 | 5.37 | 6.68 | 13.26 | 37.36 |

| | | | | | |
|---|---|---|---|---|---|
| 560 | 25.25 | 37.91 | 29.97 | 23.46 | 116.59 |
| 561 | 0 | 0 | 0 | 1.07 | 1.07 |
| 562 | 4.82 | 11.6 | 15.92 | 25.69 | 58.03 |
| 563 | 68.33 | 63.7 | 79.22 | 55.32 | 266.57 |
| 564 | 19 | 21.41 | 29.97 | 18.56 | 88.94 |
| 565 | 10.88 | 22.48 | 16.5 | 10.71 | 60.57 |
| 566 | 3.3 | 4.1 | 1.78 | 1.96 | 11.14 |
| 567 | 12.76 | 11.6 | 11.6 | 11.78 | 47.74 |
| TOTAL | 660.57 | 642.48 | 639.52 | 518.37 | 2,460.94 |

STATE: MISSISSIPPI

| ZIP CODE | QUARTER 1 | QUARTER 2 | QUARTER 3 | QUARTER 4 | TOTAL GRAMS |
|---|---|---|---|---|---|
| 386 | 218.03 | 222.71 | 261.7 | 246.67 | 949.11 |
| 387 | 32.34 | 26.36 | 30.2 | 27.79 | 116.69 |
| 388 | 596.28 | 537.9 | 478.94 | 477.87 | 2,090.99 |
| 389 | 54.06 | 57.72 | 66.6 | 63.7 | 242.08 |
| 390 | 300.55 | 311.21 | 313.57 | 275.79 | 1,201.12 |
| 391 | 167.85 | 207.68 | 214.59 | 173.91 | 764.03 |
| 392 | 248.18 | 220.08 | 229.94 | 229.89 | 928.09 |
| 393 | 227.4 | 235.65 | 220.62 | 181.77 | 865.44 |
| 394 | 574.29 | 519.02 | 471.88 | 465.1 | 2,030.29 |
| 395 | 2,647.84 | 2,420.45 | 2,301.26 | 1,992.01 | 9,361.56 |
| 396 | 165.8 | 142.47 | 145.5 | 150.5 | 604.27 |
| 397 | 75.29 | 60.17 | 62.67 | 73.78 | 271.91 |
| TOTAL | 5,307.91 | 4,961.42 | 4,797.47 | 4,358.78 | 19,425.58 |

STATE: MISSOURI

| ZIP CODE | QUARTER 1 | QUARTER 2 | QUARTER 3 | QUARTER 4 | TOTAL GRAMS |
|---|---|---|---|---|---|
| 630 | 694.29 | 664.27 | 723.56 | 681.7 | 2,763.82 |
| 631 | 1,520.72 | 1,259.42 | 1,061.96 | 1,294.93 | 5,137.03 |
| 633 | 586.29 | 612.83 | 500.16 | 481.12 | 2,180.40 |
| 634 | 85.46 | 82.39 | 68.25 | 66.1 | 302.2 |
| 635 | 3.21 | 3.66 | 15.52 | 8.92 | 31.31 |
| 636 | 246.58 | 235.07 | 222.76 | 212.32 | 916.73 |
| 637 | 149.92 | 117 | 131.54 | 120.7 | 519.16 |
| 638 | 47.19 | 36.4 | 39.07 | 44.16 | 166.82 |
| 639 | 21.41 | 13.74 | 18.07 | 11.42 | 64.64 |
| 640 | 614.12 | 635.35 | 621.88 | 590.39 | 2,461.74 |
| 641 | 524.15 | 541.91 | 483.21 | 458.85 | 2,008.12 |
| 644 | 60.84 | 32.83 | 38 | 33.1 | 164.77 |
| 645 | 310.14 | 250.41 | 233.28 | 240.33 | 1,034.16 |
| 646 | 85.11 | 92.87 | 79.84 | 90.9 | 348.72 |
| 647 | 137.65 | 141.93 | 111.24 | 122.93 | 513.75 |
| 648 | 122.31 | 135.69 | 122.84 | 126.68 | 507.52 |
| 650 | 75.2 | 79.13 | 68.16 | 80.02 | 302.51 |
| 651 | 105.98 | 116.33 | 103.17 | 98.58 | 424.06 |
| 652 | 88.63 | 111.56 | 114.63 | 104.78 | 419.6 |

DRUG CODE:9652DRUG NAME:OXYMORPHONE
STATE:MISSOURI

| ZIP CODE | QUARTER 1 | QUARTER 2 | QUARTER 3 | QUARTER 4 | TOTAL GRAMS |
|---|---|---|---|---|---|
| 653 | 58.7 | 43.18 | 54.06 | 30.55 | 186.49 |
| 654 | 50.05 | 61.91 | 62.71 | 85.2 | 259.87 |
| 655 | 96.17 | 95.32 | 82.56 | 88.54 | 362.59 |
| 656 | 138.9 | 147.46 | 139.35 | 109.82 | 535.53 |
| 657 | 438.65 | 389.67 | 311.61 | 325.88 | 1,465.81 |
| 658 | 324.91 | 304.12 | 290.16 | 315.13 | 1,234.32 |
| TOTAL | 6,586.58 | 6,204.45 | 5,697.59 | 5,823.05 | 24,311.67 |

RETAIL DRUG DISTRIBUTION BY ZIP CODE WITHIN STATE BY GRAMS WT

| STATE: | MONTANA | | | | |
|---|---|---|---|---|---|
| ZIP CODE | QUARTER 1 | QUARTER 2 | QUARTER 3 | QUARTER 4 | TOTAL GRAMS |
| 590 | 13.83 | 5.8 | 13.56 | 6.07 | 39.26 |
| 591 | 93.05 | 76.65 | 71.28 | 66.1 | 307.08 |
| 592 | 6.24 | 6.24 | 8.92 | 4.46 | 25.86 |
| 593 | 3.84 | 1.34 | 0 | 2.14 | 7.32 |
| 594 | 109.28 | 143.63 | 132.03 | 108.39 | 493.33 |
| 595 | 1.96 | 2.5 | 2.14 | 1.07 | 7.67 |
| 596 | 12.13 | 47.99 | 37.47 | 24.09 | 121.68 |
| 597 | 47.64 | 44.34 | 42.65 | 34.17 | 168.8 |
| 598 | 87.87 | 64.68 | 50.94 | 46.57 | 250.06 |
| 599 | 143.45 | 149.96 | 130.65 | 124.89 | 548.95 |
| TOTAL | 519.29 | 543.13 | 489.64 | 417.95 | 1,970.01 |
| STATE: | NEBRASKA | | | | |
| ZIP CODE | QUARTER 1 | QUARTER 2 | QUARTER 3 | QUARTER 4 | TOTAL GRAMS |
| 680 | 120.7 | 110.98 | 103.62 | 99.51 | 434.81 |
| 681 | 740.13 | 830.23 | 803.92 | 780.94 | 3,155.22 |
| 683 | 17.31 | 12.67 | 9.9 | 8.56 | 48.44 |
| 684 | 4.28 | 2.41 | 9.63 | 7.49 | 23.81 |
| 685 | 287.97 | 301.35 | 294.71 | 274.5 | 1,158.53 |
| 686 | 61.11 | 62 | 80.07 | 69.14 | 272.32 |
| 687 | 149.34 | 122.22 | 157.19 | 128.06 | 556.81 |
| 688 | 42.2 | 48.98 | 52.9 | 58.7 | 202.78 |
| 689 | 29.44 | 36.4 | 35.33 | 35.01 | 136.18 |
| 690 | 18.38 | 17.93 | 18.2 | 18.2 | 72.71 |
| 691 | 34.44 | 45.68 | 36.31 | 32.65 | 149.08 |
| 692 | 0 | 0 | 0.54 | 0.54 | 1.08 |
| 693 | 79.49 | 71.59 | 64.54 | 72.97 | 288.59 |
| TOTAL | 1,584.79 | 1,662.44 | 1,666.86 | 1,586.27 | 6,500.36 |
| STATE: | NEVADA | | | | |
| ZIP CODE | QUARTER 1 | QUARTER 2 | QUARTER 3 | QUARTER 4 | TOTAL GRAMS |
| 890 | 1,736.65 | 1,655.65 | 1,531.02 | 1,411.93 | 6,335.25 |
| 891 | 4,086.11 | 3,935.94 | 3,707.74 | 3,638.66 | 15,368.45 |
| 893 | 0.89 | 0 | 0 | 2.27 | 3.16 |
| 894 | 353.63 | 346.67 | 308.27 | 281.23 | 1,289.80 |
| 895 | 412.82 | 349.61 | 358.8 | 331.19 | 1,452.42 |
| 897 | 151.84 | 193.32 | 160.13 | 153 | 658.29 |
| 898 | 13.96 | 7.32 | 7.49 | 6.69 | 35.46 |
| TOTAL | 6,755.90 | 6,488.51 | 6,073.45 | 5,824.97 | 25,142.83 |
| STATE: | NEW HAMPSHIRE | | | | |
| ZIP CODE | QUARTER 1 | QUARTER 2 | QUARTER 3 | QUARTER 4 | TOTAL GRAMS |
| 30 | 246.8 | 261.74 | 248.63 | 248.67 | 1,005.84 |
| 31 | 164.68 | 137.93 | 123.12 | 121.78 | 547.51 |
| 32 | 149.29 | 141.84 | 133.95 | 132.25 | 557.33 |
| 33 | 137.88 | 159.46 | 131.28 | 186.32 | 614.94 |
| 34 | 23.28 | 15.79 | 17.04 | 14.99 | 71.1 |
| 35 | 12.85 | 22.57 | 25.25 | 23.37 | 84.04 |
| 37 | 28.55 | 35.15 | 27.03 | 26.85 | 117.58 |
| 38 | 609.37 | 586.38 | 478.58 | 449 | 2,123.33 |
| TOTAL | 1,372.70 | 1,360.86 | 1,184.88 | 1,203.23 | 5,121.67 |
| STATE: | NEW JERSEY | | | | |
| ZIP CODE | QUARTER 1 | QUARTER 2 | QUARTER 3 | QUARTER 4 | TOTAL GRAMS |
| 70 | 1,390.92 | 1,500.16 | 1,370.18 | 1,232.70 | 5,493.96 |
| 71 | 119.32 | 118.6 | 115.88 | 118.34 | 472.14 |
| 72 | 28.9 | 49.6 | 42.64 | 57.01 | 178.15 |
| 73 | 70.65 | 55.31 | 56.11 | 76.72 | 258.79 |

| | | | | | |
|---|---|---|---|---|---|
| 74 | 455.63 | 387.83 | 416.46 | 385.57 | 1,645.49 |
| 75 | 80.29 | 74.76 | 87.34 | 70.83 | 313.22 |
| 76 | 509.61 | 529.91 | 492.23 | 506.04 | 2,037.79 |

DRUG CODE:9652DRUG NAME:OXYMORPHONE
STATE:NEW JERSEY

| ZIP CODE | QUARTER 1 | QUARTER 2 | QUARTER 3 | QUARTER 4 | TOTAL GRAMS |
|---|---|---|---|---|---|
| 77 | 784.47 | 785.18 | 827.69 | 732.28 | 3,129.62 |
| 78 | 338.2 | 281.23 | 306.35 | 309.02 | 1,234.80 |
| 79 | 220.71 | 206.88 | 190.2 | 180.74 | 798.53 |
| 80 | 2,906.73 | 2,842.77 | 2,553.77 | 2,286.20 | 10,589.47 |
| 81 | 116.78 | 105.27 | 91.66 | 94.07 | 407.78 |
| 82 | 201.88 | 220.48 | 222.94 | 208.17 | 853.47 |
| 83 | 297.74 | 319.01 | 276.51 | 236.36 | 1,129.62 |
| 84 | 15.43 | 12.94 | 28.9 | 16.68 | 73.95 |
| 85 | 368.62 | 331.15 | 292.47 | 301.26 | 1,293.50 |
| 86 | 227.84 | 209.15 | 212.59 | 222.4 | 871.98 |
| 87 | 765.96 | 797.85 | 760.25 | 662.3 | 2,986.36 |
| 88 | 1,179.67 | 1,226.06 | 1,180.96 | 1,158.79 | 4,745.48 |
| 89 | 47.91 | 46.39 | 35.15 | 41.97 | 171.42 |
| TOTAL | 10,127.26 | 10,100.53 | 9,560.28 | 8,897.45 | 38,685.52 |

STATE: NEW MEXICO

| ZIP CODE | QUARTER 1 | QUARTER 2 | QUARTER 3 | QUARTER 4 | TOTAL GRAMS |
|---|---|---|---|---|---|
| 870 | 139.75 | 114.9 | 110.84 | 116.16 | 481.65 |
| 871 | 931.62 | 765.48 | 834.83 | 749.91 | 3,281.84 |
| 873 | 66.91 | 58.61 | 47.77 | 50.72 | 224.01 |
| 874 | 7.23 | 16.86 | 12.53 | 13.92 | 50.54 |
| 875 | 190.46 | 197.33 | 197.17 | 164.44 | 749.4 |
| 877 | 7.76 | 14.63 | 5 | 6.96 | 34.35 |
| 878 | 1.96 | 0.89 | 0.45 | 0 | 3.3 |
| 879 | 31.4 | 26.14 | 18.82 | 12.58 | 88.94 |
| 880 | 227.08 | 212.23 | 158.84 | 123.38 | 721.53 |
| 881 | 2.56 | 0.84 | 1.65 | 1.24 | 6.29 |
| 882 | 52.63 | 54.55 | 56.43 | 71.1 | 234.71 |
| 883 | 44.96 | 46.92 | 32.65 | 40.14 | 164.67 |
| 884 | 6.42 | 7.58 | 5.62 | 3.21 | 22.83 |
| TOTAL | 1,710.74 | 1,516.96 | 1,482.60 | 1,353.76 | 6,064.06 |

STATE: NEW YORK

| ZIP CODE | QUARTER 1 | QUARTER 2 | QUARTER 3 | QUARTER 4 | TOTAL GRAMS |
|---|---|---|---|---|---|
| 100 | 2,028.99 | 2,045.41 | 1,936.08 | 1,843.08 | 7,853.56 |
| 101 | 78.77 | 80.47 | 74.58 | 58.16 | 291.98 |
| 103 | 934.25 | 1,046.52 | 1,032.96 | 1,142.69 | 4,156.42 |
| 104 | 1,061.55 | 995.99 | 938.94 | 969.98 | 3,966.46 |
| 105 | 569.56 | 526.34 | 552.21 | 509.7 | 2,157.81 |
| 106 | 132.57 | 117.85 | 100.41 | 130.43 | 481.26 |
| 107 | 279.76 | 246.22 | 257.68 | 246.98 | 1,030.64 |
| 108 | 59.68 | 61.55 | 71.9 | 70.03 | 263.16 |
| 109 | 1,426.24 | 1,408.40 | 1,178.51 | 1,075.69 | 5,088.84 |
| 110 | 320.17 | 275.93 | 279.41 | 248 | 1,123.51 |
| 111 | 297.07 | 296.36 | 231.5 | 206.52 | 1,031.45 |
| 112 | 2,062.09 | 2,001.60 | 2,249.47 | 2,233.68 | 8,546.84 |
| 113 | 769.53 | 602.97 | 590.39 | 553.82 | 2,516.71 |
| 114 | 202.77 | 226.5 | 231.14 | 243.94 | 904.35 |
| 115 | 760.6 | 670.55 | 739.6 | 656.59 | 2,827.34 |
| 116 | 127.21 | 113.97 | 116.51 | 106.66 | 464.35 |
| 117 | 2,763.32 | 2,699.08 | 2,765.68 | 2,869 | 11,097.08 |
| 118 | 53.44 | 58.3 | 55 | 46.39 | 213.13 |

| | | | | | |
|---|---|---|---|---|---|
| 119 | 205.27 | 186.49 | 195.86 | 215.44 | 803.06 |
| 120 | 179.54 | 191.94 | 201.26 | 216.69 | 789.43 |
| 121 | 107.68 | 116.6 | 97.6 | 135.96 | 457.84 |
| 122 | 58.88 | 41.66 | 44.34 | 39.34 | 184.22 |
| 123 | 49.15 | 37.29 | 53.26 | 58.79 | 198.49 |
| 124 | 425.22 | 411.88 | 436.33 | 378.16 | 1,651.59 |
| 125 | 765.33 | 821.53 | 802.4 | 728.85 | 3,118.11 |
| 126 | 242.38 | 207.64 | 211.61 | 187.88 | 849.51 |
| 127 | 514.43 | 456.53 | 401.8 | 377.22 | 1,749.98 |
| 128 | 227.75 | 243.1 | 242.16 | 295.11 | 1,008.12 |
| 129 | 551.81 | 451.05 | 492.84 | 531.2 | 2,026.90 |
| 130 | 730.94 | 677.55 | 636.02 | 560.91 | 2,605.42 |
| 131 | 185.07 | 178.69 | 148.04 | 138.86 | 650.66 |
| 132 | 266.74 | 271.78 | 262.63 | 235.38 | 1,036.53 |
| 133 | 191 | 195.99 | 201.57 | 190.46 | 779.02 |
| 134 | 299.7 | 278.47 | 282.95 | 255.14 | 1,116.26 |

DRUG CODE:9652DRUG NAME:OXYMORPHONE
STATE:NEW YORK

| ZIP CODE | QUARTER 1 | QUARTER 2 | QUARTER 3 | QUARTER 4 | TOTAL GRAMS |
|---|---|---|---|---|---|
| 135 | 86.09 | 97.95 | 97.95 | 87.43 | 369.42 |
| 136 | 266.92 | 263.35 | 228.96 | 234.62 | 993.85 |
| 137 | 175.79 | 169.54 | 181.36 | 212.14 | 738.83 |
| 138 | 366.04 | 314.33 | 296 | 359.47 | 1,335.84 |
| 139 | 309.56 | 280.21 | 299.97 | 209.51 | 1,099.25 |
| 140 | 1,576.47 | 1,440.07 | 1,285.78 | 1,251.49 | 5,553.81 |
| 141 | 1,018.60 | 886.12 | 806.37 | 703.6 | 3,414.69 |
| 142 | 2,220.88 | 1,847.23 | 1,510.73 | 1,449.44 | 7,028.28 |
| 143 | 612.34 | 511.57 | 403.41 | 390.47 | 1,917.79 |
| 144 | 250.95 | 220.88 | 176.5 | 162.01 | 810.34 |
| 145 | 229.23 | 204.34 | 162.36 | 149.98 | 745.91 |
| 146 | 158.26 | 139.52 | 140.59 | 139.84 | 578.21 |
| 147 | 335.07 | 280.07 | 215.93 | 232.88 | 1,063.95 |
| 148 | 447.57 | 432.76 | 436.73 | 464.78 | 1,781.84 |
| 149 | 134.62 | 87.96 | 81.27 | 88.67 | 392.52 |
| TOTAL | 27,116.85 | 25,418.10 | 24,436.55 | 23,893.06 | 100,864.56 |

STATE: NORTH CAROLINA

| ZIP CODE | QUARTER 1 | QUARTER 2 | QUARTER 3 | QUARTER 4 | TOTAL GRAMS |
|---|---|---|---|---|---|
| 270 | 1,789.37 | 1,689.37 | 1,658.77 | 1,480.93 | 6,618.44 |
| 271 | 1,095.05 | 1,011.82 | 1,009.68 | 1,004.40 | 4,120.95 |
| 272 | 2,699.49 | 2,482.58 | 2,377.89 | 2,199.74 | 9,759.70 |
| 273 | 1,459.07 | 1,236.14 | 1,221.28 | 1,117.44 | 5,033.93 |
| 274 | 676.12 | 642.18 | 586.6 | 610.6 | 2,515.50 |
| 275 | 3,130.82 | 2,933.40 | 2,649.49 | 2,456.62 | 11,170.33 |
| 276 | 785.03 | 802.01 | 802.92 | 753.08 | 3,143.04 |
| 277 | 555.19 | 492.31 | 402.97 | 427.45 | 1,877.92 |
| 278 | 2,613.67 | 2,492.22 | 2,498.77 | 2,376.02 | 9,980.68 |
| 279 | 470.89 | 473.08 | 382.44 | 437.49 | 1,763.90 |
| 280 | 3,103.54 | 2,959.45 | 2,905.51 | 2,616.82 | 11,585.32 |
| 281 | 2,132.82 | 1,974.57 | 1,977.57 | 1,816.07 | 7,901.03 |
| 282 | 1,242.04 | 1,192.43 | 1,118.93 | 1,053.27 | 4,606.67 |
| 283 | 5,168.07 | 4,531.96 | 3,563.94 | 3,125.29 | 16,389.26 |
| 284 | 2,464.11 | 2,336.37 | 2,079.13 | 2,010.35 | 8,889.96 |
| 285 | 1,820.51 | 1,706.90 | 1,695.21 | 1,548.28 | 6,770.90 |
| 286 | 5,226.01 | 4,794.32 | 4,244.42 | 4,002.90 | 18,267.65 |
| 287 | 817.52 | 808.73 | 773.27 | 659.57 | 3,059.09 |
| 288 | 176.46 | 162.5 | 156.56 | 141.98 | 637.5 |
| 289 | 299.79 | 309.47 | 270.4 | 276.1 | 1,155.76 |
| TOTAL | 37,725.57 | 35,031.81 | 32,375.75 | 30,114.40 | 135,247.53 |

STATE: NORTH DAKOTA

| ZIP CODE | QUARTER 1 | QUARTER 2 | QUARTER 3 | QUARTER 4 | TOTAL GRAMS |
|---|---|---|---|---|---|
| 580 | 4.01 | 1.78 | 0.89 | 0.89 | 7.57 |
| 581 | 27.39 | 19.8 | 6.69 | 8.65 | 62.53 |
| 582 | 20.07 | 28.28 | 14.01 | 12.94 | 75.3 |
| 583 | 3.57 | 1.78 | 2.68 | 4.91 | 12.94 |
| 584 | 12.85 | 16.59 | 17.31 | 4.82 | 51.57 |
| 585 | 101.16 | 114.72 | 118.29 | 84.93 | 419.1 |
| 586 | 9.1 | 8.21 | 8.83 | 3.12 | 29.26 |
| 587 | 29.8 | 27.39 | 36.49 | 31.85 | 125.53 |
| 588 | 6.42 | 9.46 | 14.99 | 14.72 | 45.59 |
| TOTAL | 214.37 | 228.01 | 220.18 | 166.83 | 829.39 |

STATE: OHIO

| ZIP CODE | QUARTER 1 | QUARTER 2 | QUARTER 3 | QUARTER 4 | TOTAL GRAMS |
|---|---|---|---|---|---|
| 430 | 721.75 | 623.62 | 696.74 | 700.75 | 2,742.86 |
| 431 | 428.43 | 474.37 | 445.2 | 408.85 | 1,756.85 |
| 432 | 1,546.05 | 1,261.88 | 1,541.38 | 1,248.49 | 5,597.80 |
| 433 | 121.86 | 129.18 | 116.51 | 132.74 | 500.29 |
| 434 | 267.5 | 243.86 | 258.75 | 253.13 | 1,023.24 |
| 435 | 271.62 | 259.33 | 265.02 | 327.11 | 1,123.08 |
| 436 | 716 | 710.52 | 683.77 | 627.06 | 2,737.35 |
| 437 | 146.66 | 146.75 | 136.31 | 149.16 | 578.88 |
| 438 | 31.94 | 39.34 | 57.81 | 46.3 | 175.39 |
| 439 | 651.1 | 601.99 | 573.35 | 547.21 | 2,373.65 |
| 440 | 537.94 | 511.22 | 484.72 | 506.36 | 2,040.24 |
| 441 | 399.46 | 369.78 | 383.47 | 420.09 | 1,572.80 |
| 442 | 673.89 | 573.75 | 532.3 | 548.86 | 2,328.80 |

DRUG CODE:9652DRUG NAME:OXYMORPHONE
STATE:OHIO

| ZIP CODE | QUARTER 1 | QUARTER 2 | QUARTER 3 | QUARTER 4 | TOTAL GRAMS |
|---|---|---|---|---|---|
| 443 | 213.52 | 198.09 | 210.94 | 171.46 | 794.01 |
| 444 | 427.98 | 381.86 | 411.21 | 395.33 | 1,616.38 |
| 445 | 296.98 | 271.96 | 265.89 | 240.91 | 1,075.74 |
| 446 | 479.33 | 451.05 | 372.85 | 354.34 | 1,657.57 |
| 447 | 270.8 | 259.96 | 287.61 | 288.37 | 1,106.74 |
| 448 | 610.91 | 625.41 | 527.99 | 535.53 | 2,299.84 |
| 449 | 40.59 | 50.31 | 40.95 | 64.05 | 195.9 |
| 450 | 1,380.17 | 1,305.32 | 1,152.28 | 1,132.57 | 4,970.34 |
| 451 | 518.62 | 467.91 | 486.6 | 455.06 | 1,928.19 |
| 452 | 740.13 | 793.35 | 743.18 | 723.5 | 3,000.16 |
| 453 | 350.82 | 361.03 | 330.48 | 337.66 | 1,379.99 |
| 454 | 550.22 | 628.02 | 511.35 | 482.22 | 2,171.81 |
| 455 | 172.93 | 137.92 | 135.73 | 129.98 | 576.56 |
| 456 | 1,139.61 | 1,029.48 | 1,178.46 | 1,233.91 | 4,581.46 |
| 457 | 267.45 | 248.36 | 183.86 | 221.02 | 920.69 |
| 458 | 114.23 | 72.84 | 88.1 | 83.77 | 358.94 |
| TOTAL | 14,088.49 | 13,228.46 | 13,102.81 | 12,765.79 | 53,185.55 |

STATE: OKLAHOMA

| ZIP CODE | QUARTER 1 | QUARTER 2 | QUARTER 3 | QUARTER 4 | TOTAL GRAMS |
|---|---|---|---|---|---|
| 730 | 1,844.68 | 1,817.16 | 1,799.19 | 1,771.98 | 7,233.01 |
| 731 | 1,657.03 | 1,558.16 | 1,632.19 | 1,616.40 | 6,463.78 |
| 734 | 108.84 | 119.99 | 96.26 | 78.59 | 403.68 |
| 735 | 295.15 | 322.18 | 352.91 | 270.22 | 1,240.46 |
| 736 | 96.17 | 103.22 | 104.82 | 108.03 | 412.24 |
| 737 | 107.9 | 107.5 | 73.33 | 62.45 | 351.18 |

| | | | | | |
|---|---|---|---|---|---|
| 738 | 23.37 | 22.3 | 24.53 | 26.5 | 96.7 |
| 739 | 0 | 1.43 | 0 | 0 | 1.43 |
| 740 | 1,072.75 | 1,016.24 | 892.28 | 870.87 | 3,852.14 |
| 741 | 1,125.92 | 1,035.37 | 850.22 | 831.17 | 3,842.68 |
| 743 | 272 | 245.24 | 219.19 | 207.15 | 943.58 |
| 744 | 437.31 | 410.1 | 365.36 | 330.21 | 1,542.98 |
| 745 | 203.13 | 189.48 | 194.75 | 173.02 | 760.38 |
| 746 | 69.41 | 61.55 | 56.83 | 51.92 | 239.71 |
| 747 | 124.8 | 90.37 | 81.36 | 83.86 | 380.39 |
| 748 | 395.11 | 386.41 | 387.93 | 409.03 | 1,578.48 |
| 749 | 185.56 | 156.7 | 170.93 | 164.5 | 677.69 |
| TOTAL | 8,019.13 | 7,643.40 | 7,302.08 | 7,055.90 | 30,020.51 |

STATE: OREGON

| ZIP CODE | QUARTER 1 | QUARTER 2 | QUARTER 3 | QUARTER 4 | TOTAL GRAMS |
|---|---|---|---|---|---|
| 970 | 513.3 | 435.82 | 394.94 | 365.23 | 1,709.29 |
| 971 | 135.42 | 129.31 | 78.86 | 74 | 417.59 |
| 972 | 359 | 361.86 | 422.53 | 340.5 | 1,483.89 |
| 973 | 364.81 | 317.96 | 294.97 | 306.89 | 1,284.63 |
| 974 | 614.05 | 614.59 | 551.7 | 543.22 | 2,323.56 |
| 975 | 206.21 | 158.79 | 207.23 | 184.71 | 756.94 |
| 976 | 20.88 | 16.46 | 15.79 | 14.1 | 67.23 |
| 977 | 134.1 | 140.97 | 106.53 | 98.56 | 480.16 |
| 978 | 67.44 | 51.87 | 71.72 | 80.74 | 271.77 |
| 979 | 28.55 | 47.24 | 61.33 | 47.19 | 184.31 |
| TOTAL | 2,443.76 | 2,274.87 | 2,205.60 | 2,055.14 | 8,979.37 |

STATE: PENNSYLVANIA

| ZIP CODE | QUARTER 1 | QUARTER 2 | QUARTER 3 | QUARTER 4 | TOTAL GRAMS |
|---|---|---|---|---|---|
| 150 | 2,319.82 | 2,173.16 | 2,059.81 | 2,061.73 | 8,614.52 |
| 151 | 1,113.83 | 1,048.31 | 997.06 | 1,039.97 | 4,199.17 |
| 152 | 3,168.54 | 2,876.71 | 2,942.59 | 2,927.60 | 11,915.44 |
| 153 | 864.44 | 825.64 | 704.85 | 772.02 | 3,166.95 |
| 154 | 1,545.61 | 1,565.46 | 1,552.30 | 1,467.95 | 6,131.32 |
| 155 | 445.25 | 435.34 | 404.66 | 480.08 | 1,765.33 |
| 156 | 1,300.90 | 1,227.31 | 1,215.93 | 1,286.54 | 5,030.68 |
| 157 | 630 | 599 | 650.61 | 630.63 | 2,510.24 |
| 158 | 489.72 | 550.78 | 575.94 | 535.97 | 2,152.41 |
| 159 | 1,498.82 | 1,464.61 | 1,554.66 | 1,633.03 | 6,151.12 |
| 160 | 671.48 | 549.8 | 585.18 | 580.49 | 2,386.95 |
| 161 | 1,443.64 | 1,445.74 | 1,385.57 | 1,512.38 | 5,787.33 |
| 162 | 536.95 | 558.41 | 507.34 | 475.4 | 2,078.10 |
| 163 | 657.79 | 616.62 | 710.96 | 637.36 | 2,622.73 |

DRUG CODE:9652DRUG NAME:OXYMORPHONE
STATE:PENNSYLVANIA

| ZIP CODE | QUARTER 1 | QUARTER 2 | QUARTER 3 | QUARTER 4 | TOTAL GRAMS |
|---|---|---|---|---|---|
| 164 | 187.88 | 173.83 | 166.78 | 174.9 | 703.39 |
| 165 | 428.61 | 379.45 | 383.16 | 362.46 | 1,553.68 |
| 166 | 637.14 | 677.37 | 756.99 | 886.44 | 2,957.94 |
| 167 | 107.68 | 117 | 137.56 | 179.49 | 541.73 |
| 168 | 140.19 | 154.53 | 186.78 | 148.49 | 629.99 |
| 169 | 18.56 | 19.72 | 10.53 | 14.63 | 63.44 |
| 170 | 1,108.21 | 1,043.09 | 1,052.81 | 951.6 | 4,155.71 |
| 171 | 281.55 | 253.85 | 234.13 | 239.08 | 1,008.61 |
| 172 | 444.31 | 493.42 | 482.14 | 521.74 | 1,941.61 |
| 173 | 560.91 | 549.44 | 606.72 | 560.15 | 2,277.22 |
| 174 | 306.88 | 321.16 | 356.35 | 346.85 | 1,331.24 |
| 175 | 382.62 | 360.54 | 310.81 | 293.14 | 1,347.11 |

| | | | | | |
|---|---|---|---|---|---|
| 176 | 185.02 | 200.9 | 215 | 184.75 | 785.67 |
| 177 | 99.83 | 89.3 | 92.29 | 58.16 | 339.58 |
| 178 | 135.42 | 119.99 | 117 | 97.33 | 469.74 |
| 179 | 78.06 | 43.89 | 34.17 | 38.54 | 194.66 |
| 180 | 366.83 | 291.58 | 312.06 | 256.3 | 1,226.77 |
| 181 | 206.34 | 194.97 | 175.83 | 167.09 | 744.23 |
| 182 | 56.87 | 87.11 | 60.75 | 161.69 | 366.42 |
| 183 | 195.01 | 169.45 | 139.17 | 120.75 | 624.38 |
| 184 | 203.89 | 198.45 | 209.29 | 212.72 | 824.35 |
| 185 | 139.93 | 165.22 | 150.72 | 174.63 | 630.5 |
| 186 | 126.54 | 148.27 | 145.06 | 145.68 | 565.55 |
| 187 | 1,807.84 | 1,637.05 | 1,677.64 | 1,695.48 | 6,818.01 |
| 188 | 50.58 | 52.32 | 41.84 | 45.14 | 189.88 |
| 189 | 258.73 | 253.24 | 263.92 | 229.19 | 1,005.08 |
| 190 | 2,150.41 | 2,222.49 | 2,200.28 | 2,045.94 | 8,619.12 |
| 191 | 852.14 | 834.07 | 819.39 | 719.66 | 3,225.26 |
| 193 | 536.64 | 503.64 | 524.97 | 500.3 | 2,065.55 |
| 194 | 508.6 | 523.15 | 477.37 | 384.9 | 1,894.02 |
| 195 | 113.79 | 141.04 | 149.29 | 130.02 | 534.14 |
| 196 | 172.26 | 142.02 | 176.1 | 157.68 | 648.06 |
| TOTAL | 29,536.06 | 28,498.44 | 28,514.36 | 28,246.07 | 114,794.93 |

STATE: RHODE ISLAND

| ZIP CODE | QUARTER 1 | QUARTER 2 | QUARTER 3 | QUARTER 4 | TOTAL GRAMS |
|---|---|---|---|---|---|
| 28 | 387.67 | 352.93 | 351.59 | 325.01 | 1,417.20 |
| 29 | 264.6 | 226.05 | 230.68 | 235.37 | 956.7 |
| TOTAL | 652.27 | 578.98 | 582.27 | 560.38 | 2,373.90 |

STATE: SOUTH CAROLINA

| ZIP CODE | QUARTER 1 | QUARTER 2 | QUARTER 3 | QUARTER 4 | TOTAL GRAMS |
|---|---|---|---|---|---|
| 290 | 797.98 | 868.1 | 883.8 | 922.7 | 3,472.58 |
| 291 | 814.98 | 699.76 | 739.91 | 771.31 | 3,025.96 |
| 292 | 810.43 | 799.37 | 878.58 | 910.61 | 3,398.99 |
| 293 | 441.72 | 457.65 | 452.29 | 431.95 | 1,783.61 |
| 294 | 1,774.55 | 1,892.49 | 1,815.45 | 1,818.46 | 7,300.95 |
| 295 | 1,145.77 | 1,135.02 | 1,100.36 | 1,097.37 | 4,478.52 |
| 296 | 1,458 | 1,440.79 | 1,321.20 | 1,235.20 | 5,455.19 |
| 297 | 661.94 | 602.84 | 620.05 | 527.94 | 2,412.77 |
| 298 | 146.3 | 132.74 | 138.28 | 177.88 | 595.2 |
| 299 | 283.69 | 309.11 | 286.99 | 304.96 | 1,184.75 |
| TOTAL | 8,335.36 | 8,337.87 | 8,236.91 | 8,198.38 | 33,108.52 |

STATE: SOUTH DAKOTA

| ZIP CODE | QUARTER 1 | QUARTER 2 | QUARTER 3 | QUARTER 4 | TOTAL GRAMS |
|---|---|---|---|---|---|
| 570 | 94.47 | 92.73 | 78.55 | 77.92 | 343.67 |
| 571 | 315.82 | 248.56 | 172.91 | 163.6 | 900.89 |
| 572 | 5.35 | 9.28 | 7.49 | 6.42 | 28.54 |
| 573 | 2.32 | 7.14 | 6.24 | 5.08 | 20.78 |
| 574 | 9.28 | 10.71 | 9.81 | 19.09 | 48.89 |
| 575 | 4.1 | 6.24 | 3.21 | 0.89 | 14.44 |
| 576 | 3.57 | 5.35 | 3.57 | 3.57 | 16.06 |
| 577 | 491.99 | 453.1 | 452.25 | 406.35 | 1,803.69 |
| TOTAL | 926.9 | 833.11 | 734.03 | 682.92 | 3,176.96 |

STATE: TENNESSEE

| ZIP CODE | QUARTER 1 | QUARTER 2 | QUARTER 3 | QUARTER 4 | TOTAL GRAMS |
|---|---|---|---|---|---|
| 370 | 4,297.20 | 4,245.28 | 3,971.14 | 3,609.70 | 16,123.32 |
| 371 | 2,435.88 | 2,503.59 | 2,439.31 | 2,285.98 | 9,664.76 |

RETAIL DRUG DISTRIBUTION BY ZIP CODE WITHIN STATE BY GRAMS WT

STATE:TENNESSEE

| ZIP CODE | QUARTER 1 | QUARTER 2 | QUARTER 3 | QUARTER 4 | TOTAL GRAMS |
|---|---|---|---|---|---|
| 372 | 2,288.74 | 2,385.54 | 2,034.87 | 1,832.19 | 8,541.34 |
| 373 | 2,411.66 | 2,236.36 | 2,093.85 | 1,901.96 | 8,643.83 |
| 374 | 1,144.70 | 1,022.44 | 986.48 | 896.29 | 4,049.91 |
| 376 | 1,889.69 | 1,849.23 | 1,696.73 | 1,629.11 | 7,064.76 |
| 377 | 4,704 | 4,250.46 | 4,351.75 | 4,311.61 | 17,617.82 |
| 378 | 6,595.88 | 5,890.80 | 5,384.94 | 5,212.50 | 23,084.12 |
| 379 | 2,424.19 | 2,129.31 | 1,878.67 | 1,855.45 | 8,287.62 |
| 380 | 260.14 | 251.17 | 237.52 | 232.53 | 981.36 |
| 381 | 291.63 | 265.09 | 311.21 | 263.57 | 1,131.50 |
| 382 | 418.04 | 355.77 | 358.27 | 345.82 | 1,477.90 |
| 383 | 613.59 | 534.72 | 530.09 | 594.76 | 2,273.16 |
| 384 | 633.26 | 586.24 | 521.61 | 466.79 | 2,207.90 |
| 385 | 1,637.09 | 1,493.51 | 1,499.89 | 1,507.87 | 6,138.36 |
| TOTAL | 32,045.69 | 29,999.51 | 28,296.33 | 26,946.13 | 117,287.66 |

STATE: TEXAS

| ZIP CODE | QUARTER 1 | QUARTER 2 | QUARTER 3 | QUARTER 4 | TOTAL GRAMS |
|---|---|---|---|---|---|
| 750 | 1,752.85 | 1,772.38 | 1,860.88 | 2,017.66 | 7,403.77 |
| 751 | 355.99 | 322.09 | 324.64 | 301.98 | 1,304.70 |
| 752 | 544.41 | 524.47 | 497.7 | 544.18 | 2,110.76 |
| 754 | 163.08 | 208.53 | 176.55 | 223.43 | 771.59 |
| 755 | 3.48 | 5.66 | 4.91 | 6.33 | 20.38 |
| 756 | 57.54 | 56.2 | 39.07 | 49.96 | 202.77 |
| 757 | 89.79 | 81.23 | 71.95 | 58.79 | 301.76 |
| 758 | 47.77 | 63.61 | 51.88 | 56.56 | 219.82 |
| 759 | 110.98 | 97.77 | 94.47 | 96.35 | 399.57 |
| 760 | 1,231.06 | 1,203.26 | 1,079.76 | 1,069.40 | 4,583.48 |
| 761 | 803.74 | 791.65 | 772.02 | 794.24 | 3,161.65 |
| 762 | 251.04 | 224.59 | 264.64 | 293.23 | 1,033.50 |
| 763 | 286.01 | 297.92 | 306.57 | 279.9 | 1,170.40 |
| 764 | 56.92 | 70.3 | 59.06 | 45.32 | 231.6 |
| 765 | 140.77 | 142.29 | 137.78 | 139.17 | 560.01 |
| 766 | 27.61 | 30.33 | 21.5 | 26.23 | 105.67 |
| 767 | 46.3 | 35.57 | 18.47 | 19.98 | 120.32 |
| 768 | 40.32 | 33.36 | 45.5 | 41.48 | 160.66 |
| 769 | 11.42 | 10.88 | 38.09 | 71.28 | 131.67 |
| 770 | 1,078.28 | 918.2 | 834.2 | 854.82 | 3,685.50 |
| 773 | 544.95 | 532.66 | 615.18 | 578.64 | 2,271.43 |
| 774 | 855.03 | 861.55 | 861.19 | 876.67 | 3,454.44 |
| 775 | 986.93 | 944.51 | 865.74 | 834.47 | 3,631.65 |
| 776 | 377.67 | 363.08 | 362.06 | 330.52 | 1,433.33 |
| 777 | 145.81 | 141.98 | 146.48 | 164.82 | 599.09 |
| 778 | 168.21 | 195.8 | 161.47 | 152.56 | 678.04 |
| 779 | 188.68 | 150.23 | 147.02 | 136.36 | 622.29 |
| 780 | 310.18 | 274.68 | 262.99 | 235.96 | 1,083.81 |
| 781 | 264.06 | 278.51 | 269.19 | 308.85 | 1,120.61 |
| 782 | 2,063.87 | 2,033.68 | 1,855.36 | 1,827.57 | 7,780.48 |
| 783 | 90.01 | 81.89 | 81 | 98.67 | 351.57 |
| 784 | 221.87 | 260.14 | 222.13 | 222.71 | 926.85 |
| 785 | 29.97 | 40.5 | 50.49 | 40.06 | 161.02 |
| 786 | 1,403.23 | 1,262.59 | 1,237.94 | 1,338.34 | 5,242.10 |
| 787 | 1,208.94 | 1,279.76 | 1,071.51 | 1,112.68 | 4,672.89 |
| 788 | 33.99 | 42.29 | 51.65 | 50.36 | 178.29 |
| 789 | 26.5 | 31.58 | 19.98 | 26.94 | 105 |
| 790 | 85.73 | 122.31 | 112.94 | 118.38 | 439.36 |
| 791 | 165.04 | 154.24 | 141.13 | 151.84 | 612.25 |
| 793 | 25.07 | 26.5 | 30.42 | 24.71 | 106.7 |
| 794 | 46.3 | 39.43 | 44.61 | 49.24 | 179.58 |
| 795 | 13.11 | 6.69 | 5.08 | 3.21 | 28.09 |

| | | | | | |
|---|---|---|---|---|---|
| 796 | 12.58 | 19.8 | 22.93 | 33.72 | 89.03 |
| 797 | 134.35 | 156.47 | 165.13 | 178.42 | 634.37 |
| 798 | 14.99 | 13.38 | 11.42 | 7.32 | 47.11 |
| 799 | 209.82 | 148.49 | 144.79 | 154.87 | 657.97 |
| TOTAL | 16,726.25 | 16,353.03 | 15,659.47 | 16,048.18 | 64,786.93 |

STATE: UTAH

| ZIP CODE | QUARTER 1 | QUARTER 2 | QUARTER 3 | QUARTER 4 | TOTAL GRAMS |
|---|---|---|---|---|---|
| 840 | 2,829.88 | 2,752.45 | 2,669.31 | 2,535.88 | 10,787.52 |

DRUG CODE:9652DRUG NAME:OXYMORPHONE
STATE:UTAH

| ZIP CODE | QUARTER 1 | QUARTER 2 | QUARTER 3 | QUARTER 4 | TOTAL GRAMS |
|---|---|---|---|---|---|
| 841 | 896.3 | 886.05 | 924.5 | 937.74 | 3,644.59 |
| 843 | 218.52 | 205.54 | 193.36 | 176.68 | 794.1 |
| 844 | 560.77 | 604.17 | 593.16 | 525.27 | 2,283.37 |
| 845 | 93.76 | 66.91 | 92.73 | 83.77 | 337.17 |
| 846 | 405.77 | 502.12 | 465.41 | 377.36 | 1,750.66 |
| 847 | 311.16 | 371.69 | 351.89 | 363.44 | 1,398.18 |
| TOTAL | 5,316.16 | 5,388.93 | 5,290.36 | 5,000.14 | 20,995.59 |

STATE: VERMONT

| ZIP CODE | QUARTER 1 | QUARTER 2 | QUARTER 3 | QUARTER 4 | TOTAL GRAMS |
|---|---|---|---|---|---|
| 50 | 4.55 | 3.93 | 6.96 | 6.69 | 22.13 |
| 51 | 18.42 | 16.06 | 13.83 | 16.24 | 64.55 |
| 52 | 31.76 | 32.65 | 34.97 | 32.74 | 132.12 |
| 53 | 17.13 | 13.29 | 6.96 | 4.55 | 41.93 |
| 54 | 298.1 | 284 | 282.48 | 232.7 | 1,097.28 |
| 56 | 30.38 | 23.55 | 28.68 | 33.19 | 115.8 |
| 57 | 77.88 | 73.73 | 94.79 | 62.67 | 309.07 |
| 58 | 7.05 | 6.24 | 2.68 | 3.12 | 19.09 |
| TOTAL | 485.27 | 453.45 | 471.35 | 391.9 | 1,801.97 |

STATE: VIRGINIA

| ZIP CODE | QUARTER 1 | QUARTER 2 | QUARTER 3 | QUARTER 4 | TOTAL GRAMS |
|---|---|---|---|---|---|
| 201 | 1,235.38 | 1,156.61 | 1,143.63 | 1,065.88 | 4,601.50 |
| 220 | 920.18 | 800.53 | 926.76 | 869.84 | 3,517.31 |
| 221 | 874.35 | 1,116.70 | 880.65 | 789.89 | 3,661.59 |
| 222 | 139.21 | 127.17 | 110 | 140.91 | 517.29 |
| 223 | 260.67 | 278.34 | 262.1 | 249.79 | 1,050.90 |
| 224 | 1,181.59 | 1,236.99 | 1,154.24 | 1,163.57 | 4,736.39 |
| 225 | 192.69 | 193.85 | 213.61 | 186.63 | 786.78 |
| 226 | 446.18 | 485.17 | 518.58 | 466.48 | 1,916.41 |
| 227 | 85.64 | 92.38 | 96.12 | 76.41 | 350.55 |
| 228 | 78.59 | 110.75 | 111.56 | 81.18 | 382.08 |
| 229 | 167.18 | 171.73 | 187.25 | 157.14 | 683.3 |
| 230 | 326.37 | 303.31 | 307.24 | 277.44 | 1,214.36 |
| 231 | 498.82 | 439.09 | 415.94 | 401.31 | 1,755.16 |
| 232 | 1,118.56 | 873.63 | 663.9 | 567.15 | 3,223.24 |
| 233 | 478.92 | 433.83 | 440.57 | 418.44 | 1,771.76 |
| 234 | 993.24 | 964.33 | 914.64 | 889.25 | 3,761.46 |
| 235 | 316.29 | 336.82 | 322.07 | 304.04 | 1,279.22 |
| 236 | 312.68 | 305.72 | 298.45 | 275.17 | 1,192.02 |
| 237 | 188.68 | 195.55 | 202.86 | 186.89 | 773.98 |
| 238 | 441.23 | 457.78 | 380.44 | 424.91 | 1,704.36 |
| 239 | 86.85 | 89.08 | 99.92 | 85.82 | 361.67 |
| 240 | 552.91 | 542.16 | 448.37 | 533.79 | 2,077.23 |
| 241 | 430.3 | 394.58 | 437.22 | 345.87 | 1,607.97 |

| | | | | | |
|---|---|---|---|---|---|
| 242 | 606.46 | 514.65 | 555.51 | 531.12 | 2,207.74 |
| 243 | 251.26 | 343.68 | 500.07 | 572.42 | 1,667.43 |
| 244 | 72.8 | 96.44 | 66.82 | 101.88 | 337.94 |
| 245 | 709 | 654.18 | 540.48 | 499.67 | 2,403.33 |
| 246 | 405.73 | 394.93 | 386.28 | 348.23 | 1,535.17 |
| TOTAL | 13,371.76 | 13,109.98 | 12,585.28 | 12,011.12 | 51,078.14 |

STATE: WASHINGTON

| ZIP CODE | QUARTER 1 | QUARTER 2 | QUARTER 3 | QUARTER 4 | TOTAL GRAMS |
|---|---|---|---|---|---|
| 980 | 1,000.23 | 1,017.20 | 977.44 | 903.16 | 3,898.03 |
| 981 | 272 | 262.46 | 220.36 | 238.1 | 992.92 |
| 982 | 534.16 | 521.44 | 545.08 | 507.59 | 2,108.27 |
| 983 | 265.15 | 310.58 | 289.04 | 292.39 | 1,157.16 |
| 984 | 241.71 | 223.2 | 199.03 | 189.04 | 852.98 |
| 985 | 503.99 | 470.85 | 437.71 | 338.24 | 1,750.79 |
| 986 | 754.86 | 676.71 | 624.03 | 592.51 | 2,648.11 |
| 988 | 82.07 | 80.38 | 62.22 | 57.76 | 282.43 |
| 989 | 111.66 | 106.35 | 99.38 | 113.41 | 430.8 |
| 990 | 26.76 | 29.84 | 26.45 | 34.75 | 117.8 |
| 991 | 34.43 | 29.81 | 41.14 | 29.72 | 135.1 |
| 992 | 311.24 | 305.8 | 276.32 | 306.3 | 1,199.66 |
| 993 | 229.45 | 250.5 | 237.48 | 264.86 | 982.29 |
| 994 | 28.01 | 24.62 | 20.07 | 14.45 | 87.15 |
| TOTAL | 4,395.72 | 4,309.74 | 4,055.75 | 3,882.28 | 16,643.49 |

DRUG CODE:9652DRUG NAME:OXYMORPHONE
STATE: WEST VIRGINIA

| ZIP CODE | QUARTER 1 | QUARTER 2 | QUARTER 3 | QUARTER 4 | TOTAL GRAMS |
|---|---|---|---|---|---|
| 247 | 82.34 | 96.79 | 55.04 | 103.04 | 337.21 |
| 248 | 231.9 | 250.1 | 277.04 | 220.79 | 979.83 |
| 249 | 120.34 | 124.45 | 127.93 | 107.23 | 479.95 |
| 250 | 226.55 | 389.36 | 165.31 | 140.82 | 922.04 |
| 251 | 453.05 | 424.51 | 318.21 | 269.15 | 1,464.92 |
| 252 | 132.66 | 88.01 | 62.4 | 45.5 | 328.57 |
| 253 | 274.9 | 291.49 | 311.52 | 228.38 | 1,106.29 |
| 254 | 721.8 | 742.05 | 796.11 | 912.35 | 3,172.31 |
| 255 | 686.25 | 624.65 | 537.31 | 326.91 | 2,175.12 |
| 256 | 186.98 | 164.68 | 126.5 | 124.18 | 602.34 |
| 257 | 298.63 | 262.72 | 214.42 | 159.11 | 934.88 |
| 258 | 186.54 | 154.42 | 165.31 | 143.09 | 649.36 |
| 259 | 69.14 | 90.99 | 84.84 | 80.29 | 325.26 |
| 260 | 479.99 | 403.1 | 403.85 | 418.35 | 1,705.29 |
| 261 | 39.88 | 38.98 | 32.83 | 42.2 | 153.89 |
| 262 | 45.14 | 58.92 | 40.1 | 48.31 | 192.47 |
| 263 | 59.82 | 61.38 | 67.84 | 58.74 | 247.78 |
| 264 | 22.3 | 21.23 | 16.41 | 26.85 | 86.79 |
| 265 | 90.37 | 98.93 | 79.58 | 82.43 | 351.31 |
| 266 | 21.14 | 28.46 | 19.8 | 22.3 | 91.7 |
| 267 | 45.23 | 54.15 | 72.08 | 82.97 | 254.43 |
| 268 | 0 | 2.14 | 0 | 3.48 | 5.62 |
| TOTAL | 4,474.95 | 4,471.51 | 3,974.43 | 3,646.47 | 16,567.36 |

STATE: WISCONSIN

| ZIP CODE | QUARTER 1 | QUARTER 2 | QUARTER 3 | QUARTER 4 | TOTAL GRAMS |
|---|---|---|---|---|---|
| 530 | 524.26 | 504.54 | 411.88 | 393.43 | 1,834.11 |
| 531 | 765.48 | 728.63 | 691.56 | 666.99 | 2,852.66 |
| 532 | 690.74 | 587 | 576.81 | 576.89 | 2,431.44 |
| 534 | 131.67 | 131.58 | 151.92 | 113.56 | 528.73 |
| 535 | 140.6 | 178.83 | 163.21 | 145.92 | 628.56 |

| | | | | | |
|---|---|---|---|---|---|
| 537 | 85.01 | 95.44 | 74.2 | 84.48 | 339.13 |
| 538 | 7.7 | 8.3 | 2.16 | 3.21 | 21.37 |
| 539 | 103.31 | 82.07 | 50.32 | 62.67 | 298.37 |
| 540 | 29.44 | 18.64 | 16.24 | 21.68 | 86 |
| 541 | 83.47 | 83.38 | 83.15 | 89.99 | 339.99 |
| 542 | 65.57 | 65.03 | 50.05 | 37.02 | 217.67 |
| 543 | 89.7 | 90.99 | 71.86 | 80.29 | 332.84 |
| 544 | 205.73 | 177.18 | 215.63 | 169.07 | 767.61 |
| 545 | 30.78 | 33.28 | 35.77 | 26.32 | 126.15 |
| 546 | 13.6 | 7.76 | 6.42 | 0.59 | 28.37 |
| 547 | 79.66 | 60.66 | 63.25 | 63.8 | 267.37 |
| 548 | 57.09 | 66.19 | 28.37 | 31.22 | 182.87 |
| 549 | 717.19 | 561.06 | 550.02 | 505.74 | 2,334.01 |
| TOTAL | 3,821 | 3,480.56 | 3,242.82 | 3,072.87 | 13,617.25 |

STATE: WYOMING

| ZIP CODE | QUARTER 1 | QUARTER 2 | QUARTER 3 | QUARTER 4 | TOTAL GRAMS |
|---|---|---|---|---|---|
| 820 | 67.98 | 77.21 | 58.92 | 45.59 | 249.7 |
| 822 | 16.77 | 8.74 | 10.71 | 18.2 | 54.42 |
| 823 | 8.03 | 8.03 | 1.34 | 3.57 | 20.97 |
| 824 | 10.17 | 12.94 | 17.22 | 12.67 | 53 |
| 825 | 43.22 | 50.89 | 48.66 | 44.43 | 187.2 |
| 826 | 71.77 | 74.94 | 59.06 | 57.67 | 263.44 |
| 827 | 26.23 | 30.78 | 29.17 | 25.96 | 112.14 |
| 828 | 120.17 | 123.47 | 53.08 | 68.33 | 365.05 |
| 829 | 47.28 | 61.02 | 50.49 | 75.29 | 234.08 |
| 830 | 2.14 | 5.17 | 12.85 | 10.79 | 30.95 |
| 831 | 23.91 | 25.96 | 23.82 | 32.65 | 106.34 |
| TOTAL | 437.67 | 479.15 | 365.32 | 395.15 | 1,677.29 |

DRUG CODE:9668DRUG NAME:NOROXYMORPHONE
STATE: ALABAMA

| ZIP CODE | QUARTER 1 | QUARTER 2 | QUARTER 3 | QUARTER 4 | TOTAL GRAMS |
|---|---|---|---|---|---|
| 352 | 1.95 | 1.14 | 0.92 | 0.27 | 4.28 |
| 363 | 0.1 | 0 | 0 | 0 | 0.1 |
| TOTAL | 2.05 | 1.14 | 0.92 | 0.27 | 4.38 |

STATE: ARIZONA

DRUG CODE:9668DRUG NAME:NOROXYMORPHONE
STATE:ARIZONA

| ZIP CODE | QUARTER 1 | QUARTER 2 | QUARTER 3 | QUARTER 4 | TOTAL GRAMS |
|---|---|---|---|---|---|
| 857 | 0.77 | 0.65 | 0.27 | 0 | 1.69 |
| TOTAL | 0.77 | 0.65 | 0.27 | 0 | 1.69 |

STATE: ARKANSAS

| ZIP CODE | QUARTER 1 | QUARTER 2 | QUARTER 3 | QUARTER 4 | TOTAL GRAMS |
|---|---|---|---|---|---|
| 722 | 10.53 | 6.11 | 4.96 | 7.64 | 29.24 |
| 727 | 4.84 | 4.99 | 1.5 | 15.9 | 27.23 |
| TOTAL | 15.37 | 11.1 | 6.46 | 23.54 | 56.47 |

STATE: CALIFORNIA

| ZIP CODE | QUARTER 1 | QUARTER 2 | QUARTER 3 | QUARTER 4 | TOTAL GRAMS |
|---|---|---|---|---|---|
| 900 | 0.34 | 0.14 | 0.27 | 0 | 0.75 |
| 902 | 2.76 | 8.28 | 0 | 12.63 | 23.67 |
| 905 | 5.59 | 0.87 | 3.73 | 15.16 | 25.35 |
| 906 | 1.63 | 5.03 | 3.95 | 7.42 | 18.03 |
| 907 | 2 | 0.58 | 0.57 | 7.16 | 10.31 |
| 908 | 1.51 | 0.87 | 0.62 | 1.51 | 4.51 |

| | | | | | |
|---|---|---|---|---|---|
| 911 | 0.37 | 0 | 0 | 0 | 0.37 |
| 912 | 5.03 | 4.08 | 7.25 | 7.16 | 23.52 |
| 914 | 0.23 | 0.16 | 0.08 | 0 | 0.47 |
| 917 | 0.76 | 0.73 | 0.33 | 0.1 | 1.92 |
| 919 | 9.46 | 5.42 | 10.9 | 10.54 | 36.32 |
| 920 | 3.07 | 4.83 | 2.09 | 10.99 | 20.98 |
| 921 | 6.79 | 11.47 | 4.05 | 2.64 | 24.95 |
| 923 | 5.34 | 1.32 | 1.87 | 3.86 | 12.39 |
| 925 | 0.61 | 0.18 | 0.03 | 1.59 | 2.41 |
| 928 | 19.5 | 28.7 | 25.29 | 27.74 | 101.23 |
| 946 | 2.41 | 5.03 | 3.45 | 3.74 | 14.63 |
| TOTAL | 67.4 | 77.69 | 64.48 | 112.24 | 321.81 |

STATE: COLORADO

| ZIP CODE | QUARTER 1 | QUARTER 2 | QUARTER 3 | QUARTER 4 | TOTAL GRAMS |
|---|---|---|---|---|---|
| 809 | 0.19 | 0.14 | 0.2 | 0 | 0.53 |
| TOTAL | 0.19 | 0.14 | 0.2 | 0 | 0.53 |

STATE: CONNECTICUT

| ZIP CODE | QUARTER 1 | QUARTER 2 | QUARTER 3 | QUARTER 4 | TOTAL GRAMS |
|---|---|---|---|---|---|
| 61 | 0.24 | 0.17 | 0.03 | 0 | 0.44 |
| 63 | 0.38 | 0.17 | 0.03 | 0 | 0.58 |
| TOTAL | 0.62 | 0.34 | 0.06 | 0 | 1.02 |

STATE: DISTRICT OF COLUMBIA

| ZIP CODE | QUARTER 1 | QUARTER 2 | QUARTER 3 | QUARTER 4 | TOTAL GRAMS |
|---|---|---|---|---|---|
| 200 | 4.03 | 3.18 | 0.07 | 2 | 9.28 |
| TOTAL | 4.03 | 3.18 | 0.07 | 2 | 9.28 |

STATE: FLORIDA

| ZIP CODE | QUARTER 1 | QUARTER 2 | QUARTER 3 | QUARTER 4 | TOTAL GRAMS |
|---|---|---|---|---|---|
| 321 | 1.08 | 0.17 | 0.33 | 0 | 1.58 |
| 322 | 1.38 | 1.04 | 0.34 | 0.1 | 2.86 |
| 327 | 0.73 | 0 | 0.03 | 0 | 0.76 |
| 328 | 0.93 | 3.28 | 0.03 | 0 | 4.24 |
| 329 | 0.59 | 1.62 | 2.03 | 2.15 | 6.39 |
| 330 | 1.86 | 0.86 | 0.33 | 0 | 3.05 |
| 331 | 4.67 | 3.52 | 9.46 | 10.36 | 28.01 |
| 333 | 17.3 | 12.75 | 4.59 | 15.3 | 49.94 |
| 336 | 0.42 | 0 | 0 | 0 | 0.42 |
| 338 | 0.75 | 0.42 | 0.58 | 0.3 | 2.05 |
| 342 | 0.17 | 0.17 | 0.03 | 0 | 0.37 |
| TOTAL | 29.88 | 23.83 | 17.75 | 28.21 | 99.67 |

STATE: GEORGIA

| ZIP CODE | QUARTER 1 | QUARTER 2 | QUARTER 3 | QUARTER 4 | TOTAL GRAMS |
|---|---|---|---|---|---|
| 300 | 3.54 | 5.59 | 0.27 | 1.42 | 10.82 |
| 303 | 0.53 | 0 | 5.27 | 1.59 | 7.39 |
| 309 | 1.26 | 2.01 | 1.62 | 0.62 | 5.51 |
| TOTAL | 5.33 | 7.6 | 7.16 | 3.63 | 23.72 |

STATE: ILLINOIS

| ZIP CODE | QUARTER 1 | QUARTER 2 | QUARTER 3 | QUARTER 4 | TOTAL GRAMS |
|---|---|---|---|---|---|
| 600 | 0.93 | 0.49 | 0.64 | 0 | 2.06 |
| 601 | 2.19 | 1.01 | 0.86 | 0.57 | 4.63 |
| 604 | 0.52 | 0 | 0 | 0 | 0.52 |
| 606 | 14.01 | 8.02 | 12.89 | 10.52 | 45.44 |

RETAIL DRUG DISTRIBUTION BY ZIP CODE WITHIN STATE BY GRAMS WT

DRUG CODE:9668DRUG NAME:NOROXYMORPHONE
STATE:ILLINOIS

| ZIP CODE | QUARTER 1 | QUARTER 2 | QUARTER 3 | QUARTER 4 | TOTAL GRAMS |
|---|---|---|---|---|---|
| TOTAL | 17.65 | 9.52 | 14.39 | 11.09 | 52.65 |

STATE: INDIANA

| ZIP CODE | QUARTER 1 | QUARTER 2 | QUARTER 3 | QUARTER 4 | TOTAL GRAMS |
|---|---|---|---|---|---|
| 462 | 0.54 | 0.34 | 0.2 | 0 | 1.08 |
| 463 | 0.17 | 0.11 | 0.2 | 0 | 0.48 |
| TOTAL | 0.71 | 0.45 | 0.4 | 0 | 1.56 |

STATE: KANSAS

| ZIP CODE | QUARTER 1 | QUARTER 2 | QUARTER 3 | QUARTER 4 | TOTAL GRAMS |
|---|---|---|---|---|---|
| 662 | 0.7 | 0 | 0 | 0 | 0.7 |
| TOTAL | 0.7 | 0 | 0 | 0 | 0.7 |

STATE: LOUISIANA

| ZIP CODE | QUARTER 1 | QUARTER 2 | QUARTER 3 | QUARTER 4 | TOTAL GRAMS |
|---|---|---|---|---|---|
| 711 | 0.39 | 0 | 0 | 0.23 | 0.62 |
| TOTAL | 0.39 | 0 | 0 | 0.23 | 0.62 |

STATE: MARYLAND

| ZIP CODE | QUARTER 1 | QUARTER 2 | QUARTER 3 | QUARTER 4 | TOTAL GRAMS |
|---|---|---|---|---|---|
| 212 | 1.02 | 0.77 | 0.67 | 0 | 2.46 |
| TOTAL | 1.02 | 0.77 | 0.67 | 0 | 2.46 |

STATE: MASSACHUSETTS

| ZIP CODE | QUARTER 1 | QUARTER 2 | QUARTER 3 | QUARTER 4 | TOTAL GRAMS |
|---|---|---|---|---|---|
| 24 | 0.37 | 0 | 0.16 | 1.47 | 2 |
| TOTAL | 0.37 | 0 | 0.16 | 1.47 | 2 |

STATE: MISSISSIPPI

| ZIP CODE | QUARTER 1 | QUARTER 2 | QUARTER 3 | QUARTER 4 | TOTAL GRAMS |
|---|---|---|---|---|---|
| 392 | 8.08 | 3.7 | 3.53 | 0.88 | 16.19 |
| TOTAL | 8.08 | 3.7 | 3.53 | 0.88 | 16.19 |

STATE: MISSOURI

| ZIP CODE | QUARTER 1 | QUARTER 2 | QUARTER 3 | QUARTER 4 | TOTAL GRAMS |
|---|---|---|---|---|---|
| 631 | 7.15 | 6.79 | 21.33 | 13.92 | 49.19 |
| 633 | 5.2 | 1.19 | 0.52 | 0.71 | 7.62 |
| TOTAL | 12.35 | 7.98 | 21.85 | 14.63 | 56.81 |

STATE: NEBRASKA

| ZIP CODE | QUARTER 1 | QUARTER 2 | QUARTER 3 | QUARTER 4 | TOTAL GRAMS |
|---|---|---|---|---|---|
| 685 | 0 | 0 | 0 | 1.59 | 1.59 |
| TOTAL | 0 | 0 | 0 | 1.59 | 1.59 |

STATE: NEVADA

| ZIP CODE | QUARTER 1 | QUARTER 2 | QUARTER 3 | QUARTER 4 | TOTAL GRAMS |
|---|---|---|---|---|---|
| 891 | 0 | 8.88 | 13.99 | 20.27 | 43.14 |
| TOTAL | 0 | 8.88 | 13.99 | 20.27 | 43.14 |

STATE: NEW HAMPSHIRE

| ZIP CODE | QUARTER 1 | QUARTER 2 | QUARTER 3 | QUARTER 4 | TOTAL GRAMS |
|---|---|---|---|---|---|
| 30 | 0.39 | 0 | 0 | 0 | 0.39 |
| 37 | 1.14 | 0.87 | 0.37 | 0 | 2.38 |
| TOTAL | 1.53 | 0.87 | 0.37 | 0 | 2.77 |

STATE: NEW JERSEY

| ZIP CODE | QUARTER 1 | QUARTER 2 | QUARTER 3 | QUARTER 4 | TOTAL GRAMS |
|---|---|---|---|---|---|

| | | | | | |
|---|---|---|---|---|---|
| 80 | 0.18 | 13.6 | 4.98 | 6.85 | 25.61 |
| TOTAL | 0.18 | 13.6 | 4.98 | 6.85 | 25.61 |

STATE: NEW MEXICO

| ZIP CODE | QUARTER 1 | QUARTER 2 | QUARTER 3 | QUARTER 4 | TOTAL GRAMS |
|---|---|---|---|---|---|
| 871 | 3.24 | 0 | 0 | 0 | 3.24 |
| TOTAL | 3.24 | 0 | 0 | 0 | 3.24 |

STATE: NEW YORK

| ZIP CODE | QUARTER 1 | QUARTER 2 | QUARTER 3 | QUARTER 4 | TOTAL GRAMS |
|---|---|---|---|---|---|
| 100 | 2.57 | 2.43 | 0.76 | 0.2 | 5.96 |
| 101 | 1.5 | 1.29 | 0.46 | 0.22 | 3.47 |
| 103 | 4.44 | 0.46 | 0.03 | 0 | 4.93 |
| 105 | 0.31 | 0.16 | 0.03 | 0 | 0.5 |
| 110 | 0 | 0 | 0 | 3.18 | 3.18 |
| 112 | 6.54 | 0.28 | 0.62 | 2.25 | 9.69 |
| 114 | 0.79 | 3.39 | 1.4 | 1.49 | 7.07 |
| 115 | 3.56 | 1.25 | 1.75 | 0.76 | 7.32 |
| 146 | 0 | 0 | 0 | 3.18 | 3.18 |
| TOTAL | 19.71 | 9.26 | 5.05 | 11.28 | 45.3 |

STATE: NORTH CAROLINA

| ZIP CODE | QUARTER 1 | QUARTER 2 | QUARTER 3 | QUARTER 4 | TOTAL GRAMS |
|---|---|---|---|---|---|

DRUG CODE:9668DRUG NAME:NOROXYMORPHONE
STATE:NORTH CAROLINA

| ZIP CODE | QUARTER 1 | QUARTER 2 | QUARTER 3 | QUARTER 4 | TOTAL GRAMS |
|---|---|---|---|---|---|
| 272 | 0.21 | 0.14 | 0.2 | 0 | 0.55 |
| 282 | 0.78 | 0.55 | 0.37 | 0 | 1.7 |
| TOTAL | 0.99 | 0.69 | 0.57 | 0 | 2.25 |

STATE: OHIO

| ZIP CODE | QUARTER 1 | QUARTER 2 | QUARTER 3 | QUARTER 4 | TOTAL GRAMS |
|---|---|---|---|---|---|
| 441 | 0.13 | 3.18 | 0.03 | 0 | 3.34 |
| 447 | 1.63 | 4.08 | 1.11 | 0.56 | 7.38 |
| 452 | 0.72 | 0.6 | 0.28 | 0 | 1.6 |
| 454 | 12.03 | 11.13 | 9.09 | 8.73 | 40.98 |
| TOTAL | 14.51 | 18.99 | 10.51 | 9.29 | 53.3 |

STATE: OKLAHOMA

| ZIP CODE | QUARTER 1 | QUARTER 2 | QUARTER 3 | QUARTER 4 | TOTAL GRAMS |
|---|---|---|---|---|---|
| 731 | 1.56 | 0.49 | 0.69 | 0.07 | 2.81 |
| TOTAL | 1.56 | 0.49 | 0.69 | 0.07 | 2.81 |

STATE: OREGON

| ZIP CODE | QUARTER 1 | QUARTER 2 | QUARTER 3 | QUARTER 4 | TOTAL GRAMS |
|---|---|---|---|---|---|
| 972 | 1.7 | 0.98 | 1.07 | 0.17 | 3.92 |
| 973 | 0.28 | 0 | 0 | 0 | 0.28 |
| TOTAL | 1.98 | 0.98 | 1.07 | 0.17 | 4.2 |

STATE: PENNSYLVANIA

| ZIP CODE | QUARTER 1 | QUARTER 2 | QUARTER 3 | QUARTER 4 | TOTAL GRAMS |
|---|---|---|---|---|---|
| 181 | 0.65 | 3.62 | 0.13 | 0 | 4.4 |
| 185 | 0.39 | 0 | 0 | 0 | 0.39 |
| 190 | 0.1 | 0 | 0 | 0 | 0.1 |
| 191 | 0.57 | 0.28 | 0.28 | 0 | 1.13 |
| TOTAL | 1.71 | 3.9 | 0.41 | 0 | 6.02 |

RETAIL DRUG DISTRIBUTION BY ZIP CODE WITHIN STATE BY GRAMS WT

STATE: PUERTO RICO

| ZIP CODE | QUARTER 1 | QUARTER 2 | QUARTER 3 | QUARTER 4 | TOTAL GRAMS |
|---|---|---|---|---|---|
| 9 | 9.21 | 2.82 | 1.86 | 4.43 | 18.32 |
| TOTAL | 9.21 | 2.82 | 1.86 | 4.43 | 18.32 |

STATE: RHODE ISLAND

| ZIP CODE | QUARTER 1 | QUARTER 2 | QUARTER 3 | QUARTER 4 | TOTAL GRAMS |
|---|---|---|---|---|---|
| 28 | 0.35 | 2.96 | 0.03 | 0 | 3.34 |
| TOTAL | 0.35 | 2.96 | 0.03 | 0 | 3.34 |

STATE: SOUTH CAROLINA

| ZIP CODE | QUARTER 1 | QUARTER 2 | QUARTER 3 | QUARTER 4 | TOTAL GRAMS |
|---|---|---|---|---|---|
| 294 | 0.49 | 0 | 0 | 0 | 0.49 |
| TOTAL | 0.49 | 0 | 0 | 0 | 0.49 |

STATE: TENNESSEE

| ZIP CODE | QUARTER 1 | QUARTER 2 | QUARTER 3 | QUARTER 4 | TOTAL GRAMS |
|---|---|---|---|---|---|
| 381 | 1.26 | 0.58 | 0.74 | 0.45 | 3.03 |
| TOTAL | 1.26 | 0.58 | 0.74 | 0.45 | 3.03 |

STATE: TEXAS

| ZIP CODE | QUARTER 1 | QUARTER 2 | QUARTER 3 | QUARTER 4 | TOTAL GRAMS |
|---|---|---|---|---|---|
| 750 | 0.42 | 0.84 | 1.47 | 0 | 2.73 |
| 751 | 6.35 | 0.22 | 3.08 | 8.45 | 18.1 |
| 752 | 5.09 | 9.45 | 10.03 | 17.99 | 42.56 |
| 753 | 1.19 | 0.74 | 0.78 | 0 | 2.71 |
| 760 | 0.2 | 0 | 0 | 0 | 0.2 |
| 763 | 0.57 | 0.51 | 0.68 | 0.39 | 2.15 |
| 770 | 1.07 | 0.27 | 0.06 | 0 | 1.4 |
| 774 | 0.54 | 0.24 | 0.13 | 0 | 0.91 |
| 787 | 4.5 | 4.44 | 9.27 | 14.87 | 33.08 |
| TOTAL | 19.93 | 16.71 | 25.5 | 41.7 | 103.84 |

STATE: UTAH

| ZIP CODE | QUARTER 1 | QUARTER 2 | QUARTER 3 | QUARTER 4 | TOTAL GRAMS |
|---|---|---|---|---|---|
| 840 | 0.97 | 0.24 | 0.32 | 0 | 1.53 |
| TOTAL | 0.97 | 0.24 | 0.32 | 0 | 1.53 |

STATE: VERMONT

| ZIP CODE | QUARTER 1 | QUARTER 2 | QUARTER 3 | QUARTER 4 | TOTAL GRAMS |
|---|---|---|---|---|---|
| 50 | 0.76 | 0.17 | 0.2 | 0 | 1.13 |
| TOTAL | 0.76 | 0.17 | 0.2 | 0 | 1.13 |

STATE: WASHINGTON

| ZIP CODE | QUARTER 1 | QUARTER 2 | QUARTER 3 | QUARTER 4 | TOTAL GRAMS |
|---|---|---|---|---|---|
| 980 | 6.21 | 1.88 | 1.96 | 3.16 | 13.21 |
| 981 | 1.2 | 0.67 | 0.9 | 0.29 | 3.06 |

DRUG CODE:9668DRUG NAME:NOROXYMORPHONE
STATE:WASHINGTON

| ZIP CODE | QUARTER 1 | QUARTER 2 | QUARTER 3 | QUARTER 4 | TOTAL GRAMS |
|---|---|---|---|---|---|
| 992 | 1.09 | 0.88 | 0.79 | 0.62 | 3.38 |
| TOTAL | 8.5 | 3.43 | 3.65 | 4.07 | 19.65 |

STATE: WISCONSIN

| ZIP CODE | QUARTER 1 | QUARTER 2 | QUARTER 3 | QUARTER 4 | TOTAL GRAMS |
|---|---|---|---|---|---|
| 531 | 0.51 | 0 | 0.03 | 0 | 0.54 |
| 535 | 0.13 | 0 | 0 | 0 | 0.13 |

| | | | | | |
|---|---|---|---|---|---|
| TOTAL | 0.64 | 0 | 0.03 | 0 | 0.67 |

DRUG CODE:9737DRUG NAME:ALFENTANIL

STATE: ALABAMA

| ZIP CODE | QUARTER 1 | QUARTER 2 | QUARTER 3 | QUARTER 4 | TOTAL GRAMS |
|---|---|---|---|---|---|
| 363 | 0.04 | 0.04 | 0.02 | 0.04 | 0.14 |
| 366 | 0.03 | 0.02 | 0.02 | 0 | 0.07 |
| 368 | 0.02 | 0 | 0.02 | 0 | 0.04 |
| TOTAL | 0.09 | 0.06 | 0.06 | 0.04 | 0.25 |

STATE: ALASKA

| ZIP CODE | QUARTER 1 | QUARTER 2 | QUARTER 3 | QUARTER 4 | TOTAL GRAMS |
|---|---|---|---|---|---|
| 995 | 0.88 | 0.67 | 0.66 | 0.88 | 3.09 |
| 996 | 0.04 | 0.03 | 0.04 | 0.07 | 0.18 |
| TOTAL | 0.92 | 0.7 | 0.7 | 0.95 | 3.27 |

STATE: ARIZONA

| ZIP CODE | QUARTER 1 | QUARTER 2 | QUARTER 3 | QUARTER 4 | TOTAL GRAMS |
|---|---|---|---|---|---|
| 850 | 0.17 | 0.06 | 0.03 | 0.21 | 0.47 |
| 851 | 0.02 | 0 | 0.04 | 0 | 0.06 |
| 852 | 0.16 | 0.08 | 0.17 | 0.14 | 0.55 |
| 853 | 0.03 | 0 | 0 | 0 | 0.03 |
| 857 | 0.77 | 0.54 | 0.54 | 0.47 | 2.32 |
| 863 | 0.14 | 0.07 | 0 | 0 | 0.21 |
| TOTAL | 1.29 | 0.75 | 0.78 | 0.82 | 3.64 |

STATE: ARKANSAS

| ZIP CODE | QUARTER 1 | QUARTER 2 | QUARTER 3 | QUARTER 4 | TOTAL GRAMS |
|---|---|---|---|---|---|
| 716 | 0.01 | 0 | 0 | 0 | 0.01 |
| 718 | 0 | 0 | 0 | 0.01 | 0.01 |
| 725 | 0.1 | 0 | 0 | 0 | 0.1 |
| 726 | 0 | 0.02 | 0.01 | 0.01 | 0.04 |
| TOTAL | 0.11 | 0.02 | 0.01 | 0.02 | 0.16 |

STATE: CALIFORNIA

| ZIP CODE | QUARTER 1 | QUARTER 2 | QUARTER 3 | QUARTER 4 | TOTAL GRAMS |
|---|---|---|---|---|---|
| 900 | 1.72 | 1.47 | 1.54 | 2.01 | 6.74 |
| 902 | 0.2 | 0.3 | 0.45 | 0.57 | 1.52 |
| 904 | 0.11 | 0.19 | 0.17 | 0.12 | 0.59 |
| 905 | 0 | 0 | 0.05 | 0 | 0.05 |
| 906 | 0.31 | 0.23 | 0.45 | 0.24 | 1.23 |
| 913 | 0.66 | 0.68 | 0.29 | 0.17 | 1.8 |
| 914 | 0.02 | 0.02 | 0.05 | 0.11 | 0.2 |
| 915 | 0.03 | 0.02 | 0.05 | 0.02 | 0.12 |
| 917 | 1.25 | 1.14 | 1.11 | 0.82 | 4.32 |
| 919 | 0.23 | 0.29 | 0.22 | 0.21 | 0.95 |
| 920 | 1.36 | 0.73 | 0.66 | 0.83 | 3.58 |
| 921 | 1.15 | 0.91 | 1.22 | 1.06 | 4.34 |
| 922 | 0.62 | 0.2 | 0.34 | 0.54 | 1.7 |
| 923 | 1.27 | 2.28 | 1.07 | 1.76 | 6.38 |
| 925 | 0.25 | 0.4 | 0.21 | 0.04 | 0.9 |
| 926 | 0.19 | 0.43 | 0.67 | 0.33 | 1.62 |
| 927 | 0.11 | 0.06 | 0.13 | 0.19 | 0.49 |
| 928 | 0 | 0 | 0.03 | 0 | 0.03 |
| 930 | 0.46 | 0.46 | 0.3 | 0.28 | 1.5 |
| 931 | 0.34 | 0.28 | 0.2 | 0.23 | 1.05 |
| 932 | 0.09 | 0.1 | 0 | 0 | 0.19 |
| 934 | 0.02 | 0.05 | 0.02 | 0.07 | 0.16 |
| 935 | 0.12 | 0.27 | 0.04 | 0.03 | 0.46 |
| 937 | 0.74 | 1.21 | 1.38 | 0.83 | 4.16 |
| 939 | 0 | 0.05 | 0.03 | 0.04 | 0.12 |
| 940 | 0.24 | 0.16 | 0.13 | 0.2 | 0.73 |

| | | | | | |
|---|---|---|---|---|---|
| 941 | 0.4 | 0.05 | 0.04 | 0.08 | 0.57 |
| 943 | 0.72 | 0.52 | 0.52 | 0.96 | 2.72 |
| 944 | 0 | 0.09 | 0.03 | 0.03 | 0.15 |

DRUG CODE:9737DRUG NAME:ALFENTANIL
STATE:CALIFORNIA

| ZIP CODE | QUARTER 1 | QUARTER 2 | QUARTER 3 | QUARTER 4 | TOTAL GRAMS |
|---|---|---|---|---|---|
| 945 | 1.21 | 1.32 | 1.56 | 1.11 | 5.2 |
| 946 | 0.15 | 0.32 | 0.17 | 0.26 | 0.9 |
| 948 | 0 | 0.06 | 0 | 0 | 0.06 |
| 949 | 0.01 | 0.03 | 0.03 | 0.03 | 0.1 |
| 950 | 0.16 | 0.07 | 0 | 0.03 | 0.26 |
| 951 | 0.51 | 0.71 | 0.44 | 0.64 | 2.3 |
| 952 | 0.72 | 0.55 | 0.4 | 0.11 | 1.78 |
| 953 | 1.39 | 0.9 | 1.12 | 0.97 | 4.38 |
| 954 | 0.2 | 0.19 | 0.21 | 0.15 | 0.75 |
| 956 | 0.07 | 0.04 | 0.03 | 0.06 | 0.2 |
| 958 | 0.63 | 0.37 | 0.84 | 0.38 | 2.22 |
| 959 | 0.55 | 0.3 | 0.2 | 0.3 | 1.35 |
| 960 | 0.01 | 0.03 | 0 | 0.02 | 0.06 |
| TOTAL | 18.22 | 17.48 | 16.4 | 15.83 | 67.93 |

STATE: COLORADO

| ZIP CODE | QUARTER 1 | QUARTER 2 | QUARTER 3 | QUARTER 4 | TOTAL GRAMS |
|---|---|---|---|---|---|
| 800 | 0.58 | 0.59 | 0.67 | 0.34 | 2.18 |
| 801 | 0.03 | 0.01 | 0.04 | 0.05 | 0.13 |
| 802 | 0 | 0.01 | 0 | 0 | 0.01 |
| 805 | 0.05 | 0.02 | 0.11 | 0.09 | 0.27 |
| 808 | 0.05 | 0.04 | 0.03 | 0.02 | 0.14 |
| 811 | 0.03 | 0 | 0 | 0.01 | 0.04 |
| 812 | 0 | 0 | 0.04 | 0 | 0.04 |
| 813 | 0 | 0.03 | 0.02 | 0.01 | 0.06 |
| 814 | 0.03 | 0.01 | 0.02 | 0.02 | 0.08 |
| 815 | 0.06 | 0 | 0 | 0 | 0.06 |
| 816 | 0.02 | 0 | 0.04 | 0 | 0.06 |
| TOTAL | 0.85 | 0.71 | 0.97 | 0.54 | 3.07 |

STATE: CONNECTICUT

| ZIP CODE | QUARTER 1 | QUARTER 2 | QUARTER 3 | QUARTER 4 | TOTAL GRAMS |
|---|---|---|---|---|---|
| 64 | 0.06 | 0 | 0 | 0 | 0.06 |
| 65 | 0.65 | 0.97 | 1.11 | 0.68 | 3.41 |
| 68 | 0.33 | 0.36 | 0.4 | 0.29 | 1.38 |
| TOTAL | 1.04 | 1.33 | 1.51 | 0.97 | 4.85 |

STATE: FLORIDA

| ZIP CODE | QUARTER 1 | QUARTER 2 | QUARTER 3 | QUARTER 4 | TOTAL GRAMS |
|---|---|---|---|---|---|
| 322 | 0.18 | 0.28 | 0.53 | 0.02 | 1.01 |
| 324 | 0.42 | 0.35 | 0.58 | 0.59 | 1.94 |
| 325 | 0 | 0 | 0 | 0.01 | 0.01 |
| 326 | 3.57 | 2 | 2.61 | 3.15 | 11.33 |
| 328 | 0.26 | 0.08 | 0 | 0.04 | 0.38 |
| 329 | 0.39 | 0.57 | 0.3 | 0.32 | 1.58 |
| 330 | 0.76 | 0 | 0.02 | 0.01 | 0.79 |
| 331 | 0.26 | 0.3 | 0.2 | 0.16 | 0.92 |
| 334 | 0.07 | 0.76 | 0.3 | 0.56 | 1.69 |
| 336 | 0 | 0 | 0.02 | 0 | 0.02 |
| 337 | 0 | 0 | 0 | 0.03 | 0.03 |
| 342 | 0 | 0 | 0 | 0.02 | 0.02 |
| 344 | 0 | 0 | 0 | 0.01 | 0.01 |
| 349 | 0.09 | 0 | 0 | 0 | 0.09 |

| | | | | | |
|---|---|---|---|---|---|
| TOTAL | 6 | 4.34 | 4.56 | 4.92 | 19.82 |

STATE: GEORGIA

| ZIP CODE | QUARTER 1 | QUARTER 2 | QUARTER 3 | QUARTER 4 | TOTAL GRAMS |
|---|---|---|---|---|---|
| 300 | 0 | 0.08 | 0 | 0 | 0.08 |
| 301 | 0.01 | 0.01 | 0.01 | 0.02 | 0.05 |
| 303 | 0.99 | 0.31 | 0.58 | 0.46 | 2.34 |
| 308 | 0.1 | 0.07 | 0.1 | 0.06 | 0.33 |
| 309 | 0.79 | 0.91 | 0.95 | 0.86 | 3.51 |
| 312 | 0 | 0 | 0 | 0.28 | 0.28 |
| 315 | 0.2 | 0.11 | 0.16 | 0.2 | 0.67 |
| 317 | 0.4 | 0.21 | 0.31 | 0.28 | 1.2 |
| 319 | 0.13 | 0 | 0.04 | 0.1 | 0.27 |
| TOTAL | 2.62 | 1.7 | 2.15 | 2.26 | 8.73 |

STATE: GUAM

| ZIP CODE | QUARTER 1 | QUARTER 2 | QUARTER 3 | QUARTER 4 | TOTAL GRAMS |
|---|---|---|---|---|---|
| 969 | 0.05 | 0.06 | 0.1 | 0.03 | 0.24 |

DRUG CODE:9737DRUG NAME:ALFENTANIL
STATE:GUAM

| ZIP CODE | QUARTER 1 | QUARTER 2 | QUARTER 3 | QUARTER 4 | TOTAL GRAMS |
|---|---|---|---|---|---|
| TOTAL | 0.05 | 0.06 | 0.1 | 0.03 | 0.24 |

STATE: HAWAII

| ZIP CODE | QUARTER 1 | QUARTER 2 | QUARTER 3 | QUARTER 4 | TOTAL GRAMS |
|---|---|---|---|---|---|
| 967 | 0.23 | 0.28 | 0.15 | 0.38 | 1.04 |
| 968 | 0.15 | 0.21 | 0.1 | 0.3 | 0.76 |
| TOTAL | 0.38 | 0.49 | 0.25 | 0.68 | 1.8 |

STATE: IDAHO

| ZIP CODE | QUARTER 1 | QUARTER 2 | QUARTER 3 | QUARTER 4 | TOTAL GRAMS |
|---|---|---|---|---|---|
| 833 | 0.1 | 0.08 | 0.05 | 0.1 | 0.33 |
| 836 | 0 | 0.05 | 0.03 | 0.03 | 0.11 |
| 837 | 1.14 | 1.1 | 1.12 | 1.54 | 4.9 |
| TOTAL | 1.24 | 1.23 | 1.2 | 1.67 | 5.34 |

STATE: ILLINOIS

| ZIP CODE | QUARTER 1 | QUARTER 2 | QUARTER 3 | QUARTER 4 | TOTAL GRAMS |
|---|---|---|---|---|---|
| 600 | 0.37 | 0.18 | 0.16 | 0.32 | 1.03 |
| 601 | 0.8 | 1.18 | 0.85 | 1.35 | 4.18 |
| 604 | 0.13 | 0 | 0 | 0 | 0.13 |
| 605 | 0.05 | 0.1 | 0.03 | 0.08 | 0.26 |
| 606 | 0.68 | 1.1 | 0.33 | 0.94 | 3.05 |
| 608 | 0.16 | 0.18 | 0.12 | 0 | 0.46 |
| 610 | 0.01 | 0.5 | 0.6 | 0.4 | 1.51 |
| 611 | 1.64 | 0.95 | 0.85 | 0.75 | 4.19 |
| 612 | 0.02 | 0.02 | 0.01 | 0.02 | 0.07 |
| 619 | 0 | 0 | 0 | 0.01 | 0.01 |
| 622 | 0.13 | 0.13 | 0.07 | 0.01 | 0.34 |
| 624 | 0.02 | 0.01 | 0.02 | 0.05 | 0.1 |
| 627 | 0.09 | 0.08 | 0.06 | 0.07 | 0.3 |
| 628 | 0.04 | 0.03 | 0.11 | 0.19 | 0.37 |
| 629 | 0 | 0.05 | 0.02 | 0.03 | 0.1 |
| TOTAL | 4.14 | 4.51 | 3.23 | 4.22 | 16.1 |

STATE: INDIANA

| ZIP CODE | QUARTER 1 | QUARTER 2 | QUARTER 3 | QUARTER 4 | TOTAL GRAMS |
|---|---|---|---|---|---|
| 460 | 0.77 | 0.64 | 0.58 | 0.81 | 2.8 |

| | | | | | |
|---|---|---|---|---|---|
| 461 | 0.07 | 0 | 0 | 0 | 0.07 |
| 462 | 0.52 | 0.5 | 0.54 | 0.49 | 2.05 |
| 464 | 0 | 0.01 | 0 | 0 | 0.01 |
| 465 | 0.03 | 0 | 0 | 0 | 0.03 |
| 466 | 0.63 | 0.73 | 0.54 | 0.57 | 2.47 |
| 467 | 0 | 0.01 | 0.01 | 0.03 | 0.05 |
| 469 | 0.03 | 0.01 | 0.02 | 0.03 | 0.09 |
| 470 | 0 | 0.01 | 0.02 | 0.01 | 0.04 |
| 471 | 0 | 0.01 | 0 | 0 | 0.01 |
| 472 | 0.06 | 0.06 | 0.13 | 0.13 | 0.38 |
| 473 | 0.19 | 0.13 | 0.32 | 0 | 0.64 |
| 475 | 0 | 0 | 0 | 0.01 | 0.01 |
| 477 | 0.15 | 0.1 | 0.08 | 0.03 | 0.36 |
| 479 | 0.09 | 0.07 | 0.1 | 0.09 | 0.35 |
| TOTAL | 2.54 | 2.28 | 2.34 | 2.2 | 9.36 |

STATE: IOWA

| ZIP CODE | QUARTER 1 | QUARTER 2 | QUARTER 3 | QUARTER 4 | TOTAL GRAMS |
|---|---|---|---|---|---|
| 500 | 0.03 | 0.03 | 0.05 | 0.03 | 0.14 |
| 505 | 0 | 0.02 | 0.01 | 0.04 | 0.07 |
| 506 | 0.13 | 0 | 0 | 0.1 | 0.23 |
| 507 | 0.04 | 0 | 0.04 | 0 | 0.08 |
| 511 | 0 | 0 | 0.05 | 0.03 | 0.08 |
| 514 | 0.02 | 0 | 0 | 0 | 0.02 |
| 520 | 0 | 0.2 | 0.01 | 0 | 0.21 |
| 522 | 0.95 | 0.81 | 1.01 | 1.25 | 4.02 |
| 524 | 0.18 | 0.17 | 0.2 | 0.27 | 0.82 |
| 526 | 0.05 | 0.05 | 0.05 | 0.1 | 0.25 |
| 527 | 0.2 | 0.01 | 0.14 | 0.11 | 0.46 |
| 528 | 0.08 | 0.2 | 0.25 | 0.22 | 0.75 |
| TOTAL | 1.68 | 1.49 | 1.81 | 2.15 | 7.13 |

STATE: KANSAS

| ZIP CODE | QUARTER 1 | QUARTER 2 | QUARTER 3 | QUARTER 4 | TOTAL GRAMS |
|---|---|---|---|---|---|
| 662 | 0 | 0.05 | 0 | 0 | 0.05 |

DRUG CODE:9737DRUG NAME:ALFENTANIL
STATE:KANSAS

| ZIP CODE | QUARTER 1 | QUARTER 2 | QUARTER 3 | QUARTER 4 | TOTAL GRAMS |
|---|---|---|---|---|---|
| 669 | 0.09 | 0.15 | 0.05 | 0.05 | 0.34 |
| 671 | 0.14 | 0.1 | 0.05 | 0.02 | 0.31 |
| 674 | 0.9 | 1.1 | 1.08 | 0.97 | 4.05 |
| 675 | 0.1 | 0.27 | 0.24 | 0.3 | 0.91 |
| 676 | 0 | 0 | 0 | 0.03 | 0.03 |
| 678 | 0.29 | 0.22 | 0.33 | 0.35 | 1.19 |
| TOTAL | 1.52 | 1.89 | 1.75 | 1.72 | 6.88 |

STATE: KENTUCKY

| ZIP CODE | QUARTER 1 | QUARTER 2 | QUARTER 3 | QUARTER 4 | TOTAL GRAMS |
|---|---|---|---|---|---|
| 402 | 0.15 | 0.24 | 0.24 | 0.28 | 0.91 |
| 404 | 0.04 | 0.04 | 0.03 | 0.04 | 0.15 |
| 408 | 0.01 | 0 | 0 | 0 | 0.01 |
| 410 | 1.25 | 1.25 | 2.88 | 2.25 | 7.63 |
| 411 | 0.03 | 0.04 | 0.06 | 0.04 | 0.17 |
| 413 | 0 | 0 | 0 | 0.01 | 0.01 |
| 423 | 0.08 | 0.1 | 0 | 0 | 0.18 |
| TOTAL | 1.56 | 1.67 | 3.21 | 2.62 | 9.06 |

RETAIL DRUG DISTRIBUTION BY ZIP CODE WITHIN STATE BY GRAMS WT

STATE: LOUISIANA

| ZIP CODE | QUARTER 1 | QUARTER 2 | QUARTER 3 | QUARTER 4 | TOTAL GRAMS |
|---|---|---|---|---|---|
| 700 | 0 | 0.01 | 0.01 | 0 | 0.02 |
| 701 | 0.15 | 0.18 | 0.25 | 0.33 | 0.91 |
| 704 | 0.41 | 0.7 | 0.73 | 0.45 | 2.29 |
| 705 | 0.13 | 0.3 | 0.2 | 0.2 | 0.83 |
| 708 | 0.2 | 0.15 | 0.19 | 0.1 | 0.64 |
| 710 | 0.01 | 0.05 | 0 | 0.03 | 0.09 |
| 711 | 1.03 | 0.79 | 0.82 | 1.11 | 3.75 |
| 712 | 0.04 | 0.03 | 0.02 | 0.02 | 0.11 |
| 713 | 0.14 | 0.13 | 0.23 | 0.09 | 0.59 |
| 714 | 0.02 | 0 | 0.05 | 0 | 0.07 |
| TOTAL | 2.13 | 2.34 | 2.5 | 2.33 | 9.3 |

STATE: MAINE

| ZIP CODE | QUARTER 1 | QUARTER 2 | QUARTER 3 | QUARTER 4 | TOTAL GRAMS |
|---|---|---|---|---|---|
| 39 | 0.08 | 0.17 | 0.13 | 0.17 | 0.55 |
| 40 | 0.03 | 0.17 | 0.18 | 0.18 | 0.56 |
| 41 | 0.31 | 0.29 | 0.17 | 0.12 | 0.89 |
| 42 | 0 | 0.01 | 0.01 | 0 | 0.02 |
| 44 | 0.1 | 0.07 | 0.1 | 0.25 | 0.52 |
| 46 | 0.04 | 0.03 | 0 | 0.02 | 0.09 |
| TOTAL | 0.56 | 0.74 | 0.59 | 0.74 | 2.63 |

STATE: MARYLAND

| ZIP CODE | QUARTER 1 | QUARTER 2 | QUARTER 3 | QUARTER 4 | TOTAL GRAMS |
|---|---|---|---|---|---|
| 207 | 0.03 | 0 | 0 | 0 | 0.03 |
| 210 | 0.16 | 0.06 | 0.28 | 0.2 | 0.7 |
| 212 | 0.7 | 0.26 | 0.18 | 0.43 | 1.57 |
| 216 | 0 | 0.01 | 0 | 0 | 0.01 |
| 217 | 0 | 0 | 0 | 0.01 | 0.01 |
| TOTAL | 0.89 | 0.33 | 0.46 | 0.64 | 2.32 |

STATE: MASSACHUSETTS

| ZIP CODE | QUARTER 1 | QUARTER 2 | QUARTER 3 | QUARTER 4 | TOTAL GRAMS |
|---|---|---|---|---|---|
| 19 | 0.02 | 0 | 0 | 0 | 0.02 |
| 21 | 0.16 | 0.2 | 0.08 | 0.1 | 0.54 |
| 24 | 1.25 | 1.23 | 0.85 | 1.38 | 4.71 |
| TOTAL | 1.43 | 1.43 | 0.93 | 1.48 | 5.27 |

STATE: MICHIGAN

| ZIP CODE | QUARTER 1 | QUARTER 2 | QUARTER 3 | QUARTER 4 | TOTAL GRAMS |
|---|---|---|---|---|---|
| 481 | 0.1 | 0.16 | 0.06 | 0.25 | 0.57 |
| 483 | 0.08 | 0.04 | 0.11 | 0.16 | 0.39 |
| 485 | 0 | 0 | 0.04 | 0 | 0.04 |
| 486 | 0.05 | 0 | 0.06 | 0.05 | 0.16 |
| 488 | 0.86 | 0.49 | 0.58 | 0.6 | 2.53 |
| 489 | 0.05 | 0.04 | 0 | 0.02 | 0.11 |
| 490 | 0.15 | 0.2 | 0.18 | 0.33 | 0.86 |
| 492 | 0 | 0.03 | 0.02 | 0.05 | 0.1 |
| 497 | 0.02 | 0 | 0.05 | 0.02 | 0.09 |
| 498 | 0.1 | 0.05 | 0.07 | 0.05 | 0.27 |
| TOTAL | 1.41 | 1.01 | 1.17 | 1.53 | 5.12 |

DRUG CODE:9737DRUG NAME:ALFENTANIL

STATE: MINNESOTA

| ZIP CODE | QUARTER 1 | QUARTER 2 | QUARTER 3 | QUARTER 4 | TOTAL GRAMS |
|---|---|---|---|---|---|
| 550 | 0.07 | 0.09 | 0.08 | 0.12 | 0.36 |
| 551 | 0.84 | 0.86 | 0.64 | 0.88 | 3.22 |
| 553 | 0.02 | 0.01 | 0 | 0 | 0.03 |
| 554 | 2.33 | 2.28 | 2.09 | 2.1 | 8.8 |
| 559 | 0.05 | 0.05 | 0.08 | 0.08 | 0.26 |
| 561 | 0.03 | 0 | 0 | 0 | 0.03 |
| 562 | 0.32 | 0.19 | 0.14 | 0.23 | 0.88 |
| 565 | 0.01 | 0 | 0.01 | 0.02 | 0.04 |
| TOTAL | 3.67 | 3.48 | 3.04 | 3.43 | 13.62 |

STATE: MISSISSIPPI

| ZIP CODE | QUARTER 1 | QUARTER 2 | QUARTER 3 | QUARTER 4 | TOTAL GRAMS |
|---|---|---|---|---|---|
| 389 | 0 | 0.01 | 0.01 | 0 | 0.02 |
| 390 | 0.06 | 0.06 | 0.09 | 0.09 | 0.3 |
| 393 | 0.05 | 0.03 | 0.04 | 0.04 | 0.16 |
| 394 | 0 | 0.01 | 0 | 0.06 | 0.07 |
| 396 | 0 | 0 | 0.03 | 0 | 0.03 |
| TOTAL | 0.11 | 0.11 | 0.17 | 0.19 | 0.58 |

STATE: MISSOURI

| ZIP CODE | QUARTER 1 | QUARTER 2 | QUARTER 3 | QUARTER 4 | TOTAL GRAMS |
|---|---|---|---|---|---|
| 631 | 0.48 | 0.4 | 0.36 | 0.06 | 1.3 |
| 636 | 0.32 | 0.31 | 0.28 | 0.3 | 1.21 |
| 640 | 0.04 | 0.05 | 0.04 | 0.05 | 0.18 |
| 641 | 0 | 0.03 | 0 | 0 | 0.03 |
| 646 | 0.07 | 0.12 | 0.02 | 0.02 | 0.23 |
| 652 | 0.02 | 0.04 | 0.04 | 0.01 | 0.11 |
| 654 | 0.46 | 0.42 | 0.38 | 0.26 | 1.52 |
| 657 | 0.17 | 0.25 | 0.13 | 0.18 | 0.73 |
| TOTAL | 1.56 | 1.62 | 1.25 | 0.88 | 5.31 |

STATE: MONTANA

| ZIP CODE | QUARTER 1 | QUARTER 2 | QUARTER 3 | QUARTER 4 | TOTAL GRAMS |
|---|---|---|---|---|---|
| 590 | 0.14 | 0.08 | 0.14 | 0.1 | 0.46 |
| 592 | 0.04 | 0.04 | 0.05 | 0.06 | 0.19 |
| 596 | 0.03 | 0 | 0 | 0 | 0.03 |
| 597 | 0.15 | 0.11 | 0.16 | 0.1 | 0.52 |
| 598 | 0.2 | 0.05 | 0.07 | 0.05 | 0.37 |
| TOTAL | 0.56 | 0.28 | 0.42 | 0.31 | 1.57 |

STATE: NEBRASKA

| ZIP CODE | QUARTER 1 | QUARTER 2 | QUARTER 3 | QUARTER 4 | TOTAL GRAMS |
|---|---|---|---|---|---|
| 681 | 0.24 | 0.34 | 0.05 | 0.14 | 0.77 |
| 683 | 0.06 | 0.12 | 0.09 | 0.03 | 0.3 |
| 684 | 0.02 | 0.04 | 0.03 | 0.06 | 0.15 |
| 685 | 0 | 0.05 | 0 | 0.01 | 0.06 |
| 686 | 0.01 | 0 | 0 | 0 | 0.01 |
| 687 | 0.2 | 0.19 | 0.15 | 0.09 | 0.63 |
| 688 | 0.1 | 0.12 | 0.1 | 0.12 | 0.44 |
| 689 | 0.37 | 0.29 | 0.43 | 0.35 | 1.44 |
| 691 | 0.03 | 0.08 | 0.08 | 0.02 | 0.21 |
| 693 | 0.08 | 0 | 0.03 | 0 | 0.11 |
| TOTAL | 1.11 | 1.23 | 0.96 | 0.82 | 4.12 |

STATE: NEVADA

| ZIP CODE | QUARTER 1 | QUARTER 2 | QUARTER 3 | QUARTER 4 | TOTAL GRAMS |
|---|---|---|---|---|---|
| 890 | 0.08 | 0.08 | 0.01 | 0.2 | 0.37 |
| 891 | 1.19 | 1.35 | 0.89 | 0.8 | 4.23 |
| 894 | 0.06 | 0.09 | 0.07 | 0.07 | 0.29 |
| 895 | 0.52 | 0.55 | 0.52 | 0.38 | 1.97 |

| | | | | | |
|---|---|---|---|---|---|
| 898 | 0 | 0.15 | 0.05 | 0.03 | 0.23 |
| TOTAL | 1.85 | 2.22 | 1.54 | 1.48 | 7.09 |

STATE: NEW HAMPSHIRE

| ZIP CODE | QUARTER 1 | QUARTER 2 | QUARTER 3 | QUARTER 4 | TOTAL GRAMS |
|---|---|---|---|---|---|
| 30 | 0 | 0.01 | 0 | 0 | 0.01 |
| 32 | 0 | 0 | 0.03 | 0.01 | 0.04 |
| 38 | 0.4 | 0.36 | 0.39 | 0.37 | 1.52 |
| TOTAL | 0.4 | 0.37 | 0.42 | 0.38 | 1.57 |

STATE: NEW JERSEY

| ZIP CODE | QUARTER 1 | QUARTER 2 | QUARTER 3 | QUARTER 4 | TOTAL GRAMS |
|---|---|---|---|---|---|
| 70 | 0.03 | 0.06 | 0.05 | 0.05 | 0.19 |

DRUG CODE:9737DRUG NAME:ALFENTANIL
STATE:NEW JERSEY

| ZIP CODE | QUARTER 1 | QUARTER 2 | QUARTER 3 | QUARTER 4 | TOTAL GRAMS |
|---|---|---|---|---|---|
| 71 | 0 | 0.02 | 0 | 0 | 0.02 |
| 74 | 0.03 | 0.02 | 0.02 | 0.03 | 0.1 |
| 75 | 0 | 0 | 0 | 0.1 | 0.1 |
| 77 | 0.04 | 0.09 | 0.02 | 0.03 | 0.18 |
| 86 | 0 | 0 | 0.04 | 0 | 0.04 |
| 88 | 0.1 | 0.18 | 0.08 | 0.13 | 0.49 |
| TOTAL | 0.2 | 0.37 | 0.21 | 0.34 | 1.12 |

STATE: NEW MEXICO

| ZIP CODE | QUARTER 1 | QUARTER 2 | QUARTER 3 | QUARTER 4 | TOTAL GRAMS |
|---|---|---|---|---|---|
| 871 | 1.24 | 1.01 | 0.94 | 1.5 | 4.69 |
| 875 | 0 | 0.03 | 0 | 0.06 | 0.09 |
| TOTAL | 1.24 | 1.04 | 0.94 | 1.56 | 4.78 |

STATE: NEW YORK

| ZIP CODE | QUARTER 1 | QUARTER 2 | QUARTER 3 | QUARTER 4 | TOTAL GRAMS |
|---|---|---|---|---|---|
| 100 | 0.29 | 0.01 | 0.12 | 0 | 0.42 |
| 105 | 0.12 | 0.1 | 0.05 | 0.01 | 0.28 |
| 112 | 1.44 | 0.46 | 1.05 | 0.94 | 3.89 |
| 117 | 0.1 | 0.01 | 0.02 | 0.01 | 0.14 |
| 119 | 0 | 0.01 | 0 | 0 | 0.01 |
| 122 | 0.29 | 0.23 | 0.19 | 0.3 | 1.01 |
| 123 | 0 | 0.08 | 0 | 0.05 | 0.13 |
| 129 | 0 | 0 | 0.04 | 0 | 0.04 |
| 131 | 0 | 0.01 | 0.01 | 0 | 0.02 |
| 135 | 0 | 0 | 0.02 | 0 | 0.02 |
| 136 | 0.02 | 0 | 0.02 | 0.02 | 0.06 |
| 138 | 0.04 | 0.16 | 0 | 0 | 0.2 |
| 140 | 0 | 0.01 | 0.04 | 0.01 | 0.06 |
| 141 | 0.01 | 0.01 | 0.01 | 0 | 0.03 |
| 142 | 0.09 | 0.07 | 0.15 | 0.04 | 0.35 |
| 146 | 0.99 | 1.01 | 1.07 | 1.05 | 4.12 |
| 148 | 0.05 | 0.13 | 0.15 | 0.14 | 0.47 |
| TOTAL | 3.44 | 2.3 | 2.94 | 2.57 | 11.25 |

STATE: NORTH CAROLINA

| ZIP CODE | QUARTER 1 | QUARTER 2 | QUARTER 3 | QUARTER 4 | TOTAL GRAMS |
|---|---|---|---|---|---|
| 270 | 0.15 | 0.17 | 0.16 | 0.08 | 0.56 |
| 271 | 0.8 | 0.48 | 0.69 | 0.53 | 2.5 |
| 272 | 0.08 | 0.06 | 0.1 | 0.01 | 0.25 |
| 273 | 0.02 | 0.06 | 0 | 0.01 | 0.09 |

| | | | | | |
|---|---|---|---|---|---|
| 274 | 0.39 | 0.45 | 0.39 | 0.45 | 1.68 |
| 276 | 0.07 | 0.08 | 0.11 | 0.21 | 0.47 |
| 277 | 0.05 | 0.03 | 0.02 | 0.02 | 0.12 |
| 278 | 0 | 0.03 | 0 | 0.02 | 0.05 |
| 280 | 0.04 | 0 | 0.02 | 0 | 0.06 |
| 281 | 0 | 0.12 | 0 | 0.05 | 0.17 |
| 282 | 0.98 | 0.87 | 0.99 | 0.96 | 3.8 |
| 283 | 0.3 | 0.22 | 0.21 | 0.14 | 0.87 |
| 286 | 0.33 | 0.18 | 0.36 | 0.35 | 1.22 |
| TOTAL | 3.21 | 2.75 | 3.05 | 2.83 | 11.84 |

STATE: NORTH DAKOTA

| ZIP CODE | QUARTER 1 | QUARTER 2 | QUARTER 3 | QUARTER 4 | TOTAL GRAMS |
|---|---|---|---|---|---|
| 582 | 0 | 0 | 0.02 | 0.03 | 0.05 |
| 584 | 0.05 | 0.04 | 0.04 | 0.03 | 0.16 |
| 585 | 0.01 | 0 | 0 | 0 | 0.01 |
| TOTAL | 0.06 | 0.04 | 0.06 | 0.06 | 0.22 |

STATE: OHIO

| ZIP CODE | QUARTER 1 | QUARTER 2 | QUARTER 3 | QUARTER 4 | TOTAL GRAMS |
|---|---|---|---|---|---|
| 430 | 0 | 0.13 | 0 | 0.13 | 0.26 |
| 433 | 0.03 | 0.03 | 0.03 | 0.04 | 0.13 |
| 435 | 0.05 | 0.04 | 0.02 | 0.03 | 0.14 |
| 440 | 0.26 | 0.33 | 0.19 | 0.42 | 1.2 |
| 441 | 2.52 | 2.78 | 2.7 | 3.11 | 11.11 |
| 442 | 0.14 | 0.04 | 0.09 | 0.09 | 0.36 |
| 443 | 0.06 | 0.03 | 0.07 | 0.05 | 0.21 |
| 444 | 0.31 | 0.26 | 0.28 | 0.28 | 1.13 |
| 445 | 1.26 | 0.62 | 0.86 | 0.79 | 3.53 |
| 446 | 0.13 | 0.05 | 0.06 | 0.22 | 0.46 |

DRUG CODE:9737DRUG NAME:ALFENTANIL
STATE:OHIO

| ZIP CODE | QUARTER 1 | QUARTER 2 | QUARTER 3 | QUARTER 4 | TOTAL GRAMS |
|---|---|---|---|---|---|
| 448 | 0.02 | 0.11 | 0.02 | 0.04 | 0.19 |
| 452 | 0.01 | 0 | 0 | 0.08 | 0.09 |
| 454 | 0.44 | 0.65 | 0.53 | 0.65 | 2.27 |
| 457 | 0 | 0 | 0 | 0.03 | 0.03 |
| TOTAL | 5.23 | 5.07 | 4.85 | 5.96 | 21.11 |

STATE: OKLAHOMA

| ZIP CODE | QUARTER 1 | QUARTER 2 | QUARTER 3 | QUARTER 4 | TOTAL GRAMS |
|---|---|---|---|---|---|
| 730 | 0 | 0.01 | 0 | 0 | 0.01 |
| 731 | 0.15 | 0 | 0.15 | 0.06 | 0.36 |
| 740 | 0.07 | 0 | 0.05 | 0 | 0.12 |
| 741 | 0 | 0.12 | 0.16 | 0.22 | 0.5 |
| 744 | 0.2 | 0.54 | 0.3 | 0.25 | 1.29 |
| 748 | 1.01 | 0.2 | 0.24 | 0.35 | 1.8 |
| TOTAL | 1.43 | 0.87 | 0.9 | 0.88 | 4.08 |

STATE: OREGON

| ZIP CODE | QUARTER 1 | QUARTER 2 | QUARTER 3 | QUARTER 4 | TOTAL GRAMS |
|---|---|---|---|---|---|
| 970 | 0.38 | 0.33 | 0.39 | 0.61 | 1.71 |
| 971 | 0.04 | 0.02 | 0.06 | 0.07 | 0.19 |
| 972 | 2.61 | 2.54 | 2.94 | 2.54 | 10.63 |
| 974 | 0 | 0 | 0.07 | 0.01 | 0.08 |
| 976 | 0.64 | 0.8 | 0.68 | 0.73 | 2.85 |
| 977 | 0.12 | 0.02 | 0.02 | 0.12 | 0.28 |
| 978 | 0 | 0.02 | 0.03 | 0 | 0.05 |

RETAIL DRUG DISTRIBUTION BY ZIP CODE WITHIN STATE BY GRAMS WT

| | | | | | |
|---|---|---|---|---|---|
| TOTAL | 3.79 | 3.73 | 4.19 | 4.08 | 15.79 |

STATE: PENNSYLVANIA

| ZIP CODE | QUARTER 1 | QUARTER 2 | QUARTER 3 | QUARTER 4 | TOTAL GRAMS |
|---|---|---|---|---|---|
| 161 | 0.01 | 0.02 | 0 | 0 | 0.03 |
| 163 | 0 | 0.02 | 0.09 | 0.02 | 0.13 |
| 165 | 0.45 | 0.14 | 0.15 | 0.05 | 0.79 |
| 170 | 0.16 | 0.12 | 0.2 | 0.21 | 0.69 |
| 171 | 0.1 | 0.04 | 0.16 | 0 | 0.3 |
| 173 | 0.01 | 0 | 0 | 0 | 0.01 |
| 174 | 0.04 | 0.11 | 0.12 | 0.07 | 0.34 |
| 181 | 0.2 | 0.1 | 0 | 0.1 | 0.4 |
| TOTAL | 0.97 | 0.55 | 0.72 | 0.45 | 2.69 |

STATE: RHODE ISLAND

| ZIP CODE | QUARTER 1 | QUARTER 2 | QUARTER 3 | QUARTER 4 | TOTAL GRAMS |
|---|---|---|---|---|---|
| 28 | 0.1 | 0.06 | 0.08 | 0.05 | 0.29 |
| TOTAL | 0.1 | 0.06 | 0.08 | 0.05 | 0.29 |

STATE: SOUTH CAROLINA

| ZIP CODE | QUARTER 1 | QUARTER 2 | QUARTER 3 | QUARTER 4 | TOTAL GRAMS |
|---|---|---|---|---|---|
| 293 | 0.08 | 0 | 0.05 | 0.06 | 0.19 |
| TOTAL | 0.08 | 0 | 0.05 | 0.06 | 0.19 |

STATE: SOUTH DAKOTA

| ZIP CODE | QUARTER 1 | QUARTER 2 | QUARTER 3 | QUARTER 4 | TOTAL GRAMS |
|---|---|---|---|---|---|
| 570 | 0.06 | 0.05 | 0.06 | 0 | 0.17 |
| 572 | 0 | 0.01 | 0 | 0.05 | 0.06 |
| TOTAL | 0.06 | 0.06 | 0.06 | 0.05 | 0.23 |

STATE: TENNESSEE

| ZIP CODE | QUARTER 1 | QUARTER 2 | QUARTER 3 | QUARTER 4 | TOTAL GRAMS |
|---|---|---|---|---|---|
| 371 | 0.77 | 1 | 1.02 | 0.91 | 3.7 |
| 376 | 0 | 0.02 | 0.05 | 0.01 | 0.08 |
| 382 | 0.05 | 0.02 | 0.06 | 0.02 | 0.15 |
| 384 | 0 | 0 | 0.01 | 0 | 0.01 |
| TOTAL | 0.82 | 1.04 | 1.14 | 0.94 | 3.94 |

STATE: TEXAS

| ZIP CODE | QUARTER 1 | QUARTER 2 | QUARTER 3 | QUARTER 4 | TOTAL GRAMS |
|---|---|---|---|---|---|
| 750 | 0.06 | 0.1 | 0.04 | 0.02 | 0.22 |
| 751 | 0.25 | 0.08 | 0.21 | 0.06 | 0.6 |
| 752 | 0.11 | 0.08 | 0.15 | 0.21 | 0.55 |
| 753 | 0 | 0.01 | 0 | 0.01 | 0.02 |
| 760 | 0.25 | 0 | 0.25 | 0 | 0.5 |
| 761 | 0 | 0.02 | 0.02 | 0 | 0.04 |
| 762 | 0 | 0.03 | 0 | 0 | 0.03 |
| 770 | 0.75 | 0.61 | 0.57 | 0.61 | 2.54 |
| 773 | 0.03 | 0.18 | 0 | 0 | 0.21 |

DRUG CODE:9737DRUG NAME:ALFENTANIL
STATE:TEXAS

| ZIP CODE | QUARTER 1 | QUARTER 2 | QUARTER 3 | QUARTER 4 | TOTAL GRAMS |
|---|---|---|---|---|---|
| 774 | 0 | 0 | 0.05 | 0.04 | 0.09 |
| 775 | 0.27 | 0.25 | 0.05 | 0.1 | 0.67 |
| 777 | 0.04 | 0 | 0.02 | 0.01 | 0.07 |
| 778 | 0.02 | 0.01 | 0 | 0.01 | 0.04 |
| 779 | 0 | 0 | 0.08 | 0.03 | 0.11 |
| 782 | 1.74 | 1.51 | 1.75 | 1.71 | 6.71 |
| 785 | 0 | 0 | 0.03 | 0.03 | 0.06 |

| | | | | | |
|---|---|---|---|---|---|
| 786 | 0 | 0 | 0.01 | 0.01 | 0.02 |
| 787 | 0 | 0 | 0.02 | 0.03 | 0.05 |
| 788 | 0 | 0 | 0.01 | 0.03 | 0.04 |
| 797 | 0.02 | 0.01 | 0 | 0.04 | 0.07 |
| 799 | 3.48 | 2.55 | 2.56 | 2.76 | 11.35 |
| TOTAL | 7.02 | 5.44 | 5.82 | 5.71 | 23.99 |
| STATE: | UTAH | | | | |
| ZIP CODE | QUARTER 1 | QUARTER 2 | QUARTER 3 | QUARTER 4 | TOTAL GRAMS |
| 841 | 0 | 0 | 0 | 0.03 | 0.03 |
| 844 | 0.5 | 0.43 | 0.31 | 0.43 | 1.67 |
| 845 | 0 | 0.03 | 0 | 0.02 | 0.05 |
| 847 | 0.03 | 0.07 | 0.02 | 0.01 | 0.13 |
| TOTAL | 0.53 | 0.53 | 0.33 | 0.49 | 1.88 |
| STATE: | VERMONT | | | | |
| ZIP CODE | QUARTER 1 | QUARTER 2 | QUARTER 3 | QUARTER 4 | TOTAL GRAMS |
| 56 | 0 | 0 | 0.02 | 0 | 0.02 |
| TOTAL | 0 | 0 | 0.02 | 0 | 0.02 |
| STATE: | VIRGINIA | | | | |
| ZIP CODE | QUARTER 1 | QUARTER 2 | QUARTER 3 | QUARTER 4 | TOTAL GRAMS |
| 201 | 0 | 0 | 0.13 | 0.01 | 0.14 |
| 229 | 0.04 | 0.02 | 0.03 | 0.03 | 0.12 |
| 231 | 0.03 | 0 | 0 | 0 | 0.03 |
| 232 | 0.3 | 0.41 | 0.35 | 0.51 | 1.57 |
| 234 | 0.2 | 0.18 | 0.18 | 0.18 | 0.74 |
| 236 | 0.03 | 0.01 | 0 | 0.01 | 0.05 |
| 237 | 0 | 0 | 0.01 | 0 | 0.01 |
| 240 | 0.09 | 0.16 | 0.08 | 0.21 | 0.54 |
| 242 | 0 | 0 | 0.1 | 0.3 | 0.4 |
| 244 | 0 | 0 | 0.02 | 0 | 0.02 |
| 246 | 0 | 0 | 0.01 | 0 | 0.01 |
| TOTAL | 0.69 | 0.78 | 0.91 | 1.25 | 3.63 |
| STATE: | WASHINGTON | | | | |
| ZIP CODE | QUARTER 1 | QUARTER 2 | QUARTER 3 | QUARTER 4 | TOTAL GRAMS |
| 980 | 0.06 | 0.12 | 0.16 | 0.08 | 0.42 |
| 981 | 2.67 | 1.67 | 1.83 | 2.12 | 8.29 |
| 982 | 0.22 | 0.27 | 0.15 | 0.3 | 0.94 |
| 983 | 0.22 | 0.27 | 0.24 | 0.22 | 0.95 |
| 984 | 0.12 | 0.09 | 0.18 | 0.2 | 0.59 |
| 985 | 0 | 0.01 | 0.01 | 0 | 0.02 |
| 986 | 0.05 | 0 | 0.03 | 0.01 | 0.09 |
| 988 | 0.01 | 0.02 | 0.04 | 0.04 | 0.11 |
| 989 | 0.02 | 0.01 | 0.03 | 0.01 | 0.07 |
| 991 | 0.02 | 0.02 | 0.01 | 0.01 | 0.06 |
| 992 | 0 | 0.04 | 0 | 0.05 | 0.09 |
| 993 | 0.03 | 0.01 | 0.01 | 0.02 | 0.07 |
| TOTAL | 3.42 | 2.53 | 2.69 | 3.06 | 11.7 |
| STATE: | WISCONSIN | | | | |
| ZIP CODE | QUARTER 1 | QUARTER 2 | QUARTER 3 | QUARTER 4 | TOTAL GRAMS |
| 530 | 0.43 | 0.71 | 0.51 | 0.67 | 2.32 |
| 531 | 0 | 0.02 | 0 | 0 | 0.02 |
| 532 | 2.62 | 2.17 | 2.76 | 2.75 | 10.3 |
| 535 | 0.2 | 0.24 | 0.21 | 0.13 | 0.78 |
| 537 | 0.83 | 1.01 | 0.82 | 0.74 | 3.4 |
| 538 | 0.17 | 0.2 | 0.18 | 0.23 | 0.78 |
| 539 | 0.01 | 0 | 0.01 | 0 | 0.02 |

| | | | | | |
|---|---|---|---|---|---|
| 543 | 0.04 | 0 | 0 | 0 | 0.04 |
| 544 | 0.07 | 0.03 | 0.04 | 0.02 | 0.16 |
| 546 | 0.09 | 0.12 | 0.08 | 0.02 | 0.31 |
| 547 | 0.08 | 0.14 | 0.05 | 0.05 | 0.32 |

DRUG CODE:9737DRUG NAME:ALFENTANIL
STATE:WISCONSIN

| ZIP CODE | QUARTER 1 | QUARTER 2 | QUARTER 3 | QUARTER 4 | TOTAL GRAMS |
|---|---|---|---|---|---|
| 548 | 0 | 0 | 0 | 0.01 | 0.01 |
| 549 | 0 | 0.05 | 0 | 0.01 | 0.06 |
| TOTAL | 4.54 | 4.69 | 4.66 | 4.63 | 18.52 |

STATE: WYOMING

| ZIP CODE | QUARTER 1 | QUARTER 2 | QUARTER 3 | QUARTER 4 | TOTAL GRAMS |
|---|---|---|---|---|---|
| 820 | 0.01 | 0 | 0 | 0 | 0.01 |
| 822 | 0 | 0 | 0.01 | 0 | 0.01 |
| 823 | 0 | 0.02 | 0 | 0.02 | 0.04 |
| 826 | 0.02 | 0 | 0.01 | 0.02 | 0.05 |
| 830 | 0 | 0.03 | 0 | 0 | 0.03 |
| TOTAL | 0.03 | 0.05 | 0.02 | 0.04 | 0.14 |

DRUG CODE: 9739 DRUG NAME: REMIFENTANIL

STATE: ALABAMA

| ZIP CODE | QUARTER 1 | QUARTER 2 | QUARTER 3 | QUARTER 4 | TOTAL GRAMS |
|---|---|---|---|---|---|
| 350 | 0.01 | 0.01 | 0 | 0 | 0.02 |
| 352 | 1.61 | 2.12 | 1.85 | 2.08 | 7.66 |
| 354 | 0.02 | 0 | 0 | 0.01 | 0.03 |
| 356 | 0 | 0.02 | 0.04 | 0.08 | 0.14 |
| 358 | 0.02 | 0.02 | 0.05 | 0.06 | 0.15 |
| 361 | 0.48 | 0.49 | 0.59 | 0.58 | 2.14 |
| 363 | 0.1 | 0.08 | 0.1 | 0.12 | 0.4 |
| 366 | 0.37 | 0.62 | 0.91 | 0.19 | 2.09 |
| 368 | 0 | 0 | 0.01 | 0 | 0.01 |
| TOTAL | 2.61 | 3.36 | 3.55 | 3.12 | 12.64 |

STATE: ALASKA

| ZIP CODE | QUARTER 1 | QUARTER 2 | QUARTER 3 | QUARTER 4 | TOTAL GRAMS |
|---|---|---|---|---|---|
| 995 | 2.45 | 2.22 | 2.09 | 2.83 | 9.59 |
| 996 | 0.14 | 0.3 | 0.21 | 0.37 | 1.02 |
| 997 | 0.22 | 0.22 | 0.28 | 0.21 | 0.93 |
| TOTAL | 2.81 | 2.74 | 2.58 | 3.41 | 11.54 |

STATE: ARIZONA

| ZIP CODE | QUARTER 1 | QUARTER 2 | QUARTER 3 | QUARTER 4 | TOTAL GRAMS |
|---|---|---|---|---|---|
| 850 | 1.65 | 1.13 | 1.58 | 1.79 | 6.15 |
| 852 | 0.68 | 0.46 | 0.71 | 0.85 | 2.7 |
| 853 | 0.31 | 0.16 | 0.22 | 0.17 | 0.86 |
| 855 | 0 | 0.07 | 0 | 0.05 | 0.12 |
| 856 | 0 | 0 | 0.06 | 0 | 0.06 |
| 857 | 1.21 | 1.13 | 1.51 | 1.38 | 5.23 |
| 859 | 0.09 | 0.03 | 0.04 | 0.04 | 0.2 |
| 860 | 0.44 | 0.32 | 0.44 | 0.31 | 1.51 |
| 864 | 0.02 | 0.01 | 0 | 0 | 0.03 |
| TOTAL | 4.4 | 3.31 | 4.56 | 4.59 | 16.86 |

STATE: ARKANSAS

| ZIP CODE | QUARTER 1 | QUARTER 2 | QUARTER 3 | QUARTER 4 | TOTAL GRAMS |
|---|---|---|---|---|---|
| 716 | 0 | 0 | 0.04 | 0 | 0.04 |
| 720 | 0.14 | 0.18 | 0.07 | 0.07 | 0.46 |

| | | | | | |
|---|---|---|---|---|---|
| 722 | 1.26 | 1.14 | 1.52 | 1.66 | 5.58 |
| 724 | 0.3 | 0.2 | 0.04 | 0.06 | 0.6 |
| 725 | 0.05 | 0.01 | 0 | 0 | 0.06 |
| 727 | 0.03 | 0.01 | 0 | 0 | 0.04 |
| 728 | 0 | 0.06 | 0 | 0.06 | 0.12 |
| TOTAL | 1.78 | 1.6 | 1.67 | 1.85 | 6.9 |

STATE: CALIFORNIA

| ZIP CODE | QUARTER 1 | QUARTER 2 | QUARTER 3 | QUARTER 4 | TOTAL GRAMS |
|---|---|---|---|---|---|
| 900 | 3.52 | 4.21 | 4.18 | 4.44 | 16.35 |
| 902 | 0.29 | 0.38 | 0.35 | 0.47 | 1.49 |
| 904 | 0.65 | 1.02 | 0.66 | 0.59 | 2.92 |
| 905 | 0.38 | 0.3 | 0.34 | 0.33 | 1.35 |
| 907 | 0.14 | 0.08 | 0.11 | 0.11 | 0.44 |
| 908 | 0.32 | 0.38 | 0.36 | 0.2 | 1.26 |
| 910 | 0.04 | 0.03 | 0.02 | 0.03 | 0.12 |
| 911 | 0.26 | 0.3 | 0.22 | 0.25 | 1.03 |
| 912 | 0 | 0.01 | 0 | 0 | 0.01 |
| 913 | 0.1 | 0.11 | 0.04 | 0.07 | 0.32 |
| 914 | 0.02 | 0.02 | 0 | 0 | 0.04 |
| 917 | 0.03 | 0.01 | 0.14 | 0.08 | 0.26 |

DRUG CODE:9739DRUG NAME:REMIFENTANIL
STATE:CALIFORNIA

| ZIP CODE | QUARTER 1 | QUARTER 2 | QUARTER 3 | QUARTER 4 | TOTAL GRAMS |
|---|---|---|---|---|---|
| 919 | 0.23 | 0.17 | 0.26 | 0.25 | 0.91 |
| 920 | 0.62 | 0.98 | 0.85 | 0.82 | 3.27 |
| 921 | 1.3 | 1.14 | 1.41 | 1.19 | 5.04 |
| 922 | 0.51 | 0.44 | 0.26 | 0.35 | 1.56 |
| 923 | 0.78 | 1.12 | 1.14 | 1.12 | 4.16 |
| 924 | 0.5 | 0.35 | 0.63 | 0.47 | 1.95 |
| 925 | 0.08 | 0.23 | 0.17 | 0.27 | 0.75 |
| 926 | 1.68 | 1.49 | 1.41 | 1.7 | 6.28 |
| 927 | 0.14 | 0.11 | 0.18 | 0.17 | 0.6 |
| 928 | 1.85 | 1.96 | 2.06 | 1.99 | 7.86 |
| 930 | 0.24 | 0.26 | 0.26 | 0.27 | 1.03 |
| 931 | 0.34 | 0.46 | 0.41 | 0.51 | 1.72 |
| 932 | 0.17 | 0.42 | 0.26 | 0.21 | 1.06 |
| 933 | 0.02 | 0.02 | 0.1 | 0.16 | 0.3 |
| 934 | 0.06 | 0.02 | 0.07 | 0.05 | 0.2 |
| 935 | 0.04 | 0.04 | 0.01 | 0.02 | 0.11 |
| 936 | 0.26 | 0.2 | 0.21 | 0.33 | 1 |
| 937 | 0.72 | 1.01 | 1.39 | 1.24 | 4.36 |
| 939 | 0.05 | 0 | 0.02 | 0.04 | 0.11 |
| 940 | 1.4 | 1.27 | 1.79 | 1.51 | 5.97 |
| 941 | 3.14 | 3.09 | 3.09 | 3.64 | 12.96 |
| 943 | 4.31 | 4.86 | 5 | 5.01 | 19.18 |
| 944 | 0 | 0.03 | 0.04 | 0.06 | 0.13 |
| 945 | 1.01 | 0.76 | 1.05 | 1 | 3.82 |
| 946 | 0.19 | 0.08 | 0.08 | 0.19 | 0.54 |
| 948 | 0 | 0 | 0.01 | 0 | 0.01 |
| 949 | 0 | 0.03 | 0.02 | 0.05 | 0.1 |
| 950 | 0.27 | 0.18 | 0.26 | 0.17 | 0.88 |
| 951 | 0.98 | 1.13 | 0.98 | 1.23 | 4.32 |
| 952 | 0.19 | 0.25 | 0.14 | 0.1 | 0.68 |
| 953 | 0.02 | 0.05 | 0.05 | 0.06 | 0.18 |
| 954 | 0.07 | 0.09 | 0.05 | 0.09 | 0.3 |
| 955 | 0.02 | 0.02 | 0 | 0.05 | 0.09 |
| 956 | 0.53 | 1.14 | 0.77 | 0.86 | 3.3 |
| 957 | 0.01 | 0 | 0.01 | 0 | 0.02 |

| | | | | | |
|---|---|---|---|---|---|
| 958 | 1.71 | 2.08 | 2.13 | 1.78 | 7.7 |
| 959 | 0.07 | 0.05 | 0.02 | 0 | 0.14 |
| 960 | 0.33 | 0.29 | 0.31 | 0.2 | 1.13 |
| TOTAL | 29.59 | 32.67 | 33.32 | 33.73 | 129.31 |

STATE: COLORADO

| ZIP CODE | QUARTER 1 | QUARTER 2 | QUARTER 3 | QUARTER 4 | TOTAL GRAMS |
|---|---|---|---|---|---|
| 800 | 2.31 | 2.71 | 2.66 | 2.82 | 10.5 |
| 801 | 0.54 | 0.18 | 0.57 | 0.45 | 1.74 |
| 802 | 1.15 | 0.95 | 1.02 | 1.21 | 4.33 |
| 803 | 0.33 | 0.26 | 0.37 | 0.32 | 1.28 |
| 804 | 0.15 | 0.09 | 0.14 | 0.16 | 0.54 |
| 805 | 0.13 | 0.16 | 0.17 | 0.13 | 0.59 |
| 806 | 0.13 | 0.14 | 0.12 | 0.16 | 0.55 |
| 807 | 0.02 | 0.01 | 0.02 | 0 | 0.05 |
| 808 | 0.02 | 0.04 | 0.02 | 0.03 | 0.11 |
| 809 | 0.16 | 0.25 | 0.22 | 0.24 | 0.87 |
| 810 | 0.26 | 0.24 | 0.36 | 0.3 | 1.16 |
| 812 | 0.01 | 0 | 0.02 | 0.01 | 0.04 |
| 813 | 0.28 | 0.34 | 0.36 | 0.4 | 1.38 |
| 814 | 0.02 | 0.02 | 0.03 | 0.01 | 0.08 |
| 815 | 0.36 | 0.33 | 0.32 | 0.36 | 1.37 |
| 816 | 0.21 | 0.14 | 0.16 | 0.13 | 0.64 |
| TOTAL | 6.08 | 5.86 | 6.56 | 6.73 | 25.23 |

STATE: CONNECTICUT

| ZIP CODE | QUARTER 1 | QUARTER 2 | QUARTER 3 | QUARTER 4 | TOTAL GRAMS |
|---|---|---|---|---|---|
| 60 | 0.1 | 0.2 | 0.13 | 0.15 | 0.58 |
| 61 | 0.55 | 0.6 | 0.6 | 0.47 | 2.22 |
| 62 | 0 | 0 | 0 | 0.04 | 0.04 |
| 63 | 0.04 | 0.1 | 0.07 | 0.12 | 0.33 |
| 64 | 0.31 | 0.58 | 0.38 | 0.58 | 1.85 |
| 65 | 5.03 | 5.54 | 5.57 | 5.25 | 21.39 |

DRUG CODE:9739DRUG NAME:REMIFENTANIL
STATE:CONNECTICUT

| ZIP CODE | QUARTER 1 | QUARTER 2 | QUARTER 3 | QUARTER 4 | TOTAL GRAMS |
|---|---|---|---|---|---|
| 66 | 0.62 | 0.65 | 0.82 | 0.91 | 3 |
| 67 | 0.13 | 0.1 | 0.12 | 0.07 | 0.42 |
| 68 | 0.22 | 0.19 | 0.33 | 0.43 | 1.17 |
| 69 | 0.3 | 0.28 | 0.22 | 0.34 | 1.14 |
| TOTAL | 7.3 | 8.24 | 8.24 | 8.36 | 32.14 |

STATE: DELAWARE

| ZIP CODE | QUARTER 1 | QUARTER 2 | QUARTER 3 | QUARTER 4 | TOTAL GRAMS |
|---|---|---|---|---|---|
| 197 | 0.55 | 0.62 | 0.58 | 0.58 | 2.33 |
| 198 | 2.21 | 2.46 | 2.86 | 1.38 | 8.91 |
| 199 | 0.54 | 0.3 | 0.26 | 0.36 | 1.46 |
| TOTAL | 3.3 | 3.38 | 3.7 | 2.32 | 12.7 |

STATE: DISTRICT OF COLUMBIA

| ZIP CODE | QUARTER 1 | QUARTER 2 | QUARTER 3 | QUARTER 4 | TOTAL GRAMS |
|---|---|---|---|---|---|
| 200 | 0.59 | 0.77 | 0.94 | 0.73 | 3.03 |
| 204 | 0 | 0 | 0 | 0.06 | 0.06 |
| TOTAL | 0.59 | 0.77 | 0.94 | 0.79 | 3.09 |

STATE: FLORIDA

| ZIP CODE | QUARTER 1 | QUARTER 2 | QUARTER 3 | QUARTER 4 | TOTAL GRAMS |
|---|---|---|---|---|---|
| 320 | 0.01 | 0 | 0.04 | 0.01 | 0.06 |
| 321 | 0.02 | 0.07 | 0.14 | 0.17 | 0.4 |

| | | | | | |
|---|---|---|---|---|---|
| 322 | 1.03 | 1.1 | 1.36 | 1.49 | 4.98 |
| 323 | 0 | 0.1 | 0.04 | 0 | 0.14 |
| 324 | 0.01 | 0 | 0 | 0 | 0.01 |
| 325 | 1.13 | 0.96 | 0.82 | 0.82 | 3.73 |
| 326 | 1.87 | 1.55 | 1.84 | 1.4 | 6.66 |
| 327 | 0.8 | 0.72 | 0.7 | 1.05 | 3.27 |
| 328 | 3.07 | 2.61 | 2.44 | 2.58 | 10.7 |
| 329 | 0.09 | 0.23 | 0.23 | 0.38 | 0.93 |
| 330 | 0.43 | 0.6 | 0.61 | 0.53 | 2.17 |
| 331 | 4.15 | 4.1 | 4.95 | 5.22 | 18.42 |
| 333 | 0.59 | 0.74 | 0.79 | 0.91 | 3.03 |
| 334 | 4.2 | 4.17 | 3.51 | 3.56 | 15.44 |
| 335 | 0.28 | 0.2 | 0.12 | 0.27 | 0.87 |
| 336 | 4.17 | 5.31 | 4.46 | 4.32 | 18.26 |
| 337 | 0.51 | 0.66 | 0.7 | 0.75 | 2.62 |
| 338 | 0.23 | 0.25 | 0.23 | 0.21 | 0.92 |
| 339 | 0.3 | 0.34 | 0.43 | 0.28 | 1.35 |
| 341 | 0.03 | 0.06 | 0.07 | 0.06 | 0.22 |
| 342 | 0.3 | 0.26 | 0.28 | 0.33 | 1.17 |
| 344 | 0.08 | 0.1 | 0.09 | 0.11 | 0.38 |
| 346 | 0.4 | 0.51 | 0.53 | 0.52 | 1.96 |
| 347 | 1.02 | 0.65 | 0.92 | 0.73 | 3.32 |
| 349 | 0.34 | 0.58 | 0.57 | 0.48 | 1.97 |
| TOTAL | 25.06 | 25.87 | 25.87 | 26.18 | 102.98 |

STATE: GEORGIA

| ZIP CODE | QUARTER 1 | QUARTER 2 | QUARTER 3 | QUARTER 4 | TOTAL GRAMS |
|---|---|---|---|---|---|
| 300 | 0.87 | 0.71 | 0.72 | 1.01 | 3.31 |
| 301 | 0.1 | 0.11 | 0.09 | 0.08 | 0.38 |
| 302 | 0.28 | 0.21 | 0.29 | 0.22 | 1 |
| 303 | 0.85 | 0.99 | 0.69 | 0.77 | 3.3 |
| 305 | 0.06 | 0.04 | 0.11 | 0.13 | 0.34 |
| 306 | 0.11 | 0.07 | 0.1 | 0.11 | 0.39 |
| 307 | 0.05 | 0.06 | 0.05 | 0.05 | 0.21 |
| 309 | 1.41 | 1.3 | 1.17 | 1.78 | 5.66 |
| 310 | 0.01 | 0 | 0.02 | 0 | 0.03 |
| 312 | 0.67 | 0.59 | 0.67 | 0.69 | 2.62 |
| 314 | 0.05 | 0.03 | 0.05 | 0.08 | 0.21 |
| 315 | 0.04 | 0.02 | 0.04 | 0.04 | 0.14 |
| 316 | 0.04 | 0.02 | 0.04 | 0.03 | 0.13 |
| 317 | 0.06 | 0.13 | 0.15 | 0.17 | 0.51 |
| 319 | 0 | 0.06 | 0.54 | 0 | 0.6 |
| 398 | 0.02 | 0.02 | 0.04 | 0.04 | 0.12 |
| TOTAL | 4.62 | 4.36 | 4.77 | 5.2 | 18.95 |

STATE: GUAM

| ZIP CODE | QUARTER 1 | QUARTER 2 | QUARTER 3 | QUARTER 4 | TOTAL GRAMS |
|---|---|---|---|---|---|
| 969 | 0 | 0 | 0.5 | 0 | 0.5 |

DRUG CODE:9739DRUG NAME:REMIFENTANIL
STATE:GUAM

| ZIP CODE | QUARTER 1 | QUARTER 2 | QUARTER 3 | QUARTER 4 | TOTAL GRAMS |
|---|---|---|---|---|---|
| TOTAL | 0 | 0 | 0.5 | 0 | 0.5 |

STATE: HAWAII

| ZIP CODE | QUARTER 1 | QUARTER 2 | QUARTER 3 | QUARTER 4 | TOTAL GRAMS |
|---|---|---|---|---|---|
| 967 | 0.04 | 0 | 0.02 | 0.04 | 0.1 |
| 968 | 1.44 | 1.31 | 1.19 | 1.29 | 5.23 |
| TOTAL | 1.48 | 1.31 | 1.21 | 1.33 | 5.33 |

RETAIL DRUG DISTRIBUTION BY ZIP CODE WITHIN STATE BY GRAMS WT

STATE: IDAHO

| ZIP CODE | QUARTER 1 | QUARTER 2 | QUARTER 3 | QUARTER 4 | TOTAL GRAMS |
|---|---|---|---|---|---|
| 832 | 0.05 | 0.11 | 0.21 | 0.12 | 0.49 |
| 833 | 0.02 | 0.15 | 0.1 | 0.43 | 0.7 |
| 834 | 0.52 | 0.71 | 0.49 | 0.79 | 2.51 |
| 836 | 0.19 | 0.34 | 0.25 | 0.41 | 1.19 |
| 837 | 0.97 | 1.02 | 0.88 | 1.03 | 3.9 |
| 838 | 0 | 0 | 0.01 | 0.14 | 0.15 |
| TOTAL | 1.75 | 2.33 | 1.94 | 2.92 | 8.94 |

STATE: ILLINOIS

| ZIP CODE | QUARTER 1 | QUARTER 2 | QUARTER 3 | QUARTER 4 | TOTAL GRAMS |
|---|---|---|---|---|---|
| 600 | 1.07 | 1.05 | 1.26 | 0.97 | 4.35 |
| 601 | 2.06 | 2.03 | 2.35 | 2.77 | 9.21 |
| 602 | 0.32 | 0.39 | 0.39 | 0.42 | 1.52 |
| 603 | 0 | 0 | 0 | 0.05 | 0.05 |
| 604 | 1.04 | 0.87 | 1.08 | 1.18 | 4.17 |
| 605 | 0.55 | 1.13 | 0.64 | 0.97 | 3.29 |
| 606 | 6.25 | 4.8 | 6.15 | 5.16 | 22.36 |
| 607 | 0.12 | 0.14 | 0.24 | 0.1 | 0.6 |
| 610 | 0.03 | 0.02 | 0 | 0.01 | 0.06 |
| 611 | 0.83 | 0.94 | 0.93 | 1.3 | 4 |
| 612 | 0.21 | 0.17 | 0.15 | 0.16 | 0.69 |
| 613 | 0.01 | 0 | 0 | 0 | 0.01 |
| 614 | 0.34 | 0.28 | 0.25 | 0.27 | 1.14 |
| 616 | 0.28 | 0.23 | 0.31 | 0.44 | 1.26 |
| 617 | 0.23 | 0.17 | 0.18 | 0.13 | 0.71 |
| 618 | 0.1 | 0.08 | 0.14 | 0.09 | 0.41 |
| 619 | 0.03 | 0 | 0 | 0 | 0.03 |
| 622 | 0.23 | 0.15 | 0.2 | 0.15 | 0.73 |
| 625 | 0 | 0 | 0.06 | 0 | 0.06 |
| 627 | 0.3 | 0.42 | 0.4 | 0.33 | 1.45 |
| 629 | 0.11 | 0.18 | 0.1 | 0.18 | 0.57 |
| TOTAL | 14.11 | 13.05 | 14.83 | 14.68 | 56.67 |

STATE: INDIANA

| ZIP CODE | QUARTER 1 | QUARTER 2 | QUARTER 3 | QUARTER 4 | TOTAL GRAMS |
|---|---|---|---|---|---|
| 460 | 0.79 | 0.77 | 0.73 | 0.83 | 3.12 |
| 461 | 0.04 | 0.02 | 0.04 | 0.02 | 0.12 |
| 462 | 3.63 | 3.38 | 3.06 | 4.35 | 14.42 |
| 463 | 0.46 | 0.56 | 0.53 | 0.57 | 2.12 |
| 464 | 0.03 | 0.02 | 0.03 | 0.06 | 0.14 |
| 465 | 0.42 | 0.65 | 0.62 | 0.62 | 2.31 |
| 466 | 0.12 | 0.15 | 0.14 | 0.1 | 0.51 |
| 468 | 0.2 | 0.28 | 0.31 | 0.42 | 1.21 |
| 469 | 0.02 | 0.04 | 0.01 | 0.03 | 0.1 |
| 471 | 0.1 | 0.05 | 0.15 | 0.15 | 0.45 |
| 472 | 0.2 | 0.25 | 0.18 | 0.32 | 0.95 |
| 473 | 0.8 | 0.73 | 0.52 | 0.7 | 2.75 |
| 474 | 0.32 | 0.14 | 0.19 | 0.24 | 0.89 |
| 475 | 0.21 | 0.17 | 0.23 | 0.18 | 0.79 |
| 476 | 0.03 | 0.08 | 0.07 | 0.06 | 0.24 |
| 477 | 0.18 | 0.12 | 0.15 | 0.27 | 0.72 |
| 478 | 0.13 | 0.14 | 0.14 | 0.14 | 0.55 |
| 479 | 0.08 | 0.12 | 0.1 | 0.09 | 0.39 |
| TOTAL | 7.76 | 7.67 | 7.2 | 9.15 | 31.78 |

STATE: IOWA

| ZIP CODE | QUARTER 1 | QUARTER 2 | QUARTER 3 | QUARTER 4 | TOTAL GRAMS |
|---|---|---|---|---|---|
| 500 | 0.2 | 0.26 | 0.12 | 0.22 | 0.8 |

| | | | | | |
|---|---|---|---|---|---|
| 501 | 0.02 | 0.02 | 0 | 0 | 0.04 |
| 502 | 0 | 0.04 | 0.04 | 0 | 0.08 |
| 503 | 0.07 | 0 | 0.02 | 0.07 | 0.16 |

DRUG CODE:9739DRUG NAME:REMIFENTANIL
STATE:IOWA

| ZIP CODE | QUARTER 1 | QUARTER 2 | QUARTER 3 | QUARTER 4 | TOTAL GRAMS |
|---|---|---|---|---|---|
| 504 | 0.02 | 0 | 0.01 | 0.02 | 0.05 |
| 505 | 0.09 | 0.04 | 0.08 | 0.03 | 0.24 |
| 506 | 0.02 | 0 | 0.02 | 0 | 0.04 |
| 507 | 0.1 | 0.16 | 0.08 | 0.12 | 0.46 |
| 511 | 0.01 | 0 | 0 | 0 | 0.01 |
| 512 | 0 | 0.02 | 0.02 | 0.06 | 0.1 |
| 520 | 0.15 | 0.12 | 0.14 | 0.12 | 0.53 |
| 522 | 1.86 | 2.24 | 2.17 | 2.19 | 8.46 |
| 524 | 0.02 | 0.02 | 0.02 | 0.01 | 0.07 |
| 527 | 0.06 | 0.08 | 0.04 | 0.06 | 0.24 |
| 528 | 0.04 | 0.06 | 0.07 | 0.1 | 0.27 |
| TOTAL | 2.66 | 3.06 | 2.83 | 3 | 11.55 |

STATE: KANSAS

| ZIP CODE | QUARTER 1 | QUARTER 2 | QUARTER 3 | QUARTER 4 | TOTAL GRAMS |
|---|---|---|---|---|---|
| 660 | 0.25 | 0.09 | 0.37 | 0.34 | 1.05 |
| 661 | 0.8 | 0.69 | 0.8 | 1.16 | 3.45 |
| 662 | 0.19 | 0.35 | 0.13 | 0.4 | 1.07 |
| 664 | 0 | 0 | 0.02 | 0 | 0.02 |
| 665 | 0.03 | 0.01 | 0.03 | 0.03 | 0.1 |
| 666 | 0.22 | 0.13 | 0.3 | 0.24 | 0.89 |
| 667 | 0 | 0 | 0.01 | 0.01 | 0.02 |
| 670 | 0.05 | 0.06 | 0.02 | 0.04 | 0.17 |
| 672 | 0.25 | 0.17 | 0.27 | 0.25 | 0.94 |
| 674 | 0.06 | 0.05 | 0.04 | 0.03 | 0.18 |
| 675 | 0 | 0.02 | 0.06 | 0 | 0.08 |
| TOTAL | 1.85 | 1.57 | 2.05 | 2.5 | 7.97 |

STATE: KENTUCKY

| ZIP CODE | QUARTER 1 | QUARTER 2 | QUARTER 3 | QUARTER 4 | TOTAL GRAMS |
|---|---|---|---|---|---|
| 402 | 1.66 | 1.7 | 1.44 | 1.33 | 6.13 |
| 403 | 0 | 0 | 0 | 0.03 | 0.03 |
| 404 | 0.39 | 0.2 | 0.19 | 0.19 | 0.97 |
| 405 | 0.88 | 1.17 | 1.91 | 1.5 | 5.46 |
| 407 | 0 | 0 | 0 | 0.02 | 0.02 |
| 410 | 0.08 | 0.1 | 0.04 | 0.08 | 0.3 |
| 411 | 0.15 | 0.2 | 0.1 | 0.06 | 0.51 |
| 415 | 0 | 0 | 0.04 | 0 | 0.04 |
| 420 | 0.28 | 0.22 | 0.3 | 0.26 | 1.06 |
| 421 | 1.03 | 0.89 | 0.84 | 0.92 | 3.68 |
| 422 | 0 | 0 | 0 | 0.01 | 0.01 |
| 423 | 0.05 | 0.06 | 0.05 | 0.07 | 0.23 |
| 425 | 0 | 0 | 0.02 | 0 | 0.02 |
| TOTAL | 4.52 | 4.54 | 4.93 | 4.47 | 18.46 |

STATE: LOUISIANA

| ZIP CODE | QUARTER 1 | QUARTER 2 | QUARTER 3 | QUARTER 4 | TOTAL GRAMS |
|---|---|---|---|---|---|
| 700 | 0.36 | 0.31 | 0.31 | 0.34 | 1.32 |
| 701 | 1.81 | 1.95 | 1.87 | 2.37 | 8 |
| 703 | 0.07 | 0.02 | 0.04 | 0.01 | 0.14 |
| 704 | 0.35 | 0.35 | 0.28 | 0.38 | 1.36 |
| 705 | 0.57 | 0.55 | 0.51 | 0.6 | 2.23 |
| 706 | 0 | 0 | 0.02 | 0 | 0.02 |

| | | | | | |
|---|---|---|---|---|---|
| 708 | 0.67 | 0.69 | 0.5 | 0.45 | 2.31 |
| 711 | 0.34 | 0.38 | 0.33 | 0.31 | 1.36 |
| 712 | 0.02 | 0.11 | 0.06 | 0.04 | 0.23 |
| 713 | 0 | 0 | 0.02 | 0 | 0.02 |
| 714 | 0 | 0 | 0 | 0 | 0 |
| TOTAL | 4.19 | 4.36 | 3.94 | 4.5 | 16.99 |

STATE: MAINE

| ZIP CODE | QUARTER 1 | QUARTER 2 | QUARTER 3 | QUARTER 4 | TOTAL GRAMS |
|---|---|---|---|---|---|
| 40 | 0.04 | 0.02 | 0.02 | 0.04 | 0.12 |
| 41 | 0.25 | 0.28 | 0.43 | 0.76 | 1.72 |
| 42 | 0.25 | 0.21 | 0.22 | 0.29 | 0.97 |
| 43 | 0.06 | 0.05 | 0.02 | 0.1 | 0.23 |
| 44 | 0.41 | 0.44 | 0.45 | 0.52 | 1.82 |
| 46 | 0.01 | 0.03 | 0.04 | 0.06 | 0.14 |
| 47 | 0 | 0.02 | 0 | 0 | 0.02 |
| 49 | 0.04 | 0 | 0.04 | 0.02 | 0.1 |

DRUG CODE:9739DRUG NAME:REMIFENTANIL
STATE:MAINE

| ZIP CODE | QUARTER 1 | QUARTER 2 | QUARTER 3 | QUARTER 4 | TOTAL GRAMS |
|---|---|---|---|---|---|
| TOTAL | 1.06 | 1.05 | 1.22 | 1.79 | 5.12 |

STATE: MARYLAND

| ZIP CODE | QUARTER 1 | QUARTER 2 | QUARTER 3 | QUARTER 4 | TOTAL GRAMS |
|---|---|---|---|---|---|
| 207 | 0.08 | 0.14 | 0.22 | 0 | 0.44 |
| 208 | 1.59 | 0.68 | 1.11 | 1.09 | 4.47 |
| 209 | 0 | 0.06 | 0.02 | 0.02 | 0.1 |
| 210 | 0.13 | 0.27 | 0.13 | 0.11 | 0.64 |
| 211 | 0 | 0.05 | 0 | 0.01 | 0.06 |
| 212 | 4.79 | 5.53 | 5.74 | 5.31 | 21.37 |
| 214 | 0 | 0.02 | 0.04 | 0.02 | 0.08 |
| 216 | 0 | 0 | 0 | 0.02 | 0.02 |
| 217 | 0.07 | 0.2 | 0.06 | 0.07 | 0.4 |
| 218 | 0.24 | 0.12 | 0.24 | 0.12 | 0.72 |
| TOTAL | 6.9 | 7.07 | 7.56 | 6.77 | 28.3 |

STATE: MASSACHUSETTS

| ZIP CODE | QUARTER 1 | QUARTER 2 | QUARTER 3 | QUARTER 4 | TOTAL GRAMS |
|---|---|---|---|---|---|
| 10 | 0.13 | 0.09 | 0.09 | 0.06 | 0.37 |
| 11 | 0.16 | 0.23 | 0.17 | 0.2 | 0.76 |
| 12 | 0.04 | 0.06 | 0.08 | 0.08 | 0.26 |
| 13 | 0 | 0 | 0 | 0.05 | 0.05 |
| 14 | 0.1 | 0.11 | 0.05 | 0.08 | 0.34 |
| 15 | 0.13 | 0.01 | 0.03 | 0 | 0.17 |
| 16 | 0.8 | 1.11 | 1.02 | 1.05 | 3.98 |
| 17 | 0.27 | 0.25 | 0.24 | 0.24 | 1 |
| 18 | 0.94 | 0.94 | 0.88 | 1.05 | 3.81 |
| 19 | 0.74 | 0.48 | 0.53 | 0.56 | 2.31 |
| 20 | 0.33 | 0.35 | 0.31 | 0.3 | 1.29 |
| 21 | 13.43 | 14.58 | 13.83 | 14.56 | 56.4 |
| 22 | 0.6 | 0.85 | 0.94 | 0.89 | 3.28 |
| 23 | 0.27 | 0.22 | 0.15 | 0.27 | 0.91 |
| 24 | 0.24 | 0.28 | 0.29 | 0.2 | 1.01 |
| 25 | 0.04 | 0.05 | 0.06 | 0.04 | 0.19 |
| 26 | 0.07 | 0.1 | 0.08 | 0.1 | 0.35 |
| 27 | 0.05 | 0.09 | 0.05 | 0.04 | 0.23 |
| TOTAL | 18.34 | 19.8 | 18.8 | 19.77 | 76.71 |

STATE: MICHIGAN

| ZIP CODE | QUARTER 1 | QUARTER 2 | QUARTER 3 | QUARTER 4 | TOTAL GRAMS |
|---|---|---|---|---|---|
| 480 | 1.25 | 1.21 | 1.06 | 1.33 | 4.85 |
| 481 | 4.13 | 3.57 | 3.85 | 3.79 | 15.34 |
| 482 | 2.34 | 3.03 | 2.61 | 3.46 | 11.44 |
| 483 | 0.85 | 0.77 | 0.74 | 0.82 | 3.18 |
| 485 | 0.06 | 0.06 | 0.02 | 0.06 | 0.2 |
| 486 | 0.23 | 0.4 | 0.35 | 0.3 | 1.28 |
| 487 | 0.31 | 0.29 | 0.3 | 0.47 | 1.37 |
| 489 | 0.23 | 0.19 | 0.26 | 0.2 | 0.88 |
| 490 | 0.17 | 0.43 | 0.23 | 0.28 | 1.11 |
| 492 | 0.02 | 0.07 | 0.02 | 0.07 | 0.18 |
| 494 | 0.02 | 0.04 | 0.06 | 0.11 | 0.23 |
| 495 | 0.26 | 0.14 | 0.2 | 0.19 | 0.79 |
| 497 | 0.06 | 0.03 | 0.03 | 0.05 | 0.17 |
| 498 | 0.03 | 0.02 | 0.03 | 0.01 | 0.09 |
| TOTAL | 9.96 | 10.25 | 9.76 | 11.14 | 41.11 |

STATE: MINNESOTA

| ZIP CODE | QUARTER 1 | QUARTER 2 | QUARTER 3 | QUARTER 4 | TOTAL GRAMS |
|---|---|---|---|---|---|
| 550 | 0.01 | 0.01 | 0 | 0.02 | 0.04 |
| 551 | 0.67 | 0.7 | 0.59 | 0.9 | 2.86 |
| 553 | 0.05 | 0.04 | 0.03 | 0.05 | 0.17 |
| 554 | 4.1 | 3.58 | 3.38 | 3.99 | 15.05 |
| 558 | 0.22 | 0.37 | 0.18 | 0.45 | 1.22 |
| 559 | 1.63 | 1.67 | 1.6 | 2.27 | 7.17 |
| 562 | 0 | 0 | 0.02 | 0 | 0.02 |
| 563 | 0.02 | 0.04 | 0.06 | 0.08 | 0.2 |
| 564 | 0.01 | 0 | 0.01 | 0.01 | 0.03 |
| TOTAL | 6.71 | 6.41 | 5.87 | 7.77 | 26.76 |

STATE: MISSISSIPPI

| ZIP CODE | QUARTER 1 | QUARTER 2 | QUARTER 3 | QUARTER 4 | TOTAL GRAMS |
|---|---|---|---|---|---|

DRUG CODE:9739DRUG NAME:REMIFENTANIL
STATE:MISSISSIPPI

| ZIP CODE | QUARTER 1 | QUARTER 2 | QUARTER 3 | QUARTER 4 | TOTAL GRAMS |
|---|---|---|---|---|---|
| 392 | 0.32 | 0.38 | 0.29 | 0.34 | 1.33 |
| 394 | 0.02 | 0.04 | 0.06 | 0 | 0.12 |
| 395 | 0.16 | 0.05 | 0.21 | 0.04 | 0.46 |
| 396 | 0.02 | 0 | 0 | 0 | 0.02 |
| 397 | 0 | 0.01 | 0 | 0 | 0.01 |
| TOTAL | 0.52 | 0.48 | 0.56 | 0.38 | 1.94 |

STATE: MISSOURI

| ZIP CODE | QUARTER 1 | QUARTER 2 | QUARTER 3 | QUARTER 4 | TOTAL GRAMS |
|---|---|---|---|---|---|
| 630 | 0.04 | 0.12 | 0.15 | 0.02 | 0.33 |
| 631 | 1.84 | 2.16 | 2.17 | 2.18 | 8.35 |
| 633 | 0.02 | 0.02 | 0.01 | 0.01 | 0.06 |
| 637 | 0.16 | 0.18 | 0.31 | 0.24 | 0.89 |
| 638 | 0.02 | 0 | 0.02 | 0.02 | 0.06 |
| 640 | 0.02 | 0.07 | 0.04 | 0.05 | 0.18 |
| 641 | 1.27 | 1.4 | 1.76 | 1.43 | 5.86 |
| 645 | 0.05 | 0.05 | 0.1 | 0.05 | 0.25 |
| 648 | 0.16 | 0.04 | 0.12 | 0 | 0.32 |
| 650 | 0.15 | 0.15 | 0.2 | 0.22 | 0.72 |
| 651 | 0.18 | 0.2 | 0.1 | 0.1 | 0.58 |
| 652 | 0.7 | 0.4 | 0.69 | 0.4 | 2.19 |
| 654 | 0.08 | 0.14 | 0.18 | 0.08 | 0.48 |
| 656 | 0.04 | 0.02 | 0.08 | 0.04 | 0.18 |

| | | | | | |
|---|---|---|---|---|---|
| 658 | 0.16 | 0.14 | 0.18 | 0.06 | 0.54 |
| TOTAL | 4.89 | 5.09 | 6.11 | 4.9 | 20.99 |

STATE: MONTANA

| ZIP CODE | QUARTER 1 | QUARTER 2 | QUARTER 3 | QUARTER 4 | TOTAL GRAMS |
|---|---|---|---|---|---|
| 591 | 0.04 | 0.08 | 0.08 | 0.07 | 0.27 |
| 592 | 0.02 | 0 | 0 | 0.01 | 0.03 |
| 594 | 0.27 | 0.02 | 0.02 | 0.12 | 0.43 |
| 596 | 0.14 | 0.1 | 0.08 | 0.12 | 0.44 |
| 597 | 0.02 | 0.06 | 0.06 | 0.07 | 0.21 |
| 598 | 1.34 | 1.82 | 1.25 | 1.34 | 5.75 |
| 599 | 0.01 | 0.03 | 0.02 | 0.01 | 0.07 |
| TOTAL | 1.84 | 2.11 | 1.51 | 1.74 | 7.2 |

STATE: NEBRASKA

| ZIP CODE | QUARTER 1 | QUARTER 2 | QUARTER 3 | QUARTER 4 | TOTAL GRAMS |
|---|---|---|---|---|---|
| 680 | 0.02 | 0.04 | 0.04 | 0.05 | 0.15 |
| 681 | 0.44 | 0.78 | 0.73 | 0.61 | 2.56 |
| 685 | 0.03 | 0.01 | 0.03 | 0.07 | 0.14 |
| 687 | 0 | 0 | 0 | 0.01 | 0.01 |
| 688 | 0.23 | 0.12 | 0.09 | 0.17 | 0.61 |
| 689 | 0.02 | 0.04 | 0.02 | 0.04 | 0.12 |
| 691 | 0 | 0.01 | 0 | 0 | 0.01 |
| 693 | 0.08 | 0.12 | 0.08 | 0.1 | 0.38 |
| TOTAL | 0.82 | 1.12 | 0.99 | 1.05 | 3.98 |

STATE: NEVADA

| ZIP CODE | QUARTER 1 | QUARTER 2 | QUARTER 3 | QUARTER 4 | TOTAL GRAMS |
|---|---|---|---|---|---|
| 890 | 0.07 | 0.07 | 0.07 | 0.08 | 0.29 |
| 891 | 0.41 | 0.32 | 0.2 | 0.27 | 1.2 |
| 894 | 0.02 | 0.02 | 0.02 | 0.01 | 0.07 |
| 895 | 0.43 | 0.43 | 0.46 | 0.53 | 1.85 |
| 897 | 0.02 | 0 | 0 | 0.08 | 0.1 |
| 898 | 0 | 0 | 0.02 | 0 | 0.02 |
| TOTAL | 0.95 | 0.84 | 0.77 | 0.97 | 3.53 |

STATE: NEW HAMPSHIRE

| ZIP CODE | QUARTER 1 | QUARTER 2 | QUARTER 3 | QUARTER 4 | TOTAL GRAMS |
|---|---|---|---|---|---|
| 30 | 0.12 | 0.14 | 0.12 | 0.08 | 0.46 |
| 31 | 0.29 | 0.19 | 0.26 | 0.36 | 1.1 |
| 32 | 0.01 | 0 | 0.01 | 0 | 0.02 |
| 33 | 0.09 | 0.08 | 0.13 | 0.11 | 0.41 |
| 34 | 0 | 0.05 | 0 | 0 | 0.05 |
| 35 | 0 | 0 | 0.01 | 0 | 0.01 |
| 37 | 0.39 | 0.43 | 0.46 | 0.36 | 1.64 |
| 38 | 0.23 | 0.28 | 0.31 | 0.21 | 1.03 |
| TOTAL | 1.13 | 1.17 | 1.3 | 1.12 | 4.72 |

STATE: NEW JERSEY

DRUG CODE:9739DRUG NAME:REMIFENTANIL
STATE:NEW JERSEY

| ZIP CODE | QUARTER 1 | QUARTER 2 | QUARTER 3 | QUARTER 4 | TOTAL GRAMS |
|---|---|---|---|---|---|
| 70 | 0.53 | 0.84 | 0.66 | 0.88 | 2.91 |
| 71 | 1.28 | 1.55 | 1.4 | 1.04 | 5.27 |
| 72 | 0.06 | 0.06 | 0.06 | 0.16 | 0.34 |
| 73 | 0.03 | 0.24 | 0.02 | 0.03 | 0.32 |
| 74 | 0.4 | 0.4 | 0.4 | 0.44 | 1.64 |
| 75 | 0.54 | 0.55 | 0.55 | 0.73 | 2.37 |
| 76 | 1.02 | 0.93 | 0.91 | 1 | 3.86 |

| | | | | | |
|---|---|---|---|---|---|
| 77 | 1.3 | 0.55 | 1.07 | 1.25 | 4.17 |
| 78 | 0 | 0 | 0.03 | 0.02 | 0.05 |
| 79 | 1.19 | 1.06 | 1.14 | 1.26 | 4.65 |
| 80 | 1.39 | 1.26 | 1.29 | 1.82 | 5.76 |
| 81 | 0.62 | 0.9 | 0.97 | 0.81 | 3.3 |
| 82 | 0.59 | 0.73 | 0.61 | 0.53 | 2.46 |
| 84 | 0.28 | 0.2 | 0.32 | 0.26 | 1.06 |
| 85 | 0.35 | 0.22 | 0.25 | 0.2 | 1.02 |
| 86 | 0.3 | 0.21 | 0.31 | 0.14 | 0.96 |
| 87 | 0.08 | 0.09 | 0.03 | 0.12 | 0.32 |
| 88 | 0.43 | 0.5 | 0.57 | 0.84 | 2.34 |
| 89 | 1.4 | 1.75 | 3.32 | 0.35 | 6.82 |
| TOTAL | 11.79 | 12.04 | 13.91 | 11.88 | 49.62 |

STATE: NEW MEXICO

| ZIP CODE | QUARTER 1 | QUARTER 2 | QUARTER 3 | QUARTER 4 | TOTAL GRAMS |
|---|---|---|---|---|---|
| 871 | 0.72 | 0.61 | 0.66 | 0.84 | 2.83 |
| 873 | 0 | 0 | 0 | 0.08 | 0.08 |
| 874 | 0.1 | 0.07 | 0.02 | 0.07 | 0.26 |
| 875 | 0.09 | 0.06 | 0.13 | 0.1 | 0.38 |
| 880 | 0.11 | 0.05 | 0.09 | 0.11 | 0.36 |
| 882 | 0.03 | 0 | 0 | 0 | 0.03 |
| 883 | 0 | 0 | 0 | 0.04 | 0.04 |
| 884 | 0.04 | 0.06 | 0.06 | 0.04 | 0.2 |
| TOTAL | 1.09 | 0.85 | 0.96 | 1.28 | 4.18 |

STATE: NEW YORK

| ZIP CODE | QUARTER 1 | QUARTER 2 | QUARTER 3 | QUARTER 4 | TOTAL GRAMS |
|---|---|---|---|---|---|
| 100 | 17.64 | 20.21 | 16.25 | 19.9 | 74 |
| 101 | 0.05 | 0.04 | 0.04 | 0.03 | 0.16 |
| 103 | 0 | 0.05 | 0.2 | 0.15 | 0.4 |
| 104 | 1.57 | 1.47 | 1.44 | 1.28 | 5.76 |
| 105 | 0.91 | 1.01 | 1.14 | 0.84 | 3.9 |
| 106 | 0.28 | 0.3 | 0.39 | 0.2 | 1.17 |
| 107 | 0 | 0 | 0 | 0.02 | 0.02 |
| 109 | 0.1 | 0.08 | 0.11 | 0.1 | 0.39 |
| 110 | 3.44 | 3.46 | 4.02 | 3.33 | 14.25 |
| 112 | 2.81 | 2.59 | 2.48 | 3.57 | 11.45 |
| 113 | 0.27 | 0.16 | 0.24 | 0.25 | 0.92 |
| 114 | 0.21 | 0.15 | 0.21 | 0.12 | 0.69 |
| 115 | 0.44 | 0.71 | 0.66 | 0.8 | 2.61 |
| 117 | 0.59 | 0.84 | 0.94 | 1.24 | 3.61 |
| 119 | 0 | 0.01 | 0.01 | 0 | 0.02 |
| 121 | 0.15 | 0.3 | 0.12 | 0.03 | 0.6 |
| 122 | 0.9 | 1.06 | 1.25 | 0.86 | 4.07 |
| 123 | 0.04 | 0.19 | 0.05 | 0.08 | 0.36 |
| 124 | 0.04 | 0.04 | 0.02 | 0.02 | 0.12 |
| 125 | 0.05 | 0.02 | 0.05 | 0.04 | 0.16 |
| 126 | 0.34 | 0.23 | 0.32 | 0.12 | 1.01 |
| 127 | 0.06 | 0.04 | 0 | 0.04 | 0.14 |
| 128 | 0.04 | 0.02 | 0.04 | 0.05 | 0.15 |
| 129 | 0 | 0 | 0.01 | 0 | 0.01 |
| 131 | 0.02 | 0 | 0 | 0 | 0.02 |
| 132 | 0.93 | 0.91 | 1.19 | 1.12 | 4.15 |
| 133 | 0.15 | 0.12 | 0.1 | 0.23 | 0.6 |
| 135 | 0.15 | 0.17 | 0.12 | 0.14 | 0.58 |
| 136 | 0.07 | 0.05 | 0.07 | 0.02 | 0.21 |
| 137 | 0.2 | 0.26 | 0.28 | 0.29 | 1.03 |
| 138 | 0.01 | 0.02 | 0.01 | 0.01 | 0.05 |
| 139 | 0 | 0 | 0.02 | 0 | 0.02 |
| 140 | 0.04 | 0.03 | 0.01 | 0.03 | 0.11 |

DRUG CODE:9739DRUG NAME:REMIFENTANIL
STATE:NEW YORK

| ZIP CODE | QUARTER 1 | QUARTER 2 | QUARTER 3 | QUARTER 4 | TOTAL GRAMS |
|---|---|---|---|---|---|
| 142 | 1.03 | 0.63 | 1.04 | 1.21 | 3.91 |
| 144 | 0.02 | 0 | 0.03 | 0.02 | 0.07 |
| 146 | 0.7 | 1.61 | 0.71 | 1.26 | 4.28 |
| 148 | 0.04 | 0.06 | 0.04 | 0.03 | 0.17 |
| TOTAL | 33.29 | 36.84 | 33.61 | 37.43 | 141.17 |

STATE: NORTH CAROLINA

| ZIP CODE | QUARTER 1 | QUARTER 2 | QUARTER 3 | QUARTER 4 | TOTAL GRAMS |
|---|---|---|---|---|---|
| 270 | 0 | 0.02 | 0 | 0.01 | 0.03 |
| 271 | 1.52 | 1.74 | 1.48 | 1.52 | 6.26 |
| 272 | 0.54 | 0.59 | 0.28 | 0.48 | 1.89 |
| 273 | 0.02 | 0 | 0 | 0 | 0.02 |
| 274 | 0.1 | 0.05 | 0.1 | 0.13 | 0.38 |
| 275 | 1.56 | 2.03 | 1.74 | 1.76 | 7.09 |
| 276 | 1.89 | 2.09 | 2.03 | 2.19 | 8.2 |
| 277 | 2.49 | 2.71 | 2.96 | 2.85 | 11.01 |
| 278 | 0.62 | 0.55 | 0.86 | 0.92 | 2.95 |
| 280 | 1.07 | 1.35 | 1.1 | 1.29 | 4.81 |
| 281 | 0.16 | 0.24 | 0.21 | 0.17 | 0.78 |
| 282 | 1.14 | 1.22 | 0.93 | 1.27 | 4.56 |
| 283 | 0.32 | 0.38 | 0.56 | 0.52 | 1.78 |
| 284 | 0.01 | 0 | 0.02 | 0.01 | 0.04 |
| 285 | 0.07 | 0.09 | 0.08 | 0.05 | 0.29 |
| 286 | 1.11 | 1.05 | 1.01 | 1.11 | 4.28 |
| 287 | 0.26 | 0.26 | 0.2 | 0.07 | 0.79 |
| 288 | 0.38 | 0.35 | 0.38 | 0.45 | 1.56 |
| TOTAL | 13.26 | 14.72 | 13.94 | 14.8 | 56.72 |

STATE: NORTH DAKOTA

| ZIP CODE | QUARTER 1 | QUARTER 2 | QUARTER 3 | QUARTER 4 | TOTAL GRAMS |
|---|---|---|---|---|---|
| 581 | 0.21 | 0.27 | 0.32 | 0.34 | 1.14 |
| 585 | 0 | 0.08 | 0.06 | 0.13 | 0.27 |
| 587 | 0.07 | 0.05 | 0 | 0.02 | 0.14 |
| 588 | 0 | 0 | 0.02 | 0.02 | 0.04 |
| TOTAL | 0.28 | 0.4 | 0.4 | 0.51 | 1.59 |

STATE: OHIO

| ZIP CODE | QUARTER 1 | QUARTER 2 | QUARTER 3 | QUARTER 4 | TOTAL GRAMS |
|---|---|---|---|---|---|
| 430 | 0.14 | 0.22 | 0.27 | 0.11 | 0.74 |
| 432 | 2.25 | 2.15 | 2.9 | 3.89 | 11.19 |
| 433 | 0.04 | 0.09 | 0.01 | 0.05 | 0.19 |
| 434 | 0 | 0.01 | 0 | 0.01 | 0.02 |
| 435 | 0.02 | 0.01 | 0 | 0 | 0.03 |
| 436 | 0.33 | 0.45 | 0.41 | 0.45 | 1.64 |
| 437 | 0.4 | 0.38 | 0.22 | 0.73 | 1.73 |
| 438 | 0 | 0 | 0 | 0.02 | 0.02 |
| 440 | 0.27 | 0.18 | 0.16 | 0.31 | 0.92 |
| 441 | 6.33 | 6.43 | 6.34 | 6.09 | 25.19 |
| 442 | 0.1 | 0.13 | 0.1 | 0.17 | 0.5 |
| 443 | 0.72 | 0.85 | 0.86 | 0.71 | 3.14 |
| 444 | 0.1 | 0.1 | 0.09 | 0.08 | 0.37 |
| 445 | 2.33 | 2.11 | 2.29 | 2.23 | 8.96 |
| 446 | 0.04 | 0.02 | 0.04 | 0.03 | 0.13 |
| 447 | 0.04 | 0.05 | 0.04 | 0.07 | 0.2 |
| 448 | 0.06 | 0.03 | 0.03 | 0.03 | 0.15 |
| 449 | 0.05 | 0.02 | 0.04 | 0.05 | 0.16 |

| | | | | | |
|---|---|---|---|---|---|
| 450 | 0.53 | 0.35 | 0.38 | 0.27 | 1.53 |
| 452 | 3.19 | 3.39 | 3.16 | 3.24 | 12.98 |
| 454 | 0.62 | 1.18 | 1.06 | 1.17 | 4.03 |
| 455 | 0.02 | 0 | 0 | 0.01 | 0.03 |
| 456 | 0.22 | 0.2 | 0.22 | 0.27 | 0.91 |
| 457 | 0.21 | 0.21 | 0.35 | 0.3 | 1.07 |
| 458 | 0.02 | 0.06 | 0.13 | 0.04 | 0.25 |
| TOTAL | 18.03 | 18.62 | 19.1 | 20.33 | 76.08 |

STATE: OKLAHOMA

| ZIP CODE | QUARTER 1 | QUARTER 2 | QUARTER 3 | QUARTER 4 | TOTAL GRAMS |
|---|---|---|---|---|---|
| 730 | 0.02 | 0.11 | 0.02 | 0.02 | 0.17 |
| 731 | 2.55 | 1.22 | 1.75 | 1.85 | 7.37 |
| 734 | 0 | 0.04 | 0 | 0 | 0.04 |

DRUG CODE:9739DRUG NAME:REMIFENTANIL
STATE:OKLAHOMA

| ZIP CODE | QUARTER 1 | QUARTER 2 | QUARTER 3 | QUARTER 4 | TOTAL GRAMS |
|---|---|---|---|---|---|
| 735 | 0.02 | 0.05 | 0.04 | 0.03 | 0.14 |
| 740 | 0 | 0 | 0.02 | 0 | 0.02 |
| 741 | 0.53 | 0.47 | 0.43 | 0.53 | 1.96 |
| 743 | 0 | 0.02 | 0 | 0 | 0.02 |
| 744 | 0.03 | 0 | 0 | 0.01 | 0.04 |
| 745 | 0.01 | 0.01 | 0.02 | 0.02 | 0.06 |
| 747 | 0 | 0 | 0.03 | 0 | 0.03 |
| TOTAL | 3.16 | 1.92 | 2.31 | 2.46 | 9.85 |

STATE: OREGON

| ZIP CODE | QUARTER 1 | QUARTER 2 | QUARTER 3 | QUARTER 4 | TOTAL GRAMS |
|---|---|---|---|---|---|
| 970 | 0.66 | 0.29 | 0.26 | 0.39 | 1.6 |
| 971 | 0.04 | 0.21 | 0.07 | 0.14 | 0.46 |
| 972 | 0.57 | 0.6 | 0.4 | 0.43 | 2 |
| 973 | 0.11 | 0.1 | 0.04 | 0.15 | 0.4 |
| 974 | 1.01 | 0.9 | 0.84 | 1 | 3.75 |
| 975 | 0.1 | 0.32 | 0.15 | 0.33 | 0.9 |
| 977 | 0.8 | 0.39 | 0.37 | 0.6 | 2.16 |
| 978 | 0 | 0.01 | 0.01 | 0 | 0.02 |
| 979 | 0 | 0.01 | 0 | 0 | 0.01 |
| TOTAL | 3.29 | 2.83 | 2.14 | 3.04 | 11.3 |

STATE: PENNSYLVANIA

| ZIP CODE | QUARTER 1 | QUARTER 2 | QUARTER 3 | QUARTER 4 | TOTAL GRAMS |
|---|---|---|---|---|---|
| 150 | 0.75 | 0.45 | 0.44 | 0.41 | 2.05 |
| 151 | 0.31 | 0.46 | 0.38 | 0.25 | 1.4 |
| 152 | 7.52 | 8.55 | 8.3 | 7.53 | 31.9 |
| 153 | 0.21 | 0.14 | 0.07 | 0.05 | 0.47 |
| 155 | 0.01 | 0.01 | 0.01 | 0 | 0.03 |
| 156 | 0.21 | 0.09 | 0.19 | 0.18 | 0.67 |
| 158 | 0.07 | 0.1 | 0.12 | 0.14 | 0.43 |
| 159 | 0.16 | 0.07 | 0.14 | 0.08 | 0.45 |
| 160 | 0.35 | 0.04 | 0.11 | 0.26 | 0.76 |
| 161 | 0.03 | 0 | 0.01 | 0 | 0.04 |
| 162 | 0.02 | 0.02 | 0.02 | 0.01 | 0.07 |
| 165 | 0.26 | 0.1 | 0.22 | 0.12 | 0.7 |
| 166 | 0.01 | 0.01 | 0.01 | 0 | 0.03 |
| 168 | 0.09 | 0.04 | 0.07 | 0.09 | 0.29 |
| 170 | 0.85 | 0.83 | 0.97 | 0.79 | 3.44 |
| 171 | 0.07 | 0.07 | 0.11 | 0 | 0.25 |
| 172 | 0 | 0 | 0 | 0.04 | 0.04 |

| | | | | | |
|---|---|---|---|---|---|
| 173 | 0.03 | 0 | 0 | 0.06 | 0.09 |
| 174 | 0.08 | 0.04 | 0.04 | 0.02 | 0.18 |
| 175 | 0.02 | 0.1 | 0.2 | 0 | 0.32 |
| 176 | 0 | 0.02 | 0.05 | 0 | 0.07 |
| 178 | 1.61 | 1.45 | 1.57 | 1.19 | 5.82 |
| 179 | 0.04 | 0.04 | 0.06 | 0.04 | 0.18 |
| 180 | 0.9 | 0.92 | 1.1 | 0.85 | 3.77 |
| 181 | 0.76 | 0.81 | 1.03 | 0.94 | 3.54 |
| 182 | 0 | 0 | 0.01 | 0.01 | 0.02 |
| 183 | 0.15 | 0.2 | 0.2 | 0.27 | 0.82 |
| 184 | 0.09 | 0.08 | 0.07 | 0.06 | 0.3 |
| 185 | 0.12 | 0.04 | 0.17 | 0.34 | 0.67 |
| 186 | 0.01 | 0 | 0 | 0.01 | 0.02 |
| 187 | 0.53 | 0.38 | 0.56 | 0.55 | 2.02 |
| 188 | 0.12 | 0 | 0.05 | 0.14 | 0.31 |
| 189 | 0 | 0.02 | 0 | 0.08 | 0.1 |
| 190 | 2.26 | 1.67 | 2.18 | 1.81 | 7.92 |
| 191 | 15.57 | 17.33 | 15.56 | 16.3 | 64.76 |
| 193 | 0.34 | 0.53 | 0.82 | 0.39 | 2.08 |
| 194 | 0.3 | 0.33 | 0.34 | 0.22 | 1.19 |
| 196 | 0.08 | 0.04 | 0.11 | 0.14 | 0.37 |
| TOTAL | 33.93 | 34.98 | 35.29 | 33.37 | 137.57 |

STATE: PUERTO RICO

| ZIP CODE | QUARTER 1 | QUARTER 2 | QUARTER 3 | QUARTER 4 | TOTAL GRAMS |
|---|---|---|---|---|---|
| 6 | 0 | 0.06 | 0 | 0 | 0.06 |
| 9 | 0 | 0.1 | 0 | 0 | 0.1 |
| TOTAL | 0 | 0.16 | 0 | 0 | 0.16 |

DRUG CODE:9739DRUG NAME:REMIFENTANIL
STATE: RHODE ISLAND

| ZIP CODE | QUARTER 1 | QUARTER 2 | QUARTER 3 | QUARTER 4 | TOTAL GRAMS |
|---|---|---|---|---|---|
| 28 | 0.09 | 0 | 0.13 | 0.01 | 0.23 |
| 29 | 0.38 | 0.32 | 0.41 | 0.51 | 1.62 |
| TOTAL | 0.47 | 0.32 | 0.54 | 0.52 | 1.85 |

STATE: SOUTH CAROLINA

| ZIP CODE | QUARTER 1 | QUARTER 2 | QUARTER 3 | QUARTER 4 | TOTAL GRAMS |
|---|---|---|---|---|---|
| 290 | 0 | 0.02 | 0 | 0.02 | 0.04 |
| 291 | 0.73 | 0.67 | 0.72 | 0.64 | 2.76 |
| 292 | 0.36 | 0.42 | 0.77 | 0.68 | 2.23 |
| 293 | 0.18 | 0.26 | 0.16 | 0.24 | 0.84 |
| 294 | 1.41 | 1.62 | 1.59 | 1.57 | 6.19 |
| 295 | 0.09 | 0.17 | 0.21 | 0.16 | 0.63 |
| 296 | 1.74 | 1.87 | 1.42 | 1.58 | 6.61 |
| 297 | 0.15 | 0.17 | 0.15 | 0.1 | 0.57 |
| 299 | 0.03 | 0.02 | 0 | 0.03 | 0.08 |
| TOTAL | 4.69 | 5.22 | 5.02 | 5.02 | 19.95 |

STATE: SOUTH DAKOTA

| ZIP CODE | QUARTER 1 | QUARTER 2 | QUARTER 3 | QUARTER 4 | TOTAL GRAMS |
|---|---|---|---|---|---|
| 571 | 0.02 | 0.08 | 0.07 | 0.13 | 0.3 |
| 572 | 0 | 0.01 | 0 | 0.01 | 0.02 |
| 573 | 0.02 | 0.01 | 0.03 | 0.01 | 0.07 |
| 574 | 0.01 | 0.06 | 0 | 0.01 | 0.08 |
| 575 | 0 | 0.02 | 0 | 0 | 0.02 |
| 577 | 0.54 | 0.37 | 0.46 | 0.48 | 1.85 |
| TOTAL | 0.59 | 0.55 | 0.56 | 0.64 | 2.34 |

STATE: TENNESSEE

RETAIL DRUG DISTRIBUTION BY ZIP CODE WITHIN STATE BY GRAMS WT

| ZIP CODE | QUARTER 1 | QUARTER 2 | QUARTER 3 | QUARTER 4 | TOTAL GRAMS |
|---|---|---|---|---|---|
| 370 | 0.39 | 0.34 | 0.33 | 0.4 | 1.46 |
| 371 | 0.01 | 0.06 | 0.02 | 0.04 | 0.13 |
| 372 | 2.99 | 3.88 | 3.22 | 3.85 | 13.94 |
| 373 | 0.04 | 0.04 | 0.02 | 0.02 | 0.12 |
| 374 | 0.73 | 0 | 0.04 | 0.32 | 1.09 |
| 376 | 0.1 | 0.09 | 0.09 | 0.02 | 0.3 |
| 378 | 0.19 | 0.06 | 0.01 | 0 | 0.26 |
| 379 | 0.18 | 0.08 | 0.28 | 0.27 | 0.81 |
| 381 | 0.46 | 0.47 | 0.47 | 0.54 | 1.94 |
| 382 | 0.06 | 0 | 0 | 0.02 | 0.08 |
| 383 | 0 | 0.04 | 0.02 | 0.02 | 0.08 |
| 384 | 0.07 | 0.13 | 0.09 | 0.12 | 0.41 |
| 385 | 0.02 | 0 | 0.04 | 0.05 | 0.11 |
| TOTAL | 5.24 | 5.19 | 4.63 | 5.67 | 20.73 |

STATE: TEXAS

| ZIP CODE | QUARTER 1 | QUARTER 2 | QUARTER 3 | QUARTER 4 | TOTAL GRAMS |
|---|---|---|---|---|---|
| 750 | 0.48 | 0.71 | 0.87 | 0.94 | 3 |
| 751 | 0.13 | 0.08 | 0.1 | 0.19 | 0.5 |
| 752 | 2.7 | 2.27 | 2.98 | 1.82 | 9.77 |
| 753 | 3.17 | 2.82 | 3.67 | 3.3 | 12.96 |
| 754 | 0.08 | 0.11 | 0.09 | 0.12 | 0.4 |
| 756 | 0.01 | 0 | 0 | 0 | 0.01 |
| 757 | 0.37 | 0.25 | 0.22 | 0.57 | 1.41 |
| 759 | 0 | 0 | 0 | 0.01 | 0.01 |
| 760 | 0.78 | 0.86 | 0.64 | 0.74 | 3.02 |
| 761 | 3.61 | 4.68 | 4.06 | 4.68 | 17.03 |
| 762 | 0.05 | 0.12 | 0.11 | 0.22 | 0.5 |
| 763 | 0.06 | 0.06 | 0.01 | 0.02 | 0.15 |
| 764 | 0 | 0 | 0.05 | 0 | 0.05 |
| 765 | 0.39 | 0.5 | 0.48 | 0.37 | 1.74 |
| 767 | 0.17 | 0.12 | 0.15 | 0.19 | 0.63 |
| 769 | 0.02 | 0.02 | 0.02 | 0.02 | 0.08 |
| 770 | 4.45 | 5.22 | 5.67 | 5.69 | 21.03 |
| 773 | 0.24 | 0.35 | 0.32 | 0.34 | 1.25 |
| 774 | 0.48 | 0.42 | 0.43 | 0.49 | 1.82 |
| 775 | 1.13 | 1.8 | 1.33 | 1.28 | 5.54 |
| 778 | 0.1 | 0.13 | 0.14 | 0.1 | 0.47 |
| 779 | 0 | 0.01 | 0.02 | 0.01 | 0.04 |
| 780 | 0 | 0 | 0 | 0.01 | 0.01 |
| 781 | 0.05 | 0 | 0.05 | 0.05 | 0.15 |

DRUG CODE:9739DRUG NAME:REMIFENTANIL
STATE:TEXAS

| ZIP CODE | QUARTER 1 | QUARTER 2 | QUARTER 3 | QUARTER 4 | TOTAL GRAMS |
|---|---|---|---|---|---|
| 782 | 2.96 | 2.54 | 2.96 | 2.31 | 10.77 |
| 784 | 0.52 | 0.66 | 0.48 | 0.49 | 2.15 |
| 785 | 0.02 | 0.08 | 0.01 | 0.01 | 0.12 |
| 786 | 0.12 | 0.1 | 0.23 | 0.21 | 0.66 |
| 787 | 1.05 | 0.99 | 0.94 | 1.03 | 4.01 |
| 791 | 0.02 | 0.04 | 0.04 | 0.08 | 0.18 |
| 794 | 1.05 | 1.23 | 1.06 | 1.19 | 4.53 |
| 796 | 0.13 | 0.17 | 0.21 | 0.16 | 0.67 |
| 797 | 0.12 | 0.24 | 0.2 | 0.24 | 0.8 |
| 798 | 0.01 | 0 | 0 | 0.01 | 0.02 |
| 799 | 1.11 | 0.81 | 1.14 | 0.83 | 3.89 |
| TOTAL | 25.58 | 27.39 | 28.68 | 27.72 | 109.37 |

RETAIL DRUG DISTRIBUTION BY ZIP CODE WITHIN STATE BY GRAMS WT

STATE: UTAH

| ZIP CODE | QUARTER 1 | QUARTER 2 | QUARTER 3 | QUARTER 4 | TOTAL GRAMS |
|---|---|---|---|---|---|
| 840 | 1.72 | 1.79 | 1.58 | 2.27 | 7.36 |
| 841 | 9.3 | 10.02 | 9.57 | 9.96 | 38.85 |
| 843 | 0.05 | 0.03 | 0.04 | 0.08 | 0.2 |
| 844 | 0.55 | 0.4 | 0.36 | 0.45 | 1.76 |
| 846 | 1.9 | 2.15 | 2.39 | 2.02 | 8.46 |
| 847 | 2.34 | 2.54 | 2.48 | 2.24 | 9.6 |
| TOTAL | 15.86 | 16.93 | 16.42 | 17.02 | 66.23 |

STATE: VERMONT

| ZIP CODE | QUARTER 1 | QUARTER 2 | QUARTER 3 | QUARTER 4 | TOTAL GRAMS |
|---|---|---|---|---|---|
| 50 | 0.04 | 0.08 | 0.06 | 0.1 | 0.28 |
| 54 | 0.46 | 0.34 | 0.56 | 0.47 | 1.83 |
| 57 | 0 | 0 | 0.03 | 0.02 | 0.05 |
| TOTAL | 0.5 | 0.42 | 0.65 | 0.59 | 2.16 |

STATE: VIRGINIA

| ZIP CODE | QUARTER 1 | QUARTER 2 | QUARTER 3 | QUARTER 4 | TOTAL GRAMS |
|---|---|---|---|---|---|
| 201 | 0.08 | 0.06 | 0.08 | 0.1 | 0.32 |
| 220 | 0.38 | 0.52 | 0.22 | 0.29 | 1.41 |
| 221 | 0.09 | 0.02 | 0 | 0.02 | 0.13 |
| 222 | 0.43 | 0.21 | 0.39 | 0.44 | 1.47 |
| 223 | 0 | 0 | 0.05 | 0.01 | 0.06 |
| 224 | 0 | 0 | 0 | 0.01 | 0.01 |
| 226 | 0.14 | 0.12 | 0.2 | 0.18 | 0.64 |
| 228 | 0.01 | 0.02 | 0 | 0 | 0.03 |
| 229 | 0.3 | 0.41 | 0.35 | 0.2 | 1.26 |
| 231 | 0.01 | 0 | 0 | 0 | 0.01 |
| 232 | 2.14 | 1.84 | 1.85 | 2.4 | 8.23 |
| 233 | 0.15 | 0.09 | 0.19 | 0.11 | 0.54 |
| 234 | 0.21 | 0.31 | 0.21 | 0.26 | 0.99 |
| 235 | 0.57 | 0.57 | 0.85 | 0.71 | 2.7 |
| 236 | 0.54 | 0.48 | 0.45 | 0.51 | 1.98 |
| 237 | 0.46 | 0.24 | 0.12 | 0.12 | 0.94 |
| 238 | 0.1 | 0.05 | 0.02 | 0.15 | 0.32 |
| 239 | 0.01 | 0 | 0.01 | 0 | 0.02 |
| 240 | 0.62 | 0.45 | 0.62 | 0.6 | 2.29 |
| 241 | 0.17 | 0.03 | 0.16 | 0.23 | 0.59 |
| 242 | 0 | 0.01 | 0.02 | 0 | 0.03 |
| 244 | 0 | 0 | 0 | 0.01 | 0.01 |
| 245 | 0.27 | 0.24 | 0.22 | 0.2 | 0.93 |
| TOTAL | 6.68 | 5.67 | 6.01 | 6.55 | 24.91 |

STATE: WASHINGTON

| ZIP CODE | QUARTER 1 | QUARTER 2 | QUARTER 3 | QUARTER 4 | TOTAL GRAMS |
|---|---|---|---|---|---|
| 980 | 0.67 | 0.62 | 0.56 | 0.69 | 2.54 |
| 981 | 8.06 | 8.39 | 8.05 | 9.28 | 33.78 |
| 982 | 0.57 | 0.42 | 1.04 | 0.45 | 2.48 |
| 983 | 0.1 | 0.05 | 0.16 | 0.13 | 0.44 |
| 984 | 1.1 | 0.93 | 0.95 | 0.88 | 3.86 |
| 985 | 0.38 | 0.35 | 0.19 | 0.29 | 1.21 |
| 986 | 0.24 | 0.11 | 0.1 | 0.13 | 0.58 |
| 988 | 0.22 | 0.07 | 0.09 | 0.07 | 0.45 |
| 989 | 0.24 | 0.25 | 0.15 | 0.23 | 0.87 |
| 991 | 0.02 | 0 | 0.01 | 0 | 0.03 |
| 992 | 0.28 | 0.18 | 0.2 | 0.11 | 0.77 |

RETAIL DRUG DISTRIBUTION BY ZIP CODE WITHIN STATE BY GRAMS WT

DRUG CODE:9739DRUG NAME:REMIFENTANIL
STATE:WASHINGTON

| ZIP CODE | QUARTER 1 | QUARTER 2 | QUARTER 3 | QUARTER 4 | TOTAL GRAMS |
|---|---|---|---|---|---|
| 993 | 0.48 | 0.58 | 0.49 | 1 | 2.55 |
| TOTAL | 12.36 | 11.95 | 11.99 | 13.26 | 49.56 |

STATE: WEST VIRGINIA

| ZIP CODE | QUARTER 1 | QUARTER 2 | QUARTER 3 | QUARTER 4 | TOTAL GRAMS |
|---|---|---|---|---|---|
| 247 | 0.02 | 0 | 0.01 | 0 | 0.03 |
| 249 | 0 | 0 | 0 | 0.01 | 0.01 |
| 253 | 1.93 | 2.33 | 2.51 | 2.72 | 9.49 |
| 254 | 0 | 0.04 | 0 | 0.03 | 0.07 |
| 255 | 0.36 | 0.33 | 0.16 | 0.15 | 1 |
| 257 | 0.89 | 0.99 | 0.66 | 0.74 | 3.28 |
| 260 | 0.02 | 0.02 | 0.01 | 0 | 0.05 |
| 261 | 0.06 | 0.1 | 0.12 | 0.11 | 0.39 |
| 263 | 0 | 0 | 0 | 0.05 | 0.05 |
| 265 | 1.48 | 1.15 | 2.06 | 1.81 | 6.5 |
| TOTAL | 4.76 | 4.96 | 5.53 | 5.62 | 20.87 |

STATE: WISCONSIN

| ZIP CODE | QUARTER 1 | QUARTER 2 | QUARTER 3 | QUARTER 4 | TOTAL GRAMS |
|---|---|---|---|---|---|
| 530 | 0.32 | 0.38 | 0.25 | 0.29 | 1.24 |
| 531 | 0.1 | 0.07 | 0.1 | 0.19 | 0.46 |
| 532 | 2.46 | 2.32 | 2.85 | 2.95 | 10.58 |
| 534 | 0.09 | 0.21 | 0.19 | 0.26 | 0.75 |
| 535 | 0.29 | 0.3 | 0.28 | 0.34 | 1.21 |
| 537 | 1.32 | 1.25 | 1.05 | 1.35 | 4.97 |
| 539 | 0.04 | 0.01 | 0 | 0 | 0.05 |
| 540 | 0 | 0 | 0.02 | 0 | 0.02 |
| 543 | 0.1 | 0.08 | 0.08 | 0.15 | 0.41 |
| 544 | 0.6 | 0.51 | 0.66 | 0.54 | 2.31 |
| 545 | 0.04 | 0.04 | 0.06 | 0.02 | 0.16 |
| 546 | 0.09 | 0.08 | 0.11 | 0.07 | 0.35 |
| 547 | 0.03 | 0.03 | 0.02 | 0.02 | 0.1 |
| 548 | 0 | 0.01 | 0 | 0 | 0.01 |
| 549 | 0.37 | 0.42 | 0.43 | 0.46 | 1.68 |
| TOTAL | 5.85 | 5.71 | 6.1 | 6.64 | 24.3 |

STATE: WYOMING

| ZIP CODE | QUARTER 1 | QUARTER 2 | QUARTER 3 | QUARTER 4 | TOTAL GRAMS |
|---|---|---|---|---|---|
| 820 | 0.14 | 0.03 | 0.05 | 0.1 | 0.32 |
| 822 | 0.01 | 0 | 0 | 0.01 | 0.02 |
| 823 | 0 | 0.02 | 0.04 | 0 | 0.06 |
| 824 | 0.02 | 0.01 | 0.02 | 0.01 | 0.06 |
| 826 | 0.13 | 0.19 | 0.12 | 0.18 | 0.62 |
| 827 | 0.03 | 0.06 | 0.03 | 0.04 | 0.16 |
| 828 | 0 | 0.01 | 0 | 0 | 0.01 |
| 829 | 0 | 0.01 | 0.01 | 0.01 | 0.03 |
| 830 | 0.1 | 0.1 | 0.08 | 0.1 | 0.38 |
| 831 | 0.07 | 0.05 | 0.07 | 0.03 | 0.22 |
| TOTAL | 0.5 | 0.48 | 0.42 | 0.48 | 1.88 |

DRUG CODE:9740DRUG NAME:SUFENTANIL BASE
STATE: ALABAMA

| ZIP CODE | QUARTER 1 | QUARTER 2 | QUARTER 3 | QUARTER 4 | TOTAL GRAMS |
|---|---|---|---|---|---|
| 350 | 0 | 0 | 0 | 0 | 0 |
| 352 | 0.1 | 0.06 | 0.04 | 0.04 | 0.24 |
| 354 | 0 | 0 | 0 | 0 | 0 |
| 356 | 0.01 | 0.01 | 0.01 | 0.01 | 0.04 |

| | | | | | |
|---|---|---|---|---|---|
| 357 | 0.01 | 0 | 0 | 0 | 0.01 |
| 358 | 1.86 | 1.72 | 1.05 | 1.71 | 6.34 |
| 359 | 0.05 | 0.01 | 0.03 | 0.01 | 0.1 |
| 360 | 0.01 | 0.01 | 0.01 | 0.01 | 0.04 |
| 361 | 0.04 | 0.02 | 0.02 | 0.03 | 0.11 |
| 363 | 0.1 | 0.05 | 0.1 | 0.06 | 0.31 |
| 364 | 0 | 0 | 0 | 0 | 0 |
| 365 | 0.03 | 0.02 | 0.02 | 0.02 | 0.09 |
| 366 | 0.19 | 0.15 | 0.13 | 0.15 | 0.62 |
| 367 | 0 | 0 | 0 | 0 | 0 |
| TOTAL | 2.4 | 2.05 | 1.41 | 2.04 | 7.9 |

STATE: ALASKA

| ZIP CODE | QUARTER 1 | QUARTER 2 | QUARTER 3 | QUARTER 4 | TOTAL GRAMS |
|---|---|---|---|---|---|

DRUG CODE:9740DRUG NAME:SUFENTANIL BASE
STATE:ALASKA

| ZIP CODE | QUARTER 1 | QUARTER 2 | QUARTER 3 | QUARTER 4 | TOTAL GRAMS |
|---|---|---|---|---|---|
| 995 | 0.01 | 0.01 | 0 | 0.01 | 0.03 |
| 996 | 0 | 0 | 0 | 0 | 0 |
| 997 | 0 | 0 | 0 | 0 | 0 |
| 998 | 0 | 0 | 0 | 0 | 0 |
| TOTAL | 0.01 | 0.01 | 0 | 0.01 | 0.03 |

STATE: ARIZONA

| ZIP CODE | QUARTER 1 | QUARTER 2 | QUARTER 3 | QUARTER 4 | TOTAL GRAMS |
|---|---|---|---|---|---|
| 850 | 0.04 | 0.03 | 0.02 | 0.02 | 0.11 |
| 852 | 0.01 | 0.01 | 0.03 | 0 | 0.05 |
| 853 | 0.02 | 0 | 0.01 | 0.01 | 0.04 |
| 857 | 0.02 | 0.02 | 0.01 | 0.02 | 0.07 |
| 859 | 0 | 0 | 0 | 0 | 0 |
| 860 | 0 | 0 | 0 | 0 | 0 |
| 864 | 0.01 | 0 | 0 | 0 | 0.01 |
| 865 | 0 | 0 | 0 | 0 | 0 |
| TOTAL | 0.1 | 0.06 | 0.07 | 0.05 | 0.28 |

STATE: ARKANSAS

| ZIP CODE | QUARTER 1 | QUARTER 2 | QUARTER 3 | QUARTER 4 | TOTAL GRAMS |
|---|---|---|---|---|---|
| 716 | 0.01 | 0 | 0.01 | 0 | 0.02 |
| 717 | 0.01 | 0.01 | 0.01 | 0 | 0.03 |
| 719 | 0 | 0 | 0 | 0 | 0 |
| 720 | 0 | 0 | 0.01 | 0 | 0.01 |
| 721 | 0 | 0 | 0 | 0 | 0 |
| 722 | 0.09 | 0.06 | 0.07 | 0.09 | 0.31 |
| 723 | 0 | 0 | 0 | 0 | 0 |
| 724 | 0.04 | 0.02 | 0.03 | 0.02 | 0.11 |
| 725 | 0 | 0 | 0 | 0 | 0 |
| 727 | 0.06 | 0.02 | 0.03 | 0.01 | 0.12 |
| 728 | 0 | 0 | 0 | 0 | 0 |
| 729 | 0.03 | 0 | 0 | 0.01 | 0.04 |
| TOTAL | 0.24 | 0.11 | 0.16 | 0.13 | 0.64 |

STATE: CALIFORNIA

| ZIP CODE | QUARTER 1 | QUARTER 2 | QUARTER 3 | QUARTER 4 | TOTAL GRAMS |
|---|---|---|---|---|---|
| 900 | 0.19 | 0.07 | 0.1 | 0.13 | 0.49 |
| 902 | 0.01 | 0.01 | 0.02 | 0.01 | 0.05 |
| 904 | 0.01 | 0.02 | 0 | 0.01 | 0.04 |
| 905 | 0.01 | 0.01 | 0.01 | 0.01 | 0.04 |
| 906 | 0.01 | 0 | 0 | 0 | 0.01 |
| 907 | 0.02 | 0 | 0.01 | 0 | 0.03 |

| | | | | | |
|---|---|---|---|---|---|
| 908 | 7.35 | 3.35 | 9.36 | 10.03 | 30.09 |
| 910 | 0 | 0 | 0 | 0 | 0 |
| 911 | 0.01 | 0.01 | 0 | 0 | 0.02 |
| 913 | 0.01 | 0.01 | 0.01 | 0.01 | 0.04 |
| 917 | 0.02 | 0.01 | 0.01 | 0.01 | 0.05 |
| 919 | 0 | 0 | 0 | 0 | 0 |
| 920 | 0.02 | 0.03 | 0.02 | 0.02 | 0.09 |
| 921 | 0.06 | 0.02 | 0.06 | 0.04 | 0.18 |
| 922 | 0.02 | 0.02 | 0.02 | 0.02 | 0.08 |
| 923 | 0.03 | 0.02 | 0.01 | 0.02 | 0.08 |
| 925 | 1.19 | 1.31 | 1.38 | 1.08 | 4.96 |
| 926 | 0.04 | 0.01 | 0.02 | 0.02 | 0.09 |
| 927 | 0.01 | 0 | 0 | 0 | 0.01 |
| 928 | 0.01 | 0.01 | 0.01 | 0.02 | 0.05 |
| 930 | 0.02 | 0.01 | 0 | 0 | 0.03 |
| 931 | 0.01 | 0 | 0.01 | 0.01 | 0.03 |
| 932 | 0.02 | 0.01 | 0.01 | 0 | 0.04 |
| 933 | 0.34 | 0 | 0 | 0 | 0.34 |
| 934 | 0.02 | 0.02 | 0.03 | 0.01 | 0.08 |
| 935 | 0 | 0 | 0 | 0.01 | 0.01 |
| 936 | 0.01 | 0 | 0.05 | 0 | 0.06 |
| 937 | 0.03 | 0.01 | 0.01 | 0.02 | 0.07 |
| 939 | 0.01 | 0.01 | 0 | 0 | 0.02 |
| 940 | 0.01 | 0 | 0 | 0 | 0.01 |
| 941 | 0.16 | 0.12 | 0.13 | 0.16 | 0.57 |
| 943 | 0.14 | 0.13 | 0.12 | 0.14 | 0.53 |
| 944 | 0 | 0 | 0 | 0 | 0 |

DRUG CODE:9740DRUG NAME:SUFENTANIL BASE
STATE:CALIFORNIA

| ZIP CODE | QUARTER 1 | QUARTER 2 | QUARTER 3 | QUARTER 4 | TOTAL GRAMS |
|---|---|---|---|---|---|
| 945 | 0.02 | 0.03 | 0.01 | 0.01 | 0.07 |
| 946 | 0.01 | 0.01 | 0.01 | 0 | 0.03 |
| 948 | 0.01 | 0 | 0 | 0 | 0.01 |
| 949 | 0.01 | 0 | 0 | 0 | 0.01 |
| 950 | 0.01 | 0 | 0 | 0.01 | 0.02 |
| 951 | 0.04 | 0.05 | 0.08 | 0.09 | 0.26 |
| 952 | 0.01 | 0.01 | 0.02 | 0 | 0.04 |
| 953 | 0.02 | 0.01 | 0.01 | 0.02 | 0.06 |
| 954 | 0 | 0 | 0 | 0 | 0 |
| 955 | 0.02 | 0.02 | 0.01 | 0.01 | 0.06 |
| 956 | 0.03 | 0.04 | 0.03 | 0.05 | 0.15 |
| 958 | 0.08 | 0.06 | 0.06 | 0.05 | 0.25 |
| 959 | 0 | 0 | 0 | 0 | 0 |
| 960 | 0.02 | 0.01 | 0.01 | 0.01 | 0.05 |
| 961 | 0 | 0 | 0 | 0 | 0 |
| TOTAL | 10.07 | 5.46 | 11.64 | 12.03 | 39.2 |

STATE: COLORADO

| ZIP CODE | QUARTER 1 | QUARTER 2 | QUARTER 3 | QUARTER 4 | TOTAL GRAMS |
|---|---|---|---|---|---|
| 800 | 0.1 | 0.05 | 0.05 | 0.04 | 0.24 |
| 801 | 0.02 | 0 | 0.01 | 0.01 | 0.04 |
| 802 | 0.02 | 0.03 | 0.03 | 0.1 | 0.18 |
| 803 | 0 | 0 | 0.01 | 0 | 0.01 |
| 805 | 0 | 0 | 0 | 0 | 0 |
| 806 | 0.01 | 0.01 | 0.01 | 0.01 | 0.04 |
| 807 | 0 | 0 | 0 | 0 | 0 |
| 809 | 0.07 | 0.02 | 0.06 | 0.05 | 0.2 |
| 810 | 0 | 0 | 0 | 0 | 0 |
| 811 | 0 | 0 | 0 | 0 | 0 |

| | | | | | |
|---|---|---|---|---|---|
| 812 | 0 | 0 | 0 | 0 | 0 |
| 814 | 0 | 0 | 0 | 0 | 0 |
| 815 | 0.03 | 0.03 | 0.02 | 0.02 | 0.1 |
| 816 | 0 | 0 | 0 | 0 | 0 |
| TOTAL | 0.25 | 0.14 | 0.19 | 0.23 | 0.81 |

STATE: CONNECTICUT

| ZIP CODE | QUARTER 1 | QUARTER 2 | QUARTER 3 | QUARTER 4 | TOTAL GRAMS |
|---|---|---|---|---|---|
| 60 | 0.01 | 0 | 0 | 0.01 | 0.02 |
| 61 | 0.01 | 0.02 | 0.01 | 0.05 | 0.09 |
| 63 | 0 | 0 | 0 | 0.01 | 0.01 |
| 65 | 0.12 | 0.13 | 0.11 | 0.15 | 0.51 |
| 67 | 0 | 0 | 0 | 0 | 0 |
| 68 | 0.01 | 0 | 0.67 | 0 | 0.68 |
| TOTAL | 0.15 | 0.15 | 0.79 | 0.22 | 1.31 |

STATE: DELAWARE

| ZIP CODE | QUARTER 1 | QUARTER 2 | QUARTER 3 | QUARTER 4 | TOTAL GRAMS |
|---|---|---|---|---|---|
| 198 | 0.03 | 0 | 0.01 | 0 | 0.04 |
| 199 | 0 | 0 | 0 | 0 | 0 |
| TOTAL | 0.03 | 0 | 0.01 | 0 | 0.04 |

STATE: DISTRICT OF COLUMBIA

| ZIP CODE | QUARTER 1 | QUARTER 2 | QUARTER 3 | QUARTER 4 | TOTAL GRAMS |
|---|---|---|---|---|---|
| 200 | 0.01 | 0.01 | 0 | 0.02 | 0.04 |
| TOTAL | 0.01 | 0.01 | 0 | 0.02 | 0.04 |

STATE: FLORIDA

| ZIP CODE | QUARTER 1 | QUARTER 2 | QUARTER 3 | QUARTER 4 | TOTAL GRAMS |
|---|---|---|---|---|---|
| 320 | 0 | 0 | 0 | 0 | 0 |
| 321 | 0 | 0.01 | 0.03 | 0 | 0.04 |
| 322 | 0.1 | 0.05 | 0.06 | 0.04 | 0.25 |
| 323 | 0.02 | 0.02 | 0.01 | 0.01 | 0.06 |
| 324 | 0.08 | 0.06 | 0.04 | 0.05 | 0.23 |
| 325 | 0.04 | 0 | 0.01 | 0.03 | 0.08 |
| 326 | 0.09 | 0.08 | 0.06 | 0.05 | 0.28 |
| 327 | 0.01 | 0 | 0.01 | 0 | 0.02 |
| 328 | 0.08 | 0.07 | 0.07 | 0.08 | 0.3 |
| 329 | 0.02 | 2.02 | 0.01 | 0.68 | 2.73 |
| 330 | 0.01 | 0 | 0 | 0 | 0.01 |
| 331 | 0.14 | 0.1 | 0.08 | 0.12 | 0.44 |
| 333 | 0 | 0 | 0.01 | 0 | 0.01 |

DRUG CODE:9740DRUG NAME:SUFENTANIL BASE
STATE:FLORIDA

| ZIP CODE | QUARTER 1 | QUARTER 2 | QUARTER 3 | QUARTER 4 | TOTAL GRAMS |
|---|---|---|---|---|---|
| 334 | 0.01 | 0.01 | 0.02 | 0.01 | 0.05 |
| 336 | 1.52 | 0.83 | 1.85 | 1.4 | 5.6 |
| 337 | 0.03 | 0.02 | 0.01 | 0.02 | 0.08 |
| 339 | 0.01 | 0.03 | 0 | 0 | 0.04 |
| 342 | 0 | 0 | 0 | 0 | 0 |
| 344 | 0.01 | 0.01 | 0.03 | 0.02 | 0.07 |
| 346 | 0.04 | 0.02 | 0.02 | 0.01 | 0.09 |
| 347 | 0.03 | 0.02 | 0.02 | 0.02 | 0.09 |
| 349 | 0 | 0 | 0 | 0.01 | 0.01 |
| TOTAL | 2.24 | 3.35 | 2.34 | 2.55 | 10.48 |

STATE: GEORGIA

| ZIP CODE | QUARTER 1 | QUARTER 2 | QUARTER 3 | QUARTER 4 | TOTAL GRAMS |
|---|---|---|---|---|---|
| 300 | 0.05 | 0.05 | 0.04 | 0.05 | 0.19 |

| | | | | | |
|---|---|---|---|---|---|
| 301 | 0.09 | 0.07 | 0.09 | 0.05 | 0.3 |
| 302 | 0 | 0 | 0 | 0 | 0 |
| 303 | 0.01 | 0.01 | 0.01 | 0.01 | 0.04 |
| 304 | 0 | 0 | 0 | 0 | 0 |
| 305 | 0.05 | 0.04 | 0.04 | 0.05 | 0.18 |
| 306 | 0.02 | 0.03 | 0.03 | 0.03 | 0.11 |
| 309 | 0.04 | 0.03 | 0.03 | 0.06 | 0.16 |
| 310 | 0.07 | 0.03 | 0.03 | 0.02 | 0.15 |
| 312 | 0.08 | 0.08 | 0.07 | 0.07 | 0.3 |
| 313 | 0 | 0 | 0 | 0 | 0 |
| 315 | 0 | 0 | 0 | 0 | 0 |
| 317 | 0.03 | 0 | 0 | 0 | 0.03 |
| 319 | 0 | 0 | 0 | 0 | 0 |
| 398 | 0 | 0 | 0 | 0 | 0 |
| TOTAL | 0.44 | 0.34 | 0.34 | 0.34 | 1.46 |

STATE: HAWAII

| ZIP CODE | QUARTER 1 | QUARTER 2 | QUARTER 3 | QUARTER 4 | TOTAL GRAMS |
|---|---|---|---|---|---|
| 967 | 0 | 0 | 0 | 0 | 0 |
| 968 | 0.02 | 0.01 | 0.01 | 0.01 | 0.05 |
| TOTAL | 0.02 | 0.01 | 0.01 | 0.01 | 0.05 |

STATE: IDAHO

| ZIP CODE | QUARTER 1 | QUARTER 2 | QUARTER 3 | QUARTER 4 | TOTAL GRAMS |
|---|---|---|---|---|---|
| 832 | 0 | 0 | 0 | 0 | 0 |
| 833 | 0.01 | 0 | 0 | 0 | 0.01 |
| 834 | 0.02 | 0.01 | 0.01 | 0.01 | 0.05 |
| 835 | 0 | 0 | 0 | 0 | 0 |
| 836 | 0.01 | 0 | 0 | 0 | 0.01 |
| 837 | 0.03 | 0.03 | 0.01 | 0.01 | 0.08 |
| 838 | 0 | 0 | 0 | 0 | 0 |
| TOTAL | 0.07 | 0.04 | 0.02 | 0.02 | 0.15 |

STATE: ILLINOIS

| ZIP CODE | QUARTER 1 | QUARTER 2 | QUARTER 3 | QUARTER 4 | TOTAL GRAMS |
|---|---|---|---|---|---|
| 600 | 0.06 | 0.03 | 0.04 | 0.04 | 0.17 |
| 601 | 0.11 | 0.03 | 0.05 | 0.02 | 0.21 |
| 602 | 0.07 | 0.05 | 0.06 | 0.05 | 0.23 |
| 603 | 0.01 | 0.01 | 0.01 | 0.01 | 0.04 |
| 604 | 0.11 | 0.07 | 0.06 | 0.08 | 0.32 |
| 605 | 0.01 | 0 | 0.02 | 0.01 | 0.04 |
| 606 | 0.26 | 0.05 | 0.11 | 0.08 | 0.5 |
| 609 | 0 | 0 | 0 | 0 | 0 |
| 610 | 0 | 0 | 0 | 0 | 0 |
| 611 | 0.01 | 0 | 0 | 0.01 | 0.02 |
| 612 | 0.01 | 0.01 | 0.01 | 0.01 | 0.04 |
| 613 | 0 | 0 | 0 | 0 | 0 |
| 614 | 0 | 0 | 0 | 0 | 0 |
| 615 | 0.01 | 0 | 0 | 0.01 | 0.02 |
| 616 | 0.04 | 0.01 | 0.01 | 0.01 | 0.07 |
| 617 | 0 | 0 | 0 | 0 | 0 |
| 618 | 0.03 | 0 | 0 | 0 | 0.03 |
| 619 | 0 | 0 | 0 | 0 | 0 |
| 620 | 0 | 0 | 0 | 0 | 0 |
| 622 | 0.02 | 0.07 | 0.07 | 0.07 | 0.23 |
| 623 | 0 | 0 | 0 | 0.01 | 0.01 |

DRUG CODE:9740DRUG NAME:SUFENTANIL BASE
STATE:ILLINOIS

| ZIP CODE | QUARTER 1 | QUARTER 2 | QUARTER 3 | QUARTER 4 | TOTAL GRAMS |
|---|---|---|---|---|---|

| | | | | | |
|---|---|---|---|---|---|
| 624 | 0.01 | 0 | 0 | 0 | 0.01 |
| 625 | 0 | 0 | 0 | 0 | 0 |
| 627 | 0.03 | 0.02 | 0.01 | 0.01 | 0.07 |
| 628 | 0.01 | 0.01 | 0 | 0 | 0.02 |
| 629 | 0.07 | 0 | 0 | 0.03 | 0.1 |
| TOTAL | 0.87 | 0.36 | 0.45 | 0.45 | 2.13 |

STATE: INDIANA

| ZIP CODE | QUARTER 1 | QUARTER 2 | QUARTER 3 | QUARTER 4 | TOTAL GRAMS |
|---|---|---|---|---|---|
| 460 | 0.01 | 0.03 | 0 | 0.01 | 0.05 |
| 461 | 0 | 0 | 0 | 0 | 0 |
| 462 | 0.31 | 0.33 | 0.23 | 0.25 | 1.12 |
| 463 | 0.02 | 0.02 | 0.01 | 0.03 | 0.08 |
| 464 | 0 | 0 | 0 | 0 | 0 |
| 465 | 0.02 | 0 | 0.01 | 0 | 0.03 |
| 466 | 0 | 0.01 | 0 | 0.01 | 0.02 |
| 468 | 0.05 | 0.01 | 0.01 | 0.01 | 0.08 |
| 471 | 0.01 | 0 | 0 | 0 | 0.01 |
| 472 | 0.01 | 0 | 0 | 0 | 0.01 |
| 473 | 0 | 0 | 0 | 0 | 0 |
| 474 | 0.01 | 0.01 | 0 | 0 | 0.02 |
| 475 | 0 | 0 | 0 | 0 | 0 |
| 476 | 0 | 0 | 0 | 0 | 0 |
| 477 | 0.01 | 0.01 | 0 | 0 | 0.02 |
| 478 | 0.02 | 0.01 | 0.01 | 0.02 | 0.06 |
| 479 | 0 | 0.01 | 0 | 0 | 0.01 |
| TOTAL | 0.47 | 0.44 | 0.27 | 0.33 | 1.51 |

STATE: IOWA

| ZIP CODE | QUARTER 1 | QUARTER 2 | QUARTER 3 | QUARTER 4 | TOTAL GRAMS |
|---|---|---|---|---|---|
| 500 | 0 | 0 | 0 | 0 | 0 |
| 502 | 0 | 0 | 0 | 0 | 0 |
| 503 | 0.01 | 0 | 0.01 | 0 | 0.02 |
| 505 | 0 | 0 | 0 | 0 | 0 |
| 506 | 0 | 0 | 0 | 0 | 0 |
| 507 | 0 | 0 | 0 | 0 | 0 |
| 513 | 0.01 | 0.01 | 0 | 0.01 | 0.03 |
| 514 | 0 | 0 | 0 | 0 | 0 |
| 515 | 0 | 0 | 0 | 0 | 0 |
| 520 | 0.01 | 0 | 0 | 0 | 0.01 |
| 524 | 0.01 | 0.01 | 0.01 | 0.01 | 0.04 |
| 525 | 0 | 0 | 0 | 0 | 0 |
| 526 | 0 | 0 | 0 | 0 | 0 |
| 527 | 0 | 0 | 0 | 0 | 0 |
| 528 | 0 | 0.01 | 0.01 | 0.01 | 0.03 |
| TOTAL | 0.04 | 0.03 | 0.03 | 0.03 | 0.13 |

STATE: KANSAS

| ZIP CODE | QUARTER 1 | QUARTER 2 | QUARTER 3 | QUARTER 4 | TOTAL GRAMS |
|---|---|---|---|---|---|
| 660 | 0 | 0 | 0.01 | 0 | 0.01 |
| 661 | 0.02 | 0.01 | 0.01 | 0.02 | 0.06 |
| 662 | 0.08 | 0.06 | 0.12 | 0.05 | 0.31 |
| 664 | 0.01 | 0 | 0 | 0 | 0.01 |
| 665 | 0 | 0 | 0 | 0 | 0 |
| 666 | 0.03 | 0.03 | 0.03 | 0.03 | 0.12 |
| 667 | 0.01 | 0.01 | 0.02 | 0.02 | 0.06 |
| 668 | 0 | 0 | 0 | 0 | 0 |
| 669 | 0 | 0 | 0 | 0 | 0 |
| 670 | 0.02 | 0.01 | 0 | 0 | 0.03 |
| 671 | 0.01 | 0.01 | 0.01 | 0.01 | 0.04 |
| 672 | 0.1 | 0.06 | 0.07 | 0.07 | 0.3 |
| 674 | 0.01 | 0 | 0.01 | 0.01 | 0.03 |

| | | | | | |
|---|---|---|---|---|---|
| 675 | 0.01 | 0.01 | 0 | 0.01 | 0.03 |
| 676 | 0.01 | 0 | 0 | 0 | 0.01 |
| 678 | 0 | 0 | 0 | 0 | 0 |
| 679 | 0 | 0 | 0 | 0 | 0 |
| TOTAL | 0.31 | 0.2 | 0.28 | 0.22 | 1.01 |

STATE: KENTUCKY

| ZIP CODE | QUARTER 1 | QUARTER 2 | QUARTER 3 | QUARTER 4 | TOTAL GRAMS |
|---|---|---|---|---|---|

DRUG CODE:9740DRUG NAME:SUFENTANIL BASE
STATE:KENTUCKY

| ZIP CODE | QUARTER 1 | QUARTER 2 | QUARTER 3 | QUARTER 4 | TOTAL GRAMS |
|---|---|---|---|---|---|
| 400 | 0.02 | 0.01 | 0.01 | 0.01 | 0.05 |
| 402 | 0.05 | 0.02 | 0.04 | 0.04 | 0.15 |
| 403 | 0 | 0 | 0 | 0 | 0 |
| 404 | 0 | 0 | 0 | 0 | 0 |
| 405 | 0.11 | 0.06 | 0.06 | 0.05 | 0.28 |
| 407 | 0 | 0 | 0 | 0 | 0 |
| 409 | 0 | 0 | 0 | 0 | 0 |
| 410 | 0 | 0 | 0 | 0 | 0 |
| 411 | 0.01 | 0.02 | 0.02 | 0.01 | 0.06 |
| 413 | 0 | 0 | 0 | 0 | 0 |
| 420 | 0.15 | 0.13 | 0.12 | 0.11 | 0.51 |
| 421 | 0.03 | 0.02 | 0.04 | 0.03 | 0.12 |
| 422 | 0.01 | 0 | 0.01 | 0 | 0.02 |
| 423 | 0 | 0 | 0 | 0 | 0 |
| 424 | 0 | 0 | 0 | 0 | 0 |
| 425 | 0.01 | 0 | 0 | 0 | 0.01 |
| 427 | 0 | 0 | 0 | 0 | 0 |
| TOTAL | 0.39 | 0.26 | 0.3 | 0.25 | 1.2 |

STATE: LOUISIANA

| ZIP CODE | QUARTER 1 | QUARTER 2 | QUARTER 3 | QUARTER 4 | TOTAL GRAMS |
|---|---|---|---|---|---|
| 700 | 0.03 | 0 | 0.02 | 0.01 | 0.06 |
| 701 | 0.08 | 0.07 | 0.07 | 0.07 | 0.29 |
| 703 | 0 | 0 | 0 | 0 | 0 |
| 704 | 0.08 | 0.06 | 0.06 | 0.07 | 0.27 |
| 705 | 0.72 | 0.38 | 0.38 | 0.37 | 1.85 |
| 706 | 0 | 0.01 | 0.01 | 0 | 0.02 |
| 707 | 0.02 | 0.05 | 0.08 | 0.13 | 0.28 |
| 708 | 0.05 | 0.01 | 0.03 | 0.01 | 0.1 |
| 711 | 0.14 | 0.07 | 0.07 | 0.06 | 0.34 |
| 712 | 0.05 | 0.05 | 0.03 | 0.05 | 0.18 |
| 714 | 0 | 0 | 0 | 0 | 0 |
| TOTAL | 1.17 | 0.7 | 0.75 | 0.77 | 3.39 |

STATE: MAINE

| ZIP CODE | QUARTER 1 | QUARTER 2 | QUARTER 3 | QUARTER 4 | TOTAL GRAMS |
|---|---|---|---|---|---|
| 40 | 0 | 0 | 0 | 0 | 0 |
| 42 | 0 | 0 | 0 | 0 | 0 |
| 43 | 0 | 0 | 0 | 0 | 0 |
| 44 | 0 | 0.01 | 0 | 0 | 0.01 |
| 46 | 0 | 0 | 0 | 0 | 0 |
| 47 | 0 | 0 | 0 | 0 | 0 |
| TOTAL | 0 | 0.01 | 0 | 0 | 0.01 |

STATE: MARYLAND

| ZIP CODE | QUARTER 1 | QUARTER 2 | QUARTER 3 | QUARTER 4 | TOTAL GRAMS |
|---|---|---|---|---|---|
| 207 | 0 | 0 | 0 | 0 | 0 |
| 208 | 0.02 | 0.01 | 0.01 | 0 | 0.04 |

| | | | | | |
|---|---|---|---|---|---|
| 209 | 0 | 0 | 0 | 0 | 0 |
| 210 | 0 | 0 | 0 | 0 | 0 |
| 212 | 0.02 | 0.02 | 0.03 | 0.02 | 0.09 |
| 214 | 0 | 0 | 0 | 0 | 0 |
| 215 | 0 | 0 | 0 | 0 | 0 |
| 216 | 0 | 0 | 0 | 0 | 0 |
| 217 | 0.01 | 0 | 0 | 0 | 0.01 |
| TOTAL | 0.05 | 0.03 | 0.04 | 0.02 | 0.14 |

STATE: MASSACHUSETTS

| ZIP CODE | QUARTER 1 | QUARTER 2 | QUARTER 3 | QUARTER 4 | TOTAL GRAMS |
|---|---|---|---|---|---|
| 10 | 0.03 | 0.02 | 0.02 | 0.02 | 0.09 |
| 11 | 0 | 0 | 0 | 0 | 0 |
| 14 | 0 | 0 | 0 | 0 | 0 |
| 16 | 0.01 | 0.01 | 0 | 0.01 | 0.03 |
| 18 | 0 | 0.01 | 0.32 | 0 | 0.33 |
| 21 | 0.09 | 0.07 | 0.07 | 0.07 | 0.3 |
| 22 | 0 | 0 | 0 | 0 | 0 |
| 25 | 0 | 0 | 0 | 0 | 0 |
| 27 | 0.01 | 0.02 | 0.01 | 0.01 | 0.05 |
| TOTAL | 0.14 | 0.13 | 0.42 | 0.11 | 0.8 |

STATE: MICHIGAN

DRUG CODE:9740DRUG NAME:SUFENTANIL BASE
STATE:MICHIGAN

| ZIP CODE | QUARTER 1 | QUARTER 2 | QUARTER 3 | QUARTER 4 | TOTAL GRAMS |
|---|---|---|---|---|---|
| 480 | 0.21 | 0.1 | 0.1 | 0.12 | 0.53 |
| 481 | 0.1 | 0.05 | 0.07 | 0.05 | 0.27 |
| 482 | 0.08 | 0 | 0.02 | 0.06 | 0.16 |
| 483 | 0.04 | 0.25 | 0.27 | 0.2 | 0.76 |
| 484 | 0.02 | 0.02 | 0.04 | 0.01 | 0.09 |
| 485 | 0.01 | 0 | 0 | 0 | 0.01 |
| 486 | 0.01 | 0.01 | 0.01 | 0.01 | 0.04 |
| 488 | 0.03 | 0.02 | 0 | 0 | 0.05 |
| 489 | 0.02 | 0 | 0 | 0 | 0.02 |
| 490 | 0.02 | 0.02 | 0 | 0 | 0.04 |
| 492 | 0.01 | 0.01 | 0.01 | 0.01 | 0.04 |
| 494 | 0 | 0.01 | 0 | 0 | 0.01 |
| 495 | 0.02 | 0 | 0.01 | 0 | 0.03 |
| 496 | 0.07 | 0.06 | 0.06 | 0.06 | 0.25 |
| 497 | 0 | 0 | 0 | 0 | 0 |
| 498 | 0 | 0.01 | 0.01 | 0.01 | 0.03 |
| 499 | 0 | 0.01 | 0 | 0 | 0.01 |
| TOTAL | 0.64 | 0.57 | 0.6 | 0.53 | 2.34 |

STATE: MINNESOTA

| ZIP CODE | QUARTER 1 | QUARTER 2 | QUARTER 3 | QUARTER 4 | TOTAL GRAMS |
|---|---|---|---|---|---|
| 551 | 0 | 0 | 0 | 0 | 0 |
| 553 | 0.01 | 0 | 0 | 0 | 0.01 |
| 554 | 0.02 | 0.01 | 0.05 | 0.02 | 0.1 |
| 557 | 0 | 0 | 0 | 0 | 0 |
| 558 | 0 | 0 | 0.01 | 0 | 0.01 |
| 559 | 0.16 | 0.06 | 0.12 | 0.13 | 0.47 |
| 560 | 0 | 0 | 0 | 0 | 0 |
| 561 | 0 | 0 | 0 | 0 | 0 |
| 562 | 0.01 | 0.01 | 0.01 | 0 | 0.03 |
| 563 | 0.04 | 0.04 | 0.03 | 0.02 | 0.13 |
| 564 | 0 | 0 | 0 | 0.01 | 0.01 |

| | | | | | |
|---|---|---|---|---|---|
| 565 | 0 | 0 | 0 | 0 | 0 |
| 566 | 0 | 0 | 0 | 0 | 0 |
| TOTAL | 0.24 | 0.12 | 0.22 | 0.18 | 0.76 |

STATE: MISSISSIPPI

| ZIP CODE | QUARTER 1 | QUARTER 2 | QUARTER 3 | QUARTER 4 | TOTAL GRAMS |
|---|---|---|---|---|---|
| 386 | 0.05 | 0.02 | 0.02 | 0.02 | 0.11 |
| 391 | 7.99 | 11.36 | 10.36 | 11.71 | 41.42 |
| 392 | 0.02 | 0.01 | 0.01 | 0.01 | 0.05 |
| 393 | 0.05 | 0.05 | 0.06 | 0.02 | 0.18 |
| 394 | 0.01 | 0.01 | 0.01 | 0 | 0.03 |
| 395 | 0.01 | 0.01 | 0.01 | 0.01 | 0.04 |
| 396 | 0 | 0 | 0 | 0 | 0 |
| 397 | 0 | 0 | 0 | 0 | 0 |
| TOTAL | 8.13 | 11.46 | 10.47 | 11.77 | 41.83 |

STATE: MISSOURI

| ZIP CODE | QUARTER 1 | QUARTER 2 | QUARTER 3 | QUARTER 4 | TOTAL GRAMS |
|---|---|---|---|---|---|
| 630 | 0.02 | 0.01 | 0.01 | 0.01 | 0.05 |
| 631 | 0.13 | 0.12 | 0.13 | 0.14 | 0.52 |
| 633 | 0 | 0 | 0 | 0 | 0 |
| 634 | 0.01 | 0 | 0 | 0 | 0.01 |
| 636 | 0.02 | 0.01 | 0.01 | 0.01 | 0.05 |
| 637 | 0.1 | 0.05 | 0.03 | 0.06 | 0.24 |
| 638 | 0 | 0 | 0 | 0 | 0 |
| 639 | 0.02 | 0.03 | 0.04 | 0.03 | 0.12 |
| 640 | 0.01 | 0 | 0.01 | 0 | 0.02 |
| 641 | 0.08 | 0.05 | 0.03 | 0.03 | 0.19 |
| 645 | 0.01 | 0.04 | 0 | 0 | 0.05 |
| 648 | 0.01 | 0.02 | 0.02 | 0.01 | 0.06 |
| 652 | 0.06 | 0.05 | 0.03 | 0.02 | 0.16 |
| 653 | 0.01 | 0 | 0 | 0.01 | 0.02 |
| 654 | 0 | 0 | 0 | 0.01 | 0.01 |
| 657 | 0 | 0 | 0 | 0 | 0 |
| 658 | 0 | 0 | 0 | 0 | 0 |
| TOTAL | 0.48 | 0.38 | 0.31 | 0.33 | 1.5 |

STATE: MONTANA

DRUG CODE:9740DRUG NAME:SUFENTANIL BASE
STATE:MONTANA

| ZIP CODE | QUARTER 1 | QUARTER 2 | QUARTER 3 | QUARTER 4 | TOTAL GRAMS |
|---|---|---|---|---|---|
| 590 | 0 | 0 | 0 | 0 | 0 |
| 591 | 0.02 | 0.01 | 0.01 | 0.01 | 0.05 |
| 594 | 0 | 0 | 0 | 0 | 0 |
| 596 | 0 | 0 | 0 | 0 | 0 |
| 597 | 0.01 | 0.01 | 0 | 0.01 | 0.03 |
| 598 | 0.01 | 0.02 | 0.02 | 0.01 | 0.06 |
| 599 | 0.01 | 0.01 | 0 | 0.01 | 0.03 |
| TOTAL | 0.05 | 0.05 | 0.03 | 0.04 | 0.17 |

STATE: NEBRASKA

| ZIP CODE | QUARTER 1 | QUARTER 2 | QUARTER 3 | QUARTER 4 | TOTAL GRAMS |
|---|---|---|---|---|---|
| 680 | 0 | 0 | 0 | 0 | 0 |
| 681 | 0.08 | 0.06 | 0.05 | 0.07 | 0.26 |
| 683 | 0 | 0 | 0 | 0 | 0 |
| 684 | 0 | 0 | 0 | 0.01 | 0.01 |
| 685 | 0.01 | 0.01 | 0.01 | 0.02 | 0.05 |
| 686 | 0 | 0 | 0 | 0 | 0 |
| 687 | 0.01 | 0 | 0.01 | 0.01 | 0.03 |

| | | | | | |
|---|---|---|---|---|---|
| 688 | 0.01 | 0 | 0.01 | 0 | 0.02 |
| 689 | 0 | 0 | 0 | 0 | 0 |
| 690 | 0 | 0 | 0 | 0 | 0 |
| 691 | 0 | 0 | 0 | 0 | 0 |
| 692 | 0 | 0 | 0 | 0 | 0 |
| TOTAL | 0.11 | 0.07 | 0.08 | 0.11 | 0.37 |

STATE: NEVADA

| ZIP CODE | QUARTER 1 | QUARTER 2 | QUARTER 3 | QUARTER 4 | TOTAL GRAMS |
|---|---|---|---|---|---|
| 890 | 0.01 | 0.01 | 0.01 | 0.02 | 0.05 |
| 891 | 0.1 | 0.02 | 0.07 | 0.06 | 0.25 |
| 894 | 0 | 0 | 0 | 0 | 0 |
| 895 | 0.01 | 0.01 | 0.01 | 0.01 | 0.04 |
| 897 | 0 | 0 | 0 | 0 | 0 |
| 898 | 0.01 | 0.01 | 0 | 0 | 0.02 |
| TOTAL | 0.13 | 0.05 | 0.09 | 0.09 | 0.36 |

STATE: NEW HAMPSHIRE

| ZIP CODE | QUARTER 1 | QUARTER 2 | QUARTER 3 | QUARTER 4 | TOTAL GRAMS |
|---|---|---|---|---|---|
| 32 | 0 | 0 | 0 | 0 | 0 |
| 34 | 0 | 0 | 0 | 0 | 0 |
| 37 | 0.01 | 0 | 0 | 0 | 0.01 |
| 38 | 0.02 | 0.01 | 0.02 | 0.01 | 0.06 |
| TOTAL | 0.03 | 0.01 | 0.02 | 0.01 | 0.07 |

STATE: NEW JERSEY

| ZIP CODE | QUARTER 1 | QUARTER 2 | QUARTER 3 | QUARTER 4 | TOTAL GRAMS |
|---|---|---|---|---|---|
| 70 | 0.01 | 0 | 0.01 | 0 | 0.02 |
| 71 | 0.01 | 0.01 | 0.01 | 0.01 | 0.04 |
| 73 | 0 | 0 | 0 | 0 | 0 |
| 74 | 0.68 | 0.71 | 0.7 | 1.4 | 3.49 |
| 75 | 0.07 | 0.02 | 0.03 | 0.02 | 0.14 |
| 76 | 0.02 | 0.03 | 0.03 | 0.01 | 0.09 |
| 77 | 0.03 | 0.03 | 0.04 | 0.02 | 0.12 |
| 78 | 0 | 0 | 0 | 0 | 0 |
| 79 | 0 | 0 | 0 | 0 | 0 |
| 80 | 0 | 0 | 0 | 0 | 0 |
| 81 | 0 | 0.01 | 0 | 0 | 0.01 |
| 82 | 0.03 | 0.01 | 0.01 | 0.01 | 0.06 |
| 84 | 0 | 0 | 0 | 0 | 0 |
| 85 | 0.03 | 0 | 0 | 0 | 0.03 |
| 86 | 0 | 0 | 0 | 0 | 0 |
| 87 | 0 | 0 | 0 | 0 | 0 |
| 88 | 0.01 | 0 | 0 | 0 | 0.01 |
| 89 | 0.1 | 0.12 | 0.11 | 0.09 | 0.42 |
| TOTAL | 0.99 | 0.94 | 0.94 | 1.56 | 4.43 |

STATE: NEW MEXICO

| ZIP CODE | QUARTER 1 | QUARTER 2 | QUARTER 3 | QUARTER 4 | TOTAL GRAMS |
|---|---|---|---|---|---|
| 871 | 0.04 | 0.04 | 0.03 | 0.03 | 0.14 |
| 873 | 0 | 0 | 0 | 0 | 0 |
| 874 | 0.01 | 0 | 0 | 0 | 0.01 |
| 875 | 0 | 0 | 0 | 0 | 0 |

DRUG CODE:9740DRUG NAME:SUFENTANIL BASE
STATE:NEW MEXICO

| ZIP CODE | QUARTER 1 | QUARTER 2 | QUARTER 3 | QUARTER 4 | TOTAL GRAMS |
|---|---|---|---|---|---|
| 879 | 0 | 0 | 0 | 0 | 0 |
| 880 | 0 | 0 | 0 | 0 | 0 |

| | | | | | |
|---|---|---|---|---|---|
| 882 | 0 | 0 | 0 | 0 | 0 |
| 883 | 0 | 0 | 0 | 0 | 0 |
| TOTAL | 0.05 | 0.04 | 0.03 | 0.03 | 0.15 |

STATE: NEW YORK

| ZIP CODE | QUARTER 1 | QUARTER 2 | QUARTER 3 | QUARTER 4 | TOTAL GRAMS |
|---|---|---|---|---|---|
| 100 | 0.99 | 1.35 | 1.12 | 1.25 | 4.71 |
| 103 | 0 | 0 | 0 | 0 | 0 |
| 104 | 0.05 | 0.03 | 0.01 | 0.01 | 0.1 |
| 105 | 0 | 0 | 0 | 0 | 0 |
| 110 | 0.03 | 0.08 | 0 | 0.03 | 0.14 |
| 112 | 0.01 | 0.01 | 0.02 | 0.02 | 0.06 |
| 113 | 0.06 | 0.07 | 0.08 | 0.08 | 0.29 |
| 114 | 0.27 | 0 | 0.13 | 0 | 0.4 |
| 115 | 0 | 0 | 0.01 | 0 | 0.01 |
| 117 | 0.03 | 0.03 | 0.03 | 0.02 | 0.11 |
| 119 | 0 | 0 | 0 | 0 | 0 |
| 120 | 0 | 0 | 0 | 0 | 0 |
| 122 | 0.09 | 0.07 | 0.1 | 0.08 | 0.34 |
| 123 | 0 | 0.01 | 0 | 0 | 0.01 |
| 124 | 0 | 0 | 0 | 0 | 0 |
| 128 | 0.02 | 0.02 | 0.02 | 0.02 | 0.08 |
| 133 | 0 | 0 | 0.01 | 0.01 | 0.02 |
| 135 | 0.01 | 0 | 0 | 0 | 0.01 |
| 137 | 0.03 | 0.01 | 0.02 | 0.02 | 0.08 |
| 140 | 0 | 0 | 0 | 0 | 0 |
| 142 | 0.21 | 0.06 | 0.1 | 0.09 | 0.46 |
| 146 | 0.09 | 0.09 | 0.11 | 0.1 | 0.39 |
| TOTAL | 1.89 | 1.83 | 1.76 | 1.73 | 7.21 |

STATE: NORTH CAROLINA

| ZIP CODE | QUARTER 1 | QUARTER 2 | QUARTER 3 | QUARTER 4 | TOTAL GRAMS |
|---|---|---|---|---|---|
| 271 | 0.14 | 0.09 | 0.08 | 0.09 | 0.4 |
| 272 | 0.02 | 0.03 | 0.02 | 0.02 | 0.09 |
| 273 | 0.01 | 0 | 0 | 0 | 0.01 |
| 274 | 0.02 | 0.01 | 0.01 | 0.02 | 0.06 |
| 275 | 0.07 | 0 | 0 | 0.74 | 0.81 |
| 276 | 0.01 | 0.02 | 0.01 | 0.01 | 0.05 |
| 277 | 0.03 | 0 | 0 | 0.04 | 0.07 |
| 278 | 0.03 | 0.01 | 0.02 | 0.03 | 0.09 |
| 279 | 0.01 | 0 | 0 | 0 | 0.01 |
| 280 | 0 | 0 | 0 | 0.01 | 0.01 |
| 282 | 0.04 | 0.04 | 0.03 | 0.01 | 0.12 |
| 283 | 0.01 | 0.04 | 0.03 | 0.03 | 0.11 |
| 284 | 0.01 | 0.01 | 0.01 | 0.01 | 0.04 |
| 285 | 0 | 0 | 0.01 | 0 | 0.01 |
| 286 | 0.01 | 0.01 | 0.01 | 0 | 0.03 |
| 287 | 0.02 | 0.01 | 0.01 | 0.01 | 0.05 |
| 288 | 0.06 | 0.08 | 0.09 | 0.07 | 0.3 |
| 289 | 0 | 0 | 0 | 0 | 0 |
| TOTAL | 0.49 | 0.35 | 0.33 | 1.09 | 2.26 |

STATE: NORTH DAKOTA

| ZIP CODE | QUARTER 1 | QUARTER 2 | QUARTER 3 | QUARTER 4 | TOTAL GRAMS |
|---|---|---|---|---|---|
| 582 | 0.04 | 0.02 | 0.02 | 0.01 | 0.09 |
| 583 | 0 | 0 | 0 | 0 | 0 |
| 588 | 0 | 0 | 0 | 0 | 0 |
| TOTAL | 0.04 | 0.02 | 0.02 | 0.01 | 0.09 |

STATE: OHIO

| ZIP CODE | QUARTER 1 | QUARTER 2 | QUARTER 3 | QUARTER 4 | TOTAL GRAMS |
|---|---|---|---|---|---|

| | | | | | |
|---|---|---|---|---|---|
| 430 | 0.01 | 0 | 0.67 | 0 | 0.68 |
| 431 | 0.02 | 0.01 | 0 | 0.01 | 0.04 |
| 432 | 0.26 | 0.3 | 0.05 | 0.03 | 0.64 |
| 433 | 0 | 0 | 0 | 0 | 0 |
| 435 | 0.07 | 0.14 | 0.01 | 0.07 | 0.29 |
| 436 | 0.11 | 0.06 | 0.07 | 0.11 | 0.35 |
| 437 | 0.04 | 0.02 | 0.03 | 0.02 | 0.11 |

DRUG CODE:9740DRUG NAME:SUFENTANIL BASE
STATE:OHIO

| ZIP CODE | QUARTER 1 | QUARTER 2 | QUARTER 3 | QUARTER 4 | TOTAL GRAMS |
|---|---|---|---|---|---|
| 438 | 0 | 0 | 0 | 0 | 0 |
| 439 | 0 | 0 | 0 | 0 | 0 |
| 440 | 0.01 | 0.01 | 0.02 | 0.02 | 0.06 |
| 441 | 0.04 | 0.02 | 0.04 | 0.03 | 0.13 |
| 442 | 0 | 0 | 0 | 0 | 0 |
| 443 | 0 | 0 | 0.01 | 0 | 0.01 |
| 445 | 0.01 | 0 | 0 | 0 | 0.01 |
| 446 | 0.01 | 0.01 | 0 | 0 | 0.02 |
| 447 | 0 | 0.02 | 0 | 0 | 0.02 |
| 448 | 0.02 | 0 | 0 | 0.01 | 0.03 |
| 449 | 0.02 | 0.01 | 0.01 | 0.01 | 0.05 |
| 450 | 0.03 | 0 | 0 | 0 | 0.03 |
| 451 | 0 | 0 | 0 | 0 | 0 |
| 452 | 0.05 | 0.03 | 0.03 | 0.02 | 0.13 |
| 453 | 0 | 0 | 0.01 | 0 | 0.01 |
| 454 | 0.05 | 0.06 | 0.03 | 0.05 | 0.19 |
| 455 | 0 | 0 | 0 | 0 | 0 |
| 457 | 0.01 | 0.01 | 0.01 | 0.01 | 0.04 |
| 458 | 0.01 | 0.01 | 0.01 | 0.01 | 0.04 |
| TOTAL | 0.77 | 0.71 | 1 | 0.4 | 2.88 |

STATE: OKLAHOMA

| ZIP CODE | QUARTER 1 | QUARTER 2 | QUARTER 3 | QUARTER 4 | TOTAL GRAMS |
|---|---|---|---|---|---|
| 730 | 0.04 | 0.02 | 0.03 | 0.03 | 0.12 |
| 731 | 0.13 | 0.09 | 0.11 | 0.07 | 0.4 |
| 735 | 0.01 | 0.01 | 0.01 | 0.01 | 0.04 |
| 737 | 0 | 0 | 0 | 0 | 0 |
| 740 | 0 | 0 | 0 | 0 | 0 |
| 741 | 0.05 | 0.04 | 0.04 | 0.05 | 0.18 |
| 744 | 0.01 | 0 | 0 | 0 | 0.01 |
| TOTAL | 0.24 | 0.16 | 0.19 | 0.16 | 0.75 |

STATE: OREGON

| ZIP CODE | QUARTER 1 | QUARTER 2 | QUARTER 3 | QUARTER 4 | TOTAL GRAMS |
|---|---|---|---|---|---|
| 970 | 0.04 | 0.03 | 0.03 | 0.03 | 0.13 |
| 971 | 0.02 | 0.02 | 0.02 | 0.01 | 0.07 |
| 972 | 0.41 | 0.16 | 0.26 | 0.18 | 1.01 |
| 973 | 0.01 | 0.01 | 0.01 | 0.01 | 0.04 |
| 974 | 0.01 | 0.02 | 0 | 0 | 0.03 |
| 975 | 0 | 0 | 0 | 0 | 0 |
| 977 | 0.01 | 0 | 0 | 0 | 0.01 |
| 978 | 0 | 0 | 0 | 0 | 0 |
| TOTAL | 0.5 | 0.24 | 0.32 | 0.23 | 1.29 |

STATE: PENNSYLVANIA

| ZIP CODE | QUARTER 1 | QUARTER 2 | QUARTER 3 | QUARTER 4 | TOTAL GRAMS |
|---|---|---|---|---|---|
| 150 | 0.01 | 0.01 | 0.01 | 0.01 | 0.04 |

| | | | | | |
|---|---|---|---|---|---|
| 151 | 0 | 0 | 0 | 0 | 0 |
| 152 | 0.01 | 0.01 | 0.02 | 0.01 | 0.05 |
| 154 | 0 | 0 | 0 | 0 | 0 |
| 158 | 0 | 0 | 0 | 0 | 0 |
| 163 | 0 | 0 | 0 | 0 | 0 |
| 165 | 0.02 | 0.01 | 0.02 | 0.01 | 0.06 |
| 166 | 0.02 | 0.01 | 0.01 | 0.01 | 0.05 |
| 167 | 0 | 0 | 0 | 0 | 0 |
| 169 | 0 | 0 | 0.01 | 0 | 0.01 |
| 170 | 0.01 | 0 | 0 | 0.01 | 0.02 |
| 171 | 0 | 0 | 0 | 0 | 0 |
| 174 | 0.01 | 0.01 | 0.01 | 0 | 0.03 |
| 175 | 0.03 | 0.08 | 0.03 | 0 | 0.14 |
| 176 | 0 | 0 | 0 | 0 | 0 |
| 177 | 0 | 0 | 0 | 0 | 0 |
| 178 | 0.01 | 0.02 | 0.01 | 0.01 | 0.05 |
| 180 | 0 | 0.01 | 0 | 0 | 0.01 |
| 181 | 0.01 | 0.04 | 0.03 | 0.01 | 0.09 |
| 182 | 0.01 | 0 | 0.01 | 0 | 0.02 |
| 185 | 0.01 | 0.02 | 0.01 | 0.01 | 0.05 |
| 187 | 0.02 | 0.01 | 0.02 | 0.04 | 0.09 |
| 188 | 0.01 | 0 | 0 | 0 | 0.01 |

DRUG CODE:9740DRUG NAME:SUFENTANIL BASE
STATE:PENNSYLVANIA

| ZIP CODE | QUARTER 1 | QUARTER 2 | QUARTER 3 | QUARTER 4 | TOTAL GRAMS |
|---|---|---|---|---|---|
| 190 | 0.56 | 0.61 | 1.49 | 1.35 | 4.01 |
| 191 | 0.14 | 0.07 | 0.16 | 0.11 | 0.48 |
| 193 | 0.01 | 0 | 0 | 0 | 0.01 |
| 196 | 0.02 | 0 | 0 | 0 | 0.02 |
| TOTAL | 0.91 | 0.91 | 1.84 | 1.58 | 5.24 |

STATE: PUERTO RICO

| ZIP CODE | QUARTER 1 | QUARTER 2 | QUARTER 3 | QUARTER 4 | TOTAL GRAMS |
|---|---|---|---|---|---|
| 6 | 0 | 0.03 | 0 | 0 | 0.03 |
| 7 | 0.02 | 0.03 | 0.01 | 0.01 | 0.07 |
| 9 | 0.12 | 0.21 | 0.21 | 0.12 | 0.66 |
| TOTAL | 0.14 | 0.27 | 0.22 | 0.13 | 0.76 |

STATE: RHODE ISLAND

| ZIP CODE | QUARTER 1 | QUARTER 2 | QUARTER 3 | QUARTER 4 | TOTAL GRAMS |
|---|---|---|---|---|---|
| 28 | 0.01 | 0.01 | 0.01 | 0 | 0.03 |
| 29 | 0 | 0 | 0 | 0 | 0 |
| TOTAL | 0.01 | 0.01 | 0.01 | 0 | 0.03 |

STATE: SOUTH CAROLINA

| ZIP CODE | QUARTER 1 | QUARTER 2 | QUARTER 3 | QUARTER 4 | TOTAL GRAMS |
|---|---|---|---|---|---|
| 290 | 0 | 0 | 0 | 0 | 0 |
| 291 | 0 | 0 | 0 | 0 | 0 |
| 292 | 0.02 | 0 | 0.01 | 0 | 0.03 |
| 293 | 0.01 | 0.01 | 0.02 | 0.01 | 0.05 |
| 294 | 0.05 | 0.07 | 0.04 | 0.06 | 0.22 |
| 295 | 0.01 | 0.01 | 0.01 | 0 | 0.03 |
| 296 | 0.05 | 0.04 | 0.02 | 0.04 | 0.15 |
| 299 | 0.67 | 0.67 | 0 | 0.67 | 2.01 |
| TOTAL | 0.81 | 0.8 | 0.1 | 0.78 | 2.49 |

STATE: SOUTH DAKOTA

| ZIP CODE | QUARTER 1 | QUARTER 2 | QUARTER 3 | QUARTER 4 | TOTAL GRAMS |
|---|---|---|---|---|---|

| | | | | | |
|---|---|---|---|---|---|
| 570 | 0.02 | 0.01 | 0.01 | 0.02 | 0.06 |
| 571 | 0 | 0 | 0 | 0 | 0 |
| 574 | 0 | 0 | 0 | 0 | 0 |
| 575 | 0 | 0 | 0 | 0 | 0 |
| 577 | 0.02 | 0.01 | 0.01 | 0.01 | 0.05 |
| TOTAL | 0.04 | 0.02 | 0.02 | 0.03 | 0.11 |

STATE: TENNESSEE

| ZIP CODE | QUARTER 1 | QUARTER 2 | QUARTER 3 | QUARTER 4 | TOTAL GRAMS |
|---|---|---|---|---|---|
| 370 | 0.1 | 0.02 | 0.02 | 0.02 | 0.16 |
| 371 | 0.06 | 0.04 | 0.05 | 0.06 | 0.21 |
| 372 | 0.03 | 0.12 | 0.02 | 0.04 | 0.21 |
| 373 | 0.05 | 0.02 | 0.01 | 0 | 0.08 |
| 376 | 0.05 | 0.01 | 0.03 | 0.02 | 0.11 |
| 377 | 0 | 0 | 0 | 0 | 0 |
| 378 | 0 | 0.01 | 0.02 | 0.01 | 0.04 |
| 379 | 0.02 | 0.02 | 0.01 | 0.02 | 0.07 |
| 380 | 0.01 | 0 | 0.01 | 0.01 | 0.03 |
| 381 | 0.12 | 0.09 | 0.18 | 0.06 | 0.45 |
| 382 | 0 | 0 | 0 | 0 | 0 |
| 383 | 0.03 | 0.01 | 0.02 | 0.02 | 0.08 |
| 384 | 0.03 | 0.05 | 0 | 0.01 | 0.09 |
| 385 | 0.02 | 0.01 | 0.01 | 0.02 | 0.06 |
| TOTAL | 0.52 | 0.4 | 0.38 | 0.29 | 1.59 |

STATE: TEXAS

| ZIP CODE | QUARTER 1 | QUARTER 2 | QUARTER 3 | QUARTER 4 | TOTAL GRAMS |
|---|---|---|---|---|---|
| 750 | 0.11 | 0.06 | 0.05 | 0.07 | 0.29 |
| 751 | 0.01 | 0 | 0 | 0 | 0.01 |
| 752 | 0.1 | 0.06 | 0.06 | 0.04 | 0.26 |
| 753 | 0.04 | 0.02 | 0.02 | 0.01 | 0.09 |
| 754 | 0.01 | 0.01 | 0.01 | 0.01 | 0.04 |
| 755 | 0 | 0 | 0 | 0 | 0 |
| 756 | 0.17 | 0.1 | 0.06 | 0.18 | 0.51 |
| 757 | 0.04 | 0.02 | 0.01 | 0.02 | 0.09 |
| 758 | 0 | 0 | 0 | 0 | 0 |
| 759 | 0.01 | 0.01 | 0 | 0 | 0.02 |
| 760 | 0.02 | 0 | 0.02 | 0.01 | 0.05 |
| 761 | 0.18 | 0.11 | 0.14 | 0.29 | 0.72 |

DRUG CODE:9740DRUG NAME:SUFENTANIL BASE
STATE:TEXAS

| ZIP CODE | QUARTER 1 | QUARTER 2 | QUARTER 3 | QUARTER 4 | TOTAL GRAMS |
|---|---|---|---|---|---|
| 762 | 0.02 | 0.02 | 0.02 | 0.02 | 0.08 |
| 763 | 0.01 | 0.03 | 0.02 | 0 | 0.06 |
| 764 | 0 | 0 | 0 | 0 | 0 |
| 765 | 0.04 | 0.05 | 0.04 | 0.03 | 0.16 |
| 767 | 0 | 0 | 0 | 0 | 0 |
| 769 | 0 | 0 | 0 | 0 | 0 |
| 770 | 1.04 | 0.7 | 0.77 | 0.59 | 3.1 |
| 773 | 0.01 | 0.01 | 0 | 0.01 | 0.03 |
| 774 | 0.01 | 0.01 | 0.01 | 0.02 | 0.05 |
| 775 | 0.03 | 0.01 | 0.01 | 0.01 | 0.06 |
| 776 | 0.01 | 0 | 0 | 0 | 0.01 |
| 777 | 0 | 0 | 0 | 0 | 0 |
| 778 | 0.01 | 0.01 | 0.01 | 0.01 | 0.04 |
| 779 | 0 | 0 | 0 | 0 | 0 |
| 780 | 0.01 | 0 | 0 | 0 | 0.01 |
| 781 | 0 | 0 | 0 | 0 | 0 |
| 782 | 0.05 | 0.03 | 0.15 | 0.03 | 0.26 |

| | | | | | |
|---|---|---|---|---|---|
| 784 | 0.03 | 0.02 | 0.04 | 0.03 | 0.12 |
| 785 | 0.1 | 0.05 | 0.08 | 0.06 | 0.29 |
| 786 | 0 | 0 | 0 | 0 | 0 |
| 787 | 0.01 | 0.01 | 0.01 | 0.02 | 0.05 |
| 788 | 0 | 0 | 0 | 0 | 0 |
| 789 | 0 | 0 | 0 | 0 | 0 |
| 791 | 0 | 0 | 0.01 | 0 | 0.01 |
| 794 | 0.08 | 0.06 | 0.05 | 0.07 | 0.26 |
| 796 | 0.03 | 0.01 | 0.02 | 0.02 | 0.08 |
| 797 | 0 | 0.01 | 0 | 0 | 0.01 |
| 799 | 0.05 | 0.03 | 0.03 | 0.03 | 0.14 |
| TOTAL | 2.23 | 1.45 | 1.64 | 1.58 | 6.9 |

STATE: UTAH

| ZIP CODE | QUARTER 1 | QUARTER 2 | QUARTER 3 | QUARTER 4 | TOTAL GRAMS |
|---|---|---|---|---|---|
| 840 | 0.02 | 0.01 | 0.01 | 0.01 | 0.05 |
| 841 | 0.15 | 0.17 | 0.18 | 0.16 | 0.66 |
| 843 | 0 | 0 | 0 | 0 | 0 |
| 844 | 0.02 | 0.01 | 0.02 | 0.02 | 0.07 |
| 846 | 0 | 0.01 | 0.02 | 0 | 0.03 |
| 847 | 0.02 | 0.02 | 0.01 | 0.02 | 0.07 |
| TOTAL | 0.21 | 0.22 | 0.24 | 0.21 | 0.88 |

STATE: VERMONT

| ZIP CODE | QUARTER 1 | QUARTER 2 | QUARTER 3 | QUARTER 4 | TOTAL GRAMS |
|---|---|---|---|---|---|
| 54 | 0.02 | 0.02 | 0 | 0 | 0.04 |
| TOTAL | 0.02 | 0.02 | 0 | 0 | 0.04 |

STATE: VIRGINIA

| ZIP CODE | QUARTER 1 | QUARTER 2 | QUARTER 3 | QUARTER 4 | TOTAL GRAMS |
|---|---|---|---|---|---|
| 201 | 0 | 0 | 0 | 0 | 0 |
| 220 | 0.06 | 0.07 | 0.03 | 0.09 | 0.25 |
| 222 | 0.03 | 0.03 | 0.01 | 0.03 | 0.1 |
| 223 | 0.02 | 0.01 | 0.01 | 0.01 | 0.05 |
| 224 | 0.03 | 0.03 | 0.03 | 0.04 | 0.13 |
| 225 | 0.01 | 0.01 | 0.01 | 0.02 | 0.05 |
| 226 | 0.01 | 0 | 0 | 0 | 0.01 |
| 228 | 0 | 0.03 | 0 | 0.03 | 0.06 |
| 229 | 0.07 | 0.06 | 0.07 | 0.05 | 0.25 |
| 231 | 0.02 | 0.01 | 0.01 | 0.01 | 0.05 |
| 232 | 0.07 | 0.07 | 0.05 | 0.06 | 0.25 |
| 235 | 0 | 0 | 0 | 0 | 0 |
| 236 | 0 | 0 | 0 | 0 | 0 |
| 237 | 0.01 | 0 | 0 | 0.01 | 0.02 |
| 238 | 0.01 | 0.01 | 0 | 0 | 0.02 |
| 239 | 0 | 0 | 0 | 0 | 0 |
| 240 | 0.03 | 0.03 | 0.02 | 0.02 | 0.1 |
| 241 | 0.01 | 0 | 0 | 0.01 | 0.02 |
| 242 | 0 | 0 | 0 | 0 | 0 |
| 243 | 0 | 0 | 0 | 0 | 0 |
| 245 | 0.07 | 0.06 | 0.06 | 0.04 | 0.23 |
| TOTAL | 0.45 | 0.42 | 0.3 | 0.42 | 1.59 |

DRUG CODE:9740DRUG NAME:SUFENTANIL BASE
STATE: WASHINGTON

| ZIP CODE | QUARTER 1 | QUARTER 2 | QUARTER 3 | QUARTER 4 | TOTAL GRAMS |
|---|---|---|---|---|---|
| 980 | 0.04 | 0.02 | 0.02 | 0.02 | 0.1 |
| 981 | 0.17 | 0.1 | 0.12 | 0.18 | 0.57 |
| 982 | 0.01 | 0.02 | 0.01 | 0.01 | 0.05 |

| | | | | | |
|---|---|---|---|---|---|
| 983 | 0 | 0 | 0 | 0 | 0 |
| 984 | 0.03 | 0.01 | 0.03 | 0 | 0.07 |
| 985 | 0 | 0 | 0 | 0 | 0 |
| 986 | 0 | 0 | 0 | 0 | 0 |
| 988 | 0 | 0 | 0 | 0 | 0 |
| 989 | 0.01 | 0 | 0.01 | 0 | 0.02 |
| 991 | 0 | 0 | 0 | 0 | 0 |
| 992 | 0.05 | 0.04 | 0.04 | 0.04 | 0.17 |
| 993 | 0 | 0 | 0 | 0 | 0 |
| TOTAL | 0.31 | 0.19 | 0.23 | 0.25 | 0.98 |

STATE: WEST VIRGINIA

| ZIP CODE | QUARTER 1 | QUARTER 2 | QUARTER 3 | QUARTER 4 | TOTAL GRAMS |
|---|---|---|---|---|---|
| 253 | 0.03 | 0.03 | 0.04 | 0.02 | 0.12 |
| 254 | 0 | 0 | 0 | 0 | 0 |
| 257 | 0.01 | 0 | 0 | 0 | 0.01 |
| 261 | 0.01 | 0 | 0.01 | 0.01 | 0.03 |
| 265 | 0.02 | 0.02 | 0.02 | 0.01 | 0.07 |
| 267 | 0 | 0 | 0 | 0 | 0 |
| TOTAL | 0.07 | 0.05 | 0.07 | 0.04 | 0.23 |

STATE: WISCONSIN

| ZIP CODE | QUARTER 1 | QUARTER 2 | QUARTER 3 | QUARTER 4 | TOTAL GRAMS |
|---|---|---|---|---|---|
| 530 | 0.01 | 0 | 0.01 | 0 | 0.02 |
| 531 | 0.14 | 1.34 | 0.67 | 0 | 2.15 |
| 532 | 0.15 | 0.7 | 0.04 | 0.05 | 0.94 |
| 535 | 0 | 0.01 | 0 | 0 | 0.01 |
| 537 | 0.04 | 0.03 | 0.02 | 0.03 | 0.12 |
| 538 | 0 | 0 | 0 | 0 | 0 |
| 539 | 0 | 0 | 0 | 0 | 0 |
| 540 | 0 | 0 | 0 | 0 | 0 |
| 541 | 0 | 0 | 0 | 0 | 0 |
| 543 | 0.02 | 0.01 | 0.01 | 0.01 | 0.05 |
| 544 | 0 | 0 | 0 | 0 | 0 |
| 546 | 0.01 | 0 | 0 | 0.01 | 0.02 |
| 547 | 0 | 0 | 0 | 0 | 0 |
| 548 | 0 | 0 | 0 | 0 | 0 |
| 549 | 0.02 | 0.01 | 0.02 | 0.01 | 0.06 |
| TOTAL | 0.39 | 2.1 | 0.77 | 0.11 | 3.37 |

STATE: WYOMING

| ZIP CODE | QUARTER 1 | QUARTER 2 | QUARTER 3 | QUARTER 4 | TOTAL GRAMS |
|---|---|---|---|---|---|
| 820 | 0.01 | 0.01 | 0 | 0.01 | 0.03 |
| 826 | 0.01 | 0 | 0 | 0 | 0.01 |
| 827 | 0 | 0 | 0 | 0 | 0 |
| 828 | 0 | 0 | 0 | 0 | 0 |
| 829 | 0 | 0 | 0 | 0 | 0 |
| 831 | 0 | 0 | 0 | 0 | 0 |
| TOTAL | 0.02 | 0.01 | 0 | 0.01 | 0.04 |

DRUG CODE:9780DRUG NAME:TAPENTADOL
STATE: ALABAMA

| ZIP CODE | QUARTER 1 | QUARTER 2 | QUARTER 3 | QUARTER 4 | TOTAL GRAMS |
|---|---|---|---|---|---|
| 350 | 8,964.50 | 8,762 | 9,037.50 | 8,752.50 | 35,516.50 |
| 351 | 4,928.50 | 4,949 | 4,861 | 4,636.50 | 19,375 |
| 352 | 5,211 | 4,915 | 4,726 | 4,804.50 | 19,656.50 |
| 354 | 5,214 | 5,218.50 | 5,095.50 | 5,057 | 20,585 |
| 355 | 3,174.50 | 3,349 | 3,166 | 3,329.50 | 13,019 |
| 356 | 5,202.50 | 5,049.50 | 5,222.50 | 5,128 | 20,602.50 |
| 357 | 2,196.50 | 2,434 | 2,416 | 2,309 | 9,355.50 |

| | | | | | |
|---|---|---|---|---|---|
| 358 | 3,189 | 3,071 | 3,433.50 | 3,254 | 12,947.50 |
| 359 | 2,881.50 | 2,765 | 2,566.50 | 2,506 | 10,719 |
| 360 | 5,806 | 5,889.50 | 5,279 | 5,915 | 22,889.50 |
| 361 | 2,909 | 3,091.50 | 2,828 | 3,077.50 | 11,906 |
| 362 | 1,700.50 | 1,679 | 1,796 | 1,882 | 7,057.50 |
| 363 | 2,147 | 1,951 | 1,876 | 1,750 | 7,724 |

DRUG CODE:9780DRUG NAME:TAPENTADOL
STATE:ALABAMA

| ZIP CODE | QUARTER 1 | QUARTER 2 | QUARTER 3 | QUARTER 4 | TOTAL GRAMS |
|---|---|---|---|---|---|
| 364 | 600 | 545.5 | 564.5 | 666 | 2,376 |
| 365 | 5,177.50 | 5,465.50 | 5,928 | 5,721 | 22,292 |
| 366 | 3,375.50 | 3,488 | 3,070.50 | 3,550 | 13,484 |
| 367 | 1,014 | 843 | 697 | 868.5 | 3,422.50 |
| 368 | 2,333.50 | 2,265 | 2,173 | 2,424.50 | 9,196 |
| 369 | 135.5 | 183 | 198 | 219.5 | 736 |
| TOTAL | 66,160.50 | 65,914 | 64,934.50 | 65,851 | 262,860 |

STATE: ALASKA

| ZIP CODE | QUARTER 1 | QUARTER 2 | QUARTER 3 | QUARTER 4 | TOTAL GRAMS |
|---|---|---|---|---|---|
| 995 | 904.5 | 722 | 827 | 924 | 3,377.50 |
| 996 | 638.5 | 797 | 1,090.50 | 879 | 3,405 |
| 997 | 213 | 159 | 160.5 | 160 | 692.5 |
| 998 | 36 | 82.5 | 63 | 63 | 244.5 |
| 999 | 30 | 0 | 0 | 0 | 30 |
| TOTAL | 1,822 | 1,760.50 | 2,141 | 2,026 | 7,749.50 |

STATE: ARIZONA

| ZIP CODE | QUARTER 1 | QUARTER 2 | QUARTER 3 | QUARTER 4 | TOTAL GRAMS |
|---|---|---|---|---|---|
| 850 | 7,562.35 | 5,848 | 5,861 | 6,175 | 25,446.35 |
| 851 | 1,099.50 | 961 | 940 | 1,002 | 4,002.50 |
| 852 | 16,258 | 16,062.50 | 15,985 | 17,070 | 65,375.50 |
| 853 | 5,685 | 5,776.50 | 6,672 | 7,501.50 | 25,635 |
| 855 | 268 | 190.5 | 306 | 332.5 | 1,097 |
| 856 | 939 | 698.5 | 755.5 | 714.5 | 3,107.50 |
| 857 | 4,016.50 | 3,957 | 4,077.50 | 4,451 | 16,502 |
| 859 | 38 | 102 | 105.5 | 140 | 385.5 |
| 860 | 236 | 262 | 227 | 298.5 | 1,023.50 |
| 863 | 1,033 | 970 | 1,165 | 1,341.50 | 4,509.50 |
| 864 | 741.5 | 590.5 | 708.5 | 650.5 | 2,691 |
| TOTAL | 37,876.85 | 35,418.50 | 36,803 | 39,677 | 149,775.35 |

STATE: ARKANSAS

| ZIP CODE | QUARTER 1 | QUARTER 2 | QUARTER 3 | QUARTER 4 | TOTAL GRAMS |
|---|---|---|---|---|---|
| 716 | 827.5 | 702 | 824 | 853.5 | 3,207 |
| 717 | 1,272 | 1,387.50 | 1,259.50 | 1,301.50 | 5,220.50 |
| 718 | 305.5 | 392 | 528 | 354 | 1,579.50 |
| 719 | 1,255.50 | 1,131 | 1,281 | 1,038.50 | 4,706 |
| 720 | 4,328 | 4,802 | 4,848 | 4,526.50 | 18,504.50 |
| 721 | 3,328 | 3,214.50 | 3,428.50 | 3,233 | 13,204 |
| 722 | 1,671.35 | 1,623 | 1,774 | 1,627.50 | 6,695.85 |
| 723 | 460 | 465.5 | 451 | 438 | 1,814.50 |
| 724 | 1,805.50 | 1,913 | 1,853 | 2,070 | 7,641.50 |
| 725 | 1,203.50 | 1,143.50 | 1,175.50 | 1,211.50 | 4,734 |
| 726 | 1,672.50 | 1,619.50 | 1,446.50 | 1,464.50 | 6,203 |
| 727 | 4,759 | 5,338 | 4,849 | 5,048 | 19,994 |
| 728 | 739 | 635 | 655 | 540.5 | 2,569.50 |
| 729 | 2,913 | 3,055.50 | 2,908.50 | 2,658.50 | 11,535.50 |

| | | | | | |
|---|---|---|---|---|---|
| TOTAL | 26,540.35 | 27,422 | 27,281.50 | 26,365.50 | 107,609.35 |

STATE: CALIFORNIA

| ZIP CODE | QUARTER 1 | QUARTER 2 | QUARTER 3 | QUARTER 4 | TOTAL GRAMS |
|---|---|---|---|---|---|
| 900 | 2,136 | 2,379 | 2,210.50 | 2,818.50 | 9,544 |
| 902 | 3,224.50 | 2,732 | 3,099 | 2,510.50 | 11,566 |
| 903 | 133 | 135 | 143 | 168 | 579 |
| 904 | 850.5 | 1,255 | 1,088.50 | 1,580 | 4,774 |
| 905 | 221 | 323.5 | 196 | 223.5 | 964 |
| 906 | 925.5 | 942 | 1,077 | 1,195 | 4,139.50 |
| 907 | 1,546 | 1,466 | 1,500 | 1,688 | 6,200 |
| 908 | 945 | 826.5 | 960 | 858.5 | 3,590 |
| 910 | 1,062.50 | 899.5 | 801.5 | 959 | 3,722.50 |
| 911 | 306 | 482 | 414 | 417.5 | 1,619.50 |
| 912 | 274.5 | 223 | 212 | 272 | 981.5 |
| 913 | 4,427 | 4,598.50 | 4,919.50 | 4,787.50 | 18,732.50 |
| 914 | 1,687 | 1,306 | 1,195 | 1,182.50 | 5,370.50 |
| 915 | 167.5 | 312 | 261.5 | 277 | 1,018 |
| 916 | 157.5 | 160.5 | 260 | 191 | 769 |
| 917 | 9,582.50 | 10,653.50 | 10,161 | 9,882.50 | 40,279.50 |
| 918 | 62.5 | 81.5 | 71.5 | 87 | 302.5 |
| 919 | 1,361 | 1,141.50 | 1,080 | 1,229.50 | 4,812 |

DRUG CODE:9780DRUG NAME:TAPENTADOL
STATE:CALIFORNIA

| ZIP CODE | QUARTER 1 | QUARTER 2 | QUARTER 3 | QUARTER 4 | TOTAL GRAMS |
|---|---|---|---|---|---|
| 920 | 7,610.50 | 6,897 | 7,486.50 | 7,756.50 | 29,750.50 |
| 921 | 3,569.50 | 3,034 | 3,277.50 | 3,551.50 | 13,432.50 |
| 922 | 2,915 | 2,754.50 | 2,872 | 2,891.50 | 11,433 |
| 923 | 4,858.50 | 5,507.50 | 4,824.50 | 4,908 | 20,098.50 |
| 924 | 1,714 | 1,412.50 | 1,682 | 1,620.50 | 6,429 |
| 925 | 7,303.50 | 7,328 | 7,463 | 7,696 | 29,790.50 |
| 926 | 8,584.40 | 8,027.50 | 7,941 | 7,799.40 | 32,352.30 |
| 927 | 1,201.50 | 1,089 | 1,172 | 1,204.50 | 4,667 |
| 928 | 4,405.50 | 4,689 | 4,438 | 4,479 | 18,011.50 |
| 930 | 2,728.50 | 2,874 | 2,761 | 2,852.50 | 11,216 |
| 931 | 745.5 | 719 | 653.5 | 683 | 2,801 |
| 932 | 4,359.50 | 4,152 | 4,226.50 | 3,611 | 16,349 |
| 933 | 2,002.50 | 2,547.50 | 2,782.50 | 2,535 | 9,867.50 |
| 934 | 2,450 | 2,478.50 | 2,225.50 | 2,780 | 9,934 |
| 935 | 1,927 | 2,202 | 2,165 | 2,054.50 | 8,348.50 |
| 936 | 3,044.50 | 3,350.50 | 3,548 | 3,016 | 12,959 |
| 937 | 3,145.50 | 3,169 | 3,460 | 2,808 | 12,582.50 |
| 939 | 1,600 | 1,647.50 | 1,553.50 | 1,683.50 | 6,484.50 |
| 940 | 1,306 | 1,274 | 1,415 | 1,403 | 5,398 |
| 941 | 1,075 | 991 | 835.5 | 662.5 | 3,564 |
| 943 | 100.47 | 162.67 | 126 | 63 | 452.14 |
| 944 | 275.5 | 293 | 195 | 332.5 | 1,096 |
| 945 | 4,922 | 5,298.50 | 5,157 | 4,845 | 20,222.50 |
| 946 | 209.5 | 292 | 273.5 | 304 | 1,079 |
| 947 | 51.5 | 147 | 128 | 151 | 477.5 |
| 948 | 64 | 64 | 86.5 | 100 | 314.5 |
| 949 | 472.5 | 429.5 | 286 | 379 | 1,567 |
| 950 | 1,227.50 | 1,088.50 | 1,073.50 | 928.5 | 4,318 |
| 951 | 625.5 | 740 | 522 | 634 | 2,521.50 |
| 952 | 571 | 617.5 | 742 | 782.5 | 2,713 |
| 953 | 2,435 | 2,856.50 | 2,415.50 | 2,708.50 | 10,415.50 |
| 954 | 945.5 | 756.5 | 1,032 | 1,215.50 | 3,949.50 |
| 955 | 189 | 188 | 151 | 146.5 | 674.5 |
| 956 | 3,197.50 | 3,836.50 | 3,537 | 3,616.50 | 14,187.50 |
| 957 | 1,258 | 1,300.50 | 1,218 | 1,171.50 | 4,948 |

| | | | | | |
|---|---|---|---|---|---|
| 958 | 1,959.50 | 2,315.50 | 2,541.50 | 2,106.50 | 8,923 |
| 959 | 2,881.50 | 2,839.50 | 2,979 | 3,335.50 | 12,035.50 |
| 960 | 1,547 | 1,555 | 1,705 | 1,320 | 6,127 |
| 961 | 84 | 87 | 82.5 | 65 | 318.5 |
| TOTAL | 118,632.37 | 120,928.67 | 120,683.50 | 120,528.40 | 480,772.94 |

STATE: COLORADO

| ZIP CODE | QUARTER 1 | QUARTER 2 | QUARTER 3 | QUARTER 4 | TOTAL GRAMS |
|---|---|---|---|---|---|
| 800 | 4,312 | 4,083 | 4,036.50 | 4,320 | 16,751.50 |
| 801 | 5,607 | 5,513 | 5,851 | 5,770.50 | 22,741.50 |
| 802 | 3,472 | 3,395.50 | 3,574 | 3,514.50 | 13,956 |
| 803 | 561 | 596.5 | 553 | 544.5 | 2,255 |
| 804 | 794 | 744 | 710 | 627 | 2,875 |
| 805 | 3,453.50 | 3,228.50 | 3,261.50 | 3,707 | 13,650.50 |
| 806 | 1,613 | 1,818.50 | 1,848.50 | 1,626 | 6,906 |
| 807 | 355 | 452.5 | 431 | 503 | 1,741.50 |
| 808 | 413 | 343.5 | 513.5 | 612.5 | 1,882.50 |
| 809 | 4,076 | 3,792.50 | 3,536.50 | 3,749 | 15,154 |
| 810 | 1,342 | 1,343.50 | 1,469 | 1,634.50 | 5,789 |
| 811 | 53.5 | 42.5 | 63.5 | 77.5 | 237 |
| 812 | 354.5 | 438.5 | 399.5 | 396 | 1,588.50 |
| 813 | 357 | 361.5 | 309.5 | 381 | 1,409 |
| 814 | 104 | 76 | 85 | 152 | 417 |
| 815 | 591 | 554 | 444 | 586.5 | 2,175.50 |
| 816 | 251.5 | 237 | 225 | 241 | 954.5 |
| TOTAL | 27,710 | 27,020.50 | 27,311 | 28,442.50 | 110,484 |

STATE: CONNECTICUT

| ZIP CODE | QUARTER 1 | QUARTER 2 | QUARTER 3 | QUARTER 4 | TOTAL GRAMS |
|---|---|---|---|---|---|
| 60 | 1,842.50 | 1,449.50 | 1,957.50 | 1,873 | 7,122.50 |
| 61 | 362 | 251 | 369.5 | 380.5 | 1,363 |
| 62 | 356 | 385.5 | 730.5 | 704.5 | 2,176.50 |
| 63 | 1,667 | 2,064.50 | 2,801 | 2,775 | 9,307.50 |

DRUG CODE:9780DRUG NAME:TAPENTADOL
STATE:CONNECTICUT

| ZIP CODE | QUARTER 1 | QUARTER 2 | QUARTER 3 | QUARTER 4 | TOTAL GRAMS |
|---|---|---|---|---|---|
| 64 | 3,486.50 | 3,907 | 3,742.50 | 4,051 | 15,187 |
| 65 | 920 | 1,013.50 | 1,040 | 1,126.80 | 4,100.30 |
| 66 | 393 | 598.5 | 577 | 899 | 2,467.50 |
| 67 | 963 | 1,218 | 1,015 | 867 | 4,063 |
| 68 | 1,735.50 | 1,820.50 | 1,605.50 | 1,672.50 | 6,834 |
| 69 | 463 | 278 | 271 | 267 | 1,279 |
| TOTAL | 12,188.50 | 12,986 | 14,109.50 | 14,616.30 | 53,900.30 |

STATE: DELAWARE

| ZIP CODE | QUARTER 1 | QUARTER 2 | QUARTER 3 | QUARTER 4 | TOTAL GRAMS |
|---|---|---|---|---|---|
| 197 | 1,859.50 | 2,196.50 | 2,415.50 | 2,615.50 | 9,087 |
| 198 | 1,170 | 1,273.50 | 1,355 | 1,682 | 5,480.50 |
| 199 | 3,075.50 | 3,237.50 | 3,301 | 3,064.50 | 12,678.50 |
| TOTAL | 6,105 | 6,707.50 | 7,071.50 | 7,362 | 27,246 |

STATE: DISTRICT OF COLUMBIA

| ZIP CODE | QUARTER 1 | QUARTER 2 | QUARTER 3 | QUARTER 4 | TOTAL GRAMS |
|---|---|---|---|---|---|
| 200 | 1,448.85 | 1,474.50 | 1,456 | 1,321 | 5,700.35 |
| 203 | 0 | 0 | 0 | 45 | 45 |
| TOTAL | 1,448.85 | 1,474.50 | 1,456 | 1,366 | 5,745.35 |

STATE: FLORIDA

RETAIL DRUG DISTRIBUTION BY ZIP CODE WITHIN STATE BY GRAMS WT

| ZIP CODE | QUARTER 1 | QUARTER 2 | QUARTER 3 | QUARTER 4 | TOTAL GRAMS |
|---|---|---|---|---|---|
| 320 | 3,847.50 | 3,856.50 | 3,725 | 4,029.50 | 15,458.50 |
| 321 | 3,218 | 3,254.50 | 3,014.50 | 3,175.50 | 12,662.50 |
| 322 | 7,697 | 7,303.50 | 7,302 | 7,152 | 29,454.50 |
| 323 | 3,548.50 | 3,288 | 3,322 | 3,520.50 | 13,679 |
| 324 | 2,153 | 2,332 | 2,529.50 | 2,369 | 9,383.50 |
| 325 | 5,118 | 5,289.50 | 5,886.50 | 5,545 | 21,839 |
| 326 | 1,884.50 | 1,669.50 | 1,497.50 | 1,786 | 6,837.50 |
| 327 | 7,806.50 | 7,053.50 | 8,029 | 7,415.50 | 30,304.50 |
| 328 | 3,265 | 3,243.50 | 3,100 | 3,132 | 12,740.50 |
| 329 | 4,390.50 | 4,332 | 4,328 | 4,137.50 | 17,188 |
| 330 | 3,415 | 3,526 | 3,408 | 3,646 | 13,995 |
| 331 | 934.85 | 771.93 | 781 | 734 | 3,221.78 |
| 333 | 2,225.50 | 2,147 | 2,048.50 | 2,143 | 8,564 |
| 334 | 8,637.50 | 9,076 | 8,265.50 | 8,755 | 34,734 |
| 335 | 7,643 | 7,739 | 6,928.50 | 7,507 | 29,817.50 |
| 336 | 4,521.50 | 4,462.50 | 4,925.50 | 5,234 | 19,143.50 |
| 337 | 4,476.50 | 4,335.50 | 4,150 | 4,544.50 | 17,506.50 |
| 338 | 3,674 | 3,665 | 3,551 | 3,507 | 14,397 |
| 339 | 12,816 | 12,888 | 12,532 | 12,155 | 50,391 |
| 341 | 5,577 | 4,905 | 5,089 | 5,121 | 20,692 |
| 342 | 7,102.50 | 7,474.50 | 6,902.50 | 7,394 | 28,873.50 |
| 344 | 2,614 | 2,745 | 2,825.50 | 2,768.50 | 10,953 |
| 346 | 4,597.50 | 4,584 | 4,967 | 4,622 | 18,770.50 |
| 347 | 3,740 | 3,866.50 | 3,601 | 3,466 | 14,673.50 |
| 349 | 2,648.50 | 2,555.50 | 2,499.50 | 2,699.50 | 10,403 |
| TOTAL | 117,551.85 | 116,363.93 | 115,208.50 | 116,559 | 465,683.28 |

STATE: GEORGIA

| ZIP CODE | QUARTER 1 | QUARTER 2 | QUARTER 3 | QUARTER 4 | TOTAL GRAMS |
|---|---|---|---|---|---|
| 300 | 12,819.50 | 13,191 | 13,175.50 | 13,750 | 52,936 |
| 301 | 10,795.50 | 10,992 | 12,034 | 12,213.50 | 46,035 |
| 302 | 5,402.50 | 5,303 | 5,527 | 5,670.50 | 21,903 |
| 303 | 3,135 | 3,744.50 | 3,434 | 3,752.50 | 14,066 |
| 304 | 3,553 | 3,834.50 | 4,097 | 4,045 | 15,529.50 |
| 305 | 3,612.50 | 3,886 | 4,186.50 | 4,504 | 16,189 |
| 306 | 2,234.50 | 2,363.50 | 2,002.50 | 1,965 | 8,565.50 |
| 307 | 1,508 | 1,477.50 | 1,573.50 | 1,625 | 6,184 |
| 308 | 760.5 | 848.5 | 836 | 760.5 | 3,205.50 |
| 309 | 839.5 | 713 | 829.5 | 881.5 | 3,263.50 |
| 310 | 3,853.50 | 3,866.50 | 3,833.50 | 4,280.50 | 15,834 |
| 312 | 1,504.50 | 1,220.50 | 1,364 | 1,234 | 5,323 |
| 313 | 4,344 | 3,985 | 4,491 | 4,430.50 | 17,250.50 |
| 314 | 2,594.50 | 2,864.50 | 2,751.50 | 2,816.50 | 11,027 |
| 315 | 7,925.50 | 7,322 | 7,395.50 | 7,396.50 | 30,039.50 |
| 316 | 1,456 | 1,441 | 1,644.50 | 1,605.50 | 6,147 |
| 317 | 1,616.50 | 1,556 | 1,445 | 1,881.50 | 6,499 |
| 318 | 214 | 217 | 208 | 154.5 | 793.5 |

DRUG CODE:9780DRUG NAME:TAPENTADOL
STATE:GEORGIA

| ZIP CODE | QUARTER 1 | QUARTER 2 | QUARTER 3 | QUARTER 4 | TOTAL GRAMS |
|---|---|---|---|---|---|
| 319 | 1,683.50 | 1,357 | 1,361 | 1,344.50 | 5,746 |
| 398 | 551 | 566 | 464 | 526 | 2,107 |
| TOTAL | 70,403.50 | 70,749 | 72,653.50 | 74,837.50 | 288,643.50 |

STATE: HAWAII

| ZIP CODE | QUARTER 1 | QUARTER 2 | QUARTER 3 | QUARTER 4 | TOTAL GRAMS |
|---|---|---|---|---|---|
| 967 | 1,573.50 | 1,748.50 | 1,321 | 1,411.50 | 6,054.50 |

| | | | | | |
|---|---|---|---|---|---|
| 968 | 1,214.50 | 1,111 | 1,293 | 1,398.50 | 5,017 |
| TOTAL | 2,788 | 2,859.50 | 2,614 | 2,810 | 11,071.50 |

STATE: IDAHO

| ZIP CODE | QUARTER 1 | QUARTER 2 | QUARTER 3 | QUARTER 4 | TOTAL GRAMS |
|---|---|---|---|---|---|
| 832 | 686.5 | 684 | 771.5 | 1,280.50 | 3,422.50 |
| 833 | 411.5 | 431 | 426.5 | 492 | 1,761 |
| 834 | 546.5 | 659 | 784 | 966.5 | 2,956 |
| 835 | 207 | 275.5 | 203 | 221 | 906.5 |
| 836 | 1,555 | 1,698 | 1,745 | 1,924 | 6,922 |
| 837 | 1,393.50 | 1,570.50 | 1,541.50 | 1,540 | 6,045.50 |
| 838 | 594.5 | 608.5 | 744 | 767 | 2,714 |
| TOTAL | 5,394.50 | 5,926.50 | 6,215.50 | 7,191 | 24,727.50 |

STATE: ILLINOIS

| ZIP CODE | QUARTER 1 | QUARTER 2 | QUARTER 3 | QUARTER 4 | TOTAL GRAMS |
|---|---|---|---|---|---|
| 600 | 7,814 | 8,275.50 | 7,858.50 | 7,933 | 31,881 |
| 601 | 4,069.50 | 4,423 | 4,062 | 4,426 | 16,980.50 |
| 602 | 380.5 | 400.5 | 323 | 253.5 | 1,357.50 |
| 603 | 480 | 426 | 556.5 | 564 | 2,026.50 |
| 604 | 5,087 | 5,146 | 5,148.50 | 5,832 | 21,213.50 |
| 605 | 4,554.50 | 4,638.50 | 4,944 | 4,725 | 18,862 |
| 606 | 4,140.50 | 3,726 | 3,617.50 | 3,641 | 15,125 |
| 607 | 257 | 356 | 505.5 | 523.5 | 1,642 |
| 608 | 150 | 96 | 82 | 81 | 409 |
| 609 | 831 | 925 | 941 | 866 | 3,563 |
| 610 | 746 | 847.5 | 1,042 | 1,080 | 3,715.50 |
| 611 | 1,359.50 | 1,951 | 1,788 | 1,656.50 | 6,755 |
| 612 | 260 | 213.5 | 282.5 | 237 | 993 |
| 613 | 234.5 | 211 | 342 | 361 | 1,148.50 |
| 614 | 96 | 134 | 134 | 139 | 503 |
| 615 | 279.5 | 318.5 | 308 | 382 | 1,288 |
| 616 | 264.5 | 402.5 | 282.5 | 431.5 | 1,381 |
| 617 | 1,083 | 1,213 | 1,026.50 | 1,177.50 | 4,500 |
| 618 | 1,693.50 | 1,990 | 1,635 | 1,560.50 | 6,879 |
| 619 | 277.5 | 382 | 406 | 350 | 1,415.50 |
| 620 | 1,778 | 2,231 | 2,320.50 | 2,307 | 8,636.50 |
| 622 | 4,110.50 | 4,166 | 4,664 | 4,958 | 17,898.50 |
| 623 | 262 | 301 | 323 | 298.5 | 1,184.50 |
| 624 | 224 | 303 | 383 | 368 | 1,278 |
| 625 | 626 | 648.5 | 800 | 884 | 2,958.50 |
| 626 | 138.5 | 157 | 206 | 242 | 743.5 |
| 627 | 280 | 289 | 245 | 357 | 1,171 |
| 628 | 502 | 525.5 | 675 | 613 | 2,315.50 |
| 629 | 455 | 451 | 491 | 488 | 1,885 |
| TOTAL | 42,434 | 45,147.50 | 45,392.50 | 46,735.50 | 179,709.50 |

STATE: INDIANA

| ZIP CODE | QUARTER 1 | QUARTER 2 | QUARTER 3 | QUARTER 4 | TOTAL GRAMS |
|---|---|---|---|---|---|
| 460 | 2,085 | 1,888.50 | 1,896.50 | 1,826.50 | 7,696.50 |
| 461 | 3,294.50 | 3,263.50 | 2,954.50 | 2,902 | 12,414.50 |
| 462 | 2,731 | 2,395 | 2,732.50 | 2,827 | 10,685.50 |
| 463 | 4,040.50 | 4,010 | 3,614.50 | 3,952.50 | 15,617.50 |
| 464 | 210.5 | 186.5 | 180 | 330.5 | 907.5 |
| 465 | 2,807.50 | 2,480 | 2,425.50 | 2,512 | 10,225 |
| 466 | 793 | 786 | 747 | 693.5 | 3,019.50 |
| 467 | 3,413 | 3,523 | 3,644.50 | 3,679 | 14,259.50 |
| 468 | 4,196.50 | 4,419.50 | 4,443 | 4,501 | 17,560 |
| 469 | 1,760 | 1,721 | 1,634 | 1,768.50 | 6,883.50 |
| 470 | 366 | 270 | 558.5 | 527.5 | 1,722 |
| 471 | 1,601.50 | 1,458.50 | 1,486 | 1,496 | 6,042 |

| | | | | | |
|---|---|---|---|---|---|
| 472 | 553 | 958 | 739 | 1,131 | 3,381 |
| 473 | 1,184.50 | 1,261 | 1,424 | 1,289 | 5,158.50 |

DRUG CODE:9780DRUG NAME:TAPENTADOL
STATE:INDIANA

| ZIP CODE | QUARTER 1 | QUARTER 2 | QUARTER 3 | QUARTER 4 | TOTAL GRAMS |
|---|---|---|---|---|---|
| 474 | 576.5 | 492 | 704 | 779 | 2,551.50 |
| 475 | 880 | 902.5 | 969 | 979.5 | 3,731 |
| 476 | 1,486.50 | 1,196.50 | 1,077 | 1,179 | 4,939 |
| 477 | 2,047 | 2,158.50 | 1,777 | 2,062.50 | 8,045 |
| 478 | 4,795 | 5,517 | 5,576.50 | 5,624.50 | 21,513 |
| 479 | 871.5 | 1,231 | 1,336 | 1,565 | 5,003.50 |
| TOTAL | 39,693 | 40,118 | 39,919 | 41,625.50 | 161,355.50 |

STATE: IOWA

| ZIP CODE | QUARTER 1 | QUARTER 2 | QUARTER 3 | QUARTER 4 | TOTAL GRAMS |
|---|---|---|---|---|---|
| 500 | 1,361.50 | 1,610 | 1,553 | 1,792 | 6,316.50 |
| 501 | 896 | 977 | 1,197 | 1,536.50 | 4,606.50 |
| 502 | 1,569 | 1,579 | 1,644.50 | 1,869 | 6,661.50 |
| 503 | 2,533.50 | 2,750.50 | 2,792 | 3,188.50 | 11,264.50 |
| 504 | 115 | 127 | 161 | 120 | 523 |
| 505 | 231 | 272 | 289 | 228 | 1,020 |
| 506 | 613 | 582 | 568.5 | 441 | 2,204.50 |
| 507 | 580.5 | 471.5 | 452 | 484 | 1,988 |
| 508 | 173 | 118 | 164 | 195 | 650 |
| 510 | 274 | 320 | 181 | 235 | 1,010 |
| 511 | 273 | 172 | 129 | 90 | 664 |
| 512 | 142.5 | 160 | 162.5 | 145 | 610 |
| 513 | 45 | 85 | 60 | 77.5 | 267.5 |
| 514 | 53 | 36 | 25 | 25 | 139 |
| 515 | 392 | 532 | 561.5 | 704 | 2,189.50 |
| 516 | 231 | 288 | 318 | 241.5 | 1,078.50 |
| 520 | 672 | 681.5 | 557 | 495 | 2,405.50 |
| 521 | 27 | 40 | 41 | 18 | 126 |
| 522 | 271 | 305 | 310 | 158 | 1,044 |
| 523 | 309.5 | 328 | 335 | 432 | 1,404.50 |
| 524 | 585.5 | 667 | 495 | 807.5 | 2,555 |
| 525 | 359.5 | 303 | 384.5 | 341 | 1,388 |
| 526 | 97 | 103 | 75 | 88 | 363 |
| 527 | 264 | 235 | 209.5 | 223 | 931.5 |
| 528 | 330.5 | 424.5 | 217 | 293.5 | 1,265.50 |
| TOTAL | 12,399 | 13,167 | 12,882 | 14,228 | 52,676 |

STATE: KANSAS

| ZIP CODE | QUARTER 1 | QUARTER 2 | QUARTER 3 | QUARTER 4 | TOTAL GRAMS |
|---|---|---|---|---|---|
| 660 | 2,701 | 3,025 | 2,620.50 | 2,817.50 | 11,164 |
| 661 | 194 | 134 | 141.5 | 99.5 | 569 |
| 662 | 3,386.50 | 3,459.50 | 3,289.50 | 3,051 | 13,186.50 |
| 664 | 267 | 359 | 416 | 286 | 1,328 |
| 665 | 309 | 236 | 264 | 311 | 1,120 |
| 666 | 1,229.50 | 1,266.50 | 1,353 | 1,467.50 | 5,316.50 |
| 667 | 408.5 | 564 | 546.5 | 533.5 | 2,052.50 |
| 668 | 175 | 171 | 111 | 94 | 551 |
| 669 | 21 | 47 | 31 | 86 | 185 |
| 670 | 329.5 | 314.5 | 321.5 | 273 | 1,238.50 |
| 671 | 101 | 106 | 113 | 97 | 417 |
| 672 | 897 | 703 | 877.5 | 693 | 3,170.50 |
| 673 | 422.5 | 299 | 309 | 432 | 1,462.50 |
| 674 | 404 | 320.5 | 465 | 346 | 1,535.50 |

| | | | | | |
|---|---|---|---|---|---|
| 675 | 122.5 | 143 | 114 | 125 | 504.5 |
| 676 | 194.5 | 263 | 259.5 | 201.5 | 918.5 |
| 677 | 147.5 | 124 | 123 | 121 | 515.5 |
| 678 | 49 | 42 | 47.5 | 32 | 170.5 |
| 679 | 0 | 0 | 33 | 31 | 64 |
| TOTAL | 11,359 | 11,577 | 11,436 | 11,097.50 | 45,469.50 |

STATE: KENTUCKY

| ZIP CODE | QUARTER 1 | QUARTER 2 | QUARTER 3 | QUARTER 4 | TOTAL GRAMS |
|---|---|---|---|---|---|
| 400 | 747 | 829 | 616 | 730.5 | 2,922.50 |
| 401 | 488 | 351.5 | 390.5 | 336 | 1,566 |
| 402 | 3,338.50 | 3,197.50 | 2,768 | 2,988.30 | 12,292.30 |
| 403 | 449.5 | 417.5 | 455.5 | 741.5 | 2,064 |
| 404 | 258 | 283 | 218 | 209.5 | 968.5 |
| 405 | 491.5 | 527.5 | 607 | 659.5 | 2,285.50 |
| 406 | 22.5 | 38.5 | 76 | 112.5 | 249.5 |

DRUG CODE:9780DRUG NAME:TAPENTADOL
STATE:KENTUCKY

| ZIP CODE | QUARTER 1 | QUARTER 2 | QUARTER 3 | QUARTER 4 | TOTAL GRAMS |
|---|---|---|---|---|---|
| 407 | 359 | 284 | 249.5 | 278 | 1,170.50 |
| 408 | 31 | 40 | 20 | 40 | 131 |
| 409 | 132 | 101 | 147.5 | 179.5 | 560 |
| 410 | 2,759.50 | 2,480 | 2,769.50 | 2,741.50 | 10,750.50 |
| 411 | 248 | 221 | 228.5 | 312.5 | 1,010 |
| 412 | 107 | 86 | 125 | 126 | 444 |
| 413 | 30.5 | 25 | 34.5 | 51.5 | 141.5 |
| 414 | 11 | 0 | 0 | 9 | 20 |
| 415 | 460 | 371 | 360.5 | 349 | 1,540.50 |
| 416 | 256.5 | 193.5 | 251 | 236 | 937 |
| 417 | 116.5 | 85.5 | 106 | 66 | 374 |
| 418 | 55.5 | 78 | 42 | 80 | 255.5 |
| 420 | 1,867.50 | 1,931.50 | 1,975 | 1,806.50 | 7,580.50 |
| 421 | 577.5 | 713 | 538 | 550 | 2,378.50 |
| 422 | 547 | 519.5 | 1,218.50 | 827 | 3,112 |
| 423 | 1,739 | 1,583 | 1,643.50 | 1,984.50 | 6,950 |
| 424 | 870 | 764 | 832.5 | 930 | 3,396.50 |
| 425 | 112 | 65 | 91.5 | 34 | 302.5 |
| 426 | 39.5 | 41 | 65.5 | 43.5 | 189.5 |
| 427 | 637 | 557.5 | 557.5 | 721 | 2,473 |
| TOTAL | 16,751 | 15,784 | 16,387 | 17,143.30 | 66,065.30 |

STATE: LOUISIANA

| ZIP CODE | QUARTER 1 | QUARTER 2 | QUARTER 3 | QUARTER 4 | TOTAL GRAMS |
|---|---|---|---|---|---|
| 700 | 1,691 | 2,006.50 | 1,938.50 | 2,123 | 7,759 |
| 701 | 200 | 170.5 | 224 | 242.5 | 837 |
| 703 | 557 | 588.5 | 704 | 574 | 2,423.50 |
| 704 | 2,956 | 3,252 | 3,405 | 3,434.50 | 13,047.50 |
| 705 | 2,006 | 1,886 | 1,930 | 1,889 | 7,711 |
| 706 | 367 | 493.5 | 390.5 | 412 | 1,663 |
| 707 | 2,504 | 2,892 | 3,102.50 | 3,153.50 | 11,652 |
| 708 | 1,661 | 1,748 | 1,734 | 1,822 | 6,965 |
| 710 | 293.5 | 296.5 | 382.5 | 374.5 | 1,347 |
| 711 | 1,063.50 | 1,127.50 | 1,179.50 | 1,365.50 | 4,736 |
| 712 | 444.5 | 382.5 | 478.5 | 412 | 1,717.50 |
| 713 | 1,063.50 | 883 | 816.5 | 812 | 3,575 |
| 714 | 783 | 540 | 678.5 | 394.5 | 2,396 |
| TOTAL | 15,590 | 16,266.50 | 16,964 | 17,009 | 65,829.50 |

STATE: MAINE

| ZIP CODE | QUARTER 1 | QUARTER 2 | QUARTER 3 | QUARTER 4 | TOTAL GRAMS |
|---|---|---|---|---|---|

| | | | | | |
|---|---|---|---|---|---|
| 39 | 114 | 81 | 77 | 110.5 | 382.5 |
| 40 | 501 | 642.5 | 703.5 | 632 | 2,479 |
| 41 | 42 | 63 | 28 | 85 | 218 |
| 42 | 406 | 601.5 | 518 | 421 | 1,946.50 |
| 43 | 111 | 195 | 141 | 137 | 584 |
| 44 | 66 | 48 | 40 | 53 | 207 |
| 45 | 96 | 91.5 | 69 | 133.5 | 390 |
| 46 | 10 | 16 | 47 | 6 | 79 |
| 47 | 50 | 50 | 75 | 86.5 | 261.5 |
| 48 | 73 | 44.5 | 36 | 27 | 180.5 |
| 49 | 390 | 381.5 | 367.5 | 343.5 | 1,482.50 |
| TOTAL | 1,859 | 2,214.50 | 2,102 | 2,035 | 8,210.50 |

STATE: MARYLAND

| ZIP CODE | QUARTER 1 | QUARTER 2 | QUARTER 3 | QUARTER 4 | TOTAL GRAMS |
|---|---|---|---|---|---|
| 206 | 2,985 | 2,920 | 3,052 | 3,301.50 | 12,258.50 |
| 207 | 3,226.50 | 3,420 | 3,262.50 | 2,977.50 | 12,886.50 |
| 208 | 3,127 | 3,619 | 3,805 | 3,811 | 14,362 |
| 209 | 592.5 | 759.5 | 738.5 | 578.5 | 2,669 |
| 210 | 7,153 | 7,459 | 7,250 | 7,516 | 29,378 |
| 211 | 2,964 | 3,159.50 | 3,327.50 | 3,350 | 12,801 |
| 212 | 4,443.50 | 4,337.50 | 4,318.50 | 4,395 | 17,494.50 |
| 214 | 985.5 | 1,151.50 | 1,332.50 | 1,089.50 | 4,559 |
| 215 | 837 | 771 | 780.5 | 707.5 | 3,096 |
| 216 | 1,420.50 | 1,392 | 1,343 | 1,307.50 | 5,463 |
| 217 | 5,338.50 | 5,344 | 5,381.50 | 5,354.50 | 21,418.50 |
| 218 | 421 | 447.5 | 665.5 | 659 | 2,193 |
| 219 | 523.5 | 675 | 494.5 | 471 | 2,164 |

DRUG CODE:9780DRUG NAME:TAPENTADOL
STATE:MARYLAND

| ZIP CODE | QUARTER 1 | QUARTER 2 | QUARTER 3 | QUARTER 4 | TOTAL GRAMS |
|---|---|---|---|---|---|
| TOTAL | 34,017.50 | 35,455.50 | 35,751.50 | 35,518.50 | 140,743 |

STATE: MASSACHUSETTS

| ZIP CODE | QUARTER 1 | QUARTER 2 | QUARTER 3 | QUARTER 4 | TOTAL GRAMS |
|---|---|---|---|---|---|
| 10 | 867.5 | 926.5 | 1,110 | 1,145 | 4,049 |
| 11 | 324 | 346 | 412.5 | 503.5 | 1,586 |
| 12 | 91 | 102.5 | 140 | 100 | 433.5 |
| 13 | 98.5 | 113 | 89 | 111.5 | 412 |
| 14 | 565.5 | 486 | 482.5 | 384.5 | 1,918.50 |
| 15 | 565 | 462 | 598 | 696 | 2,321 |
| 16 | 176 | 270 | 266 | 213 | 925 |
| 17 | 482.5 | 420.5 | 603.5 | 574 | 2,080.50 |
| 18 | 8,742 | 8,902.50 | 9,327 | 9,280.50 | 36,252 |
| 19 | 1,285 | 1,078 | 953.5 | 1,176 | 4,492.50 |
| 20 | 468.5 | 419 | 462 | 438 | 1,787.50 |
| 21 | 542 | 478.5 | 574 | 603.5 | 2,198 |
| 22 | 10 | 15 | 22 | 59 | 106 |
| 23 | 904 | 845 | 827.5 | 930.5 | 3,507 |
| 24 | 366 | 450.5 | 331.5 | 405 | 1,553 |
| 25 | 272 | 247.5 | 217.5 | 185.5 | 922.5 |
| 26 | 459.5 | 414.5 | 285.5 | 299.5 | 1,459 |
| 27 | 1,693 | 1,858.50 | 1,855 | 1,818 | 7,224.50 |
| TOTAL | 17,912 | 17,835.50 | 18,557 | 18,923 | 73,227.50 |

STATE: MICHIGAN

| ZIP CODE | QUARTER 1 | QUARTER 2 | QUARTER 3 | QUARTER 4 | TOTAL GRAMS |
|---|---|---|---|---|---|
| 480 | 2,559.50 | 3,000 | 2,547 | 3,133 | 11,239.50 |

| | | | | | |
|---|---|---|---|---|---|
| 481 | 3,225 | 3,700 | 4,007.50 | 3,793.50 | 14,726 |
| 482 | 442.5 | 485 | 533 | 629 | 2,089.50 |
| 483 | 2,613.50 | 2,808.50 | 2,740 | 2,724.50 | 10,886.50 |
| 484 | 524.5 | 669 | 735 | 695.5 | 2,624 |
| 485 | 350 | 331 | 385.5 | 474.5 | 1,541 |
| 486 | 956 | 1,073 | 1,005.50 | 908.5 | 3,943 |
| 487 | 559 | 414.5 | 486 | 383 | 1,842.50 |
| 488 | 1,734 | 1,670.50 | 1,810 | 1,852.50 | 7,067 |
| 489 | 849.5 | 695.5 | 646.5 | 643 | 2,834.50 |
| 490 | 2,695.50 | 2,801 | 2,496.50 | 2,516 | 10,509 |
| 491 | 560 | 483.5 | 367 | 388 | 1,798.50 |
| 492 | 708 | 770.5 | 558.5 | 562.5 | 2,599.50 |
| 493 | 1,275.50 | 1,192.50 | 929 | 979 | 4,376 |
| 494 | 966 | 1,202.50 | 1,361.50 | 1,300 | 4,830 |
| 495 | 1,490.50 | 1,389 | 1,808 | 1,761.50 | 6,449 |
| 496 | 423 | 509.5 | 487 | 645 | 2,064.50 |
| 497 | 500.5 | 681 | 738 | 590 | 2,509.50 |
| 498 | 367 | 350 | 253.5 | 264 | 1,234.50 |
| 499 | 82 | 145 | 95 | 110 | 432 |
| TOTAL | 22,881.50 | 24,371.50 | 23,990 | 24,353 | 95,596 |

STATE: MINNESOTA

| ZIP CODE | QUARTER 1 | QUARTER 2 | QUARTER 3 | QUARTER 4 | TOTAL GRAMS |
|---|---|---|---|---|---|
| 550 | 781 | 735.5 | 894 | 734.5 | 3,145 |
| 551 | 1,175 | 1,471 | 749 | 986.5 | 4,381.50 |
| 553 | 617 | 615 | 702.5 | 817 | 2,751.50 |
| 554 | 622.5 | 452.5 | 497.5 | 460 | 2,032.50 |
| 556 | 5 | 5 | 11 | 5 | 26 |
| 557 | 47 | 27 | 0 | 6 | 80 |
| 558 | 94.5 | 66 | 144 | 136 | 440.5 |
| 559 | 229.5 | 229.5 | 211.5 | 230.5 | 901 |
| 560 | 170.5 | 339.5 | 266 | 230.5 | 1,006.50 |
| 561 | 223.5 | 182.5 | 127.5 | 218 | 751.5 |
| 562 | 273 | 274.5 | 241 | 223 | 1,011.50 |
| 563 | 187 | 245.5 | 221.5 | 220 | 874 |
| 564 | 119 | 48 | 95.5 | 160 | 422.5 |
| 565 | 168.5 | 149 | 276 | 304.5 | 898 |
| 566 | 126.5 | 99 | 135 | 118.5 | 479 |
| 567 | 307.5 | 304 | 269 | 280 | 1,160.50 |
| TOTAL | 5,147 | 5,243.50 | 4,841 | 5,130 | 20,361.50 |

STATE: MISSISSIPPI

| ZIP CODE | QUARTER 1 | QUARTER 2 | QUARTER 3 | QUARTER 4 | TOTAL GRAMS |
|---|---|---|---|---|---|

DRUG CODE:9780DRUG NAME:TAPENTADOL
STATE:MISSISSIPPI

| ZIP CODE | QUARTER 1 | QUARTER 2 | QUARTER 3 | QUARTER 4 | TOTAL GRAMS |
|---|---|---|---|---|---|
| 386 | 1,341.50 | 1,278 | 1,258.50 | 1,281.50 | 5,159.50 |
| 387 | 238 | 198.5 | 194.5 | 271.5 | 902.5 |
| 388 | 1,186 | 1,490 | 1,399.50 | 1,247 | 5,322.50 |
| 389 | 210.5 | 246.5 | 346.5 | 294.5 | 1,098 |
| 390 | 947.5 | 1,016.50 | 1,318 | 1,341.50 | 4,623.50 |
| 391 | 695 | 1,075.50 | 972 | 1,064 | 3,806.50 |
| 392 | 1,064 | 980.5 | 1,025.50 | 1,306.50 | 4,376.50 |
| 393 | 822 | 898.5 | 989 | 1,026.50 | 3,736 |
| 394 | 1,724.50 | 2,073.50 | 2,172 | 1,806.50 | 7,776.50 |
| 395 | 5,062 | 4,573.50 | 4,273.50 | 4,085 | 17,994 |
| 396 | 130 | 215 | 237.5 | 345 | 927.5 |
| 397 | 436.5 | 500.5 | 494.5 | 631 | 2,062.50 |

| | | | | | |
|---|---|---|---|---|---|
| TOTAL | 13,857.50 | 14,546.50 | 14,681 | 14,700.50 | 57,785.50 |

STATE: MISSOURI

| ZIP CODE | QUARTER 1 | QUARTER 2 | QUARTER 3 | QUARTER 4 | TOTAL GRAMS |
|---|---|---|---|---|---|
| 630 | 4,019 | 4,015 | 4,080.50 | 3,844.50 | 15,959 |
| 631 | 6,501 | 6,623.50 | 6,172.50 | 6,516.50 | 25,813.50 |
| 633 | 3,110 | 2,923 | 2,933.50 | 3,095.50 | 12,062 |
| 634 | 201.5 | 188 | 198 | 249.5 | 837 |
| 635 | 202 | 217.5 | 198 | 230 | 847.5 |
| 636 | 455 | 362 | 461 | 343.5 | 1,621.50 |
| 637 | 1,315.50 | 1,363.50 | 1,345.50 | 1,235 | 5,259.50 |
| 638 | 534.5 | 410 | 370 | 454 | 1,768.50 |
| 639 | 324 | 402 | 273 | 263.5 | 1,262.50 |
| 640 | 2,478.50 | 2,286.50 | 2,031 | 2,524 | 9,320 |
| 641 | 2,309.50 | 1,945.50 | 1,936 | 1,955 | 8,146 |
| 644 | 186 | 174 | 190 | 201 | 751 |
| 645 | 635.5 | 657.5 | 677.5 | 801.5 | 2,772 |
| 646 | 111 | 133 | 133 | 82 | 459 |
| 647 | 822 | 816.5 | 600 | 615.5 | 2,854 |
| 648 | 756 | 1,012 | 1,263.50 | 1,193.50 | 4,225 |
| 650 | 350.5 | 312 | 281 | 330 | 1,273.50 |
| 651 | 693.5 | 823 | 684 | 692 | 2,892.50 |
| 652 | 433.5 | 435 | 394.5 | 305 | 1,568 |
| 653 | 796 | 586 | 612 | 657.5 | 2,651.50 |
| 654 | 169.5 | 203.5 | 213.5 | 205.5 | 792 |
| 655 | 260.5 | 378.5 | 313 | 304.5 | 1,256.50 |
| 656 | 720 | 762.5 | 753 | 769 | 3,004.50 |
| 657 | 1,079 | 997.5 | 981.5 | 803 | 3,861 |
| 658 | 1,224.50 | 1,015.50 | 1,053.50 | 866.5 | 4,160 |
| TOTAL | 29,688 | 29,043 | 28,149 | 28,537.50 | 115,417.50 |

STATE: MONTANA

| ZIP CODE | QUARTER 1 | QUARTER 2 | QUARTER 3 | QUARTER 4 | TOTAL GRAMS |
|---|---|---|---|---|---|
| 590 | 222 | 231.5 | 214 | 205 | 872.5 |
| 591 | 476 | 502 | 559 | 530 | 2,067 |
| 592 | 0 | 0 | 0 | 17 | 17 |
| 593 | 135 | 93 | 173 | 65 | 466 |
| 594 | 358.5 | 534.5 | 522 | 600.5 | 2,015.50 |
| 595 | 84 | 71 | 124 | 151.5 | 430.5 |
| 596 | 127.5 | 186.5 | 114 | 191 | 619 |
| 597 | 534.5 | 427.5 | 411 | 426 | 1,799 |
| 598 | 313 | 354.5 | 348 | 298 | 1,313.50 |
| 599 | 186.5 | 125.5 | 91.5 | 105 | 508.5 |
| TOTAL | 2,437 | 2,526 | 2,556.50 | 2,589 | 10,108.50 |

STATE: NEBRASKA

| ZIP CODE | QUARTER 1 | QUARTER 2 | QUARTER 3 | QUARTER 4 | TOTAL GRAMS |
|---|---|---|---|---|---|
| 680 | 1,074.50 | 1,066.50 | 1,086 | 1,024 | 4,251 |
| 681 | 4,300 | 4,206.50 | 4,158.50 | 4,595 | 17,260 |
| 683 | 216 | 188 | 199 | 257 | 860 |
| 684 | 276 | 220 | 231 | 158.5 | 885.5 |
| 685 | 1,730.50 | 1,563 | 1,705.50 | 1,751.50 | 6,750.50 |
| 686 | 487.5 | 579 | 498.5 | 372 | 1,937 |
| 687 | 383.5 | 452 | 492 | 452 | 1,779.50 |
| 688 | 1,399.50 | 1,512.50 | 1,299.50 | 1,511.50 | 5,723 |
| 689 | 212 | 250 | 338 | 282.5 | 1,082.50 |
| 690 | 66 | 66 | 46.5 | 81 | 259.5 |

DRUG CODE:9780DRUG NAME:TAPENTADOL
STATE:NEBRASKA

| ZIP CODE | QUARTER 1 | QUARTER 2 | QUARTER 3 | QUARTER 4 | TOTAL GRAMS |
|---|---|---|---|---|---|

RETAIL DRUG DISTRIBUTION BY ZIP CODE WITHIN STATE BY GRAMS WT

| | | | | | |
|---|---|---|---|---|---|
| 691 | 313 | 214 | 289.5 | 307 | 1,123.50 |
| 692 | 5 | 7.5 | 5 | 12.5 | 30 |
| 693 | 67.5 | 58 | 75.5 | 88.5 | 289.5 |
| TOTAL | 10,531 | 10,383 | 10,424.50 | 10,893 | 42,231.50 |

STATE: NEVADA

| ZIP CODE | QUARTER 1 | QUARTER 2 | QUARTER 3 | QUARTER 4 | TOTAL GRAMS |
|---|---|---|---|---|---|
| 890 | 3,666 | 3,802 | 3,416 | 3,476 | 14,360 |
| 891 | 7,623 | 7,112.50 | 6,939 | 7,391.50 | 29,066 |
| 893 | 5 | 5 | 0 | 9 | 19 |
| 894 | 616 | 754 | 740 | 811.5 | 2,921.50 |
| 895 | 718.5 | 926 | 753.5 | 796 | 3,194 |
| 897 | 132 | 233 | 331 | 218 | 914 |
| 898 | 233 | 230.5 | 287.5 | 241.5 | 992.5 |
| TOTAL | 12,993.50 | 13,063 | 12,467 | 12,943.50 | 51,467 |

STATE: NEW HAMPSHIRE

| ZIP CODE | QUARTER 1 | QUARTER 2 | QUARTER 3 | QUARTER 4 | TOTAL GRAMS |
|---|---|---|---|---|---|
| 30 | 1,802.50 | 1,670.50 | 2,015.50 | 1,732.50 | 7,221 |
| 31 | 555.5 | 611 | 571 | 629.5 | 2,367 |
| 32 | 1,097 | 1,163.50 | 1,247 | 1,145 | 4,652.50 |
| 33 | 681.5 | 611.5 | 630 | 709 | 2,632 |
| 34 | 122 | 68 | 83 | 88 | 361 |
| 35 | 312.5 | 337 | 300.5 | 294.5 | 1,244.50 |
| 37 | 133.5 | 107.5 | 87.5 | 87.5 | 416 |
| 38 | 1,568 | 1,521.50 | 1,891 | 2,296.50 | 7,277 |
| TOTAL | 6,272.50 | 6,090.50 | 6,825.50 | 6,982.50 | 26,171 |

STATE: NEW JERSEY

| ZIP CODE | QUARTER 1 | QUARTER 2 | QUARTER 3 | QUARTER 4 | TOTAL GRAMS |
|---|---|---|---|---|---|
| 70 | 6,363.50 | 7,187.50 | 7,198.50 | 6,806 | 27,555.50 |
| 71 | 506 | 472 | 491.5 | 551.5 | 2,021 |
| 72 | 550.5 | 466 | 414.5 | 490 | 1,921 |
| 73 | 331.5 | 406.5 | 455.5 | 479 | 1,672.50 |
| 74 | 2,782.50 | 2,463 | 2,604 | 2,455 | 10,304.50 |
| 75 | 297.5 | 278.5 | 232.5 | 360.5 | 1,169 |
| 76 | 2,608.50 | 2,648.50 | 2,182.50 | 2,675 | 10,114.50 |
| 77 | 5,546 | 6,260.50 | 5,812.50 | 5,874.50 | 23,493.50 |
| 78 | 2,179 | 2,016 | 2,199.50 | 2,110 | 8,504.50 |
| 79 | 1,234.50 | 1,133.50 | 1,293.50 | 1,267 | 4,928.50 |
| 80 | 8,739 | 9,347.50 | 9,471 | 10,205 | 37,762.50 |
| 81 | 408.5 | 434.5 | 491.5 | 621.5 | 1,956 |
| 82 | 2,166.50 | 2,048 | 2,082 | 2,180 | 8,476.50 |
| 83 | 1,691.50 | 1,962.50 | 1,603 | 2,007 | 7,264 |
| 84 | 46 | 55 | 34 | 82 | 217 |
| 85 | 2,083.50 | 2,160.50 | 1,862.50 | 2,212 | 8,318.50 |
| 86 | 1,803 | 1,800.50 | 1,918.50 | 1,805 | 7,327 |
| 87 | 3,803 | 3,693.50 | 3,705.50 | 3,668.50 | 14,870.50 |
| 88 | 5,888.50 | 6,034 | 6,051 | 5,419 | 23,392.50 |
| 89 | 516 | 555 | 618 | 544 | 2,233 |
| TOTAL | 49,545 | 51,423 | 50,721.50 | 51,812.50 | 203,502 |

STATE: NEW MEXICO

| ZIP CODE | QUARTER 1 | QUARTER 2 | QUARTER 3 | QUARTER 4 | TOTAL GRAMS |
|---|---|---|---|---|---|
| 870 | 317.5 | 367 | 415.5 | 521.5 | 1,621.50 |
| 871 | 2,037 | 2,202.50 | 2,219.50 | 1,887 | 8,346 |
| 873 | 0 | 5 | 10 | 20 | 35 |
| 874 | 189 | 216.5 | 211 | 222 | 838.5 |
| 875 | 384.5 | 526 | 252 | 244 | 1,406.50 |
| 877 | 73.5 | 144.5 | 119 | 120 | 457 |

| | | | | | |
|---|---|---|---|---|---|
| 878 | 60 | 20 | 43 | 44 | 167 |
| 879 | 37 | 32 | 19 | 12 | 100 |
| 880 | 524 | 594 | 609 | 603.5 | 2,330.50 |
| 881 | 120 | 122 | 63 | 46 | 351 |
| 882 | 457.5 | 485.5 | 379 | 484 | 1,806 |
| 883 | 55 | 90 | 66 | 57 | 268 |
| TOTAL | 4,255 | 4,805 | 4,406 | 4,261 | 17,727 |

STATE: NEW YORK

| ZIP CODE | QUARTER 1 | QUARTER 2 | QUARTER 3 | QUARTER 4 | TOTAL GRAMS |
|---|---|---|---|---|---|
| 100 | 5,081 | 5,015 | 5,017 | 5,236 | 20,349 |

DRUG CODE:9780DRUG NAME:TAPENTADOL
STATE:NEW YORK

| ZIP CODE | QUARTER 1 | QUARTER 2 | QUARTER 3 | QUARTER 4 | TOTAL GRAMS |
|---|---|---|---|---|---|
| 101 | 110.5 | 205.5 | 77 | 156.5 | 549.5 |
| 102 | 39 | 34 | 33 | 22 | 128 |
| 103 | 2,891.50 | 3,053 | 2,816 | 2,676 | 11,436.50 |
| 104 | 2,323 | 2,406.50 | 1,980 | 1,909.50 | 8,619 |
| 105 | 2,036.50 | 1,774 | 1,640.50 | 1,821 | 7,272 |
| 106 | 315.5 | 261 | 98 | 232 | 906.5 |
| 107 | 829.5 | 967.5 | 820 | 745 | 3,362 |
| 108 | 369 | 337 | 232.5 | 326 | 1,264.50 |
| 109 | 2,298 | 2,510 | 2,240.50 | 2,058.50 | 9,107 |
| 110 | 2,298.50 | 2,145 | 2,542.50 | 2,295 | 9,281 |
| 111 | 1,242 | 1,004.50 | 1,183 | 1,159 | 4,588.50 |
| 112 | 4,310.50 | 4,171.50 | 4,005.50 | 4,780 | 17,267.50 |
| 113 | 3,464 | 3,586 | 3,484 | 3,288 | 13,822 |
| 114 | 764 | 973 | 909 | 827 | 3,473 |
| 115 | 4,493.50 | 4,732.20 | 4,943.50 | 4,781.80 | 18,951 |
| 116 | 226 | 174 | 232.5 | 191 | 823.5 |
| 117 | 12,063 | 12,486 | 13,157 | 14,642.50 | 52,348.50 |
| 118 | 832 | 666 | 686.5 | 854.5 | 3,039 |
| 119 | 1,180.50 | 1,248.50 | 1,440.50 | 1,406.50 | 5,276 |
| 120 | 814 | 911 | 1,201 | 1,059.50 | 3,985.50 |
| 121 | 317 | 469 | 451 | 302.5 | 1,539.50 |
| 122 | 1,175 | 1,126.50 | 1,193.50 | 1,158.50 | 4,653.50 |
| 123 | 416.5 | 314.5 | 344.5 | 313.5 | 1,389 |
| 124 | 643.5 | 670 | 627 | 576 | 2,516.50 |
| 125 | 2,061.50 | 2,227 | 2,025.50 | 2,450.50 | 8,764.50 |
| 126 | 471 | 324.5 | 410 | 318 | 1,523.50 |
| 127 | 478.5 | 518 | 501 | 541.5 | 2,039 |
| 128 | 1,033 | 1,220.50 | 1,090.50 | 1,260.50 | 4,604.50 |
| 129 | 1,059 | 924 | 1,010 | 958.5 | 3,951.50 |
| 130 | 6,577.50 | 5,821 | 5,907 | 6,000.50 | 24,306 |
| 131 | 1,487 | 1,464 | 1,413 | 1,347 | 5,711 |
| 132 | 3,041.50 | 2,977.50 | 2,584.50 | 2,585.50 | 11,189 |
| 133 | 1,954.50 | 2,069.50 | 2,236.50 | 2,084 | 8,344.50 |
| 134 | 2,606 | 2,520 | 2,348 | 2,355 | 9,829 |
| 135 | 1,834.50 | 1,812.50 | 1,504 | 1,464.50 | 6,615.50 |
| 136 | 4,291.50 | 3,999.50 | 4,021.50 | 4,000.50 | 16,313 |
| 137 | 654.5 | 703.5 | 654.5 | 729.5 | 2,742 |
| 138 | 650.5 | 746.5 | 645.5 | 861.5 | 2,904 |
| 139 | 728.5 | 714 | 734.5 | 942 | 3,119 |
| 140 | 6,829 | 6,480.50 | 6,354.50 | 6,123.50 | 25,787.50 |
| 141 | 3,453.50 | 2,804 | 3,095 | 3,059.50 | 12,412 |
| 142 | 8,726 | 7,893 | 7,282.50 | 7,165 | 31,066.50 |
| 143 | 1,437.50 | 1,233.50 | 1,347 | 1,339.50 | 5,357.50 |
| 144 | 1,858 | 1,822 | 1,695 | 1,712.50 | 7,087.50 |

| | | | | | |
|---|---|---|---|---|---|
| 145 | 2,003.50 | 1,838.50 | 1,731 | 1,757.50 | 7,330.50 |
| 146 | 3,273.50 | 3,279 | 3,062 | 2,580.50 | 12,195 |
| 147 | 1,140 | 748 | 778.5 | 567.5 | 3,234 |
| 148 | 1,852 | 1,916.50 | 2,032.50 | 1,975 | 7,776 |
| 149 | 280 | 324 | 289 | 231 | 1,124 |
| TOTAL | 110,315.50 | 107,622.20 | 106,108.50 | 107,228.30 | 431,274.50 |

STATE: NORTH CAROLINA

| ZIP CODE | QUARTER 1 | QUARTER 2 | QUARTER 3 | QUARTER 4 | TOTAL GRAMS |
|---|---|---|---|---|---|
| 270 | 4,017.50 | 4,324.50 | 4,522.50 | 4,716.50 | 17,581 |
| 271 | 3,820.50 | 4,231.50 | 4,614.50 | 5,019 | 17,685.50 |
| 272 | 7,556.50 | 7,166.50 | 7,279.50 | 7,587.50 | 29,590 |
| 273 | 4,828.50 | 4,872.50 | 4,296.50 | 4,338.50 | 18,336 |
| 274 | 3,314.50 | 3,624.50 | 3,477 | 3,087 | 13,503 |
| 275 | 11,663.50 | 11,891.50 | 11,310.50 | 11,456 | 46,321.50 |
| 276 | 3,484.50 | 3,569 | 3,174.50 | 3,385.50 | 13,613.50 |
| 277 | 1,680.35 | 1,844.50 | 1,910.20 | 1,747.20 | 7,182.25 |
| 278 | 4,152.50 | 3,666.50 | 3,745 | 3,832.50 | 15,396.50 |
| 279 | 1,611 | 1,794.50 | 1,646.50 | 1,978.50 | 7,030.50 |
| 280 | 7,681.50 | 7,629 | 7,916.50 | 8,653.50 | 31,880.50 |
| 281 | 6,543 | 6,034 | 6,567.50 | 6,325 | 25,469.50 |
| 282 | 4,953.50 | 5,073.50 | 5,047 | 5,272.50 | 20,346.50 |
| 283 | 7,794 | 8,389.50 | 7,929 | 8,951.50 | 33,064 |

DRUG CODE:9780DRUG NAME:TAPENTADOL
STATE:NORTH CAROLINA

| ZIP CODE | QUARTER 1 | QUARTER 2 | QUARTER 3 | QUARTER 4 | TOTAL GRAMS |
|---|---|---|---|---|---|
| 284 | 7,835 | 7,252 | 7,381.50 | 6,993 | 29,461.50 |
| 285 | 9,369.50 | 9,077 | 8,484.50 | 8,734.50 | 35,665.50 |
| 286 | 8,085 | 8,694 | 8,715.50 | 9,513 | 35,007.50 |
| 287 | 2,698 | 2,704.50 | 2,643 | 2,561.50 | 10,607 |
| 288 | 1,093 | 933.5 | 867 | 1,057.50 | 3,951 |
| 289 | 393.5 | 249 | 254 | 258.5 | 1,155 |
| TOTAL | 102,575.35 | 103,021.50 | 101,782.20 | 105,468.70 | 412,847.75 |

STATE: NORTH DAKOTA

| ZIP CODE | QUARTER 1 | QUARTER 2 | QUARTER 3 | QUARTER 4 | TOTAL GRAMS |
|---|---|---|---|---|---|
| 580 | 138 | 236 | 252.5 | 237 | 863.5 |
| 581 | 224.5 | 229 | 125 | 147 | 725.5 |
| 582 | 456 | 346 | 421.5 | 346 | 1,569.50 |
| 583 | 55 | 23 | 40 | 40 | 158 |
| 584 | 109.5 | 77 | 95.5 | 109 | 391 |
| 585 | 1,181 | 1,144.50 | 1,023 | 1,073 | 4,421.50 |
| 586 | 107 | 146.5 | 171 | 142 | 566.5 |
| 587 | 282.5 | 305 | 533.5 | 403.5 | 1,524.50 |
| 588 | 48.5 | 12 | 67.5 | 77.5 | 205.5 |
| TOTAL | 2,602 | 2,519 | 2,729.50 | 2,575 | 10,425.50 |

STATE: OHIO

| ZIP CODE | QUARTER 1 | QUARTER 2 | QUARTER 3 | QUARTER 4 | TOTAL GRAMS |
|---|---|---|---|---|---|
| 430 | 4,720 | 4,427 | 4,352.50 | 4,615.50 | 18,115 |
| 431 | 2,441 | 2,508.50 | 2,456.50 | 2,923.50 | 10,329.50 |
| 432 | 6,591.50 | 6,788.50 | 6,854.50 | 6,446 | 26,680.50 |
| 433 | 709.5 | 610 | 575.5 | 597 | 2,492 |
| 434 | 1,030 | 1,058.50 | 1,021 | 1,057.50 | 4,167 |
| 435 | 3,347 | 3,478.50 | 3,357 | 3,397.50 | 13,580 |
| 436 | 5,696 | 5,093.50 | 5,947 | 5,683.50 | 22,420 |
| 437 | 721 | 712.5 | 661.5 | 585.5 | 2,680.50 |

| | | | | | |
|---|---|---|---|---|---|
| 438 | 326 | 336.5 | 345.5 | 336.5 | 1,344.50 |
| 439 | 919.5 | 1,091 | 1,199 | 1,255.50 | 4,465 |
| 440 | 7,216.50 | 7,165.50 | 7,188.50 | 7,054.50 | 28,625 |
| 441 | 5,006 | 5,233 | 4,644.50 | 4,850.50 | 19,734 |
| 442 | 3,212.50 | 3,390.50 | 3,350.50 | 2,994 | 12,947.50 |
| 443 | 940.5 | 916 | 966.5 | 750 | 3,573 |
| 444 | 1,878 | 1,614.50 | 1,635 | 1,456.50 | 6,584 |
| 445 | 1,211.50 | 1,152.50 | 1,047 | 1,123.50 | 4,534.50 |
| 446 | 3,029.50 | 3,276 | 3,377 | 3,215 | 12,897.50 |
| 447 | 1,371.50 | 1,446 | 1,241.50 | 1,484.50 | 5,543.50 |
| 448 | 1,210 | 1,115 | 1,268 | 1,063 | 4,656 |
| 449 | 366.5 | 266 | 238 | 240 | 1,110.50 |
| 450 | 4,228 | 4,180 | 4,526 | 4,402 | 17,336 |
| 451 | 1,843.50 | 1,793 | 1,780 | 1,640 | 7,056.50 |
| 452 | 3,810.85 | 3,821.50 | 3,879 | 4,234.50 | 15,745.85 |
| 453 | 2,442.50 | 2,166 | 2,269.50 | 2,171.50 | 9,049.50 |
| 454 | 2,153.50 | 2,288 | 2,306 | 2,488.50 | 9,236 |
| 455 | 469.5 | 402.5 | 428 | 505.5 | 1,805.50 |
| 456 | 2,079.50 | 1,958 | 2,013.50 | 2,030.50 | 8,081.50 |
| 457 | 733.5 | 569 | 540 | 500 | 2,342.50 |
| 458 | 1,462.50 | 1,206.50 | 1,326.50 | 1,119 | 5,114.50 |
| TOTAL | 71,167.35 | 70,064 | 70,795 | 70,221 | 282,247.35 |

STATE: OKLAHOMA

| ZIP CODE | QUARTER 1 | QUARTER 2 | QUARTER 3 | QUARTER 4 | TOTAL GRAMS |
|---|---|---|---|---|---|
| 730 | 5,235.50 | 5,596.50 | 5,505 | 6,070 | 22,407 |
| 731 | 3,917.85 | 4,146.50 | 4,172.50 | 4,163 | 16,399.85 |
| 734 | 1,584.50 | 1,619 | 1,427 | 1,535.50 | 6,166 |
| 735 | 621 | 625.5 | 752.5 | 710 | 2,709 |
| 736 | 69 | 117.5 | 59 | 27 | 272.5 |
| 737 | 374 | 425 | 327.5 | 356 | 1,482.50 |
| 738 | 168.5 | 169.5 | 161.5 | 175.5 | 675 |
| 739 | 133 | 35 | 55 | 15 | 238 |
| 740 | 4,670.50 | 4,647.50 | 5,068.50 | 6,012.50 | 20,399 |
| 741 | 3,890.50 | 3,745 | 3,903.50 | 3,995.50 | 15,534.50 |
| 743 | 683.5 | 740.5 | 693.5 | 713.5 | 2,831 |
| 744 | 1,378 | 1,396 | 1,366 | 1,344.50 | 5,484.50 |
| 745 | 539.5 | 419 | 343 | 295.5 | 1,597 |

DRUG CODE:9780DRUG NAME:TAPENTADOL
STATE:OKLAHOMA

| ZIP CODE | QUARTER 1 | QUARTER 2 | QUARTER 3 | QUARTER 4 | TOTAL GRAMS |
|---|---|---|---|---|---|
| 746 | 253.5 | 182 | 267.5 | 282 | 985 |
| 747 | 72 | 126 | 49 | 50 | 297 |
| 748 | 1,230 | 1,191.50 | 1,050.50 | 1,414.50 | 4,886.50 |
| 749 | 359.5 | 273.5 | 301 | 382.5 | 1,316.50 |
| TOTAL | 25,180.35 | 25,455.50 | 25,502.50 | 27,542.50 | 103,680.85 |

STATE: OREGON

| ZIP CODE | QUARTER 1 | QUARTER 2 | QUARTER 3 | QUARTER 4 | TOTAL GRAMS |
|---|---|---|---|---|---|
| 970 | 1,328.50 | 1,433 | 1,648.50 | 1,590.50 | 6,000.50 |
| 971 | 421 | 550 | 461.5 | 396 | 1,828.50 |
| 972 | 888.5 | 852 | 1,001 | 1,031.50 | 3,773 |
| 973 | 1,288 | 1,192.50 | 1,417.50 | 1,775 | 5,673 |
| 974 | 1,399.50 | 1,026 | 1,271 | 1,157.50 | 4,854 |
| 975 | 1,013.50 | 1,557.50 | 1,434.50 | 1,461.50 | 5,467 |
| 976 | 207 | 202 | 182 | 207 | 798 |
| 977 | 685.5 | 681 | 636 | 473 | 2,475.50 |
| 978 | 305 | 260 | 244 | 300 | 1,109 |
| 979 | 27 | 46 | 12 | 50 | 135 |

| | | | | | |
|---|---|---|---|---|---|
| TOTAL | 7,563.50 | 7,800 | 8,308 | 8,442 | 32,113.50 |
| STATE: | PENNSYLVANIA | | | | |

| ZIP CODE | QUARTER 1 | QUARTER 2 | QUARTER 3 | QUARTER 4 | TOTAL GRAMS |
|---|---|---|---|---|---|
| 150 | 1,533.50 | 1,578 | 1,740.50 | 1,990 | 6,842 |
| 151 | 766 | 830.5 | 819 | 823.5 | 3,239 |
| 152 | 872.85 | 877.5 | 961 | 890.5 | 3,601.85 |
| 153 | 558 | 479 | 697.5 | 674.5 | 2,409 |
| 154 | 161 | 187.5 | 153 | 250 | 751.5 |
| 155 | 772.5 | 639.5 | 578 | 612.5 | 2,602.50 |
| 156 | 745.5 | 1,014 | 1,002 | 1,068.50 | 3,830 |
| 157 | 321.5 | 299 | 304 | 318.5 | 1,243 |
| 158 | 333 | 324 | 317 | 480.5 | 1,454.50 |
| 159 | 1,162 | 1,229 | 1,320 | 1,167.50 | 4,878.50 |
| 160 | 197 | 191.5 | 156 | 250.5 | 795 |
| 161 | 797.5 | 864 | 963.5 | 960.5 | 3,585.50 |
| 162 | 424 | 425.5 | 462 | 422 | 1,733.50 |
| 163 | 727 | 790 | 871.5 | 683 | 3,071.50 |
| 164 | 207.5 | 199 | 273 | 284 | 963.5 |
| 165 | 573 | 672 | 661 | 800 | 2,706 |
| 166 | 1,149.50 | 1,010 | 1,146.50 | 1,087.50 | 4,393.50 |
| 167 | 147.5 | 186.5 | 79 | 144.5 | 557.5 |
| 168 | 867.5 | 885 | 954 | 1,015 | 3,721.50 |
| 169 | 157.5 | 175 | 121.5 | 112 | 566 |
| 170 | 3,085 | 3,381.50 | 3,174.50 | 3,539.50 | 13,180.50 |
| 171 | 1,210 | 1,336 | 1,200 | 992 | 4,738 |
| 172 | 1,189 | 1,087 | 917.5 | 1,104.50 | 4,298 |
| 173 | 3,170 | 3,404.50 | 3,373 | 3,092 | 13,039.50 |
| 174 | 1,478 | 1,389 | 1,253 | 1,261 | 5,381 |
| 175 | 239.5 | 325.5 | 337.5 | 462 | 1,364.50 |
| 176 | 569.5 | 609 | 720 | 670.5 | 2,569 |
| 177 | 469 | 331 | 359.5 | 238 | 1,397.50 |
| 178 | 507 | 556.5 | 458.5 | 430 | 1,952 |
| 179 | 405 | 444 | 548.5 | 413.5 | 1,811 |
| 180 | 7,447 | 7,678 | 7,103.50 | 7,114.50 | 29,343 |
| 181 | 3,388.50 | 3,713 | 3,696.50 | 3,488.50 | 14,286.50 |
| 182 | 975.5 | 917.5 | 935 | 1,806.50 | 4,634.50 |
| 183 | 3,334.50 | 3,240 | 3,391 | 3,345 | 13,310.50 |
| 184 | 1,519.50 | 1,647 | 1,530.50 | 1,734.50 | 6,431.50 |
| 185 | 617 | 756 | 718 | 875.5 | 2,966.50 |
| 186 | 432.5 | 404 | 347 | 487.5 | 1,671 |
| 187 | 6,389 | 6,344.50 | 6,258 | 6,461 | 25,452.50 |
| 188 | 291.5 | 267.5 | 296 | 263.5 | 1,118.50 |
| 189 | 2,166 | 2,407.50 | 2,427 | 2,444.50 | 9,445 |
| 190 | 6,401.50 | 7,023 | 7,641.50 | 8,117 | 29,183 |
| 191 | 2,860.35 | 3,320 | 3,101.50 | 2,565 | 11,846.85 |
| 193 | 1,648 | 1,891.50 | 2,099.50 | 2,147 | 7,786 |
| 194 | 2,681 | 2,849 | 2,973 | 3,240.50 | 11,743.50 |
| 195 | 1,655.50 | 1,490 | 1,817.50 | 1,575.50 | 6,538.50 |
| 196 | 1,252.50 | 1,138.50 | 955.5 | 1,236 | 4,582.50 |

DRUG CODE:9780DRUG NAME:TAPENTADOL
STATE:PENNSYLVANIA

| ZIP CODE | QUARTER 1 | QUARTER 2 | QUARTER 3 | QUARTER 4 | TOTAL GRAMS |
|---|---|---|---|---|---|
| TOTAL | 67,855.70 | 70,807.50 | 71,213 | 73,140 | 283,016.20 |
| STATE: | PUERTO RICO | | | | |

| ZIP CODE | QUARTER 1 | QUARTER 2 | QUARTER 3 | QUARTER 4 | TOTAL GRAMS |
|---|---|---|---|---|---|
| 6 | 294.5 | 301.5 | 241.5 | 183 | 1,020.50 |
| 7 | 250 | 119.5 | 155 | 145 | 669.5 |

| | | | | | |
|---|---|---|---|---|---|
| 9 | 738.47 | 611 | 558.5 | 655.5 | 2,563.47 |
| TOTAL | 1,282.97 | 1,032 | 955 | 983.5 | 4,253.47 |
| STATE: | RHODE ISLAND | | | | |
| ZIP CODE | QUARTER 1 | QUARTER 2 | QUARTER 3 | QUARTER 4 | TOTAL GRAMS |
| 28 | 1,770 | 2,007.50 | 1,937.50 | 1,925.50 | 7,640.50 |
| 29 | 301.5 | 371 | 456 | 422.5 | 1,551 |
| TOTAL | 2,071.50 | 2,378.50 | 2,393.50 | 2,348 | 9,191.50 |
| STATE: | SOUTH CAROLINA | | | | |
| ZIP CODE | QUARTER 1 | QUARTER 2 | QUARTER 3 | QUARTER 4 | TOTAL GRAMS |
| 290 | 3,199 | 2,998 | 3,330 | 3,812.50 | 13,339.50 |
| 291 | 2,403 | 2,958 | 2,863 | 2,920.50 | 11,144.50 |
| 292 | 3,259 | 3,158 | 3,281 | 3,266 | 12,964 |
| 293 | 4,200.50 | 4,166.50 | 4,364 | 4,013 | 16,744 |
| 294 | 5,582.50 | 5,740 | 5,655.20 | 5,446.50 | 22,424.20 |
| 295 | 6,918.50 | 6,167.50 | 6,808 | 6,709.50 | 26,603.50 |
| 296 | 10,903.50 | 11,746.50 | 11,814.50 | 12,667.50 | 47,132 |
| 297 | 2,680 | 3,141 | 2,731.50 | 2,985 | 11,537.50 |
| 298 | 1,973 | 1,988 | 2,102 | 2,301.50 | 8,364.50 |
| 299 | 2,544 | 2,542 | 2,379.50 | 2,904.50 | 10,370 |
| TOTAL | 43,663 | 44,605.50 | 45,328.70 | 47,026.50 | 180,623.70 |
| STATE: | SOUTH DAKOTA | | | | |
| ZIP CODE | QUARTER 1 | QUARTER 2 | QUARTER 3 | QUARTER 4 | TOTAL GRAMS |
| 570 | 810.5 | 776 | 645 | 686 | 2,917.50 |
| 571 | 2,564.50 | 2,303.50 | 2,278.50 | 2,153 | 9,299.50 |
| 572 | 258 | 226 | 281 | 338 | 1,103 |
| 573 | 607.5 | 471.5 | 348 | 340 | 1,767 |
| 574 | 467 | 659.5 | 728.5 | 645 | 2,500 |
| 575 | 90 | 80 | 137.5 | 135 | 442.5 |
| 576 | 100 | 127 | 134 | 96 | 457 |
| 577 | 1,167 | 1,222.50 | 1,346 | 1,278.50 | 5,014 |
| TOTAL | 6,064.50 | 5,866 | 5,898.50 | 5,671.50 | 23,500.50 |
| STATE: | TENNESSEE | | | | |
| ZIP CODE | QUARTER 1 | QUARTER 2 | QUARTER 3 | QUARTER 4 | TOTAL GRAMS |
| 370 | 6,698 | 7,200.50 | 7,730.50 | 7,679 | 29,308 |
| 371 | 5,795 | 6,338 | 6,804 | 6,913 | 25,850 |
| 372 | 1,142.50 | 1,443 | 1,772 | 1,688.50 | 6,046 |
| 373 | 3,788 | 4,021.50 | 4,318 | 4,183.50 | 16,311 |
| 374 | 2,102 | 2,536.50 | 2,489.50 | 2,501.50 | 9,629.50 |
| 376 | 2,043 | 2,698 | 2,284.50 | 2,292 | 9,317.50 |
| 377 | 1,302.50 | 1,092 | 999.5 | 786.5 | 4,180.50 |
| 378 | 2,377 | 2,103.50 | 2,090 | 1,731 | 8,301.50 |
| 379 | 1,589 | 1,323 | 1,267.50 | 1,226.50 | 5,406 |
| 380 | 1,371 | 1,516.50 | 1,449 | 1,635.50 | 5,972 |
| 381 | 1,020.50 | 1,163 | 1,090.50 | 1,325.50 | 4,599.50 |
| 382 | 333 | 289 | 425.5 | 443.5 | 1,491 |
| 383 | 1,299 | 1,196 | 1,559.50 | 1,767.50 | 5,822 |
| 384 | 1,255.50 | 1,091.50 | 1,485 | 1,465 | 5,297 |
| 385 | 1,348.50 | 1,391 | 1,324 | 1,421.50 | 5,485 |
| TOTAL | 33,464.50 | 35,403 | 37,089 | 37,060 | 143,016.50 |
| STATE: | TEXAS | | | | |
| ZIP CODE | QUARTER 1 | QUARTER 2 | QUARTER 3 | QUARTER 4 | TOTAL GRAMS |
| 750 | 13,607 | 14,265 | 15,173 | 15,912 | 58,957 |
| 751 | 3,980.50 | 3,944 | 4,257 | 4,884.50 | 17,066 |
| 752 | 5,358.35 | 5,564.43 | 5,258 | 5,177.50 | 21,358.28 |
| 754 | 1,254 | 1,289 | 1,572 | 1,544 | 5,659 |
| 755 | 131 | 241 | 242 | 213.5 | 827.5 |

| | | | | | |
|---|---|---|---|---|---|
| 756 | 334 | 233 | 297.5 | 627 | 1,491.50 |
| 757 | 797.5 | 1,221 | 1,207.50 | 1,410 | 4,636 |
| 758 | 857 | 988.5 | 945.5 | 691.5 | 3,482.50 |
| 759 | 549.5 | 478.5 | 488 | 374 | 1,890 |
| 760 | 7,467 | 8,246.50 | 8,416.50 | 8,413.50 | 32,543.50 |

DRUG CODE:9780DRUG NAME:TAPENTADOL
STATE:TEXAS

| ZIP CODE | QUARTER 1 | QUARTER 2 | QUARTER 3 | QUARTER 4 | TOTAL GRAMS |
|---|---|---|---|---|---|
| 761 | 4,575.50 | 4,458 | 5,097 | 5,231 | 19,361.50 |
| 762 | 1,902.50 | 2,071.50 | 2,304 | 2,213.50 | 8,491.50 |
| 763 | 1,487.50 | 1,529 | 1,350 | 1,417.50 | 5,784 |
| 764 | 249 | 331.5 | 315.5 | 325.5 | 1,221.50 |
| 765 | 992 | 983.5 | 999.5 | 1,129 | 4,104 |
| 766 | 254 | 155 | 230 | 186 | 825 |
| 767 | 469 | 312.5 | 354 | 410 | 1,545.50 |
| 768 | 125.5 | 106 | 110 | 60 | 401.5 |
| 769 | 77 | 275 | 630 | 606 | 1,588 |
| 770 | 5,637.50 | 6,830 | 6,489 | 6,879 | 25,835.50 |
| 773 | 4,588 | 4,947.50 | 5,017 | 5,472.50 | 20,025 |
| 774 | 4,574.50 | 4,997 | 4,699.50 | 5,111 | 19,382 |
| 775 | 6,308.50 | 6,358 | 5,681.50 | 6,227 | 24,575 |
| 776 | 1,069.50 | 1,202 | 1,270 | 1,139 | 4,680.50 |
| 777 | 701 | 660.5 | 755 | 810.5 | 2,927 |
| 778 | 1,076.50 | 886 | 1,293.50 | 1,198 | 4,454 |
| 779 | 229 | 180.5 | 232.5 | 295.5 | 937.5 |
| 780 | 1,340.50 | 1,158 | 1,608.50 | 1,811.50 | 5,918.50 |
| 781 | 1,894.50 | 2,086 | 2,229.50 | 2,390.50 | 8,600.50 |
| 782 | 10,154 | 11,542 | 10,286 | 11,556.50 | 43,538.50 |
| 783 | 826.5 | 742 | 823 | 908 | 3,299.50 |
| 784 | 1,661.50 | 2,042.50 | 2,172.50 | 2,125.50 | 8,002 |
| 785 | 791 | 830 | 705 | 706.5 | 3,032.50 |
| 786 | 4,694.50 | 5,255.50 | 5,440 | 5,137 | 20,527 |
| 787 | 5,028 | 5,388 | 5,431.50 | 6,260.50 | 22,108 |
| 788 | 235 | 210 | 269.5 | 209 | 923.5 |
| 789 | 146 | 212 | 301 | 255 | 914 |
| 790 | 365 | 438 | 452.5 | 711.5 | 1,967 |
| 791 | 767 | 682 | 613.5 | 817 | 2,879.50 |
| 792 | 0 | 5 | 5 | 6 | 16 |
| 793 | 162 | 152 | 213 | 195.5 | 722.5 |
| 794 | 1,091.50 | 1,134 | 1,029.50 | 1,080 | 4,335 |
| 795 | 45 | 119 | 85 | 41 | 290 |
| 796 | 57 | 64 | 59 | 72 | 252 |
| 797 | 370.5 | 475.5 | 420 | 518 | 1,784 |
| 798 | 0 | 6 | 9 | 17 | 32 |
| 799 | 1,958 | 2,039.50 | 2,312 | 2,719.50 | 9,029 |
| TOTAL | 100,239.85 | 107,335.43 | 109,150 | 115,496 | 432,221.28 |

STATE: UTAH

| ZIP CODE | QUARTER 1 | QUARTER 2 | QUARTER 3 | QUARTER 4 | TOTAL GRAMS |
|---|---|---|---|---|---|
| 840 | 7,204 | 6,767.50 | 7,289 | 7,389.50 | 28,650 |
| 841 | 2,341.50 | 2,729.50 | 2,514 | 2,394 | 9,979 |
| 843 | 534.5 | 607 | 599 | 572 | 2,312.50 |
| 844 | 819 | 1,002 | 995 | 1,186 | 4,002 |
| 845 | 694.5 | 679.5 | 626.5 | 929.5 | 2,930 |
| 846 | 1,451 | 1,610.50 | 1,375.50 | 1,439.50 | 5,876.50 |
| 847 | 2,693 | 3,021 | 2,820.50 | 2,608 | 11,142.50 |
| TOTAL | 15,737.50 | 16,417 | 16,219.50 | 16,518.50 | 64,892.50 |

STATE: VERMONT

RETAIL DRUG DISTRIBUTION BY ZIP CODE WITHIN STATE BY GRAMS WT

| ZIP CODE | QUARTER 1 | QUARTER 2 | QUARTER 3 | QUARTER 4 | TOTAL GRAMS |
|---|---|---|---|---|---|
| 50 | 116 | 40 | 40 | 80 | 276 |
| 51 | 5 | 5 | 5 | 0 | 15 |
| 52 | 132 | 99 | 107 | 164 | 502 |
| 53 | 0 | 0 | 0 | 3 | 3 |
| 54 | 569 | 493.5 | 493.5 | 682 | 2,238 |
| 56 | 185 | 156 | 190 | 135 | 666 |
| 57 | 384.5 | 392 | 305 | 322 | 1,403.50 |
| 58 | 93.5 | 78 | 109.5 | 113.5 | 394.5 |
| TOTAL | 1,485 | 1,263.50 | 1,250 | 1,499.50 | 5,498 |

STATE: VIRGIN ISLANDS

| ZIP CODE | QUARTER 1 | QUARTER 2 | QUARTER 3 | QUARTER 4 | TOTAL GRAMS |
|---|---|---|---|---|---|
| 8 | 6 | 9 | 3 | 26 | 44 |
| TOTAL | 6 | 9 | 3 | 26 | 44 |

STATE: VIRGINIA

| ZIP CODE | QUARTER 1 | QUARTER 2 | QUARTER 3 | QUARTER 4 | TOTAL GRAMS |
|---|---|---|---|---|---|
| 201 | 6,281.50 | 6,358 | 6,385.50 | 6,114.50 | 25,139.50 |

DRUG CODE:9780DRUG NAME:TAPENTADOL
STATE:VIRGINIA

| ZIP CODE | QUARTER 1 | QUARTER 2 | QUARTER 3 | QUARTER 4 | TOTAL GRAMS |
|---|---|---|---|---|---|
| 220 | 2,659 | 2,862 | 2,690 | 2,984.50 | 11,195.50 |
| 221 | 1,994 | 2,173 | 2,061.50 | 1,861.50 | 8,090 |
| 222 | 1,197 | 1,126 | 1,127.50 | 1,381.50 | 4,832 |
| 223 | 1,704.50 | 2,018.25 | 1,619.50 | 1,480.75 | 6,823 |
| 224 | 2,129.50 | 2,333.50 | 2,077.50 | 1,978.50 | 8,519 |
| 225 | 1,052.50 | 844.5 | 950.5 | 1,060 | 3,907.50 |
| 226 | 1,504.50 | 1,334.50 | 1,395 | 1,663 | 5,897 |
| 227 | 167.5 | 141.5 | 190 | 270.5 | 769.5 |
| 228 | 530 | 655.5 | 638 | 603.5 | 2,427 |
| 229 | 532 | 525 | 455.5 | 591 | 2,103.50 |
| 230 | 754.5 | 661.5 | 651.5 | 650.5 | 2,718 |
| 231 | 2,406.50 | 1,969 | 2,200.50 | 1,861 | 8,437 |
| 232 | 2,967 | 2,757.50 | 2,662.50 | 2,220.50 | 10,607.50 |
| 233 | 2,633.50 | 2,598 | 2,513.50 | 2,426 | 10,171 |
| 234 | 5,743.50 | 5,890 | 5,552.50 | 6,058.50 | 23,244.50 |
| 235 | 1,299 | 1,344.50 | 1,241.50 | 1,426 | 5,311 |
| 236 | 2,617 | 2,480.50 | 2,974.50 | 2,635 | 10,707 |
| 237 | 737.5 | 933 | 962.5 | 715.5 | 3,348.50 |
| 238 | 2,072 | 1,996 | 1,790 | 1,897 | 7,755 |
| 239 | 273.5 | 304.5 | 298 | 473 | 1,349 |
| 240 | 1,785.50 | 1,913 | 1,903.75 | 1,986.50 | 7,588.75 |
| 241 | 1,483.50 | 1,716 | 1,830 | 1,804.50 | 6,834 |
| 242 | 679 | 698.5 | 728.5 | 910 | 3,016 |
| 243 | 1,118.50 | 1,013.50 | 1,053.50 | 1,177.50 | 4,363 |
| 244 | 379 | 457 | 468.5 | 563.5 | 1,868 |
| 245 | 1,990.50 | 1,783.50 | 2,056 | 2,038.50 | 7,868.50 |
| 246 | 211 | 427 | 285 | 373.5 | 1,296.50 |
| TOTAL | 48,903 | 49,314.75 | 48,762.75 | 49,206.25 | 196,186.75 |

STATE: WASHINGTON

| ZIP CODE | QUARTER 1 | QUARTER 2 | QUARTER 3 | QUARTER 4 | TOTAL GRAMS |
|---|---|---|---|---|---|
| 980 | 4,197.50 | 4,233 | 3,875.50 | 3,638 | 15,944 |
| 981 | 1,907 | 1,659 | 1,611 | 1,248.50 | 6,425.50 |
| 982 | 2,138.50 | 1,940 | 1,851 | 1,751 | 7,680.50 |
| 983 | 2,419 | 2,536 | 2,626.50 | 2,444 | 10,025.50 |
| 984 | 893 | 862 | 964 | 868.5 | 3,587.50 |

| | | | | | |
|---|---|---|---|---|---|
| 985 | 844 | 768 | 788 | 941.5 | 3,341.50 |
| 986 | 1,383.50 | 1,519.50 | 1,649.50 | 1,688 | 6,240.50 |
| 988 | 348 | 486 | 495 | 404 | 1,733 |
| 989 | 750 | 524.5 | 635 | 685 | 2,594.50 |
| 990 | 482.5 | 563 | 643 | 540 | 2,228.50 |
| 991 | 215 | 203 | 299 | 252 | 969 |
| 992 | 2,103.50 | 2,412 | 2,676.50 | 2,924 | 10,116 |
| 993 | 1,707 | 1,729.50 | 1,609 | 1,572 | 6,617.50 |
| 994 | 184 | 170.5 | 148.5 | 98.5 | 601.5 |
| TOTAL | 19,572.50 | 19,606 | 19,871.50 | 19,055 | 78,105 |

STATE: WEST VIRGINIA

| ZIP CODE | QUARTER 1 | QUARTER 2 | QUARTER 3 | QUARTER 4 | TOTAL GRAMS |
|---|---|---|---|---|---|
| 247 | 127 | 161 | 143.5 | 236.5 | 668 |
| 248 | 85 | 104 | 61 | 22 | 272 |
| 249 | 54 | 45 | 43 | 33 | 175 |
| 250 | 95 | 64 | 62 | 124 | 345 |
| 251 | 246 | 135 | 160 | 155 | 696 |
| 252 | 84 | 74 | 67 | 85.5 | 310.5 |
| 253 | 315 | 265 | 447.5 | 302 | 1,329.50 |
| 254 | 1,539 | 1,704.50 | 1,492 | 1,637.50 | 6,373 |
| 255 | 497.5 | 480 | 387 | 508 | 1,872.50 |
| 256 | 33 | 53 | 38 | 52 | 176 |
| 257 | 489 | 612 | 608.5 | 549 | 2,258.50 |
| 258 | 97 | 118 | 185 | 101.5 | 501.5 |
| 259 | 0 | 20.5 | 27 | 16 | 63.5 |
| 260 | 1,097.50 | 1,335 | 1,433 | 1,511.50 | 5,377 |
| 261 | 316.5 | 358.5 | 291.5 | 258 | 1,224.50 |
| 262 | 88.5 | 124.5 | 63 | 48 | 324 |
| 263 | 123 | 173 | 66 | 120 | 482 |
| 264 | 89 | 36 | 24 | 48 | 197 |
| 265 | 302.5 | 186.5 | 203.5 | 211 | 903.5 |

DRUG CODE:9780DRUG NAME:TAPENTADOL
STATE:WEST VIRGINIA

| ZIP CODE | QUARTER 1 | QUARTER 2 | QUARTER 3 | QUARTER 4 | TOTAL GRAMS |
|---|---|---|---|---|---|
| 266 | 52 | 64 | 60 | 35 | 211 |
| 267 | 47 | 59 | 99 | 116 | 321 |
| 268 | 90 | 80 | 50 | 55 | 275 |
| TOTAL | 5,867.50 | 6,252.50 | 6,011.50 | 6,224.50 | 24,356 |

STATE: WISCONSIN

| ZIP CODE | QUARTER 1 | QUARTER 2 | QUARTER 3 | QUARTER 4 | TOTAL GRAMS |
|---|---|---|---|---|---|
| 530 | 2,691.50 | 2,841 | 3,129.50 | 3,065.50 | 11,727.50 |
| 531 | 3,302.50 | 3,016 | 3,119.50 | 3,172 | 12,610 |
| 532 | 3,335.85 | 3,633.50 | 3,251 | 3,386 | 13,606.35 |
| 534 | 898.5 | 1,039.50 | 998 | 1,179 | 4,115 |
| 535 | 816 | 814.5 | 864 | 903 | 3,397.50 |
| 537 | 311 | 241 | 399 | 297.5 | 1,248.50 |
| 538 | 150 | 165.5 | 147.5 | 191 | 654 |
| 539 | 508 | 331.5 | 409 | 339 | 1,587.50 |
| 540 | 834.5 | 653 | 597 | 490.5 | 2,575 |
| 541 | 284 | 299.5 | 313.5 | 256 | 1,153 |
| 542 | 203 | 179.5 | 202.5 | 275.5 | 860.5 |
| 543 | 487.5 | 345 | 453 | 524.5 | 1,810 |
| 544 | 1,442 | 1,386.50 | 1,127 | 1,103.50 | 5,059 |
| 545 | 263.5 | 318 | 307 | 412 | 1,300.50 |
| 546 | 159 | 116 | 150 | 137 | 562 |
| 547 | 325.5 | 302 | 200 | 202.5 | 1,030 |

| | | | | | |
|---|---|---|---|---|---|
| 548 | 628 | 664.5 | 598 | 511 | 2,401.50 |
| 549 | 1,509.50 | 1,461.50 | 1,535.50 | 1,587 | 6,093.50 |
| TOTAL | 18,149.85 | 17,808 | 17,801 | 18,032.50 | 71,791.35 |

STATE: WYOMING

| ZIP CODE | QUARTER 1 | QUARTER 2 | QUARTER 3 | QUARTER 4 | TOTAL GRAMS |
|---|---|---|---|---|---|
| 820 | 1,329 | 941 | 960.5 | 1,111 | 4,341.50 |
| 822 | 72 | 75.5 | 45 | 59 | 251.5 |
| 823 | 25 | 35 | 10 | 30 | 100 |
| 824 | 147 | 171 | 161 | 62 | 541 |
| 825 | 31 | 22 | 29 | 21 | 103 |
| 826 | 348.5 | 208 | 259 | 288.5 | 1,104 |
| 827 | 67.5 | 56.5 | 102 | 77 | 303 |
| 828 | 45 | 84 | 59 | 48 | 236 |
| 829 | 69 | 59 | 57 | 92 | 277 |
| 830 | 13 | 10 | 0 | 5 | 28 |
| 831 | 5 | 70 | 48 | 70 | 193 |
| TOTAL | 2,152 | 1,732 | 1,730.50 | 1,863.50 | 7,478 |

DRUG CODE: 9801 DRUG NAME: FENTANYL BASE
STATE: ALABAMA

| ZIP CODE | QUARTER 1 | QUARTER 2 | QUARTER 3 | QUARTER 4 | TOTAL GRAMS |
|---|---|---|---|---|---|
| 350 | 211.73 | 214.39 | 207.36 | 195.06 | 828.54 |
| 351 | 128.4 | 120.78 | 117.95 | 116.36 | 483.49 |
| 352 | 190.68 | 174.4 | 166.46 | 164.04 | 695.58 |
| 354 | 101.47 | 102.95 | 99.62 | 91.16 | 395.2 |
| 355 | 80.59 | 75.4 | 74.85 | 73 | 303.84 |
| 356 | 165.49 | 156.14 | 150.31 | 135.48 | 607.42 |
| 357 | 48.45 | 45.86 | 46.59 | 43.05 | 183.95 |
| 358 | 93.08 | 93.83 | 84.92 | 86.52 | 358.35 |
| 359 | 179.64 | 168.79 | 157.92 | 149.42 | 655.77 |
| 360 | 107.4 | 106.65 | 100.71 | 100.32 | 415.08 |
| 361 | 101.51 | 100.69 | 104.65 | 101.77 | 408.62 |
| 362 | 74.68 | 72.93 | 65.07 | 147.29 | 359.97 |
| 363 | 96.12 | 99.32 | 99.19 | 93.76 | 388.39 |
| 364 | 39.79 | 43.75 | 41.61 | 38.15 | 163.3 |
| 365 | 142.7 | 126.15 | 130.92 | 117.72 | 517.49 |
| 366 | 99.12 | 93.26 | 89.94 | 85.61 | 367.93 |
| 367 | 35.98 | 37.29 | 36.87 | 31.96 | 142.1 |
| 368 | 63.05 | 60.5 | 61.66 | 60.68 | 245.89 |
| 369 | 9.88 | 10.32 | 9.26 | 8.18 | 37.64 |
| TOTAL | 1,969.76 | 1,903.40 | 1,845.86 | 1,839.53 | 7,558.55 |

STATE: ALASKA

| ZIP CODE | QUARTER 1 | QUARTER 2 | QUARTER 3 | QUARTER 4 | TOTAL GRAMS |
|---|---|---|---|---|---|
| 995 | 124.12 | 118.33 | 113.87 | 103.55 | 459.87 |

DRUG CODE:9801DRUG NAME:FENTANYL BASE
STATE:ALASKA

| ZIP CODE | QUARTER 1 | QUARTER 2 | QUARTER 3 | QUARTER 4 | TOTAL GRAMS |
|---|---|---|---|---|---|
| 996 | 97.2 | 95.42 | 89.85 | 87.43 | 369.9 |
| 997 | 28.17 | 32.08 | 31.55 | 27.81 | 119.61 |
| 998 | 19.44 | 17.96 | 15.55 | 14.18 | 67.13 |
| 999 | 9.34 | 11.74 | 10.99 | 11.38 | 43.45 |
| TOTAL | 278.27 | 275.53 | 261.81 | 244.35 | 1,059.96 |

STATE: AMERICAN SAMOA

| ZIP CODE | QUARTER 1 | QUARTER 2 | QUARTER 3 | QUARTER 4 | TOTAL GRAMS |
|---|---|---|---|---|---|
| 967 | 0.38 | 0 | 0.16 | 0.01 | 0.55 |

RETAIL DRUG DISTRIBUTION BY ZIP CODE WITHIN STATE BY GRAMS WT

| | | | | | |
|---|---|---|---|---|---|
| TOTAL | 0.38 | 0 | 0.16 | 0.01 | 0.55 |
| STATE: | ARIZONA | | | | |
| ZIP CODE | QUARTER 1 | QUARTER 2 | QUARTER 3 | QUARTER 4 | TOTAL GRAMS |
| 850 | 545.52 | 529.65 | 489.94 | 465.24 | 2,030.35 |
| 851 | 85.91 | 70.58 | 67.09 | 64.6 | 288.18 |
| 852 | 655.3 | 681.44 | 653.11 | 640.86 | 2,630.71 |
| 853 | 400.79 | 370.8 | 342.87 | 337.9 | 1,452.36 |
| 855 | 26.77 | 26.62 | 27.6 | 25.41 | 106.4 |
| 856 | 100.53 | 85.35 | 82.16 | 72.95 | 340.99 |
| 857 | 341.07 | 330.58 | 319.55 | 324.81 | 1,316.01 |
| 859 | 23.22 | 20.87 | 23.67 | 18.73 | 86.49 |
| 860 | 29.03 | 33 | 35.83 | 30.9 | 128.76 |
| 863 | 88.44 | 96.81 | 92.66 | 82.44 | 360.35 |
| 864 | 100.89 | 89.63 | 91.26 | 101.76 | 383.54 |
| 865 | 4.94 | 5.53 | 1.69 | 2.63 | 14.79 |
| TOTAL | 2,402.41 | 2,340.86 | 2,227.43 | 2,168.23 | 9,138.93 |
| STATE: | ARKANSAS | | | | |
| ZIP CODE | QUARTER 1 | QUARTER 2 | QUARTER 3 | QUARTER 4 | TOTAL GRAMS |
| 716 | 58.57 | 55.55 | 51.89 | 46.73 | 212.74 |
| 717 | 40.05 | 37.57 | 36 | 34.95 | 148.57 |
| 718 | 35.17 | 36.49 | 34.75 | 27.41 | 133.82 |
| 719 | 98.84 | 94.82 | 89.4 | 81.16 | 364.22 |
| 720 | 160.2 | 159.76 | 151.94 | 141.96 | 613.86 |
| 721 | 183.16 | 181.54 | 175.45 | 166.61 | 706.76 |
| 722 | 115.62 | 116.82 | 108.48 | 111.37 | 452.29 |
| 723 | 32.08 | 33.57 | 31.66 | 30.48 | 127.79 |
| 724 | 69.12 | 68.49 | 64.31 | 62.64 | 264.56 |
| 725 | 53.32 | 55.49 | 54.81 | 52.09 | 215.71 |
| 726 | 56.75 | 53.06 | 52.09 | 51.22 | 213.12 |
| 727 | 160.2 | 162.33 | 163.67 | 164.59 | 650.79 |
| 728 | 54.16 | 51.93 | 45.8 | 45.28 | 197.17 |
| 729 | 66.86 | 62.05 | 62.64 | 61.74 | 253.29 |
| TOTAL | 1,184.10 | 1,169.47 | 1,122.89 | 1,078.23 | 4,554.69 |
| STATE: | CALIFORNIA | | | | |
| ZIP CODE | QUARTER 1 | QUARTER 2 | QUARTER 3 | QUARTER 4 | TOTAL GRAMS |
| 900 | 273.33 | 274.64 | 278.54 | 260.5 | 1,087.01 |
| 902 | 107.65 | 113.13 | 99.96 | 93.94 | 414.68 |
| 903 | 7.08 | 7.07 | 8.23 | 5.19 | 27.57 |
| 904 | 47.87 | 48.9 | 48.38 | 50.25 | 195.4 |
| 905 | 46.22 | 44.64 | 37.2 | 42.58 | 170.64 |
| 906 | 125.87 | 125.58 | 97.08 | 93.48 | 442.01 |
| 907 | 90.58 | 97.92 | 81.64 | 100.9 | 371.04 |
| 908 | 145.75 | 143.83 | 145.73 | 170.51 | 605.82 |
| 910 | 65.41 | 65.57 | 60.97 | 58.52 | 250.47 |
| 911 | 31.85 | 28.21 | 29.38 | 28.98 | 118.42 |
| 912 | 29.45 | 32.12 | 32.62 | 35.24 | 129.43 |
| 913 | 261.53 | 261.04 | 248.84 | 240.03 | 1,011.44 |
| 914 | 116.18 | 131.07 | 102.28 | 99.51 | 449.04 |
| 915 | 18.33 | 17.31 | 15.5 | 16.01 | 67.15 |
| 916 | 17.41 | 16.17 | 14.79 | 14.93 | 63.3 |
| 917 | 408 | 372.52 | 385.46 | 373.4 | 1,539.38 |
| 918 | 8.25 | 8.03 | 6.66 | 5.92 | 28.86 |
| 919 | 73.14 | 68.73 | 62.97 | 58.41 | 263.25 |
| 920 | 429.49 | 426.04 | 420.7 | 410.62 | 1,686.85 |
| 921 | 493.58 | 347.05 | 411.14 | 437.47 | 1,689.24 |
| 922 | 239.05 | 226.2 | 214.47 | 308.4 | 988.12 |
| 923 | 354.98 | 356.36 | 326.04 | 319.92 | 1,357.30 |
| 924 | 84.16 | 88.12 | 77.73 | 75.99 | 326 |

RETAIL DRUG DISTRIBUTION BY ZIP CODE WITHIN STATE BY GRAMS WT

DRUG CODE:9801DRUG NAME:FENTANYL BASE
STATE:CALIFORNIA

| ZIP CODE | QUARTER 1 | QUARTER 2 | QUARTER 3 | QUARTER 4 | TOTAL GRAMS |
|---|---|---|---|---|---|
| 925 | 336.67 | 306.94 | 322.44 | 284.66 | 1,250.71 |
| 926 | 340.27 | 325.83 | 318.99 | 297.44 | 1,282.53 |
| 927 | 83.72 | 79.17 | 72.18 | 61.98 | 297.05 |
| 928 | 288.53 | 272.21 | 257.39 | 257.88 | 1,076.01 |
| 930 | 167.13 | 163.07 | 160.99 | 155.63 | 646.82 |
| 931 | 24.99 | 37.13 | 21.48 | 29.34 | 112.94 |
| 932 | 142.86 | 138.93 | 135.07 | 127.89 | 544.75 |
| 933 | 190.02 | 194.97 | 177.07 | 162.89 | 724.95 |
| 934 | 167.01 | 158.24 | 155.11 | 166.64 | 647 |
| 935 | 126.7 | 122.14 | 118.87 | 113.74 | 481.45 |
| 936 | 101.06 | 97.36 | 92.92 | 86.3 | 377.64 |
| 937 | 118.12 | 119.34 | 112.34 | 110.27 | 460.07 |
| 939 | 53.21 | 50.19 | 45.9 | 37.8 | 187.1 |
| 940 | 147.32 | 126.71 | 132.99 | 132.4 | 539.42 |
| 941 | 171.81 | 170.03 | 158.12 | 149.96 | 649.92 |
| 943 | 38.15 | 41.44 | 37.77 | 37.91 | 155.27 |
| 944 | 25.42 | 23.67 | 21.31 | 16.66 | 87.06 |
| 945 | 686.86 | 631.98 | 587.07 | 539.6 | 2,445.51 |
| 946 | 65.8 | 62.59 | 57.98 | 56.18 | 242.55 |
| 947 | 15.67 | 15.37 | 13.81 | 16.56 | 61.41 |
| 948 | 21.4 | 20.16 | 19.92 | 20.15 | 81.63 |
| 949 | 80.82 | 81.79 | 76.52 | 75.66 | 314.79 |
| 950 | 147.72 | 142.66 | 125.22 | 138.03 | 553.63 |
| 951 | 75.88 | 75.62 | 65.91 | 70.24 | 287.65 |
| 952 | 154.43 | 148.64 | 146.09 | 143.68 | 592.84 |
| 953 | 350.98 | 336.67 | 318.92 | 442.62 | 1,449.19 |
| 954 | 172.1 | 162.79 | 163.64 | 145.15 | 643.68 |
| 955 | 64.63 | 64.62 | 60.97 | 59.72 | 249.94 |
| 956 | 478.7 | 439.96 | 412.86 | 397.29 | 1,728.81 |
| 957 | 74.19 | 69.31 | 64.26 | 59.26 | 267.02 |
| 958 | 252.03 | 274.4 | 258.11 | 243.4 | 1,027.94 |
| 959 | 267.03 | 236.78 | 235.34 | 226.24 | 965.39 |
| 960 | 165.41 | 156.04 | 150.87 | 134.36 | 606.68 |
| 961 | 23.44 | 24.34 | 23.25 | 22.95 | 93.98 |
| TOTAL | 9,095.24 | 8,671.34 | 8,327.99 | 8,321.18 | 34,415.75 |

STATE: COLORADO

| ZIP CODE | QUARTER 1 | QUARTER 2 | QUARTER 3 | QUARTER 4 | TOTAL GRAMS |
|---|---|---|---|---|---|
| 800 | 267.89 | 262.39 | 243.63 | 232.09 | 1,006 |
| 801 | 204.45 | 205.09 | 180.31 | 172.99 | 762.84 |
| 802 | 250.68 | 242.89 | 248.24 | 235.57 | 977.38 |
| 803 | 41.94 | 42.21 | 38.5 | 38.48 | 161.13 |
| 804 | 63.27 | 66.97 | 64.12 | 63.99 | 258.35 |
| 805 | 212.76 | 197.03 | 186.39 | 164.01 | 760.19 |
| 806 | 69.81 | 62.06 | 60.99 | 60.33 | 253.19 |
| 807 | 37.16 | 34.57 | 30.76 | 30.67 | 133.16 |
| 808 | 34.32 | 32.93 | 31.66 | 30.02 | 128.93 |
| 809 | 196.66 | 179.96 | 172.73 | 165.1 | 714.45 |
| 810 | 108.51 | 97.47 | 95.56 | 89.27 | 390.81 |
| 811 | 23.24 | 21.8 | 21.14 | 17.9 | 84.08 |
| 812 | 31.97 | 30.3 | 27.86 | 27.19 | 117.32 |
| 813 | 19.06 | 15.85 | 18.61 | 17.84 | 71.36 |
| 814 | 19.71 | 23.53 | 21.24 | 16.61 | 81.09 |
| 815 | 65.79 | 69.69 | 64.73 | 61.58 | 261.79 |
| 816 | 25.7 | 24.1 | 21.71 | 20.72 | 92.23 |
| TOTAL | 1,672.92 | 1,608.84 | 1,528.18 | 1,444.36 | 6,254.30 |

RETAIL DRUG DISTRIBUTION BY ZIP CODE WITHIN STATE BY GRAMS WT

STATE: CONNECTICUT

| ZIP CODE | QUARTER 1 | QUARTER 2 | QUARTER 3 | QUARTER 4 | TOTAL GRAMS |
|---|---|---|---|---|---|
| 60 | 266.62 | 246.44 | 235.18 | 214.92 | 963.16 |
| 61 | 85.59 | 94.5 | 56.61 | 51.39 | 288.09 |
| 62 | 44.28 | 46.56 | 41.96 | 40.29 | 173.09 |
| 63 | 97.78 | 99.65 | 90.78 | 83.02 | 371.23 |
| 64 | 299.34 | 300.16 | 276.08 | 266.51 | 1,142.09 |
| 65 | 119.17 | 107.13 | 101.63 | 95.02 | 422.95 |
| 66 | 49.4 | 42.31 | 43.19 | 41.91 | 176.81 |
| 67 | 106.56 | 109.39 | 96.21 | 94.17 | 406.33 |
| 68 | 165.95 | 192.8 | 188.42 | 151.11 | 698.28 |

DRUG CODE:9801DRUG NAME:FENTANYL BASE
STATE:CONNECTICUT

| ZIP CODE | QUARTER 1 | QUARTER 2 | QUARTER 3 | QUARTER 4 | TOTAL GRAMS |
|---|---|---|---|---|---|
| 69 | 19.7 | 15.61 | 15.34 | 15.54 | 66.19 |
| TOTAL | 1,254.39 | 1,254.55 | 1,145.40 | 1,053.88 | 4,708.22 |

STATE: DELAWARE

| ZIP CODE | QUARTER 1 | QUARTER 2 | QUARTER 3 | QUARTER 4 | TOTAL GRAMS |
|---|---|---|---|---|---|
| 197 | 155.12 | 147.32 | 142.68 | 139.5 | 584.62 |
| 198 | 84.01 | 81.15 | 76.33 | 72.32 | 313.81 |
| 199 | 186.24 | 173.96 | 186.83 | 178.07 | 725.1 |
| TOTAL | 425.37 | 402.43 | 405.84 | 389.89 | 1,623.53 |

STATE: DISTRICT OF COLUMBIA

| ZIP CODE | QUARTER 1 | QUARTER 2 | QUARTER 3 | QUARTER 4 | TOTAL GRAMS |
|---|---|---|---|---|---|
| 200 | 95.28 | 99.55 | 89.03 | 86.21 | 370.07 |
| 204 | 6.59 | 7.89 | 6.73 | 6.36 | 27.57 |
| 205 | 0 | 0.05 | 0 | 0 | 0.05 |
| TOTAL | 101.87 | 107.49 | 95.76 | 92.57 | 397.69 |

STATE: FLORIDA

| ZIP CODE | QUARTER 1 | QUARTER 2 | QUARTER 3 | QUARTER 4 | TOTAL GRAMS |
|---|---|---|---|---|---|
| 320 | 250.97 | 254.38 | 247.53 | 251.87 | 1,004.75 |
| 321 | 272.55 | 248.77 | 241.11 | 248.51 | 1,010.94 |
| 322 | 543.48 | 516.68 | 520.73 | 481.95 | 2,062.84 |
| 323 | 97.99 | 96.59 | 88.61 | 90.19 | 373.38 |
| 324 | 136.18 | 133.54 | 133.68 | 138.87 | 542.27 |
| 325 | 188.62 | 186.09 | 171.3 | 173.54 | 719.55 |
| 326 | 131.27 | 138.88 | 132.7 | 132.37 | 535.22 |
| 327 | 408.88 | 414.93 | 397.64 | 384.87 | 1,606.32 |
| 328 | 208.67 | 200.44 | 191.86 | 184.53 | 785.5 |
| 329 | 355.95 | 389.14 | 354.86 | 354.72 | 1,454.67 |
| 330 | 332.26 | 328.55 | 309.52 | 294.05 | 1,264.38 |
| 331 | 259.26 | 249.46 | 244.92 | 233.35 | 986.99 |
| 333 | 259.43 | 264.51 | 250.09 | 251.48 | 1,025.51 |
| 334 | 548.13 | 552.25 | 502.96 | 563.94 | 2,167.28 |
| 335 | 234.45 | 224.17 | 229.29 | 226.15 | 914.06 |
| 336 | 470.06 | 436.44 | 385.23 | 412.18 | 1,703.91 |
| 337 | 380.07 | 346.83 | 336.77 | 341.22 | 1,404.89 |
| 338 | 190.22 | 189.89 | 178.6 | 179.72 | 738.43 |
| 339 | 336.04 | 319.34 | 296.22 | 303.18 | 1,254.78 |
| 341 | 107.34 | 94.22 | 92.02 | 102.48 | 396.06 |
| 342 | 396.73 | 389.09 | 367.24 | 378.4 | 1,531.46 |
| 344 | 180.59 | 176.32 | 165.79 | 177.21 | 699.91 |
| 346 | 358.54 | 343.76 | 331.12 | 326.73 | 1,360.15 |
| 347 | 172.86 | 162.08 | 156.65 | 163.29 | 654.88 |
| 349 | 193.57 | 179.88 | 165.75 | 179.28 | 718.48 |

| | | | | | |
|---|---|---|---|---|---|
| TOTAL | 7,014.11 | 6,836.23 | 6,492.19 | 6,574.08 | 26,916.61 |

STATE: GEORGIA

| ZIP CODE | QUARTER 1 | QUARTER 2 | QUARTER 3 | QUARTER 4 | TOTAL GRAMS |
|---|---|---|---|---|---|
| 300 | 627.19 | 634.54 | 610.78 | 593.79 | 2,466.30 |
| 301 | 403.76 | 395.75 | 375.13 | 379.06 | 1,553.70 |
| 302 | 274.27 | 262.26 | 268.22 | 263.25 | 1,068 |
| 303 | 191.65 | 179.39 | 192.28 | 192.93 | 756.25 |
| 304 | 111.21 | 97.92 | 98.3 | 94.95 | 402.38 |
| 305 | 231.5 | 238.06 | 226.66 | 229.8 | 926.02 |
| 306 | 125.36 | 122.89 | 116.17 | 116.81 | 481.23 |
| 307 | 89.1 | 84.5 | 86.39 | 78.5 | 338.49 |
| 308 | 66.27 | 62.75 | 60.52 | 66.94 | 256.48 |
| 309 | 122.86 | 125.4 | 125.42 | 111.57 | 485.25 |
| 310 | 238.34 | 226.67 | 224.58 | 217.8 | 907.39 |
| 312 | 84.05 | 83.38 | 80.12 | 79.74 | 327.29 |
| 313 | 82.5 | 86.4 | 91.09 | 83.37 | 343.36 |
| 314 | 82.76 | 84.35 | 79.88 | 72.69 | 319.68 |
| 315 | 144.03 | 147.51 | 147.08 | 164.21 | 602.83 |
| 316 | 69.09 | 68.34 | 71.68 | 72.21 | 281.32 |
| 317 | 137 | 136.89 | 138.72 | 136.97 | 549.58 |
| 318 | 7.34 | 7.63 | 8.41 | 8.65 | 32.03 |
| 319 | 68.39 | 67.28 | 68.28 | 61.12 | 265.07 |
| 398 | 34.34 | 34.72 | 33.7 | 35.82 | 138.58 |
| TOTAL | 3,191.01 | 3,146.63 | 3,103.41 | 3,060.18 | 12,501.23 |

STATE: GUAM

DRUG CODE:9801DRUG NAME:FENTANYL BASE
STATE:GUAM

| ZIP CODE | QUARTER 1 | QUARTER 2 | QUARTER 3 | QUARTER 4 | TOTAL GRAMS |
|---|---|---|---|---|---|
| 969 | 10.11 | 10.16 | 10.49 | 7.79 | 38.55 |
| TOTAL | 10.11 | 10.16 | 10.49 | 7.79 | 38.55 |

STATE: HAWAII

| ZIP CODE | QUARTER 1 | QUARTER 2 | QUARTER 3 | QUARTER 4 | TOTAL GRAMS |
|---|---|---|---|---|---|
| 967 | 139.07 | 133.05 | 119.7 | 114.75 | 506.57 |
| 968 | 84.45 | 89.47 | 86.33 | 80.8 | 341.05 |
| TOTAL | 223.52 | 222.52 | 206.03 | 195.55 | 847.62 |

STATE: IDAHO

| ZIP CODE | QUARTER 1 | QUARTER 2 | QUARTER 3 | QUARTER 4 | TOTAL GRAMS |
|---|---|---|---|---|---|
| 832 | 65.39 | 62 | 62.17 | 55.47 | 245.03 |
| 833 | 74.79 | 76.68 | 71.66 | 66.04 | 289.17 |
| 834 | 66.29 | 67.08 | 64.6 | 60.32 | 258.29 |
| 835 | 55.39 | 51.48 | 46.36 | 44.26 | 197.49 |
| 836 | 186.67 | 186.43 | 164.88 | 178 | 715.98 |
| 837 | 153.81 | 147.31 | 141.43 | 139.99 | 582.54 |
| 838 | 108.82 | 105.22 | 107.68 | 91.69 | 413.41 |
| TOTAL | 711.16 | 696.2 | 658.78 | 635.77 | 2,701.91 |

STATE: ILLINOIS

| ZIP CODE | QUARTER 1 | QUARTER 2 | QUARTER 3 | QUARTER 4 | TOTAL GRAMS |
|---|---|---|---|---|---|
| 600 | 452.14 | 478.86 | 463.5 | 436.58 | 1,831.08 |
| 601 | 325.48 | 318.47 | 312.03 | 292.11 | 1,248.09 |
| 602 | 16.03 | 16.17 | 14.76 | 14.67 | 61.63 |
| 603 | 12.37 | 13.78 | 12.51 | 11.23 | 49.89 |
| 604 | 415.11 | 402.9 | 401.58 | 382.21 | 1,601.80 |

| | | | | | |
|---|---|---|---|---|---|
| 605 | 238.52 | 237.96 | 231.66 | 230.3 | 938.44 |
| 606 | 330.38 | 319.09 | 315.74 | 303.4 | 1,268.61 |
| 607 | 24.29 | 20.32 | 21.01 | 22.61 | 88.23 |
| 608 | 30.11 | 21.72 | 23.82 | 20.39 | 96.04 |
| 609 | 60.35 | 57.15 | 58.51 | 59.59 | 235.6 |
| 610 | 116.14 | 111.85 | 112.49 | 115.03 | 455.51 |
| 611 | 121.42 | 123.62 | 116.18 | 121.97 | 483.19 |
| 612 | 44.23 | 43.69 | 42.4 | 39.63 | 169.95 |
| 613 | 47.96 | 46.33 | 45.29 | 43.38 | 182.96 |
| 614 | 51.38 | 48.57 | 49.7 | 49.98 | 199.63 |
| 615 | 90.08 | 86.2 | 76.82 | 75.39 | 328.49 |
| 616 | 162.43 | 155.91 | 159.05 | 143.6 | 620.99 |
| 617 | 92.45 | 91.31 | 87.62 | 86.6 | 357.98 |
| 618 | 91.61 | 87.49 | 78.99 | 83.28 | 341.37 |
| 619 | 37.06 | 35.44 | 36.16 | 35.41 | 144.07 |
| 620 | 130.2 | 125.35 | 118.99 | 114.63 | 489.17 |
| 622 | 136.71 | 133.38 | 129.67 | 122.82 | 522.58 |
| 623 | 53.96 | 53.73 | 49.37 | 48.37 | 205.43 |
| 624 | 47.84 | 48.26 | 46.95 | 47.55 | 190.6 |
| 625 | 110.74 | 111.79 | 109.12 | 104.71 | 436.36 |
| 626 | 67.52 | 66.37 | 70.81 | 69.98 | 274.68 |
| 627 | 85.39 | 87.45 | 83.59 | 76.16 | 332.59 |
| 628 | 61.06 | 59.57 | 54.11 | 55.31 | 230.05 |
| 629 | 120.35 | 119.56 | 113.43 | 113.1 | 466.44 |
| TOTAL | 3,573.31 | 3,522.29 | 3,435.86 | 3,319.99 | 13,851.45 |

STATE: INDIANA

| ZIP CODE | QUARTER 1 | QUARTER 2 | QUARTER 3 | QUARTER 4 | TOTAL GRAMS |
|---|---|---|---|---|---|
| 460 | 208.52 | 190.58 | 172.53 | 148.19 | 719.82 |
| 461 | 177.28 | 168.18 | 162.41 | 160.31 | 668.18 |
| 462 | 404.62 | 404.34 | 380.76 | 407.28 | 1,597 |
| 463 | 244.77 | 236.56 | 229.41 | 218.72 | 929.46 |
| 464 | 20.19 | 18.67 | 17.97 | 16.41 | 73.24 |
| 465 | 161.54 | 166.66 | 154.97 | 157.56 | 640.73 |
| 466 | 47.94 | 48.18 | 48.1 | 49.53 | 193.75 |
| 467 | 77.79 | 71.87 | 75.62 | 67.08 | 292.36 |
| 468 | 166.21 | 144.02 | 142.01 | 124.91 | 577.15 |
| 469 | 141.36 | 136.32 | 124.57 | 119.42 | 521.67 |
| 470 | 47.72 | 49.73 | 52 | 50.29 | 199.74 |
| 471 | 165.46 | 156.85 | 140.25 | 130.27 | 592.83 |
| 472 | 84.03 | 72.72 | 64.58 | 63.19 | 284.52 |
| 473 | 135.29 | 135.24 | 125.32 | 119.65 | 515.5 |
| 474 | 94.3 | 94.8 | 87.72 | 79.62 | 356.44 |

DRUG CODE:9801DRUG NAME:FENTANYL BASE
STATE:INDIANA

| ZIP CODE | QUARTER 1 | QUARTER 2 | QUARTER 3 | QUARTER 4 | TOTAL GRAMS |
|---|---|---|---|---|---|
| 475 | 68.95 | 64.71 | 63.56 | 65.84 | 263.06 |
| 476 | 29.4 | 35.67 | 28.14 | 28.76 | 121.97 |
| 477 | 72.71 | 68.45 | 57.59 | 54.82 | 253.57 |
| 478 | 55.61 | 56.89 | 51.76 | 44.51 | 208.77 |
| 479 | 72.66 | 76.33 | 74.3 | 73.97 | 297.26 |
| TOTAL | 2,476.35 | 2,396.77 | 2,253.57 | 2,180.33 | 9,307.02 |

STATE: IOWA

| ZIP CODE | QUARTER 1 | QUARTER 2 | QUARTER 3 | QUARTER 4 | TOTAL GRAMS |
|---|---|---|---|---|---|
| 500 | 70.23 | 73.02 | 70.75 | 71.75 | 285.75 |
| 501 | 99.02 | 98.43 | 93.13 | 91.22 | 381.8 |
| 502 | 78.51 | 79.22 | 75.14 | 72.36 | 305.23 |
| 503 | 121.6 | 118.62 | 115.36 | 103.51 | 459.09 |

| | | | | | |
|---|---|---|---|---|---|
| 504 | 45.83 | 46.47 | 44.66 | 48.94 | 185.9 |
| 505 | 49.39 | 48.16 | 48 | 45.41 | 190.96 |
| 506 | 64.14 | 61.86 | 61.43 | 54.71 | 242.14 |
| 507 | 25.03 | 23.82 | 23.61 | 22.04 | 94.5 |
| 508 | 13.88 | 14.3 | 13.08 | 12.74 | 54 |
| 510 | 14.7 | 13.54 | 14.44 | 13.31 | 55.99 |
| 511 | 23.12 | 23.54 | 24.4 | 24.34 | 95.4 |
| 512 | 16.87 | 13.07 | 15.36 | 14.16 | 59.46 |
| 513 | 13 | 12.53 | 11.27 | 12.46 | 49.26 |
| 514 | 29.39 | 27.55 | 26.4 | 25.66 | 109 |
| 515 | 39.02 | 39.32 | 37.8 | 37 | 153.14 |
| 516 | 10.52 | 9.46 | 10.3 | 8.83 | 39.11 |
| 520 | 48.65 | 47.81 | 42.44 | 42.82 | 181.72 |
| 521 | 19.91 | 19.91 | 20.91 | 18.95 | 79.68 |
| 522 | 33.7 | 35.59 | 33.01 | 34.46 | 136.76 |
| 523 | 29.48 | 31.18 | 26.64 | 27.71 | 115.01 |
| 524 | 32.97 | 35.17 | 30.15 | 30.74 | 129.03 |
| 525 | 45.5 | 45.14 | 44.86 | 45.89 | 181.39 |
| 526 | 31.99 | 30.12 | 30.38 | 29.09 | 121.58 |
| 527 | 27.65 | 26.52 | 26.85 | 24.35 | 105.37 |
| 528 | 40.82 | 38.61 | 39.14 | 37.84 | 156.41 |
| TOTAL | 1,024.92 | 1,012.96 | 979.51 | 950.29 | 3,967.68 |

STATE: KANSAS

| ZIP CODE | QUARTER 1 | QUARTER 2 | QUARTER 3 | QUARTER 4 | TOTAL GRAMS |
|---|---|---|---|---|---|
| 660 | 206 | 198.86 | 200.86 | 179.99 | 785.71 |
| 661 | 62.93 | 52.44 | 52.41 | 55.02 | 222.8 |
| 662 | 216.69 | 199.27 | 201.19 | 180.27 | 797.42 |
| 664 | 30.73 | 30.31 | 29.6 | 28.19 | 118.83 |
| 665 | 45.74 | 46.22 | 45.08 | 44.09 | 181.13 |
| 666 | 82.56 | 80.07 | 86.12 | 71.96 | 320.71 |
| 667 | 70.74 | 70.73 | 70.85 | 68.31 | 280.63 |
| 668 | 12.12 | 13 | 12.44 | 12.45 | 50.01 |
| 669 | 19.09 | 18.03 | 19.93 | 19.62 | 76.67 |
| 670 | 65.94 | 68.53 | 60.22 | 60.74 | 255.43 |
| 671 | 44.78 | 45.88 | 42.33 | 37.91 | 170.9 |
| 672 | 169.95 | 163.05 | 159.08 | 151.28 | 643.36 |
| 673 | 39.71 | 37.84 | 36.75 | 34.08 | 148.38 |
| 674 | 75.23 | 78.65 | 77.34 | 74.72 | 305.94 |
| 675 | 45.27 | 43.38 | 42.29 | 43.55 | 174.49 |
| 676 | 33.97 | 33.62 | 35.85 | 36.62 | 140.06 |
| 677 | 19.6 | 18.81 | 21.01 | 21.8 | 81.22 |
| 678 | 56.89 | 57.55 | 55.7 | 54.67 | 224.81 |
| 679 | 24.5 | 22.09 | 23.16 | 22.63 | 92.38 |
| TOTAL | 1,322.44 | 1,278.33 | 1,272.21 | 1,197.90 | 5,070.88 |

STATE: KENTUCKY

| ZIP CODE | QUARTER 1 | QUARTER 2 | QUARTER 3 | QUARTER 4 | TOTAL GRAMS |
|---|---|---|---|---|---|
| 400 | 67.18 | 63.74 | 57.14 | 58.78 | 246.84 |
| 401 | 28.13 | 27.28 | 22.68 | 22.35 | 100.44 |
| 402 | 250.56 | 244.08 | 240.55 | 231.67 | 966.86 |
| 403 | 105.31 | 102.48 | 94.91 | 91.25 | 393.95 |
| 404 | 57.99 | 57.53 | 53.32 | 49.4 | 218.24 |
| 405 | 107.09 | 113.32 | 112.62 | 96.58 | 429.61 |
| 406 | 9.84 | 8.03 | 8.21 | 6.61 | 32.69 |
| 407 | 43.12 | 39.31 | 39.95 | 39.33 | 161.71 |

DRUG CODE:9801DRUG NAME:FENTANYL BASE
STATE:KENTUCKY

| ZIP CODE | QUARTER 1 | QUARTER 2 | QUARTER 3 | QUARTER 4 | TOTAL GRAMS |
|---|---|---|---|---|---|

| | | | | | |
|---|---|---|---|---|---|
| 408 | 8.03 | 9.15 | 7.76 | 8.56 | 33.5 |
| 409 | 18.74 | 17.3 | 18.93 | 17.94 | 72.91 |
| 410 | 167.82 | 165.21 | 156.12 | 148.99 | 638.14 |
| 411 | 40.14 | 40.49 | 41.82 | 38.47 | 160.92 |
| 412 | 17.86 | 17.71 | 17.02 | 15.52 | 68.11 |
| 413 | 12.31 | 11.46 | 13.03 | 11.82 | 48.62 |
| 414 | 3.67 | 3.87 | 3.62 | 2.9 | 14.06 |
| 415 | 27.91 | 22.32 | 23.18 | 20.9 | 94.31 |
| 416 | 17.68 | 13.02 | 13.94 | 13.29 | 57.93 |
| 417 | 38.87 | 36.32 | 33.49 | 31.49 | 140.17 |
| 418 | 5.43 | 3.98 | 3.26 | 3.56 | 16.23 |
| 420 | 60.45 | 59.74 | 59.69 | 55.26 | 235.14 |
| 421 | 76.31 | 77.31 | 77.77 | 72.72 | 304.11 |
| 422 | 35.28 | 37.65 | 35.35 | 33.86 | 142.14 |
| 423 | 50.69 | 49.05 | 47.45 | 43.98 | 191.17 |
| 424 | 59.12 | 64.41 | 60.78 | 57.81 | 242.12 |
| 425 | 37.09 | 34.14 | 29.92 | 30.18 | 131.33 |
| 426 | 25 | 22.95 | 22.4 | 19.88 | 90.23 |
| 427 | 59.17 | 60 | 59.18 | 54.51 | 232.86 |
| TOTAL | 1,430.79 | 1,401.85 | 1,354.09 | 1,277.61 | 5,464.34 |

STATE: LOUISIANA

| ZIP CODE | QUARTER 1 | QUARTER 2 | QUARTER 3 | QUARTER 4 | TOTAL GRAMS |
|---|---|---|---|---|---|
| 700 | 125.3 | 122.57 | 116.84 | 119.53 | 484.24 |
| 701 | 78.55 | 83.67 | 83.55 | 73.97 | 319.74 |
| 703 | 102.32 | 99.4 | 88.61 | 86.17 | 376.5 |
| 704 | 154.75 | 158.5 | 147.59 | 140.02 | 600.86 |
| 705 | 238.72 | 243.51 | 231.63 | 238.65 | 952.51 |
| 706 | 55.54 | 55.81 | 49.35 | 49.22 | 209.92 |
| 707 | 155.42 | 155.35 | 137.22 | 137.47 | 585.46 |
| 708 | 129.67 | 127.53 | 123.16 | 109.62 | 489.98 |
| 710 | 36.41 | 35.54 | 35.99 | 34.27 | 142.21 |
| 711 | 96.32 | 93.44 | 98.33 | 93.81 | 381.9 |
| 712 | 111.51 | 105.53 | 105.33 | 107.76 | 430.13 |
| 713 | 127.37 | 123.25 | 115.15 | 106.46 | 472.23 |
| 714 | 49.62 | 44.47 | 42.14 | 41.79 | 178.02 |
| TOTAL | 1,461.50 | 1,448.57 | 1,374.89 | 1,338.74 | 5,623.70 |

STATE: MAINE

| ZIP CODE | QUARTER 1 | QUARTER 2 | QUARTER 3 | QUARTER 4 | TOTAL GRAMS |
|---|---|---|---|---|---|
| 39 | 9.15 | 8.43 | 6.49 | 6.51 | 30.58 |
| 40 | 152.69 | 145.92 | 144.15 | 116.95 | 559.71 |
| 41 | 40.03 | 41.14 | 37.6 | 30.65 | 149.42 |
| 42 | 78.11 | 71.63 | 66.06 | 69.76 | 285.56 |
| 43 | 63.39 | 68.91 | 57.74 | 56.11 | 246.15 |
| 44 | 59.66 | 55.83 | 52.86 | 48.12 | 216.47 |
| 45 | 10.3 | 11.73 | 10.41 | 9.02 | 41.46 |
| 46 | 31.66 | 26.89 | 25.62 | 20.05 | 104.22 |
| 47 | 39.68 | 36.58 | 32.29 | 26.48 | 135.03 |
| 48 | 7.41 | 7.8 | 11.47 | 9.17 | 35.85 |
| 49 | 70.76 | 70.94 | 66.01 | 55.51 | 263.22 |
| TOTAL | 562.84 | 545.8 | 510.7 | 448.33 | 2,067.67 |

STATE: MARYLAND

| ZIP CODE | QUARTER 1 | QUARTER 2 | QUARTER 3 | QUARTER 4 | TOTAL GRAMS |
|---|---|---|---|---|---|
| 206 | 113.9 | 118.02 | 108.68 | 103 | 443.6 |
| 207 | 152.68 | 159.95 | 148.86 | 141.44 | 602.93 |
| 208 | 208.58 | 187.88 | 200.16 | 175.88 | 772.5 |
| 209 | 42.39 | 43.26 | 40.52 | 42.06 | 168.23 |
| 210 | 270.67 | 282.47 | 262.78 | 250.74 | 1,066.66 |

| | | | | | |
|---|---|---|---|---|---|
| 211 | 144.45 | 140.25 | 132.99 | 127.2 | 544.89 |
| 212 | 410.04 | 398.52 | 383.09 | 371.51 | 1,563.16 |
| 214 | 29.16 | 31.77 | 29.57 | 30.88 | 121.38 |
| 215 | 74.56 | 73.21 | 73.52 | 68.73 | 290.02 |
| 216 | 53.72 | 58.14 | 57.51 | 56.84 | 226.21 |
| 217 | 201.93 | 207.62 | 198.83 | 199.02 | 807.4 |
| 218 | 67.69 | 70.33 | 68.37 | 62.03 | 268.42 |
| 219 | 44.05 | 42.51 | 42.89 | 40.18 | 169.63 |
| TOTAL | 1,813.82 | 1,813.93 | 1,747.77 | 1,669.51 | 7,045.03 |

DRUG CODE:9801DRUG NAME:FENTANYL BASE
STATE: MASSACHUSETTS

| ZIP CODE | QUARTER 1 | QUARTER 2 | QUARTER 3 | QUARTER 4 | TOTAL GRAMS |
|---|---|---|---|---|---|
| 10 | 172.44 | 168.07 | 157.94 | 155.21 | 653.66 |
| 11 | 59.45 | 54.32 | 54.17 | 47.93 | 215.87 |
| 12 | 34.3 | 32.44 | 33.14 | 29.79 | 129.67 |
| 13 | 19 | 20.06 | 17.98 | 17.15 | 74.19 |
| 14 | 45.57 | 45.9 | 41.21 | 36.64 | 169.32 |
| 15 | 72.84 | 72.96 | 67.94 | 67.47 | 281.21 |
| 16 | 66.69 | 57.64 | 56.66 | 48.39 | 229.38 |
| 17 | 82.46 | 80.45 | 79.21 | 79.74 | 321.86 |
| 18 | 239.62 | 222.14 | 220.75 | 206.65 | 889.16 |
| 19 | 101.93 | 101.02 | 95.92 | 90.48 | 389.35 |
| 20 | 63.8 | 64.8 | 60.57 | 60.41 | 249.58 |
| 21 | 203.67 | 200.94 | 190.03 | 185.94 | 780.58 |
| 22 | 26.16 | 31.06 | 24.42 | 25.92 | 107.56 |
| 23 | 132.97 | 130.3 | 122.1 | 114.92 | 500.29 |
| 24 | 47.23 | 45.22 | 47.09 | 49.82 | 189.36 |
| 25 | 28.94 | 26.02 | 25.22 | 24.9 | 105.08 |
| 26 | 33.44 | 30.99 | 27.23 | 23.65 | 115.31 |
| 27 | 192.47 | 187.89 | 175.47 | 175.98 | 731.81 |
| TOTAL | 1,622.98 | 1,572.22 | 1,497.05 | 1,440.99 | 6,133.24 |

STATE: MICHIGAN

| ZIP CODE | QUARTER 1 | QUARTER 2 | QUARTER 3 | QUARTER 4 | TOTAL GRAMS |
|---|---|---|---|---|---|
| 480 | 478.17 | 462.78 | 421.15 | 412.99 | 1,775.09 |
| 481 | 529.53 | 512.81 | 486.25 | 493.26 | 2,021.85 |
| 482 | 126.52 | 111 | 111.07 | 97.06 | 445.65 |
| 483 | 405.12 | 405.5 | 372.1 | 357.77 | 1,540.49 |
| 484 | 157.71 | 155.62 | 149.12 | 147.94 | 610.39 |
| 485 | 90.16 | 86.26 | 78.01 | 76.07 | 330.5 |
| 486 | 244.63 | 256.25 | 226.75 | 234.8 | 962.43 |
| 487 | 116.28 | 121.05 | 113.67 | 106.42 | 457.42 |
| 488 | 208.51 | 208.59 | 171.23 | 193.02 | 781.35 |
| 489 | 78.94 | 75.97 | 64.25 | 67.19 | 286.35 |
| 490 | 396.9 | 402.25 | 356.67 | 376.11 | 1,531.93 |
| 491 | 38.28 | 35.89 | 31.81 | 33.64 | 139.62 |
| 492 | 131.29 | 125.08 | 113.54 | 110.63 | 480.54 |
| 493 | 73.06 | 80.1 | 69.93 | 74.84 | 297.93 |
| 494 | 220 | 214.03 | 195.23 | 197.37 | 826.63 |
| 495 | 214.85 | 208.43 | 191.03 | 196.6 | 810.91 |
| 496 | 175.24 | 171.02 | 162.53 | 152.3 | 661.09 |
| 497 | 121.71 | 119.81 | 112.59 | 114.45 | 468.56 |
| 498 | 103.54 | 91.66 | 93.61 | 88.81 | 377.62 |
| 499 | 31.74 | 30.18 | 32.88 | 30.61 | 125.41 |
| TOTAL | 3,942.18 | 3,874.28 | 3,553.42 | 3,561.88 | 14,931.76 |

STATE: MINNESOTA

| ZIP CODE | QUARTER 1 | QUARTER 2 | QUARTER 3 | QUARTER 4 | TOTAL GRAMS |
|---|---|---|---|---|---|

| | | | | | |
|---|---|---|---|---|---|
| 550 | 137.6 | 130.89 | 126.62 | 118.43 | 513.54 |
| 551 | 177.68 | 178.99 | 175.3 | 164.58 | 696.55 |
| 553 | 160.81 | 152.75 | 154.13 | 142.37 | 610.06 |
| 554 | 277.5 | 289.74 | 243.48 | 256.85 | 1,067.57 |
| 556 | 3.85 | 3.65 | 2.4 | 2.5 | 12.4 |
| 557 | 51.21 | 47.02 | 46.17 | 38.62 | 183.02 |
| 558 | 30.21 | 33.83 | 31.93 | 27.66 | 123.63 |
| 559 | 78.17 | 80.89 | 80.06 | 70.23 | 309.35 |
| 560 | 55.35 | 51.68 | 53.39 | 47.65 | 208.07 |
| 561 | 26.09 | 23.91 | 21.78 | 20.7 | 92.48 |
| 562 | 51.94 | 53.15 | 48.77 | 44.15 | 198.01 |
| 563 | 108.6 | 99.92 | 94.25 | 81.87 | 384.64 |
| 564 | 55.75 | 48.39 | 45.05 | 38.96 | 188.15 |
| 565 | 60.01 | 54.65 | 51.3 | 47.9 | 213.86 |
| 566 | 26.36 | 27.31 | 25.19 | 21.92 | 100.78 |
| 567 | 37.25 | 35.79 | 32.41 | 29.54 | 134.99 |
| TOTAL | 1,338.38 | 1,312.56 | 1,232.23 | 1,153.93 | 5,037.10 |

STATE: MISSISSIPPI

| ZIP CODE | QUARTER 1 | QUARTER 2 | QUARTER 3 | QUARTER 4 | TOTAL GRAMS |
|---|---|---|---|---|---|
| 386 | 154.9 | 148.86 | 139.42 | 142.56 | 585.74 |
| 387 | 31.64 | 29.61 | 28.22 | 28.33 | 117.8 |
| 388 | 109.47 | 119.28 | 121.23 | 115.98 | 465.96 |

DRUG CODE:9801DRUG NAME:FENTANYL BASE
STATE:MISSISSIPPI

| ZIP CODE | QUARTER 1 | QUARTER 2 | QUARTER 3 | QUARTER 4 | TOTAL GRAMS |
|---|---|---|---|---|---|
| 389 | 30.65 | 26.77 | 28.4 | 31.77 | 117.59 |
| 390 | 80.31 | 81.11 | 78.56 | 80.08 | 320.06 |
| 391 | 298.73 | 292.3 | 276.3 | 314.66 | 1,181.99 |
| 392 | 112.8 | 110.03 | 103.64 | 105.17 | 431.64 |
| 393 | 119.19 | 118.76 | 117.68 | 115.53 | 471.16 |
| 394 | 207.82 | 204.95 | 185.98 | 183.48 | 782.23 |
| 395 | 159.4 | 155.99 | 142.7 | 122.66 | 580.75 |
| 396 | 72.99 | 71.58 | 68.91 | 73.3 | 286.78 |
| 397 | 35.96 | 36.95 | 37.49 | 35.8 | 146.2 |
| TOTAL | 1,413.86 | 1,396.19 | 1,328.53 | 1,349.32 | 5,487.90 |

STATE: MISSOURI

| ZIP CODE | QUARTER 1 | QUARTER 2 | QUARTER 3 | QUARTER 4 | TOTAL GRAMS |
|---|---|---|---|---|---|
| 630 | 259.99 | 247.74 | 230.72 | 218.71 | 957.16 |
| 631 | 453.21 | 438.03 | 403.72 | 404.81 | 1,699.77 |
| 633 | 151.98 | 150.61 | 137.74 | 133.51 | 573.84 |
| 634 | 35.17 | 31.63 | 31.48 | 29.53 | 127.81 |
| 635 | 45.51 | 40.17 | 39.87 | 35.92 | 161.47 |
| 636 | 58.4 | 62.07 | 51.83 | 53.44 | 225.74 |
| 637 | 45.11 | 41.93 | 40.89 | 41.48 | 169.41 |
| 638 | 54.06 | 50.75 | 50.39 | 47.69 | 202.89 |
| 639 | 27.12 | 28.97 | 26.29 | 22.82 | 105.2 |
| 640 | 256.91 | 253.77 | 242.34 | 221.13 | 974.15 |
| 641 | 224.55 | 219.65 | 206.03 | 216.82 | 867.05 |
| 644 | 36.26 | 37.26 | 34.62 | 32.72 | 140.86 |
| 645 | 59.93 | 59.57 | 52.84 | 50.01 | 222.35 |
| 646 | 31.78 | 27.72 | 27.97 | 29.79 | 117.26 |
| 647 | 78.1 | 75.28 | 73.32 | 73.29 | 299.99 |
| 648 | 97.09 | 94.96 | 95.38 | 92.39 | 379.82 |
| 650 | 59.67 | 56.13 | 54.66 | 48.86 | 219.32 |
| 651 | 36.62 | 35.05 | 32.36 | 31.27 | 135.3 |
| 652 | 86.94 | 84.92 | 83.21 | 87.13 | 342.2 |
| 653 | 35.41 | 35.44 | 36.04 | 36.79 | 143.68 |

| | | | | | |
|---|---|---|---|---|---|
| 654 | 48.68 | 49.19 | 46.59 | 44.84 | 189.3 |
| 655 | 55.43 | 50.08 | 48.03 | 48.31 | 201.85 |
| 656 | 102.4 | 98.19 | 97.73 | 93.33 | 391.65 |
| 657 | 158.35 | 153.15 | 145.13 | 143.66 | 600.29 |
| 658 | 181.88 | 168.9 | 168.03 | 167.33 | 686.14 |
| TOTAL | 2,680.55 | 2,591.16 | 2,457.21 | 2,405.58 | 10,134.50 |

STATE: MONTANA

| ZIP CODE | QUARTER 1 | QUARTER 2 | QUARTER 3 | QUARTER 4 | TOTAL GRAMS |
|---|---|---|---|---|---|
| 590 | 21.5 | 23.21 | 24.21 | 20.83 | 89.75 |
| 591 | 83.82 | 86.61 | 80.42 | 74.11 | 324.96 |
| 592 | 17.74 | 16.67 | 15.89 | 14.58 | 64.88 |
| 593 | 16.93 | 19.03 | 18.78 | 16.39 | 71.13 |
| 594 | 59.92 | 57.32 | 58.37 | 56.72 | 232.33 |
| 595 | 14.67 | 14.81 | 13.96 | 12.31 | 55.75 |
| 596 | 43.63 | 43.07 | 38.98 | 38.82 | 164.5 |
| 597 | 43.53 | 39.25 | 36.07 | 41.88 | 160.73 |
| 598 | 68.04 | 73.02 | 71.24 | 65.44 | 277.74 |
| 599 | 44.98 | 42.79 | 41.25 | 37.59 | 166.61 |
| TOTAL | 414.76 | 415.78 | 399.17 | 378.67 | 1,608.38 |

STATE: NEBRASKA

| ZIP CODE | QUARTER 1 | QUARTER 2 | QUARTER 3 | QUARTER 4 | TOTAL GRAMS |
|---|---|---|---|---|---|
| 680 | 73.29 | 73.2 | 73.44 | 68.58 | 288.51 |
| 681 | 179.46 | 173.12 | 153.56 | 164.24 | 670.38 |
| 683 | 30.71 | 31.62 | 27.3 | 26.64 | 116.27 |
| 684 | 20.64 | 20.56 | 18.64 | 19.16 | 79 |
| 685 | 61.6 | 64.94 | 63.61 | 58.12 | 248.27 |
| 686 | 16.86 | 15.06 | 16.35 | 14.37 | 62.64 |
| 687 | 36.61 | 39.16 | 40.35 | 37.78 | 153.9 |
| 688 | 55.57 | 68.02 | 58.48 | 62.14 | 244.21 |
| 689 | 31.73 | 34.11 | 31.04 | 29.81 | 126.69 |
| 690 | 19.72 | 19.13 | 18.9 | 18.39 | 76.14 |
| 691 | 38.04 | 41.44 | 39.42 | 39.79 | 158.69 |
| 692 | 2.81 | 2.53 | 2.59 | 3.17 | 11.1 |
| 693 | 39.96 | 40.38 | 39.89 | 35.92 | 156.15 |

DRUG CODE:9801DRUG NAME:FENTANYL BASE
STATE:NEBRASKA

| ZIP CODE | QUARTER 1 | QUARTER 2 | QUARTER 3 | QUARTER 4 | TOTAL GRAMS |
|---|---|---|---|---|---|
| TOTAL | 607 | 623.27 | 583.57 | 578.11 | 2,391.95 |

STATE: NEVADA

| ZIP CODE | QUARTER 1 | QUARTER 2 | QUARTER 3 | QUARTER 4 | TOTAL GRAMS |
|---|---|---|---|---|---|
| 890 | 191.7 | 178.72 | 181.41 | 172.33 | 724.16 |
| 891 | 397.16 | 369.5 | 358.54 | 347.24 | 1,472.44 |
| 893 | 2.11 | 1.41 | 1.74 | 1.94 | 7.2 |
| 894 | 65.95 | 66.4 | 63.09 | 57.71 | 253.15 |
| 895 | 99.06 | 98.54 | 90.76 | 83.48 | 371.84 |
| 897 | 25.84 | 25.08 | 21.39 | 22.06 | 94.37 |
| 898 | 15.96 | 13.85 | 13.33 | 11.65 | 54.79 |
| TOTAL | 797.78 | 753.5 | 730.26 | 696.41 | 2,977.95 |

STATE: NEW HAMPSHIRE

| ZIP CODE | QUARTER 1 | QUARTER 2 | QUARTER 3 | QUARTER 4 | TOTAL GRAMS |
|---|---|---|---|---|---|
| 30 | 121.22 | 127.68 | 126.9 | 116.13 | 491.93 |
| 31 | 59.59 | 57.97 | 54 | 49.99 | 221.55 |
| 32 | 38.18 | 39.27 | 33.42 | 35.27 | 146.14 |
| 33 | 58.11 | 53.52 | 57.14 | 51.55 | 220.32 |
| 34 | 16.83 | 16.24 | 14.43 | 12.16 | 59.66 |

| | | | | | |
|---|---|---|---|---|---|
| 35 | 19.95 | 21.9 | 21.51 | 22.23 | 85.59 |
| 36 | 1.98 | 1.3 | 1.67 | 1.6 | 6.55 |
| 37 | 29.91 | 31.11 | 31.65 | 25.38 | 118.05 |
| 38 | 110.62 | 102.24 | 100.99 | 92.12 | 405.97 |
| TOTAL | 456.39 | 451.23 | 441.71 | 406.43 | 1,755.76 |

STATE: NEW JERSEY

| ZIP CODE | QUARTER 1 | QUARTER 2 | QUARTER 3 | QUARTER 4 | TOTAL GRAMS |
|---|---|---|---|---|---|
| 70 | 549.65 | 575.05 | 536.13 | 573.11 | 2,233.94 |
| 71 | 48.21 | 46.69 | 43.61 | 43.35 | 181.86 |
| 72 | 49.03 | 83.88 | 40.32 | 15.85 | 189.08 |
| 73 | 18.21 | 21.47 | 21.04 | 18.27 | 78.99 |
| 74 | 167.81 | 177.12 | 164.76 | 156.41 | 666.1 |
| 75 | 28.96 | 26.16 | 28.82 | 30.68 | 114.62 |
| 76 | 160.24 | 152.81 | 207.35 | 230.83 | 751.23 |
| 77 | 223.16 | 223.78 | 211.6 | 199.21 | 857.75 |
| 78 | 95.54 | 94.64 | 93.01 | 85.43 | 368.62 |
| 79 | 79.87 | 80.21 | 79.33 | 75.76 | 315.17 |
| 80 | 521.06 | 528.98 | 496.76 | 473.55 | 2,020.35 |
| 81 | 44.75 | 38.16 | 37.65 | 190.22 | 310.78 |
| 82 | 98.72 | 102.14 | 100.33 | 89.6 | 390.79 |
| 83 | 46.87 | 44.77 | 42.96 | 40.42 | 175.02 |
| 84 | 14.52 | 13.63 | 14.54 | 12.29 | 54.98 |
| 85 | 86.83 | 81.5 | 76.85 | 77.94 | 323.12 |
| 86 | 90.08 | 86.64 | 80.93 | 78.3 | 335.95 |
| 87 | 171.55 | 161.46 | 161.07 | 147.27 | 641.35 |
| 88 | 352.81 | 351.9 | 326.72 | 323.14 | 1,354.57 |
| 89 | 38.71 | 33.72 | 30.81 | 27.38 | 130.62 |
| TOTAL | 2,886.58 | 2,924.71 | 2,794.59 | 2,889.01 | 11,494.89 |

STATE: NEW MEXICO

| ZIP CODE | QUARTER 1 | QUARTER 2 | QUARTER 3 | QUARTER 4 | TOTAL GRAMS |
|---|---|---|---|---|---|
| 870 | 35.55 | 38.19 | 35.43 | 35.93 | 145.1 |
| 871 | 274.95 | 283.78 | 272.14 | 246.59 | 1,077.46 |
| 873 | 10.38 | 9.74 | 10.66 | 8.37 | 39.15 |
| 874 | 33.04 | 30.83 | 30.37 | 32.55 | 126.79 |
| 875 | 61.51 | 56.98 | 55.35 | 56.93 | 230.77 |
| 877 | 10.23 | 9.54 | 8.36 | 8.81 | 36.94 |
| 878 | 4.33 | 3.3 | 2.93 | 3.58 | 14.14 |
| 879 | 3.79 | 3.4 | 3.24 | 3.95 | 14.38 |
| 880 | 35.62 | 36.68 | 33.27 | 33.41 | 138.98 |
| 881 | 17.26 | 14.91 | 13.33 | 14.57 | 60.07 |
| 882 | 67.06 | 66.2 | 65.17 | 64 | 262.43 |
| 883 | 20.93 | 20.11 | 19.39 | 19.68 | 80.11 |
| 884 | 3.55 | 3.88 | 4.51 | 4.98 | 16.92 |
| TOTAL | 578.2 | 577.54 | 554.15 | 533.35 | 2,243.24 |

STATE: NEW YORK

| ZIP CODE | QUARTER 1 | QUARTER 2 | QUARTER 3 | QUARTER 4 | TOTAL GRAMS |
|---|---|---|---|---|---|
| 100 | 553.72 | 519.49 | 486.61 | 450.56 | 2,010.38 |
| 101 | 10.65 | 10.71 | 11.21 | 11.27 | 43.84 |

DRUG CODE:9801DRUG NAME:FENTANYL BASE
STATE:NEW YORK

| ZIP CODE | QUARTER 1 | QUARTER 2 | QUARTER 3 | QUARTER 4 | TOTAL GRAMS |
|---|---|---|---|---|---|
| 102 | 0.05 | 0.15 | 0.29 | 0.53 | 1.02 |
| 103 | 172.81 | 170.65 | 161.69 | 166.59 | 671.74 |
| 104 | 194.92 | 207.44 | 186.62 | 170.71 | 759.69 |
| 105 | 143.54 | 139.62 | 132.47 | 128.48 | 544.11 |
| 106 | 22.55 | 22.65 | 20.96 | 21.2 | 87.36 |

| | | | | | |
|---|---|---|---|---|---|
| 107 | 46.54 | 48.82 | 46.39 | 42.29 | 184.04 |
| 108 | 19.9 | 19.74 | 17.64 | 16.01 | 73.29 |
| 109 | 209.39 | 207.64 | 196.81 | 175.49 | 789.33 |
| 110 | 95.42 | 100.13 | 93.07 | 93.02 | 381.64 |
| 111 | 29.42 | 30.53 | 27.49 | 25.07 | 112.51 |
| 112 | 298.09 | 302.19 | 290.69 | 287.56 | 1,178.53 |
| 113 | 166.71 | 141.89 | 131.13 | 122.14 | 561.87 |
| 114 | 74.16 | 63.25 | 71.81 | 70.71 | 279.93 |
| 115 | 173.09 | 168.34 | 178.77 | 165.65 | 685.85 |
| 116 | 15.81 | 16.13 | 12.17 | 13.52 | 57.63 |
| 117 | 690.87 | 674.74 | 670.5 | 592.3 | 2,628.41 |
| 118 | 30.19 | 38.72 | 32.55 | 28.09 | 129.55 |
| 119 | 76.14 | 76.15 | 73.36 | 72.63 | 298.28 |
| 120 | 121.65 | 120.1 | 111.09 | 101.29 | 454.13 |
| 121 | 51.25 | 48.11 | 48.24 | 46.26 | 193.86 |
| 122 | 88.5 | 88.86 | 89.88 | 85.14 | 352.38 |
| 123 | 30.94 | 34.56 | 33.3 | 32.79 | 131.59 |
| 124 | 71.3 | 64.27 | 64.15 | 58.24 | 257.96 |
| 125 | 133.2 | 131.1 | 119.06 | 109.02 | 492.38 |
| 126 | 38.68 | 33.51 | 35.7 | 30.12 | 138.01 |
| 127 | 51.99 | 45.61 | 44.19 | 46.25 | 188.04 |
| 128 | 100.21 | 100.51 | 105.69 | 96.41 | 402.82 |
| 129 | 74.4 | 69.48 | 62.15 | 54.4 | 260.43 |
| 130 | 149.19 | 136.59 | 119.23 | 109.03 | 514.04 |
| 131 | 58.47 | 47.03 | 40.99 | 37.97 | 184.46 |
| 132 | 161.99 | 159.37 | 139.09 | 129.86 | 590.31 |
| 133 | 64.31 | 62.43 | 55.76 | 48.58 | 231.08 |
| 134 | 95.11 | 89.66 | 89.28 | 80.26 | 354.31 |
| 135 | 41.05 | 37.73 | 33.7 | 32.51 | 144.99 |
| 136 | 86.99 | 90.69 | 77.3 | 73.34 | 328.32 |
| 137 | 41.49 | 36.16 | 38.02 | 33.31 | 148.98 |
| 138 | 108.29 | 105.2 | 105.59 | 104.48 | 423.56 |
| 139 | 37.98 | 39.17 | 33.52 | 32.81 | 143.48 |
| 140 | 166.31 | 145.27 | 143 | 132.19 | 586.77 |
| 141 | 95.92 | 87.94 | 83.44 | 72.65 | 339.95 |
| 142 | 275.64 | 273.17 | 244.5 | 224.49 | 1,017.80 |
| 143 | 42.22 | 39.6 | 33.37 | 29.88 | 145.07 |
| 144 | 95.57 | 91.28 | 88.39 | 86.69 | 361.93 |
| 145 | 91.52 | 90.48 | 90.79 | 86.37 | 359.16 |
| 146 | 191.6 | 183.25 | 176.95 | 163.48 | 715.28 |
| 147 | 81.94 | 77.62 | 73.1 | 66.25 | 298.91 |
| 148 | 152.65 | 140.15 | 128.88 | 117.19 | 538.87 |
| 149 | 42.12 | 38.32 | 34.82 | 33.08 | 148.34 |
| TOTAL | 5,866.45 | 5,666.20 | 5,385.40 | 5,008.16 | 21,926.21 |

STATE: NORTH CAROLINA

| ZIP CODE | QUARTER 1 | QUARTER 2 | QUARTER 3 | QUARTER 4 | TOTAL GRAMS |
|---|---|---|---|---|---|
| 270 | 98.21 | 99.97 | 90.9 | 90.03 | 379.11 |
| 271 | 155.73 | 151.09 | 133.05 | 124.58 | 564.45 |
| 272 | 222.45 | 232.67 | 214.14 | 202.69 | 871.95 |
| 273 | 141.58 | 135.23 | 125.07 | 125.11 | 526.99 |
| 274 | 83.49 | 77.98 | 73.08 | 74.51 | 309.06 |
| 275 | 318.27 | 321.69 | 282.5 | 278.52 | 1,200.98 |
| 276 | 146.72 | 145.05 | 149.46 | 143.26 | 584.49 |
| 277 | 108.91 | 106.13 | 127.79 | 103 | 445.83 |
| 278 | 214.33 | 204.73 | 207.64 | 201.25 | 827.95 |
| 279 | 80.6 | 75.67 | 72.88 | 70 | 299.15 |
| 280 | 297 | 294.85 | 274.12 | 248.61 | 1,114.58 |
| 281 | 209.07 | 207.75 | 190.08 | 184.06 | 790.96 |
| 282 | 188.58 | 185.27 | 180.08 | 165.12 | 719.05 |
| 283 | 379.63 | 351.91 | 298.87 | 266.68 | 1,297.09 |
| 284 | 296.12 | 269.87 | 227.33 | 239.07 | 1,032.39 |

RETAIL DRUG DISTRIBUTION BY ZIP CODE WITHIN STATE BY GRAMS WT

DRUG CODE:9801DRUG NAME:FENTANYL BASE
STATE:NORTH CAROLINA

| ZIP CODE | QUARTER 1 | QUARTER 2 | QUARTER 3 | QUARTER 4 | TOTAL GRAMS |
|---|---|---|---|---|---|
| 285 | 224.55 | 211.74 | 215.62 | 199.09 | 851 |
| 286 | 314.17 | 317.72 | 303.52 | 291.5 | 1,226.91 |
| 287 | 193.68 | 191.69 | 180.91 | 167.45 | 733.73 |
| 288 | 103.19 | 90.26 | 83.21 | 78.08 | 354.74 |
| 289 | 23.83 | 21.04 | 21.12 | 21.99 | 87.98 |
| TOTAL | 3,800.11 | 3,692.31 | 3,451.37 | 3,274.60 | 14,218.39 |

STATE: NORTH DAKOTA

| ZIP CODE | QUARTER 1 | QUARTER 2 | QUARTER 3 | QUARTER 4 | TOTAL GRAMS |
|---|---|---|---|---|---|
| 580 | 21.55 | 19.79 | 18.56 | 15.58 | 75.48 |
| 581 | 72.5 | 66.74 | 60.65 | 47.35 | 247.24 |
| 582 | 58.9 | 54.1 | 52.26 | 43.83 | 209.09 |
| 583 | 23.38 | 22.08 | 22.74 | 22.23 | 90.43 |
| 584 | 29.88 | 25.65 | 23.21 | 19.74 | 98.48 |
| 585 | 74.84 | 70.25 | 69.45 | 59.52 | 274.06 |
| 586 | 20.42 | 19.33 | 19.21 | 16.66 | 75.62 |
| 587 | 23.75 | 22.34 | 20.91 | 17.02 | 84.02 |
| 588 | 13.63 | 13.61 | 11.48 | 7.39 | 46.11 |
| TOTAL | 338.85 | 313.89 | 298.47 | 249.32 | 1,200.53 |

STATE: OHIO

| ZIP CODE | QUARTER 1 | QUARTER 2 | QUARTER 3 | QUARTER 4 | TOTAL GRAMS |
|---|---|---|---|---|---|
| 430 | 160.63 | 159.57 | 183.42 | 171.02 | 674.64 |
| 431 | 96.34 | 92.6 | 91.47 | 83.85 | 364.26 |
| 432 | 471.2 | 452.93 | 403.52 | 373.77 | 1,701.42 |
| 433 | 42.14 | 41.47 | 40.75 | 37.23 | 161.59 |
| 434 | 36.92 | 38.43 | 29.67 | 30.03 | 135.05 |
| 435 | 111.2 | 104.27 | 97.25 | 94.97 | 407.69 |
| 436 | 227.12 | 210.42 | 200.35 | 181.8 | 819.69 |
| 437 | 43.92 | 45.16 | 45.03 | 41.53 | 175.64 |
| 438 | 7.13 | 6.42 | 6.18 | 5.06 | 24.79 |
| 439 | 46.56 | 46.39 | 44.97 | 47.7 | 185.62 |
| 440 | 161.05 | 157.65 | 150.94 | 158.59 | 628.23 |
| 441 | 230.13 | 215.94 | 209.71 | 199.42 | 855.2 |
| 442 | 117.13 | 125.95 | 111.61 | 104.24 | 458.93 |
| 443 | 47.08 | 45.37 | 44.76 | 41.79 | 179 |
| 444 | 81.29 | 82.85 | 70.31 | 67.25 | 301.7 |
| 445 | 82.02 | 80.25 | 80.46 | 78.01 | 320.74 |
| 446 | 117.21 | 117.38 | 108.53 | 100.79 | 443.91 |
| 447 | 70.53 | 64.23 | 65.17 | 58.17 | 258.1 |
| 448 | 84.93 | 76.51 | 67.24 | 66.2 | 294.88 |
| 449 | 22.78 | 24.26 | 24.77 | 22.25 | 94.06 |
| 450 | 213.16 | 209.16 | 195.96 | 187.5 | 805.78 |
| 451 | 65.24 | 65.35 | 64.11 | 59.1 | 253.8 |
| 452 | 217.33 | 218.76 | 206.03 | 190.84 | 832.96 |
| 453 | 134.44 | 121.45 | 114.47 | 114.9 | 485.26 |
| 454 | 151.97 | 146.88 | 132.7 | 130.06 | 561.61 |
| 455 | 34.94 | 31.45 | 31.82 | 28.94 | 127.15 |
| 456 | 106.03 | 101.07 | 98.91 | 96.95 | 402.96 |
| 457 | 38.96 | 36.75 | 33.59 | 31.01 | 140.31 |
| 458 | 79 | 69.16 | 66.73 | 63.62 | 278.51 |
| TOTAL | 3,298.38 | 3,188.08 | 3,020.43 | 2,866.59 | 12,373.48 |

STATE: OKLAHOMA

| ZIP CODE | QUARTER 1 | QUARTER 2 | QUARTER 3 | QUARTER 4 | TOTAL GRAMS |
|---|---|---|---|---|---|

| | | | | | |
|---|---|---|---|---|---|
| 730 | 327.9 | 325.75 | 313.95 | 291.92 | 1,259.52 |
| 731 | 362.78 | 364.27 | 347.51 | 339.26 | 1,413.82 |
| 734 | 45.5 | 46.81 | 45.81 | 45.83 | 183.95 |
| 735 | 138.87 | 132.74 | 139.6 | 122.1 | 533.31 |
| 736 | 48.63 | 50.32 | 46.89 | 50.46 | 196.3 |
| 737 | 51.14 | 49.17 | 55.45 | 51.96 | 207.72 |
| 738 | 15.89 | 15.18 | 14.91 | 14.71 | 60.69 |
| 739 | 6.04 | 4.64 | 3.94 | 3.57 | 18.19 |
| 740 | 348.22 | 343.98 | 340.38 | 321.48 | 1,354.06 |
| 741 | 308.52 | 288.92 | 275.49 | 253.56 | 1,126.49 |
| 743 | 65.75 | 65.77 | 63.32 | 59.7 | 254.54 |
| 744 | 136.83 | 138.61 | 141.52 | 121.8 | 538.76 |
| 745 | 52.18 | 52.59 | 51.05 | 48.1 | 203.92 |
| 746 | 30.56 | 30.3 | 28.25 | 29.28 | 118.39 |

DRUG CODE:9801DRUG NAME:FENTANYL BASE
STATE:OKLAHOMA

| ZIP CODE | QUARTER 1 | QUARTER 2 | QUARTER 3 | QUARTER 4 | TOTAL GRAMS |
|---|---|---|---|---|---|
| 747 | 47.05 | 45.09 | 44.26 | 40.8 | 177.2 |
| 748 | 86.54 | 84.21 | 76.95 | 73.79 | 321.49 |
| 749 | 33.08 | 32.6 | 29.91 | 27.46 | 123.05 |
| TOTAL | 2,105.48 | 2,070.95 | 2,019.19 | 1,895.78 | 8,091.40 |

STATE: OREGON

| ZIP CODE | QUARTER 1 | QUARTER 2 | QUARTER 3 | QUARTER 4 | TOTAL GRAMS |
|---|---|---|---|---|---|
| 970 | 194.11 | 191.41 | 177.17 | 164.33 | 727.02 |
| 971 | 100.77 | 101.98 | 90.33 | 84.14 | 377.22 |
| 972 | 292.1 | 299.07 | 268.52 | 241.55 | 1,101.24 |
| 973 | 180.59 | 160.89 | 159.44 | 141.54 | 642.46 |
| 974 | 241.93 | 224.95 | 212.31 | 193.86 | 873.05 |
| 975 | 127.82 | 125.45 | 113.3 | 108.08 | 474.65 |
| 976 | 21.54 | 21.85 | 19.98 | 18.04 | 81.41 |
| 977 | 71.98 | 62.17 | 61.87 | 56.4 | 252.42 |
| 978 | 75.07 | 74.32 | 65.29 | 61.85 | 276.53 |
| 979 | 18.81 | 19.12 | 18.95 | 15.99 | 72.87 |
| TOTAL | 1,324.72 | 1,281.21 | 1,187.16 | 1,085.78 | 4,878.87 |

STATE: PENNSYLVANIA

| ZIP CODE | QUARTER 1 | QUARTER 2 | QUARTER 3 | QUARTER 4 | TOTAL GRAMS |
|---|---|---|---|---|---|
| 150 | 310.6 | 297.06 | 293.49 | 279.05 | 1,180.20 |
| 151 | 119.58 | 121.68 | 111.18 | 118.42 | 470.86 |
| 152 | 309.82 | 305.87 | 297.42 | 278.93 | 1,192.04 |
| 153 | 51.75 | 55.23 | 52.59 | 50.51 | 210.08 |
| 154 | 60.55 | 64.43 | 61.09 | 57.27 | 243.34 |
| 155 | 57.16 | 55.18 | 53.51 | 51.8 | 217.65 |
| 156 | 138.57 | 137.96 | 132.44 | 134.97 | 543.94 |
| 157 | 99.05 | 99.18 | 98.35 | 100.86 | 397.44 |
| 158 | 77.85 | 78.01 | 76.67 | 73.66 | 306.19 |
| 159 | 112.01 | 111.8 | 111.26 | 111.55 | 446.62 |
| 160 | 80.45 | 84.73 | 83.32 | 82.82 | 331.32 |
| 161 | 116.77 | 108.73 | 102.77 | 95.02 | 423.29 |
| 162 | 48.94 | 52.11 | 48.57 | 45.83 | 195.45 |
| 163 | 113.06 | 111.03 | 108.13 | 102.38 | 434.6 |
| 164 | 31.85 | 28.81 | 28.34 | 26.81 | 115.81 |
| 165 | 86.33 | 90.27 | 85.55 | 83.99 | 346.14 |
| 166 | 139.22 | 133.5 | 127.09 | 128.19 | 528 |
| 167 | 23.52 | 24.21 | 23.15 | 19.32 | 90.2 |
| 168 | 55.72 | 54.95 | 56.47 | 54.71 | 221.85 |

| | | | | | |
|---|---|---|---|---|---|
| 169 | 31.59 | 29.06 | 29.76 | 26.23 | 116.64 |
| 170 | 272.31 | 254.58 | 249.73 | 226.31 | 1,002.93 |
| 171 | 80.24 | 77.92 | 77.99 | 73.04 | 309.19 |
| 172 | 68.96 | 67.18 | 61.54 | 57.91 | 255.59 |
| 173 | 131.93 | 125.57 | 124.49 | 120.3 | 502.29 |
| 174 | 84.38 | 81.37 | 76.5 | 74.72 | 316.97 |
| 175 | 79.53 | 72.43 | 70.62 | 70.17 | 292.75 |
| 176 | 62.35 | 68.19 | 59.73 | 62.14 | 252.41 |
| 177 | 105.56 | 106.44 | 104.88 | 99.66 | 416.54 |
| 178 | 132.77 | 131.39 | 120.34 | 122.06 | 506.56 |
| 179 | 50.75 | 45.8 | 41.32 | 41.34 | 179.21 |
| 180 | 171.6 | 173.97 | 178.85 | 165.16 | 689.58 |
| 181 | 135.92 | 135.76 | 127.31 | 117.12 | 516.11 |
| 182 | 84.17 | 86.7 | 87.49 | 82.42 | 340.78 |
| 183 | 80.76 | 75.47 | 72.49 | 65.54 | 294.26 |
| 184 | 84.06 | 85.74 | 78.64 | 78.64 | 327.08 |
| 185 | 141.89 | 134.07 | 128.08 | 120.39 | 524.43 |
| 186 | 93.39 | 95.89 | 86.1 | 85.67 | 361.05 |
| 187 | 415.64 | 414.13 | 376.02 | 376.65 | 1,582.44 |
| 188 | 40.47 | 38.55 | 38.38 | 42.64 | 160.04 |
| 189 | 127.06 | 125.88 | 124.64 | 113.21 | 490.79 |
| 190 | 741.41 | 725.73 | 728.8 | 663.82 | 2,859.76 |
| 191 | 477.27 | 491.32 | 458.65 | 432.81 | 1,860.05 |
| 193 | 123.95 | 125.69 | 124.98 | 119.73 | 494.35 |
| 194 | 238.03 | 235.29 | 216.37 | 210.27 | 899.96 |
| 195 | 55.04 | 55.48 | 53.44 | 53.5 | 217.46 |
| 196 | 63 | 52.39 | 53.63 | 50.98 | 220 |
| TOTAL | 6,206.83 | 6,126.73 | 5,902.16 | 5,648.52 | 23,884.24 |

DRUG CODE:9801DRUG NAME:FENTANYL BASE
STATE: PUERTO RICO

| ZIP CODE | QUARTER 1 | QUARTER 2 | QUARTER 3 | QUARTER 4 | TOTAL GRAMS |
|---|---|---|---|---|---|
| 6 | 56.45 | 56.31 | 61.92 | 53.47 | 228.15 |
| 7 | 75.32 | 73.59 | 70.57 | 68.32 | 287.8 |
| 9 | 87.02 | 88.47 | 76.37 | 94.55 | 346.41 |
| TOTAL | 218.79 | 218.37 | 208.86 | 216.34 | 862.36 |

STATE: RHODE ISLAND

| ZIP CODE | QUARTER 1 | QUARTER 2 | QUARTER 3 | QUARTER 4 | TOTAL GRAMS |
|---|---|---|---|---|---|
| 28 | 152.29 | 149.79 | 148.05 | 135.78 | 585.91 |
| 29 | 86.17 | 87.64 | 82.5 | 80.02 | 336.33 |
| TOTAL | 238.46 | 237.43 | 230.55 | 215.8 | 922.24 |

STATE: SOUTH CAROLINA

| ZIP CODE | QUARTER 1 | QUARTER 2 | QUARTER 3 | QUARTER 4 | TOTAL GRAMS |
|---|---|---|---|---|---|
| 290 | 102.48 | 98.05 | 98.14 | 95.43 | 394.1 |
| 291 | 117.46 | 115.79 | 108.03 | 110.72 | 452 |
| 292 | 115.11 | 111.8 | 111.81 | 107.71 | 446.43 |
| 293 | 146.61 | 137.57 | 146.42 | 125.91 | 556.51 |
| 294 | 229.34 | 210.29 | 215.87 | 220.68 | 876.18 |
| 295 | 296.84 | 287.22 | 285.1 | 271.86 | 1,141.02 |
| 296 | 335.22 | 327.76 | 311.15 | 315.16 | 1,289.29 |
| 297 | 79.81 | 81.51 | 75.5 | 172.63 | 409.45 |
| 298 | 91.09 | 91.32 | 86.29 | 86.83 | 355.53 |
| 299 | 88.15 | 85.41 | 78.22 | 86.85 | 338.63 |
| TOTAL | 1,602.11 | 1,546.72 | 1,516.53 | 1,593.78 | 6,259.14 |

STATE: SOUTH DAKOTA

RETAIL DRUG DISTRIBUTION BY ZIP CODE WITHIN STATE BY GRAMS WT

| ZIP CODE | QUARTER 1 | QUARTER 2 | QUARTER 3 | QUARTER 4 | TOTAL GRAMS |
|---|---|---|---|---|---|
| 570 | 68.26 | 62.77 | 52.15 | 48.39 | 231.57 |
| 571 | 160.51 | 147.9 | 149.85 | 158.93 | 617.19 |
| 572 | 24.05 | 23.44 | 18.12 | 17.56 | 83.17 |
| 573 | 26.88 | 27.59 | 25.31 | 21.79 | 101.57 |
| 574 | 28.39 | 25.82 | 26.66 | 23.26 | 104.13 |
| 575 | 19.49 | 21.25 | 20.74 | 20.43 | 81.91 |
| 576 | 3 | 3.5 | 3.02 | 1.75 | 11.27 |
| 577 | 53.67 | 51.8 | 52.6 | 49.39 | 207.46 |
| TOTAL | 384.25 | 364.07 | 348.45 | 341.5 | 1,438.27 |

STATE: TENNESSEE

| ZIP CODE | QUARTER 1 | QUARTER 2 | QUARTER 3 | QUARTER 4 | TOTAL GRAMS |
|---|---|---|---|---|---|
| 370 | 299.27 | 289.69 | 281.19 | 270.64 | 1,140.79 |
| 371 | 236.04 | 236.3 | 242.1 | 220.63 | 935.07 |
| 372 | 189.02 | 181.04 | 180.47 | 170.95 | 721.48 |
| 373 | 213.8 | 202.01 | 190.89 | 177.49 | 784.19 |
| 374 | 91.34 | 83.83 | 80.97 | 76.5 | 332.64 |
| 376 | 179.34 | 173.6 | 164.14 | 153.45 | 670.53 |
| 377 | 107.4 | 94.71 | 88.54 | 83.99 | 374.64 |
| 378 | 177.74 | 172.03 | 157.44 | 180.64 | 687.85 |
| 379 | 133.92 | 117.66 | 117.94 | 209.28 | 578.8 |
| 380 | 104.18 | 107.07 | 99.25 | 97.66 | 408.16 |
| 381 | 466.88 | 421.28 | 473.8 | 341.51 | 1,703.47 |
| 382 | 40.16 | 37.91 | 37.78 | 33.96 | 149.81 |
| 383 | 102.12 | 96.26 | 92.94 | 88.35 | 379.67 |
| 384 | 63.62 | 61.77 | 58.19 | 56.17 | 239.75 |
| 385 | 130.94 | 135.82 | 125.7 | 120.52 | 512.98 |
| TOTAL | 2,535.77 | 2,410.98 | 2,391.34 | 2,281.74 | 9,619.83 |

STATE: TEXAS

| ZIP CODE | QUARTER 1 | QUARTER 2 | QUARTER 3 | QUARTER 4 | TOTAL GRAMS |
|---|---|---|---|---|---|
| 750 | 597.88 | 595.05 | 579.17 | 633.6 | 2,405.70 |
| 751 | 158.77 | 150.91 | 153.02 | 150.84 | 613.54 |
| 752 | 264.56 | 255.26 | 260.91 | 540.37 | 1,321.10 |
| 753 | 10.9 | 10.94 | 12.16 | 12.84 | 46.84 |
| 754 | 186.4 | 193 | 181.92 | 184.35 | 745.67 |
| 755 | 29.94 | 27.62 | 23.75 | 26.77 | 108.08 |
| 756 | 88.92 | 91.61 | 125.78 | 71.49 | 377.8 |
| 757 | 165.62 | 160.55 | 146.74 | 144.38 | 617.29 |
| 758 | 27.44 | 28.14 | 25.64 | 25.79 | 107.01 |
| 759 | 76.95 | 72.05 | 67.65 | 63.04 | 279.69 |
| 760 | 428.62 | 444.9 | 400.04 | 417.73 | 1,691.29 |
| 761 | 404.89 | 400.18 | 398.79 | 403 | 1,606.86 |
| 762 | 139.27 | 135.93 | 141.8 | 147.72 | 564.72 |

DRUG CODE:9801DRUG NAME:FENTANYL BASE
STATE:TEXAS

| ZIP CODE | QUARTER 1 | QUARTER 2 | QUARTER 3 | QUARTER 4 | TOTAL GRAMS |
|---|---|---|---|---|---|
| 763 | 105.77 | 108.5 | 101.84 | 98.89 | 415 |
| 764 | 57.68 | 59.43 | 58.95 | 56.59 | 232.65 |
| 765 | 106 | 111.86 | 99.93 | 99.72 | 417.51 |
| 766 | 30.52 | 28.59 | 27.81 | 26.58 | 113.5 |
| 767 | 59.54 | 54.09 | 54.27 | 49.43 | 217.33 |
| 768 | 40.98 | 41.61 | 44.94 | 44.25 | 171.78 |
| 769 | 32.62 | 28.96 | 35.18 | 32.49 | 129.25 |
| 770 | 427.11 | 407.94 | 416.51 | 634.81 | 1,886.37 |
| 772 | 0.78 | 0.65 | 0.63 | 0.57 | 2.63 |
| 773 | 281.63 | 271.58 | 279.4 | 257.43 | 1,090.04 |
| 774 | 227.96 | 214.84 | 200.89 | 204.86 | 848.55 |

| | | | | | |
|---|---|---|---|---|---|
| 775 | 265.89 | 269.72 | 270.3 | 254.92 | 1,060.83 |
| 776 | 80.39 | 77.32 | 75.7 | 69.55 | 302.96 |
| 777 | 34.18 | 32.28 | 30.41 | 32 | 128.87 |
| 778 | 79.79 | 72.69 | 73.07 | 70.62 | 296.17 |
| 779 | 64.45 | 64.51 | 61.11 | 63.24 | 253.31 |
| 780 | 92.12 | 92.97 | 86.1 | 82.64 | 353.83 |
| 781 | 104.11 | 112.02 | 101.05 | 90.59 | 407.77 |
| 782 | 546.58 | 509.5 | 509.9 | 464.54 | 2,030.52 |
| 783 | 58.08 | 56.53 | 56.62 | 50.87 | 222.1 |
| 784 | 172.91 | 158.04 | 156.44 | 114.26 | 601.65 |
| 785 | 153.8 | 154.39 | 156.04 | 148.79 | 613.02 |
| 786 | 318.98 | 312.39 | 295.82 | 276.42 | 1,203.61 |
| 787 | 284.97 | 315.27 | 303.29 | 282.23 | 1,185.76 |
| 788 | 24.49 | 24.18 | 28.19 | 23.89 | 100.75 |
| 789 | 14.89 | 13.72 | 12.44 | 12.63 | 53.68 |
| 790 | 66.78 | 66.91 | 64 | 67.54 | 265.23 |
| 791 | 117.79 | 118.2 | 118.11 | 113.12 | 467.22 |
| 792 | 11.16 | 13.35 | 16.24 | 14.02 | 54.77 |
| 793 | 42.01 | 43.35 | 40.05 | 39.86 | 165.27 |
| 794 | 121.07 | 113.12 | 114.83 | 197.93 | 546.95 |
| 795 | 39.3 | 40.37 | 39.1 | 36.98 | 155.75 |
| 796 | 74.47 | 74.62 | 71.37 | 69.55 | 290.01 |
| 797 | 69.5 | 65.29 | 58.15 | 59.08 | 252.02 |
| 798 | 4.26 | 5.09 | 4.32 | 4.2 | 17.87 |
| 799 | 153.98 | 143.55 | 146.43 | 146.23 | 590.19 |
| TOTAL | 6,946.70 | 6,843.57 | 6,726.80 | 7,113.24 | 27,630.31 |

STATE: UTAH

| ZIP CODE | QUARTER 1 | QUARTER 2 | QUARTER 3 | QUARTER 4 | TOTAL GRAMS |
|---|---|---|---|---|---|
| 840 | 332.34 | 332.71 | 350.46 | 333.11 | 1,348.62 |
| 841 | 266.75 | 268.26 | 256.49 | 244.19 | 1,035.69 |
| 843 | 51.88 | 53.68 | 61.07 | 45.62 | 212.25 |
| 844 | 80.33 | 76.68 | 73.8 | 66.74 | 297.55 |
| 845 | 36.94 | 38.48 | 37.47 | 36.72 | 149.61 |
| 846 | 92.64 | 94.18 | 93.19 | 87.94 | 367.95 |
| 847 | 131.04 | 128.74 | 123.13 | 123.87 | 506.78 |
| TOTAL | 991.92 | 992.73 | 995.61 | 938.19 | 3,918.45 |

STATE: VERMONT

| ZIP CODE | QUARTER 1 | QUARTER 2 | QUARTER 3 | QUARTER 4 | TOTAL GRAMS |
|---|---|---|---|---|---|
| 50 | 15.96 | 14.67 | 13.68 | 13.68 | 57.99 |
| 51 | 11.86 | 9.99 | 7.48 | 7.62 | 36.95 |
| 52 | 15.1 | 15.65 | 14.69 | 13.63 | 59.07 |
| 53 | 9.92 | 7.26 | 5.65 | 5.03 | 27.86 |
| 54 | 115.19 | 103.6 | 93.99 | 87.02 | 399.8 |
| 56 | 36.96 | 32.27 | 30.33 | 28.74 | 128.3 |
| 57 | 44.79 | 38.86 | 40.23 | 39.98 | 163.86 |
| 58 | 24.15 | 22.91 | 22.94 | 21.65 | 91.65 |
| TOTAL | 273.93 | 245.21 | 228.99 | 217.35 | 965.48 |

STATE: VIRGIN ISLANDS

| ZIP CODE | QUARTER 1 | QUARTER 2 | QUARTER 3 | QUARTER 4 | TOTAL GRAMS |
|---|---|---|---|---|---|
| 8 | 7.13 | 6.61 | 7.16 | 5.8 | 26.7 |
| TOTAL | 7.13 | 6.61 | 7.16 | 5.8 | 26.7 |

STATE: VIRGINIA

| ZIP CODE | QUARTER 1 | QUARTER 2 | QUARTER 3 | QUARTER 4 | TOTAL GRAMS |
|---|---|---|---|---|---|
| 201 | 151.66 | 146.29 | 131.47 | 136.69 | 566.11 |
| 220 | 78.83 | 83.32 | 76.99 | 73.32 | 312.46 |

RETAIL DRUG DISTRIBUTION BY ZIP CODE WITHIN STATE BY GRAMS WT

STATE:VIRGINIA

| ZIP CODE | QUARTER 1 | QUARTER 2 | QUARTER 3 | QUARTER 4 | TOTAL GRAMS |
|---|---|---|---|---|---|
| 221 | 52.73 | 55.82 | 49.25 | 46.82 | 204.62 |
| 222 | 24.17 | 22.45 | 17.57 | 19.56 | 83.75 |
| 223 | 33.59 | 34.17 | 36.73 | 33.52 | 138.01 |
| 224 | 57.64 | 50.3 | 48.82 | 47.74 | 204.5 |
| 225 | 19 | 17.99 | 18 | 18.34 | 73.33 |
| 226 | 78.59 | 77.17 | 72.59 | 65.36 | 293.71 |
| 227 | 13.93 | 14.51 | 18.34 | 16.8 | 63.58 |
| 228 | 54.46 | 54.01 | 52.28 | 49.23 | 209.98 |
| 229 | 89.95 | 86.93 | 81.29 | 79.81 | 337.98 |
| 230 | 77.53 | 77.49 | 75.15 | 70.77 | 300.94 |
| 231 | 120.1 | 110.11 | 109.6 | 96.96 | 436.77 |
| 232 | 210.02 | 201.39 | 194.93 | 193.6 | 799.94 |
| 233 | 88.81 | 81.73 | 76.1 | 72.58 | 319.22 |
| 234 | 183.4 | 182.14 | 171.2 | 168.45 | 705.19 |
| 235 | 71.51 | 70.93 | 63.4 | 58.67 | 264.51 |
| 236 | 116.75 | 107.2 | 110.52 | 92.52 | 426.99 |
| 237 | 54.72 | 53.8 | 52.99 | 51.23 | 212.74 |
| 238 | 87.16 | 83.42 | 77.64 | 73.99 | 322.21 |
| 239 | 23.7 | 22.48 | 22.26 | 22.53 | 90.97 |
| 240 | 125.04 | 121.35 | 116.19 | 107.75 | 470.33 |
| 241 | 114.64 | 111.4 | 100.9 | 94.42 | 421.36 |
| 242 | 108.99 | 106.57 | 101.2 | 101.45 | 418.21 |
| 243 | 54.03 | 49.86 | 47.38 | 46.19 | 197.46 |
| 244 | 56.71 | 56.42 | 51.13 | 48.84 | 213.1 |
| 245 | 111.29 | 103.35 | 97.12 | 87.39 | 399.15 |
| 246 | 26.55 | 29.75 | 26.08 | 25.69 | 108.07 |
| TOTAL | 2,285.50 | 2,212.35 | 2,097.12 | 2,000.22 | 8,595.19 |

STATE: WASHINGTON

| ZIP CODE | QUARTER 1 | QUARTER 2 | QUARTER 3 | QUARTER 4 | TOTAL GRAMS |
|---|---|---|---|---|---|
| 980 | 254.16 | 262.09 | 254.24 | 224.76 | 995.25 |
| 981 | 227.7 | 226.13 | 212.94 | 197.3 | 864.07 |
| 982 | 206.6 | 198.27 | 194.97 | 182.6 | 782.44 |
| 983 | 176.26 | 172.86 | 165.23 | 162.9 | 677.25 |
| 984 | 109.45 | 106.69 | 100.89 | 106.77 | 423.8 |
| 985 | 109.79 | 110.84 | 102.29 | 89.54 | 412.46 |
| 986 | 206.84 | 198.45 | 188.46 | 188.24 | 781.99 |
| 988 | 41.11 | 43.61 | 42.23 | 41.17 | 168.12 |
| 989 | 83.99 | 80.27 | 84.63 | 79.4 | 328.29 |
| 990 | 26.27 | 27.14 | 27.67 | 24.59 | 105.67 |
| 991 | 42.58 | 37 | 36.03 | 37.71 | 153.32 |
| 992 | 212.56 | 211.23 | 202.98 | 185.86 | 812.63 |
| 993 | 192.44 | 204.8 | 183.94 | 178.81 | 759.99 |
| 994 | 23.8 | 23.49 | 21.64 | 22.12 | 91.05 |
| TOTAL | 1,913.55 | 1,902.87 | 1,818.14 | 1,721.77 | 7,356.33 |

STATE: WEST VIRGINIA

| ZIP CODE | QUARTER 1 | QUARTER 2 | QUARTER 3 | QUARTER 4 | TOTAL GRAMS |
|---|---|---|---|---|---|
| 247 | 15.68 | 13.59 | 13.36 | 14.79 | 57.42 |
| 248 | 7.72 | 6.11 | 5.68 | 5.11 | 24.62 |
| 249 | 15 | 14.06 | 11.52 | 12.79 | 53.37 |
| 250 | 18.54 | 17.75 | 15.68 | 15.86 | 67.83 |
| 251 | 20.14 | 17.16 | 17.87 | 14.14 | 69.31 |
| 252 | 19.72 | 16.54 | 16.97 | 14.53 | 67.76 |
| 253 | 82.57 | 85.2 | 76.78 | 72.89 | 317.44 |
| 254 | 83.99 | 79.46 | 78.86 | 75.31 | 317.62 |
| 255 | 38.24 | 38.18 | 32.98 | 32.84 | 142.24 |
| 256 | 12.86 | 12.19 | 12.3 | 10.37 | 47.72 |

| | | | | | |
|---|---|---|---|---|---|
| 257 | 63.82 | 61.61 | 61.04 | 56.45 | 242.92 |
| 258 | 29.27 | 26.42 | 24.17 | 22.4 | 102.26 |
| 259 | 13.12 | 12.63 | 13.49 | 12.49 | 51.73 |
| 260 | 54.65 | 54.01 | 47.96 | 43.96 | 200.58 |
| 261 | 38.8 | 36.09 | 28.89 | 28.57 | 132.35 |
| 262 | 22.81 | 22.71 | 21.58 | 21.54 | 88.64 |
| 263 | 46.02 | 43.07 | 37.97 | 36.17 | 163.23 |
| 264 | 10.29 | 11.35 | 12.01 | 12.19 | 45.84 |
| 265 | 72.34 | 80.14 | 72.89 | 66.94 | 292.31 |
| 266 | 13.29 | 11.34 | 11.78 | 9.85 | 46.26 |

DRUG CODE:9801DRUG NAME:FENTANYL BASE
STATE:WEST VIRGINIA

| ZIP CODE | QUARTER 1 | QUARTER 2 | QUARTER 3 | QUARTER 4 | TOTAL GRAMS |
|---|---|---|---|---|---|
| 267 | 27.68 | 28.24 | 25.33 | 23.96 | 105.21 |
| 268 | 8.02 | 9.16 | 9.26 | 10.4 | 36.84 |
| TOTAL | 714.57 | 697.01 | 648.37 | 613.55 | 2,673.50 |

STATE: WISCONSIN

| ZIP CODE | QUARTER 1 | QUARTER 2 | QUARTER 3 | QUARTER 4 | TOTAL GRAMS |
|---|---|---|---|---|---|
| 530 | 197.13 | 194.37 | 178.06 | 170.67 | 740.23 |
| 531 | 212.49 | 224.78 | 193.08 | 169.45 | 799.8 |
| 532 | 312.7 | 315.85 | 287.14 | 275.81 | 1,191.50 |
| 534 | 36.21 | 33.44 | 28.28 | 30.27 | 128.2 |
| 535 | 205.54 | 195.95 | 180.74 | 167.15 | 749.38 |
| 537 | 128.8 | 134.93 | 124.28 | 124.7 | 512.71 |
| 538 | 23.96 | 24.09 | 22.22 | 22.29 | 92.56 |
| 539 | 88.54 | 80.9 | 80.2 | 75.25 | 324.89 |
| 540 | 32.91 | 30.59 | 30.15 | 24.36 | 118.01 |
| 541 | 49.64 | 48.05 | 50.02 | 41.38 | 189.09 |
| 542 | 38.08 | 31.47 | 28.24 | 25.09 | 122.88 |
| 543 | 68.98 | 60.61 | 63.46 | 64.1 | 257.15 |
| 544 | 99.33 | 98.27 | 92.16 | 88.17 | 377.93 |
| 545 | 24.14 | 23.52 | 25.21 | 20.31 | 93.18 |
| 546 | 111.25 | 110.26 | 100.58 | 103.25 | 425.34 |
| 547 | 99.71 | 94.45 | 89.02 | 89.39 | 372.57 |
| 548 | 54.56 | 51.56 | 51.39 | 48.99 | 206.5 |
| 549 | 246.26 | 232.84 | 217.24 | 207.55 | 903.89 |
| TOTAL | 2,030.23 | 1,985.93 | 1,841.47 | 1,748.18 | 7,605.81 |

STATE: WYOMING

| ZIP CODE | QUARTER 1 | QUARTER 2 | QUARTER 3 | QUARTER 4 | TOTAL GRAMS |
|---|---|---|---|---|---|
| 820 | 40.08 | 35.05 | 33.95 | 32.96 | 142.04 |
| 822 | 7.81 | 8.22 | 9.42 | 7.51 | 32.96 |
| 823 | 2.23 | 1.95 | 1.31 | 1.89 | 7.38 |
| 824 | 20.03 | 18.58 | 21.09 | 18.6 | 78.3 |
| 825 | 12.42 | 9.89 | 10.67 | 8.04 | 41.02 |
| 826 | 36.49 | 37.24 | 38.16 | 36.4 | 148.29 |
| 827 | 7.83 | 6.06 | 6.87 | 6.3 | 27.06 |
| 828 | 17.12 | 15.61 | 15.02 | 12.22 | 59.97 |
| 829 | 32.33 | 36.45 | 34.07 | 38.26 | 141.11 |
| 830 | 7.33 | 7.32 | 6.57 | 6.1 | 27.32 |
| 831 | 6.25 | 5.88 | 5.63 | 5.44 | 23.2 |
| TOTAL | 189.92 | 182.25 | 182.76 | 173.72 | 728.65 |

RETAIL DRUG DISTRIBUTION BY STATE WITHIN DRUG CODE BY GRAMS WT

DRUG CODE:1100 DRUG NAME:AMPHETAMINE

| STATE | QUARTER 1 | QUARTER 2 | QUARTER 3 | QUARTER 4 | TOTAL GRAMS |
|---|---|---|---|---|---|
| ALABAMA | 123,776.27 | 126,406.02 | 128,969.59 | 128,243.42 | 507,395.30 |
| ALASKA | 7,498.96 | 8,126.29 | 7,972.19 | 7,730.06 | 31,327.50 |
| AMERICAN SAMOA | 2.53 | 0 | 1.5 | 2.5 | 6.53 |
| ARIZONA | 73,164.39 | 75,825.33 | 76,114.49 | 78,789.71 | 303,893.92 |
| ARKANSAS | 47,407.36 | 47,995.43 | 47,742.92 | 49,542.63 | 192,688.34 |
| CALIFORNIA | 284,739.77 | 291,072.23 | 290,771.79 | 297,589.56 | 1,164,173.35 |
| COLORADO | 70,271.53 | 74,179.68 | 76,089.96 | 75,929.40 | 296,470.57 |
| CONNECTICUT | 48,508.94 | 49,896.11 | 50,892.11 | 51,304.76 | 200,601.92 |
| DELAWARE | 20,865.58 | 21,677.27 | 21,591.58 | 21,831.13 | 85,965.56 |
| DISTRICT OF COLUMBIA | 10,853.15 | 11,400.47 | 11,424.76 | 11,288.51 | 44,966.89 |
| FLORIDA | 256,942.77 | 274,657.19 | 278,083.32 | 285,591.58 | 1,095,274.86 |
| GEORGIA | 170,945.95 | 173,712.01 | 179,243.25 | 182,066.73 | 705,967.94 |
| GUAM | 397.32 | 392.44 | 490.58 | 474.36 | 1,754.70 |
| HAWAII | 9,092.13 | 8,616.45 | 9,770.49 | 8,971.32 | 36,450.39 |
| IDAHO | 22,347.51 | 23,748.55 | 23,449.30 | 23,956.38 | 93,501.74 |
| ILLINOIS | 171,041.74 | 179,991.09 | 181,512.59 | 185,495.20 | 718,040.62 |
| INDIANA | 118,326.80 | 121,457.73 | 122,249.48 | 125,424.40 | 487,458.41 |
| IOWA | 45,849.17 | 46,586.03 | 47,081.71 | 47,965.59 | 187,482.50 |
| KANSAS | 55,063.28 | 56,381.18 | 57,484.05 | 58,290.58 | 227,219.09 |
| KENTUCKY | 62,663.15 | 63,766.35 | 64,086.92 | 66,002.22 | 256,518.64 |
| LOUISIANA | 138,972.29 | 140,586.31 | 145,430.20 | 149,001.92 | 573,990.72 |
| MAINE | 20,824.87 | 21,437.91 | 21,541.96 | 21,626.84 | 85,431.58 |
| MARYLAND | 83,759.67 | 87,417.11 | 88,188.51 | 88,333.56 | 347,698.85 |
| MASSACHUSETTS | 165,957.48 | 172,463.01 | 173,880.30 | 174,311.24 | 686,612.03 |
| MICHIGAN | 227,018 | 231,940.02 | 220,976.13 | 237,790.89 | 917,725.04 |
| MINNESOTA | 103,215.99 | 105,588.22 | 106,623.93 | 106,720.83 | 422,148.97 |
| MISSISSIPPI | 58,700.18 | 58,607.44 | 60,763.94 | 60,887.35 | 238,958.91 |
| MISSOURI | 125,346.36 | 128,811.23 | 130,993.61 | 129,755.73 | 514,906.93 |
| MONTANA | 12,399.32 | 13,299.41 | 13,023.48 | 13,294.54 | 52,016.75 |
| NEBRASKA | 23,646.48 | 23,995.43 | 24,537.07 | 24,728.54 | 96,907.52 |
| NEVADA | 22,720.48 | 24,322.91 | 24,102.19 | 24,999.84 | 96,145.42 |
| NEW HAMPSHIRE | 34,534.10 | 36,016.07 | 36,633.03 | 36,766.20 | 143,949.40 |
| NEW JERSEY | 94,645.37 | 99,268.09 | 99,311.71 | 101,893.12 | 395,118.29 |
| NEW MEXICO | 19,067.11 | 19,665.60 | 18,906.15 | 19,477.13 | 77,115.99 |
| NEW YORK | 219,458.72 | 227,837.02 | 226,744.81 | 228,508.03 | 902,548.58 |
| NORTH CAROLINA | 183,241.45 | 191,132.39 | 193,254.30 | 195,934.86 | 763,563 |
| NORTH DAKOTA | 9,172.37 | 9,016.88 | 8,962.89 | 8,095.58 | 35,247.72 |
| OHIO | 202,585.12 | 198,292.10 | 201,364.85 | 209,609.30 | 811,851.37 |
| OKLAHOMA | 49,332.12 | 50,443.07 | 51,668.80 | 51,683.09 | 203,127.08 |
| OREGON | 54,476.81 | 58,498.74 | 57,264.64 | 56,897.80 | 227,137.99 |

DRUG CODE:1100 DRUG NAME:AMPHETAMINE

| STATE | QUARTER 1 | QUARTER 2 | QUARTER 3 | QUARTER 4 | TOTAL GRAMS |
|---|---|---|---|---|---|
| PENNSYLVANIA | 212,621.10 | 210,761.66 | 207,268.04 | 208,570.02 | 839,220.82 |
| PUERTO RICO | 7,872.69 | 7,884.25 | 8,329.42 | 8,484.80 | 32,571.16 |
| RHODE ISLAND | 33,625.59 | 34,957.68 | 35,221.11 | 35,451.97 | 139,256.35 |
| SOUTH CAROLINA | 127,072.51 | 131,280.66 | 134,034.83 | 133,023.91 | 525,411.91 |
| SOUTH DAKOTA | 17,108.12 | 14,413.60 | 15,975.97 | 15,362.99 | 62,860.68 |
| TENNESSEE | 119,992.33 | 124,072.24 | 127,521.19 | 127,696.95 | 499,282.71 |
| TEXAS | 366,574.35 | 384,624.58 | 388,533.08 | 394,450.93 | 1,534,182.94 |
| UTAH | 71,351.72 | 74,731.36 | 76,694.73 | 77,410.36 | 300,188.17 |
| VERMONT | 9,670.36 | 9,855.66 | 9,634.08 | 9,884.84 | 39,044.94 |
| VIRGIN ISLANDS | 308.21 | 311.32 | 296.41 | 335.32 | 1,251.26 |
| VIRGINIA | 166,902.61 | 174,212.71 | 174,476.62 | 175,604.05 | 691,195.99 |

| | | | | | |
|---|---|---|---|---|---|
| WASHINGTON | 101,510.95 | 107,161.60 | 105,543.97 | 105,761.76 | 419,978.28 |
| WEST VIRGINIA | 24,410.65 | 24,066.04 | 24,304.79 | 23,951.03 | 96,732.51 |
| WISCONSIN | 135,381.40 | 135,219.06 | 133,720.13 | 132,264.62 | 536,585.21 |
| WYOMING | 6,295.34 | 6,374.93 | 6,393.13 | 6,316.61 | 25,380.01 |
| TOTAL | 4,849,530.52 | 4,994,454.16 | 5,027,142.58 | 5,101,376.59 | 19,972,503.85 |

DRUG CODE:1105B DRUG NAME:DL-METHAMPHETAMINE RACEMIC BASE

| STATE | QUARTER 1 | QUARTER 2 | QUARTER 3 | QUARTER 4 | TOTAL GRAMS |
|---|---|---|---|---|---|
| ALABAMA | 10.45 | 8.84 | 5.63 | 8.04 | 32.96 |
| ALASKA | 0 | 0.8 | 0 | 0 | 0.8 |
| ARIZONA | 20.49 | 20.09 | 17.68 | 25.72 | 83.98 |
| ARKANSAS | 1.21 | 0.8 | 2.81 | 1.21 | 6.03 |
| CALIFORNIA | 88.01 | 95.24 | 78.36 | 90.82 | 352.43 |
| COLORADO | 6.43 | 1.61 | 0.8 | 1.61 | 10.45 |
| CONNECTICUT | 11.65 | 8.84 | 6.43 | 9.24 | 36.16 |
| DISTRICT OF COLUMBIA | 3.62 | 3.62 | 2.81 | 1.21 | 11.26 |
| FLORIDA | 13.26 | 7.23 | 6.83 | 8.44 | 35.76 |
| GEORGIA | 6.43 | 6.43 | 4.82 | 6.83 | 24.51 |
| HAWAII | 3.62 | 3.21 | 2.81 | 2.81 | 12.45 |
| IDAHO | 4.42 | 5.63 | 4.02 | 4.02 | 18.09 |
| ILLINOIS | 3.21 | 4.02 | 4.82 | 2.81 | 14.86 |
| INDIANA | 4.02 | 4.02 | 6.43 | 5.63 | 20.1 |
| IOWA | 8.04 | 9.64 | 6.83 | 8.84 | 33.35 |
| KANSAS | 1.61 | 2.01 | 1.61 | 0.8 | 6.03 |
| KENTUCKY | 4.02 | 1.61 | 2.41 | 2.41 | 10.45 |
| LOUISIANA | 3.62 | 3.62 | 1.61 | 2.41 | 11.26 |
| MAINE | 2.41 | 0 | 0.4 | 0 | 2.81 |
| MARYLAND | 2.81 | 2.81 | 1.61 | 4.82 | 12.05 |
| MASSACHUSETTS | 6.03 | 4.02 | 4.82 | 2.81 | 17.68 |
| MICHIGAN | 0.8 | 0 | 4.02 | 0 | 4.82 |
| MINNESOTA | 39.78 | 23.71 | 40.99 | 37.77 | 142.25 |
| MISSISSIPPI | 0.4 | 1.61 | 0.4 | 0.4 | 2.81 |
| MISSOURI | 28.53 | 33.35 | 15.67 | 19.69 | 97.24 |
| NEBRASKA | 4.02 | 4.02 | 3.21 | 2.01 | 13.26 |
| NEVADA | 5.22 | 2.81 | 1.61 | 4.42 | 14.06 |

DRUG CODE:1105B DRUG NAME:DL-METHAMPHETAMINE RACEMIC BASE

| STATE | QUARTER 1 | QUARTER 2 | QUARTER 3 | QUARTER 4 | TOTAL GRAMS |
|---|---|---|---|---|---|
| NEW HAMPSHIRE | 0.4 | 0.8 | 0.8 | 0.4 | 2.4 |
| NEW JERSEY | 13.66 | 11.25 | 2.81 | 5.63 | 33.35 |
| NEW MEXICO | 15.67 | 10.05 | 11.25 | 14.87 | 51.84 |
| NEW YORK | 13.66 | 15.27 | 15.67 | 18.08 | 62.68 |
| NORTH CAROLINA | 14.47 | 14.06 | 13.66 | 11.25 | 53.44 |
| NORTH DAKOTA | 1.21 | 1.21 | 1.21 | 1.21 | 4.84 |
| OHIO | 10.45 | 10.05 | 8.04 | 8.84 | 37.38 |
| OKLAHOMA | 7.23 | 8.44 | 8.04 | 5.22 | 28.93 |
| OREGON | 18.89 | 18.08 | 15.67 | 14.06 | 66.7 |
| PENNSYLVANIA | 15.67 | 14.06 | 14.87 | 10.05 | 54.65 |
| RHODE ISLAND | 2.81 | 0.8 | 0.8 | 1.21 | 5.62 |
| SOUTH CAROLINA | 3.62 | 2.81 | 4.02 | 2.81 | 13.26 |
| TENNESSEE | 2.01 | 0.8 | 0 | 0 | 2.81 |
| TEXAS | 52.24 | 47.82 | 37.77 | 35.36 | 173.19 |
| UTAH | 2.01 | 0 | 0 | 0 | 2.01 |
| VIRGINIA | 0 | 1.61 | 1.21 | 0 | 2.82 |
| WASHINGTON | 3.21 | 2.41 | 2.81 | 0.8 | 9.23 |
| WEST VIRGINIA | 2.01 | 2.81 | 2.81 | 2.01 | 9.64 |

| | | | | | |
|---|---|---|---|---|---|
| WISCONSIN | 18.49 | 18.49 | 20.09 | 19.29 | 76.36 |
| WYOMING | 0.4 | 0 | 0 | 0 | 0.4 |
| TOTAL | 482.22 | 440.41 | 390.97 | 405.86 | 1,719.46 |

DRUG CODE:1105D DRUG NAME:D-METHAMPHETAMINE

| STATE | QUARTER 1 | QUARTER 2 | QUARTER 3 | QUARTER 4 | TOTAL GRAMS |
|---|---|---|---|---|---|
| ALABAMA | 23.7 | 24.1 | 20.09 | 18.88 | 86.77 |
| ALASKA | 4.42 | 4.02 | 1.61 | 2.41 | 12.46 |
| ARIZONA | 17.67 | 16.47 | 14.46 | 18.48 | 67.08 |
| ARKANSAS | 3.21 | 4.02 | 2.81 | 4.02 | 14.06 |
| CALIFORNIA | 251.06 | 255.16 | 297.66 | 282.4 | 1,086.28 |
| COLORADO | 32.14 | 49.81 | 47 | 40.97 | 169.92 |
| CONNECTICUT | 12.45 | 18.08 | 17.27 | 18.08 | 65.88 |
| DELAWARE | 4.42 | 5.62 | 4.42 | 6.03 | 20.49 |
| DISTRICT OF COLUMBIA | 2.01 | 0 | 0 | 1.61 | 3.62 |
| FLORIDA | 16.87 | 25.31 | 21.29 | 21.29 | 84.76 |
| GEORGIA | 25.71 | 30.53 | 36.15 | 27.72 | 120.11 |
| HAWAII | 2.41 | 3.21 | 4.02 | 5.22 | 14.86 |
| IDAHO | 6.43 | 6.43 | 7.23 | 4.42 | 24.51 |
| ILLINOIS | 41.78 | 57.44 | 56.24 | 52.22 | 207.68 |
| INDIANA | 17.67 | 16.87 | 18.08 | 19.28 | 71.9 |
| IOWA | 11.65 | 14.46 | 19.28 | 13.66 | 59.05 |
| KANSAS | 12.05 | 13.66 | 15.26 | 10.44 | 51.41 |
| KENTUCKY | 5.22 | 6.03 | 4.82 | 4.82 | 20.89 |
| LOUISIANA | 8.84 | 29.73 | 14.06 | 14.86 | 67.49 |
| MAINE | 2.41 | 1.61 | 2.41 | 2.01 | 8.44 |
| MARYLAND | 51.42 | 44.59 | 42.98 | 46.6 | 185.59 |
| MASSACHUSETTS | 34.14 | 41.78 | 38.16 | 42.98 | 157.06 |

DRUG CODE:1105D DRUG NAME:D-METHAMPHETAMINE

| STATE | QUARTER 1 | QUARTER 2 | QUARTER 3 | QUARTER 4 | TOTAL GRAMS |
|---|---|---|---|---|---|
| MICHIGAN | 29.32 | 27.32 | 27.72 | 28.92 | 113.28 |
| MINNESOTA | 73.51 | 69.9 | 72.71 | 72.71 | 288.83 |
| MISSISSIPPI | 11.25 | 9.64 | 9.64 | 8.84 | 39.37 |
| MISSOURI | 46.6 | 52.22 | 42.98 | 42.18 | 183.98 |
| MONTANA | 4.02 | 3.62 | 1.61 | 0.4 | 9.65 |
| NEBRASKA | 8.84 | 7.23 | 6.83 | 5.22 | 28.12 |
| NEVADA | 18.08 | 22.5 | 24.5 | 20.09 | 85.17 |
| NEW HAMPSHIRE | 6.03 | 5.22 | 7.23 | 7.71 | 26.19 |
| NEW JERSEY | 32.94 | 37.76 | 27.72 | 32.54 | 130.96 |
| NEW MEXICO | 14.86 | 18.88 | 12.05 | 18.08 | 63.87 |
| NEW YORK | 79.62 | 77.93 | 63.87 | 75.92 | 297.34 |
| NORTH CAROLINA | 49.41 | 44.99 | 53.43 | 45.79 | 193.62 |
| NORTH DAKOTA | 1.61 | 0.8 | 1.21 | 3.62 | 7.24 |
| OHIO | 97.21 | 111.67 | 55.03 | 52.22 | 316.13 |
| OKLAHOMA | 6.43 | 8.03 | 9.24 | 12.05 | 35.75 |
| OREGON | 47.8 | 41.78 | 47 | 37.76 | 174.34 |
| PENNSYLVANIA | 107.74 | 140.6 | 120.91 | 118.1 | 487.35 |
| RHODE ISLAND | 2.01 | 2.01 | 2.41 | 2.81 | 9.24 |
| SOUTH CAROLINA | 7.23 | 12.45 | 10.85 | 18.88 | 49.41 |
| SOUTH DAKOTA | 14.06 | 8.84 | 14.06 | 12.05 | 49.01 |
| TENNESSEE | 14.46 | 10.44 | 10.44 | 10.04 | 45.38 |
| TEXAS | 67.49 | 83.15 | 80.74 | 75.92 | 307.3 |
| UTAH | 36.96 | 36.55 | 29.32 | 26.91 | 129.74 |
| VERMONT | 5.22 | 0.4 | 2.41 | 1.61 | 9.64 |
| VIRGINIA | 51.02 | 57.04 | 54.63 | 55.03 | 217.72 |

| | | | | | |
|---|---|---|---|---|---|
| WASHINGTON | 86.37 | 100.02 | 90.38 | 90.38 | 367.15 |
| WEST VIRGINIA | 0.4 | 0.4 | 1.21 | 0 | 2.01 |
| WISCONSIN | 36.15 | 49.01 | 44.19 | 54.63 | 183.98 |
| WYOMING | 6.03 | 6.03 | 5.62 | 6.83 | 24.51 |
| TOTAL | 1,550.35 | 1,715.36 | 1,615.24 | 1,595.64 | 6,476.59 |

DRUG CODE:1205 DRUG NAME:LISDEXAMFETAMINE

| STATE | QUARTER 1 | QUARTER 2 | QUARTER 3 | QUARTER 4 | TOTAL GRAMS |
|---|---|---|---|---|---|
| ALABAMA | 72,319.15 | 68,945.94 | 70,714.35 | 73,063.17 | 285,042.61 |
| ALASKA | 2,511.84 | 2,385.24 | 2,383.51 | 2,503.75 | 9,784.34 |
| ARIZONA | 33,775.49 | 33,347.70 | 33,499.74 | 35,239.24 | 135,862.17 |
| ARKANSAS | 31,502.98 | 29,453.62 | 29,783.13 | 32,120.97 | 122,860.70 |
| CALIFORNIA | 105,345.43 | 107,943.99 | 107,101.70 | 113,293.15 | 433,684.27 |
| COLORADO | 26,577.57 | 26,357.89 | 26,731.34 | 28,122.25 | 107,789.05 |
| CONNECTICUT | 34,506.21 | 35,161.78 | 34,667.50 | 36,659.63 | 140,995.12 |
| DELAWARE | 14,507.42 | 14,167.50 | 13,682.47 | 14,865.26 | 57,222.65 |
| DISTRICT OF COLUMBIA | 4,588.38 | 4,690.70 | 4,971.08 | 5,063.58 | 19,313.74 |
| FLORIDA | 139,982.29 | 136,887.72 | 133,669.38 | 143,550.90 | 554,090.29 |
| GEORGIA | 106,603.95 | 101,857.17 | 104,129.11 | 107,511.57 | 420,101.80 |
| GUAM | 21.97 | 18.5 | 8.67 | 43.94 | 93.08 |
| HAWAII | 1,203.60 | 1,253.90 | 1,493.23 | 1,237.71 | 5,188.44 |

DRUG CODE:1205 DRUG NAME:LISDEXAMFETAMINE

| STATE | QUARTER 1 | QUARTER 2 | QUARTER 3 | QUARTER 4 | TOTAL GRAMS |
|---|---|---|---|---|---|
| IDAHO | 8,184.16 | 8,432.74 | 8,052.93 | 8,677.86 | 33,347.69 |
| ILLINOIS | 70,605.09 | 70,553.06 | 69,893.45 | 73,319.84 | 284,371.44 |
| INDIANA | 84,093.32 | 81,117.26 | 82,216.23 | 85,830.51 | 333,257.32 |
| IOWA | 34,500.43 | 34,007.31 | 33,303.76 | 35,913.31 | 137,724.81 |
| KANSAS | 29,603.92 | 29,792.38 | 30,309.78 | 32,822.21 | 122,528.29 |
| KENTUCKY | 42,174.08 | 41,189.02 | 42,255.04 | 43,945.43 | 169,563.57 |
| LOUISIANA | 99,759.25 | 93,282.79 | 94,299.67 | 98,697.86 | 386,039.57 |
| MAINE | 16,243.45 | 16,978.22 | 16,361.39 | 16,767.21 | 66,350.27 |
| MARYLAND | 52,459.46 | 53,875.34 | 52,369.86 | 56,216.76 | 214,921.42 |
| MASSACHUSETTS | 32,510.03 | 33,678.95 | 32,719.88 | 35,516.73 | 134,425.59 |
| MICHIGAN | 58,318.15 | 58,512.39 | 56,142.76 | 60,224.15 | 233,197.45 |
| MINNESOTA | 30,564.73 | 31,510.50 | 31,421.47 | 33,878.97 | 127,375.67 |
| MISSISSIPPI | 38,810.17 | 35,672.82 | 37,355.09 | 40,342.71 | 152,180.79 |
| MISSOURI | 66,909.29 | 65,420.69 | 66,032.89 | 70,133.36 | 268,496.23 |
| MONTANA | 5,236.43 | 5,324.30 | 5,149.14 | 5,719.72 | 21,429.59 |
| NEBRASKA | 15,380.35 | 15,348.56 | 15,838.78 | 16,826.76 | 63,394.45 |
| NEVADA | 7,011.63 | 7,084.02 | 6,769.70 | 7,714.74 | 28,580.09 |
| NEW HAMPSHIRE | 13,578.41 | 13,128.65 | 12,843.07 | 13,667.44 | 53,217.57 |
| NEW JERSEY | 48,045.31 | 48,983.57 | 46,760.20 | 50,308.57 | 194,097.65 |
| NEW MEXICO | 7,054.55 | 6,810.02 | 6,532.53 | 6,838.34 | 27,235.44 |
| NEW YORK | 79,211.18 | 82,722.03 | 78,490.95 | 84,583.52 | 325,007.68 |
| NORTH CAROLINA | 98,890.46 | 99,690.18 | 96,090.08 | 102,123.13 | 396,793.85 |
| NORTH DAKOTA | 5,743.42 | 5,501.78 | 5,582.13 | 5,320.83 | 22,148.16 |
| OHIO | 115,095.21 | 113,163.63 | 111,496.56 | 116,771 | 456,526.40 |
| OKLAHOMA | 35,097.03 | 34,177.61 | 34,298.77 | 36,960.16 | 140,533.57 |
| OREGON | 13,858.21 | 14,690.68 | 14,432.84 | 15,194.20 | 58,175.93 |
| PENNSYLVANIA | 80,782.54 | 81,570.49 | 80,344.66 | 85,036.78 | 327,734.47 |
| PUERTO RICO | 1,790.38 | 1,644.12 | 1,628.51 | 1,971.32 | 7,034.33 |
| RHODE ISLAND | 7,147.63 | 7,312.39 | 7,317.01 | 7,614.73 | 29,391.76 |
| SOUTH CAROLINA | 60,156.51 | 58,384.63 | 58,153.97 | 61,875.78 | 238,570.89 |
| SOUTH DAKOTA | 11,927.94 | 11,679.35 | 11,753.93 | 11,928.52 | 47,289.74 |
| TENNESSEE | 72,133.01 | 70,193.48 | 71,372.80 | 74,259.84 | 287,959.13 |
| TEXAS | 274,712.54 | 265,778.01 | 263,029.14 | 283,004.81 | 1,086,524.50 |

| | | | | | |
|---|---|---|---|---|---|
| UTAH | 19,557.70 | 19,093.49 | 19,769.86 | 21,782.23 | 80,203.28 |
| VERMONT | 6,403.61 | 6,326.15 | 6,337.13 | 6,549.87 | 25,616.76 |
| VIRGIN ISLANDS | 86.72 | 80.93 | 67.06 | 60.7 | 295.41 |
| VIRGINIA | 66,898.19 | 67,298.30 | 65,503.35 | 69,474.93 | 269,174.77 |
| WASHINGTON | 31,048.59 | 31,889.73 | 31,202.37 | 33,222.83 | 127,363.52 |
| WEST VIRGINIA | 20,518.50 | 20,256.05 | 19,718.41 | 20,633.55 | 81,126.51 |
| WISCONSIN | 57,178.14 | 59,262.77 | 59,234.44 | 63,951.16 | 239,626.51 |
| WYOMING | 2,600.87 | 2,718.80 | 2,626.89 | 2,728.05 | 10,674.61 |
| TOTAL | 2,395,326.87 | 2,366,608.84 | 2,347,913.69 | 2,491,685.54 | 9,601,534.94 |

DRUG CODE:1724 DRUG NAME:METHYLPHENIDATE (DL;D;L;ISOMERS)

| STATE | QUARTER 1 | QUARTER 2 | QUARTER 3 | QUARTER 4 | TOTAL GRAMS |
|---|---|---|---|---|---|
| ALABAMA | 75,574.03 | 69,411.60 | 71,324.35 | 74,550.25 | 290,860.23 |

DRUG CODE:1724 DRUG NAME:METHYLPHENIDATE (DL;D;L;ISOMERS)

| STATE | QUARTER 1 | QUARTER 2 | QUARTER 3 | QUARTER 4 | TOTAL GRAMS |
|---|---|---|---|---|---|
| ALASKA | 8,550.69 | 8,493.39 | 7,563.16 | 8,211.35 | 32,818.59 |
| AMERICAN SAMOA | 1.3 | 1.73 | 9.08 | 2.59 | 14.7 |
| ARIZONA | 76,200.01 | 71,344.97 | 71,317.09 | 75,553.63 | 294,415.70 |
| ARKANSAS | 44,420.10 | 39,858.19 | 39,299.72 | 41,680.11 | 165,258.12 |
| CALIFORNIA | 285,417.44 | 278,577.11 | 262,107.26 | 281,028.87 | 1,107,130.68 |
| COLORADO | 62,859.68 | 60,437.31 | 59,711.56 | 62,015.55 | 245,024.10 |
| CONNECTICUT | 58,283.67 | 55,900.69 | 52,443.45 | 55,657.17 | 222,284.98 |
| DELAWARE | 20,134.19 | 19,051.66 | 17,874.70 | 19,489.29 | 76,549.84 |
| DISTRICT OF COLUMBIA | 8,389.78 | 8,002.33 | 8,428.18 | 8,027.01 | 32,847.30 |
| FLORIDA | 218,484.11 | 203,939.18 | 198,184.62 | 212,434.74 | 833,042.65 |
| GEORGIA | 175,833.72 | 158,188.98 | 162,892.57 | 167,967.33 | 664,882.60 |
| GUAM | 437.21 | 397.25 | 438.24 | 572.6 | 1,845.30 |
| HAWAII | 11,703.65 | 11,475.25 | 11,800.67 | 11,183.40 | 46,162.97 |
| IDAHO | 28,839.88 | 28,676.66 | 26,562.16 | 27,947.89 | 112,026.59 |
| ILLINOIS | 187,830.75 | 176,219.60 | 168,940.97 | 183,039.81 | 716,031.13 |
| INDIANA | 129,529.44 | 120,821.48 | 118,244.49 | 125,233.44 | 493,828.85 |
| IOWA | 87,095.53 | 82,422.54 | 79,973.49 | 84,760.34 | 334,251.90 |
| KANSAS | 52,961.60 | 49,967.11 | 49,660.58 | 52,802.36 | 205,391.65 |
| KENTUCKY | 80,526.85 | 74,039.66 | 74,992.29 | 76,689.14 | 306,247.94 |
| LOUISIANA | 93,378.61 | 80,445.61 | 82,831.03 | 89,213.67 | 345,868.92 |
| MAINE | 37,188.87 | 37,156.58 | 35,360.86 | 35,321.12 | 145,027.43 |
| MARYLAND | 104,636.26 | 101,568.86 | 98,275.46 | 102,525.06 | 407,005.64 |
| MASSACHUSETTS | 154,767.55 | 151,150.71 | 139,182.50 | 150,926.15 | 596,026.91 |
| MICHIGAN | 201,633.27 | 193,550.39 | 168,605.24 | 198,720.98 | 762,509.88 |
| MINNESOTA | 104,019.10 | 100,409.95 | 96,067.41 | 103,971.43 | 404,467.89 |
| MISSISSIPPI | 47,676.74 | 41,355.80 | 43,223.02 | 44,446.74 | 176,702.30 |
| MISSOURI | 134,958.64 | 129,411.89 | 126,801.70 | 136,688.77 | 527,861 |
| MONTANA | 18,547.58 | 17,556.11 | 16,476.37 | 18,112.03 | 70,692.09 |
| NEBRASKA | 33,016.03 | 31,353.99 | 30,785.17 | 32,461.58 | 127,616.77 |
| NEVADA | 15,335.36 | 14,464.20 | 13,666.02 | 14,450.38 | 57,915.96 |
| NEW HAMPSHIRE | 34,989.21 | 34,638.57 | 32,798.69 | 34,903.12 | 137,329.59 |
| NEW JERSEY | 95,809.55 | 95,115.20 | 84,581.27 | 93,795.26 | 369,301.28 |
| NEW MEXICO | 25,477.47 | 23,572.94 | 23,010.95 | 23,247.55 | 95,308.91 |
| NEW YORK | 240,162.23 | 241,645.58 | 215,537.81 | 235,121.76 | 932,467.38 |
| NORTH CAROLINA | 200,164.30 | 195,052.51 | 182,675.51 | 199,646.27 | 777,538.59 |
| NORTH DAKOTA | 16,458.41 | 15,618.42 | 14,634.49 | 14,351.46 | 61,062.78 |
| OHIO | 247,549.11 | 227,472.58 | 226,997.83 | 225,120.39 | 927,139.91 |
| OKLAHOMA | 52,411.12 | 48,748.72 | 47,721.19 | 49,950.20 | 198,831.23 |
| OREGON | 64,726.28 | 64,679.91 | 60,367.64 | 61,662.72 | 251,436.55 |
| PENNSYLVANIA | 243,129.94 | 227,411.59 | 218,738.55 | 220,332.30 | 909,612.38 |

| | | | | | |
|---|---|---|---|---|---|
| PUERTO RICO | 23,681.30 | 20,366.88 | 20,126.72 | 22,550.18 | 86,725.08 |
| RHODE ISLAND | 20,166.32 | 19,622.92 | 18,046.38 | 19,637.57 | 77,473.19 |
| SOUTH CAROLINA | 95,737.41 | 86,866.88 | 85,091.11 | 91,032.05 | 358,727.45 |
| SOUTH DAKOTA | 24,418.34 | 22,769.36 | 22,840.30 | 19,457.75 | 89,485.75 |
| TENNESSEE | 90,338.47 | 82,949.87 | 83,967.84 | 87,333.91 | 344,590.09 |

DRUG CODE:1724 DRUG NAME:METHYLPHENIDATE (DL;D;L;ISOMERS)

| STATE | QUARTER 1 | QUARTER 2 | QUARTER 3 | QUARTER 4 | TOTAL GRAMS |
|---|---|---|---|---|---|
| TEXAS | 360,118.49 | 330,484.31 | 315,117.14 | 347,698.23 | 1,353,418.17 |
| UTAH | 47,355.84 | 46,096.46 | 45,186.69 | 46,850.24 | 185,489.23 |
| VERMONT | 19,442.98 | 19,137.44 | 18,294.46 | 18,848.92 | 75,723.80 |
| VIRGIN ISLANDS | 215.51 | 201.95 | 183.6 | 198.22 | 799.28 |
| VIRGINIA | 140,246.29 | 136,907.99 | 127,888.78 | 137,017.93 | 542,060.99 |
| WASHINGTON | 113,512.91 | 113,115.56 | 105,641.28 | 110,968.21 | 443,237.96 |
| WEST VIRGINIA | 33,695.20 | 31,769.97 | 31,103.16 | 32,734.82 | 129,303.15 |
| WISCONSIN | 108,022.56 | 104,742.09 | 97,601.05 | 105,563.31 | 415,929.01 |
| WYOMING | 9,233.57 | 8,241.37 | 7,977.99 | 8,470.13 | 33,923.06 |
| TOTAL | 4,865,294.45 | 4,612,879.85 | 4,419,173.84 | 4,702,177.28 | 18,599,525.42 |

DRUG CODE:2012 DRUG NAME:GAMMA HYDROXYBUTYRIC ACID(FDA APPROVED-CSA

| STATE | QUARTER 1 | QUARTER 2 | QUARTER 3 | QUARTER 4 | TOTAL GRAMS |
|---|---|---|---|---|---|
| ALABAMA | 371.52 | 0 | 0 | 0 | 371.52 |
| CALIFORNIA | 4,829.76 | 3,938.11 | 2,006.21 | 2,080.51 | 12,854.59 |
| FLORIDA | 148.61 | 0 | 0 | 0 | 148.61 |
| GEORGIA | 297.22 | 1,634.69 | 668.74 | 0 | 2,600.65 |
| ILLINOIS | 668.74 | 371.52 | 1,263.17 | 743.04 | 3,046.47 |
| LOUISIANA | 0 | 0 | 222.91 | 520.13 | 743.04 |
| MASSACHUSETTS | 1,337.47 | 1,857.60 | 0 | 0 | 3,195.07 |
| MICHIGAN | 0 | 0 | 0 | 297.22 | 297.22 |
| MISSOURI | 12,981,428.93 | 4,943,965.25 | 7,389,458.50 | 0 | 25,314,852.68 |
| NEBRASKA | 222.91 | 0 | 0 | 0 | 222.91 |
| NEW YORK | 1,708.99 | 3,715.20 | 3,640.90 | 3,863.81 | 12,928.90 |
| NORTH CAROLINA | 1,560.38 | 1,114.56 | 1,188.86 | 594.43 | 4,458.23 |
| OHIO | 297.22 | 1,411.78 | 2,377.73 | 2,377.73 | 6,464.46 |
| SOUTH CAROLINA | 1,411.78 | 3,624.38 | 1,560.38 | 1,708.99 | 8,305.53 |
| TENNESSEE | 0 | 297.22 | 743.04 | 1,263.17 | 2,303.43 |
| TEXAS | 520.13 | 1,337.47 | 3,417.98 | 4,904.06 | 10,179.64 |
| VIRGINIA | 2,897.86 | 2,154.82 | 1,263.17 | 965.95 | 7,281.80 |
| WASHINGTON | 1,040.26 | 297.22 | 1,040.26 | 891.65 | 3,269.39 |
| TOTAL | 12,998,741.78 | 4,965,719.82 | 7,408,851.85 | 20,210.69 | 25,393,524.14 |

DRUG CODE:2100 DRUG NAME:BARBITURIC ACID DERIVIATIVE OR SALT [PER 21CFR 1

| STATE | QUARTER 1 | QUARTER 2 | QUARTER 3 | QUARTER 4 | TOTAL GRAMS |
|---|---|---|---|---|---|
| ALABAMA | 0 | 0 | 30 | 15 | 45 |
| ALASKA | 0 | 0 | 35 | 25 | 60 |
| ARIZONA | 0 | 175 | 395 | 10 | 580 |
| ARKANSAS | 0 | 0 | 5 | 0 | 5 |
| CALIFORNIA | 0 | 238 | 615 | 245 | 1,098 |
| COLORADO | 0 | 5 | 70 | 45 | 120 |
| CONNECTICUT | 0 | 35 | 30 | 50 | 115 |
| DELAWARE | 0 | 0 | 0 | 15 | 15 |
| DISTRICT OF COLUMBIA | 0 | 5 | 10 | 20 | 35 |
| FLORIDA | 0 | 25 | 155 | 135 | 315 |
| GEORGIA | 0 | 45 | 140 | 150 | 335 |
| IDAHO | 0 | 0 | 0 | 10 | 10 |

DRUG CODE:2100 DRUG NAME:BARBITURIC ACID DERIVIATIVE OR SALT [PER 21CFR 1

| STATE | QUARTER 1 | QUARTER 2 | QUARTER 3 | QUARTER 4 | TOTAL GRAMS |
|---|---|---|---|---|---|
| ILLINOIS | 0 | 10 | 185 | 75 | 270 |
| INDIANA | 0 | 5 | 90 | 80 | 175 |
| KANSAS | 0 | 0 | 70 | 25 | 95 |
| KENTUCKY | 0 | 10 | 25 | 50 | 85 |
| LOUISIANA | 0 | 0 | 30 | 30 | 60 |
| MAINE | 0 | 0 | 10 | 15 | 25 |
| MARYLAND | 0 | 30 | 30 | 45 | 105 |
| MASSACHUSETTS | 0 | 5 | 45 | 30 | 80 |
| MICHIGAN | 0 | 60 | 130 | 85 | 275 |
| MINNESOTA | 0 | 0 | 20 | 30 | 50 |
| MISSISSIPPI | 0 | 0 | 30 | 5 | 35 |
| MISSOURI | 0 | 5 | 40 | 65 | 110 |
| MONTANA | 0 | 5 | 5 | 15 | 25 |
| NEVADA | 0 | 15 | 25 | 5 | 45 |
| NEW HAMPSHIRE | 0 | 5 | 30 | 10 | 45 |
| NEW JERSEY | 0 | 130 | 525 | 435 | 1,090 |
| NEW MEXICO | 0 | 0 | 10 | 0 | 10 |
| NEW YORK | 0 | 57 | 320 | 330 | 707 |
| NORTH CAROLINA | 0 | 30 | 135 | 135 | 300 |
| OHIO | 0 | 15 | 110 | 150 | 275 |
| OKLAHOMA | 0 | 5 | 35 | 25 | 65 |
| OREGON | 0 | 0 | 15 | 10 | 25 |
| PENNSYLVANIA | 0 | 15 | 130 | 180 | 325 |
| RHODE ISLAND | 0 | 10 | 45 | 30 | 85 |
| SOUTH CAROLINA | 0 | 0 | 15 | 15 | 30 |
| TENNESSEE | 0 | 0 | 35 | 10 | 45 |
| TEXAS | 0 | 40 | 375 | 230 | 645 |
| VERMONT | 0 | 0 | 5 | 0 | 5 |
| VIRGINIA | 0 | 45 | 110 | 135 | 290 |
| WASHINGTON | 0 | 0 | 10 | 25 | 35 |
| WEST VIRGINIA | 0 | 0 | 0 | 40 | 40 |
| WISCONSIN | 0 | 0 | 5 | 15 | 20 |
| TOTAL | 0 | 1,025 | 4,130 | 3,050 | 8,205 |

DRUG CODE:2125 DRUG NAME:AMOBARBITAL (SCHEDULE 2)

| STATE | QUARTER 1 | QUARTER 2 | QUARTER 3 | QUARTER 4 | TOTAL GRAMS |
|---|---|---|---|---|---|
| ALABAMA | 5.47 | 0 | 0 | 0 | 5.47 |
| ARIZONA | 10.48 | 11.85 | 10.03 | 1.82 | 34.18 |
| ARKANSAS | 0.46 | 0 | 0 | 0 | 0.46 |
| CALIFORNIA | 51.49 | 28.71 | 15.04 | 7.75 | 102.99 |
| COLORADO | 26.43 | 26.43 | 11.39 | 1.82 | 66.07 |
| CONNECTICUT | 0.46 | 0.46 | 0 | 0 | 0.92 |
| DISTRICT OF COLUMBIA | 0 | 2.28 | 0 | 0 | 2.28 |
| FLORIDA | 5.92 | 13.22 | 0.91 | 8.66 | 28.71 |
| GEORGIA | 10.48 | 15.95 | 6.84 | 1.37 | 34.64 |
| IDAHO | 0.46 | 0 | 0 | 0 | 0.46 |

DRUG CODE:2125 DRUG NAME:AMOBARBITAL (SCHEDULE 2)

| STATE | QUARTER 1 | QUARTER 2 | QUARTER 3 | QUARTER 4 | TOTAL GRAMS |
|---|---|---|---|---|---|
| ILLINOIS | 10.94 | 10.94 | 7.29 | 1.37 | 30.54 |
| INDIANA | 6.38 | 1.37 | 0 | 0 | 7.75 |
| IOWA | 0 | 0.46 | 0 | 0 | 0.46 |
| KANSAS | 10.48 | 6.38 | 0.91 | 0 | 17.77 |

| | | | | | |
|---|---|---|---|---|---|
| KENTUCKY | 6.84 | 2.73 | 5.01 | 0.46 | 15.04 |
| MAINE | 1.37 | 1.37 | 0.91 | 0 | 3.65 |
| MARYLAND | 8.2 | 7.29 | 1.82 | 2.73 | 20.04 |
| MASSACHUSETTS | 7.75 | 15.49 | 5.01 | 1.37 | 29.62 |
| MICHIGAN | 8.66 | 6.38 | 3.65 | 0.91 | 19.6 |
| MINNESOTA | 5.01 | 15.95 | 0.91 | 1.82 | 23.69 |
| MISSISSIPPI | 3.65 | 0.91 | 1.37 | 0 | 5.93 |
| MISSOURI | 6.38 | 5.47 | 4.1 | 0.91 | 16.86 |
| NEBRASKA | 0.91 | 5.92 | 4.1 | 1.37 | 12.3 |
| NEVADA | 2.28 | 0 | 0.91 | 0 | 3.19 |
| NEW HAMPSHIRE | 4.1 | 5.47 | 0 | 1.37 | 10.94 |
| NEW JERSEY | 16.41 | 15.95 | 5.01 | 2.73 | 40.1 |
| NEW MEXICO | 0 | 2.28 | 0.46 | 0 | 2.74 |
| NEW YORK | 56.51 | 56.96 | 29.62 | 9.11 | 152.2 |
| NORTH CAROLINA | 12.3 | 19.14 | 0.91 | 7.75 | 40.1 |
| OHIO | 6.38 | 9.11 | 3.65 | 6.38 | 25.52 |
| OKLAHOMA | 0 | 0.46 | 0 | 0 | 0.46 |
| OREGON | 2.73 | 2.73 | 2.73 | 2.28 | 10.47 |
| PENNSYLVANIA | 15.04 | 15.04 | 9.11 | 3.65 | 42.84 |
| PUERTO RICO | 0 | 0 | 0.91 | 0 | 0.91 |
| SOUTH CAROLINA | 6.38 | 6.38 | 4.1 | 4.1 | 20.96 |
| TENNESSEE | 3.19 | 6.38 | 0 | 1.37 | 10.94 |
| TEXAS | 21.42 | 28.25 | 12.3 | 10.48 | 72.45 |
| UTAH | 7.29 | 8.66 | 4.1 | 2.28 | 22.33 |
| VERMONT | 1.82 | 0 | 0 | 0 | 1.82 |
| VIRGINIA | 5.92 | 2.28 | 4.56 | 0 | 12.76 |
| WASHINGTON | 15.95 | 22.79 | 2.28 | 7.75 | 48.77 |
| WEST VIRGINIA | 0 | 4.56 | 0.91 | 0 | 5.47 |
| WISCONSIN | 14.13 | 8.2 | 0.46 | 5.01 | 27.8 |
| TOTAL | 380.07 | 394.2 | 161.31 | 96.62 | 1,032.20 |

DRUG CODE:2165 DRUG NAME:BUTALBITAL

| STATE | QUARTER 1 | QUARTER 2 | QUARTER 3 | QUARTER 4 | TOTAL GRAMS |
|---|---|---|---|---|---|
| ALABAMA | 7,570 | 7,135 | 7,100 | 6,630 | 28,435 |
| ALASKA | 365 | 380 | 300 | 450 | 1,495 |
| ARIZONA | 12,510 | 11,615 | 11,790 | 11,415 | 47,330 |
| ARKANSAS | 4,830 | 5,050 | 4,955 | 4,850 | 19,685 |
| CALIFORNIA | 46,690 | 45,030 | 44,115 | 40,680 | 176,515 |
| COLORADO | 4,350 | 4,240 | 3,960 | 3,940 | 16,490 |
| CONNECTICUT | 6,465 | 6,325 | 6,050 | 5,820 | 24,660 |
| DELAWARE | 965 | 860 | 995 | 985 | 3,805 |
| DISTRICT OF COLUMBIA | 645 | 685 | 665 | 670 | 2,665 |

DRUG CODE:2165 DRUG NAME:BUTALBITAL

| STATE | QUARTER 1 | QUARTER 2 | QUARTER 3 | QUARTER 4 | TOTAL GRAMS |
|---|---|---|---|---|---|
| FLORIDA | 43,495 | 36,265 | 34,555 | 35,855 | 150,170 |
| GEORGIA | 15,870 | 15,960 | 15,990 | 16,030 | 63,850 |
| GUAM | 15 | 5 | 15 | 0 | 35 |
| HAWAII | 2,045 | 1,910 | 2,035 | 1,930 | 7,920 |
| IDAHO | 1,950 | 2,105 | 2,050 | 2,025 | 8,130 |
| ILLINOIS | 15,605 | 15,260 | 15,490 | 15,815 | 62,170 |
| INDIANA | 7,570 | 7,655 | 7,855 | 7,240 | 30,320 |
| IOWA | 2,210 | 2,260 | 2,005 | 2,145 | 8,620 |
| KANSAS | 3,665 | 3,545 | 3,890 | 4,355 | 15,455 |
| KENTUCKY | 4,735 | 4,710 | 4,595 | 4,705 | 18,745 |
| LOUISIANA | 8,425 | 7,895 | 7,930 | 7,935 | 32,185 |
| MAINE | 1,400 | 1,405 | 1,265 | 1,315 | 5,385 |

| | | | | | |
|---|---|---|---|---|---|
| MARYLAND | 8,000 | 8,480 | 7,975 | 7,920 | 32,375 |
| MASSACHUSETTS | 5,940 | 5,740 | 5,630 | 5,180 | 22,490 |
| MICHIGAN | 16,855 | 16,390 | 16,250 | 16,690 | 66,185 |
| MINNESOTA | 3,615 | 3,855 | 3,555 | 3,550 | 14,575 |
| MISSISSIPPI | 3,525 | 3,345 | 3,180 | 3,365 | 13,415 |
| MISSOURI | 10,235 | 10,630 | 10,605 | 10,695 | 42,165 |
| MONTANA | 1,475 | 1,600 | 1,765 | 1,560 | 6,400 |
| NEBRASKA | 2,230 | 2,220 | 1,895 | 1,945 | 8,290 |
| NEVADA | 9,315 | 8,680 | 8,550 | 8,625 | 35,170 |
| NEW HAMPSHIRE | 1,310 | 1,275 | 1,270 | 1,335 | 5,190 |
| NEW JERSEY | 12,965 | 12,170 | 11,605 | 11,080 | 47,820 |
| NEW MEXICO | 2,205 | 2,320 | 2,245 | 2,065 | 8,835 |
| NEW YORK | 13,385 | 12,460 | 11,620 | 11,060 | 48,525 |
| NORTH CAROLINA | 9,880 | 9,515 | 9,220 | 9,550 | 38,165 |
| NORTH DAKOTA | 435 | 455 | 455 | 345 | 1,690 |
| OHIO | 13,040 | 12,850 | 12,340 | 11,415 | 49,645 |
| OKLAHOMA | 9,070 | 8,560 | 8,559.95 | 8,810 | 34,999.95 |
| OREGON | 4,290 | 3,900 | 3,875 | 3,995 | 16,060 |
| PENNSYLVANIA | 21,710 | 19,190 | 18,780 | 19,320 | 79,000 |
| PUERTO RICO | 140 | 185 | 170 | 185 | 680 |
| RHODE ISLAND | 1,195 | 975 | 985 | 1,035 | 4,190 |
| SOUTH CAROLINA | 6,380 | 6,015 | 5,800 | 5,895 | 24,090 |
| SOUTH DAKOTA | 990 | 1,105 | 990 | 880 | 3,965 |
| TENNESSEE | 7,300 | 7,425 | 7,145 | 7,320 | 29,190 |
| TEXAS | 34,105 | 33,295 | 33,214.95 | 33,004.90 | 133,619.85 |
| UTAH | 4,945 | 4,760 | 4,740 | 5,055 | 19,500 |
| VERMONT | 285 | 310 | 335 | 210 | 1,140 |
| VIRGIN ISLANDS | 50 | 25 | 20 | 20 | 115 |
| VIRGINIA | 10,670 | 10,130 | 10,115 | 9,535 | 40,450 |
| WASHINGTON | 7,280 | 7,430 | 7,385 | 7,450 | 29,545 |
| WEST VIRGINIA | 1,945 | 1,700 | 1,900 | 1,920 | 7,465 |
| WISCONSIN | 4,360 | 4,415 | 3,955 | 3,860 | 16,590 |
| WYOMING | 855 | 905 | 880 | 890 | 3,530 |

DRUG CODE:2165 DRUG NAME:BUTALBITAL

| STATE | QUARTER 1 | QUARTER 2 | QUARTER 3 | QUARTER 4 | TOTAL GRAMS |
|---|---|---|---|---|---|
| TOTAL | 421,355 | 402,605 | 394,614.90 | 390,554.90 | 1,609,129.80 |

DRUG CODE:2270 DRUG NAME:PENTOBARBITAL (SCHEDULE 2)

| STATE | QUARTER 1 | QUARTER 2 | QUARTER 3 | QUARTER 4 | TOTAL GRAMS |
|---|---|---|---|---|---|
| ALABAMA | 17,489.45 | 18,217.04 | 20,550.85 | 20,455.72 | 76,713.06 |
| ALASKA | 5,021.81 | 3,222.71 | 4,896.50 | 4,798.52 | 17,939.54 |
| ARIZONA | 47,701.11 | 50,837.13 | 37,897.27 | 49,239.47 | 185,674.98 |
| ARKANSAS | 9,083.46 | 7,910.22 | 7,381.97 | 6,573.45 | 30,949.10 |
| CALIFORNIA | 150,844.67 | 162,829.07 | 190,843.75 | 175,139.41 | 679,656.90 |
| COLORADO | 38,458.51 | 35,548.61 | 37,009.22 | 31,752.81 | 142,769.15 |
| CONNECTICUT | 11,669.47 | 13,511.30 | 20,729.29 | 15,619.18 | 61,529.24 |
| DELAWARE | 1,452.03 | 2,184.06 | 1,329.28 | 2,362.52 | 7,327.89 |
| DISTRICT OF COLUMBIA | 152.66 | 246.08 | 209.62 | 284.81 | 893.17 |
| FLORIDA | 60,632.38 | 99,403.91 | 50,826.46 | 64,876.42 | 275,739.17 |
| GEORGIA | 30,796.81 | 36,037.52 | 42,020.98 | 27,735.69 | 136,591 |
| GUAM | 0 | 3,215.42 | 0 | 0 | 3,215.42 |
| HAWAII | 15,770.87 | 3,668.84 | 5,860.30 | 12,831.14 | 38,131.15 |
| IDAHO | 19,872.23 | 18,540.61 | 22,439.58 | 30,663.49 | 91,515.91 |
| ILLINOIS | 62,954.87 | 68,957.89 | 67,469.87 | 57,098.62 | 256,481.25 |
| INDIANA | 37,975.40 | 37,467.50 | 40,450.79 | 37,017.59 | 152,911.28 |
| IOWA | 21,691.23 | 11,968.96 | 22,578.84 | 17,138.78 | 73,377.81 |

| | | | | | |
|---|---|---|---|---|---|
| KANSAS | 21,923.73 | 21,831.86 | 17,475.18 | 22,912.14 | 84,142.91 |
| KENTUCKY | 22,495.01 | 28,994.53 | 31,667.80 | 26,492.68 | 109,650.02 |
| LOUISIANA | 25,022.69 | 18,894.73 | 24,193.43 | 22,280.58 | 90,391.43 |
| MAINE | 15,700.77 | 19,622.80 | 13,930.75 | 17,148.07 | 66,402.39 |
| MARYLAND | 17,821.77 | 18,327.49 | 18,560.63 | 17,364.98 | 72,074.87 |
| MASSACHUSETTS | 25,864.80 | 23,217.71 | 29,392.82 | 27,445.24 | 105,920.57 |
| MICHIGAN | 81,260.46 | 99,709.30 | 91,568.34 | 97,220.15 | 369,758.25 |
| MINNESOTA | 23,880.31 | 28,373.96 | 33,293.70 | 33,178.30 | 118,726.27 |
| MISSISSIPPI | 7,196.13 | 3,110.50 | 8,687.62 | 3,935.79 | 22,930.04 |
| MISSOURI | 33,652.70 | 30,010.20 | 41,554.73 | 39,413.53 | 144,631.16 |
| MONTANA | 11,647.69 | 10,579.71 | 25,884.22 | 21,060.63 | 69,172.25 |
| NEBRASKA | 16,982.12 | 20,440.88 | 19,965.13 | 25,221.17 | 82,609.30 |
| NEVADA | 16,105.44 | 13,254.23 | 13,750.29 | 18,045.72 | 61,155.68 |
| NEW HAMPSHIRE | 13,904.67 | 14,232.67 | 18,279.31 | 15,909.82 | 62,326.47 |
| NEW JERSEY | 18,438.21 | 18,294.79 | 25,386.92 | 20,538.48 | 82,658.40 |
| NEW MEXICO | 13,910.70 | 13,385.73 | 19,467.05 | 22,140.64 | 68,904.12 |
| NEW YORK | 57,624.50 | 70,206.87 | 73,237.51 | 81,979.40 | 283,048.28 |
| NORTH CAROLINA | 52,525.37 | 56,663.80 | 61,791.25 | 67,710.69 | 238,691.11 |
| NORTH DAKOTA | 9,788.44 | 2,500.88 | 7,289.83 | 5,369.97 | 24,949.12 |
| OHIO | 61,515.11 | 59,653.97 | 68,246.40 | 66,949.53 | 256,365.01 |
| OKLAHOMA | 17,576.07 | 12,888.73 | 19,815.37 | 17,246.60 | 67,526.77 |
| OREGON | 28,913.77 | 37,695.50 | 26,936.43 | 32,689.53 | 126,235.23 |
| PENNSYLVANIA | 66,702.67 | 56,156.10 | 64,715.40 | 67,009.70 | 254,583.87 |
| PUERTO RICO | 893.17 | 4,003.32 | 0 | 0 | 4,896.49 |
| RHODE ISLAND | 4,619.97 | 8,712.98 | 6,666.07 | 10,140.69 | 30,139.71 |

DRUG CODE:2270 DRUG NAME:PENTOBARBITAL (SCHEDULE 2)

| STATE | QUARTER 1 | QUARTER 2 | QUARTER 3 | QUARTER 4 | TOTAL GRAMS |
|---|---|---|---|---|---|
| SOUTH CAROLINA | 13,529.61 | 18,255.12 | 12,085.40 | 16,758.46 | 60,628.59 |
| SOUTH DAKOTA | 6,774.44 | 5,687.14 | 5,586.88 | 7,701.33 | 25,749.79 |
| TENNESSEE | 35,324.80 | 32,428.96 | 39,472.04 | 38,961.32 | 146,187.12 |
| TEXAS | 100,326.10 | 157,407.77 | 160,886.88 | 141,361.12 | 559,981.87 |
| UTAH | 15,440.84 | 9,771.55 | 8,612.27 | 10,820.60 | 44,645.26 |
| VERMONT | 6,841.96 | 12,702.14 | 9,205.22 | 15,369.68 | 44,119 |
| VIRGINIA | 45,446 | 44,532.98 | 45,354.76 | 41,492.20 | 176,825.94 |
| WASHINGTON | 57,384.09 | 45,265.56 | 47,334.65 | 62,841.83 | 212,826.13 |
| WEST VIRGINIA | 9,642.61 | 8,242.51 | 11,234.42 | 12,171.28 | 41,290.82 |
| WISCONSIN | 40,686.88 | 39,323.92 | 45,038.47 | 38,215.21 | 163,264.48 |
| WYOMING | 14,402.85 | 5,332.60 | 8,441.84 | 10,663.38 | 38,840.67 |
| TOTAL | 1,543,333.41 | 1,643,480.36 | 1,727,503.58 | 1,729,348.06 | 6,643,665.41 |

DRUG CODE:2315 DRUG NAME:SECOBARBITAL (SCHEDULE 2)

| STATE | QUARTER 1 | QUARTER 2 | QUARTER 3 | QUARTER 4 | TOTAL GRAMS |
|---|---|---|---|---|---|
| ARIZONA | 9.16 | 9.16 | 0 | 18.31 | 36.63 |
| ARKANSAS | 0 | 0 | 0 | 18.31 | 18.31 |
| CALIFORNIA | 64.09 | 485.22 | 1,199.31 | 1,061.98 | 2,810.60 |
| COLORADO | 18.31 | 18.31 | 9.16 | 9.16 | 54.94 |
| CONNECTICUT | 0 | 27.47 | 9.16 | 9.16 | 45.79 |
| DELAWARE | 0 | 0 | 0 | 9.16 | 9.16 |
| FLORIDA | 45.78 | 91.55 | 82.4 | 91.55 | 311.28 |
| GEORGIA | 18.31 | 18.31 | 9.16 | 18.31 | 64.09 |
| HAWAII | 18.31 | 27.47 | 9.16 | 9.16 | 64.1 |
| ILLINOIS | 82.4 | 82.4 | 91.55 | 64.09 | 320.44 |
| KANSAS | 27.47 | 27.47 | 9.16 | 9.16 | 73.26 |
| LOUISIANA | 9.16 | 27.47 | 18.31 | 0 | 54.94 |
| MARYLAND | 36.62 | 45.78 | 64.09 | 36.62 | 183.11 |

| | | | | | |
|---|---|---|---|---|---|
| MASSACHUSETTS | 18.31 | 82.4 | 73.24 | 45.78 | 219.73 |
| MICHIGAN | 0 | 0 | 9.16 | 9.16 | 18.32 |
| MISSOURI | 45.78 | 45.78 | 27.47 | 27.47 | 146.5 |
| NEVADA | 0 | 0 | 9.16 | 0 | 9.16 |
| NEW HAMPSHIRE | 9.16 | 9.16 | 18.31 | 0 | 36.63 |
| NEW JERSEY | 27.47 | 91.55 | 91.55 | 109.86 | 320.43 |
| NEW MEXICO | 0 | 9.16 | 18.31 | 9.16 | 36.63 |
| NEW YORK | 329.58 | 595.08 | 466.91 | 476.06 | 1,867.63 |
| NORTH CAROLINA | 27.47 | 9.16 | 36.62 | 9.16 | 82.41 |
| OHIO | 45.78 | 54.93 | 54.93 | 45.78 | 201.42 |
| OREGON | 164.79 | 494.37 | 201.41 | 173.95 | 1,034.52 |
| PENNSYLVANIA | 54.93 | 109.86 | 54.93 | 64.09 | 283.81 |
| RHODE ISLAND | 18.31 | 9.16 | 18.31 | 0 | 45.78 |
| TEXAS | 9.16 | 27.47 | 27.47 | 9.16 | 73.26 |
| VERMONT | 0 | 9.16 | 36.62 | 64.09 | 109.87 |
| VIRGINIA | 27.47 | 27.47 | 27.47 | 45.78 | 128.19 |
| WASHINGTON | 192.26 | 238.03 | 119.02 | 265.5 | 814.81 |
| WEST VIRGINIA | 0 | 9.16 | 0 | 0 | 9.16 |

DRUG CODE:2315 DRUG NAME:SECOBARBITAL (SCHEDULE 2)

| STATE | QUARTER 1 | QUARTER 2 | QUARTER 3 | QUARTER 4 | TOTAL GRAMS |
|---|---|---|---|---|---|
| WYOMING | 0 | 9.16 | 9.16 | 0 | 18.32 |
| TOTAL | 1,300.08 | 2,691.67 | 2,801.51 | 2,709.97 | 9,503.23 |

DRUG CODE:7379 DRUG NAME:NABILONE

| STATE | QUARTER 1 | QUARTER 2 | QUARTER 3 | QUARTER 4 | TOTAL GRAMS |
|---|---|---|---|---|---|
| ARIZONA | 0 | 0 | 0 | 0.15 | 0.15 |
| CALIFORNIA | 0 | 0 | 0 | 0.05 | 0.05 |
| COLORADO | 0.3 | 0.3 | 0.25 | 0.2 | 1.05 |
| FLORIDA | 1.2 | 0.35 | 1.2 | 0.75 | 3.5 |
| HAWAII | 0 | 0.1 | 0 | 0.1 | 0.2 |
| ILLINOIS | 0.2 | 0.1 | 0.35 | 0.15 | 0.8 |
| INDIANA | 0 | 0.1 | 0.15 | 0.05 | 0.3 |
| IOWA | 0 | 0 | 0.05 | 0 | 0.05 |
| MAINE | 0 | 0 | 0 | 0.1 | 0.1 |
| MARYLAND | 1.65 | 1.35 | 1.2 | 1.15 | 5.35 |
| MASSACHUSETTS | 0 | 0.25 | 0.35 | 0.3 | 0.9 |
| MINNESOTA | 0.05 | 0.05 | 0.15 | 0.05 | 0.3 |
| MISSOURI | 0.05 | 0.25 | 0.1 | 0.05 | 0.45 |
| NEBRASKA | 0.2 | 0.15 | 0.2 | 0.15 | 0.7 |
| NEW HAMPSHIRE | 0 | 0.1 | 0.15 | 0 | 0.25 |
| NEW JERSEY | 0 | 0 | 0.05 | 0.1 | 0.15 |
| NEW MEXICO | 0 | 0.15 | 0 | 0.15 | 0.3 |
| NEW YORK | 0.35 | 0.2 | 0.1 | 0.5 | 1.15 |
| NORTH CAROLINA | 0 | 0.15 | 0.25 | 0.15 | 0.55 |
| OHIO | 0.4 | 0.05 | 0.2 | 0.1 | 0.75 |
| PENNSYLVANIA | 0.25 | 0.35 | 0.2 | 0.25 | 1.05 |
| RHODE ISLAND | 0 | 0.05 | 0 | 0 | 0.05 |
| VIRGINIA | 0.1 | 0.35 | 0.35 | 0.2 | 1 |
| WISCONSIN | 0.05 | 0 | 0 | 0 | 0.05 |
| TOTAL | 4.8 | 4.4 | 5.3 | 4.7 | 19.2 |

DRUG CODE:7471 DRUG NAME:PHENCYCLIDINE (PCP)

| STATE | QUARTER 1 | QUARTER 2 | QUARTER 3 | QUARTER 4 | TOTAL GRAMS |
|---|---|---|---|---|---|
| MINNESOTA | 0 | 0 | 0 | 0.22 | 0.22 |

| | | | | | |
|---|---|---|---|---|---|
| TOTAL | 0 | 0 | 0 | 0.22 | 0.22 |

DRUG CODE:9041L DRUG NAME:COCAINE

| STATE | QUARTER 1 | QUARTER 2 | QUARTER 3 | QUARTER 4 | TOTAL GRAMS |
|---|---|---|---|---|---|
| ALABAMA | 55.43 | 58.57 | 60 | 74 | 248 |
| ALASKA | 17.78 | 11 | 12.57 | 21.93 | 63.28 |
| ARIZONA | 131.13 | 128.67 | 115.64 | 127.99 | 503.43 |
| ARKANSAS | 67.71 | 54.14 | 49.82 | 44.32 | 215.99 |
| CALIFORNIA | 950.44 | 941.08 | 929.26 | 944.01 | 3,764.79 |
| COLORADO | 97.1 | 97.71 | 81.28 | 99.85 | 375.94 |
| CONNECTICUT | 49.64 | 43 | 39.64 | 51.43 | 183.71 |
| DELAWARE | 23.86 | 19.14 | 11 | 20.57 | 74.57 |
| DISTRICT OF COLUMBIA | 27.43 | 19.43 | 27.43 | 23.14 | 97.43 |
| FLORIDA | 319.3 | 319.12 | 305.44 | 294.02 | 1,237.88 |

DRUG CODE:9041L DRUG NAME:COCAINE

| STATE | QUARTER 1 | QUARTER 2 | QUARTER 3 | QUARTER 4 | TOTAL GRAMS |
|---|---|---|---|---|---|
| GEORGIA | 159.85 | 176.35 | 177.52 | 216.74 | 730.46 |
| GUAM | 0 | 0 | 0.71 | 0 | 0.71 |
| HAWAII | 17.14 | 21.43 | 25 | 24.86 | 88.43 |
| IDAHO | 79.28 | 49.14 | 78.03 | 55.35 | 261.8 |
| ILLINOIS | 365.55 | 375.87 | 307.55 | 353.48 | 1,402.45 |
| INDIANA | 205.59 | 224.91 | 169.38 | 220.91 | 820.79 |
| IOWA | 93.21 | 108.78 | 91.92 | 109.71 | 403.62 |
| KANSAS | 83.71 | 105.89 | 82.35 | 94.67 | 366.62 |
| KENTUCKY | 119.53 | 122.21 | 85.6 | 110.78 | 438.12 |
| LOUISIANA | 93.57 | 92.07 | 89.57 | 110.71 | 385.92 |
| MAINE | 25.64 | 26.57 | 29.71 | 26.57 | 108.49 |
| MARYLAND | 152.45 | 153.28 | 128.99 | 124.71 | 559.43 |
| MASSACHUSETTS | 249.34 | 236.98 | 213.99 | 207.92 | 908.23 |
| MICHIGAN | 409.37 | 408.62 | 349.37 | 415.83 | 1,583.19 |
| MINNESOTA | 235.23 | 225.13 | 208.2 | 215.41 | 883.97 |
| MISSISSIPPI | 104.06 | 101.28 | 81.49 | 95.53 | 382.36 |
| MISSOURI | 188.38 | 164.85 | 185.06 | 173.02 | 711.31 |
| MONTANA | 49.57 | 48.6 | 55.07 | 54.85 | 208.09 |
| NEBRASKA | 35.28 | 34.14 | 43.46 | 37.57 | 150.45 |
| NEVADA | 40.71 | 57 | 68.92 | 50.5 | 217.13 |
| NEW HAMPSHIRE | 58.42 | 47.85 | 52.07 | 52.14 | 210.48 |
| NEW JERSEY | 179.6 | 175.02 | 169.6 | 173.06 | 697.28 |
| NEW MEXICO | 88.57 | 84.32 | 69.75 | 70.32 | 312.96 |
| NEW YORK | 381.65 | 353.94 | 390.26 | 348.19 | 1,474.04 |
| NORTH CAROLINA | 149.03 | 150.03 | 176.27 | 154.53 | 629.86 |
| NORTH DAKOTA | 42.71 | 93.89 | 22 | 25.28 | 183.88 |
| OHIO | 179.99 | 165.31 | 178.35 | 199.34 | 722.99 |
| OKLAHOMA | 46.57 | 35.78 | 31 | 40.85 | 154.2 |
| OREGON | 136.88 | 84.46 | 111.71 | 82.92 | 415.97 |
| PENNSYLVANIA | 445.94 | 456.26 | 414.01 | 420.97 | 1,737.18 |
| RHODE ISLAND | 28.18 | 23 | 20.71 | 24.36 | 96.25 |
| SOUTH CAROLINA | 42.32 | 35.35 | 30 | 34.14 | 141.81 |
| SOUTH DAKOTA | 24.71 | 56.96 | 41.85 | 22.96 | 146.48 |
| TENNESSEE | 187.49 | 198.2 | 184.92 | 192.49 | 763.1 |
| TEXAS | 373.69 | 377.23 | 350.41 | 446.97 | 1,548.30 |
| UTAH | 108.92 | 130.28 | 94.17 | 128.21 | 461.58 |
| VERMONT | 10 | 8.71 | 4 | 8.43 | 31.14 |
| VIRGINIA | 189.45 | 184.63 | 147.99 | 182.92 | 704.99 |
| WASHINGTON | 160.85 | 118.42 | 124.71 | 141.78 | 545.76 |
| WEST VIRGINIA | 50.28 | 45.14 | 58.14 | 43.57 | 197.13 |

| | | | | | |
|---|---|---|---|---|---|
| WISCONSIN | 155.42 | 171.06 | 136.21 | 166.49 | 629.18 |
| WYOMING | 17.28 | 15.28 | 25.32 | 21.14 | 79.02 |
| TOTAL | 7,505.23 | 7,436.08 | 6,937.42 | 7,381.44 | 29,260.17 |

DRUG CODE:9050 DRUG NAME:CODEINE

| STATE | QUARTER 1 | QUARTER 2 | QUARTER 3 | QUARTER 4 | TOTAL GRAMS |
|---|---|---|---|---|---|

DRUG CODE:9050 DRUG NAME:CODEINE

| STATE | QUARTER 1 | QUARTER 2 | QUARTER 3 | QUARTER 4 | TOTAL GRAMS |
|---|---|---|---|---|---|
| ALABAMA | 56,915.93 | 54,308.03 | 53,362.39 | 53,006.44 | 217,592.79 |
| ALASKA | 7,865.67 | 7,359.63 | 7,520.37 | 7,068.88 | 29,814.55 |
| AMERICAN SAMOA | 764.69 | 503.9 | 291.73 | 590.1 | 2,150.42 |
| ARIZONA | 83,767.01 | 78,144.94 | 79,191.40 | 79,062.49 | 320,165.84 |
| ARKANSAS | 78,106.94 | 78,239.34 | 77,419.83 | 75,704.17 | 309,470.28 |
| CALIFORNIA | 559,516.06 | 546,150.72 | 529,670.63 | 508,050.73 | 2,143,388.14 |
| COLORADO | 29,288.88 | 27,187.87 | 27,128.53 | 27,063.24 | 110,668.52 |
| CONNECTICUT | 29,042.10 | 28,157.52 | 27,329.06 | 26,934.62 | 111,463.30 |
| DELAWARE | 10,976.74 | 11,114.88 | 10,970.50 | 10,525.09 | 43,587.21 |
| DISTRICT OF COLUMBIA | 13,472.42 | 13,979.23 | 13,421.96 | 13,056.72 | |
| 53,930.33 | | | | | |
| FLORIDA | 189,518.48 | 173,625.41 | 163,102.78 | 171,494.95 | 697,741.62 |
| GEORGIA | 106,425.48 | 105,985.44 | 104,918.30 | 105,115.59 | 422,444.81 |
| GUAM | 1,384.25 | 1,306.60 | 1,732.30 | 1,007.81 | 5,430.96 |
| HAWAII | 15,854.62 | 14,722.08 | 14,512.39 | 13,232.63 | 58,321.72 |
| IDAHO | 15,421.05 | 15,224.86 | 14,824.80 | 13,707.14 | 59,177.85 |
| ILLINOIS | 174,229.87 | 175,503.52 | 169,230.62 | 173,824.91 | 692,788.92 |
| INDIANA | 59,015.33 | 61,397.94 | 57,949.79 | 55,718.58 | 234,081.64 |
| IOWA | 19,402.83 | 22,970.75 | 21,610.54 | 21,859.88 | 85,844 |
| KANSAS | 31,028.78 | 30,277.39 | 28,586.61 | 29,899.57 | 119,792.35 |
| KENTUCKY | 52,549.52 | 54,906.07 | 52,525.37 | 51,677.39 | 211,658.35 |
| LOUISIANA | 45,795.34 | 45,034.77 | 45,832.91 | 45,429.92 | 182,092.94 |
| MAINE | 12,175.44 | 12,947.24 | 12,474.22 | 12,175.38 | 49,772.28 |
| MARYLAND | 67,086.95 | 66,752.53 | 65,081.06 | 64,473.18 | 263,393.72 |
| MASSACHUSETTS | 56,477.91 | 54,448.78 | 52,014.62 | 49,757.98 | 212,699.29 |
| MICHIGAN | 273,097.95 | 280,248.45 | 267,810.51 | 277,520.78 | 1,098,677.69 |
| MINNESOTA | 44,320.76 | 43,239.14 | 41,747.55 | 40,472.58 | 169,780.03 |
| MISSISSIPPI | 28,189.13 | 28,359.98 | 28,066.55 | 28,602.59 | 113,218.25 |
| MISSOURI | 75,895.61 | 75,653.57 | 74,349.22 | 71,704.03 | 297,602.43 |
| MONTANA | 11,835.94 | 11,278.43 | 11,619.58 | 10,993.89 | 45,727.84 |
| NEBRASKA | 13,372.65 | 14,150.62 | 13,667.40 | 13,326.81 | 54,517.48 |
| NEVADA | 31,969.10 | 29,866.50 | 27,674.03 | 28,484.11 | 117,993.74 |
| NEW HAMPSHIRE | 9,146.99 | 8,774.92 | 8,763.17 | 8,755.11 | 35,440.19 |
| NEW JERSEY | 87,418.17 | 89,652.07 | 86,476.63 | 87,332.47 | 350,879.34 |
| NEW MEXICO | 22,117.16 | 20,455.81 | 21,127.38 | 20,623.73 | 84,324.08 |
| NEW YORK | 180,179.34 | 167,858.94 | 159,098.18 | 159,295.74 | 666,432.20 |
| NORTH CAROLINA | 53,870.54 | 51,512.90 | 50,498.46 | 52,292.66 | 208,174.56 |
| NORTH DAKOTA | 6,933.76 | 6,654.11 | 6,386.23 | 5,116.12 | 25,090.22 |
| OHIO | 119,743.37 | 114,019.85 | 111,649.68 | 111,682.83 | 457,095.73 |
| OKLAHOMA | 94,975.54 | 95,013.21 | 95,855.66 | 96,342.27 | 382,186.68 |
| OREGON | 38,406.52 | 37,358.60 | 36,435.23 | 36,005.45 | 148,205.80 |
| PENNSYLVANIA | 160,591 | 154,882.67 | 148,013.38 | 148,041.06 | 611,528.11 |
| PUERTO RICO | 12,354.42 | 13,632.48 | 13,377.92 | 13,815.56 | 53,180.38 |
| RHODE ISLAND | 9,106.41 | 8,546.82 | 8,684.66 | 8,078.82 | 34,416.71 |
| SOUTH CAROLINA | 86,228.20 | 81,643.19 | 81,013.51 | 70,850.20 | 319,735.10 |
| SOUTH DAKOTA | 8,324.82 | 8,413.64 | 7,948.46 | 7,878.32 | 32,565.24 |

RETAIL DRUG DISTRIBUTION BY STATE WITHIN DRUG CODE BY GRAMS WT

DRUG CODE:9050 DRUG NAME:CODEINE

| STATE | QUARTER 1 | QUARTER 2 | QUARTER 3 | QUARTER 4 | TOTAL GRAMS |
|---|---|---|---|---|---|
| TENNESSEE | 101,655.08 | 103,637.87 | 105,996.77 | 101,234.56 | 412,524.28 |
| TEXAS | 1,006,965.20 | 1,021,674.50 | 1,034,125.82 | 1,040,805.40 | 4,103,570.92 |
| UTAH | 18,137.64 | 17,501.25 | 17,798.59 | 17,490.20 | 70,927.68 |
| VERMONT | 4,075.17 | 3,869.39 | 3,723.96 | 3,628.38 | 15,296.90 |
| VIRGIN ISLANDS | 710.94 | 759.24 | 727.27 | 612.2 | 2,809.65 |
| VIRGINIA | 63,259.11 | 61,564.18 | 61,385.04 | 58,794.24 | 245,002.57 |
| WASHINGTON | 68,323.80 | 68,582.49 | 65,760.12 | 64,977.36 | 267,643.77 |
| WEST VIRGINIA | 31,266.96 | 31,158.31 | 31,774.17 | 31,670.98 | 125,870.42 |
| WISCONSIN | 57,748.42 | 55,766.77 | 54,799.44 | 53,386.85 | 221,701.48 |
| WYOMING | 5,033.59 | 4,758.39 | 4,590.41 | 4,527.16 | 18,909.55 |
| TOTAL | 4,441,335.58 | 4,390,737.73 | 4,311,647.69 | 4,284,779.85 | 17,428,500.85 |

DRUG CODE:9064 DRUG NAME:BUPRENORPHINE

| STATE | QUARTER 1 | QUARTER 2 | QUARTER 3 | QUARTER 4 | TOTAL GRAMS |
|---|---|---|---|---|---|
| ALABAMA | 22,519 | 22,266.21 | 22,773.07 | 23,272.19 | 90,830.47 |
| ALASKA | 1,849.08 | 1,880.45 | 1,943.54 | 2,017.84 | 7,690.91 |
| AMERICAN SAMOA | 0 | 0.01 | 0 | 0 | 0.01 |
| ARIZONA | 7,805.24 | 8,155.48 | 8,482.85 | 8,684.05 | 33,127.62 |
| ARKANSAS | 4,744.73 | 4,942.48 | 5,000.99 | 5,370.98 | 20,059.18 |
| CALIFORNIA | 32,734.39 | 34,264.25 | 34,832.23 | 35,754.13 | 137,585 |
| COLORADO | 6,473.02 | 6,717.20 | 6,806.73 | 6,940.14 | 26,937.09 |
| CONNECTICUT | 13,087.74 | 13,652.04 | 13,795.71 | 14,125.94 | 54,661.43 |
| DELAWARE | 2,785.37 | 2,901.08 | 2,999.18 | 2,980.93 | 11,666.56 |
| DISTRICT OF COLUMBIA | 1,639.16 | 1,800.75 | 1,927.91 | 2,014.34 | 7,382.16 |
| FLORIDA | 41,730.17 | 43,085.36 | 43,041.03 | 44,579.87 | 172,436.43 |
| GEORGIA | 14,067.55 | 14,316.32 | 14,436.47 | 14,716.61 | 57,536.95 |
| GUAM | 3.56 | 0.49 | 0.49 | 0.02 | 4.56 |
| HAWAII | 1,709.18 | 1,794.27 | 1,879.66 | 1,826.49 | 7,209.60 |
| IDAHO | 2,834.64 | 3,028.05 | 3,123.72 | 3,159.03 | 12,145.44 |
| ILLINOIS | 11,069.07 | 11,867.31 | 12,013.61 | 12,662.53 | 47,612.52 |
| INDIANA | 15,244.26 | 15,715.81 | 16,010.55 | 16,718.21 | 63,688.83 |
| IOWA | 1,354.15 | 1,470.96 | 1,518.75 | 1,493.66 | 5,837.52 |
| KANSAS | 2,299.43 | 2,283.26 | 2,429.10 | 2,415.71 | 9,427.50 |
| KENTUCKY | 30,516.58 | 30,935.12 | 31,023.17 | 31,919.67 | 124,394.54 |
| LOUISIANA | 14,197.22 | 14,290.58 | 17,454.66 | 17,780.74 | 63,723.20 |
| MAINE | 7,378.02 | 7,677.01 | 8,177.64 | 8,596.80 | 31,829.47 |
| MARYLAND | 21,769 | 23,089.45 | 22,979.66 | 23,593.35 | 91,431.46 |
| MASSACHUSETTS | 36,604.13 | 37,457.98 | 39,017.18 | 38,819.19 | 151,898.48 |
| MICHIGAN | 23,376.02 | 24,211.74 | 24,506.62 | 25,039.81 | 97,134.19 |
| MINNESOTA | 5,703.12 | 6,002.58 | 6,249.05 | 6,716.11 | 24,670.86 |
| MISSISSIPPI | 8,983.66 | 9,278.88 | 9,355.90 | 9,466.74 | 37,085.18 |
| MISSOURI | 7,726.79 | 8,116.40 | 8,352 | 8,627.98 | 32,823.17 |
| MONTANA | 1,546.76 | 1,738.16 | 1,868.67 | 2,073.47 | 7,227.06 |
| NEBRASKA | 1,068.52 | 1,141.63 | 1,206.76 | 1,242.60 | 4,659.51 |
| NEVADA | 2,992.99 | 3,067.61 | 3,084.02 | 3,180.68 | 12,325.30 |
| NEW HAMPSHIRE | 6,520.85 | 6,788.14 | 7,440.04 | 7,538.47 | 28,287.50 |

DRUG CODE:9064 DRUG NAME:BUPRENORPHINE

| STATE | QUARTER 1 | QUARTER 2 | QUARTER 3 | QUARTER 4 | TOTAL GRAMS |
|---|---|---|---|---|---|
| NEW JERSEY | 24,457.81 | 25,316.15 | 25,374.39 | 25,516.96 | 100,665.31 |
| NEW MEXICO | 6,030.98 | 6,240.28 | 6,580.40 | 6,574.58 | 25,426.24 |
| NEW YORK | 46,638.26 | 48,481.23 | 49,364.14 | 49,422.72 | 193,906.35 |
| NORTH CAROLINA | 25,079.21 | 26,628.36 | 27,040.24 | 29,103.70 | 107,851.51 |
| NORTH DAKOTA | 739.83 | 839.93 | 863.99 | 841.42 | 3,285.17 |

| | | | | | |
|---|---|---|---|---|---|
| OHIO | 45,036.20 | 46,884.75 | 48,250.93 | 49,305.84 | 189,477.72 |
| OKLAHOMA | 9,072.36 | 9,408.18 | 9,562.58 | 9,916.89 | 37,960.01 |
| OREGON | 8,195.18 | 8,564.76 | 8,937.70 | 8,941.38 | 34,639.02 |
| PENNSYLVANIA | 53,463.22 | 55,467.91 | 56,676 | 58,545.98 | 224,153.11 |
| PUERTO RICO | 2,359.60 | 2,464.35 | 2,590.14 | 2,553.99 | 9,968.08 |
| RHODE ISLAND | 6,630.41 | 7,216.11 | 6,760.21 | 7,055.97 | 27,662.70 |
| SOUTH CAROLINA | 9,151.44 | 9,651.32 | 9,585.56 | 9,649.54 | 38,037.86 |
| SOUTH DAKOTA | 354.26 | 385.55 | 387.12 | 393.75 | 1,520.68 |
| TENNESSEE | 30,749.73 | 31,265.03 | 31,450.89 | 31,955.99 | 125,421.64 |
| TEXAS | 24,812.23 | 25,431.29 | 25,095.48 | 25,694.10 | 101,033.10 |
| UTAH | 8,562.28 | 8,743.78 | 9,050.73 | 9,205.49 | 35,562.28 |
| VERMONT | 4,990.87 | 5,326.35 | 5,530.96 | 5,690.02 | 21,538.20 |
| VIRGIN ISLANDS | 24.39 | 25.31 | 18.9 | 20.37 | 88.97 |
| VIRGINIA | 18,508.46 | 19,232.75 | 19,464.82 | 19,565.66 | 76,771.69 |
| WASHINGTON | 15,416.87 | 16,122.88 | 16,954.84 | 17,538.37 | 66,032.96 |
| WEST VIRGINIA | 14,234.01 | 14,516.57 | 14,577.49 | 14,881.13 | 58,209.20 |
| WISCONSIN | 9,122.76 | 9,361.62 | 9,558.03 | 9,507.34 | 37,549.75 |
| WYOMING | 881.02 | 1,045.43 | 996.46 | 1,093.46 | 4,016.37 |
| TOTAL | 716,844.78 | 742,556.95 | 758,402.96 | 776,302.93 | 2,994,107.62 |

DRUG CODE:9120 DRUG NAME:DIHYDROCODEINE

| STATE | QUARTER 1 | QUARTER 2 | QUARTER 3 | QUARTER 4 | TOTAL GRAMS |
|---|---|---|---|---|---|
| ALABAMA | 669.58 | 621.22 | 772.26 | 768.58 | 2,831.64 |
| ALASKA | 0 | 0 | 1.07 | 0 | 1.07 |
| ARIZONA | 49.14 | 39.63 | 62.17 | 71.03 | 221.97 |
| ARKANSAS | 104.89 | 97.53 | 84.12 | 103.18 | 389.72 |
| CALIFORNIA | 785.1 | 2,162.17 | 945.54 | 759.68 | 4,652.49 |
| COLORADO | 0 | 2.14 | 1.07 | 0 | 3.21 |
| CONNECTICUT | 0 | 0 | 0 | 3.2 | 3.2 |
| DELAWARE | 0 | 1.39 | 1.07 | 0 | 2.46 |
| FLORIDA | 67.13 | 99.14 | 87.7 | 110.5 | 364.47 |
| GEORGIA | 386.91 | 417.23 | 388.13 | 421.92 | 1,614.19 |
| HAWAII | 7.05 | 4.7 | 9.93 | 2.88 | 24.56 |
| IDAHO | 27.24 | 104.68 | 29.91 | 51.27 | 213.1 |
| ILLINOIS | 18.44 | 24.57 | 30.98 | 139.93 | 213.92 |
| INDIANA | 17.62 | 34.61 | 13.89 | 2.14 | 68.26 |
| KANSAS | 1.07 | 1.07 | 5.34 | 0 | 7.48 |
| KENTUCKY | 34.18 | 33.11 | 19.44 | 34.18 | 120.91 |
| LOUISIANA | 349.72 | 565.77 | 695.76 | 719.25 | 2,330.50 |
| MAINE | 0 | 0 | 1.07 | 0 | 1.07 |
| MARYLAND | 1.07 | 1.07 | 16.24 | 6.41 | 24.79 |

DRUG CODE:9120 DRUG NAME:DIHYDROCODEINE

| STATE | QUARTER 1 | QUARTER 2 | QUARTER 3 | QUARTER 4 | TOTAL GRAMS |
|---|---|---|---|---|---|
| MASSACHUSETTS | 19.23 | 12.82 | 9.61 | 9.61 | 51.27 |
| MICHIGAN | 96.13 | 1.07 | 3.2 | 8.55 | 108.95 |
| MINNESOTA | 0 | 0 | 3.2 | 0 | 3.2 |
| MISSISSIPPI | 222 | 246.94 | 363.92 | 320.88 | 1,153.74 |
| MISSOURI | 20.3 | 16.02 | 19.23 | 16.02 | 71.57 |
| NEBRASKA | 2.14 | 1.07 | 3.2 | 0 | 6.41 |
| NEVADA | 37.44 | 80.7 | 67.29 | 17.2 | 202.63 |
| NEW HAMPSHIRE | 0 | 0 | 1.07 | 0 | 1.07 |
| NEW JERSEY | 5.29 | 76.78 | 60.51 | 374.4 | 516.98 |
| NEW MEXICO | 150.61 | 143.13 | 160.22 | 162.36 | 616.32 |
| NEW YORK | 9.61 | 18.16 | 73.38 | 100.43 | 201.58 |
| NORTH CAROLINA | 9.83 | 12.82 | 9.61 | 4.27 | 36.53 |

| | | | | | |
|---|---|---|---|---|---|
| NORTH DAKOTA | 0 | 0 | 0 | 2.14 | 2.14 |
| OHIO | 32.26 | 28.84 | 42.73 | 30.98 | 134.81 |
| OKLAHOMA | 233.49 | 249.5 | 293.06 | 274.78 | 1,050.83 |
| PENNSYLVANIA | 217.48 | 298.12 | 274.52 | 341.53 | 1,131.65 |
| RHODE ISLAND | 0 | 1.07 | 5.34 | 0 | 6.41 |
| SOUTH CAROLINA | 18.59 | 20.3 | 9.83 | 12.82 | 61.54 |
| TENNESSEE | 752.45 | 1,083.95 | 1,334.72 | 1,072.44 | 4,243.56 |
| TEXAS | 2,192.45 | 2,368.64 | 2,834.27 | 2,860.20 | 10,255.56 |
| VIRGINIA | 8.55 | 7.48 | 9.61 | 5.34 | 30.98 |
| WASHINGTON | 0 | 1.07 | 0 | 0 | 1.07 |
| WEST VIRGINIA | 6.62 | 6.41 | 1.28 | 17.09 | 31.4 |
| WISCONSIN | 9.61 | 3.2 | 3.2 | 5.34 | 21.35 |
| TOTAL | 6,563.22 | 8,888.12 | 8,748.69 | 8,830.53 | 33,030.56 |

DRUG CODE:9143 DRUG NAME:OXYCODONE

| STATE | QUARTER 1 | QUARTER 2 | QUARTER 3 | QUARTER 4 | TOTAL GRAMS |
|---|---|---|---|---|---|
| ALABAMA | 274,766.65 | 267,370.75 | 266,149.79 | 254,659.47 | 1,062,946.66 |
| ALASKA | 39,324.91 | 37,514.02 | 36,811.81 | 36,680.19 | 150,330.93 |
| AMERICAN SAMOA | 25.55 | 43.03 | 32.27 | 32.27 | 133.12 |
| ARIZONA | 518,795.31 | 489,849.88 | 482,606.37 | 474,475.40 | 1,965,726.96 |
| ARKANSAS | 166,499.94 | 158,926.31 | 154,602.77 | 153,731.11 | 633,760.13 |
| CALIFORNIA | 1,023,490.12 | 985,543.82 | 954,884.87 | 944,148.97 | 3,908,067.78 |
| COLORADO | 263,173.69 | 251,756.92 | 249,353.64 | 241,868.95 | 1,006,153.20 |
| CONNECTICUT | 210,072.29 | 201,850.93 | 192,483.98 | 184,881.95 | 789,289.15 |
| DELAWARE | 78,425.44 | 74,988.74 | 75,259.79 | 74,154.80 | 302,828.77 |
| DISTRICT OF COLUMBIA | 17,625.73 | 16,433.41 | 16,992.05 | 15,898.43 | 66,949.62 |
| FLORIDA | 1,023,720.44 | 1,015,520.10 | 1,018,495.71 | 1,027,646.57 | 4,085,382.82 |
| GEORGIA | 459,497.96 | 454,500.01 | 461,288.09 | 459,019.65 | 1,834,305.71 |
| GUAM | 890.2 | 1,005.84 | 988.84 | 1,080.03 | 3,964.91 |
| HAWAII | 51,303.77 | 49,443.17 | 47,928.05 | 46,319.28 | 194,994.27 |
| IDAHO | 64,794.22 | 63,950.13 | 62,606.88 | 63,382.69 | 254,733.92 |
| ILLINOIS | 175,459.42 | 172,902.68 | 172,586.89 | 172,091.64 | 693,040.63 |
| INDIANA | 273,226.27 | 261,241 | 254,600.93 | 253,597.03 | 1,042,665.23 |
| IOWA | 63,120.31 | 60,800.26 | 59,226.67 | 58,237.44 | 241,384.68 |

DRUG CODE:9143 DRUG NAME:OXYCODONE

| STATE | QUARTER 1 | QUARTER 2 | QUARTER 3 | QUARTER 4 | TOTAL GRAMS |
|---|---|---|---|---|---|
| KANSAS | 133,872.94 | 131,243 | 132,130.18 | 128,509.97 | 525,756.09 |
| KENTUCKY | 223,364.15 | 220,004.68 | 216,906.11 | 212,366 | 872,640.94 |
| LOUISIANA | 215,928.93 | 214,907.66 | 212,387.49 | 208,022.47 | 851,246.55 |
| MAINE | 77,200.83 | 73,389.75 | 68,246.96 | 62,603.63 | 281,441.17 |
| MARYLAND | 370,096.54 | 359,576.09 | 360,809.38 | 351,896.45 | 1,442,378.46 |
| MASSACHUSETTS | 301,507.71 | 286,215.19 | 272,377.99 | 260,767.34 | 1,120,868.23 |
| MICHIGAN | 355,420.82 | 340,482.20 | 328,328.15 | 325,061.90 | 1,349,293.07 |
| MINNESOTA | 168,807.13 | 162,886.42 | 160,270.78 | 157,881.88 | 649,846.21 |
| MISSISSIPPI | 111,313.64 | 110,527.81 | 111,127.38 | 112,005.93 | 444,974.76 |
| MISSOURI | 340,293.77 | 332,323.79 | 333,994.33 | 320,680.07 | 1,327,291.96 |
| MONTANA | 48,783.16 | 46,367.68 | 44,930.58 | 42,922.10 | 183,003.52 |
| NEBRASKA | 45,165.04 | 43,874.51 | 44,869.24 | 44,296.22 | 178,205.01 |
| NEVADA | 204,105.68 | 193,511.26 | 190,928.20 | 191,848.53 | 780,393.67 |
| NEW HAMPSHIRE | 78,129.58 | 73,580.13 | 71,656.32 | 67,985.33 | 291,351.36 |
| NEW JERSEY | 613,725.46 | 595,927.61 | 582,829.11 | 563,479.86 | 2,355,962.04 |
| NEW MEXICO | 116,921.61 | 111,260.35 | 108,603.28 | 106,415.62 | 443,200.86 |
| NEW YORK | 906,793.40 | 867,363.24 | 844,521.01 | 827,407.97 | 3,446,085.62 |
| NORTH CAROLINA | 661,390.54 | 635,430.64 | 622,914.61 | 611,362.80 | 2,531,098.59 |
| NORTH DAKOTA | 19,123.64 | 18,270.88 | 16,988.71 | 14,989.26 | 69,372.49 |
| OHIO | 575,724.72 | 555,201.45 | 549,662.74 | 536,263.30 | 2,216,852.21 |

| | | | | | |
|---|---|---|---|---|---|
| OKLAHOMA | 246,389.97 | 237,970.32 | 238,248.74 | 235,451.93 | 958,060.96 |
| OREGON | 216,512.20 | 206,404.94 | 202,287.19 | 191,813.75 | 817,018.08 |
| PENNSYLVANIA | 903,045.67 | 882,716.43 | 852,812.35 | 817,502.99 | 3,456,077.44 |
| PUERTO RICO | 17,586.51 | 18,176.94 | 18,577.59 | 18,718.70 | 73,059.74 |
| RHODE ISLAND | 51,606.22 | 49,395.85 | 48,278.52 | 46,858.56 | 196,139.15 |
| SOUTH CAROLINA | 264,074.19 | 256,399.60 | 257,097.90 | 253,682.59 | 1,031,254.28 |
| SOUTH DAKOTA | 23,406.84 | 21,035.29 | 21,038.91 | 19,380.09 | 84,861.13 |
| TENNESSEE | 537,692.18 | 518,737.57 | 503,158.89 | 493,895.54 | 2,053,484.18 |
| TEXAS | 335,615.43 | 334,473.66 | 335,101.75 | 334,879.80 | 1,340,070.64 |
| UTAH | 194,466.12 | 186,517.93 | 187,356.68 | 180,232.47 | 748,573.20 |
| VERMONT | 27,098.07 | 26,242.60 | 25,878.67 | 24,702.56 | 103,921.90 |
| VIRGIN ISLANDS | 742.3 | 677.08 | 587.66 | 607.78 | 2,614.82 |
| VIRGINIA | 361,767.50 | 350,970.92 | 344,677.15 | 336,984.23 | 1,394,399.80 |
| WASHINGTON | 339,475.63 | 328,334.29 | 325,220.74 | 315,512.97 | 1,308,543.63 |
| WEST VIRGINIA | 117,089.06 | 108,551.11 | 104,418.63 | 97,988.40 | 428,047.20 |
| WISCONSIN | 285,763.88 | 268,219.38 | 262,448.01 | 250,561.22 | 1,066,992.49 |
| WYOMING | 30,570.90 | 29,105.63 | 29,018.27 | 28,156.90 | 116,851.70 |
| TOTAL | 14,224,784.18 | 13,759,914.88 | 13,537,589.60 | 13,255,602.98 | 54,777,891.64 |

DRUG CODE:9150 DRUG NAME:HYDROMORPHONE

| STATE | QUARTER 1 | QUARTER 2 | QUARTER 3 | QUARTER 4 | TOTAL GRAMS |
|---|---|---|---|---|---|
| ALABAMA | 4,000.30 | 4,046.83 | 3,838.99 | 3,621.56 | 15,507.68 |
| ALASKA | 1,120.52 | 1,062.81 | 1,038.18 | 1,085.28 | 4,306.79 |
| AMERICAN SAMOA | 0.61 | 1.95 | 0.06 | 0.53 | 3.15 |
| ARIZONA | 8,289.28 | 7,964.60 | 7,773.82 | 7,681.18 | 31,708.88 |
| ARKANSAS | 4,002.69 | 4,027.72 | 3,822.85 | 3,504.40 | 15,357.66 |

DRUG CODE:9150 DRUG NAME:HYDROMORPHONE

| STATE | QUARTER 1 | QUARTER 2 | QUARTER 3 | QUARTER 4 | TOTAL GRAMS |
|---|---|---|---|---|---|
| CALIFORNIA | 47,130.62 | 45,036.10 | 42,636.08 | 42,695.94 | 177,498.74 |
| COLORADO | 8,174.39 | 7,925.12 | 7,854.74 | 7,501.41 | 31,455.66 |
| CONNECTICUT | 7,476.60 | 7,240.17 | 6,823.38 | 7,158.73 | 28,698.88 |
| DELAWARE | 2,458.29 | 2,462.87 | 2,462.45 | 2,311.01 | 9,694.62 |
| DISTRICT OF COLUMBIA | 646.07 | 593.56 | 621.29 | 623.06 | 2,483.98 |
| FLORIDA | 55,925.93 | 52,939.64 | 50,358.13 | 51,019.55 | 210,243.25 |
| GEORGIA | 8,958.63 | 9,005.50 | 8,858.23 | 8,940.95 | 35,763.31 |
| GUAM | 18.14 | 37.35 | 36 | 34.63 | 126.12 |
| HAWAII | 845.41 | 771.81 | 770.66 | 727.4 | 3,115.28 |
| IDAHO | 2,131.49 | 2,180.38 | 1,895.63 | 2,038.73 | 8,246.23 |
| ILLINOIS | 10,951.63 | 11,047.15 | 10,869.07 | 10,564.30 | 43,432.15 |
| INDIANA | 7,657.12 | 7,180.89 | 6,961.89 | 6,606.41 | 28,406.31 |
| IOWA | 3,720.22 | 3,500.90 | 3,475.35 | 3,362.03 | 14,058.50 |
| KANSAS | 3,911.26 | 4,062.56 | 3,945 | 3,847.98 | 15,766.80 |
| KENTUCKY | 3,035.43 | 3,240.22 | 3,117.11 | 3,214.33 | 12,607.09 |
| LOUISIANA | 4,081.60 | 3,939.27 | 3,646.18 | 3,589.50 | 15,256.55 |
| MAINE | 2,101.31 | 1,927.72 | 1,699.82 | 1,975.09 | 7,703.94 |
| MARYLAND | 10,143.87 | 10,025.86 | 9,739.98 | 9,214.33 | 39,124.04 |
| MASSACHUSETTS | 8,484.78 | 7,902.22 | 7,894.04 | 7,802.04 | 32,083.08 |
| MICHIGAN | 11,218.34 | 11,190.78 | 9,931.93 | 10,246.52 | 42,587.57 |
| MINNESOTA | 6,335.69 | 6,194.15 | 6,050.83 | 6,541.19 | 25,121.86 |
| MISSISSIPPI | 5,260.84 | 5,257.86 | 5,026.87 | 5,190.63 | 20,736.20 |
| MISSOURI | 7,967 | 7,645.08 | 7,467.67 | 7,347.35 | 30,427.10 |
| MONTANA | 1,786.43 | 1,786.48 | 1,816.11 | 1,837.24 | 7,226.26 |
| NEBRASKA | 1,514.13 | 1,454.52 | 1,487.72 | 1,397.06 | 5,853.43 |
| NEVADA | 3,947.43 | 3,538.97 | 3,364.79 | 3,486.30 | 14,337.49 |
| NEW HAMPSHIRE | 2,133.72 | 2,062.59 | 2,011.35 | 2,177.77 | 8,385.43 |

| | | | | | |
|---|---|---|---|---|---|
| NEW JERSEY | 10,697.41 | 10,724.73 | 10,344.04 | 9,959.20 | 41,725.38 |
| NEW MEXICO | 2,002.56 | 1,882.59 | 1,780.58 | 1,643.50 | 7,309.23 |
| NEW YORK | 23,510.83 | 22,584.15 | 22,140.43 | 22,410.43 | 90,645.84 |
| NORTH CAROLINA | 12,402.38 | 12,680.16 | 12,239.47 | 12,299.60 | 49,621.61 |
| NORTH DAKOTA | 846.41 | 826.62 | 803.03 | 718.88 | 3,194.94 |
| OHIO | 10,524.08 | 10,627.32 | 9,930.90 | 10,104.20 | 41,186.50 |
| OKLAHOMA | 4,366.18 | 4,185.59 | 3,874.87 | 4,016.80 | 16,443.44 |
| OREGON | 5,850.26 | 5,703.10 | 5,470.93 | 5,288.04 | 22,312.33 |
| PENNSYLVANIA | 17,353.96 | 17,372.89 | 16,901.20 | 16,896.22 | 68,524.27 |
| PUERTO RICO | 66.34 | 74.65 | 81.02 | 64.93 | 286.94 |
| RHODE ISLAND | 845.69 | 881.33 | 889.21 | 924.11 | 3,540.34 |
| SOUTH CAROLINA | 6,313.97 | 6,125.16 | 6,068.57 | 6,046.35 | 24,554.05 |
| SOUTH DAKOTA | 827.81 | 686.49 | 631.32 | 588.07 | 2,733.69 |
| TENNESSEE | 7,690.71 | 6,823.15 | 7,208.62 | 7,262.17 | 28,984.65 |
| TEXAS | 26,568.54 | 25,656.57 | 26,451.41 | 26,109.40 | 104,785.92 |
| UTAH | 3,031.59 | 2,928.44 | 2,860.84 | 3,142.71 | 11,963.58 |
| VERMONT | 1,392.77 | 1,323.45 | 1,377.39 | 1,332.75 | 5,426.36 |
| VIRGIN ISLANDS | 29.97 | 31.21 | 27.27 | 13.5 | 101.95 |

DRUG CODE:9150 DRUG NAME:HYDROMORPHONE

| STATE | QUARTER 1 | QUARTER 2 | QUARTER 3 | QUARTER 4 | TOTAL GRAMS |
|---|---|---|---|---|---|
| VIRGINIA | 14,282.69 | 13,512.61 | 13,577.25 | 13,195.49 | 54,568.04 |
| WASHINGTON | 12,720.01 | 12,861.70 | 12,177.04 | 11,261.96 | 49,020.71 |
| WEST VIRGINIA | 1,799.82 | 2,019.72 | 1,708.60 | 1,761.07 | 7,289.21 |
| WISCONSIN | 4,373.83 | 4,285.81 | 3,992.48 | 4,198.98 | 16,851.10 |
| WYOMING | 734.68 | 734.38 | 731.43 | 716.96 | 2,917.45 |
| TOTAL | 413,662.26 | 401,785.45 | 388,564.10 | 387,299.75 | 1,591,311.56 |

DRUG CODE:9193 DRUG NAME:HYDROCODONE

| STATE | QUARTER 1 | QUARTER 2 | QUARTER 3 | QUARTER 4 | TOTAL GRAMS |
|---|---|---|---|---|---|
| ALABAMA | 284,939.67 | 273,947.56 | 273,503.18 | 268,059.60 | 1,100,450.01 |
| ALASKA | 13,980.11 | 13,334.53 | 13,066.05 | 12,451.09 | 52,831.78 |
| AMERICAN SAMOA | 7.26 | 14.53 | 7.26 | 3.63 | 32.68 |
| ARIZONA | 125,671.65 | 115,146.88 | 113,121.24 | 114,293.24 | 468,233.01 |
| ARKANSAS | 147,260.95 | 142,464.02 | 141,420.10 | 141,605.61 | 572,750.68 |
| CALIFORNIA | 1,063,613.79 | 1,019,734.43 | 999,972.16 | 985,412.68 | 4,068,733.06 |
| COLORADO | 76,405.95 | 71,346.63 | 71,454.19 | 70,346.62 | 289,553.39 |
| CONNECTICUT | 26,241.94 | 24,242.43 | 23,324.44 | 22,854.93 | 96,663.74 |
| DELAWARE | 7,317.81 | 6,884.45 | 6,934.76 | 6,753.40 | 27,890.42 |
| DISTRICT OF COLUMBIA | 1,778.53 | 1,831.17 | 1,686.29 | 1,579.83 | 6,875.82 |
| FLORIDA | 412,305.61 | 385,200.82 | 379,385.31 | 381,595 | 1,558,486.74 |
| GEORGIA | 280,755.57 | 268,310.93 | 266,689.99 | 265,247.55 | 1,081,004.04 |
| GUAM | 215.9 | 207.3 | 223.58 | 173.35 | 820.13 |
| HAWAII | 20,634.26 | 19,576.68 | 18,896.49 | 18,643.40 | 77,750.83 |
| IDAHO | 63,464.52 | 60,726.75 | 60,684.11 | 59,981.31 | 244,856.69 |
| ILLINOIS | 351,052.88 | 338,123.13 | 333,646.91 | 335,968.29 | 1,358,791.21 |
| INDIANA | 262,812.31 | 253,624.53 | 245,138.36 | 247,798.97 | 1,009,374.17 |
| IOWA | 67,892.51 | 65,631.67 | 64,035.04 | 63,027.24 | 260,586.46 |
| KANSAS | 92,010.35 | 88,113.64 | 87,859.03 | 86,860.59 | 354,843.61 |
| KENTUCKY | 225,172.11 | 220,379.43 | 217,482.66 | 211,648.85 | 874,683.05 |
| LOUISIANA | 187,264.81 | 181,384.19 | 182,071.51 | 179,685.40 | 730,405.91 |
| MAINE | 22,789.01 | 21,160.79 | 20,137.74 | 19,345.90 | 83,433.44 |
| MARYLAND | 42,332.56 | 40,044.35 | 39,475.87 | 38,860.81 | 160,713.59 |
| MASSACHUSETTS | 37,965.13 | 35,702.58 | 34,229.48 | 32,802.29 | 140,699.48 |
| MICHIGAN | 472,380.53 | 452,913.19 | 414,419.11 | 440,427.64 | 1,780,140.47 |
| MINNESOTA | 56,155.14 | 53,607.46 | 52,321.97 | 50,315.47 | 212,400.04 |
| MISSISSIPPI | 138,168.61 | 133,135.76 | 132,185.86 | 131,703.26 | 535,193.49 |

| | | | | | |
|---|---|---|---|---|---|
| MISSOURI | 203,341.57 | 196,226.07 | 194,450.36 | 190,776.89 | 784,794.89 |
| MONTANA | 29,371.91 | 28,614.64 | 28,747.29 | 27,921.84 | 114,655.68 |
| NEBRASKA | 37,853.66 | 36,559.11 | 36,272.04 | 35,634.77 | 146,319.58 |
| NEVADA | 111,506.63 | 104,247.40 | 102,475.96 | 101,689.18 | 419,919.17 |
| NEW HAMPSHIRE | 10,939.16 | 10,234.32 | 10,145.72 | 10,051.67 | 41,370.87 |
| NEW JERSEY | 44,287.97 | 41,371.03 | 40,276.69 | 40,650.85 | 166,586.54 |
| NEW MEXICO | 43,454.57 | 39,908.68 | 39,709.08 | 38,852.08 | 161,924.41 |
| NEW YORK | 211,983.53 | 199,865.69 | 191,900.69 | 189,356.87 | 793,106.78 |
| NORTH CAROLINA | 230,385.61 | 216,698.49 | 212,530.90 | 213,244.28 | 872,859.28 |
| NORTH DAKOTA | 13,407.04 | 12,708.75 | 11,785.33 | 10,758 | 48,659.12 |

DRUG CODE:9193 DRUG NAME:HYDROCODONE

| STATE | QUARTER 1 | QUARTER 2 | QUARTER 3 | QUARTER 4 | TOTAL GRAMS |
|---|---|---|---|---|---|
| OHIO | 226,679.94 | 220,737.60 | 212,338.61 | 207,982.24 | 867,738.39 |
| OKLAHOMA | 202,627.05 | 193,533.65 | 194,098.41 | 191,532.41 | 781,791.52 |
| OREGON | 113,457.84 | 109,658.72 | 106,180.91 | 102,407.98 | 431,705.45 |
| PENNSYLVANIA | 207,471.73 | 199,070.43 | 192,683.61 | 185,658.22 | 784,883.99 |
| PUERTO RICO | 1,313.73 | 1,078.07 | 990.88 | 1,028.34 | 4,411.02 |
| RHODE ISLAND | 13,202.04 | 12,584.98 | 12,428.17 | 12,066.99 | 50,282.18 |
| SOUTH CAROLINA | 162,672.66 | 152,976.17 | 151,124.12 | 150,913.58 | 617,686.53 |
| SOUTH DAKOTA | 18,498.59 | 17,589.27 | 17,091.37 | 16,525.42 | 69,704.65 |
| TENNESSEE | 287,485.49 | 274,704.43 | 270,732.16 | 263,879.34 | 1,096,801.42 |
| TEXAS | 746,430.71 | 717,354.31 | 715,839.49 | 705,047.72 | 2,884,672.23 |
| UTAH | 68,345.84 | 65,234.01 | 64,683.78 | 62,759.48 | 261,023.11 |
| VERMONT | 6,130.77 | 5,935.17 | 5,675.48 | 5,597.59 | 23,339.01 |
| VIRGIN ISLANDS | 436.02 | 363.59 | 313.07 | 275.43 | 1,388.11 |
| VIRGINIA | 133,185.85 | 125,621.56 | 122,170.05 | 121,335.87 | 502,313.33 |
| WASHINGTON | 153,272.99 | 149,074.82 | 145,865.50 | 141,648.94 | 589,862.25 |
| WEST VIRGINIA | 76,419.42 | 72,355.17 | 69,569.37 | 67,022.10 | 285,366.06 |
| WISCONSIN | 95,261.47 | 91,111.36 | 89,598.46 | 87,402.10 | 363,373.39 |
| WYOMING | 14,181.75 | 13,512.73 | 13,178.74 | 12,974.75 | 53,847.97 |
| TOTAL | 7,944,195.01 | 7,595,720.05 | 7,446,228.93 | 7,382,440.88 | 30,368,584.87 |

DRUG CODE:9220L DRUG NAME:LEVORPHANOL

| STATE | QUARTER 1 | QUARTER 2 | QUARTER 3 | QUARTER 4 | TOTAL GRAMS |
|---|---|---|---|---|---|
| ALABAMA | 6.73 | 4.41 | 4.64 | 4.06 | 19.84 |
| ARIZONA | 1.74 | 5.11 | 9.98 | 22.75 | 39.58 |
| ARKANSAS | 0.58 | 1.97 | 2.9 | 2.44 | 7.89 |
| CALIFORNIA | 82.53 | 95.77 | 122 | 96.23 | 396.53 |
| COLORADO | 2.21 | 6.15 | 5.22 | 5.92 | 19.5 |
| CONNECTICUT | 0.35 | 2.21 | 1.86 | 0.93 | 5.35 |
| FLORIDA | 8.01 | 21.94 | 27.05 | 22.17 | 79.17 |
| GEORGIA | 0.93 | 1.16 | 1.28 | 1.16 | 4.53 |
| ILLINOIS | 7.2 | 6.38 | 6.73 | 8.71 | 29.02 |
| INDIANA | 25.65 | 24.38 | 27.16 | 24.03 | 101.22 |
| IOWA | 5.69 | 4.88 | 4.18 | 4.29 | 19.04 |
| KANSAS | 6.62 | 7.66 | 8.71 | 9.75 | 32.74 |
| LOUISIANA | 4.76 | 8.59 | 13.23 | 14.63 | 41.21 |
| MAINE | 0 | 0 | 0 | 0.23 | 0.23 |
| MARYLAND | 0.12 | 0.46 | 0.35 | 3.13 | 4.06 |
| MASSACHUSETTS | 21.82 | 18.92 | 15.9 | 18.69 | 75.33 |
| MICHIGAN | 0.23 | 1.16 | 0.93 | 1.28 | 3.6 |
| MINNESOTA | 0.35 | 0.12 | 1.63 | 0.46 | 2.56 |
| MISSISSIPPI | 1.04 | 1.39 | 0.23 | 0.35 | 3.01 |
| MISSOURI | 4.88 | 5.34 | 9.17 | 11.49 | 30.88 |
| NEBRASKA | 0.35 | 0.35 | 1.04 | 1.86 | 3.6 |

| | | | | | |
|---|---|---|---|---|---|
| NEVADA | 0.58 | 3.48 | 2.09 | 2.32 | 8.47 |
| NEW HAMPSHIRE | 1.28 | 0.93 | 0.35 | 0.81 | 3.37 |
| NEW JERSEY | 43.53 | 97.39 | 185.26 | 268.49 | 594.67 |

DRUG CODE:9220L DRUG NAME:LEVORPHANOL

| STATE | QUARTER 1 | QUARTER 2 | QUARTER 3 | QUARTER 4 | TOTAL GRAMS |
|---|---|---|---|---|---|
| NEW MEXICO | 0 | 0 | 0 | 0.12 | 0.12 |
| NEW YORK | 35.75 | 22.4 | 27.51 | 50.26 | 135.92 |
| NORTH CAROLINA | 19.85 | 17.06 | 35.06 | 40.51 | 112.48 |
| OHIO | 1.28 | 2.55 | 8.24 | 12.54 | 24.61 |
| OKLAHOMA | 0.46 | 0.7 | 0.35 | 0.35 | 1.86 |
| OREGON | 6.62 | 4.64 | 4.06 | 3.83 | 19.15 |
| PENNSYLVANIA | 9.87 | 10.45 | 8.36 | 10.1 | 38.78 |
| SOUTH CAROLINA | 0.7 | 0 | 0.46 | 0.7 | 1.86 |
| TENNESSEE | 0.35 | 0.35 | 0.12 | 0.12 | 0.94 |
| TEXAS | 22.4 | 23.56 | 36.8 | 43.41 | 126.17 |
| UTAH | 0.35 | 0 | 0 | 0 | 0.35 |
| VERMONT | 0.58 | 0.35 | 0.46 | 0.58 | 1.97 |
| VIRGINIA | 7.43 | 4.64 | 5.46 | 5.8 | 23.33 |
| WASHINGTON | 13.35 | 13 | 13 | 15.44 | 54.79 |
| WEST VIRGINIA | 0.12 | 0.35 | 0.35 | 0.23 | 1.05 |
| WISCONSIN | 3.13 | 3.48 | 2.9 | 3.37 | 12.88 |
| TOTAL | 349.42 | 423.68 | 595.02 | 713.54 | 2,081.66 |

DRUG CODE:9230 DRUG NAME:MEPERIDINE (PETHIDINE)

| STATE | QUARTER 1 | QUARTER 2 | QUARTER 3 | QUARTER 4 | TOTAL GRAMS |
|---|---|---|---|---|---|
| ALABAMA | 9,863.53 | 9,491.42 | 8,853.81 | 17,913.73 | 46,122.49 |
| ALASKA | 795.57 | 913.55 | 779.12 | 821.39 | 3,309.63 |
| AMERICAN SAMOA | 6.1 | 0 | 0 | 0 | 6.1 |
| ARIZONA | 6,303.32 | 6,517.71 | 6,245.82 | 5,702.53 | 24,769.38 |
| ARKANSAS | 9,943.79 | 9,512.27 | 9,662.76 | 9,077.54 | 38,196.36 |
| CALIFORNIA | 17,872.11 | 18,691.50 | 16,720.01 | 17,490.20 | 70,773.82 |
| COLORADO | 2,541.51 | 2,387.26 | 2,410.57 | 2,258.06 | 9,597.40 |
| CONNECTICUT | 1,461.61 | 1,278.93 | 1,280.45 | 1,335.79 | 5,356.78 |
| DELAWARE | 236.09 | 241.49 | 233.15 | 223.47 | 934.2 |
| DISTRICT OF COLUMBIA | 183.67 | 159.81 | 208.29 | 245.33 | 797.1 |
| FLORIDA | 10,869.33 | 10,812.16 | 10,126.07 | 10,255.05 | 42,062.61 |
| GEORGIA | 13,237.10 | 12,377.72 | 12,066.46 | 12,250.78 | 49,932.06 |
| GUAM | 34.32 | 70.37 | 9.59 | 43.58 | 157.86 |
| HAWAII | 495.56 | 496.97 | 389.23 | 384.66 | 1,766.42 |
| IDAHO | 968 | 921.81 | 1,001.79 | 835.38 | 3,726.98 |
| ILLINOIS | 3,948.42 | 3,816.18 | 3,615.77 | 3,391.90 | 14,772.27 |
| INDIANA | 4,153.55 | 3,557.92 | 3,503.43 | 3,812.70 | 15,027.60 |
| IOWA | 1,884.51 | 2,056.65 | 1,892.05 | 1,996.58 | 7,829.79 |
| KANSAS | 1,917.67 | 2,074.87 | 2,018.33 | 1,977.83 | 7,988.70 |
| KENTUCKY | 3,047.48 | 2,975.08 | 2,906.67 | 2,779.24 | 11,708.47 |
| LOUISIANA | 10,405.88 | 9,237.97 | 9,099.99 | 8,580.96 | 37,324.80 |
| MAINE | 594.36 | 480.96 | 455.03 | 478.45 | 2,008.80 |
| MARYLAND | 2,923.71 | 3,056.02 | 2,858.19 | 2,731.43 | 11,569.35 |
| MASSACHUSETTS | 2,743.42 | 2,883.36 | 2,641.60 | 2,597.51 | 10,865.89 |
| MICHIGAN | 3,863.64 | 3,754.71 | 3,483.63 | 3,410.18 | 14,512.16 |
| MINNESOTA | 839.36 | 843.96 | 787.44 | 857.71 | 3,328.47 |

DRUG CODE:9230 DRUG NAME:MEPERIDINE (PETHIDINE)

| STATE | QUARTER 1 | QUARTER 2 | QUARTER 3 | QUARTER 4 | TOTAL GRAMS |
|---|---|---|---|---|---|
| MISSISSIPPI | 5,884.41 | 6,028.47 | 5,533.98 | 5,280.48 | 22,727.34 |

| | | | | | |
|---|---|---|---|---|---|
| MISSOURI | 5,256.40 | 4,887.54 | 4,965.04 | 4,392.13 | 19,501.11 |
| MONTANA | 706.13 | 926.84 | 829.01 | 807.01 | 3,268.99 |
| NEBRASKA | 810.26 | 1,075.10 | 759.25 | 816.9 | 3,461.51 |
| NEVADA | 1,857.38 | 1,832.90 | 1,791.15 | 1,566.41 | 7,047.84 |
| NEW HAMPSHIRE | 741.43 | 756.35 | 730.21 | 672.8 | 2,900.79 |
| NEW JERSEY | 2,557.96 | 2,363.07 | 2,271.56 | 2,357.95 | 9,550.54 |
| NEW MEXICO | 1,945.56 | 1,968.33 | 1,931.57 | 1,635.26 | 7,480.72 |
| NEW YORK | 5,872.91 | 5,693.57 | 5,636.40 | 5,077.86 | 22,280.74 |
| NORTH CAROLINA | 7,886.66 | 7,740.66 | 7,378.01 | 6,499.65 | 29,504.98 |
| NORTH DAKOTA | 302.3 | 319.73 | 251.21 | 261.01 | 1,134.25 |
| OHIO | 5,667.73 | 5,731.53 | 5,977.79 | 5,181.91 | 22,558.96 |
| OKLAHOMA | 9,325.29 | 9,092.62 | 8,952.72 | 8,430.89 | 35,801.52 |
| OREGON | 1,812.72 | 1,846.03 | 1,826.99 | 1,776.44 | 7,262.18 |
| PENNSYLVANIA | 4,705.64 | 4,106.14 | 4,716.99 | 4,228.30 | 17,757.07 |
| PUERTO RICO | 6,561.09 | 6,693.88 | 6,497.47 | 5,488.92 | 25,241.36 |
| RHODE ISLAND | 236.29 | 220.71 | 251.75 | 242.06 | 950.81 |
| SOUTH CAROLINA | 8,645.54 | 7,721.71 | 7,290.32 | 6,832.78 | 30,490.35 |
| SOUTH DAKOTA | 710.6 | 675.09 | 631.73 | 539.79 | 2,557.21 |
| TENNESSEE | 10,540.55 | 9,215.92 | 9,354.33 | 8,516.71 | 37,627.51 |
| TEXAS | 14,760.68 | 14,745.08 | 14,129.83 | 12,976.44 | 56,612.03 |
| UTAH | 2,377.84 | 2,213.65 | 2,319.80 | 2,191.56 | 9,102.85 |
| VERMONT | 385.53 | 368.97 | 349.36 | 327.79 | 1,431.65 |
| VIRGIN ISLANDS | 41.4 | 79.42 | 77.35 | 39.22 | 237.39 |
| VIRGINIA | 7,792.67 | 7,750.60 | 7,427.66 | 6,901.45 | 29,872.38 |
| WASHINGTON | 4,505.66 | 4,639.26 | 3,872.29 | 3,911.92 | 16,929.13 |
| WEST VIRGINIA | 783.26 | 877.1 | 732.87 | 836.1 | 3,229.33 |
| WISCONSIN | 1,939.09 | 2,043.71 | 1,922.75 | 1,764.35 | 7,669.90 |
| WYOMING | 565.02 | 474.44 | 479.48 | 481.94 | 2,000.88 |
| TOTAL | 226,311.61 | 220,699.07 | 212,138.12 | 211,488.01 | 870,636.81 |

DRUG CODE:9250B DRUG NAME:METHADONE

| STATE | QUARTER 1 | QUARTER 2 | QUARTER 3 | QUARTER 4 | TOTAL GRAMS |
|---|---|---|---|---|---|
| ALABAMA | 90,150.65 | 87,839.87 | 81,598.36 | 76,436.84 | 336,025.72 |
| ALASKA | 8,581.05 | 8,547.15 | 7,450.80 | 7,626.82 | 32,205.82 |
| ARIZONA | 86,983.13 | 87,632.62 | 86,026.72 | 73,132.44 | 333,774.91 |
| ARKANSAS | 20,512.89 | 20,654.12 | 19,332.81 | 17,086.44 | 77,586.26 |
| CALIFORNIA | 378,442.79 | 373,265.70 | 365,655.83 | 283,719.23 | 1,401,083.55 |
| COLORADO | 38,408.68 | 42,118.25 | 41,172.42 | 36,894.43 | 158,593.78 |
| CONNECTICUT | 113,540.77 | 111,568.35 | 97,930.72 | 102,286.31 | 425,326.15 |
| DELAWARE | 26,996.22 | 27,744.74 | 27,469.62 | 18,972.46 | 101,183.04 |
| DISTRICT OF COLUMBIA | 10,847.80 | 11,521.62 | 10,924.87 | 7,151.94 | 40,446.23 |
| FLORIDA | 214,970.74 | 220,635.09 | 212,666.56 | 198,175.73 | 846,448.12 |
| GEORGIA | 116,386.62 | 116,562.61 | 118,829.22 | 110,386.25 | 462,164.70 |
| GUAM | 51.88 | 39.35 | 47.4 | 25.94 | 164.57 |
| HAWAII | 9,491.64 | 7,971.97 | 9,232.18 | 6,909.33 | 33,605.12 |

DRUG CODE:9250B DRUG NAME:METHADONE

| STATE | QUARTER 1 | QUARTER 2 | QUARTER 3 | QUARTER 4 | TOTAL GRAMS |
|---|---|---|---|---|---|
| IDAHO | 13,789.50 | 13,766.07 | 13,037.22 | 10,503.57 | 51,096.36 |
| ILLINOIS | 125,538.52 | 124,963.87 | 122,763.30 | 103,436.88 | 476,702.57 |
| INDIANA | 92,151.04 | 91,777.09 | 89,242.59 | 70,676.75 | 343,847.47 |
| IOWA | 13,850.88 | 15,005.44 | 13,245.35 | 11,350.20 | 53,451.87 |
| KANSAS | 24,699.57 | 25,215.37 | 23,570.75 | 22,335.67 | 95,821.36 |
| KENTUCKY | 51,685.30 | 51,552.02 | 53,083.01 | 50,893.49 | 207,213.82 |
| LOUISIANA | 37,174.75 | 40,508.28 | 37,692.52 | 37,863.80 | 153,239.35 |
| MAINE | 38,850.05 | 38,904.70 | 37,081.47 | 26,579.96 | 141,416.18 |
| MARYLAND | 187,651.24 | 188,535.65 | 181,435.89 | 154,246.97 | 711,869.75 |

| | | | | | |
|---|---|---|---|---|---|
| MASSACHUSETTS | 142,094.34 | 148,189.44 | 144,690.13 | 106,370.52 | 541,344.43 |
| MICHIGAN | 128,566.76 | 138,898.30 | 128,267.79 | 117,219.93 | 512,952.78 |
| MINNESOTA | 58,702.53 | 57,802.93 | 52,650.47 | 51,855.50 | 221,011.43 |
| MISSISSIPPI | 7,125.15 | 7,083.47 | 6,606.04 | 6,459.09 | 27,273.75 |
| MISSOURI | 42,648.71 | 44,313 | 42,209.39 | 37,482.94 | 166,654.04 |
| MONTANA | 8,043.97 | 7,254.57 | 7,462.34 | 6,956.37 | 29,717.25 |
| NEBRASKA | 9,116.31 | 8,638.79 | 8,845.80 | 8,402.62 | 35,003.52 |
| NEVADA | 40,402.91 | 39,167.37 | 36,544.02 | 38,026.98 | 154,141.28 |
| NEW HAMPSHIRE | 30,968.96 | 33,316.13 | 31,380.92 | 24,946.25 | 120,612.26 |
| NEW JERSEY | 111,253.90 | 114,236.81 | 109,004.08 | 101,690.31 | 436,185.10 |
| NEW MEXICO | 39,112.74 | 40,540.83 | 41,641.65 | 31,724.28 | 153,019.50 |
| NEW YORK | 290,628.24 | 310,553.41 | 303,484.52 | 249,598.38 | 1,154,264.55 |
| NORTH CAROLINA | 136,067.71 | 134,817.12 | 133,818.69 | 120,572.36 | 525,275.88 |
| NORTH DAKOTA | 1,425.67 | 1,246.61 | 1,451.70 | 992.96 | 5,116.94 |
| OHIO | 77,731.17 | 77,763.24 | 78,242.57 | 77,227.45 | 310,964.43 |
| OKLAHOMA | 43,826.76 | 41,654.35 | 38,338.55 | 29,341.15 | 153,160.81 |
| OREGON | 69,185.59 | 69,289.76 | 67,677.01 | 55,756.83 | 261,909.19 |
| PENNSYLVANIA | 201,406.22 | 198,758.95 | 191,704.54 | 172,560.95 | 764,430.66 |
| PUERTO RICO | 17,747.13 | 21,382.33 | 19,453.65 | 16,098.93 | 74,682.04 |
| RHODE ISLAND | 44,573.59 | 40,493.07 | 43,773.72 | 40,961.64 | 169,802.02 |
| SOUTH CAROLINA | 60,724.04 | 57,403.84 | 56,331.37 | 47,687.53 | 222,146.78 |
| SOUTH DAKOTA | 2,557.72 | 2,620.77 | 2,361.39 | 2,282.87 | 9,822.75 |
| TENNESSEE | 52,895.26 | 54,107.18 | 51,091.17 | 46,116.29 | 204,209.90 |
| TEXAS | 144,498.29 | 147,899.26 | 141,915.85 | 124,209.53 | 558,522.93 |
| UTAH | 27,818.43 | 32,177.65 | 26,724.85 | 23,855.71 | 110,576.64 |
| VERMONT | 16,978.04 | 19,299.09 | 16,904.52 | 14,998.73 | 68,180.38 |
| VIRGIN ISLANDS | 36.22 | 44.27 | 47.4 | 37.12 | 165.01 |
| VIRGINIA | 74,972.95 | 75,366.83 | 75,162.21 | 66,097.77 | 291,599.76 |
| WASHINGTON | 108,623 | 108,528.37 | 106,589.58 | 74,218.39 | 397,959.34 |
| WEST VIRGINIA | 27,482.05 | 27,732.48 | 25,623.22 | 19,701.31 | 100,539.06 |
| WISCONSIN | 70,027.36 | 68,746.26 | 68,324.78 | 48,624.28 | 255,722.68 |
| WYOMING | 1,562.25 | 1,465.74 | 1,419.95 | 1,340.53 | 5,788.47 |
| TOTAL | 3,789,559.68 | 3,837,122.15 | 3,709,233.48 | 3,190,106.95 | 14,526,022.26 |

DRUG CODE:9300 DRUG NAME:MORPHINE

| STATE | QUARTER 1 | QUARTER 2 | QUARTER 3 | QUARTER 4 | TOTAL GRAMS |
|---|---|---|---|---|---|
| ALABAMA | 87,559.97 | 85,722 | 84,192.35 | 81,395.61 | 338,869.93 |

DRUG CODE:9300 DRUG NAME:MORPHINE

| STATE | QUARTER 1 | QUARTER 2 | QUARTER 3 | QUARTER 4 | TOTAL GRAMS |
|---|---|---|---|---|---|
| ALASKA | 14,575.68 | 13,615.13 | 12,956.34 | 12,261.06 | 53,408.21 |
| AMERICAN SAMOA | 30.46 | 41.36 | 62.79 | 14.29 | 148.9 |
| ARIZONA | 212,730.17 | 200,976.47 | 197,094.19 | 189,645.98 | 800,446.81 |
| ARKANSAS | 53,290.60 | 51,436.69 | 52,219.30 | 49,452.71 | 206,399.30 |
| CALIFORNIA | 546,859.18 | 525,805.77 | 507,899.86 | 493,476.81 | 2,074,041.62 |
| COLORADO | 95,441.22 | 91,267.56 | 87,868.85 | 83,789.97 | 358,367.60 |
| CONNECTICUT | 43,162.16 | 41,322.83 | 40,286.48 | 39,673.28 | 164,444.75 |
| DELAWARE | 23,272.07 | 23,189.88 | 22,445.06 | 22,492.61 | 91,399.62 |
| DISTRICT OF COLUMBIA | 4,806.83 | 4,021.53 | 4,630.71 | 4,051.22 | 17,510.29 |
| FLORIDA | 452,558.77 | 437,377.71 | 428,194.85 | 420,521.74 | 1,738,653.07 |
| GEORGIA | 123,802.38 | 119,982.56 | 120,392.94 | 119,433.28 | 483,611.16 |
| GUAM | 1,003.37 | 1,290.67 | 1,276.39 | 1,225.44 | 4,795.87 |
| HAWAII | 27,043.16 | 26,146.20 | 25,567.12 | 24,785.60 | 103,542.08 |
| IDAHO | 32,394.45 | 32,042.87 | 30,616.17 | 30,039.31 | 125,092.80 |
| ILLINOIS | 114,128.68 | 113,278.26 | 113,216.63 | 110,151.80 | 450,775.37 |
| INDIANA | 110,039.84 | 103,584.16 | 100,444.17 | 96,118.99 | 410,187.16 |

| | | | | | |
|---|---|---|---|---|---|
| IOWA | 40,265.51 | 38,741.31 | 38,031.12 | 36,207.51 | 153,245.45 |
| KANSAS | 51,668.51 | 50,873.98 | 51,785.19 | 47,129.13 | 201,456.81 |
| KENTUCKY | 55,610.88 | 53,356.74 | 53,450.56 | 50,547.97 | 212,966.15 |
| LOUISIANA | 65,999.75 | 64,437.16 | 62,671.95 | 58,629.31 | 251,738.17 |
| MAINE | 23,946.80 | 23,795.48 | 21,781.48 | 19,454.98 | 88,978.74 |
| MARYLAND | 91,129.60 | 90,198.47 | 88,365.55 | 85,660.62 | 355,354.24 |
| MASSACHUSETTS | 78,799.20 | 75,586.77 | 72,799.50 | 71,692.67 | 298,878.14 |
| MICHIGAN | 230,877 | 223,336.24 | 220,771.25 | 211,899.40 | 886,883.89 |
| MINNESOTA | 55,061.49 | 54,552.69 | 53,711.43 | 51,280.68 | 214,606.29 |
| MISSISSIPPI | 43,699.20 | 43,388.37 | 43,305.50 | 43,184.43 | 173,577.50 |
| MISSOURI | 112,703.26 | 111,942.81 | 108,900.38 | 104,732.67 | 438,279.12 |
| MONTANA | 22,737.14 | 22,143.46 | 21,128.21 | 19,841.85 | 85,850.66 |
| NEBRASKA | 20,279.51 | 20,070.88 | 20,209.84 | 19,813.70 | 80,373.93 |
| NEVADA | 76,792.94 | 72,807.75 | 69,552.86 | 65,669.52 | 284,823.07 |
| NEW HAMPSHIRE | 20,827.92 | 20,085.82 | 19,847.24 | 18,956.44 | 79,717.42 |
| NEW JERSEY | 100,632.87 | 98,757.82 | 98,221.95 | 98,649.14 | 396,261.78 |
| NEW MEXICO | 36,307.65 | 34,218.42 | 32,301.25 | 30,702.66 | 133,529.98 |
| NEW YORK | 190,919.53 | 186,932 | 178,732.72 | 173,972.02 | 730,556.27 |
| NORTH CAROLINA | 144,656.19 | 139,634.61 | 137,422.37 | 130,778.18 | 552,491.35 |
| NORTH DAKOTA | 8,534.45 | 8,277.40 | 7,198.59 | 6,507.45 | 30,517.89 |
| OHIO | 140,275.87 | 137,919.56 | 136,397.30 | 129,154.39 | 543,747.12 |
| OKLAHOMA | 108,630.45 | 104,947.58 | 102,826.22 | 99,334.30 | 415,738.55 |
| OREGON | 97,265.96 | 93,258.76 | 88,153.64 | 81,754.40 | 360,432.76 |
| PENNSYLVANIA | 268,518.94 | 266,791.88 | 253,808.18 | 245,258.26 | 1,034,377.26 |
| PUERTO RICO | 2,876.37 | 3,220.17 | 2,925.75 | 2,897.33 | 11,919.62 |
| RHODE ISLAND | 12,892.74 | 12,776.05 | 12,939.67 | 13,334.75 | 51,943.21 |
| SOUTH CAROLINA | 74,018.27 | 72,199.07 | 71,729.79 | 71,198.10 | 289,145.23 |
| SOUTH DAKOTA | 11,949.50 | 11,024.73 | 11,025.31 | 9,661.77 | 43,661.31 |
| TENNESSEE | 206,227.90 | 196,589.50 | 186,818.56 | 174,323.12 | 763,959.08 |

DRUG CODE:9300 DRUG NAME:MORPHINE

| STATE | QUARTER 1 | QUARTER 2 | QUARTER 3 | QUARTER 4 | TOTAL GRAMS |
|---|---|---|---|---|---|
| TEXAS | 233,538.26 | 228,471.87 | 227,540.83 | 228,634.30 | 918,185.26 |
| UTAH | 58,129.17 | 56,373.91 | 55,893.29 | 53,164.46 | 223,560.83 |
| VERMONT | 11,871.04 | 11,498.71 | 10,882.67 | 10,915.41 | 45,167.83 |
| VIRGIN ISLANDS | 133.25 | 93.11 | 103.7 | 86.71 | 416.77 |
| VIRGINIA | 94,854.15 | 90,562.28 | 89,881.59 | 88,063.52 | 363,361.54 |
| WASHINGTON | 133,782.85 | 129,583.14 | 126,491.79 | 120,227.86 | 510,085.64 |
| WEST VIRGINIA | 29,110.04 | 28,658.53 | 27,267.05 | 25,854.69 | 110,890.31 |
| WISCONSIN | 102,451.84 | 99,021.25 | 95,605.97 | 88,113.03 | 385,192.09 |
| WYOMING | 11,155.59 | 10,239.70 | 10,444.80 | 9,333.30 | 41,173.39 |
| TOTAL | 5,011,830.58 | 4,857,469.63 | 4,740,289.25 | 4,575,149.73 | 19,184,739.19 |

DRUG CODE:9630 DRUG NAME:OPIUM TINCTURE (LAUDANUM)

| STATE | QUARTER 1 | QUARTER 2 | QUARTER 3 | QUARTER 4 | TOTAL GRAMS |
|---|---|---|---|---|---|
| ARIZONA | 0 | 0 | 0 | 9.46 | 9.46 |
| CALIFORNIA | 4.73 | 29.56 | 11.82 | 0 | 46.11 |
| COLORADO | 0 | 0 | 1.18 | 0 | 1.18 |
| DISTRICT OF COLUMBIA | 0 | 0 | 1.18 | 0 | 1.18 |
| FLORIDA | 14.19 | 4.73 | 4.73 | 14.19 | 37.84 |
| IDAHO | 0 | 1.18 | 0 | 0 | 1.18 |
| ILLINOIS | 10.64 | 5.9 | 0 | 17.73 | 34.27 |
| INDIANA | 15.36 | 35.47 | 14.19 | 16.55 | 81.57 |
| KANSAS | 0 | 0 | 0 | 5.91 | 5.91 |
| MAINE | 0 | 1.18 | 4.73 | 0 | 5.91 |

| | | | | | |
|---|---|---|---|---|---|
| MARYLAND | 2.36 | 7.08 | 0 | 1.18 | 10.62 |
| MASSACHUSETTS | 5.91 | 37.84 | 53.21 | 53.21 | 150.17 |
| MICHIGAN | 0 | 20.1 | 2.36 | 24.83 | 47.29 |
| MINNESOTA | 0 | 0 | 1.18 | 0 | 1.18 |
| MISSOURI | 0 | 0 | 4.73 | 21.28 | 26.01 |
| NEBRASKA | 0 | 0 | 14.19 | 0 | 14.19 |
| NEW HAMPSHIRE | 0 | 1.18 | 0 | 0 | 1.18 |
| NEW JERSEY | 2.36 | 2.36 | 1.18 | 3.54 | 9.44 |
| NEW MEXICO | 0 | 0 | 0 | 4.73 | 4.73 |
| NEW YORK | 2.36 | 0 | 1.18 | 0 | 3.54 |
| OHIO | 15.37 | 34.29 | 26.01 | 21.25 | 96.92 |
| OREGON | 0 | 0 | 0 | 4.72 | 4.72 |
| PENNSYLVANIA | 5.91 | 1.18 | 0 | 5.9 | 12.99 |
| TEXAS | 2.36 | 1.18 | 9.44 | 8.26 | 21.24 |
| VERMONT | 0 | 0 | 2.36 | 0 | 2.36 |
| VIRGINIA | 0 | 9.44 | 5.9 | 2.36 | 17.7 |
| WASHINGTON | 3.54 | 4.72 | 5.9 | 5.9 | 20.06 |
| WEST VIRGINIA | 5.91 | 4.73 | 0 | 9.45 | 20.09 |
| WISCONSIN | 0 | 0 | 0 | 1.18 | 1.18 |
| TOTAL | 91 | 202.12 | 165.47 | 231.63 | 690.22 |

DRUG CODE:9639 DRUG NAME:OPIUM POWDERED

| STATE | QUARTER 1 | QUARTER 2 | QUARTER 3 | QUARTER 4 | TOTAL GRAMS |
|---|---|---|---|---|---|
| ALABAMA | 90.36 | 89.64 | 96.84 | 102.24 | 379.08 |

DRUG CODE:9639 DRUG NAME:OPIUM POWDERED

| STATE | QUARTER 1 | QUARTER 2 | QUARTER 3 | QUARTER 4 | TOTAL GRAMS |
|---|---|---|---|---|---|
| ALASKA | 14.4 | 21.24 | 16.92 | 25.2 | 77.76 |
| ARIZONA | 240.48 | 250.92 | 308.88 | 308.16 | 1,108.44 |
| ARKANSAS | 90.36 | 104.76 | 101.52 | 94.68 | 391.32 |
| CALIFORNIA | 470.16 | 507.24 | 479.52 | 455.4 | 1,912.32 |
| COLORADO | 256.68 | 214.2 | 217.08 | 215.64 | 903.6 |
| CONNECTICUT | 107.28 | 85.68 | 82.44 | 88.92 | 364.32 |
| DELAWARE | 24.48 | 24.12 | 17.64 | 11.88 | 78.12 |
| DISTRICT OF COLUMBIA | 12.24 | 10.8 | 6.12 | 17.28 | 46.44 |
| FLORIDA | 482.4 | 463.68 | 424.44 | 501.84 | 1,872.36 |
| GEORGIA | 190.08 | 188.28 | 205.92 | 195.84 | 780.12 |
| GUAM | 0.72 | 0.72 | 1.44 | 0 | 2.88 |
| HAWAII | 25.92 | 37.44 | 20.88 | 16.56 | 100.8 |
| IDAHO | 53.64 | 45.72 | 38.88 | 43.2 | 181.44 |
| ILLINOIS | 205.56 | 188.64 | 213.84 | 198.36 | 806.4 |
| INDIANA | 241.56 | 279 | 271.44 | 239.4 | 1,031.40 |
| IOWA | 81.36 | 107.28 | 105.12 | 116.64 | 410.4 |
| KANSAS | 172.44 | 189 | 199.8 | 188.64 | 749.88 |
| KENTUCKY | 165.6 | 170.64 | 173.16 | 195.48 | 704.88 |
| LOUISIANA | 111.96 | 123.48 | 92.88 | 100.8 | 429.12 |
| MAINE | 46.8 | 39.96 | 46.8 | 31.32 | 164.88 |
| MARYLAND | 60.12 | 63.72 | 108 | 64.8 | 296.64 |
| MASSACHUSETTS | 176.04 | 143.28 | 174.96 | 203.04 | 697.32 |
| MICHIGAN | 326.88 | 389.16 | 369.36 | 382.32 | 1,467.72 |
| MINNESOTA | 404.28 | 413.64 | 453.96 | 421.2 | 1,693.08 |
| MISSISSIPPI | 47.88 | 30.24 | 44.64 | 28.08 | 150.84 |
| MISSOURI | 159.12 | 202.32 | 234 | 205.2 | 800.64 |
| MONTANA | 50.76 | 54.36 | 55.44 | 56.16 | 216.72 |
| NEBRASKA | 91.8 | 117.36 | 102.96 | 101.52 | 413.64 |
| NEVADA | 37.8 | 38.88 | 30.96 | 21.6 | 129.24 |
| NEW HAMPSHIRE | 27.36 | 48.96 | 37.08 | 36.36 | 149.76 |

| | | | | | |
|---|---|---|---|---|---|
| NEW JERSEY | 124.92 | 152.64 | 143.28 | 145.44 | 566.28 |
| NEW MEXICO | 50.04 | 59.4 | 66.96 | 50.4 | 226.8 |
| NEW YORK | 181.8 | 234 | 265.32 | 215.28 | 896.4 |
| NORTH CAROLINA | 230.04 | 232.92 | 230.76 | 234.36 | 928.08 |
| NORTH DAKOTA | 55.08 | 31.68 | 37.8 | 29.52 | 154.08 |
| OHIO | 409.68 | 444.6 | 457.56 | 415.8 | 1,727.64 |
| OKLAHOMA | 69.12 | 64.08 | 84.6 | 90.72 | 308.52 |
| OREGON | 210.96 | 202.32 | 189 | 225.36 | 827.64 |
| PENNSYLVANIA | 373.32 | 397.08 | 375.12 | 325.8 | 1,471.32 |
| RHODE ISLAND | 46.44 | 23.4 | 32.76 | 29.88 | 132.48 |
| SOUTH CAROLINA | 136.08 | 151.92 | 146.88 | 149.76 | 584.64 |
| SOUTH DAKOTA | 21.96 | 16.56 | 23.76 | 28.08 | 90.36 |
| TENNESSEE | 188.64 | 239.04 | 276.12 | 254.88 | 958.68 |
| TEXAS | 507.24 | 519.84 | 547.2 | 514.8 | 2,089.08 |
| UTAH | 53.28 | 70.2 | 60.12 | 63.72 | 247.32 |

DRUG CODE:9639 DRUG NAME:OPIUM POWDERED

| STATE | QUARTER 1 | QUARTER 2 | QUARTER 3 | QUARTER 4 | TOTAL GRAMS |
|---|---|---|---|---|---|
| VERMONT | 56.16 | 36 | 30.24 | 30.6 | 153 |
| VIRGIN ISLANDS | 0 | 0.72 | 0.72 | 0.36 | 1.8 |
| VIRGINIA | 176.04 | 177.84 | 172.8 | 189.72 | 716.4 |
| WASHINGTON | 317.88 | 368.28 | 317.16 | 306.36 | 1,309.68 |
| WEST VIRGINIA | 91.08 | 106.92 | 116.28 | 100.44 | 414.72 |
| WISCONSIN | 200.52 | 185.76 | 212.76 | 185.76 | 784.8 |
| WYOMING | 34.92 | 29.16 | 28.44 | 39.24 | 131.76 |
| TOTAL | 8,001.72 | 8,388.72 | 8,548.56 | 8,294.04 | 33,233.04 |

DRUG CODE:9652 DRUG NAME:OXYMORPHONE

| STATE | QUARTER 1 | QUARTER 2 | QUARTER 3 | QUARTER 4 | TOTAL GRAMS |
|---|---|---|---|---|---|
| ALABAMA | 10,101.34 | 10,326.86 | 9,255 | 8,203.42 | 37,886.62 |
| ALASKA | 347.29 | 328.38 | 308.4 | 305.68 | 1,289.75 |
| ARIZONA | 9,838.53 | 9,684.70 | 9,217.48 | 9,488.88 | 38,229.59 |
| ARKANSAS | 2,645.92 | 2,561 | 2,507.74 | 2,368.39 | 10,083.05 |
| CALIFORNIA | 22,021.17 | 20,671.26 | 19,724.86 | 18,766.87 | 81,184.16 |
| COLORADO | 5,249.68 | 4,864.97 | 4,686.63 | 4,319.82 | 19,121.10 |
| CONNECTICUT | 4,218.91 | 4,110.81 | 3,825.62 | 3,712.10 | 15,867.44 |
| DELAWARE | 2,258.97 | 2,223.59 | 2,222.81 | 2,140.34 | 8,845.71 |
| DISTRICT OF COLUMBIA | 152.03 | 135.79 | 171.81 | 139.93 | 599.56 |
| FLORIDA | 29,317.95 | 29,098.92 | 28,538.08 | 28,470.32 | 115,425.27 |
| GEORGIA | 8,991.78 | 8,751.09 | 8,703.13 | 8,364.66 | 34,810.66 |
| HAWAII | 722.07 | 764.42 | 701.87 | 687.35 | 2,875.71 |
| IDAHO | 1,756.41 | 1,771.42 | 1,693.97 | 1,563.13 | 6,784.93 |
| ILLINOIS | 6,236.27 | 6,153.48 | 6,068.61 | 5,810.41 | 24,268.77 |
| INDIANA | 8,468.55 | 8,015.30 | 7,641.27 | 7,451.51 | 31,576.63 |
| IOWA | 1,879.90 | 1,627.50 | 1,596.65 | 1,504.81 | 6,608.86 |
| KANSAS | 3,277.71 | 3,215.40 | 2,967.62 | 2,768.77 | 12,229.50 |
| KENTUCKY | 5,377.40 | 5,308.80 | 4,980.28 | 4,752.84 | 20,419.32 |
| LOUISIANA | 9,534.81 | 9,201.52 | 9,318.52 | 8,654.98 | 36,709.83 |
| MAINE | 1,946.45 | 1,876.62 | 1,631.18 | 1,418.56 | 6,872.81 |
| MARYLAND | 10,093.13 | 10,386.19 | 10,958.67 | 11,021.24 | 42,459.23 |
| MASSACHUSETTS | 3,241.73 | 3,137.82 | 2,815.56 | 2,758.11 | 11,953.22 |
| MICHIGAN | 20,366.96 | 19,722.63 | 20,584.29 | 20,793.48 | 81,467.36 |
| MINNESOTA | 660.58 | 642.47 | 639.55 | 518.36 | 2,460.96 |
| MISSISSIPPI | 5,307.91 | 4,961.42 | 4,797.46 | 4,358.77 | 19,425.56 |
| MISSOURI | 6,586.57 | 6,204.43 | 5,697.59 | 5,823.06 | 24,311.65 |
| MONTANA | 519.29 | 543.13 | 489.64 | 417.95 | 1,970.01 |
| NEBRASKA | 1,584.77 | 1,662.43 | 1,666.84 | 1,586.29 | 6,500.33 |

| | | | | | |
|---|---|---|---|---|---|
| NEVADA | 6,755.90 | 6,488.50 | 6,073.45 | 5,824.98 | 25,142.83 |
| NEW HAMPSHIRE | 1,372.70 | 1,360.86 | 1,184.87 | 1,203.25 | 5,121.68 |
| NEW JERSEY | 10,127.24 | 10,100.53 | 9,560.27 | 8,897.48 | 38,685.52 |
| NEW MEXICO | 1,710.75 | 1,516.98 | 1,482.60 | 1,353.75 | 6,064.08 |
| NEW YORK | 27,116.85 | 25,418.11 | 24,436.55 | 23,893.07 | 100,864.58 |
| NORTH CAROLINA | 37,725.61 | 35,031.79 | 32,375.76 | 30,114.41 | 135,247.57 |
| NORTH DAKOTA | 214.37 | 228.02 | 220.17 | 166.82 | 829.38 |

DRUG CODE:9652 DRUG NAME:OXYMORPHONE

| STATE | QUARTER 1 | QUARTER 2 | QUARTER 3 | QUARTER 4 | TOTAL GRAMS |
|---|---|---|---|---|---|
| OHIO | 14,088.50 | 13,228.47 | 13,102.83 | 12,765.79 | 53,185.59 |
| OKLAHOMA | 8,019.13 | 7,643.40 | 7,302.06 | 7,055.89 | 30,020.48 |
| OREGON | 2,443.75 | 2,274.86 | 2,205.62 | 2,055.13 | 8,979.36 |
| PENNSYLVANIA | 29,536.07 | 28,498.41 | 28,514.33 | 28,246.09 | 114,794.90 |
| RHODE ISLAND | 652.27 | 578.98 | 582.27 | 560.38 | 2,373.90 |
| SOUTH CAROLINA | 8,335.36 | 8,337.87 | 8,236.92 | 8,198.40 | 33,108.55 |
| SOUTH DAKOTA | 926.91 | 833.11 | 734.04 | 682.94 | 3,177 |
| TENNESSEE | 32,045.68 | 29,999.51 | 28,296.33 | 26,946.14 | 117,287.66 |
| TEXAS | 16,726.25 | 16,353.03 | 15,659.48 | 16,048.16 | 64,786.92 |
| UTAH | 5,316.18 | 5,388.94 | 5,290.36 | 5,000.14 | 20,995.62 |
| VERMONT | 485.26 | 453.45 | 471.34 | 391.9 | 1,801.95 |
| VIRGINIA | 13,371.78 | 13,109.97 | 12,585.27 | 12,011.11 | 51,078.13 |
| WASHINGTON | 4,395.73 | 4,309.74 | 4,055.75 | 3,882.29 | 16,643.51 |
| WEST VIRGINIA | 4,474.95 | 4,471.52 | 3,974.44 | 3,646.46 | 16,567.37 |
| WISCONSIN | 3,821.01 | 3,480.59 | 3,242.83 | 3,072.88 | 13,617.31 |
| WYOMING | 437.66 | 479.15 | 365.31 | 395.16 | 1,677.28 |
| TOTAL | 412,803.99 | 397,538.14 | 383,363.98 | 370,582.65 | 1,564,288.76 |

DRUG CODE:9668 DRUG NAME:NOROXYMORPHONE

| STATE | QUARTER 1 | QUARTER 2 | QUARTER 3 | QUARTER 4 | TOTAL GRAMS |
|---|---|---|---|---|---|
| ALABAMA | 2.05 | 1.14 | 0.92 | 0.27 | 4.38 |
| ARIZONA | 0.77 | 0.65 | 0.27 | 0 | 1.69 |
| ARKANSAS | 15.36 | 11.1 | 6.45 | 23.55 | 56.46 |
| CALIFORNIA | 67.41 | 77.7 | 64.49 | 112.25 | 321.85 |
| COLORADO | 0.19 | 0.14 | 0.2 | 0 | 0.53 |
| CONNECTICUT | 0.62 | 0.34 | 0.06 | 0 | 1.02 |
| DISTRICT OF COLUMBIA | 4.03 | 3.18 | 0.07 | 2 | 9.28 |
| FLORIDA | 29.9 | 23.82 | 17.75 | 28.21 | 99.68 |
| GEORGIA | 5.33 | 7.6 | 7.15 | 3.63 | 23.71 |
| ILLINOIS | 17.65 | 9.52 | 14.38 | 11.09 | 52.64 |
| INDIANA | 0.71 | 0.45 | 0.4 | 0 | 1.56 |
| KANSAS | 0.7 | 0 | 0 | 0 | 0.7 |
| LOUISIANA | 0.39 | 0 | 0 | 0.23 | 0.62 |
| MARYLAND | 1.02 | 0.77 | 0.67 | 0 | 2.46 |
| MASSACHUSETTS | 0.37 | 0 | 0.16 | 1.47 | 2 |
| MISSISSIPPI | 8.08 | 3.7 | 3.53 | 0.88 | 16.19 |
| MISSOURI | 12.35 | 7.98 | 21.84 | 14.63 | 56.8 |
| NEBRASKA | 0 | 0 | 0 | 1.59 | 1.59 |
| NEVADA | 0 | 8.88 | 13.99 | 20.27 | 43.14 |
| NEW HAMPSHIRE | 1.53 | 0.87 | 0.37 | 0 | 2.77 |
| NEW JERSEY | 0.18 | 13.6 | 4.98 | 6.85 | 25.61 |
| NEW MEXICO | 3.24 | 0 | 0 | 0 | 3.24 |
| NEW YORK | 19.69 | 9.26 | 5.05 | 11.28 | 45.28 |
| NORTH CAROLINA | 0.99 | 0.69 | 0.57 | 0 | 2.25 |
| OHIO | 14.52 | 18.99 | 10.51 | 9.29 | 53.31 |
| OKLAHOMA | 1.56 | 0.49 | 0.69 | 0.07 | 2.81 |

DRUG CODE:9668 DRUG NAME:NOROXYMORPHONE

| STATE | QUARTER 1 | QUARTER 2 | QUARTER 3 | QUARTER 4 | TOTAL GRAMS |
|---|---|---|---|---|---|
| OREGON | 1.98 | 0.98 | 1.07 | 0.17 | 4.2 |
| PENNSYLVANIA | 1.71 | 3.9 | 0.41 | 0 | 6.02 |
| PUERTO RICO | 9.21 | 2.82 | 1.86 | 4.43 | 18.32 |
| RHODE ISLAND | 0.35 | 2.96 | 0.03 | 0 | 3.34 |
| SOUTH CAROLINA | 0.49 | 0 | 0 | 0 | 0.49 |
| TENNESSEE | 1.26 | 0.58 | 0.74 | 0.45 | 3.03 |
| TEXAS | 19.94 | 16.71 | 25.51 | 41.71 | 103.87 |
| UTAH | 0.97 | 0.24 | 0.32 | 0 | 1.53 |
| VERMONT | 0.76 | 0.17 | 0.2 | 0 | 1.13 |
| WASHINGTON | 8.5 | 3.44 | 3.65 | 4.07 | 19.66 |
| WISCONSIN | 0.64 | 0 | 0.03 | 0 | 0.67 |
| TOTAL | 254.45 | 232.67 | 208.32 | 298.39 | 993.83 |

DRUG CODE:9737 DRUG NAME:ALFENTANIL

| STATE | QUARTER 1 | QUARTER 2 | QUARTER 3 | QUARTER 4 | TOTAL GRAMS |
|---|---|---|---|---|---|
| ALABAMA | 0.09 | 0.06 | 0.06 | 0.04 | 0.25 |
| ALASKA | 0.92 | 0.7 | 0.7 | 0.95 | 3.27 |
| ARIZONA | 1.28 | 0.75 | 0.77 | 0.82 | 3.62 |
| ARKANSAS | 0.11 | 0.02 | 0.01 | 0.02 | 0.16 |
| CALIFORNIA | 18.16 | 17.45 | 16.37 | 15.79 | 67.77 |
| COLORADO | 0.85 | 0.71 | 0.97 | 0.54 | 3.07 |
| CONNECTICUT | 1.04 | 1.32 | 1.51 | 0.96 | 4.83 |
| FLORIDA | 5.99 | 4.34 | 4.56 | 4.92 | 19.81 |
| GEORGIA | 2.62 | 1.7 | 2.15 | 2.26 | 8.73 |
| GUAM | 0.05 | 0.06 | 0.1 | 0.03 | 0.24 |
| HAWAII | 0.38 | 0.49 | 0.25 | 0.68 | 1.8 |
| IDAHO | 1.24 | 1.22 | 1.19 | 1.67 | 5.32 |
| ILLINOIS | 4.13 | 4.5 | 3.22 | 4.21 | 16.06 |
| INDIANA | 2.53 | 2.28 | 2.33 | 2.19 | 9.33 |
| IOWA | 1.67 | 1.49 | 1.81 | 2.14 | 7.11 |
| KANSAS | 1.52 | 1.89 | 1.75 | 1.72 | 6.88 |
| KENTUCKY | 1.56 | 1.67 | 3.2 | 2.62 | 9.05 |
| LOUISIANA | 2.13 | 2.33 | 2.5 | 2.32 | 9.28 |
| MAINE | 0.56 | 0.74 | 0.59 | 0.74 | 2.63 |
| MARYLAND | 0.88 | 0.33 | 0.46 | 0.64 | 2.31 |
| MASSACHUSETTS | 1.43 | 1.43 | 0.93 | 1.48 | 5.27 |
| MICHIGAN | 1.41 | 1.01 | 1.17 | 1.53 | 5.12 |
| MINNESOTA | 3.67 | 3.48 | 3.03 | 3.42 | 13.6 |
| MISSISSIPPI | 0.11 | 0.11 | 0.17 | 0.19 | 0.58 |
| MISSOURI | 1.56 | 1.62 | 1.25 | 0.88 | 5.31 |
| MONTANA | 0.55 | 0.28 | 0.41 | 0.31 | 1.55 |
| NEBRASKA | 1.1 | 1.21 | 0.96 | 0.82 | 4.09 |
| NEVADA | 1.85 | 2.22 | 1.54 | 1.47 | 7.08 |
| NEW HAMPSHIRE | 0.4 | 0.37 | 0.42 | 0.38 | 1.57 |
| NEW JERSEY | 0.2 | 0.37 | 0.21 | 0.34 | 1.12 |
| NEW MEXICO | 1.24 | 1.04 | 0.94 | 1.56 | 4.78 |

DRUG CODE:9737 DRUG NAME:ALFENTANIL

| STATE | QUARTER 1 | QUARTER 2 | QUARTER 3 | QUARTER 4 | TOTAL GRAMS |
|---|---|---|---|---|---|
| NEW YORK | 3.44 | 2.3 | 2.94 | 2.57 | 11.25 |
| NORTH CAROLINA | 3.21 | 2.75 | 3.05 | 2.83 | 11.84 |
| NORTH DAKOTA | 0.06 | 0.04 | 0.06 | 0.06 | 0.22 |
| OHIO | 5.23 | 5.06 | 4.85 | 5.94 | 21.08 |
| OKLAHOMA | 1.43 | 0.87 | 0.9 | 0.88 | 4.08 |

| | | | | | |
|---|---|---|---|---|---|
| OREGON | 3.79 | 3.72 | 4.18 | 4.07 | 15.76 |
| PENNSYLVANIA | 0.97 | 0.55 | 0.72 | 0.45 | 2.69 |
| RHODE ISLAND | 0.1 | 0.06 | 0.08 | 0.05 | 0.29 |
| SOUTH CAROLINA | 0.08 | 0 | 0.05 | 0.06 | 0.19 |
| SOUTH DAKOTA | 0.06 | 0.06 | 0.06 | 0.05 | 0.23 |
| TENNESSEE | 0.82 | 1.04 | 1.14 | 0.94 | 3.94 |
| TEXAS | 7.01 | 5.42 | 5.81 | 5.7 | 23.94 |
| UTAH | 0.53 | 0.53 | 0.33 | 0.49 | 1.88 |
| VERMONT | 0 | 0 | 0.02 | 0 | 0.02 |
| VIRGINIA | 0.69 | 0.78 | 0.91 | 1.25 | 3.63 |
| WASHINGTON | 3.42 | 2.52 | 2.68 | 3.06 | 11.68 |
| WISCONSIN | 4.54 | 4.68 | 4.65 | 4.63 | 18.5 |
| WYOMING | 0.03 | 0.05 | 0.02 | 0.04 | 0.14 |
| TOTAL | 96.64 | 87.62 | 87.98 | 90.71 | 362.95 |

DRUG CODE:9739 DRUG NAME:REMIFENTANIL

| STATE | QUARTER 1 | QUARTER 2 | QUARTER 3 | QUARTER 4 | TOTAL GRAMS |
|---|---|---|---|---|---|
| ALABAMA | 2.61 | 3.36 | 3.55 | 3.12 | 12.64 |
| ALASKA | 2.81 | 2.74 | 2.58 | 3.41 | 11.54 |
| ARIZONA | 4.4 | 3.31 | 4.56 | 4.59 | 16.86 |
| ARKANSAS | 1.78 | 1.6 | 1.67 | 1.85 | 6.9 |
| CALIFORNIA | 29.59 | 32.67 | 33.32 | 33.73 | 129.31 |
| COLORADO | 6.09 | 5.86 | 6.57 | 6.73 | 25.25 |
| CONNECTICUT | 7.3 | 8.24 | 8.24 | 8.36 | 32.14 |
| DELAWARE | 3.3 | 3.38 | 3.7 | 2.32 | 12.7 |
| DISTRICT OF COLUMBIA | 0.59 | 0.77 | 0.94 | 0.79 | 3.09 |
| FLORIDA | 25.06 | 25.87 | 25.87 | 26.18 | 102.98 |
| GEORGIA | 4.62 | 4.36 | 4.77 | 5.2 | 18.95 |
| GUAM | 0 | 0 | 0.5 | 0 | 0.5 |
| HAWAII | 1.48 | 1.31 | 1.21 | 1.33 | 5.33 |
| IDAHO | 1.75 | 2.33 | 1.94 | 2.92 | 8.94 |
| ILLINOIS | 14.11 | 13.05 | 14.83 | 14.68 | 56.67 |
| INDIANA | 7.76 | 7.67 | 7.2 | 9.15 | 31.78 |
| IOWA | 2.66 | 3.06 | 2.83 | 3 | 11.55 |
| KANSAS | 1.85 | 1.57 | 2.05 | 2.5 | 7.97 |
| KENTUCKY | 4.52 | 4.54 | 4.93 | 4.47 | 18.46 |
| LOUISIANA | 4.19 | 4.36 | 3.94 | 4.5 | 16.99 |
| MAINE | 1.06 | 1.05 | 1.22 | 1.79 | 5.12 |
| MARYLAND | 6.9 | 7.07 | 7.56 | 6.77 | 28.3 |
| MASSACHUSETTS | 18.34 | 19.79 | 18.81 | 19.76 | 76.7 |
| MICHIGAN | 9.96 | 10.25 | 9.76 | 11.14 | 41.11 |

DRUG CODE:9739 DRUG NAME:REMIFENTANIL

| STATE | QUARTER 1 | QUARTER 2 | QUARTER 3 | QUARTER 4 | TOTAL GRAMS |
|---|---|---|---|---|---|
| MINNESOTA | 6.71 | 6.41 | 5.87 | 7.77 | 26.76 |
| MISSISSIPPI | 0.52 | 0.48 | 0.56 | 0.38 | 1.94 |
| MISSOURI | 4.89 | 5.09 | 6.11 | 4.9 | 20.99 |
| MONTANA | 1.84 | 2.11 | 1.51 | 1.74 | 7.2 |
| NEBRASKA | 0.82 | 1.12 | 0.99 | 1.05 | 3.98 |
| NEVADA | 0.95 | 0.84 | 0.77 | 0.97 | 3.53 |
| NEW HAMPSHIRE | 1.13 | 1.17 | 1.3 | 1.12 | 4.72 |
| NEW JERSEY | 11.79 | 12.04 | 13.91 | 11.88 | 49.62 |
| NEW MEXICO | 1.09 | 0.85 | 0.96 | 1.28 | 4.18 |
| NEW YORK | 33.29 | 36.84 | 33.61 | 37.42 | 141.16 |
| NORTH CAROLINA | 13.26 | 14.72 | 13.93 | 14.8 | 56.71 |
| NORTH DAKOTA | 0.28 | 0.4 | 0.4 | 0.51 | 1.59 |
| OHIO | 18.03 | 18.62 | 19.1 | 20.33 | 76.08 |

| | | | | | |
|---|---|---|---|---|---|
| OKLAHOMA | 3.15 | 1.92 | 2.3 | 2.45 | 9.82 |
| OREGON | 3.29 | 2.83 | 2.14 | 3.04 | 11.3 |
| PENNSYLVANIA | 33.93 | 34.98 | 35.29 | 33.37 | 137.57 |
| PUERTO RICO | 0 | 0.16 | 0 | 0 | 0.16 |
| RHODE ISLAND | 0.47 | 0.32 | 0.54 | 0.52 | 1.85 |
| SOUTH CAROLINA | 4.69 | 5.22 | 5.02 | 5.02 | 19.95 |
| SOUTH DAKOTA | 0.59 | 0.55 | 0.56 | 0.64 | 2.34 |
| TENNESSEE | 5.24 | 5.19 | 4.63 | 5.67 | 20.73 |
| TEXAS | 25.58 | 27.39 | 28.68 | 27.72 | 109.37 |
| UTAH | 15.86 | 16.93 | 16.42 | 17.02 | 66.23 |
| VERMONT | 0.5 | 0.42 | 0.65 | 0.59 | 2.16 |
| VIRGINIA | 6.68 | 5.67 | 6.01 | 6.55 | 24.91 |
| WASHINGTON | 12.36 | 11.95 | 11.99 | 13.26 | 49.56 |
| WEST VIRGINIA | 4.76 | 4.96 | 5.53 | 5.62 | 20.87 |
| WISCONSIN | 5.85 | 5.71 | 6.1 | 6.64 | 24.3 |
| WYOMING | 0.5 | 0.48 | 0.42 | 0.48 | 1.88 |
| TOTAL | 380.78 | 393.58 | 397.85 | 411.03 | 1,583.24 |

DRUG CODE:9740 DRUG NAME:SUFENTANIL BASE

| STATE | QUARTER 1 | QUARTER 2 | QUARTER 3 | QUARTER 4 | TOTAL GRAMS |
|---|---|---|---|---|---|
| ALABAMA | 2.4 | 2.05 | 1.4 | 2.03 | 7.88 |
| ALASKA | 0.01 | 0.01 | 0.01 | 0.01 | 0.04 |
| ARIZONA | 0.1 | 0.06 | 0.08 | 0.06 | 0.3 |
| ARKANSAS | 0.24 | 0.1 | 0.16 | 0.14 | 0.64 |
| CALIFORNIA | 10.04 | 5.45 | 11.64 | 12.02 | 39.15 |
| COLORADO | 0.26 | 0.15 | 0.17 | 0.24 | 0.82 |
| CONNECTICUT | 0.15 | 0.15 | 0.79 | 0.21 | 1.3 |
| DELAWARE | 0.03 | 0 | 0.01 | 0 | 0.04 |
| DISTRICT OF COLUMBIA | 0.01 | 0.01 | 0 | 0.02 | 0.04 |
| FLORIDA | 2.23 | 3.35 | 2.32 | 2.54 | 10.44 |
| GEORGIA | 0.43 | 0.34 | 0.35 | 0.34 | 1.46 |
| HAWAII | 0.02 | 0.01 | 0.01 | 0.01 | 0.05 |
| IDAHO | 0.07 | 0.05 | 0.03 | 0.03 | 0.18 |

DRUG CODE:9740 DRUG NAME:SUFENTANIL BASE

| STATE | QUARTER 1 | QUARTER 2 | QUARTER 3 | QUARTER 4 | TOTAL GRAMS |
|---|---|---|---|---|---|
| ILLINOIS | 0.85 | 0.37 | 0.46 | 0.46 | 2.14 |
| INDIANA | 0.45 | 0.43 | 0.3 | 0.33 | 1.51 |
| IOWA | 0.05 | 0.03 | 0.04 | 0.03 | 0.15 |
| KANSAS | 0.31 | 0.2 | 0.27 | 0.24 | 1.02 |
| KENTUCKY | 0.39 | 0.25 | 0.3 | 0.25 | 1.19 |
| LOUISIANA | 1.18 | 0.7 | 0.75 | 0.78 | 3.41 |
| MAINE | 0.01 | 0.01 | 0.01 | 0 | 0.03 |
| MARYLAND | 0.04 | 0.03 | 0.05 | 0.03 | 0.15 |
| MASSACHUSETTS | 0.14 | 0.12 | 0.41 | 0.11 | 0.78 |
| MICHIGAN | 0.63 | 0.57 | 0.6 | 0.53 | 2.33 |
| MINNESOTA | 0.25 | 0.14 | 0.23 | 0.19 | 0.81 |
| MISSISSIPPI | 8.13 | 11.46 | 10.47 | 11.78 | 41.84 |
| MISSOURI | 0.48 | 0.38 | 0.32 | 0.33 | 1.51 |
| MONTANA | 0.05 | 0.05 | 0.03 | 0.05 | 0.18 |
| NEBRASKA | 0.12 | 0.09 | 0.08 | 0.1 | 0.39 |
| NEVADA | 0.14 | 0.05 | 0.08 | 0.09 | 0.36 |
| NEW HAMPSHIRE | 0.03 | 0.01 | 0.02 | 0.01 | 0.07 |
| NEW JERSEY | 0.99 | 0.95 | 0.92 | 1.57 | 4.43 |
| NEW MEXICO | 0.05 | 0.04 | 0.03 | 0.04 | 0.16 |
| NEW YORK | 1.88 | 1.84 | 1.76 | 1.72 | 7.2 |
| NORTH CAROLINA | 0.47 | 0.34 | 0.32 | 1.1 | 2.23 |

| | | | | | |
|---|---|---|---|---|---|
| NORTH DAKOTA | 0.04 | 0.02 | 0.03 | 0.01 | 0.1 |
| OHIO | 0.75 | 0.71 | 0.98 | 0.43 | 2.87 |
| OKLAHOMA | 0.25 | 0.16 | 0.19 | 0.15 | 0.75 |
| OREGON | 0.51 | 0.24 | 0.32 | 0.24 | 1.31 |
| PENNSYLVANIA | 0.9 | 0.91 | 1.83 | 1.58 | 5.22 |
| PUERTO RICO | 0.14 | 0.27 | 0.22 | 0.14 | 0.77 |
| RHODE ISLAND | 0.01 | 0.01 | 0.01 | 0 | 0.03 |
| SOUTH CAROLINA | 0.82 | 0.8 | 0.1 | 0.79 | 2.51 |
| SOUTH DAKOTA | 0.04 | 0.03 | 0.03 | 0.04 | 0.14 |
| TENNESSEE | 0.52 | 0.41 | 0.39 | 0.28 | 1.6 |
| TEXAS | 2.25 | 1.47 | 1.62 | 1.58 | 6.92 |
| UTAH | 0.21 | 0.21 | 0.25 | 0.2 | 0.87 |
| VERMONT | 0.02 | 0.02 | 0 | 0 | 0.04 |
| VIRGINIA | 0.43 | 0.43 | 0.31 | 0.4 | 1.57 |
| WASHINGTON | 0.31 | 0.19 | 0.23 | 0.25 | 0.98 |
| WEST VIRGINIA | 0.06 | 0.05 | 0.06 | 0.04 | 0.21 |
| WISCONSIN | 0.39 | 2.1 | 0.78 | 0.11 | 3.38 |
| WYOMING | 0.02 | 0.01 | 0 | 0.01 | 0.04 |
| TOTAL | 40.3 | 37.83 | 41.77 | 43.64 | 163.54 |

DRUG CODE:9780 DRUG NAME:TAPENTADOL

| STATE | QUARTER 1 | QUARTER 2 | QUARTER 3 | QUARTER 4 | TOTAL GRAMS |
|---|---|---|---|---|---|
| ALABAMA | 66,160.50 | 65,914 | 64,934.50 | 65,851 | 262,860 |
| ALASKA | 1,822 | 1,760.50 | 2,141 | 2,026 | 7,749.50 |
| ARIZONA | 37,876.85 | 35,418.50 | 36,803 | 39,677 | 149,775.35 |

DRUG CODE:9780 DRUG NAME:TAPENTADOL

| STATE | QUARTER 1 | QUARTER 2 | QUARTER 3 | QUARTER 4 | TOTAL GRAMS |
|---|---|---|---|---|---|
| ARKANSAS | 26,540.35 | 27,422 | 27,281.50 | 26,365.50 | 107,609.35 |
| CALIFORNIA | 118,632.37 | 120,928.67 | 120,683.50 | 120,528.40 | 480,772.94 |
| COLORADO | 27,710 | 27,020.50 | 27,311 | 28,442.50 | 110,484 |
| CONNECTICUT | 12,188.50 | 12,986 | 14,109.50 | 14,616.30 | 53,900.30 |
| DELAWARE | 6,105 | 6,707.50 | 7,071.50 | 7,362 | 27,246 |
| DISTRICT OF COLUMBIA | 1,448.85 | 1,474.50 | 1,456 | 1,366 | 5,745.35 |
| FLORIDA | 117,551.85 | 116,363.93 | 115,208.50 | 116,559 | 465,683.28 |
| GEORGIA | 70,403.50 | 70,749 | 72,653.50 | 74,837.50 | 288,643.50 |
| HAWAII | 2,788 | 2,859.50 | 2,614 | 2,810 | 11,071.50 |
| IDAHO | 5,394.50 | 5,926.50 | 6,215.50 | 7,191 | 24,727.50 |
| ILLINOIS | 42,434 | 45,147.50 | 45,392.50 | 46,735.50 | 179,709.50 |
| INDIANA | 39,693 | 40,118 | 39,919 | 41,625.50 | 161,355.50 |
| IOWA | 12,399 | 13,167 | 12,882 | 14,228 | 52,676 |
| KANSAS | 11,359 | 11,577 | 11,436 | 11,097.50 | 45,469.50 |
| KENTUCKY | 16,751 | 15,784 | 16,387 | 17,143.30 | 66,065.30 |
| LOUISIANA | 15,590 | 16,266.50 | 16,964 | 17,009 | 65,829.50 |
| MAINE | 1,859 | 2,214.50 | 2,102 | 2,035 | 8,210.50 |
| MARYLAND | 34,017.50 | 35,455.50 | 35,751.50 | 35,518.50 | 140,743 |
| MASSACHUSETTS | 17,912 | 17,835.50 | 18,557 | 18,923 | 73,227.50 |
| MICHIGAN | 22,881.50 | 24,371.50 | 23,990 | 24,353 | 95,596 |
| MINNESOTA | 5,147 | 5,243.50 | 4,841 | 5,130 | 20,361.50 |
| MISSISSIPPI | 13,857.50 | 14,546.50 | 14,681 | 14,700.50 | 57,785.50 |
| MISSOURI | 29,688 | 29,043 | 28,149 | 28,537.50 | 115,417.50 |
| MONTANA | 2,437 | 2,526 | 2,556.50 | 2,589 | 10,108.50 |
| NEBRASKA | 10,531 | 10,383 | 10,424.50 | 10,893 | 42,231.50 |
| NEVADA | 12,993.50 | 13,063 | 12,467 | 12,943.50 | 51,467 |
| NEW HAMPSHIRE | 6,272.50 | 6,090.50 | 6,825.50 | 6,982.50 | 26,171 |
| NEW JERSEY | 49,545 | 51,423 | 50,721.50 | 51,812.50 | 203,502 |

| | | | | | |
|---|---|---|---|---|---|
| NEW MEXICO | 4,255 | 4,805 | 4,406 | 4,261 | 17,727 |
| NEW YORK | 110,315.50 | 107,622.20 | 106,108.50 | 107,228.30 | 431,274.50 |
| NORTH CAROLINA | 102,575.35 | 103,021.50 | 101,782.20 | 105,468.70 | 412,847.75 |
| NORTH DAKOTA | 2,602 | 2,519 | 2,729.50 | 2,575 | 10,425.50 |
| OHIO | 71,167.35 | 70,064 | 70,795 | 70,221 | 282,247.35 |
| OKLAHOMA | 25,180.35 | 25,455.50 | 25,502.50 | 27,542.50 | 103,680.85 |
| OREGON | 7,563.50 | 7,800 | 8,308 | 8,442 | 32,113.50 |
| PENNSYLVANIA | 67,855.70 | 70,807.50 | 71,213 | 73,140 | 283,016.20 |
| PUERTO RICO | 1,282.97 | 1,032 | 955 | 983.5 | 4,253.47 |
| RHODE ISLAND | 2,071.50 | 2,378.50 | 2,393.50 | 2,348 | 9,191.50 |
| SOUTH CAROLINA | 43,663 | 44,605.50 | 45,328.70 | 47,026.50 | 180,623.70 |
| SOUTH DAKOTA | 6,064.50 | 5,866 | 5,898.50 | 5,671.50 | 23,500.50 |
| TENNESSEE | 33,464.50 | 35,403 | 37,089 | 37,060 | 143,016.50 |
| TEXAS | 100,239.85 | 107,335.43 | 109,150 | 115,496 | 432,221.28 |
| UTAH | 15,737.50 | 16,417 | 16,219.50 | 16,518.50 | 64,892.50 |
| VERMONT | 1,485 | 1,263.50 | 1,250 | 1,499.50 | 5,498 |
| VIRGIN ISLANDS | 6 | 9 | 3 | 26 | 44 |

DRUG CODE:9780 DRUG NAME:TAPENTADOL

| STATE | QUARTER 1 | QUARTER 2 | QUARTER 3 | QUARTER 4 | TOTAL GRAMS |
|---|---|---|---|---|---|
| VIRGINIA | 48,903 | 49,314.75 | 48,762.75 | 49,206.25 | 196,186.75 |
| WASHINGTON | 19,572.50 | 19,606 | 19,871.50 | 19,055 | 78,105 |
| WEST VIRGINIA | 5,867.50 | 6,252.50 | 6,011.50 | 6,224.50 | 24,356 |
| WISCONSIN | 18,149.85 | 17,808 | 17,801 | 18,032.50 | 71,791.35 |
| WYOMING | 2,152 | 1,732 | 1,730.50 | 1,863.50 | 7,478 |
| TOTAL | 1,530,164.19 | 1,550,904.48 | 1,555,839.15 | 1,589,778.75 | 6,226,686.57 |

DRUG CODE:9801 DRUG NAME:FENTANYL BASE

| STATE | QUARTER 1 | QUARTER 2 | QUARTER 3 | QUARTER 4 | TOTAL GRAMS |
|---|---|---|---|---|---|
| ALABAMA | 1,969.75 | 1,903.39 | 1,845.85 | 1,839.53 | 7,558.52 |
| ALASKA | 278.27 | 275.52 | 261.82 | 244.35 | 1,059.96 |
| AMERICAN SAMOA | 0.38 | 0 | 0.16 | 0.01 | 0.55 |
| ARIZONA | 2,402.41 | 2,340.86 | 2,227.44 | 2,168.23 | 9,138.94 |
| ARKANSAS | 1,184.10 | 1,169.48 | 1,122.91 | 1,078.23 | 4,554.72 |
| CALIFORNIA | 9,095.22 | 8,671.30 | 8,327.98 | 8,321.20 | 34,415.70 |
| COLORADO | 1,672.91 | 1,608.85 | 1,528.17 | 1,444.37 | 6,254.30 |
| CONNECTICUT | 1,254.39 | 1,254.56 | 1,145.41 | 1,053.89 | 4,708.25 |
| DELAWARE | 425.38 | 402.43 | 405.83 | 389.88 | 1,623.52 |
| DISTRICT OF COLUMBIA | 101.87 | 107.49 | 95.75 | 92.57 | 397.68 |
| FLORIDA | 7,014.12 | 6,836.22 | 6,492.18 | 6,574.08 | 26,916.60 |
| GEORGIA | 3,191.02 | 3,146.64 | 3,103.40 | 3,060.17 | 12,501.23 |
| GUAM | 10.11 | 10.16 | 10.49 | 7.79 | 38.55 |
| HAWAII | 223.52 | 222.53 | 206.02 | 195.55 | 847.62 |
| IDAHO | 711.16 | 696.19 | 658.78 | 635.76 | 2,701.89 |
| ILLINOIS | 3,573.30 | 3,522.27 | 3,435.88 | 3,319.95 | 13,851.40 |
| INDIANA | 2,476.35 | 2,396.75 | 2,253.59 | 2,180.33 | 9,307.02 |
| IOWA | 1,024.91 | 1,012.98 | 979.5 | 950.28 | 3,967.67 |
| KANSAS | 1,322.42 | 1,278.32 | 1,272.21 | 1,197.91 | 5,070.86 |
| KENTUCKY | 1,430.78 | 1,401.86 | 1,354.08 | 1,277.59 | 5,464.31 |
| LOUISIANA | 1,461.48 | 1,448.58 | 1,374.88 | 1,338.75 | 5,623.69 |
| MAINE | 562.83 | 545.8 | 510.69 | 448.33 | 2,067.65 |
| MARYLAND | 1,813.82 | 1,813.92 | 1,747.78 | 1,669.49 | 7,045.01 |
| MASSACHUSETTS | 1,622.99 | 1,572.21 | 1,497.04 | 1,440.99 | 6,133.23 |
| MICHIGAN | 3,942.17 | 3,874.28 | 3,553.42 | 3,561.87 | 14,931.74 |
| MINNESOTA | 1,338.36 | 1,312.57 | 1,232.23 | 1,153.93 | 5,037.09 |
| MISSISSIPPI | 1,413.86 | 1,396.19 | 1,328.52 | 1,349.31 | 5,487.88 |

| | | | | | |
|---|---|---|---|---|---|
| MISSOURI | 2,680.56 | 2,591.15 | 2,457.22 | 2,405.58 | 10,134.51 |
| MONTANA | 414.77 | 415.79 | 399.17 | 378.69 | 1,608.42 |
| NEBRASKA | 607.01 | 623.27 | 583.57 | 578.11 | 2,391.96 |
| NEVADA | 797.78 | 753.49 | 730.26 | 696.41 | 2,977.94 |
| NEW HAMPSHIRE | 456.39 | 451.22 | 441.71 | 406.43 | 1,755.75 |
| NEW JERSEY | 2,886.56 | 2,924.70 | 2,794.59 | 2,889.01 | 11,494.86 |
| NEW MEXICO | 578.21 | 577.56 | 554.15 | 533.35 | 2,243.27 |
| NEW YORK | 5,866.43 | 5,666.23 | 5,385.42 | 5,008.18 | 21,926.26 |
| NORTH CAROLINA | 3,800.08 | 3,692.32 | 3,451.39 | 3,274.62 | 14,218.41 |
| NORTH DAKOTA | 338.85 | 313.9 | 298.47 | 249.33 | 1,200.55 |

DRUG CODE:9801 DRUG NAME:FENTANYL BASE

| STATE | QUARTER 1 | QUARTER 2 | QUARTER 3 | QUARTER 4 | TOTAL GRAMS |
|---|---|---|---|---|---|
| OHIO | 3,298.37 | 3,188.10 | 3,020.43 | 2,866.57 | 12,373.47 |
| OKLAHOMA | 2,105.51 | 2,070.95 | 2,019.19 | 1,895.79 | 8,091.44 |
| OREGON | 1,324.71 | 1,281.22 | 1,187.16 | 1,085.78 | 4,878.87 |
| PENNSYLVANIA | 6,206.82 | 6,126.73 | 5,902.17 | 5,648.50 | 23,884.22 |
| PUERTO RICO | 218.78 | 218.37 | 208.87 | 216.34 | 862.36 |
| RHODE ISLAND | 238.46 | 237.43 | 230.55 | 215.81 | 922.25 |
| SOUTH CAROLINA | 1,602.12 | 1,546.72 | 1,516.53 | 1,593.78 | 6,259.15 |
| SOUTH DAKOTA | 384.26 | 364.07 | 348.46 | 341.51 | 1,438.30 |
| TENNESSEE | 2,535.77 | 2,411 | 2,391.36 | 2,281.74 | 9,619.87 |
| TEXAS | 6,946.71 | 6,843.56 | 6,726.79 | 7,113.23 | 27,630.29 |
| UTAH | 991.92 | 992.73 | 995.61 | 938.2 | 3,918.46 |
| VERMONT | 273.92 | 245.2 | 229 | 217.35 | 965.47 |
| VIRGIN ISLANDS | 7.13 | 6.61 | 7.16 | 5.8 | 26.7 |
| VIRGINIA | 2,285.49 | 2,212.36 | 2,097.13 | 2,000.23 | 8,595.21 |
| WASHINGTON | 1,913.55 | 1,902.85 | 1,818.13 | 1,721.77 | 7,356.30 |
| WEST VIRGINIA | 714.56 | 697.01 | 648.38 | 613.56 | 2,673.51 |
| WISCONSIN | 2,030.23 | 1,985.93 | 1,841.47 | 1,748.18 | 7,605.81 |
| WYOMING | 189.91 | 182.26 | 182.77 | 173.74 | 728.68 |
| TOTAL | 103,212.74 | 100,744.08 | 96,439.12 | 94,091.93 | 394,487.87 |

ARCOS 3 - REPORT 3

QUARTERLY DRUG DISTRIBUTION BY STATE PER 100K POPULATION BY GRAM WT

DRUG CODE:1100 DRUG NAME: AMPHETAMINE

| STATE | 1ST QUARTER | 2ND QUARTER | 3RD QUARTER | 4TH QUARTER | TOTAL GRAMS |
|---|---|---|---|---|---|
| ALABAMA | 2,589.60 | 2,644.62 | 2,698.26 | 2,683.06 | 10,615.55 |
| ALASKA | 1,055.85 | 1,144.18 | 1,122.48 | 1,088.39 | 4,410.89 |
| AMERICAN SAMOA | 3.86 | 0 | 2.29 | 3.81 | 9.97 |
| ARIZONA | 1,144.62 | 1,186.25 | 1,190.77 | 1,232.63 | 4,754.27 |
| ARKANSAS | 1,625.81 | 1,645.98 | 1,637.32 | 1,699.04 | 6,608.15 |
| CALIFORNIA | 764.32 | 781.32 | 780.51 | 798.81 | 3,124.97 |
| COLORADO | 1,397.27 | 1,474.98 | 1,512.96 | 1,509.77 | 5,894.99 |
| CONNECTICUT | 1,357.24 | 1,396.05 | 1,423.92 | 1,435.46 | 5,612.66 |
| DELAWARE | 2,323.73 | 2,414.13 | 2,404.58 | 2,431.26 | 9,573.71 |
| DISTRICT OF COLUMBIA | 1,803.68 | 1,894.64 | 1,898.67 | 1,876.03 | 7,473.02 |
| FLORIDA | 1,366.62 | 1,460.84 | 1,479.06 | 1,519 | 5,825.52 |
| GEORGIA | 1,764.58 | 1,793.13 | 1,850.22 | 1,879.37 | 7,287.30 |
| GUAM | 222.68 | 219.94 | 274.94 | 265.85 | 983.41 |
| HAWAII | 668.39 | 633.42 | 718.26 | 659.51 | 2,679.58 |
| IDAHO | 1,425.60 | 1,514.98 | 1,495.89 | 1,528.24 | 5,964.71 |
| ILLINOIS | 1,333.07 | 1,402.82 | 1,414.68 | 1,445.72 | 5,596.30 |
| INDIANA | 1,824.96 | 1,873.25 | 1,885.46 | 1,934.43 | 7,518.10 |
| IOWA | 1,505.05 | 1,529.24 | 1,545.51 | 1,574.52 | 6,154.32 |
| KANSAS | 1,929.93 | 1,976.13 | 2,014.78 | 2,043.05 | 7,963.89 |
| KENTUCKY | 1,444.06 | 1,469.49 | 1,476.87 | 1,521.01 | 5,911.43 |
| LOUISIANA | 3,065.54 | 3,101.14 | 3,207.99 | 3,286.78 | 12,661.45 |
| MAINE | 1,567.71 | 1,613.86 | 1,621.69 | 1,628.08 | 6,431.35 |
| MARYLAND | 1,450.75 | 1,514.10 | 1,527.46 | 1,529.97 | 6,022.27 |
| MASSACHUSETTS | 2,534.62 | 2,633.98 | 2,655.62 | 2,662.20 | 10,486.42 |
| MICHIGAN | 2,296.91 | 2,346.71 | 2,235.78 | 2,405.90 | 9,285.29 |
| MINNESOTA | 1,946.03 | 1,990.76 | 2,010.28 | 2,012.11 | 7,959.18 |
| MISSISSIPPI | 1,978.24 | 1,975.11 | 2,047.79 | 2,051.95 | 8,053.08 |
| MISSOURI | 2,092.97 | 2,150.82 | 2,187.26 | 2,166.59 | 8,597.65 |
| MONTANA | 1,253.20 | 1,344.17 | 1,316.28 | 1,343.68 | 5,257.32 |
| NEBRASKA | 1,294.75 | 1,313.85 | 1,343.51 | 1,353.99 | 5,306.10 |
| NEVADA | 841.33 | 900.66 | 892.49 | 925.73 | 3,560.21 |
| NEW HAMPSHIRE | 2,623.24 | 2,735.81 | 2,782.67 | 2,792.79 | 10,934.50 |
| NEW JERSEY | 1,076.51 | 1,129.09 | 1,129.58 | 1,158.94 | 4,494.12 |
| NEW MEXICO | 925.96 | 955.02 | 918.14 | 945.87 | 3,744.99 |
| NEW YORK | 1,132.51 | 1,175.74 | 1,170.11 | 1,179.21 | 4,657.57 |

DRUG CODE:1100 DRUG NAME: AMPHETAMINE

| STATE | 1ST QUARTER | 2ND QUARTER | 3RD QUARTER | 4TH QUARTER | TOTAL GRAMS |
|---|---|---|---|---|---|
| NORTH CAROLINA | 1,921.68 | 2,004.43 | 2,026.69 | 2,054.80 | 8,007.60 |
| NORTH DAKOTA | 1,363.74 | 1,340.62 | 1,332.59 | 1,203.64 | 5,240.59 |
| OHIO | 1,756.04 | 1,718.82 | 1,745.46 | 1,816.92 | 7,037.24 |
| OKLAHOMA | 1,315.05 | 1,344.66 | 1,377.34 | 1,377.72 | 5,414.77 |
| OREGON | 1,421.97 | 1,526.95 | 1,494.74 | 1,485.17 | 5,928.83 |
| PENNSYLVANIA | 1,673.87 | 1,659.23 | 1,631.73 | 1,641.98 | 6,606.80 |
| PUERTO RICO | 211.3 | 211.61 | 223.56 | 227.73 | 874.21 |
| RHODE ISLAND | 3,194.63 | 3,321.18 | 3,346.21 | 3,368.14 | 13,230.16 |
| SOUTH CAROLINA | 2,747.30 | 2,838.28 | 2,897.82 | 2,875.97 | 11,359.36 |
| SOUTH DAKOTA | 2,101.27 | 1,770.32 | 1,962.22 | 1,886.93 | 7,720.74 |
| TENNESSEE | 1,890.80 | 1,955.09 | 2,009.44 | 2,012.21 | 7,867.55 |
| TEXAS | 1,457.81 | 1,529.59 | 1,545.14 | 1,568.67 | 6,101.21 |
| UTAH | 2,581.57 | 2,703.85 | 2,774.89 | 2,800.78 | 10,861.09 |
| VERMONT | 1,545.42 | 1,575.04 | 1,539.63 | 1,579.70 | 6,239.79 |

| | | | | | |
|---|---|---|---|---|---|
| VIRGIN ISLANDS | 280.64 | 283.47 | 269.89 | 305.33 | 1,139.32 |
| VIRGINIA | 2,086.02 | 2,177.38 | 2,180.68 | 2,194.77 | 8,638.84 |
| WASHINGTON | 1,509.56 | 1,593.59 | 1,569.53 | 1,572.77 | 6,245.46 |
| WEST VIRGINIA | 1,317.36 | 1,298.76 | 1,311.65 | 1,292.56 | 5,220.34 |
| WISCONSIN | 2,380.55 | 2,377.69 | 2,351.34 | 2,325.74 | 9,435.32 |
| WYOMING | 1,116.94 | 1,131.06 | 1,134.29 | 1,120.71 | 4,502.99 |
| U.S. TOTAL | 1,550.24 | 1,596.58 | 1,607.03 | 1,630.76 | 6,384.61 |

DRUG CODE:1105B DRUG NAME: DL-METHAMPHETAMINE RACEMIC BASE

| STATE | 1ST QUARTER | 2ND QUARTER | 3RD QUARTER | 4TH QUARTER | TOTAL GRAMS |
|---|---|---|---|---|---|
| ALABAMA | 0.22 | 0.18 | 0.12 | 0.17 | 0.69 |
| ALASKA | 0 | 0.11 | 0 | 0 | 0.11 |
| ARIZONA | 0.32 | 0.31 | 0.28 | 0.4 | 1.31 |
| ARKANSAS | 0.04 | 0.03 | 0.1 | 0.04 | 0.21 |
| CALIFORNIA | 0.24 | 0.26 | 0.21 | 0.24 | 0.95 |
| COLORADO | 0.13 | 0.03 | 0.02 | 0.03 | 0.21 |
| CONNECTICUT | 0.33 | 0.25 | 0.18 | 0.26 | 1.01 |
| DISTRICT OF COLUMBIA | 0.6 | 0.6 | 0.47 | 0.2 | 1.87 |
| FLORIDA | 0.07 | 0.04 | 0.04 | 0.04 | 0.19 |
| GEORGIA | 0.07 | 0.07 | 0.05 | 0.07 | 0.25 |
| HAWAII | 0.27 | 0.24 | 0.21 | 0.21 | 0.92 |
| IDAHO | 0.28 | 0.36 | 0.26 | 0.26 | 1.15 |
| ILLINOIS | 0.03 | 0.03 | 0.04 | 0.02 | 0.12 |
| INDIANA | 0.06 | 0.06 | 0.1 | 0.09 | 0.31 |
| IOWA | 0.26 | 0.32 | 0.22 | 0.29 | 1.09 |
| KANSAS | 0.06 | 0.07 | 0.06 | 0.03 | 0.21 |
| KENTUCKY | 0.09 | 0.04 | 0.06 | 0.06 | 0.24 |
| LOUISIANA | 0.08 | 0.08 | 0.04 | 0.05 | 0.25 |
| MAINE | 0.18 | 0 | 0.03 | 0 | 0.21 |
| MARYLAND | 0.05 | 0.05 | 0.03 | 0.08 | 0.21 |
| MASSACHUSETTS | 0.09 | 0.06 | 0.07 | 0.04 | 0.27 |
| MICHIGAN | 0.01 | 0 | 0.04 | 0 | 0.05 |
| MINNESOTA | 0.75 | 0.45 | 0.77 | 0.71 | 2.68 |
| MISSISSIPPI | 0.01 | 0.05 | 0.01 | 0.01 | 0.09 |
| MISSOURI | 0.48 | 0.56 | 0.26 | 0.33 | 1.62 |
| NEBRASKA | 0.22 | 0.22 | 0.18 | 0.11 | 0.73 |
| NEVADA | 0.19 | 0.1 | 0.06 | 0.16 | 0.52 |
| NEW HAMPSHIRE | 0.03 | 0.06 | 0.06 | 0.03 | 0.18 |
| NEW JERSEY | 0.16 | 0.13 | 0.03 | 0.06 | 0.38 |
| NEW MEXICO | 0.76 | 0.49 | 0.55 | 0.72 | 2.52 |
| NEW YORK | 0.07 | 0.08 | 0.08 | 0.09 | 0.32 |
| NORTH CAROLINA | 0.15 | 0.15 | 0.14 | 0.12 | 0.56 |
| NORTH DAKOTA | 0.18 | 0.18 | 0.18 | 0.18 | 0.72 |
| OHIO | 0.09 | 0.09 | 0.07 | 0.08 | 0.32 |
| OKLAHOMA | 0.19 | 0.22 | 0.21 | 0.14 | 0.77 |
| OREGON | 0.49 | 0.47 | 0.41 | 0.37 | 1.74 |
| PENNSYLVANIA | 0.12 | 0.11 | 0.12 | 0.08 | 0.43 |
| RHODE ISLAND | 0.27 | 0.08 | 0.08 | 0.11 | 0.53 |
| SOUTH CAROLINA | 0.08 | 0.06 | 0.09 | 0.06 | 0.29 |
| TENNESSEE | 0.03 | 0.01 | 0 | 0 | 0.04 |
| TEXAS | 0.21 | 0.19 | 0.15 | 0.14 | 0.69 |
| UTAH | 0.07 | 0 | 0 | 0 | 0.07 |

DRUG CODE:1105B DRUG NAME: DL-METHAMPHETAMINE RACEMIC BASE

| STATE | 1ST QUARTER | 2ND QUARTER | 3RD QUARTER | 4TH QUARTER | TOTAL GRAMS |
|---|---|---|---|---|---|
| VIRGINIA | 0 | 0.02 | 0.02 | 0 | 0.04 |
| WASHINGTON | 0.05 | 0.04 | 0.04 | 0.01 | 0.14 |
| WEST VIRGINIA | 0.11 | 0.15 | 0.15 | 0.11 | 0.52 |
| WISCONSIN | 0.33 | 0.33 | 0.35 | 0.34 | 1.34 |
| WYOMING | 0.07 | 0 | 0 | 0 | 0.07 |
| U.S. TOTAL | 0.15 | 0.14 | 0.12 | 0.13 | 0.54 |

DRUG CODE:1105D DRUG NAME: D-METHAMPHETAMINE

| STATE | 1ST QUARTER | 2ND QUARTER | 3RD QUARTER | 4TH QUARTER | TOTAL GRAMS |
|---|---|---|---|---|---|
| ALABAMA | 0.5 | 0.5 | 0.42 | 0.39 | 1.82 |
| ALASKA | 0.62 | 0.57 | 0.23 | 0.34 | 1.75 |
| ARIZONA | 0.28 | 0.26 | 0.23 | 0.29 | 1.05 |
| ARKANSAS | 0.11 | 0.14 | 0.1 | 0.14 | 0.48 |
| CALIFORNIA | 0.67 | 0.68 | 0.8 | 0.76 | 2.92 |
| COLORADO | 0.64 | 0.99 | 0.93 | 0.81 | 3.38 |
| CONNECTICUT | 0.35 | 0.51 | 0.48 | 0.51 | 1.84 |
| DELAWARE | 0.49 | 0.63 | 0.49 | 0.67 | 2.28 |
| DISTRICT OF COLUMBIA | 0.33 | 0 | 0 | 0.27 | 0.6 |
| FLORIDA | 0.09 | 0.13 | 0.11 | 0.11 | 0.45 |
| GEORGIA | 0.27 | 0.32 | 0.37 | 0.29 | 1.24 |
| HAWAII | 0.18 | 0.24 | 0.3 | 0.38 | 1.09 |
| IDAHO | 0.41 | 0.41 | 0.46 | 0.28 | 1.56 |
| ILLINOIS | 0.33 | 0.45 | 0.44 | 0.41 | 1.62 |
| INDIANA | 0.27 | 0.26 | 0.28 | 0.3 | 1.11 |
| IOWA | 0.38 | 0.47 | 0.63 | 0.45 | 1.94 |
| KANSAS | 0.42 | 0.48 | 0.54 | 0.37 | 1.8 |
| KENTUCKY | 0.12 | 0.14 | 0.11 | 0.11 | 0.48 |
| LOUISIANA | 0.19 | 0.66 | 0.31 | 0.33 | 1.49 |
| MAINE | 0.18 | 0.12 | 0.18 | 0.15 | 0.64 |
| MARYLAND | 0.89 | 0.77 | 0.74 | 0.81 | 3.21 |
| MASSACHUSETTS | 0.52 | 0.64 | 0.58 | 0.66 | 2.4 |
| MICHIGAN | 0.3 | 0.28 | 0.28 | 0.29 | 1.15 |
| MINNESOTA | 1.39 | 1.32 | 1.37 | 1.37 | 5.45 |
| MISSISSIPPI | 0.38 | 0.32 | 0.32 | 0.3 | 1.33 |
| MISSOURI | 0.78 | 0.87 | 0.72 | 0.7 | 3.07 |
| MONTANA | 0.41 | 0.37 | 0.16 | 0.04 | 0.97 |
| NEBRASKA | 0.48 | 0.4 | 0.37 | 0.29 | 1.54 |
| NEVADA | 0.67 | 0.83 | 0.91 | 0.74 | 3.15 |
| NEW HAMPSHIRE | 0.46 | 0.4 | 0.55 | 0.59 | 1.99 |
| NEW JERSEY | 0.37 | 0.43 | 0.32 | 0.37 | 1.49 |
| NEW MEXICO | 0.72 | 0.92 | 0.59 | 0.88 | 3.1 |
| NEW YORK | 0.41 | 0.4 | 0.33 | 0.39 | 1.53 |
| NORTH CAROLINA | 0.52 | 0.47 | 0.56 | 0.48 | 2.03 |
| NORTH DAKOTA | 0.24 | 0.12 | 0.18 | 0.54 | 1.08 |
| OHIO | 0.84 | 0.97 | 0.48 | 0.45 | 2.74 |
| OKLAHOMA | 0.17 | 0.21 | 0.25 | 0.32 | 0.95 |
| OREGON | 1.25 | 1.09 | 1.23 | 0.99 | 4.55 |
| PENNSYLVANIA | 0.85 | 1.11 | 0.95 | 0.93 | 3.84 |
| RHODE ISLAND | 0.19 | 0.19 | 0.23 | 0.27 | 0.88 |
| SOUTH CAROLINA | 0.16 | 0.27 | 0.23 | 0.41 | 1.07 |
| SOUTH DAKOTA | 1.73 | 1.09 | 1.73 | 1.48 | 6.02 |

DRUG CODE:1105D DRUG NAME: D-METHAMPHETAMINE

| STATE | 1ST QUARTER | 2ND QUARTER | 3RD QUARTER | 4TH QUARTER | TOTAL GRAMS |
|---|---|---|---|---|---|
| TENNESSEE | 0.23 | 0.16 | 0.16 | 0.16 | 0.72 |
| TEXAS | 0.27 | 0.33 | 0.32 | 0.3 | 1.22 |
| UTAH | 1.34 | 1.32 | 1.06 | 0.97 | 4.69 |
| VERMONT | 0.83 | 0.06 | 0.39 | 0.26 | 1.54 |
| VIRGINIA | 0.64 | 0.71 | 0.68 | 0.69 | 2.72 |
| WASHINGTON | 1.28 | 1.49 | 1.34 | 1.34 | 5.46 |
| WEST VIRGINIA | 0.02 | 0.02 | 0.07 | 0 | 0.11 |
| WISCONSIN | 0.64 | 0.86 | 0.78 | 0.96 | 3.24 |
| WYOMING | 1.07 | 1.07 | 1 | 1.21 | 4.35 |
| U.S. TOTAL | 0.5 | 0.55 | 0.52 | 0.51 | 2.08 |

DRUG CODE:1205 DRUG NAME: LISDEXAMFETAMINE

| STATE | 1ST QUARTER | 2ND QUARTER | 3RD QUARTER | 4TH QUARTER | TOTAL GRAMS |
|---|---|---|---|---|---|
| ALABAMA | 1,513.04 | 1,442.46 | 1,479.46 | 1,528.60 | 5,963.56 |
| ALASKA | 353.67 | 335.84 | 335.6 | 352.53 | 1,377.63 |
| ARIZONA | 528.4 | 521.71 | 524.09 | 551.3 | 2,125.50 |
| ARKANSAS | 1,080.38 | 1,010.10 | 1,021.40 | 1,101.57 | 4,213.45 |
| CALIFORNIA | 282.78 | 289.75 | 287.49 | 304.11 | 1,164.13 |
| COLORADO | 528.47 | 524.1 | 531.52 | 559.18 | 2,143.27 |
| CONNECTICUT | 965.45 | 983.79 | 969.97 | 1,025.70 | 3,944.92 |
| DELAWARE | 1,615.64 | 1,577.79 | 1,523.77 | 1,655.50 | 6,372.70 |
| DISTRICT OF COLUMBIA | 762.54 | 779.55 | 826.14 | 841.51 | 3,209.74 |
| FLORIDA | 744.53 | 728.08 | 710.96 | 763.52 | 2,947.08 |
| GEORGIA | 1,100.41 | 1,051.41 | 1,074.86 | 1,109.78 | 4,336.47 |
| GUAM | 12.31 | 10.37 | 4.86 | 24.62 | 52.16 |
| HAWAII | 88.48 | 92.18 | 109.77 | 90.99 | 381.42 |
| IDAHO | 522.09 | 537.95 | 513.72 | 553.58 | 2,127.33 |
| ILLINOIS | 550.29 | 549.88 | 544.74 | 571.44 | 2,216.35 |
| INDIANA | 1,296.98 | 1,251.08 | 1,268.02 | 1,323.77 | 5,139.84 |
| IOWA | 1,132.52 | 1,116.33 | 1,093.23 | 1,178.89 | 4,520.97 |
| KANSAS | 1,037.60 | 1,044.20 | 1,062.34 | 1,150.40 | 4,294.54 |
| KENTUCKY | 971.89 | 949.19 | 973.76 | 1,012.72 | 3,907.56 |
| LOUISIANA | 2,200.55 | 2,057.69 | 2,080.12 | 2,177.14 | 8,515.51 |
| MAINE | 1,222.82 | 1,278.13 | 1,231.70 | 1,262.25 | 4,994.90 |
| MARYLAND | 908.62 | 933.14 | 907.06 | 973.69 | 3,722.52 |
| MASSACHUSETTS | 496.52 | 514.37 | 499.72 | 542.44 | 2,053.04 |
| MICHIGAN | 590.05 | 592.01 | 568.04 | 609.33 | 2,359.43 |
| MINNESOTA | 576.27 | 594.1 | 592.42 | 638.75 | 2,401.54 |
| MISSISSIPPI | 1,307.93 | 1,202.20 | 1,258.89 | 1,359.58 | 5,128.60 |
| MISSOURI | 1,117.22 | 1,092.36 | 1,102.58 | 1,171.05 | 4,483.21 |
| MONTANA | 529.25 | 538.13 | 520.42 | 578.09 | 2,165.88 |
| NEBRASKA | 842.14 | 840.4 | 867.24 | 921.34 | 3,471.12 |
| NEVADA | 259.64 | 262.32 | 250.68 | 285.67 | 1,058.31 |
| NEW HAMPSHIRE | 1,031.43 | 997.26 | 975.57 | 1,038.19 | 4,042.44 |
| NEW JERSEY | 546.47 | 557.14 | 531.86 | 572.22 | 2,207.69 |
| NEW MEXICO | 342.59 | 330.72 | 317.24 | 332.09 | 1,322.64 |
| NEW YORK | 408.77 | 426.88 | 405.05 | 436.49 | 1,677.19 |
| NORTH CAROLINA | 1,037.08 | 1,045.47 | 1,007.71 | 1,070.98 | 4,161.23 |
| NORTH DAKOTA | 853.93 | 818 | 829.94 | 791.09 | 3,292.96 |

| | | | | | |
|---|---|---|---|---|---|
| OHIO | 997.66 | 980.92 | 966.47 | 1,012.19 | 3,957.23 |
| OKLAHOMA | 935.58 | 911.07 | 914.3 | 985.25 | 3,746.21 |
| OREGON | 361.73 | 383.46 | 376.73 | 396.6 | 1,518.53 |
| PENNSYLVANIA | 635.96 | 642.17 | 632.52 | 669.46 | 2,580.10 |
| PUERTO RICO | 48.05 | 44.13 | 43.71 | 52.91 | 188.8 |
| RHODE ISLAND | 679.07 | 694.72 | 695.16 | 723.44 | 2,792.39 |

DRUG CODE:1205 DRUG NAME: LISDEXAMFETAMINE

| STATE | 1ST QUARTER | 2ND QUARTER | 3RD QUARTER | 4TH QUARTER | TOTAL GRAMS |
|---|---|---|---|---|---|
| SOUTH CAROLINA | 1,300.58 | 1,262.27 | 1,257.28 | 1,337.75 | 5,157.88 |
| SOUTH DAKOTA | 1,465.02 | 1,434.49 | 1,443.65 | 1,465.10 | 5,808.27 |
| TENNESSEE | 1,136.65 | 1,106.09 | 1,124.67 | 1,170.16 | 4,537.57 |
| TEXAS | 1,092.49 | 1,056.96 | 1,046.03 | 1,125.47 | 4,320.94 |
| UTAH | 707.62 | 690.82 | 715.29 | 788.1 | 2,901.83 |
| VERMONT | 1,023.36 | 1,010.99 | 1,012.74 | 1,046.74 | 4,093.83 |
| VIRGIN ISLANDS | 78.96 | 73.69 | 61.06 | 55.27 | 268.98 |
| VIRGINIA | 836.12 | 841.12 | 818.69 | 868.33 | 3,364.25 |
| WASHINGTON | 461.72 | 474.23 | 464.01 | 494.05 | 1,894.01 |
| WEST VIRGINIA | 1,107.32 | 1,093.15 | 1,064.14 | 1,113.52 | 4,378.13 |
| WISCONSIN | 1,005.42 | 1,042.08 | 1,041.58 | 1,124.52 | 4,213.59 |
| WYOMING | 461.45 | 482.38 | 466.07 | 484.02 | 1,893.92 |
| U.S. TOTAL | 765.71 | 756.53 | 750.55 | 796.51 | 3,069.30 |

DRUG CODE:1724 DRUG NAME: METHYLPHENIDATE (DL;D;L;ISOMERS)

| STATE | 1ST QUARTER | 2ND QUARTER | 3RD QUARTER | 4TH QUARTER | TOTAL GRAMS |
|---|---|---|---|---|---|
| ALABAMA | 1,581.13 | 1,452.21 | 1,492.22 | 1,559.71 | 6,085.28 |
| ALASKA | 1,203.93 | 1,195.86 | 1,064.89 | 1,156.15 | 4,620.83 |
| AMERICAN SAMOA | 1.98 | 2.64 | 13.83 | 3.95 | 22.4 |
| ARIZONA | 1,192.11 | 1,116.16 | 1,115.72 | 1,182 | 4,605.99 |
| ARKANSAS | 1,523.37 | 1,366.92 | 1,347.76 | 1,429.40 | 5,667.45 |
| CALIFORNIA | 766.14 | 747.78 | 703.57 | 754.36 | 2,971.85 |
| COLORADO | 1,249.90 | 1,201.73 | 1,187.30 | 1,233.11 | 4,872.03 |
| CONNECTICUT | 1,630.72 | 1,564.05 | 1,467.32 | 1,557.24 | 6,219.33 |
| DELAWARE | 2,242.28 | 2,121.72 | 1,990.65 | 2,170.46 | 8,525.11 |
| DISTRICT OF COLUMBIA | 1,394.29 | 1,329.90 | 1,400.67 | 1,334 | 5,458.87 |
| FLORIDA | 1,162.07 | 1,084.71 | 1,054.10 | 1,129.89 | 4,430.77 |
| GEORGIA | 1,815.03 | 1,632.89 | 1,681.45 | 1,733.83 | 6,863.20 |
| GUAM | 245.03 | 222.64 | 245.61 | 320.91 | 1,034.19 |
| HAWAII | 860.37 | 843.58 | 867.5 | 822.13 | 3,393.59 |
| IDAHO | 1,839.77 | 1,829.36 | 1,694.47 | 1,782.87 | 7,146.46 |
| ILLINOIS | 1,463.92 | 1,373.43 | 1,316.70 | 1,426.58 | 5,580.64 |
| INDIANA | 1,997.74 | 1,863.44 | 1,823.69 | 1,931.48 | 7,616.35 |
| IOWA | 2,859.01 | 2,705.61 | 2,625.22 | 2,782.35 | 10,972.19 |
| KANSAS | 1,856.27 | 1,751.32 | 1,740.57 | 1,850.69 | 7,198.85 |
| KENTUCKY | 1,855.73 | 1,706.23 | 1,728.19 | 1,767.29 | 7,057.43 |
| LOUISIANA | 2,059.80 | 1,774.52 | 1,827.14 | 1,967.93 | 7,629.40 |
| MAINE | 2,799.61 | 2,797.17 | 2,661.99 | 2,659 | 10,917.77 |
| MARYLAND | 1,812.34 | 1,759.21 | 1,702.17 | 1,775.77 | 7,049.48 |
| MASSACHUSETTS | 2,363.72 | 2,308.48 | 2,125.69 | 2,305.05 | 9,102.94 |
| MICHIGAN | 2,040.07 | 1,958.29 | 1,705.90 | 2,010.61 | 7,714.87 |
| MINNESOTA | 1,961.17 | 1,893.13 | 1,811.25 | 1,960.27 | 7,625.82 |

| | | | | | |
|---|---|---|---|---|---|
| MISSISSIPPI | 1,606.74 | 1,393.72 | 1,456.65 | 1,497.89 | 5,954.99 |
| MISSOURI | 2,253.47 | 2,160.85 | 2,117.27 | 2,282.36 | 8,813.95 |
| MONTANA | 1,874.60 | 1,774.39 | 1,665.26 | 1,830.58 | 7,144.84 |
| NEBRASKA | 1,807.77 | 1,716.77 | 1,685.62 | 1,777.41 | 6,987.56 |
| NEVADA | 567.86 | 535.6 | 506.05 | 535.09 | 2,144.60 |
| NEW HAMPSHIRE | 2,657.81 | 2,631.17 | 2,491.41 | 2,651.27 | 10,431.65 |
| NEW JERSEY | 1,089.75 | 1,081.85 | 962.04 | 1,066.84 | 4,200.47 |
| NEW MEXICO | 1,237.26 | 1,144.77 | 1,117.48 | 1,128.97 | 4,628.49 |
| NEW YORK | 1,239.35 | 1,247 | 1,112.28 | 1,213.34 | 4,811.96 |
| NORTH CAROLINA | 2,099.15 | 2,045.54 | 1,915.74 | 2,093.72 | 8,154.16 |
| NORTH DAKOTA | 2,447.02 | 2,322.13 | 2,175.84 | 2,133.76 | 9,078.74 |
| OHIO | 2,145.79 | 1,971.76 | 1,967.65 | 1,951.37 | 8,036.58 |
| OKLAHOMA | 1,397.13 | 1,299.50 | 1,272.11 | 1,331.53 | 5,300.26 |
| OREGON | 1,689.51 | 1,688.30 | 1,575.74 | 1,609.54 | 6,563.08 |
| PENNSYLVANIA | 1,914.05 | 1,790.31 | 1,722.03 | 1,734.58 | 7,160.96 |
| PUERTO RICO | 635.6 | 546.65 | 540.2 | 605.25 | 2,327.70 |

DRUG CODE:1724 DRUG NAME: METHYLPHENIDATE (DL;D;L;ISOMERS)

| STATE | 1ST QUARTER | 2ND QUARTER | 3RD QUARTER | 4TH QUARTER | TOTAL GRAMS |
|---|---|---|---|---|---|
| RHODE ISLAND | 1,915.92 | 1,864.29 | 1,714.51 | 1,865.68 | 7,360.40 |
| SOUTH CAROLINA | 2,069.84 | 1,878.06 | 1,839.66 | 1,968.11 | 7,755.66 |
| SOUTH DAKOTA | 2,999.13 | 2,796.60 | 2,805.31 | 2,389.86 | 10,990.90 |
| TENNESSEE | 1,423.53 | 1,307.10 | 1,323.14 | 1,376.18 | 5,429.95 |
| TEXAS | 1,432.14 | 1,314.28 | 1,253.17 | 1,382.74 | 5,382.33 |
| UTAH | 1,713.38 | 1,667.81 | 1,634.90 | 1,695.09 | 6,711.18 |
| VERMONT | 3,107.19 | 3,058.36 | 2,923.65 | 3,012.26 | 12,101.46 |
| VIRGIN ISLANDS | 196.23 | 183.88 | 167.17 | 180.49 | 727.77 |
| VIRGINIA | 1,752.85 | 1,711.13 | 1,598.41 | 1,712.50 | 6,774.90 |
| WASHINGTON | 1,688.04 | 1,682.13 | 1,570.98 | 1,650.20 | 6,591.35 |
| WEST VIRGINIA | 1,818.42 | 1,714.52 | 1,678.54 | 1,766.59 | 6,978.07 |
| WISCONSIN | 1,899.47 | 1,841.79 | 1,716.22 | 1,856.23 | 7,313.70 |
| WYOMING | 1,638.24 | 1,462.21 | 1,415.48 | 1,502.79 | 6,018.72 |
| U.S. TOTAL | 1,555.31 | 1,474.61 | 1,412.69 | 1,503.16 | 5,945.77 |

DRUG CODE:2012 DRUG NAME: GAMMA HYDROXYBUTYRIC ACID(FDA APPROVED-CSA III)

| STATE | 1ST QUARTER | 2ND QUARTER | 3RD QUARTER | 4TH QUARTER | TOTAL GRAMS |
|---|---|---|---|---|---|
| ALABAMA | 7.77 | 0 | 0 | 0 | 7.77 |
| CALIFORNIA | 12.96 | 10.57 | 5.39 | 5.58 | 34.51 |
| FLORIDA | 0.79 | 0 | 0 | 0 | 0.79 |
| GEORGIA | 3.07 | 16.87 | 6.9 | 0 | 26.84 |
| ILLINOIS | 5.21 | 2.9 | 9.84 | 5.79 | 23.74 |
| LOUISIANA | 0 | 0 | 4.92 | 11.47 | 16.39 |
| MASSACHUSETTS | 20.43 | 28.37 | 0 | 0 | 48.8 |
| MICHIGAN | 0 | 0 | 0 | 3.01 | 3.01 |
| MISSOURI | 216,757.17 | 82,551.77 | 123,385.35 | 0 | 422,694.29 |
| NEBRASKA | 12.21 | 0 | 0 | 0 | 12.21 |
| NEW YORK | 8.82 | 19.17 | 18.79 | 19.94 | 66.72 |
| NORTH CAROLINA | 16.36 | 11.69 | 12.47 | 6.23 | 46.75 |
| OHIO | 2.58 | 12.24 | 20.61 | 20.61 | 56.03 |
| SOUTH CAROLINA | 30.52 | 78.36 | 33.74 | 36.95 | 179.57 |

| | | | | | |
|---|---|---|---|---|---|
| TENNESSEE | 0 | 4.68 | 11.71 | 19.9 | 36.3 |
| TEXAS | 2.07 | 5.32 | 13.59 | 19.5 | 40.48 |
| VIRGINIA | 36.22 | 26.93 | 15.79 | 12.07 | 91.01 |
| WASHINGTON | 15.47 | 4.42 | 15.47 | 13.26 | 48.62 |
| U.S. TOTAL | 4,155.27 | 1,587.38 | 2,368.37 | 6.46 | 8,117.48 |

DRUG CODE:2100 DRUG NAME: BARBITURIC ACID DERIVIATIVE OR SALT [PER 21CFR 1308.13(C)(3)]

| STATE | 1ST QUARTER | 2ND QUARTER | 3RD QUARTER | 4TH QUARTER | TOTAL GRAMS |
|---|---|---|---|---|---|
| ALABAMA | 0 | 0 | 0.63 | 0.31 | 0.94 |
| ALASKA | 0 | 0 | 4.93 | 3.52 | 8.45 |
| ARIZONA | 0 | 2.74 | 6.18 | 0.16 | 9.07 |
| ARKANSAS | 0 | 0 | 0.17 | 0 | 0.17 |
| CALIFORNIA | 0 | 0.64 | 1.65 | 0.66 | 2.95 |
| COLORADO | 0 | 0.1 | 1.39 | 0.89 | 2.39 |
| CONNECTICUT | 0 | 0.98 | 0.84 | 1.4 | 3.22 |
| DELAWARE | 0 | 0 | 0 | 1.67 | 1.67 |
| DISTRICT OF COLUMBIA | 0 | 0.83 | 1.66 | 3.32 | 5.82 |
| FLORIDA | 0 | 0.13 | 0.82 | 0.72 | 1.68 |
| GEORGIA | 0 | 0.46 | 1.45 | 1.55 | 3.46 |
| IDAHO | 0 | 0 | 0 | 0.64 | 0.64 |
| ILLINOIS | 0 | 0.08 | 1.44 | 0.58 | 2.1 |
| INDIANA | 0 | 0.08 | 1.39 | 1.23 | 2.7 |
| KANSAS | 0 | 0 | 2.45 | 0.88 | 3.33 |
| KENTUCKY | 0 | 0.23 | 0.58 | 1.15 | 1.96 |
| LOUISIANA | 0 | 0 | 0.66 | 0.66 | 1.32 |
| MAINE | 0 | 0 | 0.75 | 1.13 | 1.88 |
| MARYLAND | 0 | 0.52 | 0.52 | 0.78 | 1.82 |
| MASSACHUSETTS | 0 | 0.08 | 0.69 | 0.46 | 1.22 |
| MICHIGAN | 0 | 0.61 | 1.32 | 0.86 | 2.78 |
| MINNESOTA | 0 | 0 | 0.38 | 0.57 | 0.94 |
| MISSISSIPPI | 0 | 0 | 1.01 | 0.17 | 1.18 |
| MISSOURI | 0 | 0.08 | 0.67 | 1.09 | 1.84 |
| MONTANA | 0 | 0.51 | 0.51 | 1.52 | 2.53 |
| NEVADA | 0 | 0.56 | 0.93 | 0.19 | 1.67 |
| NEW HAMPSHIRE | 0 | 0.38 | 2.28 | 0.76 | 3.42 |
| NEW JERSEY | 0 | 1.48 | 5.97 | 4.95 | 12.4 |
| NEW MEXICO | 0 | 0 | 0.49 | 0 | 0.49 |
| NEW YORK | 0 | 0.29 | 1.65 | 1.7 | 3.65 |
| NORTH CAROLINA | 0 | 0.31 | 1.42 | 1.42 | 3.15 |
| OHIO | 0 | 0.13 | 0.95 | 1.3 | 2.38 |
| OKLAHOMA | 0 | 0.13 | 0.93 | 0.67 | 1.73 |
| OREGON | 0 | 0 | 0.39 | 0.26 | 0.65 |
| PENNSYLVANIA | 0 | 0.12 | 1.02 | 1.42 | 2.56 |
| RHODE ISLAND | 0 | 0.95 | 4.28 | 2.85 | 8.08 |
| SOUTH CAROLINA | 0 | 0 | 0.32 | 0.32 | 0.65 |
| TENNESSEE | 0 | 0 | 0.55 | 0.16 | 0.71 |
| TEXAS | 0 | 0.16 | 1.49 | 0.91 | 2.57 |
| VERMONT | 0 | 0 | 0.8 | 0 | 0.8 |
| VIRGINIA | 0 | 0.56 | 1.37 | 1.69 | 3.62 |
| WASHINGTON | 0 | 0 | 0.15 | 0.37 | 0.52 |

DRUG CODE:2100 DRUG NAME: BARBITURIC ACID DERIVIATIVE OR SALT [PER 21CFR 1308.13(C)(3)]

| STATE | 1ST QUARTER | 2ND QUARTER | 3RD QUARTER | 4TH QUARTER | TOTAL GRAMS |
|---|---|---|---|---|---|
| WEST VIRGINIA | 0 | 0 | 0 | 2.16 | 2.16 |
| WISCONSIN | 0 | 0 | 0.09 | 0.26 | 0.35 |
| U.S. TOTAL | 0 | 0.33 | 1.32 | 0.97 | 2.62 |

DRUG CODE:2125 DRUG NAME: AMOBARBITAL (SCHEDULE 2)

| STATE | 1ST QUARTER | 2ND QUARTER | 3RD QUARTER | 4TH QUARTER | TOTAL GRAMS |
|---|---|---|---|---|---|
| ALABAMA | 0.11 | 0 | 0 | 0 | 0.11 |
| ARIZONA | 0.16 | 0.19 | 0.16 | 0.03 | 0.53 |
| ARKANSAS | 0.02 | 0 | 0 | 0 | 0.02 |
| CALIFORNIA | 0.14 | 0.08 | 0.04 | 0.02 | 0.28 |
| COLORADO | 0.53 | 0.53 | 0.23 | 0.04 | 1.31 |
| CONNECTICUT | 0.01 | 0.01 | 0 | 0 | 0.03 |
| DISTRICT OF COLUMBIA | 0 | 0.38 | 0 | 0 | 0.38 |
| FLORIDA | 0.03 | 0.07 | 0 | 0.05 | 0.15 |
| GEORGIA | 0.11 | 0.16 | 0.07 | 0.01 | 0.36 |
| IDAHO | 0.03 | 0 | 0 | 0 | 0.03 |
| ILLINOIS | 0.09 | 0.09 | 0.06 | 0.01 | 0.24 |
| INDIANA | 0.1 | 0.02 | 0 | 0 | 0.12 |
| IOWA | 0 | 0.01 | 0 | 0 | 0.01 |
| KANSAS | 0.37 | 0.22 | 0.03 | 0 | 0.62 |
| KENTUCKY | 0.16 | 0.06 | 0.12 | 0.01 | 0.35 |
| MAINE | 0.1 | 0.1 | 0.07 | 0 | 0.27 |
| MARYLAND | 0.14 | 0.13 | 0.03 | 0.05 | 0.35 |
| MASSACHUSETTS | 0.12 | 0.24 | 0.08 | 0.02 | 0.45 |
| MICHIGAN | 0.09 | 0.06 | 0.04 | 0.01 | 0.2 |
| MINNESOTA | 0.09 | 0.3 | 0.02 | 0.03 | 0.45 |
| MISSISSIPPI | 0.12 | 0.03 | 0.05 | 0 | 0.2 |
| MISSOURI | 0.11 | 0.09 | 0.07 | 0.02 | 0.28 |
| NEBRASKA | 0.05 | 0.32 | 0.22 | 0.07 | 0.67 |
| NEVADA | 0.08 | 0 | 0.03 | 0 | 0.12 |
| NEW HAMPSHIRE | 0.31 | 0.42 | 0 | 0.1 | 0.83 |
| NEW JERSEY | 0.19 | 0.18 | 0.06 | 0.03 | 0.46 |
| NEW MEXICO | 0 | 0.11 | 0.02 | 0 | 0.13 |
| NEW YORK | 0.29 | 0.29 | 0.15 | 0.05 | 0.79 |
| NORTH CAROLINA | 0.13 | 0.2 | 0.01 | 0.08 | 0.42 |
| OHIO | 0.06 | 0.08 | 0.03 | 0.06 | 0.22 |
| OKLAHOMA | 0 | 0.01 | 0 | 0 | 0.01 |
| OREGON | 0.07 | 0.07 | 0.07 | 0.06 | 0.27 |
| PENNSYLVANIA | 0.12 | 0.12 | 0.07 | 0.03 | 0.34 |
| PUERTO RICO | 0 | 0 | 0.02 | 0 | 0.02 |
| SOUTH CAROLINA | 0.14 | 0.14 | 0.09 | 0.09 | 0.45 |
| TENNESSEE | 0.05 | 0.1 | 0 | 0.02 | 0.17 |
| TEXAS | 0.09 | 0.11 | 0.05 | 0.04 | 0.29 |
| UTAH | 0.26 | 0.31 | 0.15 | 0.08 | 0.81 |
| VERMONT | 0.29 | 0 | 0 | 0 | 0.29 |
| VIRGINIA | 0.07 | 0.03 | 0.06 | 0 | 0.16 |
| WASHINGTON | 0.24 | 0.34 | 0.03 | 0.12 | 0.73 |
| WEST VIRGINIA | 0 | 0.25 | 0.05 | 0 | 0.3 |

DRUG CODE:2125 DRUG NAME: AMOBARBITAL (SCHEDULE 2)

| STATE | 1ST QUARTER | 2ND QUARTER | 3RD QUARTER | 4TH QUARTER | TOTAL GRAMS |
|---|---|---|---|---|---|
| WISCONSIN | 0.25 | 0.14 | 0.01 | 0.09 | 0.49 |
| U.S. TOTAL | 0.12 | 0.13 | 0.05 | 0.03 | 0.33 |

DRUG CODE:2165 DRUG NAME: BUTALBITAL

| STATE | 1ST QUARTER | 2ND QUARTER | 3RD QUARTER | 4TH QUARTER | TOTAL GRAMS |
|---|---|---|---|---|---|
| ALABAMA | 158.38 | 149.28 | 148.54 | 138.71 | 594.91 |
| ALASKA | 51.39 | 53.5 | 42.24 | 63.36 | 210.49 |
| ARIZONA | 195.71 | 181.71 | 184.45 | 178.58 | 740.45 |
| ARKANSAS | 165.64 | 173.19 | 169.93 | 166.33 | 675.09 |
| CALIFORNIA | 125.33 | 120.87 | 118.42 | 109.2 | 473.82 |
| COLORADO | 86.49 | 84.31 | 78.74 | 78.34 | 327.89 |
| CONNECTICUT | 180.88 | 176.97 | 169.27 | 162.84 | 689.96 |
| DELAWARE | 107.47 | 95.78 | 110.81 | 109.7 | 423.75 |
| DISTRICT OF COLUMBIA | 107.19 | 113.84 | 110.52 | 111.35 | 442.89 |
| FLORIDA | 231.34 | 192.89 | 183.79 | 190.7 | 798.72 |
| GEORGIA | 163.82 | 164.75 | 165.06 | 165.47 | 659.09 |
| GUAM | 8.41 | 2.8 | 8.41 | 0 | 19.62 |
| HAWAII | 150.33 | 140.41 | 149.6 | 141.88 | 582.22 |
| IDAHO | 124.4 | 134.28 | 130.77 | 129.18 | 518.63 |
| ILLINOIS | 121.62 | 118.93 | 120.73 | 123.26 | 484.54 |
| INDIANA | 116.75 | 118.06 | 121.15 | 111.66 | 467.63 |
| IOWA | 72.55 | 74.19 | 65.82 | 70.41 | 282.96 |
| KANSAS | 128.46 | 124.25 | 136.34 | 152.64 | 541.69 |
| KENTUCKY | 109.12 | 108.54 | 105.89 | 108.43 | 431.98 |
| LOUISIANA | 185.84 | 174.15 | 174.92 | 175.04 | 709.96 |
| MAINE | 105.39 | 105.77 | 95.23 | 98.99 | 405.39 |
| MARYLAND | 138.56 | 146.88 | 138.13 | 137.18 | 560.75 |
| MASSACHUSETTS | 90.72 | 87.67 | 85.99 | 79.11 | 343.48 |
| MICHIGAN | 170.53 | 165.83 | 164.41 | 168.86 | 669.64 |
| MINNESOTA | 68.16 | 72.68 | 67.03 | 66.93 | 274.8 |
| MISSISSIPPI | 118.79 | 112.73 | 107.17 | 113.4 | 452.09 |
| MISSOURI | 170.9 | 177.49 | 177.08 | 178.58 | 704.05 |
| MONTANA | 149.08 | 161.71 | 178.39 | 157.67 | 646.85 |
| NEBRASKA | 122.1 | 121.55 | 103.76 | 106.5 | 453.91 |
| NEVADA | 344.93 | 321.42 | 316.6 | 319.38 | 1,302.33 |
| NEW HAMPSHIRE | 99.51 | 96.85 | 96.47 | 101.41 | 394.24 |
| NEW JERSEY | 147.47 | 138.42 | 132 | 126.03 | 543.91 |
| NEW MEXICO | 107.08 | 112.67 | 109.02 | 100.28 | 429.05 |
| NEW YORK | 69.07 | 64.3 | 59.96 | 57.07 | 250.41 |
| NORTH CAROLINA | 103.61 | 99.79 | 96.69 | 100.15 | 400.24 |
| NORTH DAKOTA | 64.68 | 67.65 | 67.65 | 51.29 | 251.27 |
| OHIO | 113.03 | 111.39 | 106.96 | 98.95 | 430.33 |
| OKLAHOMA | 241.78 | 228.18 | 228.18 | 234.85 | 933 |
| OREGON | 111.98 | 101.8 | 101.15 | 104.28 | 419.2 |
| PENNSYLVANIA | 170.91 | 151.07 | 147.85 | 152.1 | 621.93 |
| PUERTO RICO | 3.76 | 4.97 | 4.56 | 4.97 | 18.25 |
| RHODE ISLAND | 113.53 | 92.63 | 93.58 | 98.33 | 398.07 |

DRUG CODE:2165 DRUG NAME: BUTALBITAL

| STATE | 1ST QUARTER | 2ND QUARTER | 3RD QUARTER | 4TH QUARTER | TOTAL GRAMS |
|---|---|---|---|---|---|
| SOUTH CAROLINA | 137.94 | 130.04 | 125.4 | 127.45 | 520.82 |
| SOUTH DAKOTA | 121.59 | 135.72 | 121.59 | 108.08 | 486.99 |
| TENNESSEE | 115.03 | 117 | 112.59 | 115.35 | 459.97 |
| TEXAS | 135.63 | 132.41 | 132.09 | 131.26 | 531.39 |
| UTAH | 178.91 | 172.22 | 171.5 | 182.89 | 705.53 |
| VERMONT | 45.55 | 49.54 | 53.54 | 33.56 | 182.18 |
| VIRGIN ISLANDS | 45.53 | 22.76 | 18.21 | 18.21 | 104.71 |
| VIRGINIA | 133.36 | 126.61 | 126.42 | 119.17 | 505.56 |
| WASHINGTON | 108.26 | 110.49 | 109.82 | 110.79 | 439.36 |
| WEST VIRGINIA | 104.97 | 91.74 | 102.54 | 103.62 | 402.86 |
| WISCONSIN | 76.67 | 77.63 | 69.54 | 67.87 | 291.72 |
| WYOMING | 151.7 | 160.57 | 156.13 | 157.91 | 626.3 |
| U.S. TOTAL | 134.7 | 128.7 | 126.15 | 124.85 | 514.4 |

DRUG CODE:2270 DRUG NAME: PENTOBARBITAL (SCHEDULE 2)

| STATE | 1ST QUARTER | 2ND QUARTER | 3RD QUARTER | 4TH QUARTER | TOTAL GRAMS |
|---|---|---|---|---|---|
| ALABAMA | 365.91 | 381.13 | 429.96 | 427.97 | 1,604.96 |
| ALASKA | 707.07 | 453.76 | 689.42 | 675.63 | 2,525.87 |
| ARIZONA | 746.26 | 795.32 | 592.88 | 770.33 | 2,904.79 |
| ARKANSAS | 311.51 | 271.28 | 253.16 | 225.43 | 1,061.38 |
| CALIFORNIA | 404.91 | 437.08 | 512.28 | 470.12 | 1,824.39 |
| COLORADO | 764.7 | 706.84 | 735.89 | 631.37 | 2,838.81 |
| CONNECTICUT | 326.5 | 378.03 | 579.99 | 437.01 | 1,721.53 |
| DELAWARE | 161.71 | 243.23 | 148.04 | 263.11 | 816.08 |
| DISTRICT OF COLUMBIA | 25.37 | 40.9 | 34.84 | 47.33 | 148.44 |
| FLORIDA | 322.49 | 528.71 | 270.33 | 345.06 | 1,466.60 |
| GEORGIA | 317.9 | 371.99 | 433.76 | 286.3 | 1,409.95 |
| GUAM | 0 | 1,802.06 | 0 | 0 | 1,802.06 |
| HAWAII | 1,159.37 | 269.71 | 430.81 | 943.26 | 2,803.14 |
| IDAHO | 1,267.70 | 1,182.75 | 1,431.48 | 1,956.10 | 5,838.03 |
| ILLINOIS | 490.66 | 537.45 | 525.85 | 445.02 | 1,998.98 |
| INDIANA | 585.7 | 577.86 | 623.87 | 570.92 | 2,358.36 |
| IOWA | 712.04 | 392.89 | 741.18 | 562.6 | 2,408.71 |
| KANSAS | 768.41 | 765.19 | 612.49 | 803.06 | 2,949.16 |
| KENTUCKY | 518.39 | 668.17 | 729.78 | 610.52 | 2,526.87 |
| LOUISIANA | 551.97 | 416.79 | 533.67 | 491.48 | 1,993.91 |
| MAINE | 1,181.97 | 1,477.22 | 1,048.72 | 1,290.92 | 4,998.82 |
| MARYLAND | 308.68 | 317.44 | 321.48 | 300.77 | 1,248.36 |
| MASSACHUSETTS | 395.03 | 354.6 | 448.91 | 419.16 | 1,617.69 |
| MICHIGAN | 822.17 | 1,008.83 | 926.46 | 983.65 | 3,741.11 |
| MINNESOTA | 450.24 | 534.96 | 627.72 | 625.54 | 2,238.46 |
| MISSISSIPPI | 242.51 | 104.83 | 292.78 | 132.64 | 772.76 |
| MISSOURI | 561.92 | 501.09 | 693.86 | 658.11 | 2,414.98 |
| MONTANA | 1,177.23 | 1,069.29 | 2,616.11 | 2,128.59 | 6,991.23 |
| NEBRASKA | 929.84 | 1,119.23 | 1,093.18 | 1,380.97 | 4,523.21 |
| NEVADA | 596.38 | 490.8 | 509.17 | 668.22 | 2,264.56 |
| NEW HAMPSHIRE | 1,056.21 | 1,081.12 | 1,388.51 | 1,208.52 | 4,734.36 |
| NEW JERSEY | 209.72 | 208.09 | 288.75 | 233.61 | 940.17 |
| NEW MEXICO | 675.55 | 650.05 | 945.38 | 1,075.22 | 3,346.19 |

| | | | | | |
|---|---|---|---|---|---|
| NEW YORK | 297.37 | 362.3 | 377.94 | 423.05 | 1,460.66 |
| NORTH CAROLINA | 550.84 | 594.24 | 648.01 | 710.09 | 2,503.19 |
| NORTH DAKOTA | 1,455.33 | 371.83 | 1,083.84 | 798.4 | 3,709.40 |
| OHIO | 533.22 | 517.09 | 591.57 | 580.33 | 2,222.21 |
| OKLAHOMA | 468.53 | 343.58 | 528.22 | 459.74 | 1,800.07 |
| OREGON | 754.72 | 983.94 | 703.1 | 853.27 | 3,295.04 |
| PENNSYLVANIA | 525.12 | 442.09 | 509.47 | 527.54 | 2,004.22 |
| PUERTO RICO | 23.97 | 107.45 | 0 | 0 | 131.42 |
| RHODE ISLAND | 438.92 | 827.78 | 633.32 | 963.42 | 2,863.45 |

DRUG CODE:2270 DRUG NAME: PENTOBARBITAL (SCHEDULE 2)

| STATE | 1ST QUARTER | 2ND QUARTER | 3RD QUARTER | 4TH QUARTER | TOTAL GRAMS |
|---|---|---|---|---|---|
| SOUTH CAROLINA | 292.51 | 394.67 | 261.29 | 362.32 | 1,310.79 |
| SOUTH DAKOTA | 832.06 | 698.51 | 686.2 | 945.9 | 3,162.66 |
| TENNESSEE | 556.64 | 511.01 | 621.99 | 613.94 | 2,303.57 |
| TEXAS | 398.98 | 625.99 | 639.82 | 562.17 | 2,226.96 |
| UTAH | 558.66 | 353.54 | 311.6 | 391.5 | 1,615.31 |
| VERMONT | 1,093.42 | 2,029.94 | 1,471.09 | 2,456.24 | 7,050.68 |
| VIRGINIA | 568 | 556.59 | 566.86 | 518.59 | 2,210.04 |
| WASHINGTON | 853.35 | 673.14 | 703.91 | 934.51 | 3,164.92 |
| WEST VIRGINIA | 520.38 | 444.82 | 606.28 | 656.84 | 2,228.33 |
| WISCONSIN | 715.44 | 691.47 | 791.96 | 671.98 | 2,870.84 |
| WYOMING | 2,555.39 | 946.12 | 1,497.77 | 1,891.92 | 6,891.21 |
| U.S. TOTAL | 493.35 | 525.37 | 552.23 | 552.82 | 2,123.77 |

DRUG CODE:2315 DRUG NAME: SECOBARBITAL (SCHEDULE 2)

| STATE | 1ST QUARTER | 2ND QUARTER | 3RD QUARTER | 4TH QUARTER | TOTAL GRAMS |
|---|---|---|---|---|---|
| ARIZONA | 0.14 | 0.14 | 0 | 0.29 | 0.57 |
| ARKANSAS | 0 | 0 | 0 | 0.63 | 0.63 |
| CALIFORNIA | 0.17 | 1.3 | 3.22 | 2.85 | 7.54 |
| COLORADO | 0.36 | 0.36 | 0.18 | 0.18 | 1.09 |
| CONNECTICUT | 0 | 0.77 | 0.26 | 0.26 | 1.28 |
| DELAWARE | 0 | 0 | 0 | 1.02 | 1.02 |
| FLORIDA | 0.24 | 0.49 | 0.44 | 0.49 | 1.66 |
| GEORGIA | 0.19 | 0.19 | 0.09 | 0.19 | 0.66 |
| HAWAII | 1.35 | 2.02 | 0.67 | 0.67 | 4.71 |
| ILLINOIS | 0.64 | 0.64 | 0.71 | 0.5 | 2.5 |
| KANSAS | 0.96 | 0.96 | 0.32 | 0.32 | 2.57 |
| LOUISIANA | 0.2 | 0.61 | 0.4 | 0 | 1.21 |
| MARYLAND | 0.63 | 0.79 | 1.11 | 0.63 | 3.17 |
| MASSACHUSETTS | 0.28 | 1.26 | 1.12 | 0.7 | 3.36 |
| MICHIGAN | 0 | 0 | 0.09 | 0.09 | 0.19 |
| MISSOURI | 0.76 | 0.76 | 0.46 | 0.46 | 2.45 |
| NEVADA | 0 | 0 | 0.34 | 0 | 0.34 |
| NEW HAMPSHIRE | 0.7 | 0.7 | 1.39 | 0 | 2.78 |
| NEW JERSEY | 0.31 | 1.04 | 1.04 | 1.25 | 3.64 |
| NEW MEXICO | 0 | 0.44 | 0.89 | 0.44 | 1.78 |
| NEW YORK | 1.7 | 3.07 | 2.41 | 2.46 | 9.64 |
| NORTH CAROLINA | 0.29 | 0.1 | 0.38 | 0.1 | 0.86 |
| OHIO | 0.4 | 0.48 | 0.48 | 0.4 | 1.75 |
| OREGON | 4.3 | 12.9 | 5.26 | 4.54 | 27 |
| PENNSYLVANIA | 0.43 | 0.86 | 0.43 | 0.5 | 2.23 |

| | | | | | |
|---|---|---|---|---|---|
| RHODE ISLAND | 1.74 | 0.87 | 1.74 | 0 | 4.35 |
| TEXAS | 0.04 | 0.11 | 0.11 | 0.04 | 0.29 |
| VERMONT | 0 | 1.46 | 5.85 | 10.24 | 17.56 |
| VIRGINIA | 0.34 | 0.34 | 0.34 | 0.57 | 1.6 |
| WASHINGTON | 2.86 | 3.54 | 1.77 | 3.95 | 12.12 |
| WEST VIRGINIA | 0 | 0.49 | 0 | 0 | 0.49 |
| WYOMING | 0 | 1.62 | 1.62 | 0 | 3.25 |
| U.S. TOTAL | 0.42 | 0.86 | 0.9 | 0.87 | 3.05 |

DRUG CODE:7379 DRUG NAME: NABILONE

| STATE | 1ST QUARTER | 2ND QUARTER | 3RD QUARTER | 4TH QUARTER | TOTAL GRAMS |
|---|---|---|---|---|---|
| ARIZONA | 0 | 0 | 0 | 0 | 0 |
| CALIFORNIA | 0 | 0 | 0 | 0 | 0 |
| COLORADO | 0.01 | 0.01 | 0 | 0 | 0.02 |
| FLORIDA | 0.01 | 0 | 0.01 | 0 | 0.02 |
| HAWAII | 0 | 0.01 | 0 | 0.01 | 0.01 |
| ILLINOIS | 0 | 0 | 0 | 0 | 0.01 |
| INDIANA | 0 | 0 | 0 | 0 | 0 |
| IOWA | 0 | 0 | 0 | 0 | 0 |
| MAINE | 0 | 0 | 0 | 0.01 | 0.01 |
| MARYLAND | 0.03 | 0.02 | 0.02 | 0.02 | 0.09 |
| MASSACHUSETTS | 0 | 0 | 0.01 | 0 | 0.01 |
| MINNESOTA | 0 | 0 | 0 | 0 | 0.01 |
| MISSOURI | 0 | 0 | 0 | 0 | 0.01 |
| NEBRASKA | 0.01 | 0.01 | 0.01 | 0.01 | 0.04 |
| NEW HAMPSHIRE | 0 | 0.01 | 0.01 | 0 | 0.02 |
| NEW JERSEY | 0 | 0 | 0 | 0 | 0 |
| NEW MEXICO | 0 | 0.01 | 0 | 0.01 | 0.01 |
| NEW YORK | 0 | 0 | 0 | 0 | 0.01 |
| NORTH CAROLINA | 0 | 0 | 0 | 0 | 0.01 |
| OHIO | 0 | 0 | 0 | 0 | 0.01 |
| PENNSYLVANIA | 0 | 0 | 0 | 0 | 0.01 |
| RHODE ISLAND | 0 | 0 | 0 | 0 | 0 |
| VIRGINIA | 0 | 0 | 0 | 0 | 0.01 |
| WISCONSIN | 0 | 0 | 0 | 0 | 0 |
| U.S. TOTAL | 0 | 0 | 0 | 0 | 0 |

DRUG CODE:7471 DRUG NAME: PHENCYCLIDINE (PCP)

| STATE | 1ST QUARTER | 2ND QUARTER | 3RD QUARTER | 4TH QUARTER | TOTAL GRAMS |
|---|---|---|---|---|---|
| MINNESOTA | 0 | 0 | 0 | 0 | 0 |
| U.S. TOTAL | 0 | 0 | 0 | 0 | 0 |

DRUG CODE:9041L DRUG NAME: COCAINE

| STATE | 1ST QUARTER | 2ND QUARTER | 3RD QUARTER | 4TH QUARTER | TOTAL GRAMS |
|---|---|---|---|---|---|
| ALABAMA | 1.16 | 1.23 | 1.26 | 1.55 | 5.19 |
| ALASKA | 2.5 | 1.55 | 1.77 | 3.09 | 8.91 |

| | | | | | |
|---|---|---|---|---|---|
| ARIZONA | 2.05 | 2.01 | 1.81 | 2 | 7.88 |
| ARKANSAS | 2.32 | 1.86 | 1.71 | 1.52 | 7.41 |
| CALIFORNIA | 2.55 | 2.53 | 2.49 | 2.53 | 10.11 |
| COLORADO | 1.93 | 1.94 | 1.62 | 1.99 | 7.48 |
| CONNECTICUT | 1.39 | 1.2 | 1.11 | 1.44 | 5.14 |
| DELAWARE | 2.66 | 2.13 | 1.22 | 2.29 | 8.3 |
| DISTRICT OF COLUMBIA | 4.56 | 3.23 | 4.56 | 3.85 | 16.19 |
| FLORIDA | 1.7 | 1.7 | 1.62 | 1.56 | 6.58 |
| GEORGIA | 1.65 | 1.82 | 1.83 | 2.24 | 7.54 |
| GUAM | 0 | 0 | 0.4 | 0 | 0.4 |
| HAWAII | 1.26 | 1.58 | 1.84 | 1.83 | 6.5 |
| IDAHO | 5.06 | 3.13 | 4.98 | 3.53 | 16.7 |
| ILLINOIS | 2.85 | 2.93 | 2.4 | 2.75 | 10.93 |
| INDIANA | 3.17 | 3.47 | 2.61 | 3.41 | 12.66 |
| IOWA | 3.06 | 3.57 | 3.02 | 3.6 | 13.25 |
| KANSAS | 2.93 | 3.71 | 2.89 | 3.32 | 12.85 |
| KENTUCKY | 2.75 | 2.82 | 1.97 | 2.55 | 10.1 |
| LOUISIANA | 2.06 | 2.03 | 1.98 | 2.44 | 8.51 |
| MAINE | 1.93 | 2 | 2.24 | 2 | 8.17 |
| MARYLAND | 2.64 | 2.65 | 2.23 | 2.16 | 9.69 |
| MASSACHUSETTS | 3.81 | 3.62 | 3.27 | 3.18 | 13.87 |
| MICHIGAN | 4.14 | 4.13 | 3.53 | 4.21 | 16.02 |
| MINNESOTA | 4.44 | 4.24 | 3.93 | 4.06 | 16.67 |
| MISSISSIPPI | 3.51 | 3.41 | 2.75 | 3.22 | 12.89 |
| MISSOURI | 3.15 | 2.75 | 3.09 | 2.89 | 11.88 |
| MONTANA | 5.01 | 4.91 | 5.57 | 5.54 | 21.03 |
| NEBRASKA | 1.93 | 1.87 | 2.38 | 2.06 | 8.24 |
| NEVADA | 1.51 | 2.11 | 2.55 | 1.87 | 8.04 |
| NEW HAMPSHIRE | 4.44 | 3.64 | 3.96 | 3.96 | 15.99 |
| NEW JERSEY | 2.04 | 1.99 | 1.93 | 1.97 | 7.93 |
| NEW MEXICO | 4.3 | 4.09 | 3.39 | 3.41 | 15.2 |
| NEW YORK | 1.97 | 1.83 | 2.01 | 1.8 | 7.61 |
| NORTH CAROLINA | 1.56 | 1.57 | 1.85 | 1.62 | 6.61 |
| NORTH DAKOTA | 6.35 | 13.96 | 3.27 | 3.76 | 27.34 |
| OHIO | 1.56 | 1.43 | 1.55 | 1.73 | 6.27 |
| OKLAHOMA | 1.24 | 0.95 | 0.83 | 1.09 | 4.11 |
| OREGON | 3.57 | 2.2 | 2.92 | 2.16 | 10.86 |
| PENNSYLVANIA | 3.51 | 3.59 | 3.26 | 3.31 | 13.68 |
| RHODE ISLAND | 2.68 | 2.18 | 1.97 | 2.31 | 9.14 |
| SOUTH CAROLINA | 0.91 | 0.76 | 0.65 | 0.74 | 3.07 |

DRUG CODE:9041L DRUG NAME: COCAINE

| STATE | 1ST QUARTER | 2ND QUARTER | 3RD QUARTER | 4TH QUARTER | TOTAL GRAMS |
|---|---|---|---|---|---|
| SOUTH DAKOTA | 3.04 | 7 | 5.14 | 2.82 | 17.99 |
| TENNESSEE | 2.95 | 3.12 | 2.91 | 3.03 | 12.02 |
| TEXAS | 1.49 | 1.5 | 1.39 | 1.78 | 6.16 |
| UTAH | 3.94 | 4.71 | 3.41 | 4.64 | 16.7 |
| VERMONT | 1.6 | 1.39 | 0.64 | 1.35 | 4.98 |
| VIRGINIA | 2.37 | 2.31 | 1.85 | 2.29 | 8.81 |
| WASHINGTON | 2.39 | 1.76 | 1.85 | 2.11 | 8.12 |
| WEST VIRGINIA | 2.71 | 2.44 | 3.14 | 2.35 | 10.64 |
| WISCONSIN | 2.73 | 3.01 | 2.4 | 2.93 | 11.06 |
| WYOMING | 3.07 | 2.71 | 4.49 | 3.75 | 14.02 |
| U.S. TOTAL | 2.4 | 2.38 | 2.22 | 2.36 | 9.36 |

DRUG CODE:9050 DRUG NAME: CODEINE

| STATE | 1ST QUARTER | 2ND QUARTER | 3RD QUARTER | 4TH QUARTER | TOTAL GRAMS |
|---|---|---|---|---|---|
| ALABAMA | 1,190.78 | 1,136.21 | 1,116.43 | 1,108.98 | 4,552.40 |
| ALASKA | 1,107.48 | 1,036.23 | 1,058.86 | 995.29 | 4,197.87 |
| AMERICAN SAMOA | 1,165.20 | 767.82 | 444.53 | 899.15 | 3,276.69 |
| ARIZONA | 1,310.49 | 1,222.54 | 1,238.91 | 1,236.89 | 5,008.84 |
| ARKANSAS | 2,678.64 | 2,683.18 | 2,655.08 | 2,596.24 | 10,613.13 |
| CALIFORNIA | 1,501.90 | 1,466.02 | 1,421.78 | 1,363.75 | 5,753.45 |
| COLORADO | 582.38 | 540.6 | 539.42 | 538.12 | 2,200.52 |
| CONNECTICUT | 812.57 | 787.82 | 764.64 | 753.61 | 3,118.64 |
| DELAWARE | 1,222.44 | 1,237.83 | 1,221.75 | 1,172.14 | 4,854.17 |
| DISTRICT OF COLUMBIA | 2,238.97 | 2,323.20 | 2,230.59 | 2,169.89 | 8,962.65 |
| FLORIDA | 1,008.01 | 923.48 | 867.51 | 912.14 | 3,711.13 |
| GEORGIA | 1,098.57 | 1,094.03 | 1,083.01 | 1,085.05 | 4,360.65 |
| GUAM | 775.79 | 732.28 | 970.86 | 564.82 | 3,043.75 |
| HAWAII | 1,165.52 | 1,082.27 | 1,066.85 | 972.77 | 4,287.41 |
| IDAHO | 983.75 | 971.23 | 945.71 | 874.41 | 3,775.10 |
| ILLINOIS | 1,357.92 | 1,367.85 | 1,318.96 | 1,354.76 | 5,399.49 |
| INDIANA | 910.2 | 946.94 | 893.76 | 859.35 | 3,610.25 |
| IOWA | 636.92 | 754.04 | 709.39 | 717.57 | 2,817.92 |
| KANSAS | 1,087.54 | 1,061.20 | 1,001.94 | 1,047.96 | 4,198.65 |
| KENTUCKY | 1,211 | 1,265.30 | 1,210.44 | 1,190.90 | 4,877.63 |
| LOUISIANA | 1,010.18 | 993.41 | 1,011.01 | 1,002.12 | 4,016.72 |
| MAINE | 916.58 | 974.68 | 939.07 | 916.57 | 3,746.89 |
| MARYLAND | 1,161.97 | 1,156.18 | 1,127.23 | 1,116.70 | 4,562.07 |
| MASSACHUSETTS | 862.57 | 831.58 | 794.4 | 759.94 | 3,248.49 |
| MICHIGAN | 2,763.13 | 2,835.48 | 2,709.63 | 2,807.88 | 11,116.12 |
| MINNESOTA | 835.62 | 815.23 | 787.11 | 763.07 | 3,201.03 |
| MISSISSIPPI | 949.99 | 955.75 | 945.86 | 963.93 | 3,815.53 |
| MISSOURI | 1,267.27 | 1,263.22 | 1,241.44 | 1,197.28 | 4,969.21 |
| MONTANA | 1,196.26 | 1,139.91 | 1,174.39 | 1,111.15 | 4,621.70 |
| NEBRASKA | 732.21 | 774.81 | 748.35 | 729.7 | 2,985.07 |
| NEVADA | 1,183.80 | 1,105.94 | 1,024.76 | 1,054.75 | 4,369.25 |
| NEW HAMPSHIRE | 694.81 | 666.55 | 665.66 | 665.04 | 2,692.06 |
| NEW JERSEY | 994.3 | 1,019.71 | 983.59 | 993.33 | 3,990.94 |
| NEW MEXICO | 1,074.08 | 993.4 | 1,026.01 | 1,001.55 | 4,095.03 |
| NEW YORK | 929.81 | 866.23 | 821.02 | 822.04 | 3,439.10 |
| NORTH CAROLINA | 564.95 | 540.22 | 529.58 | 548.4 | 2,183.16 |
| NORTH DAKOTA | 1,030.90 | 989.32 | 949.5 | 760.66 | 3,730.38 |
| OHIO | 1,037.95 | 988.34 | 967.79 | 968.08 | 3,962.17 |
| OKLAHOMA | 2,531.77 | 2,532.77 | 2,555.23 | 2,568.20 | 10,187.97 |
| OREGON | 1,002.50 | 975.15 | 951.04 | 939.83 | 3,868.52 |
| PENNSYLVANIA | 1,264.26 | 1,219.32 | 1,165.24 | 1,165.46 | 4,814.28 |
| PUERTO RICO | 331.59 | 365.9 | 359.06 | 370.81 | 1,427.36 |

DRUG CODE:9050 DRUG NAME: CODEINE

| STATE | 1ST QUARTER | 2ND QUARTER | 3RD QUARTER | 4TH QUARTER | TOTAL GRAMS |
|---|---|---|---|---|---|
| RHODE ISLAND | 865.16 | 812 | 825.09 | 767.54 | 3,269.79 |
| SOUTH CAROLINA | 1,864.25 | 1,765.12 | 1,751.51 | 1,531.78 | 6,912.65 |
| SOUTH DAKOTA | 1,022.48 | 1,033.39 | 976.25 | 967.64 | 3,999.76 |
| TENNESSEE | 1,601.85 | 1,633.09 | 1,670.26 | 1,595.22 | 6,500.43 |

| | | | | | |
|---|---|---|---|---|---|
| TEXAS | 4,004.54 | 4,063.04 | 4,112.56 | 4,139.12 | 16,319.27 |
| UTAH | 656.24 | 633.21 | 643.97 | 632.81 | 2,566.23 |
| VERMONT | 651.26 | 618.37 | 595.13 | 579.85 | 2,444.61 |
| VIRGIN ISLANDS | 647.34 | 691.32 | 662.21 | 557.43 | 2,558.30 |
| VIRGINIA | 790.64 | 769.45 | 767.21 | 734.83 | 3,062.14 |
| WASHINGTON | 1,016.04 | 1,019.88 | 977.91 | 966.27 | 3,980.11 |
| WEST VIRGINIA | 1,687.37 | 1,681.51 | 1,714.75 | 1,709.18 | 6,792.81 |
| WISCONSIN | 1,015.45 | 980.6 | 963.59 | 938.75 | 3,898.40 |
| WYOMING | 893.07 | 844.25 | 814.44 | 803.22 | 3,354.98 |
| U.S. TOTAL | 1,419.96 | 1,403.67 | 1,378.50 | 1,369.78 | 5,571.91 |

DRUG CODE:9064 DRUG NAME: BUPRENORPHINE

| STATE | 1ST QUARTER | 2ND QUARTER | 3RD QUARTER | 4TH QUARTER | TOTAL GRAMS |
|---|---|---|---|---|---|
| ALABAMA | 471.13 | 465.85 | 476.45 | 486.89 | 1,900.32 |
| ALASKA | 260.35 | 264.77 | 273.65 | 284.11 | 1,082.87 |
| AMERICAN SAMOA | 0 | 0.01 | 0 | 0 | 0.02 |
| ARIZONA | 122.11 | 127.59 | 132.71 | 135.86 | 518.27 |
| ARKANSAS | 162.72 | 169.5 | 171.51 | 184.2 | 687.92 |
| CALIFORNIA | 87.87 | 91.97 | 93.5 | 95.97 | 369.32 |
| COLORADO | 128.71 | 133.56 | 135.34 | 138 | 535.61 |
| CONNECTICUT | 366.18 | 381.97 | 385.99 | 395.23 | 1,529.38 |
| DELAWARE | 310.2 | 323.08 | 334.01 | 331.98 | 1,299.27 |
| DISTRICT OF COLUMBIA | 272.41 | 299.27 | 320.4 | 334.76 | 1,226.84 |
| FLORIDA | 221.95 | 229.16 | 228.93 | 237.11 | 917.15 |
| GEORGIA | 145.21 | 147.78 | 149.02 | 151.91 | 593.92 |
| GUAM | 1.99 | 0.27 | 0.28 | 0.01 | 2.56 |
| HAWAII | 125.65 | 131.9 | 138.18 | 134.27 | 530 |
| IDAHO | 180.83 | 193.17 | 199.27 | 201.52 | 774.79 |
| ILLINOIS | 86.27 | 92.49 | 93.63 | 98.69 | 371.08 |
| INDIANA | 235.11 | 242.39 | 246.93 | 257.85 | 982.28 |
| IOWA | 44.45 | 48.29 | 49.85 | 49.03 | 191.62 |
| KANSAS | 80.59 | 80.03 | 85.14 | 84.67 | 330.43 |
| KENTUCKY | 703.25 | 712.89 | 714.92 | 735.58 | 2,866.65 |
| LOUISIANA | 313.17 | 315.23 | 385.03 | 392.22 | 1,405.65 |
| MAINE | 555.42 | 577.93 | 615.62 | 647.17 | 2,396.15 |
| MARYLAND | 377.05 | 399.92 | 398.02 | 408.65 | 1,583.63 |
| MASSACHUSETTS | 559.04 | 572.08 | 595.9 | 592.87 | 2,319.90 |
| MICHIGAN | 236.51 | 244.97 | 247.95 | 253.35 | 982.78 |
| MINNESOTA | 107.53 | 113.17 | 117.82 | 126.63 | 465.14 |
| MISSISSIPPI | 302.76 | 312.7 | 315.3 | 319.04 | 1,249.80 |
| MISSOURI | 129.02 | 135.52 | 139.46 | 144.07 | 548.06 |
| MONTANA | 156.33 | 175.68 | 188.87 | 209.57 | 730.44 |
| NEBRASKA | 58.51 | 62.51 | 66.08 | 68.04 | 255.13 |
| NEVADA | 110.83 | 113.59 | 114.2 | 117.78 | 456.4 |
| NEW HAMPSHIRE | 495.33 | 515.63 | 565.15 | 572.63 | 2,148.74 |
| NEW JERSEY | 278.19 | 287.95 | 288.61 | 290.23 | 1,144.98 |
| NEW MEXICO | 292.88 | 303.05 | 319.56 | 319.28 | 1,234.78 |
| NEW YORK | 240.68 | 250.19 | 254.74 | 255.04 | 1,000.65 |
| NORTH CAROLINA | 263.01 | 279.26 | 283.57 | 305.21 | 1,131.05 |
| NORTH DAKOTA | 110 | 124.88 | 128.46 | 125.1 | 488.44 |
| OHIO | 390.38 | 406.4 | 418.25 | 427.39 | 1,642.42 |
| OKLAHOMA | 241.84 | 250.79 | 254.91 | 264.36 | 1,011.90 |
| OREGON | 213.91 | 223.56 | 233.29 | 233.39 | 904.16 |

| | | | | | |
|---|---|---|---|---|---|
| PENNSYLVANIA | 420.89 | 436.67 | 446.18 | 460.91 | 1,764.65 |
| PUERTO RICO | 63.33 | 66.14 | 69.52 | 68.55 | 267.54 |

DRUG CODE:9064 DRUG NAME: BUPRENORPHINE

| STATE | 1ST QUARTER | 2ND QUARTER | 3RD QUARTER | 4TH QUARTER | TOTAL GRAMS |
|---|---|---|---|---|---|
| RHODE ISLAND | 629.93 | 685.57 | 642.26 | 670.36 | 2,628.12 |
| SOUTH CAROLINA | 197.85 | 208.66 | 207.24 | 208.62 | 822.38 |
| SOUTH DAKOTA | 43.51 | 47.35 | 47.55 | 48.36 | 186.77 |
| TENNESSEE | 484.54 | 492.66 | 495.59 | 503.55 | 1,976.36 |
| TEXAS | 98.67 | 101.14 | 99.8 | 102.18 | 401.79 |
| UTAH | 309.79 | 316.36 | 327.46 | 333.06 | 1,286.68 |
| VERMONT | 797.59 | 851.21 | 883.91 | 909.33 | 3,442.03 |
| VIRGIN ISLANDS | 22.2 | 23.05 | 17.21 | 18.55 | 81.01 |
| VIRGINIA | 231.33 | 240.38 | 243.28 | 244.54 | 959.52 |
| WASHINGTON | 229.26 | 239.76 | 252.13 | 260.81 | 981.97 |
| WEST VIRGINIA | 768.16 | 783.41 | 786.7 | 803.09 | 3,141.36 |
| WISCONSIN | 160.41 | 164.61 | 168.07 | 167.18 | 660.28 |
| WYOMING | 156.31 | 185.48 | 176.79 | 194 | 712.6 |
| U.S. TOTAL | 229.15 | 237.37 | 242.44 | 248.16 | 957.12 |

DRUG CODE:9120 DRUG NAME: DIHYDROCODEINE

| STATE | 1ST QUARTER | 2ND QUARTER | 3RD QUARTER | 4TH QUARTER | TOTAL GRAMS |
|---|---|---|---|---|---|
| ALABAMA | 14.01 | 13 | 16.16 | 16.08 | 59.24 |
| ALASKA | 0 | 0 | 0.15 | 0 | 0.15 |
| ARIZONA | 0.77 | 0.62 | 0.97 | 1.11 | 3.47 |
| ARKANSAS | 3.6 | 3.34 | 2.88 | 3.54 | 13.37 |
| CALIFORNIA | 2.11 | 5.8 | 2.54 | 2.04 | 12.49 |
| COLORADO | 0 | 0.04 | 0.02 | 0 | 0.06 |
| CONNECTICUT | 0 | 0 | 0 | 0.09 | 0.09 |
| DELAWARE | 0 | 0.15 | 0.12 | 0 | 0.27 |
| FLORIDA | 0.36 | 0.53 | 0.47 | 0.59 | 1.94 |
| GEORGIA | 3.99 | 4.31 | 4.01 | 4.36 | 16.66 |
| HAWAII | 0.52 | 0.35 | 0.73 | 0.21 | 1.81 |
| IDAHO | 1.74 | 6.68 | 1.91 | 3.27 | 13.59 |
| ILLINOIS | 0.14 | 0.19 | 0.24 | 1.09 | 1.67 |
| INDIANA | 0.27 | 0.53 | 0.21 | 0.03 | 1.05 |
| KANSAS | 0.04 | 0.04 | 0.19 | 0 | 0.26 |
| KENTUCKY | 0.79 | 0.76 | 0.45 | 0.79 | 2.79 |
| LOUISIANA | 7.71 | 12.48 | 15.35 | 15.87 | 51.41 |
| MAINE | 0 | 0 | 0.08 | 0 | 0.08 |
| MARYLAND | 0.02 | 0.02 | 0.28 | 0.11 | 0.43 |
| MASSACHUSETTS | 0.29 | 0.2 | 0.15 | 0.15 | 0.78 |
| MICHIGAN | 0.97 | 0.01 | 0.03 | 0.09 | 1.1 |
| MINNESOTA | 0 | 0 | 0.06 | 0 | 0.06 |
| MISSISSIPPI | 7.48 | 8.32 | 12.26 | 10.81 | 38.88 |
| MISSOURI | 0.34 | 0.27 | 0.32 | 0.27 | 1.19 |
| NEBRASKA | 0.12 | 0.06 | 0.18 | 0 | 0.35 |
| NEVADA | 1.39 | 2.99 | 2.49 | 0.64 | 7.5 |
| NEW HAMPSHIRE | 0 | 0 | 0.08 | 0 | 0.08 |
| NEW JERSEY | 0.06 | 0.87 | 0.69 | 4.26 | 5.88 |
| NEW MEXICO | 7.31 | 6.95 | 7.78 | 7.88 | 29.93 |

| | | | | | |
|---|---|---|---|---|---|
| NEW YORK | 0.05 | 0.09 | 0.38 | 0.52 | 1.04 |
| NORTH CAROLINA | 0.1 | 0.13 | 0.1 | 0.04 | 0.38 |
| NORTH DAKOTA | 0 | 0 | 0 | 0.32 | 0.32 |
| OHIO | 0.28 | 0.25 | 0.37 | 0.27 | 1.17 |
| OKLAHOMA | 6.22 | 6.65 | 7.81 | 7.32 | 28.01 |
| PENNSYLVANIA | 1.71 | 2.35 | 2.16 | 2.69 | 8.91 |
| RHODE ISLAND | 0 | 0.1 | 0.51 | 0 | 0.61 |
| SOUTH CAROLINA | 0.4 | 0.44 | 0.21 | 0.28 | 1.33 |
| TENNESSEE | 11.86 | 17.08 | 21.03 | 16.9 | 66.87 |
| TEXAS | 8.72 | 9.42 | 11.27 | 11.37 | 40.78 |
| VIRGINIA | 0.11 | 0.09 | 0.12 | 0.07 | 0.39 |
| WASHINGTON | 0 | 0.02 | 0 | 0 | 0.02 |
| WEST VIRGINIA | 0.36 | 0.35 | 0.07 | 0.92 | 1.69 |

DRUG CODE:9120 DRUG NAME: DIHYDROCODEINE

| STATE | 1ST QUARTER | 2ND QUARTER | 3RD QUARTER | 4TH QUARTER | TOTAL GRAMS |
|---|---|---|---|---|---|
| WISCONSIN | 0.17 | 0.06 | 0.06 | 0.09 | 0.38 |
| U.S. TOTAL | 2.1 | 2.84 | 2.8 | 2.82 | 10.56 |

DRUG CODE:9143 DRUG NAME: OXYCODONE

| STATE | 1ST QUARTER | 2ND QUARTER | 3RD QUARTER | 4TH QUARTER | TOTAL GRAMS |
|---|---|---|---|---|---|
| ALABAMA | 5,748.57 | 5,593.84 | 5,568.29 | 5,327.90 | 22,238.61 |
| ALASKA | 5,536.92 | 5,281.95 | 5,183.08 | 5,164.54 | 21,166.48 |
| AMERICAN SAMOA | 38.93 | 65.57 | 49.18 | 49.18 | 202.86 |
| ARIZONA | 8,116.30 | 7,663.46 | 7,550.14 | 7,422.94 | 30,752.84 |
| ARKANSAS | 5,710.04 | 5,450.30 | 5,302.03 | 5,272.13 | 21,734.50 |
| CALIFORNIA | 2,747.33 | 2,645.47 | 2,563.18 | 2,534.36 | 10,490.34 |
| COLORADO | 5,232.92 | 5,005.91 | 4,958.12 | 4,809.30 | 20,006.24 |
| CONNECTICUT | 5,877.63 | 5,647.61 | 5,385.53 | 5,172.83 | 22,083.60 |
| DELAWARE | 8,733.99 | 8,351.25 | 8,381.44 | 8,258.38 | 33,725.06 |
| DISTRICT OF COLUMBIA | 2,929.21 | 2,731.06 | 2,823.90 | 2,642.15 | 11,126.32 |
| FLORIDA | 5,444.94 | 5,401.33 | 5,417.15 | 5,465.82 | 21,729.25 |
| GEORGIA | 4,743.13 | 4,691.54 | 4,761.61 | 4,738.19 | 18,934.47 |
| GUAM | 498.9 | 563.72 | 554.19 | 605.3 | 2,222.11 |
| HAWAII | 3,771.50 | 3,634.72 | 3,523.34 | 3,405.08 | 14,334.64 |
| IDAHO | 4,133.39 | 4,079.54 | 3,993.85 | 4,043.34 | 16,250.12 |
| ILLINOIS | 1,367.50 | 1,347.58 | 1,345.12 | 1,341.26 | 5,401.45 |
| INDIANA | 4,213.98 | 4,029.13 | 3,926.72 | 3,911.24 | 16,081.08 |
| IOWA | 2,071.99 | 1,995.84 | 1,944.18 | 1,911.71 | 7,923.72 |
| KANSAS | 4,692.16 | 4,599.98 | 4,631.08 | 4,504.19 | 18,427.42 |
| KENTUCKY | 5,147.39 | 5,069.97 | 4,998.57 | 4,893.94 | 20,109.87 |
| LOUISIANA | 4,763.10 | 4,740.57 | 4,684.98 | 4,588.69 | 18,777.34 |
| MAINE | 5,811.74 | 5,524.84 | 5,137.68 | 4,712.85 | 21,187.10 |
| MARYLAND | 6,410.21 | 6,227.99 | 6,249.35 | 6,094.97 | 24,982.51 |
| MASSACHUSETTS | 4,604.84 | 4,371.28 | 4,159.95 | 3,982.62 | 17,118.69 |
| MICHIGAN | 3,596.05 | 3,444.91 | 3,321.94 | 3,288.89 | 13,651.78 |
| MINNESOTA | 3,182.68 | 3,071.05 | 3,021.74 | 2,976.70 | 12,252.18 |
| MISSISSIPPI | 3,751.35 | 3,724.87 | 3,745.07 | 3,774.68 | 14,995.96 |
| MISSOURI | 5,682.05 | 5,548.97 | 5,576.86 | 5,354.55 | 22,162.43 |
| MONTANA | 4,930.51 | 4,686.37 | 4,541.13 | 4,338.13 | 18,496.13 |
| NEBRASKA | 2,472.98 | 2,402.32 | 2,456.78 | 2,425.41 | 9,757.49 |

| | | | | | |
|---|---|---|---|---|---|
| NEVADA | 7,557.93 | 7,165.62 | 7,069.97 | 7,104.05 | 28,897.57 |
| NEW HAMPSHIRE | 5,934.78 | 5,589.20 | 5,443.07 | 5,164.21 | 22,131.26 |
| NEW JERSEY | 6,980.58 | 6,778.15 | 6,629.16 | 6,409.08 | 26,796.98 |
| NEW MEXICO | 5,678.07 | 5,403.14 | 5,274.11 | 5,167.87 | 21,523.18 |
| NEW YORK | 4,679.47 | 4,476 | 4,358.12 | 4,269.81 | 17,783.40 |
| NORTH CAROLINA | 6,936.10 | 6,663.85 | 6,532.60 | 6,411.45 | 26,544 |
| NORTH DAKOTA | 2,843.28 | 2,716.49 | 2,525.86 | 2,228.58 | 10,314.22 |
| OHIO | 4,990.46 | 4,812.56 | 4,764.55 | 4,648.40 | 19,215.98 |
| OKLAHOMA | 6,568.03 | 6,343.59 | 6,351.01 | 6,276.46 | 25,539.09 |
| OREGON | 5,651.48 | 5,387.65 | 5,280.17 | 5,006.79 | 21,326.08 |
| PENNSYLVANIA | 7,109.26 | 6,949.22 | 6,713.80 | 6,435.83 | 27,208.11 |
| PUERTO RICO | 472.02 | 487.87 | 498.62 | 502.41 | 1,960.92 |

DRUG CODE:9143 DRUG NAME: OXYCODONE

| STATE | 1ST QUARTER | 2ND QUARTER | 3RD QUARTER | 4TH QUARTER | TOTAL GRAMS |
|---|---|---|---|---|---|
| RHODE ISLAND | 4,902.89 | 4,692.89 | 4,586.74 | 4,451.84 | 18,634.36 |
| SOUTH CAROLINA | 5,709.26 | 5,543.34 | 5,558.44 | 5,484.60 | 22,295.64 |
| SOUTH DAKOTA | 2,874.90 | 2,583.62 | 2,584.06 | 2,380.32 | 10,422.90 |
| TENNESSEE | 8,472.79 | 8,174.11 | 7,928.63 | 7,782.66 | 32,358.18 |
| TEXAS | 1,334.69 | 1,330.15 | 1,332.65 | 1,331.77 | 5,329.25 |
| UTAH | 7,035.97 | 6,748.40 | 6,778.74 | 6,520.98 | 27,084.09 |
| VERMONT | 4,330.56 | 4,193.84 | 4,135.68 | 3,947.73 | 16,607.81 |
| VIRGIN ISLANDS | 675.9 | 616.51 | 535.08 | 553.41 | 2,380.90 |
| VIRGINIA | 4,521.51 | 4,386.58 | 4,307.91 | 4,211.76 | 17,427.77 |
| WASHINGTON | 5,048.31 | 4,882.63 | 4,836.33 | 4,691.96 | 19,459.23 |
| WEST VIRGINIA | 6,318.91 | 5,858.15 | 5,635.13 | 5,288.11 | 23,100.30 |
| WISCONSIN | 5,024.87 | 4,716.37 | 4,614.89 | 4,405.87 | 18,762 |
| WYOMING | 5,423.97 | 5,164 | 5,148.50 | 4,995.67 | 20,732.13 |
| U.S. TOTAL | 4,547.51 | 4,398.89 | 4,327.82 | 4,237.63 | 17,511.85 |

DRUG CODE:9150 DRUG NAME: HYDROMORPHONE

| STATE | 1ST QUARTER | 2ND QUARTER | 3RD QUARTER | 4TH QUARTER | TOTAL GRAMS |
|---|---|---|---|---|---|
| ALABAMA | 83.69 | 84.67 | 80.32 | 75.77 | 324.45 |
| ALASKA | 157.77 | 149.64 | 146.18 | 152.81 | 606.39 |
| AMERICAN SAMOA | 0.93 | 2.97 | 0.1 | 0.81 | 4.81 |
| ARIZONA | 129.68 | 124.6 | 121.62 | 120.17 | 496.07 |
| ARKANSAS | 137.27 | 138.13 | 131.1 | 120.18 | 526.68 |
| CALIFORNIA | 126.51 | 120.89 | 114.45 | 114.61 | 476.46 |
| COLORADO | 162.54 | 157.58 | 156.18 | 149.16 | 625.46 |
| CONNECTICUT | 209.19 | 202.57 | 190.91 | 200.29 | 802.97 |
| DELAWARE | 273.77 | 274.28 | 274.23 | 257.37 | 1,079.66 |
| DISTRICT OF COLUMBIA | 107.37 | 98.64 | 103.25 | 103.55 | 412.81 |
| FLORIDA | 297.46 | 281.57 | 267.84 | 271.36 | 1,118.24 |
| GEORGIA | 92.47 | 92.96 | 91.44 | 92.29 | 369.16 |
| GUAM | 10.17 | 20.93 | 20.18 | 19.41 | 70.69 |
| HAWAII | 62.15 | 56.74 | 56.65 | 53.47 | 229.01 |
| IDAHO | 135.97 | 139.09 | 120.93 | 130.06 | 526.05 |
| ILLINOIS | 85.36 | 86.1 | 84.71 | 82.34 | 338.5 |
| INDIANA | 118.1 | 110.75 | 107.37 | 101.89 | 438.11 |
| IOWA | 122.12 | 114.92 | 114.08 | 110.36 | 461.49 |
| KANSAS | 137.09 | 142.39 | 138.27 | 134.87 | 552.62 |

| | | | | | |
|---|---|---|---|---|---|
| KENTUCKY | 69.95 | 74.67 | 71.83 | 74.07 | 290.53 |
| LOUISIANA | 90.03 | 86.89 | 80.43 | 79.18 | 336.54 |
| MAINE | 158.19 | 145.12 | 127.96 | 148.69 | 579.96 |
| MARYLAND | 175.7 | 173.65 | 168.7 | 159.6 | 677.64 |
| MASSACHUSETTS | 129.59 | 120.69 | 120.56 | 119.16 | 490 |
| MICHIGAN | 113.5 | 113.23 | 100.49 | 103.67 | 430.89 |
| MINNESOTA | 119.45 | 116.78 | 114.08 | 123.33 | 473.65 |
| MISSISSIPPI | 177.29 | 177.19 | 169.41 | 174.93 | 698.82 |
| MISSOURI | 133.03 | 127.65 | 124.69 | 122.68 | 508.06 |
| MONTANA | 180.55 | 180.56 | 183.55 | 185.69 | 730.36 |
| NEBRASKA | 82.91 | 79.64 | 81.46 | 76.49 | 320.5 |
| NEVADA | 146.17 | 131.05 | 124.6 | 129.1 | 530.91 |
| NEW HAMPSHIRE | 162.08 | 156.68 | 152.78 | 165.42 | 636.96 |
| NEW JERSEY | 121.67 | 121.98 | 117.65 | 113.28 | 474.59 |
| NEW MEXICO | 97.25 | 91.42 | 86.47 | 79.81 | 354.96 |
| NEW YORK | 121.33 | 116.54 | 114.25 | 115.65 | 467.77 |
| NORTH CAROLINA | 130.07 | 132.98 | 128.36 | 128.99 | 520.39 |
| NORTH DAKOTA | 125.84 | 122.9 | 119.39 | 106.88 | 475.02 |
| OHIO | 91.22 | 92.12 | 86.08 | 87.58 | 357.01 |
| OKLAHOMA | 116.39 | 111.58 | 103.29 | 107.08 | 438.33 |
| OREGON | 152.71 | 148.86 | 142.8 | 138.03 | 582.4 |
| PENNSYLVANIA | 136.62 | 136.77 | 133.06 | 133.02 | 539.46 |
| PUERTO RICO | 1.78 | 2 | 2.17 | 1.74 | 7.7 |

DRUG CODE:9150 DRUG NAME: HYDROMORPHONE

| STATE | 1ST QUARTER | 2ND QUARTER | 3RD QUARTER | 4TH QUARTER | TOTAL GRAMS |
|---|---|---|---|---|---|
| RHODE ISLAND | 80.35 | 83.73 | 84.48 | 87.8 | 336.35 |
| SOUTH CAROLINA | 136.51 | 132.43 | 131.2 | 130.72 | 530.86 |
| SOUTH DAKOTA | 101.67 | 84.32 | 77.54 | 72.23 | 335.76 |
| TENNESSEE | 121.19 | 107.52 | 113.59 | 114.44 | 456.73 |
| TEXAS | 105.66 | 102.03 | 105.19 | 103.83 | 416.72 |
| UTAH | 109.69 | 105.95 | 103.51 | 113.71 | 432.85 |
| VERMONT | 222.58 | 211.5 | 220.12 | 212.99 | 867.19 |
| VIRGIN ISLANDS | 27.29 | 28.42 | 24.83 | 12.29 | 92.83 |
| VIRGINIA | 178.51 | 168.89 | 169.69 | 164.92 | 682.01 |
| WASHINGTON | 189.16 | 191.27 | 181.08 | 167.48 | 728.98 |
| WEST VIRGINIA | 97.13 | 109 | 92.21 | 95.04 | 393.38 |
| WISCONSIN | 76.91 | 75.36 | 70.2 | 73.83 | 296.31 |
| WYOMING | 130.35 | 130.3 | 129.77 | 127.2 | 517.62 |
| U.S. TOTAL | 132.24 | 128.44 | 124.22 | 123.81 | 508.71 |

DRUG CODE:9193 DRUG NAME: HYDROCODONE

| STATE | 1ST QUARTER | 2ND QUARTER | 3RD QUARTER | 4TH QUARTER | TOTAL GRAMS |
|---|---|---|---|---|---|
| ALABAMA | 5,961.41 | 5,731.44 | 5,722.14 | 5,608.25 | 23,023.24 |
| ALASKA | 1,968.39 | 1,877.49 | 1,839.69 | 1,753.10 | 7,438.68 |
| AMERICAN SAMOA | 11.07 | 22.14 | 11.07 | 5.53 | 49.81 |
| ARIZONA | 1,966.07 | 1,801.42 | 1,769.73 | 1,788.06 | 7,325.28 |
| ARKANSAS | 5,050.24 | 4,885.73 | 4,849.93 | 4,856.30 | 19,642.21 |
| CALIFORNIA | 2,855.04 | 2,737.25 | 2,684.20 | 2,645.12 | 10,921.61 |
| COLORADO | 1,519.25 | 1,418.65 | 1,420.79 | 1,398.76 | 5,757.45 |
| CONNECTICUT | 734.23 | 678.28 | 652.6 | 639.46 | 2,704.56 |

| | | | | | |
|---|---|---|---|---|---|
| DELAWARE | 814.96 | 766.7 | 772.3 | 752.1 | 3,106.07 |
| DISTRICT OF COLUMBIA | 295.57 | 304.32 | 280.24 | 262.55 | 1,142.69 |
| FLORIDA | 2,192.96 | 2,048.80 | 2,017.87 | 2,029.62 | 8,289.25 |
| GEORGIA | 2,898.08 | 2,769.62 | 2,752.89 | 2,738 | 11,158.58 |
| GUAM | 121 | 116.18 | 125.3 | 97.15 | 459.64 |
| HAWAII | 1,516.89 | 1,439.14 | 1,389.14 | 1,370.53 | 5,715.71 |
| IDAHO | 4,048.56 | 3,873.91 | 3,871.19 | 3,826.36 | 15,620.02 |
| ILLINOIS | 2,736.05 | 2,635.28 | 2,600.39 | 2,618.49 | 10,590.21 |
| INDIANA | 4,053.37 | 3,911.66 | 3,780.78 | 3,821.82 | 15,567.63 |
| IOWA | 2,228.65 | 2,154.43 | 2,102.02 | 2,068.94 | 8,554.04 |
| KANSAS | 3,224.91 | 3,088.33 | 3,079.40 | 3,044.41 | 12,437.05 |
| KENTUCKY | 5,189.05 | 5,078.61 | 5,011.85 | 4,877.41 | 20,156.93 |
| LOUISIANA | 4,130.81 | 4,001.09 | 4,016.25 | 3,963.61 | 16,111.76 |
| MAINE | 1,715.57 | 1,593 | 1,515.98 | 1,456.37 | 6,280.93 |
| MARYLAND | 733.22 | 693.58 | 683.74 | 673.08 | 2,783.62 |
| MASSACHUSETTS | 579.83 | 545.27 | 522.78 | 500.98 | 2,148.86 |
| MICHIGAN | 4,779.42 | 4,582.45 | 4,192.98 | 4,456.13 | 18,010.98 |
| MINNESOTA | 1,058.75 | 1,010.71 | 986.48 | 948.65 | 4,004.58 |
| MISSISSIPPI | 4,656.38 | 4,486.77 | 4,454.76 | 4,438.49 | 18,036.40 |
| MISSOURI | 3,395.29 | 3,276.48 | 3,246.83 | 3,185.49 | 13,104.10 |
| MONTANA | 2,968.61 | 2,892.08 | 2,905.48 | 2,822.06 | 11,588.23 |
| NEBRASKA | 2,072.65 | 2,001.77 | 1,986.05 | 1,951.16 | 8,011.62 |
| NEVADA | 4,129.03 | 3,860.23 | 3,794.63 | 3,765.50 | 15,549.39 |
| NEW HAMPSHIRE | 830.95 | 777.41 | 770.68 | 763.53 | 3,142.56 |
| NEW JERSEY | 503.74 | 470.56 | 458.11 | 462.37 | 1,894.77 |
| NEW MEXICO | 2,110.29 | 1,938.09 | 1,928.39 | 1,886.78 | 7,863.54 |
| NEW YORK | 1,093.93 | 1,031.40 | 990.3 | 977.17 | 4,092.80 |
| NORTH CAROLINA | 2,416.09 | 2,272.55 | 2,228.84 | 2,236.32 | 9,153.80 |
| NORTH DAKOTA | 1,993.34 | 1,889.52 | 1,752.23 | 1,599.49 | 7,234.58 |
| OHIO | 1,964.89 | 1,913.38 | 1,840.58 | 1,802.82 | 7,521.68 |
| OKLAHOMA | 5,401.44 | 5,159.04 | 5,174.09 | 5,105.69 | 20,840.27 |
| OREGON | 2,961.52 | 2,862.35 | 2,771.57 | 2,673.09 | 11,268.52 |
| PENNSYLVANIA | 1,633.33 | 1,567.19 | 1,516.91 | 1,461.60 | 6,179.03 |
| PUERTO RICO | 35.26 | 28.94 | 26.6 | 27.6 | 118.39 |

DRUG CODE:9193 DRUG NAME: HYDROCODONE

| STATE | 1ST QUARTER | 2ND QUARTER | 3RD QUARTER | 4TH QUARTER | TOTAL GRAMS |
|---|---|---|---|---|---|
| RHODE ISLAND | 1,254.27 | 1,195.65 | 1,180.75 | 1,146.43 | 4,777.10 |
| SOUTH CAROLINA | 3,516.97 | 3,307.33 | 3,267.29 | 3,262.74 | 13,354.33 |
| SOUTH DAKOTA | 2,272.05 | 2,160.37 | 2,099.21 | 2,029.70 | 8,561.33 |
| TENNESSEE | 4,530.11 | 4,328.71 | 4,266.12 | 4,158.13 | 17,283.06 |
| TEXAS | 2,968.44 | 2,852.81 | 2,846.78 | 2,803.87 | 11,471.89 |
| UTAH | 2,472.82 | 2,360.23 | 2,340.32 | 2,270.70 | 9,444.07 |
| VERMONT | 979.76 | 948.5 | 907 | 894.55 | 3,729.82 |
| VIRGIN ISLANDS | 397.02 | 331.07 | 285.06 | 250.79 | 1,263.93 |
| VIRGINIA | 1,664.61 | 1,570.07 | 1,526.93 | 1,516.50 | 6,278.11 |
| WASHINGTON | 2,279.31 | 2,216.88 | 2,169.15 | 2,106.45 | 8,771.79 |
| WEST VIRGINIA | 4,124.11 | 3,904.77 | 3,754.43 | 3,616.96 | 15,400.27 |
| WISCONSIN | 1,675.08 | 1,602.10 | 1,575.50 | 1,536.88 | 6,389.56 |
| WYOMING | 2,516.16 | 2,397.46 | 2,338.21 | 2,302.01 | 9,553.85 |
| U.S. TOTAL | 2,539.52 | 2,428.11 | 2,380.33 | 2,359.93 | 9,707.89 |

DRUG CODE:9220L DRUG NAME: LEVORPHANOL

| STATE | 1ST QUARTER | 2ND QUARTER | 3RD QUARTER | 4TH QUARTER | TOTAL GRAMS |
|---|---|---|---|---|---|
| ALABAMA | 0.14 | 0.09 | 0.1 | 0.09 | 0.42 |
| ARIZONA | 0.03 | 0.08 | 0.16 | 0.36 | 0.62 |
| ARKANSAS | 0.02 | 0.07 | 0.1 | 0.08 | 0.27 |
| CALIFORNIA | 0.22 | 0.26 | 0.33 | 0.26 | 1.06 |
| COLORADO | 0.04 | 0.12 | 0.1 | 0.12 | 0.39 |
| CONNECTICUT | 0.01 | 0.06 | 0.05 | 0.03 | 0.15 |
| FLORIDA | 0.04 | 0.12 | 0.14 | 0.12 | 0.42 |
| GEORGIA | 0.01 | 0.01 | 0.01 | 0.01 | 0.05 |
| ILLINOIS | 0.06 | 0.05 | 0.05 | 0.07 | 0.23 |
| INDIANA | 0.4 | 0.38 | 0.42 | 0.37 | 1.56 |
| IOWA | 0.19 | 0.16 | 0.14 | 0.14 | 0.62 |
| KANSAS | 0.23 | 0.27 | 0.31 | 0.34 | 1.15 |
| LOUISIANA | 0.1 | 0.19 | 0.29 | 0.32 | 0.91 |
| MAINE | 0 | 0 | 0 | 0.02 | 0.02 |
| MARYLAND | 0 | 0.01 | 0.01 | 0.05 | 0.07 |
| MASSACHUSETTS | 0.33 | 0.29 | 0.24 | 0.29 | 1.15 |
| MICHIGAN | 0 | 0.01 | 0.01 | 0.01 | 0.04 |
| MINNESOTA | 0.01 | 0 | 0.03 | 0.01 | 0.05 |
| MISSISSIPPI | 0.04 | 0.05 | 0.01 | 0.01 | 0.1 |
| MISSOURI | 0.08 | 0.09 | 0.15 | 0.19 | 0.52 |
| NEBRASKA | 0.02 | 0.02 | 0.06 | 0.1 | 0.2 |
| NEVADA | 0.02 | 0.13 | 0.08 | 0.09 | 0.31 |
| NEW HAMPSHIRE | 0.1 | 0.07 | 0.03 | 0.06 | 0.26 |
| NEW JERSEY | 0.5 | 1.11 | 2.11 | 3.05 | 6.76 |
| NEW MEXICO | 0 | 0 | 0 | 0.01 | 0.01 |
| NEW YORK | 0.18 | 0.12 | 0.14 | 0.26 | 0.7 |
| NORTH CAROLINA | 0.21 | 0.18 | 0.37 | 0.42 | 1.18 |
| OHIO | 0.01 | 0.02 | 0.07 | 0.11 | 0.21 |
| OKLAHOMA | 0.01 | 0.02 | 0.01 | 0.01 | 0.05 |
| OREGON | 0.17 | 0.12 | 0.11 | 0.1 | 0.5 |
| PENNSYLVANIA | 0.08 | 0.08 | 0.07 | 0.08 | 0.31 |
| SOUTH CAROLINA | 0.02 | 0 | 0.01 | 0.02 | 0.04 |
| TENNESSEE | 0.01 | 0.01 | 0 | 0 | 0.01 |
| TEXAS | 0.09 | 0.09 | 0.15 | 0.17 | 0.5 |
| UTAH | 0.01 | 0 | 0 | 0 | 0.01 |
| VERMONT | 0.09 | 0.06 | 0.07 | 0.09 | 0.32 |
| VIRGINIA | 0.09 | 0.06 | 0.07 | 0.07 | 0.29 |
| WASHINGTON | 0.2 | 0.19 | 0.19 | 0.23 | 0.81 |
| WEST VIRGINIA | 0.01 | 0.02 | 0.02 | 0.01 | 0.06 |
| WISCONSIN | 0.06 | 0.06 | 0.05 | 0.06 | 0.23 |
| U.S. TOTAL | 0.11 | 0.14 | 0.19 | 0.23 | 0.67 |

DRUG CODE:9230 DRUG NAME: MEPERIDINE (PETHIDINE)

| STATE | 1ST QUARTER | 2ND QUARTER | 3RD QUARTER | 4TH QUARTER | TOTAL GRAMS |
|---|---|---|---|---|---|
| ALABAMA | 206.36 | 198.58 | 185.24 | 374.78 | 964.96 |
| ALASKA | 112.02 | 128.63 | 109.7 | 115.65 | 465.99 |
| AMERICAN SAMOA | 9.3 | 0 | 0 | 0 | 9.3 |
| ARIZONA | 98.61 | 101.97 | 97.71 | 89.21 | 387.5 |
| ARKANSAS | 341.02 | 326.22 | 331.38 | 311.31 | 1,309.93 |
| CALIFORNIA | 47.97 | 50.17 | 44.88 | 46.95 | 189.98 |
| COLORADO | 50.54 | 47.47 | 47.93 | 44.9 | 190.83 |

| | | | | | |
|---|---|---|---|---|---|
| CONNECTICUT | 40.89 | 35.78 | 35.83 | 37.37 | 149.88 |
| DELAWARE | 26.29 | 26.89 | 25.96 | 24.89 | 104.04 |
| DISTRICT OF COLUMBIA | 30.52 | 26.56 | 34.62 | 40.77 | 132.47 |
| FLORIDA | 57.81 | 57.51 | 53.86 | 54.54 | 223.72 |
| GEORGIA | 136.64 | 127.77 | 124.56 | 126.46 | 515.42 |
| GUAM | 19.23 | 39.44 | 5.37 | 24.42 | 88.47 |
| HAWAII | 36.43 | 36.53 | 28.61 | 28.28 | 129.86 |
| IDAHO | 61.75 | 58.8 | 63.91 | 53.29 | 237.75 |
| ILLINOIS | 30.77 | 29.74 | 28.18 | 26.44 | 115.13 |
| INDIANA | 64.06 | 54.87 | 54.03 | 58.8 | 231.77 |
| IOWA | 61.86 | 67.51 | 62.11 | 65.54 | 257.02 |
| KANSAS | 67.21 | 72.72 | 70.74 | 69.32 | 280 |
| KENTUCKY | 70.23 | 68.56 | 66.98 | 64.05 | 269.82 |
| LOUISIANA | 229.54 | 203.78 | 200.73 | 189.28 | 823.33 |
| MAINE | 44.74 | 36.21 | 34.26 | 36.02 | 151.22 |
| MARYLAND | 50.64 | 52.93 | 49.5 | 47.31 | 200.39 |
| MASSACHUSETTS | 41.9 | 44.04 | 40.34 | 39.67 | 165.95 |
| MICHIGAN | 39.09 | 37.99 | 35.25 | 34.5 | 146.83 |
| MINNESOTA | 15.83 | 15.91 | 14.85 | 16.17 | 62.75 |
| MISSISSIPPI | 198.31 | 203.16 | 186.5 | 177.96 | 765.93 |
| MISSOURI | 87.77 | 81.61 | 82.9 | 73.34 | 325.62 |
| MONTANA | 71.37 | 93.68 | 83.79 | 81.56 | 330.4 |
| NEBRASKA | 44.36 | 58.87 | 41.57 | 44.73 | 189.53 |
| NEVADA | 68.78 | 67.87 | 66.33 | 58 | 260.98 |
| NEW HAMPSHIRE | 56.32 | 57.45 | 55.47 | 51.11 | 220.35 |
| NEW JERSEY | 29.09 | 26.88 | 25.84 | 26.82 | 108.63 |
| NEW MEXICO | 94.48 | 95.59 | 93.8 | 79.41 | 363.29 |
| NEW YORK | 30.31 | 29.38 | 29.09 | 26.2 | 114.98 |
| NORTH CAROLINA | 82.71 | 81.18 | 77.37 | 68.16 | 309.42 |
| NORTH DAKOTA | 44.95 | 47.54 | 37.35 | 38.81 | 168.64 |
| OHIO | 49.13 | 49.68 | 51.82 | 44.92 | 195.54 |
| OKLAHOMA | 248.58 | 242.38 | 238.65 | 224.74 | 954.36 |
| OREGON | 47.32 | 48.19 | 47.69 | 46.37 | 189.56 |
| PENNSYLVANIA | 37.05 | 32.33 | 37.13 | 33.29 | 139.79 |
| PUERTO RICO | 176.1 | 179.66 | 174.39 | 147.32 | 677.48 |

DRUG CODE:9230 DRUG NAME: MEPERIDINE (PETHIDINE)

| STATE | 1ST QUARTER | 2ND QUARTER | 3RD QUARTER | 4TH QUARTER | TOTAL GRAMS |
|---|---|---|---|---|---|
| RHODE ISLAND | 22.45 | 20.97 | 23.92 | 23 | 90.33 |
| SOUTH CAROLINA | 186.92 | 166.94 | 157.62 | 147.72 | 659.2 |
| SOUTH DAKOTA | 87.28 | 82.92 | 77.59 | 66.3 | 314.08 |
| TENNESSEE | 166.09 | 145.22 | 147.4 | 134.2 | 592.92 |
| TEXAS | 58.7 | 58.64 | 56.19 | 51.61 | 225.14 |
| UTAH | 86.03 | 80.09 | 83.93 | 79.29 | 329.35 |
| VERMONT | 61.61 | 58.97 | 55.83 | 52.38 | 228.79 |
| VIRGIN ISLANDS | 37.69 | 72.31 | 70.43 | 35.71 | 216.14 |
| VIRGINIA | 97.4 | 96.87 | 92.83 | 86.26 | 373.36 |
| WASHINGTON | 67 | 68.99 | 57.58 | 58.17 | 251.75 |
| WEST VIRGINIA | 42.27 | 47.33 | 39.55 | 45.12 | 174.28 |
| WISCONSIN | 34.1 | 35.94 | 33.81 | 31.02 | 134.87 |
| WYOMING | 100.25 | 84.18 | 85.07 | 85.51 | 355 |
| U.S. TOTAL | 72.36 | 70.56 | 67.82 | 67.61 | 278.35 |

ARCOS 3 - REPORT 3

QUARTERLY DRUG DISTRIBUTION BY STATE PER 100K POPULATION BY GRAM WT

DRUG CODE:9250B DRUG NAME: METHADONE

| STATE | 1ST QUARTER | 2ND QUARTER | 3RD QUARTER | 4TH QUARTER | TOTAL GRAMS |
|---|---|---|---|---|---|
| ALABAMA | 1,886.10 | 1,837.76 | 1,707.17 | 1,599.19 | 7,030.22 |
| ALASKA | 1,208.21 | 1,203.43 | 1,049.07 | 1,073.85 | 4,534.56 |
| ARIZONA | 1,360.81 | 1,370.97 | 1,345.85 | 1,144.12 | 5,221.75 |
| ARKANSAS | 703.48 | 708.32 | 663.01 | 585.97 | 2,660.78 |
| CALIFORNIA | 1,015.85 | 1,001.95 | 981.52 | 761.58 | 3,760.90 |
| COLORADO | 763.71 | 837.47 | 818.67 | 733.6 | 3,153.46 |
| CONNECTICUT | 3,176.77 | 3,121.58 | 2,740.01 | 2,861.88 | 11,900.24 |
| DELAWARE | 3,006.48 | 3,089.84 | 3,059.20 | 2,112.90 | 11,268.43 |
| DISTRICT OF COLUMBIA | 1,802.79 | 1,914.77 | 1,815.60 | 1,188.58 | 6,721.74 |
| FLORIDA | 1,143.38 | 1,173.51 | 1,131.13 | 1,054.05 | 4,502.07 |
| GEORGIA | 1,201.39 | 1,203.21 | 1,226.60 | 1,139.45 | 4,770.66 |
| GUAM | 29.07 | 22.06 | 26.57 | 14.54 | 92.23 |
| HAWAII | 697.76 | 586.04 | 678.69 | 507.93 | 2,470.42 |
| IDAHO | 879.67 | 878.17 | 831.68 | 670.05 | 3,259.57 |
| ILLINOIS | 978.43 | 973.95 | 956.8 | 806.17 | 3,715.35 |
| INDIANA | 1,421.25 | 1,415.48 | 1,376.39 | 1,090.05 | 5,303.18 |
| IOWA | 454.67 | 492.57 | 434.79 | 372.58 | 1,754.62 |
| KANSAS | 865.7 | 883.78 | 826.14 | 782.85 | 3,358.48 |
| KENTUCKY | 1,191.08 | 1,188.01 | 1,223.29 | 1,172.83 | 4,775.21 |
| LOUISIANA | 820.02 | 893.56 | 831.45 | 835.22 | 3,380.25 |
| MAINE | 2,924.66 | 2,928.77 | 2,791.52 | 2,000.96 | 10,645.91 |
| MARYLAND | 3,250.19 | 3,265.51 | 3,142.53 | 2,671.61 | 12,329.84 |
| MASSACHUSETTS | 2,170.16 | 2,263.25 | 2,209.81 | 1,624.57 | 8,267.79 |
| MICHIGAN | 1,300.80 | 1,405.34 | 1,297.78 | 1,186 | 5,189.92 |
| MINNESOTA | 1,106.78 | 1,089.81 | 992.67 | 977.68 | 4,166.94 |
| MISSISSIPPI | 240.12 | 238.72 | 222.63 | 217.68 | 919.14 |
| MISSOURI | 712.13 | 739.92 | 704.79 | 625.87 | 2,782.70 |
| MONTANA | 813 | 733.22 | 754.22 | 703.08 | 3,003.52 |
| NEBRASKA | 499.16 | 473.01 | 484.35 | 460.08 | 1,916.59 |
| NEVADA | 1,496.10 | 1,450.35 | 1,353.21 | 1,408.12 | 5,707.77 |
| NEW HAMPSHIRE | 2,352.42 | 2,530.72 | 2,383.72 | 1,894.93 | 9,161.79 |
| NEW JERSEY | 1,265.41 | 1,299.34 | 1,239.82 | 1,156.64 | 4,961.22 |
| NEW MEXICO | 1,899.43 | 1,968.79 | 2,022.25 | 1,540.63 | 7,431.09 |
| NEW YORK | 1,499.78 | 1,602.60 | 1,566.12 | 1,288.04 | 5,956.54 |
| NORTH CAROLINA | 1,426.96 | 1,413.85 | 1,403.38 | 1,264.46 | 5,508.64 |
| NORTH DAKOTA | 211.97 | 185.35 | 215.84 | 147.63 | 760.78 |
| OHIO | 673.78 | 674.06 | 678.22 | 669.42 | 2,695.48 |
| OKLAHOMA | 1,168.29 | 1,110.38 | 1,021.99 | 782.15 | 4,082.82 |
| OREGON | 1,805.91 | 1,808.63 | 1,766.53 | 1,455.38 | 6,836.44 |
| PENNSYLVANIA | 1,585.58 | 1,564.74 | 1,509.20 | 1,358.49 | 6,018.01 |
| PUERTO RICO | 476.33 | 573.9 | 522.13 | 432.09 | 2,004.46 |
| RHODE ISLAND | 4,234.75 | 3,847.08 | 4,158.76 | 3,891.59 | 16,132.18 |

DRUG CODE:9250B DRUG NAME: METHADONE

| STATE | 1ST QUARTER | 2ND QUARTER | 3RD QUARTER | 4TH QUARTER | TOTAL GRAMS |
|---|---|---|---|---|---|
| SOUTH CAROLINA | 1,312.85 | 1,241.07 | 1,217.88 | 1,031 | 4,802.80 |
| SOUTH DAKOTA | 314.15 | 321.89 | 290.03 | 280.39 | 1,206.46 |
| TENNESSEE | 833.51 | 852.6 | 805.08 | 726.69 | 3,217.88 |
| TEXAS | 574.65 | 588.17 | 564.38 | 493.96 | 2,221.16 |
| UTAH | 1,006.50 | 1,164.22 | 966.93 | 863.12 | 4,000.77 |
| VERMONT | 2,713.27 | 3,084.20 | 2,701.52 | 2,396.96 | 10,895.94 |

| | | | | | |
|---|---|---|---|---|---|
| VIRGIN ISLANDS | 32.98 | 40.31 | 43.16 | 33.8 | 150.25 |
| VIRGINIA | 937.04 | 941.96 | 939.41 | 826.12 | 3,644.53 |
| WASHINGTON | 1,615.32 | 1,613.92 | 1,585.08 | 1,103.69 | 5,918.02 |
| WEST VIRGINIA | 1,483.12 | 1,496.63 | 1,382.80 | 1,063.21 | 5,425.76 |
| WISCONSIN | 1,231.36 | 1,208.83 | 1,201.42 | 855.01 | 4,496.63 |
| WYOMING | 277.18 | 260.06 | 251.93 | 237.84 | 1,027.01 |
| U.S. TOTAL | 1,211.42 | 1,226.62 | 1,185.73 | 1,019.78 | 4,643.55 |

DRUG CODE:9300 DRUG NAME: MORPHINE

| STATE | 1ST QUARTER | 2ND QUARTER | 3RD QUARTER | 4TH QUARTER | TOTAL GRAMS |
|---|---|---|---|---|---|
| ALABAMA | 1,831.90 | 1,793.45 | 1,761.44 | 1,702.93 | 7,089.72 |
| ALASKA | 2,052.25 | 1,917 | 1,824.24 | 1,726.35 | 7,519.84 |
| AMERICAN SAMOA | 46.41 | 63.02 | 95.68 | 21.77 | 226.88 |
| ARIZONA | 3,328.06 | 3,144.18 | 3,083.44 | 2,966.92 | 12,522.60 |
| ARKANSAS | 1,827.58 | 1,764 | 1,790.84 | 1,695.96 | 7,078.36 |
| CALIFORNIA | 1,467.92 | 1,411.41 | 1,363.34 | 1,324.63 | 5,567.31 |
| COLORADO | 1,897.74 | 1,814.75 | 1,747.17 | 1,666.07 | 7,125.74 |
| CONNECTICUT | 1,207.64 | 1,156.18 | 1,127.18 | 1,110.02 | 4,601.02 |
| DELAWARE | 2,591.74 | 2,582.58 | 2,499.63 | 2,504.93 | 10,178.88 |
| DISTRICT OF COLUMBIA | 798.84 | 668.34 | 769.58 | 673.27 | 2,910.03 |
| FLORIDA | 2,407.06 | 2,326.32 | 2,277.47 | 2,236.66 | 9,247.51 |
| GEORGIA | 1,277.94 | 1,238.51 | 1,242.75 | 1,232.84 | 4,992.04 |
| GUAM | 562.33 | 723.35 | 715.35 | 686.79 | 2,687.82 |
| HAWAII | 1,988.03 | 1,922.09 | 1,879.52 | 1,822.07 | 7,611.70 |
| IDAHO | 2,066.52 | 2,044.10 | 1,953.08 | 1,916.28 | 7,979.98 |
| ILLINOIS | 889.5 | 882.87 | 882.39 | 858.51 | 3,513.27 |
| INDIANA | 1,697.15 | 1,597.58 | 1,549.16 | 1,482.45 | 6,326.34 |
| IOWA | 1,321.76 | 1,271.73 | 1,248.41 | 1,188.55 | 5,030.45 |
| KANSAS | 1,810.95 | 1,783.10 | 1,815.04 | 1,651.85 | 7,060.94 |
| KENTUCKY | 1,281.54 | 1,229.60 | 1,231.76 | 1,164.87 | 4,907.77 |
| LOUISIANA | 1,455.86 | 1,421.40 | 1,382.46 | 1,293.28 | 5,553 |
| MAINE | 1,802.73 | 1,791.34 | 1,639.73 | 1,464.59 | 6,698.39 |
| MARYLAND | 1,578.40 | 1,562.27 | 1,530.52 | 1,483.67 | 6,154.86 |
| MASSACHUSETTS | 1,203.48 | 1,154.41 | 1,111.85 | 1,094.94 | 4,564.68 |
| MICHIGAN | 2,335.95 | 2,259.66 | 2,233.70 | 2,143.94 | 8,973.25 |
| MINNESOTA | 1,038.13 | 1,028.53 | 1,012.67 | 966.84 | 4,046.18 |
| MISSISSIPPI | 1,472.69 | 1,462.22 | 1,459.43 | 1,455.35 | 5,849.68 |
| MISSOURI | 1,881.86 | 1,869.16 | 1,818.36 | 1,748.77 | 7,318.16 |
| MONTANA | 2,298.04 | 2,238.04 | 2,135.42 | 2,005.41 | 8,676.91 |
| NEBRASKA | 1,110.39 | 1,098.97 | 1,106.58 | 1,084.89 | 4,400.82 |
| NEVADA | 2,843.60 | 2,696.03 | 2,575.51 | 2,431.71 | 10,546.85 |
| NEW HAMPSHIRE | 1,582.10 | 1,525.73 | 1,507.61 | 1,439.94 | 6,055.39 |
| NEW JERSEY | 1,144.61 | 1,123.28 | 1,117.19 | 1,122.05 | 4,507.13 |
| NEW MEXICO | 1,763.21 | 1,661.75 | 1,568.65 | 1,491.01 | 6,484.62 |
| NEW YORK | 985.23 | 964.66 | 922.34 | 897.78 | 3,770.01 |
| NORTH CAROLINA | 1,517.03 | 1,464.37 | 1,441.17 | 1,371.49 | 5,794.06 |
| NORTH DAKOTA | 1,268.89 | 1,230.67 | 1,070.28 | 967.52 | 4,537.36 |
| OHIO | 1,215.93 | 1,195.51 | 1,182.31 | 1,119.53 | 4,713.27 |
| OKLAHOMA | 2,895.77 | 2,797.59 | 2,741.05 | 2,647.96 | 11,082.37 |
| OREGON | 2,538.87 | 2,434.27 | 2,301.02 | 2,133.98 | 9,408.14 |
| PENNSYLVANIA | 2,113.93 | 2,100.33 | 1,998.12 | 1,930.81 | 8,143.18 |
| PUERTO RICO | 77.2 | 86.43 | 78.53 | 77.76 | 319.92 |

DRUG CODE:9300 DRUG NAME: MORPHINE

| STATE | 1ST QUARTER | 2ND QUARTER | 3RD QUARTER | 4TH QUARTER | TOTAL GRAMS |
|---|---|---|---|---|---|
| RHODE ISLAND | 1,224.89 | 1,213.80 | 1,229.34 | 1,266.88 | 4,934.91 |
| SOUTH CAROLINA | 1,600.27 | 1,560.94 | 1,550.79 | 1,539.30 | 6,251.30 |
| SOUTH DAKOTA | 1,467.67 | 1,354.09 | 1,354.16 | 1,186.69 | 5,362.61 |
| TENNESSEE | 3,249.68 | 3,097.80 | 2,943.83 | 2,746.93 | 12,038.24 |
| TEXAS | 928.75 | 908.6 | 904.89 | 909.24 | 3,651.48 |
| UTAH | 2,103.17 | 2,039.66 | 2,022.27 | 1,923.54 | 8,088.64 |
| VERMONT | 1,897.12 | 1,837.61 | 1,739.16 | 1,744.40 | 7,218.29 |
| VIRGIN ISLANDS | 121.33 | 84.78 | 94.42 | 78.95 | 379.48 |
| VIRGINIA | 1,185.53 | 1,131.88 | 1,123.38 | 1,100.65 | 4,541.44 |
| WASHINGTON | 1,989.47 | 1,927.02 | 1,881.05 | 1,787.90 | 7,585.44 |
| WEST VIRGINIA | 1,570.97 | 1,546.61 | 1,471.51 | 1,395.29 | 5,984.39 |
| WISCONSIN | 1,801.51 | 1,741.19 | 1,681.14 | 1,549.38 | 6,773.22 |
| WYOMING | 1,979.25 | 1,816.75 | 1,853.14 | 1,655.94 | 7,305.09 |
| U.S. TOTAL | 1,602.27 | 1,552.94 | 1,515.49 | 1,462.68 | 6,133.38 |

DRUG CODE:9630 DRUG NAME: OPIUM TINCTURE (LAUDANUM)

| STATE | 1ST QUARTER | 2ND QUARTER | 3RD QUARTER | 4TH QUARTER | TOTAL GRAMS |
|---|---|---|---|---|---|
| ARIZONA | 0 | 0 | 0 | 0.15 | 0.15 |
| CALIFORNIA | 0.01 | 0.08 | 0.03 | 0 | 0.12 |
| COLORADO | 0 | 0 | 0.02 | 0 | 0.02 |
| DISTRICT OF COLUMBIA | 0 | 0 | 0.2 | 0 | 0.2 |
| FLORIDA | 0.08 | 0.03 | 0.03 | 0.08 | 0.2 |
| IDAHO | 0 | 0.08 | 0 | 0 | 0.08 |
| ILLINOIS | 0.08 | 0.05 | 0 | 0.14 | 0.27 |
| INDIANA | 0.24 | 0.55 | 0.22 | 0.26 | 1.26 |
| KANSAS | 0 | 0 | 0 | 0.21 | 0.21 |
| MAINE | 0 | 0.09 | 0.36 | 0 | 0.44 |
| MARYLAND | 0.04 | 0.12 | 0 | 0.02 | 0.18 |
| MASSACHUSETTS | 0.09 | 0.58 | 0.81 | 0.81 | 2.29 |
| MICHIGAN | 0 | 0.2 | 0.02 | 0.25 | 0.48 |
| MINNESOTA | 0 | 0 | 0.02 | 0 | 0.02 |
| MISSOURI | 0 | 0 | 0.08 | 0.36 | 0.43 |
| NEBRASKA | 0 | 0 | 0.78 | 0 | 0.78 |
| NEW HAMPSHIRE | 0 | 0.09 | 0 | 0 | 0.09 |
| NEW JERSEY | 0.03 | 0.03 | 0.01 | 0.04 | 0.11 |
| NEW MEXICO | 0 | 0 | 0 | 0.23 | 0.23 |
| NEW YORK | 0.01 | 0 | 0.01 | 0 | 0.02 |
| OHIO | 0.13 | 0.3 | 0.23 | 0.18 | 0.84 |
| OREGON | 0 | 0 | 0 | 0.12 | 0.12 |
| PENNSYLVANIA | 0.05 | 0.01 | 0 | 0.05 | 0.1 |
| TEXAS | 0.01 | 0 | 0.04 | 0.03 | 0.08 |
| VERMONT | 0 | 0 | 0.38 | 0 | 0.38 |
| VIRGINIA | 0 | 0.12 | 0.07 | 0.03 | 0.22 |
| WASHINGTON | 0.05 | 0.07 | 0.09 | 0.09 | 0.3 |
| WEST VIRGINIA | 0.32 | 0.26 | 0 | 0.51 | 1.08 |
| WISCONSIN | 0 | 0 | 0 | 0.02 | 0.02 |
| U.S. TOTAL | 0.03 | 0.06 | 0.05 | 0.07 | 0.21 |

DRUG CODE:9639 DRUG NAME: OPIUM POWDERED

| STATE | 1ST QUARTER | 2ND QUARTER | 3RD QUARTER | 4TH QUARTER | TOTAL GRAMS |
|---|---|---|---|---|---|
| ALABAMA | 1.89 | 1.88 | 2.03 | 2.14 | 7.93 |
| ALASKA | 2.03 | 2.99 | 2.38 | 3.55 | 10.95 |
| ARIZONA | 3.76 | 3.93 | 4.83 | 4.82 | 17.34 |
| ARKANSAS | 3.1 | 3.59 | 3.48 | 3.25 | 13.42 |
| CALIFORNIA | 1.26 | 1.36 | 1.29 | 1.22 | 5.13 |
| COLORADO | 5.1 | 4.26 | 4.32 | 4.29 | 17.97 |
| CONNECTICUT | 3 | 2.4 | 2.31 | 2.49 | 10.19 |
| DELAWARE | 2.73 | 2.69 | 1.96 | 1.32 | 8.7 |
| DISTRICT OF COLUMBIA | 2.03 | 1.79 | 1.02 | 2.87 | 7.72 |
| FLORIDA | 2.57 | 2.47 | 2.26 | 2.67 | 9.96 |
| GEORGIA | 1.96 | 1.94 | 2.13 | 2.02 | 8.05 |
| GUAM | 0.4 | 0.4 | 0.81 | 0 | 1.61 |
| HAWAII | 1.91 | 2.75 | 1.53 | 1.22 | 7.41 |
| IDAHO | 3.42 | 2.92 | 2.48 | 2.76 | 11.57 |
| ILLINOIS | 1.6 | 1.47 | 1.67 | 1.55 | 6.28 |
| INDIANA | 3.73 | 4.3 | 4.19 | 3.69 | 15.91 |
| IOWA | 2.67 | 3.52 | 3.45 | 3.83 | 13.47 |
| KANSAS | 6.04 | 6.62 | 7 | 6.61 | 26.28 |
| KENTUCKY | 3.82 | 3.93 | 3.99 | 4.5 | 16.24 |
| LOUISIANA | 2.47 | 2.72 | 2.05 | 2.22 | 9.47 |
| MAINE | 3.52 | 3.01 | 3.52 | 2.36 | 12.41 |
| MARYLAND | 1.04 | 1.1 | 1.87 | 1.12 | 5.14 |
| MASSACHUSETTS | 2.69 | 2.19 | 2.67 | 3.1 | 10.65 |
| MICHIGAN | 3.31 | 3.94 | 3.74 | 3.87 | 14.85 |
| MINNESOTA | 7.62 | 7.8 | 8.56 | 7.94 | 31.92 |
| MISSISSIPPI | 1.61 | 1.02 | 1.5 | 0.95 | 5.08 |
| MISSOURI | 2.66 | 3.38 | 3.91 | 3.43 | 13.37 |
| MONTANA | 5.13 | 5.49 | 5.6 | 5.68 | 21.9 |
| NEBRASKA | 5.03 | 6.43 | 5.64 | 5.56 | 22.65 |
| NEVADA | 1.4 | 1.44 | 1.15 | 0.8 | 4.79 |
| NEW HAMPSHIRE | 2.08 | 3.72 | 2.82 | 2.76 | 11.38 |
| NEW JERSEY | 1.42 | 1.74 | 1.63 | 1.65 | 6.44 |
| NEW MEXICO | 2.43 | 2.88 | 3.25 | 2.45 | 11.01 |
| NEW YORK | 0.94 | 1.21 | 1.37 | 1.11 | 4.63 |
| NORTH CAROLINA | 2.41 | 2.44 | 2.42 | 2.46 | 9.73 |
| NORTH DAKOTA | 8.19 | 4.71 | 5.62 | 4.39 | 22.91 |
| OHIO | 3.55 | 3.85 | 3.97 | 3.6 | 14.98 |
| OKLAHOMA | 1.84 | 1.71 | 2.26 | 2.42 | 8.22 |
| OREGON | 5.51 | 5.28 | 4.93 | 5.88 | 21.6 |
| PENNSYLVANIA | 2.94 | 3.13 | 2.95 | 2.56 | 11.58 |
| RHODE ISLAND | 4.41 | 2.22 | 3.11 | 2.84 | 12.59 |
| SOUTH CAROLINA | 2.94 | 3.28 | 3.18 | 3.24 | 12.64 |

DRUG CODE:9639 DRUG NAME: OPIUM POWDERED

| STATE | 1ST QUARTER | 2ND QUARTER | 3RD QUARTER | 4TH QUARTER | TOTAL GRAMS |
|---|---|---|---|---|---|
| SOUTH DAKOTA | 2.7 | 2.03 | 2.92 | 3.45 | 11.1 |
| TENNESSEE | 2.97 | 3.77 | 4.35 | 4.02 | 15.11 |
| TEXAS | 2.02 | 2.07 | 2.18 | 2.05 | 8.31 |
| UTAH | 1.93 | 2.54 | 2.18 | 2.31 | 8.95 |
| VERMONT | 8.97 | 5.75 | 4.83 | 4.89 | 24.45 |
| VIRGIN ISLANDS | 0 | 0.66 | 0.66 | 0.33 | 1.64 |

| | | | | | |
|---|---|---|---|---|---|
| VIRGINIA | 2.2 | 2.22 | 2.16 | 2.37 | 8.95 |
| WASHINGTON | 4.73 | 5.48 | 4.72 | 4.56 | 19.48 |
| WEST VIRGINIA | 4.92 | 5.77 | 6.28 | 5.42 | 22.38 |
| WISCONSIN | 3.53 | 3.27 | 3.74 | 3.27 | 13.8 |
| WYOMING | 6.2 | 5.17 | 5.05 | 6.96 | 23.38 |
| U.S. TOTAL | 2.56 | 2.68 | 2.73 | 2.65 | 10.62 |

DRUG CODE:9652 DRUG NAME: OXYMORPHONE

| STATE | 1ST QUARTER | 2ND QUARTER | 3RD QUARTER | 4TH QUARTER | TOTAL GRAMS |
|---|---|---|---|---|---|
| ALABAMA | 211.34 | 216.06 | 193.63 | 171.63 | 792.65 |
| ALASKA | 48.9 | 46.24 | 43.42 | 43.04 | 181.6 |
| ARIZONA | 153.92 | 151.51 | 144.2 | 148.45 | 598.08 |
| ARKANSAS | 90.74 | 87.83 | 86 | 81.22 | 345.79 |
| CALIFORNIA | 59.11 | 55.49 | 52.95 | 50.38 | 217.92 |
| COLORADO | 104.38 | 96.73 | 93.19 | 85.89 | 380.2 |
| CONNECTICUT | 118.04 | 115.02 | 107.04 | 103.86 | 443.96 |
| DELAWARE | 251.57 | 247.63 | 247.55 | 238.36 | 985.12 |
| DISTRICT OF COLUMBIA | 25.27 | 22.57 | 28.55 | 23.25 | 99.64 |
| FLORIDA | 155.94 | 154.77 | 151.79 | 151.43 | 613.92 |
| GEORGIA | 92.82 | 90.33 | 89.84 | 86.34 | 359.33 |
| HAWAII | 53.08 | 56.2 | 51.6 | 50.53 | 211.4 |
| IDAHO | 112.05 | 113 | 108.06 | 99.72 | 432.83 |
| ILLINOIS | 48.6 | 47.96 | 47.3 | 45.29 | 189.15 |
| INDIANA | 130.61 | 123.62 | 117.85 | 114.92 | 487.01 |
| IOWA | 61.71 | 53.42 | 52.41 | 49.4 | 216.94 |
| KANSAS | 114.88 | 112.7 | 104.01 | 97.04 | 428.64 |
| KENTUCKY | 123.92 | 122.34 | 114.77 | 109.53 | 470.56 |
| LOUISIANA | 210.32 | 202.97 | 205.55 | 190.92 | 809.77 |
| MAINE | 146.53 | 141.27 | 122.8 | 106.79 | 517.39 |
| MARYLAND | 174.82 | 179.89 | 189.81 | 190.89 | 735.41 |
| MASSACHUSETTS | 49.51 | 47.92 | 43 | 42.12 | 182.56 |
| MICHIGAN | 206.07 | 199.55 | 208.27 | 210.38 | 824.26 |
| MINNESOTA | 12.45 | 12.11 | 12.06 | 9.77 | 46.4 |
| MISSISSIPPI | 178.88 | 167.2 | 161.68 | 146.89 | 654.66 |
| MISSOURI | 109.98 | 103.6 | 95.14 | 97.23 | 405.94 |
| MONTANA | 52.48 | 54.89 | 49.49 | 42.24 | 199.11 |
| NEBRASKA | 86.77 | 91.03 | 91.27 | 86.86 | 355.92 |
| NEVADA | 250.17 | 240.27 | 224.9 | 215.7 | 931.03 |
| NEW HAMPSHIRE | 104.27 | 103.37 | 90 | 91.4 | 389.05 |
| NEW JERSEY | 115.19 | 114.88 | 108.74 | 101.2 | 440.01 |
| NEW MEXICO | 83.08 | 73.67 | 72 | 65.74 | 294.49 |
| NEW YORK | 139.94 | 131.17 | 126.1 | 123.3 | 520.51 |
| NORTH CAROLINA | 395.63 | 367.38 | 339.53 | 315.81 | 1,418.36 |
| NORTH DAKOTA | 31.87 | 33.9 | 32.73 | 24.8 | 123.31 |
| OHIO | 122.12 | 114.67 | 113.58 | 110.66 | 461.02 |
| OKLAHOMA | 213.77 | 203.75 | 194.65 | 188.09 | 800.26 |
| OREGON | 63.79 | 59.38 | 57.57 | 53.64 | 234.38 |
| PENNSYLVANIA | 232.52 | 224.35 | 224.48 | 222.37 | 903.73 |
| RHODE ISLAND | 61.97 | 55.01 | 55.32 | 53.24 | 225.53 |
| SOUTH CAROLINA | 180.21 | 180.26 | 178.08 | 177.25 | 715.8 |
| SOUTH DAKOTA | 113.85 | 102.32 | 90.16 | 83.88 | 390.21 |

DRUG CODE:9652 DRUG NAME: OXYMORPHONE

| STATE | 1ST QUARTER | 2ND QUARTER | 3RD QUARTER | 4TH QUARTER | TOTAL GRAMS |
|---|---|---|---|---|---|
| TENNESSEE | 504.97 | 472.72 | 445.89 | 424.61 | 1,848.18 |
| TEXAS | 66.52 | 65.03 | 62.28 | 63.82 | 257.65 |
| UTAH | 192.34 | 194.98 | 191.41 | 180.91 | 759.64 |
| VERMONT | 77.55 | 72.47 | 75.33 | 62.63 | 287.97 |
| VIRGINIA | 167.13 | 163.85 | 157.3 | 150.12 | 638.39 |
| WASHINGTON | 65.37 | 64.09 | 60.31 | 57.73 | 247.5 |
| WEST VIRGINIA | 241.5 | 241.31 | 214.49 | 196.79 | 894.09 |
| WISCONSIN | 67.19 | 61.2 | 57.02 | 54.03 | 239.45 |
| WYOMING | 77.65 | 85.01 | 64.82 | 70.11 | 297.59 |
| U.S. TOTAL | 131.96 | 127.08 | 122.55 | 118.46 | 500.05 |

DRUG CODE:9668 DRUG NAME: NOROXYMORPHONE

| STATE | 1ST QUARTER | 2ND QUARTER | 3RD QUARTER | 4TH QUARTER | TOTAL GRAMS |
|---|---|---|---|---|---|
| ALABAMA | 0.04 | 0.02 | 0.02 | 0.01 | 0.09 |
| ARIZONA | 0.01 | 0.01 | 0 | 0 | 0.03 |
| ARKANSAS | 0.53 | 0.38 | 0.22 | 0.81 | 1.94 |
| CALIFORNIA | 0.18 | 0.21 | 0.17 | 0.3 | 0.86 |
| COLORADO | 0 | 0 | 0 | 0 | 0.01 |
| CONNECTICUT | 0.02 | 0.01 | 0 | 0 | 0.03 |
| DISTRICT OF COLUMBIA | 0.67 | 0.53 | 0.01 | 0.33 | 1.54 |
| FLORIDA | 0.16 | 0.13 | 0.09 | 0.15 | 0.53 |
| GEORGIA | 0.06 | 0.08 | 0.07 | 0.04 | 0.24 |
| ILLINOIS | 0.14 | 0.07 | 0.11 | 0.09 | 0.41 |
| INDIANA | 0.01 | 0.01 | 0.01 | 0 | 0.02 |
| KANSAS | 0.02 | 0 | 0 | 0 | 0.02 |
| LOUISIANA | 0.01 | 0 | 0 | 0.01 | 0.01 |
| MARYLAND | 0.02 | 0.01 | 0.01 | 0 | 0.04 |
| MASSACHUSETTS | 0.01 | 0 | 0 | 0.02 | 0.03 |
| MISSISSIPPI | 0.27 | 0.12 | 0.12 | 0.03 | 0.55 |
| MISSOURI | 0.21 | 0.13 | 0.36 | 0.24 | 0.95 |
| NEBRASKA | 0 | 0 | 0 | 0.09 | 0.09 |
| NEVADA | 0 | 0.33 | 0.52 | 0.75 | 1.6 |
| NEW HAMPSHIRE | 0.12 | 0.07 | 0.03 | 0 | 0.21 |
| NEW JERSEY | 0 | 0.15 | 0.06 | 0.08 | 0.29 |
| NEW MEXICO | 0.16 | 0 | 0 | 0 | 0.16 |
| NEW YORK | 0.1 | 0.05 | 0.03 | 0.06 | 0.23 |
| NORTH CAROLINA | 0.01 | 0.01 | 0.01 | 0 | 0.02 |
| OHIO | 0.13 | 0.16 | 0.09 | 0.08 | 0.46 |
| OKLAHOMA | 0.04 | 0.01 | 0.02 | 0 | 0.07 |
| OREGON | 0.05 | 0.03 | 0.03 | 0 | 0.11 |
| PENNSYLVANIA | 0.01 | 0.03 | 0 | 0 | 0.05 |
| PUERTO RICO | 0.25 | 0.08 | 0.05 | 0.12 | 0.49 |
| RHODE ISLAND | 0.03 | 0.28 | 0 | 0 | 0.32 |
| SOUTH CAROLINA | 0.01 | 0 | 0 | 0 | 0.01 |
| TENNESSEE | 0.02 | 0.01 | 0.01 | 0.01 | 0.05 |
| TEXAS | 0.08 | 0.07 | 0.1 | 0.17 | 0.41 |
| UTAH | 0.04 | 0.01 | 0.01 | 0 | 0.06 |
| VERMONT | 0.12 | 0.03 | 0.03 | 0 | 0.18 |
| WASHINGTON | 0.13 | 0.05 | 0.05 | 0.06 | 0.29 |
| WISCONSIN | 0.01 | 0 | 0 | 0 | 0.01 |
| U.S. TOTAL | 0.08 | 0.07 | 0.07 | 0.1 | 0.32 |

DRUG CODE:9737 DRUG NAME: ALFENTANIL

| STATE | 1ST QUARTER | 2ND QUARTER | 3RD QUARTER | 4TH QUARTER | TOTAL GRAMS |
|---|---|---|---|---|---|
| ALABAMA | 0 | 0 | 0 | 0 | 0.01 |
| ALASKA | 0.13 | 0.1 | 0.1 | 0.13 | 0.46 |
| ARIZONA | 0.02 | 0.01 | 0.01 | 0.01 | 0.06 |
| ARKANSAS | 0 | 0 | 0 | 0 | 0.01 |
| CALIFORNIA | 0.05 | 0.05 | 0.04 | 0.04 | 0.18 |
| COLORADO | 0.02 | 0.01 | 0.02 | 0.01 | 0.06 |
| CONNECTICUT | 0.03 | 0.04 | 0.04 | 0.03 | 0.13 |
| FLORIDA | 0.03 | 0.02 | 0.02 | 0.03 | 0.11 |
| GEORGIA | 0.03 | 0.02 | 0.02 | 0.02 | 0.09 |
| GUAM | 0.03 | 0.03 | 0.06 | 0.02 | 0.13 |
| HAWAII | 0.03 | 0.04 | 0.02 | 0.05 | 0.13 |
| IDAHO | 0.08 | 0.08 | 0.08 | 0.11 | 0.34 |
| ILLINOIS | 0.03 | 0.04 | 0.03 | 0.03 | 0.13 |
| INDIANA | 0.04 | 0.04 | 0.04 | 0.03 | 0.14 |
| IOWA | 0.05 | 0.05 | 0.06 | 0.07 | 0.23 |
| KANSAS | 0.05 | 0.07 | 0.06 | 0.06 | 0.24 |
| KENTUCKY | 0.04 | 0.04 | 0.07 | 0.06 | 0.21 |
| LOUISIANA | 0.05 | 0.05 | 0.06 | 0.05 | 0.2 |
| MAINE | 0.04 | 0.06 | 0.04 | 0.06 | 0.2 |
| MARYLAND | 0.02 | 0.01 | 0.01 | 0.01 | 0.04 |
| MASSACHUSETTS | 0.02 | 0.02 | 0.01 | 0.02 | 0.08 |
| MICHIGAN | 0.01 | 0.01 | 0.01 | 0.02 | 0.05 |
| MINNESOTA | 0.07 | 0.07 | 0.06 | 0.06 | 0.26 |
| MISSISSIPPI | 0 | 0 | 0.01 | 0.01 | 0.02 |
| MISSOURI | 0.03 | 0.03 | 0.02 | 0.01 | 0.09 |
| MONTANA | 0.06 | 0.03 | 0.04 | 0.03 | 0.16 |
| NEBRASKA | 0.06 | 0.07 | 0.05 | 0.04 | 0.22 |
| NEVADA | 0.07 | 0.08 | 0.06 | 0.05 | 0.26 |
| NEW HAMPSHIRE | 0.03 | 0.03 | 0.03 | 0.03 | 0.12 |
| NEW JERSEY | 0 | 0 | 0 | 0 | 0.01 |
| NEW MEXICO | 0.06 | 0.05 | 0.05 | 0.08 | 0.23 |
| NEW YORK | 0.02 | 0.01 | 0.02 | 0.01 | 0.06 |
| NORTH CAROLINA | 0.03 | 0.03 | 0.03 | 0.03 | 0.12 |
| NORTH DAKOTA | 0.01 | 0.01 | 0.01 | 0.01 | 0.03 |
| OHIO | 0.05 | 0.04 | 0.04 | 0.05 | 0.18 |
| OKLAHOMA | 0.04 | 0.02 | 0.02 | 0.02 | 0.11 |
| OREGON | 0.1 | 0.1 | 0.11 | 0.11 | 0.41 |
| PENNSYLVANIA | 0.01 | 0 | 0.01 | 0 | 0.02 |
| RHODE ISLAND | 0.01 | 0.01 | 0.01 | 0 | 0.03 |
| SOUTH CAROLINA | 0 | 0 | 0 | 0 | 0 |
| SOUTH DAKOTA | 0.01 | 0.01 | 0.01 | 0.01 | 0.03 |
| TENNESSEE | 0.01 | 0.02 | 0.02 | 0.01 | 0.06 |

DRUG CODE:9737 DRUG NAME: ALFENTANIL

| STATE | 1ST QUARTER | 2ND QUARTER | 3RD QUARTER | 4TH QUARTER | TOTAL GRAMS |
|---|---|---|---|---|---|
| TEXAS | 0.03 | 0.02 | 0.02 | 0.02 | 0.1 |
| UTAH | 0.02 | 0.02 | 0.01 | 0.02 | 0.07 |
| VERMONT | 0 | 0 | 0 | 0 | 0 |
| VIRGINIA | 0.01 | 0.01 | 0.01 | 0.02 | 0.05 |

| | | | | | |
|---|---|---|---|---|---|
| WASHINGTON | 0.05 | 0.04 | 0.04 | 0.05 | 0.17 |
| WISCONSIN | 0.08 | 0.08 | 0.08 | 0.08 | 0.33 |
| WYOMING | 0.01 | 0.01 | 0 | 0.01 | 0.02 |
| U.S. TOTAL | 0.03 | 0.03 | 0.03 | 0.03 | 0.12 |

DRUG CODE:9739 DRUG NAME: REMIFENTANIL

| STATE | 1ST QUARTER | 2ND QUARTER | 3RD QUARTER | 4TH QUARTER | TOTAL GRAMS |
|---|---|---|---|---|---|
| ALABAMA | 0.05 | 0.07 | 0.07 | 0.07 | 0.26 |
| ALASKA | 0.4 | 0.39 | 0.36 | 0.48 | 1.62 |
| ARIZONA | 0.07 | 0.05 | 0.07 | 0.07 | 0.26 |
| ARKANSAS | 0.06 | 0.05 | 0.06 | 0.06 | 0.24 |
| CALIFORNIA | 0.08 | 0.09 | 0.09 | 0.09 | 0.35 |
| COLORADO | 0.12 | 0.12 | 0.13 | 0.13 | 0.5 |
| CONNECTICUT | 0.2 | 0.23 | 0.23 | 0.23 | 0.9 |
| DELAWARE | 0.37 | 0.38 | 0.41 | 0.26 | 1.41 |
| DISTRICT OF COLUMBIA | 0.1 | 0.13 | 0.16 | 0.13 | 0.51 |
| FLORIDA | 0.13 | 0.14 | 0.14 | 0.14 | 0.55 |
| GEORGIA | 0.05 | 0.04 | 0.05 | 0.05 | 0.2 |
| GUAM | 0 | 0 | 0.28 | 0 | 0.28 |
| HAWAII | 0.11 | 0.1 | 0.09 | 0.1 | 0.39 |
| IDAHO | 0.11 | 0.15 | 0.12 | 0.19 | 0.57 |
| ILLINOIS | 0.11 | 0.1 | 0.12 | 0.11 | 0.44 |
| INDIANA | 0.12 | 0.12 | 0.11 | 0.14 | 0.49 |
| IOWA | 0.09 | 0.1 | 0.09 | 0.1 | 0.38 |
| KANSAS | 0.06 | 0.06 | 0.07 | 0.09 | 0.28 |
| KENTUCKY | 0.1 | 0.1 | 0.11 | 0.1 | 0.43 |
| LOUISIANA | 0.09 | 0.1 | 0.09 | 0.1 | 0.37 |
| MAINE | 0.08 | 0.08 | 0.09 | 0.13 | 0.39 |
| MARYLAND | 0.12 | 0.12 | 0.13 | 0.12 | 0.49 |
| MASSACHUSETTS | 0.28 | 0.3 | 0.29 | 0.3 | 1.17 |
| MICHIGAN | 0.1 | 0.1 | 0.1 | 0.11 | 0.42 |
| MINNESOTA | 0.13 | 0.12 | 0.11 | 0.15 | 0.5 |
| MISSISSIPPI | 0.02 | 0.02 | 0.02 | 0.01 | 0.07 |
| MISSOURI | 0.08 | 0.08 | 0.1 | 0.08 | 0.35 |
| MONTANA | 0.19 | 0.21 | 0.15 | 0.18 | 0.73 |
| NEBRASKA | 0.04 | 0.06 | 0.05 | 0.06 | 0.22 |
| NEVADA | 0.04 | 0.03 | 0.03 | 0.04 | 0.13 |
| NEW HAMPSHIRE | 0.09 | 0.09 | 0.1 | 0.09 | 0.36 |
| NEW JERSEY | 0.13 | 0.14 | 0.16 | 0.14 | 0.56 |
| NEW MEXICO | 0.05 | 0.04 | 0.05 | 0.06 | 0.2 |
| NEW YORK | 0.17 | 0.19 | 0.17 | 0.19 | 0.73 |
| NORTH CAROLINA | 0.14 | 0.15 | 0.15 | 0.16 | 0.59 |
| NORTH DAKOTA | 0.04 | 0.06 | 0.06 | 0.08 | 0.24 |
| OHIO | 0.16 | 0.16 | 0.17 | 0.18 | 0.66 |
| OKLAHOMA | 0.08 | 0.05 | 0.06 | 0.07 | 0.26 |
| OREGON | 0.09 | 0.07 | 0.06 | 0.08 | 0.29 |
| PENNSYLVANIA | 0.27 | 0.28 | 0.28 | 0.26 | 1.08 |
| PUERTO RICO | 0 | 0 | 0 | 0 | 0 |
| RHODE ISLAND | 0.04 | 0.03 | 0.05 | 0.05 | 0.18 |

DRUG CODE:9739 DRUG NAME: REMIFENTANIL

| STATE | 1ST QUARTER | 2ND QUARTER | 3RD QUARTER | 4TH QUARTER | TOTAL GRAMS |
|---|---|---|---|---|---|

| | | | | | |
|---|---|---|---|---|---|
| SOUTH CAROLINA | 0.1 | 0.11 | 0.11 | 0.11 | 0.43 |
| SOUTH DAKOTA | 0.07 | 0.07 | 0.07 | 0.08 | 0.29 |
| TENNESSEE | 0.08 | 0.08 | 0.07 | 0.09 | 0.33 |
| TEXAS | 0.1 | 0.11 | 0.11 | 0.11 | 0.43 |
| UTAH | 0.57 | 0.61 | 0.59 | 0.62 | 2.4 |
| VERMONT | 0.08 | 0.07 | 0.1 | 0.09 | 0.35 |
| VIRGINIA | 0.08 | 0.07 | 0.08 | 0.08 | 0.31 |
| WASHINGTON | 0.18 | 0.18 | 0.18 | 0.2 | 0.74 |
| WEST VIRGINIA | 0.26 | 0.27 | 0.3 | 0.3 | 1.13 |
| WISCONSIN | 0.1 | 0.1 | 0.11 | 0.12 | 0.43 |
| WYOMING | 0.09 | 0.09 | 0.07 | 0.09 | 0.33 |
| U.S. TOTAL | 0.12 | 0.13 | 0.13 | 0.13 | 0.51 |

DRUG CODE:9740 DRUG NAME: SUFENTANIL BASE

| STATE | 1ST QUARTER | 2ND QUARTER | 3RD QUARTER | 4TH QUARTER | TOTAL GRAMS |
|---|---|---|---|---|---|
| ALABAMA | 0.05 | 0.04 | 0.03 | 0.04 | 0.16 |
| ALASKA | 0 | 0 | 0 | 0 | 0 |
| ARIZONA | 0 | 0 | 0 | 0 | 0 |
| ARKANSAS | 0.01 | 0 | 0.01 | 0 | 0.02 |
| CALIFORNIA | 0.03 | 0.01 | 0.03 | 0.03 | 0.11 |
| COLORADO | 0.01 | 0 | 0 | 0 | 0.02 |
| CONNECTICUT | 0 | 0 | 0.02 | 0.01 | 0.04 |
| DELAWARE | 0 | 0 | 0 | 0 | 0 |
| DISTRICT OF COLUMBIA | 0 | 0 | 0 | 0 | 0.01 |
| FLORIDA | 0.01 | 0.02 | 0.01 | 0.01 | 0.06 |
| GEORGIA | 0 | 0 | 0 | 0 | 0.02 |
| HAWAII | 0 | 0 | 0 | 0 | 0 |
| IDAHO | 0 | 0 | 0 | 0 | 0.01 |
| ILLINOIS | 0.01 | 0 | 0 | 0 | 0.02 |
| INDIANA | 0.01 | 0.01 | 0 | 0.01 | 0.02 |
| IOWA | 0 | 0 | 0 | 0 | 0 |
| KANSAS | 0.01 | 0.01 | 0.01 | 0.01 | 0.04 |
| KENTUCKY | 0.01 | 0.01 | 0.01 | 0.01 | 0.03 |
| LOUISIANA | 0.03 | 0.02 | 0.02 | 0.02 | 0.08 |
| MAINE | 0 | 0 | 0 | 0 | 0 |
| MARYLAND | 0 | 0 | 0 | 0 | 0 |
| MASSACHUSETTS | 0 | 0 | 0.01 | 0 | 0.01 |
| MICHIGAN | 0.01 | 0.01 | 0.01 | 0.01 | 0.02 |
| MINNESOTA | 0 | 0 | 0 | 0 | 0.02 |
| MISSISSIPPI | 0.27 | 0.39 | 0.35 | 0.4 | 1.41 |
| MISSOURI | 0.01 | 0.01 | 0.01 | 0.01 | 0.03 |
| MONTANA | 0.01 | 0 | 0 | 0 | 0.02 |
| NEBRASKA | 0.01 | 0 | 0 | 0.01 | 0.02 |
| NEVADA | 0.01 | 0 | 0 | 0 | 0.01 |
| NEW HAMPSHIRE | 0 | 0 | 0 | 0 | 0.01 |
| NEW JERSEY | 0.01 | 0.01 | 0.01 | 0.02 | 0.05 |
| NEW MEXICO | 0 | 0 | 0 | 0 | 0.01 |
| NEW YORK | 0.01 | 0.01 | 0.01 | 0.01 | 0.04 |
| NORTH CAROLINA | 0 | 0 | 0 | 0.01 | 0.02 |
| NORTH DAKOTA | 0.01 | 0 | 0 | 0 | 0.02 |
| OHIO | 0.01 | 0.01 | 0.01 | 0 | 0.02 |
| OKLAHOMA | 0.01 | 0 | 0.01 | 0 | 0.02 |
| OREGON | 0.01 | 0.01 | 0.01 | 0.01 | 0.03 |
| PENNSYLVANIA | 0.01 | 0.01 | 0.01 | 0.01 | 0.04 |

| | | | | | |
|---|---|---|---|---|---|
| PUERTO RICO | 0 | 0.01 | 0.01 | 0 | 0.02 |
| RHODE ISLAND | 0 | 0 | 0 | 0 | 0 |
| SOUTH CAROLINA | 0.02 | 0.02 | 0 | 0.02 | 0.05 |

DRUG CODE:9740 DRUG NAME: SUFENTANIL BASE

| STATE | 1ST QUARTER | 2ND QUARTER | 3RD QUARTER | 4TH QUARTER | TOTAL GRAMS |
|---|---|---|---|---|---|
| SOUTH DAKOTA | 0.01 | 0 | 0 | 0 | 0.02 |
| TENNESSEE | 0.01 | 0.01 | 0.01 | 0 | 0.03 |
| TEXAS | 0.01 | 0.01 | 0.01 | 0.01 | 0.03 |
| UTAH | 0.01 | 0.01 | 0.01 | 0.01 | 0.03 |
| VERMONT | 0 | 0 | 0 | 0 | 0.01 |
| VIRGINIA | 0.01 | 0.01 | 0 | 0.01 | 0.02 |
| WASHINGTON | 0 | 0 | 0 | 0 | 0.01 |
| WEST VIRGINIA | 0 | 0 | 0 | 0 | 0.01 |
| WISCONSIN | 0.01 | 0.04 | 0.01 | 0 | 0.06 |
| WYOMING | 0 | 0 | 0 | 0 | 0.01 |
| U.S. TOTAL | 0.01 | 0.01 | 0.01 | 0.01 | 0.04 |

DRUG CODE:9780 DRUG NAME: TAPENTADOL

| STATE | 1ST QUARTER | 2ND QUARTER | 3RD QUARTER | 4TH QUARTER | TOTAL GRAMS |
|---|---|---|---|---|---|
| ALABAMA | 1,384.19 | 1,379.03 | 1,358.54 | 1,377.71 | 5,499.47 |
| ALASKA | 256.54 | 247.88 | 301.45 | 285.26 | 1,091.12 |
| ARIZONA | 592.56 | 554.11 | 575.77 | 620.73 | 2,343.16 |
| ARKANSAS | 910.19 | 940.42 | 935.61 | 904.19 | 3,690.41 |
| CALIFORNIA | 318.44 | 324.61 | 323.95 | 323.53 | 1,290.53 |
| COLORADO | 550.98 | 537.27 | 543.05 | 565.55 | 2,196.85 |
| CONNECTICUT | 341.02 | 363.34 | 394.77 | 408.95 | 1,508.08 |
| DELAWARE | 679.89 | 746.99 | 787.53 | 819.88 | 3,034.30 |
| DISTRICT OF COLUMBIA | 240.78 | 245.05 | 241.97 | 227.01 | 954.82 |
| FLORIDA | 625.23 | 618.91 | 612.77 | 619.95 | 2,476.87 |
| GEORGIA | 726.73 | 730.3 | 749.96 | 772.5 | 2,979.50 |
| HAWAII | 204.95 | 210.21 | 192.16 | 206.57 | 813.9 |
| IDAHO | 344.13 | 378.07 | 396.5 | 458.73 | 1,577.43 |
| ILLINOIS | 330.72 | 351.87 | 353.78 | 364.25 | 1,400.63 |
| INDIANA | 612.19 | 618.74 | 615.67 | 641.99 | 2,488.59 |
| IOWA | 407.01 | 432.22 | 422.87 | 467.05 | 1,729.15 |
| KANSAS | 398.13 | 405.77 | 400.82 | 388.96 | 1,593.68 |
| KENTUCKY | 386.02 | 363.74 | 377.64 | 395.06 | 1,522.46 |
| LOUISIANA | 343.89 | 358.82 | 374.2 | 375.2 | 1,452.11 |
| MAINE | 139.95 | 166.71 | 158.24 | 153.2 | 618.09 |
| MARYLAND | 589.2 | 614.1 | 619.23 | 615.19 | 2,437.72 |
| MASSACHUSETTS | 273.56 | 272.4 | 283.42 | 289.01 | 1,118.38 |
| MICHIGAN | 231.51 | 246.58 | 242.72 | 246.4 | 967.21 |
| MINNESOTA | 97.04 | 98.86 | 91.27 | 96.72 | 383.89 |
| MISSISSIPPI | 467.01 | 490.23 | 494.76 | 495.42 | 1,947.41 |
| MISSOURI | 495.71 | 484.94 | 470.02 | 476.5 | 1,927.18 |
| MONTANA | 246.31 | 255.3 | 258.39 | 261.67 | 1,021.66 |
| NEBRASKA | 576.62 | 568.51 | 570.79 | 596.44 | 2,312.36 |
| NEVADA | 481.14 | 483.72 | 461.65 | 479.29 | 1,905.80 |
| NEW HAMPSHIRE | 476.46 | 462.64 | 518.47 | 530.4 | 1,987.97 |
| NEW JERSEY | 563.53 | 584.89 | 576.91 | 589.32 | 2,314.65 |

| | | | | | |
|---|---|---|---|---|---|
| NEW MEXICO | 206.64 | 233.35 | 213.97 | 206.93 | 860.88 |
| NEW YORK | 569.28 | 555.38 | 547.57 | 553.35 | 2,225.58 |
| NORTH CAROLINA | 1,075.72 | 1,080.40 | 1,067.40 | 1,106.07 | 4,329.59 |
| NORTH DAKOTA | 386.86 | 374.52 | 405.82 | 382.85 | 1,550.05 |
| OHIO | 616.89 | 607.32 | 613.66 | 608.69 | 2,446.56 |
| OKLAHOMA | 671.23 | 678.57 | 679.82 | 734.2 | 2,763.83 |
| OREGON | 197.43 | 203.6 | 216.86 | 220.36 | 838.24 |
| PENNSYLVANIA | 534.2 | 557.43 | 560.63 | 575.8 | 2,228.06 |
| PUERTO RICO | 34.43 | 27.7 | 25.63 | 26.4 | 114.16 |
| RHODE ISLAND | 196.8 | 225.97 | 227.4 | 223.07 | 873.25 |
| SOUTH CAROLINA | 943.99 | 964.37 | 980 | 1,016.71 | 3,905.07 |

DRUG CODE:9780 DRUG NAME: TAPENTADOL

| STATE | 1ST QUARTER | 2ND QUARTER | 3RD QUARTER | 4TH QUARTER | TOTAL GRAMS |
|---|---|---|---|---|---|
| SOUTH DAKOTA | 744.86 | 720.48 | 724.47 | 696.59 | 2,886.40 |
| TENNESSEE | 527.32 | 557.87 | 584.44 | 583.98 | 2,253.61 |
| TEXAS | 398.64 | 426.86 | 434.07 | 459.31 | 1,718.88 |
| UTAH | 569.4 | 593.98 | 586.84 | 597.66 | 2,347.87 |
| VERMONT | 237.32 | 201.92 | 199.76 | 239.64 | 878.64 |
| VIRGIN ISLANDS | 5.46 | 8.19 | 2.73 | 23.67 | 40.06 |
| VIRGINIA | 611.21 | 616.36 | 609.46 | 615 | 2,452.02 |
| WASHINGTON | 291.06 | 291.56 | 295.51 | 283.37 | 1,161.49 |
| WEST VIRGINIA | 316.65 | 337.43 | 324.42 | 335.92 | 1,314.41 |
| WISCONSIN | 319.15 | 313.14 | 313.01 | 317.08 | 1,262.38 |
| WYOMING | 381.81 | 307.3 | 307.03 | 330.63 | 1,326.77 |
| U.S. TOTAL | 489.14 | 495.77 | 497.35 | 508.2 | 1,990.46 |

DRUG CODE:9801 DRUG NAME: FENTANYL BASE

| STATE | 1ST QUARTER | 2ND QUARTER | 3RD QUARTER | 4TH QUARTER | TOTAL GRAMS |
|---|---|---|---|---|---|
| ALABAMA | 41.21 | 39.82 | 38.62 | 38.49 | 158.14 |
| ALASKA | 39.18 | 38.79 | 36.86 | 34.4 | 149.24 |
| AMERICAN SAMOA | 0.58 | 0 | 0.25 | 0.02 | 0.84 |
| ARIZONA | 37.58 | 36.62 | 34.85 | 33.92 | 142.97 |
| ARKANSAS | 40.61 | 40.11 | 38.51 | 36.98 | 156.2 |
| CALIFORNIA | 24.41 | 23.28 | 22.35 | 22.34 | 92.38 |
| COLORADO | 33.26 | 31.99 | 30.39 | 28.72 | 124.36 |
| CONNECTICUT | 35.1 | 35.1 | 32.05 | 29.49 | 131.73 |
| DELAWARE | 47.37 | 44.82 | 45.2 | 43.42 | 180.81 |
| DISTRICT OF COLUMBIA | 16.93 | 17.86 | 15.91 | 15.38 | 66.09 |
| FLORIDA | 37.31 | 36.36 | 34.53 | 34.97 | 143.16 |
| GEORGIA | 32.94 | 32.48 | 32.03 | 31.59 | 129.04 |
| GUAM | 5.67 | 5.69 | 5.88 | 4.37 | 21.61 |
| HAWAII | 16.43 | 16.36 | 15.15 | 14.38 | 62.31 |
| IDAHO | 45.37 | 44.41 | 42.03 | 40.56 | 172.36 |
| ILLINOIS | 27.85 | 27.45 | 26.78 | 25.88 | 107.96 |
| INDIANA | 38.19 | 36.97 | 34.76 | 33.63 | 143.54 |
| IOWA | 33.64 | 33.25 | 32.15 | 31.19 | 130.24 |
| KANSAS | 46.35 | 44.8 | 44.59 | 41.99 | 177.73 |
| KENTUCKY | 32.97 | 32.31 | 31.2 | 29.44 | 125.92 |
| LOUISIANA | 32.24 | 31.95 | 30.33 | 29.53 | 124.05 |
| MAINE | 42.37 | 41.09 | 38.45 | 33.75 | 155.66 |
| MARYLAND | 31.42 | 31.42 | 30.27 | 28.92 | 122.02 |

| | | | | | |
|---|---|---|---|---|---|
| MASSACHUSETTS | 24.79 | 24.01 | 22.86 | 22.01 | 93.67 |
| MICHIGAN | 39.89 | 39.2 | 35.95 | 36.04 | 151.08 |
| MINNESOTA | 25.23 | 24.75 | 23.23 | 21.76 | 94.97 |
| MISSISSIPPI | 47.65 | 47.05 | 44.77 | 45.47 | 184.95 |
| MISSOURI | 44.76 | 43.27 | 41.03 | 40.17 | 169.22 |
| MONTANA | 41.92 | 42.02 | 40.34 | 38.27 | 162.56 |
| NEBRASKA | 33.24 | 34.13 | 31.95 | 31.65 | 130.97 |
| NEVADA | 29.54 | 27.9 | 27.04 | 25.79 | 110.27 |
| NEW HAMPSHIRE | 34.67 | 34.28 | 33.55 | 30.87 | 133.37 |
| NEW JERSEY | 32.83 | 33.27 | 31.79 | 32.86 | 130.74 |
| NEW MEXICO | 28.08 | 28.05 | 26.91 | 25.9 | 108.94 |
| NEW YORK | 30.27 | 29.24 | 27.79 | 25.84 | 113.15 |
| NORTH CAROLINA | 39.85 | 38.72 | 36.2 | 34.34 | 149.11 |
| NORTH DAKOTA | 50.38 | 46.67 | 44.38 | 37.07 | 178.5 |
| OHIO | 28.59 | 27.63 | 26.18 | 24.85 | 107.25 |
| OKLAHOMA | 56.13 | 55.21 | 53.83 | 50.54 | 215.69 |
| OREGON | 34.58 | 33.44 | 30.99 | 28.34 | 127.35 |
| PENNSYLVANIA | 48.86 | 48.23 | 46.47 | 44.47 | 188.03 |
| PUERTO RICO | 5.87 | 5.86 | 5.61 | 5.81 | 23.15 |

DRUG CODE:9801 DRUG NAME: FENTANYL BASE

| STATE | 1ST QUARTER | 2ND QUARTER | 3RD QUARTER | 4TH QUARTER | TOTAL GRAMS |
|---|---|---|---|---|---|
| RHODE ISLAND | 22.66 | 22.56 | 21.9 | 20.5 | 87.62 |
| SOUTH CAROLINA | 34.64 | 33.44 | 32.79 | 34.46 | 135.32 |
| SOUTH DAKOTA | 47.2 | 44.72 | 42.8 | 41.95 | 176.66 |
| TENNESSEE | 39.96 | 37.99 | 37.68 | 35.95 | 151.59 |
| TEXAS | 27.63 | 27.22 | 26.75 | 28.29 | 109.88 |
| UTAH | 35.89 | 35.92 | 36.02 | 33.94 | 141.77 |
| VERMONT | 43.77 | 39.19 | 36.6 | 34.74 | 154.29 |
| VIRGIN ISLANDS | 6.5 | 6.01 | 6.52 | 5.28 | 24.31 |
| VIRGINIA | 28.56 | 27.65 | 26.21 | 25 | 107.43 |
| WASHINGTON | 28.46 | 28.3 | 27.04 | 25.6 | 109.39 |
| WEST VIRGINIA | 38.56 | 37.62 | 34.99 | 33.11 | 144.28 |
| WISCONSIN | 35.7 | 34.92 | 32.38 | 30.74 | 133.74 |
| WYOMING | 33.69 | 32.34 | 32.43 | 30.83 | 129.28 |
| U.S. TOTAL | 33 | 32.21 | 30.83 | 30.08 | 126.12 |

CUMULATIVE DISTRIBUTION BY STATE IN GRAMS PER 100,000 POPULATION

DRUG CODE:1100 DRUG NAME:AMPHETAMINE

| RANK | STATE | 2010 POP | TOTAL GRAMS | GRAMS/100K POP |
|---|---|---|---|---|
| 1 | RHODE ISLAND | 1,052,567 | 139,256.35 | 13,230.16 |
| 2 | LOUISIANA | 4,533,372 | 573,990.72 | 12,661.45 |
| 3 | SOUTH CAROLINA | 4,625,364 | 525,411.90 | 11,359.36 |
| 4 | NEW HAMPSHIRE | 1,316,470 | 143,949.41 | 10,934.50 |
| 5 | UTAH | 2,763,885 | 300,188.17 | 10,861.09 |
| 6 | ALABAMA | 4,779,736 | 507,395.30 | 10,615.55 |
| 7 | MASSACHUSETTS | 6,547,629 | 686,612.02 | 10,486.42 |
| 8 | DELAWARE | 897,934 | 85,965.56 | 9,573.71 |
| 9 | WISCONSIN | 5,686,986 | 536,585.21 | 9,435.32 |
| 10 | MICHIGAN | 9,883,640 | 917,725.03 | 9,285.29 |
| 11 | VIRGINIA | 8,001,024 | 691,196 | 8,638.84 |
| 12 | MISSOURI | 5,988,927 | 514,906.93 | 8,597.65 |
| 13 | MISSISSIPPI | 2,967,297 | 238,958.91 | 8,053.08 |
| 14 | NORTH CAROLINA | 9,535,483 | 763,562.99 | 8,007.60 |
| 15 | KANSAS | 2,853,118 | 227,219.10 | 7,963.89 |
| 16 | MINNESOTA | 5,303,925 | 422,148.97 | 7,959.18 |
| 17 | TENNESSEE | 6,346,105 | 499,282.71 | 7,867.55 |
| 18 | SOUTH DAKOTA | 814,180 | 62,860.68 | 7,720.74 |
| 19 | INDIANA | 6,483,802 | 487,458.41 | 7,518.10 |
| 20 | DISTRICT OF COLUMBIA | 601,723 | 44,966.88 | 7,473.02 |
| 21 | GEORGIA | 9,687,653 | 705,967.94 | 7,287.30 |
| 22 | OHIO | 11,536,504 | 811,851.36 | 7,037.24 |
| 23 | ARKANSAS | 2,915,918 | 192,688.33 | 6,608.15 |
| 24 | PENNSYLVANIA | 12,702,379 | 839,220.82 | 6,606.80 |
| 25 | MAINE | 1,328,361 | 85,431.59 | 6,431.35 |
| 26 | WASHINGTON | 6,724,540 | 419,978.28 | 6,245.46 |
| 27 | VERMONT | 625,741 | 39,044.93 | 6,239.79 |
| 28 | IOWA | 3,046,355 | 187,482.50 | 6,154.32 |
| 29 | TEXAS | 25,145,561 | 1,534,182.94 | 6,101.21 |
| 30 | MARYLAND | 5,773,552 | 347,698.85 | 6,022.27 |
| 31 | IDAHO | 1,567,582 | 93,501.74 | 5,964.71 |
| 32 | OREGON | 3,831,074 | 227,137.99 | 5,928.83 |
| 33 | KENTUCKY | 4,339,367 | 256,518.64 | 5,911.43 |
| 34 | COLORADO | 5,029,196 | 296,470.56 | 5,894.99 |
| 35 | FLORIDA | 18,801,310 | 1,095,274.86 | 5,825.52 |
| 36 | CONNECTICUT | 3,574,097 | 200,601.92 | 5,612.66 |
| 37 | ILLINOIS | 12,830,632 | 718,040.62 | 5,596.30 |
| 38 | OKLAHOMA | 3,751,351 | 203,127.08 | 5,414.77 |
| 39 | NEBRASKA | 1,826,341 | 96,907.52 | 5,306.10 |
| 40 | MONTANA | 989,415 | 52,016.75 | 5,257.32 |
| 41 | NORTH DAKOTA | 672,591 | 35,247.71 | 5,240.59 |
| 42 | WEST VIRGINIA | 1,852,994 | 96,732.51 | 5,220.34 |
| 43 | ARIZONA | 6,392,017 | 303,893.93 | 4,754.27 |
| 44 | NEW YORK | 19,378,102 | 902,548.59 | 4,657.57 |
| 45 | WYOMING | 563,626 | 25,380.01 | 4,502.99 |
| 46 | NEW JERSEY | 8,791,894 | 395,118.29 | 4,494.12 |
| 47 | ALASKA | 710,231 | 31,327.50 | 4,410.89 |
| 48 | NEW MEXICO | 2,059,179 | 77,115.99 | 3,744.99 |
| 49 | NEVADA | 2,700,551 | 96,145.41 | 3,560.21 |
| 50 | CALIFORNIA | 37,253,956 | 1,164,173.35 | 3,124.97 |
| 51 | HAWAII | 1,360,301 | 36,450.39 | 2,679.58 |
| 52 | VIRGIN ISLANDS | 109,825 | 1,251.26 | 1,139.32 |
| 53 | GUAM | 178,430 | 1,754.70 | 983.41 |

DRUG CODE:1100 DRUG NAME:AMPHETAMINE

| RANK | STATE | 2010 POP | TOTAL GRAMS | GRAMS/100K POP |
|---|---|---|---|---|
| 54 | PUERTO RICO | 3,725,789 | 32,571.15 | 874.21 |
| 55 | AMERICAN SAMOA | 65,628 | 6.54 | 9.97 |
| U.S. GRAMS / PER 100K: | | 312,825,210 | 19,972,503.78 | 6,384.56 |

DRUG CODE:1105B DRUG NAME:DL-METHAMPHETAMINE RACEMIC BASE

| RANK | STATE | 2010 POP | TOTAL GRAMS | GRAMS/100K POP |
|---|---|---|---|---|
| 1 | MINNESOTA | 5,303,925 | 142.25 | 2.68 |
| 2 | NEW MEXICO | 2,059,179 | 51.84 | 2.52 |
| 3 | DISTRICT OF COLUMBIA | 601,723 | 11.25 | 1.87 |
| 4 | OREGON | 3,831,074 | 66.71 | 1.74 |
| 5 | MISSOURI | 5,988,927 | 97.25 | 1.62 |
| 6 | WISCONSIN | 5,686,986 | 76.35 | 1.34 |
| 7 | ARIZONA | 6,392,017 | 83.99 | 1.31 |
| 8 | IDAHO | 1,567,582 | 18.08 | 1.15 |
| 9 | IOWA | 3,046,355 | 33.35 | 1.09 |
| 10 | CONNECTICUT | 3,574,097 | 36.17 | 1.01 |
| 11 | CALIFORNIA | 37,253,956 | 352.42 | 0.95 |
| 12 | HAWAII | 1,360,301 | 12.46 | 0.92 |
| 13 | OKLAHOMA | 3,751,351 | 28.93 | 0.77 |
| 14 | NEBRASKA | 1,826,341 | 13.26 | 0.73 |
| 15 | NORTH DAKOTA | 672,591 | 4.82 | 0.72 |
| 16 | ALABAMA | 4,779,736 | 32.95 | 0.69 |
| 17 | TEXAS | 25,145,561 | 173.2 | 0.69 |
| 18 | NORTH CAROLINA | 9,535,483 | 53.45 | 0.56 |
| 19 | RHODE ISLAND | 1,052,567 | 5.63 | 0.53 |
| 20 | WEST VIRGINIA | 1,852,994 | 9.64 | 0.52 |
| 21 | NEVADA | 2,700,551 | 14.06 | 0.52 |
| 22 | PENNSYLVANIA | 12,702,379 | 54.65 | 0.43 |
| 23 | NEW JERSEY | 8,791,894 | 33.35 | 0.38 |
| 24 | OHIO | 11,536,504 | 37.37 | 0.32 |
| 25 | NEW YORK | 19,378,102 | 62.69 | 0.32 |
| 26 | INDIANA | 6,483,802 | 20.09 | 0.31 |
| 27 | SOUTH CAROLINA | 4,625,364 | 13.26 | 0.29 |
| 28 | MASSACHUSETTS | 6,547,629 | 17.68 | 0.27 |
| 29 | LOUISIANA | 4,533,372 | 11.25 | 0.25 |
| 30 | GEORGIA | 9,687,653 | 24.51 | 0.25 |
| 31 | KENTUCKY | 4,339,367 | 10.45 | 0.24 |
| 32 | MAINE | 1,328,361 | 2.81 | 0.21 |
| 33 | MARYLAND | 5,773,552 | 12.06 | 0.21 |
| 34 | ARKANSAS | 2,915,918 | 6.03 | 0.21 |
| 35 | COLORADO | 5,029,196 | 10.45 | 0.21 |
| 36 | KANSAS | 2,853,118 | 6.03 | 0.21 |
| 37 | FLORIDA | 18,801,310 | 35.76 | 0.19 |
| 38 | NEW HAMPSHIRE | 1,316,470 | 2.41 | 0.18 |
| 39 | WASHINGTON | 6,724,540 | 9.24 | 0.14 |
| 40 | ILLINOIS | 12,830,632 | 14.87 | 0.12 |
| 41 | ALASKA | 710,231 | 0.8 | 0.11 |
| 42 | MISSISSIPPI | 2,967,297 | 2.81 | 0.09 |
| 43 | UTAH | 2,763,885 | 2.01 | 0.07 |
| 44 | WYOMING | 563,626 | 0.4 | 0.07 |
| 45 | MICHIGAN | 9,883,640 | 4.82 | 0.05 |
| 46 | VIRGINIA | 8,001,024 | 2.81 | 0.04 |
| 47 | TENNESSEE | 6,346,105 | 2.81 | 0.04 |
| U.S. GRAMS / PER 100K: | | 312,825,210 | 1,719.52 | 0.55 |

CUMULATIVE DISTRIBUTION BY STATE IN GRAMS PER 100,000 POPULATION

DRUG CODE:1105D DRUG NAME:D-METHAMPHETAMINE

| RANK | STATE | 2010 POP | TOTAL GRAMS | GRAMS/100K POP |
|---|---|---|---|---|
| 1 | SOUTH DAKOTA | 814,180 | 49.01 | 6.02 |
| 2 | WASHINGTON | 6,724,540 | 367.15 | 5.46 |
| 3 | MINNESOTA | 5,303,925 | 288.82 | 5.45 |
| 4 | UTAH | 2,763,885 | 129.75 | 4.69 |

DRUG CODE:1105D DRUG NAME:D-METHAMPHETAMINE

| RANK | STATE | 2010 POP | TOTAL GRAMS | GRAMS/100K POP |
|---|---|---|---|---|
| 5 | OREGON | 3,831,074 | 174.34 | 4.55 |
| 6 | WYOMING | 563,626 | 24.5 | 4.35 |
| 7 | PENNSYLVANIA | 12,702,379 | 487.34 | 3.84 |
| 8 | COLORADO | 5,029,196 | 169.92 | 3.38 |
| 9 | WISCONSIN | 5,686,986 | 183.98 | 3.24 |
| 10 | MARYLAND | 5,773,552 | 185.59 | 3.21 |
| 11 | NEVADA | 2,700,551 | 85.16 | 3.15 |
| 12 | NEW MEXICO | 2,059,179 | 63.87 | 3.1 |
| 13 | MISSOURI | 5,988,927 | 183.98 | 3.07 |
| 14 | CALIFORNIA | 37,253,956 | 1,086.28 | 2.92 |
| 15 | OHIO | 11,536,504 | 316.14 | 2.74 |
| 16 | VIRGINIA | 8,001,024 | 217.72 | 2.72 |
| 17 | MASSACHUSETTS | 6,547,629 | 157.06 | 2.4 |
| 18 | DELAWARE | 897,934 | 20.49 | 2.28 |
| 19 | NORTH CAROLINA | 9,535,483 | 193.62 | 2.03 |
| 20 | NEW HAMPSHIRE | 1,316,470 | 26.19 | 1.99 |
| 21 | IOWA | 3,046,355 | 59.05 | 1.94 |
| 22 | CONNECTICUT | 3,574,097 | 65.88 | 1.84 |
| 23 | ALABAMA | 4,779,736 | 86.77 | 1.82 |
| 24 | KANSAS | 2,853,118 | 51.42 | 1.8 |
| 25 | ALASKA | 710,231 | 12.45 | 1.75 |
| 26 | ILLINOIS | 12,830,632 | 207.68 | 1.62 |
| 27 | IDAHO | 1,567,582 | 24.5 | 1.56 |
| 28 | NEBRASKA | 1,826,341 | 28.12 | 1.54 |
| 29 | VERMONT | 625,741 | 9.64 | 1.54 |
| 30 | NEW YORK | 19,378,102 | 297.34 | 1.53 |
| 31 | NEW JERSEY | 8,791,894 | 130.95 | 1.49 |
| 32 | LOUISIANA | 4,533,372 | 67.49 | 1.49 |
| 33 | MISSISSIPPI | 2,967,297 | 39.37 | 1.33 |
| 34 | GEORGIA | 9,687,653 | 120.11 | 1.24 |
| 35 | TEXAS | 25,145,561 | 307.3 | 1.22 |
| 36 | MICHIGAN | 9,883,640 | 113.28 | 1.15 |
| 37 | INDIANA | 6,483,802 | 71.9 | 1.11 |
| 38 | HAWAII | 1,360,301 | 14.86 | 1.09 |
| 39 | NORTH DAKOTA | 672,591 | 7.23 | 1.08 |
| 40 | SOUTH CAROLINA | 4,625,364 | 49.41 | 1.07 |
| 41 | ARIZONA | 6,392,017 | 67.08 | 1.05 |
| 42 | MONTANA | 989,415 | 9.64 | 0.97 |
| 43 | OKLAHOMA | 3,751,351 | 35.75 | 0.95 |
| 44 | RHODE ISLAND | 1,052,567 | 9.24 | 0.88 |
| 45 | TENNESSEE | 6,346,105 | 45.39 | 0.72 |
| 46 | MAINE | 1,328,361 | 8.44 | 0.64 |
| 47 | DISTRICT OF COLUMBIA | 601,723 | 3.62 | 0.6 |
| 48 | ARKANSAS | 2,915,918 | 14.06 | 0.48 |
| 49 | KENTUCKY | 4,339,367 | 20.89 | 0.48 |
| 50 | FLORIDA | 18,801,310 | 84.76 | 0.45 |
| 51 | WEST VIRGINIA | 1,852,994 | 2.01 | 0.11 |
| U.S. GRAMS / PER 100K: | | 312,825,210 | 6,476.53 | 2.07 |

CUMULATIVE DISTRIBUTION BY STATE IN GRAMS PER 100,000 POPULATION

DRUG CODE:1205 DRUG NAME:LISDEXAMFETAMINE

| RANK | STATE | 2010 POP | TOTAL GRAMS | GRAMS/100K POP |
|---|---|---|---|---|
| 1 | LOUISIANA | 4,533,372 | 386,039.57 | 8,515.51 |
| 2 | DELAWARE | 897,934 | 57,222.65 | 6,372.70 |
| 3 | ALABAMA | 4,779,736 | 285,042.61 | 5,963.56 |
| 4 | SOUTH DAKOTA | 814,180 | 47,289.74 | 5,808.27 |
| 5 | SOUTH CAROLINA | 4,625,364 | 238,570.89 | 5,157.88 |
| 6 | INDIANA | 6,483,802 | 333,257.31 | 5,139.84 |
| 7 | MISSISSIPPI | 2,967,297 | 152,180.78 | 5,128.60 |
| 8 | MAINE | 1,328,361 | 66,350.27 | 4,994.90 |
| 9 | TENNESSEE | 6,346,105 | 287,959.13 | 4,537.57 |

DRUG CODE:1205 DRUG NAME:LISDEXAMFETAMINE

| RANK | STATE | 2010 POP | TOTAL GRAMS | GRAMS/100K POP |
|---|---|---|---|---|
| 10 | IOWA | 3,046,355 | 137,724.81 | 4,520.97 |
| 11 | MISSOURI | 5,988,927 | 268,496.23 | 4,483.21 |
| 12 | WEST VIRGINIA | 1,852,994 | 81,126.51 | 4,378.13 |
| 13 | GEORGIA | 9,687,653 | 420,101.80 | 4,336.47 |
| 14 | TEXAS | 25,145,561 | 1,086,524.50 | 4,320.94 |
| 15 | KANSAS | 2,853,118 | 122,528.30 | 4,294.54 |
| 16 | WISCONSIN | 5,686,986 | 239,626.50 | 4,213.59 |
| 17 | ARKANSAS | 2,915,918 | 122,860.70 | 4,213.45 |
| 18 | NORTH CAROLINA | 9,535,483 | 396,793.85 | 4,161.23 |
| 19 | VERMONT | 625,741 | 25,616.77 | 4,093.83 |
| 20 | NEW HAMPSHIRE | 1,316,470 | 53,217.57 | 4,042.44 |
| 21 | OHIO | 11,536,504 | 456,526.40 | 3,957.23 |
| 22 | CONNECTICUT | 3,574,097 | 140,995.12 | 3,944.92 |
| 23 | KENTUCKY | 4,339,367 | 169,563.56 | 3,907.56 |
| 24 | OKLAHOMA | 3,751,351 | 140,533.57 | 3,746.21 |
| 25 | MARYLAND | 5,773,552 | 214,921.41 | 3,722.52 |
| 26 | NEBRASKA | 1,826,341 | 63,394.45 | 3,471.12 |
| 27 | VIRGINIA | 8,001,024 | 269,174.77 | 3,364.25 |
| 28 | NORTH DAKOTA | 672,591 | 22,148.17 | 3,292.96 |
| 29 | DISTRICT OF COLUMBIA | 601,723 | 19,313.74 | 3,209.74 |
| 30 | FLORIDA | 18,801,310 | 554,090.29 | 2,947.08 |
| 31 | UTAH | 2,763,885 | 80,203.28 | 2,901.83 |
| 32 | RHODE ISLAND | 1,052,567 | 29,391.76 | 2,792.39 |
| 33 | PENNSYLVANIA | 12,702,379 | 327,734.46 | 2,580.10 |
| 34 | MINNESOTA | 5,303,925 | 127,375.66 | 2,401.54 |
| 35 | MICHIGAN | 9,883,640 | 233,197.45 | 2,359.43 |
| 36 | ILLINOIS | 12,830,632 | 284,371.44 | 2,216.35 |
| 37 | NEW JERSEY | 8,791,894 | 194,097.65 | 2,207.69 |
| 38 | MONTANA | 989,415 | 21,429.59 | 2,165.88 |
| 39 | COLORADO | 5,029,196 | 107,789.06 | 2,143.27 |
| 40 | IDAHO | 1,567,582 | 33,347.70 | 2,127.33 |
| 41 | ARIZONA | 6,392,017 | 135,862.17 | 2,125.50 |
| 42 | MASSACHUSETTS | 6,547,629 | 134,425.59 | 2,053.04 |
| 43 | WASHINGTON | 6,724,540 | 127,363.52 | 1,894.01 |
| 44 | WYOMING | 563,626 | 10,674.62 | 1,893.92 |
| 45 | NEW YORK | 19,378,102 | 325,007.68 | 1,677.19 |
| 46 | OREGON | 3,831,074 | 58,175.94 | 1,518.53 |
| 47 | ALASKA | 710,231 | 9,784.34 | 1,377.63 |
| 48 | NEW MEXICO | 2,059,179 | 27,235.45 | 1,322.64 |
| 49 | CALIFORNIA | 37,253,956 | 433,684.26 | 1,164.13 |
| 50 | NEVADA | 2,700,551 | 28,580.09 | 1,058.31 |

| | | | | |
|---|---|---|---|---|
| 51 | HAWAII | 1,360,301 | 5,188.45 | 381.42 |
| 52 | VIRGIN ISLANDS | 109,825 | 295.41 | 268.98 |
| 53 | PUERTO RICO | 3,725,789 | 7,034.32 | 188.8 |
| 54 | GUAM | 178,430 | 93.07 | 52.16 |

| | | | |
|---|---|---|---|
| U.S. GRAMS / PER 100K: | 312,825,210 | 9,601,534.91 | 3,069.30 |

DRUG CODE:1724 DRUG NAME:METHYLPHENIDATE (DL;D;L;ISOMERS)

| RANK | STATE | 2010 POP | TOTAL GRAMS | GRAMS/100K POP |
|---|---|---|---|---|
| 1 | VERMONT | 625,741 | 75,723.80 | 12,101.46 |
| 2 | SOUTH DAKOTA | 814,180 | 89,485.74 | 10,990.90 |
| 3 | IOWA | 3,046,355 | 334,251.90 | 10,972.19 |
| 4 | MAINE | 1,328,361 | 145,027.42 | 10,917.77 |
| 5 | NEW HAMPSHIRE | 1,316,470 | 137,329.59 | 10,431.65 |
| 6 | MASSACHUSETTS | 6,547,629 | 596,026.91 | 9,102.94 |
| 7 | NORTH DAKOTA | 672,591 | 61,062.77 | 9,078.74 |
| 8 | MISSOURI | 5,988,927 | 527,861.01 | 8,813.95 |
| 9 | DELAWARE | 897,934 | 76,549.84 | 8,525.11 |
| 10 | NORTH CAROLINA | 9,535,483 | 777,538.58 | 8,154.16 |
| 11 | OHIO | 11,536,504 | 927,139.90 | 8,036.58 |

DRUG CODE:1724 DRUG NAME:METHYLPHENIDATE (DL;D;L;ISOMERS)

| RANK | STATE | 2010 POP | TOTAL GRAMS | GRAMS/100K POP |
|---|---|---|---|---|
| 12 | SOUTH CAROLINA | 4,625,364 | 358,727.46 | 7,755.66 |
| 13 | MICHIGAN | 9,883,640 | 762,509.88 | 7,714.87 |
| 14 | LOUISIANA | 4,533,372 | 345,868.92 | 7,629.40 |
| 15 | MINNESOTA | 5,303,925 | 404,467.88 | 7,625.82 |
| 16 | INDIANA | 6,483,802 | 493,828.86 | 7,616.35 |
| 17 | RHODE ISLAND | 1,052,567 | 77,473.19 | 7,360.40 |
| 18 | WISCONSIN | 5,686,986 | 415,929.01 | 7,313.70 |
| 19 | KANSAS | 2,853,118 | 205,391.66 | 7,198.85 |
| 20 | PENNSYLVANIA | 12,702,379 | 909,612.38 | 7,160.96 |
| 21 | IDAHO | 1,567,582 | 112,026.60 | 7,146.46 |
| 22 | MONTANA | 989,415 | 70,692.09 | 7,144.84 |
| 23 | KENTUCKY | 4,339,367 | 306,247.94 | 7,057.43 |
| 24 | MARYLAND | 5,773,552 | 407,005.63 | 7,049.48 |
| 25 | NEBRASKA | 1,826,341 | 127,616.76 | 6,987.56 |
| 26 | WEST VIRGINIA | 1,852,994 | 129,303.15 | 6,978.07 |
| 27 | GEORGIA | 9,687,653 | 664,882.60 | 6,863.20 |
| 28 | VIRGINIA | 8,001,024 | 542,060.99 | 6,774.90 |
| 29 | UTAH | 2,763,885 | 185,489.22 | 6,711.18 |
| 30 | WASHINGTON | 6,724,540 | 443,237.96 | 6,591.35 |
| 31 | OREGON | 3,831,074 | 251,436.54 | 6,563.08 |
| 32 | CONNECTICUT | 3,574,097 | 222,284.98 | 6,219.33 |
| 33 | ALABAMA | 4,779,736 | 290,860.23 | 6,085.28 |
| 34 | WYOMING | 563,626 | 33,923.06 | 6,018.72 |
| 35 | MISSISSIPPI | 2,967,297 | 176,702.31 | 5,954.99 |
| 36 | ARKANSAS | 2,915,918 | 165,258.11 | 5,667.45 |
| 37 | ILLINOIS | 12,830,632 | 716,031.14 | 5,580.64 |
| 38 | DISTRICT OF COLUMBIA | 601,723 | 32,847.30 | 5,458.87 |
| 39 | TENNESSEE | 6,346,105 | 344,590.08 | 5,429.95 |
| 40 | TEXAS | 25,145,561 | 1,353,418.17 | 5,382.33 |
| 41 | OKLAHOMA | 3,751,351 | 198,831.22 | 5,300.26 |
| 42 | COLORADO | 5,029,196 | 245,024.10 | 4,872.03 |
| 43 | NEW YORK | 19,378,102 | 932,467.39 | 4,811.96 |
| 44 | NEW MEXICO | 2,059,179 | 95,308.90 | 4,628.49 |

| | | | | |
|---|---|---|---|---|
| 45 | ALASKA | 710,231 | 32,818.59 | 4,620.83 |
| 46 | ARIZONA | 6,392,017 | 294,415.69 | 4,605.99 |
| 47 | FLORIDA | 18,801,310 | 833,042.66 | 4,430.77 |
| 48 | NEW JERSEY | 8,791,894 | 369,301.28 | 4,200.47 |
| 49 | HAWAII | 1,360,301 | 46,162.97 | 3,393.59 |
| 50 | CALIFORNIA | 37,253,956 | 1,107,130.68 | 2,971.85 |
| 51 | PUERTO RICO | 3,725,789 | 86,725.09 | 2,327.70 |
| 52 | NEVADA | 2,700,551 | 57,915.97 | 2,144.60 |
| 53 | GUAM | 178,430 | 1,845.31 | 1,034.19 |
| 54 | VIRGIN ISLANDS | 109,825 | 799.27 | 727.77 |
| 55 | AMERICAN SAMOA | 65,628 | 14.7 | 22.4 |
| U.S. GRAMS / PER 100K: | | 312,825,210 | 18,599,525.40 | 5,945.66 |

DRUG CODE:2012 DRUG NAME:

| RANK | STATE | 2010 POP | TOTAL GRAMS | GRAMS/100K POP |
|---|---|---|---|---|
| 1 | MISSOURI | 5,988,927 | 25,314,852.67 | 422,694.29 |
| 2 | SOUTH CAROLINA | 4,625,364 | 8,305.54 | 179.57 |
| 3 | VIRGINIA | 8,001,024 | 7,281.79 | 91.01 |
| 4 | NEW YORK | 19,378,102 | 12,928.90 | 66.72 |
| 5 | OHIO | 11,536,504 | 6,464.45 | 56.03 |
| 6 | MASSACHUSETTS | 6,547,629 | 3,195.07 | 48.8 |
| 7 | WASHINGTON | 6,724,540 | 3,269.38 | 48.62 |
| 8 | NORTH CAROLINA | 9,535,483 | 4,458.24 | 46.75 |
| 9 | TEXAS | 25,145,561 | 10,179.65 | 40.48 |
| 10 | TENNESSEE | 6,346,105 | 2,303.42 | 36.3 |
| 11 | CALIFORNIA | 37,253,956 | 12,854.59 | 34.51 |
| 12 | GEORGIA | 9,687,653 | 2,600.64 | 26.84 |

DRUG CODE:2012 DRUG NAME:GAMMA HYDROXYBUTYRIC ACID(FDA APPROVED- CSA III)

| RANK | STATE | 2010 POP | TOTAL GRAMS | GRAMS/100K POP |
|---|---|---|---|---|
| 13 | ILLINOIS | 12,830,632 | 3,046.46 | 23.74 |
| 14 | LOUISIANA | 4,533,372 | 743.04 | 16.39 |
| 15 | NEBRASKA | 1,826,341 | 222.91 | 12.21 |
| 16 | ALABAMA | 4,779,736 | 371.52 | 7.77 |
| 17 | MICHIGAN | 9,883,640 | 297.22 | 3.01 |
| 18 | FLORIDA | 18,801,310 | 148.61 | 0.79 |
| U.S. GRAMS / PER 100K: | | 312,825,210 | 25,393,524.10 | 8,117.48 |

DRUG CODE:2100 DRUG NAME:BARBITURIC ACID DERIVIATIVE OR SALT [PER 21CFR 1308.13(C)(3)]

| RANK | STATE | 2010 POP | TOTAL GRAMS | GRAMS/100K POP |
|---|---|---|---|---|
| 1 | NEW JERSEY | 8,791,894 | 1,090 | 12.4 |
| 2 | ARIZONA | 6,392,017 | 580 | 9.07 |
| 3 | ALASKA | 710,231 | 60 | 8.45 |
| 4 | RHODE ISLAND | 1,052,567 | 85 | 8.08 |
| 5 | DISTRICT OF COLUMBIA | 601,723 | 35 | 5.82 |
| 6 | NEW YORK | 19,378,102 | 707 | 3.65 |
| 7 | VIRGINIA | 8,001,024 | 290 | 3.62 |
| 8 | GEORGIA | 9,687,653 | 335 | 3.46 |
| 9 | NEW HAMPSHIRE | 1,316,470 | 45 | 3.42 |
| 10 | KANSAS | 2,853,118 | 95 | 3.33 |
| 11 | CONNECTICUT | 3,574,097 | 115 | 3.22 |

| | | | | |
|---|---|---|---|---|
| 12 | NORTH CAROLINA | 9,535,483 | 300 | 3.15 |
| 13 | CALIFORNIA | 37,253,956 | 1,098 | 2.95 |
| 14 | MICHIGAN | 9,883,640 | 275 | 2.78 |
| 15 | INDIANA | 6,483,802 | 175 | 2.7 |
| 16 | TEXAS | 25,145,561 | 645 | 2.57 |
| 17 | PENNSYLVANIA | 12,702,379 | 325 | 2.56 |
| 18 | MONTANA | 989,415 | 25 | 2.53 |
| 19 | COLORADO | 5,029,196 | 120 | 2.39 |
| 20 | OHIO | 11,536,504 | 275 | 2.38 |
| 21 | WEST VIRGINIA | 1,852,994 | 40 | 2.16 |
| 22 | ILLINOIS | 12,830,632 | 270 | 2.1 |
| 23 | KENTUCKY | 4,339,367 | 85 | 1.96 |
| 24 | MAINE | 1,328,361 | 25 | 1.88 |
| 25 | MISSOURI | 5,988,927 | 110 | 1.84 |
| 26 | MARYLAND | 5,773,552 | 105 | 1.82 |
| 27 | OKLAHOMA | 3,751,351 | 65 | 1.73 |
| 28 | FLORIDA | 18,801,310 | 315 | 1.68 |
| 29 | DELAWARE | 897,934 | 15 | 1.67 |
| 30 | NEVADA | 2,700,551 | 45 | 1.67 |
| 31 | LOUISIANA | 4,533,372 | 60 | 1.32 |
| 32 | MASSACHUSETTS | 6,547,629 | 80 | 1.22 |
| 33 | MISSISSIPPI | 2,967,297 | 35 | 1.18 |
| 34 | MINNESOTA | 5,303,925 | 50 | 0.94 |
| 35 | ALABAMA | 4,779,736 | 45 | 0.94 |
| 36 | VERMONT | 625,741 | 5 | 0.8 |
| 37 | TENNESSEE | 6,346,105 | 45 | 0.71 |
| 38 | SOUTH CAROLINA | 4,625,364 | 30 | 0.65 |
| 39 | OREGON | 3,831,074 | 25 | 0.65 |
| 40 | IDAHO | 1,567,582 | 10 | 0.64 |
| 41 | WASHINGTON | 6,724,540 | 35 | 0.52 |
| 42 | NEW MEXICO | 2,059,179 | 10 | 0.49 |
| 43 | WISCONSIN | 5,686,986 | 20 | 0.35 |
| 44 | ARKANSAS | 2,915,918 | 5 | 0.17 |
| U.S. GRAMS / PER 100K: | | 312,825,210 | 8,205 | 2.62 |

DRUG CODE:2125 DRUG NAME:AMOBARBITAL (SCHEDULE 2)

| RANK | STATE | 2010 POP | TOTAL GRAMS | GRAMS/100K POP |
|---|---|---|---|---|
| 1 | COLORADO | 5,029,196 | 66.08 | 1.31 |
| 2 | NEW HAMPSHIRE | 1,316,470 | 10.94 | 0.83 |
| 3 | UTAH | 2,763,885 | 22.33 | 0.81 |

DRUG CODE:2125 DRUG NAME:AMOBARBITAL (SCHEDULE 2)

| RANK | STATE | 2010 POP | TOTAL GRAMS | GRAMS/100K POP |
|---|---|---|---|---|
| 4 | NEW YORK | 19,378,102 | 152.2 | 0.79 |
| 5 | WASHINGTON | 6,724,540 | 48.76 | 0.73 |
| 6 | NEBRASKA | 1,826,341 | 12.3 | 0.67 |
| 7 | KANSAS | 2,853,118 | 17.77 | 0.62 |
| 8 | ARIZONA | 6,392,017 | 34.18 | 0.53 |
| 9 | WISCONSIN | 5,686,986 | 27.8 | 0.49 |
| 10 | NEW JERSEY | 8,791,894 | 40.1 | 0.46 |
| 11 | SOUTH CAROLINA | 4,625,364 | 20.96 | 0.45 |
| 12 | MINNESOTA | 5,303,925 | 23.7 | 0.45 |
| 13 | MASSACHUSETTS | 6,547,629 | 29.62 | 0.45 |
| 14 | NORTH CAROLINA | 9,535,483 | 40.1 | 0.42 |
| 15 | DISTRICT OF COLUMBIA | 601,723 | 2.28 | 0.38 |

| | | | | |
|---|---|---|---|---|
| 16 | GEORGIA | 9,687,653 | 34.63 | 0.36 |
| 17 | MARYLAND | 5,773,552 | 20.05 | 0.35 |
| 18 | KENTUCKY | 4,339,367 | 15.04 | 0.35 |
| 19 | PENNSYLVANIA | 12,702,379 | 42.84 | 0.34 |
| 20 | WEST VIRGINIA | 1,852,994 | 5.47 | 0.3 |
| 21 | VERMONT | 625,741 | 1.82 | 0.29 |
| 22 | TEXAS | 25,145,561 | 72.46 | 0.29 |
| 23 | MISSOURI | 5,988,927 | 16.86 | 0.28 |
| 24 | CALIFORNIA | 37,253,956 | 102.99 | 0.28 |
| 25 | MAINE | 1,328,361 | 3.65 | 0.27 |
| 26 | OREGON | 3,831,074 | 10.48 | 0.27 |
| 27 | ILLINOIS | 12,830,632 | 30.53 | 0.24 |
| 28 | OHIO | 11,536,504 | 25.52 | 0.22 |
| 29 | MICHIGAN | 9,883,640 | 19.6 | 0.2 |
| 30 | MISSISSIPPI | 2,967,297 | 5.92 | 0.2 |
| 31 | TENNESSEE | 6,346,105 | 10.94 | 0.17 |
| 32 | VIRGINIA | 8,001,024 | 12.76 | 0.16 |
| 33 | FLORIDA | 18,801,310 | 28.71 | 0.15 |
| 34 | NEW MEXICO | 2,059,179 | 2.73 | 0.13 |
| 35 | NEVADA | 2,700,551 | 3.19 | 0.12 |
| 36 | INDIANA | 6,483,802 | 7.75 | 0.12 |
| 37 | ALABAMA | 4,779,736 | 5.47 | 0.11 |
| 38 | CONNECTICUT | 3,574,097 | 0.91 | 0.03 |
| 39 | IDAHO | 1,567,582 | 0.46 | 0.03 |
| 40 | PUERTO RICO | 3,725,789 | 0.91 | 0.02 |
| 41 | ARKANSAS | 2,915,918 | 0.46 | 0.02 |
| 42 | IOWA | 3,046,355 | 0.46 | 0.01 |
| 43 | OKLAHOMA | 3,751,351 | 0.46 | 0.01 |
| U.S. GRAMS / PER 100K: | | 312,825,210 | 1,032.16 | 0.33 |

DRUG CODE:2165 DRUG NAME:BUTALBITAL

| RANK | STATE | 2010 POP | TOTAL GRAMS | GRAMS/100K POP |
|---|---|---|---|---|
| 1 | NEVADA | 2,700,551 | 35,170 | 1,302.33 |
| 2 | OKLAHOMA | 3,751,351 | 34,999.95 | 933 |
| 3 | FLORIDA | 18,801,310 | 150,170 | 798.72 |
| 4 | ARIZONA | 6,392,017 | 47,330 | 740.45 |
| 5 | LOUISIANA | 4,533,372 | 32,185 | 709.96 |
| 6 | UTAH | 2,763,885 | 19,500 | 705.53 |
| 7 | MISSOURI | 5,988,927 | 42,165 | 704.05 |
| 8 | CONNECTICUT | 3,574,097 | 24,660 | 689.96 |
| 9 | ARKANSAS | 2,915,918 | 19,685 | 675.09 |
| 10 | MICHIGAN | 9,883,640 | 66,185 | 669.64 |
| 11 | GEORGIA | 9,687,653 | 63,850 | 659.09 |
| 12 | MONTANA | 989,415 | 6,400 | 646.85 |
| 13 | WYOMING | 563,626 | 3,530 | 626.3 |
| 14 | PENNSYLVANIA | 12,702,379 | 79,000 | 621.93 |
| 15 | ALABAMA | 4,779,736 | 28,435 | 594.91 |
| 16 | HAWAII | 1,360,301 | 7,920 | 582.22 |

DRUG CODE:2165 DRUG NAME:BUTALBITAL

| RANK | STATE | 2010 POP | TOTAL GRAMS | GRAMS/100K POP |
|---|---|---|---|---|
| 17 | MARYLAND | 5,773,552 | 32,375 | 560.75 |
| 18 | NEW JERSEY | 8,791,894 | 47,820 | 543.91 |
| 19 | KANSAS | 2,853,118 | 15,455 | 541.69 |
| 20 | TEXAS | 25,145,561 | 133,619.85 | 531.39 |

| | | | | |
|---|---|---|---|---|
| 21 | SOUTH CAROLINA | 4,625,364 | 24,090 | 520.82 |
| 22 | IDAHO | 1,567,582 | 8,130 | 518.63 |
| 23 | VIRGINIA | 8,001,024 | 40,450 | 505.56 |
| 24 | SOUTH DAKOTA | 814,180 | 3,965 | 486.99 |
| 25 | ILLINOIS | 12,830,632 | 62,170 | 484.54 |
| 26 | CALIFORNIA | 37,253,956 | 176,515 | 473.82 |
| 27 | INDIANA | 6,483,802 | 30,320 | 467.63 |
| 28 | TENNESSEE | 6,346,105 | 29,190 | 459.97 |
| 29 | NEBRASKA | 1,826,341 | 8,290 | 453.91 |
| 30 | MISSISSIPPI | 2,967,297 | 13,415 | 452.09 |
| 31 | DISTRICT OF COLUMBIA | 601,723 | 2,665 | 442.89 |
| 32 | WASHINGTON | 6,724,540 | 29,545 | 439.36 |
| 33 | KENTUCKY | 4,339,367 | 18,745 | 431.98 |
| 34 | OHIO | 11,536,504 | 49,645 | 430.33 |
| 35 | NEW MEXICO | 2,059,179 | 8,835 | 429.05 |
| 36 | DELAWARE | 897,934 | 3,805 | 423.75 |
| 37 | OREGON | 3,831,074 | 16,060 | 419.2 |
| 38 | MAINE | 1,328,361 | 5,385 | 405.39 |
| 39 | WEST VIRGINIA | 1,852,994 | 7,465 | 402.86 |
| 40 | NORTH CAROLINA | 9,535,483 | 38,165 | 400.24 |
| 41 | RHODE ISLAND | 1,052,567 | 4,190 | 398.07 |
| 42 | NEW HAMPSHIRE | 1,316,470 | 5,190 | 394.24 |
| 43 | MASSACHUSETTS | 6,547,629 | 22,490 | 343.48 |
| 44 | COLORADO | 5,029,196 | 16,490 | 327.89 |
| 45 | WISCONSIN | 5,686,986 | 16,590 | 291.72 |
| 46 | IOWA | 3,046,355 | 8,620 | 282.96 |
| 47 | MINNESOTA | 5,303,925 | 14,575 | 274.8 |
| 48 | NORTH DAKOTA | 672,591 | 1,690 | 251.27 |
| 49 | NEW YORK | 19,378,102 | 48,525 | 250.41 |
| 50 | ALASKA | 710,231 | 1,495 | 210.49 |
| 51 | VERMONT | 625,741 | 1,140 | 182.18 |
| 52 | VIRGIN ISLANDS | 109,825 | 115 | 104.71 |
| 53 | GUAM | 178,430 | 35 | 19.62 |
| 54 | PUERTO RICO | 3,725,789 | 680 | 18.25 |
| U.S. GRAMS / PER 100K: | | 312,825,210 | 1,609,129.80 | 514.39 |

DRUG CODE:2270 DRUG NAME:PENTOBARBITAL (SCHEDULE 2)

| RANK | STATE | 2010 POP | TOTAL GRAMS | GRAMS/100K POP |
|---|---|---|---|---|
| 1 | VERMONT | 625,741 | 44,119 | 7,050.68 |
| 2 | MONTANA | 989,415 | 69,172.25 | 6,991.23 |
| 3 | WYOMING | 563,626 | 38,840.68 | 6,891.21 |
| 4 | IDAHO | 1,567,582 | 91,515.91 | 5,838.03 |
| 5 | MAINE | 1,328,361 | 66,402.40 | 4,998.82 |
| 6 | NEW HAMPSHIRE | 1,316,470 | 62,326.47 | 4,734.36 |
| 7 | NEBRASKA | 1,826,341 | 82,609.30 | 4,523.21 |
| 8 | MICHIGAN | 9,883,640 | 369,758.25 | 3,741.11 |
| 9 | NORTH DAKOTA | 672,591 | 24,949.12 | 3,709.40 |
| 10 | NEW MEXICO | 2,059,179 | 68,904.12 | 3,346.19 |
| 11 | OREGON | 3,831,074 | 126,235.24 | 3,295.04 |
| 12 | WASHINGTON | 6,724,540 | 212,826.14 | 3,164.92 |
| 13 | SOUTH DAKOTA | 814,180 | 25,749.78 | 3,162.66 |
| 14 | KANSAS | 2,853,118 | 84,142.91 | 2,949.16 |
| 15 | ARIZONA | 6,392,017 | 185,674.98 | 2,904.79 |
| 16 | WISCONSIN | 5,686,986 | 163,264.48 | 2,870.84 |
| 17 | RHODE ISLAND | 1,052,567 | 30,139.72 | 2,863.45 |
| 18 | COLORADO | 5,029,196 | 142,769.15 | 2,838.81 |

DRUG CODE:2270 DRUG NAME:PENTOBARBITAL (SCHEDULE 2)

| RANK | STATE | 2010 POP | TOTAL GRAMS | GRAMS/100K POP |
|---|---|---|---|---|
| 19 | HAWAII | 1,360,301 | 38,131.15 | 2,803.14 |
| 20 | KENTUCKY | 4,339,367 | 109,650.01 | 2,526.87 |
| 21 | ALASKA | 710,231 | 17,939.54 | 2,525.87 |
| 22 | NORTH CAROLINA | 9,535,483 | 238,691.10 | 2,503.19 |
| 23 | MISSOURI | 5,988,927 | 144,631.15 | 2,414.98 |
| 24 | IOWA | 3,046,355 | 73,377.80 | 2,408.71 |
| 25 | INDIANA | 6,483,802 | 152,911.28 | 2,358.36 |
| 26 | TENNESSEE | 6,346,105 | 146,187.12 | 2,303.57 |
| 27 | NEVADA | 2,700,551 | 61,155.68 | 2,264.56 |
| 28 | MINNESOTA | 5,303,925 | 118,726.28 | 2,238.46 |
| 29 | WEST VIRGINIA | 1,852,994 | 41,290.82 | 2,228.33 |
| 30 | TEXAS | 25,145,561 | 559,981.86 | 2,226.96 |
| 31 | OHIO | 11,536,504 | 256,365.01 | 2,222.21 |
| 32 | VIRGINIA | 8,001,024 | 176,825.95 | 2,210.04 |
| 33 | PENNSYLVANIA | 12,702,379 | 254,583.88 | 2,004.22 |
| 34 | ILLINOIS | 12,830,632 | 256,481.24 | 1,998.98 |
| 35 | LOUISIANA | 4,533,372 | 90,391.44 | 1,993.91 |
| 36 | CALIFORNIA | 37,253,956 | 679,656.90 | 1,824.39 |
| 37 | GUAM | 178,430 | 3,215.42 | 1,802.06 |
| 38 | OKLAHOMA | 3,751,351 | 67,526.78 | 1,800.07 |
| 39 | CONNECTICUT | 3,574,097 | 61,529.25 | 1,721.53 |
| 40 | MASSACHUSETTS | 6,547,629 | 105,920.58 | 1,617.69 |
| 41 | UTAH | 2,763,885 | 44,645.27 | 1,615.31 |
| 42 | ALABAMA | 4,779,736 | 76,713.06 | 1,604.96 |
| 43 | FLORIDA | 18,801,310 | 275,739.17 | 1,466.60 |
| 44 | NEW YORK | 19,378,102 | 283,048.28 | 1,460.66 |
| 45 | GEORGIA | 9,687,653 | 136,590.99 | 1,409.95 |
| 46 | SOUTH CAROLINA | 4,625,364 | 60,628.59 | 1,310.79 |
| 47 | MARYLAND | 5,773,552 | 72,074.88 | 1,248.36 |
| 48 | ARKANSAS | 2,915,918 | 30,949.10 | 1,061.38 |
| 49 | NEW JERSEY | 8,791,894 | 82,658.41 | 940.17 |
| 50 | DELAWARE | 897,934 | 7,327.89 | 816.08 |
| 51 | MISSISSIPPI | 2,967,297 | 22,930.04 | 772.76 |
| 52 | DISTRICT OF COLUMBIA | 601,723 | 893.17 | 148.44 |
| 53 | PUERTO RICO | 3,725,789 | 4,896.50 | 131.42 |
| U.S. GRAMS / PER 100K: | | 312,825,210 | 6,643,665.49 | 2,123.76 |

DRUG CODE:2315 DRUG NAME:SECOBARBITAL (SCHEDULE 2)

| RANK | STATE | 2010 POP | TOTAL GRAMS | GRAMS/100K POP |
|---|---|---|---|---|
| 1 | OREGON | 3,831,074 | 1,034.52 | 27 |
| 2 | VERMONT | 625,741 | 109.86 | 17.56 |
| 3 | WASHINGTON | 6,724,540 | 814.8 | 12.12 |
| 4 | NEW YORK | 19,378,102 | 1,867.62 | 9.64 |
| 5 | CALIFORNIA | 37,253,956 | 2,810.59 | 7.54 |
| 6 | HAWAII | 1,360,301 | 64.09 | 4.71 |
| 7 | RHODE ISLAND | 1,052,567 | 45.78 | 4.35 |
| 8 | NEW JERSEY | 8,791,894 | 320.43 | 3.64 |
| 9 | MASSACHUSETTS | 6,547,629 | 219.72 | 3.36 |
| 10 | WYOMING | 563,626 | 18.31 | 3.25 |
| 11 | MARYLAND | 5,773,552 | 183.1 | 3.17 |
| 12 | NEW HAMPSHIRE | 1,316,470 | 36.62 | 2.78 |
| 13 | KANSAS | 2,853,118 | 73.24 | 2.57 |
| 14 | ILLINOIS | 12,830,632 | 320.43 | 2.5 |

| | | | | |
|---|---|---|---|---|
| 15 | MISSOURI | 5,988,927 | 146.48 | 2.45 |
| 16 | PENNSYLVANIA | 12,702,379 | 283.81 | 2.23 |
| 17 | NEW MEXICO | 2,059,179 | 36.62 | 1.78 |
| 18 | OHIO | 11,536,504 | 201.41 | 1.75 |
| 19 | FLORIDA | 18,801,310 | 311.27 | 1.66 |
| 20 | VIRGINIA | 8,001,024 | 128.17 | 1.6 |
| 21 | CONNECTICUT | 3,574,097 | 45.78 | 1.28 |

DRUG CODE:2315 DRUG NAME:SECOBARBITAL (SCHEDULE 2)

| RANK | STATE | 2010 POP | TOTAL GRAMS | GRAMS/100K POP |
|---|---|---|---|---|
| 22 | LOUISIANA | 4,533,372 | 54.93 | 1.21 |
| 23 | COLORADO | 5,029,196 | 54.93 | 1.09 |
| 24 | DELAWARE | 897,934 | 9.16 | 1.02 |
| 25 | NORTH CAROLINA | 9,535,483 | 82.4 | 0.86 |
| 26 | GEORGIA | 9,687,653 | 64.09 | 0.66 |
| 27 | ARKANSAS | 2,915,918 | 18.31 | 0.63 |
| 28 | ARIZONA | 6,392,017 | 36.62 | 0.57 |
| 29 | WEST VIRGINIA | 1,852,994 | 9.16 | 0.49 |
| 30 | NEVADA | 2,700,551 | 9.16 | 0.34 |
| 31 | TEXAS | 25,145,561 | 73.24 | 0.29 |
| 32 | MICHIGAN | 9,883,640 | 18.31 | 0.19 |
| U.S. GRAMS / PER 100K: | | 312,825,210 | 9,502.89 | 3.04 |

DRUG CODE:7379 DRUG NAME:NABILONE

| RANK | STATE | 2010 POP | TOTAL GRAMS | GRAMS/100K POP |
|---|---|---|---|---|
| 1 | MARYLAND | 5,773,552 | 5.35 | 0.09 |
| 2 | NEBRASKA | 1,826,341 | 0.7 | 0.04 |
| 3 | COLORADO | 5,029,196 | 1.05 | 0.02 |
| 4 | FLORIDA | 18,801,310 | 3.5 | 0.02 |
| 5 | NEW HAMPSHIRE | 1,316,470 | 0.25 | 0.02 |
| 6 | NEW YORK | 19,378,102 | 1.15 | 0.01 |
| 7 | OHIO | 11,536,504 | 0.75 | 0.01 |
| 8 | MAINE | 1,328,361 | 0.1 | 0.01 |
| 9 | VIRGINIA | 8,001,024 | 1 | 0.01 |
| 10 | MISSOURI | 5,988,927 | 0.45 | 0.01 |
| 11 | NEW MEXICO | 2,059,179 | 0.3 | 0.01 |
| 12 | HAWAII | 1,360,301 | 0.2 | 0.01 |
| 13 | MINNESOTA | 5,303,925 | 0.3 | 0.01 |
| 14 | PENNSYLVANIA | 12,702,379 | 1.05 | 0.01 |
| 15 | NORTH CAROLINA | 9,535,483 | 0.55 | 0.01 |
| 16 | MASSACHUSETTS | 6,547,629 | 0.9 | 0.01 |
| 17 | ILLINOIS | 12,830,632 | 0.8 | 0.01 |
| 18 | IOWA | 3,046,355 | 0.05 | 0 |
| 19 | NEW JERSEY | 8,791,894 | 0.15 | 0 |
| 20 | ARIZONA | 6,392,017 | 0.15 | 0 |
| 21 | CALIFORNIA | 37,253,956 | 0.05 | 0 |
| 22 | RHODE ISLAND | 1,052,567 | 0.05 | 0 |
| 23 | INDIANA | 6,483,802 | 0.3 | 0 |
| 24 | WISCONSIN | 5,686,986 | 0.05 | 0 |
| U.S. GRAMS / PER 100K: | | 312,825,210 | 19.2 | 0.01 |

DRUG CODE:7471 DRUG NAME:PHENCYCLIDINE (PCP)

| RANK | STATE | 2010 POP | TOTAL GRAMS | GRAMS/100K POP |
|---|---|---|---|---|
| 1 | MINNESOTA | 5,303,925 | 0.22 | 0 |
| U.S. GRAMS / PER 100K: | | 312,825,210 | 0.22 | 0 |

CUMULATIVE DISTRIBUTION BY STATE IN GRAMS PER 100,000 POPULATION

DRUG CODE:9041L DRUG NAME:COCAINE

| RANK | STATE | 2010 POP | TOTAL GRAMS | GRAMS/100K POP |
|---|---|---|---|---|
| 1 | NORTH DAKOTA | 672,591 | 183.88 | 27.34 |
| 2 | MONTANA | 989,415 | 208.09 | 21.03 |
| 3 | SOUTH DAKOTA | 814,180 | 146.49 | 17.99 |
| 4 | UTAH | 2,763,885 | 461.58 | 16.7 |
| 5 | IDAHO | 1,567,582 | 261.8 | 16.7 |
| 6 | MINNESOTA | 5,303,925 | 883.98 | 16.67 |
| 7 | DISTRICT OF COLUMBIA | 601,723 | 97.42 | 16.19 |
| 8 | MICHIGAN | 9,883,640 | 1,583.18 | 16.02 |
| 9 | NEW HAMPSHIRE | 1,316,470 | 210.49 | 15.99 |
| 10 | NEW MEXICO | 2,059,179 | 312.94 | 15.2 |
| 11 | WYOMING | 563,626 | 79.03 | 14.02 |
| 12 | MASSACHUSETTS | 6,547,629 | 908.23 | 13.87 |
| 13 | PENNSYLVANIA | 12,702,379 | 1,737.17 | 13.68 |
| 14 | IOWA | 3,046,355 | 403.62 | 13.25 |

DRUG CODE:9041L DRUG NAME:COCAINE

| RANK | STATE | 2010 POP | TOTAL GRAMS | GRAMS/100K POP |
|---|---|---|---|---|
| 15 | MISSISSIPPI | 2,967,297 | 382.37 | 12.89 |
| 16 | KANSAS | 2,853,118 | 366.62 | 12.85 |
| 17 | INDIANA | 6,483,802 | 820.8 | 12.66 |
| 18 | TENNESSEE | 6,346,105 | 763.09 | 12.02 |
| 19 | MISSOURI | 5,988,927 | 711.31 | 11.88 |
| 20 | WISCONSIN | 5,686,986 | 629.17 | 11.06 |
| 21 | ILLINOIS | 12,830,632 | 1,402.45 | 10.93 |
| 22 | OREGON | 3,831,074 | 415.97 | 10.86 |
| 23 | WEST VIRGINIA | 1,852,994 | 197.13 | 10.64 |
| 24 | CALIFORNIA | 37,253,956 | 3,764.79 | 10.11 |
| 25 | KENTUCKY | 4,339,367 | 438.11 | 10.1 |
| 26 | MARYLAND | 5,773,552 | 559.43 | 9.69 |
| 27 | RHODE ISLAND | 1,052,567 | 96.24 | 9.14 |
| 28 | ALASKA | 710,231 | 63.28 | 8.91 |
| 29 | VIRGINIA | 8,001,024 | 704.99 | 8.81 |
| 30 | LOUISIANA | 4,533,372 | 385.9 | 8.51 |
| 31 | DELAWARE | 897,934 | 74.57 | 8.3 |
| 32 | NEBRASKA | 1,826,341 | 150.45 | 8.24 |
| 33 | MAINE | 1,328,361 | 108.49 | 8.17 |
| 34 | WASHINGTON | 6,724,540 | 545.75 | 8.12 |
| 35 | NEVADA | 2,700,551 | 217.13 | 8.04 |
| 36 | NEW JERSEY | 8,791,894 | 697.28 | 7.93 |
| 37 | ARIZONA | 6,392,017 | 503.43 | 7.88 |
| 38 | NEW YORK | 19,378,102 | 1,474.05 | 7.61 |
| 39 | GEORGIA | 9,687,653 | 730.45 | 7.54 |
| 40 | COLORADO | 5,029,196 | 375.94 | 7.48 |
| 41 | ARKANSAS | 2,915,918 | 215.99 | 7.41 |
| 42 | NORTH CAROLINA | 9,535,483 | 629.85 | 6.61 |
| 43 | FLORIDA | 18,801,310 | 1,237.89 | 6.58 |
| 44 | HAWAII | 1,360,301 | 88.42 | 6.5 |
| 45 | OHIO | 11,536,504 | 722.99 | 6.27 |
| 46 | TEXAS | 25,145,561 | 1,548.29 | 6.16 |
| 47 | ALABAMA | 4,779,736 | 247.98 | 5.19 |
| 48 | CONNECTICUT | 3,574,097 | 183.7 | 5.14 |
| 49 | VERMONT | 625,741 | 31.14 | 4.98 |
| 50 | OKLAHOMA | 3,751,351 | 154.2 | 4.11 |
| 51 | SOUTH CAROLINA | 4,625,364 | 141.81 | 3.07 |
| 52 | GUAM | 178,430 | 0.71 | 0.4 |
| U.S. GRAMS / PER 100K: | | 312,825,210 | 29,260.13 | 9.35 |

CUMULATIVE DISTRIBUTION BY STATE IN GRAMS PER 100,000 POPULATION

DRUG CODE:9050 DRUG NAME:CODEINE

| RANK | STATE | 2010 POP | TOTAL GRAMS | GRAMS/100K POP |
|---|---|---|---|---|
| 1 | TEXAS | 25,145,561 | 4,103,570.92 | 16,319.27 |
| 2 | MICHIGAN | 9,883,640 | 1,098,677.69 | 11,116.12 |
| 3 | ARKANSAS | 2,915,918 | 309,470.29 | 10,613.13 |
| 4 | OKLAHOMA | 3,751,351 | 382,186.69 | 10,187.97 |
| 5 | DISTRICT OF COLUMBIA | 601,723 | 53,930.33 | 8,962.65 |
| 6 | SOUTH CAROLINA | 4,625,364 | 319,735.11 | 6,912.65 |
| 7 | WEST VIRGINIA | 1,852,994 | 125,870.43 | 6,792.81 |
| 8 | TENNESSEE | 6,346,105 | 412,524.28 | 6,500.43 |
| 9 | CALIFORNIA | 37,253,956 | 2,143,388.13 | 5,753.45 |
| 10 | ILLINOIS | 12,830,632 | 692,788.92 | 5,399.49 |
| 11 | ARIZONA | 6,392,017 | 320,165.84 | 5,008.84 |
| 12 | MISSOURI | 5,988,927 | 297,602.43 | 4,969.21 |
| 13 | KENTUCKY | 4,339,367 | 211,658.36 | 4,877.63 |
| 14 | DELAWARE | 897,934 | 43,587.21 | 4,854.17 |
| 15 | PENNSYLVANIA | 12,702,379 | 611,528.12 | 4,814.28 |
| 16 | MONTANA | 989,415 | 45,727.83 | 4,621.70 |
| 17 | MARYLAND | 5,773,552 | 263,393.72 | 4,562.07 |
| 18 | ALABAMA | 4,779,736 | 217,592.79 | 4,552.40 |

DRUG CODE:9050 DRUG NAME:CODEINE

| RANK | STATE | 2010 POP | TOTAL GRAMS | GRAMS/100K POP |
|---|---|---|---|---|
| 19 | NEVADA | 2,700,551 | 117,993.75 | 4,369.25 |
| 20 | GEORGIA | 9,687,653 | 422,444.81 | 4,360.65 |
| 21 | HAWAII | 1,360,301 | 58,321.71 | 4,287.41 |
| 22 | KANSAS | 2,853,118 | 119,792.35 | 4,198.65 |
| 23 | ALASKA | 710,231 | 29,814.55 | 4,197.87 |
| 24 | NEW MEXICO | 2,059,179 | 84,324.07 | 4,095.03 |
| 25 | LOUISIANA | 4,533,372 | 182,092.95 | 4,016.72 |
| 26 | SOUTH DAKOTA | 814,180 | 32,565.25 | 3,999.76 |
| 27 | NEW JERSEY | 8,791,894 | 350,879.33 | 3,990.94 |
| 28 | WASHINGTON | 6,724,540 | 267,643.78 | 3,980.11 |
| 29 | OHIO | 11,536,504 | 457,095.73 | 3,962.17 |
| 30 | WISCONSIN | 5,686,986 | 221,701.48 | 3,898.40 |
| 31 | OREGON | 3,831,074 | 148,205.79 | 3,868.52 |
| 32 | MISSISSIPPI | 2,967,297 | 113,218.24 | 3,815.53 |
| 33 | IDAHO | 1,567,582 | 59,177.85 | 3,775.10 |
| 34 | MAINE | 1,328,361 | 49,772.28 | 3,746.89 |
| 35 | NORTH DAKOTA | 672,591 | 25,090.21 | 3,730.38 |
| 36 | FLORIDA | 18,801,310 | 697,741.62 | 3,711.13 |
| 37 | INDIANA | 6,483,802 | 234,081.64 | 3,610.25 |
| 38 | NEW YORK | 19,378,102 | 666,432.20 | 3,439.10 |
| 39 | WYOMING | 563,626 | 18,909.55 | 3,354.98 |
| 40 | AMERICAN SAMOA | 65,628 | 2,150.43 | 3,276.69 |
| 41 | RHODE ISLAND | 1,052,567 | 34,416.71 | 3,269.79 |
| 42 | MASSACHUSETTS | 6,547,629 | 212,699.29 | 3,248.49 |
| 43 | MINNESOTA | 5,303,925 | 169,780.02 | 3,201.03 |
| 44 | CONNECTICUT | 3,574,097 | 111,463.30 | 3,118.64 |
| 45 | VIRGINIA | 8,001,024 | 245,002.58 | 3,062.14 |
| 46 | GUAM | 178,430 | 5,430.96 | 3,043.75 |
| 47 | NEBRASKA | 1,826,341 | 54,517.49 | 2,985.07 |
| 48 | IOWA | 3,046,355 | 85,843.99 | 2,817.92 |
| 49 | NEW HAMPSHIRE | 1,316,470 | 35,440.19 | 2,692.06 |
| 50 | UTAH | 2,763,885 | 70,927.68 | 2,566.23 |

| | | | | |
|---|---|---|---|---|
| 51 | VIRGIN ISLANDS | 109,825 | 2,809.65 | 2,558.30 |
| 52 | VERMONT | 625,741 | 15,296.90 | 2,444.61 |
| 53 | COLORADO | 5,029,196 | 110,668.52 | 2,200.52 |
| 54 | NORTH CAROLINA | 9,535,483 | 208,174.55 | 2,183.16 |
| 55 | PUERTO RICO | 3,725,789 | 53,180.38 | 1,427.36 |
| U.S. GRAMS / PER 100K: | | 312,825,210 | 17,428,500.87 | 5,571.32 |

DRUG CODE:9064 DRUG NAME:BUPRENORPHINE

| RANK | STATE | 2010 POP | TOTAL GRAMS | GRAMS/100K POP |
|---|---|---|---|---|
| 1 | VERMONT | 625,741 | 21,538.21 | 3,442.03 |
| 2 | WEST VIRGINIA | 1,852,994 | 58,209.20 | 3,141.36 |
| 3 | KENTUCKY | 4,339,367 | 124,394.55 | 2,866.65 |
| 4 | RHODE ISLAND | 1,052,567 | 27,662.71 | 2,628.12 |
| 5 | MAINE | 1,328,361 | 31,829.46 | 2,396.15 |
| 6 | MASSACHUSETTS | 6,547,629 | 151,898.48 | 2,319.90 |
| 7 | NEW HAMPSHIRE | 1,316,470 | 28,287.51 | 2,148.74 |
| 8 | TENNESSEE | 6,346,105 | 125,421.64 | 1,976.36 |
| 9 | ALABAMA | 4,779,736 | 90,830.47 | 1,900.32 |
| 10 | PENNSYLVANIA | 12,702,379 | 224,153.12 | 1,764.65 |
| 11 | OHIO | 11,536,504 | 189,477.72 | 1,642.42 |
| 12 | MARYLAND | 5,773,552 | 91,431.46 | 1,583.63 |
| 13 | CONNECTICUT | 3,574,097 | 54,661.42 | 1,529.38 |
| 14 | LOUISIANA | 4,533,372 | 63,723.20 | 1,405.65 |
| 15 | DELAWARE | 897,934 | 11,666.55 | 1,299.27 |
| 16 | UTAH | 2,763,885 | 35,562.28 | 1,286.68 |
| 17 | MISSISSIPPI | 2,967,297 | 37,085.17 | 1,249.80 |
| 18 | NEW MEXICO | 2,059,179 | 25,426.25 | 1,234.78 |
| 19 | DISTRICT OF COLUMBIA | 601,723 | 7,382.17 | 1,226.84 |

DRUG CODE:9064 DRUG NAME:BUPRENORPHINE

| RANK | STATE | 2010 POP | TOTAL GRAMS | GRAMS/100K POP |
|---|---|---|---|---|
| 20 | NEW JERSEY | 8,791,894 | 100,665.32 | 1,144.98 |
| 21 | NORTH CAROLINA | 9,535,483 | 107,851.50 | 1,131.05 |
| 22 | ALASKA | 710,231 | 7,690.90 | 1,082.87 |
| 23 | OKLAHOMA | 3,751,351 | 37,960.01 | 1,011.90 |
| 24 | NEW YORK | 19,378,102 | 193,906.35 | 1,000.65 |
| 25 | MICHIGAN | 9,883,640 | 97,134.19 | 982.78 |
| 26 | INDIANA | 6,483,802 | 63,688.83 | 982.28 |
| 27 | WASHINGTON | 6,724,540 | 66,032.95 | 981.97 |
| 28 | VIRGINIA | 8,001,024 | 76,771.69 | 959.52 |
| 29 | FLORIDA | 18,801,310 | 172,436.44 | 917.15 |
| 30 | OREGON | 3,831,074 | 34,639.03 | 904.16 |
| 31 | SOUTH CAROLINA | 4,625,364 | 38,037.86 | 822.38 |
| 32 | IDAHO | 1,567,582 | 12,145.43 | 774.79 |
| 33 | MONTANA | 989,415 | 7,227.06 | 730.44 |
| 34 | WYOMING | 563,626 | 4,016.37 | 712.6 |
| 35 | ARKANSAS | 2,915,918 | 20,059.19 | 687.92 |
| 36 | WISCONSIN | 5,686,986 | 37,549.75 | 660.28 |
| 37 | GEORGIA | 9,687,653 | 57,536.95 | 593.92 |
| 38 | MISSOURI | 5,988,927 | 32,823.17 | 548.06 |
| 39 | COLORADO | 5,029,196 | 26,937.09 | 535.61 |
| 40 | HAWAII | 1,360,301 | 7,209.61 | 530 |
| 41 | ARIZONA | 6,392,017 | 33,127.63 | 518.27 |
| 42 | NORTH DAKOTA | 672,591 | 3,285.17 | 488.44 |
| 43 | MINNESOTA | 5,303,925 | 24,670.85 | 465.14 |

| | | | | |
|---|---|---|---|---|
| 44 | NEVADA | 2,700,551 | 12,325.29 | 456.4 |
| 45 | TEXAS | 25,145,561 | 101,033.10 | 401.79 |
| 46 | ILLINOIS | 12,830,632 | 47,612.52 | 371.08 |
| 47 | CALIFORNIA | 37,253,956 | 137,585 | 369.32 |
| 48 | KANSAS | 2,853,118 | 9,427.50 | 330.43 |
| 49 | PUERTO RICO | 3,725,789 | 9,968.08 | 267.54 |
| 50 | NEBRASKA | 1,826,341 | 4,659.52 | 255.13 |
| 51 | IOWA | 3,046,355 | 5,837.53 | 191.62 |
| 52 | SOUTH DAKOTA | 814,180 | 1,520.67 | 186.77 |
| 53 | VIRGIN ISLANDS | 109,825 | 88.96 | 81.01 |
| 54 | GUAM | 178,430 | 4.56 | 2.56 |
| 55 | AMERICAN SAMOA | 65,628 | 0.01 | 0.02 |

U.S. GRAMS / PER 100K: 312,825,210 2,994,107.63 957.12

DRUG CODE:9120 DRUG NAME:DIHYDROCODEINE

| RANK | STATE | 2010 POP | TOTAL GRAMS | GRAMS/100K POP |
|---|---|---|---|---|
| 1 | TENNESSEE | 6,346,105 | 4,243.56 | 66.87 |
| 2 | ALABAMA | 4,779,736 | 2,831.64 | 59.24 |
| 3 | LOUISIANA | 4,533,372 | 2,330.49 | 51.41 |
| 4 | TEXAS | 25,145,561 | 10,255.56 | 40.78 |
| 5 | MISSISSIPPI | 2,967,297 | 1,153.73 | 38.88 |
| 6 | NEW MEXICO | 2,059,179 | 616.33 | 29.93 |
| 7 | OKLAHOMA | 3,751,351 | 1,050.83 | 28.01 |
| 8 | GEORGIA | 9,687,653 | 1,614.19 | 16.66 |
| 9 | IDAHO | 1,567,582 | 213.1 | 13.59 |
| 10 | ARKANSAS | 2,915,918 | 389.73 | 13.37 |
| 11 | CALIFORNIA | 37,253,956 | 4,652.48 | 12.49 |
| 12 | PENNSYLVANIA | 12,702,379 | 1,131.65 | 8.91 |
| 13 | NEVADA | 2,700,551 | 202.63 | 7.5 |
| 14 | NEW JERSEY | 8,791,894 | 516.98 | 5.88 |
| 15 | ARIZONA | 6,392,017 | 221.96 | 3.47 |
| 16 | KENTUCKY | 4,339,367 | 120.92 | 2.79 |
| 17 | FLORIDA | 18,801,310 | 364.47 | 1.94 |
| 18 | HAWAII | 1,360,301 | 24.57 | 1.81 |
| 19 | WEST VIRGINIA | 1,852,994 | 31.4 | 1.69 |
| 20 | ILLINOIS | 12,830,632 | 213.91 | 1.67 |

DRUG CODE:9120 DRUG NAME:DIHYDROCODEINE

| RANK | STATE | 2010 POP | TOTAL GRAMS | GRAMS/100K POP |
|---|---|---|---|---|
| 21 | SOUTH CAROLINA | 4,625,364 | 61.53 | 1.33 |
| 22 | MISSOURI | 5,988,927 | 71.57 | 1.19 |
| 23 | OHIO | 11,536,504 | 134.8 | 1.17 |
| 24 | MICHIGAN | 9,883,640 | 108.95 | 1.1 |
| 25 | INDIANA | 6,483,802 | 68.26 | 1.05 |
| 26 | NEW YORK | 19,378,102 | 201.58 | 1.04 |
| 27 | MASSACHUSETTS | 6,547,629 | 51.27 | 0.78 |
| 28 | RHODE ISLAND | 1,052,567 | 6.41 | 0.61 |
| 29 | MARYLAND | 5,773,552 | 24.78 | 0.43 |
| 30 | VIRGINIA | 8,001,024 | 30.98 | 0.39 |
| 31 | NORTH CAROLINA | 9,535,483 | 36.53 | 0.38 |
| 32 | WISCONSIN | 5,686,986 | 21.36 | 0.38 |
| 33 | NEBRASKA | 1,826,341 | 6.41 | 0.35 |
| 34 | NORTH DAKOTA | 672,591 | 2.14 | 0.32 |
| 35 | DELAWARE | 897,934 | 2.46 | 0.27 |
| 36 | KANSAS | 2,853,118 | 7.48 | 0.26 |

| | | | | |
|---|---|---|---|---|
| 37 | ALASKA | 710,231 | 1.07 | 0.15 |
| 38 | CONNECTICUT | 3,574,097 | 3.2 | 0.09 |
| 39 | MAINE | 1,328,361 | 1.07 | 0.08 |
| 40 | NEW HAMPSHIRE | 1,316,470 | 1.07 | 0.08 |
| 41 | MINNESOTA | 5,303,925 | 3.2 | 0.06 |
| 42 | COLORADO | 5,029,196 | 3.2 | 0.06 |
| 43 | WASHINGTON | 6,724,540 | 1.07 | 0.02 |

U.S. GRAMS / PER 100K: 312,825,210 33,030.52 10.56

DRUG CODE:9143 DRUG NAME:OXYCODONE

| RANK | STATE | 2010 POP | TOTAL GRAMS | GRAMS/100K POP |
|---|---|---|---|---|
| 1 | DELAWARE | 897,934 | 302,828.77 | 33,725.06 |
| 2 | TENNESSEE | 6,346,105 | 2,053,484.17 | 32,358.18 |
| 3 | ARIZONA | 6,392,017 | 1,965,726.95 | 30,752.84 |
| 4 | NEVADA | 2,700,551 | 780,393.67 | 28,897.57 |
| 5 | PENNSYLVANIA | 12,702,379 | 3,456,077.45 | 27,208.11 |
| 6 | UTAH | 2,763,885 | 748,573.20 | 27,084.09 |
| 7 | NEW JERSEY | 8,791,894 | 2,355,962.03 | 26,796.98 |
| 8 | NORTH CAROLINA | 9,535,483 | 2,531,098.59 | 26,544 |
| 9 | OKLAHOMA | 3,751,351 | 958,060.96 | 25,539.09 |
| 10 | MARYLAND | 5,773,552 | 1,442,378.45 | 24,982.51 |
| 11 | WEST VIRGINIA | 1,852,994 | 428,047.21 | 23,100.30 |
| 12 | SOUTH CAROLINA | 4,625,364 | 1,031,254.28 | 22,295.64 |
| 13 | ALABAMA | 4,779,736 | 1,062,946.66 | 22,238.61 |
| 14 | MISSOURI | 5,988,927 | 1,327,291.96 | 22,162.43 |
| 15 | NEW HAMPSHIRE | 1,316,470 | 291,351.36 | 22,131.26 |
| 16 | CONNECTICUT | 3,574,097 | 789,289.15 | 22,083.60 |
| 17 | ARKANSAS | 2,915,918 | 633,760.13 | 21,734.50 |
| 18 | FLORIDA | 18,801,310 | 4,085,382.83 | 21,729.25 |
| 19 | NEW MEXICO | 2,059,179 | 443,200.86 | 21,523.18 |
| 20 | OREGON | 3,831,074 | 817,018.07 | 21,326.08 |
| 21 | MAINE | 1,328,361 | 281,441.17 | 21,187.10 |
| 22 | ALASKA | 710,231 | 150,330.93 | 21,166.48 |
| 23 | WYOMING | 563,626 | 116,851.70 | 20,732.13 |
| 24 | KENTUCKY | 4,339,367 | 872,640.94 | 20,109.87 |
| 25 | COLORADO | 5,029,196 | 1,006,153.19 | 20,006.24 |
| 26 | WASHINGTON | 6,724,540 | 1,308,543.62 | 19,459.23 |
| 27 | OHIO | 11,536,504 | 2,216,852.22 | 19,215.98 |
| 28 | GEORGIA | 9,687,653 | 1,834,305.71 | 18,934.47 |
| 29 | LOUISIANA | 4,533,372 | 851,246.54 | 18,777.34 |
| 30 | WISCONSIN | 5,686,986 | 1,066,992.50 | 18,762 |
| 31 | RHODE ISLAND | 1,052,567 | 196,139.15 | 18,634.36 |
| 32 | MONTANA | 989,415 | 183,003.52 | 18,496.13 |
| 33 | KANSAS | 2,853,118 | 525,756.09 | 18,427.42 |

DRUG CODE:9143 DRUG NAME:OXYCODONE

| RANK | STATE | 2010 POP | TOTAL GRAMS | GRAMS/100K POP |
|---|---|---|---|---|
| 34 | NEW YORK | 19,378,102 | 3,446,085.62 | 17,783.40 |
| 35 | VIRGINIA | 8,001,024 | 1,394,399.81 | 17,427.77 |
| 36 | MASSACHUSETTS | 6,547,629 | 1,120,868.23 | 17,118.69 |
| 37 | VERMONT | 625,741 | 103,921.91 | 16,607.81 |
| 38 | IDAHO | 1,567,582 | 254,733.92 | 16,250.12 |
| 39 | INDIANA | 6,483,802 | 1,042,665.24 | 16,081.08 |
| 40 | MISSISSIPPI | 2,967,297 | 444,974.76 | 14,995.96 |
| 41 | HAWAII | 1,360,301 | 194,994.28 | 14,334.64 |

| | | | | |
|---|---|---|---|---|
| 42 | MICHIGAN | 9,883,640 | 1,349,293.07 | 13,651.78 |
| 43 | MINNESOTA | 5,303,925 | 649,846.21 | 12,252.18 |
| 44 | DISTRICT OF COLUMBIA | 601,723 | 66,949.62 | 11,126.32 |
| 45 | CALIFORNIA | 37,253,956 | 3,908,067.77 | 10,490.34 |
| 46 | SOUTH DAKOTA | 814,180 | 84,861.13 | 10,422.90 |
| 47 | NORTH DAKOTA | 672,591 | 69,372.48 | 10,314.22 |
| 48 | NEBRASKA | 1,826,341 | 178,205.01 | 9,757.49 |
| 49 | IOWA | 3,046,355 | 241,384.68 | 7,923.72 |
| 50 | ILLINOIS | 12,830,632 | 693,040.63 | 5,401.45 |
| 51 | TEXAS | 25,145,561 | 1,340,070.64 | 5,329.25 |
| 52 | VIRGIN ISLANDS | 109,825 | 2,614.82 | 2,380.90 |
| 53 | GUAM | 178,430 | 3,964.91 | 2,222.11 |
| 54 | PUERTO RICO | 3,725,789 | 73,059.73 | 1,960.92 |
| 55 | AMERICAN SAMOA | 65,628 | 133.13 | 202.86 |
| U.S. GRAMS / PER 100K: | | 312,825,210 | 54,777,891.63 | 17,510.70 |

DRUG CODE:9150 DRUG NAME:HYDROMORPHONE

| RANK | STATE | 2010 POP | TOTAL GRAMS | GRAMS/100K POP |
|---|---|---|---|---|
| 1 | FLORIDA | 18,801,310 | 210,243.26 | 1,118.24 |
| 2 | DELAWARE | 897,934 | 9,694.62 | 1,079.66 |
| 3 | VERMONT | 625,741 | 5,426.36 | 867.19 |
| 4 | CONNECTICUT | 3,574,097 | 28,698.88 | 802.97 |
| 5 | MONTANA | 989,415 | 7,226.26 | 730.36 |
| 6 | WASHINGTON | 6,724,540 | 49,020.70 | 728.98 |
| 7 | MISSISSIPPI | 2,967,297 | 20,736.20 | 698.82 |
| 8 | VIRGINIA | 8,001,024 | 54,568.03 | 682.01 |
| 9 | MARYLAND | 5,773,552 | 39,124.04 | 677.64 |
| 10 | NEW HAMPSHIRE | 1,316,470 | 8,385.43 | 636.96 |
| 11 | COLORADO | 5,029,196 | 31,455.65 | 625.46 |
| 12 | ALASKA | 710,231 | 4,306.79 | 606.39 |
| 13 | OREGON | 3,831,074 | 22,312.33 | 582.4 |
| 14 | MAINE | 1,328,361 | 7,703.94 | 579.96 |
| 15 | KANSAS | 2,853,118 | 15,766.80 | 552.62 |
| 16 | PENNSYLVANIA | 12,702,379 | 68,524.28 | 539.46 |
| 17 | NEVADA | 2,700,551 | 14,337.49 | 530.91 |
| 18 | SOUTH CAROLINA | 4,625,364 | 24,554.06 | 530.86 |
| 19 | ARKANSAS | 2,915,918 | 15,357.65 | 526.68 |
| 20 | IDAHO | 1,567,582 | 8,246.24 | 526.05 |
| 21 | NORTH CAROLINA | 9,535,483 | 49,621.61 | 520.39 |
| 22 | WYOMING | 563,626 | 2,917.45 | 517.62 |
| 23 | MISSOURI | 5,988,927 | 30,427.08 | 508.06 |
| 24 | ARIZONA | 6,392,017 | 31,708.88 | 496.07 |
| 25 | MASSACHUSETTS | 6,547,629 | 32,083.07 | 490 |
| 26 | CALIFORNIA | 37,253,956 | 177,498.75 | 476.46 |
| 27 | NORTH DAKOTA | 672,591 | 3,194.94 | 475.02 |
| 28 | NEW JERSEY | 8,791,894 | 41,725.37 | 474.59 |
| 29 | MINNESOTA | 5,303,925 | 25,121.86 | 473.65 |
| 30 | NEW YORK | 19,378,102 | 90,645.84 | 467.77 |
| 31 | IOWA | 3,046,355 | 14,058.50 | 461.49 |
| 32 | TENNESSEE | 6,346,105 | 28,984.64 | 456.73 |
| 33 | OKLAHOMA | 3,751,351 | 16,443.44 | 438.33 |
| 34 | INDIANA | 6,483,802 | 28,406.31 | 438.11 |

DRUG CODE:9150 DRUG NAME:HYDROMORPHONE

| RANK | STATE | 2010 POP | TOTAL GRAMS | GRAMS/100K POP |
|---|---|---|---|---|
| 35 | UTAH | 2,763,885 | 11,963.58 | 432.85 |

| | | | | |
|---|---|---|---|---|
| 36 | MICHIGAN | 9,883,640 | 42,587.58 | 430.89 |
| 37 | TEXAS | 25,145,561 | 104,785.93 | 416.72 |
| 38 | DISTRICT OF COLUMBIA | 601,723 | 2,483.98 | 412.81 |
| 39 | WEST VIRGINIA | 1,852,994 | 7,289.22 | 393.38 |
| 40 | GEORGIA | 9,687,653 | 35,763.31 | 369.16 |
| 41 | OHIO | 11,536,504 | 41,186.50 | 357.01 |
| 42 | NEW MEXICO | 2,059,179 | 7,309.23 | 354.96 |
| 43 | ILLINOIS | 12,830,632 | 43,432.15 | 338.5 |
| 44 | LOUISIANA | 4,533,372 | 15,256.55 | 336.54 |
| 45 | RHODE ISLAND | 1,052,567 | 3,540.34 | 336.35 |
| 46 | SOUTH DAKOTA | 814,180 | 2,733.70 | 335.76 |
| 47 | ALABAMA | 4,779,736 | 15,507.69 | 324.45 |
| 48 | NEBRASKA | 1,826,341 | 5,853.43 | 320.5 |
| 49 | WISCONSIN | 5,686,986 | 16,851.09 | 296.31 |
| 50 | KENTUCKY | 4,339,367 | 12,607.09 | 290.53 |
| 51 | HAWAII | 1,360,301 | 3,115.28 | 229.01 |
| 52 | VIRGIN ISLANDS | 109,825 | 101.95 | 92.83 |
| 53 | GUAM | 178,430 | 126.13 | 70.69 |
| 54 | PUERTO RICO | 3,725,789 | 286.95 | 7.7 |
| 55 | AMERICAN SAMOA | 65,628 | 3.16 | 4.81 |

U.S. GRAMS / PER 100K: 312,825,210 1,591,311.59 508.69

DRUG CODE:9193 DRUG NAME:HYDROCODONE

| RANK | STATE | 2010 POP | TOTAL GRAMS | GRAMS/100K POP |
|---|---|---|---|---|
| 1 | ALABAMA | 4,779,736 | 1,100,450.01 | 23,023.24 |
| 2 | OKLAHOMA | 3,751,351 | 781,791.52 | 20,840.27 |
| 3 | KENTUCKY | 4,339,367 | 874,683.05 | 20,156.93 |
| 4 | ARKANSAS | 2,915,918 | 572,750.68 | 19,642.21 |
| 5 | MISSISSIPPI | 2,967,297 | 535,193.49 | 18,036.40 |
| 6 | MICHIGAN | 9,883,640 | 1,780,140.48 | 18,010.98 |
| 7 | TENNESSEE | 6,346,105 | 1,096,801.43 | 17,283.06 |
| 8 | LOUISIANA | 4,533,372 | 730,405.90 | 16,111.76 |
| 9 | IDAHO | 1,567,582 | 244,856.68 | 15,620.02 |
| 10 | INDIANA | 6,483,802 | 1,009,374.18 | 15,567.63 |
| 11 | NEVADA | 2,700,551 | 419,919.17 | 15,549.39 |
| 12 | WEST VIRGINIA | 1,852,994 | 285,366.06 | 15,400.27 |
| 13 | SOUTH CAROLINA | 4,625,364 | 617,686.53 | 13,354.33 |
| 14 | MISSOURI | 5,988,927 | 784,794.89 | 13,104.10 |
| 15 | KANSAS | 2,853,118 | 354,843.61 | 12,437.05 |
| 16 | MONTANA | 989,415 | 114,655.69 | 11,588.23 |
| 17 | TEXAS | 25,145,561 | 2,884,672.22 | 11,471.89 |
| 18 | OREGON | 3,831,074 | 431,705.45 | 11,268.52 |
| 19 | GEORGIA | 9,687,653 | 1,081,004.03 | 11,158.58 |
| 20 | CALIFORNIA | 37,253,956 | 4,068,733.07 | 10,921.61 |
| 21 | ILLINOIS | 12,830,632 | 1,358,791.21 | 10,590.21 |
| 22 | WYOMING | 563,626 | 53,847.97 | 9,553.85 |
| 23 | UTAH | 2,763,885 | 261,023.11 | 9,444.07 |
| 24 | NORTH CAROLINA | 9,535,483 | 872,859.27 | 9,153.80 |
| 25 | WASHINGTON | 6,724,540 | 589,862.25 | 8,771.79 |
| 26 | SOUTH DAKOTA | 814,180 | 69,704.64 | 8,561.33 |
| 27 | IOWA | 3,046,355 | 260,586.46 | 8,554.04 |
| 28 | FLORIDA | 18,801,310 | 1,558,486.74 | 8,289.25 |
| 29 | NEBRASKA | 1,826,341 | 146,319.58 | 8,011.62 |
| 30 | NEW MEXICO | 2,059,179 | 161,924.40 | 7,863.54 |
| 31 | OHIO | 11,536,504 | 867,738.40 | 7,521.68 |
| 32 | ALASKA | 710,231 | 52,831.78 | 7,438.68 |
| 33 | ARIZONA | 6,392,017 | 468,233.01 | 7,325.28 |

| | | | | |
|---|---|---|---|---|
| 34 | NORTH DAKOTA | 672,591 | 48,659.11 | 7,234.58 |
| 35 | WISCONSIN | 5,686,986 | 363,373.40 | 6,389.56 |

DRUG CODE:9193 DRUG NAME:HYDROCODONE

| RANK | STATE | 2010 POP | TOTAL GRAMS | GRAMS/100K POP |
|---|---|---|---|---|
| 36 | MAINE | 1,328,361 | 83,433.44 | 6,280.93 |
| 37 | VIRGINIA | 8,001,024 | 502,313.33 | 6,278.11 |
| 38 | PENNSYLVANIA | 12,702,379 | 784,883.99 | 6,179.03 |
| 39 | COLORADO | 5,029,196 | 289,553.38 | 5,757.45 |
| 40 | HAWAII | 1,360,301 | 77,750.83 | 5,715.71 |
| 41 | RHODE ISLAND | 1,052,567 | 50,282.18 | 4,777.10 |
| 42 | NEW YORK | 19,378,102 | 793,106.78 | 4,092.80 |
| 43 | MINNESOTA | 5,303,925 | 212,400.04 | 4,004.58 |
| 44 | VERMONT | 625,741 | 23,339.01 | 3,729.82 |
| 45 | NEW HAMPSHIRE | 1,316,470 | 41,370.88 | 3,142.56 |
| 46 | DELAWARE | 897,934 | 27,890.42 | 3,106.07 |
| 47 | MARYLAND | 5,773,552 | 160,713.59 | 2,783.62 |
| 48 | CONNECTICUT | 3,574,097 | 96,663.75 | 2,704.56 |
| 49 | MASSACHUSETTS | 6,547,629 | 140,699.48 | 2,148.86 |
| 50 | NEW JERSEY | 8,791,894 | 166,586.54 | 1,894.77 |
| 51 | VIRGIN ISLANDS | 109,825 | 1,388.11 | 1,263.93 |
| 52 | DISTRICT OF COLUMBIA | 601,723 | 6,875.82 | 1,142.69 |
| 53 | GUAM | 178,430 | 820.13 | 459.64 |
| 54 | PUERTO RICO | 3,725,789 | 4,411.01 | 118.39 |
| 55 | AMERICAN SAMOA | 65,628 | 32.69 | 49.81 |
| U.S. GRAMS / PER 100K: | | 312,825,210 | 30,368,584.86 | 9,707.84 |

DRUG CODE:9220L DRUG NAME:LEVORPHANOL

| RANK | STATE | 2010 POP | TOTAL GRAMS | GRAMS/100K POP |
|---|---|---|---|---|
| 1 | NEW JERSEY | 8,791,894 | 594.68 | 6.76 |
| 2 | INDIANA | 6,483,802 | 101.22 | 1.56 |
| 3 | NORTH CAROLINA | 9,535,483 | 112.48 | 1.18 |
| 4 | MASSACHUSETTS | 6,547,629 | 75.34 | 1.15 |
| 5 | KANSAS | 2,853,118 | 32.73 | 1.15 |
| 6 | CALIFORNIA | 37,253,956 | 396.53 | 1.06 |
| 7 | LOUISIANA | 4,533,372 | 41.21 | 0.91 |
| 8 | WASHINGTON | 6,724,540 | 54.79 | 0.81 |
| 9 | NEW YORK | 19,378,102 | 135.93 | 0.7 |
| 10 | ARIZONA | 6,392,017 | 39.58 | 0.62 |
| 11 | IOWA | 3,046,355 | 19.04 | 0.62 |
| 12 | MISSOURI | 5,988,927 | 30.88 | 0.52 |
| 13 | OREGON | 3,831,074 | 19.15 | 0.5 |
| 14 | TEXAS | 25,145,561 | 126.18 | 0.5 |
| 15 | FLORIDA | 18,801,310 | 79.17 | 0.42 |
| 16 | ALABAMA | 4,779,736 | 19.85 | 0.42 |
| 17 | COLORADO | 5,029,196 | 19.5 | 0.39 |
| 18 | VERMONT | 625,741 | 1.97 | 0.32 |
| 19 | NEVADA | 2,700,551 | 8.47 | 0.31 |
| 20 | PENNSYLVANIA | 12,702,379 | 38.77 | 0.31 |
| 21 | VIRGINIA | 8,001,024 | 23.33 | 0.29 |
| 22 | ARKANSAS | 2,915,918 | 7.89 | 0.27 |
| 23 | NEW HAMPSHIRE | 1,316,470 | 3.37 | 0.26 |
| 24 | WISCONSIN | 5,686,986 | 12.88 | 0.23 |
| 25 | ILLINOIS | 12,830,632 | 29.02 | 0.23 |
| 26 | OHIO | 11,536,504 | 24.61 | 0.21 |

| | | | | |
|---|---|---|---|---|
| 27 | NEBRASKA | 1,826,341 | 3.6 | 0.2 |
| 28 | CONNECTICUT | 3,574,097 | 5.34 | 0.15 |
| 29 | MISSISSIPPI | 2,967,297 | 3.02 | 0.1 |
| 30 | MARYLAND | 5,773,552 | 4.06 | 0.07 |
| 31 | WEST VIRGINIA | 1,852,994 | 1.04 | 0.06 |
| 32 | OKLAHOMA | 3,751,351 | 1.86 | 0.05 |
| 33 | MINNESOTA | 5,303,925 | 2.55 | 0.05 |
| 34 | GEORGIA | 9,687,653 | 4.53 | 0.05 |
| 35 | MICHIGAN | 9,883,640 | 3.6 | 0.04 |
| 36 | SOUTH CAROLINA | 4,625,364 | 1.86 | 0.04 |

DRUG CODE:9220L DRUG NAME:LEVORPHANOL

| RANK | STATE | 2010 POP | TOTAL GRAMS | GRAMS/100K POP |
|---|---|---|---|---|
| 37 | MAINE | 1,328,361 | 0.23 | 0.02 |
| 38 | NEW MEXICO | 2,059,179 | 0.12 | 0.01 |
| 39 | UTAH | 2,763,885 | 0.35 | 0.01 |
| 40 | TENNESSEE | 6,346,105 | 0.93 | 0.01 |
| U.S. GRAMS / PER 100K: | | 312,825,210 | 2,081.66 | 0.67 |

DRUG CODE:9230 DRUG NAME:MEPERIDINE (PETHIDINE)

| RANK | STATE | 2010 POP | TOTAL GRAMS | GRAMS/100K POP |
|---|---|---|---|---|
| 1 | ARKANSAS | 2,915,918 | 38,196.36 | 1,309.93 |
| 2 | ALABAMA | 4,779,736 | 46,122.48 | 964.96 |
| 3 | OKLAHOMA | 3,751,351 | 35,801.53 | 954.36 |
| 4 | LOUISIANA | 4,533,372 | 37,324.80 | 823.33 |
| 5 | MISSISSIPPI | 2,967,297 | 22,727.35 | 765.93 |
| 6 | PUERTO RICO | 3,725,789 | 25,241.36 | 677.48 |
| 7 | SOUTH CAROLINA | 4,625,364 | 30,490.34 | 659.2 |
| 8 | TENNESSEE | 6,346,105 | 37,627.51 | 592.92 |
| 9 | GEORGIA | 9,687,653 | 49,932.07 | 515.42 |
| 10 | ALASKA | 710,231 | 3,309.63 | 465.99 |
| 11 | ARIZONA | 6,392,017 | 24,769.38 | 387.5 |
| 12 | VIRGINIA | 8,001,024 | 29,872.38 | 373.36 |
| 13 | NEW MEXICO | 2,059,179 | 7,480.72 | 363.29 |
| 14 | WYOMING | 563,626 | 2,000.88 | 355 |
| 15 | MONTANA | 989,415 | 3,269 | 330.4 |
| 16 | UTAH | 2,763,885 | 9,102.86 | 329.35 |
| 17 | MISSOURI | 5,988,927 | 19,501.12 | 325.62 |
| 18 | SOUTH DAKOTA | 814,180 | 2,557.20 | 314.08 |
| 19 | NORTH CAROLINA | 9,535,483 | 29,504.98 | 309.42 |
| 20 | KANSAS | 2,853,118 | 7,988.69 | 280 |
| 21 | KENTUCKY | 4,339,367 | 11,708.47 | 269.82 |
| 22 | NEVADA | 2,700,551 | 7,047.84 | 260.98 |
| 23 | IOWA | 3,046,355 | 7,829.80 | 257.02 |
| 24 | WASHINGTON | 6,724,540 | 16,929.13 | 251.75 |
| 25 | IDAHO | 1,567,582 | 3,726.97 | 237.75 |
| 26 | INDIANA | 6,483,802 | 15,027.60 | 231.77 |
| 27 | VERMONT | 625,741 | 1,431.66 | 228.79 |
| 28 | TEXAS | 25,145,561 | 56,612.03 | 225.14 |
| 29 | FLORIDA | 18,801,310 | 42,062.60 | 223.72 |
| 30 | NEW HAMPSHIRE | 1,316,470 | 2,900.79 | 220.35 |
| 31 | VIRGIN ISLANDS | 109,825 | 237.37 | 216.14 |
| 32 | MARYLAND | 5,773,552 | 11,569.35 | 200.39 |
| 33 | OHIO | 11,536,504 | 22,558.96 | 195.54 |

| | | | | |
|---|---|---|---|---|
| 34 | COLORADO | 5,029,196 | 9,597.39 | 190.83 |
| 35 | CALIFORNIA | 37,253,956 | 70,773.82 | 189.98 |
| 36 | OREGON | 3,831,074 | 7,262.19 | 189.56 |
| 37 | NEBRASKA | 1,826,341 | 3,461.51 | 189.53 |
| 38 | WEST VIRGINIA | 1,852,994 | 3,229.32 | 174.28 |
| 39 | NORTH DAKOTA | 672,591 | 1,134.26 | 168.64 |
| 40 | MASSACHUSETTS | 6,547,629 | 10,865.88 | 165.95 |
| 41 | MAINE | 1,328,361 | 2,008.81 | 151.22 |
| 42 | CONNECTICUT | 3,574,097 | 5,356.78 | 149.88 |
| 43 | MICHIGAN | 9,883,640 | 14,512.15 | 146.83 |
| 44 | PENNSYLVANIA | 12,702,379 | 17,757.07 | 139.79 |
| 45 | WISCONSIN | 5,686,986 | 7,669.90 | 134.87 |
| 46 | DISTRICT OF COLUMBIA | 601,723 | 797.1 | 132.47 |
| 47 | HAWAII | 1,360,301 | 1,766.42 | 129.86 |
| 48 | ILLINOIS | 12,830,632 | 14,772.26 | 115.13 |
| 49 | NEW YORK | 19,378,102 | 22,280.75 | 114.98 |
| 50 | NEW JERSEY | 8,791,894 | 9,550.55 | 108.63 |
| 51 | DELAWARE | 897,934 | 934.2 | 104.04 |
| 52 | RHODE ISLAND | 1,052,567 | 950.81 | 90.33 |

DRUG CODE:9230 DRUG NAME:MEPERIDINE (PETHIDINE)

| RANK | STATE | 2010 POP | TOTAL GRAMS | GRAMS/100K POP |
|---|---|---|---|---|
| 53 | GUAM | 178,430 | 157.85 | 88.47 |
| 54 | MINNESOTA | 5,303,925 | 3,328.48 | 62.75 |
| 55 | AMERICAN SAMOA | 65,628 | 6.1 | 9.3 |
| U.S. GRAMS / PER 100K: | | 312,825,210 | 870,636.82 | 278.31 |

DRUG CODE:9250B DRUG NAME:METHADONE

| RANK | STATE | 2010 POP | TOTAL GRAMS | GRAMS/100K POP |
|---|---|---|---|---|
| 1 | RHODE ISLAND | 1,052,567 | 169,802.02 | 16,132.18 |
| 2 | MARYLAND | 5,773,552 | 711,869.76 | 12,329.84 |
| 3 | CONNECTICUT | 3,574,097 | 425,326.15 | 11,900.24 |
| 4 | DELAWARE | 897,934 | 101,183.03 | 11,268.43 |
| 5 | VERMONT | 625,741 | 68,180.38 | 10,895.94 |
| 6 | MAINE | 1,328,361 | 141,416.18 | 10,645.91 |
| 7 | NEW HAMPSHIRE | 1,316,470 | 120,612.25 | 9,161.79 |
| 8 | MASSACHUSETTS | 6,547,629 | 541,344.43 | 8,267.79 |
| 9 | NEW MEXICO | 2,059,179 | 153,019.50 | 7,431.09 |
| 10 | ALABAMA | 4,779,736 | 336,025.72 | 7,030.22 |
| 11 | OREGON | 3,831,074 | 261,909.20 | 6,836.44 |
| 12 | DISTRICT OF COLUMBIA | 601,723 | 40,446.24 | 6,721.74 |
| 13 | PENNSYLVANIA | 12,702,379 | 764,430.66 | 6,018.01 |
| 14 | NEW YORK | 19,378,102 | 1,154,264.55 | 5,956.54 |
| 15 | WASHINGTON | 6,724,540 | 397,959.34 | 5,918.02 |
| 16 | NEVADA | 2,700,551 | 154,141.28 | 5,707.77 |
| 17 | NORTH CAROLINA | 9,535,483 | 525,275.89 | 5,508.64 |
| 18 | WEST VIRGINIA | 1,852,994 | 100,539.06 | 5,425.76 |
| 19 | INDIANA | 6,483,802 | 343,847.46 | 5,303.18 |
| 20 | ARIZONA | 6,392,017 | 333,774.91 | 5,221.75 |
| 21 | MICHIGAN | 9,883,640 | 512,952.79 | 5,189.92 |
| 22 | NEW JERSEY | 8,791,894 | 436,185.10 | 4,961.22 |
| 23 | SOUTH CAROLINA | 4,625,364 | 222,146.78 | 4,802.80 |
| 24 | KENTUCKY | 4,339,367 | 207,213.82 | 4,775.21 |
| 25 | GEORGIA | 9,687,653 | 462,164.69 | 4,770.66 |
| 26 | ALASKA | 710,231 | 32,205.82 | 4,534.56 |

| | | | | |
|---|---|---|---|---|
| 27 | FLORIDA | 18,801,310 | 846,448.12 | 4,502.07 |
| 28 | WISCONSIN | 5,686,986 | 255,722.68 | 4,496.63 |
| 29 | MINNESOTA | 5,303,925 | 221,011.42 | 4,166.94 |
| 30 | OKLAHOMA | 3,751,351 | 153,160.81 | 4,082.82 |
| 31 | UTAH | 2,763,885 | 110,576.64 | 4,000.77 |
| 32 | CALIFORNIA | 37,253,956 | 1,401,083.55 | 3,760.90 |
| 33 | ILLINOIS | 12,830,632 | 476,702.56 | 3,715.35 |
| 34 | VIRGINIA | 8,001,024 | 291,599.76 | 3,644.53 |
| 35 | LOUISIANA | 4,533,372 | 153,239.35 | 3,380.25 |
| 36 | KANSAS | 2,853,118 | 95,821.37 | 3,358.48 |
| 37 | IDAHO | 1,567,582 | 51,096.36 | 3,259.57 |
| 38 | TENNESSEE | 6,346,105 | 204,209.90 | 3,217.88 |
| 39 | COLORADO | 5,029,196 | 158,593.77 | 3,153.46 |
| 40 | MONTANA | 989,415 | 29,717.24 | 3,003.52 |
| 41 | MISSOURI | 5,988,927 | 166,654.04 | 2,782.70 |
| 42 | OHIO | 11,536,504 | 310,964.42 | 2,695.48 |
| 43 | ARKANSAS | 2,915,918 | 77,586.26 | 2,660.78 |
| 44 | HAWAII | 1,360,301 | 33,605.11 | 2,470.42 |
| 45 | TEXAS | 25,145,561 | 558,522.94 | 2,221.16 |
| 46 | PUERTO RICO | 3,725,789 | 74,682.04 | 2,004.46 |
| 47 | NEBRASKA | 1,826,341 | 35,003.52 | 1,916.59 |
| 48 | IOWA | 3,046,355 | 53,451.87 | 1,754.62 |
| 49 | SOUTH DAKOTA | 814,180 | 9,822.75 | 1,206.46 |
| 50 | WYOMING | 563,626 | 5,788.47 | 1,027.01 |
| 51 | MISSISSIPPI | 2,967,297 | 27,273.74 | 919.14 |
| 52 | NORTH DAKOTA | 672,591 | 5,116.95 | 760.78 |
| 53 | VIRGIN ISLANDS | 109,825 | 165.02 | 150.25 |

DRUG CODE:9250B DRUG NAME:METHADONE

| RANK | STATE | 2010 POP | TOTAL GRAMS | GRAMS/100K POP |
|---|---|---|---|---|
| 54 | GUAM | 178,430 | 164.57 | 92.23 |

| | | | | |
|---|---|---|---|---|
| U.S. GRAMS / PER 100K: | | 312,825,210 | 14,526,022.25 | 4,643.49 |

DRUG CODE:9300 DRUG NAME:MORPHINE

| RANK | STATE | 2010 POP | TOTAL GRAMS | GRAMS/100K POP |
|---|---|---|---|---|
| 1 | ARIZONA | 6,392,017 | 800,446.82 | 12,522.60 |
| 2 | TENNESSEE | 6,346,105 | 763,959.07 | 12,038.24 |
| 3 | OKLAHOMA | 3,751,351 | 415,738.56 | 11,082.37 |
| 4 | NEVADA | 2,700,551 | 284,823.07 | 10,546.85 |
| 5 | DELAWARE | 897,934 | 91,399.62 | 10,178.88 |
| 6 | OREGON | 3,831,074 | 360,432.77 | 9,408.14 |
| 7 | FLORIDA | 18,801,310 | 1,738,653.07 | 9,247.51 |
| 8 | MICHIGAN | 9,883,640 | 886,883.89 | 8,973.25 |
| 9 | MONTANA | 989,415 | 85,850.66 | 8,676.91 |
| 10 | PENNSYLVANIA | 12,702,379 | 1,034,377.25 | 8,143.18 |
| 11 | UTAH | 2,763,885 | 223,560.82 | 8,088.64 |
| 12 | IDAHO | 1,567,582 | 125,092.80 | 7,979.98 |
| 13 | HAWAII | 1,360,301 | 103,542.08 | 7,611.70 |
| 14 | WASHINGTON | 6,724,540 | 510,085.64 | 7,585.44 |
| 15 | ALASKA | 710,231 | 53,408.22 | 7,519.84 |
| 16 | MISSOURI | 5,988,927 | 438,279.12 | 7,318.16 |
| 17 | WYOMING | 563,626 | 41,173.40 | 7,305.09 |
| 18 | VERMONT | 625,741 | 45,167.82 | 7,218.29 |
| 19 | COLORADO | 5,029,196 | 358,367.59 | 7,125.74 |
| 20 | ALABAMA | 4,779,736 | 338,869.93 | 7,089.72 |

| | | | | |
|---|---|---|---|---|
| 21 | ARKANSAS | 2,915,918 | 206,399.29 | 7,078.36 |
| 22 | KANSAS | 2,853,118 | 201,456.82 | 7,060.94 |
| 23 | WISCONSIN | 5,686,986 | 385,192.10 | 6,773.22 |
| 24 | MAINE | 1,328,361 | 88,978.74 | 6,698.39 |
| 25 | NEW MEXICO | 2,059,179 | 133,529.97 | 6,484.62 |
| 26 | INDIANA | 6,483,802 | 410,187.16 | 6,326.34 |
| 27 | SOUTH CAROLINA | 4,625,364 | 289,145.23 | 6,251.30 |
| 28 | MARYLAND | 5,773,552 | 355,354.24 | 6,154.86 |
| 29 | NEW HAMPSHIRE | 1,316,470 | 79,717.42 | 6,055.39 |
| 30 | WEST VIRGINIA | 1,852,994 | 110,890.31 | 5,984.39 |
| 31 | MISSISSIPPI | 2,967,297 | 173,577.48 | 5,849.68 |
| 32 | NORTH CAROLINA | 9,535,483 | 552,491.34 | 5,794.06 |
| 33 | CALIFORNIA | 37,253,956 | 2,074,041.62 | 5,567.31 |
| 34 | LOUISIANA | 4,533,372 | 251,738.17 | 5,553 |
| 35 | SOUTH DAKOTA | 814,180 | 43,661.31 | 5,362.61 |
| 36 | IOWA | 3,046,355 | 153,245.45 | 5,030.45 |
| 37 | GEORGIA | 9,687,653 | 483,611.16 | 4,992.04 |
| 38 | RHODE ISLAND | 1,052,567 | 51,943.21 | 4,934.91 |
| 39 | KENTUCKY | 4,339,367 | 212,966.15 | 4,907.77 |
| 40 | OHIO | 11,536,504 | 543,747.13 | 4,713.27 |
| 41 | CONNECTICUT | 3,574,097 | 164,444.74 | 4,601.02 |
| 42 | MASSACHUSETTS | 6,547,629 | 298,878.14 | 4,564.68 |
| 43 | VIRGINIA | 8,001,024 | 363,361.54 | 4,541.44 |
| 44 | NORTH DAKOTA | 672,591 | 30,517.90 | 4,537.36 |
| 45 | NEW JERSEY | 8,791,894 | 396,261.77 | 4,507.13 |
| 46 | NEBRASKA | 1,826,341 | 80,373.93 | 4,400.82 |
| 47 | MINNESOTA | 5,303,925 | 214,606.29 | 4,046.18 |
| 48 | NEW YORK | 19,378,102 | 730,556.27 | 3,770.01 |
| 49 | TEXAS | 25,145,561 | 918,185.26 | 3,651.48 |
| 50 | ILLINOIS | 12,830,632 | 450,775.38 | 3,513.27 |
| 51 | DISTRICT OF COLUMBIA | 601,723 | 17,510.30 | 2,910.03 |
| 52 | GUAM | 178,430 | 4,795.87 | 2,687.82 |
| 53 | VIRGIN ISLANDS | 109,825 | 416.76 | 379.48 |
| 54 | PUERTO RICO | 3,725,789 | 11,919.62 | 319.92 |
| 55 | AMERICAN SAMOA | 65,628 | 148.9 | 226.88 |

DRUG CODE:9300 DRUG NAME:MORPHINE

| RANK | STATE | 2010 POP | TOTAL GRAMS | GRAMS/100K POP |
|---|---|---|---|---|
| U.S. GRAMS / PER 100K: | | 312,825,210 | 19,184,739.20 | 6,132.73 |

DRUG CODE:9630 DRUG NAME:OPIUM TINCTURE (LAUDANUM)

| RANK | STATE | 2010 POP | TOTAL GRAMS | GRAMS/100K POP |
|---|---|---|---|---|
| 1 | MASSACHUSETTS | 6,547,629 | 150.17 | 2.29 |
| 2 | INDIANA | 6,483,802 | 81.57 | 1.26 |
| 3 | WEST VIRGINIA | 1,852,994 | 20.09 | 1.08 |
| 4 | OHIO | 11,536,504 | 96.92 | 0.84 |
| 5 | NEBRASKA | 1,826,341 | 14.19 | 0.78 |
| 6 | MICHIGAN | 9,883,640 | 47.29 | 0.48 |
| 7 | MAINE | 1,328,361 | 5.91 | 0.44 |
| 8 | MISSOURI | 5,988,927 | 26.01 | 0.43 |
| 9 | VERMONT | 625,741 | 2.36 | 0.38 |
| 10 | WASHINGTON | 6,724,540 | 20.06 | 0.3 |
| 11 | ILLINOIS | 12,830,632 | 34.27 | 0.27 |
| 12 | NEW MEXICO | 2,059,179 | 4.73 | 0.23 |
| 13 | VIRGINIA | 8,001,024 | 17.7 | 0.22 |

| | | | | |
|---|---|---|---|---|
| 14 | KANSAS | 2,853,118 | 5.91 | 0.21 |
| 15 | FLORIDA | 18,801,310 | 37.84 | 0.2 |
| 16 | DISTRICT OF COLUMBIA | 601,723 | 1.18 | 0.2 |
| 17 | MARYLAND | 5,773,552 | 10.62 | 0.18 |
| 18 | ARIZONA | 6,392,017 | 9.46 | 0.15 |
| 19 | CALIFORNIA | 37,253,956 | 46.11 | 0.12 |
| 20 | OREGON | 3,831,074 | 4.72 | 0.12 |
| 21 | NEW JERSEY | 8,791,894 | 9.44 | 0.11 |
| 22 | PENNSYLVANIA | 12,702,379 | 12.99 | 0.1 |
| 23 | NEW HAMPSHIRE | 1,316,470 | 1.18 | 0.09 |
| 24 | TEXAS | 25,145,561 | 21.24 | 0.08 |
| 25 | IDAHO | 1,567,582 | 1.18 | 0.08 |
| 26 | COLORADO | 5,029,196 | 1.18 | 0.02 |
| 27 | MINNESOTA | 5,303,925 | 1.18 | 0.02 |
| 28 | WISCONSIN | 5,686,986 | 1.18 | 0.02 |
| 29 | NEW YORK | 19,378,102 | 3.54 | 0.02 |
| U.S. GRAMS / PER 100K: | | 312,825,210 | 690.22 | 0.22 |

DRUG CODE:9639 DRUG NAME:OPIUM POWDERED

| RANK | STATE | 2010 POP | TOTAL GRAMS | GRAMS/100K POP |
|---|---|---|---|---|
| 1 | MINNESOTA | 5,303,925 | 1,693.08 | 31.92 |
| 2 | KANSAS | 2,853,118 | 749.88 | 26.28 |
| 3 | VERMONT | 625,741 | 153 | 24.45 |
| 4 | WYOMING | 563,626 | 131.76 | 23.38 |
| 5 | NORTH DAKOTA | 672,591 | 154.08 | 22.91 |
| 6 | NEBRASKA | 1,826,341 | 413.64 | 22.65 |
| 7 | WEST VIRGINIA | 1,852,994 | 414.72 | 22.38 |
| 8 | MONTANA | 989,415 | 216.72 | 21.9 |
| 9 | OREGON | 3,831,074 | 827.64 | 21.6 |
| 10 | WASHINGTON | 6,724,540 | 1,309.68 | 19.48 |
| 11 | COLORADO | 5,029,196 | 903.6 | 17.97 |
| 12 | ARIZONA | 6,392,017 | 1,108.44 | 17.34 |
| 13 | KENTUCKY | 4,339,367 | 704.88 | 16.24 |
| 14 | INDIANA | 6,483,802 | 1,031.40 | 15.91 |
| 15 | TENNESSEE | 6,346,105 | 958.68 | 15.11 |
| 16 | OHIO | 11,536,504 | 1,727.64 | 14.98 |
| 17 | MICHIGAN | 9,883,640 | 1,467.72 | 14.85 |
| 18 | WISCONSIN | 5,686,986 | 784.8 | 13.8 |
| 19 | IOWA | 3,046,355 | 410.4 | 13.47 |
| 20 | ARKANSAS | 2,915,918 | 391.32 | 13.42 |
| 21 | MISSOURI | 5,988,927 | 800.64 | 13.37 |
| 22 | SOUTH CAROLINA | 4,625,364 | 584.64 | 12.64 |
| 23 | RHODE ISLAND | 1,052,567 | 132.48 | 12.59 |
| 24 | MAINE | 1,328,361 | 164.88 | 12.41 |

DRUG CODE:9639 DRUG NAME:OPIUM POWDERED

| RANK | STATE | 2010 POP | TOTAL GRAMS | GRAMS/100K POP |
|---|---|---|---|---|
| 25 | PENNSYLVANIA | 12,702,379 | 1,471.32 | 11.58 |
| 26 | IDAHO | 1,567,582 | 181.44 | 11.57 |
| 27 | NEW HAMPSHIRE | 1,316,470 | 149.76 | 11.38 |
| 28 | SOUTH DAKOTA | 814,180 | 90.36 | 11.1 |
| 29 | NEW MEXICO | 2,059,179 | 226.8 | 11.01 |
| 30 | ALASKA | 710,231 | 77.76 | 10.95 |
| 31 | MASSACHUSETTS | 6,547,629 | 697.32 | 10.65 |

| | | | | |
|---|---|---|---|---|
| 32 | CONNECTICUT | 3,574,097 | 364.32 | 10.19 |
| 33 | FLORIDA | 18,801,310 | 1,872.36 | 9.96 |
| 34 | NORTH CAROLINA | 9,535,483 | 928.08 | 9.73 |
| 35 | LOUISIANA | 4,533,372 | 429.12 | 9.47 |
| 36 | UTAH | 2,763,885 | 247.32 | 8.95 |
| 37 | VIRGINIA | 8,001,024 | 716.4 | 8.95 |
| 38 | DELAWARE | 897,934 | 78.12 | 8.7 |
| 39 | TEXAS | 25,145,561 | 2,089.08 | 8.31 |
| 40 | OKLAHOMA | 3,751,351 | 308.52 | 8.22 |
| 41 | GEORGIA | 9,687,653 | 780.12 | 8.05 |
| 42 | ALABAMA | 4,779,736 | 379.08 | 7.93 |
| 43 | DISTRICT OF COLUMBIA | 601,723 | 46.44 | 7.72 |
| 44 | HAWAII | 1,360,301 | 100.8 | 7.41 |
| 45 | NEW JERSEY | 8,791,894 | 566.28 | 6.44 |
| 46 | ILLINOIS | 12,830,632 | 806.4 | 6.28 |
| 47 | MARYLAND | 5,773,552 | 296.64 | 5.14 |
| 48 | CALIFORNIA | 37,253,956 | 1,912.32 | 5.13 |
| 49 | MISSISSIPPI | 2,967,297 | 150.84 | 5.08 |
| 50 | NEVADA | 2,700,551 | 129.24 | 4.79 |
| 51 | NEW YORK | 19,378,102 | 896.4 | 4.63 |
| 52 | VIRGIN ISLANDS | 109,825 | 1.8 | 1.64 |
| 53 | GUAM | 178,430 | 2.88 | 1.61 |
| U.S. GRAMS / PER 100K: | | 312,825,210 | 33,233.04 | 10.62 |

DRUG CODE:9652 DRUG NAME:OXYMORPHONE

| RANK | STATE | 2010 POP | TOTAL GRAMS | GRAMS/100K POP |
|---|---|---|---|---|
| 1 | TENNESSEE | 6,346,105 | 117,287.66 | 1,848.18 |
| 2 | NORTH CAROLINA | 9,535,483 | 135,247.57 | 1,418.36 |
| 3 | DELAWARE | 897,934 | 8,845.71 | 985.12 |
| 4 | NEVADA | 2,700,551 | 25,142.83 | 931.03 |
| 5 | PENNSYLVANIA | 12,702,379 | 114,794.89 | 903.73 |
| 6 | WEST VIRGINIA | 1,852,994 | 16,567.37 | 894.09 |
| 7 | MICHIGAN | 9,883,640 | 81,467.36 | 824.26 |
| 8 | LOUISIANA | 4,533,372 | 36,709.83 | 809.77 |
| 9 | OKLAHOMA | 3,751,351 | 30,020.47 | 800.26 |
| 10 | ALABAMA | 4,779,736 | 37,886.62 | 792.65 |
| 11 | UTAH | 2,763,885 | 20,995.61 | 759.64 |
| 12 | MARYLAND | 5,773,552 | 42,459.23 | 735.41 |
| 13 | SOUTH CAROLINA | 4,625,364 | 33,108.55 | 715.8 |
| 14 | MISSISSIPPI | 2,967,297 | 19,425.57 | 654.66 |
| 15 | VIRGINIA | 8,001,024 | 51,078.13 | 638.39 |
| 16 | FLORIDA | 18,801,310 | 115,425.27 | 613.92 |
| 17 | ARIZONA | 6,392,017 | 38,229.60 | 598.08 |
| 18 | NEW YORK | 19,378,102 | 100,864.58 | 520.51 |
| 19 | MAINE | 1,328,361 | 6,872.81 | 517.39 |
| 20 | INDIANA | 6,483,802 | 31,576.64 | 487.01 |
| 21 | KENTUCKY | 4,339,367 | 20,419.32 | 470.56 |
| 22 | OHIO | 11,536,504 | 53,185.59 | 461.02 |
| 23 | CONNECTICUT | 3,574,097 | 15,867.44 | 443.96 |
| 24 | NEW JERSEY | 8,791,894 | 38,685.52 | 440.01 |
| 25 | IDAHO | 1,567,582 | 6,784.93 | 432.83 |
| 26 | KANSAS | 2,853,118 | 12,229.49 | 428.64 |
| 27 | MISSOURI | 5,988,927 | 24,311.65 | 405.94 |

DRUG CODE:9652 DRUG NAME:OXYMORPHONE

| RANK | STATE | 2010 POP | TOTAL GRAMS | GRAMS/100K POP |
|---|---|---|---|---|
| 28 | SOUTH DAKOTA | 814,180 | 3,176.99 | 390.21 |
| 29 | NEW HAMPSHIRE | 1,316,470 | 5,121.68 | 389.05 |
| 30 | COLORADO | 5,029,196 | 19,121.10 | 380.2 |
| 31 | GEORGIA | 9,687,653 | 34,810.66 | 359.33 |
| 32 | NEBRASKA | 1,826,341 | 6,500.33 | 355.92 |
| 33 | ARKANSAS | 2,915,918 | 10,083.05 | 345.79 |
| 34 | WYOMING | 563,626 | 1,677.28 | 297.59 |
| 35 | NEW MEXICO | 2,059,179 | 6,064.09 | 294.49 |
| 36 | VERMONT | 625,741 | 1,801.95 | 287.97 |
| 37 | TEXAS | 25,145,561 | 64,786.91 | 257.65 |
| 38 | WASHINGTON | 6,724,540 | 16,643.52 | 247.5 |
| 39 | WISCONSIN | 5,686,986 | 13,617.31 | 239.45 |
| 40 | OREGON | 3,831,074 | 8,979.36 | 234.38 |
| 41 | RHODE ISLAND | 1,052,567 | 2,373.90 | 225.53 |
| 42 | CALIFORNIA | 37,253,956 | 81,184.15 | 217.92 |
| 43 | IOWA | 3,046,355 | 6,608.87 | 216.94 |
| 44 | HAWAII | 1,360,301 | 2,875.71 | 211.4 |
| 45 | MONTANA | 989,415 | 1,970.01 | 199.11 |
| 46 | ILLINOIS | 12,830,632 | 24,268.77 | 189.15 |
| 47 | MASSACHUSETTS | 6,547,629 | 11,953.21 | 182.56 |
| 48 | ALASKA | 710,231 | 1,289.75 | 181.6 |
| 49 | NORTH DAKOTA | 672,591 | 829.39 | 123.31 |
| 50 | DISTRICT OF COLUMBIA | 601,723 | 599.55 | 99.64 |
| 51 | MINNESOTA | 5,303,925 | 2,460.96 | 46.4 |
| U.S. GRAMS / PER 100K: | | 312,825,210 | 1,564,288.76 | 500.05 |

DRUG CODE:9668 DRUG NAME:NOROXYMORPHONE

| RANK | STATE | 2010 POP | TOTAL GRAMS | GRAMS/100K POP |
|---|---|---|---|---|
| 1 | ARKANSAS | 2,915,918 | 56.46 | 1.94 |
| 2 | NEVADA | 2,700,551 | 43.13 | 1.6 |
| 3 | DISTRICT OF COLUMBIA | 601,723 | 9.28 | 1.54 |
| 4 | MISSOURI | 5,988,927 | 56.8 | 0.95 |
| 5 | CALIFORNIA | 37,253,956 | 321.85 | 0.86 |
| 6 | MISSISSIPPI | 2,967,297 | 16.18 | 0.55 |
| 7 | FLORIDA | 18,801,310 | 99.67 | 0.53 |
| 8 | PUERTO RICO | 3,725,789 | 18.31 | 0.49 |
| 9 | OHIO | 11,536,504 | 53.3 | 0.46 |
| 10 | ILLINOIS | 12,830,632 | 52.65 | 0.41 |
| 11 | TEXAS | 25,145,561 | 103.87 | 0.41 |
| 12 | RHODE ISLAND | 1,052,567 | 3.34 | 0.32 |
| 13 | WASHINGTON | 6,724,540 | 19.67 | 0.29 |
| 14 | NEW JERSEY | 8,791,894 | 25.6 | 0.29 |
| 15 | GEORGIA | 9,687,653 | 23.72 | 0.24 |
| 16 | NEW YORK | 19,378,102 | 45.28 | 0.23 |
| 17 | NEW HAMPSHIRE | 1,316,470 | 2.78 | 0.21 |
| 18 | VERMONT | 625,741 | 1.13 | 0.18 |
| 19 | NEW MEXICO | 2,059,179 | 3.24 | 0.16 |
| 20 | OREGON | 3,831,074 | 4.2 | 0.11 |
| 21 | ALABAMA | 4,779,736 | 4.38 | 0.09 |
| 22 | NEBRASKA | 1,826,341 | 1.59 | 0.09 |
| 23 | OKLAHOMA | 3,751,351 | 2.81 | 0.07 |
| 24 | UTAH | 2,763,885 | 1.53 | 0.06 |
| 25 | PENNSYLVANIA | 12,702,379 | 6.02 | 0.05 |
| 26 | TENNESSEE | 6,346,105 | 3.03 | 0.05 |
| 27 | MARYLAND | 5,773,552 | 2.46 | 0.04 |
| 28 | ARIZONA | 6,392,017 | 1.69 | 0.03 |

| | | | | |
|---|---|---|---|---|
| 29 | MASSACHUSETTS | 6,547,629 | 2 | 0.03 |
| 30 | CONNECTICUT | 3,574,097 | 1.02 | 0.03 |
| 31 | NORTH CAROLINA | 9,535,483 | 2.25 | 0.02 |
| 32 | INDIANA | 6,483,802 | 1.56 | 0.02 |

DRUG CODE:9668 DRUG NAME:NOROXYMORPHONE

| RANK | STATE | 2010 POP | TOTAL GRAMS | GRAMS/100K POP |
|---|---|---|---|---|
| 33 | KANSAS | 2,853,118 | 0.7 | 0.02 |
| 34 | WISCONSIN | 5,686,986 | 0.67 | 0.01 |
| 35 | COLORADO | 5,029,196 | 0.53 | 0.01 |
| 36 | SOUTH CAROLINA | 4,625,364 | 0.49 | 0.01 |
| 37 | LOUISIANA | 4,533,372 | 0.62 | 0.01 |
| U.S. GRAMS / PER 100K: | | 312,825,210 | 993.83 | 0.32 |

DRUG CODE:9737 DRUG NAME:ALFENTANIL

| RANK | STATE | 2010 POP | TOTAL GRAMS | GRAMS/100K POP |
|---|---|---|---|---|
| 1 | ALASKA | 710,231 | 3.27 | 0.46 |
| 2 | OREGON | 3,831,074 | 15.75 | 0.41 |
| 3 | IDAHO | 1,567,582 | 5.32 | 0.34 |
| 4 | WISCONSIN | 5,686,986 | 18.49 | 0.33 |
| 5 | NEVADA | 2,700,551 | 7.07 | 0.26 |
| 6 | MINNESOTA | 5,303,925 | 13.59 | 0.26 |
| 7 | KANSAS | 2,853,118 | 6.87 | 0.24 |
| 8 | NEW MEXICO | 2,059,179 | 4.78 | 0.23 |
| 9 | IOWA | 3,046,355 | 7.1 | 0.23 |
| 10 | NEBRASKA | 1,826,341 | 4.08 | 0.22 |
| 11 | KENTUCKY | 4,339,367 | 9.05 | 0.21 |
| 12 | MAINE | 1,328,361 | 2.63 | 0.2 |
| 13 | LOUISIANA | 4,533,372 | 9.28 | 0.2 |
| 14 | CALIFORNIA | 37,253,956 | 67.76 | 0.18 |
| 15 | OHIO | 11,536,504 | 21.08 | 0.18 |
| 16 | WASHINGTON | 6,724,540 | 11.67 | 0.17 |
| 17 | MONTANA | 989,415 | 1.55 | 0.16 |
| 18 | INDIANA | 6,483,802 | 9.33 | 0.14 |
| 19 | GUAM | 178,430 | 0.24 | 0.13 |
| 20 | ILLINOIS | 12,830,632 | 16.05 | 0.13 |
| 21 | CONNECTICUT | 3,574,097 | 4.83 | 0.13 |
| 22 | HAWAII | 1,360,301 | 1.78 | 0.13 |
| 23 | NEW HAMPSHIRE | 1,316,470 | 1.57 | 0.12 |
| 24 | NORTH CAROLINA | 9,535,483 | 11.83 | 0.12 |
| 25 | FLORIDA | 18,801,310 | 19.8 | 0.11 |
| 26 | OKLAHOMA | 3,751,351 | 4.07 | 0.11 |
| 27 | TEXAS | 25,145,561 | 23.93 | 0.1 |
| 28 | GEORGIA | 9,687,653 | 8.72 | 0.09 |
| 29 | MISSOURI | 5,988,927 | 5.31 | 0.09 |
| 30 | MASSACHUSETTS | 6,547,629 | 5.25 | 0.08 |
| 31 | UTAH | 2,763,885 | 1.88 | 0.07 |
| 32 | NEW YORK | 19,378,102 | 11.24 | 0.06 |
| 33 | TENNESSEE | 6,346,105 | 3.94 | 0.06 |
| 34 | ARIZONA | 6,392,017 | 3.62 | 0.06 |
| 35 | COLORADO | 5,029,196 | 3.06 | 0.06 |
| 36 | VIRGINIA | 8,001,024 | 3.62 | 0.05 |
| 37 | MICHIGAN | 9,883,640 | 5.11 | 0.05 |
| 38 | MARYLAND | 5,773,552 | 2.3 | 0.04 |
| 39 | SOUTH DAKOTA | 814,180 | 0.23 | 0.03 |

| | | | | |
|---|---|---|---|---|
| 40 | RHODE ISLAND | 1,052,567 | 0.29 | 0.03 |
| 41 | NORTH DAKOTA | 672,591 | 0.22 | 0.03 |
| 42 | PENNSYLVANIA | 12,702,379 | 2.69 | 0.02 |
| 43 | MISSISSIPPI | 2,967,297 | 0.58 | 0.02 |
| 44 | WYOMING | 563,626 | 0.14 | 0.02 |
| 45 | NEW JERSEY | 8,791,894 | 1.12 | 0.01 |
| 46 | ALABAMA | 4,779,736 | 0.25 | 0.01 |
| 47 | ARKANSAS | 2,915,918 | 0.16 | 0.01 |
| 48 | SOUTH CAROLINA | 4,625,364 | 0.19 | 0 |
| 49 | VERMONT | 625,741 | 0.02 | 0 |
| U.S. GRAMS / PER 100K: | | 312,825,210 | 362.62 | 0.12 |

DRUG CODE:9739 DRUG NAME:REMIFENTANIL

DRUG CODE:9739 DRUG NAME:REMIFENTANIL

| RANK | STATE | 2010 POP | TOTAL GRAMS | GRAMS/100K POP |
|---|---|---|---|---|
| 1 | UTAH | 2,763,885 | 66.23 | 2.4 |
| 2 | ALASKA | 710,231 | 11.54 | 1.62 |
| 3 | DELAWARE | 897,934 | 12.7 | 1.41 |
| 4 | MASSACHUSETTS | 6,547,629 | 76.7 | 1.17 |
| 5 | WEST VIRGINIA | 1,852,994 | 20.87 | 1.13 |
| 6 | PENNSYLVANIA | 12,702,379 | 137.57 | 1.08 |
| 7 | CONNECTICUT | 3,574,097 | 32.14 | 0.9 |
| 8 | WASHINGTON | 6,724,540 | 49.56 | 0.74 |
| 9 | NEW YORK | 19,378,102 | 141.16 | 0.73 |
| 10 | MONTANA | 989,415 | 7.2 | 0.73 |
| 11 | OHIO | 11,536,504 | 76.08 | 0.66 |
| 12 | NORTH CAROLINA | 9,535,483 | 56.71 | 0.59 |
| 13 | IDAHO | 1,567,582 | 8.94 | 0.57 |
| 14 | NEW JERSEY | 8,791,894 | 49.62 | 0.56 |
| 15 | FLORIDA | 18,801,310 | 102.98 | 0.55 |
| 16 | DISTRICT OF COLUMBIA | 601,723 | 3.09 | 0.51 |
| 17 | COLORADO | 5,029,196 | 25.25 | 0.5 |
| 18 | MINNESOTA | 5,303,925 | 26.75 | 0.5 |
| 19 | MARYLAND | 5,773,552 | 28.3 | 0.49 |
| 20 | INDIANA | 6,483,802 | 31.78 | 0.49 |
| 21 | ILLINOIS | 12,830,632 | 56.66 | 0.44 |
| 22 | TEXAS | 25,145,561 | 109.37 | 0.43 |
| 23 | SOUTH CAROLINA | 4,625,364 | 19.95 | 0.43 |
| 24 | KENTUCKY | 4,339,367 | 18.46 | 0.43 |
| 25 | WISCONSIN | 5,686,986 | 24.29 | 0.43 |
| 26 | MICHIGAN | 9,883,640 | 41.11 | 0.42 |
| 27 | MAINE | 1,328,361 | 5.12 | 0.39 |
| 28 | HAWAII | 1,360,301 | 5.33 | 0.39 |
| 29 | IOWA | 3,046,355 | 11.55 | 0.38 |
| 30 | LOUISIANA | 4,533,372 | 16.99 | 0.37 |
| 31 | NEW HAMPSHIRE | 1,316,470 | 4.72 | 0.36 |
| 32 | CALIFORNIA | 37,253,956 | 129.31 | 0.35 |
| 33 | MISSOURI | 5,988,927 | 20.99 | 0.35 |
| 34 | VERMONT | 625,741 | 2.17 | 0.35 |
| 35 | WYOMING | 563,626 | 1.88 | 0.33 |
| 36 | TENNESSEE | 6,346,105 | 20.73 | 0.33 |
| 37 | VIRGINIA | 8,001,024 | 24.91 | 0.31 |
| 38 | OREGON | 3,831,074 | 11.3 | 0.29 |
| 39 | SOUTH DAKOTA | 814,180 | 2.34 | 0.29 |

| | | | | |
|---|---|---|---|---|
| 40 | GUAM | 178,430 | 0.5 | 0.28 |
| 41 | KANSAS | 2,853,118 | 7.97 | 0.28 |
| 42 | ALABAMA | 4,779,736 | 12.64 | 0.26 |
| 43 | ARIZONA | 6,392,017 | 16.86 | 0.26 |
| 44 | OKLAHOMA | 3,751,351 | 9.83 | 0.26 |
| 45 | NORTH DAKOTA | 672,591 | 1.59 | 0.24 |
| 46 | ARKANSAS | 2,915,918 | 6.9 | 0.24 |
| 47 | NEBRASKA | 1,826,341 | 3.97 | 0.22 |
| 48 | NEW MEXICO | 2,059,179 | 4.18 | 0.2 |
| 49 | GEORGIA | 9,687,653 | 18.95 | 0.2 |
| 50 | RHODE ISLAND | 1,052,567 | 1.85 | 0.18 |
| 51 | NEVADA | 2,700,551 | 3.53 | 0.13 |
| 52 | MISSISSIPPI | 2,967,297 | 1.94 | 0.07 |
| 53 | PUERTO RICO | 3,725,789 | 0.16 | 0 |
| U.S. GRAMS / PER 100K: | | 312,825,210 | 1,583.20 | 0.51 |

DRUG CODE:9740 DRUG NAME:SUFENTANIL BASE

| RANK | STATE | 2010 POP | TOTAL GRAMS | GRAMS/100K POP |
|---|---|---|---|---|
| 1 | MISSISSIPPI | 2,967,297 | 41.85 | 1.41 |
| 2 | ALABAMA | 4,779,736 | 7.89 | 0.16 |
| 3 | CALIFORNIA | 37,253,956 | 39.15 | 0.11 |

DRUG CODE:9740 DRUG NAME:SUFENTANIL BASE

| RANK | STATE | 2010 POP | TOTAL GRAMS | GRAMS/100K POP |
|---|---|---|---|---|
| 4 | LOUISIANA | 4,533,372 | 3.42 | 0.08 |
| 5 | FLORIDA | 18,801,310 | 10.44 | 0.06 |
| 6 | WISCONSIN | 5,686,986 | 3.38 | 0.06 |
| 7 | SOUTH CAROLINA | 4,625,364 | 2.5 | 0.05 |
| 8 | NEW JERSEY | 8,791,894 | 4.44 | 0.05 |
| 9 | KANSAS | 2,853,118 | 1.02 | 0.04 |
| 10 | CONNECTICUT | 3,574,097 | 1.31 | 0.04 |
| 11 | PENNSYLVANIA | 12,702,379 | 5.22 | 0.04 |
| 12 | NEW YORK | 19,378,102 | 7.2 | 0.04 |
| 13 | TENNESSEE | 6,346,105 | 1.59 | 0.03 |
| 14 | MISSOURI | 5,988,927 | 1.51 | 0.03 |
| 15 | UTAH | 2,763,885 | 0.87 | 0.03 |
| 16 | OREGON | 3,831,074 | 1.31 | 0.03 |
| 17 | KENTUCKY | 4,339,367 | 1.2 | 0.03 |
| 18 | TEXAS | 25,145,561 | 6.92 | 0.03 |
| 19 | VIRGINIA | 8,001,024 | 1.58 | 0.02 |
| 20 | INDIANA | 6,483,802 | 1.51 | 0.02 |
| 21 | NORTH DAKOTA | 672,591 | 0.1 | 0.02 |
| 22 | SOUTH DAKOTA | 814,180 | 0.14 | 0.02 |
| 23 | MICHIGAN | 9,883,640 | 2.34 | 0.02 |
| 24 | ILLINOIS | 12,830,632 | 2.15 | 0.02 |
| 25 | COLORADO | 5,029,196 | 0.82 | 0.02 |
| 26 | ARKANSAS | 2,915,918 | 0.64 | 0.02 |
| 27 | GEORGIA | 9,687,653 | 1.47 | 0.02 |
| 28 | OHIO | 11,536,504 | 2.88 | 0.02 |
| 29 | MONTANA | 989,415 | 0.18 | 0.02 |
| 30 | OKLAHOMA | 3,751,351 | 0.75 | 0.02 |
| 31 | PUERTO RICO | 3,725,789 | 0.76 | 0.02 |
| 32 | NEBRASKA | 1,826,341 | 0.39 | 0.02 |
| 33 | MINNESOTA | 5,303,925 | 0.8 | 0.02 |
| 34 | NORTH CAROLINA | 9,535,483 | 2.23 | 0.02 |

| | | | | |
|---|---|---|---|---|
| 35 | NEW HAMPSHIRE | 1,316,470 | 0.07 | 0.01 |
| 36 | NEW MEXICO | 2,059,179 | 0.17 | 0.01 |
| 37 | WYOMING | 563,626 | 0.04 | 0.01 |
| 38 | IDAHO | 1,567,582 | 0.17 | 0.01 |
| 39 | MASSACHUSETTS | 6,547,629 | 0.79 | 0.01 |
| 40 | WEST VIRGINIA | 1,852,994 | 0.21 | 0.01 |
| 41 | WASHINGTON | 6,724,540 | 0.98 | 0.01 |
| 42 | DISTRICT OF COLUMBIA | 601,723 | 0.04 | 0.01 |
| 43 | VERMONT | 625,741 | 0.04 | 0.01 |
| 44 | NEVADA | 2,700,551 | 0.36 | 0.01 |
| 45 | MAINE | 1,328,361 | 0.03 | 0 |
| 46 | MARYLAND | 5,773,552 | 0.15 | 0 |
| 47 | RHODE ISLAND | 1,052,567 | 0.03 | 0 |
| 48 | DELAWARE | 897,934 | 0.04 | 0 |
| 49 | ALASKA | 710,231 | 0.03 | 0 |
| 50 | IOWA | 3,046,355 | 0.15 | 0 |
| 51 | ARIZONA | 6,392,017 | 0.29 | 0 |
| 52 | HAWAII | 1,360,301 | 0.05 | 0 |
| U.S. GRAMS / PER 100K: | | 312,825,210 | 163.58 | 0.05 |

DRUG CODE:9780 DRUG NAME:TAPENTADOL

| RANK | STATE | 2010 POP | TOTAL GRAMS | GRAMS/100K POP |
|---|---|---|---|---|
| 1 | ALABAMA | 4,779,736 | 262,860 | 5,499.47 |
| 2 | NORTH CAROLINA | 9,535,483 | 412,847.75 | 4,329.59 |
| 3 | SOUTH CAROLINA | 4,625,364 | 180,623.70 | 3,905.07 |
| 4 | ARKANSAS | 2,915,918 | 107,609.35 | 3,690.41 |
| 5 | DELAWARE | 897,934 | 27,246 | 3,034.30 |
| 6 | GEORGIA | 9,687,653 | 288,643.50 | 2,979.50 |
| 7 | SOUTH DAKOTA | 814,180 | 23,500.50 | 2,886.40 |

DRUG CODE:9780 DRUG NAME:TAPENTADOL

| RANK | STATE | 2010 POP | TOTAL GRAMS | GRAMS/100K POP |
|---|---|---|---|---|
| 8 | OKLAHOMA | 3,751,351 | 103,680.85 | 2,763.83 |
| 9 | INDIANA | 6,483,802 | 161,355.50 | 2,488.59 |
| 10 | FLORIDA | 18,801,310 | 465,683.29 | 2,476.87 |
| 11 | VIRGINIA | 8,001,024 | 196,186.75 | 2,452.02 |
| 12 | OHIO | 11,536,504 | 282,247.35 | 2,446.56 |
| 13 | MARYLAND | 5,773,552 | 140,743 | 2,437.72 |
| 14 | UTAH | 2,763,885 | 64,892.50 | 2,347.87 |
| 15 | ARIZONA | 6,392,017 | 149,775.35 | 2,343.16 |
| 16 | NEW JERSEY | 8,791,894 | 203,502 | 2,314.65 |
| 17 | NEBRASKA | 1,826,341 | 42,231.50 | 2,312.36 |
| 18 | TENNESSEE | 6,346,105 | 143,016.50 | 2,253.61 |
| 19 | PENNSYLVANIA | 12,702,379 | 283,016.20 | 2,228.06 |
| 20 | NEW YORK | 19,378,102 | 431,274.50 | 2,225.58 |
| 21 | COLORADO | 5,029,196 | 110,484 | 2,196.85 |
| 22 | NEW HAMPSHIRE | 1,316,470 | 26,171 | 1,987.97 |
| 23 | MISSISSIPPI | 2,967,297 | 57,785.50 | 1,947.41 |
| 24 | MISSOURI | 5,988,927 | 115,417.50 | 1,927.18 |
| 25 | NEVADA | 2,700,551 | 51,467 | 1,905.80 |
| 26 | IOWA | 3,046,355 | 52,676 | 1,729.15 |
| 27 | TEXAS | 25,145,561 | 432,221.29 | 1,718.88 |
| 28 | KANSAS | 2,853,118 | 45,469.50 | 1,593.68 |
| 29 | IDAHO | 1,567,582 | 24,727.50 | 1,577.43 |
| 30 | NORTH DAKOTA | 672,591 | 10,425.50 | 1,550.05 |

| | | | | |
|---|---|---|---|---|
| 31 | KENTUCKY | 4,339,367 | 66,065.30 | 1,522.46 |
| 32 | CONNECTICUT | 3,574,097 | 53,900.30 | 1,508.08 |
| 33 | LOUISIANA | 4,533,372 | 65,829.50 | 1,452.11 |
| 34 | ILLINOIS | 12,830,632 | 179,709.50 | 1,400.63 |
| 35 | WYOMING | 563,626 | 7,478 | 1,326.77 |
| 36 | WEST VIRGINIA | 1,852,994 | 24,356 | 1,314.41 |
| 37 | CALIFORNIA | 37,253,956 | 480,772.94 | 1,290.53 |
| 38 | WISCONSIN | 5,686,986 | 71,791.35 | 1,262.38 |
| 39 | WASHINGTON | 6,724,540 | 78,105 | 1,161.49 |
| 40 | MASSACHUSETTS | 6,547,629 | 73,227.50 | 1,118.38 |
| 41 | ALASKA | 710,231 | 7,749.50 | 1,091.12 |
| 42 | MONTANA | 989,415 | 10,108.50 | 1,021.66 |
| 43 | MICHIGAN | 9,883,640 | 95,596 | 967.21 |
| 44 | DISTRICT OF COLUMBIA | 601,723 | 5,745.35 | 954.82 |
| 45 | VERMONT | 625,741 | 5,498 | 878.64 |
| 46 | RHODE ISLAND | 1,052,567 | 9,191.50 | 873.25 |
| 47 | NEW MEXICO | 2,059,179 | 17,727 | 860.88 |
| 48 | OREGON | 3,831,074 | 32,113.50 | 838.24 |
| 49 | HAWAII | 1,360,301 | 11,071.50 | 813.9 |
| 50 | MAINE | 1,328,361 | 8,210.50 | 618.09 |
| 51 | MINNESOTA | 5,303,925 | 20,361.50 | 383.89 |
| 52 | PUERTO RICO | 3,725,789 | 4,253.47 | 114.16 |
| 53 | VIRGIN ISLANDS | 109,825 | 44 | 40.06 |
| U.S. GRAMS / PER 100K: | | 312,825,210 | 6,226,686.59 | 1,990.47 |

DRUG CODE:9801 DRUG NAME:FENTANYL BASE

| RANK | STATE | 2010 POP | TOTAL GRAMS | GRAMS/100K POP |
|---|---|---|---|---|
| 1 | OKLAHOMA | 3,751,351 | 8,091.43 | 215.69 |
| 2 | PENNSYLVANIA | 12,702,379 | 23,884.22 | 188.03 |
| 3 | MISSISSIPPI | 2,967,297 | 5,487.89 | 184.95 |
| 4 | DELAWARE | 897,934 | 1,623.51 | 180.81 |
| 5 | NORTH DAKOTA | 672,591 | 1,200.56 | 178.5 |
| 6 | KANSAS | 2,853,118 | 5,070.86 | 177.73 |
| 7 | SOUTH DAKOTA | 814,180 | 1,438.30 | 176.66 |
| 8 | IDAHO | 1,567,582 | 2,701.89 | 172.36 |
| 9 | MISSOURI | 5,988,927 | 10,134.51 | 169.22 |
| 10 | MONTANA | 989,415 | 1,608.43 | 162.56 |

DRUG CODE:9801 DRUG NAME:FENTANYL BASE

| RANK | STATE | 2010 POP | TOTAL GRAMS | GRAMS/100K POP |
|---|---|---|---|---|
| 11 | ALABAMA | 4,779,736 | 7,558.52 | 158.14 |
| 12 | ARKANSAS | 2,915,918 | 4,554.72 | 156.2 |
| 13 | MAINE | 1,328,361 | 2,067.66 | 155.66 |
| 14 | VERMONT | 625,741 | 965.47 | 154.29 |
| 15 | TENNESSEE | 6,346,105 | 9,619.86 | 151.59 |
| 16 | MICHIGAN | 9,883,640 | 14,931.75 | 151.08 |
| 17 | ALASKA | 710,231 | 1,059.96 | 149.24 |
| 18 | NORTH CAROLINA | 9,535,483 | 14,218.40 | 149.11 |
| 19 | WEST VIRGINIA | 1,852,994 | 2,673.51 | 144.28 |
| 20 | INDIANA | 6,483,802 | 9,307.01 | 143.54 |
| 21 | FLORIDA | 18,801,310 | 26,916.60 | 143.16 |
| 22 | ARIZONA | 6,392,017 | 9,138.94 | 142.97 |
| 23 | UTAH | 2,763,885 | 3,918.46 | 141.77 |
| 24 | SOUTH CAROLINA | 4,625,364 | 6,259.16 | 135.32 |
| 25 | WISCONSIN | 5,686,986 | 7,605.81 | 133.74 |

CUMULATIVE DISTRIBUTION BY STATE IN GRAMS PER 100,000 POPULATION

| | | | | |
|---|---|---|---|---|
| 26 | NEW HAMPSHIRE | 1,316,470 | 1,755.75 | 133.37 |
| 27 | CONNECTICUT | 3,574,097 | 4,708.25 | 131.73 |
| 28 | NEBRASKA | 1,826,341 | 2,391.96 | 130.97 |
| 29 | NEW JERSEY | 8,791,894 | 11,494.86 | 130.74 |
| 30 | IOWA | 3,046,355 | 3,967.67 | 130.24 |
| 31 | WYOMING | 563,626 | 728.68 | 129.28 |
| 32 | GEORGIA | 9,687,653 | 12,501.23 | 129.04 |
| 33 | OREGON | 3,831,074 | 4,878.87 | 127.35 |
| 34 | KENTUCKY | 4,339,367 | 5,464.31 | 125.92 |
| 35 | COLORADO | 5,029,196 | 6,254.30 | 124.36 |
| 36 | LOUISIANA | 4,533,372 | 5,623.70 | 124.05 |
| 37 | MARYLAND | 5,773,552 | 7,045.01 | 122.02 |
| 38 | NEW YORK | 19,378,102 | 21,926.26 | 113.15 |
| 39 | NEVADA | 2,700,551 | 2,977.95 | 110.27 |
| 40 | TEXAS | 25,145,561 | 27,630.29 | 109.88 |
| 41 | WASHINGTON | 6,724,540 | 7,356.30 | 109.39 |
| 42 | NEW MEXICO | 2,059,179 | 2,243.27 | 108.94 |
| 43 | ILLINOIS | 12,830,632 | 13,851.40 | 107.96 |
| 44 | VIRGINIA | 8,001,024 | 8,595.21 | 107.43 |
| 45 | OHIO | 11,536,504 | 12,373.47 | 107.25 |
| 46 | MINNESOTA | 5,303,925 | 5,037.09 | 94.97 |
| 47 | MASSACHUSETTS | 6,547,629 | 6,133.24 | 93.67 |
| 48 | CALIFORNIA | 37,253,956 | 34,415.70 | 92.38 |
| 49 | RHODE ISLAND | 1,052,567 | 922.24 | 87.62 |
| 50 | DISTRICT OF COLUMBIA | 601,723 | 397.68 | 66.09 |
| 51 | HAWAII | 1,360,301 | 847.62 | 62.31 |
| 52 | VIRGIN ISLANDS | 109,825 | 26.7 | 24.31 |
| 53 | PUERTO RICO | 3,725,789 | 862.36 | 23.15 |
| 54 | GUAM | 178,430 | 38.56 | 21.61 |
| 55 | AMERICAN SAMOA | 65,628 | 0.55 | 0.84 |
| U.S. GRAMS / PER 100K: | | 312,825,210 | 394,487.93 | 126.1 |

STATISTICAL SUMMARY FOR RETAIL DRUG PURCHASES BY GRAMS WT

STATE:ALABAMA BUSINESS ACTIVITY:A - PHARMACIES

| DRUG NAME | DRUG CODE | BUYERS | TOTAL GRAMS | AVG GRAMS |
|---|---|---|---|---|
| AMPHETAMINE | 1100 | 1,365 | 499,133.36 | 365.67 |
| DL-METHAMPHETAMINE RACEMIC BASE | 1105B | 9 | 32.95 | 3.66 |
| D-METHAMPHETAMINE | 1105D | 27 | 86.77 | 3.21 |
| LISDEXAMFETAMINE | 1205 | 1,331 | 281,761.32 | 211.69 |
| METHYLPHENIDATE (DL;D;L;ISOMERS) | 1724 | 1,343 | 283,227.95 | 210.89 |
| BARBITURIC ACID DERIVIATIVE OR SALT [PER 21C | 2100 | 7 | 45 | 6.43 |
| BUTALBITAL | 2165 | 846 | 28,350 | 33.51 |
| CODEINE | 9050 | 1,355 | 209,741.85 | 154.79 |
| BUPRENORPHINE | 9064 | 1,295 | 89,762.30 | 69.31 |
| DIHYDROCODEINE | 9120 | 529 | 2,665.06 | 5.04 |
| OXYCODONE | 9143 | 1,374 | 1,022,679.04 | 744.31 |
| HYDROMORPHONE | 9150 | 1,160 | 12,108.90 | 10.44 |
| HYDROCODONE | 9193 | 1,393 | 1,045,556.13 | 750.58 |
| LEVORPHANOL | 9220L | 21 | 19.85 | 0.95 |
| MEPERIDINE (PETHIDINE) | 9230 | 978 | 23,217.63 | 23.74 |
| METHADONE | 9250B | 1,190 | 94,384.51 | 79.31 |
| MORPHINE | 9300 | 1,338 | 306,154.66 | 228.82 |
| OPIUM POWDERED | 9639 | 60 | 131.4 | 2.19 |
| OXYMORPHONE | 9652 | 999 | 37,764.33 | 37.8 |
| TAPENTADOL | 9780 | 1,162 | 261,324 | 224.89 |
| FENTANYL BASE | 9801 | 1,318 | 6,938.80 | 5.26 |

STATE:ALABAMA BUSINESS ACTIVITY:B - HOSPITALS

| DRUG NAME | DRUG CODE | BUYERS | TOTAL GRAMS | AVG GRAMS |
|---|---|---|---|---|
| AMPHETAMINE | 1100 | 70 | 8,261.94 | 118.03 |
| LISDEXAMFETAMINE | 1205 | 28 | 3,281.30 | 117.19 |
| METHYLPHENIDATE (DL;D;L;ISOMERS) | 1724 | 80 | 7,585.24 | 94.82 |
| GAMMA HYDROXYBUTYRIC ACID(FDA APPROVED | 2012 | 1 | 371.52 | 371.52 |
| AMOBARBITAL (SCHEDULE 2) | 2125 | 1 | 5.47 | 5.47 |

| | | | | |
|---|---|---|---|---|
| BUTALBITAL | 2165 | 8 | 85 | 10.63 |
| PENTOBARBITAL (SCHEDULE 2) | 2270 | 6 | 291.65 | 48.61 |
| COCAINE | 9041L | 43 | 160.85 | 3.74 |
| CODEINE | 9050 | 122 | 4,771.02 | 39.11 |
| BUPRENORPHINE | 9064 | 86 | 640.89 | 7.45 |
| OXYCODONE | 9143 | 140 | 35,674.15 | 254.82 |
| HYDROMORPHONE | 9150 | 127 | 3,122.07 | 24.58 |
| HYDROCODONE | 9193 | 145 | 52,073.42 | 359.13 |
| MEPERIDINE (PETHIDINE) | 9230 | 121 | 20,685.03 | 170.95 |
| METHADONE | 9250B | 113 | 4,553.82 | 40.3 |
| MORPHINE | 9300 | 144 | 31,601.15 | 219.45 |
| OPIUM POWDERED | 9639 | 49 | 242.28 | 4.94 |
| OXYMORPHONE | 9652 | 17 | 120.7 | 7.1 |
| ALFENTANIL | 9737 | 3 | 0.25 | 0.08 |
| REMIFENTANIL | 9739 | 17 | 10.65 | 0.63 |
| SUFENTANIL BASE | 9740 | 35 | 1.61 | 0.05 |
| TAPENTADOL | 9780 | 29 | 1,536 | 52.97 |
| FENTANYL BASE | 9801 | 132 | 568.61 | 4.31 |

STATE:ALABAMA BUSINESS ACTIVITY:C - PRACTITIONERS

| DRUG NAME | DRUG CODE | BUYERS | TOTAL GRAMS | AVG GRAMS |
|---|---|---|---|---|
| METHYLPHENIDATE (DL;D;L;ISOMERS) | 1724 | 1 | 47.04 | 47.04 |
| PENTOBARBITAL (SCHEDULE 2) | 2270 | 73 | 46,386.23 | 635.43 |
| COCAINE | 9041L | 11 | 87.14 | 7.92 |
| CODEINE | 9050 | 129 | 3,076.42 | 23.85 |
| BUPRENORPHINE | 9064 | 248 | 113.27 | 0.46 |
| DIHYDROCODEINE | 9120 | 266 | 157.46 | 0.59 |
| OXYCODONE | 9143 | 40 | 4,593.47 | 114.84 |
| HYDROMORPHONE | 9150 | 114 | 275.65 | 2.42 |
| HYDROCODONE | 9193 | 142 | 2,820.46 | 19.86 |
| MEPERIDINE (PETHIDINE) | 9230 | 130 | 2,219.82 | 17.08 |
| METHADONE | 9250B | 7 | 58.87 | 8.41 |

| | | | | |
|---|---|---|---|---|
| MORPHINE | 9300 | 147 | 1,113.74 | 7.58 |
| OPIUM POWDERED | 9639 | 2 | 5.4 | 2.7 |
| OXYMORPHONE | 9652 | 2 | 1.59 | 0.79 |
| NOROXYMORPHONE | 9668 | 3 | 4.38 | 1.46 |
| REMIFENTANIL | 9739 | 1 | 1.99 | 1.99 |
| SUFENTANIL BASE | 9740 | 9 | 6.27 | 0.7 |
| FENTANYL BASE | 9801 | 213 | 50.94 | 0.24 |

STATE:ALABAMA BUSINESS ACTIVITY:D - TEACHING INSTITUTIONS

| DRUG NAME | DRUG CODE | BUYERS | TOTAL GRAMS | AVG GRAMS |
|---|---|---|---|---|
| BUPRENORPHINE | 9064 | 1 | 0.03 | 0.03 |
| HYDROMORPHONE | 9150 | 1 | 1.06 | 1.06 |
| MORPHINE | 9300 | 1 | 0.38 | 0.38 |
| FENTANYL BASE | 9801 | 1 | 0.17 | 0.17 |

STATE:ALABAMA BUSINESS ACTIVITY:M - MID-LEVEL PRACTITIONERS

| DRUG NAME | DRUG CODE | BUYERS | TOTAL GRAMS | AVG GRAMS |
|---|---|---|---|---|
| PENTOBARBITAL (SCHEDULE 2) | 2270 | 20 | 30,035.19 | 1,501.76 |
| CODEINE | 9050 | 6 | 3.51 | 0.58 |
| DIHYDROCODEINE | 9120 | 17 | 9.12 | 0.54 |

STATE:ALABAMA BUSINESS ACTIVITY:N-U NARCOTIC TREATMENT PROGRAMS

| DRUG NAME | DRUG CODE | BUYERS | TOTAL GRAMS | AVG GRAMS |
|---|---|---|---|---|
| BUPRENORPHINE | 9064 | 9 | 313.98 | 34.89 |
| METHADONE | 9250B | 22 | 237,028.52 | 10,774.02 |

STATE:ALASKA BUSINESS ACTIVITY:A - PHARMACIES

| DRUG NAME | DRUG CODE | BUYERS | TOTAL GRAMS | AVG GRAMS |
|---|---|---|---|---|
| AMPHETAMINE | 1100 | 110 | 25,025.91 | 227.51 |
| DL-METHAMPHETAMINE RACEMIC BASE | 1105B | 1 | 0.8 | 0.8 |
| D-METHAMPHETAMINE | 1105D | 3 | 12.45 | 4.15 |
| LISDEXAMFETAMINE | 1205 | 100 | 8,990.61 | 89.91 |
| METHYLPHENIDATE (DL;D;L;ISOMERS) | 1724 | 111 | 24,855.55 | 223.92 |
| BARBITURIC ACID DERIVIATIVE OR SALT [PER 21C | 2100 | 3 | 60 | 20 |
| BUTALBITAL | 2165 | 55 | 1,490 | 27.09 |
| COCAINE | 9041L | 1 | 0.29 | 0.29 |
| CODEINE | 9050 | 110 | 21,039.73 | 191.27 |
| BUPRENORPHINE | 9064 | 106 | 7,132.59 | 67.29 |
| DIHYDROCODEINE | 9120 | 1 | 1.07 | 1.07 |
| OXYCODONE | 9143 | 114 | 133,418.43 | 1,170.34 |
| HYDROMORPHONE | 9150 | 103 | 3,583.19 | 34.79 |
| HYDROCODONE | 9193 | 113 | 45,929.16 | 406.45 |
| MEPERIDINE (PETHIDINE) | 9230 | 69 | 2,738.91 | 39.69 |
| METHADONE | 9250B | 99 | 18,009.73 | 181.92 |
| MORPHINE | 9300 | 111 | 43,658.60 | 393.32 |
| OPIUM POWDERED | 9639 | 10 | 15.12 | 1.51 |
| OXYMORPHONE | 9652 | 42 | 1,264.33 | 30.1 |
| ALFENTANIL | 9737 | 2 | 1.01 | 0.51 |
| REMIFENTANIL | 9739 | 3 | 0.38 | 0.13 |
| TAPENTADOL | 9780 | 63 | 7,144.50 | 113.4 |
| FENTANYL BASE | 9801 | 111 | 840.21 | 7.57 |

STATE:ALASKA BUSINESS ACTIVITY:B - HOSPITALS

| DRUG NAME | DRUG CODE | BUYERS | TOTAL GRAMS | AVG GRAMS |
|---|---|---|---|---|
| AMPHETAMINE | 1100 | 26 | 6,301.59 | 242.37 |
| LISDEXAMFETAMINE | 1205 | 17 | 793.73 | 46.69 |
| METHYLPHENIDATE (DL;D;L;ISOMERS) | 1724 | 31 | 7,963.04 | 256.87 |
| BUTALBITAL | 2165 | 1 | 5 | 5 |

| | | | | |
|---|---|---|---|---|
| PENTOBARBITAL (SCHEDULE 2) | 2270 | 2 | 1,313.33 | 656.66 |
| COCAINE | 9041L | 12 | 50.28 | 4.19 |
| CODEINE | 9050 | 40 | 8,695.66 | 217.39 |
| BUPRENORPHINE | 9064 | 28 | 508.76 | 18.17 |
| OXYCODONE | 9143 | 47 | 16,824.65 | 357.97 |
| HYDROMORPHONE | 9150 | 37 | 674.64 | 18.23 |
| HYDROCODONE | 9193 | 50 | 6,677.63 | 133.55 |
| MEPERIDINE (PETHIDINE) | 9230 | 33 | 519.09 | 15.73 |
| METHADONE | 9250B | 25 | 1,351.08 | 54.04 |
| MORPHINE | 9300 | 47 | 9,694.70 | 206.27 |
| OPIUM POWDERED | 9639 | 13 | 62.64 | 4.82 |
| OXYMORPHONE | 9652 | 3 | 15.79 | 5.26 |
| ALFENTANIL | 9737 | 2 | 2.07 | 1.04 |
| REMIFENTANIL | 9739 | 15 | 10.65 | 0.71 |
| SUFENTANIL BASE | 9740 | 6 | 0.03 | 0.01 |
| TAPENTADOL | 9780 | 6 | 605 | 100.83 |
| FENTANYL BASE | 9801 | 48 | 207.25 | 4.32 |

STATE:ALASKA BUSINESS ACTIVITY:C - PRACTITIONERS

| DRUG NAME | DRUG CODE | BUYERS | TOTAL GRAMS | AVG GRAMS |
|---|---|---|---|---|
| PENTOBARBITAL (SCHEDULE 2) | 2270 | 26 | 16,359.63 | 629.22 |
| COCAINE | 9041L | 2 | 12.71 | 6.36 |
| CODEINE | 9050 | 9 | 67 | 7.44 |
| BUPRENORPHINE | 9064 | 54 | 6.59 | 0.12 |
| OXYCODONE | 9143 | 8 | 85.61 | 10.7 |
| HYDROMORPHONE | 9150 | 46 | 48.97 | 1.06 |
| HYDROCODONE | 9193 | 44 | 218.03 | 4.96 |
| MEPERIDINE (PETHIDINE) | 9230 | 12 | 49.89 | 4.16 |
| METHADONE | 9250B | 2 | 1.43 | 0.72 |
| MORPHINE | 9300 | 40 | 54.61 | 1.37 |
| OXYMORPHONE | 9652 | 1 | 9.63 | 9.63 |
| ALFENTANIL | 9737 | 2 | 0.19 | 0.1 |

| | | | | |
|---|---|---|---|---|
| REMIFENTANIL | 9739 | 5 | 0.47 | 0.09 |
| FENTANYL BASE | 9801 | 73 | 12.5 | 0.17 |

STATE:ALASKA BUSINESS ACTIVITY:M - MID-LEVEL PRACTITIONERS

| DRUG NAME | DRUG CODE | BUYERS | TOTAL GRAMS | AVG GRAMS |
|---|---|---|---|---|
| PENTOBARBITAL (SCHEDULE 2) | 2270 | 2 | 266.58 | 133.29 |
| CODEINE | 9050 | 2 | 12.16 | 6.08 |
| OXYCODONE | 9143 | 2 | 2.24 | 1.12 |
| HYDROCODONE | 9193 | 4 | 6.96 | 1.74 |
| MEPERIDINE (PETHIDINE) | 9230 | 1 | 1.74 | 1.74 |
| MORPHINE | 9300 | 3 | 0.32 | 0.11 |
| REMIFENTANIL | 9739 | 3 | 0.04 | 0.01 |
| FENTANYL BASE | 9801 | 2 | 0 | 0 |

STATE:ALASKA BUSINESS ACTIVITY:N-U NARCOTIC TREATMENT PROGRAMS

| DRUG NAME | DRUG CODE | BUYERS | TOTAL GRAMS | AVG GRAMS |
|---|---|---|---|---|
| BUPRENORPHINE | 9064 | 1 | 42.96 | 42.96 |
| METHADONE | 9250B | 4 | 12,843.58 | 3,210.90 |

STATE:AMERICAN SAMOA BUSINESS ACTIVITY:B - HOSPITALS

| DRUG NAME | DRUG CODE | BUYERS | TOTAL GRAMS | AVG GRAMS |
|---|---|---|---|---|
| AMPHETAMINE | 1100 | 1 | 6.54 | 6.54 |
| METHYLPHENIDATE (DL;D;L;ISOMERS) | 1724 | 1 | 14.7 | 14.7 |
| CODEINE | 9050 | 1 | 2,150.43 | 2,150.43 |
| OXYCODONE | 9143 | 1 | 133.13 | 133.13 |
| HYDROMORPHONE | 9150 | 1 | 3.16 | 3.16 |
| HYDROCODONE | 9193 | 1 | 32.69 | 32.69 |
| MEPERIDINE (PETHIDINE) | 9230 | 1 | 6.1 | 6.1 |

| | | | | |
|---|---|---|---|---|
| MORPHINE | 9300 | 1 | 148.9 | 148.9 |
| FENTANYL BASE | 9801 | 1 | 0.55 | 0.55 |

STATE:AMERICAN SAMOA BUSINESS ACTIVITY:C - PRACTITIONERS

| DRUG NAME | DRUG CODE | BUYERS | TOTAL GRAMS | AVG GRAMS |
|---|---|---|---|---|
| BUPRENORPHINE | 9064 | 2 | 0.01 | 0.01 |

STATE:ARIZONA BUSINESS ACTIVITY:A - PHARMACIES

| DRUG NAME | DRUG CODE | BUYERS | TOTAL GRAMS | AVG GRAMS |
|---|---|---|---|---|
| AMPHETAMINE | 1100 | 1,102 | 296,052.47 | 268.65 |
| DL-METHAMPHETAMINE RACEMIC BASE | 1105B | 12 | 83.99 | 7 |
| D-METHAMPHETAMINE | 1105D | 20 | 67.08 | 3.35 |
| LISDEXAMFETAMINE | 1205 | 1,037 | 134,111.68 | 129.33 |
| METHYLPHENIDATE (DL;D;L;ISOMERS) | 1724 | 1,096 | 277,369.65 | 253.07 |
| BARBITURIC ACID DERIVIATIVE OR SALT [PER 21C | 2100 | 16 | 580 | 36.25 |
| BUTALBITAL | 2165 | 840 | 47,260 | 56.26 |
| PENTOBARBITAL (SCHEDULE 2) | 2270 | 1 | 88.86 | 88.86 |
| SECOBARBITAL (SCHEDULE 2) | 2315 | 4 | 36.62 | 9.16 |
| NABILONE | 7379 | 1 | 0.15 | 0.15 |
| COCAINE | 9041L | 3 | 15.46 | 5.15 |
| CODEINE | 9050 | 1,108 | 286,938.76 | 258.97 |
| BUPRENORPHINE | 9064 | 1,050 | 30,734.22 | 29.27 |
| DIHYDROCODEINE | 9120 | 15 | 212.56 | 14.17 |
| OXYCODONE | 9143 | 1,125 | 1,879,425.14 | 1,670.60 |
| HYDROMORPHONE | 9150 | 1,057 | 28,104.77 | 26.59 |
| HYDROCODONE | 9193 | 1,125 | 439,249.04 | 390.44 |
| LEVORPHANOL | 9220L | 24 | 39.58 | 1.65 |
| MEPERIDINE (PETHIDINE) | 9230 | 635 | 15,822.08 | 24.92 |
| METHADONE | 9250B | 976 | 63,291.23 | 64.85 |
| MORPHINE | 9300 | 1,122 | 728,473.26 | 649.26 |

| | | | | |
|---|---|---|---|---|
| OPIUM TINCTURE (LAUDANUM) | 9630 | 1 | 9.46 | 9.46 |
| OPIUM POWDERED | 9639 | 121 | 570.24 | 4.71 |
| OXYMORPHONE | 9652 | 961 | 38,116.49 | 39.66 |
| REMIFENTANIL | 9739 | 1 | 0.03 | 0.03 |
| TAPENTADOL | 9780 | 885 | 149,351.50 | 168.76 |
| FENTANYL BASE | 9801 | 1,073 | 8,428.55 | 7.86 |

STATE:ARIZONA BUSINESS ACTIVITY:B - HOSPITALS

| DRUG NAME | DRUG CODE | BUYERS | TOTAL GRAMS | AVG GRAMS |
|---|---|---|---|---|
| AMPHETAMINE | 1100 | 93 | 7,841.46 | 84.32 |
| LISDEXAMFETAMINE | 1205 | 36 | 1,750.49 | 48.62 |
| METHYLPHENIDATE (DL;D;L;ISOMERS) | 1724 | 107 | 17,032.04 | 159.18 |
| AMOBARBITAL (SCHEDULE 2) | 2125 | 4 | 34.18 | 8.54 |
| BUTALBITAL | 2165 | 10 | 70 | 7 |
| PENTOBARBITAL (SCHEDULE 2) | 2270 | 16 | 2,342.75 | 146.42 |
| COCAINE | 9041L | 59 | 342.26 | 5.8 |
| CODEINE | 9050 | 141 | 29,013.86 | 205.77 |
| BUPRENORPHINE | 9064 | 66 | 1,846.61 | 27.98 |
| OXYCODONE | 9143 | 155 | 83,391.28 | 538.01 |
| HYDROMORPHONE | 9150 | 119 | 3,359.60 | 28.23 |
| HYDROCODONE | 9193 | 153 | 27,414.92 | 179.18 |
| MEPERIDINE (PETHIDINE) | 9230 | 94 | 5,684.90 | 60.48 |
| METHADONE | 9250B | 107 | 6,304.48 | 58.92 |
| MORPHINE | 9300 | 153 | 71,358.40 | 466.39 |
| OPIUM POWDERED | 9639 | 58 | 525.6 | 9.06 |
| OXYMORPHONE | 9652 | 12 | 96.17 | 8.01 |
| ALFENTANIL | 9737 | 7 | 1.47 | 0.21 |
| REMIFENTANIL | 9739 | 30 | 13.87 | 0.46 |
| SUFENTANIL BASE | 9740 | 35 | 0.29 | 0.01 |
| TAPENTADOL | 9780 | 15 | 423.85 | 28.26 |
| FENTANYL BASE | 9801 | 134 | 640.9 | 4.78 |

STATE:ARIZONA BUSINESS ACTIVITY:C - PRACTITIONERS

| DRUG NAME | DRUG CODE | BUYERS | TOTAL GRAMS | AVG GRAMS |
|---|---|---|---|---|
| METHYLPHENIDATE (DL;D;L;ISOMERS) | 1724 | 1 | 14.01 | 14.01 |
| PENTOBARBITAL (SCHEDULE 2) | 2270 | 170 | 178,089.40 | 1,047.58 |
| COCAINE | 9041L | 24 | 145.7 | 6.07 |
| CODEINE | 9050 | 139 | 4,107.65 | 29.55 |
| BUPRENORPHINE | 9064 | 590 | 81.62 | 0.14 |
| DIHYDROCODEINE | 9120 | 3 | 8.76 | 2.92 |
| OXYCODONE | 9143 | 82 | 2,910.08 | 35.49 |
| HYDROMORPHONE | 9150 | 433 | 244.45 | 0.56 |
| HYDROCODONE | 9193 | 354 | 1,504.89 | 4.25 |
| MEPERIDINE (PETHIDINE) | 9230 | 304 | 3,262.39 | 10.73 |
| METHADONE | 9250B | 12 | 10.11 | 0.84 |
| MORPHINE | 9300 | 334 | 615.13 | 1.84 |
| OPIUM POWDERED | 9639 | 8 | 12.6 | 1.58 |
| OXYMORPHONE | 9652 | 18 | 16.94 | 0.94 |
| NOROXYMORPHONE | 9668 | 1 | 1.69 | 1.69 |
| ALFENTANIL | 9737 | 28 | 2.15 | 0.08 |
| REMIFENTANIL | 9739 | 28 | 2.96 | 0.11 |
| SUFENTANIL BASE | 9740 | 1 | 0 | 0 |
| FENTANYL BASE | 9801 | 700 | 69.49 | 0.1 |

STATE:ARIZONA BUSINESS ACTIVITY:M - MID-LEVEL PRACTITIONERS

| DRUG NAME | DRUG CODE | BUYERS | TOTAL GRAMS | AVG GRAMS |
|---|---|---|---|---|
| PENTOBARBITAL (SCHEDULE 2) | 2270 | 4 | 5,153.97 | 1,288.49 |
| CODEINE | 9050 | 35 | 105.58 | 3.02 |
| BUPRENORPHINE | 9064 | 1 | 2.82 | 2.82 |
| DIHYDROCODEINE | 9120 | 2 | 0.64 | 0.32 |
| OXYCODONE | 9143 | 1 | 0.45 | 0.45 |
| HYDROMORPHONE | 9150 | 1 | 0.05 | 0.05 |

| | | | | |
|---|---|---|---|---|
| HYDROCODONE | 9193 | 2 | 64.17 | 32.09 |
| MORPHINE | 9300 | 1 | 0.02 | 0.02 |

STATE:ARIZONA BUSINESS ACTIVITY:N-U NARCOTIC TREATMENT PROGRAMS

| DRUG NAME | DRUG CODE | BUYERS | TOTAL GRAMS | AVG GRAMS |
|---|---|---|---|---|
| BUPRENORPHINE | 9064 | 10 | 462.35 | 46.24 |
| METHADONE | 9250B | 36 | 264,169.09 | 7,338.03 |

STATE:ARKANSAS BUSINESS ACTIVITY:A - PHARMACIES

| DRUG NAME | DRUG CODE | BUYERS | TOTAL GRAMS | AVG GRAMS |
|---|---|---|---|---|
| AMPHETAMINE | 1100 | 792 | 188,616.84 | 238.15 |
| DL-METHAMPHETAMINE RACEMIC BASE | 1105B | 5 | 6.03 | 1.21 |
| D-METHAMPHETAMINE | 1105D | 13 | 14.06 | 1.08 |
| LISDEXAMFETAMINE | 1205 | 774 | 121,987.77 | 157.61 |
| METHYLPHENIDATE (DL;D;L;ISOMERS) | 1724 | 785 | 160,777 | 204.81 |
| BARBITURIC ACID DERIVIATIVE OR SALT [PER 21C | 2100 | 1 | 5 | 5 |
| BUTALBITAL | 2165 | 503 | 19,600 | 38.97 |
| SECOBARBITAL (SCHEDULE 2) | 2315 | 1 | 18.31 | 18.31 |
| COCAINE | 9041L | 1 | 4.46 | 4.46 |
| CODEINE | 9050 | 783 | 302,840.94 | 386.77 |
| BUPRENORPHINE | 9064 | 721 | 19,120.73 | 26.52 |
| DIHYDROCODEINE | 9120 | 93 | 366.38 | 3.94 |
| OXYCODONE | 9143 | 798 | 601,540.30 | 753.81 |
| HYDROMORPHONE | 9150 | 742 | 13,110.70 | 17.67 |
| HYDROCODONE | 9193 | 802 | 539,410.93 | 672.58 |
| LEVORPHANOL | 9220L | 4 | 7.89 | 1.97 |
| MEPERIDINE (PETHIDINE) | 9230 | 697 | 30,293.45 | 43.46 |
| METHADONE | 9250B | 718 | 42,883.44 | 59.73 |
| MORPHINE | 9300 | 786 | 185,736.67 | 236.31 |
| OPIUM POWDERED | 9639 | 31 | 61.56 | 1.99 |

| | | | | |
|---|---|---|---|---|
| OXYMORPHONE | 9652 | 513 | 10,031.40 | 19.55 |
| TAPENTADOL | 9780 | 667 | 106,348.50 | 159.44 |
| FENTANYL BASE | 9801 | 770 | 4,069.44 | 5.28 |

STATE:ARKANSAS BUSINESS ACTIVITY:B - HOSPITALS

| DRUG NAME | DRUG CODE | BUYERS | TOTAL GRAMS | AVG GRAMS |
|---|---|---|---|---|
| AMPHETAMINE | 1100 | 46 | 4,071.50 | 88.51 |
| LISDEXAMFETAMINE | 1205 | 24 | 870.62 | 36.28 |
| METHYLPHENIDATE (DL;D;L;ISOMERS) | 1724 | 70 | 4,481.12 | 64.02 |
| AMOBARBITAL (SCHEDULE 2) | 2125 | 1 | 0.46 | 0.46 |
| BUTALBITAL | 2165 | 5 | 85 | 17 |
| PENTOBARBITAL (SCHEDULE 2) | 2270 | 3 | 26.43 | 8.81 |
| COCAINE | 9041L | 32 | 185.24 | 5.79 |
| CODEINE | 9050 | 93 | 6,044.56 | 65 |
| BUPRENORPHINE | 9064 | 42 | 872.4 | 20.77 |
| OXYCODONE | 9143 | 118 | 32,135.77 | 272.34 |
| HYDROMORPHONE | 9150 | 110 | 2,210.89 | 20.1 |
| HYDROCODONE | 9193 | 120 | 33,166.01 | 276.38 |
| MEPERIDINE (PETHIDINE) | 9230 | 107 | 7,293.74 | 68.17 |
| METHADONE | 9250B | 80 | 3,905.95 | 48.82 |
| MORPHINE | 9300 | 120 | 20,495.06 | 170.79 |
| OPIUM POWDERED | 9639 | 34 | 326.16 | 9.59 |
| OXYMORPHONE | 9652 | 7 | 51.65 | 7.38 |
| ALFENTANIL | 9737 | 4 | 0.16 | 0.04 |
| REMIFENTANIL | 9739 | 11 | 6.69 | 0.61 |
| SUFENTANIL BASE | 9740 | 26 | 0.64 | 0.02 |
| TAPENTADOL | 9780 | 28 | 1,254.50 | 44.8 |
| FENTANYL BASE | 9801 | 121 | 463.83 | 3.83 |

STATE:ARKANSAS BUSINESS ACTIVITY:C - PRACTITIONERS

| DRUG NAME | DRUG CODE | BUYERS | TOTAL GRAMS | AVG GRAMS |
|---|---|---|---|---|
| LISDEXAMFETAMINE | 1205 | 1 | 2.31 | 2.31 |
| PENTOBARBITAL (SCHEDULE 2) | 2270 | 47 | 23,011.26 | 489.6 |
| COCAINE | 9041L | 2 | 26.28 | 13.14 |
| CODEINE | 9050 | 31 | 584.79 | 18.86 |
| BUPRENORPHINE | 9064 | 130 | 8.46 | 0.07 |
| DIHYDROCODEINE | 9120 | 30 | 7.57 | 0.25 |
| OXYCODONE | 9143 | 8 | 84.06 | 10.51 |
| HYDROMORPHONE | 9150 | 55 | 36.06 | 0.66 |
| HYDROCODONE | 9193 | 78 | 173.74 | 2.23 |
| MEPERIDINE (PETHIDINE) | 9230 | 44 | 609.18 | 13.84 |
| METHADONE | 9250B | 1 | 0.89 | 0.89 |
| MORPHINE | 9300 | 86 | 167.57 | 1.95 |
| OPIUM POWDERED | 9639 | 2 | 3.6 | 1.8 |
| NOROXYMORPHONE | 9668 | 2 | 56.46 | 28.23 |
| REMIFENTANIL | 9739 | 3 | 0.21 | 0.07 |
| TAPENTADOL | 9780 | 1 | 6.35 | 6.35 |
| FENTANYL BASE | 9801 | 144 | 21.44 | 0.15 |

STATE:ARKANSAS BUSINESS ACTIVITY:D - TEACHING INSTITUTIONS

| DRUG NAME | DRUG CODE | BUYERS | TOTAL GRAMS | AVG GRAMS |
|---|---|---|---|---|
| BUPRENORPHINE | 9064 | 1 | 0 | 0 |

STATE:ARKANSAS BUSINESS ACTIVITY:M - MID-LEVEL PRACTITIONERS

| DRUG NAME | DRUG CODE | BUYERS | TOTAL GRAMS | AVG GRAMS |
|---|---|---|---|---|
| PENTOBARBITAL (SCHEDULE 2) | 2270 | 7 | 7,911.41 | 1,130.20 |
| DIHYDROCODEINE | 9120 | 19 | 15.78 | 0.83 |

STATE:ARKANSAS BUSINESS ACTIVITY:N-U NARCOTIC TREATMENT PROGRAMS

STATISTICAL SUMMARY FOR RETAIL DRUG PURCHASES BY GRAMS WT

| DRUG NAME | DRUG CODE | BUYERS | TOTAL GRAMS | AVG GRAMS |
|---|---|---|---|---|
| BUPRENORPHINE | 9064 | 1 | 57.6 | 57.6 |
| METHADONE | 9250B | 6 | 30,795.98 | 5,132.66 |

STATE:CALIFORNIA BUSINESS ACTIVITY:A - PHARMACIES

| DRUG NAME | DRUG CODE | BUYERS | TOTAL GRAMS | AVG GRAMS |
|---|---|---|---|---|
| AMPHETAMINE | 1100 | 5,423 | 1,123,817.86 | 207.23 |
| DL-METHAMPHETAMINE RACEMIC BASE | 1105B | 54 | 352.42 | 6.53 |
| D-METHAMPHETAMINE | 1105D | 310 | 1,072.14 | 3.46 |
| LISDEXAMFETAMINE | 1205 | 4,771 | 427,563.34 | 89.62 |
| METHYLPHENIDATE (DL;D;L;ISOMERS) | 1724 | 5,374 | 1,062,158.76 | 197.65 |
| BARBITURIC ACID DERIVIATIVE OR SALT [PER 21C | 2100 | 98 | 1,070 | 10.92 |
| BUTALBITAL | 2165 | 3,344 | 175,375 | 52.44 |
| PENTOBARBITAL (SCHEDULE 2) | 2270 | 1 | 444.31 | 444.31 |
| SECOBARBITAL (SCHEDULE 2) | 2315 | 55 | 2,783.12 | 50.6 |
| COCAINE | 9041L | 43 | 481.4 | 11.2 |
| CODEINE | 9050 | 6,114 | 1,994,231.82 | 326.17 |
| BUPRENORPHINE | 9064 | 5,061 | 131,711.27 | 26.02 |
| DIHYDROCODEINE | 9120 | 53 | 1,556.95 | 29.38 |
| OXYCODONE | 9143 | 5,792 | 3,725,454.20 | 643.21 |
| HYDROMORPHONE | 9150 | 5,194 | 151,530.79 | 29.17 |
| HYDROCODONE | 9193 | 6,067 | 3,846,456.56 | 634 |
| LEVORPHANOL | 9220L | 217 | 392.12 | 1.81 |
| MEPERIDINE (PETHIDINE) | 9230 | 1,149 | 34,371.85 | 29.91 |
| METHADONE | 9250B | 4,746 | 422,872.59 | 89.1 |
| MORPHINE | 9300 | 5,446 | 1,880,203.48 | 345.24 |
| OPIUM TINCTURE (LAUDANUM) | 9630 | 8 | 46.11 | 5.76 |
| OPIUM POWDERED | 9639 | 276 | 616.68 | 2.23 |
| OXYMORPHONE | 9652 | 2,685 | 79,974.86 | 29.79 |
| NOROXYMORPHONE | 9668 | 2 | 4.77 | 2.39 |
| ALFENTANIL | 9737 | 14 | 4.88 | 0.35 |

| | | | | |
|---|---|---|---|---|
| REMIFENTANIL | 9739 | 13 | 0.99 | 0.08 |
| SUFENTANIL BASE | 9740 | 8 | 35.25 | 4.41 |
| TAPENTADOL | 9780 | 3,697 | 477,364 | 129.12 |
| FENTANYL BASE | 9801 | 5,187 | 30,620.81 | 5.9 |

STATE:CALIFORNIA BUSINESS ACTIVITY:B - HOSPITALS

| DRUG NAME | DRUG CODE | BUYERS | TOTAL GRAMS | AVG GRAMS |
|---|---|---|---|---|
| AMPHETAMINE | 1100 | 260 | 39,339.90 | 151.31 |
| D-METHAMPHETAMINE | 1105D | 2 | 14.06 | 7.03 |
| LISDEXAMFETAMINE | 1205 | 90 | 6,120.92 | 68.01 |
| METHYLPHENIDATE (DL;D;L;ISOMERS) | 1724 | 382 | 44,934.74 | 117.63 |
| GAMMA HYDROXYBUTYRIC ACID(FDA APPROVED | 2012 | 1 | 1,486.08 | 1,486.08 |
| BARBITURIC ACID DERIVIATIVE OR SALT [PER 21C | 2100 | 2 | 20 | 10 |
| AMOBARBITAL (SCHEDULE 2) | 2125 | 25 | 102.99 | 4.12 |
| BUTALBITAL | 2165 | 68 | 1,080 | 15.88 |
| PENTOBARBITAL (SCHEDULE 2) | 2270 | 73 | 15,102.81 | 206.89 |
| SECOBARBITAL (SCHEDULE 2) | 2315 | 3 | 27.47 | 9.16 |
| NABILONE | 7379 | 1 | 0.05 | 0.05 |
| COCAINE | 9041L | 407 | 2,996.27 | 7.36 |
| CODEINE | 9050 | 599 | 106,071.07 | 177.08 |
| BUPRENORPHINE | 9064 | 313 | 4,655.29 | 14.87 |
| OXYCODONE | 9143 | 644 | 181,018.59 | 281.08 |
| HYDROMORPHONE | 9150 | 687 | 25,028.96 | 36.43 |
| HYDROCODONE | 9193 | 744 | 205,005.63 | 275.55 |
| LEVORPHANOL | 9220L | 8 | 4.41 | 0.55 |
| MEPERIDINE (PETHIDINE) | 9230 | 656 | 27,379.65 | 41.74 |
| METHADONE | 9250B | 518 | 54,840.81 | 105.87 |
| MORPHINE | 9300 | 781 | 191,805.65 | 245.59 |
| OPIUM POWDERED | 9639 | 295 | 1,287.72 | 4.37 |
| OXYMORPHONE | 9652 | 65 | 1,141.29 | 17.56 |
| NOROXYMORPHONE | 9668 | 2 | 9.63 | 4.81 |
| ALFENTANIL | 9737 | 127 | 52.32 | 0.41 |

| | | | | |
|---|---|---|---|---|
| REMIFENTANIL | 9739 | 191 | 124.97 | 0.65 |
| SUFENTANIL BASE | 9740 | 151 | 3.89 | 0.03 |
| TAPENTADOL | 9780 | 77 | 3,408.94 | 44.27 |
| FENTANYL BASE | 9801 | 842 | 3,442.23 | 4.09 |

STATE:CALIFORNIA BUSINESS ACTIVITY:C - PRACTITIONERS

| DRUG NAME | DRUG CODE | BUYERS | TOTAL GRAMS | AVG GRAMS |
|---|---|---|---|---|
| AMPHETAMINE | 1100 | 16 | 1,015.58 | 63.47 |
| D-METHAMPHETAMINE | 1105D | 1 | 0.08 | 0.08 |
| METHYLPHENIDATE (DL;D;L;ISOMERS) | 1724 | 2 | 37.18 | 18.59 |
| GAMMA HYDROXYBUTYRIC ACID(FDA APPROVED | 2012 | 3 | 11,368.51 | 3,789.50 |
| BUTALBITAL | 2165 | 5 | 60 | 12 |
| PENTOBARBITAL (SCHEDULE 2) | 2270 | 520 | 645,182.28 | 1,240.74 |
| COCAINE | 9041L | 113 | 287.12 | 2.54 |
| CODEINE | 9050 | 865 | 42,506.02 | 49.14 |
| BUPRENORPHINE | 9064 | 2,594 | 988.39 | 0.38 |
| DIHYDROCODEINE | 9120 | 60 | 3,095.53 | 51.59 |
| OXYCODONE | 9143 | 242 | 1,594.98 | 6.59 |
| HYDROMORPHONE | 9150 | 1,668 | 938.9 | 0.56 |
| HYDROCODONE | 9193 | 2,114 | 17,270.88 | 8.17 |
| MEPERIDINE (PETHIDINE) | 9230 | 881 | 9,022.31 | 10.24 |
| METHADONE | 9250B | 109 | 367.24 | 3.37 |
| MORPHINE | 9300 | 1,165 | 2,032.02 | 1.74 |
| OPIUM POWDERED | 9639 | 8 | 7.92 | 0.99 |
| OXYMORPHONE | 9652 | 93 | 68 | 0.73 |
| NOROXYMORPHONE | 9668 | 28 | 307.45 | 10.98 |
| ALFENTANIL | 9737 | 52 | 10.57 | 0.2 |
| REMIFENTANIL | 9739 | 69 | 3.35 | 0.05 |
| SUFENTANIL BASE | 9740 | 8 | 0.01 | 0 |
| FENTANYL BASE | 9801 | 2,385 | 352.67 | 0.15 |

STATISTICAL SUMMARY FOR RETAIL DRUG PURCHASES BY GRAMS WT

STATE:CALIFORNIA BUSINESS ACTIVITY:D - TEACHING INSTITUTIONS

| DRUG NAME | DRUG CODE | BUYERS | TOTAL GRAMS | AVG GRAMS |
|---|---|---|---|---|
| BUPRENORPHINE | 9064 | 3 | 0.03 | 0.01 |
| HYDROMORPHONE | 9150 | 2 | 0.11 | 0.05 |
| MORPHINE | 9300 | 1 | 0.47 | 0.47 |

STATE:CALIFORNIA BUSINESS ACTIVITY:M - MID-LEVEL PRACTITIONERS

| DRUG NAME | DRUG CODE | BUYERS | TOTAL GRAMS | AVG GRAMS |
|---|---|---|---|---|
| BARBITURIC ACID DERIVIATIVE OR SALT [PER 21C | 2100 | 1 | 8 | 8 |
| PENTOBARBITAL (SCHEDULE 2) | 2270 | 6 | 18,927.50 | 3,154.58 |
| CODEINE | 9050 | 75 | 579.23 | 7.72 |

STATE:CALIFORNIA BUSINESS ACTIVITY:N-U NARCOTIC TREATMENT PROGRAMS

| DRUG NAME | DRUG CODE | BUYERS | TOTAL GRAMS | AVG GRAMS |
|---|---|---|---|---|
| BUPRENORPHINE | 9064 | 27 | 230.02 | 8.52 |
| METHADONE | 9250B | 153 | 923,002.91 | 6,032.70 |

STATE:COLORADO BUSINESS ACTIVITY:A - PHARMACIES

| DRUG NAME | DRUG CODE | BUYERS | TOTAL GRAMS | AVG GRAMS |
|---|---|---|---|---|
| AMPHETAMINE | 1100 | 803 | 253,933.41 | 316.23 |
| DL-METHAMPHETAMINE RACEMIC BASE | 1105B | 5 | 9.64 | 1.93 |
| D-METHAMPHETAMINE | 1105D | 62 | 167.51 | 2.7 |
| LISDEXAMFETAMINE | 1205 | 763 | 101,976.26 | 133.65 |
| METHYLPHENIDATE (DL;D;L;ISOMERS) | 1724 | 802 | 203,104.25 | 253.25 |
| BARBITURIC ACID DERIVIATIVE OR SALT [PER 21C | 2100 | 12 | 120 | 10 |
| BUTALBITAL | 2165 | 467 | 15,710 | 33.64 |
| SECOBARBITAL (SCHEDULE 2) | 2315 | 2 | 45.78 | 22.89 |

| | | | | |
|---|---|---|---|---|
| NABILONE | 7379 | 2 | 1 | 0.5 |
| COCAINE | 9041L | 5 | 3.71 | 0.74 |
| CODEINE | 9050 | 792 | 93,584.28 | 118.16 |
| BUPRENORPHINE | 9064 | 772 | 23,697.98 | 30.7 |
| DIHYDROCODEINE | 9120 | 3 | 3.2 | 1.07 |
| OXYCODONE | 9143 | 814 | 867,954.27 | 1,066.28 |
| HYDROMORPHONE | 9150 | 791 | 24,993.26 | 31.6 |
| HYDROCODONE | 9193 | 816 | 246,313.94 | 301.86 |
| LEVORPHANOL | 9220L | 15 | 19.5 | 1.3 |
| MEPERIDINE (PETHIDINE) | 9230 | 322 | 8,144.06 | 25.29 |
| METHADONE | 9250B | 689 | 39,102.54 | 56.75 |
| MORPHINE | 9300 | 802 | 280,505.28 | 349.76 |
| OPIUM TINCTURE (LAUDANUM) | 9630 | 1 | 1.18 | 1.18 |
| OPIUM POWDERED | 9639 | 91 | 209.52 | 2.3 |
| OXYMORPHONE | 9652 | 582 | 18,254.60 | 31.37 |
| SUFENTANIL BASE | 9740 | 1 | 0.07 | 0.07 |
| TAPENTADOL | 9780 | 661 | 105,799.50 | 160.06 |
| FENTANYL BASE | 9801 | 782 | 5,111.37 | 6.54 |

STATE:COLORADO BUSINESS ACTIVITY:B - HOSPITALS

| DRUG NAME | DRUG CODE | BUYERS | TOTAL GRAMS | AVG GRAMS |
|---|---|---|---|---|
| AMPHETAMINE | 1100 | 109 | 42,536.88 | 390.25 |
| DL-METHAMPHETAMINE RACEMIC BASE | 1105B | 1 | 0.8 | 0.8 |
| D-METHAMPHETAMINE | 1105D | 2 | 2.41 | 1.21 |
| LISDEXAMFETAMINE | 1205 | 67 | 5,812.80 | 86.76 |
| METHYLPHENIDATE (DL;D;L;ISOMERS) | 1724 | 127 | 41,919.85 | 330.08 |
| AMOBARBITAL (SCHEDULE 2) | 2125 | 5 | 66.08 | 13.22 |
| BUTALBITAL | 2165 | 35 | 780 | 22.29 |
| PENTOBARBITAL (SCHEDULE 2) | 2270 | 14 | 1,734.39 | 123.89 |
| SECOBARBITAL (SCHEDULE 2) | 2315 | 1 | 9.16 | 9.16 |
| NABILONE | 7379 | 1 | 0.05 | 0.05 |
| COCAINE | 9041L | 56 | 244.81 | 4.37 |

| | | | | |
|---|---|---|---|---|
| CODEINE | 9050 | 160 | 16,140.66 | 100.88 |
| BUPRENORPHINE | 9064 | 99 | 3,009.42 | 30.4 |
| OXYCODONE | 9143 | 184 | 137,325.12 | 746.33 |
| HYDROMORPHONE | 9150 | 192 | 6,161.86 | 32.09 |
| HYDROCODONE | 9193 | 203 | 42,110.81 | 207.44 |
| MEPERIDINE (PETHIDINE) | 9230 | 96 | 1,063.01 | 11.07 |
| METHADONE | 9250B | 121 | 11,267.31 | 93.12 |
| MORPHINE | 9300 | 199 | 77,342.02 | 388.65 |
| OPIUM POWDERED | 9639 | 63 | 600.48 | 9.53 |
| OXYMORPHONE | 9652 | 54 | 864.98 | 16.02 |
| ALFENTANIL | 9737 | 14 | 2.73 | 0.2 |
| REMIFENTANIL | 9739 | 57 | 23.81 | 0.42 |
| SUFENTANIL BASE | 9740 | 32 | 0.73 | 0.02 |
| TAPENTADOL | 9780 | 60 | 4,684.50 | 78.08 |
| FENTANYL BASE | 9801 | 195 | 1,054.25 | 5.41 |

STATE:COLORADO BUSINESS ACTIVITY:C - PRACTITIONERS

| DRUG NAME | DRUG CODE | BUYERS | TOTAL GRAMS | AVG GRAMS |
|---|---|---|---|---|
| AMPHETAMINE | 1100 | 1 | 0.28 | 0.28 |
| PENTOBARBITAL (SCHEDULE 2) | 2270 | 167 | 123,706.30 | 740.76 |
| COCAINE | 9041L | 18 | 127.42 | 7.08 |
| CODEINE | 9050 | 71 | 935.36 | 13.17 |
| BUPRENORPHINE | 9064 | 687 | 69.88 | 0.1 |
| OXYCODONE | 9143 | 124 | 873.8 | 7.05 |
| HYDROMORPHONE | 9150 | 539 | 299.95 | 0.56 |
| HYDROCODONE | 9193 | 394 | 1,128.62 | 2.86 |
| MEPERIDINE (PETHIDINE) | 9230 | 118 | 390.32 | 3.31 |
| METHADONE | 9250B | 28 | 55.18 | 1.97 |
| MORPHINE | 9300 | 340 | 510.66 | 1.5 |
| OPIUM POWDERED | 9639 | 13 | 93.6 | 7.2 |
| OXYMORPHONE | 9652 | 16 | 1.53 | 0.1 |
| NOROXYMORPHONE | 9668 | 1 | 0.53 | 0.53 |

| | | | | |
|---|---|---|---|---|
| ALFENTANIL | 9737 | 7 | 0.33 | 0.05 |
| REMIFENTANIL | 9739 | 27 | 1.44 | 0.05 |
| SUFENTANIL BASE | 9740 | 10 | 0.02 | 0 |
| FENTANYL BASE | 9801 | 607 | 87.42 | 0.14 |

STATE:COLORADO BUSINESS ACTIVITY:M - MID-LEVEL PRACTITIONERS

| DRUG NAME | DRUG CODE | BUYERS | TOTAL GRAMS | AVG GRAMS |
|---|---|---|---|---|
| PENTOBARBITAL (SCHEDULE 2) | 2270 | 18 | 17,328.45 | 962.69 |
| CODEINE | 9050 | 19 | 8.23 | 0.43 |
| HYDROMORPHONE | 9150 | 7 | 0.57 | 0.08 |
| MORPHINE | 9300 | 27 | 9.63 | 0.36 |
| FENTANYL BASE | 9801 | 44 | 1.26 | 0.03 |

STATE:COLORADO BUSINESS ACTIVITY:N-U NARCOTIC TREATMENT PROGRAMS

| DRUG NAME | DRUG CODE | BUYERS | TOTAL GRAMS | AVG GRAMS |
|---|---|---|---|---|
| BUPRENORPHINE | 9064 | 5 | 159.8 | 31.96 |
| METHADONE | 9250B | 17 | 108,168.74 | 6,362.87 |

STATE:CONNECTICUT BUSINESS ACTIVITY:A - PHARMACIES

| DRUG NAME | DRUG CODE | BUYERS | TOTAL GRAMS | AVG GRAMS |
|---|---|---|---|---|
| AMPHETAMINE | 1100 | 708 | 196,427.29 | 277.44 |
| DL-METHAMPHETAMINE RACEMIC BASE | 1105B | 7 | 21.7 | 3.1 |
| D-METHAMPHETAMINE | 1105D | 21 | 65.88 | 3.14 |
| LISDEXAMFETAMINE | 1205 | 693 | 139,641.79 | 201.5 |
| METHYLPHENIDATE (DL;D;L;ISOMERS) | 1724 | 705 | 217,729.82 | 308.84 |
| BARBITURIC ACID DERIVIATIVE OR SALT [PER 21C | 2100 | 13 | 115 | 8.85 |
| BUTALBITAL | 2165 | 459 | 24,555 | 53.5 |
| SECOBARBITAL (SCHEDULE 2) | 2315 | 2 | 45.78 | 22.89 |

| | | | | |
|---|---|---|---|---|
| CODEINE | 9050 | 703 | 106,175.71 | 151.03 |
| BUPRENORPHINE | 9064 | 689 | 52,535.72 | 76.25 |
| DIHYDROCODEINE | 9120 | 1 | 3.2 | 3.2 |
| OXYCODONE | 9143 | 715 | 764,387.69 | 1,069.07 |
| HYDROMORPHONE | 9150 | 683 | 24,422.91 | 35.76 |
| HYDROCODONE | 9193 | 709 | 93,196.81 | 131.45 |
| LEVORPHANOL | 9220L | 11 | 5.22 | 0.47 |
| MEPERIDINE (PETHIDINE) | 9230 | 165 | 4,519.38 | 27.39 |
| METHADONE | 9250B | 526 | 27,402.72 | 52.1 |
| MORPHINE | 9300 | 676 | 149,625.74 | 221.34 |
| OPIUM POWDERED | 9639 | 56 | 242.64 | 4.33 |
| OXYMORPHONE | 9652 | 465 | 15,710.68 | 33.79 |
| SUFENTANIL BASE | 9740 | 1 | 0.67 | 0.67 |
| TAPENTADOL | 9780 | 500 | 53,164.50 | 106.33 |
| FENTANYL BASE | 9801 | 680 | 4,239.82 | 6.24 |

STATE:CONNECTICUT BUSINESS ACTIVITY:B - HOSPITALS

| DRUG NAME | DRUG CODE | BUYERS | TOTAL GRAMS | AVG GRAMS |
|---|---|---|---|---|
| AMPHETAMINE | 1100 | 42 | 4,161.49 | 99.08 |
| DL-METHAMPHETAMINE RACEMIC BASE | 1105B | 1 | 14.47 | 14.47 |
| LISDEXAMFETAMINE | 1205 | 18 | 1,353.33 | 75.19 |
| METHYLPHENIDATE (DL;D;L;ISOMERS) | 1724 | 53 | 4,555.16 | 85.95 |
| AMOBARBITAL (SCHEDULE 2) | 2125 | 1 | 0.91 | 0.91 |
| BUTALBITAL | 2165 | 5 | 105 | 21 |
| PENTOBARBITAL (SCHEDULE 2) | 2270 | 6 | 676.71 | 112.79 |
| COCAINE | 9041L | 27 | 155.56 | 5.76 |
| CODEINE | 9050 | 54 | 5,061.64 | 93.73 |
| BUPRENORPHINE | 9064 | 50 | 2,008.19 | 40.16 |
| OXYCODONE | 9143 | 66 | 23,151.41 | 350.78 |
| HYDROMORPHONE | 9150 | 65 | 4,113.41 | 63.28 |
| HYDROCODONE | 9193 | 58 | 2,679.38 | 46.2 |
| LEVORPHANOL | 9220L | 1 | 0.12 | 0.12 |

| | | | | |
|---|---|---|---|---|
| MEPERIDINE (PETHIDINE) | 9230 | 41 | 460.7 | 11.24 |
| METHADONE | 9250B | 54 | 4,510.24 | 83.52 |
| MORPHINE | 9300 | 65 | 14,073.03 | 216.51 |
| OPIUM POWDERED | 9639 | 30 | 120.96 | 4.03 |
| OXYMORPHONE | 9652 | 10 | 156.39 | 15.64 |
| NOROXYMORPHONE | 9668 | 1 | 0.44 | 0.44 |
| ALFENTANIL | 9737 | 3 | 3.68 | 1.23 |
| REMIFENTANIL | 9739 | 23 | 31.76 | 1.38 |
| SUFENTANIL BASE | 9740 | 11 | 0.64 | 0.06 |
| TAPENTADOL | 9780 | 8 | 735.8 | 91.98 |
| FENTANYL BASE | 9801 | 68 | 429.03 | 6.31 |

STATE:CONNECTICUT BUSINESS ACTIVITY:C - PRACTITIONERS

| DRUG NAME | DRUG CODE | BUYERS | TOTAL GRAMS | AVG GRAMS |
|---|---|---|---|---|
| AMPHETAMINE | 1100 | 1 | 13.14 | 13.14 |
| PENTOBARBITAL (SCHEDULE 2) | 2270 | 73 | 60,852.53 | 833.6 |
| COCAINE | 9041L | 3 | 28.14 | 9.38 |
| CODEINE | 9050 | 46 | 224.43 | 4.88 |
| BUPRENORPHINE | 9064 | 319 | 33.4 | 0.1 |
| OXYCODONE | 9143 | 43 | 1,750.05 | 40.7 |
| HYDROMORPHONE | 9150 | 176 | 162.57 | 0.92 |
| HYDROCODONE | 9193 | 102 | 787.55 | 7.72 |
| MEPERIDINE (PETHIDINE) | 9230 | 31 | 376.71 | 12.15 |
| METHADONE | 9250B | 15 | 97.22 | 6.48 |
| MORPHINE | 9300 | 152 | 745.97 | 4.91 |
| OPIUM POWDERED | 9639 | 2 | 0.72 | 0.36 |
| OXYMORPHONE | 9652 | 5 | 0.37 | 0.07 |
| NOROXYMORPHONE | 9668 | 1 | 0.58 | 0.58 |
| ALFENTANIL | 9737 | 3 | 1.15 | 0.38 |
| REMIFENTANIL | 9739 | 8 | 0.38 | 0.05 |
| FENTANYL BASE | 9801 | 236 | 39.32 | 0.17 |

STATE:CONNECTICUT BUSINESS ACTIVITY:M - MID-LEVEL PRACTITIONERS

| DRUG NAME | DRUG CODE | BUYERS | TOTAL GRAMS | AVG GRAMS |
|---|---|---|---|---|
| CODEINE | 9050 | 3 | 1.52 | 0.51 |
| FENTANYL BASE | 9801 | 1 | 0.09 | 0.09 |

STATE:CONNECTICUT BUSINESS ACTIVITY:N-U NARCOTIC TREATMENT PROGRAMS

| DRUG NAME | DRUG CODE | BUYERS | TOTAL GRAMS | AVG GRAMS |
|---|---|---|---|---|
| BUPRENORPHINE | 9064 | 4 | 84.12 | 21.03 |
| METHADONE | 9250B | 37 | 393,315.98 | 10,630.16 |

STATE:DELAWARE BUSINESS ACTIVITY:A - PHARMACIES

| DRUG NAME | DRUG CODE | BUYERS | TOTAL GRAMS | AVG GRAMS |
|---|---|---|---|---|
| AMPHETAMINE | 1100 | 212 | 84,260.46 | 397.45 |
| D-METHAMPHETAMINE | 1105D | 5 | 20.49 | 4.1 |
| LISDEXAMFETAMINE | 1205 | 205 | 56,801.22 | 277.08 |
| METHYLPHENIDATE (DL;D;L;ISOMERS) | 1724 | 206 | 74,400.86 | 361.17 |
| BARBITURIC ACID DERIVIATIVE OR SALT [PER 21C | 2100 | 1 | 15 | 15 |
| BUTALBITAL | 2165 | 120 | 3,800 | 31.67 |
| SECOBARBITAL (SCHEDULE 2) | 2315 | 1 | 9.16 | 9.16 |
| CODEINE | 9050 | 209 | 41,838.74 | 200.19 |
| BUPRENORPHINE | 9064 | 202 | 8,975.69 | 44.43 |
| DIHYDROCODEINE | 9120 | 2 | 2.14 | 1.07 |
| OXYCODONE | 9143 | 211 | 292,441.94 | 1,385.98 |
| HYDROMORPHONE | 9150 | 201 | 9,035.72 | 44.95 |
| HYDROCODONE | 9193 | 210 | 26,642.96 | 126.87 |
| MEPERIDINE (PETHIDINE) | 9230 | 40 | 662.78 | 16.57 |
| METHADONE | 9250B | 180 | 16,721.70 | 92.9 |
| MORPHINE | 9300 | 201 | 84,186.54 | 418.84 |

| | | | | |
|---|---|---|---|---|
| OPIUM POWDERED | 9639 | 12 | 47.16 | 3.93 |
| OXYMORPHONE | 9652 | 155 | 8,804.76 | 56.8 |
| TAPENTADOL | 9780 | 162 | 27,158 | 167.64 |
| FENTANYL BASE | 9801 | 205 | 1,518.53 | 7.41 |

STATE:DELAWARE BUSINESS ACTIVITY:B - HOSPITALS

| DRUG NAME | DRUG CODE | BUYERS | TOTAL GRAMS | AVG GRAMS |
|---|---|---|---|---|
| AMPHETAMINE | 1100 | 11 | 1,705.10 | 155.01 |
| LISDEXAMFETAMINE | 1205 | 7 | 421.43 | 60.2 |
| METHYLPHENIDATE (DL;D;L;ISOMERS) | 1724 | 16 | 2,130.30 | 133.14 |
| BUTALBITAL | 2165 | 1 | 5 | 5 |
| PENTOBARBITAL (SCHEDULE 2) | 2270 | 2 | 313.52 | 156.76 |
| COCAINE | 9041L | 12 | 74.57 | 6.21 |
| CODEINE | 9050 | 17 | 1,615.28 | 95.02 |
| BUPRENORPHINE | 9064 | 16 | 633.41 | 39.59 |
| OXYCODONE | 9143 | 26 | 10,382.59 | 399.33 |
| HYDROMORPHONE | 9150 | 30 | 648.03 | 21.6 |
| HYDROCODONE | 9193 | 23 | 1,209.74 | 52.6 |
| MEPERIDINE (PETHIDINE) | 9230 | 18 | 267.72 | 14.87 |
| METHADONE | 9250B | 17 | 1,847.29 | 108.66 |
| MORPHINE | 9300 | 27 | 7,129.85 | 264.07 |
| OPIUM POWDERED | 9639 | 7 | 30.96 | 4.42 |
| OXYMORPHONE | 9652 | 6 | 40.93 | 6.82 |
| REMIFENTANIL | 9739 | 7 | 12.7 | 1.81 |
| SUFENTANIL BASE | 9740 | 4 | 0.04 | 0.01 |
| TAPENTADOL | 9780 | 3 | 88 | 29.33 |
| FENTANYL BASE | 9801 | 34 | 98.29 | 2.89 |

STATE:DELAWARE BUSINESS ACTIVITY:C - PRACTITIONERS

| DRUG NAME | DRUG CODE | BUYERS | TOTAL GRAMS | AVG GRAMS |
|---|---|---|---|---|

| | | | | |
|---|---|---|---|---|
| METHYLPHENIDATE (DL;D;L;ISOMERS) | 1724 | 1 | 18.68 | 18.68 |
| PENTOBARBITAL (SCHEDULE 2) | 2270 | 9 | 7,014.37 | 779.37 |
| CODEINE | 9050 | 18 | 130.55 | 7.25 |
| BUPRENORPHINE | 9064 | 62 | 3.77 | 0.06 |
| DIHYDROCODEINE | 9120 | 1 | 0.32 | 0.32 |
| OXYCODONE | 9143 | 5 | 3.77 | 0.75 |
| HYDROMORPHONE | 9150 | 31 | 10.55 | 0.34 |
| HYDROCODONE | 9193 | 16 | 37.72 | 2.36 |
| MEPERIDINE (PETHIDINE) | 9230 | 2 | 3.7 | 1.85 |
| METHADONE | 9250B | 3 | 3.67 | 1.22 |
| MORPHINE | 9300 | 9 | 26.3 | 2.92 |
| OXYMORPHONE | 9652 | 1 | 0.02 | 0.02 |
| FENTANYL BASE | 9801 | 51 | 4.58 | 0.09 |

STATE:DELAWARE BUSINESS ACTIVITY:M - MID-LEVEL PRACTITIONERS

| DRUG NAME | DRUG CODE | BUYERS | TOTAL GRAMS | AVG GRAMS |
|---|---|---|---|---|
| CODEINE | 9050 | 2 | 2.64 | 1.32 |
| OXYCODONE | 9143 | 1 | 0.47 | 0.47 |
| HYDROMORPHONE | 9150 | 1 | 0.32 | 0.32 |
| METHADONE | 9250B | 1 | 0 | 0 |
| MORPHINE | 9300 | 2 | 56.93 | 28.47 |
| FENTANYL BASE | 9801 | 1 | 2.11 | 2.11 |

STATE:DELAWARE BUSINESS ACTIVITY:N-U NARCOTIC TREATMENT PROGRAMS

| DRUG NAME | DRUG CODE | BUYERS | TOTAL GRAMS | AVG GRAMS |
|---|---|---|---|---|
| BUPRENORPHINE | 9064 | 11 | 2,053.68 | 186.7 |
| METHADONE | 9250B | 12 | 82,610.36 | 6,884.20 |

STATE:DISTRICT OF COLUMBIA BUSINESS ACTIVITY:A - PHARMACIES

| DRUG NAME | DRUG CODE | BUYERS | TOTAL GRAMS | AVG GRAMS |
|---|---|---|---|---|
| AMPHETAMINE | 1100 | 137 | 42,570.46 | 310.73 |
| DL-METHAMPHETAMINE RACEMIC BASE | 1105B | 1 | 11.25 | 11.25 |
| D-METHAMPHETAMINE | 1105D | 4 | 3.62 | 0.9 |
| LISDEXAMFETAMINE | 1205 | 128 | 19,027.01 | 148.65 |
| METHYLPHENIDATE (DL;D;L;ISOMERS) | 1724 | 137 | 29,541.24 | 215.63 |
| BARBITURIC ACID DERIVIATIVE OR SALT [PER 21C | 2100 | 3 | 35 | 11.67 |
| BUTALBITAL | 2165 | 80 | 2,660 | 33.25 |
| CODEINE | 9050 | 146 | 51,328.42 | 351.56 |
| BUPRENORPHINE | 9064 | 124 | 7,282.24 | 58.73 |
| OXYCODONE | 9143 | 142 | 56,540.54 | 398.17 |
| HYDROMORPHONE | 9150 | 117 | 1,421.69 | 12.15 |
| HYDROCODONE | 9193 | 140 | 6,189.03 | 44.21 |
| MEPERIDINE (PETHIDINE) | 9230 | 21 | 631.51 | 30.07 |
| METHADONE | 9250B | 50 | 1,562.96 | 31.26 |
| MORPHINE | 9300 | 105 | 10,432.65 | 99.36 |
| OPIUM TINCTURE (LAUDANUM) | 9630 | 1 | 1.18 | 1.18 |
| OPIUM POWDERED | 9639 | 8 | 8.28 | 1.04 |
| OXYMORPHONE | 9652 | 30 | 581.34 | 19.38 |
| TAPENTADOL | 9780 | 77 | 5,487 | 71.26 |
| FENTANYL BASE | 9801 | 103 | 238 | 2.31 |

STATE:DISTRICT OF COLUMBIA BUSINESS ACTIVITY:B - HOSPITALS

| DRUG NAME | DRUG CODE | BUYERS | TOTAL GRAMS | AVG GRAMS |
|---|---|---|---|---|
| AMPHETAMINE | 1100 | 11 | 2,396.06 | 217.82 |
| LISDEXAMFETAMINE | 1205 | 4 | 286.74 | 71.68 |
| METHYLPHENIDATE (DL;D;L;ISOMERS) | 1724 | 16 | 3,306.06 | 206.63 |
| AMOBARBITAL (SCHEDULE 2) | 2125 | 1 | 2.28 | 2.28 |
| BUTALBITAL | 2165 | 1 | 5 | 5 |
| PENTOBARBITAL (SCHEDULE 2) | 2270 | 5 | 893.17 | 178.63 |
| COCAINE | 9041L | 7 | 97.28 | 13.9 |

| | | | | |
|---|---|---|---|---|
| CODEINE | 9050 | 17 | 2,545.22 | 149.72 |
| BUPRENORPHINE | 9064 | 21 | 98.81 | 4.71 |
| OXYCODONE | 9143 | 21 | 10,401.01 | 495.29 |
| HYDROMORPHONE | 9150 | 24 | 1,059.10 | 44.13 |
| HYDROCODONE | 9193 | 19 | 669.08 | 35.21 |
| MEPERIDINE (PETHIDINE) | 9230 | 10 | 132.9 | 13.29 |
| METHADONE | 9250B | 14 | 1,810.40 | 129.31 |
| MORPHINE | 9300 | 21 | 7,071.56 | 336.74 |
| OPIUM POWDERED | 9639 | 5 | 38.16 | 7.63 |
| OXYMORPHONE | 9652 | 4 | 18.22 | 4.55 |
| REMIFENTANIL | 9739 | 6 | 3.09 | 0.52 |
| SUFENTANIL BASE | 9740 | 3 | 0.04 | 0.01 |
| TAPENTADOL | 9780 | 5 | 258.35 | 51.67 |
| FENTANYL BASE | 9801 | 24 | 159.2 | 6.63 |

STATE:DISTRICT OF COLUMBIA BUSINESS ACTIVITY:C - PRACTITIONERS

| DRUG NAME | DRUG CODE | BUYERS | TOTAL GRAMS | AVG GRAMS |
|---|---|---|---|---|
| AMPHETAMINE | 1100 | 1 | 0.37 | 0.37 |
| COCAINE | 9041L | 1 | 0.14 | 0.14 |
| CODEINE | 9050 | 15 | 55.81 | 3.72 |
| BUPRENORPHINE | 9064 | 18 | 1.12 | 0.06 |
| OXYCODONE | 9143 | 4 | 8.07 | 2.02 |
| HYDROMORPHONE | 9150 | 4 | 3.19 | 0.8 |
| HYDROCODONE | 9193 | 6 | 17.71 | 2.95 |
| MEPERIDINE (PETHIDINE) | 9230 | 5 | 32.68 | 6.54 |
| MORPHINE | 9300 | 7 | 6.09 | 0.87 |
| NOROXYMORPHONE | 9668 | 1 | 9.28 | 9.28 |
| FENTANYL BASE | 9801 | 14 | 0.41 | 0.03 |

STATE:DISTRICT OF COLUMBIA BUSINESS ACTIVITY:D - TEACHING INSTITUTIONS

| DRUG NAME | DRUG CODE | BUYERS | TOTAL GRAMS | AVG GRAMS |
|---|---|---|---|---|
| BUPRENORPHINE | 9064 | 1 | 0.01 | 0.01 |

STATE:DISTRICT OF COLUMBIA BUSINESS ACTIVITY:M - MID-LEVEL PRACTITIONERS

| DRUG NAME | DRUG CODE | BUYERS | TOTAL GRAMS | AVG GRAMS |
|---|---|---|---|---|
| CODEINE | 9050 | 2 | 0.87 | 0.43 |
| FENTANYL BASE | 9801 | 1 | 0.06 | 0.06 |

STATE:DISTRICT OF COLUMBIA BUSINESS ACTIVITY:N-U NARCOTIC TREATMENT
PROGRAMS

| DRUG NAME | DRUG CODE | BUYERS | TOTAL GRAMS | AVG GRAMS |
|---|---|---|---|---|
| METHADONE | 9250B | 5 | 37,072.88 | 7,414.58 |

STATE:FLORIDA BUSINESS ACTIVITY:A - PHARMACIES

| DRUG NAME | DRUG CODE | BUYERS | TOTAL GRAMS | AVG GRAMS |
|---|---|---|---|---|
| AMPHETAMINE | 1100 | 4,401 | 1,066,088.57 | 242.24 |
| DL-METHAMPHETAMINE RACEMIC BASE | 1105B | 23 | 35.76 | 1.55 |
| D-METHAMPHETAMINE | 1105D | 47 | 81.14 | 1.73 |
| LISDEXAMFETAMINE | 1205 | 4,041 | 546,535.74 | 135.25 |
| METHYLPHENIDATE (DL;D;L;ISOMERS) | 1724 | 4,280 | 800,182.91 | 186.96 |
| GAMMA HYDROXYBUTYRIC ACID(FDA APPROVED | 2012 | 1 | 148.61 | 148.61 |
| BARBITURIC ACID DERIVIATIVE OR SALT [PER 21C | 2100 | 41 | 315 | 7.68 |
| BUTALBITAL | 2165 | 3,036 | 144,290 | 47.53 |
| PENTOBARBITAL (SCHEDULE 2) | 2270 | 1 | 5.47 | 5.47 |
| SECOBARBITAL (SCHEDULE 2) | 2315 | 11 | 311.27 | 28.3 |
| NABILONE | 7379 | 14 | 3.5 | 0.25 |
| COCAINE | 9041L | 11 | 8.79 | 0.8 |
| CODEINE | 9050 | 4,556 | 667,608.66 | 146.53 |

| | | | | |
|---|---|---|---|---|
| BUPRENORPHINE | 9064 | 4,017 | 164,685.28 | 41 |
| DIHYDROCODEINE | 9120 | 74 | 340.21 | 4.6 |
| OXYCODONE | 9143 | 4,605 | 3,919,498.45 | 851.14 |
| HYDROMORPHONE | 9150 | 4,132 | 194,646.92 | 47.11 |
| HYDROCODONE | 9193 | 4,563 | 1,473,196.61 | 322.86 |
| LEVORPHANOL | 9220L | 25 | 79.17 | 3.17 |
| MEPERIDINE (PETHIDINE) | 9230 | 1,637 | 30,345.19 | 18.54 |
| METHADONE | 9250B | 3,493 | 248,155.20 | 71.04 |
| MORPHINE | 9300 | 4,303 | 1,602,173.23 | 372.34 |
| OPIUM TINCTURE (LAUDANUM) | 9630 | 3 | 37.84 | 12.61 |
| OPIUM POWDERED | 9639 | 192 | 520.92 | 2.71 |
| OXYMORPHONE | 9652 | 2,922 | 114,977.01 | 39.35 |
| ALFENTANIL | 9737 | 3 | 1.73 | 0.58 |
| REMIFENTANIL | 9739 | 3 | 0.19 | 0.06 |
| SUFENTANIL BASE | 9740 | 4 | 7.83 | 1.96 |
| TAPENTADOL | 9780 | 3,050 | 459,699 | 150.72 |
| FENTANYL BASE | 9801 | 4,213 | 24,325.08 | 5.77 |

STATE:FLORIDA BUSINESS ACTIVITY:B - HOSPITALS

| DRUG NAME | DRUG CODE | BUYERS | TOTAL GRAMS | AVG GRAMS |
|---|---|---|---|---|
| AMPHETAMINE | 1100 | 187 | 29,171.82 | 156 |
| D-METHAMPHETAMINE | 1105D | 1 | 3.62 | 3.62 |
| LISDEXAMFETAMINE | 1205 | 78 | 7,434.37 | 95.31 |
| METHYLPHENIDATE (DL;D;L;ISOMERS) | 1724 | 267 | 32,541.89 | 121.88 |
| AMOBARBITAL (SCHEDULE 2) | 2125 | 5 | 28.71 | 5.74 |
| BUTALBITAL | 2165 | 42 | 5,810 | 138.33 |
| PENTOBARBITAL (SCHEDULE 2) | 2270 | 40 | 6,170.63 | 154.27 |
| COCAINE | 9041L | 242 | 1,165.85 | 4.82 |
| CODEINE | 9050 | 427 | 28,744.70 | 67.32 |
| BUPRENORPHINE | 9064 | 236 | 5,223.72 | 22.13 |
| OXYCODONE | 9143 | 557 | 155,427.82 | 279.04 |
| HYDROMORPHONE | 9150 | 572 | 15,113.54 | 26.42 |

| | | | | |
|---|---|---|---|---|
| HYDROCODONE | 9193 | 554 | 82,312.32 | 148.58 |
| MEPERIDINE (PETHIDINE) | 9230 | 461 | 8,600.92 | 18.66 |
| METHADONE | 9250B | 322 | 13,383.41 | 41.56 |
| MORPHINE | 9300 | 627 | 134,679.75 | 214.8 |
| OPIUM POWDERED | 9639 | 231 | 1,351.44 | 5.85 |
| OXYMORPHONE | 9652 | 34 | 407.6 | 11.99 |
| ALFENTANIL | 9737 | 27 | 16.03 | 0.59 |
| REMIFENTANIL | 9739 | 165 | 102.05 | 0.62 |
| SUFENTANIL BASE | 9740 | 98 | 2.51 | 0.03 |
| TAPENTADOL | 9780 | 87 | 5,984.29 | 68.78 |
| FENTANYL BASE | 9801 | 759 | 2,502.85 | 3.3 |

STATE:FLORIDA BUSINESS ACTIVITY:C - PRACTITIONERS

| DRUG NAME | DRUG CODE | BUYERS | TOTAL GRAMS | AVG GRAMS |
|---|---|---|---|---|
| AMPHETAMINE | 1100 | 10 | 14.47 | 1.45 |
| LISDEXAMFETAMINE | 1205 | 1 | 120.19 | 120.19 |
| METHYLPHENIDATE (DL;D;L;ISOMERS) | 1724 | 5 | 317.86 | 63.57 |
| BUTALBITAL | 2165 | 1 | 70 | 70 |
| PENTOBARBITAL (SCHEDULE 2) | 2270 | 233 | 157,848.67 | 677.46 |
| COCAINE | 9041L | 42 | 63.25 | 1.51 |
| CODEINE | 9050 | 104 | 1,388.26 | 13.35 |
| BUPRENORPHINE | 9064 | 1,817 | 1,103.40 | 0.61 |
| DIHYDROCODEINE | 9120 | 48 | 24.26 | 0.51 |
| OXYCODONE | 9143 | 111 | 10,456.56 | 94.2 |
| HYDROMORPHONE | 9150 | 773 | 482.8 | 0.62 |
| HYDROCODONE | 9193 | 896 | 2,977.81 | 3.32 |
| MEPERIDINE (PETHIDINE) | 9230 | 308 | 3,116.48 | 10.12 |
| METHADONE | 9250B | 18 | 54.2 | 3.01 |
| MORPHINE | 9300 | 480 | 1,800.09 | 3.75 |
| OXYMORPHONE | 9652 | 13 | 40.66 | 3.13 |
| NOROXYMORPHONE | 9668 | 19 | 99.67 | 5.25 |
| ALFENTANIL | 9737 | 6 | 2.05 | 0.34 |

| | | | | |
|---|---|---|---|---|
| REMIFENTANIL | 9739 | 16 | 0.74 | 0.05 |
| SUFENTANIL BASE | 9740 | 8 | 0.11 | 0.01 |
| FENTANYL BASE | 9801 | 1,189 | 88.67 | 0.07 |

STATE:FLORIDA BUSINESS ACTIVITY:D - TEACHING INSTITUTIONS

| DRUG NAME | DRUG CODE | BUYERS | TOTAL GRAMS | AVG GRAMS |
|---|---|---|---|---|
| BUPRENORPHINE | 9064 | 1 | 0 | 0 |

STATE:FLORIDA BUSINESS ACTIVITY:M - MID-LEVEL PRACTITIONERS

| DRUG NAME | DRUG CODE | BUYERS | TOTAL GRAMS | AVG GRAMS |
|---|---|---|---|---|
| PENTOBARBITAL (SCHEDULE 2) | 2270 | 52 | 111,714.40 | 2,148.35 |
| BUPRENORPHINE | 9064 | 2 | 0.05 | 0.02 |

STATE:FLORIDA BUSINESS ACTIVITY:N-U NARCOTIC TREATMENT PROGRAMS

| DRUG NAME | DRUG CODE | BUYERS | TOTAL GRAMS | AVG GRAMS |
|---|---|---|---|---|
| BUPRENORPHINE | 9064 | 51 | 1,423.99 | 27.92 |
| METHADONE | 9250B | 56 | 584,855.32 | 10,443.84 |

STATE:GEORGIA BUSINESS ACTIVITY:A - PHARMACIES

| DRUG NAME | DRUG CODE | BUYERS | TOTAL GRAMS | AVG GRAMS |
|---|---|---|---|---|
| AMPHETAMINE | 1100 | 2,278 | 686,476.32 | 301.35 |
| DL-METHAMPHETAMINE RACEMIC BASE | 1105B | 9 | 15.27 | 1.7 |
| D-METHAMPHETAMINE | 1105D | 39 | 120.11 | 3.08 |
| LISDEXAMFETAMINE | 1205 | 2,196 | 415,020.30 | 188.99 |
| METHYLPHENIDATE (DL;D;L;ISOMERS) | 1724 | 2,266 | 642,485.69 | 283.53 |
| BARBITURIC ACID DERIVIATIVE OR SALT [PER 21C | 2100 | 33 | 325 | 9.85 |

| | | | | |
|---|---|---|---|---|
| BUTALBITAL | 2165 | 1,613 | 63,600 | 39.43 |
| SECOBARBITAL (SCHEDULE 2) | 2315 | 3 | 64.09 | 21.36 |
| COCAINE | 9041L | 7 | 20.61 | 2.94 |
| CODEINE | 9050 | 2,282 | 402,505.48 | 176.38 |
| BUPRENORPHINE | 9064 | 2,133 | 54,960.40 | 25.77 |
| DIHYDROCODEINE | 9120 | 335 | 1,431.33 | 4.27 |
| OXYCODONE | 9143 | 2,298 | 1,758,257.94 | 765.13 |
| HYDROMORPHONE | 9150 | 2,066 | 30,328.57 | 14.68 |
| HYDROCODONE | 9193 | 2,310 | 1,016,134.24 | 439.88 |
| LEVORPHANOL | 9220L | 4 | 4.53 | 1.13 |
| MEPERIDINE (PETHIDINE) | 9230 | 1,522 | 39,545.51 | 25.98 |
| METHADONE | 9250B | 1,941 | 108,476.32 | 55.89 |
| MORPHINE | 9300 | 2,226 | 439,600.72 | 197.48 |
| OPIUM POWDERED | 9639 | 129 | 223.92 | 1.74 |
| OXYMORPHONE | 9652 | 1,418 | 34,629.31 | 24.42 |
| ALFENTANIL | 9737 | 1 | 0.18 | 0.18 |
| REMIFENTANIL | 9739 | 1 | 0.08 | 0.08 |
| SUFENTANIL BASE | 9740 | 4 | 0.19 | 0.05 |
| TAPENTADOL | 9780 | 1,795 | 285,409.50 | 159 |
| FENTANYL BASE | 9801 | 2,196 | 11,097.65 | 5.05 |

STATE:GEORGIA BUSINESS ACTIVITY:B - HOSPITALS

| DRUG NAME | DRUG CODE | BUYERS | TOTAL GRAMS | AVG GRAMS |
|---|---|---|---|---|
| AMPHETAMINE | 1100 | 97 | 19,262.40 | 198.58 |
| DL-METHAMPHETAMINE RACEMIC BASE | 1105B | 1 | 9.24 | 9.24 |
| LISDEXAMFETAMINE | 1205 | 54 | 5,063 | 93.76 |
| METHYLPHENIDATE (DL;D;L;ISOMERS) | 1724 | 145 | 22,370.97 | 154.28 |
| BARBITURIC ACID DERIVIATIVE OR SALT [PER 21C | 2100 | 1 | 10 | 10 |
| AMOBARBITAL (SCHEDULE 2) | 2125 | 5 | 34.63 | 6.93 |
| BUTALBITAL | 2165 | 20 | 250 | 12.5 |
| PENTOBARBITAL (SCHEDULE 2) | 2270 | 13 | 1,563.51 | 120.27 |
| COCAINE | 9041L | 88 | 633.78 | 7.2 |

| | | | | |
|---|---|---|---|---|
| CODEINE | 9050 | 194 | 15,998.33 | 82.47 |
| BUPRENORPHINE | 9064 | 108 | 1,385.86 | 12.83 |
| DIHYDROCODEINE | 9120 | 1 | 6.41 | 6.41 |
| OXYCODONE | 9143 | 224 | 69,133.16 | 308.63 |
| HYDROMORPHONE | 9150 | 205 | 5,188.60 | 25.31 |
| HYDROCODONE | 9193 | 220 | 58,424.52 | 265.57 |
| MEPERIDINE (PETHIDINE) | 9230 | 177 | 8,808.60 | 49.77 |
| METHADONE | 9250B | 163 | 5,057.06 | 31.02 |
| MORPHINE | 9300 | 220 | 42,143.96 | 191.56 |
| OPIUM POWDERED | 9639 | 90 | 530.64 | 5.9 |
| OXYMORPHONE | 9652 | 16 | 140.79 | 8.8 |
| NOROXYMORPHONE | 9668 | 1 | 5.52 | 5.52 |
| ALFENTANIL | 9737 | 7 | 5.51 | 0.79 |
| REMIFENTANIL | 9739 | 54 | 17.91 | 0.33 |
| SUFENTANIL BASE | 9740 | 41 | 1.05 | 0.03 |
| TAPENTADOL | 9780 | 59 | 3,234 | 54.81 |
| FENTANYL BASE | 9801 | 211 | 1,302.61 | 6.17 |

STATE:GEORGIA BUSINESS ACTIVITY:C - PRACTITIONERS

| DRUG NAME | DRUG CODE | BUYERS | TOTAL GRAMS | AVG GRAMS |
|---|---|---|---|---|
| AMPHETAMINE | 1100 | 6 | 229.21 | 38.2 |
| LISDEXAMFETAMINE | 1205 | 1 | 18.5 | 18.5 |
| METHYLPHENIDATE (DL;D;L;ISOMERS) | 1724 | 3 | 8.65 | 2.88 |
| GAMMA HYDROXYBUTYRIC ACID(FDA APPROVED | 2012 | 1 | 2,600.64 | 2,600.64 |
| PENTOBARBITAL (SCHEDULE 2) | 2270 | 157 | 135,027.49 | 860.05 |
| COCAINE | 9041L | 37 | 76.07 | 2.06 |
| CODEINE | 9050 | 166 | 3,908.38 | 23.54 |
| BUPRENORPHINE | 9064 | 797 | 60.77 | 0.08 |
| DIHYDROCODEINE | 9120 | 180 | 124.66 | 0.69 |
| OXYCODONE | 9143 | 116 | 6,914.60 | 59.61 |
| HYDROMORPHONE | 9150 | 392 | 246.14 | 0.63 |
| HYDROCODONE | 9193 | 483 | 6,445.27 | 13.34 |

| | | | | |
|---|---|---|---|---|
| MEPERIDINE (PETHIDINE) | 9230 | 238 | 1,577.96 | 6.63 |
| METHADONE | 9250B | 7 | 34.44 | 4.92 |
| MORPHINE | 9300 | 326 | 1,866.48 | 5.73 |
| OPIUM POWDERED | 9639 | 8 | 25.56 | 3.2 |
| OXYMORPHONE | 9652 | 14 | 40.55 | 2.9 |
| NOROXYMORPHONE | 9668 | 5 | 18.2 | 3.64 |
| ALFENTANIL | 9737 | 6 | 3.03 | 0.5 |
| REMIFENTANIL | 9739 | 6 | 0.96 | 0.16 |
| SUFENTANIL BASE | 9740 | 4 | 0.23 | 0.06 |
| FENTANYL BASE | 9801 | 771 | 100.98 | 0.13 |

STATE:GEORGIA BUSINESS ACTIVITY:D - TEACHING INSTITUTIONS

| DRUG NAME | DRUG CODE | BUYERS | TOTAL GRAMS | AVG GRAMS |
|---|---|---|---|---|
| METHYLPHENIDATE (DL;D;L;ISOMERS) | 1724 | 1 | 17.29 | 17.29 |

STATE:GEORGIA BUSINESS ACTIVITY:M - MID-LEVEL PRACTITIONERS

| DRUG NAME | DRUG CODE | BUYERS | TOTAL GRAMS | AVG GRAMS |
|---|---|---|---|---|
| CODEINE | 9050 | 17 | 32.6 | 1.92 |
| DIHYDROCODEINE | 9120 | 41 | 51.78 | 1.26 |

STATE:GEORGIA BUSINESS ACTIVITY:N-U NARCOTIC TREATMENT PROGRAMS

| DRUG NAME | DRUG CODE | BUYERS | TOTAL GRAMS | AVG GRAMS |
|---|---|---|---|---|
| BUPRENORPHINE | 9064 | 27 | 1,129.91 | 41.85 |
| METHADONE | 9250B | 68 | 348,596.87 | 5,126.42 |

STATE:GUAM BUSINESS ACTIVITY:A - PHARMACIES

| DRUG NAME | DRUG CODE | BUYERS | TOTAL GRAMS | AVG GRAMS |
|---|---|---|---|---|
| AMPHETAMINE | 1100 | 10 | 1,104.08 | 110.41 |
| LISDEXAMFETAMINE | 1205 | 4 | 55.5 | 13.87 |
| METHYLPHENIDATE (DL;D;L;ISOMERS) | 1724 | 9 | 829.89 | 92.21 |
| BUTALBITAL | 2165 | 5 | 35 | 7 |
| CODEINE | 9050 | 18 | 4,424.62 | 245.81 |
| BUPRENORPHINE | 9064 | 2 | 4.48 | 2.24 |
| OXYCODONE | 9143 | 11 | 3,112.56 | 282.96 |
| HYDROMORPHONE | 9150 | 5 | 95.04 | 19.01 |
| HYDROCODONE | 9193 | 11 | 518.05 | 47.1 |
| MEPERIDINE (PETHIDINE) | 9230 | 2 | 39.22 | 19.61 |
| METHADONE | 9250B | 2 | 155.63 | 77.81 |
| MORPHINE | 9300 | 9 | 4,043.43 | 449.27 |
| FENTANYL BASE | 9801 | 10 | 20 | 2 |

STATE:GUAM BUSINESS ACTIVITY:B - HOSPITALS

| DRUG NAME | DRUG CODE | BUYERS | TOTAL GRAMS | AVG GRAMS |
|---|---|---|---|---|
| AMPHETAMINE | 1100 | 4 | 650.62 | 162.65 |
| LISDEXAMFETAMINE | 1205 | 2 | 37.58 | 18.79 |
| METHYLPHENIDATE (DL;D;L;ISOMERS) | 1724 | 5 | 1,015.42 | 203.08 |
| COCAINE | 9041L | 1 | 0.71 | 0.71 |
| CODEINE | 9050 | 6 | 955.51 | 159.25 |
| BUPRENORPHINE | 9064 | 2 | 0.01 | 0.01 |
| OXYCODONE | 9143 | 7 | 852.35 | 121.76 |
| HYDROMORPHONE | 9150 | 8 | 30.85 | 3.86 |
| HYDROCODONE | 9193 | 6 | 291.78 | 48.63 |
| MEPERIDINE (PETHIDINE) | 9230 | 5 | 67.11 | 13.42 |
| METHADONE | 9250B | 1 | 8.94 | 8.94 |
| MORPHINE | 9300 | 7 | 751.63 | 107.38 |
| OPIUM POWDERED | 9639 | 2 | 2.88 | 1.44 |
| ALFENTANIL | 9737 | 1 | 0.03 | 0.03 |
| REMIFENTANIL | 9739 | 1 | 0.5 | 0.5 |

| | | | | |
|---|---|---|---|---|
| FENTANYL BASE | 9801 | 7 | 18.31 | 2.62 |

STATE:GUAM BUSINESS ACTIVITY:C - PRACTITIONERS

| DRUG NAME | DRUG CODE | BUYERS | TOTAL GRAMS | AVG GRAMS |
|---|---|---|---|---|
| CODEINE | 9050 | 3 | 50.83 | 16.94 |
| BUPRENORPHINE | 9064 | 3 | 0.07 | 0.02 |
| HYDROMORPHONE | 9150 | 2 | 0.23 | 0.12 |
| HYDROCODONE | 9193 | 2 | 10.29 | 5.15 |
| MEPERIDINE (PETHIDINE) | 9230 | 3 | 51.53 | 17.18 |
| MORPHINE | 9300 | 3 | 0.81 | 0.27 |
| ALFENTANIL | 9737 | 1 | 0.21 | 0.21 |
| FENTANYL BASE | 9801 | 5 | 0.25 | 0.05 |

STATE:GUAM BUSINESS ACTIVITY:M - MID-LEVEL PRACTITIONERS

| DRUG NAME | DRUG CODE | BUYERS | TOTAL GRAMS | AVG GRAMS |
|---|---|---|---|---|
| PENTOBARBITAL (SCHEDULE 2) | 2270 | 1 | 3,215.42 | 3,215.42 |

STATE:HAWAII BUSINESS ACTIVITY:A - PHARMACIES

| DRUG NAME | DRUG CODE | BUYERS | TOTAL GRAMS | AVG GRAMS |
|---|---|---|---|---|
| AMPHETAMINE | 1100 | 224 | 27,924.75 | 124.66 |
| DL-METHAMPHETAMINE RACEMIC BASE | 1105B | 5 | 12.46 | 2.49 |
| D-METHAMPHETAMINE | 1105D | 4 | 14.86 | 3.72 |
| LISDEXAMFETAMINE | 1205 | 154 | 4,158.85 | 27.01 |
| METHYLPHENIDATE (DL;D;L;ISOMERS) | 1724 | 222 | 33,554.06 | 151.14 |
| BUTALBITAL | 2165 | 131 | 7,915 | 60.42 |
| PENTOBARBITAL (SCHEDULE 2) | 2270 | 1 | 4.56 | 4.56 |
| SECOBARBITAL (SCHEDULE 2) | 2315 | 2 | 64.09 | 32.04 |
| NABILONE | 7379 | 1 | 0.1 | 0.1 |

| | | | | |
|---|---|---|---|---|
| COCAINE | 9041L | 4 | 8.43 | 2.11 |
| CODEINE | 9050 | 241 | 53,325.48 | 221.27 |
| BUPRENORPHINE | 9064 | 207 | 7,005.71 | 33.84 |
| DIHYDROCODEINE | 9120 | 1 | 7.48 | 7.48 |
| OXYCODONE | 9143 | 235 | 172,813.09 | 735.37 |
| HYDROMORPHONE | 9150 | 158 | 2,357.52 | 14.92 |
| HYDROCODONE | 9193 | 236 | 70,269.49 | 297.75 |
| MEPERIDINE (PETHIDINE) | 9230 | 50 | 1,242.54 | 24.85 |
| METHADONE | 9250B | 166 | 15,363.73 | 92.55 |
| MORPHINE | 9300 | 217 | 92,910.79 | 428.16 |
| OPIUM POWDERED | 9639 | 6 | 30.24 | 5.04 |
| OXYMORPHONE | 9652 | 75 | 2,386.81 | 31.82 |
| SUFENTANIL BASE | 9740 | 1 | 0 | 0 |
| TAPENTADOL | 9780 | 101 | 10,745.50 | 106.39 |
| FENTANYL BASE | 9801 | 193 | 667.6 | 3.46 |

STATE:HAWAII BUSINESS ACTIVITY:B - HOSPITALS

| DRUG NAME | DRUG CODE | BUYERS | TOTAL GRAMS | AVG GRAMS |
|---|---|---|---|---|
| AMPHETAMINE | 1100 | 20 | 8,525.64 | 426.28 |
| LISDEXAMFETAMINE | 1205 | 11 | 1,029.60 | 93.6 |
| METHYLPHENIDATE (DL;D;L;ISOMERS) | 1724 | 22 | 12,608.91 | 573.13 |
| BUTALBITAL | 2165 | 1 | 5 | 5 |
| PENTOBARBITAL (SCHEDULE 2) | 2270 | 3 | 819.35 | 273.12 |
| NABILONE | 7379 | 1 | 0.1 | 0.1 |
| COCAINE | 9041L | 14 | 78.71 | 5.62 |
| CODEINE | 9050 | 39 | 4,651.68 | 119.27 |
| BUPRENORPHINE | 9064 | 24 | 199.14 | 8.3 |
| OXYCODONE | 9143 | 42 | 21,104.04 | 502.48 |
| HYDROMORPHONE | 9150 | 38 | 731 | 19.24 |
| HYDROCODONE | 9193 | 39 | 5,963.16 | 152.9 |
| MEPERIDINE (PETHIDINE) | 9230 | 29 | 372.89 | 12.86 |
| METHADONE | 9250B | 29 | 1,317.90 | 45.44 |

| | | | | |
|---|---|---|---|---|
| MORPHINE | 9300 | 44 | 10,230.71 | 232.52 |
| OPIUM POWDERED | 9639 | 15 | 68.76 | 4.58 |
| OXYMORPHONE | 9652 | 6 | 485.12 | 80.85 |
| ALFENTANIL | 9737 | 5 | 1.53 | 0.31 |
| REMIFENTANIL | 9739 | 8 | 5.33 | 0.67 |
| SUFENTANIL BASE | 9740 | 5 | 0.04 | 0.01 |
| TAPENTADOL | 9780 | 8 | 326 | 40.75 |
| FENTANYL BASE | 9801 | 44 | 175.05 | 3.98 |

STATE:HAWAII BUSINESS ACTIVITY:C - PRACTITIONERS

| DRUG NAME | DRUG CODE | BUYERS | TOTAL GRAMS | AVG GRAMS |
|---|---|---|---|---|
| PENTOBARBITAL (SCHEDULE 2) | 2270 | 14 | 37,307.25 | 2,664.80 |
| COCAINE | 9041L | 1 | 1.29 | 1.29 |
| CODEINE | 9050 | 15 | 344.55 | 22.97 |
| BUPRENORPHINE | 9064 | 80 | 4.76 | 0.06 |
| DIHYDROCODEINE | 9120 | 4 | 16.88 | 4.22 |
| OXYCODONE | 9143 | 17 | 1,077.14 | 63.36 |
| HYDROMORPHONE | 9150 | 42 | 26.77 | 0.64 |
| HYDROCODONE | 9193 | 63 | 1,518.18 | 24.1 |
| MEPERIDINE (PETHIDINE) | 9230 | 20 | 150.99 | 7.55 |
| METHADONE | 9250B | 5 | 8.59 | 1.72 |
| MORPHINE | 9300 | 25 | 400.28 | 16.01 |
| OPIUM POWDERED | 9639 | 1 | 1.8 | 1.8 |
| OXYMORPHONE | 9652 | 5 | 3.77 | 0.75 |
| ALFENTANIL | 9737 | 2 | 0.26 | 0.13 |
| FENTANYL BASE | 9801 | 51 | 4.95 | 0.1 |

STATE:HAWAII BUSINESS ACTIVITY:M - MID-LEVEL PRACTITIONERS

| DRUG NAME | DRUG CODE | BUYERS | TOTAL GRAMS | AVG GRAMS |
|---|---|---|---|---|
| DIHYDROCODEINE | 9120 | 1 | 0.21 | 0.21 |

| | | | | |
|---|---|---|---|---|
| MORPHINE | 9300 | 1 | 0.3 | 0.3 |
| FENTANYL BASE | 9801 | 1 | 0.02 | 0.02 |

STATE:HAWAII BUSINESS ACTIVITY:N-U NARCOTIC TREATMENT PROGRAMS

| DRUG NAME | DRUG CODE | BUYERS | TOTAL GRAMS | AVG GRAMS |
|---|---|---|---|---|
| METHADONE | 9250B | 4 | 16,914.89 | 4,228.72 |

STATE:IDAHO BUSINESS ACTIVITY:A - PHARMACIES

| DRUG NAME | DRUG CODE | BUYERS | TOTAL GRAMS | AVG GRAMS |
|---|---|---|---|---|
| AMPHETAMINE | 1100 | 317 | 92,084.39 | 290.49 |
| DL-METHAMPHETAMINE RACEMIC BASE | 1105B | 2 | 18.08 | 9.04 |
| D-METHAMPHETAMINE | 1105D | 14 | 24.5 | 1.75 |
| LISDEXAMFETAMINE | 1205 | 301 | 33,243.64 | 110.44 |
| METHYLPHENIDATE (DL;D;L;ISOMERS) | 1724 | 318 | 109,931.20 | 345.7 |
| BARBITURIC ACID DERIVIATIVE OR SALT [PER 21C | 2100 | 1 | 10 | 10 |
| BUTALBITAL | 2165 | 193 | 8,120 | 42.07 |
| COCAINE | 9041L | 4 | 76.57 | 19.14 |
| CODEINE | 9050 | 318 | 55,588.41 | 174.81 |
| BUPRENORPHINE | 9064 | 302 | 11,590.76 | 38.38 |
| DIHYDROCODEINE | 9120 | 2 | 2.14 | 1.07 |
| OXYCODONE | 9143 | 320 | 241,177.24 | 753.68 |
| HYDROMORPHONE | 9150 | 307 | 7,266.75 | 23.67 |
| HYDROCODONE | 9193 | 322 | 232,911.02 | 723.33 |
| MEPERIDINE (PETHIDINE) | 9230 | 128 | 2,595.44 | 20.28 |
| METHADONE | 9250B | 311 | 29,410.29 | 94.57 |
| MORPHINE | 9300 | 321 | 113,648.82 | 354.05 |
| OPIUM TINCTURE (LAUDANUM) | 9630 | 1 | 1.18 | 1.18 |
| OPIUM POWDERED | 9639 | 26 | 31.32 | 1.2 |
| OXYMORPHONE | 9652 | 234 | 6,751.81 | 28.85 |
| REMIFENTANIL | 9739 | 1 | 0.01 | 0.01 |

| | | | | |
|---|---|---|---|---|
| TAPENTADOL | 9780 | 242 | 24,421.50 | 100.92 |
| FENTANYL BASE | 9801 | 315 | 2,434.35 | 7.73 |

STATE:IDAHO BUSINESS ACTIVITY:B - HOSPITALS

| DRUG NAME | DRUG CODE | BUYERS | TOTAL GRAMS | AVG GRAMS |
|---|---|---|---|---|
| AMPHETAMINE | 1100 | 21 | 1,417.35 | 67.49 |
| LISDEXAMFETAMINE | 1205 | 8 | 104.06 | 13.01 |
| METHYLPHENIDATE (DL;D;L;ISOMERS) | 1724 | 37 | 2,095.40 | 56.63 |
| AMOBARBITAL (SCHEDULE 2) | 2125 | 1 | 0.46 | 0.46 |
| BUTALBITAL | 2165 | 2 | 10 | 5 |
| PENTOBARBITAL (SCHEDULE 2) | 2270 | 3 | 187.75 | 62.58 |
| COCAINE | 9041L | 20 | 172.81 | 8.64 |
| CODEINE | 9050 | 50 | 2,269.52 | 45.39 |
| BUPRENORPHINE | 9064 | 35 | 368.05 | 10.52 |
| OXYCODONE | 9143 | 63 | 13,399.05 | 212.68 |
| HYDROMORPHONE | 9150 | 55 | 926.9 | 16.85 |
| HYDROCODONE | 9193 | 64 | 11,432.64 | 178.64 |
| MEPERIDINE (PETHIDINE) | 9230 | 42 | 837.08 | 19.93 |
| METHADONE | 9250B | 44 | 1,708.39 | 38.83 |
| MORPHINE | 9300 | 60 | 11,199.25 | 186.65 |
| OPIUM POWDERED | 9639 | 19 | 150.12 | 7.9 |
| OXYMORPHONE | 9652 | 7 | 32.56 | 4.65 |
| ALFENTANIL | 9737 | 7 | 5.2 | 0.74 |
| REMIFENTANIL | 9739 | 19 | 8.54 | 0.45 |
| SUFENTANIL BASE | 9740 | 16 | 0.17 | 0.01 |
| TAPENTADOL | 9780 | 10 | 306 | 30.6 |
| FENTANYL BASE | 9801 | 60 | 242.18 | 4.04 |

STATE:IDAHO BUSINESS ACTIVITY:C - PRACTITIONERS

| DRUG NAME | DRUG CODE | BUYERS | TOTAL GRAMS | AVG GRAMS |
|---|---|---|---|---|

| | | | | |
|---|---|---|---|---|
| PENTOBARBITAL (SCHEDULE 2) | 2270 | 72 | 77,532.75 | 1,076.84 |
| COCAINE | 9041L | 2 | 12.43 | 6.21 |
| CODEINE | 9050 | 36 | 1,253.48 | 34.82 |
| BUPRENORPHINE | 9064 | 125 | 8.94 | 0.07 |
| DIHYDROCODEINE | 9120 | 9 | 210.96 | 23.44 |
| OXYCODONE | 9143 | 15 | 157.63 | 10.51 |
| HYDROMORPHONE | 9150 | 88 | 52.43 | 0.6 |
| HYDROCODONE | 9193 | 122 | 508.78 | 4.17 |
| MEPERIDINE (PETHIDINE) | 9230 | 41 | 293.59 | 7.16 |
| METHADONE | 9250B | 2 | 14.67 | 7.33 |
| MORPHINE | 9300 | 77 | 244.62 | 3.18 |
| OXYMORPHONE | 9652 | 3 | 0.55 | 0.18 |
| ALFENTANIL | 9737 | 1 | 0.12 | 0.12 |
| REMIFENTANIL | 9739 | 4 | 0.39 | 0.1 |
| FENTANYL BASE | 9801 | 122 | 25.27 | 0.21 |

STATE:IDAHO BUSINESS ACTIVITY:D - TEACHING INSTITUTIONS

| DRUG NAME | DRUG CODE | BUYERS | TOTAL GRAMS | AVG GRAMS |
|---|---|---|---|---|
| BUPRENORPHINE | 9064 | 1 | 0 | 0 |
| HYDROMORPHONE | 9150 | 1 | 0.07 | 0.07 |

STATE:IDAHO BUSINESS ACTIVITY:M - MID-LEVEL PRACTITIONERS

| DRUG NAME | DRUG CODE | BUYERS | TOTAL GRAMS | AVG GRAMS |
|---|---|---|---|---|
| PENTOBARBITAL (SCHEDULE 2) | 2270 | 8 | 13,795.41 | 1,724.43 |
| CODEINE | 9050 | 6 | 66.44 | 11.07 |
| HYDROMORPHONE | 9150 | 1 | 0.09 | 0.09 |
| HYDROCODONE | 9193 | 2 | 4.24 | 2.12 |
| MEPERIDINE (PETHIDINE) | 9230 | 1 | 0.87 | 0.87 |
| MORPHINE | 9300 | 2 | 0.11 | 0.05 |
| FENTANYL BASE | 9801 | 3 | 0.09 | 0.03 |

STATE:IDAHO BUSINESS ACTIVITY:N-U NARCOTIC TREATMENT PROGRAMS

| DRUG NAME | DRUG CODE | BUYERS | TOTAL GRAMS | AVG GRAMS |
|---|---|---|---|---|
| BUPRENORPHINE | 9064 | 3 | 177.67 | 59.22 |
| METHADONE | 9250B | 3 | 19,963.01 | 6,654.34 |

STATE:ILLINOIS BUSINESS ACTIVITY:A - PHARMACIES

| DRUG NAME | DRUG CODE | BUYERS | TOTAL GRAMS | AVG GRAMS |
|---|---|---|---|---|
| AMPHETAMINE | 1100 | 2,264 | 710,076.51 | 313.64 |
| DL-METHAMPHETAMINE RACEMIC BASE | 1105B | 5 | 12.86 | 2.57 |
| D-METHAMPHETAMINE | 1105D | 55 | 204.87 | 3.72 |
| LISDEXAMFETAMINE | 1205 | 2,007 | 282,441.74 | 140.73 |
| METHYLPHENIDATE (DL;D;L;ISOMERS) | 1724 | 2,257 | 705,682.79 | 312.66 |
| BARBITURIC ACID DERIVIATIVE OR SALT [PER 21C | 2100 | 18 | 265 | 14.72 |
| BUTALBITAL | 2165 | 1,395 | 61,855 | 44.34 |
| SECOBARBITAL (SCHEDULE 2) | 2315 | 10 | 320.43 | 32.04 |
| NABILONE | 7379 | 7 | 0.8 | 0.11 |
| COCAINE | 9041L | 5 | 45.1 | 9.02 |
| CODEINE | 9050 | 2,366 | 674,985.05 | 285.29 |
| BUPRENORPHINE | 9064 | 1,997 | 45,044.56 | 22.56 |
| DIHYDROCODEINE | 9120 | 20 | 69.43 | 3.47 |
| OXYCODONE | 9143 | 2,207 | 659,746.47 | 298.93 |
| HYDROMORPHONE | 9150 | 1,964 | 35,440.24 | 18.04 |
| HYDROCODONE | 9193 | 2,359 | 1,284,628.42 | 544.56 |
| LEVORPHANOL | 9220L | 28 | 29.02 | 1.04 |
| MEPERIDINE (PETHIDINE) | 9230 | 326 | 8,451.37 | 25.92 |
| METHADONE | 9250B | 1,588 | 55,923.16 | 35.22 |
| MORPHINE | 9300 | 2,111 | 407,033.93 | 192.82 |
| OPIUM TINCTURE (LAUDANUM) | 9630 | 9 | 34.27 | 3.81 |
| OPIUM POWDERED | 9639 | 140 | 282.6 | 2.02 |

| | | | | |
|---|---|---|---|---|
| OXYMORPHONE | 9652 | 1,132 | 23,698.37 | 20.93 |
| REMIFENTANIL | 9739 | 1 | 0.01 | 0.01 |
| TAPENTADOL | 9780 | 1,441 | 177,993 | 123.52 |
| FENTANYL BASE | 9801 | 2,138 | 12,441.10 | 5.82 |

STATE:ILLINOIS BUSINESS ACTIVITY:B - HOSPITALS

| DRUG NAME | DRUG CODE | BUYERS | TOTAL GRAMS | AVG GRAMS |
|---|---|---|---|---|
| AMPHETAMINE | 1100 | 134 | 7,960.03 | 59.4 |
| DL-METHAMPHETAMINE RACEMIC BASE | 1105B | 1 | 2.01 | 2.01 |
| D-METHAMPHETAMINE | 1105D | 2 | 2.81 | 1.41 |
| LISDEXAMFETAMINE | 1205 | 49 | 1,929.70 | 39.38 |
| METHYLPHENIDATE (DL;D;L;ISOMERS) | 1724 | 198 | 10,328.03 | 52.16 |
| GAMMA HYDROXYBUTYRIC ACID(FDA APPROVED | 2012 | 1 | 3,046.46 | 3,046.46 |
| BARBITURIC ACID DERIVIATIVE OR SALT [PER 21C | 2100 | 1 | 5 | 5 |
| AMOBARBITAL (SCHEDULE 2) | 2125 | 11 | 30.53 | 2.78 |
| BUTALBITAL | 2165 | 30 | 315 | 10.5 |
| PENTOBARBITAL (SCHEDULE 2) | 2270 | 23 | 3,690.26 | 160.45 |
| COCAINE | 9041L | 136 | 990.54 | 7.28 |
| CODEINE | 9050 | 240 | 13,920.84 | 58 |
| BUPRENORPHINE | 9064 | 145 | 1,164.64 | 8.03 |
| OXYCODONE | 9143 | 224 | 31,073.83 | 138.72 |
| HYDROMORPHONE | 9150 | 231 | 7,653.02 | 33.13 |
| HYDROCODONE | 9193 | 241 | 67,678.50 | 280.82 |
| MEPERIDINE (PETHIDINE) | 9230 | 184 | 4,947.40 | 26.89 |
| METHADONE | 9250B | 184 | 7,355.34 | 39.97 |
| MORPHINE | 9300 | 233 | 43,059.26 | 184.8 |
| OPIUM POWDERED | 9639 | 124 | 516.24 | 4.16 |
| OXYMORPHONE | 9652 | 34 | 569.51 | 16.75 |
| ALFENTANIL | 9737 | 22 | 7.52 | 0.34 |
| REMIFENTANIL | 9739 | 82 | 56.2 | 0.69 |
| SUFENTANIL BASE | 9740 | 78 | 2.14 | 0.03 |
| TAPENTADOL | 9780 | 46 | 1,716.50 | 37.32 |

| | | | | |
|---|---|---|---|---|
| FENTANYL BASE | 9801 | 232 | 1,307.99 | 5.64 |

STATE:ILLINOIS BUSINESS ACTIVITY:C - PRACTITIONERS

| DRUG NAME | DRUG CODE | BUYERS | TOTAL GRAMS | AVG GRAMS |
|---|---|---|---|---|
| AMPHETAMINE | 1100 | 2 | 4.08 | 2.04 |
| METHYLPHENIDATE (DL;D;L;ISOMERS) | 1724 | 1 | 19.8 | 19.8 |
| PENTOBARBITAL (SCHEDULE 2) | 2270 | 300 | 230,091.20 | 766.97 |
| COCAINE | 9041L | 45 | 366.8 | 8.15 |
| CODEINE | 9050 | 297 | 3,876.27 | 13.05 |
| BUPRENORPHINE | 9064 | 858 | 75.35 | 0.09 |
| DIHYDROCODEINE | 9120 | 8 | 144.48 | 18.06 |
| OXYCODONE | 9143 | 56 | 2,220.34 | 39.65 |
| HYDROMORPHONE | 9150 | 411 | 324.66 | 0.79 |
| HYDROCODONE | 9193 | 581 | 6,474.72 | 11.14 |
| MEPERIDINE (PETHIDINE) | 9230 | 146 | 1,373.49 | 9.41 |
| METHADONE | 9250B | 27 | 36.31 | 1.34 |
| MORPHINE | 9300 | 279 | 628.89 | 2.25 |
| OPIUM POWDERED | 9639 | 9 | 7.56 | 0.84 |
| OXYMORPHONE | 9652 | 5 | 0.89 | 0.18 |
| NOROXYMORPHONE | 9668 | 7 | 52.65 | 7.52 |
| ALFENTANIL | 9737 | 8 | 8.53 | 1.07 |
| REMIFENTANIL | 9739 | 10 | 0.44 | 0.04 |
| SUFENTANIL BASE | 9740 | 1 | 0 | 0 |
| FENTANYL BASE | 9801 | 609 | 101.24 | 0.17 |

STATE:ILLINOIS BUSINESS ACTIVITY:D - TEACHING INSTITUTIONS

| DRUG NAME | DRUG CODE | BUYERS | TOTAL GRAMS | AVG GRAMS |
|---|---|---|---|---|
| METHYLPHENIDATE (DL;D;L;ISOMERS) | 1724 | 2 | 0.52 | 0.26 |
| PENTOBARBITAL (SCHEDULE 2) | 2270 | 2 | 8,003.91 | 4,001.96 |
| CODEINE | 9050 | 1 | 2.39 | 2.39 |

| | | | | |
|---|---|---|---|---|
| BUPRENORPHINE | 9064 | 2 | 0.49 | 0.25 |
| HYDROMORPHONE | 9150 | 1 | 14.23 | 14.23 |
| HYDROCODONE | 9193 | 1 | 9.57 | 9.57 |
| METHADONE | 9250B | 1 | 17.53 | 17.53 |
| MORPHINE | 9300 | 1 | 53.3 | 53.3 |
| REMIFENTANIL | 9739 | 1 | 0.01 | 0.01 |
| FENTANYL BASE | 9801 | 1 | 1.06 | 1.06 |

STATE:ILLINOIS BUSINESS ACTIVITY:M - MID-LEVEL PRACTITIONERS

| DRUG NAME | DRUG CODE | BUYERS | TOTAL GRAMS | AVG GRAMS |
|---|---|---|---|---|
| PENTOBARBITAL (SCHEDULE 2) | 2270 | 11 | 14,695.87 | 1,335.99 |
| CODEINE | 9050 | 6 | 4.38 | 0.73 |

STATE:ILLINOIS BUSINESS ACTIVITY:N-U NARCOTIC TREATMENT PROGRAMS

| DRUG NAME | DRUG CODE | BUYERS | TOTAL GRAMS | AVG GRAMS |
|---|---|---|---|---|
| BUPRENORPHINE | 9064 | 9 | 1,327.47 | 147.5 |
| METHADONE | 9250B | 72 | 413,370.21 | 5,741.25 |

STATE:INDIANA BUSINESS ACTIVITY:A - PHARMACIES

| DRUG NAME | DRUG CODE | BUYERS | TOTAL GRAMS | AVG GRAMS |
|---|---|---|---|---|
| AMPHETAMINE | 1100 | 1,212 | 479,839.90 | 395.91 |
| DL-METHAMPHETAMINE RACEMIC BASE | 1105B | 7 | 20.09 | 2.87 |
| D-METHAMPHETAMINE | 1105D | 21 | 71.9 | 3.42 |
| LISDEXAMFETAMINE | 1205 | 1,196 | 329,459.19 | 275.47 |
| METHYLPHENIDATE (DL;D;L;ISOMERS) | 1724 | 1,216 | 486,109.88 | 399.76 |
| BARBITURIC ACID DERIVIATIVE OR SALT [PER 21C | 2100 | 16 | 175 | 10.94 |
| BUTALBITAL | 2165 | 734 | 29,970 | 40.83 |
| NABILONE | 7379 | 2 | 0.3 | 0.15 |

| | | | | |
|---|---|---|---|---|
| COCAINE | 9041L | 3 | 10.18 | 3.39 |
| CODEINE | 9050 | 1,195 | 227,357.70 | 190.26 |
| BUPRENORPHINE | 9064 | 1,146 | 58,931.47 | 51.42 |
| DIHYDROCODEINE | 9120 | 8 | 16.02 | 2 |
| OXYCODONE | 9143 | 1,226 | 999,374.16 | 815.15 |
| HYDROMORPHONE | 9150 | 1,088 | 22,615.86 | 20.79 |
| HYDROCODONE | 9193 | 1,231 | 955,540.23 | 776.23 |
| LEVORPHANOL | 9220L | 45 | 100.99 | 2.24 |
| MEPERIDINE (PETHIDINE) | 9230 | 389 | 7,027.78 | 18.07 |
| METHADONE | 9250B | 1,055 | 85,182.92 | 80.74 |
| MORPHINE | 9300 | 1,196 | 373,154.37 | 312 |
| OPIUM TINCTURE (LAUDANUM) | 9630 | 13 | 81.57 | 6.27 |
| OPIUM POWDERED | 9639 | 100 | 307.08 | 3.07 |
| OXYMORPHONE | 9652 | 865 | 31,460.98 | 36.37 |
| REMIFENTANIL | 9739 | 1 | 0.1 | 0.1 |
| SUFENTANIL BASE | 9740 | 1 | 0.03 | 0.03 |
| TAPENTADOL | 9780 | 935 | 159,839.50 | 170.95 |
| FENTANYL BASE | 9801 | 1,179 | 8,190.21 | 6.95 |

STATE:INDIANA BUSINESS ACTIVITY:B - HOSPITALS

| DRUG NAME | DRUG CODE | BUYERS | TOTAL GRAMS | AVG GRAMS |
|---|---|---|---|---|
| AMPHETAMINE | 1100 | 122 | 7,618.51 | 62.45 |
| LISDEXAMFETAMINE | 1205 | 72 | 3,798.12 | 52.75 |
| METHYLPHENIDATE (DL;D;L;ISOMERS) | 1724 | 142 | 7,718.98 | 54.36 |
| AMOBARBITAL (SCHEDULE 2) | 2125 | 2 | 7.75 | 3.87 |
| BUTALBITAL | 2165 | 28 | 345 | 12.32 |
| PENTOBARBITAL (SCHEDULE 2) | 2270 | 14 | 6,229.42 | 444.96 |
| COCAINE | 9041L | 122 | 808.84 | 6.63 |
| CODEINE | 9050 | 192 | 6,151.78 | 32.04 |
| BUPRENORPHINE | 9064 | 124 | 1,999.53 | 16.13 |
| OXYCODONE | 9143 | 243 | 43,023.81 | 177.05 |
| HYDROMORPHONE | 9150 | 246 | 5,673.10 | 23.06 |

| | | | | |
|---|---|---|---|---|
| HYDROCODONE | 9193 | 257 | 53,207.03 | 207.03 |
| LEVORPHANOL | 9220L | 1 | 0.23 | 0.23 |
| MEPERIDINE (PETHIDINE) | 9230 | 218 | 7,767.03 | 35.63 |
| METHADONE | 9250B | 161 | 3,964.28 | 24.62 |
| MORPHINE | 9300 | 267 | 36,580.63 | 137.01 |
| OPIUM POWDERED | 9639 | 118 | 724.32 | 6.14 |
| OXYMORPHONE | 9652 | 27 | 95.16 | 3.52 |
| ALFENTANIL | 9737 | 33 | 9.1 | 0.28 |
| REMIFENTANIL | 9739 | 82 | 31.61 | 0.39 |
| SUFENTANIL BASE | 9740 | 47 | 1.48 | 0.03 |
| TAPENTADOL | 9780 | 35 | 1,516 | 43.31 |
| FENTANYL BASE | 9801 | 298 | 1,072.41 | 3.6 |

STATE:INDIANA BUSINESS ACTIVITY:C - PRACTITIONERS

| DRUG NAME | DRUG CODE | BUYERS | TOTAL GRAMS | AVG GRAMS |
|---|---|---|---|---|
| BUTALBITAL | 2165 | 1 | 5 | 5 |
| PENTOBARBITAL (SCHEDULE 2) | 2270 | 173 | 129,240.95 | 747.06 |
| COCAINE | 9041L | 5 | 1.79 | 0.36 |
| CODEINE | 9050 | 61 | 541.05 | 8.87 |
| BUPRENORPHINE | 9064 | 432 | 28.64 | 0.07 |
| DIHYDROCODEINE | 9120 | 3 | 52.23 | 17.41 |
| OXYCODONE | 9143 | 16 | 267.27 | 16.7 |
| HYDROMORPHONE | 9150 | 217 | 117.35 | 0.54 |
| HYDROCODONE | 9193 | 266 | 608.15 | 2.29 |
| MEPERIDINE (PETHIDINE) | 9230 | 79 | 232.8 | 2.95 |
| METHADONE | 9250B | 3 | 1.97 | 0.66 |
| MORPHINE | 9300 | 170 | 452.17 | 2.66 |
| OXYMORPHONE | 9652 | 3 | 20.5 | 6.83 |
| NOROXYMORPHONE | 9668 | 2 | 1.56 | 0.78 |
| ALFENTANIL | 9737 | 3 | 0.23 | 0.08 |
| REMIFENTANIL | 9739 | 2 | 0.07 | 0.04 |
| FENTANYL BASE | 9801 | 278 | 44.4 | 0.16 |

STATE:INDIANA BUSINESS ACTIVITY:M - MID-LEVEL PRACTITIONERS

| DRUG NAME | DRUG CODE | BUYERS | TOTAL GRAMS | AVG GRAMS |
|---|---|---|---|---|
| PENTOBARBITAL (SCHEDULE 2) | 2270 | 22 | 17,440.91 | 792.77 |
| CODEINE | 9050 | 7 | 31.11 | 4.44 |
| HYDROCODONE | 9193 | 1 | 18.77 | 18.77 |

STATE:INDIANA BUSINESS ACTIVITY:N-U NARCOTIC TREATMENT PROGRAMS

| DRUG NAME | DRUG CODE | BUYERS | TOTAL GRAMS | AVG GRAMS |
|---|---|---|---|---|
| BUPRENORPHINE | 9064 | 9 | 2,729.19 | 303.24 |
| METHADONE | 9250B | 14 | 254,698.29 | 18,192.73 |

STATE:IOWA BUSINESS ACTIVITY:A - PHARMACIES

| DRUG NAME | DRUG CODE | BUYERS | TOTAL GRAMS | AVG GRAMS |
|---|---|---|---|---|
| AMPHETAMINE | 1100 | 695 | 182,424.02 | 262.48 |
| DL-METHAMPHETAMINE RACEMIC BASE | 1105B | 4 | 33.35 | 8.34 |
| D-METHAMPHETAMINE | 1105D | 15 | 56.64 | 3.78 |
| LISDEXAMFETAMINE | 1205 | 678 | 135,554.62 | 199.93 |
| METHYLPHENIDATE (DL;D;L;ISOMERS) | 1724 | 703 | 326,496.10 | 464.43 |
| BUTALBITAL | 2165 | 302 | 8,565 | 28.36 |
| NABILONE | 7379 | 1 | 0.05 | 0.05 |
| COCAINE | 9041L | 2 | 0.29 | 0.14 |
| CODEINE | 9050 | 693 | 82,491.86 | 119.04 |
| BUPRENORPHINE | 9064 | 542 | 5,186.81 | 9.57 |
| OXYCODONE | 9143 | 705 | 225,339.08 | 319.63 |
| HYDROMORPHONE | 9150 | 654 | 11,551.40 | 17.66 |
| HYDROCODONE | 9193 | 714 | 242,632.60 | 339.82 |
| LEVORPHANOL | 9220L | 14 | 18.92 | 1.35 |

| | | | | |
|---|---|---|---|---|
| MEPERIDINE (PETHIDINE) | 9230 | 134 | 2,511.34 | 18.74 |
| METHADONE | 9250B | 563 | 19,304.46 | 34.29 |
| MORPHINE | 9300 | 686 | 136,634.82 | 199.18 |
| OPIUM POWDERED | 9639 | 66 | 93.6 | 1.42 |
| OXYMORPHONE | 9652 | 333 | 6,562.87 | 19.71 |
| ALFENTANIL | 9737 | 1 | 0.03 | 0.03 |
| TAPENTADOL | 9780 | 473 | 51,845 | 109.61 |
| FENTANYL BASE | 9801 | 679 | 3,601.54 | 5.3 |

STATE:IOWA BUSINESS ACTIVITY:B - HOSPITALS

| DRUG NAME | DRUG CODE | BUYERS | TOTAL GRAMS | AVG GRAMS |
|---|---|---|---|---|
| AMPHETAMINE | 1100 | 52 | 5,058.47 | 97.28 |
| D-METHAMPHETAMINE | 1105D | 1 | 2.41 | 2.41 |
| LISDEXAMFETAMINE | 1205 | 38 | 2,168.45 | 57.06 |
| METHYLPHENIDATE (DL;D;L;ISOMERS) | 1724 | 74 | 7,755.80 | 104.81 |
| AMOBARBITAL (SCHEDULE 2) | 2125 | 1 | 0.46 | 0.46 |
| BUTALBITAL | 2165 | 9 | 55 | 6.11 |
| PENTOBARBITAL (SCHEDULE 2) | 2270 | 9 | 2,362.35 | 262.48 |
| COCAINE | 9041L | 69 | 373.76 | 5.42 |
| CODEINE | 9050 | 117 | 3,119.64 | 26.66 |
| BUPRENORPHINE | 9064 | 55 | 368.26 | 6.7 |
| OXYCODONE | 9143 | 139 | 15,183.15 | 109.23 |
| HYDROMORPHONE | 9150 | 142 | 2,474.34 | 17.42 |
| HYDROCODONE | 9193 | 146 | 17,527.34 | 120.05 |
| LEVORPHANOL | 9220L | 1 | 0.12 | 0.12 |
| MEPERIDINE (PETHIDINE) | 9230 | 123 | 4,064.70 | 33.05 |
| METHADONE | 9250B | 83 | 1,641.34 | 19.78 |
| MORPHINE | 9300 | 150 | 16,498.76 | 109.99 |
| OPIUM POWDERED | 9639 | 62 | 316.8 | 5.11 |
| OXYMORPHONE | 9652 | 12 | 46 | 3.83 |
| ALFENTANIL | 9737 | 23 | 6.68 | 0.29 |
| REMIFENTANIL | 9739 | 25 | 11.54 | 0.46 |

| | | | | |
|---|---|---|---|---|
| SUFENTANIL BASE | 9740 | 26 | 0.15 | 0.01 |
| TAPENTADOL | 9780 | 29 | 831 | 28.66 |
| FENTANYL BASE | 9801 | 148 | 336.68 | 2.27 |

STATE:IOWA BUSINESS ACTIVITY:C - PRACTITIONERS

| DRUG NAME | DRUG CODE | BUYERS | TOTAL GRAMS | AVG GRAMS |
|---|---|---|---|---|
| LISDEXAMFETAMINE | 1205 | 1 | 1.73 | 1.73 |
| PENTOBARBITAL (SCHEDULE 2) | 2270 | 73 | 68,705.06 | 941.17 |
| COCAINE | 9041L | 4 | 29.57 | 7.39 |
| CODEINE | 9050 | 17 | 232.49 | 13.68 |
| BUPRENORPHINE | 9064 | 241 | 9.54 | 0.04 |
| OXYCODONE | 9143 | 11 | 862.45 | 78.4 |
| HYDROMORPHONE | 9150 | 86 | 32.51 | 0.38 |
| HYDROCODONE | 9193 | 102 | 426.51 | 4.18 |
| MEPERIDINE (PETHIDINE) | 9230 | 21 | 1,253.76 | 59.7 |
| MORPHINE | 9300 | 74 | 111.87 | 1.51 |
| OXYMORPHONE | 9652 | 1 | 0.01 | 0.01 |
| ALFENTANIL | 9737 | 1 | 0.39 | 0.39 |
| REMIFENTANIL | 9739 | 1 | 0.01 | 0.01 |
| FENTANYL BASE | 9801 | 93 | 29.45 | 0.32 |

STATE:IOWA BUSINESS ACTIVITY:D - TEACHING INSTITUTIONS

| DRUG NAME | DRUG CODE | BUYERS | TOTAL GRAMS | AVG GRAMS |
|---|---|---|---|---|
| BUPRENORPHINE | 9064 | 2 | 0.01 | 0 |
| HYDROMORPHONE | 9150 | 1 | 0.25 | 0.25 |

STATE:IOWA BUSINESS ACTIVITY:M - MID-LEVEL PRACTITIONERS

| DRUG NAME | DRUG CODE | BUYERS | TOTAL GRAMS | AVG GRAMS |
|---|---|---|---|---|

| | | | | |
|---|---|---|---|---|
| PENTOBARBITAL (SCHEDULE 2) | 2270 | 5 | 2,310.40 | 462.08 |

STATE:IOWA BUSINESS ACTIVITY:N-U NARCOTIC TREATMENT PROGRAMS

| DRUG NAME | DRUG CODE | BUYERS | TOTAL GRAMS | AVG GRAMS |
|---|---|---|---|---|
| BUPRENORPHINE | 9064 | 5 | 272.9 | 54.58 |
| METHADONE | 9250B | 7 | 32,506.07 | 4,643.72 |

STATE:KANSAS BUSINESS ACTIVITY:A - PHARMACIES

| DRUG NAME | DRUG CODE | BUYERS | TOTAL GRAMS | AVG GRAMS |
|---|---|---|---|---|
| AMPHETAMINE | 1100 | 655 | 217,573.97 | 332.17 |
| DL-METHAMPHETAMINE RACEMIC BASE | 1105B | 2 | 1.61 | 0.8 |
| D-METHAMPHETAMINE | 1105D | 19 | 51.42 | 2.71 |
| LISDEXAMFETAMINE | 1205 | 633 | 120,767.40 | 190.79 |
| METHYLPHENIDATE (DL;D;L;ISOMERS) | 1724 | 661 | 196,156.94 | 296.76 |
| BARBITURIC ACID DERIVIATIVE OR SALT [PER 21C | 2100 | 9 | 95 | 10.56 |
| BUTALBITAL | 2165 | 342 | 15,290 | 44.71 |
| SECOBARBITAL (SCHEDULE 2) | 2315 | 5 | 73.24 | 14.65 |
| COCAINE | 9041L | 4 | 74.35 | 18.59 |
| CODEINE | 9050 | 634 | 113,025.16 | 178.27 |
| BUPRENORPHINE | 9064 | 564 | 8,900.16 | 15.78 |
| DIHYDROCODEINE | 9120 | 3 | 7.48 | 2.49 |
| OXYCODONE | 9143 | 658 | 487,616.97 | 741.06 |
| HYDROMORPHONE | 9150 | 602 | 13,403.66 | 22.27 |
| HYDROCODONE | 9193 | 665 | 331,381.47 | 498.32 |
| LEVORPHANOL | 9220L | 14 | 32.73 | 2.34 |
| MEPERIDINE (PETHIDINE) | 9230 | 285 | 5,706.23 | 20.02 |
| METHADONE | 9250B | 555 | 33,282.32 | 59.97 |
| MORPHINE | 9300 | 647 | 179,853.20 | 277.98 |
| OPIUM TINCTURE (LAUDANUM) | 9630 | 2 | 5.91 | 2.96 |
| OPIUM POWDERED | 9639 | 89 | 327.6 | 3.68 |

| | | | | |
|---|---|---|---|---|
| OXYMORPHONE | 9652 | 402 | 11,870.28 | 29.53 |
| SUFENTANIL BASE | 9740 | 4 | 0.31 | 0.08 |
| TAPENTADOL | 9780 | 392 | 44,797 | 114.28 |
| FENTANYL BASE | 9801 | 640 | 4,517.28 | 7.06 |

STATE:KANSAS BUSINESS ACTIVITY:B - HOSPITALS

| DRUG NAME | DRUG CODE | BUYERS | TOTAL GRAMS | AVG GRAMS |
|---|---|---|---|---|
| AMPHETAMINE | 1100 | 49 | 9,643.37 | 196.8 |
| DL-METHAMPHETAMINE RACEMIC BASE | 1105B | 1 | 4.42 | 4.42 |
| LISDEXAMFETAMINE | 1205 | 27 | 1,760.89 | 65.22 |
| METHYLPHENIDATE (DL;D;L;ISOMERS) | 1724 | 81 | 9,234.71 | 114.01 |
| AMOBARBITAL (SCHEDULE 2) | 2125 | 2 | 17.77 | 8.89 |
| BUTALBITAL | 2165 | 4 | 165 | 41.25 |
| PENTOBARBITAL (SCHEDULE 2) | 2270 | 4 | 11,351.03 | 2,837.76 |
| COCAINE | 9041L | 50 | 241.56 | 4.83 |
| CODEINE | 9050 | 154 | 6,196.67 | 40.24 |
| BUPRENORPHINE | 9064 | 42 | 347.6 | 8.28 |
| OXYCODONE | 9143 | 185 | 36,713.70 | 198.45 |
| HYDROMORPHONE | 9150 | 163 | 2,300.43 | 14.11 |
| HYDROCODONE | 9193 | 192 | 23,184.71 | 120.75 |
| MEPERIDINE (PETHIDINE) | 9230 | 146 | 2,151.08 | 14.73 |
| METHADONE | 9250B | 87 | 2,236.80 | 25.71 |
| MORPHINE | 9300 | 196 | 21,428.75 | 109.33 |
| OPIUM POWDERED | 9639 | 67 | 421.56 | 6.29 |
| OXYMORPHONE | 9652 | 15 | 359.2 | 23.95 |
| ALFENTANIL | 9737 | 17 | 6.64 | 0.39 |
| REMIFENTANIL | 9739 | 28 | 7.49 | 0.27 |
| SUFENTANIL BASE | 9740 | 48 | 0.72 | 0.01 |
| TAPENTADOL | 9780 | 25 | 672.5 | 26.9 |
| FENTANYL BASE | 9801 | 193 | 542.24 | 2.81 |

STATISTICAL SUMMARY FOR RETAIL DRUG PURCHASES BY GRAMS WT

STATE:KANSAS BUSINESS ACTIVITY:C - PRACTITIONERS

| DRUG NAME | DRUG CODE | BUYERS | TOTAL GRAMS | AVG GRAMS |
|---|---|---|---|---|
| AMPHETAMINE | 1100 | 1 | 1.76 | 1.76 |
| PENTOBARBITAL (SCHEDULE 2) | 2270 | 107 | 72,791.88 | 680.3 |
| COCAINE | 9041L | 3 | 50.71 | 16.9 |
| CODEINE | 9050 | 15 | 570.52 | 38.03 |
| BUPRENORPHINE | 9064 | 253 | 20.2 | 0.08 |
| OXYCODONE | 9143 | 10 | 1,425.42 | 142.54 |
| HYDROMORPHONE | 9150 | 115 | 62.08 | 0.54 |
| HYDROCODONE | 9193 | 102 | 277.43 | 2.72 |
| MEPERIDINE (PETHIDINE) | 9230 | 24 | 131.38 | 5.47 |
| MORPHINE | 9300 | 57 | 168.75 | 2.96 |
| OPIUM POWDERED | 9639 | 1 | 0.72 | 0.72 |
| NOROXYMORPHONE | 9668 | 1 | 0.7 | 0.7 |
| ALFENTANIL | 9737 | 1 | 0.23 | 0.23 |
| REMIFENTANIL | 9739 | 1 | 0.48 | 0.48 |
| SUFENTANIL BASE | 9740 | 1 | 0 | 0 |
| FENTANYL BASE | 9801 | 100 | 11.01 | 0.11 |

STATE:KANSAS BUSINESS ACTIVITY:D - TEACHING INSTITUTIONS

| DRUG NAME | DRUG CODE | BUYERS | TOTAL GRAMS | AVG GRAMS |
|---|---|---|---|---|
| BUPRENORPHINE | 9064 | 1 | 0 | 0 |

STATE:KANSAS BUSINESS ACTIVITY:M - MID-LEVEL PRACTITIONERS

| DRUG NAME | DRUG CODE | BUYERS | TOTAL GRAMS | AVG GRAMS |
|---|---|---|---|---|
| HYDROMORPHONE | 9150 | 17 | 0.63 | 0.04 |
| MORPHINE | 9300 | 31 | 6.13 | 0.2 |
| FENTANYL BASE | 9801 | 44 | 0.34 | 0.01 |

STATISTICAL SUMMARY FOR RETAIL DRUG PURCHASES BY GRAMS WT

STATE:KANSAS BUSINESS ACTIVITY:N-U NARCOTIC TREATMENT PROGRAMS

| DRUG NAME | DRUG CODE | BUYERS | TOTAL GRAMS | AVG GRAMS |
|---|---|---|---|---|
| BUPRENORPHINE | 9064 | 6 | 159.54 | 26.59 |
| METHADONE | 9250B | 10 | 60,302.24 | 6,030.22 |

STATE:KENTUCKY BUSINESS ACTIVITY:A - PHARMACIES

| DRUG NAME | DRUG CODE | BUYERS | TOTAL GRAMS | AVG GRAMS |
|---|---|---|---|---|
| AMPHETAMINE | 1100 | 1,123 | 248,475.23 | 221.26 |
| DL-METHAMPHETAMINE RACEMIC BASE | 1105B | 3 | 10.45 | 3.48 |
| D-METHAMPHETAMINE | 1105D | 12 | 20.89 | 1.74 |
| LISDEXAMFETAMINE | 1205 | 1,062 | 167,490.60 | 157.71 |
| METHYLPHENIDATE (DL;D;L;ISOMERS) | 1724 | 1,111 | 296,773.12 | 267.12 |
| BARBITURIC ACID DERIVIATIVE OR SALT [PER 21C | 2100 | 12 | 85 | 7.08 |
| BUTALBITAL | 2165 | 533 | 18,645 | 34.98 |
| COCAINE | 9041L | 3 | 9.75 | 3.25 |
| CODEINE | 9050 | 1,123 | 205,074.68 | 182.61 |
| BUPRENORPHINE | 9064 | 1,047 | 122,356.60 | 116.86 |
| DIHYDROCODEINE | 9120 | 23 | 120.7 | 5.25 |
| OXYCODONE | 9143 | 1,141 | 834,695.85 | 731.55 |
| HYDROMORPHONE | 9150 | 868 | 9,549.67 | 11 |
| HYDROCODONE | 9193 | 1,150 | 840,005.74 | 730.44 |
| MEPERIDINE (PETHIDINE) | 9230 | 327 | 5,667.69 | 17.33 |
| METHADONE | 9250B | 919 | 58,378.29 | 63.52 |
| MORPHINE | 9300 | 1,103 | 188,578.07 | 170.97 |
| OPIUM POWDERED | 9639 | 123 | 309.24 | 2.51 |
| OXYMORPHONE | 9652 | 724 | 20,283.86 | 28.02 |
| REMIFENTANIL | 9739 | 1 | 0.12 | 0.12 |
| SUFENTANIL BASE | 9740 | 2 | 0 | 0 |
| TAPENTADOL | 9780 | 654 | 64,788 | 99.06 |
| FENTANYL BASE | 9801 | 1,083 | 4,769.78 | 4.4 |

STATISTICAL SUMMARY FOR RETAIL DRUG PURCHASES BY GRAMS WT

STATE:KENTUCKY BUSINESS ACTIVITY:B - HOSPITALS

| DRUG NAME | DRUG CODE | BUYERS | TOTAL GRAMS | AVG GRAMS |
|---|---|---|---|---|
| AMPHETAMINE | 1100 | 53 | 8,043.41 | 151.76 |
| LISDEXAMFETAMINE | 1205 | 27 | 2,072.96 | 76.78 |
| METHYLPHENIDATE (DL;D;L;ISOMERS) | 1724 | 75 | 9,474.83 | 126.33 |
| AMOBARBITAL (SCHEDULE 2) | 2125 | 4 | 15.04 | 3.76 |
| BUTALBITAL | 2165 | 9 | 100 | 11.11 |
| PENTOBARBITAL (SCHEDULE 2) | 2270 | 3 | 280.26 | 93.42 |
| COCAINE | 9041L | 69 | 425.08 | 6.16 |
| CODEINE | 9050 | 137 | 6,011.70 | 43.88 |
| BUPRENORPHINE | 9064 | 86 | 1,220.59 | 14.19 |
| OXYCODONE | 9143 | 147 | 37,530.47 | 255.31 |
| HYDROMORPHONE | 9150 | 138 | 2,797.75 | 20.27 |
| HYDROCODONE | 9193 | 148 | 34,167.14 | 230.86 |
| MEPERIDINE (PETHIDINE) | 9230 | 121 | 5,394.37 | 44.58 |
| METHADONE | 9250B | 101 | 3,667.34 | 36.31 |
| MORPHINE | 9300 | 146 | 23,963.73 | 164.14 |
| OPIUM POWDERED | 9639 | 72 | 378.36 | 5.26 |
| OXYMORPHONE | 9652 | 20 | 135.47 | 6.77 |
| ALFENTANIL | 9737 | 8 | 1.37 | 0.17 |
| REMIFENTANIL | 9739 | 30 | 18.18 | 0.61 |
| SUFENTANIL BASE | 9740 | 36 | 1.05 | 0.03 |
| TAPENTADOL | 9780 | 21 | 1,267.30 | 60.35 |
| FENTANYL BASE | 9801 | 143 | 660.91 | 4.62 |

STATE:KENTUCKY BUSINESS ACTIVITY:C - PRACTITIONERS

| DRUG NAME | DRUG CODE | BUYERS | TOTAL GRAMS | AVG GRAMS |
|---|---|---|---|---|
| PENTOBARBITAL (SCHEDULE 2) | 2270 | 97 | 84,470.78 | 870.83 |
| COCAINE | 9041L | 3 | 3.29 | 1.1 |

| | | | | |
|---|---|---|---|---|
| CODEINE | 9050 | 14 | 565.34 | 40.38 |
| BUPRENORPHINE | 9064 | 264 | 10.84 | 0.04 |
| DIHYDROCODEINE | 9120 | 1 | 0.21 | 0.21 |
| OXYCODONE | 9143 | 12 | 414.62 | 34.55 |
| HYDROMORPHONE | 9150 | 99 | 259.46 | 2.62 |
| HYDROCODONE | 9193 | 129 | 510.17 | 3.95 |
| MEPERIDINE (PETHIDINE) | 9230 | 60 | 637.7 | 10.63 |
| METHADONE | 9250B | 1 | 2.15 | 2.15 |
| MORPHINE | 9300 | 151 | 424.35 | 2.81 |
| OPIUM POWDERED | 9639 | 1 | 17.28 | 17.28 |
| ALFENTANIL | 9737 | 2 | 7.68 | 3.84 |
| REMIFENTANIL | 9739 | 2 | 0.12 | 0.06 |
| SUFENTANIL BASE | 9740 | 3 | 0.14 | 0.05 |
| TAPENTADOL | 9780 | 1 | 10 | 10 |
| FENTANYL BASE | 9801 | 257 | 33.56 | 0.13 |

STATE:KENTUCKY BUSINESS ACTIVITY:D - TEACHING INSTITUTIONS

| DRUG NAME | DRUG CODE | BUYERS | TOTAL GRAMS | AVG GRAMS |
|---|---|---|---|---|
| BUPRENORPHINE | 9064 | 1 | 0.01 | 0.01 |

STATE:KENTUCKY BUSINESS ACTIVITY:M - MID-LEVEL PRACTITIONERS

| DRUG NAME | DRUG CODE | BUYERS | TOTAL GRAMS | AVG GRAMS |
|---|---|---|---|---|
| PENTOBARBITAL (SCHEDULE 2) | 2270 | 23 | 24,898.98 | 1,082.56 |
| CODEINE | 9050 | 1 | 6.63 | 6.63 |
| HYDROMORPHONE | 9150 | 1 | 0.21 | 0.21 |
| MEPERIDINE (PETHIDINE) | 9230 | 1 | 8.72 | 8.72 |
| REMIFENTANIL | 9739 | 1 | 0.04 | 0.04 |
| FENTANYL BASE | 9801 | 1 | 0.05 | 0.05 |

STATISTICAL SUMMARY FOR RETAIL DRUG PURCHASES BY GRAMS WT

STATE:KENTUCKY BUSINESS ACTIVITY:N-U NARCOTIC TREATMENT PROGRAMS

| DRUG NAME | DRUG CODE | BUYERS | TOTAL GRAMS | AVG GRAMS |
|---|---|---|---|---|
| BUPRENORPHINE | 9064 | 10 | 806.52 | 80.65 |
| METHADONE | 9250B | 20 | 145,166.04 | 7,258.30 |

STATE:LOUISIANA BUSINESS ACTIVITY:A - PHARMACIES

| DRUG NAME | DRUG CODE | BUYERS | TOTAL GRAMS | AVG GRAMS |
|---|---|---|---|---|
| AMPHETAMINE | 1100 | 1,210 | 568,366.11 | 469.72 |
| DL-METHAMPHETAMINE RACEMIC BASE | 1105B | 8 | 11.25 | 1.41 |
| D-METHAMPHETAMINE | 1105D | 18 | 47.4 | 2.63 |
| LISDEXAMFETAMINE | 1205 | 1,190 | 384,163.06 | 322.83 |
| METHYLPHENIDATE (DL;D;L;ISOMERS) | 1724 | 1,205 | 340,359.76 | 282.46 |
| BARBITURIC ACID DERIVIATIVE OR SALT [PER 21C | 2100 | 8 | 55 | 6.88 |
| BUTALBITAL | 2165 | 797 | 32,030 | 40.19 |
| SECOBARBITAL (SCHEDULE 2) | 2315 | 2 | 54.93 | 27.47 |
| COCAINE | 9041L | 5 | 74.21 | 14.84 |
| CODEINE | 9050 | 1,210 | 177,833.50 | 146.97 |
| BUPRENORPHINE | 9064 | 1,090 | 62,738.14 | 57.56 |
| DIHYDROCODEINE | 9120 | 250 | 2,228.18 | 8.91 |
| OXYCODONE | 9143 | 1,230 | 814,167.02 | 661.92 |
| HYDROMORPHONE | 9150 | 1,007 | 12,228.42 | 12.14 |
| HYDROCODONE | 9193 | 1,242 | 688,482.04 | 554.33 |
| LEVORPHANOL | 9220L | 18 | 41.21 | 2.29 |
| MEPERIDINE (PETHIDINE) | 9230 | 937 | 26,875.43 | 28.68 |
| METHADONE | 9250B | 748 | 40,314.36 | 53.9 |
| MORPHINE | 9300 | 1,117 | 228,511.89 | 204.58 |
| OPIUM POWDERED | 9639 | 58 | 158.76 | 2.74 |
| OXYMORPHONE | 9652 | 684 | 36,588.41 | 53.49 |
| ALFENTANIL | 9737 | 1 | 0.83 | 0.83 |
| REMIFENTANIL | 9739 | 4 | 0.29 | 0.07 |
| SUFENTANIL BASE | 9740 | 6 | 1.96 | 0.33 |

| | | | | |
|---|---|---|---|---|
| TAPENTADOL | 9780 | 684 | 64,900 | 94.88 |
| FENTANYL BASE | 9801 | 1,127 | 5,094.02 | 4.52 |

STATE:LOUISIANA BUSINESS ACTIVITY:B - HOSPITALS

| DRUG NAME | DRUG CODE | BUYERS | TOTAL GRAMS | AVG GRAMS |
|---|---|---|---|---|
| AMPHETAMINE | 1100 | 49 | 5,621.24 | 114.72 |
| D-METHAMPHETAMINE | 1105D | 1 | 20.09 | 20.09 |
| LISDEXAMFETAMINE | 1205 | 28 | 1,876.51 | 67.02 |
| METHYLPHENIDATE (DL;D;L;ISOMERS) | 1724 | 105 | 5,509.17 | 52.47 |
| BARBITURIC ACID DERIVIATIVE OR SALT [PER 21C | 2100 | 1 | 5 | 5 |
| BUTALBITAL | 2165 | 12 | 150 | 12.5 |
| PENTOBARBITAL (SCHEDULE 2) | 2270 | 13 | 3,435.07 | 264.24 |
| COCAINE | 9041L | 56 | 274.27 | 4.9 |
| CODEINE | 9050 | 139 | 3,602.88 | 25.92 |
| BUPRENORPHINE | 9064 | 80 | 624.31 | 7.8 |
| OXYCODONE | 9143 | 188 | 33,702.50 | 179.27 |
| HYDROMORPHONE | 9150 | 187 | 2,985.04 | 15.96 |
| HYDROCODONE | 9193 | 201 | 41,456.62 | 206.25 |
| MEPERIDINE (PETHIDINE) | 9230 | 175 | 9,258.90 | 52.91 |
| METHADONE | 9250B | 113 | 2,926.31 | 25.9 |
| MORPHINE | 9300 | 196 | 23,022.24 | 117.46 |
| OPIUM POWDERED | 9639 | 64 | 249.48 | 3.9 |
| OXYMORPHONE | 9652 | 22 | 121.41 | 5.52 |
| ALFENTANIL | 9737 | 15 | 5.48 | 0.37 |
| REMIFENTANIL | 9739 | 34 | 16.61 | 0.49 |
| SUFENTANIL BASE | 9740 | 39 | 1.45 | 0.04 |
| TAPENTADOL | 9780 | 17 | 924.5 | 54.38 |
| FENTANYL BASE | 9801 | 192 | 498.55 | 2.6 |

STATE:LOUISIANA BUSINESS ACTIVITY:C - PRACTITIONERS

| DRUG NAME | DRUG CODE | BUYERS | TOTAL GRAMS | AVG GRAMS |
|---|---|---|---|---|
| AMPHETAMINE | 1100 | 1 | 3.38 | 3.38 |
| GAMMA HYDROXYBUTYRIC ACID(FDA APPROVED | 2012 | 1 | 743.04 | 743.04 |
| BUTALBITAL | 2165 | 1 | 5 | 5 |
| PENTOBARBITAL (SCHEDULE 2) | 2270 | 75 | 51,678.36 | 689.04 |
| COCAINE | 9041L | 14 | 37.43 | 2.67 |
| CODEINE | 9050 | 62 | 653.26 | 10.54 |
| BUPRENORPHINE | 9064 | 285 | 24.62 | 0.09 |
| DIHYDROCODEINE | 9120 | 132 | 90.13 | 0.68 |
| OXYCODONE | 9143 | 21 | 3,377.02 | 160.81 |
| HYDROMORPHONE | 9150 | 116 | 43.09 | 0.37 |
| HYDROCODONE | 9193 | 206 | 467.24 | 2.27 |
| MEPERIDINE (PETHIDINE) | 9230 | 132 | 1,190.47 | 9.02 |
| METHADONE | 9250B | 3 | 10.73 | 3.58 |
| MORPHINE | 9300 | 143 | 204.04 | 1.43 |
| OPIUM POWDERED | 9639 | 4 | 20.88 | 5.22 |
| NOROXYMORPHONE | 9668 | 1 | 0.62 | 0.62 |
| ALFENTANIL | 9737 | 5 | 2.97 | 0.59 |
| REMIFENTANIL | 9739 | 4 | 0.09 | 0.02 |
| SUFENTANIL BASE | 9740 | 1 | 0 | 0 |
| TAPENTADOL | 9780 | 1 | 5 | 5 |
| FENTANYL BASE | 9801 | 235 | 31.11 | 0.13 |

STATE:LOUISIANA BUSINESS ACTIVITY:D - TEACHING INSTITUTIONS

| DRUG NAME | DRUG CODE | BUYERS | TOTAL GRAMS | AVG GRAMS |
|---|---|---|---|---|
| FENTANYL BASE | 9801 | 1 | 0.02 | 0.02 |

STATE:LOUISIANA BUSINESS ACTIVITY:M - MID-LEVEL PRACTITIONERS

| DRUG NAME | DRUG CODE | BUYERS | TOTAL GRAMS | AVG GRAMS |
|---|---|---|---|---|
| PENTOBARBITAL (SCHEDULE 2) | 2270 | 18 | 35,278.02 | 1,959.89 |

| | | | | |
|---|---|---|---|---|
| CODEINE | 9050 | 3 | 3.32 | 1.11 |
| DIHYDROCODEINE | 9120 | 24 | 12.18 | 0.51 |

STATE:LOUISIANA BUSINESS ACTIVITY:N-U NARCOTIC TREATMENT PROGRAMS

| DRUG NAME | DRUG CODE | BUYERS | TOTAL GRAMS | AVG GRAMS |
|---|---|---|---|---|
| BUPRENORPHINE | 9064 | 5 | 336.13 | 67.23 |
| METHADONE | 9250B | 10 | 109,987.95 | 10,998.79 |

STATE:MAINE BUSINESS ACTIVITY:A - PHARMACIES

| DRUG NAME | DRUG CODE | BUYERS | TOTAL GRAMS | AVG GRAMS |
|---|---|---|---|---|
| AMPHETAMINE | 1100 | 300 | 84,076.98 | 280.26 |
| DL-METHAMPHETAMINE RACEMIC BASE | 1105B | 2 | 2.81 | 1.41 |
| D-METHAMPHETAMINE | 1105D | 5 | 8.44 | 1.69 |
| LISDEXAMFETAMINE | 1205 | 298 | 65,921.32 | 221.21 |
| METHYLPHENIDATE (DL;D;L;ISOMERS) | 1724 | 303 | 142,140.49 | 469.11 |
| BARBITURIC ACID DERIVIATIVE OR SALT [PER 21C | 2100 | 4 | 25 | 6.25 |
| BUTALBITAL | 2165 | 160 | 5,385 | 33.66 |
| NABILONE | 7379 | 1 | 0.1 | 0.1 |
| COCAINE | 9041L | 1 | 2.86 | 2.86 |
| CODEINE | 9050 | 303 | 47,884.66 | 158.04 |
| BUPRENORPHINE | 9064 | 305 | 31,082.33 | 101.91 |
| DIHYDROCODEINE | 9120 | 1 | 1.07 | 1.07 |
| OXYCODONE | 9143 | 304 | 267,581.76 | 880.2 |
| HYDROMORPHONE | 9150 | 301 | 6,545.46 | 21.75 |
| HYDROCODONE | 9193 | 305 | 78,167.31 | 256.29 |
| LEVORPHANOL | 9220L | 1 | 0.23 | 0.23 |
| MEPERIDINE (PETHIDINE) | 9230 | 79 | 1,794.85 | 22.72 |
| METHADONE | 9250B | 293 | 38,977.95 | 133.03 |
| MORPHINE | 9300 | 304 | 81,640.54 | 268.55 |
| OPIUM TINCTURE (LAUDANUM) | 9630 | 2 | 5.91 | 2.96 |

| | | | | |
|---|---|---|---|---|
| OPIUM POWDERED | 9639 | 22 | 32.76 | 1.49 |
| OXYMORPHONE | 9652 | 201 | 6,865.82 | 34.16 |
| TAPENTADOL | 9780 | 117 | 8,165.50 | 69.79 |
| FENTANYL BASE | 9801 | 300 | 1,845.62 | 6.15 |

STATE:MAINE BUSINESS ACTIVITY:B - HOSPITALS

| DRUG NAME | DRUG CODE | BUYERS | TOTAL GRAMS | AVG GRAMS |
|---|---|---|---|---|
| AMPHETAMINE | 1100 | 25 | 1,354.61 | 54.18 |
| LISDEXAMFETAMINE | 1205 | 15 | 428.95 | 28.6 |
| METHYLPHENIDATE (DL;D;L;ISOMERS) | 1724 | 39 | 2,886.94 | 74.02 |
| AMOBARBITAL (SCHEDULE 2) | 2125 | 1 | 3.65 | 3.65 |
| PENTOBARBITAL (SCHEDULE 2) | 2270 | 2 | 287.09 | 143.55 |
| COCAINE | 9041L | 24 | 105.64 | 4.4 |
| CODEINE | 9050 | 48 | 1,702.97 | 35.48 |
| BUPRENORPHINE | 9064 | 39 | 737.56 | 18.91 |
| OXYCODONE | 9143 | 47 | 13,841.25 | 294.49 |
| HYDROMORPHONE | 9150 | 41 | 1,130.23 | 27.57 |
| HYDROCODONE | 9193 | 46 | 5,165.44 | 112.29 |
| MEPERIDINE (PETHIDINE) | 9230 | 36 | 209.6 | 5.82 |
| METHADONE | 9250B | 38 | 1,319.15 | 34.71 |
| MORPHINE | 9300 | 44 | 7,278.95 | 165.43 |
| OPIUM POWDERED | 9639 | 23 | 132.12 | 5.74 |
| OXYMORPHONE | 9652 | 3 | 6.87 | 2.29 |
| ALFENTANIL | 9737 | 7 | 2.02 | 0.29 |
| REMIFENTANIL | 9739 | 14 | 5.1 | 0.36 |
| SUFENTANIL BASE | 9740 | 8 | 0.03 | 0 |
| TAPENTADOL | 9780 | 1 | 45 | 45 |
| FENTANYL BASE | 9801 | 44 | 169.65 | 3.86 |

STATE:MAINE BUSINESS ACTIVITY:C - PRACTITIONERS

| DRUG NAME | DRUG CODE | BUYERS | TOTAL GRAMS | AVG GRAMS |
|---|---|---|---|---|
| PENTOBARBITAL (SCHEDULE 2) | 2270 | 88 | 64,154.88 | 729.03 |
| CODEINE | 9050 | 6 | 165.42 | 27.57 |
| BUPRENORPHINE | 9064 | 156 | 9.56 | 0.06 |
| OXYCODONE | 9143 | 5 | 18.15 | 3.63 |
| HYDROMORPHONE | 9150 | 70 | 28.26 | 0.4 |
| HYDROCODONE | 9193 | 60 | 100.18 | 1.67 |
| MEPERIDINE (PETHIDINE) | 9230 | 6 | 4.36 | 0.73 |
| MORPHINE | 9300 | 50 | 59.26 | 1.19 |
| OXYMORPHONE | 9652 | 2 | 0.12 | 0.06 |
| ALFENTANIL | 9737 | 3 | 0.61 | 0.2 |
| REMIFENTANIL | 9739 | 1 | 0.02 | 0.02 |
| FENTANYL BASE | 9801 | 53 | 52.39 | 0.99 |

STATE:MAINE BUSINESS ACTIVITY:D - TEACHING INSTITUTIONS

| DRUG NAME | DRUG CODE | BUYERS | TOTAL GRAMS | AVG GRAMS |
|---|---|---|---|---|
| BUPRENORPHINE | 9064 | 1 | 0.01 | 0.01 |

STATE:MAINE BUSINESS ACTIVITY:M - MID-LEVEL PRACTITIONERS

| DRUG NAME | DRUG CODE | BUYERS | TOTAL GRAMS | AVG GRAMS |
|---|---|---|---|---|
| PENTOBARBITAL (SCHEDULE 2) | 2270 | 3 | 1,960.42 | 653.47 |
| CODEINE | 9050 | 3 | 19.23 | 6.41 |
| HYDROCODONE | 9193 | 1 | 0.51 | 0.51 |

STATE:MAINE BUSINESS ACTIVITY:N-U NARCOTIC TREATMENT PROGRAMS

| DRUG NAME | DRUG CODE | BUYERS | TOTAL GRAMS | AVG GRAMS |
|---|---|---|---|---|
| METHADONE | 9250B | 10 | 101,119.08 | 10,111.91 |

STATISTICAL SUMMARY FOR RETAIL DRUG PURCHASES BY GRAMS WT

STATE:MARYLAND BUSINESS ACTIVITY:A - PHARMACIES

| DRUG NAME | DRUG CODE | BUYERS | TOTAL GRAMS | AVG GRAMS |
|---|---|---|---|---|
| AMPHETAMINE | 1100 | 1,262 | 338,606.68 | 268.31 |
| DL-METHAMPHETAMINE RACEMIC BASE | 1105B | 3 | 12.06 | 4.02 |
| D-METHAMPHETAMINE | 1105D | 40 | 185.18 | 4.63 |
| LISDEXAMFETAMINE | 1205 | 1,233 | 213,226.40 | 172.93 |
| METHYLPHENIDATE (DL;D;L;ISOMERS) | 1724 | 1,266 | 391,988.72 | 309.63 |
| BARBITURIC ACID DERIVIATIVE OR SALT [PER 21C | 2100 | 14 | 105 | 7.5 |
| BUTALBITAL | 2165 | 763 | 32,315 | 42.35 |
| SECOBARBITAL (SCHEDULE 2) | 2315 | 6 | 183.1 | 30.52 |
| NABILONE | 7379 | 18 | 5.35 | 0.3 |
| COCAINE | 9041L | 11 | 127.31 | 11.57 |
| CODEINE | 9050 | 1,276 | 248,917.17 | 195.08 |
| BUPRENORPHINE | 9064 | 1,213 | 84,677.14 | 69.81 |
| DIHYDROCODEINE | 9120 | 18 | 24.57 | 1.36 |
| OXYCODONE | 9143 | 1,295 | 1,379,431.12 | 1,065.20 |
| HYDROMORPHONE | 9150 | 1,210 | 33,025.38 | 27.29 |
| HYDROCODONE | 9193 | 1,284 | 154,299.96 | 120.17 |
| LEVORPHANOL | 9220L | 16 | 4.06 | 0.25 |
| MEPERIDINE (PETHIDINE) | 9230 | 393 | 9,913.13 | 25.22 |
| METHADONE | 9250B | 1,002 | 72,978.93 | 72.83 |
| MORPHINE | 9300 | 1,236 | 330,659.01 | 267.52 |
| OPIUM TINCTURE (LAUDANUM) | 9630 | 6 | 10.62 | 1.77 |
| OPIUM POWDERED | 9639 | 56 | 120.24 | 2.15 |
| OXYMORPHONE | 9652 | 926 | 41,542.91 | 44.86 |
| ALFENTANIL | 9737 | 1 | 0.69 | 0.69 |
| REMIFENTANIL | 9739 | 4 | 0.3 | 0.08 |
| TAPENTADOL | 9780 | 914 | 138,158 | 151.16 |
| FENTANYL BASE | 9801 | 1,207 | 6,287.45 | 5.21 |

STATE:MARYLAND BUSINESS ACTIVITY:B - HOSPITALS

| DRUG NAME | DRUG CODE | BUYERS | TOTAL GRAMS | AVG GRAMS |
|---|---|---|---|---|
| AMPHETAMINE | 1100 | 55 | 9,092.17 | 165.31 |
| D-METHAMPHETAMINE | 1105D | 1 | 0.4 | 0.4 |
| LISDEXAMFETAMINE | 1205 | 26 | 1,695.01 | 65.19 |
| METHYLPHENIDATE (DL;D;L;ISOMERS) | 1724 | 67 | 15,016.92 | 224.13 |
| AMOBARBITAL (SCHEDULE 2) | 2125 | 6 | 20.05 | 3.34 |
| BUTALBITAL | 2165 | 4 | 60 | 15 |
| PENTOBARBITAL (SCHEDULE 2) | 2270 | 9 | 4,773.46 | 530.38 |
| COCAINE | 9041L | 60 | 405.76 | 6.76 |
| CODEINE | 9050 | 87 | 6,101.64 | 70.13 |
| BUPRENORPHINE | 9064 | 70 | 1,634.19 | 23.35 |
| OXYCODONE | 9143 | 124 | 60,648.36 | 489.1 |
| HYDROMORPHONE | 9150 | 122 | 5,927.80 | 48.59 |
| HYDROCODONE | 9193 | 103 | 4,577.13 | 44.44 |
| MEPERIDINE (PETHIDINE) | 9230 | 98 | 1,313.50 | 13.4 |
| METHADONE | 9250B | 68 | 5,658.89 | 83.22 |
| MORPHINE | 9300 | 115 | 23,494.46 | 204.3 |
| OPIUM POWDERED | 9639 | 32 | 176.4 | 5.51 |
| OXYMORPHONE | 9652 | 13 | 239.31 | 18.41 |
| ALFENTANIL | 9737 | 6 | 1.61 | 0.27 |
| REMIFENTANIL | 9739 | 28 | 27.13 | 0.97 |
| SUFENTANIL BASE | 9740 | 11 | 0.15 | 0.01 |
| TAPENTADOL | 9780 | 14 | 2,027 | 144.79 |
| FENTANYL BASE | 9801 | 191 | 677.76 | 3.55 |

STATE:MARYLAND BUSINESS ACTIVITY:C - PRACTITIONERS

| DRUG NAME | DRUG CODE | BUYERS | TOTAL GRAMS | AVG GRAMS |
|---|---|---|---|---|
| PENTOBARBITAL (SCHEDULE 2) | 2270 | 79 | 52,325.75 | 662.35 |
| COCAINE | 9041L | 15 | 26.36 | 1.76 |
| CODEINE | 9050 | 168 | 8,367.07 | 49.8 |
| BUPRENORPHINE | 9064 | 530 | 671.71 | 1.27 |

| | | | | |
|---|---|---|---|---|
| DIHYDROCODEINE | 9120 | 1 | 0.21 | 0.21 |
| OXYCODONE | 9143 | 73 | 2,298.08 | 31.48 |
| HYDROMORPHONE | 9150 | 244 | 170.86 | 0.7 |
| HYDROCODONE | 9193 | 272 | 1,836.21 | 6.75 |
| MEPERIDINE (PETHIDINE) | 9230 | 69 | 338.36 | 4.9 |
| METHADONE | 9250B | 11 | 78.13 | 7.1 |
| MORPHINE | 9300 | 176 | 1,200.77 | 6.82 |
| OXYMORPHONE | 9652 | 24 | 677.01 | 28.21 |
| NOROXYMORPHONE | 9668 | 2 | 2.46 | 1.23 |
| REMIFENTANIL | 9739 | 7 | 0.87 | 0.12 |
| TAPENTADOL | 9780 | 7 | 558 | 79.71 |
| FENTANYL BASE | 9801 | 373 | 79.79 | 0.21 |

STATE:MARYLAND BUSINESS ACTIVITY:D - TEACHING INSTITUTIONS

| DRUG NAME | DRUG CODE | BUYERS | TOTAL GRAMS | AVG GRAMS |
|---|---|---|---|---|
| MEPERIDINE (PETHIDINE) | 9230 | 1 | 4.36 | 4.36 |
| FENTANYL BASE | 9801 | 1 | 0.01 | 0.01 |

STATE:MARYLAND BUSINESS ACTIVITY:M - MID-LEVEL PRACTITIONERS

| DRUG NAME | DRUG CODE | BUYERS | TOTAL GRAMS | AVG GRAMS |
|---|---|---|---|---|
| PENTOBARBITAL (SCHEDULE 2) | 2270 | 11 | 14,975.67 | 1,361.42 |
| CODEINE | 9050 | 16 | 7.84 | 0.49 |
| BUPRENORPHINE | 9064 | 1 | 0.6 | 0.6 |
| OXYCODONE | 9143 | 1 | 0.9 | 0.9 |
| HYDROCODONE | 9193 | 1 | 0.3 | 0.3 |

STATE:MARYLAND BUSINESS ACTIVITY:N-U NARCOTIC TREATMENT PROGRAMS

| DRUG NAME | DRUG CODE | BUYERS | TOTAL GRAMS | AVG GRAMS |
|---|---|---|---|---|

| | | | | |
|---|---|---|---|---|
| BUPRENORPHINE | 9064 | 29 | 4,447.81 | 153.37 |
| METHADONE | 9250B | 74 | 633,153.81 | 8,556.13 |

STATE:MASSACHUSETTS BUSINESS ACTIVITY:A - PHARMACIES

| DRUG NAME | DRUG CODE | BUYERS | TOTAL GRAMS | AVG GRAMS |
|---|---|---|---|---|
| AMPHETAMINE | 1100 | 1,183 | 657,235.37 | 555.57 |
| DL-METHAMPHETAMINE RACEMIC BASE | 1105B | 4 | 17.28 | 4.32 |
| D-METHAMPHETAMINE | 1105D | 30 | 157.06 | 5.24 |
| LISDEXAMFETAMINE | 1205 | 1,122 | 130,955.84 | 116.72 |
| METHYLPHENIDATE (DL;D;L;ISOMERS) | 1724 | 1,184 | 565,965.65 | 478.01 |
| BARBITURIC ACID DERIVIATIVE OR SALT [PER 21C | 2100 | 9 | 75 | 8.33 |
| BUTALBITAL | 2165 | 699 | 21,900 | 31.33 |
| SECOBARBITAL (SCHEDULE 2) | 2315 | 13 | 219.72 | 16.9 |
| NABILONE | 7379 | 2 | 0.55 | 0.28 |
| CODEINE | 9050 | 1,172 | 200,813.18 | 171.34 |
| BUPRENORPHINE | 9064 | 1,134 | 143,095.49 | 126.19 |
| DIHYDROCODEINE | 9120 | 2 | 51.27 | 25.64 |
| OXYCODONE | 9143 | 1,194 | 1,037,016.83 | 868.52 |
| HYDROMORPHONE | 9150 | 1,114 | 23,537.85 | 21.13 |
| HYDROCODONE | 9193 | 1,193 | 135,339.49 | 113.44 |
| LEVORPHANOL | 9220L | 40 | 73.94 | 1.85 |
| MEPERIDINE (PETHIDINE) | 9230 | 239 | 8,125.32 | 34 |
| METHADONE | 9250B | 1,012 | 76,396.96 | 75.49 |
| MORPHINE | 9300 | 1,139 | 266,586.38 | 234.05 |
| OPIUM TINCTURE (LAUDANUM) | 9630 | 6 | 150.17 | 25.03 |
| OPIUM POWDERED | 9639 | 114 | 457.2 | 4.01 |
| OXYMORPHONE | 9652 | 337 | 11,020.56 | 32.7 |
| SUFENTANIL BASE | 9740 | 1 | 0.32 | 0.32 |
| TAPENTADOL | 9780 | 544 | 70,811 | 130.17 |
| FENTANYL BASE | 9801 | 1,080 | 5,321.79 | 4.93 |

STATISTICAL SUMMARY FOR RETAIL DRUG PURCHASES BY GRAMS WT

STATE:MASSACHUSETTS BUSINESS ACTIVITY:B - HOSPITALS

| DRUG NAME | DRUG CODE | BUYERS | TOTAL GRAMS | AVG GRAMS |
|---|---|---|---|---|
| AMPHETAMINE | 1100 | 145 | 29,372.99 | 202.57 |
| DL-METHAMPHETAMINE RACEMIC BASE | 1105B | 1 | 0.4 | 0.4 |
| LISDEXAMFETAMINE | 1205 | 64 | 3,469.76 | 54.21 |
| METHYLPHENIDATE (DL;D;L;ISOMERS) | 1724 | 159 | 30,061.26 | 189.06 |
| GAMMA HYDROXYBUTYRIC ACID(FDA APPROVED | 2012 | 1 | 3,195.07 | 3,195.07 |
| BARBITURIC ACID DERIVIATIVE OR SALT [PER 21C | 2100 | 1 | 5 | 5 |
| AMOBARBITAL (SCHEDULE 2) | 2125 | 9 | 29.62 | 3.29 |
| BUTALBITAL | 2165 | 24 | 590 | 24.58 |
| PENTOBARBITAL (SCHEDULE 2) | 2270 | 13 | 11,468.60 | 882.2 |
| NABILONE | 7379 | 1 | 0.35 | 0.35 |
| COCAINE | 9041L | 78 | 880.37 | 11.29 |
| CODEINE | 9050 | 163 | 11,047.40 | 67.78 |
| BUPRENORPHINE | 9064 | 156 | 8,308.75 | 53.26 |
| OXYCODONE | 9143 | 205 | 83,766.77 | 408.62 |
| HYDROMORPHONE | 9150 | 182 | 8,440.28 | 46.38 |
| HYDROCODONE | 9193 | 175 | 5,034.69 | 28.77 |
| LEVORPHANOL | 9220L | 2 | 1.39 | 0.7 |
| MEPERIDINE (PETHIDINE) | 9230 | 120 | 2,245.33 | 18.71 |
| METHADONE | 9250B | 157 | 12,056.67 | 76.79 |
| MORPHINE | 9300 | 197 | 32,190.06 | 163.4 |
| OPIUM POWDERED | 9639 | 53 | 236.88 | 4.47 |
| OXYMORPHONE | 9652 | 17 | 928.2 | 54.6 |
| ALFENTANIL | 9737 | 4 | 5.25 | 1.31 |
| REMIFENTANIL | 9739 | 68 | 74.38 | 1.09 |
| SUFENTANIL BASE | 9740 | 17 | 0.47 | 0.03 |
| TAPENTADOL | 9780 | 19 | 2,416.50 | 127.18 |
| FENTANYL BASE | 9801 | 188 | 772.11 | 4.11 |

STATE:MASSACHUSETTS BUSINESS ACTIVITY:C - PRACTITIONERS

| DRUG NAME | DRUG CODE | BUYERS | TOTAL GRAMS | AVG GRAMS |
|---|---|---|---|---|
| AMPHETAMINE | 1100 | 1 | 3.67 | 3.67 |
| PENTOBARBITAL (SCHEDULE 2) | 2270 | 174 | 94,451.98 | 542.83 |
| COCAINE | 9041L | 6 | 27.86 | 4.64 |
| CODEINE | 9050 | 27 | 837.84 | 31.03 |
| BUPRENORPHINE | 9064 | 537 | 69.96 | 0.13 |
| OXYCODONE | 9143 | 25 | 84.63 | 3.39 |
| HYDROMORPHONE | 9150 | 237 | 104.95 | 0.44 |
| HYDROCODONE | 9193 | 132 | 325.3 | 2.46 |
| MEPERIDINE (PETHIDINE) | 9230 | 38 | 495.23 | 13.03 |
| METHADONE | 9250B | 19 | 70.84 | 3.73 |
| MORPHINE | 9300 | 129 | 101.7 | 0.79 |
| OPIUM POWDERED | 9639 | 2 | 3.24 | 1.62 |
| OXYMORPHONE | 9652 | 19 | 4.45 | 0.23 |
| NOROXYMORPHONE | 9668 | 2 | 2 | 1 |
| REMIFENTANIL | 9739 | 22 | 2.32 | 0.11 |
| FENTANYL BASE | 9801 | 303 | 39.35 | 0.13 |

STATE:MASSACHUSETTS BUSINESS ACTIVITY:D - TEACHING INSTITUTIONS

| DRUG NAME | DRUG CODE | BUYERS | TOTAL GRAMS | AVG GRAMS |
|---|---|---|---|---|
| BUPRENORPHINE | 9064 | 1 | 0 | 0 |

STATE:MASSACHUSETTS BUSINESS ACTIVITY:M - MID-LEVEL PRACTITIONERS

| DRUG NAME | DRUG CODE | BUYERS | TOTAL GRAMS | AVG GRAMS |
|---|---|---|---|---|
| CODEINE | 9050 | 2 | 0.87 | 0.43 |
| BUPRENORPHINE | 9064 | 1 | 1.9 | 1.9 |

STATE:MASSACHUSETTS BUSINESS ACTIVITY:N-U NARCOTIC TREATMENT PROGRAMS

| DRUG NAME | DRUG CODE | BUYERS | TOTAL GRAMS | AVG GRAMS |
|---|---|---|---|---|
| BUPRENORPHINE | 9064 | 28 | 422.38 | 15.09 |
| METHADONE | 9250B | 69 | 452,819.96 | 6,562.61 |

STATE:MICHIGAN BUSINESS ACTIVITY:A - PHARMACIES

| DRUG NAME | DRUG CODE | BUYERS | TOTAL GRAMS | AVG GRAMS |
|---|---|---|---|---|
| AMPHETAMINE | 1100 | 2,433 | 899,195.53 | 369.58 |
| DL-METHAMPHETAMINE RACEMIC BASE | 1105B | 7 | 4.82 | 0.69 |
| D-METHAMPHETAMINE | 1105D | 31 | 113.28 | 3.65 |
| LISDEXAMFETAMINE | 1205 | 2,217 | 228,968.07 | 103.28 |
| METHYLPHENIDATE (DL;D;L;ISOMERS) | 1724 | 2,388 | 743,966.29 | 311.54 |
| BARBITURIC ACID DERIVIATIVE OR SALT [PER 21C | 2100 | 26 | 275 | 10.58 |
| BUTALBITAL | 2165 | 1,549 | 65,435 | 42.24 |
| SECOBARBITAL (SCHEDULE 2) | 2315 | 2 | 18.31 | 9.16 |
| COCAINE | 9041L | 6 | 18.71 | 3.12 |
| CODEINE | 9050 | 2,519 | 1,066,998.24 | 423.58 |
| BUPRENORPHINE | 9064 | 2,211 | 94,235.53 | 42.62 |
| DIHYDROCODEINE | 9120 | 8 | 108.95 | 13.62 |
| OXYCODONE | 9143 | 2,484 | 1,277,851.39 | 514.43 |
| HYDROMORPHONE | 9150 | 2,066 | 35,811.54 | 17.33 |
| HYDROCODONE | 9193 | 2,534 | 1,686,150.49 | 665.41 |
| LEVORPHANOL | 9220L | 3 | 3.6 | 1.2 |
| MEPERIDINE (PETHIDINE) | 9230 | 364 | 9,345.86 | 25.68 |
| METHADONE | 9250B | 1,956 | 169,561.96 | 86.69 |
| MORPHINE | 9300 | 2,364 | 811,650.57 | 343.34 |
| OPIUM TINCTURE (LAUDANUM) | 9630 | 6 | 47.29 | 7.88 |
| OPIUM POWDERED | 9639 | 156 | 430.2 | 2.76 |
| OXYMORPHONE | 9652 | 1,329 | 80,929.53 | 60.9 |
| REMIFENTANIL | 9739 | 5 | 3.09 | 0.62 |
| SUFENTANIL BASE | 9740 | 3 | 0.78 | 0.26 |
| TAPENTADOL | 9780 | 1,161 | 93,610.50 | 80.63 |
| FENTANYL BASE | 9801 | 2,277 | 13,489.65 | 5.92 |

STATISTICAL SUMMARY FOR RETAIL DRUG PURCHASES BY GRAMS WT

STATE:MICHIGAN BUSINESS ACTIVITY:B - HOSPITALS

| DRUG NAME | DRUG CODE | BUYERS | TOTAL GRAMS | AVG GRAMS |
|---|---|---|---|---|
| AMPHETAMINE | 1100 | 117 | 18,480.19 | 157.95 |
| LISDEXAMFETAMINE | 1205 | 56 | 4,229.38 | 75.52 |
| METHYLPHENIDATE (DL;D;L;ISOMERS) | 1724 | 151 | 18,531.48 | 122.73 |
| GAMMA HYDROXYBUTYRIC ACID(FDA APPROVED | 2012 | 1 | 297.22 | 297.22 |
| AMOBARBITAL (SCHEDULE 2) | 2125 | 8 | 19.6 | 2.45 |
| BUTALBITAL | 2165 | 26 | 665 | 25.58 |
| PENTOBARBITAL (SCHEDULE 2) | 2270 | 26 | 2,260.73 | 86.95 |
| COCAINE | 9041L | 108 | 1,406.20 | 13.02 |
| CODEINE | 9050 | 190 | 28,691.24 | 151.01 |
| BUPRENORPHINE | 9064 | 124 | 2,536.77 | 20.46 |
| OXYCODONE | 9143 | 187 | 68,715.88 | 367.46 |
| HYDROMORPHONE | 9150 | 182 | 6,573.50 | 36.12 |
| HYDROCODONE | 9193 | 196 | 91,441.79 | 466.54 |
| MEPERIDINE (PETHIDINE) | 9230 | 130 | 3,464.37 | 26.65 |
| METHADONE | 9250B | 158 | 11,161.38 | 70.64 |
| MORPHINE | 9300 | 188 | 74,834.92 | 398.06 |
| OPIUM POWDERED | 9639 | 105 | 1,030.68 | 9.82 |
| OXYMORPHONE | 9652 | 29 | 537.22 | 18.52 |
| ALFENTANIL | 9737 | 9 | 2.55 | 0.28 |
| REMIFENTANIL | 9739 | 54 | 37.83 | 0.7 |
| SUFENTANIL BASE | 9740 | 39 | 1.55 | 0.04 |
| TAPENTADOL | 9780 | 29 | 1,985.50 | 68.47 |
| FENTANYL BASE | 9801 | 185 | 1,309.87 | 7.08 |

STATE:MICHIGAN BUSINESS ACTIVITY:C - PRACTITIONERS

| DRUG NAME | DRUG CODE | BUYERS | TOTAL GRAMS | AVG GRAMS |
|---|---|---|---|---|
| AMPHETAMINE | 1100 | 3 | 49.31 | 16.44 |

| | | | | |
|---|---|---|---|---|
| METHYLPHENIDATE (DL;D;L;ISOMERS) | 1724 | 1 | 12.11 | 12.11 |
| BUTALBITAL | 2165 | 2 | 85 | 42.5 |
| PENTOBARBITAL (SCHEDULE 2) | 2270 | 395 | 322,267.02 | 815.87 |
| COCAINE | 9041L | 30 | 158.28 | 5.28 |
| CODEINE | 9050 | 122 | 2,417.45 | 19.82 |
| BUPRENORPHINE | 9064 | 663 | 270.74 | 0.41 |
| OXYCODONE | 9143 | 33 | 2,725.80 | 82.6 |
| HYDROMORPHONE | 9150 | 383 | 201.8 | 0.53 |
| HYDROCODONE | 9193 | 276 | 2,526.14 | 9.15 |
| MEPERIDINE (PETHIDINE) | 9230 | 125 | 1,701.93 | 13.62 |
| METHADONE | 9250B | 6 | 20.75 | 3.46 |
| MORPHINE | 9300 | 293 | 391.22 | 1.34 |
| OPIUM POWDERED | 9639 | 7 | 6.84 | 0.98 |
| OXYMORPHONE | 9652 | 4 | 0.61 | 0.15 |
| ALFENTANIL | 9737 | 6 | 2.56 | 0.43 |
| REMIFENTANIL | 9739 | 2 | 0.19 | 0.1 |
| SUFENTANIL BASE | 9740 | 3 | 0.01 | 0 |
| FENTANYL BASE | 9801 | 522 | 129.03 | 0.25 |

STATE:MICHIGAN BUSINESS ACTIVITY:D - TEACHING INSTITUTIONS

| DRUG NAME | DRUG CODE | BUYERS | TOTAL GRAMS | AVG GRAMS |
|---|---|---|---|---|
| PENTOBARBITAL (SCHEDULE 2) | 2270 | 1 | 13,329.23 | 13,329.23 |
| CODEINE | 9050 | 1 | 550.21 | 550.21 |
| BUPRENORPHINE | 9064 | 1 | 2.22 | 2.22 |
| HYDROMORPHONE | 9150 | 1 | 0.74 | 0.74 |
| HYDROCODONE | 9193 | 1 | 22.06 | 22.06 |
| METHADONE | 9250B | 1 | 42.39 | 42.39 |
| MORPHINE | 9300 | 2 | 7.19 | 3.59 |
| SUFENTANIL BASE | 9740 | 1 | 0 | 0 |
| FENTANYL BASE | 9801 | 2 | 3.2 | 1.6 |

STATISTICAL SUMMARY FOR RETAIL DRUG PURCHASES BY GRAMS WT

STATE:MICHIGAN BUSINESS ACTIVITY:M - MID-LEVEL PRACTITIONERS

| DRUG NAME | DRUG CODE | BUYERS | TOTAL GRAMS | AVG GRAMS |
|---|---|---|---|---|
| PENTOBARBITAL (SCHEDULE 2) | 2270 | 33 | 31,901.28 | 966.71 |
| CODEINE | 9050 | 5 | 20.55 | 4.11 |

STATE:MICHIGAN BUSINESS ACTIVITY:N-U NARCOTIC TREATMENT PROGRAMS

| DRUG NAME | DRUG CODE | BUYERS | TOTAL GRAMS | AVG GRAMS |
|---|---|---|---|---|
| BUPRENORPHINE | 9064 | 3 | 88.92 | 29.64 |
| METHADONE | 9250B | 43 | 332,166.30 | 7,724.80 |

STATE:MINNESOTA BUSINESS ACTIVITY:A - PHARMACIES

| DRUG NAME | DRUG CODE | BUYERS | TOTAL GRAMS | AVG GRAMS |
|---|---|---|---|---|
| AMPHETAMINE | 1100 | 1,048 | 414,393.75 | 395.41 |
| DL-METHAMPHETAMINE RACEMIC BASE | 1105B | 10 | 73.54 | 7.35 |
| D-METHAMPHETAMINE | 1105D | 41 | 286.81 | 7 |
| LISDEXAMFETAMINE | 1205 | 1,008 | 125,526.32 | 124.53 |
| METHYLPHENIDATE (DL;D;L;ISOMERS) | 1724 | 1,043 | 395,970.25 | 379.65 |
| BARBITURIC ACID DERIVIATIVE OR SALT [PER 21C | 2100 | 2 | 50 | 25 |
| BUTALBITAL | 2165 | 473 | 14,480 | 30.61 |
| NABILONE | 7379 | 1 | 0.3 | 0.3 |
| COCAINE | 9041L | 2 | 7.86 | 3.93 |
| CODEINE | 9050 | 1,040 | 162,506.19 | 156.26 |
| BUPRENORPHINE | 9064 | 867 | 21,554.59 | 24.86 |
| DIHYDROCODEINE | 9120 | 2 | 3.2 | 1.6 |
| OXYCODONE | 9143 | 1,063 | 612,286.47 | 576 |
| HYDROMORPHONE | 9150 | 990 | 19,650.39 | 19.85 |
| HYDROCODONE | 9193 | 1,062 | 197,094.23 | 185.59 |
| LEVORPHANOL | 9220L | 7 | 2.55 | 0.36 |
| MEPERIDINE (PETHIDINE) | 9230 | 163 | 2,051.73 | 12.59 |

| | | | | |
|---|---|---|---|---|
| METHADONE | 9250B | 868 | 36,879.69 | 42.49 |
| MORPHINE | 9300 | 1,023 | 192,495.01 | 188.17 |
| OPIUM TINCTURE (LAUDANUM) | 9630 | 1 | 1.18 | 1.18 |
| OPIUM POWDERED | 9639 | 156 | 780.12 | 5 |
| OXYMORPHONE | 9652 | 248 | 2,433.87 | 9.81 |
| ALFENTANIL | 9737 | 3 | 0.83 | 0.28 |
| SUFENTANIL BASE | 9740 | 1 | 0.03 | 0.03 |
| TAPENTADOL | 9780 | 327 | 20,211.50 | 61.81 |
| FENTANYL BASE | 9801 | 1,000 | 4,405.26 | 4.41 |

STATE:MINNESOTA BUSINESS ACTIVITY:B - HOSPITALS

| DRUG NAME | DRUG CODE | BUYERS | TOTAL GRAMS | AVG GRAMS |
|---|---|---|---|---|
| AMPHETAMINE | 1100 | 85 | 7,755.22 | 91.24 |
| DL-METHAMPHETAMINE RACEMIC BASE | 1105B | 2 | 68.72 | 34.36 |
| D-METHAMPHETAMINE | 1105D | 3 | 2.01 | 0.67 |
| LISDEXAMFETAMINE | 1205 | 51 | 1,849.34 | 36.26 |
| METHYLPHENIDATE (DL;D;L;ISOMERS) | 1724 | 84 | 8,452.67 | 100.63 |
| AMOBARBITAL (SCHEDULE 2) | 2125 | 5 | 23.7 | 4.74 |
| BUTALBITAL | 2165 | 12 | 95 | 7.92 |
| PENTOBARBITAL (SCHEDULE 2) | 2270 | 14 | 15,388.53 | 1,099.18 |
| PHENCYCLIDINE (PCP) | 7471 | 1 | 0.22 | 0.22 |
| COCAINE | 9041L | 88 | 731.35 | 8.31 |
| CODEINE | 9050 | 132 | 6,885.42 | 52.16 |
| BUPRENORPHINE | 9064 | 63 | 1,094.39 | 17.37 |
| OXYCODONE | 9143 | 149 | 37,150.14 | 249.33 |
| HYDROMORPHONE | 9150 | 152 | 5,309.03 | 34.93 |
| HYDROCODONE | 9193 | 152 | 14,678.59 | 96.57 |
| MEPERIDINE (PETHIDINE) | 9230 | 122 | 1,023.58 | 8.39 |
| METHADONE | 9250B | 101 | 3,217.77 | 31.86 |
| MORPHINE | 9300 | 155 | 21,841.84 | 140.92 |
| OPIUM POWDERED | 9639 | 90 | 903.6 | 10.04 |
| OXYMORPHONE | 9652 | 13 | 27 | 2.08 |

| | | | | |
|---|---|---|---|---|
| ALFENTANIL | 9737 | 19 | 10.93 | 0.58 |
| REMIFENTANIL | 9739 | 26 | 25.88 | 1 |
| SUFENTANIL BASE | 9740 | 29 | 0.76 | 0.03 |
| TAPENTADOL | 9780 | 10 | 150 | 15 |
| FENTANYL BASE | 9801 | 149 | 584.05 | 3.92 |

STATE:MINNESOTA BUSINESS ACTIVITY:C - PRACTITIONERS

| DRUG NAME | DRUG CODE | BUYERS | TOTAL GRAMS | AVG GRAMS |
|---|---|---|---|---|
| METHYLPHENIDATE (DL;D;L;ISOMERS) | 1724 | 1 | 44.96 | 44.96 |
| PENTOBARBITAL (SCHEDULE 2) | 2270 | 132 | 103,337.75 | 782.86 |
| COCAINE | 9041L | 23 | 144.78 | 6.29 |
| CODEINE | 9050 | 35 | 377.36 | 10.78 |
| BUPRENORPHINE | 9064 | 500 | 27.39 | 0.05 |
| OXYCODONE | 9143 | 54 | 409.59 | 7.59 |
| HYDROMORPHONE | 9150 | 359 | 162.23 | 0.45 |
| HYDROCODONE | 9193 | 234 | 626.22 | 2.68 |
| MEPERIDINE (PETHIDINE) | 9230 | 52 | 253.17 | 4.87 |
| METHADONE | 9250B | 3 | 7.24 | 2.41 |
| MORPHINE | 9300 | 204 | 269.25 | 1.32 |
| OPIUM POWDERED | 9639 | 6 | 9.36 | 1.56 |
| OXYMORPHONE | 9652 | 3 | 0.09 | 0.03 |
| ALFENTANIL | 9737 | 8 | 1.83 | 0.23 |
| REMIFENTANIL | 9739 | 5 | 0.87 | 0.17 |
| SUFENTANIL BASE | 9740 | 3 | 0.01 | 0 |
| FENTANYL BASE | 9801 | 343 | 47.74 | 0.14 |

STATE:MINNESOTA BUSINESS ACTIVITY:D - TEACHING INSTITUTIONS

| DRUG NAME | DRUG CODE | BUYERS | TOTAL GRAMS | AVG GRAMS |
|---|---|---|---|---|
| BUPRENORPHINE | 9064 | 7 | 0.07 | 0.01 |
| HYDROMORPHONE | 9150 | 2 | 0.21 | 0.11 |

| | | | | |
|---|---|---|---|---|
| MORPHINE | 9300 | 1 | 0.19 | 0.19 |

STATE:MINNESOTA BUSINESS ACTIVITY:M - MID-LEVEL PRACTITIONERS

| DRUG NAME | DRUG CODE | BUYERS | TOTAL GRAMS | AVG GRAMS |
|---|---|---|---|---|
| CODEINE | 9050 | 2 | 11.05 | 5.53 |
| HYDROCODONE | 9193 | 1 | 1 | 1 |
| MORPHINE | 9300 | 1 | 0 | 0 |
| FENTANYL BASE | 9801 | 3 | 0.04 | 0.01 |

STATE:MINNESOTA BUSINESS ACTIVITY:N-U NARCOTIC TREATMENT PROGRAMS

| DRUG NAME | DRUG CODE | BUYERS | TOTAL GRAMS | AVG GRAMS |
|---|---|---|---|---|
| BUPRENORPHINE | 9064 | 10 | 1,994.41 | 199.44 |
| METHADONE | 9250B | 19 | 180,906.71 | 9,521.41 |

STATE:MISSISSIPPI BUSINESS ACTIVITY:A - PHARMACIES

| DRUG NAME | DRUG CODE | BUYERS | TOTAL GRAMS | AVG GRAMS |
|---|---|---|---|---|
| AMPHETAMINE | 1100 | 760 | 232,822.18 | 306.34 |
| DL-METHAMPHETAMINE RACEMIC BASE | 1105B | 2 | 2.81 | 1.41 |
| D-METHAMPHETAMINE | 1105D | 6 | 39.37 | 6.56 |
| LISDEXAMFETAMINE | 1205 | 739 | 148,367.05 | 200.77 |
| METHYLPHENIDATE (DL;D;L;ISOMERS) | 1724 | 765 | 171,384.76 | 224.03 |
| BARBITURIC ACID DERIVIATIVE OR SALT [PER 21C | 2100 | 6 | 35 | 5.83 |
| BUTALBITAL | 2165 | 409 | 13,255 | 32.41 |
| COCAINE | 9041L | 3 | 73.39 | 24.46 |
| CODEINE | 9050 | 762 | 107,504.42 | 141.08 |
| BUPRENORPHINE | 9064 | 704 | 36,356.46 | 51.64 |
| DIHYDROCODEINE | 9120 | 283 | 1,086.32 | 3.84 |
| OXYCODONE | 9143 | 783 | 425,724.07 | 543.71 |

| | | | | |
|---|---|---|---|---|
| HYDROMORPHONE | 9150 | 611 | 18,771.45 | 30.72 |
| HYDROCODONE | 9193 | 792 | 505,867.84 | 638.72 |
| LEVORPHANOL | 9220L | 5 | 3.02 | 0.6 |
| MEPERIDINE (PETHIDINE) | 9230 | 570 | 14,886.42 | 26.12 |
| METHADONE | 9250B | 505 | 20,584.79 | 40.76 |
| MORPHINE | 9300 | 741 | 159,472 | 215.21 |
| OPIUM POWDERED | 9639 | 30 | 52.92 | 1.76 |
| OXYMORPHONE | 9652 | 496 | 19,336.67 | 38.99 |
| SUFENTANIL BASE | 9740 | 1 | 41.42 | 41.42 |
| TAPENTADOL | 9780 | 513 | 56,347.50 | 109.84 |
| FENTANYL BASE | 9801 | 749 | 5,081.42 | 6.78 |

STATE:MISSISSIPPI BUSINESS ACTIVITY:B - HOSPITALS

| DRUG NAME | DRUG CODE | BUYERS | TOTAL GRAMS | AVG GRAMS |
|---|---|---|---|---|
| AMPHETAMINE | 1100 | 57 | 6,136.73 | 107.66 |
| LISDEXAMFETAMINE | 1205 | 34 | 3,813.73 | 112.17 |
| METHYLPHENIDATE (DL;D;L;ISOMERS) | 1724 | 65 | 5,317.55 | 81.81 |
| AMOBARBITAL (SCHEDULE 2) | 2125 | 1 | 5.92 | 5.92 |
| BUTALBITAL | 2165 | 16 | 160 | 10 |
| PENTOBARBITAL (SCHEDULE 2) | 2270 | 3 | 98.89 | 32.96 |
| COCAINE | 9041L | 31 | 275.41 | 8.88 |
| CODEINE | 9050 | 114 | 5,655.06 | 49.61 |
| BUPRENORPHINE | 9064 | 60 | 713.78 | 11.9 |
| DIHYDROCODEINE | 9120 | 6 | 27.77 | 4.63 |
| OXYCODONE | 9143 | 135 | 15,949.44 | 118.14 |
| HYDROMORPHONE | 9150 | 121 | 1,951.65 | 16.13 |
| HYDROCODONE | 9193 | 145 | 29,196.35 | 201.35 |
| MEPERIDINE (PETHIDINE) | 9230 | 120 | 7,076.19 | 58.97 |
| METHADONE | 9250B | 60 | 910.41 | 15.17 |
| MORPHINE | 9300 | 143 | 14,001.14 | 97.91 |
| OPIUM POWDERED | 9639 | 30 | 96.84 | 3.23 |
| OXYMORPHONE | 9652 | 17 | 88.9 | 5.23 |

| | | | | |
|---|---|---|---|---|
| ALFENTANIL | 9737 | 6 | 0.58 | 0.1 |
| REMIFENTANIL | 9739 | 7 | 1.82 | 0.26 |
| SUFENTANIL BASE | 9740 | 21 | 0.43 | 0.02 |
| TAPENTADOL | 9780 | 19 | 1,438 | 75.68 |
| FENTANYL BASE | 9801 | 140 | 395.53 | 2.83 |

STATE:MISSISSIPPI BUSINESS ACTIVITY:C - PRACTITIONERS

| DRUG NAME | DRUG CODE | BUYERS | TOTAL GRAMS | AVG GRAMS |
|---|---|---|---|---|
| PENTOBARBITAL (SCHEDULE 2) | 2270 | 24 | 22,831.15 | 951.3 |
| COCAINE | 9041L | 3 | 33.57 | 11.19 |
| CODEINE | 9050 | 7 | 58.33 | 8.33 |
| BUPRENORPHINE | 9064 | 110 | 14.91 | 0.14 |
| DIHYDROCODEINE | 9120 | 84 | 28.98 | 0.34 |
| OXYCODONE | 9143 | 10 | 3,301.25 | 330.13 |
| HYDROMORPHONE | 9150 | 30 | 13.02 | 0.43 |
| HYDROCODONE | 9193 | 40 | 129.3 | 3.23 |
| MEPERIDINE (PETHIDINE) | 9230 | 44 | 764.74 | 17.38 |
| METHADONE | 9250B | 2 | 0.72 | 0.36 |
| MORPHINE | 9300 | 53 | 104.34 | 1.97 |
| OPIUM POWDERED | 9639 | 1 | 1.08 | 1.08 |
| NOROXYMORPHONE | 9668 | 1 | 16.18 | 16.18 |
| REMIFENTANIL | 9739 | 5 | 0.12 | 0.02 |
| FENTANYL BASE | 9801 | 103 | 10.93 | 0.11 |

STATE:MISSISSIPPI BUSINESS ACTIVITY:D - TEACHING INSTITUTIONS

| DRUG NAME | DRUG CODE | BUYERS | TOTAL GRAMS | AVG GRAMS |
|---|---|---|---|---|
| BUPRENORPHINE | 9064 | 1 | 0.02 | 0.02 |
| HYDROMORPHONE | 9150 | 1 | 0.07 | 0.07 |

STATE:MISSISSIPPI BUSINESS ACTIVITY:M - MID-LEVEL PRACTITIONERS

| DRUG NAME | DRUG CODE | BUYERS | TOTAL GRAMS | AVG GRAMS |
|---|---|---|---|---|
| CODEINE | 9050 | 1 | 0.43 | 0.43 |
| DIHYDROCODEINE | 9120 | 42 | 10.66 | 0.25 |

STATE:MISSISSIPPI BUSINESS ACTIVITY:N-U NARCOTIC TREATMENT PROGRAMS

| DRUG NAME | DRUG CODE | BUYERS | TOTAL GRAMS | AVG GRAMS |
|---|---|---|---|---|
| METHADONE | 9250B | 1 | 5,777.82 | 5,777.82 |

STATE:MISSOURI BUSINESS ACTIVITY:A - PHARMACIES

| DRUG NAME | DRUG CODE | BUYERS | TOTAL GRAMS | AVG GRAMS |
|---|---|---|---|---|
| AMPHETAMINE | 1100 | 1,344 | 505,995.67 | 376.48 |
| DL-METHAMPHETAMINE RACEMIC BASE | 1105B | 18 | 97.25 | 5.4 |
| D-METHAMPHETAMINE | 1105D | 14 | 183.98 | 13.14 |
| LISDEXAMFETAMINE | 1205 | 1,293 | 265,756.62 | 205.53 |
| METHYLPHENIDATE (DL;D;L;ISOMERS) | 1724 | 1,343 | 517,387.64 | 385.25 |
| GAMMA HYDROXYBUTYRIC ACID(FDA APPROVED 25,314,852.67 | 2012 | 1 | 25,314,852.67 | |
| BARBITURIC ACID DERIVIATIVE OR SALT [PER 21C | 2100 | 7 | 110 | 15.71 |
| BUTALBITAL | 2165 | 782 | 41,985 | 53.69 |
| SECOBARBITAL (SCHEDULE 2) | 2315 | 4 | 146.48 | 36.62 |
| NABILONE | 7379 | 3 | 0.45 | 0.15 |
| COCAINE | 9041L | 3 | 8.43 | 2.81 |
| CODEINE | 9050 | 1,331 | 287,222.59 | 215.79 |
| BUPRENORPHINE | 9064 | 1,226 | 30,734.79 | 25.07 |
| DIHYDROCODEINE | 9120 | 8 | 70.5 | 8.81 |
| OXYCODONE | 9143 | 1,352 | 1,259,524.49 | 931.6 |
| HYDROMORPHONE | 9150 | 1,215 | 26,369.89 | 21.7 |
| HYDROCODONE | 9193 | 1,360 | 748,416.86 | 550.31 |

| | | | | |
|---|---|---|---|---|
| LEVORPHANOL | 9220L | 27 | 30.88 | 1.14 |
| MEPERIDINE (PETHIDINE) | 9230 | 582 | 13,297.24 | 22.85 |
| METHADONE | 9250B | 1,153 | 58,883.27 | 51.07 |
| MORPHINE | 9300 | 1,320 | 396,782.63 | 300.59 |
| OPIUM TINCTURE (LAUDANUM) | 9630 | 6 | 26.01 | 4.34 |
| OPIUM POWDERED | 9639 | 126 | 407.88 | 3.24 |
| OXYMORPHONE | 9652 | 867 | 24,122.79 | 27.82 |
| REMIFENTANIL | 9739 | 3 | 0.17 | 0.06 |
| SUFENTANIL BASE | 9740 | 1 | 0 | 0 |
| TAPENTADOL | 9780 | 941 | 113,648.50 | 120.77 |
| FENTANYL BASE | 9801 | 1,304 | 9,267.68 | 7.11 |

STATE:MISSOURI BUSINESS ACTIVITY:B - HOSPITALS

| DRUG NAME | DRUG CODE | BUYERS | TOTAL GRAMS | AVG GRAMS |
|---|---|---|---|---|
| AMPHETAMINE | 1100 | 96 | 8,889.12 | 92.59 |
| LISDEXAMFETAMINE | 1205 | 45 | 2,739.62 | 60.88 |
| METHYLPHENIDATE (DL;D;L;ISOMERS) | 1724 | 119 | 10,473.37 | 88.01 |
| AMOBARBITAL (SCHEDULE 2) | 2125 | 5 | 16.86 | 3.37 |
| BUTALBITAL | 2165 | 13 | 180 | 13.85 |
| PENTOBARBITAL (SCHEDULE 2) | 2270 | 14 | 1,117.38 | 79.81 |
| COCAINE | 9041L | 105 | 700.88 | 6.68 |
| CODEINE | 9050 | 158 | 8,813.33 | 55.78 |
| BUPRENORPHINE | 9064 | 86 | 1,012.17 | 11.77 |
| DIHYDROCODEINE | 9120 | 1 | 1.07 | 1.07 |
| OXYCODONE | 9143 | 215 | 66,114.90 | 307.51 |
| HYDROMORPHONE | 9150 | 177 | 3,793.27 | 21.43 |
| HYDROCODONE | 9193 | 232 | 34,917.14 | 150.5 |
| MEPERIDINE (PETHIDINE) | 9230 | 174 | 5,823.12 | 33.47 |
| METHADONE | 9250B | 120 | 4,730.97 | 39.42 |
| MORPHINE | 9300 | 230 | 40,782.20 | 177.31 |
| OPIUM POWDERED | 9639 | 87 | 392.76 | 4.51 |
| OXYMORPHONE | 9652 | 25 | 172.35 | 6.89 |

| | | | | |
|---|---|---|---|---|
| ALFENTANIL | 9737 | 13 | 5.31 | 0.41 |
| REMIFENTANIL | 9739 | 42 | 20.78 | 0.49 |
| SUFENTANIL BASE | 9740 | 41 | 1.46 | 0.04 |
| TAPENTADOL | 9780 | 34 | 1,769 | 52.03 |
| FENTANYL BASE | 9801 | 248 | 845.89 | 3.41 |

STATE:MISSOURI BUSINESS ACTIVITY:C - PRACTITIONERS

| DRUG NAME | DRUG CODE | BUYERS | TOTAL GRAMS | AVG GRAMS |
|---|---|---|---|---|
| AMPHETAMINE | 1100 | 3 | 22.14 | 7.38 |
| PENTOBARBITAL (SCHEDULE 2) | 2270 | 171 | 134,627.62 | 787.3 |
| COCAINE | 9041L | 2 | 2 | 1 |
| CODEINE | 9050 | 42 | 1,497.99 | 35.67 |
| BUPRENORPHINE | 9064 | 436 | 58.45 | 0.13 |
| OXYCODONE | 9143 | 19 | 1,652.57 | 86.98 |
| HYDROMORPHONE | 9150 | 143 | 256.12 | 1.79 |
| HYDROCODONE | 9193 | 207 | 1,454.01 | 7.02 |
| MEPERIDINE (PETHIDINE) | 9230 | 36 | 379.67 | 10.55 |
| METHADONE | 9250B | 2 | 0.89 | 0.45 |
| MORPHINE | 9300 | 140 | 682.78 | 4.88 |
| OXYMORPHONE | 9652 | 5 | 16.51 | 3.3 |
| NOROXYMORPHONE | 9668 | 8 | 56.8 | 7.1 |
| REMIFENTANIL | 9739 | 1 | 0.04 | 0.04 |
| SUFENTANIL BASE | 9740 | 4 | 0.05 | 0.01 |
| FENTANYL BASE | 9801 | 183 | 18.84 | 0.1 |

STATE:MISSOURI BUSINESS ACTIVITY:D - TEACHING INSTITUTIONS

| DRUG NAME | DRUG CODE | BUYERS | TOTAL GRAMS | AVG GRAMS |
|---|---|---|---|---|
| PENTOBARBITAL (SCHEDULE 2) | 2270 | 1 | 8,886.15 | 8,886.15 |
| CODEINE | 9050 | 1 | 68.51 | 68.51 |
| BUPRENORPHINE | 9064 | 2 | 0.47 | 0.23 |

| | | | | |
|---|---|---|---|---|
| HYDROMORPHONE | 9150 | 1 | 7.74 | 7.74 |
| HYDROCODONE | 9193 | 1 | 6.88 | 6.88 |
| MORPHINE | 9300 | 1 | 13.95 | 13.95 |
| FENTANYL BASE | 9801 | 4 | 1.59 | 0.4 |

STATE:MISSOURI BUSINESS ACTIVITY:M - MID-LEVEL PRACTITIONERS

| DRUG NAME | DRUG CODE | BUYERS | TOTAL GRAMS | AVG GRAMS |
|---|---|---|---|---|
| HYDROMORPHONE | 9150 | 2 | 0.06 | 0.03 |
| MEPERIDINE (PETHIDINE) | 9230 | 1 | 1.09 | 1.09 |
| MORPHINE | 9300 | 46 | 17.56 | 0.38 |
| FENTANYL BASE | 9801 | 49 | 0.52 | 0.01 |

STATE:MISSOURI BUSINESS ACTIVITY:N-U NARCOTIC TREATMENT PROGRAMS

| DRUG NAME | DRUG CODE | BUYERS | TOTAL GRAMS | AVG GRAMS |
|---|---|---|---|---|
| BUPRENORPHINE | 9064 | 13 | 1,017.29 | 78.25 |
| METHADONE | 9250B | 14 | 103,038.90 | 7,359.92 |

STATE:MONTANA BUSINESS ACTIVITY:A - PHARMACIES

| DRUG NAME | DRUG CODE | BUYERS | TOTAL GRAMS | AVG GRAMS |
|---|---|---|---|---|
| AMPHETAMINE | 1100 | 237 | 48,154.51 | 203.18 |
| D-METHAMPHETAMINE | 1105D | 4 | 9.64 | 2.41 |
| LISDEXAMFETAMINE | 1205 | 231 | 20,268.19 | 87.74 |
| METHYLPHENIDATE (DL;D;L;ISOMERS) | 1724 | 240 | 64,442.07 | 268.51 |
| BARBITURIC ACID DERIVIATIVE OR SALT [PER 21C | 2100 | 1 | 25 | 25 |
| BUTALBITAL | 2165 | 129 | 6,170 | 47.83 |
| COCAINE | 9041L | 2 | 0.57 | 0.29 |
| CODEINE | 9050 | 243 | 40,313.42 | 165.9 |
| BUPRENORPHINE | 9064 | 221 | 6,020.64 | 27.24 |

| | | | | |
|---|---|---|---|---|
| OXYCODONE | 9143 | 242 | 167,168.24 | 690.78 |
| HYDROMORPHONE | 9150 | 235 | 5,999.82 | 25.53 |
| HYDROCODONE | 9193 | 242 | 96,790.88 | 399.96 |
| MEPERIDINE (PETHIDINE) | 9230 | 122 | 2,104.24 | 17.25 |
| METHADONE | 9250B | 215 | 9,681.88 | 45.03 |
| MORPHINE | 9300 | 245 | 72,414.54 | 295.57 |
| OPIUM POWDERED | 9639 | 22 | 34.2 | 1.55 |
| OXYMORPHONE | 9652 | 122 | 1,942.06 | 15.92 |
| TAPENTADOL | 9780 | 136 | 9,833.50 | 72.31 |
| FENTANYL BASE | 9801 | 244 | 1,321.03 | 5.41 |

STATE:MONTANA BUSINESS ACTIVITY:B - HOSPITALS

| DRUG NAME | DRUG CODE | BUYERS | TOTAL GRAMS | AVG GRAMS |
|---|---|---|---|---|
| AMPHETAMINE | 1100 | 37 | 3,862.24 | 104.38 |
| LISDEXAMFETAMINE | 1205 | 16 | 1,161.40 | 72.59 |
| METHYLPHENIDATE (DL;D;L;ISOMERS) | 1724 | 41 | 6,250.02 | 152.44 |
| BUTALBITAL | 2165 | 10 | 230 | 23 |
| PENTOBARBITAL (SCHEDULE 2) | 2270 | 3 | 26.43 | 8.81 |
| COCAINE | 9041L | 27 | 205.24 | 7.6 |
| CODEINE | 9050 | 61 | 5,261.84 | 86.26 |
| BUPRENORPHINE | 9064 | 29 | 683.03 | 23.55 |
| OXYCODONE | 9143 | 80 | 15,831.70 | 197.9 |
| HYDROMORPHONE | 9150 | 68 | 1,194.06 | 17.56 |
| HYDROCODONE | 9193 | 77 | 17,731 | 230.27 |
| MEPERIDINE (PETHIDINE) | 9230 | 60 | 1,005.28 | 16.75 |
| METHADONE | 9250B | 33 | 1,252.70 | 37.96 |
| MORPHINE | 9300 | 82 | 13,329.38 | 162.55 |
| OPIUM POWDERED | 9639 | 22 | 182.52 | 8.3 |
| OXYMORPHONE | 9652 | 4 | 27.92 | 6.98 |
| ALFENTANIL | 9737 | 11 | 1.55 | 0.14 |
| REMIFENTANIL | 9739 | 16 | 7.2 | 0.45 |
| SUFENTANIL BASE | 9740 | 16 | 0.15 | 0.01 |

| | | | | |
|---|---|---|---|---|
| TAPENTADOL | 9780 | 8 | 275 | 34.38 |
| FENTANYL BASE | 9801 | 79 | 273.43 | 3.46 |

STATE:MONTANA BUSINESS ACTIVITY:C - PRACTITIONERS

| DRUG NAME | DRUG CODE | BUYERS | TOTAL GRAMS | AVG GRAMS |
|---|---|---|---|---|
| PENTOBARBITAL (SCHEDULE 2) | 2270 | 80 | 65,412.27 | 817.65 |
| COCAINE | 9041L | 1 | 2.29 | 2.29 |
| CODEINE | 9050 | 8 | 152.58 | 19.07 |
| BUPRENORPHINE | 9064 | 123 | 4.02 | 0.03 |
| OXYCODONE | 9143 | 3 | 3.59 | 1.2 |
| HYDROMORPHONE | 9150 | 79 | 32.39 | 0.41 |
| HYDROCODONE | 9193 | 62 | 133.81 | 2.16 |
| MEPERIDINE (PETHIDINE) | 9230 | 12 | 159.48 | 13.29 |
| METHADONE | 9250B | 1 | 0.27 | 0.27 |
| MORPHINE | 9300 | 76 | 105.61 | 1.39 |
| OXYMORPHONE | 9652 | 2 | 0.03 | 0.01 |
| SUFENTANIL BASE | 9740 | 2 | 0.03 | 0.01 |
| FENTANYL BASE | 9801 | 80 | 13.95 | 0.17 |

STATE:MONTANA BUSINESS ACTIVITY:D - TEACHING INSTITUTIONS

| DRUG NAME | DRUG CODE | BUYERS | TOTAL GRAMS | AVG GRAMS |
|---|---|---|---|---|
| BUPRENORPHINE | 9064 | 1 | 0 | 0 |

STATE:MONTANA BUSINESS ACTIVITY:M - MID-LEVEL PRACTITIONERS

| DRUG NAME | DRUG CODE | BUYERS | TOTAL GRAMS | AVG GRAMS |
|---|---|---|---|---|
| PENTOBARBITAL (SCHEDULE 2) | 2270 | 4 | 3,733.55 | 933.39 |
| MORPHINE | 9300 | 2 | 1.13 | 0.56 |
| FENTANYL BASE | 9801 | 2 | 0.02 | 0.01 |

STATE:MONTANA BUSINESS ACTIVITY:N-U NARCOTIC TREATMENT PROGRAMS

| DRUG NAME | DRUG CODE | BUYERS | TOTAL GRAMS | AVG GRAMS |
|---|---|---|---|---|
| BUPRENORPHINE | 9064 | 4 | 519.37 | 129.84 |
| METHADONE | 9250B | 4 | 18,782.40 | 4,695.60 |

STATE:NEBRASKA BUSINESS ACTIVITY:A - PHARMACIES

| DRUG NAME | DRUG CODE | BUYERS | TOTAL GRAMS | AVG GRAMS |
|---|---|---|---|---|
| AMPHETAMINE | 1100 | 449 | 93,506.88 | 208.26 |
| DL-METHAMPHETAMINE RACEMIC BASE | 1105B | 7 | 13.26 | 1.89 |
| D-METHAMPHETAMINE | 1105D | 10 | 28.12 | 2.81 |
| LISDEXAMFETAMINE | 1205 | 429 | 62,888.03 | 146.59 |
| METHYLPHENIDATE (DL;D;L;ISOMERS) | 1724 | 453 | 123,159.27 | 271.87 |
| BUTALBITAL | 2165 | 224 | 8,260 | 36.88 |
| NABILONE | 7379 | 1 | 0.7 | 0.7 |
| CODEINE | 9050 | 447 | 49,769.88 | 111.34 |
| BUPRENORPHINE | 9064 | 369 | 4,369.29 | 11.84 |
| DIHYDROCODEINE | 9120 | 1 | 6.41 | 6.41 |
| OXYCODONE | 9143 | 461 | 166,056.27 | 360.21 |
| HYDROMORPHONE | 9150 | 410 | 4,804.84 | 11.72 |
| HYDROCODONE | 9193 | 466 | 135,914.19 | 291.66 |
| LEVORPHANOL | 9220L | 7 | 3.6 | 0.51 |
| MEPERIDINE (PETHIDINE) | 9230 | 126 | 2,045.63 | 16.24 |
| METHADONE | 9250B | 375 | 13,416 | 35.78 |
| MORPHINE | 9300 | 458 | 68,721.01 | 150.05 |
| OPIUM TINCTURE (LAUDANUM) | 9630 | 1 | 14.19 | 14.19 |
| OPIUM POWDERED | 9639 | 59 | 119.88 | 2.03 |
| OXYMORPHONE | 9652 | 254 | 6,455.10 | 25.41 |
| ALFENTANIL | 9737 | 2 | 0.34 | 0.17 |
| TAPENTADOL | 9780 | 348 | 40,979 | 117.76 |

| | | | | |
|---|---|---|---|---|
| FENTANYL BASE | 9801 | 446 | 2,127.22 | 4.77 |

STATE:NEBRASKA BUSINESS ACTIVITY:B - HOSPITALS

| DRUG NAME | DRUG CODE | BUYERS | TOTAL GRAMS | AVG GRAMS |
|---|---|---|---|---|
| AMPHETAMINE | 1100 | 37 | 3,400.65 | 91.91 |
| LISDEXAMFETAMINE | 1205 | 25 | 506.42 | 20.26 |
| METHYLPHENIDATE (DL;D;L;ISOMERS) | 1724 | 60 | 4,457.49 | 74.29 |
| AMOBARBITAL (SCHEDULE 2) | 2125 | 2 | 12.3 | 6.15 |
| BUTALBITAL | 2165 | 6 | 30 | 5 |
| PENTOBARBITAL (SCHEDULE 2) | 2270 | 7 | 493.07 | 70.44 |
| COCAINE | 9041L | 48 | 145.45 | 3.03 |
| CODEINE | 9050 | 95 | 4,175.41 | 43.95 |
| BUPRENORPHINE | 9064 | 40 | 283.24 | 7.08 |
| OXYCODONE | 9143 | 122 | 11,481.64 | 94.11 |
| HYDROMORPHONE | 9150 | 99 | 971.75 | 9.82 |
| HYDROCODONE | 9193 | 125 | 10,192.74 | 81.54 |
| MEPERIDINE (PETHIDINE) | 9230 | 107 | 1,292.35 | 12.08 |
| METHADONE | 9250B | 54 | 1,798.92 | 33.31 |
| MORPHINE | 9300 | 124 | 11,555.80 | 93.19 |
| OPIUM POWDERED | 9639 | 61 | 287.28 | 4.71 |
| OXYMORPHONE | 9652 | 12 | 45.23 | 3.77 |
| ALFENTANIL | 9737 | 24 | 3.22 | 0.13 |
| REMIFENTANIL | 9739 | 20 | 3.97 | 0.2 |
| SUFENTANIL BASE | 9740 | 35 | 0.39 | 0.01 |
| TAPENTADOL | 9780 | 49 | 1,182.50 | 24.13 |
| FENTANYL BASE | 9801 | 130 | 253.58 | 1.95 |

STATE:NEBRASKA BUSINESS ACTIVITY:C - PRACTITIONERS

| DRUG NAME | DRUG CODE | BUYERS | TOTAL GRAMS | AVG GRAMS |
|---|---|---|---|---|
| GAMMA HYDROXYBUTYRIC ACID(FDA APPROVED | 2012 | 1 | 222.91 | 222.91 |

| | | | | |
|---|---|---|---|---|
| PENTOBARBITAL (SCHEDULE 2) | 2270 | 108 | 81,849.64 | 757.87 |
| COCAINE | 9041L | 1 | 5 | 5 |
| CODEINE | 9050 | 13 | 539.71 | 41.52 |
| BUPRENORPHINE | 9064 | 151 | 6.98 | 0.05 |
| OXYCODONE | 9143 | 9 | 666.66 | 74.07 |
| HYDROMORPHONE | 9150 | 84 | 76.11 | 0.91 |
| HYDROCODONE | 9193 | 75 | 212.65 | 2.84 |
| MEPERIDINE (PETHIDINE) | 9230 | 29 | 123.54 | 4.26 |
| METHADONE | 9250B | 1 | 4.47 | 4.47 |
| MORPHINE | 9300 | 48 | 83.45 | 1.74 |
| OPIUM POWDERED | 9639 | 1 | 6.48 | 6.48 |
| NOROXYMORPHONE | 9668 | 1 | 1.59 | 1.59 |
| ALFENTANIL | 9737 | 2 | 0.52 | 0.26 |
| TAPENTADOL | 9780 | 2 | 70 | 35 |
| FENTANYL BASE | 9801 | 101 | 10.94 | 0.11 |

STATE:NEBRASKA BUSINESS ACTIVITY:D - TEACHING INSTITUTIONS

| DRUG NAME | DRUG CODE | BUYERS | TOTAL GRAMS | AVG GRAMS |
|---|---|---|---|---|
| PENTOBARBITAL (SCHEDULE 2) | 2270 | 1 | 266.58 | 266.58 |
| BUPRENORPHINE | 9064 | 1 | 0 | 0 |

STATE:NEBRASKA BUSINESS ACTIVITY:M - MID-LEVEL PRACTITIONERS

| DRUG NAME | DRUG CODE | BUYERS | TOTAL GRAMS | AVG GRAMS |
|---|---|---|---|---|
| CODEINE | 9050 | 2 | 32.49 | 16.24 |
| OXYCODONE | 9143 | 1 | 0.45 | 0.45 |
| HYDROMORPHONE | 9150 | 7 | 0.74 | 0.11 |
| MORPHINE | 9300 | 20 | 13.66 | 0.68 |
| FENTANYL BASE | 9801 | 28 | 0.22 | 0.01 |

STATE:NEBRASKA BUSINESS ACTIVITY:N-U NARCOTIC TREATMENT PROGRAMS

| DRUG NAME | DRUG CODE | BUYERS | TOTAL GRAMS | AVG GRAMS |
|---|---|---|---|---|
| METHADONE | 9250B | 3 | 19,784.13 | 6,594.71 |

STATE:NEVADA BUSINESS ACTIVITY:A - PHARMACIES

| DRUG NAME | DRUG CODE | BUYERS | TOTAL GRAMS | AVG GRAMS |
|---|---|---|---|---|
| AMPHETAMINE | 1100 | 461 | 93,146.95 | 202.05 |
| DL-METHAMPHETAMINE RACEMIC BASE | 1105B | 5 | 14.06 | 2.81 |
| D-METHAMPHETAMINE | 1105D | 26 | 85.16 | 3.28 |
| LISDEXAMFETAMINE | 1205 | 426 | 27,907.20 | 65.51 |
| METHYLPHENIDATE (DL;D;L;ISOMERS) | 1724 | 451 | 53,799.52 | 119.29 |
| BARBITURIC ACID DERIVIATIVE OR SALT [PER 21C | 2100 | 6 | 45 | 7.5 |
| BUTALBITAL | 2165 | 387 | 34,400 | 88.89 |
| SECOBARBITAL (SCHEDULE 2) | 2315 | 1 | 9.16 | 9.16 |
| COCAINE | 9041L | 2 | 32.14 | 16.07 |
| CODEINE | 9050 | 475 | 112,925.72 | 237.74 |
| BUPRENORPHINE | 9064 | 434 | 11,226.13 | 25.87 |
| DIHYDROCODEINE | 9120 | 17 | 184.79 | 10.87 |
| OXYCODONE | 9143 | 479 | 726,683.62 | 1,517.08 |
| HYDROMORPHONE | 9150 | 451 | 11,728.44 | 26.01 |
| HYDROCODONE | 9193 | 482 | 387,446.64 | 803.83 |
| LEVORPHANOL | 9220L | 9 | 8.47 | 0.94 |
| MEPERIDINE (PETHIDINE) | 9230 | 192 | 4,002.80 | 20.85 |
| METHADONE | 9250B | 453 | 52,801.80 | 116.56 |
| MORPHINE | 9300 | 476 | 247,865.42 | 520.73 |
| OPIUM POWDERED | 9639 | 26 | 42.84 | 1.65 |
| OXYMORPHONE | 9652 | 413 | 25,015.29 | 60.57 |
| SUFENTANIL BASE | 9740 | 1 | 0.01 | 0.01 |
| TAPENTADOL | 9780 | 369 | 50,597 | 137.12 |
| FENTANYL BASE | 9801 | 456 | 2,461.43 | 5.4 |

STATE:NEVADA BUSINESS ACTIVITY:B - HOSPITALS

| DRUG NAME | DRUG CODE | BUYERS | TOTAL GRAMS | AVG GRAMS |
|---|---|---|---|---|
| AMPHETAMINE | 1100 | 36 | 2,950.85 | 81.97 |
| LISDEXAMFETAMINE | 1205 | 13 | 650.36 | 50.03 |
| METHYLPHENIDATE (DL;D;L;ISOMERS) | 1724 | 50 | 4,085.54 | 81.71 |
| AMOBARBITAL (SCHEDULE 2) | 2125 | 1 | 3.19 | 3.19 |
| BUTALBITAL | 2165 | 7 | 165 | 23.57 |
| PENTOBARBITAL (SCHEDULE 2) | 2270 | 8 | 1,164.77 | 145.6 |
| COCAINE | 9041L | 30 | 183.99 | 6.13 |
| CODEINE | 9050 | 59 | 4,126.08 | 69.93 |
| BUPRENORPHINE | 9064 | 26 | 698.08 | 26.85 |
| OXYCODONE | 9143 | 94 | 47,656.95 | 506.99 |
| HYDROMORPHONE | 9150 | 82 | 2,394.63 | 29.2 |
| HYDROCODONE | 9193 | 89 | 30,598.48 | 343.8 |
| MEPERIDINE (PETHIDINE) | 9230 | 73 | 2,057.85 | 28.19 |
| METHADONE | 9250B | 57 | 4,186.53 | 73.45 |
| MORPHINE | 9300 | 99 | 35,421.01 | 357.79 |
| OPIUM POWDERED | 9639 | 26 | 86.4 | 3.32 |
| OXYMORPHONE | 9652 | 10 | 102.95 | 10.29 |
| ALFENTANIL | 9737 | 22 | 6.97 | 0.32 |
| REMIFENTANIL | 9739 | 18 | 3.39 | 0.19 |
| SUFENTANIL BASE | 9740 | 21 | 0.36 | 0.02 |
| TAPENTADOL | 9780 | 10 | 760 | 76 |
| FENTANYL BASE | 9801 | 112 | 479.79 | 4.28 |

STATE:NEVADA BUSINESS ACTIVITY:C - PRACTITIONERS

| DRUG NAME | DRUG CODE | BUYERS | TOTAL GRAMS | AVG GRAMS |
|---|---|---|---|---|
| AMPHETAMINE | 1100 | 2 | 47.61 | 23.8 |
| LISDEXAMFETAMINE | 1205 | 1 | 22.53 | 22.53 |
| METHYLPHENIDATE (DL;D;L;ISOMERS) | 1724 | 1 | 27.45 | 27.45 |

| | | | | |
|---|---|---|---|---|
| BUTALBITAL | 2165 | 1 | 165 | 165 |
| PENTOBARBITAL (SCHEDULE 2) | 2270 | 44 | 52,881.08 | 1,201.84 |
| COCAINE | 9041L | 2 | 1 | 0.5 |
| CODEINE | 9050 | 19 | 680.49 | 35.82 |
| BUPRENORPHINE | 9064 | 182 | 266.68 | 1.47 |
| DIHYDROCODEINE | 9120 | 58 | 13.96 | 0.24 |
| OXYCODONE | 9143 | 25 | 3,569.79 | 142.79 |
| HYDROMORPHONE | 9150 | 146 | 175.23 | 1.2 |
| HYDROCODONE | 9193 | 127 | 879.23 | 6.92 |
| MEPERIDINE (PETHIDINE) | 9230 | 70 | 987.19 | 14.1 |
| METHADONE | 9250B | 11 | 202.22 | 18.38 |
| MORPHINE | 9300 | 83 | 546.06 | 6.58 |
| OXYMORPHONE | 9652 | 4 | 9.07 | 2.27 |
| NOROXYMORPHONE | 9668 | 1 | 43.13 | 43.13 |
| ALFENTANIL | 9737 | 1 | 0.1 | 0.1 |
| REMIFENTANIL | 9739 | 4 | 0.14 | 0.04 |
| FENTANYL BASE | 9801 | 173 | 26.98 | 0.16 |

STATE:NEVADA BUSINESS ACTIVITY:D - TEACHING INSTITUTIONS

| DRUG NAME | DRUG CODE | BUYERS | TOTAL GRAMS | AVG GRAMS |
|---|---|---|---|---|
| MORPHINE | 9300 | 1 | 0.19 | 0.19 |
| FENTANYL BASE | 9801 | 1 | 0 | 0 |

STATE:NEVADA BUSINESS ACTIVITY:M - MID-LEVEL PRACTITIONERS

| DRUG NAME | DRUG CODE | BUYERS | TOTAL GRAMS | AVG GRAMS |
|---|---|---|---|---|
| METHYLPHENIDATE (DL;D;L;ISOMERS) | 1724 | 1 | 3.46 | 3.46 |
| BUTALBITAL | 2165 | 2 | 440 | 220 |
| PENTOBARBITAL (SCHEDULE 2) | 2270 | 7 | 7,109.83 | 1,015.69 |
| CODEINE | 9050 | 6 | 261.45 | 43.58 |
| DIHYDROCODEINE | 9120 | 18 | 3.88 | 0.22 |

| | | | | |
|---|---|---|---|---|
| OXYCODONE | 9143 | 2 | 2,483.31 | 1,241.65 |
| HYDROMORPHONE | 9150 | 2 | 39.19 | 19.59 |
| HYDROCODONE | 9193 | 2 | 994.82 | 497.41 |
| METHADONE | 9250B | 2 | 475.37 | 237.69 |
| MORPHINE | 9300 | 2 | 990.38 | 495.19 |
| OXYMORPHONE | 9652 | 2 | 15.52 | 7.76 |
| TAPENTADOL | 9780 | 1 | 110 | 110 |
| FENTANYL BASE | 9801 | 2 | 9.75 | 4.87 |

STATE:NEVADA BUSINESS ACTIVITY:N-U NARCOTIC TREATMENT PROGRAMS

| DRUG NAME | DRUG CODE | BUYERS | TOTAL GRAMS | AVG GRAMS |
|---|---|---|---|---|
| BUPRENORPHINE | 9064 | 6 | 134.39 | 22.4 |
| METHADONE | 9250B | 14 | 96,475.35 | 6,891.10 |

STATE:NEW HAMPSHIRE BUSINESS ACTIVITY:A - PHARMACIES

| DRUG NAME | DRUG CODE | BUYERS | TOTAL GRAMS | AVG GRAMS |
|---|---|---|---|---|
| AMPHETAMINE | 1100 | 273 | 141,303.90 | 517.6 |
| DL-METHAMPHETAMINE RACEMIC BASE | 1105B | 3 | 2.41 | 0.8 |
| D-METHAMPHETAMINE | 1105D | 8 | 26.11 | 3.26 |
| LISDEXAMFETAMINE | 1205 | 271 | 52,790.36 | 194.8 |
| METHYLPHENIDATE (DL;D;L;ISOMERS) | 1724 | 272 | 134,832.49 | 495.71 |
| BARBITURIC ACID DERIVIATIVE OR SALT [PER 21C | 2100 | 5 | 45 | 9 |
| BUTALBITAL | 2165 | 167 | 5,180 | 31.02 |
| SECOBARBITAL (SCHEDULE 2) | 2315 | 1 | 36.62 | 36.62 |
| NABILONE | 7379 | 1 | 0.25 | 0.25 |
| COCAINE | 9041L | 2 | 5.36 | 2.68 |
| CODEINE | 9050 | 271 | 34,716.38 | 128.1 |
| BUPRENORPHINE | 9064 | 269 | 27,304.10 | 101.5 |
| DIHYDROCODEINE | 9120 | 1 | 1.07 | 1.07 |
| OXYCODONE | 9143 | 273 | 278,685.79 | 1,020.83 |

| | | | | |
|---|---|---|---|---|
| HYDROMORPHONE | 9150 | 270 | 7,358 | 27.25 |
| HYDROCODONE | 9193 | 276 | 39,607.34 | 143.5 |
| LEVORPHANOL | 9220L | 9 | 3.37 | 0.37 |
| MEPERIDINE (PETHIDINE) | 9230 | 79 | 2,485.84 | 31.47 |
| METHADONE | 9250B | 258 | 25,606.31 | 99.25 |
| MORPHINE | 9300 | 273 | 73,804.91 | 270.35 |
| OPIUM TINCTURE (LAUDANUM) | 9630 | 1 | 1.18 | 1.18 |
| OPIUM POWDERED | 9639 | 21 | 47.88 | 2.28 |
| OXYMORPHONE | 9652 | 194 | 5,121.55 | 26.4 |
| TAPENTADOL | 9780 | 197 | 25,976.50 | 131.86 |
| FENTANYL BASE | 9801 | 263 | 1,588.16 | 6.04 |

STATE:NEW HAMPSHIRE BUSINESS ACTIVITY:B - HOSPITALS

| DRUG NAME | DRUG CODE | BUYERS | TOTAL GRAMS | AVG GRAMS |
|---|---|---|---|---|
| AMPHETAMINE | 1100 | 22 | 2,645.50 | 120.25 |
| LISDEXAMFETAMINE | 1205 | 11 | 427.22 | 38.84 |
| METHYLPHENIDATE (DL;D;L;ISOMERS) | 1724 | 33 | 2,497.01 | 75.67 |
| AMOBARBITAL (SCHEDULE 2) | 2125 | 1 | 10.94 | 10.94 |
| BUTALBITAL | 2165 | 1 | 10 | 10 |
| PENTOBARBITAL (SCHEDULE 2) | 2270 | 3 | 136.71 | 45.57 |
| COCAINE | 9041L | 27 | 201.7 | 7.47 |
| CODEINE | 9050 | 37 | 663.77 | 17.94 |
| BUPRENORPHINE | 9064 | 23 | 547.29 | 23.8 |
| OXYCODONE | 9143 | 47 | 12,650.33 | 269.16 |
| HYDROMORPHONE | 9150 | 41 | 992.44 | 24.21 |
| HYDROCODONE | 9193 | 42 | 1,741.02 | 41.45 |
| MEPERIDINE (PETHIDINE) | 9230 | 34 | 374.75 | 11.02 |
| METHADONE | 9250B | 33 | 1,574.23 | 47.7 |
| MORPHINE | 9300 | 45 | 5,862.69 | 130.28 |
| OPIUM POWDERED | 9639 | 20 | 101.88 | 5.09 |
| ALFENTANIL | 9737 | 6 | 1.57 | 0.26 |
| REMIFENTANIL | 9739 | 17 | 4.71 | 0.28 |

| | | | | |
|---|---|---|---|---|
| SUFENTANIL BASE | 9740 | 5 | 0.07 | 0.01 |
| TAPENTADOL | 9780 | 3 | 194.5 | 64.83 |
| FENTANYL BASE | 9801 | 48 | 153.14 | 3.19 |

STATE:NEW HAMPSHIRE BUSINESS ACTIVITY:C - PRACTITIONERS

| DRUG NAME | DRUG CODE | BUYERS | TOTAL GRAMS | AVG GRAMS |
|---|---|---|---|---|
| PENTOBARBITAL (SCHEDULE 2) | 2270 | 81 | 62,189.76 | 767.77 |
| COCAINE | 9041L | 2 | 3.43 | 1.71 |
| CODEINE | 9050 | 2 | 50.09 | 25.05 |
| BUPRENORPHINE | 9064 | 175 | 8.57 | 0.05 |
| OXYCODONE | 9143 | 4 | 15.24 | 3.81 |
| HYDROMORPHONE | 9150 | 85 | 34.99 | 0.41 |
| HYDROCODONE | 9193 | 19 | 21.92 | 1.15 |
| MEPERIDINE (PETHIDINE) | 9230 | 10 | 40.2 | 4.02 |
| MORPHINE | 9300 | 64 | 49.8 | 0.78 |
| OXYMORPHONE | 9652 | 5 | 0.13 | 0.03 |
| NOROXYMORPHONE | 9668 | 2 | 2.78 | 1.39 |
| REMIFENTANIL | 9739 | 1 | 0.01 | 0.01 |
| FENTANYL BASE | 9801 | 91 | 14.44 | 0.16 |

STATE:NEW HAMPSHIRE BUSINESS ACTIVITY:D - TEACHING INSTITUTIONS

| DRUG NAME | DRUG CODE | BUYERS | TOTAL GRAMS | AVG GRAMS |
|---|---|---|---|---|
| D-METHAMPHETAMINE | 1105D | 1 | 0.08 | 0.08 |
| METHYLPHENIDATE (DL;D;L;ISOMERS) | 1724 | 1 | 0.09 | 0.09 |

STATE:NEW HAMPSHIRE BUSINESS ACTIVITY:M - MID-LEVEL PRACTITIONERS

| DRUG NAME | DRUG CODE | BUYERS | TOTAL GRAMS | AVG GRAMS |
|---|---|---|---|---|
| CODEINE | 9050 | 1 | 9.95 | 9.95 |

| | | | | |
|---|---|---|---|---|
| HYDROCODONE | 9193 | 1 | 0.61 | 0.61 |
| MORPHINE | 9300 | 1 | 0.02 | 0.02 |
| FENTANYL BASE | 9801 | 1 | 0.01 | 0.01 |

STATE:NEW HAMPSHIRE BUSINESS ACTIVITY:N-U NARCOTIC TREATMENT PROGRAMS

| DRUG NAME | DRUG CODE | BUYERS | TOTAL GRAMS | AVG GRAMS |
|---|---|---|---|---|
| BUPRENORPHINE | 9064 | 5 | 427.55 | 85.51 |
| METHADONE | 9250B | 8 | 93,431.71 | 11,678.96 |

STATE:NEW JERSEY BUSINESS ACTIVITY:A - PHARMACIES

| DRUG NAME | DRUG CODE | BUYERS | TOTAL GRAMS | AVG GRAMS |
|---|---|---|---|---|
| AMPHETAMINE | 1100 | 2,009 | 391,994.39 | 195.12 |
| DL-METHAMPHETAMINE RACEMIC BASE | 1105B | 14 | 33.35 | 2.38 |
| D-METHAMPHETAMINE | 1105D | 49 | 130.95 | 2.67 |
| LISDEXAMFETAMINE | 1205 | 1,825 | 192,658.76 | 105.57 |
| METHYLPHENIDATE (DL;D;L;ISOMERS) | 1724 | 1,986 | 365,660.08 | 184.12 |
| BARBITURIC ACID DERIVIATIVE OR SALT [PER 21C | 2100 | 56 | 1,090 | 19.46 |
| BUTALBITAL | 2165 | 1,077 | 47,495 | 44.1 |
| SECOBARBITAL (SCHEDULE 2) | 2315 | 15 | 320.43 | 21.36 |
| NABILONE | 7379 | 3 | 0.15 | 0.05 |
| COCAINE | 9041L | 9 | 129.88 | 14.43 |
| CODEINE | 9050 | 2,035 | 336,204.52 | 165.21 |
| BUPRENORPHINE | 9064 | 1,858 | 98,916.86 | 53.24 |
| DIHYDROCODEINE | 9120 | 62 | 472.13 | 7.61 |
| OXYCODONE | 9143 | 2,070 | 2,313,888.60 | 1,117.82 |
| HYDROMORPHONE | 9150 | 1,832 | 36,015.64 | 19.66 |
| HYDROCODONE | 9193 | 2,023 | 164,078.13 | 81.11 |
| LEVORPHANOL | 9220L | 14 | 594.68 | 42.48 |
| MEPERIDINE (PETHIDINE) | 9230 | 312 | 7,901.89 | 25.33 |
| METHADONE | 9250B | 1,244 | 53,080.67 | 42.67 |

| | | | | |
|---|---|---|---|---|
| MORPHINE | 9300 | 1,869 | 378,704.55 | 202.62 |
| OPIUM TINCTURE (LAUDANUM) | 9630 | 6 | 9.44 | 1.57 |
| OPIUM POWDERED | 9639 | 99 | 439.2 | 4.44 |
| OXYMORPHONE | 9652 | 1,173 | 38,623.96 | 32.93 |
| ALFENTANIL | 9737 | 2 | 0.51 | 0.25 |
| REMIFENTANIL | 9739 | 4 | 0.35 | 0.09 |
| SUFENTANIL BASE | 9740 | 2 | 3.43 | 1.72 |
| TAPENTADOL | 9780 | 1,464 | 200,886.50 | 137.22 |
| FENTANYL BASE | 9801 | 1,883 | 10,513.11 | 5.58 |

STATE:NEW JERSEY BUSINESS ACTIVITY:B - HOSPITALS

| DRUG NAME | DRUG CODE | BUYERS | TOTAL GRAMS | AVG GRAMS |
|---|---|---|---|---|
| AMPHETAMINE | 1100 | 49 | 3,118.62 | 63.65 |
| LISDEXAMFETAMINE | 1205 | 26 | 1,438.89 | 55.34 |
| METHYLPHENIDATE (DL;D;L;ISOMERS) | 1724 | 96 | 3,641.19 | 37.93 |
| AMOBARBITAL (SCHEDULE 2) | 2125 | 10 | 40.1 | 4.01 |
| BUTALBITAL | 2165 | 7 | 325 | 46.43 |
| PENTOBARBITAL (SCHEDULE 2) | 2270 | 14 | 1,090.03 | 77.86 |
| COCAINE | 9041L | 66 | 378.62 | 5.74 |
| CODEINE | 9050 | 112 | 5,187.31 | 46.32 |
| BUPRENORPHINE | 9064 | 82 | 742.86 | 9.06 |
| DIHYDROCODEINE | 9120 | 1 | 1.07 | 1.07 |
| OXYCODONE | 9143 | 126 | 41,725.10 | 331.15 |
| HYDROMORPHONE | 9150 | 114 | 5,586.32 | 49 |
| HYDROCODONE | 9193 | 93 | 1,640.88 | 17.64 |
| MEPERIDINE (PETHIDINE) | 9230 | 82 | 1,290.91 | 15.74 |
| METHADONE | 9250B | 104 | 3,386.55 | 32.56 |
| MORPHINE | 9300 | 121 | 17,411.67 | 143.9 |
| OPIUM POWDERED | 9639 | 42 | 124.56 | 2.97 |
| OXYMORPHONE | 9652 | 7 | 59.77 | 8.54 |
| ALFENTANIL | 9737 | 4 | 0.43 | 0.11 |
| REMIFENTANIL | 9739 | 49 | 47.69 | 0.97 |

| | | | | |
|---|---|---|---|---|
| SUFENTANIL BASE | 9740 | 24 | 0.99 | 0.04 |
| TAPENTADOL | 9780 | 33 | 2,565.50 | 77.74 |
| FENTANYL BASE | 9801 | 121 | 901.17 | 7.45 |

STATE:NEW JERSEY BUSINESS ACTIVITY:C - PRACTITIONERS

| DRUG NAME | DRUG CODE | BUYERS | TOTAL GRAMS | AVG GRAMS |
|---|---|---|---|---|
| AMPHETAMINE | 1100 | 2 | 5.28 | 2.64 |
| PENTOBARBITAL (SCHEDULE 2) | 2270 | 80 | 71,437.07 | 892.96 |
| COCAINE | 9041L | 44 | 188.77 | 4.29 |
| CODEINE | 9050 | 285 | 6,344.19 | 22.26 |
| BUPRENORPHINE | 9064 | 543 | 403.84 | 0.74 |
| DIHYDROCODEINE | 9120 | 79 | 42.73 | 0.54 |
| OXYCODONE | 9143 | 114 | 348.34 | 3.06 |
| HYDROMORPHONE | 9150 | 286 | 123.41 | 0.43 |
| HYDROCODONE | 9193 | 262 | 867.53 | 3.31 |
| MEPERIDINE (PETHIDINE) | 9230 | 132 | 357.75 | 2.71 |
| METHADONE | 9250B | 17 | 67.44 | 3.97 |
| MORPHINE | 9300 | 203 | 145.37 | 0.72 |
| OPIUM POWDERED | 9639 | 4 | 2.52 | 0.63 |
| OXYMORPHONE | 9652 | 6 | 1.78 | 0.3 |
| NOROXYMORPHONE | 9668 | 2 | 25.6 | 12.8 |
| ALFENTANIL | 9737 | 2 | 0.18 | 0.09 |
| REMIFENTANIL | 9739 | 22 | 1.58 | 0.07 |
| SUFENTANIL BASE | 9740 | 2 | 0.01 | 0 |
| TAPENTADOL | 9780 | 2 | 50 | 25 |
| FENTANYL BASE | 9801 | 495 | 80.55 | 0.16 |

STATE:NEW JERSEY BUSINESS ACTIVITY:D - TEACHING INSTITUTIONS

| DRUG NAME | DRUG CODE | BUYERS | TOTAL GRAMS | AVG GRAMS |
|---|---|---|---|---|
| BUPRENORPHINE | 9064 | 3 | 0.02 | 0.01 |

| | | | | |
|---|---|---|---|---|
| MORPHINE | 9300 | 1 | 0.19 | 0.19 |
| FENTANYL BASE | 9801 | 1 | 0.03 | 0.03 |

STATE:NEW JERSEY BUSINESS ACTIVITY:M - MID-LEVEL PRACTITIONERS

| DRUG NAME | DRUG CODE | BUYERS | TOTAL GRAMS | AVG GRAMS |
|---|---|---|---|---|
| PENTOBARBITAL (SCHEDULE 2) | 2270 | 15 | 10,131.30 | 675.42 |
| CODEINE | 9050 | 25 | 3,143.32 | 125.73 |
| DIHYDROCODEINE | 9120 | 4 | 1.06 | 0.26 |

STATE:NEW JERSEY BUSINESS ACTIVITY:N-U NARCOTIC TREATMENT PROGRAMS

| DRUG NAME | DRUG CODE | BUYERS | TOTAL GRAMS | AVG GRAMS |
|---|---|---|---|---|
| BUPRENORPHINE | 9064 | 13 | 601.75 | 46.29 |
| METHADONE | 9250B | 31 | 379,650.44 | 12,246.79 |

STATE:NEW MEXICO BUSINESS ACTIVITY:A - PHARMACIES

| DRUG NAME | DRUG CODE | BUYERS | TOTAL GRAMS | AVG GRAMS |
|---|---|---|---|---|
| AMPHETAMINE | 1100 | 305 | 74,589.59 | 244.56 |
| DL-METHAMPHETAMINE RACEMIC BASE | 1105B | 16 | 51.84 | 3.24 |
| D-METHAMPHETAMINE | 1105D | 24 | 63.87 | 2.66 |
| LISDEXAMFETAMINE | 1205 | 295 | 26,853.90 | 91.03 |
| METHYLPHENIDATE (DL;D;L;ISOMERS) | 1724 | 314 | 88,346.16 | 281.36 |
| BARBITURIC ACID DERIVIATIVE OR SALT [PER 21C | 2100 | 1 | 10 | 10 |
| BUTALBITAL | 2165 | 203 | 8,815 | 43.42 |
| SECOBARBITAL (SCHEDULE 2) | 2315 | 2 | 36.62 | 18.31 |
| NABILONE | 7379 | 1 | 0.3 | 0.3 |
| COCAINE | 9041L | 1 | 0.14 | 0.14 |
| CODEINE | 9050 | 315 | 71,757.11 | 227.8 |
| BUPRENORPHINE | 9064 | 298 | 23,602.97 | 79.2 |

| | | | | |
|---|---|---|---|---|
| DIHYDROCODEINE | 9120 | 27 | 616.33 | 22.83 |
| OXYCODONE | 9143 | 316 | 413,497.74 | 1,308.54 |
| HYDROMORPHONE | 9150 | 285 | 6,395.91 | 22.44 |
| HYDROCODONE | 9193 | 318 | 147,266.34 | 463.1 |
| LEVORPHANOL | 9220L | 1 | 0.12 | 0.12 |
| MEPERIDINE (PETHIDINE) | 9230 | 197 | 6,441.69 | 32.7 |
| METHADONE | 9250B | 268 | 11,468.98 | 42.79 |
| MORPHINE | 9300 | 312 | 113,241.13 | 362.95 |
| OPIUM TINCTURE (LAUDANUM) | 9630 | 1 | 4.73 | 4.73 |
| OPIUM POWDERED | 9639 | 19 | 108.72 | 5.72 |
| OXYMORPHONE | 9652 | 209 | 6,048.71 | 28.94 |
| TAPENTADOL | 9780 | 195 | 17,686 | 90.7 |
| FENTANYL BASE | 9801 | 302 | 1,842.05 | 6.1 |

STATE:NEW MEXICO BUSINESS ACTIVITY:B - HOSPITALS

| DRUG NAME | DRUG CODE | BUYERS | TOTAL GRAMS | AVG GRAMS |
|---|---|---|---|---|
| AMPHETAMINE | 1100 | 43 | 2,526.40 | 58.75 |
| LISDEXAMFETAMINE | 1205 | 8 | 381.55 | 47.69 |
| METHYLPHENIDATE (DL;D;L;ISOMERS) | 1724 | 61 | 6,962.74 | 114.14 |
| AMOBARBITAL (SCHEDULE 2) | 2125 | 1 | 2.73 | 2.73 |
| BUTALBITAL | 2165 | 3 | 20 | 6.67 |
| PENTOBARBITAL (SCHEDULE 2) | 2270 | 3 | 191.39 | 63.8 |
| COCAINE | 9041L | 25 | 312.8 | 12.51 |
| CODEINE | 9050 | 84 | 12,529.19 | 149.16 |
| BUPRENORPHINE | 9064 | 43 | 1,648.09 | 38.33 |
| OXYCODONE | 9143 | 87 | 29,507.21 | 339.16 |
| HYDROMORPHONE | 9150 | 63 | 820.31 | 13.02 |
| HYDROCODONE | 9193 | 94 | 14,502.20 | 154.28 |
| MEPERIDINE (PETHIDINE) | 9230 | 57 | 993.27 | 17.43 |
| METHADONE | 9250B | 45 | 2,744.56 | 60.99 |
| MORPHINE | 9300 | 102 | 20,030.29 | 196.38 |
| OPIUM POWDERED | 9639 | 23 | 118.08 | 5.13 |

| | | | | |
|---|---|---|---|---|
| OXYMORPHONE | 9652 | 5 | 15 | 3 |
| ALFENTANIL | 9737 | 8 | 4.78 | 0.6 |
| REMIFENTANIL | 9739 | 17 | 4.16 | 0.24 |
| SUFENTANIL BASE | 9740 | 14 | 0.17 | 0.01 |
| TAPENTADOL | 9780 | 3 | 41 | 13.67 |
| FENTANYL BASE | 9801 | 100 | 381.63 | 3.82 |

STATE:NEW MEXICO BUSINESS ACTIVITY:C - PRACTITIONERS

| DRUG NAME | DRUG CODE | BUYERS | TOTAL GRAMS | AVG GRAMS |
|---|---|---|---|---|
| PENTOBARBITAL (SCHEDULE 2) | 2270 | 65 | 56,259.35 | 865.53 |
| CODEINE | 9050 | 11 | 24.48 | 2.23 |
| BUPRENORPHINE | 9064 | 130 | 6.36 | 0.05 |
| OXYCODONE | 9143 | 5 | 195.91 | 39.18 |
| HYDROMORPHONE | 9150 | 76 | 78.07 | 1.03 |
| HYDROCODONE | 9193 | 44 | 147.72 | 3.36 |
| MEPERIDINE (PETHIDINE) | 9230 | 8 | 43.58 | 5.45 |
| METHADONE | 9250B | 4 | 4.02 | 1.01 |
| MORPHINE | 9300 | 50 | 218.31 | 4.37 |
| OXYMORPHONE | 9652 | 5 | 0.25 | 0.05 |
| NOROXYMORPHONE | 9668 | 1 | 3.24 | 3.24 |
| REMIFENTANIL | 9739 | 1 | 0.02 | 0.02 |
| FENTANYL BASE | 9801 | 96 | 18.34 | 0.19 |

STATE:NEW MEXICO BUSINESS ACTIVITY:D - TEACHING INSTITUTIONS

| DRUG NAME | DRUG CODE | BUYERS | TOTAL GRAMS | AVG GRAMS |
|---|---|---|---|---|
| BUPRENORPHINE | 9064 | 1 | 0.02 | 0.02 |

STATE:NEW MEXICO BUSINESS ACTIVITY:M - MID-LEVEL PRACTITIONERS

| DRUG NAME | DRUG CODE | BUYERS | TOTAL GRAMS | AVG GRAMS |
|---|---|---|---|---|
| PENTOBARBITAL (SCHEDULE 2) | 2270 | 9 | 12,453.37 | 1,383.71 |
| CODEINE | 9050 | 5 | 13.28 | 2.66 |
| BUPRENORPHINE | 9064 | 2 | 0.63 | 0.31 |
| HYDROMORPHONE | 9150 | 10 | 14.94 | 1.49 |
| HYDROCODONE | 9193 | 3 | 8.14 | 2.71 |
| MEPERIDINE (PETHIDINE) | 9230 | 1 | 2.18 | 2.18 |
| MORPHINE | 9300 | 43 | 40.24 | 0.94 |
| OXYMORPHONE | 9652 | 1 | 0.13 | 0.13 |
| FENTANYL BASE | 9801 | 60 | 1.26 | 0.02 |

STATE:NEW MEXICO BUSINESS ACTIVITY:N-U NARCOTIC TREATMENT PROGRAMS

| DRUG NAME | DRUG CODE | BUYERS | TOTAL GRAMS | AVG GRAMS |
|---|---|---|---|---|
| BUPRENORPHINE | 9064 | 3 | 168.19 | 56.06 |
| METHADONE | 9250B | 15 | 138,801.94 | 9,253.46 |

STATE:NEW YORK BUSINESS ACTIVITY:A - PHARMACIES

| DRUG NAME | DRUG CODE | BUYERS | TOTAL GRAMS | AVG GRAMS |
|---|---|---|---|---|
| AMPHETAMINE | 1100 | 4,664 | 893,824.44 | 191.64 |
| DL-METHAMPHETAMINE RACEMIC BASE | 1105B | 23 | 62.69 | 2.73 |
| D-METHAMPHETAMINE | 1105D | 81 | 293.24 | 3.62 |
| LISDEXAMFETAMINE | 1205 | 3,875 | 323,568.35 | 83.5 |
| METHYLPHENIDATE (DL;D;L;ISOMERS) | 1724 | 4,593 | 919,510.10 | 200.2 |
| BARBITURIC ACID DERIVIATIVE OR SALT [PER 21C | 2100 | 52 | 705 | 13.56 |
| BUTALBITAL | 2165 | 1,540 | 48,415 | 31.44 |
| SECOBARBITAL (SCHEDULE 2) | 2315 | 47 | 1,867.62 | 39.74 |
| NABILONE | 7379 | 6 | 0.8 | 0.13 |
| COCAINE | 9041L | 5 | 29.43 | 5.89 |
| CODEINE | 9050 | 4,920 | 645,139.91 | 131.13 |
| BUPRENORPHINE | 9064 | 4,008 | 187,343.39 | 46.74 |

| | | | | |
|---|---|---|---|---|
| DIHYDROCODEINE | 9120 | 37 | 198.68 | 5.37 |
| OXYCODONE | 9143 | 5,022 | 3,335,189.79 | 664.12 |
| HYDROMORPHONE | 9150 | 4,078 | 73,707.39 | 18.07 |
| HYDROCODONE | 9193 | 4,861 | 759,132.36 | 156.17 |
| LEVORPHANOL | 9220L | 96 | 130.13 | 1.36 |
| MEPERIDINE (PETHIDINE) | 9230 | 547 | 16,209.25 | 29.63 |
| METHADONE | 9250B | 3,253 | 164,571.84 | 50.59 |
| MORPHINE | 9300 | 4,235 | 658,272.27 | 155.44 |
| OPIUM TINCTURE (LAUDANUM) | 9630 | 3 | 3.54 | 1.18 |
| OPIUM POWDERED | 9639 | 147 | 365.76 | 2.49 |
| OXYMORPHONE | 9652 | 2,453 | 100,250.32 | 40.87 |
| REMIFENTANIL | 9739 | 2 | 0.07 | 0.04 |
| SUFENTANIL BASE | 9740 | 2 | 0.41 | 0.2 |
| TAPENTADOL | 9780 | 2,951 | 423,081 | 143.37 |
| FENTANYL BASE | 9801 | 4,317 | 18,997.34 | 4.4 |

STATE:NEW YORK BUSINESS ACTIVITY:B - HOSPITALS

| DRUG NAME | DRUG CODE | BUYERS | TOTAL GRAMS | AVG GRAMS |
|---|---|---|---|---|
| AMPHETAMINE | 1100 | 178 | 8,712.57 | 48.95 |
| LISDEXAMFETAMINE | 1205 | 77 | 1,439.32 | 18.69 |
| METHYLPHENIDATE (DL;D;L;ISOMERS) | 1724 | 253 | 12,935.33 | 51.13 |
| AMOBARBITAL (SCHEDULE 2) | 2125 | 24 | 152.2 | 6.34 |
| BUTALBITAL | 2165 | 3 | 110 | 36.67 |
| PENTOBARBITAL (SCHEDULE 2) | 2270 | 30 | 16,393.35 | 546.45 |
| NABILONE | 7379 | 1 | 0.35 | 0.35 |
| COCAINE | 9041L | 154 | 1,220.21 | 7.92 |
| CODEINE | 9050 | 298 | 18,182.41 | 61.01 |
| BUPRENORPHINE | 9064 | 221 | 4,790.66 | 21.68 |
| OXYCODONE | 9143 | 333 | 109,821.74 | 329.8 |
| HYDROMORPHONE | 9150 | 289 | 16,660.71 | 57.65 |
| HYDROCODONE | 9193 | 255 | 32,629.93 | 127.96 |
| LEVORPHANOL | 9220L | 1 | 5.8 | 5.8 |

| | | | | |
|---|---|---|---|---|
| MEPERIDINE (PETHIDINE) | 9230 | 205 | 4,602.70 | 22.45 |
| METHADONE | 9250B | 261 | 26,274.59 | 100.67 |
| MORPHINE | 9300 | 328 | 71,715.17 | 218.64 |
| OPIUM POWDERED | 9639 | 83 | 518.4 | 6.25 |
| OXYMORPHONE | 9652 | 46 | 611.17 | 13.29 |
| NOROXYMORPHONE | 9668 | 1 | 1.6 | 1.6 |
| ALFENTANIL | 9737 | 26 | 9.85 | 0.38 |
| REMIFENTANIL | 9739 | 112 | 138.78 | 1.24 |
| SUFENTANIL BASE | 9740 | 44 | 6.79 | 0.15 |
| TAPENTADOL | 9780 | 84 | 8,193.50 | 97.54 |
| FENTANYL BASE | 9801 | 297 | 2,760.30 | 9.29 |

STATE:NEW YORK BUSINESS ACTIVITY:C - PRACTITIONERS

| DRUG NAME | DRUG CODE | BUYERS | TOTAL GRAMS | AVG GRAMS |
|---|---|---|---|---|
| AMPHETAMINE | 1100 | 3 | 11.58 | 3.86 |
| D-METHAMPHETAMINE | 1105D | 2 | 4.1 | 2.05 |
| METHYLPHENIDATE (DL;D;L;ISOMERS) | 1724 | 2 | 21.96 | 10.98 |
| GAMMA HYDROXYBUTYRIC ACID(FDA APPROVED | 2012 | 1 | 12,928.90 | 12,928.90 |
| BARBITURIC ACID DERIVIATIVE OR SALT [PER 21C | 2100 | 1 | 2 | 2 |
| PENTOBARBITAL (SCHEDULE 2) | 2270 | 288 | 255,445.73 | 886.96 |
| COCAINE | 9041L | 53 | 224.41 | 4.23 |
| CODEINE | 9050 | 133 | 3,023.02 | 22.73 |
| BUPRENORPHINE | 9064 | 1,105 | 159.84 | 0.14 |
| DIHYDROCODEINE | 9120 | 10 | 2.64 | 0.26 |
| OXYCODONE | 9143 | 174 | 1,074.08 | 6.17 |
| HYDROMORPHONE | 9150 | 587 | 277.7 | 0.47 |
| HYDROCODONE | 9193 | 456 | 1,344.12 | 2.95 |
| MEPERIDINE (PETHIDINE) | 9230 | 217 | 1,468.80 | 6.77 |
| METHADONE | 9250B | 28 | 89.31 | 3.19 |
| MORPHINE | 9300 | 430 | 567.8 | 1.32 |
| OPIUM POWDERED | 9639 | 1 | 12.24 | 12.24 |
| OXYMORPHONE | 9652 | 11 | 3.1 | 0.28 |

| | | | | |
|---|---|---|---|---|
| NOROXYMORPHONE | 9668 | 12 | 43.68 | 3.64 |
| ALFENTANIL | 9737 | 4 | 1.39 | 0.35 |
| REMIFENTANIL | 9739 | 31 | 2.32 | 0.07 |
| SUFENTANIL BASE | 9740 | 4 | 0.01 | 0 |
| FENTANYL BASE | 9801 | 1,145 | 168.49 | 0.15 |

STATE:NEW YORK BUSINESS ACTIVITY:D - TEACHING INSTITUTIONS

| DRUG NAME | DRUG CODE | BUYERS | TOTAL GRAMS | AVG GRAMS |
|---|---|---|---|---|
| PENTOBARBITAL (SCHEDULE 2) | 2270 | 1 | 88.86 | 88.86 |
| BUPRENORPHINE | 9064 | 7 | 0.04 | 0.01 |
| HYDROMORPHONE | 9150 | 1 | 0.04 | 0.04 |
| MORPHINE | 9300 | 2 | 1.03 | 0.52 |

STATE:NEW YORK BUSINESS ACTIVITY:M - MID-LEVEL PRACTITIONERS

| DRUG NAME | DRUG CODE | BUYERS | TOTAL GRAMS | AVG GRAMS |
|---|---|---|---|---|
| PENTOBARBITAL (SCHEDULE 2) | 2270 | 8 | 11,120.34 | 1,390.04 |
| CODEINE | 9050 | 3 | 86.86 | 28.95 |
| BUPRENORPHINE | 9064 | 2 | 0.52 | 0.26 |
| DIHYDROCODEINE | 9120 | 1 | 0.27 | 0.27 |
| HYDROCODONE | 9193 | 1 | 0.36 | 0.36 |
| FENTANYL BASE | 9801 | 1 | 0.13 | 0.13 |

STATE:NEW YORK BUSINESS ACTIVITY:N-U NARCOTIC TREATMENT PROGRAMS

| DRUG NAME | DRUG CODE | BUYERS | TOTAL GRAMS | AVG GRAMS |
|---|---|---|---|---|
| BUPRENORPHINE | 9064 | 28 | 1,611.91 | 57.57 |
| METHADONE | 9250B | 104 | 963,328.81 | 9,262.78 |

STATISTICAL SUMMARY FOR RETAIL DRUG PURCHASES BY GRAMS WT

STATE:NORTH CAROLINA BUSINESS ACTIVITY:A - PHARMACIES

| DRUG NAME | DRUG CODE | BUYERS | TOTAL GRAMS | AVG GRAMS |
|---|---|---|---|---|
| AMPHETAMINE | 1100 | 2,128 | 735,634.42 | 345.69 |
| DL-METHAMPHETAMINE RACEMIC BASE | 1105B | 13 | 53.45 | 4.11 |
| D-METHAMPHETAMINE | 1105D | 59 | 192.01 | 3.25 |
| LISDEXAMFETAMINE | 1205 | 2,066 | 388,125.99 | 187.86 |
| METHYLPHENIDATE (DL;D;L;ISOMERS) | 1724 | 2,122 | 745,235.79 | 351.19 |
| BARBITURIC ACID DERIVIATIVE OR SALT [PER 21C | 2100 | 25 | 300 | 12 |
| BUTALBITAL | 2165 | 1,176 | 37,910 | 32.24 |
| SECOBARBITAL (SCHEDULE 2) | 2315 | 5 | 82.4 | 16.48 |
| NABILONE | 7379 | 1 | 0.55 | 0.55 |
| COCAINE | 9041L | 10 | 81.89 | 8.19 |
| CODEINE | 9050 | 2,082 | 195,209.32 | 93.76 |
| BUPRENORPHINE | 9064 | 1,972 | 95,082.77 | 48.22 |
| DIHYDROCODEINE | 9120 | 17 | 36.32 | 2.14 |
| OXYCODONE | 9143 | 2,147 | 2,404,512.62 | 1,119.94 |
| HYDROMORPHONE | 9150 | 2,037 | 40,617.02 | 19.94 |
| HYDROCODONE | 9193 | 2,160 | 815,317.08 | 377.46 |
| LEVORPHANOL | 9220L | 195 | 111.67 | 0.57 |
| MEPERIDINE (PETHIDINE) | 9230 | 1,177 | 25,514.25 | 21.68 |
| METHADONE | 9250B | 1,928 | 125,873.74 | 65.29 |
| MORPHINE | 9300 | 2,106 | 490,056.16 | 232.7 |
| OPIUM POWDERED | 9639 | 167 | 268.2 | 1.61 |
| OXYMORPHONE | 9652 | 1,818 | 133,559.17 | 73.46 |
| ALFENTANIL | 9737 | 3 | 0.37 | 0.12 |
| REMIFENTANIL | 9739 | 5 | 0.16 | 0.03 |
| SUFENTANIL BASE | 9740 | 1 | 0.8 | 0.8 |
| TAPENTADOL | 9780 | 1,860 | 399,415.50 | 214.74 |
| FENTANYL BASE | 9801 | 2,070 | 12,689.75 | 6.13 |

STATE:NORTH CAROLINA BUSINESS ACTIVITY:B - HOSPITALS

STATISTICAL SUMMARY FOR RETAIL DRUG PURCHASES BY GRAMS WT

| DRUG NAME | DRUG CODE | BUYERS | TOTAL GRAMS | AVG GRAMS |
|---|---|---|---|---|
| AMPHETAMINE | 1100 | 136 | 27,927.94 | 205.35 |
| D-METHAMPHETAMINE | 1105D | 1 | 1.61 | 1.61 |
| LISDEXAMFETAMINE | 1205 | 81 | 8,658.20 | 106.89 |
| METHYLPHENIDATE (DL;D;L;ISOMERS) | 1724 | 169 | 32,267.48 | 190.93 |
| GAMMA HYDROXYBUTYRIC ACID(FDA APPROVED | 2012 | 1 | 2,229.12 | 2,229.12 |
| AMOBARBITAL (SCHEDULE 2) | 2125 | 5 | 40.1 | 8.02 |
| BUTALBITAL | 2165 | 13 | 255 | 19.62 |
| PENTOBARBITAL (SCHEDULE 2) | 2270 | 35 | 43,358.83 | 1,238.82 |
| COCAINE | 9041L | 110 | 542.25 | 4.93 |
| CODEINE | 9050 | 184 | 12,603.41 | 68.5 |
| BUPRENORPHINE | 9064 | 200 | 2,332.81 | 11.66 |
| OXYCODONE | 9143 | 254 | 124,771.79 | 491.23 |
| HYDROMORPHONE | 9150 | 298 | 8,817.50 | 29.59 |
| HYDROCODONE | 9193 | 290 | 56,907.24 | 196.23 |
| LEVORPHANOL | 9220L | 3 | 0.81 | 0.27 |
| MEPERIDINE (PETHIDINE) | 9230 | 187 | 3,478.07 | 18.6 |
| METHADONE | 9250B | 169 | 10,489.75 | 62.07 |
| MORPHINE | 9300 | 312 | 61,611.73 | 197.47 |
| OPIUM POWDERED | 9639 | 101 | 659.88 | 6.53 |
| OXYMORPHONE | 9652 | 81 | 1,656.31 | 20.45 |
| NOROXYMORPHONE | 9668 | 1 | 0.55 | 0.55 |
| ALFENTANIL | 9737 | 25 | 11.46 | 0.46 |
| REMIFENTANIL | 9739 | 75 | 51.95 | 0.69 |
| SUFENTANIL BASE | 9740 | 49 | 1.42 | 0.03 |
| TAPENTADOL | 9780 | 80 | 13,432.25 | 167.9 |
| FENTANYL BASE | 9801 | 312 | 1,485.68 | 4.76 |

STATE:NORTH CAROLINA BUSINESS ACTIVITY:C - PRACTITIONERS

| DRUG NAME | DRUG CODE | BUYERS | TOTAL GRAMS | AVG GRAMS |
|---|---|---|---|---|
| AMPHETAMINE | 1100 | 1 | 0.63 | 0.63 |
| LISDEXAMFETAMINE | 1205 | 1 | 9.65 | 9.65 |

| | | | | |
|---|---|---|---|---|
| METHYLPHENIDATE (DL;D;L;ISOMERS) | 1724 | 1 | 35.32 | 35.32 |
| GAMMA HYDROXYBUTYRIC ACID(FDA APPROVED | 2012 | 2 | 2,229.12 | 1,114.56 |
| PENTOBARBITAL (SCHEDULE 2) | 2270 | 159 | 117,059.41 | 736.22 |
| COCAINE | 9041L | 3 | 5.71 | 1.9 |
| CODEINE | 9050 | 54 | 345.59 | 6.4 |
| BUPRENORPHINE | 9064 | 909 | 48.32 | 0.05 |
| DIHYDROCODEINE | 9120 | 1 | 0.21 | 0.21 |
| OXYCODONE | 9143 | 19 | 1,814.17 | 95.48 |
| HYDROMORPHONE | 9150 | 377 | 185.56 | 0.49 |
| HYDROCODONE | 9193 | 277 | 634.95 | 2.29 |
| MEPERIDINE (PETHIDINE) | 9230 | 108 | 512.66 | 4.75 |
| METHADONE | 9250B | 4 | 17.89 | 4.47 |
| MORPHINE | 9300 | 203 | 766.48 | 3.78 |
| OXYMORPHONE | 9652 | 7 | 32.09 | 4.58 |
| NOROXYMORPHONE | 9668 | 1 | 1.69 | 1.69 |
| REMIFENTANIL | 9739 | 24 | 4.6 | 0.19 |
| SUFENTANIL BASE | 9740 | 1 | 0.01 | 0.01 |
| FENTANYL BASE | 9801 | 414 | 41.98 | 0.1 |

STATE:NORTH CAROLINA BUSINESS ACTIVITY:D - TEACHING INSTITUTIONS

| DRUG NAME | DRUG CODE | BUYERS | TOTAL GRAMS | AVG GRAMS |
|---|---|---|---|---|
| PENTOBARBITAL (SCHEDULE 2) | 2270 | 1 | 0.91 | 0.91 |
| BUPRENORPHINE | 9064 | 3 | 0.02 | 0.01 |
| HYDROMORPHONE | 9150 | 2 | 0.11 | 0.05 |
| MORPHINE | 9300 | 1 | 0.19 | 0.19 |
| FENTANYL BASE | 9801 | 1 | 0.03 | 0.03 |

STATE:NORTH CAROLINA BUSINESS ACTIVITY:M - MID-LEVEL PRACTITIONERS

| DRUG NAME | DRUG CODE | BUYERS | TOTAL GRAMS | AVG GRAMS |
|---|---|---|---|---|
| PENTOBARBITAL (SCHEDULE 2) | 2270 | 44 | 78,271.95 | 1,778.91 |

| | | | | |
|---|---|---|---|---|
| CODEINE | 9050 | 41 | 16.24 | 0.4 |
| BUPRENORPHINE | 9064 | 2 | 0.08 | 0.04 |
| HYDROMORPHONE | 9150 | 8 | 1.43 | 0.18 |
| MORPHINE | 9300 | 80 | 56.79 | 0.71 |
| FENTANYL BASE | 9801 | 69 | 0.96 | 0.01 |

STATE:NORTH CAROLINA BUSINESS ACTIVITY:N-U NARCOTIC TREATMENT PROGRAMS

| DRUG NAME | DRUG CODE | BUYERS | TOTAL GRAMS | AVG GRAMS |
|---|---|---|---|---|
| BUPRENORPHINE | 9064 | 37 | 10,387.50 | 280.74 |
| METHADONE | 9250B | 50 | 388,894.51 | 7,777.89 |

STATE:NORTH DAKOTA BUSINESS ACTIVITY:A - PHARMACIES

| DRUG NAME | DRUG CODE | BUYERS | TOTAL GRAMS | AVG GRAMS |
|---|---|---|---|---|
| AMPHETAMINE | 1100 | 169 | 32,813.77 | 194.16 |
| D-METHAMPHETAMINE | 1105D | 3 | 7.23 | 2.41 |
| LISDEXAMFETAMINE | 1205 | 167 | 20,732.98 | 124.15 |
| METHYLPHENIDATE (DL;D;L;ISOMERS) | 1724 | 172 | 54,590.24 | 317.39 |
| BUTALBITAL | 2165 | 62 | 1,640 | 26.45 |
| CODEINE | 9050 | 171 | 20,333.54 | 118.91 |
| BUPRENORPHINE | 9064 | 130 | 3,179.18 | 24.46 |
| DIHYDROCODEINE | 9120 | 1 | 2.14 | 2.14 |
| OXYCODONE | 9143 | 174 | 57,825.72 | 332.33 |
| HYDROMORPHONE | 9150 | 155 | 2,258.65 | 14.57 |
| HYDROCODONE | 9193 | 175 | 38,509.54 | 220.05 |
| MEPERIDINE (PETHIDINE) | 9230 | 51 | 757.33 | 14.85 |
| METHADONE | 9250B | 139 | 4,205.65 | 30.26 |
| MORPHINE | 9300 | 171 | 23,006.21 | 134.54 |
| OPIUM POWDERED | 9639 | 11 | 48.96 | 4.45 |
| OXYMORPHONE | 9652 | 56 | 778.45 | 13.9 |
| TAPENTADOL | 9780 | 105 | 8,661.50 | 82.49 |

| | | | | |
|---|---|---|---|---|
| FENTANYL BASE | 9801 | 170 | 990.92 | 5.83 |

STATE:NORTH DAKOTA BUSINESS ACTIVITY:B - HOSPITALS

| DRUG NAME | DRUG CODE | BUYERS | TOTAL GRAMS | AVG GRAMS |
|---|---|---|---|---|
| AMPHETAMINE | 1100 | 20 | 2,433.95 | 121.7 |
| DL-METHAMPHETAMINE RACEMIC BASE | 1105B | 1 | 4.82 | 4.82 |
| LISDEXAMFETAMINE | 1205 | 16 | 1,415.19 | 88.45 |
| METHYLPHENIDATE (DL;D;L;ISOMERS) | 1724 | 25 | 6,472.53 | 258.9 |
| BUTALBITAL | 2165 | 3 | 50 | 16.67 |
| PENTOBARBITAL (SCHEDULE 2) | 2270 | 3 | 156.76 | 52.25 |
| COCAINE | 9041L | 13 | 182.45 | 14.03 |
| CODEINE | 9050 | 41 | 4,599.08 | 112.17 |
| BUPRENORPHINE | 9064 | 16 | 103.54 | 6.47 |
| OXYCODONE | 9143 | 50 | 10,948.96 | 218.98 |
| HYDROMORPHONE | 9150 | 52 | 907.83 | 17.46 |
| HYDROCODONE | 9193 | 55 | 9,906.46 | 180.12 |
| MEPERIDINE (PETHIDINE) | 9230 | 35 | 333.89 | 9.54 |
| METHADONE | 9250B | 24 | 621.88 | 25.91 |
| MORPHINE | 9300 | 57 | 7,420.16 | 130.18 |
| OPIUM POWDERED | 9639 | 16 | 103.68 | 6.48 |
| OXYMORPHONE | 9652 | 3 | 50.94 | 16.98 |
| ALFENTANIL | 9737 | 4 | 0.22 | 0.06 |
| REMIFENTANIL | 9739 | 6 | 1.59 | 0.27 |
| SUFENTANIL BASE | 9740 | 3 | 0.1 | 0.03 |
| TAPENTADOL | 9780 | 7 | 1,764 | 252 |
| FENTANYL BASE | 9801 | 53 | 203.46 | 3.84 |

STATE:NORTH DAKOTA BUSINESS ACTIVITY:C - PRACTITIONERS

| DRUG NAME | DRUG CODE | BUYERS | TOTAL GRAMS | AVG GRAMS |
|---|---|---|---|---|
| PENTOBARBITAL (SCHEDULE 2) | 2270 | 24 | 24,792.36 | 1,033.01 |

| | | | | |
|---|---|---|---|---|
| COCAINE | 9041L | 3 | 1.43 | 0.48 |
| CODEINE | 9050 | 7 | 157.59 | 22.51 |
| BUPRENORPHINE | 9064 | 52 | 2.45 | 0.05 |
| OXYCODONE | 9143 | 8 | 597.8 | 74.73 |
| HYDROMORPHONE | 9150 | 32 | 28.45 | 0.89 |
| HYDROCODONE | 9193 | 30 | 243.11 | 8.1 |
| MEPERIDINE (PETHIDINE) | 9230 | 17 | 42.16 | 2.48 |
| METHADONE | 9250B | 2 | 3.22 | 1.61 |
| MORPHINE | 9300 | 37 | 91.47 | 2.47 |
| OPIUM POWDERED | 9639 | 1 | 1.44 | 1.44 |
| FENTANYL BASE | 9801 | 50 | 6.18 | 0.12 |

STATE:NORTH DAKOTA BUSINESS ACTIVITY:M - MID-LEVEL PRACTITIONERS

| DRUG NAME | DRUG CODE | BUYERS | TOTAL GRAMS | AVG GRAMS |
|---|---|---|---|---|
| MEPERIDINE (PETHIDINE) | 9230 | 1 | 0.87 | 0.87 |
| MORPHINE | 9300 | 2 | 0.06 | 0.03 |

STATE:NORTH DAKOTA BUSINESS ACTIVITY:N-U NARCOTIC TREATMENT PROGRAMS

| DRUG NAME | DRUG CODE | BUYERS | TOTAL GRAMS | AVG GRAMS |
|---|---|---|---|---|
| METHADONE | 9250B | 1 | 286.21 | 286.21 |

STATE:OHIO BUSINESS ACTIVITY:A - PHARMACIES

| DRUG NAME | DRUG CODE | BUYERS | TOTAL GRAMS | AVG GRAMS |
|---|---|---|---|---|
| AMPHETAMINE | 1100 | 2,262 | 798,847.61 | 353.16 |
| DL-METHAMPHETAMINE RACEMIC BASE | 1105B | 7 | 35.36 | 5.05 |
| D-METHAMPHETAMINE | 1105D | 36 | 315.74 | 8.77 |
| LISDEXAMFETAMINE | 1205 | 2,203 | 453,105.53 | 205.68 |
| METHYLPHENIDATE (DL;D;L;ISOMERS) | 1724 | 2,253 | 911,169.43 | 404.42 |

| | | | | |
|---|---|---|---|---|
| BARBITURIC ACID DERIVIATIVE OR SALT [PER 21C | 2100 | 22 | 275 | 12.5 |
| BUTALBITAL | 2165 | 1,251 | 49,435 | 39.52 |
| SECOBARBITAL (SCHEDULE 2) | 2315 | 8 | 183.1 | 22.89 |
| NABILONE | 7379 | 4 | 0.75 | 0.19 |
| COCAINE | 9041L | 4 | 12.36 | 3.09 |
| CODEINE | 9050 | 2,251 | 443,677.84 | 197.1 |
| BUPRENORPHINE | 9064 | 2,100 | 180,232.73 | 85.83 |
| DIHYDROCODEINE | 9120 | 9 | 134.59 | 14.95 |
| OXYCODONE | 9143 | 2,275 | 2,108,791.69 | 926.94 |
| HYDROMORPHONE | 9150 | 2,000 | 32,047.95 | 16.02 |
| HYDROCODONE | 9193 | 2,286 | 837,455.52 | 366.34 |
| LEVORPHANOL | 9220L | 7 | 24.61 | 3.52 |
| MEPERIDINE (PETHIDINE) | 9230 | 579 | 11,354.49 | 19.61 |
| METHADONE | 9250B | 1,959 | 103,218.50 | 52.69 |
| MORPHINE | 9300 | 2,209 | 488,663.10 | 221.21 |
| OPIUM TINCTURE (LAUDANUM) | 9630 | 22 | 96.92 | 4.41 |
| OPIUM POWDERED | 9639 | 269 | 822.96 | 3.06 |
| OXYMORPHONE | 9652 | 1,536 | 52,518.15 | 34.19 |
| ALFENTANIL | 9737 | 2 | 0.3 | 0.15 |
| SUFENTANIL BASE | 9740 | 4 | 1.44 | 0.36 |
| TAPENTADOL | 9780 | 1,755 | 279,024 | 158.99 |
| FENTANYL BASE | 9801 | 2,162 | 10,832.32 | 5.01 |

STATE:OHIO BUSINESS ACTIVITY:B - HOSPITALS

| DRUG NAME | DRUG CODE | BUYERS | TOTAL GRAMS | AVG GRAMS |
|---|---|---|---|---|
| AMPHETAMINE | 1100 | 167 | 13,003.40 | 77.86 |
| DL-METHAMPHETAMINE RACEMIC BASE | 1105B | 1 | 2.01 | 2.01 |
| D-METHAMPHETAMINE | 1105D | 1 | 0.4 | 0.4 |
| LISDEXAMFETAMINE | 1205 | 74 | 3,388.82 | 45.79 |
| METHYLPHENIDATE (DL;D;L;ISOMERS) | 1724 | 225 | 15,970.47 | 70.98 |
| GAMMA HYDROXYBUTYRIC ACID(FDA APPROVED | 2012 | 2 | 6,464.45 | 3,232.22 |
| AMOBARBITAL (SCHEDULE 2) | 2125 | 6 | 25.52 | 4.25 |

| | | | | |
|---|---|---|---|---|
| BUTALBITAL | 2165 | 16 | 210 | 13.13 |
| PENTOBARBITAL (SCHEDULE 2) | 2270 | 33 | 25,454.95 | 771.36 |
| SECOBARBITAL (SCHEDULE 2) | 2315 | 1 | 18.31 | 18.31 |
| COCAINE | 9041L | 175 | 707.35 | 4.04 |
| CODEINE | 9050 | 293 | 12,921.64 | 44.1 |
| BUPRENORPHINE | 9064 | 229 | 5,057.35 | 22.08 |
| OXYCODONE | 9143 | 380 | 105,733.14 | 278.25 |
| HYDROMORPHONE | 9150 | 389 | 8,963.45 | 23.04 |
| HYDROCODONE | 9193 | 389 | 29,499.21 | 75.83 |
| MEPERIDINE (PETHIDINE) | 9230 | 321 | 10,692.14 | 33.31 |
| METHADONE | 9250B | 255 | 13,744.48 | 53.9 |
| MORPHINE | 9300 | 403 | 54,386.88 | 134.96 |
| OPIUM POWDERED | 9639 | 187 | 904.68 | 4.84 |
| OXYMORPHONE | 9652 | 64 | 650.73 | 10.17 |
| ALFENTANIL | 9737 | 30 | 20.67 | 0.69 |
| REMIFENTANIL | 9739 | 115 | 74.72 | 0.65 |
| SUFENTANIL BASE | 9740 | 61 | 1.44 | 0.02 |
| TAPENTADOL | 9780 | 67 | 3,223.35 | 48.11 |
| FENTANYL BASE | 9801 | 462 | 1,453.58 | 3.15 |

STATE:OHIO BUSINESS ACTIVITY:C - PRACTITIONERS

| DRUG NAME | DRUG CODE | BUYERS | TOTAL GRAMS | AVG GRAMS |
|---|---|---|---|---|
| AMPHETAMINE | 1100 | 1 | 0.35 | 0.35 |
| LISDEXAMFETAMINE | 1205 | 2 | 32.05 | 16.02 |
| PENTOBARBITAL (SCHEDULE 2) | 2270 | 239 | 205,937.97 | 861.67 |
| COCAINE | 9041L | 6 | 3.29 | 0.55 |
| CODEINE | 9050 | 63 | 496.25 | 7.88 |
| BUPRENORPHINE | 9064 | 869 | 143.76 | 0.17 |
| DIHYDROCODEINE | 9120 | 1 | 0.21 | 0.21 |
| OXYCODONE | 9143 | 35 | 2,327.39 | 66.5 |
| HYDROMORPHONE | 9150 | 313 | 174.56 | 0.56 |
| HYDROCODONE | 9193 | 291 | 783.66 | 2.69 |

| | | | | |
|---|---|---|---|---|
| MEPERIDINE (PETHIDINE) | 9230 | 67 | 511.9 | 7.64 |
| METHADONE | 9250B | 8 | 7.87 | 0.98 |
| MORPHINE | 9300 | 187 | 638.18 | 3.41 |
| OXYMORPHONE | 9652 | 11 | 16.71 | 1.52 |
| NOROXYMORPHONE | 9668 | 5 | 53.3 | 10.66 |
| ALFENTANIL | 9737 | 3 | 0.11 | 0.04 |
| REMIFENTANIL | 9739 | 12 | 1.32 | 0.11 |
| FENTANYL BASE | 9801 | 381 | 85.13 | 0.22 |

STATE:OHIO BUSINESS ACTIVITY:D - TEACHING INSTITUTIONS

| DRUG NAME | DRUG CODE | BUYERS | TOTAL GRAMS | AVG GRAMS |
|---|---|---|---|---|
| PENTOBARBITAL (SCHEDULE 2) | 2270 | 1 | 2.28 | 2.28 |
| BUPRENORPHINE | 9064 | 3 | 0.26 | 0.09 |
| HYDROMORPHONE | 9150 | 2 | 0.12 | 0.06 |
| MORPHINE | 9300 | 1 | 0.12 | 0.12 |
| REMIFENTANIL | 9739 | 1 | 0.04 | 0.04 |
| FENTANYL BASE | 9801 | 3 | 0.77 | 0.26 |

STATE:OHIO BUSINESS ACTIVITY:M - MID-LEVEL PRACTITIONERS

| DRUG NAME | DRUG CODE | BUYERS | TOTAL GRAMS | AVG GRAMS |
|---|---|---|---|---|
| PENTOBARBITAL (SCHEDULE 2) | 2270 | 22 | 24,969.81 | 1,134.99 |
| BUPRENORPHINE | 9064 | 3 | 0.04 | 0.01 |
| HYDROMORPHONE | 9150 | 4 | 0.42 | 0.1 |
| MEPERIDINE (PETHIDINE) | 9230 | 1 | 0.44 | 0.44 |
| MORPHINE | 9300 | 93 | 58.85 | 0.63 |
| FENTANYL BASE | 9801 | 115 | 1.66 | 0.01 |

STATE:OHIO BUSINESS ACTIVITY:N-U NARCOTIC TREATMENT PROGRAMS

| DRUG NAME | DRUG CODE | BUYERS | TOTAL GRAMS | AVG GRAMS |
|---|---|---|---|---|
| BUPRENORPHINE | 9064 | 14 | 4,043.58 | 288.83 |
| METHADONE | 9250B | 24 | 193,993.57 | 8,083.07 |

STATE:OKLAHOMA BUSINESS ACTIVITY:A - PHARMACIES

| DRUG NAME | DRUG CODE | BUYERS | TOTAL GRAMS | AVG GRAMS |
|---|---|---|---|---|
| AMPHETAMINE | 1100 | 897 | 197,019.65 | 219.64 |
| DL-METHAMPHETAMINE RACEMIC BASE | 1105B | 7 | 28.93 | 4.13 |
| D-METHAMPHETAMINE | 1105D | 16 | 35.35 | 2.21 |
| LISDEXAMFETAMINE | 1205 | 869 | 138,186.13 | 159.02 |
| METHYLPHENIDATE (DL;D;L;ISOMERS) | 1724 | 903 | 188,834.78 | 209.12 |
| BARBITURIC ACID DERIVIATIVE OR SALT [PER 21C | 2100 | 5 | 65 | 13 |
| BUTALBITAL | 2165 | 626 | 34,680 | 55.4 |
| COCAINE | 9041L | 1 | 0.29 | 0.29 |
| CODEINE | 9050 | 926 | 338,673.14 | 365.74 |
| BUPRENORPHINE | 9064 | 850 | 37,036.58 | 43.57 |
| DIHYDROCODEINE | 9120 | 230 | 973.09 | 4.23 |
| OXYCODONE | 9143 | 918 | 920,194.51 | 1,002.39 |
| HYDROMORPHONE | 9150 | 798 | 14,099.65 | 17.67 |
| HYDROCODONE | 9193 | 931 | 734,739.09 | 789.19 |
| LEVORPHANOL | 9220L | 3 | 1.86 | 0.62 |
| MEPERIDINE (PETHIDINE) | 9230 | 692 | 29,071.39 | 42.01 |
| METHADONE | 9250B | 749 | 38,136.68 | 50.92 |
| MORPHINE | 9300 | 911 | 379,541.72 | 416.62 |
| OPIUM POWDERED | 9639 | 45 | 131.76 | 2.93 |
| OXYMORPHONE | 9652 | 670 | 29,731.20 | 44.37 |
| REMIFENTANIL | 9739 | 1 | 0.09 | 0.09 |
| SUFENTANIL BASE | 9740 | 2 | 0.09 | 0.04 |
| TAPENTADOL | 9780 | 668 | 102,275.50 | 153.11 |
| FENTANYL BASE | 9801 | 897 | 7,380.79 | 8.23 |

STATISTICAL SUMMARY FOR RETAIL DRUG PURCHASES BY GRAMS WT

STATE:OKLAHOMA BUSINESS ACTIVITY:B - HOSPITALS

| DRUG NAME | DRUG CODE | BUYERS | TOTAL GRAMS | AVG GRAMS |
|---|---|---|---|---|
| AMPHETAMINE | 1100 | 70 | 6,106.58 | 87.24 |
| D-METHAMPHETAMINE | 1105D | 1 | 0.4 | 0.4 |
| LISDEXAMFETAMINE | 1205 | 38 | 2,347.43 | 61.77 |
| METHYLPHENIDATE (DL;D;L;ISOMERS) | 1724 | 88 | 9,996.43 | 113.6 |
| AMOBARBITAL (SCHEDULE 2) | 2125 | 1 | 0.46 | 0.46 |
| BUTALBITAL | 2165 | 15 | 319.95 | 21.33 |
| PENTOBARBITAL (SCHEDULE 2) | 2270 | 4 | 608.36 | 152.09 |
| COCAINE | 9041L | 42 | 150.92 | 3.59 |
| CODEINE | 9050 | 160 | 41,044.73 | 256.53 |
| BUPRENORPHINE | 9064 | 65 | 737.4 | 11.34 |
| DIHYDROCODEINE | 9120 | 3 | 4.27 | 1.42 |
| OXYCODONE | 9143 | 199 | 36,908.94 | 185.47 |
| HYDROMORPHONE | 9150 | 170 | 2,251.76 | 13.25 |
| HYDROCODONE | 9193 | 203 | 44,750.91 | 220.45 |
| MEPERIDINE (PETHIDINE) | 9230 | 170 | 6,523.05 | 38.37 |
| METHADONE | 9250B | 85 | 2,322.04 | 27.32 |
| MORPHINE | 9300 | 204 | 35,024.25 | 171.69 |
| OPIUM POWDERED | 9639 | 39 | 175.32 | 4.5 |
| OXYMORPHONE | 9652 | 20 | 273.89 | 13.69 |
| ALFENTANIL | 9737 | 10 | 4.07 | 0.41 |
| REMIFENTANIL | 9739 | 28 | 9.74 | 0.35 |
| SUFENTANIL BASE | 9740 | 34 | 0.66 | 0.02 |
| TAPENTADOL | 9780 | 25 | 1,405.35 | 56.21 |
| FENTANYL BASE | 9801 | 206 | 641.28 | 3.11 |

STATE:OKLAHOMA BUSINESS ACTIVITY:C - PRACTITIONERS

| DRUG NAME | DRUG CODE | BUYERS | TOTAL GRAMS | AVG GRAMS |
|---|---|---|---|---|
| AMPHETAMINE | 1100 | 1 | 0.84 | 0.84 |
| PENTOBARBITAL (SCHEDULE 2) | 2270 | 80 | 56,072.76 | 700.91 |

| | | | | |
|---|---|---|---|---|
| COCAINE | 9041L | 4 | 3 | 0.75 |
| CODEINE | 9050 | 61 | 2,295.25 | 37.63 |
| BUPRENORPHINE | 9064 | 273 | 47.85 | 0.18 |
| DIHYDROCODEINE | 9120 | 127 | 42.8 | 0.34 |
| OXYCODONE | 9143 | 13 | 957.51 | 73.65 |
| HYDROMORPHONE | 9150 | 97 | 84.52 | 0.87 |
| HYDROCODONE | 9193 | 69 | 2,293.75 | 33.24 |
| MEPERIDINE (PETHIDINE) | 9230 | 40 | 207.09 | 5.18 |
| METHADONE | 9250B | 6 | 58.4 | 9.73 |
| MORPHINE | 9300 | 84 | 1,126.68 | 13.41 |
| OPIUM POWDERED | 9639 | 1 | 1.44 | 1.44 |
| OXYMORPHONE | 9652 | 1 | 15.39 | 15.39 |
| NOROXYMORPHONE | 9668 | 2 | 2.81 | 1.41 |
| FENTANYL BASE | 9801 | 163 | 62.27 | 0.38 |

STATE:OKLAHOMA BUSINESS ACTIVITY:D - TEACHING INSTITUTIONS

| DRUG NAME | DRUG CODE | BUYERS | TOTAL GRAMS | AVG GRAMS |
|---|---|---|---|---|
| BUPRENORPHINE | 9064 | 2 | 0.16 | 0.08 |
| HYDROMORPHONE | 9150 | 2 | 7.09 | 3.55 |
| HYDROCODONE | 9193 | 1 | 7.77 | 7.77 |
| METHADONE | 9250B | 1 | 20.84 | 20.84 |
| MORPHINE | 9300 | 1 | 31.48 | 31.48 |
| FENTANYL BASE | 9801 | 1 | 1.03 | 1.03 |

STATE:OKLAHOMA BUSINESS ACTIVITY:M - MID-LEVEL PRACTITIONERS

| DRUG NAME | DRUG CODE | BUYERS | TOTAL GRAMS | AVG GRAMS |
|---|---|---|---|---|
| PENTOBARBITAL (SCHEDULE 2) | 2270 | 10 | 10,845.66 | 1,084.57 |
| CODEINE | 9050 | 23 | 173.57 | 7.55 |
| DIHYDROCODEINE | 9120 | 75 | 30.67 | 0.41 |
| HYDROMORPHONE | 9150 | 13 | 0.42 | 0.03 |

| | | | | |
|---|---|---|---|---|
| MORPHINE | 9300 | 47 | 14.43 | 0.31 |
| FENTANYL BASE | 9801 | 69 | 6.06 | 0.09 |

STATE:OKLAHOMA BUSINESS ACTIVITY:N-U NARCOTIC TREATMENT PROGRAMS

| DRUG NAME | DRUG CODE | BUYERS | TOTAL GRAMS | AVG GRAMS |
|---|---|---|---|---|
| BUPRENORPHINE | 9064 | 8 | 138.02 | 17.25 |
| METHADONE | 9250B | 18 | 112,622.85 | 6,256.82 |

STATE:OREGON BUSINESS ACTIVITY:A - PHARMACIES

| DRUG NAME | DRUG CODE | BUYERS | TOTAL GRAMS | AVG GRAMS |
|---|---|---|---|---|
| AMPHETAMINE | 1100 | 678 | 223,697.23 | 329.94 |
| DL-METHAMPHETAMINE RACEMIC BASE | 1105B | 6 | 59.07 | 9.85 |
| D-METHAMPHETAMINE | 1105D | 37 | 173.94 | 4.7 |
| LISDEXAMFETAMINE | 1205 | 636 | 57,777.63 | 90.85 |
| METHYLPHENIDATE (DL;D;L;ISOMERS) | 1724 | 677 | 246,323.96 | 363.85 |
| BARBITURIC ACID DERIVIATIVE OR SALT [PER 21C | 2100 | 5 | 25 | 5 |
| BUTALBITAL | 2165 | 429 | 15,655 | 36.49 |
| SECOBARBITAL (SCHEDULE 2) | 2315 | 39 | 1,034.52 | 26.53 |
| COCAINE | 9041L | 11 | 108.99 | 9.91 |
| CODEINE | 9050 | 681 | 140,890.05 | 206.89 |
| BUPRENORPHINE | 9064 | 654 | 29,253.82 | 44.73 |
| OXYCODONE | 9143 | 687 | 774,184.94 | 1,126.91 |
| HYDROMORPHONE | 9150 | 673 | 18,942.10 | 28.15 |
| HYDROCODONE | 9193 | 693 | 403,043.79 | 581.59 |
| LEVORPHANOL | 9220L | 15 | 19.15 | 1.28 |
| MEPERIDINE (PETHIDINE) | 9230 | 253 | 5,644.49 | 22.31 |
| METHADONE | 9250B | 665 | 83,378.83 | 125.38 |
| MORPHINE | 9300 | 694 | 326,138.23 | 469.94 |
| OPIUM TINCTURE (LAUDANUM) | 9630 | 3 | 4.72 | 1.57 |
| OPIUM POWDERED | 9639 | 68 | 149.04 | 2.19 |

| | | | | |
|---|---|---|---|---|
| OXYMORPHONE | 9652 | 416 | 8,964.27 | 21.55 |
| ALFENTANIL | 9737 | 2 | 1.75 | 0.88 |
| REMIFENTANIL | 9739 | 3 | 0.16 | 0.05 |
| SUFENTANIL BASE | 9740 | 1 | 0 | 0 |
| TAPENTADOL | 9780 | 382 | 32,085.50 | 83.99 |
| FENTANYL BASE | 9801 | 666 | 4,422.94 | 6.64 |

STATE:OREGON BUSINESS ACTIVITY:B - HOSPITALS

| DRUG NAME | DRUG CODE | BUYERS | TOTAL GRAMS | AVG GRAMS |
|---|---|---|---|---|
| AMPHETAMINE | 1100 | 46 | 3,437.25 | 74.72 |
| DL-METHAMPHETAMINE RACEMIC BASE | 1105B | 1 | 7.64 | 7.64 |
| D-METHAMPHETAMINE | 1105D | 1 | 0.4 | 0.4 |
| LISDEXAMFETAMINE | 1205 | 24 | 398.31 | 16.6 |
| METHYLPHENIDATE (DL;D;L;ISOMERS) | 1724 | 53 | 5,112.58 | 96.46 |
| AMOBARBITAL (SCHEDULE 2) | 2125 | 2 | 10.48 | 5.24 |
| BUTALBITAL | 2165 | 8 | 405 | 50.63 |
| PENTOBARBITAL (SCHEDULE 2) | 2270 | 5 | 1,720.27 | 344.05 |
| COCAINE | 9041L | 45 | 273.7 | 6.08 |
| CODEINE | 9050 | 91 | 4,777.73 | 52.5 |
| BUPRENORPHINE | 9064 | 53 | 813.42 | 15.35 |
| OXYCODONE | 9143 | 87 | 42,612.36 | 489.8 |
| HYDROMORPHONE | 9150 | 80 | 3,214.15 | 40.18 |
| HYDROCODONE | 9193 | 96 | 28,046.81 | 292.15 |
| MEPERIDINE (PETHIDINE) | 9230 | 65 | 1,143.39 | 17.59 |
| METHADONE | 9250B | 68 | 5,492.86 | 80.78 |
| MORPHINE | 9300 | 88 | 34,057.72 | 387.02 |
| OPIUM POWDERED | 9639 | 49 | 658.44 | 13.44 |
| OXYMORPHONE | 9652 | 7 | 12.33 | 1.76 |
| ALFENTANIL | 9737 | 16 | 13.44 | 0.84 |
| REMIFENTANIL | 9739 | 28 | 6.63 | 0.24 |
| SUFENTANIL BASE | 9740 | 30 | 1.3 | 0.04 |
| TAPENTADOL | 9780 | 3 | 28 | 9.33 |

| | | | | |
|---|---|---|---|---|
| FENTANYL BASE | 9801 | 82 | 404.83 | 4.94 |

STATE:OREGON BUSINESS ACTIVITY:C - PRACTITIONERS

| DRUG NAME | DRUG CODE | BUYERS | TOTAL GRAMS | AVG GRAMS |
|---|---|---|---|---|
| AMPHETAMINE | 1100 | 2 | 3.52 | 1.76 |
| PENTOBARBITAL (SCHEDULE 2) | 2270 | 128 | 114,729.27 | 896.32 |
| COCAINE | 9041L | 17 | 33.28 | 1.96 |
| CODEINE | 9050 | 40 | 2,496.25 | 62.41 |
| BUPRENORPHINE | 9064 | 560 | 90.22 | 0.16 |
| OXYCODONE | 9143 | 53 | 220.77 | 4.17 |
| HYDROMORPHONE | 9150 | 370 | 155.8 | 0.42 |
| HYDROCODONE | 9193 | 285 | 605.77 | 2.13 |
| MEPERIDINE (PETHIDINE) | 9230 | 86 | 474.31 | 5.52 |
| METHADONE | 9250B | 7 | 26.12 | 3.73 |
| MORPHINE | 9300 | 213 | 236.54 | 1.11 |
| OPIUM POWDERED | 9639 | 7 | 20.16 | 2.88 |
| OXYMORPHONE | 9652 | 18 | 2.77 | 0.15 |
| NOROXYMORPHONE | 9668 | 3 | 4.2 | 1.4 |
| ALFENTANIL | 9737 | 3 | 0.57 | 0.19 |
| REMIFENTANIL | 9739 | 35 | 4.51 | 0.13 |
| SUFENTANIL BASE | 9740 | 4 | 0.02 | 0 |
| FENTANYL BASE | 9801 | 356 | 51.04 | 0.14 |

STATE:OREGON BUSINESS ACTIVITY:D - TEACHING INSTITUTIONS

| DRUG NAME | DRUG CODE | BUYERS | TOTAL GRAMS | AVG GRAMS |
|---|---|---|---|---|
| BUPRENORPHINE | 9064 | 1 | 0.02 | 0.02 |
| HYDROMORPHONE | 9150 | 1 | 0.02 | 0.02 |
| MORPHINE | 9300 | 1 | 0.28 | 0.28 |
| FENTANYL BASE | 9801 | 1 | 0.03 | 0.03 |

STATE:OREGON BUSINESS ACTIVITY:M - MID-LEVEL PRACTITIONERS

| DRUG NAME | DRUG CODE | BUYERS | TOTAL GRAMS | AVG GRAMS |
|---|---|---|---|---|
| PENTOBARBITAL (SCHEDULE 2) | 2270 | 9 | 9,785.70 | 1,087.30 |
| CODEINE | 9050 | 3 | 41.77 | 13.92 |
| BUPRENORPHINE | 9064 | 1 | 0.31 | 0.31 |
| HYDROMORPHONE | 9150 | 4 | 0.27 | 0.07 |
| HYDROCODONE | 9193 | 5 | 9.08 | 1.82 |
| FENTANYL BASE | 9801 | 1 | 0.04 | 0.04 |

STATE:OREGON BUSINESS ACTIVITY:N-U NARCOTIC TREATMENT PROGRAMS

| DRUG NAME | DRUG CODE | BUYERS | TOTAL GRAMS | AVG GRAMS |
|---|---|---|---|---|
| BUPRENORPHINE | 9064 | 12 | 4,481.23 | 373.44 |
| METHADONE | 9250B | 15 | 173,011.39 | 11,534.09 |

STATE:PENNSYLVANIA BUSINESS ACTIVITY:A - PHARMACIES

| DRUG NAME | DRUG CODE | BUYERS | TOTAL GRAMS | AVG GRAMS |
|---|---|---|---|---|
| AMPHETAMINE | 1100 | 2,995 | 827,536.77 | 276.31 |
| DL-METHAMPHETAMINE RACEMIC BASE | 1105B | 8 | 54.65 | 6.83 |
| D-METHAMPHETAMINE | 1105D | 77 | 487.26 | 6.33 |
| LISDEXAMFETAMINE | 1205 | 2,797 | 324,876.91 | 116.15 |
| METHYLPHENIDATE (DL;D;L;ISOMERS) | 1724 | 2,976 | 896,238.48 | 301.16 |
| BARBITURIC ACID DERIVIATIVE OR SALT [PER 21C | 2100 | 30 | 325 | 10.83 |
| BUTALBITAL | 2165 | 1,650 | 78,500 | 47.58 |
| PENTOBARBITAL (SCHEDULE 2) | 2270 | 1 | 41.01 | 41.01 |
| SECOBARBITAL (SCHEDULE 2) | 2315 | 7 | 283.81 | 40.54 |
| NABILONE | 7379 | 4 | 1.05 | 0.26 |
| COCAINE | 9041L | 10 | 42.28 | 4.23 |
| CODEINE | 9050 | 3,056 | 588,139.84 | 192.45 |

| | | | | |
|---|---|---|---|---|
| BUPRENORPHINE | 9064 | 2,807 | 217,488.42 | 77.48 |
| DIHYDROCODEINE | 9120 | 52 | 178.92 | 3.44 |
| OXYCODONE | 9143 | 3,074 | 3,312,413.83 | 1,077.56 |
| HYDROMORPHONE | 9150 | 2,708 | 57,124.23 | 21.09 |
| HYDROCODONE | 9193 | 3,059 | 751,130.13 | 245.55 |
| LEVORPHANOL | 9220L | 40 | 31.69 | 0.79 |
| MEPERIDINE (PETHIDINE) | 9230 | 590 | 14,796 | 25.08 |
| METHADONE | 9250B | 2,360 | 171,754.05 | 72.78 |
| MORPHINE | 9300 | 2,985 | 955,357.04 | 320.05 |
| OPIUM TINCTURE (LAUDANUM) | 9630 | 6 | 12.99 | 2.17 |
| OPIUM POWDERED | 9639 | 183 | 757.8 | 4.14 |
| OXYMORPHONE | 9652 | 2,118 | 113,408.93 | 53.55 |
| REMIFENTANIL | 9739 | 4 | 1.26 | 0.32 |
| SUFENTANIL BASE | 9740 | 1 | 4.01 | 4.01 |
| TAPENTADOL | 9780 | 1,913 | 280,945 | 146.86 |
| FENTANYL BASE | 9801 | 2,938 | 21,923.86 | 7.46 |

STATE:PENNSYLVANIA BUSINESS ACTIVITY:B - HOSPITALS

| DRUG NAME | DRUG CODE | BUYERS | TOTAL GRAMS | AVG GRAMS |
|---|---|---|---|---|
| AMPHETAMINE | 1100 | 167 | 11,664.33 | 69.85 |
| LISDEXAMFETAMINE | 1205 | 56 | 2,843.67 | 50.78 |
| METHYLPHENIDATE (DL;D;L;ISOMERS) | 1724 | 232 | 13,333.92 | 57.47 |
| AMOBARBITAL (SCHEDULE 2) | 2125 | 8 | 42.84 | 5.35 |
| BUTALBITAL | 2165 | 19 | 490 | 25.79 |
| PENTOBARBITAL (SCHEDULE 2) | 2270 | 27 | 2,969.34 | 109.98 |
| COCAINE | 9041L | 159 | 1,455.91 | 9.16 |
| CODEINE | 9050 | 287 | 18,272.27 | 63.67 |
| BUPRENORPHINE | 9064 | 191 | 5,909.49 | 30.94 |
| OXYCODONE | 9143 | 330 | 141,541.08 | 428.91 |
| HYDROMORPHONE | 9150 | 302 | 11,148.81 | 36.92 |
| HYDROCODONE | 9193 | 296 | 32,083.84 | 108.39 |
| LEVORPHANOL | 9220L | 1 | 7.08 | 7.08 |

| | | | | |
|---|---|---|---|---|
| MEPERIDINE (PETHIDINE) | 9230 | 230 | 2,487.09 | 10.81 |
| METHADONE | 9250B | 241 | 15,854.89 | 65.79 |
| MORPHINE | 9300 | 328 | 78,515.51 | 239.38 |
| OPIUM POWDERED | 9639 | 162 | 711.72 | 4.39 |
| OXYMORPHONE | 9652 | 74 | 1,367.75 | 18.48 |
| ALFENTANIL | 9737 | 8 | 1.82 | 0.23 |
| REMIFENTANIL | 9739 | 116 | 133.93 | 1.15 |
| SUFENTANIL BASE | 9740 | 43 | 1.2 | 0.03 |
| TAPENTADOL | 9780 | 66 | 2,071.20 | 31.38 |
| FENTANYL BASE | 9801 | 362 | 1,852.15 | 5.12 |

STATE:PENNSYLVANIA BUSINESS ACTIVITY:C - PRACTITIONERS

| DRUG NAME | DRUG CODE | BUYERS | TOTAL GRAMS | AVG GRAMS |
|---|---|---|---|---|
| AMPHETAMINE | 1100 | 3 | 19.72 | 6.57 |
| LISDEXAMFETAMINE | 1205 | 1 | 13.87 | 13.87 |
| METHYLPHENIDATE (DL;D;L;ISOMERS) | 1724 | 1 | 39.55 | 39.55 |
| BUTALBITAL | 2165 | 2 | 10 | 5 |
| PENTOBARBITAL (SCHEDULE 2) | 2270 | 275 | 251,573.52 | 914.81 |
| COCAINE | 9041L | 28 | 238.98 | 8.54 |
| CODEINE | 9050 | 184 | 4,683.03 | 25.45 |
| BUPRENORPHINE | 9064 | 949 | 515.88 | 0.54 |
| DIHYDROCODEINE | 9120 | 86 | 949.72 | 11.04 |
| OXYCODONE | 9143 | 64 | 2,121.20 | 33.14 |
| HYDROMORPHONE | 9150 | 464 | 250.85 | 0.54 |
| HYDROCODONE | 9193 | 437 | 1,626.88 | 3.72 |
| MEPERIDINE (PETHIDINE) | 9230 | 110 | 473.99 | 4.31 |
| METHADONE | 9250B | 25 | 184.51 | 7.38 |
| MORPHINE | 9300 | 279 | 482.86 | 1.73 |
| OPIUM POWDERED | 9639 | 2 | 1.8 | 0.9 |
| OXYMORPHONE | 9652 | 9 | 17.76 | 1.97 |
| NOROXYMORPHONE | 9668 | 5 | 6.02 | 1.2 |
| ALFENTANIL | 9737 | 6 | 0.87 | 0.15 |

| | | | | |
|---|---|---|---|---|
| REMIFENTANIL | 9739 | 27 | 2.29 | 0.08 |
| SUFENTANIL BASE | 9740 | 2 | 0.02 | 0.01 |
| FENTANYL BASE | 9801 | 627 | 103.54 | 0.17 |

STATE:PENNSYLVANIA BUSINESS ACTIVITY:D - TEACHING INSTITUTIONS

| DRUG NAME | DRUG CODE | BUYERS | TOTAL GRAMS | AVG GRAMS |
|---|---|---|---|---|
| D-METHAMPHETAMINE | 1105D | 1 | 0.08 | 0.08 |
| METHYLPHENIDATE (DL;D;L;ISOMERS) | 1724 | 1 | 0.43 | 0.43 |
| CODEINE | 9050 | 1 | 423.42 | 423.42 |
| BUPRENORPHINE | 9064 | 3 | 1.39 | 0.46 |
| OXYCODONE | 9143 | 1 | 1.34 | 1.34 |
| HYDROMORPHONE | 9150 | 2 | 0.39 | 0.2 |
| HYDROCODONE | 9193 | 1 | 43.14 | 43.14 |
| METHADONE | 9250B | 1 | 46.87 | 46.87 |
| MORPHINE | 9300 | 3 | 21.85 | 7.28 |
| OXYMORPHONE | 9652 | 1 | 0.45 | 0.45 |
| REMIFENTANIL | 9739 | 2 | 0.09 | 0.05 |
| FENTANYL BASE | 9801 | 4 | 4.66 | 1.17 |

STATE:PENNSYLVANIA BUSINESS ACTIVITY:M - MID-LEVEL PRACTITIONERS

| DRUG NAME | DRUG CODE | BUYERS | TOTAL GRAMS | AVG GRAMS |
|---|---|---|---|---|
| CODEINE | 9050 | 9 | 9.57 | 1.06 |
| DIHYDROCODEINE | 9120 | 6 | 3.01 | 0.5 |

STATE:PENNSYLVANIA BUSINESS ACTIVITY:N-U NARCOTIC TREATMENT PROGRAMS

| DRUG NAME | DRUG CODE | BUYERS | TOTAL GRAMS | AVG GRAMS |
|---|---|---|---|---|
| BUPRENORPHINE | 9064 | 6 | 237.95 | 39.66 |
| METHADONE | 9250B | 80 | 576,590.34 | 7,207.38 |

STATE:PUERTO RICO BUSINESS ACTIVITY:A - PHARMACIES

| DRUG NAME | DRUG CODE | BUYERS | TOTAL GRAMS | AVG GRAMS |
|---|---|---|---|---|
| AMPHETAMINE | 1100 | 623 | 32,480.74 | 52.14 |
| LISDEXAMFETAMINE | 1205 | 266 | 7,034.32 | 26.44 |
| METHYLPHENIDATE (DL;D;L;ISOMERS) | 1724 | 680 | 86,453.30 | 127.14 |
| BUTALBITAL | 2165 | 33 | 675 | 20.45 |
| CODEINE | 9050 | 1,033 | 48,284.82 | 46.74 |
| BUPRENORPHINE | 9064 | 308 | 9,425.79 | 30.6 |
| OXYCODONE | 9143 | 814 | 65,692.29 | 80.7 |
| HYDROMORPHONE | 9150 | 26 | 177.55 | 6.83 |
| HYDROCODONE | 9193 | 581 | 4,180.84 | 7.2 |
| MEPERIDINE (PETHIDINE) | 9230 | 716 | 9,389.43 | 13.11 |
| METHADONE | 9250B | 16 | 220.11 | 13.76 |
| MORPHINE | 9300 | 187 | 6,489.82 | 34.7 |
| TAPENTADOL | 9780 | 86 | 3,874.50 | 45.05 |
| FENTANYL BASE | 9801 | 672 | 671.74 | 1 |

STATE:PUERTO RICO BUSINESS ACTIVITY:B - HOSPITALS

| DRUG NAME | DRUG CODE | BUYERS | TOTAL GRAMS | AVG GRAMS |
|---|---|---|---|---|
| AMPHETAMINE | 1100 | 8 | 90.41 | 11.3 |
| METHYLPHENIDATE (DL;D;L;ISOMERS) | 1724 | 15 | 271.79 | 18.12 |
| AMOBARBITAL (SCHEDULE 2) | 2125 | 1 | 0.91 | 0.91 |
| BUTALBITAL | 2165 | 1 | 5 | 5 |
| PENTOBARBITAL (SCHEDULE 2) | 2270 | 2 | 2,226.09 | 1,113.05 |
| CODEINE | 9050 | 77 | 4,895.55 | 63.58 |
| BUPRENORPHINE | 9064 | 16 | 533.59 | 33.35 |
| OXYCODONE | 9143 | 70 | 7,367.44 | 105.25 |
| HYDROMORPHONE | 9150 | 5 | 109.24 | 21.85 |
| HYDROCODONE | 9193 | 26 | 229.46 | 8.83 |

| | | | | |
|---|---|---|---|---|
| MEPERIDINE (PETHIDINE) | 9230 | 92 | 15,746.26 | 171.16 |
| METHADONE | 9250B | 3 | 190.95 | 63.65 |
| MORPHINE | 9300 | 89 | 5,427.73 | 60.99 |
| REMIFENTANIL | 9739 | 2 | 0.16 | 0.08 |
| SUFENTANIL BASE | 9740 | 14 | 0.76 | 0.05 |
| TAPENTADOL | 9780 | 6 | 378.97 | 63.16 |
| FENTANYL BASE | 9801 | 85 | 189.96 | 2.23 |

STATE:PUERTO RICO BUSINESS ACTIVITY:C - PRACTITIONERS

| DRUG NAME | DRUG CODE | BUYERS | TOTAL GRAMS | AVG GRAMS |
|---|---|---|---|---|
| PENTOBARBITAL (SCHEDULE 2) | 2270 | 1 | 533.17 | 533.17 |
| BUPRENORPHINE | 9064 | 19 | 8.69 | 0.46 |
| HYDROMORPHONE | 9150 | 1 | 0.16 | 0.16 |
| HYDROCODONE | 9193 | 1 | 0.72 | 0.72 |
| MEPERIDINE (PETHIDINE) | 9230 | 6 | 105.67 | 17.61 |
| MORPHINE | 9300 | 3 | 2.07 | 0.69 |
| NOROXYMORPHONE | 9668 | 2 | 18.31 | 9.15 |
| FENTANYL BASE | 9801 | 13 | 0.66 | 0.05 |

STATE:PUERTO RICO BUSINESS ACTIVITY:M - MID-LEVEL PRACTITIONERS

| DRUG NAME | DRUG CODE | BUYERS | TOTAL GRAMS | AVG GRAMS |
|---|---|---|---|---|
| PENTOBARBITAL (SCHEDULE 2) | 2270 | 2 | 2,137.23 | 1,068.62 |

STATE:PUERTO RICO BUSINESS ACTIVITY:N-U NARCOTIC TREATMENT PROGRAMS

| DRUG NAME | DRUG CODE | BUYERS | TOTAL GRAMS | AVG GRAMS |
|---|---|---|---|---|
| METHADONE | 9250B | 6 | 74,270.98 | 12,378.50 |

STATISTICAL SUMMARY FOR RETAIL DRUG PURCHASES BY GRAMS WT

STATE:RHODE ISLAND BUSINESS ACTIVITY:A - PHARMACIES

| DRUG NAME | DRUG CODE | BUYERS | TOTAL GRAMS | AVG GRAMS |
|---|---|---|---|---|
| AMPHETAMINE | 1100 | 195 | 137,070.60 | 702.93 |
| DL-METHAMPHETAMINE RACEMIC BASE | 1105B | 3 | 5.63 | 1.88 |
| D-METHAMPHETAMINE | 1105D | 4 | 9.24 | 2.31 |
| LISDEXAMFETAMINE | 1205 | 187 | 29,098.09 | 155.6 |
| METHYLPHENIDATE (DL;D;L;ISOMERS) | 1724 | 194 | 74,691.90 | 385.01 |
| BARBITURIC ACID DERIVIATIVE OR SALT [PER 21C | 2100 | 6 | 85 | 14.17 |
| BUTALBITAL | 2165 | 105 | 4,185 | 39.86 |
| SECOBARBITAL (SCHEDULE 2) | 2315 | 1 | 45.78 | 45.78 |
| NABILONE | 7379 | 1 | 0.05 | 0.05 |
| COCAINE | 9041L | 1 | 0.89 | 0.89 |
| CODEINE | 9050 | 195 | 32,374.05 | 166.02 |
| BUPRENORPHINE | 9064 | 192 | 26,488.26 | 137.96 |
| DIHYDROCODEINE | 9120 | 2 | 6.41 | 3.2 |
| OXYCODONE | 9143 | 197 | 184,131.34 | 934.68 |
| HYDROMORPHONE | 9150 | 176 | 2,840.95 | 16.14 |
| HYDROCODONE | 9193 | 197 | 47,546.16 | 241.35 |
| MEPERIDINE (PETHIDINE) | 9230 | 30 | 589.79 | 19.66 |
| METHADONE | 9250B | 152 | 7,230.51 | 47.57 |
| MORPHINE | 9300 | 197 | 46,128.32 | 234.15 |
| OPIUM POWDERED | 9639 | 19 | 31.68 | 1.67 |
| OXYMORPHONE | 9652 | 98 | 2,371.83 | 24.2 |
| TAPENTADOL | 9780 | 115 | 9,096.50 | 79.1 |
| FENTANYL BASE | 9801 | 184 | 745.48 | 4.05 |

STATE:RHODE ISLAND BUSINESS ACTIVITY:B - HOSPITALS

| DRUG NAME | DRUG CODE | BUYERS | TOTAL GRAMS | AVG GRAMS |
|---|---|---|---|---|
| AMPHETAMINE | 1100 | 20 | 2,185.75 | 109.29 |
| LISDEXAMFETAMINE | 1205 | 7 | 293.67 | 41.95 |
| METHYLPHENIDATE (DL;D;L;ISOMERS) | 1724 | 19 | 2,781.29 | 146.38 |

| | | | | |
|---|---|---|---|---|
| BUTALBITAL | 2165 | 1 | 5 | 5 |
| PENTOBARBITAL (SCHEDULE 2) | 2270 | 2 | 817.07 | 408.54 |
| COCAINE | 9041L | 11 | 80.07 | 7.28 |
| CODEINE | 9050 | 21 | 2,038.24 | 97.06 |
| BUPRENORPHINE | 9064 | 14 | 599.45 | 42.82 |
| OXYCODONE | 9143 | 21 | 12,002.43 | 571.54 |
| HYDROMORPHONE | 9150 | 18 | 683.81 | 37.99 |
| HYDROCODONE | 9193 | 21 | 2,650.21 | 126.2 |
| MEPERIDINE (PETHIDINE) | 9230 | 14 | 333.24 | 23.8 |
| METHADONE | 9250B | 16 | 1,373.62 | 85.85 |
| MORPHINE | 9300 | 19 | 5,790.21 | 304.75 |
| OPIUM POWDERED | 9639 | 10 | 93.6 | 9.36 |
| OXYMORPHONE | 9652 | 1 | 1.34 | 1.34 |
| ALFENTANIL | 9737 | 2 | 0.29 | 0.15 |
| REMIFENTANIL | 9739 | 6 | 1.65 | 0.28 |
| SUFENTANIL BASE | 9740 | 4 | 0.03 | 0.01 |
| TAPENTADOL | 9780 | 2 | 95 | 47.5 |
| FENTANYL BASE | 9801 | 18 | 167.69 | 9.32 |

STATE:RHODE ISLAND BUSINESS ACTIVITY:C - PRACTITIONERS

| DRUG NAME | DRUG CODE | BUYERS | TOTAL GRAMS | AVG GRAMS |
|---|---|---|---|---|
| PENTOBARBITAL (SCHEDULE 2) | 2270 | 34 | 29,322.65 | 862.43 |
| COCAINE | 9041L | 3 | 15.28 | 5.09 |
| CODEINE | 9050 | 1 | 2.21 | 2.21 |
| BUPRENORPHINE | 9064 | 76 | 437.89 | 5.76 |
| OXYCODONE | 9143 | 4 | 5.38 | 1.34 |
| HYDROMORPHONE | 9150 | 45 | 15.58 | 0.35 |
| HYDROCODONE | 9193 | 32 | 85.81 | 2.68 |
| MEPERIDINE (PETHIDINE) | 9230 | 11 | 27.78 | 2.53 |
| METHADONE | 9250B | 2 | 48.3 | 24.15 |
| MORPHINE | 9300 | 20 | 24.68 | 1.23 |
| OPIUM POWDERED | 9639 | 1 | 7.2 | 7.2 |

| | | | | |
|---|---|---|---|---|
| OXYMORPHONE | 9652 | 5 | 0.74 | 0.15 |
| NOROXYMORPHONE | 9668 | 1 | 3.34 | 3.34 |
| REMIFENTANIL | 9739 | 1 | 0.2 | 0.2 |
| FENTANYL BASE | 9801 | 44 | 9.07 | 0.21 |

STATE:RHODE ISLAND BUSINESS ACTIVITY:M - MID-LEVEL PRACTITIONERS

| DRUG NAME | DRUG CODE | BUYERS | TOTAL GRAMS | AVG GRAMS |
|---|---|---|---|---|
| CODEINE | 9050 | 1 | 2.21 | 2.21 |

STATE:RHODE ISLAND BUSINESS ACTIVITY:N-U NARCOTIC TREATMENT PROGRAMS

| DRUG NAME | DRUG CODE | BUYERS | TOTAL GRAMS | AVG GRAMS |
|---|---|---|---|---|
| BUPRENORPHINE | 9064 | 11 | 137.1 | 12.46 |
| METHADONE | 9250B | 18 | 161,149.59 | 8,952.76 |

STATE:SOUTH CAROLINA BUSINESS ACTIVITY:A - PHARMACIES

| DRUG NAME | DRUG CODE | BUYERS | TOTAL GRAMS | AVG GRAMS |
|---|---|---|---|---|
| AMPHETAMINE | 1100 | 1,123 | 515,862.33 | 459.36 |
| DL-METHAMPHETAMINE RACEMIC BASE | 1105B | 1 | 0.8 | 0.8 |
| D-METHAMPHETAMINE | 1105D | 21 | 49.41 | 2.35 |
| LISDEXAMFETAMINE | 1205 | 1,083 | 237,117.54 | 218.95 |
| METHYLPHENIDATE (DL;D;L;ISOMERS) | 1724 | 1,115 | 348,402.86 | 312.47 |
| BARBITURIC ACID DERIVIATIVE OR SALT [PER 21C | 2100 | 6 | 30 | 5 |
| BUTALBITAL | 2165 | 661 | 24,055 | 36.39 |
| COCAINE | 9041L | 2 | 0.96 | 0.48 |
| CODEINE | 9050 | 1,113 | 290,264.36 | 260.79 |
| BUPRENORPHINE | 9064 | 1,009 | 35,586.30 | 35.27 |
| DIHYDROCODEINE | 9120 | 18 | 60.89 | 3.38 |
| OXYCODONE | 9143 | 1,119 | 997,095.88 | 891.06 |

| | | | | |
|---|---|---|---|---|
| HYDROMORPHONE | 9150 | 1,045 | 20,817.65 | 19.92 |
| HYDROCODONE | 9193 | 1,132 | 599,011.87 | 529.16 |
| LEVORPHANOL | 9220L | 11 | 1.86 | 0.17 |
| MEPERIDINE (PETHIDINE) | 9230 | 836 | 25,232.76 | 30.18 |
| METHADONE | 9250B | 902 | 43,724.44 | 48.47 |
| MORPHINE | 9300 | 1,106 | 262,667.49 | 237.49 |
| OPIUM POWDERED | 9639 | 56 | 191.88 | 3.43 |
| OXYMORPHONE | 9652 | 864 | 33,051.64 | 38.25 |
| SUFENTANIL BASE | 9740 | 1 | 2 | 2 |
| TAPENTADOL | 9780 | 949 | 179,015 | 188.64 |
| FENTANYL BASE | 9801 | 1,074 | 5,598.11 | 5.21 |

STATE:SOUTH CAROLINA BUSINESS ACTIVITY:B - HOSPITALS

| DRUG NAME | DRUG CODE | BUYERS | TOTAL GRAMS | AVG GRAMS |
|---|---|---|---|---|
| AMPHETAMINE | 1100 | 63 | 9,548.66 | 151.57 |
| DL-METHAMPHETAMINE RACEMIC BASE | 1105B | 1 | 12.46 | 12.46 |
| LISDEXAMFETAMINE | 1205 | 28 | 1,453.34 | 51.91 |
| METHYLPHENIDATE (DL;D;L;ISOMERS) | 1724 | 74 | 10,324.59 | 139.52 |
| GAMMA HYDROXYBUTYRIC ACID(FDA APPROVED | 2012 | 1 | 2,303.42 | 2,303.42 |
| AMOBARBITAL (SCHEDULE 2) | 2125 | 2 | 20.96 | 10.48 |
| BUTALBITAL | 2165 | 4 | 30 | 7.5 |
| PENTOBARBITAL (SCHEDULE 2) | 2270 | 11 | 7,942.85 | 722.08 |
| COCAINE | 9041L | 54 | 121.85 | 2.26 |
| CODEINE | 9050 | 102 | 29,173.39 | 286.01 |
| BUPRENORPHINE | 9064 | 68 | 874.54 | 12.86 |
| OXYCODONE | 9143 | 143 | 32,900.94 | 230.08 |
| HYDROMORPHONE | 9150 | 157 | 3,640.34 | 23.19 |
| HYDROCODONE | 9193 | 152 | 18,405.05 | 121.09 |
| MEPERIDINE (PETHIDINE) | 9230 | 124 | 4,668.23 | 37.65 |
| METHADONE | 9250B | 82 | 2,753.23 | 33.58 |
| MORPHINE | 9300 | 166 | 26,007.32 | 156.67 |
| OPIUM POWDERED | 9639 | 63 | 392.76 | 6.23 |

| | | | | |
|---|---|---|---|---|
| OXYMORPHONE | 9652 | 11 | 41.35 | 3.76 |
| ALFENTANIL | 9737 | 1 | 0.19 | 0.19 |
| REMIFENTANIL | 9739 | 29 | 19.66 | 0.68 |
| SUFENTANIL BASE | 9740 | 27 | 0.5 | 0.02 |
| TAPENTADOL | 9780 | 35 | 1,608.70 | 45.96 |
| FENTANYL BASE | 9801 | 176 | 642.43 | 3.65 |

STATE:SOUTH CAROLINA BUSINESS ACTIVITY:C - PRACTITIONERS

| DRUG NAME | DRUG CODE | BUYERS | TOTAL GRAMS | AVG GRAMS |
|---|---|---|---|---|
| AMPHETAMINE | 1100 | 1 | 0.91 | 0.91 |
| GAMMA HYDROXYBUTYRIC ACID(FDA APPROVED | 2012 | 1 | 6,002.11 | 6,002.11 |
| BUTALBITAL | 2165 | 1 | 5 | 5 |
| PENTOBARBITAL (SCHEDULE 2) | 2270 | 58 | 52,685.74 | 908.37 |
| COCAINE | 9041L | 4 | 19 | 4.75 |
| CODEINE | 9050 | 20 | 295.41 | 14.77 |
| BUPRENORPHINE | 9064 | 368 | 45.03 | 0.12 |
| DIHYDROCODEINE | 9120 | 2 | 0.64 | 0.32 |
| OXYCODONE | 9143 | 12 | 1,257.47 | 104.79 |
| HYDROMORPHONE | 9150 | 134 | 96 | 0.72 |
| HYDROCODONE | 9193 | 83 | 269.61 | 3.25 |
| MEPERIDINE (PETHIDINE) | 9230 | 84 | 589.35 | 7.02 |
| MORPHINE | 9300 | 97 | 447.21 | 4.61 |
| OXYMORPHONE | 9652 | 2 | 15.57 | 7.78 |
| NOROXYMORPHONE | 9668 | 1 | 0.49 | 0.49 |
| REMIFENTANIL | 9739 | 6 | 0.29 | 0.05 |
| FENTANYL BASE | 9801 | 211 | 18.17 | 0.09 |

STATE:SOUTH CAROLINA BUSINESS ACTIVITY:D - TEACHING INSTITUTIONS

| DRUG NAME | DRUG CODE | BUYERS | TOTAL GRAMS | AVG GRAMS |
|---|---|---|---|---|
| BUPRENORPHINE | 9064 | 1 | 0.04 | 0.04 |

| | | | | |
|---|---|---|---|---|
| HYDROMORPHONE | 9150 | 1 | 0.07 | 0.07 |

STATE:SOUTH CAROLINA BUSINESS ACTIVITY:M - MID-LEVEL PRACTITIONERS

| DRUG NAME | DRUG CODE | BUYERS | TOTAL GRAMS | AVG GRAMS |
|---|---|---|---|---|
| CODEINE | 9050 | 5 | 1.95 | 0.39 |
| MORPHINE | 9300 | 71 | 23.21 | 0.33 |
| FENTANYL BASE | 9801 | 54 | 0.45 | 0.01 |

STATE:SOUTH CAROLINA BUSINESS ACTIVITY:N-U NARCOTIC TREATMENT PROGRAMS

| DRUG NAME | DRUG CODE | BUYERS | TOTAL GRAMS | AVG GRAMS |
|---|---|---|---|---|
| BUPRENORPHINE | 9064 | 11 | 1,531.94 | 139.27 |
| METHADONE | 9250B | 22 | 175,669.10 | 7,984.96 |

STATE:SOUTH DAKOTA BUSINESS ACTIVITY:A - PHARMACIES

| DRUG NAME | DRUG CODE | BUYERS | TOTAL GRAMS | AVG GRAMS |
|---|---|---|---|---|
| AMPHETAMINE | 1100 | 195 | 60,115.58 | 308.29 |
| D-METHAMPHETAMINE | 1105D | 6 | 47.4 | 7.9 |
| LISDEXAMFETAMINE | 1205 | 196 | 46,791.41 | 238.73 |
| METHYLPHENIDATE (DL;D;L;ISOMERS) | 1724 | 196 | 84,902.72 | 433.18 |
| BUTALBITAL | 2165 | 94 | 3,940 | 41.91 |
| COCAINE | 9041L | 1 | 0.14 | 0.14 |
| CODEINE | 9050 | 197 | 27,605.67 | 140.13 |
| BUPRENORPHINE | 9064 | 150 | 1,360.56 | 9.07 |
| OXYCODONE | 9143 | 199 | 74,698.63 | 375.37 |
| HYDROMORPHONE | 9150 | 165 | 1,893.32 | 11.47 |
| HYDROCODONE | 9193 | 201 | 55,660.32 | 276.92 |
| MEPERIDINE (PETHIDINE) | 9230 | 61 | 1,579.48 | 25.89 |
| METHADONE | 9250B | 155 | 5,681.23 | 36.65 |

| | | | | |
|---|---|---|---|---|
| MORPHINE | 9300 | 192 | 31,997.52 | 166.65 |
| OPIUM POWDERED | 9639 | 28 | 29.16 | 1.04 |
| OXYMORPHONE | 9652 | 82 | 3,054.86 | 37.25 |
| SUFENTANIL BASE | 9740 | 1 | 0.01 | 0.01 |
| TAPENTADOL | 9780 | 145 | 21,935.50 | 151.28 |
| FENTANYL BASE | 9801 | 199 | 1,203.57 | 6.05 |

STATE:SOUTH DAKOTA BUSINESS ACTIVITY:B - HOSPITALS

| DRUG NAME | DRUG CODE | BUYERS | TOTAL GRAMS | AVG GRAMS |
|---|---|---|---|---|
| AMPHETAMINE | 1100 | 26 | 2,744.37 | 105.55 |
| D-METHAMPHETAMINE | 1105D | 2 | 1.61 | 0.8 |
| LISDEXAMFETAMINE | 1205 | 15 | 498.32 | 33.22 |
| METHYLPHENIDATE (DL;D;L;ISOMERS) | 1724 | 31 | 4,583.02 | 147.84 |
| BUTALBITAL | 2165 | 3 | 25 | 8.33 |
| PENTOBARBITAL (SCHEDULE 2) | 2270 | 3 | 236.96 | 78.99 |
| COCAINE | 9041L | 21 | 146.2 | 6.96 |
| CODEINE | 9050 | 63 | 4,833.60 | 76.72 |
| BUPRENORPHINE | 9064 | 20 | 158.05 | 7.9 |
| OXYCODONE | 9143 | 70 | 9,268.32 | 132.4 |
| HYDROMORPHONE | 9150 | 59 | 768.8 | 13.03 |
| HYDROCODONE | 9193 | 74 | 13,990.48 | 189.06 |
| MEPERIDINE (PETHIDINE) | 9230 | 57 | 790.56 | 13.87 |
| METHADONE | 9250B | 24 | 642.27 | 26.76 |
| MORPHINE | 9300 | 76 | 11,384.84 | 149.8 |
| OPIUM POWDERED | 9639 | 21 | 60.48 | 2.88 |
| OXYMORPHONE | 9652 | 9 | 106.74 | 11.86 |
| ALFENTANIL | 9737 | 3 | 0.23 | 0.08 |
| REMIFENTANIL | 9739 | 9 | 2.2 | 0.24 |
| SUFENTANIL BASE | 9740 | 10 | 0.13 | 0.01 |
| TAPENTADOL | 9780 | 15 | 1,565 | 104.33 |
| FENTANYL BASE | 9801 | 75 | 229.06 | 3.05 |

STATE:SOUTH DAKOTA BUSINESS ACTIVITY:C - PRACTITIONERS

| DRUG NAME | DRUG CODE | BUYERS | TOTAL GRAMS | AVG GRAMS |
|---|---|---|---|---|
| PENTOBARBITAL (SCHEDULE 2) | 2270 | 17 | 25,335.10 | 1,490.30 |
| COCAINE | 9041L | 1 | 0.14 | 0.14 |
| CODEINE | 9050 | 5 | 125.98 | 25.2 |
| BUPRENORPHINE | 9064 | 64 | 2.06 | 0.03 |
| OXYCODONE | 9143 | 6 | 894.19 | 149.03 |
| HYDROMORPHONE | 9150 | 36 | 71.31 | 1.98 |
| HYDROCODONE | 9193 | 29 | 53.84 | 1.86 |
| MEPERIDINE (PETHIDINE) | 9230 | 13 | 187.15 | 14.4 |
| METHADONE | 9250B | 1 | 0.36 | 0.36 |
| MORPHINE | 9300 | 27 | 277.14 | 10.26 |
| OPIUM POWDERED | 9639 | 1 | 0.72 | 0.72 |
| OXYMORPHONE | 9652 | 1 | 15.39 | 15.39 |
| REMIFENTANIL | 9739 | 2 | 0.14 | 0.07 |
| FENTANYL BASE | 9801 | 41 | 5.53 | 0.13 |

STATE:SOUTH DAKOTA BUSINESS ACTIVITY:D - TEACHING INSTITUTIONS

| DRUG NAME | DRUG CODE | BUYERS | TOTAL GRAMS | AVG GRAMS |
|---|---|---|---|---|
| AMPHETAMINE | 1100 | 1 | 0.73 | 0.73 |
| PENTOBARBITAL (SCHEDULE 2) | 2270 | 1 | 177.72 | 177.72 |
| BUPRENORPHINE | 9064 | 1 | 0 | 0 |

STATE:SOUTH DAKOTA BUSINESS ACTIVITY:M - MID-LEVEL PRACTITIONERS

| DRUG NAME | DRUG CODE | BUYERS | TOTAL GRAMS | AVG GRAMS |
|---|---|---|---|---|
| HYDROMORPHONE | 9150 | 3 | 0.27 | 0.09 |
| MORPHINE | 9300 | 4 | 1.8 | 0.45 |
| FENTANYL BASE | 9801 | 4 | 0.15 | 0.04 |

STATE:SOUTH DAKOTA BUSINESS ACTIVITY:N-U NARCOTIC TREATMENT PROGRAMS

| DRUG NAME | DRUG CODE | BUYERS | TOTAL GRAMS | AVG GRAMS |
|---|---|---|---|---|
| METHADONE | 9250B | 1 | 3,498.89 | 3,498.89 |

STATE:TENNESSEE BUSINESS ACTIVITY:A - PHARMACIES

| DRUG NAME | DRUG CODE | BUYERS | TOTAL GRAMS | AVG GRAMS |
|---|---|---|---|---|
| AMPHETAMINE | 1100 | 1,586 | 493,655.62 | 311.26 |
| DL-METHAMPHETAMINE RACEMIC BASE | 1105B | 3 | 2.81 | 0.94 |
| D-METHAMPHETAMINE | 1105D | 13 | 45.39 | 3.49 |
| LISDEXAMFETAMINE | 1205 | 1,537 | 284,507.29 | 185.11 |
| METHYLPHENIDATE (DL;D;L;ISOMERS) | 1724 | 1,568 | 338,449.26 | 215.85 |
| BARBITURIC ACID DERIVIATIVE OR SALT [PER 21C | 2100 | 7 | 45 | 6.43 |
| BUTALBITAL | 2165 | 901 | 29,065 | 32.26 |
| COCAINE | 9041L | 6 | 10.25 | 1.71 |
| CODEINE | 9050 | 1,540 | 405,872.34 | 263.55 |
| BUPRENORPHINE | 9064 | 1,448 | 122,921.79 | 84.89 |
| DIHYDROCODEINE | 9120 | 310 | 4,123.10 | 13.3 |
| OXYCODONE | 9143 | 1,599 | 1,939,796.93 | 1,213.13 |
| HYDROMORPHONE | 9150 | 1,385 | 23,127.10 | 16.7 |
| HYDROCODONE | 9193 | 1,605 | 1,029,704.64 | 641.56 |
| LEVORPHANOL | 9220L | 3 | 0.93 | 0.31 |
| MEPERIDINE (PETHIDINE) | 9230 | 1,100 | 32,030.78 | 29.12 |
| METHADONE | 9250B | 1,131 | 49,744.29 | 43.98 |
| MORPHINE | 9300 | 1,580 | 700,962.98 | 443.65 |
| OPIUM POWDERED | 9639 | 126 | 407.88 | 3.24 |
| OXYMORPHONE | 9652 | 1,267 | 116,415.75 | 91.88 |
| REMIFENTANIL | 9739 | 1 | 0.04 | 0.04 |
| SUFENTANIL BASE | 9740 | 2 | 0.05 | 0.03 |
| TAPENTADOL | 9780 | 1,198 | 141,460.50 | 118.08 |

| | | | | |
|---|---|---|---|---|
| FENTANYL BASE | 9801 | 1,541 | 8,475.65 | 5.5 |

STATE:TENNESSEE BUSINESS ACTIVITY:B - HOSPITALS

| DRUG NAME | DRUG CODE | BUYERS | TOTAL GRAMS | AVG GRAMS |
|---|---|---|---|---|
| AMPHETAMINE | 1100 | 90 | 5,080.67 | 56.45 |
| LISDEXAMFETAMINE | 1205 | 42 | 2,374.83 | 56.54 |
| METHYLPHENIDATE (DL;D;L;ISOMERS) | 1724 | 111 | 5,531.98 | 49.84 |
| GAMMA HYDROXYBUTYRIC ACID(FDA APPROVED | 2012 | 1 | 2,303.42 | 2,303.42 |
| AMOBARBITAL (SCHEDULE 2) | 2125 | 3 | 10.94 | 3.65 |
| BUTALBITAL | 2165 | 14 | 125 | 8.93 |
| PENTOBARBITAL (SCHEDULE 2) | 2270 | 18 | 2,974.35 | 165.24 |
| COCAINE | 9041L | 88 | 544.72 | 6.19 |
| CODEINE | 9050 | 162 | 4,774.50 | 29.47 |
| BUPRENORPHINE | 9064 | 112 | 1,436.57 | 12.83 |
| DIHYDROCODEINE | 9120 | 3 | 6.41 | 2.14 |
| OXYCODONE | 9143 | 185 | 111,737.25 | 603.99 |
| HYDROMORPHONE | 9150 | 179 | 5,637.22 | 31.49 |
| HYDROCODONE | 9193 | 197 | 65,570.71 | 332.85 |
| MEPERIDINE (PETHIDINE) | 9230 | 146 | 4,600.39 | 31.51 |
| METHADONE | 9250B | 142 | 4,081.28 | 28.74 |
| MORPHINE | 9300 | 190 | 62,284.35 | 327.81 |
| OPIUM POWDERED | 9639 | 102 | 494.64 | 4.85 |
| OXYMORPHONE | 9652 | 46 | 869.59 | 18.9 |
| ALFENTANIL | 9737 | 5 | 1.98 | 0.4 |
| REMIFENTANIL | 9739 | 46 | 19.15 | 0.42 |
| SUFENTANIL BASE | 9740 | 49 | 1.46 | 0.03 |
| TAPENTADOL | 9780 | 40 | 1,526 | 38.15 |
| FENTANYL BASE | 9801 | 177 | 1,089.06 | 6.15 |

STATE:TENNESSEE BUSINESS ACTIVITY:C - PRACTITIONERS

| DRUG NAME | DRUG CODE | BUYERS | TOTAL GRAMS | AVG GRAMS |
|---|---|---|---|---|
| AMPHETAMINE | 1100 | 9 | 546.42 | 60.71 |
| LISDEXAMFETAMINE | 1205 | 8 | 1,077 | 134.63 |
| METHYLPHENIDATE (DL;D;L;ISOMERS) | 1724 | 4 | 608.84 | 152.21 |
| PENTOBARBITAL (SCHEDULE 2) | 2270 | 144 | 107,101.10 | 743.76 |
| COCAINE | 9041L | 22 | 208.13 | 9.46 |
| CODEINE | 9050 | 41 | 1,505.71 | 36.72 |
| BUPRENORPHINE | 9064 | 504 | 32.05 | 0.06 |
| DIHYDROCODEINE | 9120 | 109 | 68.47 | 0.63 |
| OXYCODONE | 9143 | 77 | 1,949.99 | 25.32 |
| HYDROMORPHONE | 9150 | 261 | 220.31 | 0.84 |
| HYDROCODONE | 9193 | 288 | 1,524.57 | 5.29 |
| MEPERIDINE (PETHIDINE) | 9230 | 151 | 978.8 | 6.48 |
| METHADONE | 9250B | 7 | 6.62 | 0.95 |
| MORPHINE | 9300 | 188 | 678.09 | 3.61 |
| OPIUM POWDERED | 9639 | 8 | 56.16 | 7.02 |
| OXYMORPHONE | 9652 | 6 | 2.32 | 0.39 |
| NOROXYMORPHONE | 9668 | 2 | 3.03 | 1.51 |
| ALFENTANIL | 9737 | 1 | 1.96 | 1.96 |
| REMIFENTANIL | 9739 | 10 | 1.54 | 0.15 |
| SUFENTANIL BASE | 9740 | 5 | 0.07 | 0.01 |
| TAPENTADOL | 9780 | 1 | 30 | 30 |
| FENTANYL BASE | 9801 | 411 | 53.84 | 0.13 |

STATE:TENNESSEE BUSINESS ACTIVITY:D - TEACHING INSTITUTIONS

| DRUG NAME | DRUG CODE | BUYERS | TOTAL GRAMS | AVG GRAMS |
|---|---|---|---|---|
| PENTOBARBITAL (SCHEDULE 2) | 2270 | 1 | 888.62 | 888.62 |
| BUPRENORPHINE | 9064 | 1 | 0.07 | 0.07 |

STATE:TENNESSEE BUSINESS ACTIVITY:M - MID-LEVEL PRACTITIONERS

| DRUG NAME | DRUG CODE | BUYERS | TOTAL GRAMS | AVG GRAMS |
|---|---|---|---|---|
| PENTOBARBITAL (SCHEDULE 2) | 2270 | 29 | 35,223.05 | 1,214.59 |
| CODEINE | 9050 | 9 | 371.73 | 41.3 |
| DIHYDROCODEINE | 9120 | 83 | 45.59 | 0.55 |
| HYDROCODONE | 9193 | 2 | 1.51 | 0.76 |
| MEPERIDINE (PETHIDINE) | 9230 | 8 | 17.54 | 2.19 |
| MORPHINE | 9300 | 70 | 33.64 | 0.48 |
| FENTANYL BASE | 9801 | 66 | 1.31 | 0.02 |

STATE:TENNESSEE BUSINESS ACTIVITY:N-U NARCOTIC TREATMENT PROGRAMS

| DRUG NAME | DRUG CODE | BUYERS | TOTAL GRAMS | AVG GRAMS |
|---|---|---|---|---|
| BUPRENORPHINE | 9064 | 11 | 1,031.16 | 93.74 |
| METHADONE | 9250B | 12 | 150,377.71 | 12,531.48 |

STATE:TEXAS BUSINESS ACTIVITY:A - PHARMACIES

| DRUG NAME | DRUG CODE | BUYERS | TOTAL GRAMS | AVG GRAMS |
|---|---|---|---|---|
| AMPHETAMINE | 1100 | 4,698 | 1,500,008.67 | 319.29 |
| DL-METHAMPHETAMINE RACEMIC BASE | 1105B | 61 | 166.37 | 2.73 |
| D-METHAMPHETAMINE | 1105D | 121 | 307.3 | 2.54 |
| LISDEXAMFETAMINE | 1205 | 4,555 | 1,078,296.98 | 236.73 |
| METHYLPHENIDATE (DL;D;L;ISOMERS) | 1724 | 4,697 | 1,302,051.19 | 277.21 |
| BARBITURIC ACID DERIVIATIVE OR SALT [PER 21C | 2100 | 62 | 640 | 10.32 |
| BUTALBITAL | 2165 | 3,048 | 132,865 | 43.59 |
| PENTOBARBITAL (SCHEDULE 2) | 2270 | 1 | 7.75 | 7.75 |
| SECOBARBITAL (SCHEDULE 2) | 2315 | 6 | 64.09 | 10.68 |
| COCAINE | 9041L | 10 | 47.03 | 4.7 |
| CODEINE | 9050 | 5,080 | 3,967,977.36 | 781.1 |
| BUPRENORPHINE | 9064 | 4,346 | 96,208.64 | 22.14 |
| DIHYDROCODEINE | 9120 | 1,557 | 9,636.94 | 6.19 |
| OXYCODONE | 9143 | 4,594 | 1,267,597.71 | 275.92 |

| | | | | |
|---|---|---|---|---|
| HYDROMORPHONE | 9150 | 3,915 | 87,382.22 | 22.32 |
| HYDROCODONE | 9193 | 5,069 | 2,698,345.92 | 532.32 |
| LEVORPHANOL | 9220L | 97 | 126.18 | 1.3 |
| MEPERIDINE (PETHIDINE) | 9230 | 1,234 | 23,514.81 | 19.06 |
| METHADONE | 9250B | 3,558 | 166,329.51 | 46.75 |
| MORPHINE | 9300 | 4,290 | 802,211.73 | 187 |
| OPIUM TINCTURE (LAUDANUM) | 9630 | 5 | 20.06 | 4.01 |
| OPIUM POWDERED | 9639 | 162 | 317.88 | 1.96 |
| OXYMORPHONE | 9652 | 2,416 | 63,876.55 | 26.44 |
| ALFENTANIL | 9737 | 1 | 0.17 | 0.17 |
| REMIFENTANIL | 9739 | 3 | 0.06 | 0.02 |
| SUFENTANIL BASE | 9740 | 9 | 2.73 | 0.3 |
| TAPENTADOL | 9780 | 3,037 | 426,793.50 | 140.53 |
| FENTANYL BASE | 9801 | 4,406 | 23,450.25 | 5.32 |

STATE:TEXAS BUSINESS ACTIVITY:B - HOSPITALS

| DRUG NAME | DRUG CODE | BUYERS | TOTAL GRAMS | AVG GRAMS |
|---|---|---|---|---|
| AMPHETAMINE | 1100 | 294 | 34,061.22 | 115.85 |
| DL-METHAMPHETAMINE RACEMIC BASE | 1105B | 1 | 6.83 | 6.83 |
| LISDEXAMFETAMINE | 1205 | 118 | 8,227.52 | 69.72 |
| METHYLPHENIDATE (DL;D;L;ISOMERS) | 1724 | 432 | 51,365.27 | 118.9 |
| GAMMA HYDROXYBUTYRIC ACID(FDA APPROVED | 2012 | 1 | 2,452.03 | 2,452.03 |
| BARBITURIC ACID DERIVIATIVE OR SALT [PER 21C | 2100 | 1 | 5 | 5 |
| AMOBARBITAL (SCHEDULE 2) | 2125 | 15 | 72.46 | 4.83 |
| BUTALBITAL | 2165 | 52 | 754.85 | 14.52 |
| PENTOBARBITAL (SCHEDULE 2) | 2270 | 33 | 6,521.52 | 197.62 |
| SECOBARBITAL (SCHEDULE 2) | 2315 | 1 | 9.16 | 9.16 |
| COCAINE | 9041L | 298 | 1,452.91 | 4.88 |
| CODEINE | 9050 | 935 | 132,176.77 | 141.37 |
| BUPRENORPHINE | 9064 | 296 | 3,505.22 | 11.84 |
| DIHYDROCODEINE | 9120 | 2 | 13.89 | 6.94 |
| OXYCODONE | 9143 | 754 | 68,471.40 | 90.81 |

| | | | | |
|---|---|---|---|---|
| HYDROMORPHONE | 9150 | 1,133 | 16,982.03 | 14.99 |
| HYDROCODONE | 9193 | 1,202 | 184,384.61 | 153.4 |
| MEPERIDINE (PETHIDINE) | 9230 | 797 | 29,373.99 | 36.86 |
| METHADONE | 9250B | 509 | 14,261.75 | 28.02 |
| MORPHINE | 9300 | 1,312 | 114,903.46 | 87.58 |
| OPIUM TINCTURE (LAUDANUM) | 9630 | 1 | 1.18 | 1.18 |
| OPIUM POWDERED | 9639 | 322 | 1,771.20 | 5.5 |
| OXYMORPHONE | 9652 | 57 | 901.7 | 15.82 |
| NOROXYMORPHONE | 9668 | 1 | 2.74 | 2.74 |
| ALFENTANIL | 9737 | 58 | 21.68 | 0.37 |
| REMIFENTANIL | 9739 | 182 | 108.14 | 0.59 |
| SUFENTANIL BASE | 9740 | 204 | 4.19 | 0.02 |
| TAPENTADOL | 9780 | 107 | 5,407.79 | 50.54 |
| FENTANYL BASE | 9801 | 1,263 | 4,027.56 | 3.19 |

STATE:TEXAS BUSINESS ACTIVITY:C - PRACTITIONERS

| DRUG NAME | DRUG CODE | BUYERS | TOTAL GRAMS | AVG GRAMS |
|---|---|---|---|---|
| AMPHETAMINE | 1100 | 5 | 113.05 | 22.61 |
| METHYLPHENIDATE (DL;D;L;ISOMERS) | 1724 | 2 | 1.72 | 0.86 |
| GAMMA HYDROXYBUTYRIC ACID(FDA APPROVED | 2012 | 3 | 7,727.62 | 2,575.87 |
| PENTOBARBITAL (SCHEDULE 2) | 2270 | 243 | 211,320.33 | 869.63 |
| COCAINE | 9041L | 17 | 48.35 | 2.84 |
| CODEINE | 9050 | 364 | 3,370.96 | 9.26 |
| BUPRENORPHINE | 9064 | 1,833 | 632.92 | 0.35 |
| DIHYDROCODEINE | 9120 | 595 | 458.58 | 0.77 |
| OXYCODONE | 9143 | 38 | 4,001.53 | 105.3 |
| HYDROMORPHONE | 9150 | 772 | 410.47 | 0.53 |
| HYDROCODONE | 9193 | 882 | 1,940.50 | 2.2 |
| MEPERIDINE (PETHIDINE) | 9230 | 423 | 3,705.25 | 8.76 |
| METHADONE | 9250B | 28 | 57.06 | 2.04 |
| MORPHINE | 9300 | 569 | 897.93 | 1.58 |
| OXYMORPHONE | 9652 | 18 | 7.87 | 0.44 |

| | | | | |
|---|---|---|---|---|
| NOROXYMORPHONE | 9668 | 12 | 101.13 | 8.43 |
| ALFENTANIL | 9737 | 12 | 2.08 | 0.17 |
| REMIFENTANIL | 9739 | 26 | 1.1 | 0.04 |
| SUFENTANIL BASE | 9740 | 3 | 0.01 | 0 |
| TAPENTADOL | 9780 | 1 | 20 | 20 |
| FENTANYL BASE | 9801 | 1,310 | 114.74 | 0.09 |

STATE:TEXAS BUSINESS ACTIVITY:D - TEACHING INSTITUTIONS

| DRUG NAME | DRUG CODE | BUYERS | TOTAL GRAMS | AVG GRAMS |
|---|---|---|---|---|
| PENTOBARBITAL (SCHEDULE 2) | 2270 | 3 | 3,554.46 | 1,184.82 |
| BUPRENORPHINE | 9064 | 3 | 0.68 | 0.23 |
| HYDROMORPHONE | 9150 | 2 | 5.28 | 2.64 |
| HYDROCODONE | 9193 | 1 | 1.19 | 1.19 |
| MEPERIDINE (PETHIDINE) | 9230 | 2 | 10.89 | 5.45 |
| METHADONE | 9250B | 1 | 17.89 | 17.89 |
| MORPHINE | 9300 | 3 | 4.72 | 1.57 |
| OXYMORPHONE | 9652 | 1 | 0.8 | 0.8 |
| REMIFENTANIL | 9739 | 1 | 0.07 | 0.07 |
| FENTANYL BASE | 9801 | 3 | 2.41 | 0.8 |

STATE:TEXAS BUSINESS ACTIVITY:M - MID-LEVEL PRACTITIONERS

| DRUG NAME | DRUG CODE | BUYERS | TOTAL GRAMS | AVG GRAMS |
|---|---|---|---|---|
| PENTOBARBITAL (SCHEDULE 2) | 2270 | 172 | 338,577.81 | 1,968.48 |
| CODEINE | 9050 | 45 | 45.84 | 1.02 |
| DIHYDROCODEINE | 9120 | 217 | 146.16 | 0.67 |
| HYDROMORPHONE | 9150 | 41 | 5.92 | 0.14 |
| MEPERIDINE (PETHIDINE) | 9230 | 5 | 7.08 | 1.42 |
| MORPHINE | 9300 | 310 | 167.44 | 0.54 |
| FENTANYL BASE | 9801 | 533 | 35.33 | 0.07 |

STATISTICAL SUMMARY FOR RETAIL DRUG PURCHASES BY GRAMS WT

STATE:TEXAS BUSINESS ACTIVITY:N-U NARCOTIC TREATMENT PROGRAMS

| DRUG NAME | DRUG CODE | BUYERS | TOTAL GRAMS | AVG GRAMS |
|---|---|---|---|---|
| BUPRENORPHINE | 9064 | 22 | 685.64 | 31.17 |
| METHADONE | 9250B | 91 | 377,856.72 | 4,152.27 |

STATE:UTAH BUSINESS ACTIVITY:A - PHARMACIES

| DRUG NAME | DRUG CODE | BUYERS | TOTAL GRAMS | AVG GRAMS |
|---|---|---|---|---|
| AMPHETAMINE | 1100 | 539 | 295,355.68 | 547.97 |
| DL-METHAMPHETAMINE RACEMIC BASE | 1105B | 3 | 2.01 | 0.67 |
| D-METHAMPHETAMINE | 1105D | 30 | 129.75 | 4.32 |
| LISDEXAMFETAMINE | 1205 | 520 | 79,338.44 | 152.57 |
| METHYLPHENIDATE (DL;D;L;ISOMERS) | 1724 | 532 | 180,863.75 | 339.97 |
| BUTALBITAL | 2165 | 411 | 19,495 | 47.43 |
| COCAINE | 9041L | 5 | 2.89 | 0.58 |
| CODEINE | 9050 | 533 | 68,901.99 | 129.27 |
| BUPRENORPHINE | 9064 | 523 | 34,540.58 | 66.04 |
| OXYCODONE | 9143 | 539 | 714,822.92 | 1,326.20 |
| HYDROMORPHONE | 9150 | 514 | 10,115.97 | 19.68 |
| HYDROCODONE | 9193 | 540 | 245,902.30 | 455.37 |
| LEVORPHANOL | 9220L | 2 | 0.35 | 0.17 |
| MEPERIDINE (PETHIDINE) | 9230 | 292 | 7,061.48 | 24.18 |
| METHADONE | 9250B | 506 | 45,502.42 | 89.93 |
| MORPHINE | 9300 | 531 | 203,934.91 | 384.06 |
| OPIUM POWDERED | 9639 | 19 | 37.8 | 1.99 |
| OXYMORPHONE | 9652 | 444 | 20,921.13 | 47.12 |
| REMIFENTANIL | 9739 | 5 | 1.09 | 0.22 |
| SUFENTANIL BASE | 9740 | 3 | 0.01 | 0 |
| TAPENTADOL | 9780 | 420 | 62,248 | 148.21 |
| FENTANYL BASE | 9801 | 528 | 3,595.48 | 6.81 |

STATISTICAL SUMMARY FOR RETAIL DRUG PURCHASES BY GRAMS WT

STATE:UTAH BUSINESS ACTIVITY:B - HOSPITALS

| DRUG NAME | DRUG CODE | BUYERS | TOTAL GRAMS | AVG GRAMS |
|---|---|---|---|---|
| AMPHETAMINE | 1100 | 33 | 4,828.74 | 146.33 |
| LISDEXAMFETAMINE | 1205 | 17 | 864.84 | 50.87 |
| METHYLPHENIDATE (DL;D;L;ISOMERS) | 1724 | 42 | 4,591.80 | 109.33 |
| AMOBARBITAL (SCHEDULE 2) | 2125 | 3 | 22.33 | 7.44 |
| BUTALBITAL | 2165 | 1 | 5 | 5 |
| PENTOBARBITAL (SCHEDULE 2) | 2270 | 3 | 459.8 | 153.27 |
| COCAINE | 9041L | 52 | 428.4 | 8.24 |
| CODEINE | 9050 | 43 | 1,777.27 | 41.33 |
| BUPRENORPHINE | 9064 | 42 | 592.28 | 14.1 |
| OXYCODONE | 9143 | 85 | 32,085.09 | 377.47 |
| HYDROMORPHONE | 9150 | 80 | 1,780.80 | 22.26 |
| HYDROCODONE | 9193 | 89 | 14,815.51 | 166.47 |
| MEPERIDINE (PETHIDINE) | 9230 | 76 | 1,722.74 | 22.67 |
| METHADONE | 9250B | 56 | 1,650.88 | 29.48 |
| MORPHINE | 9300 | 90 | 19,529.76 | 217 |
| OPIUM POWDERED | 9639 | 36 | 209.52 | 5.82 |
| OXYMORPHONE | 9652 | 10 | 74.4 | 7.44 |
| ALFENTANIL | 9737 | 6 | 1.88 | 0.31 |
| REMIFENTANIL | 9739 | 39 | 64.99 | 1.67 |
| SUFENTANIL BASE | 9740 | 26 | 0.86 | 0.03 |
| TAPENTADOL | 9780 | 20 | 2,644.50 | 132.23 |
| FENTANYL BASE | 9801 | 98 | 308.88 | 3.15 |

STATE:UTAH BUSINESS ACTIVITY:C - PRACTITIONERS

| DRUG NAME | DRUG CODE | BUYERS | TOTAL GRAMS | AVG GRAMS |
|---|---|---|---|---|
| AMPHETAMINE | 1100 | 1 | 3.75 | 3.75 |
| METHYLPHENIDATE (DL;D;L;ISOMERS) | 1724 | 1 | 33.68 | 33.68 |
| PENTOBARBITAL (SCHEDULE 2) | 2270 | 39 | 33,680.67 | 863.61 |

| | | | | |
|---|---|---|---|---|
| COCAINE | 9041L | 3 | 30.28 | 10.09 |
| CODEINE | 9050 | 11 | 226.32 | 20.57 |
| BUPRENORPHINE | 9064 | 167 | 18.22 | 0.11 |
| OXYCODONE | 9143 | 10 | 1,665.19 | 166.52 |
| HYDROMORPHONE | 9150 | 87 | 66.73 | 0.77 |
| HYDROCODONE | 9193 | 77 | 305.3 | 3.96 |
| MEPERIDINE (PETHIDINE) | 9230 | 33 | 294.24 | 8.92 |
| METHADONE | 9250B | 9 | 6.8 | 0.76 |
| MORPHINE | 9300 | 99 | 96.12 | 0.97 |
| OXYMORPHONE | 9652 | 2 | 0.08 | 0.04 |
| NOROXYMORPHONE | 9668 | 1 | 1.53 | 1.53 |
| REMIFENTANIL | 9739 | 5 | 0.07 | 0.01 |
| FENTANYL BASE | 9801 | 186 | 13.48 | 0.07 |

STATE:UTAH BUSINESS ACTIVITY:D - TEACHING INSTITUTIONS

| DRUG NAME | DRUG CODE | BUYERS | TOTAL GRAMS | AVG GRAMS |
|---|---|---|---|---|
| BUPRENORPHINE | 9064 | 2 | 0.01 | 0 |
| HYDROMORPHONE | 9150 | 1 | 0.07 | 0.07 |

STATE:UTAH BUSINESS ACTIVITY:M - MID-LEVEL PRACTITIONERS

| DRUG NAME | DRUG CODE | BUYERS | TOTAL GRAMS | AVG GRAMS |
|---|---|---|---|---|
| PENTOBARBITAL (SCHEDULE 2) | 2270 | 11 | 10,504.80 | 954.98 |
| CODEINE | 9050 | 1 | 22.1 | 22.1 |
| BUPRENORPHINE | 9064 | 1 | 0.01 | 0.01 |
| MEPERIDINE (PETHIDINE) | 9230 | 4 | 24.4 | 6.1 |
| MORPHINE | 9300 | 1 | 0.03 | 0.03 |
| REMIFENTANIL | 9739 | 1 | 0.08 | 0.08 |
| FENTANYL BASE | 9801 | 10 | 0.62 | 0.06 |

STATE:UTAH BUSINESS ACTIVITY:N-U NARCOTIC TREATMENT PROGRAMS

| DRUG NAME | DRUG CODE | BUYERS | TOTAL GRAMS | AVG GRAMS |
|---|---|---|---|---|
| BUPRENORPHINE | 9064 | 10 | 411.18 | 41.12 |
| METHADONE | 9250B | 13 | 63,416.54 | 4,878.20 |

STATE:VERMONT BUSINESS ACTIVITY:A - PHARMACIES

| DRUG NAME | DRUG CODE | BUYERS | TOTAL GRAMS | AVG GRAMS |
|---|---|---|---|---|
| AMPHETAMINE | 1100 | 147 | 38,068.21 | 258.97 |
| D-METHAMPHETAMINE | 1105D | 2 | 5.62 | 2.81 |
| LISDEXAMFETAMINE | 1205 | 146 | 25,301.12 | 173.3 |
| METHYLPHENIDATE (DL;D;L;ISOMERS) | 1724 | 147 | 73,597.17 | 500.66 |
| BARBITURIC ACID DERIVIATIVE OR SALT [PER 21C | 2100 | 1 | 5 | 5 |
| BUTALBITAL | 2165 | 55 | 1,140 | 20.73 |
| SECOBARBITAL (SCHEDULE 2) | 2315 | 3 | 109.86 | 36.62 |
| COCAINE | 9041L | 1 | 0.14 | 0.14 |
| CODEINE | 9050 | 141 | 14,676.33 | 104.09 |
| BUPRENORPHINE | 9064 | 147 | 16,580.54 | 112.79 |
| OXYCODONE | 9143 | 148 | 95,881.05 | 647.84 |
| HYDROMORPHONE | 9150 | 147 | 4,613.12 | 31.38 |
| HYDROCODONE | 9193 | 149 | 22,198.47 | 148.98 |
| LEVORPHANOL | 9220L | 1 | 0.23 | 0.23 |
| MEPERIDINE (PETHIDINE) | 9230 | 22 | 485.21 | 22.05 |
| METHADONE | 9250B | 141 | 15,853.33 | 112.43 |
| MORPHINE | 9300 | 149 | 39,536.81 | 265.35 |
| OPIUM TINCTURE (LAUDANUM) | 9630 | 1 | 2.36 | 2.36 |
| OPIUM POWDERED | 9639 | 13 | 61.92 | 4.76 |
| OXYMORPHONE | 9652 | 59 | 1,792.94 | 30.39 |
| TAPENTADOL | 9780 | 64 | 5,358 | 83.72 |
| FENTANYL BASE | 9801 | 145 | 894.35 | 6.17 |

STATISTICAL SUMMARY FOR RETAIL DRUG PURCHASES BY GRAMS WT

STATE:VERMONT BUSINESS ACTIVITY:B - HOSPITALS

| DRUG NAME | DRUG CODE | BUYERS | TOTAL GRAMS | AVG GRAMS |
|---|---|---|---|---|
| AMPHETAMINE | 1100 | 12 | 976.72 | 81.39 |
| LISDEXAMFETAMINE | 1205 | 7 | 315.64 | 45.09 |
| METHYLPHENIDATE (DL;D;L;ISOMERS) | 1724 | 15 | 2,126.63 | 141.78 |
| AMOBARBITAL (SCHEDULE 2) | 2125 | 1 | 1.82 | 1.82 |
| PENTOBARBITAL (SCHEDULE 2) | 2270 | 1 | 63.8 | 63.8 |
| COCAINE | 9041L | 8 | 30.71 | 3.84 |
| CODEINE | 9050 | 15 | 502.91 | 33.53 |
| BUPRENORPHINE | 9064 | 16 | 1,144.30 | 71.52 |
| OXYCODONE | 9143 | 18 | 8,037.27 | 446.51 |
| HYDROMORPHONE | 9150 | 17 | 794.82 | 46.75 |
| HYDROCODONE | 9193 | 17 | 1,107.47 | 65.15 |
| LEVORPHANOL | 9220L | 1 | 1.74 | 1.74 |
| MEPERIDINE (PETHIDINE) | 9230 | 14 | 945.58 | 67.54 |
| METHADONE | 9250B | 16 | 840.47 | 52.53 |
| MORPHINE | 9300 | 17 | 5,593.94 | 329.06 |
| OPIUM POWDERED | 9639 | 12 | 91.08 | 7.59 |
| OXYMORPHONE | 9652 | 1 | 8.92 | 8.92 |
| ALFENTANIL | 9737 | 1 | 0.02 | 0.02 |
| REMIFENTANIL | 9739 | 4 | 2.15 | 0.54 |
| SUFENTANIL BASE | 9740 | 1 | 0.04 | 0.04 |
| TAPENTADOL | 9780 | 1 | 140 | 140 |
| FENTANYL BASE | 9801 | 16 | 66.85 | 4.18 |

STATE:VERMONT BUSINESS ACTIVITY:C - PRACTITIONERS

| DRUG NAME | DRUG CODE | BUYERS | TOTAL GRAMS | AVG GRAMS |
|---|---|---|---|---|
| PENTOBARBITAL (SCHEDULE 2) | 2270 | 69 | 43,072.26 | 624.24 |
| COCAINE | 9041L | 1 | 0.29 | 0.29 |
| CODEINE | 9050 | 3 | 7.74 | 2.58 |
| BUPRENORPHINE | 9064 | 69 | 8.47 | 0.12 |

| | | | | |
|---|---|---|---|---|
| OXYCODONE | 9143 | 1 | 3.59 | 3.59 |
| HYDROMORPHONE | 9150 | 50 | 18.41 | 0.37 |
| HYDROCODONE | 9193 | 19 | 33.07 | 1.74 |
| MEPERIDINE (PETHIDINE) | 9230 | 1 | 0.87 | 0.87 |
| MORPHINE | 9300 | 23 | 37.07 | 1.61 |
| OXYMORPHONE | 9652 | 1 | 0.09 | 0.09 |
| NOROXYMORPHONE | 9668 | 1 | 1.13 | 1.13 |
| REMIFENTANIL | 9739 | 1 | 0.02 | 0.02 |
| FENTANYL BASE | 9801 | 34 | 3.7 | 0.11 |

STATE:VERMONT BUSINESS ACTIVITY:D - TEACHING INSTITUTIONS

| DRUG NAME | DRUG CODE | BUYERS | TOTAL GRAMS | AVG GRAMS |
|---|---|---|---|---|
| D-METHAMPHETAMINE | 1105D | 1 | 4.02 | 4.02 |
| PENTOBARBITAL (SCHEDULE 2) | 2270 | 1 | 94.33 | 94.33 |
| BUPRENORPHINE | 9064 | 1 | 0.01 | 0.01 |
| FENTANYL BASE | 9801 | 1 | 0.58 | 0.58 |

STATE:VERMONT BUSINESS ACTIVITY:M - MID-LEVEL PRACTITIONERS

| DRUG NAME | DRUG CODE | BUYERS | TOTAL GRAMS | AVG GRAMS |
|---|---|---|---|---|
| PENTOBARBITAL (SCHEDULE 2) | 2270 | 4 | 888.62 | 222.15 |
| CODEINE | 9050 | 8 | 109.93 | 13.74 |

STATE:VERMONT BUSINESS ACTIVITY:N-U NARCOTIC TREATMENT PROGRAMS

| DRUG NAME | DRUG CODE | BUYERS | TOTAL GRAMS | AVG GRAMS |
|---|---|---|---|---|
| BUPRENORPHINE | 9064 | 9 | 3,804.89 | 422.77 |
| METHADONE | 9250B | 9 | 51,486.58 | 5,720.73 |

STATISTICAL SUMMARY FOR RETAIL DRUG PURCHASES BY GRAMS WT

STATE:VIRGIN ISLANDS BUSINESS ACTIVITY:A - PHARMACIES

| DRUG NAME | DRUG CODE | BUYERS | TOTAL GRAMS | AVG GRAMS |
|---|---|---|---|---|
| AMPHETAMINE | 1100 | 21 | 1,251.26 | 59.58 |
| LISDEXAMFETAMINE | 1205 | 11 | 295.41 | 26.86 |
| METHYLPHENIDATE (DL;D;L;ISOMERS) | 1724 | 19 | 798.84 | 42.04 |
| BUTALBITAL | 2165 | 6 | 115 | 19.17 |
| CODEINE | 9050 | 22 | 2,665.99 | 121.18 |
| BUPRENORPHINE | 9064 | 10 | 88.82 | 8.88 |
| OXYCODONE | 9143 | 22 | 2,571.34 | 116.88 |
| HYDROMORPHONE | 9150 | 15 | 72.42 | 4.83 |
| HYDROCODONE | 9193 | 22 | 1,359.29 | 61.79 |
| MEPERIDINE (PETHIDINE) | 9230 | 9 | 214.61 | 23.85 |
| METHADONE | 9250B | 7 | 165.02 | 23.57 |
| MORPHINE | 9300 | 15 | 364.57 | 24.3 |
| TAPENTADOL | 9780 | 2 | 44 | 22 |
| FENTANYL BASE | 9801 | 15 | 23.29 | 1.55 |

STATE:VIRGIN ISLANDS BUSINESS ACTIVITY:B - HOSPITALS

| DRUG NAME | DRUG CODE | BUYERS | TOTAL GRAMS | AVG GRAMS |
|---|---|---|---|---|
| METHYLPHENIDATE (DL;D;L;ISOMERS) | 1724 | 1 | 0.43 | 0.43 |
| CODEINE | 9050 | 2 | 137.03 | 68.51 |
| BUPRENORPHINE | 9064 | 2 | 0 | 0 |
| OXYCODONE | 9143 | 2 | 43.48 | 21.74 |
| HYDROMORPHONE | 9150 | 2 | 29.32 | 14.66 |
| HYDROCODONE | 9193 | 2 | 28.21 | 14.11 |
| MEPERIDINE (PETHIDINE) | 9230 | 1 | 15.8 | 15.8 |
| MORPHINE | 9300 | 2 | 43.72 | 21.86 |
| OPIUM POWDERED | 9639 | 1 | 1.8 | 1.8 |
| FENTANYL BASE | 9801 | 3 | 3.31 | 1.1 |

STATE:VIRGIN ISLANDS BUSINESS ACTIVITY:C - PRACTITIONERS

| DRUG NAME | DRUG CODE | BUYERS | TOTAL GRAMS | AVG GRAMS |
|---|---|---|---|---|
| CODEINE | 9050 | 2 | 6.63 | 3.32 |
| BUPRENORPHINE | 9064 | 4 | 0.14 | 0.04 |
| HYDROMORPHONE | 9150 | 2 | 0.21 | 0.11 |
| HYDROCODONE | 9193 | 1 | 0.61 | 0.61 |
| MEPERIDINE (PETHIDINE) | 9230 | 1 | 6.97 | 6.97 |
| MORPHINE | 9300 | 3 | 8.47 | 2.82 |
| FENTANYL BASE | 9801 | 4 | 0.1 | 0.03 |

STATE:VIRGINIA BUSINESS ACTIVITY:A - PHARMACIES

| DRUG NAME | DRUG CODE | BUYERS | TOTAL GRAMS | AVG GRAMS |
|---|---|---|---|---|
| AMPHETAMINE | 1100 | 1,606 | 663,016.71 | 412.84 |
| DL-METHAMPHETAMINE RACEMIC BASE | 1105B | 3 | 2.81 | 0.94 |
| D-METHAMPHETAMINE | 1105D | 60 | 217.72 | 3.63 |
| LISDEXAMFETAMINE | 1205 | 1,558 | 262,001.31 | 168.17 |
| METHYLPHENIDATE (DL;D;L;ISOMERS) | 1724 | 1,600 | 507,567.67 | 317.23 |
| BARBITURIC ACID DERIVIATIVE OR SALT [PER 21C | 2100 | 26 | 290 | 11.15 |
| BUTALBITAL | 2165 | 1,059 | 40,395 | 38.14 |
| SECOBARBITAL (SCHEDULE 2) | 2315 | 5 | 128.17 | 25.63 |
| NABILONE | 7379 | 3 | 1 | 0.33 |
| COCAINE | 9041L | 6 | 71.78 | 11.96 |
| CODEINE | 9050 | 1,590 | 229,983.78 | 144.64 |
| BUPRENORPHINE | 9064 | 1,496 | 73,246.31 | 48.96 |
| DIHYDROCODEINE | 9120 | 14 | 30.98 | 2.21 |
| OXYCODONE | 9143 | 1,621 | 1,311,307.06 | 808.95 |
| HYDROMORPHONE | 9150 | 1,567 | 46,660.16 | 29.78 |
| HYDROCODONE | 9193 | 1,620 | 474,154.50 | 292.69 |
| LEVORPHANOL | 9220L | 38 | 23.33 | 0.61 |
| MEPERIDINE (PETHIDINE) | 9230 | 875 | 24,397.21 | 27.88 |
| METHADONE | 9250B | 1,342 | 77,621.49 | 57.84 |

| | | | | |
|---|---|---|---|---|
| MORPHINE | 9300 | 1,584 | 322,152.27 | 203.38 |
| OPIUM TINCTURE (LAUDANUM) | 9630 | 5 | 17.7 | 3.54 |
| OPIUM POWDERED | 9639 | 108 | 261.72 | 2.42 |
| OXYMORPHONE | 9652 | 1,185 | 50,904.25 | 42.96 |
| REMIFENTANIL | 9739 | 1 | 0.03 | 0.03 |
| TAPENTADOL | 9780 | 1,280 | 192,481 | 150.38 |
| FENTANYL BASE | 9801 | 1,555 | 7,618.83 | 4.9 |

STATE:VIRGINIA BUSINESS ACTIVITY:B - HOSPITALS

| DRUG NAME | DRUG CODE | BUYERS | TOTAL GRAMS | AVG GRAMS |
|---|---|---|---|---|
| AMPHETAMINE | 1100 | 91 | 28,075.94 | 308.53 |
| LISDEXAMFETAMINE | 1205 | 43 | 7,031.43 | 163.52 |
| METHYLPHENIDATE (DL;D;L;ISOMERS) | 1724 | 108 | 34,169.10 | 316.38 |
| GAMMA HYDROXYBUTYRIC ACID(FDA APPROVED | 2012 | 1 | 7,281.79 | 7,281.79 |
| AMOBARBITAL (SCHEDULE 2) | 2125 | 3 | 12.76 | 4.25 |
| BUTALBITAL | 2165 | 7 | 50 | 7.14 |
| PENTOBARBITAL (SCHEDULE 2) | 2270 | 18 | 4,939.33 | 274.41 |
| COCAINE | 9041L | 101 | 526.47 | 5.21 |
| CODEINE | 9050 | 140 | 8,708.85 | 62.21 |
| BUPRENORPHINE | 9064 | 103 | 1,047.71 | 10.17 |
| OXYCODONE | 9143 | 175 | 81,371.22 | 464.98 |
| HYDROMORPHONE | 9150 | 183 | 7,654.08 | 41.83 |
| HYDROCODONE | 9193 | 173 | 25,859.08 | 149.47 |
| MEPERIDINE (PETHIDINE) | 9230 | 142 | 4,609.67 | 32.46 |
| METHADONE | 9250B | 112 | 4,837.32 | 43.19 |
| MORPHINE | 9300 | 191 | 40,595.36 | 212.54 |
| OPIUM POWDERED | 9639 | 79 | 446.04 | 5.65 |
| OXYMORPHONE | 9652 | 17 | 90.43 | 5.32 |
| ALFENTANIL | 9737 | 18 | 3.49 | 0.19 |
| REMIFENTANIL | 9739 | 51 | 24.06 | 0.47 |
| SUFENTANIL BASE | 9740 | 36 | 1.58 | 0.04 |
| TAPENTADOL | 9780 | 36 | 2,986.50 | 82.96 |

| | | | | |
|---|---|---|---|---|
| FENTANYL BASE | 9801 | 195 | 912.27 | 4.68 |

STATE:VIRGINIA BUSINESS ACTIVITY:C - PRACTITIONERS

| DRUG NAME | DRUG CODE | BUYERS | TOTAL GRAMS | AVG GRAMS |
|---|---|---|---|---|
| AMPHETAMINE | 1100 | 2 | 103.35 | 51.67 |
| LISDEXAMFETAMINE | 1205 | 1 | 142.04 | 142.04 |
| METHYLPHENIDATE (DL;D;L;ISOMERS) | 1724 | 2 | 324.22 | 162.11 |
| BUTALBITAL | 2165 | 1 | 5 | 5 |
| PENTOBARBITAL (SCHEDULE 2) | 2270 | 161 | 128,162.20 | 796.04 |
| COCAINE | 9041L | 23 | 105.56 | 4.59 |
| CODEINE | 9050 | 176 | 5,971.31 | 33.93 |
| BUPRENORPHINE | 9064 | 812 | 46.9 | 0.06 |
| OXYCODONE | 9143 | 42 | 1,721.53 | 40.99 |
| HYDROMORPHONE | 9150 | 380 | 244.11 | 0.64 |
| HYDROCODONE | 9193 | 239 | 2,297.38 | 9.61 |
| MEPERIDINE (PETHIDINE) | 9230 | 134 | 865.51 | 6.46 |
| METHADONE | 9250B | 10 | 158.85 | 15.88 |
| MORPHINE | 9300 | 224 | 597.67 | 2.67 |
| OPIUM POWDERED | 9639 | 1 | 8.64 | 8.64 |
| OXYMORPHONE | 9652 | 16 | 82.84 | 5.18 |
| ALFENTANIL | 9737 | 2 | 0.14 | 0.07 |
| REMIFENTANIL | 9739 | 9 | 0.8 | 0.09 |
| SUFENTANIL BASE | 9740 | 1 | 0 | 0 |
| TAPENTADOL | 9780 | 6 | 719.25 | 119.88 |
| FENTANYL BASE | 9801 | 467 | 63.18 | 0.14 |

STATE:VIRGINIA BUSINESS ACTIVITY:D - TEACHING INSTITUTIONS

| DRUG NAME | DRUG CODE | BUYERS | TOTAL GRAMS | AVG GRAMS |
|---|---|---|---|---|
| PENTOBARBITAL (SCHEDULE 2) | 2270 | 2 | 6,669.17 | 3,334.58 |
| COCAINE | 9041L | 2 | 1.18 | 0.59 |

| | | | | |
|---|---|---|---|---|
| CODEINE | 9050 | 2 | 298.71 | 149.36 |
| BUPRENORPHINE | 9064 | 1 | 0.54 | 0.54 |
| HYDROMORPHONE | 9150 | 2 | 9.68 | 4.84 |
| HYDROCODONE | 9193 | 1 | 2.37 | 2.37 |
| METHADONE | 9250B | 1 | 4.65 | 4.65 |
| MORPHINE | 9300 | 2 | 13.61 | 6.81 |
| OXYMORPHONE | 9652 | 1 | 0.61 | 0.61 |
| REMIFENTANIL | 9739 | 1 | 0.02 | 0.02 |
| FENTANYL BASE | 9801 | 1 | 0.75 | 0.75 |

STATE:VIRGINIA BUSINESS ACTIVITY:M - MID-LEVEL PRACTITIONERS

| DRUG NAME | DRUG CODE | BUYERS | TOTAL GRAMS | AVG GRAMS |
|---|---|---|---|---|
| PENTOBARBITAL (SCHEDULE 2) | 2270 | 42 | 37,055.25 | 882.27 |
| CODEINE | 9050 | 39 | 39.92 | 1.02 |
| MORPHINE | 9300 | 11 | 2.63 | 0.24 |
| FENTANYL BASE | 9801 | 14 | 0.18 | 0.01 |

STATE:VIRGINIA BUSINESS ACTIVITY:N-U NARCOTIC TREATMENT PROGRAMS

| DRUG NAME | DRUG CODE | BUYERS | TOTAL GRAMS | AVG GRAMS |
|---|---|---|---|---|
| BUPRENORPHINE | 9064 | 20 | 2,430.23 | 121.51 |
| METHADONE | 9250B | 36 | 208,977.45 | 5,804.93 |

STATE:WASHINGTON BUSINESS ACTIVITY:A - PHARMACIES

| DRUG NAME | DRUG CODE | BUYERS | TOTAL GRAMS | AVG GRAMS |
|---|---|---|---|---|
| AMPHETAMINE | 1100 | 1,263 | 403,754.32 | 319.68 |
| DL-METHAMPHETAMINE RACEMIC BASE | 1105B | 4 | 9.24 | 2.31 |
| D-METHAMPHETAMINE | 1105D | 95 | 367.15 | 3.86 |
| LISDEXAMFETAMINE | 1205 | 1,185 | 124,020.37 | 104.66 |

| | | | | |
|---|---|---|---|---|
| METHYLPHENIDATE (DL;D;L;ISOMERS) | 1724 | 1,263 | 417,744.69 | 330.76 |
| BARBITURIC ACID DERIVIATIVE OR SALT [PER 21C | 2100 | 2 | 35 | 17.5 |
| BUTALBITAL | 2165 | 787 | 29,245 | 37.16 |
| SECOBARBITAL (SCHEDULE 2) | 2315 | 18 | 439.44 | 24.41 |
| COCAINE | 9041L | 4 | 18.75 | 4.69 |
| CODEINE | 9050 | 1,282 | 250,322.10 | 195.26 |
| BUPRENORPHINE | 9064 | 1,224 | 61,939.21 | 50.6 |
| DIHYDROCODEINE | 9120 | 1 | 1.07 | 1.07 |
| OXYCODONE | 9143 | 1,283 | 1,204,886.74 | 939.12 |
| HYDROMORPHONE | 9150 | 1,248 | 40,942.65 | 32.81 |
| HYDROCODONE | 9193 | 1,295 | 546,627.49 | 422.11 |
| LEVORPHANOL | 9220L | 42 | 53.75 | 1.28 |
| MEPERIDINE (PETHIDINE) | 9230 | 543 | 13,519.03 | 24.9 |
| METHADONE | 9250B | 1,224 | 130,894.19 | 106.94 |
| MORPHINE | 9300 | 1,278 | 450,621.88 | 352.6 |
| OPIUM TINCTURE (LAUDANUM) | 9630 | 8 | 20.06 | 2.51 |
| OPIUM POWDERED | 9639 | 110 | 568.44 | 5.17 |
| OXYMORPHONE | 9652 | 793 | 16,439.04 | 20.73 |
| TAPENTADOL | 9780 | 812 | 76,382.50 | 94.07 |
| FENTANYL BASE | 9801 | 1,210 | 6,388.43 | 5.28 |

STATE:WASHINGTON BUSINESS ACTIVITY:B - HOSPITALS

| DRUG NAME | DRUG CODE | BUYERS | TOTAL GRAMS | AVG GRAMS |
|---|---|---|---|---|
| AMPHETAMINE | 1100 | 94 | 16,222.06 | 172.58 |
| LISDEXAMFETAMINE | 1205 | 35 | 3,343.15 | 95.52 |
| METHYLPHENIDATE (DL;D;L;ISOMERS) | 1724 | 104 | 25,448.04 | 244.69 |
| GAMMA HYDROXYBUTYRIC ACID(FDA APPROVED | 2012 | 1 | 3,269.38 | 3,269.38 |
| AMOBARBITAL (SCHEDULE 2) | 2125 | 8 | 48.76 | 6.09 |
| BUTALBITAL | 2165 | 17 | 300 | 17.65 |
| PENTOBARBITAL (SCHEDULE 2) | 2270 | 12 | 4,402.06 | 366.84 |
| SECOBARBITAL (SCHEDULE 2) | 2315 | 4 | 375.36 | 93.84 |
| COCAINE | 9041L | 70 | 525.43 | 7.51 |

| | | | | |
|---|---|---|---|---|
| CODEINE | 9050 | 177 | 12,436.35 | 70.26 |
| BUPRENORPHINE | 9064 | 88 | 2,324.14 | 26.41 |
| OXYCODONE | 9143 | 224 | 103,453.34 | 461.85 |
| HYDROMORPHONE | 9150 | 205 | 7,807.55 | 38.09 |
| HYDROCODONE | 9193 | 218 | 42,116.02 | 193.19 |
| LEVORPHANOL | 9220L | 2 | 1.04 | 0.52 |
| MEPERIDINE (PETHIDINE) | 9230 | 184 | 2,370.07 | 12.88 |
| METHADONE | 9250B | 120 | 12,153.11 | 101.28 |
| MORPHINE | 9300 | 227 | 58,864.58 | 259.32 |
| OPIUM POWDERED | 9639 | 73 | 739.8 | 10.13 |
| OXYMORPHONE | 9652 | 22 | 197.51 | 8.98 |
| ALFENTANIL | 9737 | 27 | 10.21 | 0.38 |
| REMIFENTANIL | 9739 | 58 | 47.16 | 0.81 |
| SUFENTANIL BASE | 9740 | 30 | 0.96 | 0.03 |
| TAPENTADOL | 9780 | 21 | 1,722.50 | 82.02 |
| FENTANYL BASE | 9801 | 237 | 906.04 | 3.82 |

STATE:WASHINGTON BUSINESS ACTIVITY:C - PRACTITIONERS

| DRUG NAME | DRUG CODE | BUYERS | TOTAL GRAMS | AVG GRAMS |
|---|---|---|---|---|
| AMPHETAMINE | 1100 | 2 | 1.9 | 0.95 |
| METHYLPHENIDATE (DL;D;L;ISOMERS) | 1724 | 1 | 45.22 | 45.22 |
| PENTOBARBITAL (SCHEDULE 2) | 2270 | 170 | 185,578.03 | 1,091.64 |
| COCAINE | 9041L | 3 | 1.57 | 0.52 |
| CODEINE | 9050 | 72 | 4,743.66 | 65.88 |
| BUPRENORPHINE | 9064 | 756 | 113.93 | 0.15 |
| OXYCODONE | 9143 | 34 | 195.92 | 5.76 |
| HYDROMORPHONE | 9150 | 529 | 270.01 | 0.51 |
| HYDROCODONE | 9193 | 387 | 1,118.74 | 2.89 |
| MEPERIDINE (PETHIDINE) | 9230 | 154 | 1,039.15 | 6.75 |
| METHADONE | 9250B | 31 | 313.93 | 10.13 |
| MORPHINE | 9300 | 316 | 596.83 | 1.89 |
| OPIUM POWDERED | 9639 | 2 | 1.44 | 0.72 |

| | | | | |
|---|---|---|---|---|
| OXYMORPHONE | 9652 | 37 | 6.97 | 0.19 |
| NOROXYMORPHONE | 9668 | 3 | 19.67 | 6.56 |
| ALFENTANIL | 9737 | 5 | 1.47 | 0.29 |
| REMIFENTANIL | 9739 | 46 | 2.38 | 0.05 |
| SUFENTANIL BASE | 9740 | 5 | 0.01 | 0 |
| FENTANYL BASE | 9801 | 667 | 61.59 | 0.09 |

STATE:WASHINGTON BUSINESS ACTIVITY:D - TEACHING INSTITUTIONS

| DRUG NAME | DRUG CODE | BUYERS | TOTAL GRAMS | AVG GRAMS |
|---|---|---|---|---|
| BUPRENORPHINE | 9064 | 1 | 0 | 0 |
| HYDROMORPHONE | 9150 | 1 | 0.04 | 0.04 |

STATE:WASHINGTON BUSINESS ACTIVITY:M - MID-LEVEL PRACTITIONERS

| DRUG NAME | DRUG CODE | BUYERS | TOTAL GRAMS | AVG GRAMS |
|---|---|---|---|---|
| PENTOBARBITAL (SCHEDULE 2) | 2270 | 18 | 22,846.05 | 1,269.22 |
| CODEINE | 9050 | 4 | 141.67 | 35.42 |
| OXYCODONE | 9143 | 2 | 7.62 | 3.81 |
| HYDROMORPHONE | 9150 | 5 | 0.46 | 0.09 |
| MEPERIDINE (PETHIDINE) | 9230 | 1 | 0.87 | 0.87 |
| METHADONE | 9250B | 1 | 1.79 | 1.79 |
| MORPHINE | 9300 | 2 | 2.35 | 1.17 |
| REMIFENTANIL | 9739 | 1 | 0.02 | 0.02 |
| FENTANYL BASE | 9801 | 13 | 0.24 | 0.02 |

STATE:WASHINGTON BUSINESS ACTIVITY:N-U NARCOTIC TREATMENT PROGRAMS

| DRUG NAME | DRUG CODE | BUYERS | TOTAL GRAMS | AVG GRAMS |
|---|---|---|---|---|
| BUPRENORPHINE | 9064 | 15 | 1,655.66 | 110.38 |
| METHADONE | 9250B | 23 | 254,596.33 | 11,069.41 |

STATISTICAL SUMMARY FOR RETAIL DRUG PURCHASES BY GRAMS WT

STATE:WEST VIRGINIA BUSINESS ACTIVITY:A - PHARMACIES

| DRUG NAME | DRUG CODE | BUYERS | TOTAL GRAMS | AVG GRAMS |
|---|---|---|---|---|
| AMPHETAMINE | 1100 | 561 | 95,457.05 | 170.16 |
| DL-METHAMPHETAMINE RACEMIC BASE | 1105B | 2 | 9.64 | 4.82 |
| D-METHAMPHETAMINE | 1105D | 2 | 2.01 | 1 |
| LISDEXAMFETAMINE | 1205 | 550 | 80,525.86 | 146.41 |
| METHYLPHENIDATE (DL;D;L;ISOMERS) | 1724 | 568 | 126,922.93 | 223.46 |
| BARBITURIC ACID DERIVIATIVE OR SALT [PER 21C | 2100 | 1 | 40 | 40 |
| BUTALBITAL | 2165 | 232 | 7,405 | 31.92 |
| SECOBARBITAL (SCHEDULE 2) | 2315 | 1 | 9.16 | 9.16 |
| CODEINE | 9050 | 563 | 122,616.29 | 217.79 |
| BUPRENORPHINE | 9064 | 501 | 54,196.35 | 108.18 |
| DIHYDROCODEINE | 9120 | 2 | 30.98 | 15.49 |
| OXYCODONE | 9143 | 577 | 407,342.79 | 705.97 |
| HYDROMORPHONE | 9150 | 437 | 5,665.06 | 12.96 |
| HYDROCODONE | 9193 | 577 | 260,633.62 | 451.7 |
| LEVORPHANOL | 9220L | 2 | 1.04 | 0.52 |
| MEPERIDINE (PETHIDINE) | 9230 | 121 | 1,848.89 | 15.28 |
| METHADONE | 9250B | 388 | 19,528.06 | 50.33 |
| MORPHINE | 9300 | 553 | 95,569.71 | 172.82 |
| OPIUM TINCTURE (LAUDANUM) | 9630 | 4 | 20.09 | 5.02 |
| OPIUM POWDERED | 9639 | 38 | 136.8 | 3.6 |
| OXYMORPHONE | 9652 | 330 | 16,532.75 | 50.1 |
| REMIFENTANIL | 9739 | 1 | 0.15 | 0.15 |
| TAPENTADOL | 9780 | 270 | 24,268.50 | 89.88 |
| FENTANYL BASE | 9801 | 533 | 2,356.22 | 4.42 |

STATE:WEST VIRGINIA BUSINESS ACTIVITY:B - HOSPITALS

| DRUG NAME | DRUG CODE | BUYERS | TOTAL GRAMS | AVG GRAMS |
|---|---|---|---|---|

| | | | | |
|---|---|---|---|---|
| AMPHETAMINE | 1100 | 36 | 1,275.47 | 35.43 |
| LISDEXAMFETAMINE | 1205 | 17 | 600.65 | 35.33 |
| METHYLPHENIDATE (DL;D;L;ISOMERS) | 1724 | 48 | 2,380.23 | 49.59 |
| AMOBARBITAL (SCHEDULE 2) | 2125 | 4 | 5.47 | 1.37 |
| BUTALBITAL | 2165 | 6 | 60 | 10 |
| PENTOBARBITAL (SCHEDULE 2) | 2270 | 2 | 159.5 | 79.75 |
| COCAINE | 9041L | 38 | 197.13 | 5.19 |
| CODEINE | 9050 | 72 | 3,161.62 | 43.91 |
| BUPRENORPHINE | 9064 | 52 | 2,248.93 | 43.25 |
| OXYCODONE | 9143 | 75 | 20,703.52 | 276.05 |
| HYDROMORPHONE | 9150 | 64 | 1,602.74 | 25.04 |
| HYDROCODONE | 9193 | 75 | 24,510.92 | 326.81 |
| MEPERIDINE (PETHIDINE) | 9230 | 55 | 1,089.57 | 19.81 |
| METHADONE | 9250B | 52 | 1,158.96 | 22.29 |
| MORPHINE | 9300 | 71 | 15,267.50 | 215.04 |
| OPIUM POWDERED | 9639 | 37 | 277.92 | 7.51 |
| OXYMORPHONE | 9652 | 8 | 34.61 | 4.33 |
| REMIFENTANIL | 9739 | 23 | 20.5 | 0.89 |
| SUFENTANIL BASE | 9740 | 9 | 0.21 | 0.02 |
| TAPENTADOL | 9780 | 8 | 87.5 | 10.94 |
| FENTANYL BASE | 9801 | 71 | 310.08 | 4.37 |

STATE:WEST VIRGINIA BUSINESS ACTIVITY:C - PRACTITIONERS

| DRUG NAME | DRUG CODE | BUYERS | TOTAL GRAMS | AVG GRAMS |
|---|---|---|---|---|
| PENTOBARBITAL (SCHEDULE 2) | 2270 | 36 | 31,713.66 | 880.93 |
| CODEINE | 9050 | 35 | 90.57 | 2.59 |
| BUPRENORPHINE | 9064 | 94 | 20.37 | 0.22 |
| DIHYDROCODEINE | 9120 | 2 | 0.43 | 0.21 |
| OXYCODONE | 9143 | 1 | 0.9 | 0.9 |
| HYDROMORPHONE | 9150 | 38 | 21.41 | 0.56 |
| HYDROCODONE | 9193 | 75 | 221.53 | 2.95 |
| MEPERIDINE (PETHIDINE) | 9230 | 15 | 290.86 | 19.39 |

| | | | | |
|---|---|---|---|---|
| MORPHINE | 9300 | 33 | 32.25 | 0.98 |
| REMIFENTANIL | 9739 | 1 | 0.22 | 0.22 |
| FENTANYL BASE | 9801 | 55 | 6.34 | 0.12 |

STATE:WEST VIRGINIA BUSINESS ACTIVITY:M - MID-LEVEL PRACTITIONERS

| DRUG NAME | DRUG CODE | BUYERS | TOTAL GRAMS | AVG GRAMS |
|---|---|---|---|---|
| PENTOBARBITAL (SCHEDULE 2) | 2270 | 17 | 9,417.67 | 553.98 |
| CODEINE | 9050 | 5 | 1.95 | 0.39 |
| MORPHINE | 9300 | 40 | 20.85 | 0.52 |
| FENTANYL BASE | 9801 | 43 | 0.87 | 0.02 |

STATE:WEST VIRGINIA BUSINESS ACTIVITY:N-U NARCOTIC TREATMENT PROGRAMS

| DRUG NAME | DRUG CODE | BUYERS | TOTAL GRAMS | AVG GRAMS |
|---|---|---|---|---|
| BUPRENORPHINE | 9064 | 7 | 1,743.54 | 249.08 |
| METHADONE | 9250B | 9 | 79,852.03 | 8,872.45 |

STATE:WISCONSIN BUSINESS ACTIVITY:A - PHARMACIES

| DRUG NAME | DRUG CODE | BUYERS | TOTAL GRAMS | AVG GRAMS |
|---|---|---|---|---|
| AMPHETAMINE | 1100 | 1,108 | 524,860.77 | 473.7 |
| DL-METHAMPHETAMINE RACEMIC BASE | 1105B | 10 | 69.92 | 6.99 |
| D-METHAMPHETAMINE | 1105D | 44 | 179.56 | 4.08 |
| LISDEXAMFETAMINE | 1205 | 1,075 | 237,036.03 | 220.5 |
| METHYLPHENIDATE (DL;D;L;ISOMERS) | 1724 | 1,104 | 406,169.95 | 367.91 |
| BARBITURIC ACID DERIVIATIVE OR SALT [PER 21C | 2100 | 2 | 20 | 10 |
| BUTALBITAL | 2165 | 558 | 16,295 | 29.2 |
| NABILONE | 7379 | 1 | 0.05 | 0.05 |
| COCAINE | 9041L | 2 | 1.86 | 0.93 |
| CODEINE | 9050 | 1,112 | 212,763.45 | 191.33 |

| | | | | |
|---|---|---|---|---|
| BUPRENORPHINE | 9064 | 1,024 | 33,149.73 | 32.37 |
| DIHYDROCODEINE | 9120 | 7 | 21.36 | 3.05 |
| OXYCODONE | 9143 | 1,114 | 978,494.49 | 878.36 |
| HYDROMORPHONE | 9150 | 991 | 12,587.27 | 12.7 |
| HYDROCODONE | 9193 | 1,119 | 342,630.01 | 306.19 |
| LEVORPHANOL | 9220L | 11 | 12.88 | 1.17 |
| MEPERIDINE (PETHIDINE) | 9230 | 165 | 2,918.54 | 17.69 |
| METHADONE | 9250B | 960 | 53,008.85 | 55.22 |
| MORPHINE | 9300 | 1,103 | 342,584.18 | 310.59 |
| OPIUM POWDERED | 9639 | 131 | 252.72 | 1.93 |
| OXYMORPHONE | 9652 | 654 | 13,296.65 | 20.33 |
| SUFENTANIL BASE | 9740 | 2 | 2.81 | 1.41 |
| TAPENTADOL | 9780 | 708 | 69,446 | 98.09 |
| FENTANYL BASE | 9801 | 1,083 | 6,786.49 | 6.27 |

STATE:WISCONSIN BUSINESS ACTIVITY:B - HOSPITALS

| DRUG NAME | DRUG CODE | BUYERS | TOTAL GRAMS | AVG GRAMS |
|---|---|---|---|---|
| AMPHETAMINE | 1100 | 117 | 11,716.93 | 100.14 |
| DL-METHAMPHETAMINE RACEMIC BASE | 1105B | 1 | 6.43 | 6.43 |
| D-METHAMPHETAMINE | 1105D | 1 | 4.42 | 4.42 |
| LISDEXAMFETAMINE | 1205 | 68 | 2,590.47 | 38.1 |
| METHYLPHENIDATE (DL;D;L;ISOMERS) | 1724 | 136 | 9,758.63 | 71.75 |
| AMOBARBITAL (SCHEDULE 2) | 2125 | 7 | 27.8 | 3.97 |
| BUTALBITAL | 2165 | 12 | 295 | 24.58 |
| PENTOBARBITAL (SCHEDULE 2) | 2270 | 14 | 7,052.86 | 503.78 |
| COCAINE | 9041L | 91 | 506.9 | 5.57 |
| CODEINE | 9050 | 178 | 8,399.80 | 47.19 |
| BUPRENORPHINE | 9064 | 100 | 2,353.05 | 23.53 |
| OXYCODONE | 9143 | 178 | 86,286.80 | 484.76 |
| HYDROMORPHONE | 9150 | 167 | 4,045.92 | 24.23 |
| HYDROCODONE | 9193 | 174 | 20,026.38 | 115.09 |
| MEPERIDINE (PETHIDINE) | 9230 | 129 | 3,986.07 | 30.9 |

| | | | | |
|---|---|---|---|---|
| METHADONE | 9250B | 131 | 4,261.05 | 32.53 |
| MORPHINE | 9300 | 179 | 42,027.15 | 234.79 |
| OPIUM TINCTURE (LAUDANUM) | 9630 | 1 | 1.18 | 1.18 |
| OPIUM POWDERED | 9639 | 109 | 498.96 | 4.58 |
| OXYMORPHONE | 9652 | 30 | 306.01 | 10.2 |
| ALFENTANIL | 9737 | 33 | 13.84 | 0.42 |
| REMIFENTANIL | 9739 | 60 | 23.13 | 0.39 |
| SUFENTANIL BASE | 9740 | 42 | 0.55 | 0.01 |
| TAPENTADOL | 9780 | 37 | 2,339.35 | 63.23 |
| FENTANYL BASE | 9801 | 169 | 768.82 | 4.55 |

STATE:WISCONSIN BUSINESS ACTIVITY:C - PRACTITIONERS

| DRUG NAME | DRUG CODE | BUYERS | TOTAL GRAMS | AVG GRAMS |
|---|---|---|---|---|
| AMPHETAMINE | 1100 | 2 | 7.51 | 3.75 |
| METHYLPHENIDATE (DL;D;L;ISOMERS) | 1724 | 1 | 0.43 | 0.43 |
| PENTOBARBITAL (SCHEDULE 2) | 2270 | 203 | 142,862.99 | 703.76 |
| COCAINE | 9041L | 26 | 120.42 | 4.63 |
| CODEINE | 9050 | 64 | 531.6 | 8.31 |
| BUPRENORPHINE | 9064 | 482 | 48.89 | 0.1 |
| OXYCODONE | 9143 | 56 | 2,210.76 | 39.48 |
| HYDROMORPHONE | 9150 | 391 | 217.86 | 0.56 |
| HYDROCODONE | 9193 | 218 | 717.01 | 3.29 |
| MEPERIDINE (PETHIDINE) | 9230 | 105 | 765.29 | 7.29 |
| METHADONE | 9250B | 13 | 42.66 | 3.28 |
| MORPHINE | 9300 | 235 | 580.68 | 2.47 |
| OPIUM POWDERED | 9639 | 8 | 33.12 | 4.14 |
| OXYMORPHONE | 9652 | 13 | 14.65 | 1.13 |
| NOROXYMORPHONE | 9668 | 2 | 0.67 | 0.34 |
| ALFENTANIL | 9737 | 15 | 4.65 | 0.31 |
| REMIFENTANIL | 9739 | 17 | 1.16 | 0.07 |
| SUFENTANIL BASE | 9740 | 2 | 0.02 | 0.01 |
| TAPENTADOL | 9780 | 1 | 6 | 6 |

| | | | | |
|---|---|---|---|---|
| FENTANYL BASE | 9801 | 332 | 50.5 | 0.15 |

STATE:WISCONSIN BUSINESS ACTIVITY:M - MID-LEVEL PRACTITIONERS

| DRUG NAME | DRUG CODE | BUYERS | TOTAL GRAMS | AVG GRAMS |
|---|---|---|---|---|
| PENTOBARBITAL (SCHEDULE 2) | 2270 | 19 | 13,348.63 | 702.56 |
| CODEINE | 9050 | 1 | 6.63 | 6.63 |
| BUPRENORPHINE | 9064 | 1 | 0.24 | 0.24 |
| OXYCODONE | 9143 | 1 | 0.45 | 0.45 |
| HYDROMORPHONE | 9150 | 1 | 0.04 | 0.04 |
| MORPHINE | 9300 | 1 | 0.08 | 0.08 |
| FENTANYL BASE | 9801 | 1 | 0 | 0 |

STATE:WISCONSIN BUSINESS ACTIVITY:N-U NARCOTIC TREATMENT PROGRAMS

| DRUG NAME | DRUG CODE | BUYERS | TOTAL GRAMS | AVG GRAMS |
|---|---|---|---|---|
| BUPRENORPHINE | 9064 | 8 | 1,997.83 | 249.73 |
| METHADONE | 9250B | 18 | 198,410.12 | 11,022.78 |

STATE:WYOMING BUSINESS ACTIVITY:A - PHARMACIES

| DRUG NAME | DRUG CODE | BUYERS | TOTAL GRAMS | AVG GRAMS |
|---|---|---|---|---|
| AMPHETAMINE | 1100 | 122 | 23,735.74 | 194.56 |
| DL-METHAMPHETAMINE RACEMIC BASE | 1105B | 1 | 0.4 | 0.4 |
| D-METHAMPHETAMINE | 1105D | 7 | 24.5 | 3.5 |
| LISDEXAMFETAMINE | 1205 | 121 | 10,318.51 | 85.28 |
| METHYLPHENIDATE (DL;D;L;ISOMERS) | 1724 | 122 | 31,482.82 | 258.06 |
| BUTALBITAL | 2165 | 78 | 3,505 | 44.94 |
| SECOBARBITAL (SCHEDULE 2) | 2315 | 1 | 18.31 | 18.31 |
| COCAINE | 9041L | 2 | 23.43 | 11.71 |
| CODEINE | 9050 | 120 | 16,710.80 | 139.26 |

| | | | | |
|---|---|---|---|---|
| BUPRENORPHINE | 9064 | 117 | 3,760.33 | 32.14 |
| OXYCODONE | 9143 | 123 | 108,110.22 | 878.94 |
| HYDROMORPHONE | 9150 | 118 | 2,588.30 | 21.93 |
| HYDROCODONE | 9193 | 123 | 47,349.24 | 384.95 |
| MEPERIDINE (PETHIDINE) | 9230 | 61 | 1,666.31 | 27.32 |
| METHADONE | 9250B | 109 | 5,071.61 | 46.53 |
| MORPHINE | 9300 | 122 | 31,835.84 | 260.95 |
| OPIUM POWDERED | 9639 | 11 | 15.12 | 1.37 |
| OXYMORPHONE | 9652 | 68 | 1,676.84 | 24.66 |
| REMIFENTANIL | 9739 | 1 | 0.08 | 0.08 |
| TAPENTADOL | 9780 | 71 | 7,435.50 | 104.73 |
| FENTANYL BASE | 9801 | 120 | 635.52 | 5.3 |

STATE:WYOMING BUSINESS ACTIVITY:B - HOSPITALS

| DRUG NAME | DRUG CODE | BUYERS | TOTAL GRAMS | AVG GRAMS |
|---|---|---|---|---|
| AMPHETAMINE | 1100 | 16 | 1,644.27 | 102.77 |
| LISDEXAMFETAMINE | 1205 | 9 | 356.11 | 39.57 |
| METHYLPHENIDATE (DL;D;L;ISOMERS) | 1724 | 22 | 2,440.24 | 110.92 |
| BUTALBITAL | 2165 | 1 | 25 | 25 |
| PENTOBARBITAL (SCHEDULE 2) | 2270 | 1 | 2.73 | 2.73 |
| COCAINE | 9041L | 23 | 55.6 | 2.42 |
| CODEINE | 9050 | 39 | 2,059.75 | 52.81 |
| BUPRENORPHINE | 9064 | 17 | 254 | 14.94 |
| OXYCODONE | 9143 | 42 | 8,736.36 | 208.01 |
| HYDROMORPHONE | 9150 | 40 | 312.09 | 7.8 |
| HYDROCODONE | 9193 | 43 | 6,371.74 | 148.18 |
| MEPERIDINE (PETHIDINE) | 9230 | 37 | 323.46 | 8.74 |
| METHADONE | 9250B | 19 | 708.81 | 37.31 |
| MORPHINE | 9300 | 44 | 9,273.22 | 210.75 |
| OPIUM POWDERED | 9639 | 14 | 116.64 | 8.33 |
| OXYMORPHONE | 9652 | 1 | 0.45 | 0.45 |
| ALFENTANIL | 9737 | 7 | 0.14 | 0.02 |

| | | | | |
|---|---|---|---|---|
| REMIFENTANIL | 9739 | 16 | 1.8 | 0.11 |
| SUFENTANIL BASE | 9740 | 9 | 0.04 | 0 |
| TAPENTADOL | 9780 | 3 | 42.5 | 14.17 |
| FENTANYL BASE | 9801 | 44 | 91.67 | 2.08 |

STATE:WYOMING BUSINESS ACTIVITY:C - PRACTITIONERS

| DRUG NAME | DRUG CODE | BUYERS | TOTAL GRAMS | AVG GRAMS |
|---|---|---|---|---|
| PENTOBARBITAL (SCHEDULE 2) | 2270 | 40 | 38,482.50 | 962.06 |
| CODEINE | 9050 | 8 | 133.04 | 16.63 |
| BUPRENORPHINE | 9064 | 63 | 2.04 | 0.03 |
| OXYCODONE | 9143 | 1 | 4.84 | 4.84 |
| HYDROMORPHONE | 9150 | 35 | 17.06 | 0.49 |
| HYDROCODONE | 9193 | 22 | 126.8 | 5.76 |
| MEPERIDINE (PETHIDINE) | 9230 | 4 | 11.11 | 2.78 |
| METHADONE | 9250B | 1 | 8.05 | 8.05 |
| MORPHINE | 9300 | 36 | 64.34 | 1.79 |
| FENTANYL BASE | 9801 | 34 | 1.49 | 0.04 |

STATE:WYOMING BUSINESS ACTIVITY:D - TEACHING INSTITUTIONS

| DRUG NAME | DRUG CODE | BUYERS | TOTAL GRAMS | AVG GRAMS |
|---|---|---|---|---|
| PENTOBARBITAL (SCHEDULE 2) | 2270 | 1 | 88.86 | 88.86 |
| BUPRENORPHINE | 9064 | 1 | 0 | 0 |

STATE:WYOMING BUSINESS ACTIVITY:M - MID-LEVEL PRACTITIONERS

| DRUG NAME | DRUG CODE | BUYERS | TOTAL GRAMS | AVG GRAMS |
|---|---|---|---|---|
| PENTOBARBITAL (SCHEDULE 2) | 2270 | 3 | 266.58 | 88.86 |
| CODEINE | 9050 | 2 | 5.97 | 2.98 |
| OXYCODONE | 9143 | 1 | 0.27 | 0.27 |

| | | | | |
|---|---|---|---|---|
| HYDROCODONE | 9193 | 1 | 0.18 | 0.18 |

UNITED STATES SUMMARY FOR RETAIL DRUG PURCHASES BY GRAMS WT

ENTIRE UNITED STATES BUSINESS ACTIVITY: A - PHARMACIES

| DRUG NAME | DRUG CODE | BUYERS | TOTAL GRAMS | AVG GRAMS |
|---|---|---|---|---|
| AMPHETAMINE | 1100 | 64,685 | 19,459,361.45 | 300.83 |
| DL-METHAMPHETAMINE RACEMIC BASE | 1105B | 408 | 1,579.27 | 3.87 |
| D-METHAMPHETAMINE | 1105D | 1,711 | 6,411.53 | 3.75 |
| LISDEXAMFETAMINE | 1205 | 60,254 | 9,480,895.90 | 157.35 |
| METHYLPHENIDATE (DL;D;L;ISOMERS) | 1724 | 64,316 | 17,972,772.61 | 279.44 |
| GAMMA HYDROXYBUTYRIC ACID(FDA APPROVED-CSA III) | 2012 | 2 | 25,315,001.28 | 12,657,500.64 |
| BARBITURIC ACID DERIVIATIVE OR SALT [PER 21CFR 1308.13(C)(3)] | 2100 | 681 | 8,145 | 11.96 |
| BUTALBITAL | 2165 | 37,579 | 1,592,805 | 42.39 |
| PENTOBARBITAL (SCHEDULE 2) | 2270 | 6 | 591.95 | 98.66 |
| SECOBARBITAL (SCHEDULE 2) | 2315 | 283 | 9,063.45 | 32.03 |
| NABILONE | 7379 | 79 | 18.3 | 0.23 |
| COCAINE | 9041L | 224 | 1,693.61 | 7.56 |
| CODEINE | 9050 | 66,753 | 16,602,557.32 | 248.72 |
| BUPRENORPHINE | 9064 | 59,192 | 2,844,643.52 | 48.06 |
| DIHYDROCODEINE | 9120 | 4,100 | 27,095.59 | 6.61 |
| OXYCODONE | 9143 | 66,283 | 52,050,579.29 | 785.28 |
| HYDROMORPHONE | 9150 | 58,087 | 1,342,091.34 | 23.1 |
| HYDROCODONE | 9193 | 66,910 | 28,615,716.36 | 427.67 |
| LEVORPHANOL | 9220L | 1,142 | 2,058.91 | 1.8 |
| MEPERIDINE (PETHIDINE) | 9230 | 23,185 | 582,601.56 | 25.13 |
| METHADONE | 9250B | 51,761 | 3,372,211.67 | 65.15 |
| MORPHINE | 9300 | 62,481 | 17,287,250.62 | 276.68 |
| OPIUM TINCTURE (LAUDANUM) | 9630 | 132 | 687.86 | 5.21 |
| OPIUM POWDERED | 9639 | 4,211 | 12,119.40 | 2.88 |
| OXYMORPHONE | 9652 | 39,317 | 1,548,716 | 39.39 |
| NOROXYMORPHONE | 9668 | 2 | 4.77 | 2.39 |
| ALFENTANIL | 9737 | 38 | 13.61 | 0.36 |
| REMIFENTANIL | 9739 | 72 | 9.3 | 0.13 |
| SUFENTANIL BASE | 9740 | 71 | 106.65 | 1.5 |
| TAPENTADOL | 9780 | 43,658 | 6,129,817.50 | 140.41 |
| FENTANYL BASE | 9801 | 61,998 | 350,397.31 | 5.65 |

UNITED STATES SUMMARY FOR RETAIL DRUG PURCHASES BY GRAMS WT

ENTIRE UNITED STATES BUSINESS ACTIVITY: B - HOSPITALS

| DRUG NAME | DRUG CODE | BUYERS | TOTAL GRAMS | AVG GRAMS |
|---|---|---|---|---|
| AMPHETAMINE | 1100 | 3,945 | 510,917.78 | 129.51 |
| DL-METHAMPHETAMINE RACEMIC BASE | 1105B | 14 | 140.25 | 10.02 |
| D-METHAMPHETAMINE | 1105D | 20 | 56.64 | 2.83 |
| LISDEXAMFETAMINE | 1205 | 1,847 | 119,199.14 | 64.54 |
| METHYLPHENIDATE (DL;D;L;ISOMERS) | 1724 | 5,262 | 625,072.32 | 118.79 |
| GAMMA HYDROXYBUTYRIC ACID(FDA APPROVED-CSA III) | 2012 | 13 | 34,699.97 | 2,669.23 |
| BARBITURIC ACID DERIVIATIVE OR SALT [PER 21CFR 1308.13(C)(3)] | 2100 | 7 | 50 | 7.14 |
| AMOBARBITAL (SCHEDULE 2) | 2125 | 212 | 1,032.16 | 4.87 |
| BUTALBITAL | 2165 | 616 | 15,474.80 | 25.12 |
| PENTOBARBITAL (SCHEDULE 2) | 2270 | 628 | 225,742.27 | 359.46 |
| SECOBARBITAL (SCHEDULE 2) | 2315 | 10 | 439.44 | 43.94 |
| NABILONE | 7379 | 5 | 0.9 | 0.18 |
| PHENCYCLIDINE (PCP) | 7471 | 1 | 0.22 | 0.22 |
| COCAINE | 9041L | 3,792 | 24,555.61 | 6.48 |
| CODEINE | 9050 | 7,659 | 702,050.18 | 91.66 |
| BUPRENORPHINE | 9064 | 4,420 | 84,632.22 | 19.15 |
| DIHYDROCODEINE | 9120 | 17 | 60.89 | 3.58 |
| OXYCODONE | 9143 | 8,714 | 2,647,424.10 | 303.81 |
| HYDROMORPHONE | 9150 | 8,710 | 241,154.57 | 27.69 |
| HYDROCODONE | 9193 | 9,200 | 1,681,992.77 | 182.83 |
| LEVORPHANOL | 9220L | 21 | 22.75 | 1.08 |
| MEPERIDINE (PETHIDINE) | 9230 | 7,152 | 243,768.93 | 34.08 |
| METHADONE | 9250B | 5,750 | 311,371.43 | 54.15 |
| MORPHINE | 9300 | 9,742 | 1,872,037.94 | 192.16 |
| OPIUM TINCTURE (LAUDANUM) | 9630 | 2 | 2.36 | 1.18 |
| OPIUM POWDERED | 9639 | 3,548 | 20,742.12 | 5.85 |
| OXYMORPHONE | 9652 | 1,034 | 14,405.83 | 13.93 |
| NOROXYMORPHONE | 9668 | 7 | 20.48 | 2.93 |
| ALFENTANIL | 9737 | 744 | 289.93 | 0.39 |
| REMIFENTANIL | 9739 | 2,266 | 1,530.25 | 0.68 |
| SUFENTANIL BASE | 9740 | 1,743 | 49.87 | 0.03 |
| TAPENTADOL | 9780 | 1,463 | 95,284.49 | 65.13 |
| FENTANYL BASE | 9801 | 10,022 | 41,436.45 | 4.13 |

UNITED STATES SUMMARY FOR RETAIL DRUG PURCHASES BY GRAMS WT

ENTIRE UNITED STATES BUSINESS ACTIVITY: C - PRACTITIONERS

| DRUG NAME | DRUG CODE | BUYERS | TOTAL GRAMS | AVG GRAMS |
|---|---|---|---|---|
| AMPHETAMINE | 1100 | 83 | 2,223.82 | 26.79 |
| D-METHAMPHETAMINE | 1105D | 3 | 4.18 | 1.39 |
| LISDEXAMFETAMINE | 1205 | 18 | 1,439.88 | 79.99 |
| METHYLPHENIDATE (DL;D;L;ISOMERS) | 1724 | 32 | 1,658.68 | 51.83 |
| GAMMA HYDROXYBUTYRIC ACID(FDA APPROVED-CSA III) | 2012 | 13 | 43,822.85 | 3,370.99 |
| BARBITURIC ACID DERIVIATIVE OR SALT [PER 21CFR 1308.13(C)(3)] | 2100 | 1 | 2 | 2 |
| BUTALBITAL | 2165 | 15 | 410 | 27.33 |
| PENTOBARBITAL (SCHEDULE 2) | 2270 | 6,313 | 5,384,783.13 | 852.97 |
| COCAINE | 9041L | 676 | 3,009.74 | 4.45 |
| CODEINE | 9050 | 4,196 | 117,093.57 | 27.91 |
| BUPRENORPHINE | 9064 | 24,057 | 6,898.75 | 0.29 |
| DIHYDROCODEINE | 9120 | 1,900 | 5,543.05 | 2.92 |
| OXYCODONE | 9143 | 1,968 | 77,390.74 | 39.32 |
| HYDROMORPHONE | 9150 | 12,620 | 7,952.29 | 0.63 |
| HYDROCODONE | 9193 | 12,205 | 69,672.09 | 5.71 |
| MEPERIDINE (PETHIDINE) | 9230 | 5,009 | 44,185.28 | 8.82 |
| METHADONE | 9250B | 500 | 2,244.61 | 4.49 |
| MORPHINE | 9300 | 8,938 | 23,782.95 | 2.66 |
| OPIUM POWDERED | 9639 | 113 | 371.52 | 3.29 |
| OXYMORPHONE | 9652 | 417 | 1,149.40 | 2.76 |
| NOROXYMORPHONE | 9668 | 145 | 968.57 | 6.68 |
| ALFENTANIL | 9737 | 201 | 59.09 | 0.29 |
| REMIFENTANIL | 9739 | 511 | 43.24 | 0.08 |
| SUFENTANIL BASE | 9740 | 87 | 7.06 | 0.08 |
| TAPENTADOL | 9780 | 23 | 1,474.60 | 64.11 |
| FENTANYL BASE | 9801 | 17,979 | 2,573.98 | 0.14 |

UNITED STATES SUMMARY FOR RETAIL DRUG PURCHASES BY GRAMS WT

ENTIRE UNITED STATES BUSINESS ACTIVITY: D - TEACHING INSTITUTIONS

| DRUG NAME | DRUG CODE | BUYERS | TOTAL GRAMS | AVG GRAMS |
|---|---|---|---|---|
| AMPHETAMINE | 1100 | 1 | 0.73 | 0.73 |
| D-METHAMPHETAMINE | 1105D | 3 | 4.18 | 1.39 |
| METHYLPHENIDATE (DL;D;L;ISOMERS) | 1724 | 5 | 18.33 | 3.67 |
| PENTOBARBITAL (SCHEDULE 2) | 2270 | 17 | 42,051.08 | 2,473.59 |
| COCAINE | 9041L | 2 | 1.18 | 0.59 |
| CODEINE | 9050 | 6 | 1,343.24 | 223.87 |
| BUPRENORPHINE | 9064 | 64 | 6.68 | 0.1 |
| OXYCODONE | 9143 | 1 | 1.34 | 1.34 |
| HYDROMORPHONE | 9150 | 28 | 47.39 | 1.69 |
| HYDROCODONE | 9193 | 7 | 92.98 | 13.28 |
| MEPERIDINE (PETHIDINE) | 9230 | 3 | 15.25 | 5.08 |
| METHADONE | 9250B | 6 | 150.17 | 25.03 |
| MORPHINE | 9300 | 23 | 149.12 | 6.48 |
| OXYMORPHONE | 9652 | 3 | 1.86 | 0.62 |
| REMIFENTANIL | 9739 | 6 | 0.23 | 0.04 |
| SUFENTANIL BASE | 9740 | 1 | 0 | 0 |
| FENTANYL BASE | 9801 | 27 | 16.36 | 0.61 |

UNITED STATES SUMMARY FOR RETAIL DRUG PURCHASES BY GRAMS WT

ENTIRE UNITED STATES BUSINESS ACTIVITY: M - MID-LEVEL PRACTITIONERS

| DRUG NAME | DRUG CODE | BUYERS | TOTAL GRAMS | AVG GRAMS |
|---|---|---|---|---|
| METHYLPHENIDATE (DL;D;L;ISOMERS) | 1724 | 1 | 3.46 | 3.46 |
| BARBITURIC ACID DERIVIATIVE OR SALT [PER 21CFR 1308.13(C)(3)] | 2100 | 1 | 8 | 8 |
| BUTALBITAL | 2165 | 2 | 440 | 220 |
| PENTOBARBITAL (SCHEDULE 2) | 2270 | 689 | 990,497.05 | 1,437.59 |
| CODEINE | 9050 | 451 | 5,456.56 | 12.1 |
| BUPRENORPHINE | 9064 | 17 | 7.19 | 0.42 |
| DIHYDROCODEINE | 9120 | 550 | 331 | 0.6 |
| OXYCODONE | 9143 | 12 | 2,496.15 | 208.01 |
| HYDROMORPHONE | 9150 | 128 | 66.01 | 0.52 |
| HYDROCODONE | 9193 | 27 | 1,110.66 | 41.14 |
| MEPERIDINE (PETHIDINE) | 9230 | 25 | 65.8 | 2.63 |
| METHADONE | 9250B | 4 | 477.17 | 119.29 |
| MORPHINE | 9300 | 914 | 1,518.57 | 1.66 |
| OXYMORPHONE | 9652 | 3 | 15.66 | 5.22 |
| REMIFENTANIL | 9739 | 6 | 0.18 | 0.03 |
| TAPENTADOL | 9780 | 1 | 110 | 110 |
| FENTANYL BASE | 9801 | 1,236 | 63.84 | 0.05 |

ENTIRE UNITED STATES BUSINESS ACTIVITY: N-U NARCOTIC TREATMENT PROGRAMS

| DRUG NAME | DRUG CODE | BUYERS | TOTAL GRAMS | AVG GRAMS |
|---|---|---|---|---|
| BUPRENORPHINE | 9064 | 536 | 57,919.26 | 108.06 |
| METHADONE | 9250B | 1,411 | 10,839,567.21 | 7,682.19 |
